# Identification of candidate biomarkers and molecular networks associated with Pulmonary Arterial Hypertension using machine learning and plasma multi-Omics analysis

**DOI:** 10.1101/2024.12.17.24319117

**Authors:** V.O. Kheyfets, A.J. Sweatt, H. Zhang, T. Nemkov, M. Aizin, P. Heerdt, M. Dzieciatkowska, D. Stephenson, I.S. LaCroix, A. D’Alessandro, W. M. Oldham, K.C. Hansen, R.T. Zamanian, K.R. Stenmark

**Author notes:** Corresponding Author:Vitaly O. Kheyfets, University of Colorado Anschutz Medical Campus, School of Medicine Research Complex 2 - 12705 E. Montview BLVD, Office 6122, Aurora, CO 80045, /.

## Abstract

**Background:** Pulmonary arterial hypertension (PAH) is a rare but severe and life-threatening condition that primarily affects the pulmonary blood vessels and the right ventricle of the heart. The limited availability of human tissue for research—most of which represents only end-stage disease—has led to a reliance on preclinical animal models. However, these models often fail to capture the heterogeneity and complexity of the human condition. Analyzing the molecular signatures in patient plasma provides a unique opportunity to gain insights into PAH pathobiology, explore disease heterogeneity absent in animal models, and identify potential therapeutic targets.

**Objective:** This study aims to characterize the circulating peptides, metabolites, and lipids most relevant to PAH by leveraging unbiased mass spectrometry and advanced computational tools. Building on prior research that identified individual circulating factors, this work seeks to integrate these molecular layers to better understand their interactions and collective contribution to PAH pathobiology.

**Methods:** Peripheral blood samples were collected from 402 patients with PAH and 76 healthy individuals. Various types of molecules in the blood – peptides, metabolites, and lipids- were measured. Statistical and machine learning methods were used to identify differences between PAH patients and healthy individuals, and further to understand how these molecules might interact with each other. A survival model was also trained to examine the association between the blood molecular signature and patient outcomes.

**Results:** Differential abundance analysis revealed 832 peptides (from 291 proteins), 45 metabolites, and 222 lipids significantly altered in PAH compared to controls. Machine learning-based feature selection identified 11 key molecules, including 2-Hydroxyglutarate, that together achieved a classification accuracy of 98.6% for PAH in a multivariate model tested on a withheld cohort. Latent network discovery uncovered 7 distinct networks, highlighting interacting molecules from pathways—such as hypoxia, glycolysis, fatty acid metabolism, and complement activation—that we and others have previously linked to vascular lesions in PAH patients.

A survival model incorporating 155 molecular features predicted outcomes in PAH patients with a c-index of 0.762, independent of traditional clinical parameters. This model stratified patients into risk categories consistent with established markers of cardiac function, exercise tolerance, and the REVEAL 2.0 risk score.

**Conclusion:** This study underscores the utility of integrated omics in unraveling PAH pathobiology in human subjects. Our findings highlight the central role of hypoxia signaling pathways interacting with disrupted fatty acid metabolism, complement activation, inflammation, and mitochondrial dysfunction. These interactions, revealed through latent network analysis, emphasize the metabolic and immune dysregulation underlying PAH. Furthermore, many of the molecules identified in the circulation were consistent with pathways enriched in pulmonary vascular lesions, reinforcing their biological relevance. Circulating plasma molecules from these networks demonstrated strong prognostic capabilities, comparable to current clinical risk scores, offering insights into disease progression and potential for future clinical application.

## 1. Introduction

Pulmonary arterial hypertension (PAH) is a rare and progressive cardiopulmonary disease that currently has no cure [1]. Limited access to lung tissue for study has made it challenging to fully understand the biological processes initiating or driving PAH, leading researchers to rely heavily on data from pre-clinical animal models. However, while the use of these models has offered enormous insight into disease pathobiology, they overlook its heterogeneity and underlying causes, which are crucial for treatment discovery and predicting outcomes [2–5].

Also, while animal models have been shown to offer a reliable surrogate for some aspects of the human disease [6], there are well-known limitations of extrapolating findings from rodents to the human condition [7]. This underscores the importance of studying circulating factors in human blood to complement preclinical findings and deepen our understanding of PAH pathobiology.

When measuring blood biomarkers, the challenge lies not only in the vast number of potential targets but also in the uncertainty surrounding which biological pathways to explore. Previous studies have characterized the transcriptomics and circulating inflammatory markers of PAH by analyzing patient blood, uncovering heterogeneity that would not be possible to observe in preclinical models [3, 4]. These findings suggest the potential importance of circulating biomarkers in understanding PAH pathobiology. However, most studies have focused on a single omics layer—such as targeted proteins, metabolites, or lipids—limiting their ability to capture the broader, integrated molecular networks underlying the disease [8–15]. While these previous single-omics approaches are invaluable for understanding specific biological aspects, they may miss interactions between different types of molecules, and thus might not capture the full complexity the system driving disease progression.

Multi-omics approaches, which simultaneously analyze proteins, metabolites, and lipids within the same subjects, provide a comprehensive view of PAH pathobiology. Until now, no study has applied an untargeted approach to examine all three molecular domains concurrently in a single cohort of PAH patients. By integrating these analyses with advanced machine learning algorithms, we can identify key molecules from an extensive pool of biochemical compounds across all omics domains. Furthermore, this approach enables us to explore the interactions among these molecules, uncovering hidden drivers of PAH and offering new insights into disease mechanisms [2].

In this study, we hypothesized that integrating data from proteins, metabolites, and lipids would help identify the molecular signature of PAH in human subjects and shed light on how these molecules interact with each other in the progression of this disease.

## 2. Methods

### 2.1 Study Population

The current study considered patient data from a cohort of 402 PAH patients and 76 control (CTL) subjects evaluated at Stanford University (Stanford, CA) between 2008 and 2014. PAH was confirmed hemodynamically as a mean pulmonary arterial pressure ≥ 25 mmHg, pulmonary vascular resistance ≥ 240 dyn·s·cm^-5^, and pulmonary arterial wedge pressure ≤ 15 mmHg. Additional inclusion/exclusion criteria can be found in ref. [3]. Samples for CTL subjects were acquired for the Stanford Cardiovascular Institute Biomarker and Phenotype Core Laboratory biorepository (IRB #40869). These subjects were healthy volunteers, collected

between 2009 and 2013, who passed a screening to confirm no cardiopulmonary symptoms, atherosclerosis, cardiovascular disease, lung disease, diabetes, systemic inflammatory disease, recent infections, or Alzheimer’s disease. Smokers or subjects with body mass index above 35 were also excluded.

### 2.2 Mass Spectrometry (MS)-based proteomics, metabolomics, and lipidomics analyses

The plasma fraction of each sample was flash frozen at the time of collection at Stanford University, maintained at -40°C, and eventually shipped (without thawing) to the University of Colorado (Anschutz Medical Campus, Aurora, CO). All MS analysis was performed at the University of Colorado. Details about sample preparation, MS configuration, and identification libraries are described in Supplemental Material 1.

Notably, the MS-based proteomics approach used in this study, which did not employ enrichment techniques, had inherent limitations in detecting low-abundance molecules.

Consequently, important signaling molecules such as cytokines and members of the TGF-beta pathway were not captured in our analysis.

### 2.3 Data analysis

Data analysis was performed using MATLAB 2021b (Mathworks, MA), Python 3.9.16, and R 4.2.2. Untargeted proteomics, metabolomics, and lipidomics identified a total of 15,781 potential circulating biomarkers from blood plasma samples. Subsequently, a global data adjustment was employed to remove systematic biases (e.g., differences in the amount of material loaded for each sample) [16]. The normalizer library [17] (NormalizerDE 1.13.2) was used to perform an exhaustive comparative evaluation of various normalization schemes. Based on the lowest intragroup post-event variability, we chose Variance Stabilizing Normalization (VSN) as the optimal normalization method for this data. Any peptide, metabolite, or lipid that was missing in more than 5% of the patients was removed. Such a conservative approach was chosen to avoid imputing more than 1% of the data, which we felt would lead to misleading conclusions, while recognizing that it might overlook important features. The remaining missing values (0.25% of the data) were imputed using kNN (k = √478). This resulted in a 478x3961 final dataset (3300 peptides, 121 metabolites, and 540 lipids).

The overall data analysis pipeline is centered around 5 objectives (see Fig. 1): (1) Identify differentially expressed molecules of PAH, which was intended as a housekeeping step to compare the molecules identified by our MS and data analysis pipeline against existing PAH literature. (2) Identify circulating molecules that are most predictive of PAH. While differential expression analysis identifies molecules with altered abundance in disease, those that serve as effective classifiers provide deeper insights by highlighting molecules that are not only elevated but also specific to the disease state. This distinction underscores their potential as biomarkers uniquely linked to the pathobiology of the condition. (3) Within the molecules identified in Step 2, analyze their association to other clinical markers (e.g., patient demographics, hemodynamics, pulmonary function). (4) Identify interactions between the three omics domains to gain deeper biological insight into PAH pathogenesis in human disease. (5) Evaluate the association between blood molecular signature and patient prognosis in PAH.

*(1) Differential expression analysis.* To identify differentially expressed blood molecules of PAH, we performed repeated t-tests to compare expression levels between PAH and CTL subjects. If variances were equal, a standard t-test was used; otherwise, Welch’s t-test (for unequal variances) was applied. To control for false discoveries, we applied the Benjamini-Hochberg correction. This analysis aimed to compare the biomarkers identified through our untargeted mass spectrometry (MS) approach with those reported in the broader PAH literature.
*(2) Recursive feature elimination (RFE) wrapped in a random forest classification model (RFE-RF).* The purpose of this step was to identify the most differentiative molecules of PAH from an untargeted pool of molecular compounds. To this end, it was necessary to first remove highly correlated features because correlated classifiers individually receive smaller importance due to their shared responsibility in the classification. Our approach for correcting correlation bias consisted of finding highly correlated network groups and then summarizing them using the feature with the highest variance. To this end, a logical adjacency matrix was created using pairwise linear correlation threshold of r > 0.85. This threshold was chosen so that the 1^st^ principal component of a connected network explained at least 70% of the variance for all networks. This reduced our dataset to 3282 uncorrelated features.

**Figure 1.**
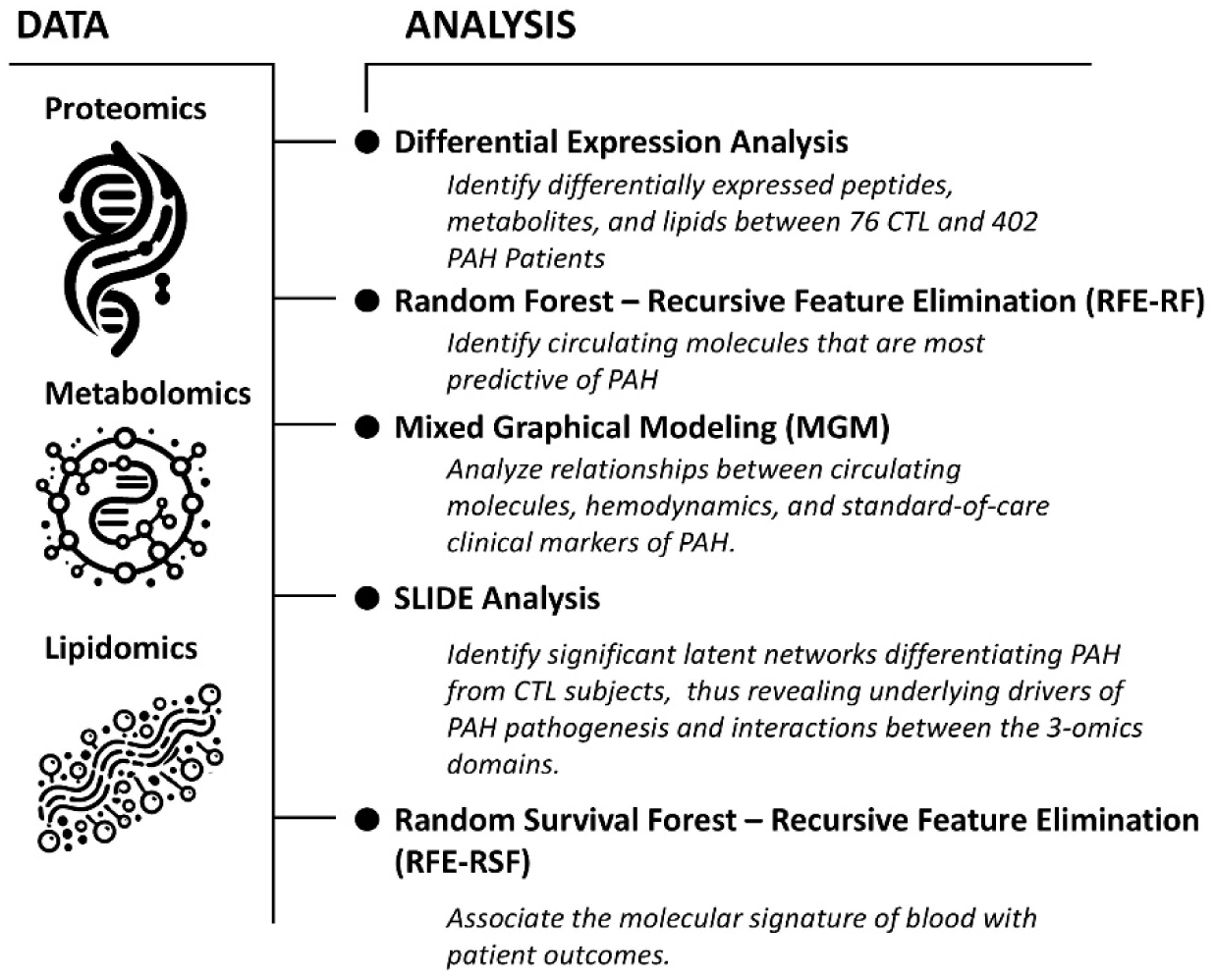
Overview of collected data and analytical approach.

Before implementing the RFE-RF algorithm, we randomly divided the patients into 70% for model training and 30% for testing. Using the training cohort, we applied RFE based on a technique outlined in refs.[11, 18] (1% of the least important features removed at each iteration). The RFE algorithm was run 100 times on the training cohort, where the subset of the most differentiating features was identified at the minimum of the out-of-bag (OOB) classification error. For every RFE simulation, hyperparameter tuning was performed using Bayesian optimization before RFE and for the final trained RF model (preset 500 trees; optimized minimum leaf size; optimized maximum number of splits).

a. *(3) Mixed Graphical Models.* To evaluate potential interactions between predictive circulating molecules, patient demographics, hemodynamics, lung function, clinical bloodwork, and disease course (e.g., time from diagnosis), we trained a 2^nd^ order (pairwise comparisons) mixed graphical model using the mgm-package in R (L1-penalized regression, which is weighted by the extended Bayesian information criterion, γ = 0.25). These undirected graphic models allow us to determine interactions between variables, including between continuous (e.g., pulmonary vascular resistance, PVR) and categorical (e.g., Ethnicity) variables. The available PAH clinical data, not including the multi-omics data, had 4% missing values, which were imputed using kNN (k =√402).

The interaction parameters of the pairwise models were visualized using the qgraph (R) package.

a. *(4) SLIDE: significant latent factor interaction discovery and exploration* (SLIDE). This data reduction approach was used to identify stand-alone and interacting latent networks that can differentiate PAH from CTL subjects while offering insight into the interactions between circulating proteomics, metabolomics, and lipidomics. SLIDE incorporates both linear and nonlinear relationships and comes with statistical guarantees regarding latent factor identifiability.

Latent variable models are based on the primary assumption that the available high-dimensional multi-omics dataset arises from a lower dimensional description. Each lower dimensional description is a linear combination of some subset of measured compounds (e.g., a combination of measured proteins, metabolites, and lipids), and are termed in this manuscript as *latent networks*. By inspecting these networks, we get an idea of which multi-omics features are co-regulated when differentiating healthy CTL from PAH patients.

After performing hyperparameter tuning, the final analysis (performed in R 4.2.2) was based on an optimal delta = 0.1; lambda = 1 (500 iterations, 500 sample cross-validation iterations; with 4-fold cross-validation).

a. *(5) Recursive feature elimination (RFE) wrapped in a random survival forest prognostic model (RFE-RSF).* Our objective was to identify a blood molecular signature associated with patient outcomes, specifically one that could differentiate between low-risk and high-risk patients based on their survival profiles. Beginning with molecules included within underlying drivers (a.k.a latent networks) discovered by the SLIDE algorithm, we performed a recursive feature elimination wrapped in an random survival forest (RSF) machine learning regression algorithm, which accounts for censored patients (e.g., patients lost to follow-up) [19].

Before performing feature selection by RFE-RSF, we divided the patients into 70% for model training and 30% for testing based on the training/testing cohorts from RFE-RF but excluding CTL subjects as they do not contribute to survival models. The RFE-RSF algorithm was run 64 times, where each simulation revealed a general trend of increasing concordance index (c-index) at each iteration until roughly 155 features remained, at which point the c-index began to decline. At each iteration of the RFE-RSF algorithm (RSF, 1000 trees, minimum of 10 samples in each split, minimum of 15 samples in each leaf) [19], the least important 3% of features were eliminated until fewer than 100 features remained, after which one feature was eliminated per iteration. For each RFE-RSF simulation, the training cohort was further divided with 30% of the training data randomly reserved for validating that simulation. The testing cohort was withheld from the RFE-RSF algorithm to evaluate a final RSF model on a reduced feature subset.

## 3. Results

Table 1 shows a breakdown of the demographic data for the CTL and PAH patients included in the study. Within the PAH patients, Kaplan-Meyer curves (not shown) revealed statistical differences (p < 0.05) in survival based on etiology, but no differences based on sex or ethnicity. There were also no differences in survival based on number of concurrent PAH-specific treatments (e.g., treatment naive vs. three PAH-specific treatments), and no relationship between the time since diagnosis and the number of PAH related therapies.

**Table 1.** Study cohort demographics in CTL and PAH patients. Continuous data is compared using a *t-test* and categorical data by x^2^ test.

|  | PAH | CTL | P-value |
| --- | --- | --- | --- |
| **N** | 402 | 76 |  |
| **Age (years ± SD)** | 52.3 ± 15.7 | 55.5 ± 16.2 | 0.11 |
| **% Female Patients** | 75% | 61% |  |
| **Race** |  |  |  |
| White | 211 | 57 | 0.02 |
| Black | 30 | 4 |  |
| Asian or Pacific Islander | 64 | 9 |  |
| Hispanic | 61 | 4 |  |
| Native American | 2 | 0 |  |
| Other | 31 | 2 |  |
| Unknown | 3 | 0 |  |
| **Etiology** |  |  |  |
| Congenital Heart Disease, CHD | 51 |  |  |
| Connective Tissue Disease, CTD | 122 |  |  |
| Chronic Thromboembolic PH, CTPH | 5 |  |  |
| Drug and Toxin, D&T | 84 |  |  |
| Hereditary PAH, HPAH | 3 |  |  |
| Idiopathic PAH, IPAH | 112 |  |  |
| Portopulmonary Hypertension, PoPH | 25 |  |  |
| **Hemodynamics** |  |  |  |
| mPAP (mmHg) | 48.3 ± 15.2 | | |
| PVR (Wood units) | 10.7 ± 5.92 | | |
| Cardiac Index ($\text{m}^2 \cdot \text{min}$) | 2.11 ± 0.61 | | |
| Mean Right Atrial Pressure (mmHg) | 9.14 ± 5.16 | | |
| PCWP (mmHg) | 11.7 ± 4.57 | | |
| **Timing from Diagnosis (years ± SD)** | 3.29 ± 5.94 | | |
| **Therapy** |  |  |  |
| Treatment Naive | 180 (45%) |  |  |
| Monotherapy | 100 (25%) |  |  |
| Dual Therapy | 83 (21%) |  |  |
| Triple Therapy | 39 (10%) |  |  |

### 3.1 Differential abundance patterns across metabolomic, lipidomic, and proteomic domains between PAH patients and CTL subjects

Overall, we found 832 peptides (from 291 proteins), 45 metabolites, and 222 lipids are up/down- regulated in PAH patients, relative to CTL subjects. In Fig. 2, volcano plots depict differences between CTL and PAH patients for circulating metabolites (Fig. 2a) and lipids (Fig. 2b). Fig. 2c and d illustrate volcano plots for circulating protein abundance (Fig. 2c) and the corresponding peptides (Fig. 2d).

**Figure 2.**
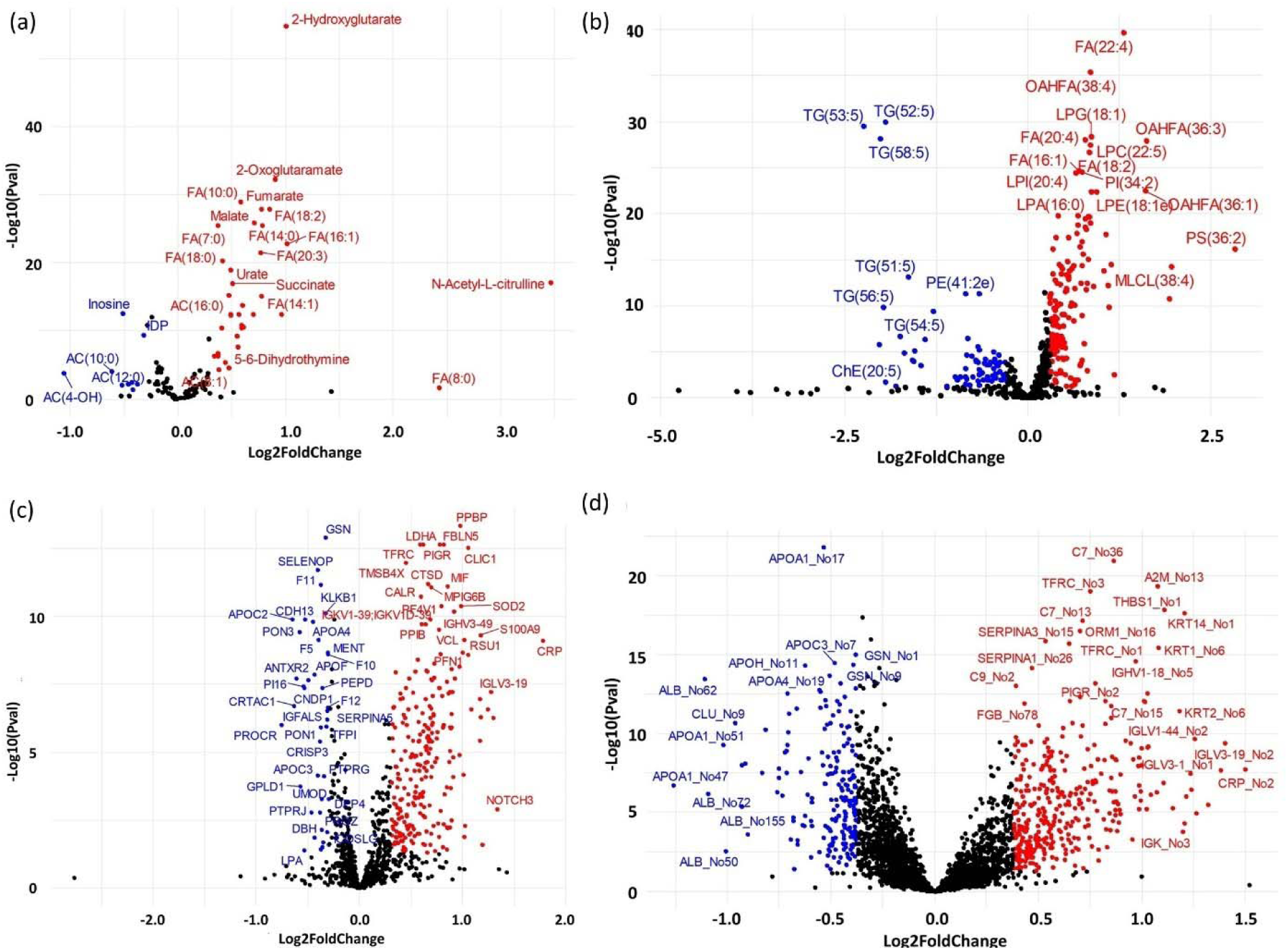
Volcano plots showing differentially expressed metabolites (a), lipids (b), proteins (c), and peptides (d) in PAH vs. CTL subjects. Each numbered peptide (e.g., APOA1_No17) can be cross-referenced using Supplement Material 2. Pval = adjusted p-value after Benjamini- Hochberg correction for false discovery.

In this study, we chose to investigate peptides instead of proteins for multiple reasons.

Peptide abundance represents a more unprocessed and raw form of the data while also revealing a molecular signature that accounts for post-translational modifications. Supplemental Material 2 lists all peptide names, which can be cross-referenced to the peptide sequence and description.

However, we also explored protein abundance to validate that the pathological insights derived from this dataset are consistent with prior studies. Our findings in the proteomics domain reveal well-established PAH molecules including activin (INHBE; p-value = 2.24e-7; 25% increase in PAH, but data point not labeled in Fig. 2c), complement (C7, CRP), platelet activation (PPBP) and protease inhibition (SERPINs). In the metabolomic and lipidomic domains, our MS platform also detected compounds consistent with previous studies, which included markers of hypoxia activation (e.g., 2-Hydroxyglutarate [20]), urate [4, 21], malate [22], and dysregulation of fatty acid metabolism [23].

### 3.2 Molecules Differentiating PAH from CTL Subjects

While differential expression analysis reveals molecules with altered abundance, molecules that serve as classifiers of PAH go a step further by pinpointing those specifically associated with the disease state. These molecules are not only elevated but also distinguishable as key indicators of the condition, offering valuable insights into its unique pathobiology. To this end, one hundred simulations of the RFE-RF algorithm were executed using the training subset of the patient cohort. This analysis identified 11 molecules that were influential in at least 80% of RFE simulations (see Table 2). When combined into a final RF algorithm, these molecules formed a predictive model achieving a classification accuracy [= (TP + TN) / (TP + TN + FP + FN), where TP = true positive, TN = true negative, FP = false positive, and FN = false negative] of 98.6% in the testing cohort, which was entirely withheld during both the RFE process and the training of the final RF model.

**Table 2.**
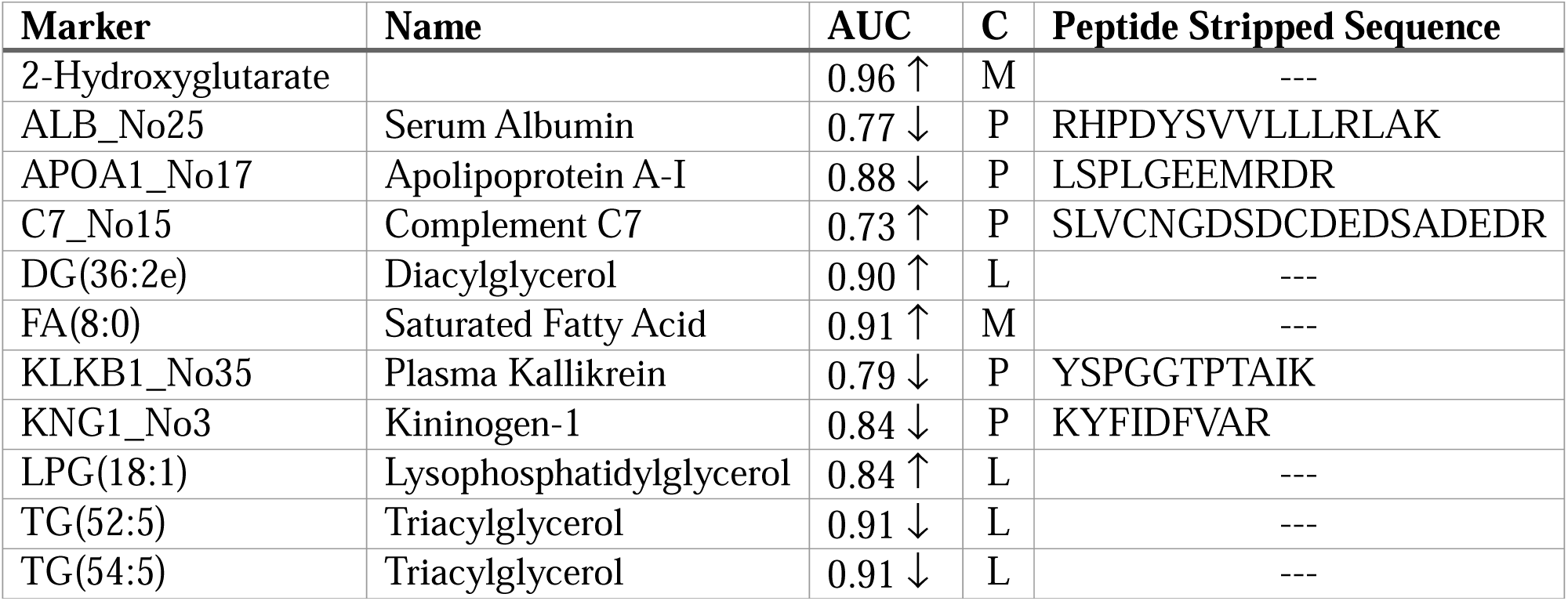
List of markers identified by RFE-RF as being able to differentiate between PAH vs. CTL subjects. AUC = area under receiver operating curve for entire study cohort, with an arrow showing an increase/decrease (↑/↓) in PAH. *C* column indicates category, which lists each marker as a peptide (P), metabolite (M), or lipid (L).

The molecule that consistently appeared as the best classifier of PAH in 100% of the RFE simulations was 2-Hydroxyglutarate. Within the training cohort used for the RFE algorithm, 2- Hydroxyglutarate demonstrated excellent classification performance as a univariate classifier, achieving a sensitivity of 90.8% and a specificity of 96.2%. However, in a blinded testing cohort that was entirely excluded from the RFE algorithm, 2-Hydroxyglutarate stratified PAH patients with 89.1% sensitivity and 87.5% specificity (see Fig. 3a and b).

**Figure 3.**
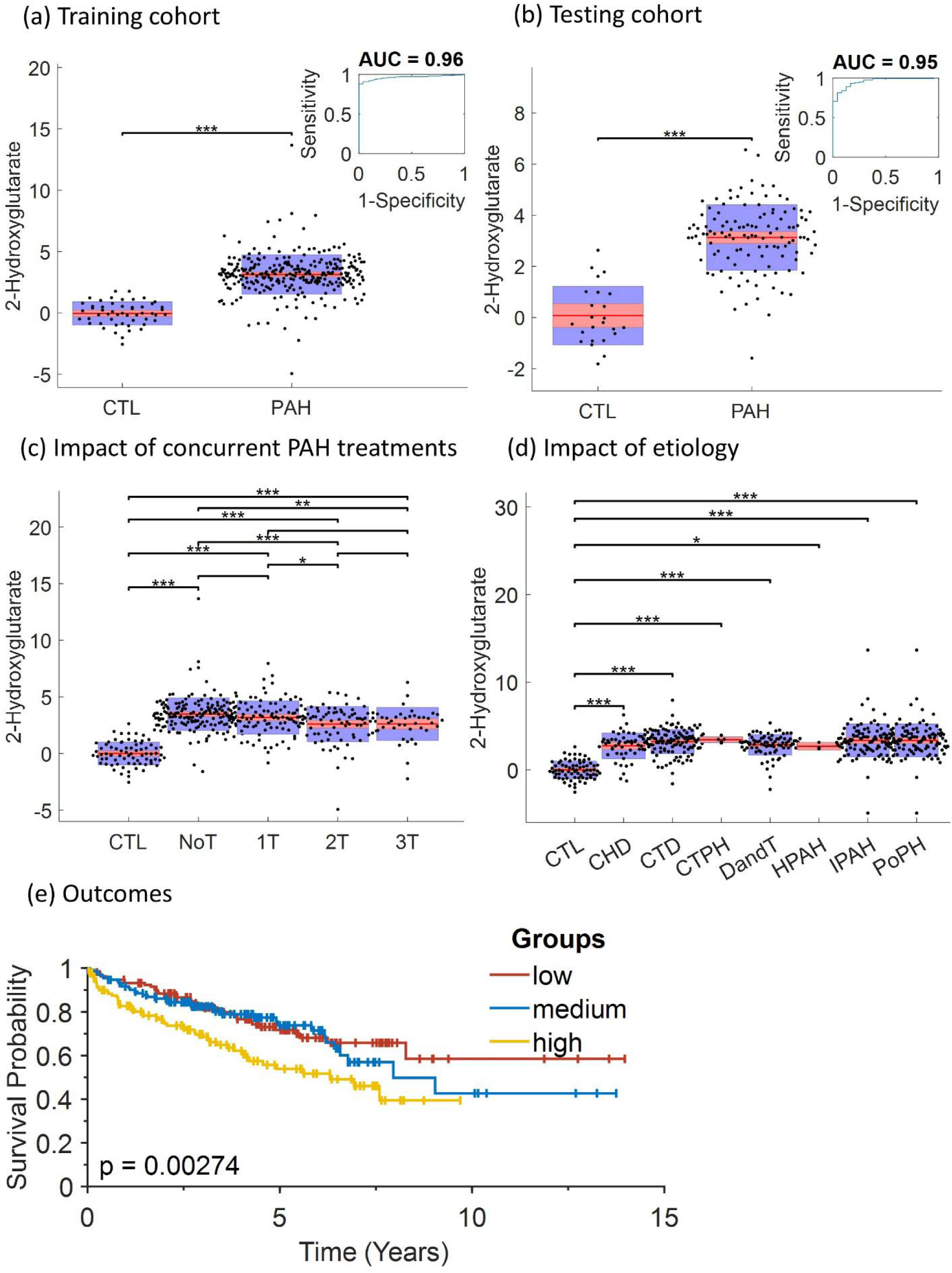
(a and b) 2-Hydroxyglutarate compared between PAH vs. CTL subjects in the training and testing subsets, respectively. (c) 2-Hydroxyglutarate levels compared between CTL and PAH patients categorized by the number of concurrent PAH-specific treatments. NoT = treatment- naïve patients; 1T = receiving single PAH treatment; 2T = dual PAH treatment; and 3T = tripe- combination PAH treatment. (d) 2-Hydroxyglutarate levels compared between CTL and sub- etiologies of PAH (acronyms defined in Table 1). (e) Survival curve for all patients after being strategies based on low (bottom 33^rd^ percentile), medium, or high (upper 67^th^ percentile) plasma 2-Hydroxyglutarate levels.

To confirm that the elevated levels of 2-Hydroxyglutarate were not a result of PAH-specific treatment, Fig. 3c demonstrates that levels are highest in treatment-naive patients and decrease proportionally with treatment intensity. Furthermore, elevated levels of 2- Hydroxyglutarate appear to be consistent across multiple sub-etiologies of PAH (see Fig. 3d), and patients with the highest circulating levels are at a statistically higher risk of poor outcomes (see Fig. 3e).

Fig 4 shows a bird-eye view of the partial relationships (corrected for confounding variables) between different routine clinical markers (demographics, hemodynamics, clinical blood and urine markers, exercise tolerance) and the 11 circulating molecules identified by the RFE algorithm. This analysis revealed that the multi-omics molecular signature is weakly correlated with demographic and exercise tolerance variables, but not associated with hemodynamics, lung function, or other well-established routine clinical blood and urine biomarkers of PAH. In fact, when considering the various clinical modalities utilized for patient evaluation (e.g., hemodynamics, lung function, blood and urine biomarkers), there appears to be little correlation among measurements from different modalities.

**Figure 4.**
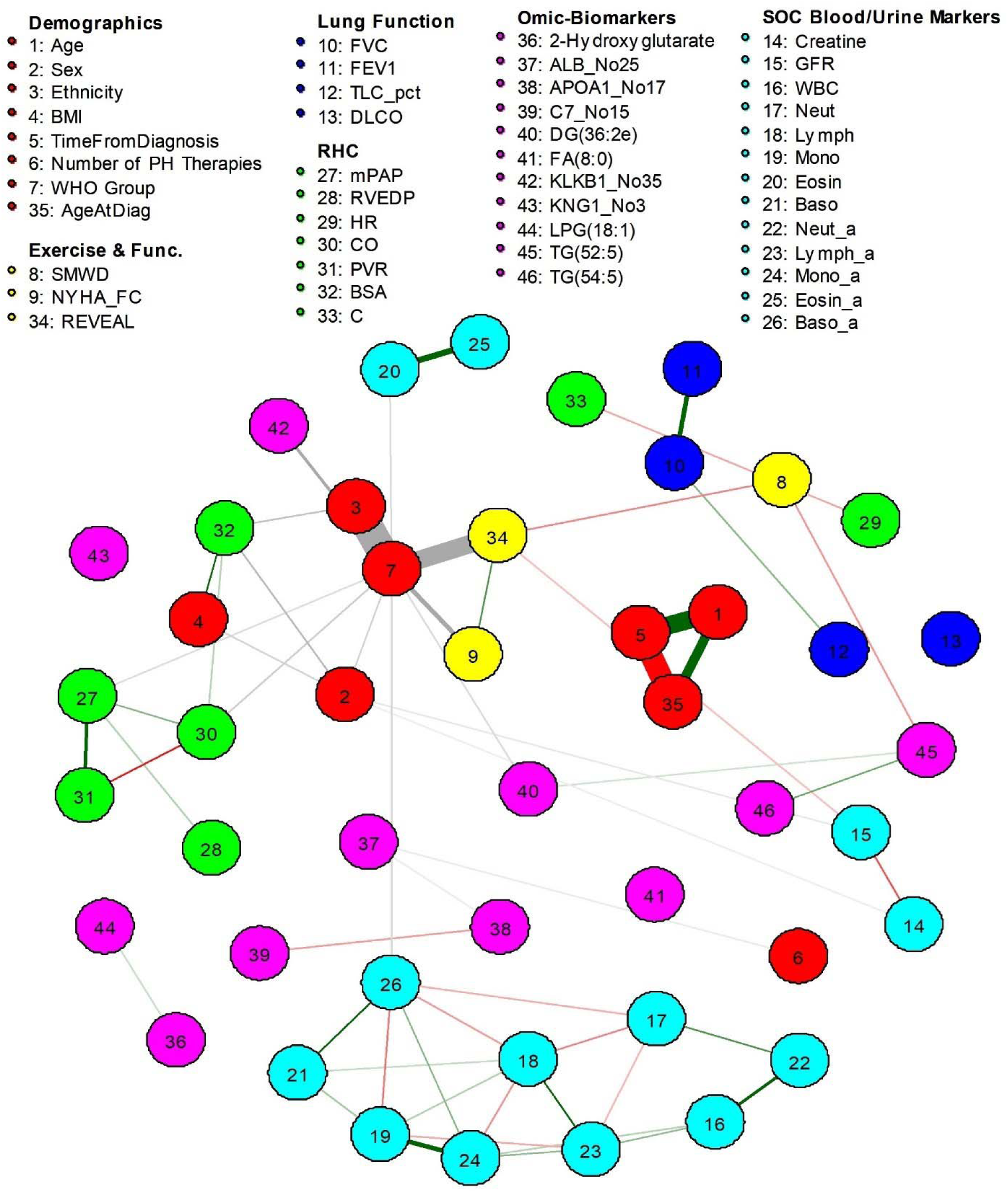
Association network of the mixed graphical model created using the Fruchterman-Reingold algorithm, which attempts to keep the distance between nodes equal and minimize edge-crossings. Between continuous variables, green and red edges indicate a positive and negative relationship, respectively, while edge thickness indicates the strength of the relationship. Associations involving categorical variables are represented with gray edges, with the width of the edge indicated by the strength of the association, but without any consideration of the direction.

### 3.3 Latent Network Discovery

While the RFE-RF analysis revealed predictive biomarkers of PAH, it did not offer any insight into the crosstalk between the metabolome, lipidome, and proteome. Furthermore, since RFE analysis is biased toward identifying only the best classifiers of PAH, it may overlook important molecules that, while not individually predictive, are part of biologically meaningful pathways.

To explore how PAH predictive molecular candidates interact with other compounds across the three omics domains, we employed latent network discovery (SLIDE analysis [24]). SLIDE is designed to identify latent networks, potentially capturing markers that are not immediately apparent as top predictors in traditional feature selection methods like RFE-RF.

The underlying assumption behind latent models is that there is some reduced combination of compounds (represented by a network or multiple networks) that can differentiate between CTL and PAH patients. In fact, our analysis showed that there are 7 stand- alone latent networks that can differentiate between CTL and PAH, and an additional 37 interactors that can predict PAH when interacting with the stand-alone networks (see Fig. 5a, all networks available in Supplemental Material 3). Fig 5b shows the model fit results in very good computational performance as indicated by the red dashed line (true performance of the significant latent networks) with a higher AUC than blue (random latent factors used for false discovery control) or green density (knockoffs paired with significant latent networks) plots.

**Figure 5.**
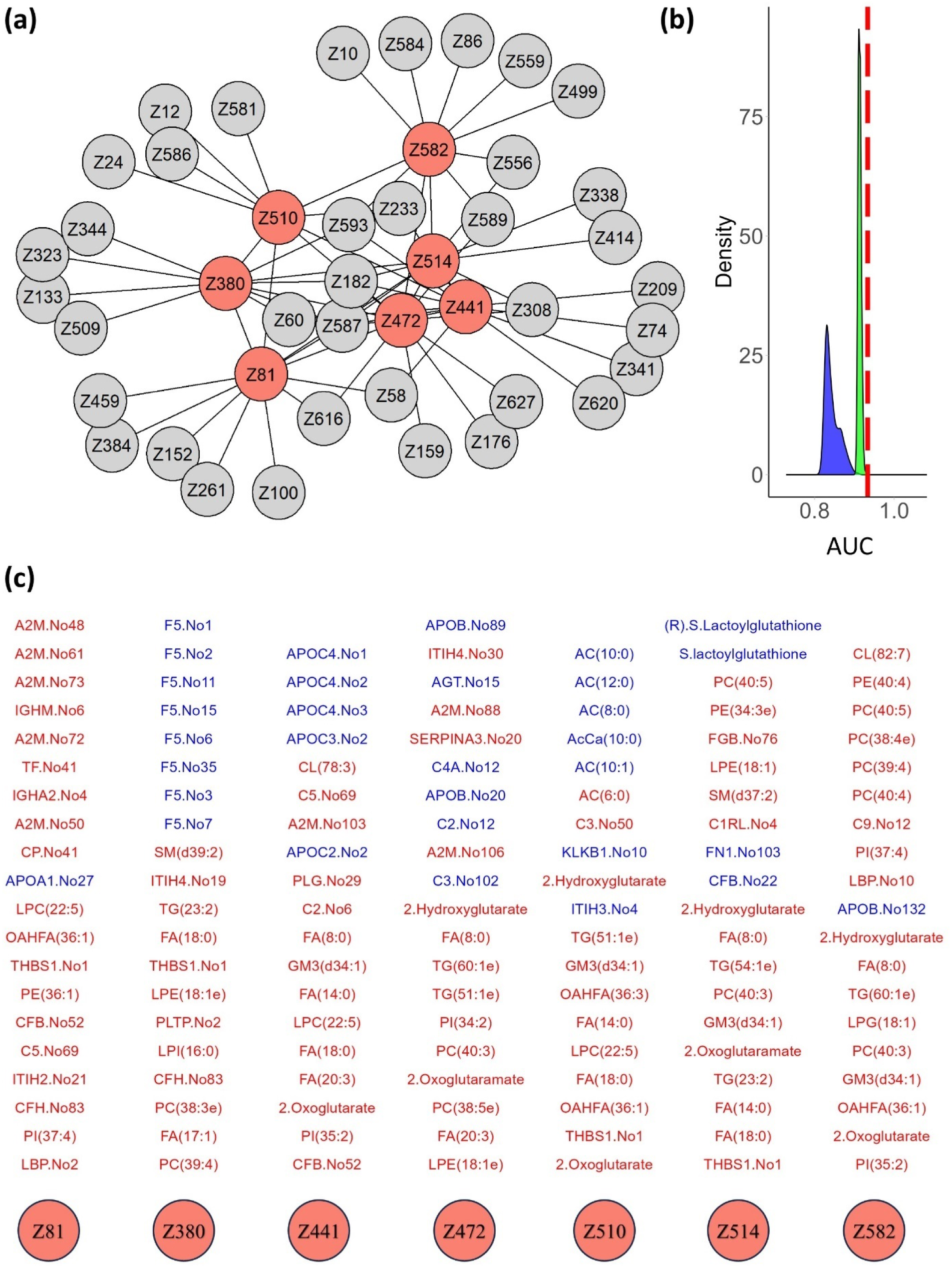
(a) Network showing interaction between underlying drivers of PAH (the network within each node is shown in Supplemental Material 3). Salmon color nodes represent latent networks that can classify PAH on their own (stand-alone). Grey nodes indicate interacting networks. (b) SLIDE model performance. Red line shows actual performance of the significant underlying drivers. Blue density shows the performance of the “knockoffs” (a fake driver introduced into the algorithm), while green density shows the performance of the true marginal latent variables paired with knockoffs. AUC = area under curve. (c) Table showing the compounds constituting the 7 latent networks identified by SLIDE analysis. Red markers are elevated in PAH, while blue markers are decreased, relative to CTL subjects.

Fig. 5c shows all the circulating blood compounds involved in the 7 stand-alone latent networks that can significantly distinguish PAH from CTL subjects. Three selected networks are shown in Fig. 6a-c, revealing notable interaction between circulating peptides, metabolites, and lipids. Out of the 7 significant latent networks, 5 included 2-Hydroxygluterate (and/or 2- Oxyglutarate), which appears to have a role in mediating crosstalk between HIF signaling, immune response and inflammation (e.g., complement immune system, Thrombospondin-1), and altered fatty acid lipid metabolism (e.g., acylcarnitines, triglycerides, gangliosides).

**Figure 6.**
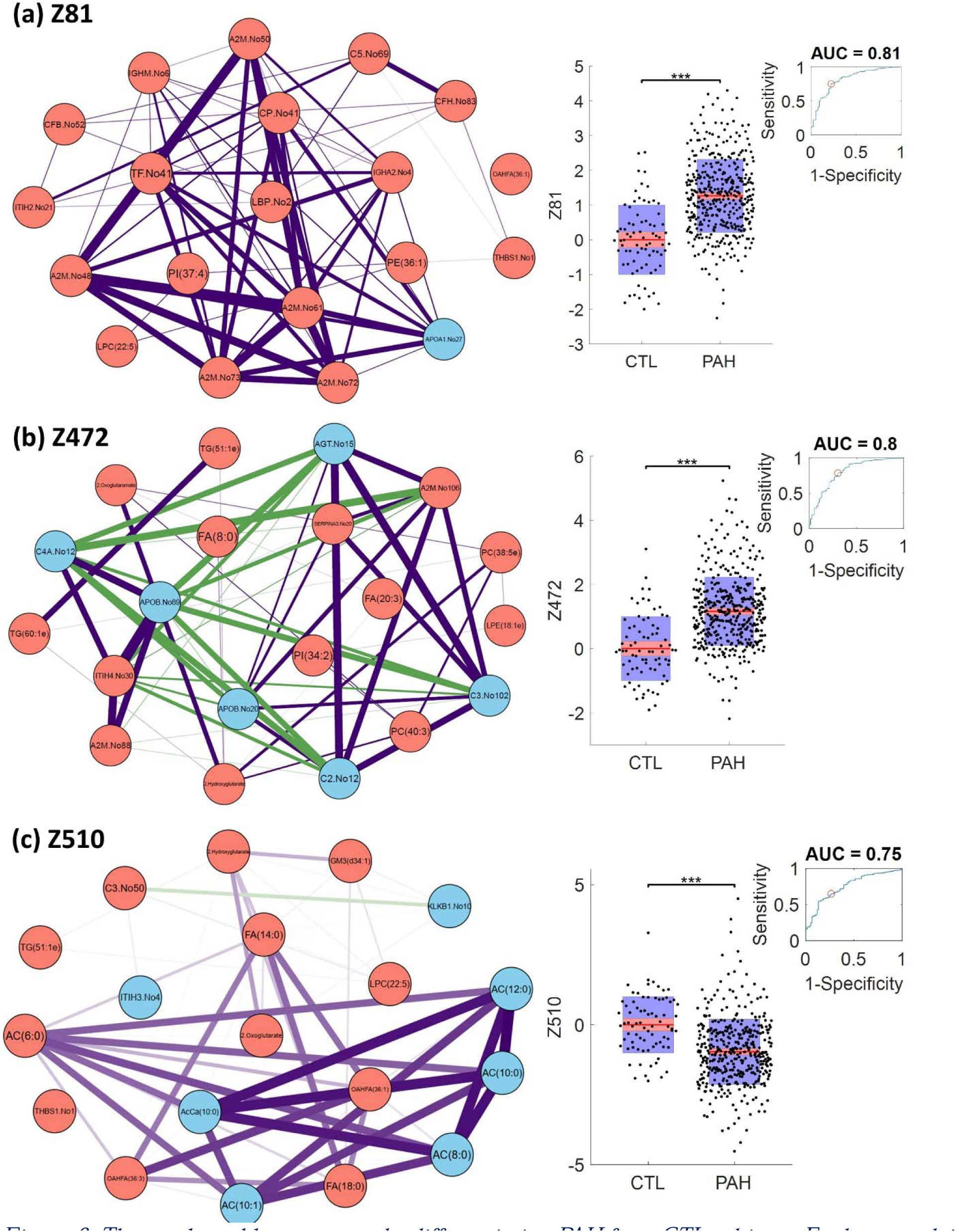
Three selected latent networks differentiating PAH from CTL subjects. Each network is accompanied by a bar plot, with a receiver operating curve insert, showing how each latent variable compares between control (CTL) and PAH patients. Note: All other networks in Fig. 5 can be found in Supplemental Material 3.

### 3.4 The Circulating Molecular Signature of PAH is Associated with Poor Outcomes

Finally, we sought to further explore the association between circulating multi-omics and outcomes in PAH. Of the 402 PAH patients included in the study, the mean time from sample collection to the event (defined as death or censoring) was 3.81 years. A total of 122 patients died, while 132 were censored at various time points before reaching 3.81 years, with some censored up to 14 years.

Combining all molecular compounds involved in the significant networks identified by SLIDE analysis consisted of 473 unique circulating molecules involved in PAH pathogenesis, which was narrowed down to 399 by removing highly correlated features (see Methods, section 2.3). To further reduce the dimensionality of this dataset and identify the least number of markers needed for an optimal model, the RFE-RSF algorithm was run 64 times, where each simulation showed that the c-index increases as you iteratively remove the least influential features (see Fig. 7a). Each RFE-RSF simulation randomly divided the PAH patients from the training cohort into 70% for training and the c-index was computed for the remaining 30% of the training cohort at each iteration of feature elimination. Therefore, the resulting c-index computed from each RSF simulation at the first iteration of RFE (when all simulations considered the same 399 features) is equivalent to a 64-fold cross validation range, which turned out to be between 0.62 and 0.78. All simulations displayed roughly the same behavior when iteratively removing non-influential features, which resulted in a maximum c-index between ∼0.75 and 0.85 at approximately 76 features. However, since not all simulations agreed on which 76 molecules were the most predictive, we retained only those molecules that appeared in at least 20% of the simulations, resulting in a final set of 155 circulating molecules that -combined into a multivariate model- were associated with outcomes in PAH. After training the final RSF model on the entire training cohort using these 155 features, the model exhibited strong performance (c-index = 0.762) in the testing cohort, which had been withheld from the RFE-RSF process (same testing cohort used for RFE-RF). Including potential confounders into the model, such as age, sex, etiology, ethnicity, the REVEAL risk score, and/or time from diagnosis did not improve accuracy, thus suggesting that the molecular signature of blood is in itself predictive and is not interacting with other confounders (e.g., no molecules were found to be associated with outcomes in men but not women). Figure 7b presents the feature importance plot for the final RSF model, highlighting peptides from the APOA1, KRT1, and FGB proteins, along with GM3(d34:1) and OAHFA(36:1) lipids, as the most influential predictors of outcomes (see Supplemental Material 4 for full list).

**Figure 7.**
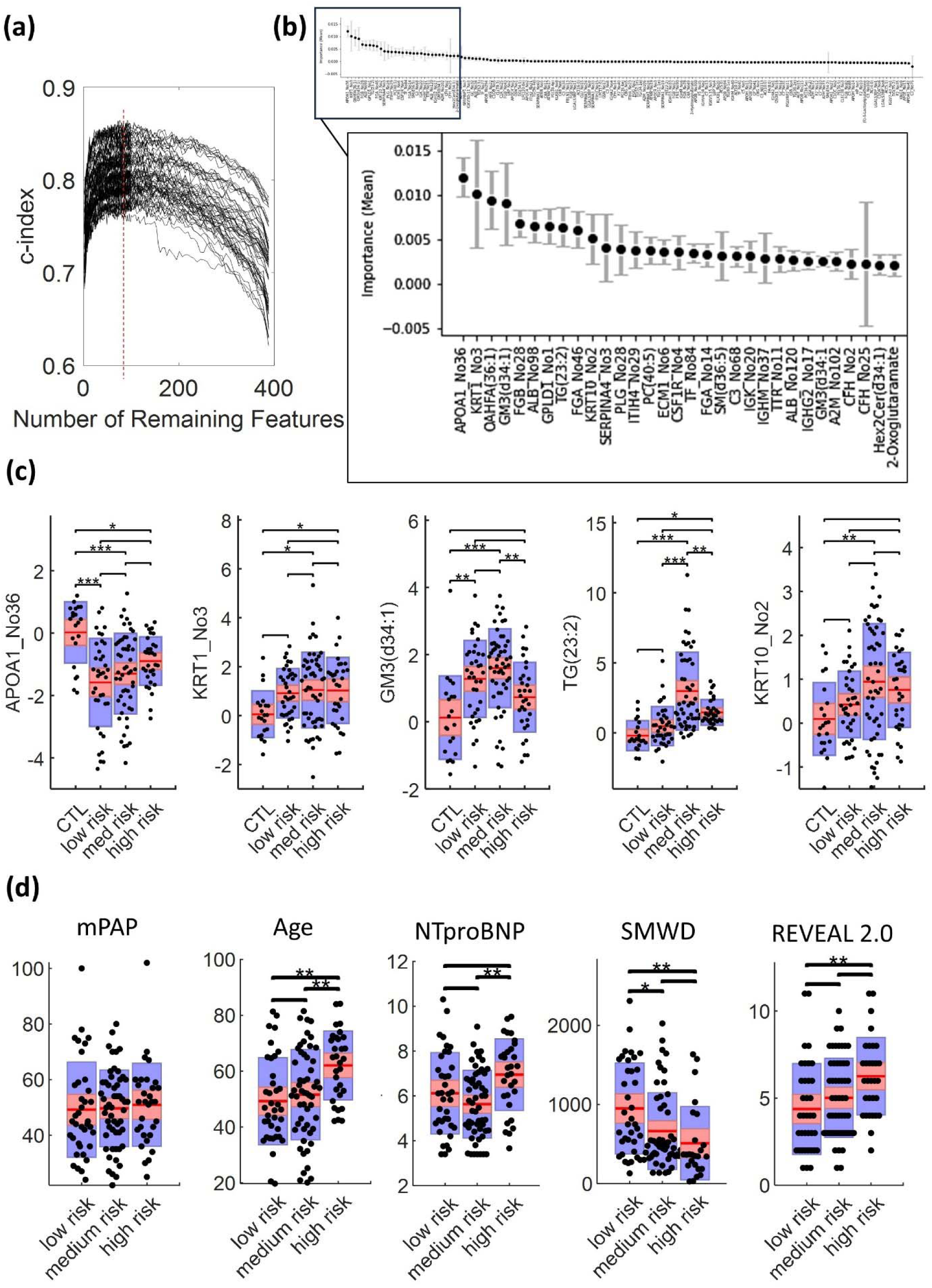
(a) RFE-RSF results for 64 independent simulations, all showing an increase in prognostic performance as the features are recursively decreased from 399 to roughly 75, at which point the performance begins to decrease. (b) Feature importance plot from a final trained RSF model to predict outcomes in the training cohort (see Supplemental Material 4). The model performed with a c-index = 0.762 in the testing cohort, which was withheld from model training. (c) Selected markers compared between CTL subjects and PAH subjects categorized as low, medium, or high risk by the trained RSF model from the testing cohort. (d) Demographic and clinical markers compared between patients categorized as low, medium, or high. Note: mPAP: mean pulmonary arterial pressure; SMWD: six-minute walk distance.

The 5-year hazard risk estimated by the final RSF model enabled the classification of patients into low (lower 33rd percentile), medium, and high risk (upper 66th percentile) categories for adverse outcomes. Figure 7c compares key markers identified by the RSF model between CTL subjects and PAH patients stratified by predicted risk levels in the testing cohort. Notably, patients assigned by the model into the high-risk category tended to be older, exhibited lower exercise tolerance, elevated NTproBNP levels, and had higher REVEAL 2.0 risk scores (see Fig. 7d).

## 4. Discussion

In this study, we conducted a comprehensive analysis of plasma samples from 402 individuals with PAH and 76 healthy control (CTL) subjects. Our investigation employed unbiased metabolomic (study of metabolites), lipidomic (study of lipids), and proteomic (study of proteins) analyses. Using a combination of well-established and cutting-edge machine learning methods, we discovered the following:

a. The molecules we identified as being significantly altered in PAH (either increased or decreased) are consistent with findings in the broader scientific literature. These include specific proteins (e.g., A2M, APOA1, C7, C9, complement peptides, SERPINs, and certain immunoglobulins [14]), metabolites (e.g., malate, fumarate, and 2-Oxoglutarate [25]), and lipids (e.g., fatty acids) [12]. Taken together, these molecules, if presumed to originate from the lung, align with our previous transcriptomic analyses of individual PAH lesions [26] and are implicated in key pathways, including hypoxia, fatty acid metabolism, complement activation, and glycolysis.
b. The metabolite 2-Hydroxyglutarate emerged as an excellent predictor of PAH, consistently elevated across various subtypes, highlighting hypoxia signaling as a pervasive feature of the disease. Notably, elevated levels were also associated with poor outcomes, but decreased with increasing PAH-specific treatments, suggesting its potential as a marker of disease severity and treatment response.
c. We uncovered hidden (latent) networks of molecules interacting with each other, revealing a major co-regulation between the proteome (peptides), metabolome (metabolites), and lipidome (lipids) in PAH. These findings highlight the interconnected nature of molecular pathways driving the disease.
d. When combining the multi-Omics data from the circulation into a survival model, we demonstrated that the abundance of certain molecules in the circulation is related to well-established clinical markers of the disease and even poor outcomes.

### 4.1 Circulating Omics-biomarkers differentiating PAH from CTL subjects

One of the most significant findings in our study was that elevated levels of 2-Hydroxyglutarate were the most impactful for distinguishing PAH patients from healthy CTL subjects. This molecule plays a role in the tricarboxylic acid (TCA) cycle, a critical pathway for energy production in cells, is linked with metabolic reprogramming in PAH [27] as well as hypoxic activation of lactate dehydrogenase [28]. 2-Hydroxyglutarate is produced through the reduction of 2-Oxoglutarate (another metabolite found to be elevated in PAH patients). Mechanistically, 2- Hydroxyglutarate enhances the activity of hypoxia-inducible factors (HIFs) [29], which are proteins activated under low oxygen conditions and well-known to contribute to the development and progression of PAH [30, 31].

While previous research, including our own, has characterized hypoxia-like metabolic reprogramming as a hallmark of pulmonary vascular pathobiology in PAH [20, 32], this current study marks the first report of elevated circulating levels of 2-Hydroxyglutarate across multiple PAH subtypes. Notably, levels decreased toward those of healthy controls in patients receiving PAH-specific treatments, with a greater decrease observed in those undergoing multiple therapies. It’s also noteworthy that patients with the lowest levels of 2-Hydroxyglutarate tended to have the best outcomes, but there was no significant association with outcomes and the number of PAH-specific treatments, thus highlighting its potentially critical role in the pathogenesis of disease progression. Future animal studies will investigate the cellular origin of 2-Hydroxyglutarate and examine how current and emerging PAH treatments affect these cells.

Beyond 2-Hydroxyglutarate, several other circulating molecules showed strong predictive power for PAH across our machine learning simulations. Many of these molecules, such as saturated fatty acids and triacylglycerols [32], have already been reported in PAH research. For instance, lower levels of Apolipoprotein A1 (APOA1) in PAH patients have been associated with increased resistance in pulmonary blood vessels and worse clinical outcomes in related diseases [33, 34]. In our analysis, APOA1 peptides were consistently decreased in PAH patients, both at the peptide and whole protein levels.

### 4.2 Interaction between the proteome, metabolome, and lipidome in PAH

Although there have been several studies separately exploring the proteomics, metabolomics, or lipidomics in PAH patients, to our knowledge, this is the first study to perform untargeted analysis across all three omics-domains simultaneously. This allowed for a unique opportunity to explore interaction across these omics-domains, particularly within latent networks, differentiating PAH from CTL subjects. This analysis confirmed earlier results, highlighting the central role of hypoxia and metabolic reprogramming in PAH. Circulating molecules like 2-Hydroxyglutarate and 2-Oxoglutarate were part of five of the seven major latent networks driving PAH pathogenesis. Within these networks, these metabolic intermediates interacted with various lipids, peptides, and other molecules with striking alignment with well-known cellular pathways of metabolic reprogramming [35]. We also observed interactions between 2-Hydroxyglutarate and markers of gangliosides, a type of lipid known to influence vascular diseases [36].

The identified networks highlighted a broad range of disrupted pathways in PAH, reflecting systemic dysregulation often appearing in the same networks with 2-Hydroxyglutarate. Notably, molecules involved in fatty acid metabolism, such as acylcarnitines (e.g., AC(10:0), AC(12:0), AC(8:0)), pointed to significant impairments in energy production and mitochondrial function. These findings align with previous reports of energy deficits in PAH [21, 22] and our previous data exploring the transcriptomics signature of vascular lesions in the human lung [26].

The abundance of proteins related to the complement cascade (e.g., C3, C7, CFB) and/or immunoglobulins (e.g., IGHA2, IGMH), which appear in each of the 7 stand-alone latent networks, highlight their role in immune surveillance and point to its important supporting role in almost all pathogenic pathways of PAH [14, 26, 37, 38]. Other inflammatory markers (e.g., THBS1, ITIH2,3,4) that appear in multiple latent networks highlight the significant inflammatory and immune responses in PAH [3]. These components, combined with elevated acute-phase proteins like A2M (Alpha-2-macroglobulin), suggest a proinflammatory state that contributes to vascular remodeling and disease progression [38], and occurs in concert with the aforementioned metabolic reprogramming pathways.

### 4.3 Prognosticating PAH based on multi-omics

When training a survival model on all biomarkers implicated in the latent networks of PAH, the model predicted outcomes in alignment with well-established clinical risk scores, suggesting that a broader molecular signature within the circulation captures critical aspects of the disease. Interestingly, 2-Hydroxyglutarate, while significant, was not the most predictive marker in the survival model.

This observation highlights the power of circulating biomarkers as a window into the pathobiology of PAH. The identified molecular signature includes markers of hypoxic signaling, triglycerides, and peptides from key systems such as the complement cascade, as well as well-characterized proteins like Apolipoprotein A-I [33] and Albumin. These findings align with and expand upon the broader PAH literature, which has highlighted the roles of Apolipoprotein A-I, gangliosides [36], and keratins [38] in disease mechanisms. These circulating molecules collectively reflect the intricate interplay of metabolic, inflammatory, and vascular remodeling pathways driving PAH. They offer a non-invasive means of studying its pathogenesis in humans while generating hypotheses for further exploration in animal models.

While this molecular signature may one day contribute to prognostic tools, our current focus is on its potential to reveal insights into PAH pathobiology. The fact that these circulating molecules track with cardiac function, exercise tolerance, and outcomes reinforces their value in understanding the systemic dysregulation inherent to this disease. Future studies integrating these findings with tissue-level analyses from human and large animal models will be crucial for deciphering how these molecular changes in the circulation are linked to specific cellular and organ-level dysfunctions in PAH.

### 4.4 Limitations, omics data integration and interpretation challenges, and future work

One consideration of the current study is the presence of patients with documented chronic obstructive pulmonary disease (COPD) and CTEPH. Technically, these patients could be considered as WHO group 3 and 4 PH, respectively, but were initially diagnosed with PAH in their clinical record because it was deemed that their hemodynamics were disproportionate to pulmonary function tests or interstitial changes. Therefore, we categorized these patients as PAH based on their official clinical diagnosis and considered COPD or CTEPH to be a secondary diagnosis.

From an analytical standpoint, it is worth noting that the methods used herein could not discriminate between the D(R)- or L(S)- enantiomers of 2-Hydroxyglutarate, the former deriving from aberrant activity of mutant isocitrate dehydrogenase – hence, unlikely to be relevant in the setting of PH – while the latter being a product of the non-canonical activity of lactate dehydrogenase [39], consistent with a higher glycolytic reprogramming of PH adventitia associated endothelial and immune cells [40].

The REVEAL 2.0 risk score was calculated without considering systolic blood pressure or the presence of pericardial effusion, which was not available in the patient record for the majority of subjects in the study cohort.

Aggregating peptide quantification into protein quantification is not always straight forward. The protein parsimony problem can yield varied results based on data fidelity and the employed strategy [41]. Furthermore, the peptide abundance available from shotgun proteomics offers insight beyond protein abundance, as it can include post-translational modifications, information about isoforms, and is the norm for modern clinical diagnostics aimed at protein quantification by LC-MS [42, 43]. Furthermore, latent network analysis highlights the advantages of looking at the proteome at the peptide level. We see that, in some cases, many peptides from the same protein are correlated with each other, but certain individual peptides are also correlated with other peptides, metabolites, or lipids (e.g., Z81 – F5, Z380 – A2M), thus offering a window into the interaction between the three omics domains that might not have been visible at the protein level. Therefore, our study focused on circulating peptide levels. Although the focus of the current study was not on biomarker discovery, the use of peptides, instead of proteins, for biomarkers is becoming recognized as the future of clinical chemistry and growing in the literature across multiple disciplines [42]. In a similar fashion, peptide probes are being developed for peptide-based Enzyme-Linked Immunosorbent Assay (ELISA) [42].

One puzzling result of the current analysis is the modest overlap between differentiating markers found by RFE-RF versus SLIDE. In fact, only 6 out of the 11 markers identified by RF-RFE were found to be included in latent factors: 2-Hydroxyglutarate, C7_No15, FA(8:0), KLKB1_No35, LPG(18:1), TG(52:5). A plausible explanation for this discrepancy is that SLIDE prioritizes markers that participate in latent variable interactions, which may involve complex networks or relationships between features. As a result, markers that are highly predictive on their own may not be part of these larger interacting networks, which SLIDE seeks to uncover. Specifically, SLIDE is designed to identify latent factors with strong biological inference, focusing on relationships that extend beyond mere prediction, potentially capturing markers that are not immediately apparent as top predictors in traditional feature selection methods like RFE-RF [24].

## Conclusion

This study confirmed and extended previous findings from in vitro and preclinical models by demonstrating, in humans, the critical role of metabolic reprogramming in the pathobiology of PAH. By leveraging metabolomics, lipidomics, and proteomics, we identified key molecules, such as 2-Hydroxyglutarate, that could predict PAH across multiple subtypes with excellent accuracy. This metabolite’s association with hypoxia signaling and its decrease with increasing PAH-specific treatments highlight its potential role in tracking disease severity and progression.

Furthermore, many of the molecules identified in circulation align with pathways enriched in pulmonary vascular lesions associated with PAH, including hypoxia signaling, immune activation, and lipid metabolism. Latent network analysis revealed intricate interactions between the proteome, metabolome, and lipidome, underscoring the interconnected nature of these pathways in driving systemic dysregulation. These findings validate and extend previous preclinical studies by uncovering human-specific interactions and generating hypotheses not apparent in animal models.

While animal models remain indispensable for studying PAH pathobiology, our work emphasizes the importance of human studies in revealing clinically relevant mechanisms and therapeutic targets. The integration of human and animal research will be critical to bridging gaps in understanding, enabling the translation of discoveries into treatments most likely to succeed in patients. This study highlights the power of circulating multi-omics to provide a holistic view of disease pathogenesis, linking metabolic and inflammatory pathways to outcomes and advancing our understanding of PAH in ways that may ultimately inform precision medicine.

Author contributions

KRS conceived the study; VOK developed and executed data analysis approach; VOK and KRS compiled original manuscript; VOK, KRS, PH, WMO, RTZ, KH, AD edited the manuscript; AJS, RTZ, HZ, TN, MD, DS, ISL, AD, WMO collected and organized patient data and/or generated omics data.

Grant Support

NIH P01HL152961 (VOK, KRS, and KCH); NIH R01HL152250 (VOK); K08HL128802 (WMO)

## Abbreviations

PAH: pulmonary arterial hypertension
SLIDE: Significant latent factor interaction discovery and exploration (tool)
RFE: recursive feature elimination
RF: random forest
RSF: random survival forest
MGM: mixed graphical modeling
c-index: concordance index.

## Data Availability

All data produced in the present study are not yet available.

## References

1. Levine, D.J., Pulmonary Arterial Hypertension: Updates in Epidemiology and Evaluation of Patients. American Journal of Managed Care, 2021. 27(3): p. S35–S41.

2. Oldham, W.M., A.R. Hemnes, M.A. Aldred, J. Barnard, E.L. Brittain, S.Y. Chan, F. Cheng, M.H. Cho, A.A. Desai, J.G.N. Garcia, M.W. Geraci, S.D. Ghiassian, K.T. Hall, E.M. Horn, M. Jain, R.S. Kelly, J.A. Leopold, S. Lindstrom, B.D. Modena, W.C. Nichols, C.J. Rhodes, W. Sun, A.J. Sweatt, R.R. Vanderpool, M.R. Wilkins, B. Wilmot, R.T. Zamanian, J.P. Fessel, N.R. Aggarwal, J. Loscalzo, and L. Xiao, NHLBI-CMREF Workshop Report on Pulmonary Vascular Disease Classification: JACC State-of-the-Art Review. J Am Coll Cardiol, 2021. 77(16): p. 2040–2052.

3. Sweatt, A.J., H.K. Hedlin, V. Balasubramanian, A. Hsi, L.K. Blum, W.H. Robinson, F. Haddad, P.M. Hickey, R. Condliffe, A. Lawrie, M.R. Nicolls, M. Rabinovitch, P. Khatri, and R.T. Zamanian, Discovery of Distinct Immune Phenotypes Using Machine Learning in Pulmonary Arterial Hypertension. Circ Res, 2019. 124(6): p. 904–919.

4. Kariotis, S., E. Jammeh, E.M. Swietlik, J.A. Pickworth, C.J. Rhodes, P. Otero, J. Wharton, J. Iremonger, M.J. Dunning, D. Pandya, T.S. Mascarenhas, N. Errington, A.A.R. Thompson, C.E. Romanoski, F. Rischard, J.G.N. Garcia, J.X.J. Yuan, T.-H.S. An, A.A. Desai, G. Coghlan, J. Lordan, P.A. Corris, L.S. Howard, R. Condliffe, D.G. Kiely, C. Church, J. Pepke-Zaba, M. Toshner, S. Wort, S. Gräf, N.W. Morrell, M.R. Wilkins, A. Lawrie, D. Wang, M. Bleda, C. Hadinnapola, M. Haimel, K. Auckland, T. Tilly, J.M. Martin, K. Yates, C.M. Treacy, M. Day, A. Greenhalgh, D. Shipley, A.J. Peacock, V. Irvine, F. Kennedy, S. Moledina, L. Macdonald, E. Tamvaki, A. Barnes, V. Cookson, L. Chentouf, S. Ali, S. Othman, L. Ranganathan, J.S.R. Gibbs, R. Dacosta, J. Pinguel, N. Dormand, A. Parker, D. Stokes, D. Ghedia, Y. Tan, T. Ngcozana, I. Wanjiku, G. Polwarth, R.V. Mackenzie Ross, J. Suntharalingam, M. Grover, A. Kirby, A. Grove, K. White, A. Seatter, A. Creaser-Myers, S. Walker, S. Roney, C.A. Elliot, A. Charalampopoulos, I. Sabroe, A. Hameed, I. Armstrong, N. Hamilton, A.M.K. Rothman, A.J. Swift, J.M. Wild, F. Soubrier, M. Eyries, M. Humbert, D. Montani, B. Girerd, L. Scelsi, S. Ghio, H. Gall, A. Ghofrani, H.J. Bogaard, A.V. Noordegraaf, A.C. Houweling, A.H.I.T. Veld and G. Schotte, Biological heterogeneity in idiopathic pulmonary arterial hypertension identified through unsupervised transcriptomic profiling of whole blood. Nature Communications, 2021. 12(1).

5. Gu, S., K. Goel, L.M. Forbes, V.O. Kheyfets, Y.A. Yu, R.M. Tuder, and K.R. Stenmark, Tensions in Taxonomies: Current Understanding and Future Directions in the Pathobiologic Basis and Treatment of Group 1 and Group 3 Pulmonary Hypertension. Compr Physiol, 2023. 13(1): p. 4295–4319.

6 Dufva, M.J., U. Truong, R. Shandas, and V.O. Kheyfets. Left Ventricular Torsion Rates by CMR correlate with invasively-derived hemodynamic data in Pediatric Pulmonary Hypertension. in Society for Cardiovascular Magnetic Resonance Annual Scientific Sessions. 2016. Los Angeles, CA.

7. Stenmark, K.R., B. Meyrick, N. Galie, W.J. Mooi, and I.F. McMurtry, Animal models of pulmonary arterial hypertension: the hope for etiological discovery and pharmacological cure. Am J Physiol Lung Cell Mol Physiol, 2009. 297(6): p. L1013–32.

8. Sweatt, A.J., K. Miyagawa, C.J. Rhodes, S. Taylor, P.A. Del Rosario, A. Hsi, F. Haddad, E. Spiekerkoetter, M. Bental-Roof, R.D. Bland, E.M. Swietlik, S. Gräf, M.R. Wilkins, N.W. Morrell, M.R. Nicolls, M. Rabinovitch, and R.T. Zamanian, Severe Pulmonary Arterial Hypertension Is Characterized by Increased Neutrophil Elastase and Relative Elafin Deficiency. Chest, 2021. 160(4): p. 1442–1458.

9. Rhodes, C.J., J. Wharton, E.M. Swietlik, L. Harbaum, B. Girerd, J.G. Coghlan, J. Lordan, C. Church, J. Pepke-Zaba, M. Toshner, S.J. Wort, D.G. Kiely, R. Condliffe, A. Lawrie, S. Gräf, D. Montani, A. Boucly, O. Sitbon, M. Humbert, L.S. Howard, N.W. Morrell, and M.R. Wilkins, Using the Plasma Proteome for Risk Stratifying Patients with Pulmonary Arterial Hypertension. Am J Respir Crit Care Med, 2022.

10. Pi, H., L. Xia, D.D. Ralph, S.G. Rayner, A. Shojaie, P.J. Leary, and S.A. Gharib, Metabolomic Signatures Associated With Pulmonary Arterial Hypertension Outcomes. Circ Res, 2023. 132(3): p. 254–266.

11. Kheyfets, V.O., A.J. Sweatt, M. Gomberg-Maitland, D.D. Ivy, R. Condliffe, D.G. Kiely, A. Lawrie, B.A. Maron, R.T. Zamanian, and K.R. Stenmark, Computational platform for doctor-artificial intelligence cooperation in pulmonary arterial hypertension prognostication: a pilot study. ERJ Open Res, 2023. 9(1).

12. Bordag, N., B.M. Nagy, E. Zugner, H. Ludwig, V. Foris, C. Nagaraj, V. Biasin, U. Bodenhofer, C. Magnes, B.A. Maron, S. Ulrich, T.J. Lange, K. Hotzenecker, T. Pieber, H. Olschewski, and A. Olschewski, Lipidomics for diagnosis and prognosis of pulmonary hypertension. medRxiv, 2023.

13. Rhodes, C.J., J. Wharton, P. Ghataorhe, G. Watson, B. Girerd, L.S. Howard, J.S.R. Gibbs, R. Condliffe, C.A. Elliot, D.G. Kiely, G. Simonneau, D. Montani, O. Sitbon, H. Gall, R.T. Schermuly, H.A. Ghofrani, A. Lawrie, M. Humbert, and M.R. Wilkins, Plasma proteome analysis in patients with pulmonary arterial hypertension: an observational cohort study. Lancet Respir Med, 2017. 5(9): p. 717–726.

14. Frid, M.G., B.A. McKeon, J.M. Thurman, B.A. Maron, M. Li, H. Zhang, S. Kumar, T. Sullivan, J. Laskowsky, M.A. Fini, S. Hu, R.M. Tuder, A. Gandjeva, M.R. Wilkins, C.J. Rhodes, P. Ghataorhe, J.A. Leopold, R.S. Wang, V.M. Holers, and K.R. Stenmark, Immunoglobulin-driven Complement Activation Regulates Proinflammatory Remodeling in Pulmonary Hypertension. Am J Respir Crit Care Med, 2020. 201(2): p. 224–239.

15. Harbaum, L., C.J. Rhodes, J. Wharton, A. Lawrie, J.H. Karnes, A.A. Desai, W.C. Nichols, M. Humbert, D. Montani, B. Girerd, O. Sitbon, M. Boehm, T. Novoyatleva, R.T. Schermuly, H.A. Ghofrani, M. Toshner, D.G. Kiely, L.S. Howard, E.M. Swietlik, S. Gräf, M. Pietzner, N.W. Morrell, and M.R. Wilkins, Mining the Plasma Proteome for Insights into the Molecular Pathology of Pulmonary Arterial Hypertension. Am J Respir Crit Care Med, 2022.

16. Karpievitch, Y.V., A.R. Dabney, and R.D. Smith, Normalization and missing value imputation for label-free LC-MS analysis. BMC Bioinformatics, 2012. 13 **Suppl 16**(Suppl 16): p. S5.

17. Chawade, A., E. Alexandersson, and F. Levander, Normalyzer: a tool for rapid evaluation of normalization methods for omics data sets. J Proteome Res, 2014. 13(6): p. 3114–20.

18. Jiang, H., Y. Deng, H.S. Chen, L. Tao, Q. Sha, J. Chen, C.J. Tsai, and S. Zhang, Joint analysis of two microarray gene-expression data sets to select lung adenocarcinoma marker genes. BMC Bioinformatics, 2004. 5: p. 81.

19. Ishwaran, H., U.B. Kogalur, E.H. Blackstone, and M.S. Lauer, Random survival forests. The Annals of Applied Statistics, 2008. 2(3): p. 841–860, 20.

20. Zhang, H., A. D’Alessandro, M. Li, J.A. Reisz, S. Riddle, A. Muralidhar, T. Bull, L. Zhao, E. Gerasimovskaya, and K.R. Stenmark, Histone deacetylase inhibitors synergize with sildenafil to suppress purine metabolism and proliferation in pulmonary hypertension. Vascul Pharmacol, 2023. 149: p. 107157.

21. Rhodes, C.J., J. Wharton, and M.R. Wilkins, Metabolomic Insights in Pulmonary Arterial Hypertension. Advances in Pulmonary Hypertension, 2018. 17(3): p. 103–109.

22. Rhodes, C.J., P. Ghataorhe, J. Wharton, K.C. Rue-Albrecht, C. Hadinnapola, G. Watson, M. Bleda, M. Haimel, G. Coghlan, P.A. Corris, L.S. Howard, D.G. Kiely, A.J. Peacock, J. Pepke-Zaba, M.R. Toshner, S.J. Wort, J.S. Gibbs, A. Lawrie, S. Graf, N.W. Morrell, and M.R. Wilkins, Plasma Metabolomics Implicates Modified Transfer RNAs and Altered Bioenergetics in the Outcomes of Pulmonary Arterial Hypertension. Circulation, 2017. 135(5): p. 460–475.

23. Bordag, N., B.M. Nagy, E. Zügner, H. Ludwig, V. Foris, C. Nagaraj, V. Biasin, U. Bodenhofer, C. Magnes, B.A. Maron, S. Ulrich, T.J. Lange, K. Hötzenecker, T. Pieber, H. Olschewski, and A. Olschewski, Lipidomics for diagnosis and prognosis of pulmonary hypertension. medRxiv, 2023.

24. Rahimikollu, J., H. Xiao, A. Rosengart, A.B.I. Rosen, T. Tabib, P.M. Zdinak, K. He, X. Bing, F. Bunea, M. Wegkamp, A.C. Poholek, A.V. Joglekar, R.A. Lafyatis, and J. Das, SLIDE: Significant Latent Factor Interaction Discovery and Exploration across biological domains. Nature Methods, 2024. 21(5): p. 835–845.

25. Wawrzyniak, R., P. Grešner, E. Lewicka, S. Macioszek, A. Furga, B. Zieba, J.M. M, and A. Da Browska-Kugacka, Metabolomics Meets Clinics: A Multivariate Analysis of Plasma and Urine Metabolic Signatures in Pulmonary Arterial Hypertension. J Proteome Res, 2023.

26. Tuder, R.M., A. Gandjeva, S. Williams, S. Kumar, V.O. Kheyfets, K.M. Hatton-Jones, J.R. Starr, J. Yun, J. Hong, N.P. West, and K.R. Stenmark, Digital Spatial Profiling Identifies Distinct Molecular Signatures of Vascular Lesions in Pulmonary Arterial Hypertension. Am J Respir Crit Care Med, 2024. 210(3): p. 329–342.

27. D’Alessandro, A., K.C. El Kasmi, L. Plecita-Hlavata, P. Jezek, M. Li, H. Zhang, S.A. Gupte, and K.R. Stenmark, Hallmarks of Pulmonary Hypertension: Mesenchymal and Inflammatory Cell Metabolic Reprogramming. Antioxid Redox Signal, 2018. 28(3): p. 230–250.

28. Intlekofer, A.M., R.G. Dematteo, S. Venneti, L.W. Finley, C. Lu, A.R. Judkins, A.S. Rustenburg, P.B. Grinaway, J.D. Chodera, J.R. Cross, and C.B. Thompson, Hypoxia Induces Production of L-2-Hydroxyglutarate. Cell Metab, 2015. 22(2): p. 304–11.

29. Oldham, W.M., C.B. Clish, Y. Yang, and J. Loscalzo, Hypoxia-Mediated Increases in L-2- hydroxyglutarate Coordinate the Metabolic Response to Reductive Stress. Cell Metab, 2015. 22(2): p. 291–303.

30. Hu, C.J., A. Laux, A. Gandjeva, L. Wang, M. Li, D. Brown, S. Riddle, V.O. Kheyfets, R.M. Tuder, H. Zhang, and K.R. Stenmark, The Effect of HIF Inhibition on the Phenotype of Fibroblasts in Human and Bovine Pulmonary Hypertension. Am J Respir Cell Mol Biol, 2023.

31. Kim, H., Y. Liu, J. Kim, Y. Kim, T. Klouda, S. Fisch, S.H. Baek, T. Liu, S. Dahlberg, C.J. Hu, W. Tian, X. Jiang, K. Kosmas, H.A. Christou, B.D. Korman, S.O. Vargas, J.C. Wu, K.R. Stenmark, V.J. Perez, M.R. Nicolls, B.A. Raby, and K. Yuan, *Pericytes contribute to pulmonary vascular remodeling via HIF2*α *signaling*. EMBO Rep, 2024.

32. Bassareo, P.P. and M. D’Alto, Metabolomics in Pulmonary Hypertension-A Useful Tool to Provide Insights into the Dark Side of a Tricky Pathology. Int J Mol Sci, 2023. 24(17).

33. Hemnes, A.R., J.M. Luther, C.J. Rhodes, J.P. Burgess, J. Carlson, R. Fan, J.P. Fessel, N. Fortune, R.E. Gerszten, S.J. Halliday, R. Hekmat, L. Howard, J.H. Newman, K.D. Niswender, M.E. Pugh, I.M. Robbins, Q. Sheng, C.A. Shibao, Y. Shyr, S. Sumner, M. Talati, J. Wharton, M.R. Wilkins, F. Ye, C. Yu, J. West, and E.L. Brittain, Human PAH is characterized by a pattern of lipid-related insulin resistance. JCI Insight, 2019. 4(1).

34. Yu, W., X. Dujiang, W. Yi, D. Guanwen, Z. Mengyu, P. Chang, Z. Aikai, Z. Juan, Z. Linlin, and Z. Hang, Apolipoprotein A1 is associated with pulmonary vascular resistance and adverse clinical outcomes in patients with pulmonary hypertension secondary to heart failure. Pulm Circ, 2022. 12(3): p. e12096.

35. Ferreira, A.V., J. Domínguez-Andrés, L.M. Merlo Pich, L.A.B. Joosten, and M.G. Netea, Metabolic Regulation in the Induction of Trained Immunity. Semin Immunopathol, 2024. 46(3-4): p. 7.

36. Sasaki, N. and M. Toyoda, Vascular Diseases and Gangliosides. Int J Mol Sci, 2019. 20(24).

37. Ito, S., H. Hashimoto, H. Yamakawa, D. Kusumoto, Y. Akiba, T. Nakamura, M. Momoi, J. Komuro, T. Katsuki, M. Kimura, Y. Kishino, S. Kashimura, A. Kunitomi, M. Lachmann, M. Shimojima, G. Yozu, C. Motoda, T. Seki, T. Yamamoto, Y. Shinya, T. Hiraide, M. Kataoka, T. Kawakami, K. Suzuki, K. Ito, H. Yada, M. Abe, M. Osaka, H. Tsuru, M. Yoshida, K. Sakimura, Y. Fukumoto, M. Yuzaki, K. Fukuda, and S. Yuasa, The complement C3-complement factor D-C3a receptor signalling axis regulates cardiac remodelling in right ventricular failure. Nat Commun, 2022. 13(1): p. 5409.

38. Zhang, L., S. Chen, X. Zeng, D. Lin, Y. Li, L. Gui, and M.J. Lin, Revealing the pathogenic changes of PAH based on multiomics characteristics. J Transl Med, 2019. 17(1): p. 231.

39. Intlekofer, A.M., B. Wang, H. Liu, H. Shah, C. Carmona-Fontaine, A.S. Rustenburg, S. Salah, M.R. Gunner, J.D. Chodera, J.R. Cross, and C.B. Thompson, L-2- Hydroxyglutarate production arises from noncanonical enzyme function at acidic pH. Nat Chem Biol, 2017. 13(5): p. 494–500.

40. Li, M., S. Riddle, H. Zhang, A. D’Alessandro, A. Flockton, N.J. Serkova, K.C. Hansen, R. Moldvan, B.A. McKeon, M. Frid, S. Kumar, H. Li, H. Liu, A. Caanovas, J.F. Medrano, M.G. Thomas, D. Iloska, L. Plecita-Hlavata, P. Jezek, S. Pullamsetti, M.A. Fini, K.C. El Kasmi, Q. Zhang, and K.R. Stenmark, Metabolic Reprogramming Regulates the Proliferative and Inflammatory Phenotype of Adventitial Fibroblasts in Pulmonary Hypertension Through the Transcriptional Corepressor C-Terminal Binding Protein-1. Circulation, 2016. 134(15): p. 1105–1121.

41. Matzke, M.M., J.N. Brown, M.A. Gritsenko, T.O. Metz, J.G. Pounds, K.D. Rodland, A.K. Shukla, R.D. Smith, K.M. Waters, J.E. McDermott, and B.J. Webb-Robertson, A comparative analysis of computational approaches to relative protein quantification using peptide peak intensities in label-free LC-MS proteomics experiments. Proteomics, 2013. 13(3-4): p. 493–503.

42. Pandey, S., G. Malviya, and M. Chottova Dvorakova, Role of Peptides in Diagnostics. Int J Mol Sci, 2021. 22(16).

43. Neubert, H., C.M. Shuford, T.V. Olah, F. Garofolo, G.A. Schultz, B.R. Jones, L. Amaravadi, O.F. Laterza, K. Xu, and B.L. Ackermann, Protein Biomarker Quantification by Immunoaffinity Liquid Chromatography–Tandem Mass Spectrometry: Current State and Future Vision. Clinical Chemistry, 2020. 66(2): p. 282–301.

