## Supplemental Material 1 for "Identification of candidate biomarkers and molecular networks associated with Pulmonary Arterial Hypertension using machine learning and plasma multi-Omics analysis"

| Feature | Importance_mean | Importance_std |
| --- | --- | --- |
| APOA1_No36 | 0.012032005 | 0.002240004 |
| KRT1_No3 | 0.010144928 | 0.006088749 |
| OAHFA(36:1) | 0.009450483 | 0.00327057 |
| GM3(d34:1) | 0.009027778 | 0.004615617 |
| FGB_No28 | 0.006808575 | 0.001545764 |
| ALB_No98 | 0.006582126 | 0.001806807 |
| GPLD1_No1 | 0.006551932 | 0.002000743 |
| TG(23:2) | 0.006446256 | 0.002232361 |
| FGA_No46 | 0.006099034 | 0.002032949 |
| KRT10_No2 | 0.005147947 | 0.002811945 |
| SERPINA4_No3 | 0.004136473 | 0.003783322 |
| PLG_No28 | 0.003940217 | 0.002814781 |
| ITIH4_No29 | 0.003879831 | 0.002071925 |
| PC(40:5) | 0.003789251 | 0.001530204 |
| ECM1_No6 | 0.003698671 | 0.001365389 |
| CSF1R_No4 | 0.003592995 | 0.001923027 |
| TF_No84 | 0.003517512 | 0.001096559 |
| FGA_No14 | 0.003321256 | 0.001250379 |
| SM(d36:5) | 0.00321558 | 0.002748911 |
| C3_No68 | 0.003185386 | 0.001153089 |
| IGK_No20 | 0.00317029 | 0.001724586 |
| IGHM_No37 | 0.002974034 | 0.002781877 |
| TTR_No11 | 0.002868357 | 0.001431395 |
| ALB_No120 | 0.002687198 | 0.001133353 |
| IGHG2_No17 | 0.002657005 | 0.00101091 |
| GM3(d34:1 | 0.002641908 | 0.000616933 |
| A2M_No102 | 0.002626812 | 0.001028788 |
| CFH_No2 | 0.002324879 | 0.001612867 |
| CFH_No25 | 0.002279589 | 0.006923804 |
| Hex2Cer(d34:1) | 0.002204106 | 0.001107726 |
| 2-Oxoglutaramate | 0.00218901 | 0.001177149 |
| A2M_No44 | 0.001751208 | 0.004340481 |
| SERPINF1_No7 | 0.001388889 | 0.000834558 |
| IGKV3D-11_No3 | 0.001388889 | 0.000809607 |
| GC_No32 | 0.001192633 | 0.000557554 |
| ALB_No34 | 0.001086957 | 0.000811013 |
| TG(52:5) | 0.001056763 | 0.000959556 |
| TF_No77 | 0.000920894 | 0.000684195 |
| APOB_No294 | 0.000785024 | 0.000705838 |
| HPX_No22 | 0.000467995 | 0.000506131 |
| C1QB_No5 | 0.000467995 | 0.000658739 |
| CL(78:3) | 0.000452899 | 0.001096975 |
| C1R_No25 | 0.000437802 | 0.000506131 |

| Feature | Importance_mean | Importance_std |
|---|---|---|
| FGB_No43 | 0.000422705 | 0.000752714 |
| LBP_No2 | 0.000377415 | 0.000410672 |
| APOC4_No1 | 0.000332126 | 0.000660811 |
| GSN_No1 | 0.000317029 | 0.000420001 |
| DG(34:1) | 0.000301932 | 0.00044272 |
| LPC(20:0e) | 0.000301932 | 0.001272064 |
| APOC4_No2 | 0.000286836 | 0.000335539 |
| ITIH2_No21 | 0.000166063 | 0.000280001 |
| TG(36:4e) | 0.000166063 | 0.000674128 |
| SERPINA1_No60 | 0.000166063 | 0.000506131 |
| ATRN_No5 | 0.000150966 | 0.000552627 |
| C4A_No12 | 0.00013587 | 0.000328677 |
| ATRN_No17 | 0.000120773 | 0.000377717 |
| PE(38:4p) | 9.06E-05 | 0.000420001 |
| KLKB1_No8 | 9.06E-05 | 0.000644751 |
| C4B_No6 | 7.55E-05 | 0.000426997 |
| C3_No34 | 6.04E-05 | 0.000210272 |
| FBLN1_No13 | 6.04E-05 | 0.000391931 |
| FGB_No72 | 6.04E-05 | 0.000314503 |
| LGALS3BP_No11 | 6.04E-05 | 0.000492437 |
| AC(16:0) | 4.53E-05 | 0.000188557 |
| CLU_No15 | 4.53E-05 | 0.000776559 |
| SERPINA3_No24 | 1.51E-05 | 0.000409004 |
| LGALS3BP_No9 | 1.51E-05 | 0.000391931 |
| SERPINA3_No33 | 1.51E-05 | 0.000400559 |
| ITIH3_No15 | 1.51E-05 | 0.000425393 |
| LPC(17:0) | 1.51E-05 | 0.00062135 |
| GSN_No17 | -7.40E-18 | 0.000233876 |
| C3_No8 | -1.51E-05 | 0.000254413 |
| IGHA2_No4 | -1.51E-05 | 0.000557554 |
| CFH_No14 | -3.02E-05 | 0.000459394 |
| F2_No40 | -3.02E-05 | 0.000459394 |
| PC(36:6e) | -3.02E-05 | 0.000757243 |
| C8G_No5 | -6.04E-05 | 0.000280001 |
| ITIH4_No23 | -6.04E-05 | 0.000291955 |
| FGG_No2 | -7.55E-05 | 0.000178626 |
| TG(52:1e) | -7.55E-05 | 0.000337571 |
| TG(34:1e) | -7.55E-05 | 0.000402262 |
| A2M_No106 | -9.06E-05 | 0.000110937 |
| SERPINA1_No24 | -9.06E-05 | 0.000338918 |
| ATRN_No19 | -0.000105676 | 0.000216677 |
| APOC4_No3 | -0.000105676 | 0.000560001 |
| TG(51:1e) | -0.000105676 | 0.000386662 |

| Feature | Importance_mean | Importance_std |
| --- | --- | --- |
| SERPINA1_No19 | -0.000105676 | 0.000296601 |
| PE(22:2) | -0.000105676 | 0.000451891 |
| IGHG3_No10 | -0.000105676 | 0.000386662 |
| FGB_No57 | -0.000120773 | 0.000272577 |
| FN1_No90 | -0.000120773 | 0.000296601 |
| IGHM_No6 | -0.000150966 | 0.000228952 |
| FGB_No78 | -0.000150966 | 0.000316669 |
| C4A_No49 | -0.000166063 | 0.000210272 |
| 2-Hydroxyglutarate | -0.000166063 | 0.000280001 |
| APOB_No300 | -0.000166063 | 0.000291955 |
| F5_No35 | -0.000181159 | 0.000276726 |
| IGHV4-34_No2 | -0.000181159 | 0.000288815 |
| C7_No13 | -0.000211353 | 0.000240601 |
| IGKV3-15_No3 | -0.000211353 | 0.000325191 |
| LBP_No4 | -0.000226449 | 0.000286438 |
| KLKB1_No20 | -0.000226449 | 0.000387839 |
| AC(5:0) | -0.000256643 | 0.000216677 |
| APOB_No94 | -0.000256643 | 0.000246218 |
| TG(60:1e) | -0.000271739 | 0.000439101 |
| IGHM_No40 | -0.000271739 | 0.000300419 |
| AC(10:1) | -0.000271739 | 0.000300419 |
| TF_No41 | -0.000271739 | 0.000332811 |
| C5_No5 | -0.000271739 | 0.000380722 |
| APOH_No11 | -0.000286836 | 0.000478351 |
| APOB_No20 | -0.000286836 | 0.000267513 |
| CP_No41 | -0.000286836 | 0.000240601 |
| C8G_No2 | -0.000286836 | 0.000210272 |
| C3_No113 | -0.000301932 | 0.000514172 |
| AC(10:0) | -0.000301932 | 0.000479778 |
| FA(8:0) | -0.000317029 | 0.000258853 |
| IGHA2_No1 | -0.000317029 | 0.000595121 |
| QSOX1_No16 | -0.000317029 | 0.000318105 |
| C2_No12 | -0.000332126 | 0.000259732 |
| C7_No31 | -0.000332126 | 0.000246218 |
| PGLYRP2_No11 | -0.000332126 | 0.00035916 |
| C8A_No4 | -0.000332126 | 0.000231919 |
| TG(36:2e) | -0.000347222 | 0.000329369 |
| AGT_No15 | -0.000347222 | 0.000200279 |
| APOB_No115 | -0.000362319 | 0.000386072 |
| PC(36:5e) | -0.000362319 | 0.000215623 |
| DG(36:2) | -0.000362319 | 0.000386072 |
| A2M_No72 | -0.000377415 | 0.000196836 |
| F2_No12 | -0.000377415 | 0.000228952 |

| Feature | Importance_mean | Importance_std |
| --- | --- | --- |
| C7_No10 | -0.000392512 | 0.000225945 |
| A2M_No73 | -0.000392512 | 0.000325191 |
| SM(d34:2) | -0.000392512 | 0.004114874 |
| APOA1_No27 | -0.000392512 | 0.000240601 |
| C1QC_No2 | -0.000392512 | 0.000409004 |
| PE(36:5) | -0.000407609 | 0.000300419 |
| CLEC3B_No3 | -0.000422705 | 0.000307912 |
| FGB_No82 | -0.000437802 | 0.000703898 |
| C3_No88 | -0.000437802 | 0.000425393 |
| APOH_No14 | -0.000437802 | 0.000174756 |
| HP_No1 | -0.000452899 | 0.00034093 |
| F2_No10 | -0.000452899 | 0.000298134 |
| (R)-S-Lactoylglutathione | -0.000483092 | 0.000386662 |
| C3_No102 | -0.000483092 | 0.000246218 |
| AC(4:0) | -0.000483092 | 0.000216677 |
| LGALS3BP_No1 | -0.000528382 | 0.00038489 |
| C1QA_No6 | -0.000543478 | 0.000559594 |
| LGALS3BP_No8 | -0.000543478 | 0.000271739 |
| AC(5:1) | -0.000558575 | 0.00035916 |
| IGLV7-43_No1 | -0.000558575 | 0.000162596 |
| DG(35:2) | -0.000573671 | 0.000386662 |
| F2_No35 | -0.000588768 | 0.000271739 |
| KNG1_No9 | -0.000603865 | 0.000305683 |
| APOE_No13 | -0.000679348 | 0.00036979 |
| APOA1_No42 | -0.000815217 | 0.000394828 |
| C3_No76 | -0.00200785 | 0.004287705 |
