## Supplemental Material 2 for "Identification of candidate biomarkers and molecular networks associated with Pulmonary Arterial Hypertension using machine learning and plasma multi-Omics analysis"

### Supplemental Material 1

#### Mass Spectrometry and Multi-Omics Analysis for the primary cohort

*Proteomics* – Plasma samples were prepared as previously described [1]. Briefly, approximately 50ug plasma proteins were digested with trypsin using S-Trap 96-well plates (Protifi, Huntington, NY, USA) following the manufacturer procedures. Peptides were separated using an Evosep One system using the 30 runs per day method (Evosep, Odense, Denmark) and a Pepsep column. The system was coupled to a timsTOF MS Pro (Bruker Daltonics, Bremen Germany) via electro spray ionization. The MS was operated in diaPASEF mode. Raw DIA files were searched using Spectronaut v 17.0 (Biognosys, Schlieren, Switzerland), using a project-specific spectral library from DDA runs of 24 fractions and searched with the Pulsar search engine.

*Metabolomics and Lipidomics* –High throughput metabolomics and lipidomics were performed via ultra-high performance liquid chromatography coupled to high-resolution mass spectrometry, consistent with previously published methods [2]. A volume of 480 uL of ice cold 5:3:2 MeOH:MeCN:water (v/v/v) or 180 uL pure methanol was added to 20 uL of each sample for metabolomics and lipidomics, respectively, prior to vortexing for 30 min at 4ºC and centrifugation for 10 min at 18,000*g* at 4ºC. Blanks and quality control samples (generated by mixing 5 uL of all sample extracts) were injected every 20 samples and used to monitor instrument performance throughout the analysis. Metabolites were resolved on a Phenomenex Kinetex C18 column (2.1 x 30 mm, 1.7 um) at 45 °C using a 1-minute ballistic gradient method in positive and negative ion modes (separate runs) over the scan range 65-975 m/z exactly as previously described [3]. The UHPLC was coupled online to a Q Exactive mass spectrometer (Thermo Fisher). The Q Exactive MS was operated in negative ion mode, scanning in Full MS mode (2 μscans) from 90 to 900 m/z at 70,000 resolution, with 4 kV spray voltage, 45 sheath gas, 15 auxiliary gas. Following data acquisition, .raw files were converted to .mzXML using RawConverter then metabolites assigned and peaks integrated using ElMaven (Elucidata) in conjunction with an in-house standard library [4].

Untargeted lipidomics analyses were performed via UHPLC-MS/MS, as previously described [5]. Briefly, lipidomics analysis employed a Vanquish UHPLC system (Thermo Fisher Scientific) coupled to a Q Exactive mass spectrometer (Thermo Fisher Scientific). A volume of 5 μL injections of the samples were resolved across a 2.1 x 30 mm, 1.7 um particle size Kinetex C18 column (Phenomenex) using a 5 minute, reversed-phase gradient adapted from a previous method. The Q Exactive was run independently in positive and negative ion mode, scanning using data dependent $MS^2$ (top 10) from 125-1500 m/z at 17,500 resolution, with 45 Arb sheath gas, 25 Arb auxiliary gas, and 4 kV spray voltage. Calibration was performed prior to the run using the PierceTM Positive and Negative Ion Calibration Solutions (Thermo Fisher Scientific). Run order of samples was randomized and technical replicates were injected after every 4 samples to assess quality control. Lipid assignments and peak integration were performed using LipidSearch v 5.0 (Thermo Fisher Scientific).

1. LaCroix, I.S., M. Cohen, E.E. Moore, M. Dzieciatkowska, T. Nemkov, T.R. Schaid, Jr., M. Debot, K. Jones, C.C. Silliman, K.C. Hansen, and A. D'Alessandro, *Omics Markers of Red Blood Cell Transfusion in Trauma.* Int J Mol Sci, 2022. **23**(22).
2. D'Alessandro, A., *High-Throughput Metabolomics: Methods and Protocols*. Methods in Molecular Biology. Springer Nature.
3. Nemkov, T., T. Yoshida, M. Nikulina, and A. D'Alessandro, *High-Throughput Metabolomics Platform for the Rapid Data-Driven Development of Novel Additive Solutions for Blood Storage.* Front Physiol, 2022. **13**: p. 833242.
4. Nemkov, T., K.C. Hansen, and A. D'Alessandro, *A three-minute method for high-throughput quantitative metabolomics and quantitative tracing experiments of central carbon and nitrogen pathways.* Rapid Commun Mass Spectrom, 2017. **31**(8): p. 663-673.
5. Reisz, J.A., C. Zheng, A. D'Alessandro, and T. Nemkov, *Untargeted and Semi-targeted Lipid Analysis of Biological Samples Using Mass Spectrometry-Based Metabolomics.* Methods Mol Biol, 2019. **1978**: p. 121-135.
