## Supplemental Material 3 for "Identification of candidate biomarkers and molecular networks associated with Pulmonary Arterial Hypertension using machine learning and plasma multi-Omics analysis"

| | A | B | C |
|---|---|---|---|
| 1 | **Peptide Name** | **Peptide Sequence** | **Peptide Description** |
| 2 | 'IGKV3-7_No1' | 'EIVMTQSPPTLSLSPGER' | 'Probable non-functional immunoglobulin kappa variable 3-7' |
| 3 | 'IGKV3-7_No2' | 'ASQSVSSSYLTWYQQKPGQAPR' | 'Probable non-functional immunoglobulin kappa variable 3-7' |
| 4 | IGKV3-7_No3' | 'EIVMTQSPPTLSLSPGER' | 'Probable non-functional immunoglobulin kappa variable 3-7' |
| 5 | 'IGLV4-69_No1' | 'LTCTLSSGHSSYAIAWHQQQPEK' | 'Immunoglobulin lambda variable 4-69' |
| 6 | 'IGLV4-69_No2' | 'GDGIPDRFSGSSSGAER' | 'Immunoglobulin lambda variable 4-69' |
| 7 | 'IGLV4-69_No3' | 'LTCTLSSGHSSYAIAWHQQQPEKGPR' | 'Immunoglobulin lambda variable 4-69' |
| 8 | 'IGLV4-69_No4' | 'FSGSSSGAER' | 'Immunoglobulin lambda variable 4-69' |
| 9 | 'IGLV4-69_No5' | 'LNSDGSHSKGDGIPDRFSGSSSGAER' | 'Immunoglobulin lambda variable 4-69' |
| 10 | 'IGLV8-61_No1' | 'FSGSILGNK' | 'Immunoglobulin lambda variable 8-61' |
| 11 | 'IGLV8-61_No2' | 'TLIYSTNTR' | 'Immunoglobulin lambda variable 8-61' |
| 12 | 'IGLV8-61_No3' | 'SSGVPDRFSGSILGNK' | 'Immunoglobulin lambda variable 8-61' |
| 13 | 'IGLV4-60_No1' | 'GSGVPDRFSGSSSGADR' | 'Immunoglobulin lambda variable 4-60' |
| 14 | 'IGLV4-60_No2' | 'LTCTLSSGHSSYIIAWHQQQPGK' | 'Immunoglobulin lambda variable 4-60' |
| 15 | 'IGLV4-60_No3' | 'FSGSSSGADR' | 'Immunoglobulin lambda variable 4-60' |
| 16 | 'IGLV10-54' | 'NNNRPSGISER' | 'Immunoglobulin lambda variable 10-54' |
| 17 | 'IGLV1-50_No1' | 'VTISCTGSSSNIGAGYVVHWYQQLPGTAPK' | 'Probable non-functional immunoglobulin lambda variable 1-50' |
| 18 | 'IGLV1-50_No2' | 'LLIYGNSNRPSGVPDQFSGSK' | 'Probable non-functional immunoglobulin lambda variable 1-50' |
| 19 | 'IGLV5-48' | 'IYWYQQKPGSPPR' | 'Probable non-functional immunoglobulin lambda variable 5-48' |
| 20 | 'IGLV7-46_No1' | 'TLIYDTSNK' | 'Immunoglobulin lambda variable 7-46' |
| 21 | 'IGLV7-46_No2' | 'TLIYDTSNKHSWTPAR' | 'Immunoglobulin lambda variable 7-46' |
| 22 | 'IGLV5-37' | 'YLLYYYSDSDKGQGSGVPSR' | 'Immunoglobulin lambda variable 5-37' |
| 23 | 'IGLV2-33' | 'LLIYNVNTR' | 'Probable non-functional immunoglobulin lambda variable 2-33' |
| 24 | 'IGLV2-18_No1' | 'VSWYQQPPGTAPK' | 'Immunoglobulin lambda variable 2-18' |
| 25 | 'IGLV2-18_No2' | 'LMIYEVSNRPSGVPDR' | 'Immunoglobulin lambda variable 2-18' |
| 26 | 'IGLV2-18_No3' | 'LMIYEVSNRPSGVPDR' | 'Immunoglobulin lambda variable 2-18' |
| 27 | 'IGLV2-18_No4' | 'LMIYEVSNRPSGVPDRFSGSK' | 'Immunoglobulin lambda variable 2-18' |
| 28 | 'IGLV2-18_No5' | 'LMIYEVSNRPSGVPDRFSGSK' | 'Immunoglobulin lambda variable 2-18' |
| 29 | 'IGLV3-16_No1' | 'ITCSGEALPK' | 'Immunoglobulin lambda variable 3-16' |
| 30 | 'IGLV3-16_No2' | 'PGQFPVLVIYK' | 'Immunoglobulin lambda variable 3-16' |
| 31 | 'IGLV3-12' | 'FSGSNPGNTATLTISR' | 'Immunoglobulin lambda variable 3-12' |
| 32 | 'IGLV3-10_No1' | 'SGQAPVLVIYEDSK' | 'Immunoglobulin lambda variable 3-10' |
| 33 | 'IGLV3-10_No2' | 'RPSGIPER' | 'Immunoglobulin lambda variable 3-10' |
| 34 | 'IGLV3-10_No3' | 'YAYWYQQK' | 'Immunoglobulin lambda variable 3-10' |
| 35 | 'IGLV3-10_No4' | 'SGQAPVLVIYEDSKRPSGIPER' | 'Immunoglobulin lambda variable 3-10' |
| 36 | 'IGLV3-10_No5' | 'SGQAPVLVIYEDSKR' | 'Immunoglobulin lambda variable 3-10' |
| 37 | 'IGLV3-10_No6' | 'ITCSGDALPKK' | 'Immunoglobulin lambda variable 3-10' |
| 38 | 'IGLV3-9_No1' | 'PGQAPVLVIYR' | 'Immunoglobulin lambda variable 3-9' |
| 39 | 'IGLV3-9_No2' | 'NVHWYQQKPGQAPVLVIYR' | 'Immunoglobulin lambda variable 3-9' |
| 40 | 'IGLV3-9_No3' | 'NVHWYQQK' | 'Immunoglobulin lambda variable 3-9' |
| 41 | 'IGKV2-28_No1' | 'SSQSLLHSNGYNYLDWYLQKPGQSPQLLIYLGSNR' | 'Immunoglobulin kappa variable 2-28' |
| 42 | 'IGKV2-28_No2' | 'PGQSPQLLIYLGSNR' | 'Immunoglobulin kappa variable 2-28' |
| 43 | 'IGKV2-28_No3' | 'ASGVPDRFSGSGSGTDFTLK' | 'Immunoglobulin kappa variable 2-28' |
| 44 | 'IGKV2-28_No4' | 'SSQSLLHSNGYNYLDWYLQK' | 'Immunoglobulin kappa variable 2-28' |
| 45 | 'IGKV2-28_No5' | 'ASGVPDRFSGSGSGTDFTLKISR' | 'Immunoglobulin kappa variable 2-28' |

| | A | B | C |
|---|---|---|---|
| 1 | **Peptide Name** | **Peptide Sequence** | **Peptide Description** |
| 46 | 'IGKV2-28_No6' | 'VEAEDVGVYYCMQALQTP' | 'Immunoglobulin kappa variable 2-28' |
| 47 | 'IGHV3-64_No1' | 'AEDMAVYYCAR' | 'Immunoglobulin heavy variable 3-64' |
| 48 | 'IGHV3-64_No2' | 'AEDMAVYYCAR' | 'Immunoglobulin heavy variable 3-64' |
| 49 | 'IGHV3-64_No3' | 'GLEYVSAISSNGGSTYYADSVK' | 'Immunoglobulin heavy variable 3-64' |
| 50 | 'IGHV3-64_No4' | 'NTLYLQMGSLRAEDMAVYYCAR' | 'Immunoglobulin heavy variable 3-64' |
| 51 | 'IGHV3-64_No5' | 'NTLYLQMGSLR' | 'Immunoglobulin heavy variable 3-64' |
| 52 | 'IGHV3-64_No6' | 'DNSKNTLYLQMGSLR' | 'Immunoglobulin heavy variable 3-64' |
| 53 | 'IGHV3-64_No7' | 'NTLYLQMGSLR' | 'Immunoglobulin heavy variable 3-64' |
| 54 | 'IGHV3-64_No8' | 'DNSKNTLYLQMGSLR' | 'Immunoglobulin heavy variable 3-64' |
| 55 | 'IGHV4-4_No1' | 'GLEWIGEIYHSGSTNYNPSLK' | 'Immunoglobulin heavy variable 4-4' |
| 56 | 'IGHV4-4_No2' | 'VTISVDKSKNQFSLK' | 'Immunoglobulin heavy variable 4-4' |
| 57 | 'IGHV4-4_No3' | 'GLEWIGEIYHSGSTNYNPSLKSR' | 'Immunoglobulin heavy variable 4-4' |
| 58 | 'IGHV4-4_No4' | 'SKNQFSLK' | 'Immunoglobulin heavy variable 4-4' |
| 59 | 'IGKV2D-29' | 'PGQPPQLLIYEVSNR' | 'Immunoglobulin kappa variable 2D-29' |
| 60 | 'IGKV1D-17_No1' | 'ARQGISNYLAWFQQKPGK' | 'Immunoglobulin kappa variable 1D-17' |
| 61 | 'IGKV1D-17_No2' | 'QGISNYLAWFQQKPGK' | 'Immunoglobulin kappa variable 1D-17' |
| 62 | 'IGKV1D-17_No3' | 'QGISNYLAWFQQKPGK' | 'Immunoglobulin kappa variable 1D-17' |
| 63 | 'IGKV1D-17_No4' | 'HLIYAASSLQSGVPSR' | 'Immunoglobulin kappa variable 1D-17' |
| 64 | 'IGKV1-27_No1' | 'ASQGISNYLAWYQQKPGK' | 'Immunoglobulin kappa variable 1-27' |
| 65 | 'IGKV1-27_No2' | 'ASQGISNYLAWYQQK' | 'Immunoglobulin kappa variable 1-27' |
| 66 | 'IGKV1-27_No3' | 'FSGSGSGTDFTLTISSLQPEDVATYYCQK' | 'Immunoglobulin kappa variable 1-27' |
| 67 | 'IGKV1-27_No4' | 'VPKLLIYAASTLQSGVPSR' | 'Immunoglobulin kappa variable 1-27' |
| 68 | 'IGKV1-27_No5' | 'ASQGISNYLAWYQQKPGKVPK' | 'Immunoglobulin kappa variable 1-27' |
| 69 | 'IGKV2D-30' | 'VSNWDSGVPDR' | 'Immunoglobulin kappa variable 2D-30' |
| 70 | 'IGKV1-37_No1' | 'LLIYSASNLQSGVPSR' | 'Probable non-functional immunoglobulinn kappa variable 1-37' |
| 71 | 'IGKV1-37_No2' | 'VSQGISSYLNWYR' | 'Probable non-functional immunoglobulinn kappa variable 1-37' |
| 72 | 'IGLV5-45_No1' | 'IYWYQQKPGSPPQYLLR' | 'Immunoglobulin lambda variable 5-45' |
| 73 | 'IGLV5-45_No2' | 'PGSPPQYLLR' | 'Immunoglobulin lambda variable 5-45' |
| 74 | 'IGHV4-30-2_No1' | 'GLEWIGYIYHSGSTYYNPSLK' | 'Immunoglobulin heavy variable 4-30-2' |
| 75 | 'IGHV4-30-2_No2' | 'GLEWIGYIYHSGSTYYNPSLKSR' | 'Immunoglobulin heavy variable 4-30-2' |
| 76 | 'IGHV4-30-2_No3' | 'SRVTISVDR' | 'Immunoglobulin heavy variable 4-30-2' |
| 77 | 'IGKV3D-15' | 'LLIYGASIR' | 'Immunoglobulin kappa variable 3D-15' |
| 78 | 'IGKV1D-8_No1' | 'APELLIYAASTLQSGVPSR' | 'Immunoglobulin kappa variable 1D-8' |
| 79 | 'IGKV1D-8_No2' | 'MSQGISSYLAWYQQK' | 'Immunoglobulin kappa variable 1D-8' |
| 80 | 'IGKV1D-8_No3' | 'MSQGISSYLAWYQQKPGK' | 'Immunoglobulin kappa variable 1D-8' |
| 81 | 'IGKV1D-8_No4' | 'MSQGISSYLAWYQQK' | 'Immunoglobulin kappa variable 1D-8' |
| 82 | 'IGKV1D-8_No5' | 'MSQGISSYLAWYQQKPGK' | 'Immunoglobulin kappa variable 1D-8' |
| 83 | 'IGKV2-40_No1' | 'VEAEDVGVYYCMQR' | 'Immunoglobulin kappa variable 2-40' |
| 84 | 'IGKV2-40_No2' | 'VEAEDVGVYYCMQR' | 'Immunoglobulin kappa variable 2-40' |
| 85 | 'IGKV2-40_No3' | 'SSQSLLDSDDGNTYLDWYLQK' | 'Immunoglobulin kappa variable 2-40' |
| 86 | 'IGKV2-40_No4' | 'PGQSPQLLIYTLSYR' | 'Immunoglobulin kappa variable 2-40' |
| 87 | 'IGKV2-40_No5' | 'ISRVEAEDVGVYYCMQR' | 'Immunoglobulin kappa variable 2-40' |
| 88 | 'IGKV2-40_No6' | 'ISRVEAEDVGVYYCMQR' | 'Immunoglobulin kappa variable 2-40' |
| 89 | 'IGKV3D-11_No1' | 'LLIYDASNR' | 'Immunoglobulin kappa variable 3D-11' |

| | A | B | C |
|---|---|---|---|
| 1 | **Peptide Name** | **Peptide Sequence** | **Peptide Description** |
| 90 | 'IGKV3D-11_No2' | 'ASQSVSSYLAWYQQKPGQAPR' | 'Immunoglobulin kappa variable 3D-11' |
| 91 | 'IGKV3D-11_No3' | 'ASQSVSSYLAWYQQK' | 'Immunoglobulin kappa variable 3D-11' |
| 92 | 'IGKV3D-11_No4' | 'LLIYDASNRATGIPAR' | 'Immunoglobulin kappa variable 3D-11' |
| 93 | 'IGLV5-52_No1' | 'FSGSNDASANAGILR' | 'Immunoglobulin lambda variable 5-52' |
| 94 | 'IGLV5-52_No2' | 'ISGLQPEDEADYYCGTWHSNSK' | 'Immunoglobulin lambda variable 5-52' |
| 95 | 'IGHV1-45_No1' | 'SEDTAMYYCAR' | 'Immunoglobulin heavy variable 1-45' |
| 96 | 'IGHV1-45_No2' | 'SMSTAYMELSSLR' | 'Immunoglobulin heavy variable 1-45' |
| 97 | 'IGHV1-45_No3' | 'SEDTAMYYCAR' | 'Immunoglobulin heavy variable 1-45' |
| 98 | 'IGHV1-45_No4' | 'SMSTAYMELSSLR' | 'Immunoglobulin heavy variable 1-45' |
| 99 | 'IGHV1-45_No5' | 'ASGYTFTYR' | 'Immunoglobulin heavy variable 1-45' |
| 100 | 'IGHV3-49_No1' | 'SIAYLQMNSLK' | 'Immunoglobulin heavy variable 3-49' |
| 101 | 'IGHV3-49_No2' | 'QAPGKGLEWVGFIR' | 'Immunoglobulin heavy variable 3-49' |
| 102 | 'IGHV3-49_No3' | 'GLEWVGFIR' | 'Immunoglobulin heavy variable 3-49' |
| 103 | 'IGHV3-49_No4' | 'LSCTASGFTFGDYAMSWVR' | 'Immunoglobulin heavy variable 3-49' |
| 104 | 'IGHV3-49_No5' | 'QAPGKGLEWVGFIR' | 'Immunoglobulin heavy variable 3-49' |
| 105 | 'IGHV3-49_No6' | 'SIAYLQMNSLK' | 'Immunoglobulin heavy variable 3-49' |
| 106 | 'IGHV3-49_No7' | 'LSCTASGFTFGDYAMSWVR' | 'Immunoglobulin heavy variable 3-49' |
| 107 | 'IGHV3-49_No8' | 'AYGGTTEYAASVK' | 'Immunoglobulin heavy variable 3-49' |
| 108 | 'IGHV3-49_No9' | 'SKAYGGTTEYAASVK' | 'Immunoglobulin heavy variable 3-49' |
| 109 | 'IGKV6D-21' | 'YASQSISGVPSR' | 'Immunoglobulin kappa variable 6D-21' |
| 110 | 'IGKJ1_No1' | 'WTFGQGTK' | 'Immunoglobulin kappa joining 1' |
| 111 | 'IGKJ1_No2' | 'WTFGQGTKVEIK' | 'Immunoglobulin kappa joining 1' |
| 112 | 'IGLV1-36_No1' | 'LLIYYDDLLPSGVSDR' | 'Immunoglobulin lambda variable 1-36' |
| 113 | 'IGLV1-36_No2' | 'LLIYYDDLLPSGVSDRFSGSK' | 'Immunoglobulin lambda variable 1-36' |
| 114 | 'IGLV1-36_No3' | 'VTISCSGSSSNIGNNAVNWYQQLPGK' | 'Immunoglobulin lambda variable 1-36' |
| 115 | 'IGHV6-1_No1' | 'WYNDYAVSVK' | 'Immunoglobulin heavy variable 6-1' |
| 116 | 'IGHV6-1_No2' | 'GLEWLGR' | 'Immunoglobulin heavy variable 6-1' |
| 117 | 'IGHV6-1_No3' | 'SKWYNDYAVSVK' | 'Immunoglobulin heavy variable 6-1' |
| 118 | 'IGHV6-1_No4' | 'ITINPDTSKNQFSLQLNSVTPEDTAVYYCAR' | 'Immunoglobulin heavy variable 6-1' |
| 119 | 'IGHV6-1_No5' | 'NQFSLQLNSVTPEDTAVYYCAR' | 'Immunoglobulin heavy variable 6-1' |
| 120 | 'IGHV6-1_No6' | 'QSPSRGLEWLGR' | 'Immunoglobulin heavy variable 6-1' |
| 121 | 'IGHV3-15_No1' | 'NTLYLQMNSLK' | 'Immunoglobulin heavy variable 3-15' |
| 122 | 'IGHV3-15_No2' | 'TDGGTTDYAAPVK' | 'Immunoglobulin heavy variable 3-15' |
| 123 | 'IGHV3-15_No3' | 'NTLYLQMNSLK' | 'Immunoglobulin heavy variable 3-15' |
| 124 | 'IGHV3-15_No4' | 'LSCAASGFTFSNAWMSWVR' | 'Immunoglobulin heavy variable 3-15' |
| 125 | 'IGHV3-15_No5' | 'LSCAASGFTFSNAWMSWVR' | 'Immunoglobulin heavy variable 3-15' |
| 126 | 'IGHV3-15_No6' | 'DDSKNTLYLQMNSLK' | 'Immunoglobulin heavy variable 3-15' |
| 127 | 'IGHV3-15_No7' | 'FTISRDDSKNTLYLQMNSLK' | 'Immunoglobulin heavy variable 3-15' |
| 128 | 'IGHV3-15_No8' | 'SKTDGGTTDYAAPVKGR' | 'Immunoglobulin heavy variable 3-15' |
| 129 | 'IGHV3-15_No9' | 'SKTDGGTTDYAAPVK' | 'Immunoglobulin heavy variable 3-15' |
| 130 | 'IGHV3-15_No10' | 'DDSKNTLYLQMNSLK' | 'Immunoglobulin heavy variable 3-15' |
| 131 | 'IGHV3-15_No11' | 'TDGGTTDYAAPVKGR' | 'Immunoglobulin heavy variable 3-15' |
| 132 | 'IGHV3-15_No12' | 'FTISRDDSKNTLYLQMNSLK' | 'Immunoglobulin heavy variable 3-15' |
| 133 | 'IGHV3-15_No13' | 'QAPGKGLEWVGRIK' | 'Immunoglobulin heavy variable 3-15' |

| | A | B | C |
|---|---|---|---|
| 1 | **Peptide Name** | **Peptide Sequence** | **Peptide Description** |
| 134 | 'IGHV3-21_No1' | 'LSCAASGFTFSSYSMNWVR' | 'Immunoglobulin heavy variable 3-21' |
| 135 | 'IGHV3-21_No2' | 'LSCAASGFTFSSYSMNWVR' | 'Immunoglobulin heavy variable 3-21' |
| 136 | 'IGHV3-21_No3' | 'QAPGKGLEWVSSISSSSSYIYYADSVK' | 'Immunoglobulin heavy variable 3-21' |
| 137 | 'IGHV2-26_No1' | 'MGVSWIR' | 'Immunoglobulin heavy variable 2-26' |
| 138 | 'IGHV2-26_No2' | 'ALEWLAHIFSNDEK' | 'Immunoglobulin heavy variable 2-26' |
| 139 | 'IGHV2-26_No3' | 'MGVSWIR' | 'Immunoglobulin heavy variable 2-26' |
| 140 | 'IGHV2-26_No4' | 'ESGPVLVKPTETLTLTCTVSGFSLSNAR' | 'Immunoglobulin heavy variable 2-26' |
| 141 | 'IGHV2-26_No5' | 'ALEWLAHIFSNDEKSYSTSLK' | 'Immunoglobulin heavy variable 2-26' |
| 142 | 'IGHV2-26_No6' | 'SQVVLTMTNMDPVDTATYYCAR' | 'Immunoglobulin heavy variable 2-26' |
| 143 | 'IGHV2-26_No7' | 'DTSKSQVVLTMTNMDPVDTATYYCAR' | 'Immunoglobulin heavy variable 2-26' |
| 144 | 'IGHV2-26_No8' | 'MGVSWIRQPPGK' | 'Immunoglobulin heavy variable 2-26' |
| 145 | 'IGHV3-73_No1' | 'TEDTAVYYCTR' | 'Immunoglobulin heavy variable 3-73' |
| 146 | 'IGHV3-73_No2' | 'GLEWVGR' | 'Immunoglobulin heavy variable 3-73' |
| 147 | 'IGHV3-73_No3' | 'LSCAASGFTFSGSAMHWVR' | 'Immunoglobulin heavy variable 3-73' |
| 148 | 'IGHV3-73_No4' | 'NTAYLQMNSLK' | 'Immunoglobulin heavy variable 3-73' |
| 149 | 'IGHV3-73_No5' | 'ANSYATAYAASVK' | 'Immunoglobulin heavy variable 3-73' |
| 150 | 'IGHV3-73_No6' | 'DDSKNTAYLQMNSLK' | 'Immunoglobulin heavy variable 3-73' |
| 151 | 'IGHV3-73_No7' | 'QASGKGLEWVGR' | 'Immunoglobulin heavy variable 3-73' |
| 152 | 'IGHV3-73_No8' | 'NTAYLQMNSLK' | 'Immunoglobulin heavy variable 3-73' |
| 153 | 'IGHV3-73_No9' | 'LSCAASGFTFSGSAMHWVR' | 'Immunoglobulin heavy variable 3-73' |
| 154 | 'IGHV3-73_No10' | 'FTISRDDSK' | 'Immunoglobulin heavy variable 3-73' |
| 155 | 'IGHV3-73_No11' | 'DDSKNTAYLQMNSLK' | 'Immunoglobulin heavy variable 3-73' |
| 156 | 'IGHV3-73_No12' | 'SKANSYATAYAASVK' | 'Immunoglobulin heavy variable 3-73' |
| 157 | 'IGHV3-74_No1' | 'INSDGSSTSYADSVK' | 'Immunoglobulin heavy variable 3-74' |
| 158 | 'IGHV3-74_No2' | 'GLVWVSR' | 'Immunoglobulin heavy variable 3-74' |
| 159 | 'IGHV3-74_No3' | 'LSCAASGFTFSSYWMHWVR' | 'Immunoglobulin heavy variable 3-74' |
| 160 | 'IGHV3-74_No4' | 'DNAKNTLYLQMNSLR' | 'Immunoglobulin heavy variable 3-74' |
| 161 | 'IGHV3-74_No5' | 'QAPGKGLVWVSR' | 'Immunoglobulin heavy variable 3-74' |
| 162 | 'IGHV3-74_No6' | 'QAPGKGLVWVSR' | 'Immunoglobulin heavy variable 3-74' |
| 163 | 'IGHV3-74_No7' | 'INSDGSSTSYADSVKGR' | 'Immunoglobulin heavy variable 3-74' |
| 164 | 'IGHV3-74_No8' | 'LSCAASGFTFSSYWMHWVR' | 'Immunoglobulin heavy variable 3-74' |
| 165 | 'IGHV3-74_No9' | 'DNAKNTLYLQMNSLR' | 'Immunoglobulin heavy variable 3-74' |
| 166 | 'IGHV3-74_No10' | 'DNAKNTLYLQMNSLRAEDTAVYYCAR' | 'Immunoglobulin heavy variable 3-74' |
| 167 | 'IGHV3-74_No11' | 'DNAKNTLYLQMNSLRAEDTAVYYCAR' | 'Immunoglobulin heavy variable 3-74' |
| 168 | 'IGHV3-43_No1' | 'TEDTALYYCAK' | 'Immunoglobulin heavy variable 3-43' |
| 169 | 'IGHV3-43_No2' | 'LSCAASGFTFDDYTMHWVR' | 'Immunoglobulin heavy variable 3-43' |
| 170 | 'IGHV3-43_No3' | 'DNSKNSLYLQMNSLR' | 'Immunoglobulin heavy variable 3-43' |
| 171 | 'IGHV3-43_No4' | 'LSCAASGFTFDDYTMHWVR' | 'Immunoglobulin heavy variable 3-43' |
| 172 | 'IGHV3-43_No5' | 'NSLYLQMNSLRTEDTALYYCAK' | 'Immunoglobulin heavy variable 3-43' |
| 173 | 'IGHV3-43_No6' | 'NSLYLQMNSLRTEDTALYYCAK' | 'Immunoglobulin heavy variable 3-43' |
| 174 | 'IGHV3-43_No7' | 'DNSKNSLYLQMNSLR' | 'Immunoglobulin heavy variable 3-43' |
| 175 | 'IGLV9-49_No1' | 'VDWYQQRPGK' | 'Immunoglobulin lambda variable 9-49' |
| 176 | 'IGLV9-49_No2' | 'FSVLGSGLNR' | 'Immunoglobulin lambda variable 9-49' |
| 177 | 'IGLV9-49_No3' | 'VDWYQQR' | 'Immunoglobulin lambda variable 9-49' |

| | A | B | C |
|---|---|---|---|
| 1 | **Peptide Name** | **Peptide Sequence** | **Peptide Description** |
| 178 | 'IGLV9-49_No4' | 'VGTGGIVGSKGDGIPDR' | 'Immunoglobulin lambda variable 9-49' |
| 179 | 'IGLV9-49_No5' | 'VDWYQQRPGKGPR' | 'Immunoglobulin lambda variable 9-49' |
| 180 | 'IGLV9-49_No6' | 'VGTGGIVGSKGDGIPDRFSVLGSGLNR' | 'Immunoglobulin lambda variable 9-49' |
| 181 | 'IGLV9-49_No7' | 'VGTGGIVGSK' | 'Immunoglobulin lambda variable 9-49' |
| 182 | 'IGLV9-49_No8' | 'GDGIPDRFSVLGSGLNR' | 'Immunoglobulin lambda variable 9-49' |
| 183 | 'IGHV3-72_No1' | 'LSCAASGFTFSDHYMDWVR' | 'Immunoglobulin heavy variable 3-72' |
| 184 | 'IGHV3-72_No2' | 'NSLYLQMNSLK' | 'Immunoglobulin heavy variable 3-72' |
| 185 | 'IGHV3-72_No3' | 'TEDTAVYYCAR' | 'Immunoglobulin heavy variable 3-72' |
| 186 | 'IGHV3-72_No4' | 'NSLYLQMNSLK' | 'Immunoglobulin heavy variable 3-72' |
| 187 | 'IGHV3-72_No5' | 'QAPGKGLEWVGR' | 'Immunoglobulin heavy variable 3-72' |
| 188 | 'IGHV3-72_No6' | 'QAPGKGLEWVGR' | 'Immunoglobulin heavy variable 3-72' |
| 189 | 'IGHV3-72_No7' | 'LSCAASGFTFSDHYMDWVR' | 'Immunoglobulin heavy variable 3-72' |
| 190 | 'IGHV3-72_No8' | 'ANSYTTEYAASVK' | 'Immunoglobulin heavy variable 3-72' |
| 191 | 'IGHV3-72_No9' | 'DDSKNSLYLQMNSLK' | 'Immunoglobulin heavy variable 3-72' |
| 192 | 'IGHV3-72_No10' | 'NSLYLQMNSLKTEDTAVYYCAR' | 'Immunoglobulin heavy variable 3-72' |
| 193 | 'IGHV3-72_No11' | 'NKANSYTTEYAASVK' | 'Immunoglobulin heavy variable 3-72' |
| 194 | 'IGHV3-72_No12' | 'DDSKNSLYLQMNSLK' | 'Immunoglobulin heavy variable 3-72' |
| 195 | 'IGHV3-72_No13' | 'NKANSYTTEYAASVKGR' | 'Immunoglobulin heavy variable 3-72' |
| 196 | 'IGHV3-72_No14' | 'ANSYTTEYAASVKGR' | 'Immunoglobulin heavy variable 3-72' |
| 197 | 'IGHV3-72_No15' | 'NSLYLQMNSLKTEDTAVYYCAR' | 'Immunoglobulin heavy variable 3-72' |
| 198 | 'IGKV1D-43_No1' | 'MTQSPFSLSASVGDR' | 'Immunoglobulin kappa variable 1D-43' |
| 199 | 'IGKV1D-43_No2' | 'MTQSPFSLSASVGDR' | 'Immunoglobulin kappa variable 1D-43' |
| 200 | 'IGKV1D-13_No1' | 'LLIYDASSLESGVPSR' | 'Immunoglobulin kappa variable 1D-13' |
| 201 | 'IGKV1D-13_No2' | 'APKLLIYDASSLESGVPSR' | 'Immunoglobulin kappa variable 1D-13' |
| 202 | 'IGKV1D-13_No3' | 'ASQGISSALAWYQQKPGK' | 'Immunoglobulin kappa variable 1D-13' |
| 203 | 'IGKV1D-13_No4' | 'VTITCRASQGISSALAWYQQKPGK' | 'Immunoglobulin kappa variable 1D-13' |
| 204 | 'IGHV1-69D_No1' | 'VTITADESTSTAYMELSSLR' | 'Immunoglobulin heavy variable 1-69D' |
| 205 | 'IGHV1-69D_No2' | 'VTITADESTSTAYMELSSLR' | 'Immunoglobulin heavy variable 1-69D' |
| 206 | 'IGHV1-69D_No3' | 'VTITADESTSTAYMELSSLRSEDTAVYYCAR' | 'Immunoglobulin heavy variable 1-69D' |
| 207 | 'IGHV1-69D_No4' | 'VTITADESTSTAYMELSSLRSEDTAVYYCAR' | 'Immunoglobulin heavy variable 1-69D' |
| 208 | 'IGKV6-21_No1' | 'YASQSFSGVPSR' | 'Immunoglobulin kappa variable 6-21' |
| 209 | 'IGKV6-21_No2' | 'ASQSIGSSLHWYQQKPDQSPK' | 'Immunoglobulin kappa variable 6-21' |
| 210 | 'IGKV6-21_No3' | 'ASQSIGSSLHWYQQK' | 'Immunoglobulin kappa variable 6-21' |
| 211 | 'IGKV6-21_No4' | 'EKVTITCR' | 'Immunoglobulin kappa variable 6-21' |
| 212 | 'IGKV3D-20_No1' | 'LLIYDASSR' | 'Immunoglobulin kappa variable 3D-20' |
| 213 | 'IGKV3D-20_No2' | 'ATLSCGASQSVSSSYLAWYQQKPGLAPR' | 'Immunoglobulin kappa variable 3D-20' |
| 214 | 'IGHV1-3_No1' | 'VTITRDTSASTAYMELSSLR' | 'Immunoglobulin heavy variable 1-3' |
| 215 | 'IGHV1-3_No2' | 'ASGYTFTSYAMHWVR' | 'Immunoglobulin heavy variable 1-3' |
| 216 | 'IGHV1-3_No3' | 'VTITRDTSASTAYMELSSLR' | 'Immunoglobulin heavy variable 1-3' |
| 217 | 'IGHV1-3_No4' | 'DTSASTAYMELSSLR' | 'Immunoglobulin heavy variable 1-3' |
| 218 | 'IGHV1-3_No5' | 'DTSASTAYMELSSLR' | 'Immunoglobulin heavy variable 1-3' |
| 219 | 'IGHV1-3_No6' | 'LEWMGWINAGNGNTK' | 'Immunoglobulin heavy variable 1-3' |
| 220 | 'IGHV1-3_No7' | 'ASGYTFTSYAMHWVR' | 'Immunoglobulin heavy variable 1-3' |
| 221 | 'IGHV1-3_No8' | 'LEWMGWINAGNGNTK' | 'Immunoglobulin heavy variable 1-3' |

| | A | B | C |
|---|---|---|---|
| 1 | **Peptide Name** | **Peptide Sequence** | **Peptide Description** |
| 222 | 'IGHV1-3_No9' | 'DTSASTAYMELSSLRSEDTAVYYCAR' | 'Immunoglobulin heavy variable 1-3' |
| 223 | 'IGHV1-3_No10' | 'DTSASTAYMELSSLRSEDTAVYYCAR' | 'Immunoglobulin heavy variable 1-3' |
| 224 | 'IGHV3-16_No1' | 'LSCAASGFTFSNSDMNWARK' | 'Probable non-functional immunoglobulin heavy variable 3-16' |
| 225 | 'IGHV3-16_No2' | 'LSCAASGFTFSNSDMNWARK' | 'Probable non-functional immunoglobulin heavy variable 3-16' |
| 226 | 'IGHV3-16_No3' | 'LSCAASGFTFSNSDMNWAR' | 'Probable non-functional immunoglobulin heavy variable 3-16' |
| 227 | 'IGHV1-18_No1' | 'VTMTTDTSTSTAYMELR' | 'Immunoglobulin heavy variable 1-18' |
| 228 | 'IGHV1-18_No2' | 'VTMTTDTSTSTAYMELR' | 'Immunoglobulin heavy variable 1-18' |
| 229 | 'IGHV1-18_No3' | 'SLRSDDTAVYYCAR' | 'Immunoglobulin heavy variable 1-18' |
| 230 | 'IGHV1-18_No4' | 'VTMTTDTSTSTAYMELR' | 'Immunoglobulin heavy variable 1-18' |
| 231 | 'IGHV1-18_No5' | 'ASGYTFTSYGISWVR' | 'Immunoglobulin heavy variable 1-18' |
| 232 | 'IGHV1-18_No6' | 'QAPGQGLEWMGWISAYNGNTNYAQK' | 'Immunoglobulin heavy variable 1-18' |
| 233 | 'IGHV1-18_No7' | 'VTMTTDTSTSTAYMELR' | 'Immunoglobulin heavy variable 1-18' |
| 234 | 'IGHV3-20_No1' | 'LSCAASGFTFDDYGMSWVR' | 'Immunoglobulin heavy variable 3-20' |
| 235 | 'IGHV3-20_No2' | 'AEDTALYHCAR' | 'Immunoglobulin heavy variable 3-20' |
| 236 | 'IGHV3-20_No3' | 'GLEWVSGINWNGGSTGYADSVK' | 'Immunoglobulin heavy variable 3-20' |
| 237 | 'IGHV3-20_No4' | 'LSCAASGFTFDDYGMSWVR' | 'Immunoglobulin heavy variable 3-20' |
| 238 | 'IGHV3-20_No5' | 'NSLYLQMNSLRAEDTALYHCAR' | 'Immunoglobulin heavy variable 3-20' |
| 239 | 'IGHV1-24_No1' | 'VSGYTLTELSMHWVR' | 'Immunoglobulin heavy variable 1-24' |
| 240 | 'IGHV1-24_No2' | 'VTMTEDTSTDTAYMELSSLR' | 'Immunoglobulin heavy variable 1-24' |
| 241 | 'IGHV1-24_No3' | 'VTMTEDTSTDTAYMELSSLR' | 'Immunoglobulin heavy variable 1-24' |
| 242 | 'IGHV1-24_No4' | 'GLEWMGGFDPEDGETIYAQK' | 'Immunoglobulin heavy variable 1-24' |
| 243 | 'IGHV1-24_No5' | 'GLEWMGGFDPEDGETIYAQK' | 'Immunoglobulin heavy variable 1-24' |
| 244 | 'IGHV1-24_No6' | 'VTMTEDTSTDTAYMELSSLR' | 'Immunoglobulin heavy variable 1-24' |
| 245 | 'IGHV1-24_No7' | 'VTMTEDTSTDTAYMELSSLR' | 'Immunoglobulin heavy variable 1-24' |
| 246 | 'IGHV1-24_No8' | 'VSGYTLTELSMHWVR' | 'Immunoglobulin heavy variable 1-24' |
| 247 | 'IGHV1-24_No9' | 'GLEWMGGFDPEDGETIYAQKFQGR' | 'Immunoglobulin heavy variable 1-24' |
| 248 | 'IGHV1-24_No10' | 'QAPGKGLEWMGGFDPEDGETIYAQK' | 'Immunoglobulin heavy variable 1-24' |
| 249 | 'IGHV4-28_No1' | 'VTMSVDTSKNQFSLK' | 'Immunoglobulin heavy variable 4-28' |
| 250 | 'IGHV4-28_No2' | 'VTMSVDTSKNQFSLK' | 'Immunoglobulin heavy variable 4-28' |
| 251 | 'IGHV4-28_No3' | 'LSSVTAVDTAVYYCAR' | 'Immunoglobulin heavy variable 4-28' |
| 252 | 'IGHV4-28_No4' | 'VTMSVDTSK' | 'Immunoglobulin heavy variable 4-28' |
| 253 | 'IGHV4-28_No5' | 'VTMSVDTSK' | 'Immunoglobulin heavy variable 4-28' |
| 254 | 'IGHV4-28_No6' | 'SRVTMSVDTSKNQFSLK' | 'Immunoglobulin heavy variable 4-28' |
| 255 | 'IGHV4-28_No7' | 'SRVTMSVDTSKNQFSLK' | 'Immunoglobulin heavy variable 4-28' |
| 256 | 'IGHV4-28_No8' | 'SRVTMSVDTSK' | 'Immunoglobulin heavy variable 4-28' |
| 257 | 'IGHV3-35_No1' | 'AEDTAVYYCVR' | 'Probable non-functional immunoglobulin heavy variable 3-35' |
| 258 | 'IGHV3-35_No2' | 'NTLYLQTNSLRAEDTAVYYCVR' | 'Probable non-functional immunoglobulin heavy variable 3-35' |
| 259 | 'IGHV3-35_No3' | 'NTLYLQTNSLR' | 'Probable non-functional immunoglobulin heavy variable 3-35' |
| 260 | 'IGHV3-35_No4' | 'THYADSVK' | 'Probable non-functional immunoglobulin heavy variable 3-35' |
| 261 | 'IGHV3-38_No1' | 'NTLYLQMNNLR' | 'Probable non-functional immunoglobulin heavy variable 3-38' |
| 262 | 'IGHV3-38_No2' | 'NTLYLQMNNLR' | 'Probable non-functional immunoglobulin heavy variable 3-38' |
| 263 | 'IGHV3-38_No3' | 'AEGTAVYYCAR' | 'Probable non-functional immunoglobulin heavy variable 3-38' |
| 264 | 'IGHV3-38_No4' | 'LSCAASGFTVSSNEMSWIR' | 'Probable non-functional immunoglobulin heavy variable 3-38' |
| 265 | 'IGHV3-38_No5' | 'DNSKNTLYLQMNNLR' | 'Probable non-functional immunoglobulin heavy variable 3-38' |

| | A | B | C |
|---|---|---|---|
| 1 | **Peptide Name** | **Peptide Sequence** | **Peptide Description** |
| 266 | 'IGHV3-38_No6' | 'LSCAASGFTVSSNEMSWIR' | 'Probable non-functional immunoglobulin heavy variable 3-38' |
| 267 | 'IGHV3-38_No7' | 'DNSKNTLYLQMNNLR' | 'Probable non-functional immunoglobulin heavy variable 3-38' |
| 268 | 'IGHV5-51_No1' | 'YSPSFQGQVTISADK' | 'Immunoglobulin heavy variable 5-51' |
| 269 | 'IGHV5-51_No2' | 'SISTAYLQWSSLK' | 'Immunoglobulin heavy variable 5-51' |
| 270 | 'IGHV5-51_No3' | 'GLEWMGIIYPGDSDTR' | 'Immunoglobulin heavy variable 5-51' |
| 271 | 'IGHV5-51_No4' | 'ASDTAMYYCAR' | 'Immunoglobulin heavy variable 5-51' |
| 272 | 'IGHV5-51_No5' | 'GLEWMGIIYPGDSDTR' | 'Immunoglobulin heavy variable 5-51' |
| 273 | 'IGHV5-51_No6' | 'ASDTAMYYCAR' | 'Immunoglobulin heavy variable 5-51' |
| 274 | 'IGHV5-51_No7' | 'SISTAYLQWSSLKASDTAMYYCAR' | 'Immunoglobulin heavy variable 5-51' |
| 275 | 'IGHV5-51_No8' | 'QMPGKGLEWMGIIYPGDSDTR' | 'Immunoglobulin heavy variable 5-51' |
| 276 | 'IGHV5-51_No9' | 'QMPGKGLEWMGIIYPGDSDTR' | 'Immunoglobulin heavy variable 5-51' |
| 277 | 'IGHV5-51_No10' | 'QMPGKGLEWMGIIYPGDSDTR' | 'Immunoglobulin heavy variable 5-51' |
| 278 | 'IGHV5-51_No11' | 'QMPGKGLEWMGIIYPGDSDTR' | 'Immunoglobulin heavy variable 5-51' |
| 279 | 'IGHV5-51_No12' | 'QMPGKGLEWMGIIYPGDSDTR' | 'Immunoglobulin heavy variable 5-51' |
| 280 | 'IGHV1-58_No1' | 'ASGFTFTSSAVQWVR' | 'Immunoglobulin heavy variable 1-58' |
| 281 | 'IGHV1-58_No2' | 'DMSTSTAYMELSSLR' | 'Immunoglobulin heavy variable 1-58' |
| 282 | 'IGHV1-58_No3' | 'DMSTSTAYMELSSLR' | 'Immunoglobulin heavy variable 1-58' |
| 283 | 'IGHV1-58_No4' | 'DMSTSTAYMELSSLR' | 'Immunoglobulin heavy variable 1-58' |
| 284 | 'IGHV2-70D_No1' | 'IDWDDDKFYSTSLK' | 'Immunoglobulin heavy variable 2-70D' |
| 285 | 'IGHV2-70D_No2' | 'DTSKNQVVLTMTNMDPVDTATYYCAR' | 'Immunoglobulin heavy variable 2-70D' |
| 286 | 'IGHV2-70D_No3' | 'ALEWLAR' | 'Immunoglobulin heavy variable 2-70D' |
| 287 | 'IGHV2-70D_No4' | 'ESGPALVKPTQTLTLTCTFSGFSLSTSGMR' | 'Immunoglobulin heavy variable 2-70D' |
| 288 | 'IGHV2-70D_No5' | 'NQVVLTMTNMDPVDTATYYCAR' | 'Immunoglobulin heavy variable 2-70D' |
| 289 | 'IGHV2-70D_No6' | 'ESGPALVKPTQTLTLTCTFSGFSLSTSGMR' | 'Immunoglobulin heavy variable 2-70D' |
| 290 | 'IGHV2-70D_No7' | 'IDWDDDKFYSTSLKTR' | 'Immunoglobulin heavy variable 2-70D' |
| 291 | 'IGHV2-70D_No8' | 'DTSKNQVVLTMTNMDPVDTATYYCAR' | 'Immunoglobulin heavy variable 2-70D' |
| 292 | 'IGHV2-70D_No9' | 'VSWIRQPPGK' | 'Immunoglobulin heavy variable 2-70D' |
| 293 | 'IGHV2-70D_No10' | 'LTISKDTSK' | 'Immunoglobulin heavy variable 2-70D' |
| 294 | 'IGKV3D-7_No1' | 'ASQSVSSSYLSWYQQKPGQAPR' | 'Immunoglobulin kappa variable 3D-7' |
| 295 | 'IGKV3D-7_No2' | 'ASQSVSSSYLSWYQQK' | 'Immunoglobulin kappa variable 3D-7' |
| 296 | 'IGKV1-8_No1' | 'ASQGISSYLAWYQQKPGK' | 'Immunoglobulin kappa variable 1-8' |
| 297 | 'IGKV1-8_No2' | 'LLIYAASTLQSGVPSR' | 'Immunoglobulin kappa variable 1-8' |
| 298 | 'IGKV1-8_No3' | 'MTQSPSSFSASTGDR' | 'Immunoglobulin kappa variable 1-8' |
| 299 | 'IGKV1-8_No4' | 'MTQSPSSFSASTGDR' | 'Immunoglobulin kappa variable 1-8' |
| 300 | 'IGKV1-8_No5' | 'ASQGISSYLAWYQQK' | 'Immunoglobulin kappa variable 1-8' |
| 301 | 'IGKV1-8_No6' | 'ASQGISSYLAWYQQKPGKAPK' | 'Immunoglobulin kappa variable 1-8' |
| 302 | 'IGKV1-8_No7' | 'APKLLIYAASTLQSGVPSR' | 'Immunoglobulin kappa variable 1-8' |
| 303 | 'IGKV1-8_No8' | 'MTQSPSSFSASTGDRVTITCR' | 'Immunoglobulin kappa variable 1-8' |
| 304 | 'IGKV1-8_No9' | 'MTQSPSSFSASTGDRVTITCR' | 'Immunoglobulin kappa variable 1-8' |
| 305 | 'IGKV2-24_No1' | 'FSGSGAGTDFTLK' | 'Immunoglobulin kappa variable 2-24' |
| 306 | 'IGKV2-24_No2' | 'SSQSLVHSDGNTYLSWLQQR' | 'Immunoglobulin kappa variable 2-24' |
| 307 | 'IGKV2-24_No3' | 'SSQSLVHSDGNTYLSWLQQRPGQPPR' | 'Immunoglobulin kappa variable 2-24' |
| 308 | 'IGKV2-24_No4' | 'FSGVPDRFSGSGAGTDFTLK' | 'Immunoglobulin kappa variable 2-24' |
| 309 | 'IGKV2-24_No5' | 'ISNRFSGVPDR' | 'Immunoglobulin kappa variable 2-24' |

| | A | B | C |
|---|---|---|---|
| 1 | **Peptide Name** | **Peptide Sequence** | **Peptide Description** |
| 310 | 'IGKV2-24_No6' | 'FSGSGAGTDFTLKISR' | 'Immunoglobulin kappa variable 2-24' |
| 311 | 'IGKV1-6_No1' | 'NDLGWYQQK' | 'Immunoglobulin kappa variable 1-6' |
| 312 | 'IGKV1-6_No2' | 'NDLGWYQQKPGK' | 'Immunoglobulin kappa variable 1-6' |
| 313 | 'IGKV1-6_No3' | 'NDLGWYQQKPGKAPK' | 'Immunoglobulin kappa variable 1-6' |
| 314 | 'IGKV1-6_No4' | 'LLIYAASSLQSGVPSR' | 'Immunoglobulin kappa variable 1-6' |
| 315 | 'IGKV1-6_No5' | 'APKLLIYAASSLQSGVPSR' | 'Immunoglobulin kappa variable 1-6' |
| 316 | 'IGKV1-6_No6' | 'ASQGIRNDLGWYQQKPGK' | 'Immunoglobulin kappa variable 1-6' |
| 317 | 'IGKV1-6_No7' | 'CAIQMTQSPSSLSASVGDR' | 'Immunoglobulin kappa variable 1-6' |
| 318 | 'IGKV1-6_No8' | 'CAIQMTQSPSSLSASVGDR' | 'Immunoglobulin kappa variable 1-6' |
| 319 | 'IGKV1-6_No9' | 'ASQGIRNDLGWYQQK' | 'Immunoglobulin kappa variable 1-6' |
| 320 | 'IGKV1-12_No1' | 'ASQGISSWLAWYQQKPGK' | 'Immunoglobulin kappa variable 1-12' |
| 321 | 'IGKV1-12_No2' | 'ASQGISSWLAWYQQK' | 'Immunoglobulin kappa variable 1-12' |
| 322 | 'IGKV1-12_No3' | 'VTITCRASQGISSWLAWYQQKPGK' | 'Immunoglobulin kappa variable 1-12' |
| 323 | 'IGKV1-12_No4' | 'CDIQMTQSPSSVSASVGDR' | 'Immunoglobulin kappa variable 1-12' |
| 324 | 'IGKV1-12_No5' | 'CDIQMTQSPSSVSASVGDR' | 'Immunoglobulin kappa variable 1-12' |
| 325 | 'IGHV1-69-2_No1' | 'VTITADTSTDTAYMELSSLR' | 'Immunoglobulin heavy variable 1-69-2' |
| 326 | 'IGHV1-69-2_No2' | 'VTITADTSTDTAYMELSSLR' | 'Immunoglobulin heavy variable 1-69-2' |
| 327 | 'IGHV1-69-2_No3' | 'GLEWMGLVDPEDGETIYAEK' | 'Immunoglobulin heavy variable 1-69-2' |
| 328 | 'IGHV1-69-2_No4' | 'VSGYTFTDYYMHWVQQAPGK' | 'Immunoglobulin heavy variable 1-69-2' |
| 329 | 'IGHV1-69-2_No5' | 'VSGYTFTDYYMHWVQQAPGK' | 'Immunoglobulin heavy variable 1-69-2' |
| 330 | 'IGHV1-69-2_No6' | 'GLEWMGLVDPEDGETIYAEK' | 'Immunoglobulin heavy variable 1-69-2' |
| 331 | 'IGHV1-69-2_No7' | 'GLEWMGLVDPEDGETIYAEKFQGR' | 'Immunoglobulin heavy variable 1-69-2' |
| 332 | 'IGLV5-39_No1' | 'SGINVGTYR' | 'Immunoglobulin lambda variable 5-39' |
| 333 | 'IGLV5-39_No2' | 'IYWYQQKPGSLPR' | 'Immunoglobulin lambda variable 5-39' |
| 334 | 'IGLV5-39_No3' | 'YKSDSDKQQGSGVPSR' | 'Immunoglobulin lambda variable 5-39' |
| 335 | 'IGLV5-39_No4' | 'IYWYQQK' | 'Immunoglobulin lambda variable 5-39' |
| 336 | 'IGLV5-39_No5' | 'SDSDKQQGSGVPSR' | 'Immunoglobulin lambda variable 5-39' |
| 337 | 'IGHV7-4-1_No1' | 'ASGYTFTSYAMNWVR' | 'Immunoglobulin heavy variable 7-4-1' |
| 338 | 'IGHV7-4-1_No2' | 'QAPGQGLEWMGWINTNTGNPTYAQGFTGR' | 'Immunoglobulin heavy variable 7-4-1' |
| 339 | 'IGHV7-4-1_No3' | 'ASGYTFTSYAMNWVR' | 'Immunoglobulin heavy variable 7-4-1' |
| 340 | 'IGHV7-4-1_No4' | 'QAPGQGLEWMGWINTNTGNPTYAQGFTGR' | 'Immunoglobulin heavy variable 7-4-1' |
| 341 | 'IGHV3-64D_No1' | 'AEDTAVYYCVK' | 'Immunoglobulin heavy variable 3-64D' |
| 342 | 'IGHV3-64D_No2' | 'NTLYLQMSSLR' | 'Immunoglobulin heavy variable 3-64D' |
| 343 | 'IGHV3-64D_No3' | 'LSCSASGFTFSSYAMHWVR' | 'Immunoglobulin heavy variable 3-64D' |
| 344 | 'IGHV3-64D_No4' | 'LSCSASGFTFSSYAMHWVR' | 'Immunoglobulin heavy variable 3-64D' |
| 345 | 'IGHV3-64D_No5' | 'NTLYLQMSSLR' | 'Immunoglobulin heavy variable 3-64D' |
| 346 | 'IGHV3-64D_No6' | 'DNSKNTLYLQMSSLR' | 'Immunoglobulin heavy variable 3-64D' |
| 347 | 'IGHV3-64D_No7' | 'DNSKNTLYLQMSSLR' | 'Immunoglobulin heavy variable 3-64D' |
| 348 | 'IGHV3-64D_No8' | 'NTLYLQMSSLRAEDTAVYYCVK' | 'Immunoglobulin heavy variable 3-64D' |
| 349 | 'IGHV5-10-1_No1' | 'GLEWMGR' | 'Immunoglobulin heavy variable 5-10-1' |
| 350 | 'IGHV5-10-1_No2' | 'IDPSDSYTNYSPSFQGHVTISADK' | 'Immunoglobulin heavy variable 5-10-1' |
| 351 | 'IGHV5-10-1_No3' | 'QMPGKGLEWMGR' | 'Immunoglobulin heavy variable 5-10-1' |
| 352 | 'IGHV5-10-1_No4' | 'QMPGKGLEWMGR' | 'Immunoglobulin heavy variable 5-10-1' |
| 353 | 'IGLC7_No1' | 'ADGSPVKVGVETTKPSK' | 'Immunoglobulin lambda constant 7' |

| | A | B | C |
|---|---|---|---|
| 1 | **Peptide Name** | **Peptide Sequence** | **Peptide Description** |
| 354 | 'IGLC7_No2' | 'VTHEGSTVEK' | 'Immunoglobulin lambda constant 7' |
| 355 | 'IGLC7_No3' | 'VTHEGSTVEKTVAPAECS' | 'Immunoglobulin lambda constant 7' |
| 356 | 'IGLC7_No4' | 'SYSCRVTHEGSTVEK' | 'Immunoglobulin lambda constant 7' |
| 357 | 'IGLC7_No5' | 'VGVETTKPSK' | 'Immunoglobulin lambda constant 7' |
| 358 | 'SSC5D_No1' | 'SNCDHSEDAGLVCTGPAPR' | 'Soluble scavenger receptor cysteine-rich domain-containing protein SSC5D' |
| 359 | 'SSC5D_No2' | 'AHICSHEEDAGVVCAGQR' | 'Soluble scavenger receptor cysteine-rich domain-containing protein SSC5D' |
| 360 | 'SSC5D_No3' | 'ELGCGGPQQPDPAAGR' | 'Soluble scavenger receptor cysteine-rich domain-containing protein SSC5D' |
| 361 | 'SSC5D_No4' | 'WGTVCDDGWDLRDAAVACR' | 'Soluble scavenger receptor cysteine-rich domain-containing protein SSC5D' |
| 362 | 'SSC5D_No5' | 'WGTVCDDGWDLR' | 'Soluble scavenger receptor cysteine-rich domain-containing protein SSC5D' |
| 363 | 'SSC5D_No6' | 'GSEASLSDCPSGAWGK' | 'Soluble scavenger receptor cysteine-rich domain-containing protein SSC5D' |
| 364 | 'SSC5D_No7' | 'QALLLGLTQLVEAAR' | 'Soluble scavenger receptor cysteine-rich domain-containing protein SSC5D' |
| 365 | 'SSC5D_No8' | 'WGSVCDDAWDLR' | 'Soluble scavenger receptor cysteine-rich domain-containing protein SSC5D' |
| 366 | 'IGKV2-29_No1' | 'FSGSGSGTDFTLK' | 'Immunoglobulin kappa variable 2-29' |
| 367 | 'IGKV2-29_No2' | 'TYLYWYLQK' | 'Immunoglobulin kappa variable 2-29' |
| 368 | 'IGKV2-29_No3' | 'FSGSGSGTDFTLKISR' | 'Immunoglobulin kappa variable 2-29' |
| 369 | 'IGKV2-29_No4' | 'TYLYWYLQKPGQSPQLLIYEVSSR' | 'Immunoglobulin kappa variable 2-29' |
| 370 | 'IGKV2-29_No5' | 'PGQSPQLLIYEVSSR' | 'Immunoglobulin kappa variable 2-29' |
| 371 | 'IGKV2-29_No6' | 'FSGVPDRFSGSGSGTDFTLK' | 'Immunoglobulin kappa variable 2-29' |
| 372 | 'IGKV2-29_No7' | 'SSQSLLHSDGK' | 'Immunoglobulin kappa variable 2-29' |
| 373 | 'FITM1' | 'IQALLGCLLK' | 'Fat storage-inducing transmembrane protein 1' |
| 374 | 'GRXCR2' | 'IIIYTNNLK' | 'Glutaredoxin domain-containing cysteine-rich protein 2' |
| 375 | 'KIF2A_No1' | 'IDILTELR' | 'Kinesin-like protein KIF2A' |
| 376 | 'KIF2A_No2' | 'GSLDYRPLTTADPIDEHR' | 'Kinesin-like protein KIF2A' |
| 377 | 'PDLIM1_No1' | 'VTPPEGYEVVTVFPK' | 'PDZ and LIM domain protein 1' |
| 378 | 'PDLIM1_No2' | 'GHFFVEDQIYCEK' | 'PDZ and LIM domain protein 1' |
| 379 | 'PDLIM1_No3' | 'MNLASEPQEVLHIGSAHNR' | 'PDZ and LIM domain protein 1' |
| 380 | 'PDLIM1_No4' | 'SAMPFTASPASSTTAR' | 'PDZ and LIM domain protein 1' |
| 381 | 'PDLIM1_No5' | 'HRHPECYVCTDCGTNLK' | 'PDZ and LIM domain protein 1' |
| 382 | 'PDLIM1_No6' | 'LVGGKDFEQPLAISR' | 'PDZ and LIM domain protein 1' |
| 383 | 'PDLIM1_No7' | 'VWSPLVTEEGKR' | 'PDZ and LIM domain protein 1' |
| 384 | 'PDLIM1_No8' | 'TTQQIDLQGPGPWGFR' | 'PDZ and LIM domain protein 1' |
| 385 | 'PDLIM1_No9' | 'SAMPFTASPASSTTAR' | 'PDZ and LIM domain protein 1' |
| 386 | 'PDLIM1_No10' | 'QSTSFLVLQEILESEEKGDPNKPSGFR' | 'PDZ and LIM domain protein 1' |
| 387 | 'PDLIM1_No11' | 'AALANLCIGDVITAIDGENTSNMTHLEAQNR' | 'PDZ and LIM domain protein 1' |
| 388 | 'PDLIM1_No12' | 'HPECYVCTDCGTNLK' | 'PDZ and LIM domain protein 1' |
| 389 | 'PDLIM1_No13' | 'CGTGIVGVFVK' | 'PDZ and LIM domain protein 1' |
| 390 | 'PDLIM1_No14' | 'MNLASEPQEVLHIGSAHNR' | 'PDZ and LIM domain protein 1' |
| 391 | 'PDLIM1_No15' | 'IKGCTDNLTLTVAR' | 'PDZ and LIM domain protein 1' |
| 392 | 'PDLIM1_No16' | 'GCTDNLTLTVAR' | 'PDZ and LIM domain protein 1' |
| 393 | 'PDLIM1_No17' | 'ERVTPPEGYEVVTVFPK' | 'PDZ and LIM domain protein 1' |
| 394 | 'PDLIM1_No18' | 'TAASGVEANSRPLDHAQPPSSLVIDKESEVYK' | 'PDZ and LIM domain protein 1' |
| 395 | 'PDLIM1_No19' | 'SEHKVWSPLVTEEGKR' | 'PDZ and LIM domain protein 1' |
| 396 | 'PDLIM1_No20' | 'VAASIGNAQKLPMCDK' | 'PDZ and LIM domain protein 1' |
| 397 | 'PDLIM1_No21' | 'AALANLCIGDVITAIDGENTSNMTHLEAQNR' | 'PDZ and LIM domain protein 1' |

| | A | B | C |
|---|---|---|---|
| 1 | **Peptide Name** | **Peptide Sequence** | **Peptide Description** |
| 398 | 'SNAP23_No1' | 'TTWGDGGENSPCNVVSK' | 'Synaptosomal-associated protein 23' |
| 399 | 'SNAP23_No2' | 'ILGLAIESQDAGIK' | 'Synaptosomal-associated protein 23' |
| 400 | 'SNAP23_No3' | 'TITMLDEQKEQLNR' | 'Synaptosomal-associated protein 23' |
| 401 | 'SNAP23_No4' | 'DMALNIGNEIDAQNPQIK' | 'Synaptosomal-associated protein 23' |
| 402 | 'SNAP23_No5' | 'IEEGLDQINK' | 'Synaptosomal-associated protein 23' |
| 403 | 'SNAP23_No6' | 'TITMLDEQKEQLNR' | 'Synaptosomal-associated protein 23' |
| 404 | 'MASP2_No1' | 'LASPGFPGEYANDQER' | 'Mannan-binding lectin serine protease 2' |
| 405 | 'MASP2_No2' | 'WPEPVFGR' | 'Mannan-binding lectin serine protease 2' |
| 406 | 'MASP2_No3' | 'TCSALCSGQVFTQR' | 'Mannan-binding lectin serine protease 2' |
| 407 | 'MASP2_No4' | 'APGKDTFYSLGSSLDITFR' | 'Mannan-binding lectin serine protease 2' |
| 408 | 'MASP2_No5' | 'SNTVTITFVTDESGDHTGWK' | 'Mannan-binding lectin serine protease 2' |
| 409 | 'MASP2_No6' | 'NLMYVDIPIVDHQK' | 'Mannan-binding lectin serine protease 2' |
| 410 | 'MASP2_No7' | 'NLMYVDIPIVDHQK' | 'Mannan-binding lectin serine protease 2' |
| 411 | 'MASP2_No8' | 'TDDIGTASGWGLTQR' | 'Mannan-binding lectin serine protease 2' |
| 412 | 'MASP2_No9' | 'LASPGFPGEYANDQERR' | 'Mannan-binding lectin serine protease 2' |
| 413 | 'MASP2_No10' | 'YVCEADGFWTSSK' | 'Mannan-binding lectin serine protease 2' |
| 414 | 'MASP2_No11' | 'GDSGGALVFLDSETER' | 'Mannan-binding lectin serine protease 2' |
| 415 | 'MASP2_No12' | 'VLATLCGQESTDTER' | 'Mannan-binding lectin serine protease 2' |
| 416 | 'MASP2_No13' | 'RWTLTAPPGYR' | 'Mannan-binding lectin serine protease 2' |
| 417 | 'MASP2_No14' | 'GSVTANMLCAGLESGGK' | 'Mannan-binding lectin serine protease 2' |
| 418 | 'MASP2_No15' | 'WTLTAPPGYR' | 'Mannan-binding lectin serine protease 2' |
| 419 | 'MASP2_No16' | 'AGYVLHR' | 'Mannan-binding lectin serine protease 2' |
| 420 | 'MASP2_No17' | 'CTAAYEKPPYPR' | 'Mannan-binding lectin serine protease 2' |
| 421 | 'MASP2_No18' | 'VLATLCGQESTDTERAPGK' | 'Mannan-binding lectin serine protease 2' |
| 422 | 'MASP2_No19' | 'VNDGKYVCEADGFWTSSKGEK' | 'Mannan-binding lectin serine protease 2' |
| 423 | 'RAB27B_No1' | 'ADLPDQREVNER' | 'Ras-related protein Rab-27B' |
| 424 | 'RAB27B_No2' | 'AVETLLDLIMKR' | 'Ras-related protein Rab-27B' |
| 425 | 'RAB27B_No3' | 'AVETLLDLIMK' | 'Ras-related protein Rab-27B' |
| 426 | 'RAB27B_No4' | 'VHLQLWDTAGQER' | 'Ras-related protein Rab-27B' |
| 427 | 'CLIC1_No1' | 'LAALNPESNTAGLDIFAK' | 'Chloride intracellular channel protein 1' |
| 428 | 'CLIC1_No2' | 'NSNPALNDNLEK' | 'Chloride intracellular channel protein 1' |
| 429 | 'CLIC1_No3' | 'YLSNAYAR' | 'Chloride intracellular channel protein 1' |
| 430 | 'CLIC1_No4' | 'FLDGNELTLADCNLLPK' | 'Chloride intracellular channel protein 1' |
| 431 | 'CLIC1_No5' | 'GFTIPEAFR' | 'Chloride intracellular channel protein 1' |
| 432 | 'CLIC1_No6' | 'KFLDGNELTLADCNLLPK' | 'Chloride intracellular channel protein 1' |
| 433 | 'CLIC1_No7' | 'AEEQPQVELFVK' | 'Chloride intracellular channel protein 1' |
| 434 | 'CLIC1_No8' | 'VLDNYLTSPLPEEVDETSAEDEGVSQRK' | 'Chloride intracellular channel protein 1' |
| 435 | 'QSOX1_No1' | 'AHFSPSNIILDFPAAGSAAR' | 'Sulfhydryl oxidase 1' |
| 436 | 'QSOX1_No2' | 'GYVHYFFGCR' | 'Sulfhydryl oxidase 1' |
| 437 | 'QSOX1_No3' | 'DFNIPGFPTVR' | 'Sulfhydryl oxidase 1' |
| 438 | 'QSOX1_No4' | 'NNEEYLALIFEK' | 'Sulfhydryl oxidase 1' |
| 439 | 'QSOX1_No5' | 'FPVLEGQR' | 'Sulfhydryl oxidase 1' |
| 440 | 'QSOX1_No6' | 'AWRPALYLAALDCAEETNSAVCR' | 'Sulfhydryl oxidase 1' |
| 441 | 'QSOX1_No7' | 'DCASHFEQMAAASMHR' | 'Sulfhydryl oxidase 1' |

| | A | B | C |
|---|---|---|---|
| 1 | **Peptide Name** | **Peptide Sequence** | **Peptide Description** |
| 442 | 'QSOX1_No8' | 'AAPGQEPPEHMAELQR' | 'Sulfhydryl oxidase 1' |
| 443 | 'QSOX1_No9' | 'LAGAPSEDPQFPK' | 'Sulfhydryl oxidase 1' |
| 444 | 'QSOX1_No10' | 'EVALDLSQHK' | 'Sulfhydryl oxidase 1' |
| 445 | 'QSOX1_No11' | 'VPVLMESR' | 'Sulfhydryl oxidase 1' |
| 446 | 'QSOX1_No12' | 'SALYSPSDPLTLLQADTVR' | 'Sulfhydryl oxidase 1' |
| 447 | 'QSOX1_No13' | 'VNWIGCQGSEPHFR' | 'Sulfhydryl oxidase 1' |
| 448 | 'QSOX1_No14' | 'KVNWIGCQGSEPHFR' | 'Sulfhydryl oxidase 1' |
| 449 | 'QSOX1_No15' | 'VGSPNAAVLWLWSSHNR' | 'Sulfhydryl oxidase 1' |
| 450 | 'QSOX1_No16' | 'IEVGRFPVLEGQR' | 'Sulfhydryl oxidase 1' |
| 451 | 'QSOX1_No17' | 'KFGVTDFPSCYLLFR' | 'Sulfhydryl oxidase 1' |
| 452 | 'QSOX1_No18' | 'VLNTEANVVR' | 'Sulfhydryl oxidase 1' |
| 453 | 'QSOX1_No19' | 'LIDALESHHDTWPPACPPLEPAKLEEIDGFFAR' | 'Sulfhydryl oxidase 1' |
| 454 | 'QSOX1_No20' | 'PLVQNFLHSVNEWLK' | 'Sulfhydryl oxidase 1' |
| 455 | 'QSOX1_No21' | 'AAPGQEPPEHMAELQR' | 'Sulfhydryl oxidase 1' |
| 456 | 'QSOX1_No22' | 'IPYSFFK' | 'Sulfhydryl oxidase 1' |
| 457 | 'QSOX1_No23' | 'DCASHFEQMAAASMHR' | 'Sulfhydryl oxidase 1' |
| 458 | 'QSOX1_No24' | 'IYMADLESALHYILR' | 'Sulfhydryl oxidase 1' |
| 459 | 'QSOX1_No25' | 'RVLNTEANVVR' | 'Sulfhydryl oxidase 1' |
| 460 | 'QSOX1_No26' | 'DCASHFEQMAAASMHR' | 'Sulfhydryl oxidase 1' |
| 461 | 'QSOX1_No27' | 'NKIPYSFFK' | 'Sulfhydryl oxidase 1' |
| 462 | 'QSOX1_No28' | 'LEEIDGFFAR' | 'Sulfhydryl oxidase 1' |
| 463 | 'QSOX1_No29' | 'IYMADLESALHYILR' | 'Sulfhydryl oxidase 1' |
| 464 | 'QSOX1_No30' | 'DCASHFEQMAAASMHR' | 'Sulfhydryl oxidase 1' |
| 465 | 'QSOX1_No31' | 'VPVLMESR' | 'Sulfhydryl oxidase 1' |
| 466 | 'QSOX1_No32' | 'LWGPLEVR' | 'Sulfhydryl oxidase 1' |
| 467 | 'QSOX1_No33' | 'DVQNVAAAPELAMGALELESR' | 'Sulfhydryl oxidase 1' |
| 468 | 'QSOX1_No34' | 'VLNTEANVVRK' | 'Sulfhydryl oxidase 1' |
| 469 | 'QSOX1_No35' | 'RDVQNVAAAPELAMGALELESR' | 'Sulfhydryl oxidase 1' |
| 470 | 'QSOX1_No36' | 'SFYTAYLQR' | 'Sulfhydryl oxidase 1' |
| 471 | 'DNM1L_No1' | 'YIETSELCGGAR' | 'Dynamin-1-like protein' |
| 472 | 'DNM1L_No2' | 'LYTDFDEIRQEIENETER' | 'Dynamin-1-like protein' |
| 473 | 'DNM1L_No3' | 'AVMHFLVNHVK' | 'Dynamin-1-like protein' |
| 474 | 'DNM1L_No4' | 'NVASGGGGVGDGVQEPTTGNWR' | 'Dynamin-1-like protein' |
| 475 | 'DNM1L_No5' | 'DTLQSELVGQLYK' | 'Dynamin-1-like protein' |
| 476 | 'DNM1L_No6' | 'IIQHCSNYSTQELLR' | 'Dynamin-1-like protein' |
| 477 | 'NDUFA4' | 'LALFNPDVCWDR' | 'Cytochrome c oxidase subunit NDUFA4' |
| 478 | 'CHL1_No1' | 'TTEEDAGSYSCWVENAIGK' | 'Neural cell adhesion molecule L1-like protein' |
| 479 | 'CHL1_No2' | 'TTVILPLAPFVR' | 'Neural cell adhesion molecule L1-like protein' |
| 480 | 'CHL1_No3' | 'GDLYFANVEEKDSR' | 'Neural cell adhesion molecule L1-like protein' |
| 481 | 'CHL1_No4' | 'ACTSQGCGKPITEESSTLGEGSK' | 'Neural cell adhesion molecule L1-like protein' |
| 482 | 'CHL1_No5' | 'KTTVILPLAPFVR' | 'Neural cell adhesion molecule L1-like protein' |
| 483 | 'CHL1_No6' | 'KPQSAVYSTGSNGILLCEAEGEPQPTIK' | 'Neural cell adhesion molecule L1-like protein' |
| 484 | 'CHL1_No7' | 'LHMLELHCESK' | 'Neural cell adhesion molecule L1-like protein' |
| 485 | 'CHL1_No8' | 'SQPSQPSDHHETPPAAPDRNPQNIR' | 'Neural cell adhesion molecule L1-like protein' |

| | A | B | C |
|---|---|---|---|
| 1 | **Peptide Name** | **Peptide Sequence** | **Peptide Description** |
| 486 | 'CHL1_No9' | 'CTASNFLGTATHDFHVIVEEPPR' | 'Neural cell adhesion molecule L1-like protein' |
| 487 | 'CHL1_No10' | 'LGIAMSEEIEFIVPSVPK' | 'Neural cell adhesion molecule L1-like protein' |
| 488 | 'CHL1_No11' | 'IDPLEVEEGDPIVLPCNPPK' | 'Neural cell adhesion molecule L1-like protein' |
| 489 | 'CHL1_No12' | 'DGNPFYFTDHR' | 'Neural cell adhesion molecule L1-like protein' |
| 490 | 'CHL1_No13' | 'SQPSQPSDHHETPPAAPDR' | 'Neural cell adhesion molecule L1-like protein' |
| 491 | 'CHL1_No14' | 'EKIDPLEVEEGDPIVLPCNPPK' | 'Neural cell adhesion molecule L1-like protein' |
| 492 | 'CHL1_No15' | 'NDYCCFAAFPR' | 'Neural cell adhesion molecule L1-like protein' |
| 493 | 'CHL1_No16' | 'VMTPAVYAPYDVK' | 'Neural cell adhesion molecule L1-like protein' |
| 494 | 'CHL1_No17' | 'GDLYFANVEEK' | 'Neural cell adhesion molecule L1-like protein' |
| 495 | 'CHL1_No18' | 'VDKDTATLSWGLPK' | 'Neural cell adhesion molecule L1-like protein' |
| 496 | 'CHL1_No19' | 'IPNEGHISHFQGK' | 'Neural cell adhesion molecule L1-like protein' |
| 497 | 'CHL1_No20' | 'VIAVNEVGR' | 'Neural cell adhesion molecule L1-like protein' |
| 498 | 'CHL1_No21' | 'GYQINWWK' | 'Neural cell adhesion molecule L1-like protein' |
| 499 | 'CHL1_No22' | 'CTASNFLGTATHDFHVIVEEPPRWTK' | 'Neural cell adhesion molecule L1-like protein' |
| 500 | 'CHL1_No23' | 'GNPEPTFSWTK' | 'Neural cell adhesion molecule L1-like protein' |
| 501 | 'CHL1_No24' | 'VIKVDKDTATLSWGLPK' | 'Neural cell adhesion molecule L1-like protein' |
| 502 | 'CHL1_No25' | 'GAGPESEPYIFQTPEGVPEQPTFLK' | 'Neural cell adhesion molecule L1-like protein' |
| 503 | 'CHL1_No26' | 'TLKIENVSYQDKGNYR' | 'Neural cell adhesion molecule L1-like protein' |
| 504 | 'CHL1_No27' | 'SMEQNGPGLEYR' | 'Neural cell adhesion molecule L1-like protein' |
| 505 | 'CHL1_No28' | 'LLLPPTESGSESSITILK' | 'Neural cell adhesion molecule L1-like protein' |
| 506 | 'CHL1_No29' | 'TLKIENVSYQDK' | 'Neural cell adhesion molecule L1-like protein' |
| 507 | 'CHL1_No30' | 'IENVSYQDKGNYR' | 'Neural cell adhesion molecule L1-like protein' |
| 508 | 'PODXL' | 'LASVPGSQTVVVK' | 'Podocalyxin' |
| 509 | 'ISLR_No1' | 'LPGLPEGAFR' | 'Immunoglobulin superfamily containing leucine-rich repeat protein' |
| 510 | 'ISLR_No2' | 'ALPGTPVASSQPR' | 'Immunoglobulin superfamily containing leucine-rich repeat protein' |
| 511 | 'ISLR_No3' | 'EVPLLQSLWLAHNEIR' | 'Immunoglobulin superfamily containing leucine-rich repeat protein' |
| 512 | 'ISLR_No4' | 'MDSNELTFIPR' | 'Immunoglobulin superfamily containing leucine-rich repeat protein' |
| 513 | 'ISLR_No5' | 'TVAAGALASLSHLK' | 'Immunoglobulin superfamily containing leucine-rich repeat protein' |
| 514 | 'ISLR_No6' | 'TWALTTAVSIPEQDNIACTSPHVLK' | 'Immunoglobulin superfamily containing leucine-rich repeat protein' |
| 515 | 'ISLR_No7' | 'YGFQIADCAYR' | 'Immunoglobulin superfamily containing leucine-rich repeat protein' |
| 516 | 'TPP1_No1' | 'LFGGNFAHQASVAR' | 'Tripeptidyl-peptidase 1' |
| 517 | 'TPP1_No2' | 'LSELVQAVSDPSSPQYGK' | 'Tripeptidyl-peptidase 1' |
| 518 | 'TPP1_No3' | 'LYQQHGAGLFDVTR' | 'Tripeptidyl-peptidase 1' |
| 519 | 'NRP1_No1' | 'FVTAVGTQGAISK' | 'Neuropilin-1' |
| 520 | 'NRP1_No2' | 'SFEGNNNYDTPELR' | 'Neuropilin-1' |
| 521 | 'NRP1_No3' | 'IAPPPVVSSGPFLFIK' | 'Neuropilin-1' |
| 522 | 'NRP1_No4' | 'EGNKPVLFQGNTNPTDVVVAVFPKPLITR' | 'Neuropilin-1' |
| 523 | 'NRP1_No5' | 'YDRLEIWDGFPDVGPHIGR' | 'Neuropilin-1' |
| 524 | 'NRP1_No6' | 'IKPATWETGISMR' | 'Neuropilin-1' |
| 525 | 'NRP1_No7' | 'FVSDYETHGAGFSIR' | 'Neuropilin-1' |
| 526 | 'NRP1_No8' | 'SSSGILSMVFYTDSAIAK' | 'Neuropilin-1' |
| 527 | 'NRP1_No9' | 'IESPGYLTSPGYPHSYHPSEK' | 'Neuropilin-1' |
| 528 | 'NRP1_No10' | 'LNYPENGWTPGEDSYR' | 'Neuropilin-1' |
| 529 | 'NRP1_No11' | 'IKPATWETGISMR' | 'Neuropilin-1' |

| | A | B | C |
|---|---|---|---|
| 1 | **Peptide Name** | **Peptide Sequence** | **Peptide Description** |
| 530 | 'NRP1_No12' | 'CEWLIQAPDPYQR' | 'Neuropilin-1' |
| 531 | 'NRP1_No13' | 'IMINFNPHFDLEDR' | 'Neuropilin-1' |
| 532 | 'NRP1_No14' | 'YDYVEVFDGENENGHFR' | 'Neuropilin-1' |
| 533 | 'NRP1_No15' | 'LEIWDGFPDVGPHIGR' | 'Neuropilin-1' |
| 534 | 'NRP1_No16' | 'IMINFNPHFDLEDR' | 'Neuropilin-1' |
| 535 | 'NRP1_No17' | 'EGNKPVLFQGNTNPTDVVVAVFPK' | 'Neuropilin-1' |
| 536 | 'NRP1_No18' | 'IDVSSNGEDWITIK' | 'Neuropilin-1' |
| 537 | 'NRP1_No19' | 'SSSGILSMVFYTDSAIAK' | 'Neuropilin-1' |
| 538 | 'APOL1_No1' | 'VTEPISAESGEQVER' | 'Apolipoprotein L1' |
| 539 | 'APOL1_No2' | 'VQQNVPSGTDTGDPQSK' | 'Apolipoprotein L1' |
| 540 | 'APOL1_No3' | 'ANLQSVPHASASR' | 'Apolipoprotein L1' |
| 541 | 'APOL1_No4' | 'VAQELEEKLNILNNNYK' | 'Apolipoprotein L1' |
| 542 | 'APOL1_No5' | 'WWTQAQAHDLVIK' | 'Apolipoprotein L1' |
| 543 | 'APOL1_No6' | 'ANLQSVPHASASRPR' | 'Apolipoprotein L1' |
| 544 | 'APOL1_No7' | 'VTEPISAESGEQVERVNEPSILEMSR' | 'Apolipoprotein L1' |
| 545 | 'APOL1_No8' | 'VNEPSILEMSR' | 'Apolipoprotein L1' |
| 546 | 'APOL1_No9' | 'KWWTQAQAHDLVIK' | 'Apolipoprotein L1' |
| 547 | 'APOL1_No10' | 'VTEPISAESGEQVERVNEPSILEMSR' | 'Apolipoprotein L1' |
| 548 | 'APOL1_No11' | 'NWFLKEFPR' | 'Apolipoprotein L1' |
| 549 | 'APOL1_No12' | 'VNEPSILEMSR' | 'Apolipoprotein L1' |
| 550 | 'APOL1_No13' | 'LKSELEDNIR' | 'Apolipoprotein L1' |
| 551 | 'APOL1_No14' | 'LKSELEDNIRR' | 'Apolipoprotein L1' |
| 552 | 'APOL1_No15' | 'NEADELRK' | 'Apolipoprotein L1' |
| 553 | 'APOL1_No16' | 'NEADELRKALDNLAR' | 'Apolipoprotein L1' |
| 554 | 'APOL1_No17' | 'VAQELEEK' | 'Apolipoprotein L1' |
| 555 | 'APOL1_No18' | 'SETAEELKK' | 'Apolipoprotein L1' |
| 556 | 'APOL1_No19' | 'VQQNVPSGTDTGDPQSKPLGDWAAGTMDPESSIFIEDAIK' | 'Apolipoprotein L1' |
| 557 | 'APOL1_No20' | 'ALADGVQK' | 'Apolipoprotein L1' |
| 558 | 'APOL1_No21' | 'LNILNNNYK' | 'Apolipoprotein L1' |
| 559 | 'APOL1_No22' | 'DKNWHDKGQQYR' | 'Apolipoprotein L1' |
| 560 | 'APOL1_No23' | 'KVAQELEEKLNILNNNYK' | 'Apolipoprotein L1' |
| 561 | 'APOL1_No24' | 'NWHDKGQQYR' | 'Apolipoprotein L1' |
| 562 | 'APOL1_No25' | 'SELEDNIRR' | 'Apolipoprotein L1' |
| 563 | 'APOL1_No26' | 'PLGDWAAGTMDPESSIFIEDAIK' | 'Apolipoprotein L1' |
| 564 | 'PSMA7_No1' | 'LYQTDPSGTYHAWK' | 'Proteasome subunit alpha type-7' |
| 565 | 'PSMA7_No2' | 'ICALDDNVCMAFAGLTADAR' | 'Proteasome subunit alpha type-7' |
| 566 | 'PSMA7_No3' | 'GRDIVVLGVEK' | 'Proteasome subunit alpha type-7' |
| 567 | 'PSMA7_No4' | 'YVAEIEKEKEENEK' | 'Proteasome subunit alpha type-7' |
| 568 | 'PSMA7_No5' | 'LTVEDPVTVEYITR' | 'Proteasome subunit alpha type-7' |
| 569 | 'MYL12B_No1' | 'ELLTTMGDRFTDEEVDELYR' | 'Myosin regulatory light chain 12B' |
| 570 | 'MYL12B_No2' | 'ATSNVFAMFDQSQIQEFK' | 'Myosin regulatory light chain 12B' |
| 571 | 'MYL12B_No3' | 'FTDEEVDELYR' | 'Myosin regulatory light chain 12B' |
| 572 | 'MYL12B_No4' | 'DGFIDKEDLHDMLASLGK' | 'Myosin regulatory light chain 12B' |
| 573 | 'MYL12B_No5' | 'EAFNMIDQNR' | 'Myosin regulatory light chain 12B' |

| | A | B | C |
|---|---|---|---|
| 1 | **Peptide Name** | **Peptide Sequence** | **Peptide Description** |
| 574 | 'MYL12B_No6' | 'LNGTDPEDVIR' | 'Myosin regulatory light chain 12B' |
| 575 | 'MYL12B_No7' | 'DGFIDKEDLHDMLASLGK' | 'Myosin regulatory light chain 12B' |
| 576 | 'MYL12B_No8' | 'EAFNMIDQNR' | 'Myosin regulatory light chain 12B' |
| 577 | 'MYL12B_No9' | 'ATSNVFAMFDQSQIQEFK' | 'Myosin regulatory light chain 12B' |
| 578 | 'MYL12B_No10' | 'NAFACFDEEATGTIQEDYLR' | 'Myosin regulatory light chain 12B' |
| 579 | 'ARPC1B_No1' | 'STVLSLDWHPNNVLLAAGSCDFK' | 'Actin-related protein 2/3 complex subunit 1B' |
| 580 | 'ARPC1B_No2' | 'AYHSFLVEPISCHAWNK' | 'Actin-related protein 2/3 complex subunit 1B' |
| 581 | 'ARPC1B_No3' | 'ASSEGGTAAGAGLDSLHK' | 'Actin-related protein 2/3 complex subunit 1B' |
| 582 | 'ARPC1B_No4' | 'EVEERPAPTPWGSK' | 'Actin-related protein 2/3 complex subunit 1B' |
| 583 | 'ARPC1B_No5' | 'VAWVSHDSTVCLADADKK' | 'Actin-related protein 2/3 complex subunit 1B' |
| 584 | 'ARPC1B_No6' | 'EHNGQVTGIDWAPESNR' | 'Actin-related protein 2/3 complex subunit 1B' |
| 585 | 'ARPC1B_No7' | 'TQIAICPNNHEVHIYEK' | 'Actin-related protein 2/3 complex subunit 1B' |
| 586 | 'ARPC1B_No8' | 'AYHSFLVEPISCHAWNKDR' | 'Actin-related protein 2/3 complex subunit 1B' |
| 587 | 'ARPC2_No1' | 'DNTINLIHTFR' | 'Actin-related protein 2/3 complex subunit 2' |
| 588 | 'ARPC2_No2' | 'ELQAHGADELLKR' | 'Actin-related protein 2/3 complex subunit 2' |
| 589 | 'ARPC2_No3' | 'DSIVHQAGMLK' | 'Actin-related protein 2/3 complex subunit 2' |
| 590 | 'ARPC2_No4' | 'YFQFQEEGKEGENR' | 'Actin-related protein 2/3 complex subunit 2' |
| 591 | 'ARPC2_No5' | 'DSIVHQAGMLKR' | 'Actin-related protein 2/3 complex subunit 2' |
| 592 | 'ARPC2_No6' | 'ELQAHGADELLK' | 'Actin-related protein 2/3 complex subunit 2' |
| 593 | 'ARPC2_No7' | 'DYLHYHIK' | 'Actin-related protein 2/3 complex subunit 2' |
| 594 | 'ARPC2_No8' | 'MILLEVNNR' | 'Actin-related protein 2/3 complex subunit 2' |
| 595 | 'ARPC2_No9' | 'AVIHYRDDETMYVESK' | 'Actin-related protein 2/3 complex subunit 2' |
| 596 | 'ARPC3_No1' | 'LIGNMALLPIR' | 'Actin-related protein 2/3 complex subunit 3' |
| 597 | 'ARPC3_No2' | 'LIGNMALLPIR' | 'Actin-related protein 2/3 complex subunit 3' |
| 598 | 'ADAMDEC1_No1' | 'GEEITTKPENMEHCYYK' | 'ADAM DEC1' |
| 599 | 'ADAMDEC1_No2' | 'VGLAASNSLCSPSSVAVIEAK' | 'ADAM DEC1' |
| 600 | 'ADAMDEC1_No3' | 'LKPGTDCGGDAPNHTTE' | 'ADAM DEC1' |
| 601 | 'ADAMDEC1_No4' | 'GEEITTKPENMEHCYYK' | 'ADAM DEC1' |
| 602 | 'STX7_No1' | 'TLNQLGTPQDSPELR' | 'Syntaxin-7' |
| 603 | 'STX7_No2' | 'ITQCSVEIQR' | 'Syntaxin-7' |
| 604 | 'CCL16' | 'ALNCHLPAIIFVTK' | 'C-C motif chemokine 16' |
| 605 | 'PLXNB1_No1' | 'VYLGPGSDGHPYSTQSIQQGSAVSR' | 'Plexin-B1' |
| 606 | 'PLXNB1_No2' | 'GASAYFLFLR' | 'Plexin-B1' |
| 607 | 'PLXNB1_No3' | 'DPYCGWCVLLGR' | 'Plexin-B1' |
| 608 | 'PLXNB1_No4' | 'YGLIQAAAVATSR' | 'Plexin-B1' |
| 609 | 'PLXNB1_No5' | 'VVVGDQPCHLLPEQQSEQLR' | 'Plexin-B1' |
| 610 | 'PLXNB1_No6' | 'DQHYYSYVELPLACEGGR' | 'Plexin-B1' |
| 611 | 'PLXNB1_No7' | 'ALWPPDPQAAFSYEETAK' | 'Plexin-B1' |
| 612 | 'PLXNB1_No8' | 'CETSPRPTPATLPVAVWFGATER' | 'Plexin-B1' |
| 613 | 'PLXNB1_No9' | 'LSEYSHHFVSAFAR' | 'Plexin-B1' |
| 614 | 'PLXNB1_No10' | 'VPVASCAQHLDCASCLAHR' | 'Plexin-B1' |
| 615 | 'TGFB1I1_No1' | 'DFLQLFAPR' | 'Transforming growth factor beta-1-induced transcript 1 protein' |
| 616 | 'TGFB1I1_No2' | 'DGAPFCPECYFER' | 'Transforming growth factor beta-1-induced transcript 1 protein' |
| 617 | 'TXNL1' | 'VGVKPVGSDPDFQPELSGAGSR' | 'Thioredoxin-like protein 1' |

| | A | B | C |
|---|---|---|---|
| 1 | **Peptide Name** | **Peptide Sequence** | **Peptide Description** |
| 618 | 'AKR7A2_No1' | 'ALQAAYGASAPSVTSAALR' | 'Aflatoxin B1 aldehyde reductase member 2' |
| 619 | 'AKR7A2_No2' | 'FYAYNPLAGGLLTGK' | 'Aflatoxin B1 aldehyde reductase member 2' |
| 620 | 'TGOLN2_No1' | 'SGAEEQGPIDGPSK' | 'Trans-Golgi network integral membrane protein 2' |
| 621 | 'TGOLN2_No2' | 'DHSKPISNPSDNKELPK' | 'Trans-Golgi network integral membrane protein 2' |
| 622 | 'TGOLN2_No3' | 'SGAEDQTPKDVPNKSGAEK' | 'Trans-Golgi network integral membrane protein 2' |
| 623 | 'TGOLN2_No4' | 'SSAEAQTPEDTPNKSGAEAK' | 'Trans-Golgi network integral membrane protein 2' |
| 624 | 'ACTN4_No1' | 'LSGSNPYTTVTPQIINSK' | 'Alpha-actinin-4' |
| 625 | 'ACTN4_No2' | 'DGLAFNALIHR' | 'Alpha-actinin-4' |
| 626 | 'ACTN4_No3' | 'ACLISLGYDVENDR' | 'Alpha-actinin-4' |
| 627 | 'ACTN4_No4' | 'MLDAEDIVNTARPDEK' | 'Alpha-actinin-4' |
| 628 | 'ACTN4_No5' | 'ASFNHFDKDHGGALGPEEFK' | 'Alpha-actinin-4' |
| 629 | 'ACTN4_No6' | 'VLAVNQENEHLMEDYEK' | 'Alpha-actinin-4' |
| 630 | 'ACTN4_No7' | 'AGTQIENIDEDFRDGLK' | 'Alpha-actinin-4' |
| 631 | 'ACTN4_No8' | 'HRDYETATLSDIK' | 'Alpha-actinin-4' |
| 632 | 'ACTN4_No9' | 'DDPVTNLNNAFEVAEK' | 'Alpha-actinin-4' |
| 633 | 'ACTN4_No10' | 'MLDAEDIVNTARPDEK' | 'Alpha-actinin-4' |
| 634 | 'ACTN4_No11' | 'ASIHEAWTDGKEAMLK' | 'Alpha-actinin-4' |
| 635 | 'ACTN4_No12' | 'ETTDTDTADQVIASFK' | 'Alpha-actinin-4' |
| 636 | 'ACTN4_No13' | 'ICDQWDALGSLTHSR' | 'Alpha-actinin-4' |
| 637 | 'ACTN4_No14' | 'QLEAIDQLHLEYAK' | 'Alpha-actinin-4' |
| 638 | 'KRT86_No1' | 'ATAENEFVALKK' | 'Keratin, type II cuticular Hb6' |
| 639 | 'KRT86_No2' | 'LASELNHVQEVLEGYKK' | 'Keratin, type II cuticular Hb6' |
| 640 | 'KRT86_No3' | 'LEAAVAQSEQQGEAALSDAR' | 'Keratin, type II cuticular Hb6' |
| 641 | 'KRT86_No4' | 'TKEEINELNR' | 'Keratin, type II cuticular Hb6' |
| 642 | 'KRT86_No5' | 'VSSVPSNSNVVVGTTNACAPSAR' | 'Keratin, type II cuticular Hb6' |
| 643 | 'KRT86_No6' | 'FAAFIDK' | 'Keratin, type II cuticular Hb6' |
| 644 | 'KRT86_No7' | 'DLNMDCIIAEIK' | 'Keratin, type II cuticular Hb6' |
| 645 | 'KRT86_No8' | 'LAELEGALQK' | 'Keratin, type II cuticular Hb6' |
| 646 | 'KRT86_No9' | 'LASELNHVQEVLEGYK' | 'Keratin, type II cuticular Hb6' |
| 647 | 'KRT86_No10' | 'FAAFIDKVR' | 'Keratin, type II cuticular Hb6' |
| 648 | 'KRT86_No11' | 'LLEGEEQR' | 'Keratin, type II cuticular Hb6' |
| 649 | 'KRT86_No12' | 'GGVVCGDLCASTTAPVVSTR' | 'Keratin, type II cuticular Hb6' |
| 650 | 'KRT86_No13' | 'GLTGGFGSHSVCGGFR' | 'Keratin, type II cuticular Hb6' |
| 651 | 'KRT86_No14' | 'VLQSHISDTSVVVK' | 'Keratin, type II cuticular Hb6' |
| 652 | 'CD5L_No1' | 'FWGFHDCTHQEDVAVICSG' | 'CD5 antigen-like' |
| 653 | 'CD5L_No2' | 'VLIQSVSCTGTEDTLAQCEQEEVYDCSHDEDAGASCENPESSFSPVPE[illegible] | 'CD5 antigen-like' |
| 654 | 'CD5L_No3' | 'LVGGDNLCSGR' | 'CD5 antigen-like' |
| 655 | 'CD5L_No4' | 'EATLQDCPSGPWGK' | 'CD5 antigen-like' |
| 656 | 'CD5L_No5' | 'GVWGSVCDDNWGEKEDQVVCK' | 'CD5 antigen-like' |
| 657 | 'CD5L_No6' | 'IWLDNVR' | 'CD5 antigen-like' |
| 658 | 'CD5L_No7' | 'KPIWLSQMSCSGR' | 'CD5 antigen-like' |
| 659 | 'CD5L_No8' | 'HQNQWYTVCQTGWSLR' | 'CD5 antigen-like' |
| 660 | 'CD5L_No9' | 'GQWGTVCDDGWDIK' | 'CD5 antigen-like' |
| 661 | 'CD5L_No10' | 'GQWGTVCDDGWDIKDVAVLCR' | 'CD5 antigen-like' |

| | A | B | C |
|---|---|---|---|
| 1 | **Peptide Name** | **Peptide Sequence** | **Peptide Description** |
| 662 | 'CD5L_No11' | 'KPIWLSQMSCSGR' | 'CD5 antigen-like' |
| 663 | 'CD5L_No12' | 'NTCNHDEDTWVECEDPFDLR' | 'CD5 antigen-like' |
| 664 | 'CD5L_No13' | 'LVGGDNLCSGRLEVLHK' | 'CD5 antigen-like' |
| 665 | 'CD5L_No14' | 'CSGEEQSLEQCQHR' | 'CD5 antigen-like' |
| 666 | 'CD5L_No15' | 'NTCNHDEDTWVECEDPFDLRLVGGDNLCSGRLEVLHK' | 'CD5 antigen-like' |
| 667 | 'CD5L_No16' | 'CYGPGVGR' | 'CD5 antigen-like' |
| 668 | 'CD5L_No17' | 'VEVKHQNQWYTVCQTGWSLR' | 'CD5 antigen-like' |
| 669 | 'CD5L_No18' | 'ELGCGAASGTPSGILYEPPAEK' | 'CD5 antigen-like' |
| 670 | 'CD5L_No19' | 'ELGCGAASGTPSGILYEPPAEKEQK' | 'CD5 antigen-like' |
| 671 | 'CD5L_No20' | 'KCYGPGVGR' | 'CD5 antigen-like' |
| 672 | 'CD5L_No21' | 'AVLTQKR' | 'CD5 antigen-like' |
| 673 | 'CD5L_No22' | 'VEVEQKGQWGTVCDDGWDIKDVAVLCR' | 'CD5 antigen-like' |
| 674 | 'CD5L_No23' | 'GVWGSVCDDNWGEKEDQVVCKQLGCGK' | 'CD5 antigen-like' |
| 675 | 'CD5L_No24' | 'LEVLHKGVWGSVCDDNWGEKEDQVVCK' | 'CD5 antigen-like' |
| 676 | 'CD5L_No25' | 'NTCNHDEDTWVECEDPFDLRLVGGDNLCSGR' | 'CD5 antigen-like' |
| 677 | 'CD5L_No26' | 'CEGRVEVEQK' | 'CD5 antigen-like' |
| 678 | 'CD5L_No27' | 'LADGPGHCK' | 'CD5 antigen-like' |
| 679 | 'CD5L_No28' | 'GVWGSVCDDNWGEK' | 'CD5 antigen-like' |
| 680 | 'CD5L_No29' | 'HQNQWYTVCQTGWSLRAAK' | 'CD5 antigen-like' |
| 681 | 'GMFG_No1' | 'SDSLVVCEVDPELTEK' | 'Glia maturation factor gamma' |
| 682 | 'GMFG_No2' | 'SDSLVVCEVDPELTEKLR' | 'Glia maturation factor gamma' |
| 683 | 'GMFG_No3' | 'QMVVLEEEFQNISPEELK' | 'Glia maturation factor gamma' |
| 684 | 'PPP1R12B' | 'FSSGLFNK' | 'Protein phosphatase 1 regulatory subunit 12B' |
| 685 | 'NRP2_No1' | 'VFQANNDATEVVLNK' | 'Neuropilin-2' |
| 686 | 'NRP2_No2' | 'YYLVHQEPLENFQCNVPLGMESGR' | 'Neuropilin-2' |
| 687 | 'NRP2_No3' | 'IANEQISASSTYSDGR' | 'Neuropilin-2' |
| 688 | 'NRP2_No4' | 'HCGNIAPPTIISSGSMLYIK' | 'Neuropilin-2' |
| 689 | 'DIAPH1_No1' | 'LIEECISQIVLHK' | 'Protein diaphanous homolog 1' |
| 690 | 'DIAPH1_No2' | 'TAQNLSIFLGSFR' | 'Protein diaphanous homolog 1' |
| 691 | 'DIAPH1_No3' | 'SPDELPSAGGDGGK' | 'Protein diaphanous homolog 1' |
| 692 | 'DIAPH1_No4' | 'ELGEYFLFDPK' | 'Protein diaphanous homolog 1' |
| 693 | 'DIAPH1_No5' | 'LQFSEQVENIKPEIVSVTAACEELRK' | 'Protein diaphanous homolog 1' |
| 694 | 'FCMR_No1' | 'EMAGSGTCGTVVSTTNFIK' | 'Fas apoptotic inhibitory molecule 3' |
| 695 | 'FCMR_No2' | 'EMAGSGTCGTVVSTTNFIK' | 'Fas apoptotic inhibitory molecule 3' |
| 696 | 'FCGR3B_No1' | 'AVVFLEPQWYSVLEKDSVTLK' | 'Low affinity immunoglobulin gamma Fc region receptor III-B' |
| 697 | 'FCGR3B_No2' | 'AVVFLEPQWYSVLEK' | 'Low affinity immunoglobulin gamma Fc region receptor III-B' |
| 698 | 'WDR1_No1' | 'VFASLPQVER' | 'WD repeat-containing protein 1' |
| 699 | 'WDR1_No2' | 'LATGSDDNCAAFFEGPPFK' | 'WD repeat-containing protein 1' |
| 700 | 'WDR1_No3' | 'NIDNPALADIYTEHAHQVVVAK' | 'WD repeat-containing protein 1' |
| 701 | 'WDR1_No4' | 'LYSILGTTLKDEGK' | 'WD repeat-containing protein 1' |
| 702 | 'WDR1_No5' | 'IKDIAWTEDSKR' | 'WD repeat-containing protein 1' |
| 703 | 'WDR1_No6' | 'MTVDESGQLISCSMDDTVR' | 'WD repeat-containing protein 1' |
| 704 | 'WDR1_No7' | 'LHHVSSLAWLDEHTLVTTSHDASVK' | 'WD repeat-containing protein 1' |
| 705 | 'WDR1_No8' | 'VVTVFSVADGYSENNVFYGHHAK' | 'WD repeat-containing protein 1' |

| | A | B | C |
|---|---|---|---|
| 1 | **Peptide Name** | **Peptide Sequence** | **Peptide Description** |
| 706 | 'WDR1_No9' | 'AHDGGIYAISWSPDSTHLLSASGDK' | 'WD repeat-containing protein 1' |
| 707 | 'WDR1_No10' | 'SYIYSGSHDGHINYWDSETGENDSFAGK' | 'WD repeat-containing protein 1' |
| 708 | 'WDR1_No11' | 'GPVTDVAYSHDGAFLAVCDASK' | 'WD repeat-containing protein 1' |
| 709 | 'WDR1_No12' | 'MTVDESGQLISCSMDDTVR' | 'WD repeat-containing protein 1' |
| 710 | 'WDR1_No13' | 'CFSIDNPGYEPEVVAVHPGGDTVAIGGVDGNVR' | 'WD repeat-containing protein 1' |
| 711 | 'WDR1_No14' | 'FGAVFLWDSGSSVGEITGHNK' | 'WD repeat-containing protein 1' |
| 712 | 'ROCK2' | 'LEGWLSLPVR' | 'Rho-associated protein kinase 2' |
| 713 | 'ICOSLG_No1' | 'GLYDVVSVLR' | 'ICOS ligand' |
| 714 | 'ICOSLG_No2' | 'AMVGSDVELSCACPEGSR' | 'ICOS ligand' |
| 715 | 'ICOSLG_No3' | 'AMVGSDVELSCACPEGSR' | 'ICOS ligand' |
| 716 | 'PDCD6_No1' | 'LSDQFHDILIR' | 'Programmed cell death protein 6' |
| 717 | 'PDCD6_No2' | 'TYDRDNSGMIDKNELK' | 'Programmed cell death protein 6' |
| 718 | 'PDCD6_No3' | 'AGVNFSEFTGVWK' | 'Programmed cell death protein 6' |
| 719 | 'ERN1' | 'LTPTLYVGK' | 'Serine/threonine-protein kinase/endoribonuclease IRE1' |
| 720 | 'STX11_No1' | 'ELSEAAEAQHGPHSAVAR' | 'Syntaxin-11' |
| 721 | 'STX11_No2' | 'QADTLNVIELNVQK' | 'Syntaxin-11' |
| 722 | 'PGLYRP1_No1' | 'YVVVSHTAGSSCNTPASCQQQAR' | 'Peptidoglycan recognition protein 1' |
| 723 | 'PGLYRP1_No2' | 'ALASECAQHLSLPLR' | 'Peptidoglycan recognition protein 1' |
| 724 | 'PGLYRP1_No3' | 'AAQGLLACGVAQGALR' | 'Peptidoglycan recognition protein 1' |
| 725 | 'FCN3_No1' | 'YGIDWASGR' | 'Ficolin-3' |
| 726 | 'FCN3_No2' | 'ELLSQGATLSGWYHLCLPEGR' | 'Ficolin-3' |
| 727 | 'FCN3_No3' | 'TFAHYATFR' | 'Ficolin-3' |
| 728 | 'FCN3_No4' | 'VVLLPSCPGAPGSPGEK' | 'Ficolin-3' |
| 729 | 'FCN3_No5' | 'QDGSVDFFR' | 'Ficolin-3' |
| 730 | 'FCN3_No6' | 'LLGEVDHYQLALGK' | 'Ficolin-3' |
| 731 | 'FCN3_No7' | 'YAVSEAAAHKYGIDWASGR' | 'Ficolin-3' |
| 732 | 'FCN3_No8' | 'AGFGNQESEFWLGNENLHQLTLQGNWELR' | 'Ficolin-3' |
| 733 | 'FCN3_No9' | 'GEPGDPVNLLR' | 'Ficolin-3' |
| 734 | 'FCN3_No10' | 'FSEGTAGDSLSLHSGR' | 'Ficolin-3' |
| 735 | 'FCN3_No11' | 'RQDGSVDFFR' | 'Ficolin-3' |
| 736 | 'FCN3_No12' | 'MGPKGEPGDPVNLLR' | 'Ficolin-3' |
| 737 | 'FCN3_No13' | 'ALPVFCDMDTEGGGWLVFQR' | 'Ficolin-3' |
| 738 | 'FCN3_No14' | 'YAVSEAAAHK' | 'Ficolin-3' |
| 739 | 'FCN3_No15' | 'PFTTYDADHDSSNSNCAVIVHGAWWYASCYR' | 'Ficolin-3' |
| 740 | 'FCN3_No16' | 'ALPVFCDMDTEGGGWLVFQR' | 'Ficolin-3' |
| 741 | 'FCN3_No17' | 'SNLNGRYAVSEAAAHK' | 'Ficolin-3' |
| 742 | 'FCN3_No18' | 'SNLNGRYAVSEAAAHKYGIDWASGR' | 'Ficolin-3' |
| 743 | 'IDH1_No1' | 'IIWELIK' | 'Isocitrate dehydrogenase [NADP] cytoplasmic' |
| 744 | 'IDH1_No2' | 'SDYLNTFEFMDKLGENLK' | 'Isocitrate dehydrogenase [NADP] cytoplasmic' |
| 745 | 'IDH1_No3' | 'SEGGFIWACK' | 'Isocitrate dehydrogenase [NADP] cytoplasmic' |
| 746 | 'IDH1_No4' | 'SIEDFAHSSFQMALSK' | 'Isocitrate dehydrogenase [NADP] cytoplasmic' |
| 747 | 'IDH1_No5' | 'FKDIFQEIYDK' | 'Isocitrate dehydrogenase [NADP] cytoplasmic' |
| 748 | 'ATRN_No1' | 'NQECIALPENICGIGWHLVGNSCLK' | 'Attractin' |
| 749 | 'ATRN_No2' | 'LTGSSGFVTDGPGNYK' | 'Attractin' |

| | A | B | C |
|---|---|---|---|
| 1 | **Peptide Name** | **Peptide Sequence** | **Peptide Description** |
| 750 | 'ATRN_No3' | 'LTLTPWVGLR' | 'Attractin' |
| 751 | 'ATRN_No4' | 'GVKGDECQLCEVENR' | 'Attractin' |
| 752 | 'ATRN_No5' | 'EEYSNLKLPR' | 'Attractin' |
| 753 | 'ATRN_No6' | 'YGHSLALYK' | 'Attractin' |
| 754 | 'ATRN_No7' | 'HCETCISGFYGDPTNGGK' | 'Attractin' |
| 755 | 'ATRN_No8' | 'TACGDCTSGSSECMWCSNMK' | 'Attractin' |
| 756 | 'ATRN_No9' | 'YDVDTQMWTILK' | 'Attractin' |
| 757 | 'ATRN_No10' | 'CNPGTGQCVCPAGWVGEQCQHCGGR' | 'Attractin' |
| 758 | 'ATRN_No11' | 'LTLTPWVGLRK' | 'Attractin' |
| 759 | 'ATRN_No12' | 'CTWLIEGQPNR' | 'Attractin' |
| 760 | 'ATRN_No13' | 'AKEQYAVVGHSAHIVTLK' | 'Attractin' |
| 761 | 'ATRN_No14' | 'WSVLPRPDLHHDVNR' | 'Attractin' |
| 762 | 'ATRN_No15' | 'YGHSLALYKDK' | 'Attractin' |
| 763 | 'ATRN_No16' | 'CVWNTGSSQCISWALATDEQEEKLKSECFSK' | 'Attractin' |
| 764 | 'ATRN_No17' | 'EQYAVVGHSAHIVTLK' | 'Attractin' |
| 765 | 'ATRN_No18' | 'GDECQLCEVENR' | 'Attractin' |
| 766 | 'ATRN_No19' | 'TLDHDRCDQHTDCYSCTANTNDCHWCNDHCVPR' | 'Attractin' |
| 767 | 'ATRN_No20' | 'CVWNTGSSQCISWALATDEQEEKLK' | 'Attractin' |
| 768 | 'ATRN_No21' | 'LADDLYRYDVDTQMWTILKDSR' | 'Attractin' |
| 769 | 'ATRN_No22' | 'TNIKEYKDSFSNEK' | 'Attractin' |
| 770 | 'ATRN_No23' | 'CFSSDFMAYDIACDR' | 'Attractin' |
| 771 | 'ATRN_No24' | 'YDVDTQMWTILK' | 'Attractin' |
| 772 | 'ATRN_No25' | 'SEAACLAAGPGIR' | 'Attractin' |
| 773 | 'ATRN_No26' | 'SCALDQNCQWEPR' | 'Attractin' |
| 774 | 'ATRN_No27' | 'NHNALLASLTTQK' | 'Attractin' |
| 775 | 'ATRN_No28' | 'DLDMFINASK' | 'Attractin' |
| 776 | 'ATRN_No29' | 'CFSSDFMAYDIACDR' | 'Attractin' |
| 777 | 'ATRN_No30' | 'SVNNVVVR' | 'Attractin' |
| 778 | 'ATRN_No31' | 'CDQHTDCYSCTANTNDCHWCNDHCVPR' | 'Attractin' |
| 779 | 'ATRN_No32' | 'LTGSSGFVTDGPGNYKYK' | 'Attractin' |
| 780 | 'ATRN_No33' | 'YENCPKDNPMYYCNKK' | 'Attractin' |
| 781 | 'ATRN_No34' | 'DLDMFINASK' | 'Attractin' |
| 782 | 'ATRN_No35' | 'GVKGDECQLCEVENRYQGNPLR' | 'Attractin' |
| 783 | 'ATRN_No36' | 'LADDLYR' | 'Attractin' |
| 784 | 'ATRN_No37' | 'LADDLYRYDVDTQMWTILK' | 'Attractin' |
| 785 | 'ATRN_No38' | 'DSFSNEKFDFR' | 'Attractin' |
| 786 | 'ATRN_No39' | 'NTWSILHTQGALVQGGYGHSSVYDHR' | 'Attractin' |
| 787 | 'ATRN_No40' | 'TACGDCTSGSSECMWCSNMK' | 'Attractin' |
| 788 | 'ATRN_No41' | 'ISNSSDTVECECSENWKGEACDIPHCTDNCGFPHR' | 'Attractin' |
| 789 | 'ATRN_No42' | 'ALYVHGGYK' | 'Attractin' |
| 790 | 'ATRN_No43' | 'IYMYGGK' | 'Attractin' |
| 791 | 'ATRN_No44' | 'GDECQLCEVENRYQGNPLR' | 'Attractin' |
| 792 | 'ATRN_No45' | 'CIEGSYKGPVK' | 'Attractin' |
| 793 | 'ATRN_No46' | 'YQGNPLR' | 'Attractin' |

| | A | B | C |
|---|---|---|---|
| 1 | **Peptide Name** | **Peptide Sequence** | **Peptide Description** |
| 794 | 'ATRN_No47' | 'YENCPKDNPMYYCNKK' | 'Attractin' |
| 795 | 'ATRN_No48' | 'NHSCSEGQISIFR' | 'Attractin' |
| 796 | 'ATRN_No49' | 'TKCTWLIEGQPNR' | 'Attractin' |
| 797 | 'ATRN_No50' | 'YGHSLALYKDKIYMYGGK' | 'Attractin' |
| 798 | 'ATRN_No51' | 'YENCPKDNPMYYCNK' | 'Attractin' |
| 799 | 'ATRN_No52' | 'FRLTGSSGFVTDGPGNYK' | 'Attractin' |
| 800 | 'ATRN_No53' | 'GEACDIPHCTDNCGFPHR' | 'Attractin' |
| 801 | 'ATRN_No54' | 'ECDRPCVNGGR' | 'Attractin' |
| 802 | 'ATRN_No55' | 'LADDLYRYDVDTQMWTILK' | 'Attractin' |
| 803 | 'ATRN_No56' | 'FRLTGSSGFVTDGPGNYKYK' | 'Attractin' |
| 804 | 'ATRN_No57' | 'ITTAKENYDNAKLFCR' | 'Attractin' |
| 805 | 'ATRN_No58' | 'YENCPKDNPMYYCNK' | 'Attractin' |
| 806 | 'ATRN_No59' | 'LADDLYRYDVDTQMWTILKDSR' | 'Attractin' |
| 807 | 'ATRN_No60' | 'ITTAKENYDNAK' | 'Attractin' |
| 808 | 'ATRN_No61' | 'EYKDSFSNEK' | 'Attractin' |
| 809 | 'ARL6IP5_No1' | 'AWDDFFPGSDR' | 'PRA1 family protein 3' |
| 810 | 'ARL6IP5_No2' | 'TPMGIVLDALEQQEEGINR' | 'PRA1 family protein 3' |
| 811 | 'KRT34_No1' | 'LNVEVDTAPTVDLNQVLNETR' | 'Keratin, type I cuticular Ha4' |
| 812 | 'KRT34_No2' | 'LLVESDINSIRR' | 'Keratin, type I cuticular Ha4' |
| 813 | 'ENDOD1_No1' | 'STLLPPEASEGSSSFLGK' | 'Endonuclease domain-containing 1 protein' |
| 814 | 'ENDOD1_No2' | 'QALNTDYLDSDYQR' | 'Endonuclease domain-containing 1 protein' |
| 815 | 'ENDOD1_No3' | 'WYVNLHSLMDR' | 'Endonuclease domain-containing 1 protein' |
| 816 | 'ENDOD1_No4' | 'DRIPVYSAFR' | 'Endonuclease domain-containing 1 protein' |
| 817 | 'APOM_No1' | 'AFLLTPR' | 'Apolipoprotein M' |
| 818 | 'APOM_No2' | 'EFPEVHLGQWYFIAGAAPTK' | 'Apolipoprotein M' |
| 819 | 'APOM_No3' | 'SLTSCLDSK' | 'Apolipoprotein M' |
| 820 | 'APOM_No4' | 'MKDGLCVPR' | 'Apolipoprotein M' |
| 821 | 'APOM_No5' | 'KWIYHLTEGSTDLR' | 'Apolipoprotein M' |
| 822 | 'APOM_No6' | 'MKDGLCVPR' | 'Apolipoprotein M' |
| 823 | 'APOM_No7' | 'WIYHLTEGSTDLR' | 'Apolipoprotein M' |
| 824 | 'APOM_No8' | 'EELATFDPVDNIVFNMAAGSAPMQLHLR' | 'Apolipoprotein M' |
| 825 | 'APOM_No9' | 'EELATFDPVDNIVFNMAAGSAPMQLHLR' | 'Apolipoprotein M' |
| 826 | 'APOM_No10' | 'SPHPPEKCVEEFKSLTSCLDSK' | 'Apolipoprotein M' |
| 827 | 'APOM_No11' | 'WIYHLTEGSTDLRTEGRPDMK' | 'Apolipoprotein M' |
| 828 | 'APOM_No12' | 'WIYHLTEGSTDLRTEGRPDMK' | 'Apolipoprotein M' |
| 829 | 'APOM_No13' | 'NQEACELSNN' | 'Apolipoprotein M' |
| 830 | 'APOM_No14' | 'SPHPPEKCVEEFK' | 'Apolipoprotein M' |
| 831 | 'APOM_No15' | 'SLTSCLDSKAFLLTPR' | 'Apolipoprotein M' |
| 832 | 'APOM_No16' | 'TEGRPDMK' | 'Apolipoprotein M' |
| 833 | 'H6PD_No1' | 'QLVFHIGHGDLGSPAVLVSR' | 'GDH/6PGL endoplasmic bifunctional protein' |
| 834 | 'H6PD_No2' | 'QAVAQILPFR' | 'GDH/6PGL endoplasmic bifunctional protein' |
| 835 | 'H6PD_No3' | 'ELQKPDSFHSLTPTFAAVLVHIDNLR' | 'GDH/6PGL endoplasmic bifunctional protein' |
| 836 | 'H6PD_No4' | 'LANDIEATAVR' | 'GDH/6PGL endoplasmic bifunctional protein' |
| 837 | 'H6PD_No5' | 'DAHSVLLSHIFHGR' | 'GDH/6PGL endoplasmic bifunctional protein' |

| | A | B | C |
|---|---|---|---|
| 1 | **Peptide Name** | **Peptide Sequence** | **Peptide Description** |
| 838 | 'H6PD_No6' | 'WEGVPFILMSGK' | 'GDH/6PGL endoplasmic bifunctional protein' |
| 839 | 'H6PD_No7' | 'MSLSLPLINR' | 'GDH/6PGL endoplasmic bifunctional protein' |
| 840 | 'H6PD_No8' | 'DIEAQLQHAGLR' | 'GDH/6PGL endoplasmic bifunctional protein' |
| 841 | 'H6PD_No9' | 'LFGSPLSDYYAYSPVR' | 'GDH/6PGL endoplasmic bifunctional protein' |
| 842 | 'H6PD_No10' | 'LCAEEDQGAQIYAR' | 'GDH/6PGL endoplasmic bifunctional protein' |
| 843 | 'H6PD_No11' | 'GHSFSFHGAALTAPK' | 'GDH/6PGL endoplasmic bifunctional protein' |
| 844 | 'H6PD_No12' | 'HWAAAQSQCLPR' | 'GDH/6PGL endoplasmic bifunctional protein' |
| 845 | 'H6PD_No13' | 'IPYYNIHPMPVHLQQR' | 'GDH/6PGL endoplasmic bifunctional protein' |
| 846 | 'H6PD_No14' | 'TSFYEEYGVIR' | 'GDH/6PGL endoplasmic bifunctional protein' |
| 847 | 'H6PD_No15' | 'ESPLVSAWSEELISK' | 'GDH/6PGL endoplasmic bifunctional protein' |
| 848 | 'H6PD_No16' | 'TAEDYQALNKDIEAQLQHAGLR' | 'GDH/6PGL endoplasmic bifunctional protein' |
| 849 | 'VNN1_No1' | 'TTNLNTCGDSAETASTR' | 'Pantetheinase' |
| 850 | 'VNN1_No2' | 'MSENIPNEVYALGAFDGLHTVEGR' | 'Pantetheinase' |
| 851 | 'VNN1_No3' | 'GTVFFDEFTFVK' | 'Pantetheinase' |
| 852 | 'VNN1_No4' | 'YQYNTDVVFDSQGK' | 'Pantetheinase' |
| 853 | 'VNN1_No5' | 'DLCCHLSYK' | 'Pantetheinase' |
| 854 | 'VNN1_No6' | 'VNFLASNIHYPSKK' | 'Pantetheinase' |
| 855 | 'VNN1_No7' | 'LFSLKPTSGPVLTVTLFGR' | 'Pantetheinase' |
| 856 | 'VNN1_No8' | 'VNFLASNIHYPSK' | 'Pantetheinase' |
| 857 | 'VNN1_No9' | 'MSENIPNEVYALGAFDGLHTVEGR' | 'Pantetheinase' |
| 858 | 'VNN1_No10' | 'YYLQICTLLK' | 'Pantetheinase' |
| 859 | 'VNN1_No11' | 'KPCDTSDPQCPPDGR' | 'Pantetheinase' |
| 860 | 'CAVIN2_No1' | 'LVNMLDAVQENQHK' | 'Caveolae-associated protein 2' |
| 861 | 'CAVIN2_No2' | 'VLIFQEENEIPASVFVK' | 'Caveolae-associated protein 2' |
| 862 | 'CAVIN2_No3' | 'SDGDPVQPAVLQVHQTS' | 'Caveolae-associated protein 2' |
| 863 | 'CAVIN2_No4' | 'RLENNHAQLLR' | 'Caveolae-associated protein 2' |
| 864 | 'CAVIN2_No5' | 'QPVSGAVEGKEELPDENK' | 'Caveolae-associated protein 2' |
| 865 | 'CAVIN2_No6' | 'YEGSYALTSEEAER' | 'Caveolae-associated protein 2' |
| 866 | 'CAVIN2_No7' | 'QEKPSSPSPMPSSTPSPSLNLGNTEEAIR' | 'Caveolae-associated protein 2' |
| 867 | 'CAVIN2_No8' | 'GEDAAQAEKFQHPGSDMR' | 'Caveolae-associated protein 2' |
| 868 | 'CAVIN2_No9' | 'LVNMLDAVQENQHK' | 'Caveolae-associated protein 2' |
| 869 | 'CAVIN2_No10' | 'GEDAAQAEKFQHPGSDMR' | 'Caveolae-associated protein 2' |
| 870 | 'CAVIN2_No11' | 'YEGSYALTSEEAERSDGDPVQPAVLQVHQTS' | 'Caveolae-associated protein 2' |
| 871 | 'MPIG6B_No1' | 'TVLHVLGDR' | 'Megakaryocyte and platelet inhibitory receptor G6b' |
| 872 | 'MPIG6B_No2' | 'WVWAPSFPACK' | 'Megakaryocyte and platelet inhibitory receptor G6b' |
| 873 | 'EFEMP2_No1' | 'FSCHCPQGYQLLATR' | 'EGF-containing fibulin-like extracellular matrix protein 2' |
| 874 | 'EFEMP2_No2' | 'SCVDVNECDMGAPCEQR' | 'EGF-containing fibulin-like extracellular matrix protein 2' |
| 875 | 'EFEMP2_No3' | 'IGPECVDIDECR' | 'EGF-containing fibulin-like extracellular matrix protein 2' |
| 876 | 'EFEMP2_No4' | 'IGPECVDIDECRYR' | 'EGF-containing fibulin-like extracellular matrix protein 2' |
| 877 | 'EFEMP2_No5' | 'LCQDIDECESGAHQCSEAQTCVNFHGGYR' | 'EGF-containing fibulin-like extracellular matrix protein 2' |
| 878 | 'EFEMP2_No6' | 'CVEPYIQVSENR' | 'EGF-containing fibulin-like extracellular matrix protein 2' |
| 879 | 'EFEMP2_No7' | 'QINNVSAMLVLAR' | 'EGF-containing fibulin-like extracellular matrix protein 2' |
| 880 | 'EFEMP2_No8' | 'DVNECLTIPEACKGEMK' | 'EGF-containing fibulin-like extracellular matrix protein 2' |
| 881 | 'LDHA_No1' | 'VIGSGCNLDSAR' | 'L-lactate dehydrogenase A chain' |

| | A | B | C |
|---|---|---|---|
| 1 | **Peptide Name** | **Peptide Sequence** | **Peptide Description** |
| 882 | 'LDHA_No2' | 'VTLTSEEEAR' | 'L-lactate dehydrogenase A chain' |
| 883 | 'LDHA_No3' | 'FIIPNVVK' | 'L-lactate dehydrogenase A chain' |
| 884 | 'LDHA_No4' | 'TLHPDLGTDKDKEQWK' | 'L-lactate dehydrogenase A chain' |
| 885 | 'LDHA_No5' | 'GEMMDLQHGSLFLR' | 'L-lactate dehydrogenase A chain' |
| 886 | 'LDHA_No6' | 'ATLKDQLIYNLLK' | 'L-lactate dehydrogenase A chain' |
| 887 | 'LDHA_No7' | 'DQLIYNLLK' | 'L-lactate dehydrogenase A chain' |
| 888 | 'LDHA_No8' | 'DQLIYNLLKEEQTPQNK' | 'L-lactate dehydrogenase A chain' |
| 889 | 'LDHA_No9' | 'ATLKDQLIYNLLKEEQTPQNK' | 'L-lactate dehydrogenase A chain' |
| 890 | 'LDHA_No10' | 'SADTLWGIQKELQF' | 'L-lactate dehydrogenase A chain' |
| 891 | 'LDHA_No11' | 'IVSGKDYNVTANSK' | 'L-lactate dehydrogenase A chain' |
| 892 | 'ALDH1A1_No1' | 'IFVEESIYDEFVR' | 'Retinal dehydrogenase 1' |
| 893 | 'ALDH1A1_No2' | 'LYSNAYLNDLAGCIK' | 'Retinal dehydrogenase 1' |
| 894 | 'ALDH1A1_No3' | 'ANNTFYGLSAGVFTK' | 'Retinal dehydrogenase 1' |
| 895 | 'ALDH1A1_No4' | 'ELGEYGFHEYTEVK' | 'Retinal dehydrogenase 1' |
| 896 | 'ALDH1A1_No5' | 'TIPIDGNFFTYTR' | 'Retinal dehydrogenase 1' |
| 897 | 'CYB5R3_No1' | 'STPAITLESPDIK' | 'NADH-cytochrome b5 reductase 3' |
| 898 | 'CYB5R3_No2' | 'LIDREIISHDTR' | 'NADH-cytochrome b5 reductase 3' |
| 899 | 'CYB5R3_No3' | 'SVGMIAGGTGITPMLQVIR' | 'NADH-cytochrome b5 reductase 3' |
| 900 | 'CYB5R3_No4' | 'IDGNLVVRPYTPISSDDDKGFVDLVIK' | 'NADH-cytochrome b5 reductase 3' |
| 901 | 'CYB5R3_No5' | 'STPAITLESPDIKYPLR' | 'NADH-cytochrome b5 reductase 3' |
| 902 | 'CYB5R3_No6' | 'APEAWDYGQGFVNEEMIR' | 'NADH-cytochrome b5 reductase 3' |
| 903 | 'GSR_No1' | 'LGGTCVNVGCVPK' | 'Glutathione reductase, mitochondrial' |
| 904 | 'GSR_No2' | 'ALLTPVAIAAGR' | 'Glutathione reductase, mitochondrial' |
| 905 | 'GSR_No3' | 'TYSTSFTPMYHAVTK' | 'Glutathione reductase, mitochondrial' |
| 906 | 'GSR_No4' | 'VVGIHMQGLGCDEMLQGFAVAVK' | 'Glutathione reductase, mitochondrial' |
| 907 | 'GSR_No5' | 'GHAAFTSDPKPTIEVSGK' | 'Glutathione reductase, mitochondrial' |
| 908 | 'GSR_No6' | 'LNAIYQNNLTK' | 'Glutathione reductase, mitochondrial' |
| 909 | 'GSR_No7' | 'TYSTSFTPMYHAVTK' | 'Glutathione reductase, mitochondrial' |
| 910 | 'GSR_No8' | 'ADFDNTVAIHPTSSEELVTLR' | 'Glutathione reductase, mitochondrial' |
| 911 | 'GSR_No9' | 'LDYNNIPTVVFSHPPIGTVGLTEDEAIHK' | 'Glutathione reductase, mitochondrial' |
| 912 | 'SOD1_No1' | 'GDGPVQGIINFEQK' | 'Superoxide dismutase [Cu-Zn]' |
| 913 | 'SOD1_No2' | 'HVGDLGNVTADKDGVADVSIEDSVISLSGDHCIIGR' | 'Superoxide dismutase [Cu-Zn]' |
| 914 | 'SOD1_No3' | 'TLVVHEKADDLGKGGNEESTK' | 'Superoxide dismutase [Cu-Zn]' |
| 915 | 'SOD1_No4' | 'AVCVLKGDGPVQGIINFEQK' | 'Superoxide dismutase [Cu-Zn]' |
| 916 | 'CP_No1' | 'ALYLQYTDETFR' | 'Ceruloplasmin' |
| 917 | 'CP_No2' | 'DIASGLIGPLIICK' | 'Ceruloplasmin' |
| 918 | 'CP_No3' | 'VNKDDEEFIESNK' | 'Ceruloplasmin' |
| 919 | 'CP_No4' | 'MFTTAPDQVDKEDEDFQESNK' | 'Ceruloplasmin' |
| 920 | 'CP_No5' | 'HYYIGIIETTWDYASDHGEK' | 'Ceruloplasmin' |
| 921 | 'CP_No6' | 'HYYIGIIETTWDYASDHGEKK' | 'Ceruloplasmin' |
| 922 | 'CP_No7' | 'KAEEEHLGILGPQLHADVGDKVK' | 'Ceruloplasmin' |
| 923 | 'CP_No8' | 'EYTDASFTNR' | 'Ceruloplasmin' |
| 924 | 'CP_No9' | 'WYLFGMGNEVDVHAAFFHGQALTNK' | 'Ceruloplasmin' |
| 925 | 'CP_No10' | 'SVPPSASHVAPTETFTYEWTVPK' | 'Ceruloplasmin' |

| | A | B | C |
|---|---|---|---|
| 1 | **Peptide Name** | **Peptide Sequence** | **Peptide Description** |
| 926 | 'CP_No11' | 'DLYSGLIGPLIVCR' | 'Ceruloplasmin' |
| 927 | 'CP_No12' | 'PYTFHSHGITYYK' | 'Ceruloplasmin' |
| 928 | 'CP_No13' | 'TTIEKPVWLGFLGPIIK' | 'Ceruloplasmin' |
| 929 | 'CP_No14' | 'GPEEEHLGILGPVIWAEVGDTIR' | 'Ceruloplasmin' |
| 930 | 'CP_No15' | 'QSEDSTFYLGER' | 'Ceruloplasmin' |
| 931 | 'CP_No16' | 'EVGPTNADPVCLAK' | 'Ceruloplasmin' |
| 932 | 'CP_No17' | 'NNEGTYYSPNYNPQSR' | 'Ceruloplasmin' |
| 933 | 'CP_No18' | 'TPGIWLLHCHVTDHIHAGMETTYTVLQNEDTK' | 'Ceruloplasmin' |
| 934 | 'CP_No19' | 'RQSEDSTFYLGER' | 'Ceruloplasmin' |
| 935 | 'CP_No20' | 'PVWLGFLGPIIK' | 'Ceruloplasmin' |
| 936 | 'CP_No21' | 'EYTDASFTNRK' | 'Ceruloplasmin' |
| 937 | 'CP_No22' | 'AEEEHLGILGPQLHADVGDK' | 'Ceruloplasmin' |
| 938 | 'CP_No23' | 'ERGPEEEHLGILGPVIWAEVGDTIR' | 'Ceruloplasmin' |
| 939 | 'CP_No24' | 'LISVDTEHSNIYLQNGPDR' | 'Ceruloplasmin' |
| 940 | 'CP_No25' | 'KLISVDTEHSNIYLQNGPDR' | 'Ceruloplasmin' |
| 941 | 'CP_No26' | 'QYTDSTFRVPVER' | 'Ceruloplasmin' |
| 942 | 'CP_No27' | 'MHSMNGFMYGNQPGLTMCK' | 'Ceruloplasmin' |
| 943 | 'CP_No28' | 'DTANLFPQTSLTLHMWPDTEGTFNVECLTTDHYTGGMK' | 'Ceruloplasmin' |
| 944 | 'CP_No29' | 'DTANLFPQTSLTLHMWPDTEGTFNVECLTTDHYTGGMK' | 'Ceruloplasmin' |
| 945 | 'CP_No30' | 'MHSMNGFMYGNQPGLTMCK' | 'Ceruloplasmin' |
| 946 | 'CP_No31' | 'MFTTAPDQVDKEDEDFQESNK' | 'Ceruloplasmin' |
| 947 | 'CP_No32' | 'MYYSAVDPTKDIFTGLIGPMK' | 'Ceruloplasmin' |
| 948 | 'CP_No33' | 'MHSMNGFMYGNQPGLTMCK' | 'Ceruloplasmin' |
| 949 | 'CP_No34' | 'VKWYLFGMGNEVDVHAAFFHGQALTNK' | 'Ceruloplasmin' |
| 950 | 'CP_No35' | 'MHSMNGFMYGNQPGLTMCK' | 'Ceruloplasmin' |
| 951 | 'CP_No36' | 'QYTDSTFR' | 'Ceruloplasmin' |
| 952 | 'CP_No37' | 'MYYSAVDPTKDIFTGLIGPMK' | 'Ceruloplasmin' |
| 953 | 'CP_No38' | 'TYCSEPEKVDKDNEDFQESNR' | 'Ceruloplasmin' |
| 954 | 'CP_No39' | 'AETGDKVYVHLK' | 'Ceruloplasmin' |
| 955 | 'CP_No40' | 'DTANLFPQTSLTLHMWPDTEGTFNVECLTTDHYTGGMK' | 'Ceruloplasmin' |
| 956 | 'CP_No41' | 'YTVNQCR' | 'Ceruloplasmin' |
| 957 | 'CP_No42' | 'KALYLQYTDETFR' | 'Ceruloplasmin' |
| 958 | 'CP_No43' | 'IYHSHIDAPK' | 'Ceruloplasmin' |
| 959 | 'CP_No44' | 'MYYSAVDPTK' | 'Ceruloplasmin' |
| 960 | 'CP_No45' | 'MHSMNGFMYGNQPGLTMCK' | 'Ceruloplasmin' |
| 961 | 'CP_No46' | 'NMATRPYSIHAHGVQTESSTVTPTLPGETLTYVWK' | 'Ceruloplasmin' |
| 962 | 'CP_No47' | 'RDTANLFPQTSLTLHMWPDTEGTFNVECLTTDHYTGGMK' | 'Ceruloplasmin' |
| 963 | 'CP_No48' | 'WYLFGMGNEVDVHAAFFHGQALTNK' | 'Ceruloplasmin' |
| 964 | 'CP_No49' | 'QYTDSTFRVPVER' | 'Ceruloplasmin' |
| 965 | 'CP_No50' | 'TPGIWLLHCHVTDHIHAGMETTYTVLQNEDTK' | 'Ceruloplasmin' |
| 966 | 'CP_No51' | 'GAYPLSIEPIGVR' | 'Ceruloplasmin' |
| 967 | 'CP_No52' | 'MHSMNGFMYGNQPGLTMCK' | 'Ceruloplasmin' |
| 968 | 'CP_No53' | 'IYHSHIDAPKDIASGLIGPLIICK' | 'Ceruloplasmin' |
| 969 | 'CP_No54' | 'MHSMNGFMYGNQPGLTMCK' | 'Ceruloplasmin' |

| | A | B | C |
|---|---|---|---|
| 1 | **Peptide Name** | **Peptide Sequence** | **Peptide Description** |
| 970 | 'CP_No55' | 'DIFTGLIGPMK' | 'Ceruloplasmin' |
| 971 | 'CP_No56' | 'MYYSAVDPTKDIFTGLIGPMK' | 'Ceruloplasmin' |
| 972 | 'CP_No57' | 'DTANLFPQTSLTLHMWPDTEGTFNVECLTTDHYTGGMK' | 'Ceruloplasmin' |
| 973 | 'CP_No58' | 'MHSMNGFMYGNQPGLTMCK' | 'Ceruloplasmin' |
| 974 | 'CP_No59' | 'MHSMNGFMYGNQPGLTMCK' | 'Ceruloplasmin' |
| 975 | 'CP_No60' | 'MHSMNGFMYGNQPGLTMCK' | 'Ceruloplasmin' |
| 976 | 'CP_No61' | 'TYSDHPEKVNKDDEEFIESNK' | 'Ceruloplasmin' |
| 977 | 'CP_No62' | 'KAEEEHLGILGPQLHADVGDK' | 'Ceruloplasmin' |
| 978 | 'CP_No63' | 'SVPPSASHVAPTETFTYEWTVPKEVGPTNADPVCLAK' | 'Ceruloplasmin' |
| 979 | 'CP_No64' | 'KERGPEEEHLGILGPVIWAEVGDTIR' | 'Ceruloplasmin' |
| 980 | 'CP_No65' | 'MHSMNGFMYGNQPGLTMCK' | 'Ceruloplasmin' |
| 981 | 'CP_No66' | 'MHSMNGFMYGNQPGLTMCK' | 'Ceruloplasmin' |
| 982 | 'CP_No67' | 'MYSVNGYTFGSLPGLSMCAEDR' | 'Ceruloplasmin' |
| 983 | 'CP_No68' | 'NMATRPYSIHAHGVQTESSTVTPTLPGETLTYVWKIPER' | 'Ceruloplasmin' |
| 984 | 'CP_No69' | 'DIFTGLIGPMK' | 'Ceruloplasmin' |
| 985 | 'CP_No70' | 'AGLQAFFQVQECNK' | 'Ceruloplasmin' |
| 986 | 'CP_No71' | 'MHSMNGFMYGNQPGLTMCK' | 'Ceruloplasmin' |
| 987 | 'CP_No72' | 'IYHSHIDAPKDIASGLIGPLIICKK' | 'Ceruloplasmin' |
| 988 | 'CP_No73' | 'MHSMNGFMYGNQPGLTMCK' | 'Ceruloplasmin' |
| 989 | 'CP_No74' | 'MHSMNGFMYGNQPGLTMCK' | 'Ceruloplasmin' |
| 990 | 'CP_No75' | 'VNKDDEEFIESNKMHAINGR' | 'Ceruloplasmin' |
| 991 | 'CP_No76' | 'PYSIHAHGVQTESSTVTPTLPGETLTYVWK' | 'Ceruloplasmin' |
| 992 | 'CP_No77' | 'DIASGLIGPLIICKK' | 'Ceruloplasmin' |
| 993 | 'CP_No78' | 'VTFHNKGAYPLSIEPIGVR' | 'Ceruloplasmin' |
| 994 | 'CP_No79' | 'TPGIWLLHCHVTDHIHAGMETTYTVLQNEDTKSG' | 'Ceruloplasmin' |
| 995 | 'CP_No80' | 'MYSVNGYTFGSLPGLSMCAEDR' | 'Ceruloplasmin' |
| 996 | 'CP_No81' | 'MYYSAVDPTKDIFTGLIGPMK' | 'Ceruloplasmin' |
| 997 | 'CP_No82' | 'VDKDNEDFQESNR' | 'Ceruloplasmin' |
| 998 | 'CP_No83' | 'NLASRPYTFHSHGITYYK' | 'Ceruloplasmin' |
| 999 | 'CP_No84' | 'ADDKVYPGEQYTYMLLATEEQSPGEGDGNCVTR' | 'Ceruloplasmin' |
| 1000 | 'CP_No85' | 'FNKNNEGTYYSPNYNPQSR' | 'Ceruloplasmin' |
| 1001 | 'CP_No86' | 'QSEDSTFYLGER' | 'Ceruloplasmin' |
| 1002 | 'CP_No87' | 'MYSVNGYTFGSLPGLSMCAEDR' | 'Ceruloplasmin' |
| 1003 | 'CP_No88' | 'MFTTAPDQVDKEDEDFQESNKMHSMNGFMYGNQPGLTMCK' | 'Ceruloplasmin' |
| 1004 | 'CP_No89' | 'MYYSAVDPTK' | 'Ceruloplasmin' |
| 1005 | 'CP_No90' | 'NMATRPYSIHAHGVQTESSTVTPTLPGETLTYVWK' | 'Ceruloplasmin' |
| 1006 | 'CP_No91' | 'VKWYLFGMGNEVDVHAAFFHGQALTNK' | 'Ceruloplasmin' |
| 1007 | 'CP_No92' | 'KLVYREYTDASFTNR' | 'Ceruloplasmin' |
| 1008 | 'CP_No93' | 'MFTTAPDQVDKEDEDFQESNKMHSMNGFMYGNQPGLTMCK' | 'Ceruloplasmin' |
| 1009 | 'CP_No94' | 'AEEEHLGILGPQLHADVGDKVK' | 'Ceruloplasmin' |
| 1010 | 'CP_No95' | 'MYSVNGYTFGSLPGLSMCAEDRVK' | 'Ceruloplasmin' |
| 1011 | 'CP_No96' | 'QKDVDKEFYLFPTVFDENESLLLEDNIR' | 'Ceruloplasmin' |
| 1012 | 'CP_No97' | 'MYSVNGYTFGSLPGLSMCAEDRVK' | 'Ceruloplasmin' |
| 1013 | 'CP_No98' | 'LVYREYTDASFTNR' | 'Ceruloplasmin' |

| | A | B | C |
|---|---|---|---|
| 1 | **Peptide Name** | **Peptide Sequence** | **Peptide Description** |
| 1014 | 'CP_No99' | 'VNKDDEEFIESNKMHAINGR' | 'Ceruloplasmin' |
| 1015 | 'CP_No100' | 'KLISVDTEHSNIYLQNGPDRIGR' | 'Ceruloplasmin' |
| 1016 | 'CP_No101' | 'MYSVNGYTFGSLPGLSMCAEDRVK' | 'Ceruloplasmin' |
| 1017 | 'CP_No102' | 'VYPGEQYTYMLLATEEQSPGEGDGNCVTR' | 'Ceruloplasmin' |
| 1018 | 'CP_No103' | 'RDTANLFPQTSLTLHMWPDTEGTFNVECLTTDHYTGGMK' | 'Ceruloplasmin' |
| 1019 | 'CP_No104' | 'LVYREYTDASFTNRK' | 'Ceruloplasmin' |
| 1020 | 'CP_No105' | 'MHSMNGFMYGNQPGLTMCK' | 'Ceruloplasmin' |
| 1021 | 'CP_No106' | 'ADDKVYPGEQYTYMLLATEEQSPGEGDGNCVTR' | 'Ceruloplasmin' |
| 1022 | 'CP_No107' | 'LISVDTEHSNIYLQNGPDRIGR' | 'Ceruloplasmin' |
| 1023 | 'CP_No108' | 'VVYRQYTDSTFRVPVER' | 'Ceruloplasmin' |
| 1024 | 'CP_No109' | 'MFTTAPDQVDKEDEDFQESNKMHSMNGFMYGNQPGLTMCK' | 'Ceruloplasmin' |
| 1025 | 'CP_No110' | 'GVYSSDVFDIFPGTYQTLEMFPR' | 'Ceruloplasmin' |
| 1026 | 'CP_No111' | 'MFTTAPDQVDKEDEDFQESNKMHSMNGFMYGNQPGLTMCK' | 'Ceruloplasmin' |
| 1027 | 'CP_No112' | 'QYTDSTFRVPVERK' | 'Ceruloplasmin' |
| 1028 | 'CP_No113' | 'TYCSEPEKVDK' | 'Ceruloplasmin' |
| 1029 | 'CP_No114' | 'MFTTAPDQVDKEDEDFQESNKMHSMNGFMYGNQPGLTMCK' | 'Ceruloplasmin' |
| 1030 | 'CP_No115' | 'QKDVDKEFYLFPTVFDENESLLLEDNIR' | 'Ceruloplasmin' |
| 1031 | 'CP_No116' | 'DNEDFQESNR' | 'Ceruloplasmin' |
| 1032 | 'CP_No117' | 'MFTTAPDQVDKEDEDFQESNKMHSMNGFMYGNQPGLTMCK' | 'Ceruloplasmin' |
| 1033 | 'CP_No118' | 'TYCSEPEK' | 'Ceruloplasmin' |
| 1034 | 'CP_No119' | 'GSLHANGR' | 'Ceruloplasmin' |
| 1035 | 'CP_No120' | 'TYSDHPEK' | 'Ceruloplasmin' |
| 1036 | 'CP_No121' | 'TPGIWLLHCHVTDHIHAGMETTYTVLQNEDTKSG' | 'Ceruloplasmin' |
| 1037 | 'CP_No122' | 'MYYSAVDPTKDIFTGLIGPMKICK' | 'Ceruloplasmin' |
| 1038 | 'CP_No123' | 'EYTDASFTNRKER' | 'Ceruloplasmin' |
| 1039 | 'CP_No124' | 'SGAGTEDSACIPWAYYSTVDQVK' | 'Ceruloplasmin' |
| 1040 | 'CP_No125' | 'MYSVNGYTFGSLPGLSMCAEDRVK' | 'Ceruloplasmin' |
| 1041 | 'CP_No126' | 'QKYTVNQCR' | 'Ceruloplasmin' |
| 1042 | 'CP_No127' | 'QYTDSTFRVPVERK' | 'Ceruloplasmin' |
| 1043 | 'CP_No128' | 'EHEGAIYPDNTTDFQR' | 'Ceruloplasmin' |
| 1044 | 'CP_No129' | 'VVYRQYTDSTFR' | 'Ceruloplasmin' |
| 1045 | 'CP_No130' | 'QKYTVNQCR' | 'Ceruloplasmin' |
| 1046 | 'CP_No131' | 'DDEEFIESNK' | 'Ceruloplasmin' |
| 1047 | 'CP_No132' | 'VYPGEQYTYMLLATEEQSPGEGDGNCVTR' | 'Ceruloplasmin' |
| 1048 | 'CP_No133' | 'LYKKALYLQYTDETFR' | 'Ceruloplasmin' |
| 1049 | 'CP_No134' | 'MYSVNGYTFGSLPGLSMCAEDR' | 'Ceruloplasmin' |
| 1050 | 'CP_No135' | 'MHAINGR' | 'Ceruloplasmin' |
| 1051 | 'CP_No136' | 'GAYPLSIEPIGVRFNK' | 'Ceruloplasmin' |
| 1052 | 'CP_No137' | 'DDEEFIESNKMHAINGR' | 'Ceruloplasmin' |
| 1053 | 'CP_No138' | 'EWEKELHHLQEQNVSNAFLDKGEFYIGSK' | 'Ceruloplasmin' |
| 1054 | 'CP_No139' | 'ELHHLQEQNVSNAFLDK' | 'Ceruloplasmin' |
| 1055 | 'CP_No140' | 'KVVYRQYTDSTFR' | 'Ceruloplasmin' |
| 1056 | 'CP_No141' | 'MYYSAVDPTKDIFTGLIGPMKICK' | 'Ceruloplasmin' |
| 1057 | 'CP_No142' | 'DDEEFIESNKMHAINGR' | 'Ceruloplasmin' |

| | A | B | C |
|---|---|---|---|
| 1 | **Peptide Name** | **Peptide Sequence** | **Peptide Description** |
| 1058 | 'CP_No143' | 'ELHHLQEQNVSNAFLDKGEFYIGSK' | 'Ceruloplasmin' |
| 1059 | 'CP_No144' | 'DSLDKEK' | 'Ceruloplasmin' |
| 1060 | 'CP_No145' | 'AGLQAFFQVQECNKSSSK' | 'Ceruloplasmin' |
| 1061 | 'F13A1_No1' | 'STVLTIPEIIIK' | 'Coagulation factor XIII A chain' |
| 1062 | 'F13A1_No2' | 'LIASMSSDSLR' | 'Coagulation factor XIII A chain' |
| 1063 | 'F13A1_No3' | 'MYVAVWTPYGVLR' | 'Coagulation factor XIII A chain' |
| 1064 | 'F13A1_No4' | 'ETFDVTLEPLSFK' | 'Coagulation factor XIII A chain' |
| 1065 | 'F13A1_No5' | 'LALETALMYGAK' | 'Coagulation factor XIII A chain' |
| 1066 | 'F13A1_No6' | 'NVWVHLDGPGVTR' | 'Coagulation factor XIII A chain' |
| 1067 | 'F13A1_No7' | 'HVYGELDVQIQR' | 'Coagulation factor XIII A chain' |
| 1068 | 'F13A1_No8' | 'QIGGDGMMDITDTYK' | 'Coagulation factor XIII A chain' |
| 1069 | 'F13A1_No9' | 'LIASMSSDSLR' | 'Coagulation factor XIII A chain' |
| 1070 | 'F13A1_No10' | 'QIGGDGMMDITDTYK' | 'Coagulation factor XIII A chain' |
| 1071 | 'F13A1_No11' | 'EIRPNSTVQWEEVCRPWVSGHR' | 'Coagulation factor XIII A chain' |
| 1072 | 'F13A1_No12' | 'GVNLQEFLNVTSVHLFK' | 'Coagulation factor XIII A chain' |
| 1073 | 'F13A1_No13' | 'SNVDMDFEVENAVLGK' | 'Coagulation factor XIII A chain' |
| 1074 | 'F13A1_No14' | 'NVWVHLDGPGVTRPMK' | 'Coagulation factor XIII A chain' |
| 1075 | 'F13A1_No15' | 'GTYIPVPIVSELQSGK' | 'Coagulation factor XIII A chain' |
| 1076 | 'F13A1_No16' | 'NVWVHLDGPGVTRPMK' | 'Coagulation factor XIII A chain' |
| 1077 | 'F13A1_No17' | 'MYVAVWTPYGVLR' | 'Coagulation factor XIII A chain' |
| 1078 | 'F13A1_No18' | 'WDTNKVDHHTDKYENNK' | 'Coagulation factor XIII A chain' |
| 1079 | 'F13A1_No19' | 'DGTHVVENVDATHIGK' | 'Coagulation factor XIII A chain' |
| 1080 | 'F13A1_No20' | 'KLIASMSSDSLR' | 'Coagulation factor XIII A chain' |
| 1081 | 'F13A1_No21' | 'KETFDVTLEPLSFK' | 'Coagulation factor XIII A chain' |
| 1082 | 'F13A1_No22' | 'LALETALMYGAK' | 'Coagulation factor XIII A chain' |
| 1083 | 'F13A1_No23' | 'KDGTHVVENVDATHIGK' | 'Coagulation factor XIII A chain' |
| 1084 | 'F13A1_No24' | 'CGPASVQAIK' | 'Coagulation factor XIII A chain' |
| 1085 | 'F13A1_No25' | 'SNVDMDFEVENAVLGKDFK' | 'Coagulation factor XIII A chain' |
| 1086 | 'F13A1_No26' | 'SNVDMDFEVENAVLGK' | 'Coagulation factor XIII A chain' |
| 1087 | 'F13A1_No27' | 'NVWVHLDGPGVTRPMKK' | 'Coagulation factor XIII A chain' |
| 1088 | 'F13A1_No28' | 'KLIASMSSDSLR' | 'Coagulation factor XIII A chain' |
| 1089 | 'F13A1_No29' | 'GVNLQEFLNVTSVHLFKER' | 'Coagulation factor XIII A chain' |
| 1090 | 'F13A1_No30' | 'KETFDVTLEPLSFKK' | 'Coagulation factor XIII A chain' |
| 1091 | 'F13A1_No31' | 'ETFDVTLEPLSFKK' | 'Coagulation factor XIII A chain' |
| 1092 | 'F13A1_No32' | 'RAVPPNNSNAAEDDLPTVELQGVVPR' | 'Coagulation factor XIII A chain' |
| 1093 | 'F13A1_No33' | 'FQEGQEEERLALETALMYGAK' | 'Coagulation factor XIII A chain' |
| 1094 | 'F13A1_No34' | 'IVTNYFSAHDNDANLQMDIFLEEDGNVNSK' | 'Coagulation factor XIII A chain' |
| 1095 | 'F13A1_No35' | 'DLFRVEYVIGR' | 'Coagulation factor XIII A chain' |
| 1096 | 'F13A1_No36' | 'FQEGQEEERLALETALMYGAK' | 'Coagulation factor XIII A chain' |
| 1097 | 'F13A1_No37' | 'VDHHTDKYENNKLIVR' | 'Coagulation factor XIII A chain' |
| 1098 | 'F13A1_No38' | 'GTYIPVPIVSELQSGKWGAK' | 'Coagulation factor XIII A chain' |
| 1099 | 'F13A1_No39' | 'SNVDMDFEVENAVLGKDFK' | 'Coagulation factor XIII A chain' |
| 1100 | 'F13A1_No40' | 'QIGGDGMMDITDTYK' | 'Coagulation factor XIII A chain' |
| 1101 | 'F13A1_No41' | 'AVPPNNSNAAEDDLPTVELQGVVPR' | 'Coagulation factor XIII A chain' |

| | A | B | C |
|---|---|---|---|
| 1 | **Peptide Name** | **Peptide Sequence** | **Peptide Description** |
| 1102 | 'F13A1_No42' | 'AQMDLSGRGNPIKVSR' | 'Coagulation factor XIII A chain' |
| 1103 | 'F13A1_No43' | 'GTQVVGSDMTVTVQFTNPLK' | 'Coagulation factor XIII A chain' |
| 1104 | 'F13A1_No44' | 'IVTNYFSAHDNDANLQMDIFLEEDGNVNSK' | 'Coagulation factor XIII A chain' |
| 1105 | 'F13A1_No45' | 'VEYVIGRYPQENK' | 'Coagulation factor XIII A chain' |
| 1106 | 'F13A1_No46' | 'NVWVHLDGPGVTRPMKK' | 'Coagulation factor XIII A chain' |
| 1107 | 'F13A1_No47' | 'GTQVVGSDMTVTVQFTNPLK' | 'Coagulation factor XIII A chain' |
| 1108 | 'F13A1_No48' | 'AQMDLSGRGNPIK' | 'Coagulation factor XIII A chain' |
| 1109 | 'F13A1_No49' | 'QKSTVLTIPEIIIK' | 'Coagulation factor XIII A chain' |
| 1110 | 'F13A1_No50' | 'KDGTHVVENVDATHIGKLIVTK' | 'Coagulation factor XIII A chain' |
| 1111 | 'F13A1_No51' | 'AQMDLSGRGNPIKVSR' | 'Coagulation factor XIII A chain' |
| 1112 | 'F13A1_No52' | 'STVLTIPEIIIKVR' | 'Coagulation factor XIII A chain' |
| 1113 | 'PNP_No1' | 'LGADAVGMSTVPEVIVAR' | 'Purine nucleoside phosphorylase' |
| 1114 | 'PNP_No2' | 'LVFGFLNGR' | 'Purine nucleoside phosphorylase' |
| 1115 | 'PNP_No3' | 'VFHLLGVDTLVVTNAAGGLNPK' | 'Purine nucleoside phosphorylase' |
| 1116 | 'PNP_No4' | 'FEVGDIMLIR' | 'Purine nucleoside phosphorylase' |
| 1117 | 'PNP_No5' | 'FGDRFPAMSDAYDR' | 'Purine nucleoside phosphorylase' |
| 1118 | 'PNP_No6' | 'NTAEWLLSHTK' | 'Purine nucleoside phosphorylase' |
| 1119 | 'PNP_No7' | 'LGADAVGMSTVPEVIVAR' | 'Purine nucleoside phosphorylase' |
| 1120 | 'PNP_No8' | 'FHMYEGYPLWK' | 'Purine nucleoside phosphorylase' |
| 1121 | 'PNP_No9' | 'FGDRFPAMSDAYDR' | 'Purine nucleoside phosphorylase' |
| 1122 | 'PNP_No10' | 'FHMYEGYPLWK' | 'Purine nucleoside phosphorylase' |
| 1123 | 'EGFR_No1' | 'IPLENLQIIR' | 'Epidermal growth factor receptor' |
| 1124 | 'EGFR_No2' | 'GDSFTHTPPLDPQELDILK' | 'Epidermal growth factor receptor' |
| 1125 | 'EGFR_No3' | 'TDLHAFENLEIIR' | 'Epidermal growth factor receptor' |
| 1126 | 'EGFR_No4' | 'LTQLGTFEDHFLSLQR' | 'Epidermal growth factor receptor' |
| 1127 | 'EGFR_No5' | 'NLQEILHGAVR' | 'Epidermal growth factor receptor' |
| 1128 | 'EGFR_No6' | 'SPSDCCHNQCAAGCTGPR' | 'Epidermal growth factor receptor' |
| 1129 | 'EGFR_No7' | 'GPDNCIQCAHYIDGPHCVK' | 'Epidermal growth factor receptor' |
| 1130 | 'EGFR_No8' | 'ACGADSYEMEEDGVRK' | 'Epidermal growth factor receptor' |
| 1131 | 'PGK1_No1' | 'GCITIIGGGDTATCCAK' | 'Phosphoglycerate kinase 1' |
| 1132 | 'PGK1_No2' | 'ITLPVDFVTADKFDENAK' | 'Phosphoglycerate kinase 1' |
| 1133 | 'PGK1_No3' | 'VLNNMEIGTSLFDEEGAK' | 'Phosphoglycerate kinase 1' |
| 1134 | 'PGK1_No4' | 'AHSSMVGVNLPQK' | 'Phosphoglycerate kinase 1' |
| 1135 | 'PGK1_No5' | 'ALESPERPFLAILGGAK' | 'Phosphoglycerate kinase 1' |
| 1136 | 'PGK1_No6' | 'LTLDKLDVK' | 'Phosphoglycerate kinase 1' |
| 1137 | 'PGK1_No7' | 'ACANPAAGSVILLENLR' | 'Phosphoglycerate kinase 1' |
| 1138 | 'PGK1_No8' | 'LGDVYVNDAFGTAHR' | 'Phosphoglycerate kinase 1' |
| 1139 | 'PGK1_No9' | 'ALMDEVVK' | 'Phosphoglycerate kinase 1' |
| 1140 | 'PGK1_No10' | 'VDFNVPMK' | 'Phosphoglycerate kinase 1' |
| 1141 | 'PGK1_No11' | 'SLLGKDVLFLKDCVGPEVEK' | 'Phosphoglycerate kinase 1' |
| 1142 | 'PGK1_No12' | 'WNTEDKVSHVSTGGGASLELLEGK' | 'Phosphoglycerate kinase 1' |
| 1143 | 'PGK1_No13' | 'VLNNMEIGTSLFDEEGAK' | 'Phosphoglycerate kinase 1' |
| 1144 | 'AK1_No1' | 'KVNAEGSVDSVFSQVCTHLDALK' | 'Adenylate kinase isoenzyme 1' |
| 1145 | 'AK1_No2' | 'YGYTHLSTGDLLR' | 'Adenylate kinase isoenzyme 1' |

| | A | B | C |
|---|---|---|---|
| 1 | **Peptide Name** | **Peptide Sequence** | **Peptide Description** |
| 1146 | 'AK1_No3' | 'ATEPVIAFYEKR' | 'Adenylate kinase isoenzyme 1' |
| 1147 | 'AK1_No4' | 'GQLVPLETVLDMLR' | 'Adenylate kinase isoenzyme 1' |
| 1148 | 'AK1_No5' | 'IGQPTLLLYVDAGPETMTQR' | 'Adenylate kinase isoenzyme 1' |
| 1149 | 'F2_No1' | 'YGFYTHVFR' | 'Prothrombin' |
| 1150 | 'F2_No2' | 'LAVTTHGLPCLAWASAQAK' | 'Prothrombin' |
| 1151 | 'F2_No3' | 'SEGSSVNLSPPLEQCVPDR' | 'Prothrombin' |
| 1152 | 'F2_No4' | 'RQECSIPVCGQDQVTVAMTPR' | 'Prothrombin' |
| 1153 | 'F2_No5' | 'HQDFNSAVQLVENFCR' | 'Prothrombin' |
| 1154 | 'F2_No6' | 'TFGSGEADCGLRPLFEK' | 'Prothrombin' |
| 1155 | 'F2_No7' | 'GDACEGDSGGPFVMK' | 'Prothrombin' |
| 1156 | 'F2_No8' | 'LAACLEGNCAEGLGTNYR' | 'Prothrombin' |
| 1157 | 'F2_No9' | 'ITDNMFCAGYKPDEGKR' | 'Prothrombin' |
| 1158 | 'F2_No10' | 'QECSIPVCGQDQVTVAMTPR' | 'Prothrombin' |
| 1159 | 'F2_No11' | 'WYQMGIVSWGEGCDR' | 'Prothrombin' |
| 1160 | 'F2_No12' | 'WYQMGIVSWGEGCDRDGK' | 'Prothrombin' |
| 1161 | 'F2_No13' | 'LKKPVAFSDYIHPVCLPDR' | 'Prothrombin' |
| 1162 | 'F2_No14' | 'ENLDRDIALMK' | 'Prothrombin' |
| 1163 | 'F2_No15' | 'DKLAACLEGNCAEGLGTNYR' | 'Prothrombin' |
| 1164 | 'F2_No16' | 'IVEGSDAEIGMSPWQVMLFR' | 'Prothrombin' |
| 1165 | 'F2_No17' | 'VTGWGNLKETWTANVGK' | 'Prothrombin' |
| 1166 | 'F2_No18' | 'GQPSVLQVVNLPIVERPVCK' | 'Prothrombin' |
| 1167 | 'F2_No19' | 'ETAASLLQAGYK' | 'Prothrombin' |
| 1168 | 'F2_No20' | 'IVEGSDAEIGMSPWQVMLFR' | 'Prothrombin' |
| 1169 | 'F2_No21' | 'TFGSGEADCGLRPLFEKK' | 'Prothrombin' |
| 1170 | 'F2_No22' | 'ITDNMFCAGYKPDEGKR' | 'Prothrombin' |
| 1171 | 'F2_No23' | 'SLEDKTERELLESYIDGR' | 'Prothrombin' |
| 1172 | 'F2_No24' | 'NPDGDEEGVWCYVAGK' | 'Prothrombin' |
| 1173 | 'F2_No25' | 'ENLDRDIALMK' | 'Prothrombin' |
| 1174 | 'F2_No26' | 'ITDNMFCAGYKPDEGK' | 'Prothrombin' |
| 1175 | 'F2_No27' | 'IVEGSDAEIGMSPWQVMLFRK' | 'Prothrombin' |
| 1176 | 'F2_No28' | 'IVEGSDAEIGMSPWQVMLFR' | 'Prothrombin' |
| 1177 | 'F2_No29' | 'WYQMGIVSWGEGCDRDGK' | 'Prothrombin' |
| 1178 | 'F2_No30' | 'KPVAFSDYIHPVCLPDR' | 'Prothrombin' |
| 1179 | 'F2_No31' | 'ITDNMFCAGYKPDEGK' | 'Prothrombin' |
| 1180 | 'F2_No32' | 'RGDACEGDSGGPFVMK' | 'Prothrombin' |
| 1181 | 'F2_No33' | 'GQPSVLQVVNLPIVER' | 'Prothrombin' |
| 1182 | 'F2_No34' | 'YTACETAR' | 'Prothrombin' |
| 1183 | 'F2_No35' | 'WYQMGIVSWGEGCDR' | 'Prothrombin' |
| 1184 | 'F2_No36' | 'YNWRENLDRDIALMK' | 'Prothrombin' |
| 1185 | 'F2_No37' | 'SGIECQLWR' | 'Prothrombin' |
| 1186 | 'F2_No38' | 'YNWRENLDRDIALMK' | 'Prothrombin' |
| 1187 | 'F2_No39' | 'HQDFNSAVQLVENFCRNPDGDEEGVWCYVAGK' | 'Prothrombin' |
| 1188 | 'F2_No40' | 'QECSIPVCGQDQVTVAMTPR' | 'Prothrombin' |
| 1189 | 'F2_No41' | 'SPQELLCGASLISDR' | 'Prothrombin' |

| | A | B | C |
|---|---|---|---|
| 1 | **Peptide Name** | **Peptide Sequence** | **Peptide Description** |
| 1190 | 'F2_No42' | 'QECSIPVCGQDQVTVAMTPR' | 'Prothrombin' |
| 1191 | 'F2_No43' | 'KSPQELLCGASLISDR' | 'Prothrombin' |
| 1192 | 'F2_No44' | 'YNWRENLDR' | 'Prothrombin' |
| 1193 | 'F2_No45' | 'ETAASLLQAGYKGR' | 'Prothrombin' |
| 1194 | 'F2_No46' | 'ELLESYIDGR' | 'Prothrombin' |
| 1195 | 'F2_No47' | 'QECSIPVCGQDQVTVAMTPR' | 'Prothrombin' |
| 1196 | 'F2_No48' | 'RQECSIPVCGQDQVTVAMTPR' | 'Prothrombin' |
| 1197 | 'F2_No49' | 'TATSEYQTFFNPR' | 'Prothrombin' |
| 1198 | 'F2_No50' | 'GDACEGDSGGPFVMK' | 'Prothrombin' |
| 1199 | 'F2_No51' | 'NPDSSTTGPWCYTTDPTVRR' | 'Prothrombin' |
| 1200 | 'F2_No52' | 'SEGSSVNLSPPLEQCVPDRGQQYQGR' | 'Prothrombin' |
| 1201 | 'F2_No53' | 'ANTFLEEVRK' | 'Prothrombin' |
| 1202 | 'F2_No54' | 'RGDACEGDSGGPFVMK' | 'Prothrombin' |
| 1203 | 'F2_No55' | 'NPDSSTTGPWCYTTDPTVR' | 'Prothrombin' |
| 1204 | 'F2_No56' | 'KPVAFSDYIHPVCLPDRETAASLLQAGYK' | 'Prothrombin' |
| 1205 | 'F2_No57' | 'GQPSVLQVVNLPIVERPVCKDSTR' | 'Prothrombin' |
| 1206 | 'F2_No58' | 'NIEKISMLEK' | 'Prothrombin' |
| 1207 | 'F2_No59' | 'IVEGSDAEIGMSPWQVMLFR' | 'Prothrombin' |
| 1208 | 'F2_No60' | 'DGKYGFYTHVFR' | 'Prothrombin' |
| 1209 | 'F2_No61' | 'PVAFSDYIHPVCLPDRETAASLLQAGYK' | 'Prothrombin' |
| 1210 | 'F2_No62' | 'IRITDNMFCAGYKPDEGK' | 'Prothrombin' |
| 1211 | 'F2_No63' | 'ETWTANVGK' | 'Prothrombin' |
| 1212 | 'F2_No64' | 'IVEGSDAEIGMSPWQVMLFRK' | 'Prothrombin' |
| 1213 | 'F2_No65' | 'VIDQFGE' | 'Prothrombin' |
| 1214 | 'F2_No66' | 'ALSKHQDFNSAVQLVENFCR' | 'Prothrombin' |
| 1215 | 'F2_No67' | 'SLEDKTER' | 'Prothrombin' |
| 1216 | 'F2_No68' | 'WVLTAAHCLLYPPWDK' | 'Prothrombin' |
| 1217 | 'F2_No69' | 'VTGWGNLK' | 'Prothrombin' |
| 1218 | 'F2_No70' | 'NIEKISMLEKIYIHPR' | 'Prothrombin' |
| 1219 | 'F2_No71' | 'GRVTGWGNLKETWTANVGK' | 'Prothrombin' |
| 1220 | 'F2_No72' | 'ETWTANVGKGQPSVLQVVNLPIVERPVCK' | 'Prothrombin' |
| 1221 | 'F2_No73' | 'TERELLESYIDGR' | 'Prothrombin' |
| 1222 | 'F2_No74' | 'IVEGSDAEIGMSPWQVMLFRK' | 'Prothrombin' |
| 1223 | 'F2_No75' | 'IVEGSDAEIGMSPWQVMLFRK' | 'Prothrombin' |
| 1224 | 'F2_No76' | 'NIEKISMLEKIYIHPR' | 'Prothrombin' |
| 1225 | 'F2_No77' | 'PVAFSDYIHPVCLPDR' | 'Prothrombin' |
| 1226 | 'F2_No78' | 'IRITDNMFCAGYKPDEGK' | 'Prothrombin' |
| 1227 | 'F2_No79' | 'NIEKISMLEK' | 'Prothrombin' |
| 1228 | 'F2_No80' | 'GQQYQGR' | 'Prothrombin' |
| 1229 | 'C1R_No1' | 'FCGQLGSPLGNPPGKK' | 'Complement C1r subcomponent' |
| 1230 | 'C1R_No2' | 'FCGQLGSPLGNPPGK' | 'Complement C1r subcomponent' |
| 1231 | 'C1R_No3' | 'VLNYVDWIKK' | 'Complement C1r subcomponent' |
| 1232 | 'C1R_No4' | 'QGYQLIEGNQVLHSFTAVCQDDGTWHR' | 'Complement C1r subcomponent' |
| 1233 | 'C1R_No5' | 'LFGEVTSPLFPK' | 'Complement C1r subcomponent' |

| | A | B | C |
|---|---|---|---|
| 1 | **Peptide Name** | **Peptide Sequence** | **Peptide Description** |
| 1234 | 'C1R_No6' | 'MGNFPWQVFTNIHGR' | 'Complement C1r subcomponent' |
| 1235 | 'C1R_No7' | 'TLDEFTIIQNLQPQYQFR' | 'Complement C1r subcomponent' |
| 1236 | 'C1R_No8' | 'MDVFSQNMFCAGHPSLK' | 'Complement C1r subcomponent' |
| 1237 | 'C1R_No9' | 'QDACQGDSGGVFAVR' | 'Complement C1r subcomponent' |
| 1238 | 'C1R_No10' | 'YTTTMGVNTYK' | 'Complement C1r subcomponent' |
| 1239 | 'C1R_No11' | 'IQYYCHEPYYK' | 'Complement C1r subcomponent' |
| 1240 | 'C1R_No12' | 'LFGEVTSPLFPKPYPNNFETTTVITVPTGYR' | 'Complement C1r subcomponent' |
| 1241 | 'C1R_No13' | 'DPNTDRWVATGIVSWGIGCSR' | 'Complement C1r subcomponent' |
| 1242 | 'C1R_No14' | 'QRPPDLDTSSNAVDLLFFTDESGDSR' | 'Complement C1r subcomponent' |
| 1243 | 'C1R_No15' | 'VKLVFQQFDLEPSEGCFYDYVK' | 'Complement C1r subcomponent' |
| 1244 | 'C1R_No16' | 'MGNFPWQVFTNIHGR' | 'Complement C1r subcomponent' |
| 1245 | 'C1R_No17' | 'VLNYVDWIKKEMEEED' | 'Complement C1r subcomponent' |
| 1246 | 'C1R_No18' | 'MDVFSQNMFCAGHPSLK' | 'Complement C1r subcomponent' |
| 1247 | 'C1R_No19' | 'ESEQGVYTCTAQGIWKNEQKGEK' | 'Complement C1r subcomponent' |
| 1248 | 'C1R_No20' | 'MDVFSQNMFCAGHPSLK' | 'Complement C1r subcomponent' |
| 1249 | 'C1R_No21' | 'QDACQGDSGGVFAVRDPNTDR' | 'Complement C1r subcomponent' |
| 1250 | 'C1R_No22' | 'YTTEIIKCPQPK' | 'Complement C1r subcomponent' |
| 1251 | 'C1R_No23' | 'ESEQGVYTCTAQGIWKNEQK' | 'Complement C1r subcomponent' |
| 1252 | 'C1R_No24' | 'MDVFSQNMFCAGHPSLK' | 'Complement C1r subcomponent' |
| 1253 | 'C1R_No25' | 'QGYQLIEGNQVLHSFTAVCQDDGTWHR' | 'Complement C1r subcomponent' |
| 1254 | 'C1R_No26' | 'FVRLPVANPQACENWLR' | 'Complement C1r subcomponent' |
| 1255 | 'C1R_No27' | 'FLEPFDIDDHQQVHCPYDQLQIYANGK' | 'Complement C1r subcomponent' |
| 1256 | 'C1R_No28' | 'VLNYVDWIK' | 'Complement C1r subcomponent' |
| 1257 | 'C1R_No29' | 'ESEQGVYTCTAQGIWK' | 'Complement C1r subcomponent' |
| 1258 | 'C1R_No30' | 'WVATGIVSWGIGCSR' | 'Complement C1r subcomponent' |
| 1259 | 'C1R_No31' | 'LPVANPQACENWLR' | 'Complement C1r subcomponent' |
| 1260 | 'C1R_No32' | 'QDACQGDSGGVFAVR' | 'Complement C1r subcomponent' |
| 1261 | 'C1R_No33' | 'DYFIATCK' | 'Complement C1r subcomponent' |
| 1262 | 'C1R_No34' | 'LVFQQFDLEPSEGCFYDYVK' | 'Complement C1r subcomponent' |
| 1263 | 'C1R_No35' | 'PVNPVEQR' | 'Complement C1r subcomponent' |
| 1264 | 'C1R_No36' | 'CLPVCGKPVNPVEQR' | 'Complement C1r subcomponent' |
| 1265 | 'C1R_No37' | 'PYPNNFETTTVITVPTGYR' | 'Complement C1r subcomponent' |
| 1266 | 'C1R_No38' | 'MDVFSQNMFCAGHPSLKQDACQGDSGGVFAVR' | 'Complement C1r subcomponent' |
| 1267 | 'C1R_No39' | 'QDACQGDSGGVFAVRDPNTDR' | 'Complement C1r subcomponent' |
| 1268 | 'C1R_No40' | 'GFLAYYQAVDLDECASR' | 'Complement C1r subcomponent' |
| 1269 | 'C1R_No41' | 'YTTEIIK' | 'Complement C1r subcomponent' |
| 1270 | 'C1R_No42' | 'VLNYVDWIKKEMEEED' | 'Complement C1r subcomponent' |
| 1271 | 'C1R_No43' | 'VSVHPDYR' | 'Complement C1r subcomponent' |
| 1272 | 'C1R_No44' | 'MDVFSQNMFCAGHPSLKQDACQGDSGGVFAVR' | 'Complement C1r subcomponent' |
| 1273 | 'C1R_No45' | 'QRPPDLDTSSNAVDLLFFTDESGDSRGWK' | 'Complement C1r subcomponent' |
| 1274 | 'C1R_No46' | 'LRYTTEIIK' | 'Complement C1r subcomponent' |
| 1275 | 'C1R_No47' | 'YTTTMGVNTYK' | 'Complement C1r subcomponent' |
| 1276 | 'C1R_No48' | 'LRYTTEIIKCPQPK' | 'Complement C1r subcomponent' |
| 1277 | 'C1R_No49' | 'LFGEVTSPLFPKPYPNNFETTTVITVPTGYRVK' | 'Complement C1r subcomponent' |

| | A | B | C |
|---|---|---|---|
| 1 | **Peptide Name** | **Peptide Sequence** | **Peptide Description** |
| 1278 | 'C1R_No50' | 'NLPNGDFRYTTTMGVNTYK' | 'Complement C1r subcomponent' |
| 1279 | 'C1R_No51' | 'ARIQYYCHEPYYK' | 'Complement C1r subcomponent' |
| 1280 | 'C1R_No52' | 'QRPPDLDTSSNAVDLLFFTDESGDSR' | 'Complement C1r subcomponent' |
| 1281 | 'C1R_No53' | 'LGNHPIR' | 'Complement C1r subcomponent' |
| 1282 | 'C1R_No54' | 'NLPNGDFRYTTTMGVNTYKAR' | 'Complement C1r subcomponent' |
| 1283 | 'C1R_No55' | 'LPVANPQACENWLRGK' | 'Complement C1r subcomponent' |
| 1284 | 'HP_No1' | 'SCAVAEYGVYVK' | 'Haptoglobin' |
| 1285 | 'HP_No2' | 'DIAPTLTLYVGKK' | 'Haptoglobin' |
| 1286 | 'HP_No3' | 'VTSIQDWVQK' | 'Haptoglobin' |
| 1287 | 'HP_No4' | 'DIAPTLTLYVGK' | 'Haptoglobin' |
| 1288 | 'HP_No5' | 'AVGDKLPECEAVCGKPK' | 'Haptoglobin' |
| 1289 | 'HP_No6' | 'LRTEGDGVYTLNDKK' | 'Haptoglobin' |
| 1290 | 'HP_No7' | 'LRTEGDGVYTLNNEK' | 'Haptoglobin' |
| 1291 | 'HP_No8' | 'TEGDGVYTLNNEK' | 'Haptoglobin' |
| 1292 | 'HP_No9' | 'SPVGVQPILNEHTFCAGMSK' | 'Haptoglobin' |
| 1293 | 'HP_No10' | 'AVGDKLPECEAVCGK' | 'Haptoglobin' |
| 1294 | 'HP_No11' | 'ILGGHLDAK' | 'Haptoglobin' |
| 1295 | 'HP_No12' | 'TEGDGVYTLNDKK' | 'Haptoglobin' |
| 1296 | 'HP_No13' | 'VVLHPNYSQVDIGLIK' | 'Haptoglobin' |
| 1297 | 'HP_No14' | 'LPECEAVCGKPK' | 'Haptoglobin' |
| 1298 | 'HP_No15' | 'SPVGVQPILNEHTFCAGMSK' | 'Haptoglobin' |
| 1299 | 'HP_No16' | 'TEGDGVYTLNDK' | 'Haptoglobin' |
| 1300 | 'HP_No17' | 'VMPICLPSK' | 'Haptoglobin' |
| 1301 | 'HP_No18' | 'VMPICLPSKDYAEVGR' | 'Haptoglobin' |
| 1302 | 'HP_No19' | 'LPECEAVCGK' | 'Haptoglobin' |
| 1303 | 'HP_No20' | 'TEGDGVYTLNNEKQWINK' | 'Haptoglobin' |
| 1304 | 'HP_No21' | 'NPANPVQR' | 'Haptoglobin' |
| 1305 | 'HP_No22' | 'LRTEGDGVYTLNDK' | 'Haptoglobin' |
| 1306 | 'HP_No23' | 'HYEGSTVPEK' | 'Haptoglobin' |
| 1307 | 'HP_No24' | 'DYAEVGR' | 'Haptoglobin' |
| 1308 | 'HP_No25' | 'NANFKFTDHLK' | 'Haptoglobin' |
| 1309 | 'HP_No26' | 'TEGDGVYTLNDKKQWINK' | 'Haptoglobin' |
| 1310 | 'HP_No27' | 'KQLVEIEK' | 'Haptoglobin' |
| 1311 | 'HP_No28' | 'YVMLPVADQDQCIR' | 'Haptoglobin' |
| 1312 | 'HP_No29' | 'VMPICLPSK' | 'Haptoglobin' |
| 1313 | 'HP_No30' | 'AVGDKLPECEADDGCPKPPEIAHGYVEHSVR' | 'Haptoglobin' |
| 1314 | 'HP_No31' | 'LRTEGDGVYTLNNEKQWINK' | 'Haptoglobin' |
| 1315 | 'HP_No32' | 'LPECEADDGCPKPPEIAHGYVEHSVR' | 'Haptoglobin' |
| 1316 | 'HP_No33' | 'HYEGSTVPEKK' | 'Haptoglobin' |
| 1317 | 'HP_No34' | 'YVMLPVADQDQCIRHYEGSTVPEKK' | 'Haptoglobin' |
| 1318 | 'HP_No35' | 'VMPICLPSKDYAEVGR' | 'Haptoglobin' |
| 1319 | 'HP_No36' | 'YVMLPVADQDQCIRHYEGSTVPEKK' | 'Haptoglobin' |
| 1320 | 'HP_No37' | 'YVMLPVADQDQCIRHYEGSTVPEK' | 'Haptoglobin' |
| 1321 | 'HP_No38' | 'YVMLPVADQDQCIR' | 'Haptoglobin' |

| | A | B | C |
|---|---|---|---|
| 1 | **Peptide Name** | **Peptide Sequence** | **Peptide Description** |
| 1322 | 'HP_No39' | 'GSFPWQAK' | 'Haptoglobin' |
| 1323 | 'HP_No40' | 'VMPICLPSKDYAEVGRVGYVSGWGR' | 'Haptoglobin' |
| 1324 | 'HP_No41' | 'QKVSVNER' | 'Haptoglobin' |
| 1325 | 'HP_No42' | 'VGYVSGWGR' | 'Haptoglobin' |
| 1326 | 'HP_No43' | 'QLVEIEKVVLHPNYSQVDIGLIKLK' | 'Haptoglobin' |
| 1327 | 'HP_No44' | 'VTSIQDWVQKTIAEN' | 'Haptoglobin' |
| 1328 | 'HP_No45' | 'VVLHPNYSQVDIGLIKLK' | 'Haptoglobin' |
| 1329 | 'HP_No46' | 'YVMLPVADQDQCIRHYEGSTVPEK' | 'Haptoglobin' |
| 1330 | 'HP_No47' | 'FTDHLKYVMLPVADQDQCIR' | 'Haptoglobin' |
| 1331 | 'HP_No48' | 'FTDHLKYVMLPVADQDQCIR' | 'Haptoglobin' |
| 1332 | 'HP_No49' | 'PPEIAHGYVEHSVR' | 'Haptoglobin' |
| 1333 | 'HP_No50' | 'ILGGHLDAKGSFPWQAK' | 'Haptoglobin' |
| 1334 | 'HP_No51' | 'TPKSPVGVQPILNEHTFCAGMSK' | 'Haptoglobin' |
| 1335 | 'HP_No52' | 'QKVSVNER' | 'Haptoglobin' |
| 1336 | 'HP_No53' | 'LPECEAVCGKPKNPANPVQR' | 'Haptoglobin' |
| 1337 | 'HP_No54' | 'KQLVEIEKVVLHPNYSQVDIGLIK' | 'Haptoglobin' |
| 1338 | 'HP_No55' | 'QLVEIEKVVLHPNYSQVDIGLIK' | 'Haptoglobin' |
| 1339 | 'HP_No56' | 'NYYKLRTEGDGVYTLNNEK' | 'Haptoglobin' |
| 1340 | 'HP_No57' | 'NANFKFTDHLKYVMLPVADQDQCIR' | 'Haptoglobin' |
| 1341 | 'HP_No58' | 'TPKSPVGVQPILNEHTFCAGMSK' | 'Haptoglobin' |
| 1342 | 'HP_No59' | 'NANFKFTDHLKYVMLPVADQDQCIR' | 'Haptoglobin' |
| 1343 | 'HP_No60' | 'VMPICLPSKDYAEVGRVGYVSGWGR' | 'Haptoglobin' |
| 1344 | 'HP_No61' | 'LPECEADDGCPKPPEIAHGYVEHSVRYQCK' | 'Haptoglobin' |
| 1345 | 'HP_No62' | 'DIAPTLTLYVGKKQLVEIEK' | 'Haptoglobin' |
| 1346 | 'HP_No63' | 'QLVEIEKVVLHPNYSQVDIGLIK' | 'Haptoglobin' |
| 1347 | 'HP_No64' | 'NLFLNHSENATAK' | 'Haptoglobin' |
| 1348 | 'HP_No65' | 'NYYKLRTEGDGVYTLNDK' | 'Haptoglobin' |
| 1349 | 'HP_No66' | 'VGYVSGWGRNANFK' | 'Haptoglobin' |
| 1350 | 'HP_No67' | 'YQCKNYYK' | 'Haptoglobin' |
| 1351 | 'HP_No68' | 'DYAEVGRVGYVSGWGR' | 'Haptoglobin' |
| 1352 | 'HP_No69' | 'QLVEIEKVVLHPNYSQVDIGLIKLK' | 'Haptoglobin' |
| 1353 | 'HP_No70' | 'SCAVAEYGVYVKVTSIQDWVQK' | 'Haptoglobin' |
| 1354 | 'HP_No71' | 'LKQKVSVNER' | 'Haptoglobin' |
| 1355 | 'HP_No72' | 'VSVNERVMPICLPSKDYAEVGR' | 'Haptoglobin' |
| 1356 | 'HP_No73' | 'HYEGSTVPEKKTPK' | 'Haptoglobin' |
| 1357 | 'HP_No74' | 'VSVNERVMPICLPSKDYAEVGR' | 'Haptoglobin' |
| 1358 | 'HP_No75' | 'TEGDGVYTLNNEKQWINKAVGDK' | 'Haptoglobin' |
| 1359 | 'HPR_No1' | 'YVMLPVADQYDCITHYEGSTCPK' | 'Haptoglobin-related protein' |
| 1360 | 'HPR_No2' | 'VVLHPNYHQVDIGLIK' | 'Haptoglobin-related protein' |
| 1361 | 'HPR_No3' | 'VGYVSGWGQSDNFK' | 'Haptoglobin-related protein' |
| 1362 | 'HPR_No4' | 'SPVGVQPILNEHTFCVGMSK' | 'Haptoglobin-related protein' |
| 1363 | 'HPR_No5' | 'YVMLPVADQYDCITHYEGSTCPK' | 'Haptoglobin-related protein' |
| 1364 | 'HPR_No6' | 'SPVGVQPILNEHTFCVGMSK' | 'Haptoglobin-related protein' |
| 1365 | 'HPR_No7' | 'QLVEIEKVVLHPNYHQVDIGLIK' | 'Haptoglobin-related protein' |

| | A | B | C |
|---|---|---|---|
| 1 | **Peptide Name** | **Peptide Sequence** | **Peptide Description** |
| 1366 | 'HPR_No8' | 'QLFALYSGNDVTDISDDR' | 'Haptoglobin-related protein' |
| 1367 | 'HPR_No9' | 'VMPICLPSKNYAEVGR' | 'Haptoglobin-related protein' |
| 1368 | 'HPR_No10' | 'VTSIQHWVQK' | 'Haptoglobin-related protein' |
| 1369 | 'HPR_No11' | 'FPKPPEIANGYVEHLFR' | 'Haptoglobin-related protein' |
| 1370 | 'HPR_No12' | 'NYAEVGR' | 'Haptoglobin-related protein' |
| 1371 | 'HPR_No13' | 'QLVEIEKVVLHPNYHQVDIGLIK' | 'Haptoglobin-related protein' |
| 1372 | 'HPR_No14' | 'VGYVSGWGQSDNFKLTDHLK' | 'Haptoglobin-related protein' |
| 1373 | 'HPR_No15' | 'VMPICLPSKNYAEVGR' | 'Haptoglobin-related protein' |
| 1374 | 'HPR_No16' | 'QKVLVNER' | 'Haptoglobin-related protein' |
| 1375 | 'F9_No1' | 'FGSGYVSGWGR' | 'Coagulation factor IX' |
| 1376 | 'F9_No2' | 'IIPHHNYNAAINK' | 'Coagulation factor IX' |
| 1377 | 'F9_No3' | 'VDAFCGGSIVNEK' | 'Coagulation factor IX' |
| 1378 | 'F9_No4' | 'NCELDVTCNIK' | 'Coagulation factor IX' |
| 1379 | 'F9_No5' | 'FTIYNNMFCAGFHEGGR' | 'Coagulation factor IX' |
| 1380 | 'F9_No6' | 'SCEPAVPFPCGR' | 'Coagulation factor IX' |
| 1381 | 'F9_No7' | 'SALVLQYLR' | 'Coagulation factor IX' |
| 1382 | 'F9_No8' | 'VVGGEDAKPGQFPWQVVLNGK' | 'Coagulation factor IX' |
| 1383 | 'F9_No9' | 'VVCSCTEGYR' | 'Coagulation factor IX' |
| 1384 | 'F9_No10' | 'ITVVAGEHNIEETEHTEQKR' | 'Coagulation factor IX' |
| 1385 | 'F9_No11' | 'WIVTAAHCVETGVK' | 'Coagulation factor IX' |
| 1386 | 'F9_No12' | 'NSADNKVVCSCTEGYR' | 'Coagulation factor IX' |
| 1387 | 'F9_No13' | 'ITVVAGEHNIEETEHTEQK' | 'Coagulation factor IX' |
| 1388 | 'F9_No14' | 'DDINSYECWCPFGFEGK' | 'Coagulation factor IX' |
| 1389 | 'F9_No15' | 'NCELDVTCNIKNGR' | 'Coagulation factor IX' |
| 1390 | 'F9_No16' | 'FTIYNNMFCAGFHEGGR' | 'Coagulation factor IX' |
| 1391 | 'F9_No17' | 'SALVLQYLRVPLVDR' | 'Coagulation factor IX' |
| 1392 | 'F9_No18' | 'PGQFPWQVVLNGK' | 'Coagulation factor IX' |
| 1393 | 'F9_No19' | 'VSVSQTSK' | 'Coagulation factor IX' |
| 1394 | 'F10_No1' | 'ACIPTGPYPCGK' | 'Coagulation factor X' |
| 1395 | 'F10_No2' | 'MNVAPACLPERDWAESTLMTQK' | 'Coagulation factor X' |
| 1396 | 'F10_No3' | 'MNVAPACLPERDWAESTLMTQK' | 'Coagulation factor X' |
| 1397 | 'F10_No4' | 'LSSSFIITQNMFCAGYDTK' | 'Coagulation factor X' |
| 1398 | 'F10_No5' | 'TGIVSGFGR' | 'Coagulation factor X' |
| 1399 | 'F10_No6' | 'LSSSFIITQNMFCAGYDTKQEDACQGDSGGPHVTR' | 'Coagulation factor X' |
| 1400 | 'F10_No7' | 'FKDTYFVTGIVSWGEGCAR' | 'Coagulation factor X' |
| 1401 | 'F10_No8' | 'ETYDFDIAVLR' | 'Coagulation factor X' |
| 1402 | 'F10_No9' | 'QEDACQGDSGGPHVTR' | 'Coagulation factor X' |
| 1403 | 'F10_No10' | 'NTEQEEGGEAVHEVEVVIK' | 'Coagulation factor X' |
| 1404 | 'F10_No11' | 'MLEVPYVDR' | 'Coagulation factor X' |
| 1405 | 'F10_No12' | 'MNVAPACLPERDWAESTLMTQK' | 'Coagulation factor X' |
| 1406 | 'F10_No13' | 'VGDRNTEQEEGGEAVHEVEVVIK' | 'Coagulation factor X' |
| 1407 | 'F10_No14' | 'QEDACQGDSGGPHVTR' | 'Coagulation factor X' |
| 1408 | 'F10_No15' | 'KLCSLDNGDCDQFCHEEQNSVVCSCAR' | 'Coagulation factor X' |
| 1409 | 'F10_No16' | 'LKTPITFR' | 'Coagulation factor X' |

| | A | B | C |
|---|---|---|---|
| 1 | **Peptide Name** | **Peptide Sequence** | **Peptide Description** |
| 1410 | 'F10_No17' | 'YKDGDQCETSPCQNQGK' | 'Coagulation factor X' |
| 1411 | 'F10_No18' | 'LSSSFIITQNMFCAGYDTK' | 'Coagulation factor X' |
| 1412 | 'F10_No19' | 'MNVAPACLPER' | 'Coagulation factor X' |
| 1413 | 'F10_No20' | 'MLEVPYVDR' | 'Coagulation factor X' |
| 1414 | 'F10_No21' | 'DGDQCETSPCQNQGK' | 'Coagulation factor X' |
| 1415 | 'F10_No22' | 'MLEVPYVDRNSCK' | 'Coagulation factor X' |
| 1416 | 'F10_No23' | 'FTKETYDFDIAVLR' | 'Coagulation factor X' |
| 1417 | 'F10_No24' | 'GYTLADNGKACIPTGPYPCGK' | 'Coagulation factor X' |
| 1418 | 'F10_No25' | 'VGDRNTEQEEGGEAVHEVEVVIKHNR' | 'Coagulation factor X' |
| 1419 | 'F10_No26' | 'LSSSFIITQNMFCAGYDTKQEDACQGDSGGPHVTR' | 'Coagulation factor X' |
| 1420 | 'F10_No27' | 'DWAESTLMTQK' | 'Coagulation factor X' |
| 1421 | 'F10_No28' | 'DWAESTLMTQK' | 'Coagulation factor X' |
| 1422 | 'F10_No29' | 'CKDGLGEYTCTCLEGFEGKNCELFTR' | 'Coagulation factor X' |
| 1423 | 'F10_No30' | 'NTEQEEGGEAVHEVEVVIKHNR' | 'Coagulation factor X' |
| 1424 | 'F10_No31' | 'ANSFLEEMKK' | 'Coagulation factor X' |
| 1425 | 'F10_No32' | 'LCSLDNGDCDQFCHEEQNSVVCSCAR' | 'Coagulation factor X' |
| 1426 | 'F10_No33' | 'MLEVPYVDRNSCK' | 'Coagulation factor X' |
| 1427 | 'F10_No34' | 'LKMLEVPYVDR' | 'Coagulation factor X' |
| 1428 | 'CFD_No1' | 'DSCKGDSGGPLVCGGVLEGVVTSGSR' | 'Complement factor D' |
| 1429 | 'CFD_No2' | 'VQVLLGAHSLSQPEPSKR' | 'Complement factor D' |
| 1430 | 'CFD_No3' | 'RPDSLQHVLLPVLDR' | 'Complement factor D' |
| 1431 | 'CFD_No4' | 'ATLGPAVRPLPWQR' | 'Complement factor D' |
| 1432 | 'CFD_No5' | 'DVAPGTLCDVAGWGIVNHAGR' | 'Complement factor D' |
| 1433 | 'CFD_No6' | 'VQVLLGAHSLSQPEPSK' | 'Complement factor D' |
| 1434 | 'CFD_No7' | 'GDSGGPLVCGGVLEGVVTSGSR' | 'Complement factor D' |
| 1435 | 'CFD_No8' | 'AVPHPDSQPDTIDHDLLLLQLSEK' | 'Complement factor D' |
| 1436 | 'CFD_No9' | 'VDRDVAPGTLCDVAGWGIVNHAGR' | 'Complement factor D' |
| 1437 | 'CFD_No10' | 'KPGIYTR' | 'Complement factor D' |
| 1438 | 'CFD_No11' | 'VASYAAWIDSVLA' | 'Complement factor D' |
| 1439 | 'PLG_No1' | 'VILGAHQEVNLEPHVQEIEVSR' | 'Plasminogen' |
| 1440 | 'PLG_No2' | 'TECFITGWGETQGTFGAGLLK' | 'Plasminogen' |
| 1441 | 'PLG_No3' | 'FVTWIEGVMR' | 'Plasminogen' |
| 1442 | 'PLG_No4' | 'YILQGVTSWGLGCARPNKPGVYVR' | 'Plasminogen' |
| 1443 | 'PLG_No5' | 'LSSPAVITDKVIPACLPSPNYVVADR' | 'Plasminogen' |
| 1444 | 'PLG_No6' | 'ELRPWCFTTDPNKR' | 'Plasminogen' |
| 1445 | 'PLG_No7' | 'VVGGCVAHPHSWPWQVSLR' | 'Plasminogen' |
| 1446 | 'PLG_No8' | 'NPDADKGPWCFTTDPSVR' | 'Plasminogen' |
| 1447 | 'PLG_No9' | 'DKYILQGVTSWGLGCAR' | 'Plasminogen' |
| 1448 | 'PLG_No10' | 'NPDGDVGGPWCYTTNPR' | 'Plasminogen' |
| 1449 | 'PLG_No11' | 'MRDVVLFEK' | 'Plasminogen' |
| 1450 | 'PLG_No12' | 'NPDNDPQGPWCYTTDPEKR' | 'Plasminogen' |
| 1451 | 'PLG_No13' | 'ATTVTGTPCQDWAAQEPHR' | 'Plasminogen' |
| 1452 | 'PLG_No14' | 'GNVAVTVSGHTCQHWSAQTPHTHNR' | 'Plasminogen' |
| 1453 | 'PLG_No15' | 'LFLEPTRK' | 'Plasminogen' |

| | A | B | C |
|---|---|---|---|
| 1 | **Peptide Name** | **Peptide Sequence** | **Peptide Description** |
| 1454 | 'PLG_No16' | 'LYDYCDVPQCAAPSFDCGKPQVEPK' | 'Plasminogen' |
| 1455 | 'PLG_No17' | 'KQLGAGSIEECAAK' | 'Plasminogen' |
| 1456 | 'PLG_No18' | 'QLGAGSIEECAAK' | 'Plasminogen' |
| 1457 | 'PLG_No19' | 'MRDVVLFEK' | 'Plasminogen' |
| 1458 | 'PLG_No20' | 'TPENYPNAGLTMNYCR' | 'Plasminogen' |
| 1459 | 'PLG_No21' | 'CTTPPPSSGPTYQCLK' | 'Plasminogen' |
| 1460 | 'PLG_No22' | 'TPENYPNAGLTMNYCR' | 'Plasminogen' |
| 1461 | 'PLG_No23' | 'YILQGVTSWGLGCAR' | 'Plasminogen' |
| 1462 | 'PLG_No24' | 'LFLEPTR' | 'Plasminogen' |
| 1463 | 'PLG_No25' | 'FSPATHPSEGLEENYCR' | 'Plasminogen' |
| 1464 | 'PLG_No26' | 'NPDNDPQGPWCYTTDPEK' | 'Plasminogen' |
| 1465 | 'PLG_No27' | 'ELRPWCFTTDPNK' | 'Plasminogen' |
| 1466 | 'PLG_No28' | 'FGMHFCGGTLISPEWVLTAAHCLEK' | 'Plasminogen' |
| 1467 | 'PLG_No29' | 'WELCDIPR' | 'Plasminogen' |
| 1468 | 'PLG_No30' | 'YEFLNGR' | 'Plasminogen' |
| 1469 | 'PLG_No31' | 'DVVLFEK' | 'Plasminogen' |
| 1470 | 'PLG_No32' | 'TMSGLECQAWDSQSPHAHGYIPSK' | 'Plasminogen' |
| 1471 | 'PLG_No33' | 'QLGAGSIEECAAKCEEDEEFTCR' | 'Plasminogen' |
| 1472 | 'PLG_No34' | 'TMSGLECQAWDSQSPHAHGYIPSK' | 'Plasminogen' |
| 1473 | 'PLG_No35' | 'FVTWIEGVMR' | 'Plasminogen' |
| 1474 | 'PLG_No36' | 'FGMHFCGGTLISPEWVLTAAHCLEK' | 'Plasminogen' |
| 1475 | 'PLG_No37' | 'CQSWSSMTPHR' | 'Plasminogen' |
| 1476 | 'PLG_No38' | 'TMSGLECQAWDSQSPHAHGYIPSKFPNK' | 'Plasminogen' |
| 1477 | 'PLG_No39' | 'VCNRYEFLNGR' | 'Plasminogen' |
| 1478 | 'PLG_No40' | 'WEYCNLK' | 'Plasminogen' |
| 1479 | 'PLG_No41' | 'NLDENYCRNPDGK' | 'Plasminogen' |
| 1480 | 'PLG_No42' | 'NLDENYCR' | 'Plasminogen' |
| 1481 | 'PLG_No43' | 'CEEDEEFTCR' | 'Plasminogen' |
| 1482 | 'PLG_No44' | 'HSIFTPETNPR' | 'Plasminogen' |
| 1483 | 'PLG_No45' | 'CQSWSSMTPHR' | 'Plasminogen' |
| 1484 | 'PLG_No46' | 'APWCHTTNSQVR' | 'Plasminogen' |
| 1485 | 'PLG_No47' | 'TMSGLECQAWDSQSPHAHGYIPSKFPNK' | 'Plasminogen' |
| 1486 | 'PLG_No48' | 'QLGAGSIEECAAK' | 'Plasminogen' |
| 1487 | 'PLG_No49' | 'RWELCDIPR' | 'Plasminogen' |
| 1488 | 'PLG_No50' | 'TPENFPCK' | 'Plasminogen' |
| 1489 | 'PLG_No51' | 'WEYCNLKK' | 'Plasminogen' |
| 1490 | 'PLG_No52' | 'LSSPAVITDK' | 'Plasminogen' |
| 1491 | 'PLG_No53' | 'EQQCVIMAENRK' | 'Plasminogen' |
| 1492 | 'PLG_No54' | 'VQSTELCAGHLAGGTDSCQGDSGGPLVCFEKDK' | 'Plasminogen' |
| 1493 | 'PLG_No55' | 'PNKPGVYVR' | 'Plasminogen' |
| 1494 | 'PLG_No56' | 'TPENYPNAGLTMNYCRNPDADKGPWCFTTDPSVR' | 'Plasminogen' |
| 1495 | 'PLG_No57' | 'NPDGKRAPWCHTTNSQVR' | 'Plasminogen' |
| 1496 | 'PLG_No58' | 'MRDVVLFEKK' | 'Plasminogen' |
| 1497 | 'PLG_No59' | 'EAQLPVIENK' | 'Plasminogen' |

| | A | B | C |
|---|---|---|---|
| 1 | **Peptide Name** | **Peptide Sequence** | **Peptide Description** |
| 1498 | 'PLG_No60' | 'FSPATHPSEGLEENYCRNPDNDPQGPWCYTTDPEKR' | 'Plasminogen' |
| 1499 | 'PLG_No61' | 'KQLGAGSIEECAAKCEEDEEFTCR' | 'Plasminogen' |
| 1500 | 'PLG_No62' | 'EQQCVIMAENRK' | 'Plasminogen' |
| 1501 | 'PLG_No63' | 'AFQYHSKEQQCVIMAENRK' | 'Plasminogen' |
| 1502 | 'PLG_No64' | 'GTGENYRGNVAVTVSGHTCQHWSAQTPHTHNR' | 'Plasminogen' |
| 1503 | 'PLG_No65' | 'KLYDYCDVPQCAAPSFDCGKPQVEPK' | 'Plasminogen' |
| 1504 | 'PLG_No66' | 'NYCRNPDGDVGGPWCYTTNPR' | 'Plasminogen' |
| 1505 | 'PLG_No67' | 'KCQSWSSMTPHR' | 'Plasminogen' |
| 1506 | 'PLG_No68' | 'RAPWCHTTNSQVR' | 'Plasminogen' |
| 1507 | 'PLG_No69' | 'YDYCDILECEEECMHCSGENYDGKISK' | 'Plasminogen' |
| 1508 | 'PLG_No70' | 'CSGTEASVVAPPPVVLLPDVETPSEEDCMFGNGK' | 'Plasminogen' |
| 1509 | 'PLG_No71' | 'VIPACLPSPNYVVADR' | 'Plasminogen' |
| 1510 | 'PLG_No72' | 'EQQCVIMAENR' | 'Plasminogen' |
| 1511 | 'PLG_No73' | 'TMSGLECQAWDSQSPHAHGYIPSKFPNKNLK' | 'Plasminogen' |
| 1512 | 'PLG_No74' | 'EQQCVIMAENR' | 'Plasminogen' |
| 1513 | 'PLG_No75' | 'VIPACLPSPNYVVADRTECFITGWGETQGTFGAGLLK' | 'Plasminogen' |
| 1514 | 'PLG_No76' | 'KCQSWSSMTPHR' | 'Plasminogen' |
| 1515 | 'PLG_No77' | 'KVYLSECK' | 'Plasminogen' |
| 1516 | 'PLG_No78' | 'MRDVVLFEKK' | 'Plasminogen' |
| 1517 | 'PLG_No79' | 'NPDGDVGGPWCYTTNPRK' | 'Plasminogen' |
| 1518 | 'PLG_No80' | 'KSSIIIR' | 'Plasminogen' |
| 1519 | 'PLG_No81' | 'RYDYCDILECEEECMHCSGENYDGKISK' | 'Plasminogen' |
| 1520 | 'PLG_No82' | 'SPRPSSYKVILGAHQEVNLEPHVQEIEVSR' | 'Plasminogen' |
| 1521 | 'PLG_No83' | 'RATTVTGTPCQDWAAQEPHRHSIFTPETNPR' | 'Plasminogen' |
| 1522 | 'PLG_No84' | 'TMSGLECQAWDSQSPHAHGYIPSKFPNKNLK' | 'Plasminogen' |
| 1523 | 'PLG_No85' | 'ATTVTGTPCQDWAAQEPHRHSIFTPETNPR' | 'Plasminogen' |
| 1524 | 'PLG_No86' | 'WSSTSPHRPR' | 'Plasminogen' |
| 1525 | 'PLG_No87' | 'QLGAGSIEECAAKCEEDEEFTCR' | 'Plasminogen' |
| 1526 | 'PLG_No88' | 'VYLSECK' | 'Plasminogen' |
| 1527 | 'PLG_No89' | 'NLDENYCRNPDGKR' | 'Plasminogen' |
| 1528 | 'PLG_No90' | 'AFQYHSKEQQCVIMAENRK' | 'Plasminogen' |
| 1529 | 'PLG_No91' | 'NPDADKGPWCFTTDPSVRWEYCNLK' | 'Plasminogen' |
| 1530 | 'PLG_No92' | 'YDYCDILECEEECMHCSGENYDGKISK' | 'Plasminogen' |
| 1531 | 'PLG_No93' | 'PWCFTTDPNKR' | 'Plasminogen' |
| 1532 | 'PLG_No94' | 'RATTVTGTPCQDWAAQEPHR' | 'Plasminogen' |
| 1533 | 'PLG_No95' | 'AFQYHSKEQQCVIMAENR' | 'Plasminogen' |
| 1534 | 'PLG_No96' | 'TPENYPNAGLTMNYCRNPDADKGPWCFTTDPSVR' | 'Plasminogen' |
| 1535 | 'PLG_No97' | 'EAQLPVIENKVCNRYEFLNGR' | 'Plasminogen' |
| 1536 | 'PLG_No98' | 'KLYDYCDVPQCAAPSFDCGK' | 'Plasminogen' |
| 1537 | 'PLG_No99' | 'VSRFVTWIEGVMR' | 'Plasminogen' |
| 1538 | 'PLG_No100' | 'NPDRELRPWCFTTDPNK' | 'Plasminogen' |
| 1539 | 'PLG_No101' | 'HQKTPENYPNAGLTMNYCR' | 'Plasminogen' |
| 1540 | 'PLG_No102' | 'LFLEPTRKDIALLK' | 'Plasminogen' |
| 1541 | 'PLG_No103' | 'PSSYKVILGAHQEVNLEPHVQEIEVSR' | 'Plasminogen' |

| | A | B | C |
|---|---|---|---|
| 1 | **Peptide Name** | **Peptide Sequence** | **Peptide Description** |
| 1542 | 'PLG_No104' | 'TRFGMHFCGGTLISPEWVLTAAHCLEK' | 'Plasminogen' |
| 1543 | 'PLG_No105' | 'NYCRNPDGDVGGPWCYTTNPRK' | 'Plasminogen' |
| 1544 | 'PLG_No106' | 'IPSCDSSPVSTEQLAPTAPPELTPVVQDCYHGDGQSYR' | 'Plasminogen' |
| 1545 | 'PLG_No107' | 'AFQYHSKEQQCVIMAENR' | 'Plasminogen' |
| 1546 | 'PLG_No108' | 'RYDYCDILECEEECMHCSGENYDGK' | 'Plasminogen' |
| 1547 | 'PLG_No109' | 'KCSGTEASVVAPPPVVLLPDVETPSEEDCMFGNGK' | 'Plasminogen' |
| 1548 | 'PLG_No110' | 'EAQLPVIENKVCNR' | 'Plasminogen' |
| 1549 | 'PLG_No111' | 'GPWCFTTDPSVR' | 'Plasminogen' |
| 1550 | 'PLG_No112' | 'LYDYCDVPQCAAPSFDCGKPQVEPKK' | 'Plasminogen' |
| 1551 | 'PLG_No113' | 'YDYCDILECEEECMHCSGENYDGK' | 'Plasminogen' |
| 1552 | 'PLG_No114' | 'DIALLKLSSPAVITDK' | 'Plasminogen' |
| 1553 | 'PLG_No115' | 'HQKTPENYPNAGLTMNYCR' | 'Plasminogen' |
| 1554 | 'PLG_No116' | 'AFQYHSK' | 'Plasminogen' |
| 1555 | 'PLG_No117' | 'RAPWCHTTNSQVRWEYCK' | 'Plasminogen' |
| 1556 | 'PLG_No118' | 'YDYCDILECEEECMHCSGENYDGK' | 'Plasminogen' |
| 1557 | 'PLG_No119' | 'TPENFPCKNLDENYCR' | 'Plasminogen' |
| 1558 | 'PLG_No120' | 'VQSTELCAGHLAGGTDSCQGDSGGPLVCFEK' | 'Plasminogen' |
| 1559 | 'PLG_No121' | 'NGITCQK' | 'Plasminogen' |
| 1560 | 'PLG_No122' | 'AGLEKNYCRNPDGDVGGPWCYTTNPR' | 'Plasminogen' |
| 1561 | 'PLG_No123' | 'KCSGTEASVVAPPPVVLLPDVETPSEEDCMFGNGK' | 'Plasminogen' |
| 1562 | 'PLG_No124' | 'CSGTEASVVAPPPVVLLPDVETPSEEDCMFGNGK' | 'Plasminogen' |
| 1563 | 'PLG_No125' | 'TPENFPCKNLDENYCRNPDGK' | 'Plasminogen' |
| 1564 | 'PLG_No126' | 'RYDYCDILECEEECMHCSGENYDGKISK' | 'Plasminogen' |
| 1565 | 'PLG_No127' | 'TRFGMHFCGGTLISPEWVLTAAHCLEK' | 'Plasminogen' |
| 1566 | 'PLG_No128' | 'VSRFVTWIEGVMR' | 'Plasminogen' |
| 1567 | 'PLG_No129' | 'FVTWIEGVMRNN' | 'Plasminogen' |
| 1568 | 'PLG_No130' | 'FVTWIEGVMRNN' | 'Plasminogen' |
| 1569 | 'PLG_No131' | 'SPRPSSYK' | 'Plasminogen' |
| 1570 | 'PLG_No132' | 'LYDYCDVPQCAAPSFDCGK' | 'Plasminogen' |
| 1571 | 'F12_No1' | 'LHEAFSPVSYQHDLALLR' | 'Coagulation factor XII' |
| 1572 | 'F12_No2' | 'TTLSGAPCQPWASEATYR' | 'Coagulation factor XII' |
| 1573 | 'F12_No3' | 'GRPGPQPWCATTPNFDQDQR' | 'Coagulation factor XII' |
| 1574 | 'F12_No4' | 'NWGLGGHAFCR' | 'Coagulation factor XII' |
| 1575 | 'F12_No5' | 'LTLQGIISWGSGCGDR' | 'Coagulation factor XII' |
| 1576 | 'F12_No6' | 'NKPGVYTDVAYYLAWIR' | 'Coagulation factor XII' |
| 1577 | 'F12_No7' | 'VVGGLVALR' | 'Coagulation factor XII' |
| 1578 | 'F12_No8' | 'FFHKNEIWYR' | 'Coagulation factor XII' |
| 1579 | 'F12_No9' | 'NPDNDIRPWCFVLNR' | 'Coagulation factor XII' |
| 1580 | 'F12_No10' | 'RLTLQGIISWGSGCGDR' | 'Coagulation factor XII' |
| 1581 | 'F12_No11' | 'CFEPQLLR' | 'Coagulation factor XII' |
| 1582 | 'F12_No12' | 'EQPPSLTR' | 'Coagulation factor XII' |
| 1583 | 'F12_No13' | 'EKCFEPQLLR' | 'Coagulation factor XII' |
| 1584 | 'F12_No14' | 'WGYCLEPK' | 'Coagulation factor XII' |
| 1585 | 'F12_No15' | 'GGTCVNMPSGPHCLCPQHLTGNHCQK' | 'Coagulation factor XII' |

| | A | B | C |
|---|---|---|---|
| 1 | **Peptide Name** | **Peptide Sequence** | **Peptide Description** |
| 1586 | 'F12_No16' | 'PAPEDLTVVLGQER' | 'Coagulation factor XII' |
| 1587 | 'F12_No17' | 'AEEHTVVLTVTGEPCHFPFQYHR' | 'Coagulation factor XII' |
| 1588 | 'F12_No18' | 'YKAEEHTVVLTVTGEPCHFPFQYHR' | 'Coagulation factor XII' |
| 1589 | 'F12_No19' | 'NHSCEPCQTLAVR' | 'Coagulation factor XII' |
| 1590 | 'F12_No20' | 'GGTCVNMPSGPHCLCPQHLTGNHCQK' | 'Coagulation factor XII' |
| 1591 | 'F12_No21' | 'LCHCPVGYTGAFCDVDTK' | 'Coagulation factor XII' |
| 1592 | 'F12_No22' | 'NPDNDIRPWCFVLNRDR' | 'Coagulation factor XII' |
| 1593 | 'F12_No23' | 'RNHSCEPCQTLAVR' | 'Coagulation factor XII' |
| 1594 | 'F12_No24' | 'LQEDADGSCALLSPYVQPVCLPSGAAR' | 'Coagulation factor XII' |
| 1595 | 'F12_No25' | 'NKPGVYTDVAYYLAWIREHTVS' | 'Coagulation factor XII' |
| 1596 | 'F12_No26' | 'PGPQPWCATTPNFDQDQR' | 'Coagulation factor XII' |
| 1597 | 'F12_No27' | 'SYRLHEAFSPVSYQHDLALLR' | 'Coagulation factor XII' |
| 1598 | 'F12_No28' | 'FFHKNEIWYRTEQAAVAR' | 'Coagulation factor XII' |
| 1599 | 'F12_No29' | 'CSAPDVHGSSILPGMLCAGFLEGGTDACQGDSGGPLVCEDQAAER' | 'Coagulation factor XII' |
| 1600 | 'F12_No30' | 'NEIWYRTEQAAVAR' | 'Coagulation factor XII' |
| 1601 | 'F12_No31' | 'TNPCLHGGR' | 'Coagulation factor XII' |
| 1602 | 'F12_No32' | 'NGPLSCGQR' | 'Coagulation factor XII' |
| 1603 | 'F12_No33' | 'TEQAAVAR' | 'Coagulation factor XII' |
| 1604 | 'F12_No34' | 'LASQACR' | 'Coagulation factor XII' |
| 1605 | 'F12_No35' | 'ASCYDGR' | 'Coagulation factor XII' |
| 1606 | 'F12_No36' | 'WGYCLEPKK' | 'Coagulation factor XII' |
| 1607 | 'F12_No37' | 'ASCYDGRGLSYR' | 'Coagulation factor XII' |
| 1608 | 'F12_No38' | 'TPPQSQTPGALPAKR' | 'Coagulation factor XII' |
| 1609 | 'CFB_No1' | 'YGLVTYATYPK' | 'Complement factor B' |
| 1610 | 'CFB_No2' | 'DLEIEVVLFHPNYNINGK' | 'Complement factor B' |
| 1611 | 'CFB_No3' | 'VSEADSSNADWVTK' | 'Complement factor B' |
| 1612 | 'CFB_No4' | 'YGQTIRPICLPCTEGTTR' | 'Complement factor B' |
| 1613 | 'CFB_No5' | 'GHESCMGAVVSEYFVLTAAHCFTVDDKEHSIK' | 'Complement factor B' |
| 1614 | 'CFB_No6' | 'LPPTTTCQQQKEELLPAQDIK' | 'Complement factor B' |
| 1615 | 'CFB_No7' | 'VKDISEVVTPR' | 'Complement factor B' |
| 1616 | 'CFB_No8' | 'ALFVSEEEKK' | 'Complement factor B' |
| 1617 | 'CFB_No9' | 'EKLQDEDLGFL' | 'Complement factor B' |
| 1618 | 'CFB_No10' | 'PICLPCTEGTTR' | 'Complement factor B' |
| 1619 | 'CFB_No11' | 'KCLVNLIEK' | 'Complement factor B' |
| 1620 | 'CFB_No12' | 'LEDSVTYHCSR' | 'Complement factor B' |
| 1621 | 'CFB_No13' | 'LQDEDLGFL' | 'Complement factor B' |
| 1622 | 'CFB_No14' | 'CLVNLIEK' | 'Complement factor B' |
| 1623 | 'CFB_No15' | 'KEAGIPEFYDYDVALIK' | 'Complement factor B' |
| 1624 | 'CFB_No16' | 'QLNEINYEDHK' | 'Complement factor B' |
| 1625 | 'CFB_No17' | 'FLCTGGVSPYADPNTCRGDSGGPLIVHK' | 'Complement factor B' |
| 1626 | 'CFB_No18' | 'HVIILMTDGLHNMGGDPITVIDEIR' | 'Complement factor B' |
| 1627 | 'CFB_No19' | 'AIHCPRPHDFENGEYWPR' | 'Complement factor B' |
| 1628 | 'CFB_No20' | 'WSGQTAICDNGAGYCSNPGIPIGTRK' | 'Complement factor B' |
| 1629 | 'CFB_No21' | 'DFHINLFQVLPWLK' | 'Complement factor B' |

| | A | B | C |
|---|---|---|---|
| 1 | **Peptide Name** | **Peptide Sequence** | **Peptide Description** |
| 1630 | 'CFB_No22' | 'QLNEINYEDHKLK' | 'Complement factor B' |
| 1631 | 'CFB_No23' | 'DAQYAPGYDK' | 'Complement factor B' |
| 1632 | 'CFB_No24' | 'VASYGVKPR' | 'Complement factor B' |
| 1633 | 'CFB_No25' | 'EAGIPEFYDYDVALIK' | 'Complement factor B' |
| 1634 | 'CFB_No26' | 'GDSGGPLIVHK' | 'Complement factor B' |
| 1635 | 'CFB_No27' | 'FIQVGVISWGVVDVCK' | 'Complement factor B' |
| 1636 | 'CFB_No28' | 'NPREDYLDVYVFGVGPLVNQVNINALASKK' | 'Complement factor B' |
| 1637 | 'CFB_No29' | 'PHDFENGEYWPR' | 'Complement factor B' |
| 1638 | 'CFB_No30' | 'PQGSCSLEGVEIK' | 'Complement factor B' |
| 1639 | 'CFB_No31' | 'GHESCMGAVVSEYFVLTAAHCFTVDDKEHSIK' | 'Complement factor B' |
| 1640 | 'CFB_No32' | 'LLQEGQALEYVCPSGFYPYPVQTR' | 'Complement factor B' |
| 1641 | 'CFB_No33' | 'HVIILMTDGLHNMGGDPITVIDEIR' | 'Complement factor B' |
| 1642 | 'CFB_No34' | 'ISVIRPSK' | 'Complement factor B' |
| 1643 | 'CFB_No35' | 'QLNEINYEDHKLK' | 'Complement factor B' |
| 1644 | 'CFB_No36' | 'DAQYAPGYDKVK' | 'Complement factor B' |
| 1645 | 'CFB_No37' | 'FLCTGGVSPYADPNTCRGDSGGPLIVHKR' | 'Complement factor B' |
| 1646 | 'CFB_No38' | 'HVIILMTDGLHNMGGDPITVIDEIR' | 'Complement factor B' |
| 1647 | 'CFB_No39' | 'ALRLPPTTTCQQQKEELLPAQDIK' | 'Complement factor B' |
| 1648 | 'CFB_No40' | 'DLEIEVVLFHPNYNINGKK' | 'Complement factor B' |
| 1649 | 'CFB_No41' | 'DLLYIGKDR' | 'Complement factor B' |
| 1650 | 'CFB_No42' | 'STGSWSTLKTQDQK' | 'Complement factor B' |
| 1651 | 'CFB_No43' | 'LPPTTTCQQQK' | 'Complement factor B' |
| 1652 | 'CFB_No44' | 'FLCTGGVSPYADPNTCR' | 'Complement factor B' |
| 1653 | 'CFB_No45' | 'VGSQYRLEDSVTYHCSR' | 'Complement factor B' |
| 1654 | 'CFB_No46' | 'ALFVSEEEKKLTR' | 'Complement factor B' |
| 1655 | 'CFB_No47' | 'KVGSQYRLEDSVTYHCSR' | 'Complement factor B' |
| 1656 | 'CFB_No48' | 'HVIILMTDGLHNMGGDPITVIDEIR' | 'Complement factor B' |
| 1657 | 'CFB_No49' | 'GSCERDAQYAPGYDK' | 'Complement factor B' |
| 1658 | 'CFB_No50' | 'ALRLPPTTTCQQQK' | 'Complement factor B' |
| 1659 | 'CFB_No51' | 'DAQYAPGYDKVKDISEVVTPR' | 'Complement factor B' |
| 1660 | 'CFB_No52' | 'QLNEINYEDHK' | 'Complement factor B' |
| 1661 | 'CFB_No53' | 'LKYGQTIRPICLPCTEGTTR' | 'Complement factor B' |
| 1662 | 'CFB_No54' | 'KNPREDYLDVYVFGVGPLVNQVNINALASK' | 'Complement factor B' |
| 1663 | 'CFB_No55' | 'DLLYIGK' | 'Complement factor B' |
| 1664 | 'CFB_No56' | 'ALFVSEEEK' | 'Complement factor B' |
| 1665 | 'CFB_No57' | 'GDSGGPLIVHKR' | 'Complement factor B' |
| 1666 | 'CFB_No58' | 'RDLEIEVVLFHPNYNINGK' | 'Complement factor B' |
| 1667 | 'CFB_No59' | 'EAGIPEFYDYDVALIKLK' | 'Complement factor B' |
| 1668 | 'CFB_No60' | 'VSEADSSNADWVTKQLNEINYEDHK' | 'Complement factor B' |
| 1669 | 'CFB_No61' | 'GSCERDAQYAPGYDKVK' | 'Complement factor B' |
| 1670 | 'CFB_No62' | 'VSEADSSNADWVTKQLNEINYEDHKLK' | 'Complement factor B' |
| 1671 | 'CFB_No63' | 'GTDYHKQPWQAK' | 'Complement factor B' |
| 1672 | 'CFB_No64' | 'PQGSCSLEGVEIKGGSFR' | 'Complement factor B' |
| 1673 | 'CFB_No65' | 'GHESCMGAVVSEYFVLTAAHCFTVDDK' | 'Complement factor B' |

| | A | B | C |
|---|---|---|---|
| 1 | **Peptide Name** | **Peptide Sequence** | **Peptide Description** |
| 1674 | 'CFB_No66' | 'DNEQHVFK' | 'Complement factor B' |
| 1675 | 'CFB_No67' | 'STGSWSTLKTQDQKTVR' | 'Complement factor B' |
| 1676 | 'CFB_No68' | 'DISEVVTPR' | 'Complement factor B' |
| 1677 | 'CFB_No69' | 'CLVNLIEKVASYGVKPR' | 'Complement factor B' |
| 1678 | 'CFB_No70' | 'EELLPAQDIK' | 'Complement factor B' |
| 1679 | 'CFB_No71' | 'FIQVGVISWGVVDVCKNQK' | 'Complement factor B' |
| 1680 | 'CFB_No72' | 'EAGIPEFYDYDVALIKLKNK' | 'Complement factor B' |
| 1681 | 'CFB_No73' | 'IWVKVSEADSSNADWVTK' | 'Complement factor B' |
| 1682 | 'CFB_No74' | 'KDNEQHVFK' | 'Complement factor B' |
| 1683 | 'CFB_No75' | 'SRFIQVGVISWGVVDVCK' | 'Complement factor B' |
| 1684 | 'CFB_No76' | 'VSVGGEKR' | 'Complement factor B' |
| 1685 | 'CFB_No77' | 'DFHINLFQVLPWLKEK' | 'Complement factor B' |
| 1686 | 'CFB_No78' | 'EELLPAQDIKALFVSEEEKK' | 'Complement factor B' |
| 1687 | 'CFB_No79' | 'STGSWSTLK' | 'Complement factor B' |
| 1688 | 'CFB_No80' | 'TCRSTGSWSTLKTQDQK' | 'Complement factor B' |
| 1689 | 'CFB_No81' | 'KVGSQYR' | 'Complement factor B' |
| 1690 | 'CFB_No82' | 'QLNEINYEDHKLKSGTNTK' | 'Complement factor B' |
| 1691 | 'CA1_No1' | 'SLLSNVEGDNAVPMQHNNRPTQPLK' | 'Carbonic anhydrase 1' |
| 1692 | 'CA1_No2' | 'ADGLAVIGVLMK' | 'Carbonic anhydrase 1' |
| 1693 | 'CA1_No3' | 'LYPIANGNNQSPVDIK' | 'Carbonic anhydrase 1' |
| 1694 | 'CA1_No4' | 'HDTSLKPISVSYNPATAK' | 'Carbonic anhydrase 1' |
| 1695 | 'CA1_No5' | 'SLLSNVEGDNAVPMQHNNRPTQPLK' | 'Carbonic anhydrase 1' |
| 1696 | 'CA1_No6' | 'VLDALQAIK' | 'Carbonic anhydrase 1' |
| 1697 | 'CA1_No7' | 'ADGLAVIGVLMK' | 'Carbonic anhydrase 1' |
| 1698 | 'CA1_No8' | 'YSAELHVAHWNSAK' | 'Carbonic anhydrase 1' |
| 1699 | 'CA1_No9' | 'EIINVGHSFHVNFEDNDNR' | 'Carbonic anhydrase 1' |
| 1700 | 'CA1_No10' | 'ESISVSSEQLAQFR' | 'Carbonic anhydrase 1' |
| 1701 | 'CA1_No11' | 'ASPDWGYDDKNGPEQWSK' | 'Carbonic anhydrase 1' |
| 1702 | 'CA1_No12' | 'TSETKHDTSLKPISVSYNPATAK' | 'Carbonic anhydrase 1' |
| 1703 | 'CA1_No13' | 'YSSLAEAASKADGLAVIGVLMK' | 'Carbonic anhydrase 1' |
| 1704 | 'CA1_No14' | 'EIINVGHSFHVNFEDNDNRSVLK' | 'Carbonic anhydrase 1' |
| 1705 | 'CA2_No1' | 'YDPSLKPLSVSYDQATSLR' | 'Carbonic anhydrase 2' |
| 1706 | 'CA2_No2' | 'QSPVDIDTHTAK' | 'Carbonic anhydrase 2' |
| 1707 | 'CA2_No3' | 'VVDVLDSIK' | 'Carbonic anhydrase 2' |
| 1708 | 'CA2_No4' | 'LNFNGEGEPEELMVDNWRPAQPLK' | 'Carbonic anhydrase 2' |
| 1709 | 'CA2_No5' | 'SADFTNFDPR' | 'Carbonic anhydrase 2' |
| 1710 | 'CA2_No6' | 'YAAELHLVHWNTK' | 'Carbonic anhydrase 2' |
| 1711 | 'CA2_No7' | 'ILNNGHAFNVEFDDSQDKAVLK' | 'Carbonic anhydrase 2' |
| 1712 | 'CA2_No8' | 'AVQQPDGLAVLGIFLK' | 'Carbonic anhydrase 2' |
| 1713 | 'CA2_No9' | 'HNGPEHWHKDFPIAK' | 'Carbonic anhydrase 2' |
| 1714 | 'CA2_No10' | 'QSPVDIDTHTAKYDPSLKPLSVSYDQATSLR' | 'Carbonic anhydrase 2' |
| 1715 | 'CA2_No11' | 'QSPVDIDTHTAK' | 'Carbonic anhydrase 2' |
| 1716 | 'CA2_No12' | 'KYAAELHLVHWNTK' | 'Carbonic anhydrase 2' |
| 1717 | 'CA2_No13' | 'KLNFNGEGEPEELMVDNWRPAQPLK' | 'Carbonic anhydrase 2' |

| | A | B | C |
|---|---|---|---|
| 1 | **Peptide Name** | **Peptide Sequence** | **Peptide Description** |
| 1718 | 'CA2_No14' | 'KLNFNGEGEPEELMVDNWRPAQPLK' | 'Carbonic anhydrase 2' |
| 1719 | 'SERPINC1_No1' | 'ANRPFLVFIR' | 'Antithrombin-III' |
| 1720 | 'SERPINC1_No2' | 'SKLPGIVAEGRDDLYVSDAFHK' | 'Antithrombin-III' |
| 1721 | 'SERPINC1_No3' | 'DIPMNPMCIYR' | 'Antithrombin-III' |
| 1722 | 'SERPINC1_No4' | 'FATTFYQHLADSK' | 'Antithrombin-III' |
| 1723 | 'SERPINC1_No5' | 'TSDQIHFFFAK' | 'Antithrombin-III' |
| 1724 | 'SERPINC1_No6' | 'DDLYVSDAFHK' | 'Antithrombin-III' |
| 1725 | 'SERPINC1_No7' | 'ATEDEGSEQKIPEATNR' | 'Antithrombin-III' |
| 1726 | 'SERPINC1_No8' | 'LPGIVAEGRDDLYVSDAFHK' | 'Antithrombin-III' |
| 1727 | 'SERPINC1_No9' | 'SKLPGIVAEGR' | 'Antithrombin-III' |
| 1728 | 'SERPINC1_No10' | 'LPGIVAEGR' | 'Antithrombin-III' |
| 1729 | 'SERPINC1_No11' | 'LQPLDFKENAEQSR' | 'Antithrombin-III' |
| 1730 | 'SERPINC1_No12' | 'IEDGFSLK' | 'Antithrombin-III' |
| 1731 | 'SERPINC1_No13' | 'LQPLDFK' | 'Antithrombin-III' |
| 1732 | 'SERPINC1_No14' | 'GDDITMVLILPKPEK' | 'Antithrombin-III' |
| 1733 | 'SERPINC1_No15' | 'GDDITMVLILPKPEK' | 'Antithrombin-III' |
| 1734 | 'SERPINC1_No16' | 'EVPLNTIIFMGR' | 'Antithrombin-III' |
| 1735 | 'SERPINC1_No17' | 'ITDVIPSEAINELTVLVLVNTIYFK' | 'Antithrombin-III' |
| 1736 | 'SERPINC1_No18' | 'FSPENTR' | 'Antithrombin-III' |
| 1737 | 'SERPINC1_No19' | 'KATEDEGSEQKIPEATNR' | 'Antithrombin-III' |
| 1738 | 'SERPINC1_No20' | 'DIPMNPMCIYR' | 'Antithrombin-III' |
| 1739 | 'SERPINC1_No21' | 'AFLEVNEEGSEAAASTAVVIAGR' | 'Antithrombin-III' |
| 1740 | 'SERPINC1_No22' | 'ELFYKADGESCSASMMYQEGK' | 'Antithrombin-III' |
| 1741 | 'SERPINC1_No23' | 'DIPMNPMCIYR' | 'Antithrombin-III' |
| 1742 | 'SERPINC1_No24' | 'ELFYKADGESCSASMMYQEGK' | 'Antithrombin-III' |
| 1743 | 'SERPINC1_No25' | 'ELFYKADGESCSASMMYQEGK' | 'Antithrombin-III' |
| 1744 | 'SERPINC1_No26' | 'RVWELSK' | 'Antithrombin-III' |
| 1745 | 'SERPINC1_No27' | 'EQLQDMGLVDLFSPEK' | 'Antithrombin-III' |
| 1746 | 'SERPINC1_No28' | 'FRIEDGFSLK' | 'Antithrombin-III' |
| 1747 | 'SERPINC1_No29' | 'ATEDEGSEQKIPEATNRR' | 'Antithrombin-III' |
| 1748 | 'SERPINC1_No30' | 'EVPLNTIIFMGR' | 'Antithrombin-III' |
| 1749 | 'SERPINC1_No31' | 'FATTFYQHLADSKNDNDNIFLSPLSISTAFAMTK' | 'Antithrombin-III' |
| 1750 | 'SERPINC1_No32' | 'IEDGFSLKEQLQDMGLVDLFSPEK' | 'Antithrombin-III' |
| 1751 | 'SERPINC1_No33' | 'PFLVFIR' | 'Antithrombin-III' |
| 1752 | 'SERPINC1_No34' | 'ADGESCSASMMYQEGK' | 'Antithrombin-III' |
| 1753 | 'SERPINC1_No35' | 'VAEGTQVLELPFKGDDITMVLILPKPEK' | 'Antithrombin-III' |
| 1754 | 'SERPINC1_No36' | 'ELFYKADGESCSASMMYQEGKFR' | 'Antithrombin-III' |
| 1755 | 'SERPINC1_No37' | 'VAEGTQVLELPFK' | 'Antithrombin-III' |
| 1756 | 'SERPINC1_No38' | 'EQLQDMGLVDLFSPEK' | 'Antithrombin-III' |
| 1757 | 'SERPINC1_No39' | 'FATTFYQHLADSKNDNDNIFLSPLSISTAFAMTK' | 'Antithrombin-III' |
| 1758 | 'SERPINC1_No40' | 'ADGESCSASMMYQEGK' | 'Antithrombin-III' |
| 1759 | 'SERPINC1_No41' | 'ADGESCSASMMYQEGK' | 'Antithrombin-III' |
| 1760 | 'SERPINC1_No42' | 'ADGESCSASMMYQEGKFR' | 'Antithrombin-III' |
| 1761 | 'SERPINC1_No43' | 'VAEGTQVLELPFKGDDITMVLILPKPEK' | 'Antithrombin-III' |

| | A | B | C |
|---|---|---|---|
| 1 | **Peptide Name** | **Peptide Sequence** | **Peptide Description** |
| 1762 | 'SERPINC1_No44' | 'ADGESCSASMMYQEGK' | 'Antithrombin-III' |
| 1763 | 'SERPINC1_No45' | 'FSPENTRK' | 'Antithrombin-III' |
| 1764 | 'SERPINC1_No46' | 'KELFYKADGESCSASMMYQEGK' | 'Antithrombin-III' |
| 1765 | 'SERPINC1_No47' | 'SLNPNRVTFK' | 'Antithrombin-III' |
| 1766 | 'SERPINC1_No48' | 'NDNDNIFLSPLSISTAFAMTK' | 'Antithrombin-III' |
| 1767 | 'SERPINC1_No49' | 'RVAEGTQVLELPFK' | 'Antithrombin-III' |
| 1768 | 'SERPINC1_No50' | 'NDNDNIFLSPLSISTAFAMTK' | 'Antithrombin-III' |
| 1769 | 'SERPINC1_No51' | 'ADGESCSASMMYQEGKFR' | 'Antithrombin-III' |
| 1770 | 'SERPINC1_No52' | 'FDTISEK' | 'Antithrombin-III' |
| 1771 | 'SERPINC1_No53' | 'IPEATNR' | 'Antithrombin-III' |
| 1772 | 'SERPINC1_No54' | 'DIPMNPMCIYRSPEKK' | 'Antithrombin-III' |
| 1773 | 'SERPINC1_No55' | 'ADGESCSASMMYQEGKFR' | 'Antithrombin-III' |
| 1774 | 'SERPINC1_No56' | 'SKFSPENTR' | 'Antithrombin-III' |
| 1775 | 'SERPINC1_No57' | 'ADGESCSASMMYQEGKFR' | 'Antithrombin-III' |
| 1776 | 'SERPINC1_No58' | 'IEDGFSLKEQLQDMGLVDLFSPEK' | 'Antithrombin-III' |
| 1777 | 'SERPINC1_No59' | 'LVSANRLFGDK' | 'Antithrombin-III' |
| 1778 | 'SERPINC1_No60' | 'DIPMNPMCIYRSPEK' | 'Antithrombin-III' |
| 1779 | 'SERPINC1_No61' | 'KELFYKADGESCSASMMYQEGK' | 'Antithrombin-III' |
| 1780 | 'SERPINC1_No62' | 'ELFYKADGESCSASMMYQEGK' | 'Antithrombin-III' |
| 1781 | 'SERPINC1_No63' | 'DIPMNPMCIYRSPEKK' | 'Antithrombin-III' |
| 1782 | 'SERPINC1_No64' | 'EQLQDMGLVDLFSPEKSK' | 'Antithrombin-III' |
| 1783 | 'SERPINC1_No65' | 'FSPENTRKELFYK' | 'Antithrombin-III' |
| 1784 | 'SERPINC1_No66' | 'DIPMNPMCIYRSPEKK' | 'Antithrombin-III' |
| 1785 | 'SERPINC1_No67' | 'TSDQIHFFFAKLNCR' | 'Antithrombin-III' |
| 1786 | 'SERPINC1_No68' | 'AFLEVNEEGSEAAASTAVVIAGRSLNPNR' | 'Antithrombin-III' |
| 1787 | 'SERPINC1_No69' | 'FDTISEKTSDQIHFFFAK' | 'Antithrombin-III' |
| 1788 | 'SERPINC1_No70' | 'ELFYKADGESCSASMMYQEGKFR' | 'Antithrombin-III' |
| 1789 | 'SERPINC1_No71' | 'DIPMNPMCIYRSPEK' | 'Antithrombin-III' |
| 1790 | 'SERPINC1_No72' | 'EQLQDMGLVDLFSPEKSK' | 'Antithrombin-III' |
| 1791 | 'SERPINC1_No73' | 'ANSRFATTFYQHLADSK' | 'Antithrombin-III' |
| 1792 | 'SERPINC1_No74' | 'DIPMNPMCIYRSPEKK' | 'Antithrombin-III' |
| 1793 | 'SERPINA1_No1' | 'LYHSEAFTVNFGDTEEAKK' | 'Alpha-1-antitrypsin' |
| 1794 | 'SERPINA1_No2' | 'FNKPFVFLMIEQNTK' | 'Alpha-1-antitrypsin' |
| 1795 | 'SERPINA1_No3' | 'LSITGTYDLK' | 'Alpha-1-antitrypsin' |
| 1796 | 'SERPINA1_No4' | 'LVDKFLEDVKK' | 'Alpha-1-antitrypsin' |
| 1797 | 'SERPINA1_No5' | 'LQHLENELTHDIITK' | 'Alpha-1-antitrypsin' |
| 1798 | 'SERPINA1_No6' | 'KLSSWVLLMK' | 'Alpha-1-antitrypsin' |
| 1799 | 'SERPINA1_No7' | 'VFSNGADLSGVTEEAPLKLSK' | 'Alpha-1-antitrypsin' |
| 1800 | 'SERPINA1_No8' | 'SVLGQLGITK' | 'Alpha-1-antitrypsin' |
| 1801 | 'SERPINA1_No9' | 'KLSSWVLLMK' | 'Alpha-1-antitrypsin' |
| 1802 | 'SERPINA1_No10' | 'TLNQPDSQLQLTTGNGLFLSEGLK' | 'Alpha-1-antitrypsin' |
| 1803 | 'SERPINA1_No11' | 'LSSWVLLMK' | 'Alpha-1-antitrypsin' |
| 1804 | 'SERPINA1_No12' | 'LGMFNIQHCK' | 'Alpha-1-antitrypsin' |
| 1805 | 'SERPINA1_No13' | 'LGMFNIQHCK' | 'Alpha-1-antitrypsin' |

| | A | B | C |
|---|---|---|---|
| 1 | **Peptide Name** | **Peptide Sequence** | **Peptide Description** |
| 1806 | 'SERPINA1_No14' | 'LVDKFLEDVK' | 'Alpha-1-antitrypsin' |
| 1807 | 'SERPINA1_No15' | 'QINDYVEK' | 'Alpha-1-antitrypsin' |
| 1808 | 'SERPINA1_No16' | 'RLGMFNIQHCK' | 'Alpha-1-antitrypsin' |
| 1809 | 'SERPINA1_No17' | 'WERPFEVK' | 'Alpha-1-antitrypsin' |
| 1810 | 'SERPINA1_No18' | 'FLENEDRR' | 'Alpha-1-antitrypsin' |
| 1811 | 'SERPINA1_No19' | 'FLEDVKK' | 'Alpha-1-antitrypsin' |
| 1812 | 'SERPINA1_No20' | 'SASLHLPK' | 'Alpha-1-antitrypsin' |
| 1813 | 'SERPINA1_No21' | 'RLGMFNIQHCK' | 'Alpha-1-antitrypsin' |
| 1814 | 'SERPINA1_No22' | 'GTEAAGAMFLEAIPMSIPPEVK' | 'Alpha-1-antitrypsin' |
| 1815 | 'SERPINA1_No23' | 'LSSWVLLMK' | 'Alpha-1-antitrypsin' |
| 1816 | 'SERPINA1_No24' | 'DTEEEDFHVDQVTTVK' | 'Alpha-1-antitrypsin' |
| 1817 | 'SERPINA1_No25' | 'KQINDYVEK' | 'Alpha-1-antitrypsin' |
| 1818 | 'SERPINA1_No26' | 'GKWERPFEVK' | 'Alpha-1-antitrypsin' |
| 1819 | 'SERPINA1_No27' | 'GTEAAGAMFLEAIPMSIPPEVK' | 'Alpha-1-antitrypsin' |
| 1820 | 'SERPINA1_No28' | 'AVLTIDEKGTEAAGAMFLEAIPMSIPPEVK' | 'Alpha-1-antitrypsin' |
| 1821 | 'SERPINA1_No29' | 'QINDYVEKGTQGK' | 'Alpha-1-antitrypsin' |
| 1822 | 'SERPINA1_No30' | 'ELDRDTVFALVNYIFFK' | 'Alpha-1-antitrypsin' |
| 1823 | 'SERPINA1_No31' | 'LGMFNIQHCKK' | 'Alpha-1-antitrypsin' |
| 1824 | 'SERPINA1_No32' | 'GTEAAGAMFLEAIPMSIPPEVK' | 'Alpha-1-antitrypsin' |
| 1825 | 'SERPINA1_No33' | 'WERPFEVKDTEEEDFHVDQVTTVK' | 'Alpha-1-antitrypsin' |
| 1826 | 'SERPINA1_No34' | 'PFEVKDTEEEDFHVDQVTTVK' | 'Alpha-1-antitrypsin' |
| 1827 | 'SERPINA1_No35' | 'AVLTIDEK' | 'Alpha-1-antitrypsin' |
| 1828 | 'SERPINA1_No36' | 'SASLHLPKLSITGTYDLKSVLGQLGITK' | 'Alpha-1-antitrypsin' |
| 1829 | 'SERPINA1_No37' | 'FLENEDR' | 'Alpha-1-antitrypsin' |
| 1830 | 'SERPINA1_No38' | 'KLYHSEAFTVNFGDTEEAKK' | 'Alpha-1-antitrypsin' |
| 1831 | 'SERPINA1_No39' | 'ITPNLAEFAFSLYR' | 'Alpha-1-antitrypsin' |
| 1832 | 'SERPINA1_No40' | 'LSITGTYDLKSVLGQLGITK' | 'Alpha-1-antitrypsin' |
| 1833 | 'SERPINA1_No41' | 'SVLGQLGITKVFSNGADLSGVTEEAPLK' | 'Alpha-1-antitrypsin' |
| 1834 | 'SERPINA1_No42' | 'LQHLENELTHDIITKFLENEDRR' | 'Alpha-1-antitrypsin' |
| 1835 | 'SERPINA1_No43' | 'AVLTIDEKGTEAAGAMFLEAIPMSIPPEVK' | 'Alpha-1-antitrypsin' |
| 1836 | 'SERPINA1_No44' | 'FNKPFVFLMIEQNTKSPLFMGK' | 'Alpha-1-antitrypsin' |
| 1837 | 'SERPINA1_No45' | 'KLYHSEAFTVNFGDTEEAK' | 'Alpha-1-antitrypsin' |
| 1838 | 'SERPINA1_No46' | 'YLGNATAIFFLPDEGK' | 'Alpha-1-antitrypsin' |
| 1839 | 'SERPINA1_No47' | 'FNKPFVFLMIEQNTKSPLFMGK' | 'Alpha-1-antitrypsin' |
| 1840 | 'SERPINA1_No48' | 'AVLTIDEKGTEAAGAMFLEAIPMSIPPEVK' | 'Alpha-1-antitrypsin' |
| 1841 | 'SERPINA1_No49' | 'FNKPFVFLMIEQNTKSPLFMGK' | 'Alpha-1-antitrypsin' |
| 1842 | 'SERPINA1_No50' | 'LGMFNIQHCKK' | 'Alpha-1-antitrypsin' |
| 1843 | 'SERPINA1_No51' | 'PFVFLMIEQNTK' | 'Alpha-1-antitrypsin' |
| 1844 | 'SERPINA1_No52' | 'DTVFALVNYIFFK' | 'Alpha-1-antitrypsin' |
| 1845 | 'SERPINA1_No53' | 'LYHSEAFTVNFGDTEEAKKQINDYVEK' | 'Alpha-1-antitrypsin' |
| 1846 | 'SERPINA1_No54' | 'TDTSHHDQDHPTFNK' | 'Alpha-1-antitrypsin' |
| 1847 | 'SERPINA1_No55' | 'VFSNGADLSGVTEEAPLKLSKAVHK' | 'Alpha-1-antitrypsin' |
| 1848 | 'SERPINA1_No56' | 'TLNQPDSQLQLTTGNGLFLSEGLKLVDK' | 'Alpha-1-antitrypsin' |
| 1849 | 'SERPINA1_No57' | 'RLGMFNIQHCKK' | 'Alpha-1-antitrypsin' |

| | A | B | C |
|---|---|---|---|
| 1 | **Peptide Name** | **Peptide Sequence** | **Peptide Description** |
| 1850 | 'SERPINA1_No58' | 'IVDLVKELDR' | 'Alpha-1-antitrypsin' |
| 1851 | 'SERPINA1_No59' | 'DTEEEDFHVDQVTTVKVPMMK' | 'Alpha-1-antitrypsin' |
| 1852 | 'SERPINA1_No60' | 'LYHSEAFTVNFGDTEEAK' | 'Alpha-1-antitrypsin' |
| 1853 | 'SERPINA1_No61' | 'GTEAAGAMFLEAIPMSIPPEVK' | 'Alpha-1-antitrypsin' |
| 1854 | 'SERPINA1_No62' | 'AVHKAVLTIDEKGTEAAGAMFLEAIPMSIPPEVK' | 'Alpha-1-antitrypsin' |
| 1855 | 'SERPINA1_No63' | 'KQINDYVEKGTQGK' | 'Alpha-1-antitrypsin' |
| 1856 | 'SERPINA1_No64' | 'SVLGQLGITKVFSNGADLSGVTEEAPLKLSK' | 'Alpha-1-antitrypsin' |
| 1857 | 'SERPINA1_No65' | 'FNKPFVFLMIEQNTK' | 'Alpha-1-antitrypsin' |
| 1858 | 'SERPINA1_No66' | 'AVLTIDEKGTEAAGAMFLEAIPMSIPPEVK' | 'Alpha-1-antitrypsin' |
| 1859 | 'SERPINA1_No67' | 'QINDYVEK' | 'Alpha-1-antitrypsin' |
| 1860 | 'SERPINA1_No68' | 'PFVFLMIEQNTK' | 'Alpha-1-antitrypsin' |
| 1861 | 'SERPINA1_No69' | 'VFSNGADLSGVTEEAPLK' | 'Alpha-1-antitrypsin' |
| 1862 | 'SERPINA1_No70' | 'TDTSHHDQDHPTFNKITPNLAEFAFSLYR' | 'Alpha-1-antitrypsin' |
| 1863 | 'SERPINA1_No71' | 'QINDYVEKGTQGK' | 'Alpha-1-antitrypsin' |
| 1864 | 'SERPINA1_No72' | 'FNKPFVFLMIEQNTKSPLFMGK' | 'Alpha-1-antitrypsin' |
| 1865 | 'SERPINA1_No73' | 'LQHLENELTHDIITKFLENEDR' | 'Alpha-1-antitrypsin' |
| 1866 | 'SERPINA1_No74' | 'DTEEEDFHVDQVTTVKVPMMK' | 'Alpha-1-antitrypsin' |
| 1867 | 'SERPINA1_No75' | 'QINDYVEKGTQGKIVDLVK' | 'Alpha-1-antitrypsin' |
| 1868 | 'SERPINA1_No76' | 'SASLHLPKLSITGTYDLK' | 'Alpha-1-antitrypsin' |
| 1869 | 'SERPINA1_No77' | 'QINDYVEKGTQGKIVDLVK' | 'Alpha-1-antitrypsin' |
| 1870 | 'SERPINA1_No78' | 'DTEEEDFHVDQVTTVKVPMMK' | 'Alpha-1-antitrypsin' |
| 1871 | 'SERPINA1_No79' | 'DTEEEDFHVDQVTTVKVPMMK' | 'Alpha-1-antitrypsin' |
| 1872 | 'SERPINA1_No80' | 'SPLFMGKVVNPTQK' | 'Alpha-1-antitrypsin' |
| 1873 | 'SERPINA1_No81' | 'AVHKAVLTIDEK' | 'Alpha-1-antitrypsin' |
| 1874 | 'SERPINA1_No82' | 'DTEEEDFHVDQVTTVKVPMMKR' | 'Alpha-1-antitrypsin' |
| 1875 | 'SERPINA1_No83' | 'FLENEDRRSASLHLPK' | 'Alpha-1-antitrypsin' |
| 1876 | 'SERPINA1_No84' | 'GTQGKIVDLVK' | 'Alpha-1-antitrypsin' |
| 1877 | 'SERPINA1_No85' | 'DTEEEDFHVDQVTTVKVPMMKR' | 'Alpha-1-antitrypsin' |
| 1878 | 'SERPINA1_No86' | 'SPLFMGKVVNPTQK' | 'Alpha-1-antitrypsin' |
| 1879 | 'SERPINA3_No1' | 'ITLLSALVETR' | 'Alpha-1-antichymotrypsin' |
| 1880 | 'SERPINA3_No2' | 'EQLSLLDRFTEDAKR' | 'Alpha-1-antichymotrypsin' |
| 1881 | 'SERPINA3_No3' | 'AVLDVFEEGTEASAATAVK' | 'Alpha-1-antichymotrypsin' |
| 1882 | 'SERPINA3_No4' | 'LYGSEAFATDFQDSAAAK' | 'Alpha-1-antichymotrypsin' |
| 1883 | 'SERPINA3_No5' | 'EQLSLLDRFTEDAK' | 'Alpha-1-antichymotrypsin' |
| 1884 | 'SERPINA3_No6' | 'LYGSEAFATDFQDSAAAKK' | 'Alpha-1-antichymotrypsin' |
| 1885 | 'SERPINA3_No7' | 'FNRPFLMIIVPTDTQNIFFMSK' | 'Alpha-1-antichymotrypsin' |
| 1886 | 'SERPINA3_No8' | 'AKWEMPFDPQDTHQSR' | 'Alpha-1-antichymotrypsin' |
| 1887 | 'SERPINA3_No9' | 'ADLSGITGAR' | 'Alpha-1-antichymotrypsin' |
| 1888 | 'SERPINA3_No10' | 'WEMPFDPQDTHQSR' | 'Alpha-1-antichymotrypsin' |
| 1889 | 'SERPINA3_No11' | 'MEEVEAMLLPETLKR' | 'Alpha-1-antichymotrypsin' |
| 1890 | 'SERPINA3_No12' | 'WEMPFDPQDTHQSR' | 'Alpha-1-antichymotrypsin' |
| 1891 | 'SERPINA3_No13' | 'MEEVEAMLLPETLKR' | 'Alpha-1-antichymotrypsin' |
| 1892 | 'SERPINA3_No14' | 'KLINDYVK' | 'Alpha-1-antichymotrypsin' |
| 1893 | 'SERPINA3_No15' | 'MEEVEAMLLPETLKR' | 'Alpha-1-antichymotrypsin' |

| | A | B | C |
|---|---|---|---|
| 1 | **Peptide Name** | **Peptide Sequence** | **Peptide Description** |
| 1894 | 'SERPINA3_No16' | 'DYNLNDILLQLGIEEAFTSK' | 'Alpha-1-antichymotrypsin' |
| 1895 | 'SERPINA3_No17' | 'AKWEMPFDPQDTHQSR' | 'Alpha-1-antichymotrypsin' |
| 1896 | 'SERPINA3_No18' | 'MEEVEAMLLPETLK' | 'Alpha-1-antichymotrypsin' |
| 1897 | 'SERPINA3_No19' | 'GKITDLIK' | 'Alpha-1-antichymotrypsin' |
| 1898 | 'SERPINA3_No20' | 'DLDSQTMMVLVNYIFFK' | 'Alpha-1-antichymotrypsin' |
| 1899 | 'SERPINA3_No21' | 'WVMVPMMSLHHLTIPYFRDEELSCTVVELK' | 'Alpha-1-antichymotrypsin' |
| 1900 | 'SERPINA3_No22' | 'GTHVDLGLASANVDFAFSLYK' | 'Alpha-1-antichymotrypsin' |
| 1901 | 'SERPINA3_No23' | 'WVMVPMMSLHHLTIPYFRDEELSCTVVELK' | 'Alpha-1-antichymotrypsin' |
| 1902 | 'SERPINA3_No24' | 'FNRPFLMIIVPTDTQNIFFMSK' | 'Alpha-1-antichymotrypsin' |
| 1903 | 'SERPINA3_No25' | 'WVMVPMMSLHHLTIPYFRDEELSCTVVELK' | 'Alpha-1-antichymotrypsin' |
| 1904 | 'SERPINA3_No26' | 'WVMVPMMSLHHLTIPYFRDEELSCTVVELK' | 'Alpha-1-antichymotrypsin' |
| 1905 | 'SERPINA3_No27' | 'WVMVPMMSLHHLTIPYFRDEELSCTVVELK' | 'Alpha-1-antichymotrypsin' |
| 1906 | 'SERPINA3_No28' | 'DEELSCTVVELK' | 'Alpha-1-antichymotrypsin' |
| 1907 | 'SERPINA3_No29' | 'FNRPFLMIIVPTDTQNIFFMSK' | 'Alpha-1-antichymotrypsin' |
| 1908 | 'SERPINA3_No30' | 'WVMVPMMSLHHLTIPYFR' | 'Alpha-1-antichymotrypsin' |
| 1909 | 'SERPINA3_No31' | 'WVMVPMMSLHHLTIPYFRDEELSCTVVELK' | 'Alpha-1-antichymotrypsin' |
| 1910 | 'SERPINA3_No32' | 'MEEVEAMLLPETLKR' | 'Alpha-1-antichymotrypsin' |
| 1911 | 'SERPINA3_No33' | 'NLAVSQVVHK' | 'Alpha-1-antichymotrypsin' |
| 1912 | 'SERPINA3_No34' | 'WVMVPMMSLHHLTIPYFR' | 'Alpha-1-antichymotrypsin' |
| 1913 | 'SERPINA3_No35' | 'WVMVPMMSLHHLTIPYFR' | 'Alpha-1-antichymotrypsin' |
| 1914 | 'SERPINA3_No36' | 'WRDSLEFREIGELYLPK' | 'Alpha-1-antichymotrypsin' |
| 1915 | 'SERPINA3_No37' | 'FNRPFLMIIVPTDTQNIFFMSK' | 'Alpha-1-antichymotrypsin' |
| 1916 | 'SERPINA3_No38' | 'WVMVPMMSLHHLTIPYFR' | 'Alpha-1-antichymotrypsin' |
| 1917 | 'SERPINA3_No39' | 'MEEVEAMLLPETLK' | 'Alpha-1-antichymotrypsin' |
| 1918 | 'SERPINA3_No40' | 'WVMVPMMSLHHLTIPYFRDEELSCTVVELK' | 'Alpha-1-antichymotrypsin' |
| 1919 | 'SERPINA3_No41' | 'WVMVPMMSLHHLTIPYFR' | 'Alpha-1-antichymotrypsin' |
| 1920 | 'SERPINA3_No42' | 'NLAVSQVVHKAVLDVFEEGTEASAATAVK' | 'Alpha-1-antichymotrypsin' |
| 1921 | 'SERPINA3_No43' | 'DLDSQTMMVLVNYIFFK' | 'Alpha-1-antichymotrypsin' |
| 1922 | 'SERPINA3_No44' | 'EIGELYLPK' | 'Alpha-1-antichymotrypsin' |
| 1923 | 'SERPINA3_No45' | 'WRDSLEFR' | 'Alpha-1-antichymotrypsin' |
| 1924 | 'SERPINA3_No46' | 'RLYGSEAFATDFQDSAAAKK' | 'Alpha-1-antichymotrypsin' |
| 1925 | 'SERPINA3_No47' | 'EQLSLLDR' | 'Alpha-1-antichymotrypsin' |
| 1926 | 'SERPINA3_No48' | 'MEEVEAMLLPETLK' | 'Alpha-1-antichymotrypsin' |
| 1927 | 'SERPINA3_No49' | 'LINDYVK' | 'Alpha-1-antichymotrypsin' |
| 1928 | 'SERPINA3_No50' | 'MEEVEAMLLPETLK' | 'Alpha-1-antichymotrypsin' |
| 1929 | 'SERPINA3_No51' | 'WVMVPMMSLHHLTIPYFRDEELSCTVVELK' | 'Alpha-1-antichymotrypsin' |
| 1930 | 'SERPINA3_No52' | 'TLNQSSDELQLSMGNAMFVK' | 'Alpha-1-antichymotrypsin' |
| 1931 | 'SERPINA3_No53' | 'RLYGSEAFATDFQDSAAAK' | 'Alpha-1-antichymotrypsin' |
| 1932 | 'SERPINA3_No54' | 'WVMVPMMSLHHLTIPYFR' | 'Alpha-1-antichymotrypsin' |
| 1933 | 'SERPINA3_No55' | 'DSLEFREIGELYLPK' | 'Alpha-1-antichymotrypsin' |
| 1934 | 'SERPINA3_No56' | 'DLDSQTMMVLVNYIFFK' | 'Alpha-1-antichymotrypsin' |
| 1935 | 'SERPINA3_No57' | 'WVMVPMMSLHHLTIPYFR' | 'Alpha-1-antichymotrypsin' |
| 1936 | 'SERPINA3_No58' | 'TLNQSSDELQLSMGNAMFVK' | 'Alpha-1-antichymotrypsin' |
| 1937 | 'SERPINA3_No59' | 'FTEDAKR' | 'Alpha-1-antichymotrypsin' |

| | A | B | C |
|---|---|---|---|
| 1 | **Peptide Name** | **Peptide Sequence** | **Peptide Description** |
| 1938 | 'SERPINA3_No60' | 'TLNQSSDELQLSMGNAMFVK' | 'Alpha-1-antichymotrypsin' |
| 1939 | 'SERPINA3_No61' | 'TLNQSSDELQLSMGNAMFVK' | 'Alpha-1-antichymotrypsin' |
| 1940 | 'SERPINA3_No62' | 'ITLLSALVETRTIVR' | 'Alpha-1-antichymotrypsin' |
| 1941 | 'SERPINA3_No63' | 'RWRDSLEFR' | 'Alpha-1-antichymotrypsin' |
| 1942 | 'SERPINA3_No64' | 'QLVLKAPDK' | 'Alpha-1-antichymotrypsin' |
| 1943 | 'SERPINA3_No65' | 'QLVLKAPDK' | 'Alpha-1-antichymotrypsin' |
| 1944 | 'SERPINA3_No66' | 'FNLTETSEAEIHQSFQHLLR' | 'Alpha-1-antichymotrypsin' |
| 1945 | 'SERPINA3_No67' | 'AKWEMPFDPQDTHQSRFYLSK' | 'Alpha-1-antichymotrypsin' |
| 1946 | 'SERPINA3_No68' | 'WEMPFDPQDTHQSRFYLSK' | 'Alpha-1-antichymotrypsin' |
| 1947 | 'SERPINA3_No69' | 'EIGELYLPKFSISR' | 'Alpha-1-antichymotrypsin' |
| 1948 | 'SERPINA3_No70' | 'ADLSGITGARNLAVSQVVHKAVLDVFEEGTEASAATAVK' | 'Alpha-1-antichymotrypsin' |
| 1949 | 'AGT_No1' | 'SLDFTELDVAAEK' | 'Angiotensinogen' |
| 1950 | 'AGT_No2' | 'QPFVQGLALYTPVVLPR' | 'Angiotensinogen' |
| 1951 | 'AGT_No3' | 'SLDFTELDVAAEKIDR' | 'Angiotensinogen' |
| 1952 | 'AGT_No4' | 'ALQDQLVLVAAK' | 'Angiotensinogen' |
| 1953 | 'AGT_No5' | 'FMQAVTGWK' | 'Angiotensinogen' |
| 1954 | 'AGT_No6' | 'LQAILGVPWK' | 'Angiotensinogen' |
| 1955 | 'AGT_No7' | 'IDRFMQAVTGWK' | 'Angiotensinogen' |
| 1956 | 'AGT_No8' | 'PKDPTFIPAPIQAK' | 'Angiotensinogen' |
| 1957 | 'AGT_No9' | 'VLSALQAVQGLLVAQGR' | 'Angiotensinogen' |
| 1958 | 'AGT_No10' | 'LDTEDKLR' | 'Angiotensinogen' |
| 1959 | 'AGT_No11' | 'FMQAVTGWK' | 'Angiotensinogen' |
| 1960 | 'AGT_No12' | 'TGCSLMGASVDSTLAFNTYVHFQGK' | 'Angiotensinogen' |
| 1961 | 'AGT_No13' | 'AAMVGMLANFLGFR' | 'Angiotensinogen' |
| 1962 | 'AGT_No14' | 'VEGLTFQQNSLNWMK' | 'Angiotensinogen' |
| 1963 | 'AGT_No15' | 'ADSQAQLLLSTVVGVFTAPGLHLK' | 'Angiotensinogen' |
| 1964 | 'AGT_No16' | 'TSPVDEKALQDQLVLVAAK' | 'Angiotensinogen' |
| 1965 | 'AGT_No17' | 'TGCSLMGASVDSTLAFNTYVHFQGK' | 'Angiotensinogen' |
| 1966 | 'AGT_No18' | 'AAMVGMLANFLGFR' | 'Angiotensinogen' |
| 1967 | 'AGT_No19' | 'ANAGKPKDPTFIPAPIQAK' | 'Angiotensinogen' |
| 1968 | 'AGT_No20' | 'AAMVGMLANFLGFR' | 'Angiotensinogen' |
| 1969 | 'AGT_No21' | 'ALQDQLVLVAAKLDTEDKLR' | 'Angiotensinogen' |
| 1970 | 'AGT_No22' | 'QPFVQGLALYTPVVLPR' | 'Angiotensinogen' |
| 1971 | 'AGT_No23' | 'VYIHPFHLVIHNESTCEQLAK' | 'Angiotensinogen' |
| 1972 | 'AGT_No24' | 'VEGLTFQQNSLNWMK' | 'Angiotensinogen' |
| 1973 | 'AGT_No25' | 'DPTFIPAPIQAK' | 'Angiotensinogen' |
| 1974 | 'AGT_No26' | 'AAMVGMLANFLGFR' | 'Angiotensinogen' |
| 1975 | 'AGT_No27' | 'LQAILGVPWKDK' | 'Angiotensinogen' |
| 1976 | 'AGT_No28' | 'PFLFAVYDQSATALHFLGR' | 'Angiotensinogen' |
| 1977 | 'AGT_No29' | 'IDRFMQAVTGWK' | 'Angiotensinogen' |
| 1978 | 'A2M_No1' | 'LLIYAVLPTGDVIGDSAK' | 'Alpha-2-macroglobulin' |
| 1979 | 'A2M_No2' | 'IAQWQSFQLEGGLK' | 'Alpha-2-macroglobulin' |
| 1980 | 'A2M_No3' | 'MVSGFIPLKPTVK' | 'Alpha-2-macroglobulin' |
| 1981 | 'A2M_No4' | 'LLLQQVSLPELPGEYSMK' | 'Alpha-2-macroglobulin' |

| | A | B | C |
|---|---|---|---|
| 1 | **Peptide Name** | **Peptide Sequence** | **Peptide Description** |
| 1982 | 'A2M_No5' | 'GHFSISIPVK' | 'Alpha-2-macroglobulin' |
| 1983 | 'A2M_No6' | 'AIGYLNTGYQR' | 'Alpha-2-macroglobulin' |
| 1984 | 'A2M_No7' | 'DTVIKPLLVEPEGLEK' | 'Alpha-2-macroglobulin' |
| 1985 | 'A2M_No8' | 'VGFYESDVMGR' | 'Alpha-2-macroglobulin' |
| 1986 | 'A2M_No9' | 'NEDSLVFVQTDK' | 'Alpha-2-macroglobulin' |
| 1987 | 'A2M_No10' | 'TEVSSNHVLIYLDK' | 'Alpha-2-macroglobulin' |
| 1988 | 'A2M_No11' | 'VDLSFSPSQSLPASHAHLR' | 'Alpha-2-macroglobulin' |
| 1989 | 'A2M_No12' | 'VSVQLEASPAFLAVPVEK' | 'Alpha-2-macroglobulin' |
| 1990 | 'A2M_No13' | 'NALFCLESAWK' | 'Alpha-2-macroglobulin' |
| 1991 | 'A2M_No14' | 'YDVENCLANK' | 'Alpha-2-macroglobulin' |
| 1992 | 'A2M_No15' | 'LLIYAVLPTGDVIGDSAKYDVENCLANKVDLSFSPSQSLPASHAHLR' | 'Alpha-2-macroglobulin' |
| 1993 | 'A2M_No16' | 'YSDASDCHGEDSQAFCEK' | 'Alpha-2-macroglobulin' |
| 1994 | 'A2M_No17' | 'FEVQVTVPK' | 'Alpha-2-macroglobulin' |
| 1995 | 'A2M_No18' | 'VGFYESDVMGR' | 'Alpha-2-macroglobulin' |
| 1996 | 'A2M_No19' | 'VVSMDENFHPLNELIPLVYIQDPK' | 'Alpha-2-macroglobulin' |
| 1997 | 'A2M_No20' | 'FSGQLNSHGCFYQQVK' | 'Alpha-2-macroglobulin' |
| 1998 | 'A2M_No21' | 'LSFYYLIMAK' | 'Alpha-2-macroglobulin' |
| 1999 | 'A2M_No22' | 'GGVEDEVTLSAYITIALLEIPLTVTHPVVR' | 'Alpha-2-macroglobulin' |
| 2000 | 'A2M_No23' | 'KLSFYYLIMAK' | 'Alpha-2-macroglobulin' |
| 2001 | 'A2M_No24' | 'PQYMVLVPSLLHTETTEK' | 'Alpha-2-macroglobulin' |
| 2002 | 'A2M_No25' | 'VTAAPQSVCALR' | 'Alpha-2-macroglobulin' |
| 2003 | 'A2M_No26' | 'SFVHLEPMSHELPCGHTQTVQAHYILNGGTLLGLK' | 'Alpha-2-macroglobulin' |
| 2004 | 'A2M_No27' | 'SFVHLEPMSHELPCGHTQTVQAHYILNGGTLLGLKK' | 'Alpha-2-macroglobulin' |
| 2005 | 'A2M_No28' | 'DMYSFLEDMGLK' | 'Alpha-2-macroglobulin' |
| 2006 | 'A2M_No29' | 'PVPGHVTVSICR' | 'Alpha-2-macroglobulin' |
| 2007 | 'A2M_No30' | 'PQYMVLVPSLLHTETTEK' | 'Alpha-2-macroglobulin' |
| 2008 | 'A2M_No31' | 'LVHVEEPHTETVR' | 'Alpha-2-macroglobulin' |
| 2009 | 'A2M_No32' | 'LPPNVVEESAR' | 'Alpha-2-macroglobulin' |
| 2010 | 'A2M_No33' | 'ATVLNYLPK' | 'Alpha-2-macroglobulin' |
| 2011 | 'A2M_No34' | 'HYDGSYSTFGER' | 'Alpha-2-macroglobulin' |
| 2012 | 'A2M_No35' | 'YGAATFTR' | 'Alpha-2-macroglobulin' |
| 2013 | 'A2M_No36' | 'MVSGFIPLKPTVK' | 'Alpha-2-macroglobulin' |
| 2014 | 'A2M_No37' | 'KYSDASDCHGEDSQAFCEK' | 'Alpha-2-macroglobulin' |
| 2015 | 'A2M_No38' | 'MCPQLQQYEMHGPEGLR' | 'Alpha-2-macroglobulin' |
| 2016 | 'A2M_No39' | 'LVHVEEPHTETVRK' | 'Alpha-2-macroglobulin' |
| 2017 | 'A2M_No40' | 'MCPQLQQYEMHGPEGLR' | 'Alpha-2-macroglobulin' |
| 2018 | 'A2M_No41' | 'KDTVIKPLLVEPEGLEK' | 'Alpha-2-macroglobulin' |
| 2019 | 'A2M_No42' | 'MCPQLQQYEMHGPEGLR' | 'Alpha-2-macroglobulin' |
| 2020 | 'A2M_No43' | 'MCPQLQQYEMHGPEGLR' | 'Alpha-2-macroglobulin' |
| 2021 | 'A2M_No44' | 'DLTGFPGPLNDQDNEDCINR' | 'Alpha-2-macroglobulin' |
| 2022 | 'A2M_No45' | 'DMYSFLEDMGLK' | 'Alpha-2-macroglobulin' |
| 2023 | 'A2M_No46' | 'KLSFYYLIMAK' | 'Alpha-2-macroglobulin' |
| 2024 | 'A2M_No47' | 'QQNAQGGFSSTQDTVVALHALSK' | 'Alpha-2-macroglobulin' |
| 2025 | 'A2M_No48' | 'SDIAPVAR' | 'Alpha-2-macroglobulin' |

| | A | B | C |
|---|---|---|---|
| 1 | **Peptide Name** | **Peptide Sequence** | **Peptide Description** |
| 2026 | 'A2M_No49' | 'DMYSFLEDMGLK' | 'Alpha-2-macroglobulin' |
| 2027 | 'A2M_No50' | 'SIYKPGQTVK' | 'Alpha-2-macroglobulin' |
| 2028 | 'A2M_No51' | 'LLLQQVSLPELPGEYSMK' | 'Alpha-2-macroglobulin' |
| 2029 | 'A2M_No52' | 'NQGNTWLTAFVLK' | 'Alpha-2-macroglobulin' |
| 2030 | 'A2M_No53' | 'SSGSLLNNAIK' | 'Alpha-2-macroglobulin' |
| 2031 | 'A2M_No54' | 'SSSNEEVMFLTVQVK' | 'Alpha-2-macroglobulin' |
| 2032 | 'A2M_No55' | 'GVPIPNKVIFIR' | 'Alpha-2-macroglobulin' |
| 2033 | 'A2M_No56' | 'GHFSISIPVKSDIAPVAR' | 'Alpha-2-macroglobulin' |
| 2034 | 'A2M_No57' | 'SFVHLEPMSHELPCGHTQTVQAHYILNGGTLLGLK' | 'Alpha-2-macroglobulin' |
| 2035 | 'A2M_No58' | 'NEDSLVFVQTDKSIYKPGQTVK' | 'Alpha-2-macroglobulin' |
| 2036 | 'A2M_No59' | 'LSFVKVDSHFR' | 'Alpha-2-macroglobulin' |
| 2037 | 'A2M_No60' | 'PLLVEPEGLEK' | 'Alpha-2-macroglobulin' |
| 2038 | 'A2M_No61' | 'FQVDNNNR' | 'Alpha-2-macroglobulin' |
| 2039 | 'A2M_No62' | 'QTVSWAVTPK' | 'Alpha-2-macroglobulin' |
| 2040 | 'A2M_No63' | 'AVDQSVLLMKPDAELSASSVYNLLPEK' | 'Alpha-2-macroglobulin' |
| 2041 | 'A2M_No64' | 'ALLAYAFALAGNQDK' | 'Alpha-2-macroglobulin' |
| 2042 | 'A2M_No65' | 'EQAPHCICANGR' | 'Alpha-2-macroglobulin' |
| 2043 | 'A2M_No66' | 'DNSVHWERPQKPK' | 'Alpha-2-macroglobulin' |
| 2044 | 'A2M_No67' | 'SASNMAIVDVKMVSGFIPLKPTVK' | 'Alpha-2-macroglobulin' |
| 2045 | 'A2M_No68' | 'TEHPFTVEEFVLPKFEVQVTVPK' | 'Alpha-2-macroglobulin' |
| 2046 | 'A2M_No69' | 'AGAFCLSEDAGLGISSTASLR' | 'Alpha-2-macroglobulin' |
| 2047 | 'A2M_No70' | 'TGKAAQVTIQSSGTFSSK' | 'Alpha-2-macroglobulin' |
| 2048 | 'A2M_No71' | 'TGTHGLLVKQEDMK' | 'Alpha-2-macroglobulin' |
| 2049 | 'A2M_No72' | 'TGTHGLLVK' | 'Alpha-2-macroglobulin' |
| 2050 | 'A2M_No73' | 'SLNEEAVKK' | 'Alpha-2-macroglobulin' |
| 2051 | 'A2M_No74' | 'SKAIGYLNTGYQR' | 'Alpha-2-macroglobulin' |
| 2052 | 'A2M_No75' | 'SGGRTEHPFTVEEFVLPK' | 'Alpha-2-macroglobulin' |
| 2053 | 'A2M_No76' | 'AFQPFFVELTMPYSVIR' | 'Alpha-2-macroglobulin' |
| 2054 | 'A2M_No77' | 'SASNMAIVDVK' | 'Alpha-2-macroglobulin' |
| 2055 | 'A2M_No78' | 'YDVENCLANKVDLSFSPSQSLPASHAHLR' | 'Alpha-2-macroglobulin' |
| 2056 | 'A2M_No79' | 'TGTHGLLVKQEDMK' | 'Alpha-2-macroglobulin' |
| 2057 | 'A2M_No80' | 'SSSNEEVMFLTVQVK' | 'Alpha-2-macroglobulin' |
| 2058 | 'A2M_No81' | 'TITKLSFVK' | 'Alpha-2-macroglobulin' |
| 2059 | 'A2M_No82' | 'VYDYYETDEFAIAEYNAPCSK' | 'Alpha-2-macroglobulin' |
| 2060 | 'A2M_No83' | 'TEHPFTVEEFVLPK' | 'Alpha-2-macroglobulin' |
| 2061 | 'A2M_No84' | 'YNILPEKEEFPFALGVQTLPQTCDEPK' | 'Alpha-2-macroglobulin' |
| 2062 | 'A2M_No85' | 'SLNEEAVK' | 'Alpha-2-macroglobulin' |
| 2063 | 'A2M_No86' | 'LLIYAVLPTGDVIGDSAKYDVENCLANK' | 'Alpha-2-macroglobulin' |
| 2064 | 'A2M_No87' | 'SSSNEEVMFLTVQVKGPTQEFK' | 'Alpha-2-macroglobulin' |
| 2065 | 'A2M_No88' | 'VSNQTLSLFFTVLQDVPVR' | 'Alpha-2-macroglobulin' |
| 2066 | 'A2M_No89' | 'SPCYGYQWVSEEHEEAHHTAYLVFSPSK' | 'Alpha-2-macroglobulin' |
| 2067 | 'A2M_No90' | 'AAQVTIQSSGTFSSK' | 'Alpha-2-macroglobulin' |
| 2068 | 'A2M_No91' | 'DTVIKPLLVEPEGLEKETTFNSLLCPSGGEVSEELSLK' | 'Alpha-2-macroglobulin' |
| 2069 | 'A2M_No92' | 'QGIPFFGQVR' | 'Alpha-2-macroglobulin' |

| | A | B | C |
|---|---|---|---|
| 1 | **Peptide Name** | **Peptide Sequence** | **Peptide Description** |
| 2070 | 'A2M_No93' | 'VTGEGCVYLQTSLK' | 'Alpha-2-macroglobulin' |
| 2071 | 'A2M_No94' | 'QLNYKHYDGSYSTFGER' | 'Alpha-2-macroglobulin' |
| 2072 | 'A2M_No95' | 'LHTEAQIQEEGTVVELTGR' | 'Alpha-2-macroglobulin' |
| 2073 | 'A2M_No96' | 'TITKLSFVKVDSHFR' | 'Alpha-2-macroglobulin' |
| 2074 | 'A2M_No97' | 'DMYSFLEDMGLK' | 'Alpha-2-macroglobulin' |
| 2075 | 'A2M_No98' | 'HNVYINGITYTPVSSTNEK' | 'Alpha-2-macroglobulin' |
| 2076 | 'A2M_No99' | 'FRVVSMDENFHPLNELIPLVYIQDPK' | 'Alpha-2-macroglobulin' |
| 2077 | 'A2M_No100' | 'VVSMDENFHPLNELIPLVYIQDPK' | 'Alpha-2-macroglobulin' |
| 2078 | 'A2M_No101' | 'SFVHLEPMSHELPCGHTQTVQAHYILNGGTLLGLKK' | 'Alpha-2-macroglobulin' |
| 2079 | 'A2M_No102' | 'EEFPFALGVQTLPQTCDEPK' | 'Alpha-2-macroglobulin' |
| 2080 | 'A2M_No103' | 'SASNMAIVDVK' | 'Alpha-2-macroglobulin' |
| 2081 | 'A2M_No104' | 'QLNYKHYDGSYSTFGER' | 'Alpha-2-macroglobulin' |
| 2082 | 'A2M_No105' | 'IAQWQSFQLEGGLKQFSFPLSSEPFQGSYKVVVQK' | 'Alpha-2-macroglobulin' |
| 2083 | 'A2M_No106' | 'AYIFIDEAHITQALIWLSQR' | 'Alpha-2-macroglobulin' |
| 2084 | 'A2M_No107' | 'QEDMKGHFSISIPVKSDIAPVAR' | 'Alpha-2-macroglobulin' |
| 2085 | 'A2M_No108' | 'QTVSWAVTPK' | 'Alpha-2-macroglobulin' |
| 2086 | 'A2M_No109' | 'ETTFNSLLCPSGGEVSEELSLK' | 'Alpha-2-macroglobulin' |
| 2087 | 'A2M_No110' | 'ALLAYAFALAGNQDKRK' | 'Alpha-2-macroglobulin' |
| 2088 | 'A2M_No111' | 'SLFTDLEAENDVLHCVAFAVPK' | 'Alpha-2-macroglobulin' |
| 2089 | 'A2M_No112' | 'AHTSFQISLSVSYTGSR' | 'Alpha-2-macroglobulin' |
| 2090 | 'A2M_No113' | 'QFSFPLSSEPFQGSYK' | 'Alpha-2-macroglobulin' |
| 2091 | 'A2M_No114' | 'ALLAYAFALAGNQDKR' | 'Alpha-2-macroglobulin' |
| 2092 | 'A2M_No115' | 'GPTQEFK' | 'Alpha-2-macroglobulin' |
| 2093 | 'A2M_No116' | 'SASNMAIVDVKMVSGFIPLKPTVK' | 'Alpha-2-macroglobulin' |
| 2094 | 'A2M_No117' | 'ETTFNSLLCPSGGEVSEELSLKLPPNVVEESAR' | 'Alpha-2-macroglobulin' |
| 2095 | 'A2M_No118' | 'QFSFPLSSEPFQGSYKVVVQKK' | 'Alpha-2-macroglobulin' |
| 2096 | 'A2M_No119' | 'SASNMAIVDVKMVSGFIPLKPTVK' | 'Alpha-2-macroglobulin' |
| 2097 | 'A2M_No120' | 'VNYKDRSPCYGYQWVSEEHEEAHHTAYLVFSPSK' | 'Alpha-2-macroglobulin' |
| 2098 | 'A2M_No121' | 'QQNAQGGFSSTQDTVVALHALSK' | 'Alpha-2-macroglobulin' |
| 2099 | 'A2M_No122' | 'VSVQLEASPAFLAVPVEKEQAPHCICANGR' | 'Alpha-2-macroglobulin' |
| 2100 | 'A2M_No123' | 'LSFVKVDSHFRQGIPFFGQVR' | 'Alpha-2-macroglobulin' |
| 2101 | 'A2M_No124' | 'LVDGKGVPIPNKVIFIR' | 'Alpha-2-macroglobulin' |
| 2102 | 'A2M_No125' | 'WITKQQNAQGGFSSTQDTVVALHALSK' | 'Alpha-2-macroglobulin' |
| 2103 | 'A2M_No126' | 'QEDMKGHFSISIPVK' | 'Alpha-2-macroglobulin' |
| 2104 | 'A2M_No127' | 'AVDQSVLLMKPDAELSASSVYNLLPEK' | 'Alpha-2-macroglobulin' |
| 2105 | 'A2M_No128' | 'TKVFQLK' | 'Alpha-2-macroglobulin' |
| 2106 | 'A2M_No129' | 'FRVVSMDENFHPLNELIPLVYIQDPK' | 'Alpha-2-macroglobulin' |
| 2107 | 'A2M_No130' | 'DLKPAIVK' | 'Alpha-2-macroglobulin' |
| 2108 | 'A2M_No131' | 'TAQEGDHGSHVYTK' | 'Alpha-2-macroglobulin' |
| 2109 | 'A2M_No132' | 'IAQWQSFQLEGGLKQFSFPLSSEPFQGSYK' | 'Alpha-2-macroglobulin' |
| 2110 | 'A2M_No133' | 'HNVYINGITYTPVSSTNEKDMYSFLEDMGLK' | 'Alpha-2-macroglobulin' |
| 2111 | 'A2M_No134' | 'QFSFPLSSEPFQGSYK' | 'Alpha-2-macroglobulin' |
| 2112 | 'A2M_No135' | 'LSFYYLIMAK' | 'Alpha-2-macroglobulin' |
| 2113 | 'A2M_No136' | 'TTVMVKNEDSLVFVQTDK' | 'Alpha-2-macroglobulin' |

| | A | B | C |
|---|---|---|---|
| 1 | **Peptide Name** | **Peptide Sequence** | **Peptide Description** |
| 2114 | 'A2M_No137' | 'SGGRTEHPFTVEEFVLPKFEVQVTVPK' | 'Alpha-2-macroglobulin' |
| 2115 | 'A2M_No138' | 'DRSPCYGYQWVSEEHEEAHHTAYLVFSPSK' | 'Alpha-2-macroglobulin' |
| 2116 | 'A2M_No139' | 'NQGNTWLTAFVLKTFAQAR' | 'Alpha-2-macroglobulin' |
| 2117 | 'A2M_No140' | 'QGIPFFGQVR' | 'Alpha-2-macroglobulin' |
| 2118 | 'A2M_No141' | 'AFQPFFVELTMPYSVIR' | 'Alpha-2-macroglobulin' |
| 2119 | 'A2M_No142' | 'QFSFPLSSEPFQGSYKVVVQK' | 'Alpha-2-macroglobulin' |
| 2120 | 'A2M_No143' | 'QSSEITR' | 'Alpha-2-macroglobulin' |
| 2121 | 'A2M_No144' | 'HNVYINGITYTPVSSTNEKDMYSFLEDMGLK' | 'Alpha-2-macroglobulin' |
| 2122 | 'A2M_No145' | 'QLNYKHYDGSYSTFGERYGR' | 'Alpha-2-macroglobulin' |
| 2123 | 'A2M_No146' | 'DLKPAIVKVYDYYETDEFAIAEYNAPCSK' | 'Alpha-2-macroglobulin' |
| 2124 | 'A2M_No147' | 'AIGYLNTGYQRQLNYKHYDGSYSTFGER' | 'Alpha-2-macroglobulin' |
| 2125 | 'A2M_No148' | 'HNVYINGITYTPVSSTNEKDMYSFLEDMGLK' | 'Alpha-2-macroglobulin' |
| 2126 | 'A2M_No149' | 'TAQEGDHGSHVYTKALLAYAFALAGNQDKR' | 'Alpha-2-macroglobulin' |
| 2127 | 'A2M_No150' | 'LVDGKGVPIPNK' | 'Alpha-2-macroglobulin' |
| 2128 | 'A2M_No151' | 'SIYKPGQTVKFR' | 'Alpha-2-macroglobulin' |
| 2129 | 'A2M_No152' | 'QEDMKGHFSISIPVKSDIAPVAR' | 'Alpha-2-macroglobulin' |
| 2130 | 'A2M_No153' | 'TAQEGDHGSHVYTKALLAYAFALAGNQDK' | 'Alpha-2-macroglobulin' |
| 2131 | 'A2M_No154' | 'GEAFTLKATVLNYLPK' | 'Alpha-2-macroglobulin' |
| 2132 | 'A2M_No155' | 'VDSHFRQGIPFFGQVR' | 'Alpha-2-macroglobulin' |
| 2133 | 'A2M_No156' | 'VVSMDENFHPLNELIPLVYIQDPKGNR' | 'Alpha-2-macroglobulin' |
| 2134 | 'A2M_No157' | 'QEDMKGHFSISIPVKSDIAPVAR' | 'Alpha-2-macroglobulin' |
| 2135 | 'A2M_No158' | 'YGRNQGNTWLTAFVLK' | 'Alpha-2-macroglobulin' |
| 2136 | 'A2M_No159' | 'SSSNEEVMFLTVQVKGPTQEFK' | 'Alpha-2-macroglobulin' |
| 2137 | 'A2M_No160' | 'HNVYINGITYTPVSSTNEKDMYSFLEDMGLK' | 'Alpha-2-macroglobulin' |
| 2138 | 'A2M_No161' | 'QEDMKGHFSISIPVK' | 'Alpha-2-macroglobulin' |
| 2139 | 'A2M_No162' | 'QQNAQGGFSSTQDTVVALHALSKYGAATFTR' | 'Alpha-2-macroglobulin' |
| 2140 | 'A2M_No163' | 'FRVVSMDENFHPLNELIPLVYIQDPKGNR' | 'Alpha-2-macroglobulin' |
| 2141 | 'A2M_No164' | 'QQNAQGGFSSTQDTVVALHALSKYGAATFTR' | 'Alpha-2-macroglobulin' |
| 2142 | 'A2M_No165' | 'TTVMVKNEDSLVFVQTDK' | 'Alpha-2-macroglobulin' |
| 2143 | 'A2M_No166' | 'SSSNEEVMFLTVQVKGPTQEFKK' | 'Alpha-2-macroglobulin' |
| 2144 | 'A2M_No167' | 'EVLKSLNEEAVKK' | 'Alpha-2-macroglobulin' |
| 2145 | 'A2M_No168' | 'QFSFPLSSEPFQGSYKVVVQKK' | 'Alpha-2-macroglobulin' |
| 2146 | 'A2M_No169' | 'MCPQLQQYEMHGPEGLRVGFYESDVMGR' | 'Alpha-2-macroglobulin' |
| 2147 | 'A2M_No170' | 'YSDASDCHGEDSQAFCEKFSGQLNSHGCFYQQVK' | 'Alpha-2-macroglobulin' |
| 2148 | 'A2M_No171' | 'QEDMKGHFSISIPVKSDIAPVAR' | 'Alpha-2-macroglobulin' |
| 2149 | 'A2M_No172' | 'CIRVSVQLEASPAFLAVPVEK' | 'Alpha-2-macroglobulin' |
| 2150 | 'A2M_No173' | 'QQNAQGGFSSTQDTVVALHALSKYGAATFTRTGK' | 'Alpha-2-macroglobulin' |
| 2151 | 'A2M_No174' | 'HYDGSYSTFGERYGR' | 'Alpha-2-macroglobulin' |
| 2152 | 'A2M_No175' | 'KYSDASDCHGEDSQAFCEKFSGQLNSHGCFYQQVK' | 'Alpha-2-macroglobulin' |
| 2153 | 'A2M_No176' | 'MCPQLQQYEMHGPEGLRVGFYESDVMGR' | 'Alpha-2-macroglobulin' |
| 2154 | 'A2M_No177' | 'MCPQLQQYEMHGPEGLRVGFYESDVMGR' | 'Alpha-2-macroglobulin' |
| 2155 | 'A2M_No178' | 'KEYEMKLHTEAQIQEEGTVVELTGR' | 'Alpha-2-macroglobulin' |
| 2156 | 'A2M_No179' | 'GGIVRTGTHGLLVKQEDMK' | 'Alpha-2-macroglobulin' |
| 2157 | 'A2M_No180' | 'QEDMKGHFSISIPVK' | 'Alpha-2-macroglobulin' |

| | A | B | C |
|---|---|---|---|
| 1 | **Peptide Name** | **Peptide Sequence** | **Peptide Description** |
| 2158 | 'A2M_No181' | 'DNSVHWERPQK' | 'Alpha-2-macroglobulin' |
| 2159 | 'A2M_No182' | 'EYEMKLHTEAQIQEEGTVVELTGR' | 'Alpha-2-macroglobulin' |
| 2160 | 'A2M_No183' | 'GGIVRTGTHGLLVK' | 'Alpha-2-macroglobulin' |
| 2161 | 'A2M_No184' | 'MCPQLQQYEMHGPEGLRVGFYESDVMGR' | 'Alpha-2-macroglobulin' |
| 2162 | 'A2M_No185' | 'FQVDNNNRLLLQQVSLPELPGEYSMK' | 'Alpha-2-macroglobulin' |
| 2163 | 'A2M_No186' | 'NALFCLESAWKTAQEGDHGSHVYTK' | 'Alpha-2-macroglobulin' |
| 2164 | 'A2M_No187' | 'GPTQEFKK' | 'Alpha-2-macroglobulin' |
| 2165 | 'A2M_No188' | 'FSGQLNSHGCFYQQVKTK' | 'Alpha-2-macroglobulin' |
| 2166 | 'A2M_No189' | 'QSSEITRTITKLSFVK' | 'Alpha-2-macroglobulin' |
| 2167 | 'A2M_No190' | 'VYDYYETDEFAIAEYNAPCSKDLGNA' | 'Alpha-2-macroglobulin' |
| 2168 | 'A2M_No191' | 'QEDMKGHFSISIPVK' | 'Alpha-2-macroglobulin' |
| 2169 | 'A2M_No192' | 'PKMCPQLQQYEMHGPEGLR' | 'Alpha-2-macroglobulin' |
| 2170 | 'A2M_No193' | 'QKDNGCFR' | 'Alpha-2-macroglobulin' |
| 2171 | 'A2M_No194' | 'SLNEEAVKKDNSVHWER' | 'Alpha-2-macroglobulin' |
| 2172 | 'A2M_No195' | 'EVLKSLNEEAVK' | 'Alpha-2-macroglobulin' |
| 2173 | 'A2M_No196' | 'KSGGRTEHPFTVEEFVLPK' | 'Alpha-2-macroglobulin' |
| 2174 | 'A2M_No197' | 'TGKAAQVTIQSSGTFSSKFQVDNNNR' | 'Alpha-2-macroglobulin' |
| 2175 | 'A2M_No198' | 'MVSGFIPLK' | 'Alpha-2-macroglobulin' |
| 2176 | 'A2M_No199' | 'QKDNGCFR' | 'Alpha-2-macroglobulin' |
| 2177 | 'A2M_No200' | 'VVSMDENFHPLNELIPLVYIQDPKGNR' | 'Alpha-2-macroglobulin' |
| 2178 | 'A2M_No201' | 'VGFYESDVMGRGHAR' | 'Alpha-2-macroglobulin' |
| 2179 | 'A2M_No202' | 'LHTEAQIQEEGTVVELTGRQSSEITR' | 'Alpha-2-macroglobulin' |
| 2180 | 'A2M_No203' | 'FQVDNNNRLLLQQVSLPELPGEYSMK' | 'Alpha-2-macroglobulin' |
| 2181 | 'A2M_No204' | 'LHTEAQIQEEGTVVELTGRQSSEITRTITK' | 'Alpha-2-macroglobulin' |
| 2182 | 'A2M_No205' | 'QFSFPLSSEPFQGSYKVVVQK' | 'Alpha-2-macroglobulin' |
| 2183 | 'A2M_No206' | 'PLLVEPEGLEKETTFNSLLCPSGGEVSEELSLK' | 'Alpha-2-macroglobulin' |
| 2184 | 'A2M_No207' | 'FRVVSMDENFHPLNELIPLVYIQDPKGNR' | 'Alpha-2-macroglobulin' |
| 2185 | 'A2M_No208' | 'VGFYESDVMGRGHAR' | 'Alpha-2-macroglobulin' |
| 2186 | 'A2M_No209' | 'RTTVMVKNEDSLVFVQTDK' | 'Alpha-2-macroglobulin' |
| 2187 | 'A2M_No210' | 'SSSNEEVMFLTVQVKGPTQEFKK' | 'Alpha-2-macroglobulin' |
| 2188 | 'A2M_No211' | 'RTTVMVKNEDSLVFVQTDK' | 'Alpha-2-macroglobulin' |
| 2189 | 'A2M_No212' | 'KEYEMKLHTEAQIQEEGTVVELTGR' | 'Alpha-2-macroglobulin' |
| 2190 | 'A2M_No213' | 'ATVLNYLPKCIR' | 'Alpha-2-macroglobulin' |
| 2191 | 'A2M_No214' | 'QSSEITRTITKLSFVK' | 'Alpha-2-macroglobulin' |
| 2192 | 'A2M_No215' | 'GGIVRTGTHGLLVKQEDMK' | 'Alpha-2-macroglobulin' |
| 2193 | 'A2M_No216' | 'EYEMKLHTEAQIQEEGTVVELTGR' | 'Alpha-2-macroglobulin' |
| 2194 | 'A2M_No217' | 'GPTQEFKKR' | 'Alpha-2-macroglobulin' |
| 2195 | 'A2M_No218' | 'PDAELSASSVYNLLPEK' | 'Alpha-2-macroglobulin' |
| 2196 | 'A2M_No219' | 'QLNYKHYDGSYSTFGERYGR' | 'Alpha-2-macroglobulin' |
| 2197 | 'A2M_No220' | 'KDNSVHWER' | 'Alpha-2-macroglobulin' |
| 2198 | 'A2M_No221' | 'PKMCPQLQQYEMHGPEGLR' | 'Alpha-2-macroglobulin' |
| 2199 | 'A2M_No222' | 'GEAFTLKATVLNYLPKCIR' | 'Alpha-2-macroglobulin' |
| 2200 | 'A2M_No223' | 'EQAPHCICANGRQTVSWAVTPK' | 'Alpha-2-macroglobulin' |
| 2201 | 'A2M_No224' | 'SNHVSRTEVSSNHVLIYLDK' | 'Alpha-2-macroglobulin' |

| | A | B | C |
|---|---|---|---|
| 1 | **Peptide Name** | **Peptide Sequence** | **Peptide Description** |
| 2202 | 'C3_No1' | 'SGIPIVTSPYQIHFTK' | 'Complement C3' |
| 2203 | 'C3_No2' | 'IHWESASLLR' | 'Complement C3' |
| 2204 | 'C3_No3' | 'LSINTHPSQKPLSITVR' | 'Complement C3' |
| 2205 | 'C3_No4' | 'LVAYYTLIGASGQR' | 'Complement C3' |
| 2206 | 'C3_No5' | 'DICEEQVNSLPGSITK' | 'Complement C3' |
| 2207 | 'C3_No6' | 'TIYTPGSTVLYR' | 'Complement C3' |
| 2208 | 'C3_No7' | 'IPIEDGSGEVVLSR' | 'Complement C3' |
| 2209 | 'C3_No8' | 'VELLHNPAFCSLATTK' | 'Complement C3' |
| 2210 | 'C3_No9' | 'VPVAVQGEDTVQSLTQGDGVAK' | 'Complement C3' |
| 2211 | 'C3_No10' | 'VYAYYNLEESCTR' | 'Complement C3' |
| 2212 | 'C3_No11' | 'GQGTLSVVTMYHAK' | 'Complement C3' |
| 2213 | 'C3_No12' | 'SLYVSATVILHSGSDMVQAER' | 'Complement C3' |
| 2214 | 'C3_No13' | 'FYYIYNEK' | 'Complement C3' |
| 2215 | 'C3_No14' | 'AGDFLEANYMNLQR' | 'Complement C3' |
| 2216 | 'C3_No15' | 'NTLIIYLDK' | 'Complement C3' |
| 2217 | 'C3_No16' | 'SSLSVPYVIVPLKTGLQEVEVK' | 'Complement C3' |
| 2218 | 'C3_No17' | 'VFLDCCNYITELR' | 'Complement C3' |
| 2219 | 'C3_No18' | 'DSITTWEILAVSMSDKK' | 'Complement C3' |
| 2220 | 'C3_No19' | 'SEETKENEGFTVTAEGK' | 'Complement C3' |
| 2221 | 'C3_No20' | 'GQGTLSVVTMYHAK' | 'Complement C3' |
| 2222 | 'C3_No21' | 'FISLGEACK' | 'Complement C3' |
| 2223 | 'C3_No22' | 'VEGTAFVIFGIQDGEQR' | 'Complement C3' |
| 2224 | 'C3_No23' | 'QKPDGVFQEDAPVIHQEMIGGLR' | 'Complement C3' |
| 2225 | 'C3_No24' | 'VVLVSLQSGYLFIQTDK' | 'Complement C3' |
| 2226 | 'C3_No25' | 'DSCVGSLVVK' | 'Complement C3' |
| 2227 | 'C3_No26' | 'YFKPGMPFDLMVFVTNPDGSPAYR' | 'Complement C3' |
| 2228 | 'C3_No27' | 'SDDKVTLEERLDK' | 'Complement C3' |
| 2229 | 'C3_No28' | 'QCQDLGAFTESMVVFGCPN' | 'Complement C3' |
| 2230 | 'C3_No29' | 'QLYNVEATSYALLALLQLK' | 'Complement C3' |
| 2231 | 'C3_No30' | 'AFSDRNTLIIYLDK' | 'Complement C3' |
| 2232 | 'C3_No31' | 'LPYSVVR' | 'Complement C3' |
| 2233 | 'C3_No32' | 'SNLDEDIIAEENIVSR' | 'Complement C3' |
| 2234 | 'C3_No33' | 'ISLPESLKR' | 'Complement C3' |
| 2235 | 'C3_No34' | 'AAVYHHFISDGVR' | 'Complement C3' |
| 2236 | 'C3_No35' | 'VSHSEDDCLAFK' | 'Complement C3' |
| 2237 | 'C3_No36' | 'QKPDGVFQEDAPVIHQEMIGGLR' | 'Complement C3' |
| 2238 | 'C3_No37' | 'AAVYHHFISDGVRK' | 'Complement C3' |
| 2239 | 'C3_No38' | 'LDKACEPGVDYVYK' | 'Complement C3' |
| 2240 | 'C3_No39' | 'SGQSEDRQPVPGQQMTLK' | 'Complement C3' |
| 2241 | 'C3_No40' | 'QELSEAEQATR' | 'Complement C3' |
| 2242 | 'C3_No41' | 'YYTYLIMNK' | 'Complement C3' |
| 2243 | 'C3_No42' | 'EVVADSVWVDVK' | 'Complement C3' |
| 2244 | 'C3_No43' | 'YYTYLIMNK' | 'Complement C3' |
| 2245 | 'C3_No44' | 'VTIKPAPETEK' | 'Complement C3' |

| | A | B | C |
|---|---|---|---|
| 1 | **Peptide Name** | **Peptide Sequence** | **Peptide Description** |
| 2246 | 'C3_No45' | 'EDIPPADLSDQVPDTESETR' | 'Complement C3' |
| 2247 | 'C3_No46' | 'LMNIFLK' | 'Complement C3' |
| 2248 | 'C3_No47' | 'FISLGEACKK' | 'Complement C3' |
| 2249 | 'C3_No48' | 'KQELSEAEQATR' | 'Complement C3' |
| 2250 | 'C3_No49' | 'YISKYELDK' | 'Complement C3' |
| 2251 | 'C3_No50' | 'QGALELIKK' | 'Complement C3' |
| 2252 | 'C3_No51' | 'YELDKAFSDR' | 'Complement C3' |
| 2253 | 'C3_No52' | 'AKDQLTCNKFDLK' | 'Complement C3' |
| 2254 | 'C3_No53' | 'QPVPGQQMTLK' | 'Complement C3' |
| 2255 | 'C3_No54' | 'RQGALELIK' | 'Complement C3' |
| 2256 | 'C3_No55' | 'ENEGFTVTAEGK' | 'Complement C3' |
| 2257 | 'C3_No56' | 'SDDKVTLEER' | 'Complement C3' |
| 2258 | 'C3_No57' | 'AELQCPQPAAR' | 'Complement C3' |
| 2259 | 'C3_No58' | 'LKGPLLNK' | 'Complement C3' |
| 2260 | 'C3_No59' | 'GVFVLNKK' | 'Complement C3' |
| 2261 | 'C3_No60' | 'ILLQGTPVAQMTEDAVDAER' | 'Complement C3' |
| 2262 | 'C3_No61' | 'QPSSAFAAFVK' | 'Complement C3' |
| 2263 | 'C3_No62' | 'KHYLMWGLSSDFWGEKPNLSYIIGK' | 'Complement C3' |
| 2264 | 'C3_No63' | 'SEFPESWLWNVEDLKEPPKNGISTK' | 'Complement C3' |
| 2265 | 'C3_No64' | 'FYHPEKEDGKLNK' | 'Complement C3' |
| 2266 | 'C3_No65' | 'NTMILEICTR' | 'Complement C3' |
| 2267 | 'C3_No66' | 'YISKYELDKAFSDR' | 'Complement C3' |
| 2268 | 'C3_No67' | 'SLYVSATVILHSGSDMVQAER' | 'Complement C3' |
| 2269 | 'C3_No68' | 'NNNEKDMALTAFVLISLQEAK' | 'Complement C3' |
| 2270 | 'C3_No69' | 'DYAGVFSDAGLTFTSSSGQQTAQR' | 'Complement C3' |
| 2271 | 'C3_No70' | 'NNNEKDMALTAFVLISLQEAK' | 'Complement C3' |
| 2272 | 'C3_No71' | 'EGVQKEDIPPADLSDQVPDTESETR' | 'Complement C3' |
| 2273 | 'C3_No72' | 'SYTVAIAGYALAQMGR' | 'Complement C3' |
| 2274 | 'C3_No73' | 'ADIGCTPGSGK' | 'Complement C3' |
| 2275 | 'C3_No74' | 'ASHLGLAR' | 'Complement C3' |
| 2276 | 'C3_No75' | 'TMQALPYSTVGNSNNYLHLSVLR' | 'Complement C3' |
| 2277 | 'C3_No76' | 'YYGGGYGSTQATFMVFQALAQYQK' | 'Complement C3' |
| 2278 | 'C3_No77' | 'VRVELLHNPAFCSLATTK' | 'Complement C3' |
| 2279 | 'C3_No78' | 'YFKPGMPFDLMVFVTNPDGSPAYR' | 'Complement C3' |
| 2280 | 'C3_No79' | 'CAEENCFIQK' | 'Complement C3' |
| 2281 | 'C3_No80' | 'HYLMWGLSSDFWGEKPNLSYIIGK' | 'Complement C3' |
| 2282 | 'C3_No81' | 'IFTVNHK' | 'Complement C3' |
| 2283 | 'C3_No82' | 'HQQTVTIPPK' | 'Complement C3' |
| 2284 | 'C3_No83' | 'KVLLDGVQNPR' | 'Complement C3' |
| 2285 | 'C3_No84' | 'VFLDCCNYITELRR' | 'Complement C3' |
| 2286 | 'C3_No85' | 'TELRPGETLNVNFLLR' | 'Complement C3' |
| 2287 | 'C3_No86' | 'GLEVTITAR' | 'Complement C3' |
| 2288 | 'C3_No87' | 'HLIVTPSGCGEQNMIGMTPTVIAVHYLDETEQWEK' | 'Complement C3' |
| 2289 | 'C3_No88' | 'HYLMWGLSSDFWGEKPNLSYIIGK' | 'Complement C3' |

| | A | B | C |
|---|---|---|---|
| 1 | **Peptide Name** | **Peptide Sequence** | **Peptide Description** |
| 2290 | 'C3_No89' | 'HLIVTPSGCGEQNMIGMTPTVIAVHYLDETEQWEK' | 'Complement C3' |
| 2291 | 'C3_No90' | 'KVFLDCCNYITELRR' | 'Complement C3' |
| 2292 | 'C3_No91' | 'NTMILEICTR' | 'Complement C3' |
| 2293 | 'C3_No92' | 'IPIEDGSGEVVLSRK' | 'Complement C3' |
| 2294 | 'C3_No93' | 'VQLSNDFDEYIMAIEQTIK' | 'Complement C3' |
| 2295 | 'C3_No94' | 'QGALELIK' | 'Complement C3' |
| 2296 | 'C3_No95' | 'IRYYTYLIMNK' | 'Complement C3' |
| 2297 | 'C3_No96' | 'NEQVEIR' | 'Complement C3' |
| 2298 | 'C3_No97' | 'QPSSAFAAFVK' | 'Complement C3' |
| 2299 | 'C3_No98' | 'KVFLDCCNYITELR' | 'Complement C3' |
| 2300 | 'C3_No99' | 'IWDVVEKADIGCTPGSGK' | 'Complement C3' |
| 2301 | 'C3_No100' | 'FLYGKKVEGTAFVIFGIQDGEQR' | 'Complement C3' |
| 2302 | 'C3_No101' | 'EALKLEEK' | 'Complement C3' |
| 2303 | 'C3_No102' | 'QLYNVEATSYALLALLQLK' | 'Complement C3' |
| 2304 | 'C3_No103' | 'HLIVTPSGCGEQNMIGMTPTVIAVHYLDETEQWEK' | 'Complement C3' |
| 2305 | 'C3_No104' | 'VSHSEDDCLAFKVHQYFNVELIQPGAVK' | 'Complement C3' |
| 2306 | 'C3_No105' | 'QCQDLGAFTESMVVFGCPN' | 'Complement C3' |
| 2307 | 'C3_No106' | 'KHYLMWGLSSDFWGEKPNLSYIIGK' | 'Complement C3' |
| 2308 | 'C3_No107' | 'FYHPEKEDGK' | 'Complement C3' |
| 2309 | 'C3_No108' | 'QLANGVDR' | 'Complement C3' |
| 2310 | 'C3_No109' | 'DFDFVPPVVR' | 'Complement C3' |
| 2311 | 'C3_No110' | 'PGETLNVNFLLR' | 'Complement C3' |
| 2312 | 'C3_No111' | 'VLLDGVQNPR' | 'Complement C3' |
| 2313 | 'C3_No112' | 'QGALELIKK' | 'Complement C3' |
| 2314 | 'C3_No113' | 'SYTVAIAGYALAQMGR' | 'Complement C3' |
| 2315 | 'C3_No114' | 'VVLVSLQSGYLFIQTDKTIYTPGSTVLYR' | 'Complement C3' |
| 2316 | 'C3_No115' | 'VFSLAVNLIAIDSQVLCGAVK' | 'Complement C3' |
| 2317 | 'C3_No116' | 'YYGGGYGSTQATFMVFQALAQYQK' | 'Complement C3' |
| 2318 | 'C3_No117' | 'FVTVQATFGTQVVEKVVLVSLQSGYLFIQTDK' | 'Complement C3' |
| 2319 | 'C3_No118' | 'EYVLPSFEVIVEPTEK' | 'Complement C3' |
| 2320 | 'C3_No119' | 'VELLHNPAFCSLATTKR' | 'Complement C3' |
| 2321 | 'C3_No120' | 'DTWVEHWPEEDECQDEENQK' | 'Complement C3' |
| 2322 | 'C3_No121' | 'QPSSAFAAFVKR' | 'Complement C3' |
| 2323 | 'C3_No122' | 'QPVPGQQMTLK' | 'Complement C3' |
| 2324 | 'C3_No123' | 'LESEETMVLEAHDAQGDVPVTVTVHDFPGK' | 'Complement C3' |
| 2325 | 'C3_No124' | 'DQLTCNKFDLK' | 'Complement C3' |
| 2326 | 'C3_No125' | 'VQLSNDFDEYIMAIEQTIK' | 'Complement C3' |
| 2327 | 'C3_No126' | 'LESEETMVLEAHDAQGDVPVTVTVHDFPGKK' | 'Complement C3' |
| 2328 | 'C3_No127' | 'TKKQELSEAEQATR' | 'Complement C3' |
| 2329 | 'C3_No128' | 'TRFISLGEACK' | 'Complement C3' |
| 2330 | 'C3_No129' | 'IRYYTYLIMNK' | 'Complement C3' |
| 2331 | 'C3_No130' | 'VHQYFNVELIQPGAVK' | 'Complement C3' |
| 2332 | 'C3_No131' | 'VVLVAVDKGVFVLNKK' | 'Complement C3' |
| 2333 | 'C3_No132' | 'EPGQDLVVLPLSITTDFIPSFR' | 'Complement C3' |

| | A | B | C |
|---|---|---|---|
| 1 | **Peptide Name** | **Peptide Sequence** | **Peptide Description** |
| 2334 | 'C3_No133' | 'VLLDGVQNPRAEDLVGK' | 'Complement C3' |
| 2335 | 'C3_No134' | 'RIPIEDGSGEVVLSRK' | 'Complement C3' |
| 2336 | 'C3_No135' | 'DMALTAFVLISLQEAK' | 'Complement C3' |
| 2337 | 'C3_No136' | 'KGYTQQLAFR' | 'Complement C3' |
| 2338 | 'C3_No137' | 'DAPDHQELNLDVSLQLPSR' | 'Complement C3' |
| 2339 | 'C3_No138' | 'VVLVAVDKGVFVLNK' | 'Complement C3' |
| 2340 | 'C3_No139' | 'YELDKAFSDRNTLIIYLDK' | 'Complement C3' |
| 2341 | 'C3_No140' | 'SSLSVPYVIVPLK' | 'Complement C3' |
| 2342 | 'C3_No141' | 'DSITTWEILAVSMSDKK' | 'Complement C3' |
| 2343 | 'C3_No142' | 'KCCEDGMRENPMR' | 'Complement C3' |
| 2344 | 'C3_No143' | 'CCEDGMRENPMR' | 'Complement C3' |
| 2345 | 'C3_No144' | 'DSITTWEILAVSMSDK' | 'Complement C3' |
| 2346 | 'C3_No145' | 'TFISPIK' | 'Complement C3' |
| 2347 | 'C3_No146' | 'TGLQEVEVK' | 'Complement C3' |
| 2348 | 'C3_No147' | 'KLVLSSEK' | 'Complement C3' |
| 2349 | 'C3_No148' | 'SGSDEVQVGQQR' | 'Complement C3' |
| 2350 | 'C3_No149' | 'PNLSYIIGK' | 'Complement C3' |
| 2351 | 'C3_No150' | 'LESEETMVLEAHDAQGDVPVTVTVHDFPGK' | 'Complement C3' |
| 2352 | 'C3_No151' | 'KVEGTAFVIFGIQDGEQR' | 'Complement C3' |
| 2353 | 'C3_No152' | 'YFKPGMPFDLMVFVTNPDGSPAYR' | 'Complement C3' |
| 2354 | 'C3_No153' | 'TRFISLGEACKK' | 'Complement C3' |
| 2355 | 'C3_No154' | 'GYTQQLAFRQPSSAFAAFVK' | 'Complement C3' |
| 2356 | 'C3_No155' | 'PDGVFQEDAPVIHQEMIGGLR' | 'Complement C3' |
| 2357 | 'C3_No156' | 'AGDFLEANYMNLQR' | 'Complement C3' |
| 2358 | 'C3_No157' | 'LPYSVVRNEQVEIR' | 'Complement C3' |
| 2359 | 'C3_No158' | 'LESEETMVLEAHDAQGDVPVTVTVHDFPGKK' | 'Complement C3' |
| 2360 | 'C3_No159' | 'TLDPERLGR' | 'Complement C3' |
| 2361 | 'C3_No160' | 'AYYENSPQQVFSTEFEVK' | 'Complement C3' |
| 2362 | 'C3_No161' | 'LKGPLLNKFLTTAK' | 'Complement C3' |
| 2363 | 'C3_No162' | 'TMQALPYSTVGNSNNYLHLSVLR' | 'Complement C3' |
| 2364 | 'C3_No163' | 'APSTWLTAYVVK' | 'Complement C3' |
| 2365 | 'C3_No164' | 'VRVELLHNPAFCSLATTKR' | 'Complement C3' |
| 2366 | 'C3_No165' | 'ENEGFTVTAEGKGQGTLSVVTMYHAK' | 'Complement C3' |
| 2367 | 'C3_No166' | 'SEETKENEGFTVTAEGKGQGTLSVVTMYHAK' | 'Complement C3' |
| 2368 | 'C3_No167' | 'SVQLTEK' | 'Complement C3' |
| 2369 | 'C3_No168' | 'GYTQQLAFR' | 'Complement C3' |
| 2370 | 'C3_No169' | 'SEFPESWLWNVEDLKEPPK' | 'Complement C3' |
| 2371 | 'C3_No170' | 'SGQSEDRQPVPGQQMTLK' | 'Complement C3' |
| 2372 | 'C3_No171' | 'EVVADSVWVDVKDSCVGSLVVK' | 'Complement C3' |
| 2373 | 'C3_No172' | 'HLIVTPSGCGEQNMIGMTPTVIAVHYLDETEQWEK' | 'Complement C3' |
| 2374 | 'C3_No173' | 'HQQTVTIPPKSSLSVPYVIVPLKTGLQEVEVK' | 'Complement C3' |
| 2375 | 'C3_No174' | 'VVLVAVDK' | 'Complement C3' |
| 2376 | 'C3_No175' | 'QCQDLGAFTESMVVFGCPN' | 'Complement C3' |
| 2377 | 'C3_No176' | 'LMNIFLK' | 'Complement C3' |

| | A | B | C |
|---|---|---|---|
| 1 | **Peptide Name** | **Peptide Sequence** | **Peptide Description** |
| 2378 | 'C3_No177' | 'QKPDGVFQEDAPVIHQEMIGGLR' | 'Complement C3' |
| 2379 | 'C3_No178' | 'KVLLDGVQNPRAEDLVGK' | 'Complement C3' |
| 2380 | 'C3_No179' | 'PDGVFQEDAPVIHQEMIGGLR' | 'Complement C3' |
| 2381 | 'C3_No180' | 'HQQTVTIPPKSSLSVPYVIVPLK' | 'Complement C3' |
| 2382 | 'C3_No181' | 'AVLYNYRQNQELK' | 'Complement C3' |
| 2383 | 'C3_No182' | 'NTLIIYLDKVSHSEDDCLAFK' | 'Complement C3' |
| 2384 | 'C3_No183' | 'YRGDQDATMSILDISMMTGFAPDTDDLK' | 'Complement C3' |
| 2385 | 'C3_No184' | 'FVTVQATFGTQVVEK' | 'Complement C3' |
| 2386 | 'C3_No185' | 'ISLPESLK' | 'Complement C3' |
| 2387 | 'C3_No186' | 'YFKPGMPFDLMVFVTNPDGSPAYR' | 'Complement C3' |
| 2388 | 'C3_No187' | 'VTLEERLDK' | 'Complement C3' |
| 2389 | 'C3_No188' | 'TGLQEVEVKAAVYHHFISDGVRK' | 'Complement C3' |
| 2390 | 'C3_No189' | 'SGQSEDRQPVPGQQMTLKIEGDHGAR' | 'Complement C3' |
| 2391 | 'C3_No190' | 'SSLSVPYVIVPLKTGLQEVEVKAAVYHHFISDGVR' | 'Complement C3' |
| 2392 | 'C3_No191' | 'YRGDQDATMSILDISMMTGFAPDTDDLK' | 'Complement C3' |
| 2393 | 'C3_No192' | 'ILLQGTPVAQMTEDAVDAERLK' | 'Complement C3' |
| 2394 | 'C3_No193' | 'NKFVTVQATFGTQVVEK' | 'Complement C3' |
| 2395 | 'C3_No194' | 'ACEPGVDYVYKTR' | 'Complement C3' |
| 2396 | 'C3_No195' | 'IHWESASLLRSEETKENEGFTVTAEGK' | 'Complement C3' |
| 2397 | 'C3_No196' | 'LMNIFLKDSITTWEILAVSMSDKK' | 'Complement C3' |
| 2398 | 'C3_No197' | 'LCRDELCR' | 'Complement C3' |
| 2399 | 'C3_No198' | 'ILLQGTPVAQMTEDAVDAER' | 'Complement C3' |
| 2400 | 'C3_No199' | 'ADIGCTPGSGKDYAGVFSDAGLTFTSSSGQQTAQR' | 'Complement C3' |
| 2401 | 'C3_No200' | 'RAPSTWLTAYVVK' | 'Complement C3' |
| 2402 | 'C3_No201' | 'TVMVNIENPEGIPVK' | 'Complement C3' |
| 2403 | 'C3_No202' | 'TVMVNIENPEGIPVK' | 'Complement C3' |
| 2404 | 'C3_No203' | 'QKPDGVFQEDAPVIHQEMIGGLR' | 'Complement C3' |
| 2405 | 'C3_No204' | 'SVQLTEKR' | 'Complement C3' |
| 2406 | 'C3_No205' | 'RIPIEDGSGEVVLSR' | 'Complement C3' |
| 2407 | 'C3_No206' | 'SGQSEDRQPVPGQQMTLKIEGDHGAR' | 'Complement C3' |
| 2408 | 'C3_No207' | 'YRGDQDATMSILDISMMTGFAPDTDDLKQLANGVDR' | 'Complement C3' |
| 2409 | 'C3_No208' | 'DMALTAFVLISLQEAK' | 'Complement C3' |
| 2410 | 'C3_No209' | 'AFSDRNTLIIYLDKVSHSEDDCLAFK' | 'Complement C3' |
| 2411 | 'C3_No210' | 'PGMPFDLMVFVTNPDGSPAYR' | 'Complement C3' |
| 2412 | 'C3_No211' | 'LMNIFLKDSITTWEILAVSMSDKK' | 'Complement C3' |
| 2413 | 'C3_No212' | 'LDKACEPGVDYVYKTR' | 'Complement C3' |
| 2414 | 'C3_No213' | 'CAEENCFIQKSDDKVTLEER' | 'Complement C3' |
| 2415 | 'C3_No214' | 'QPSSAFAAFVKR' | 'Complement C3' |
| 2416 | 'C3_No215' | 'IWDVVEK' | 'Complement C3' |
| 2417 | 'C3_No216' | 'KGYTQQLAFRQPSSAFAAFVK' | 'Complement C3' |
| 2418 | 'C3_No217' | 'ACEPGVDYVYK' | 'Complement C3' |
| 2419 | 'C3_No218' | 'YRGDQDATMSILDISMMTGFAPDTDDLK' | 'Complement C3' |
| 2420 | 'C3_No219' | 'PGMPFDLMVFVTNPDGSPAYR' | 'Complement C3' |
| 2421 | 'C3_No220' | 'QLANGVDRYISK' | 'Complement C3' |

| | A | B | C |
|---|---|---|---|
| 1 | **Peptide Name** | **Peptide Sequence** | **Peptide Description** |
| 2422 | 'C3_No221' | 'SSKITHRIHWESASLLR' | 'Complement C3' |
| 2423 | 'C3_No222' | 'TVLTPATNHMGNVTFTIPANR' | 'Complement C3' |
| 2424 | 'C3_No223' | 'IFTVNHKLLPVGR' | 'Complement C3' |
| 2425 | 'C3_No224' | 'ILLQGTPVAQMTEDAVDAERLK' | 'Complement C3' |
| 2426 | 'C3_No225' | 'QVREPGQDLVVLPLSITTDFIPSFR' | 'Complement C3' |
| 2427 | 'C3_No226' | 'LGREGVQKEDIPPADLSDQVPDTESETR' | 'Complement C3' |
| 2428 | 'C3_No227' | 'RQGALELIKK' | 'Complement C3' |
| 2429 | 'C3_No228' | 'AGDFLEANYMNLQRSYTVAIAGYALAQMGR' | 'Complement C3' |
| 2430 | 'C3_No229' | 'AEDLVGKSLYVSATVILHSGSDMVQAER' | 'Complement C3' |
| 2431 | 'C3_No230' | 'RPQDAKNTMILEICTR' | 'Complement C3' |
| 2432 | 'C3_No231' | 'AVLYNYRQNQELKVR' | 'Complement C3' |
| 2433 | 'C3_No232' | 'ITHRIHWESASLLR' | 'Complement C3' |
| 2434 | 'C3_No233' | 'FYYIYNEKGLEVTITAR' | 'Complement C3' |
| 2435 | 'C3_No234' | 'QNQELKVRVELLHNPAFCSLATTK' | 'Complement C3' |
| 2436 | 'C3_No235' | 'SLKVVPEGIR' | 'Complement C3' |
| 2437 | 'C3_No236' | 'QLANGVDRYISKYELDK' | 'Complement C3' |
| 2438 | 'C3_No237' | 'SEETKENEGFTVTAEGKGQGTLSVVTMYHAK' | 'Complement C3' |
| 2439 | 'C3_No238' | 'RHQQTVTIPPK' | 'Complement C3' |
| 2440 | 'C3_No239' | 'CCEDGMRENPMR' | 'Complement C3' |
| 2441 | 'C3_No240' | 'KCCEDGMRENPMR' | 'Complement C3' |
| 2442 | 'C3_No241' | 'TIYTPGSTVLYRIFTVNHK' | 'Complement C3' |
| 2443 | 'C3_No242' | 'LNKLCRDELCR' | 'Complement C3' |
| 2444 | 'C3_No243' | 'TGLQEVEVKAAVYHHFISDGVR' | 'Complement C3' |
| 2445 | 'C3_No244' | 'ACEPGVDYVYKTRLVK' | 'Complement C3' |
| 2446 | 'C3_No245' | 'QGALELIK' | 'Complement C3' |
| 2447 | 'C3_No246' | 'CCEDGMR' | 'Complement C3' |
| 2448 | 'C3_No247' | 'LSINTHPSQK' | 'Complement C3' |
| 2449 | 'C3_No248' | 'DKNRWEDPGK' | 'Complement C3' |
| 2450 | 'C3_No249' | 'AHEAKIRYYTYLIMNK' | 'Complement C3' |
| 2451 | 'C3_No250' | 'TFISPIKCR' | 'Complement C3' |
| 2452 | 'C3_No251' | 'QLANGVDRYISKYELDK' | 'Complement C3' |
| 2453 | 'C3_No252' | 'VELLHNPAFCSLATTKRR' | 'Complement C3' |
| 2454 | 'C3_No253' | 'QLANGVDRYISK' | 'Complement C3' |
| 2455 | 'C3_No254' | 'QDSLSSQNQLGVLPLSWDIPELVNMGQWK' | 'Complement C3' |
| 2456 | 'C3_No255' | 'DAPDHQELNLDVSLQLPSRSSK' | 'Complement C3' |
| 2457 | 'C3_No256' | 'QCQDLGAFTESMVVFGCPN' | 'Complement C3' |
| 2458 | 'C3_No257' | 'QPVPGQQMTLK' | 'Complement C3' |
| 2459 | 'C3_No258' | 'YRGDQDATMSILDISMMTGFAPDTDDLK' | 'Complement C3' |
| 2460 | 'C3_No259' | 'QPVPGQQMTLK' | 'Complement C3' |
| 2461 | 'C3_No260' | 'VTLEERLDKACEPGVDYVYK' | 'Complement C3' |
| 2462 | 'C3_No261' | 'QDSLSSQNQLGVLPLSWDIPELVNMGQWK' | 'Complement C3' |
| 2463 | 'C3_No262' | 'GDQDATMSILDISMMTGFAPDTDDLK' | 'Complement C3' |
| 2464 | 'C3_No263' | 'YYTYLIMNKGR' | 'Complement C3' |
| 2465 | 'C3_No264' | 'QPVPGQQMTLKIEGDHGAR' | 'Complement C3' |

| | A | B | C |
|---|---|---|---|
| 1 | **Peptide Name** | **Peptide Sequence** | **Peptide Description** |
| 2466 | 'C3_No265' | 'LMNIFLKDSITTWEILAVSMSDKK' | 'Complement C3' |
| 2467 | 'C3_No266' | 'AEDLVGKSLYVSATVILHSGSDMVQAER' | 'Complement C3' |
| 2468 | 'C3_No267' | 'AKDQLTCNK' | 'Complement C3' |
| 2469 | 'C3_No268' | 'DQLTCNK' | 'Complement C3' |
| 2470 | 'C3_No269' | 'SGIPIVTSPYQIHFTKTPK' | 'Complement C3' |
| 2471 | 'C3_No270' | 'QVREPGQDLVVLPLSITTDFIPSFR' | 'Complement C3' |
| 2472 | 'C3_No271' | 'FDLKVTIKPAPETEK' | 'Complement C3' |
| 2473 | 'C3_No272' | 'SEFPESWLWNVEDLK' | 'Complement C3' |
| 2474 | 'C3_No273' | 'ENEGFTVTAEGKGQGTLSVVTMYHAK' | 'Complement C3' |
| 2475 | 'C3_No274' | 'VLLDGVQNPRAEDLVGKSLYVSATVILHSGSDMVQAER' | 'Complement C3' |
| 2476 | 'C3_No275' | 'EALKLEEKK' | 'Complement C3' |
| 2477 | 'C3_No276' | 'IEGDHGARVVLVAVDKGVFVLNK' | 'Complement C3' |
| 2478 | 'C3_No277' | 'DICEEQVNSLPGSITKAGDFLEANYMNLQR' | 'Complement C3' |
| 2479 | 'C3_No278' | 'IRAYYENSPQQVFSTEFEVK' | 'Complement C3' |
| 2480 | 'C3_No279' | 'QPVPGQQMTLKIEGDHGAR' | 'Complement C3' |
| 2481 | 'C3_No280' | 'IHWESASLLRSEETK' | 'Complement C3' |
| 2482 | 'C3_No281' | 'NEQVEIRAVLYNYR' | 'Complement C3' |
| 2483 | 'C3_No282' | 'IEGDHGAR' | 'Complement C3' |
| 2484 | 'C3_No283' | 'QPVPGQQMTLKIEGDHGAR' | 'Complement C3' |
| 2485 | 'C3_No284' | 'TELRPGETLNVNFLLRMDR' | 'Complement C3' |
| 2486 | 'C3_No285' | 'TLDPERLGREGVQK' | 'Complement C3' |
| 2487 | 'C3_No286' | 'QELSEAEQATR' | 'Complement C3' |
| 2488 | 'C3_No287' | 'VLLDGVQNPRAEDLVGKSLYVSATVILHSGSDMVQAER' | 'Complement C3' |
| 2489 | 'C3_No288' | 'KCCEDGMR' | 'Complement C3' |
| 2490 | 'C3_No289' | 'QNQELKVR' | 'Complement C3' |
| 2491 | 'C3_No290' | 'MDKVGKYPK' | 'Complement C3' |
| 2492 | 'C3_No291' | 'GPLLNKFLTTAK' | 'Complement C3' |
| 2493 | 'C3_No292' | 'FLTTAKDK' | 'Complement C3' |
| 2494 | 'C3_No293' | 'NEQVEIRAVLYNYRQNQELK' | 'Complement C3' |
| 2495 | 'C3_No294' | 'QNQELKVRVELLHNPAFCSLATTK' | 'Complement C3' |
| 2496 | 'C3_No295' | 'GPLLNKFLTTAKDK' | 'Complement C3' |
| 2497 | 'C3_No296' | 'TVLTPATNHMGNVTFTIPANR' | 'Complement C3' |
| 2498 | 'C3_No297' | 'YYTYLIMNKGR' | 'Complement C3' |
| 2499 | 'C3_No298' | 'CAEENCFIQKSDDK' | 'Complement C3' |
| 2500 | 'C3_No299' | 'MDKVGKYPK' | 'Complement C3' |
| 2501 | 'C5_No1' | 'DGHVILQLNSIPSSDFLCVR' | 'Complement C5' |
| 2502 | 'C5_No2' | 'AFTECCVVASQLR' | 'Complement C5' |
| 2503 | 'C5_No3' | 'LNLVATPLFLKPGIPYPIK' | 'Complement C5' |
| 2504 | 'C5_No4' | 'QLPGGQNPVSYVYLEVVSK' | 'Complement C5' |
| 2505 | 'C5_No5' | 'FQNSAILTIQPK' | 'Complement C5' |
| 2506 | 'C5_No6' | 'EYVLPHFSVSIEPEYNFIGYK' | 'Complement C5' |
| 2507 | 'C5_No7' | 'AIAYSSLSQSYLYIDWTDNHK' | 'Complement C5' |
| 2508 | 'C5_No8' | 'SDLGCGAGGGLNNANVFHLAGLTFLTNANADDSQENDEPCKEILRPR' | 'Complement C5' |
| 2509 | 'C5_No9' | 'ALLVGEHLNIIVTPK' | 'Complement C5' |

| | A | B | C |
|---|---|---|---|
| 1 | **Peptide Name** | **Peptide Sequence** | **Peptide Description** |
| 2510 | 'C5_No10' | 'TLLPVSKPEIR' | 'Complement C5' |
| 2511 | 'C5_No11' | 'IDTQDIEASHYR' | 'Complement C5' |
| 2512 | 'C5_No12' | 'ITHYNYLILSK' | 'Complement C5' |
| 2513 | 'C5_No13' | 'AFDICPLVK' | 'Complement C5' |
| 2514 | 'C5_No14' | 'IPLDLVPK' | 'Complement C5' |
| 2515 | 'C5_No15' | 'EGMLSIMSYR' | 'Complement C5' |
| 2516 | 'C5_No16' | 'MSAVEGICTSESPVIDHQGTK' | 'Complement C5' |
| 2517 | 'C5_No17' | 'FSYSSGHVHLSSENK' | 'Complement C5' |
| 2518 | 'C5_No18' | 'TSTSEEVCSFYLK' | 'Complement C5' |
| 2519 | 'C5_No19' | 'SYFPESWLWEVHLVPR' | 'Complement C5' |
| 2520 | 'C5_No20' | 'KAFDICPLVK' | 'Complement C5' |
| 2521 | 'C5_No21' | 'VFQFLEK' | 'Complement C5' |
| 2522 | 'C5_No22' | 'QTACKPEIAYAYK' | 'Complement C5' |
| 2523 | 'C5_No23' | 'GGSASTWLTAFALR' | 'Complement C5' |
| 2524 | 'C5_No24' | 'LKEGMLSIMSYR' | 'Complement C5' |
| 2525 | 'C5_No25' | 'IVACASYKPSR' | 'Complement C5' |
| 2526 | 'C5_No26' | 'QCTMFYSTSNIK' | 'Complement C5' |
| 2527 | 'C5_No27' | 'NFEITIK' | 'Complement C5' |
| 2528 | 'C5_No28' | 'YGMWTIK' | 'Complement C5' |
| 2529 | 'C5_No29' | 'VFKDVFLEMNIPYSVVR' | 'Complement C5' |
| 2530 | 'C5_No30' | 'VFKDVFLEMNIPYSVVR' | 'Complement C5' |
| 2531 | 'C5_No31' | 'VYSLNDDLKPAK' | 'Complement C5' |
| 2532 | 'C5_No32' | 'VYSLNDDLKPAKR' | 'Complement C5' |
| 2533 | 'C5_No33' | 'DVFLEMNIPYSVVR' | 'Complement C5' |
| 2534 | 'C5_No34' | 'ENSLYLTAFTVIGIRK' | 'Complement C5' |
| 2535 | 'C5_No35' | 'VTCTNAELVK' | 'Complement C5' |
| 2536 | 'C5_No36' | 'YGMWTIK' | 'Complement C5' |
| 2537 | 'C5_No37' | 'ALVEGVDQLFTDYQIK' | 'Complement C5' |
| 2538 | 'C5_No38' | 'DVFLEMNIPYSVVR' | 'Complement C5' |
| 2539 | 'C5_No39' | 'GIYGTISR' | 'Complement C5' |
| 2540 | 'C5_No40' | 'MSAVEGICTSESPVIDHQGTK' | 'Complement C5' |
| 2541 | 'C5_No41' | 'QCTMFYSTSNIK' | 'Complement C5' |
| 2542 | 'C5_No42' | 'TGEAVAEKDSEITFIKK' | 'Complement C5' |
| 2543 | 'C5_No43' | 'EKFSDASYQSINIPVTQNMVPSSR' | 'Complement C5' |
| 2544 | 'C5_No44' | 'KQTACKPEIAYAYK' | 'Complement C5' |
| 2545 | 'C5_No45' | 'WLSEEQR' | 'Complement C5' |
| 2546 | 'C5_No46' | 'DNLQHKDSSVPNTGTAR' | 'Complement C5' |
| 2547 | 'C5_No47' | 'QCTMFYSTSNIK' | 'Complement C5' |
| 2548 | 'C5_No48' | 'CCYDGACVNNDETCEQR' | 'Complement C5' |
| 2549 | 'C5_No49' | 'YVLSPYK' | 'Complement C5' |
| 2550 | 'C5_No50' | 'TSGMQFCVK' | 'Complement C5' |
| 2551 | 'C5_No51' | 'MVETTAYALLTSLNLK' | 'Complement C5' |
| 2552 | 'C5_No52' | 'PGIPYPIK' | 'Complement C5' |
| 2553 | 'C5_No53' | 'SPYIDKITHYNYLILSK' | 'Complement C5' |

| | A | B | C |
|---|---|---|---|
| 1 | **Peptide Name** | **Peptide Sequence** | **Peptide Description** |
| 2554 | 'C5_No54' | 'ATLLDIYK' | 'Complement C5' |
| 2555 | 'C5_No55' | 'NADYSYSVWK' | 'Complement C5' |
| 2556 | 'C5_No56' | 'YKATLLDIYK' | 'Complement C5' |
| 2557 | 'C5_No57' | 'TGEAVAEKDSEITFIK' | 'Complement C5' |
| 2558 | 'C5_No58' | 'LSMDIDVSYK' | 'Complement C5' |
| 2559 | 'C5_No59' | 'KCCYDGACVNNDETCEQR' | 'Complement C5' |
| 2560 | 'C5_No60' | 'SIVSALKR' | 'Complement C5' |
| 2561 | 'C5_No61' | 'MVETTAYALLTSLNLK' | 'Complement C5' |
| 2562 | 'C5_No62' | 'LQGTLPVEARENSLYLTAFTVIGIRK' | 'Complement C5' |
| 2563 | 'C5_No63' | 'TDAPDLPEENQAR' | 'Complement C5' |
| 2564 | 'C5_No64' | 'RMPITYDNGFLFIHTDKPVYTPDQSVK' | 'Complement C5' |
| 2565 | 'C5_No65' | 'LKEGMLSIMSYR' | 'Complement C5' |
| 2566 | 'C5_No66' | 'QYLIMGK' | 'Complement C5' |
| 2567 | 'C5_No67' | 'RMPITYDNGFLFIHTDKPVYTPDQSVK' | 'Complement C5' |
| 2568 | 'C5_No68' | 'LKEGMLSIMSYR' | 'Complement C5' |
| 2569 | 'C5_No69' | 'VSITSITVENVFVK' | 'Complement C5' |
| 2570 | 'C5_No70' | 'CVEADCGQMQEELDLTISAETRK' | 'Complement C5' |
| 2571 | 'C5_No71' | 'LQGTLPVEAR' | 'Complement C5' |
| 2572 | 'C5_No72' | 'ELSYYSLEDLNNK' | 'Complement C5' |
| 2573 | 'C5_No73' | 'QTACKPEIAYAYK' | 'Complement C5' |
| 2574 | 'C5_No74' | 'MPITYDNGFLFIHTDKPVYTPDQSVK' | 'Complement C5' |
| 2575 | 'C5_No75' | 'MPITYDNGFLFIHTDKPVYTPDQSVK' | 'Complement C5' |
| 2576 | 'C5_No76' | 'ADNFLLENTLPAQSTFTLAISAYALSLGDKTHPQFR' | 'Complement C5' |
| 2577 | 'C5_No77' | 'EESSSGSSHAVMDISLPTGISANEEDLK' | 'Complement C5' |
| 2578 | 'C5_No78' | 'QYLIMGK' | 'Complement C5' |
| 2579 | 'C5_No79' | 'FSYSSGHVHLSSENKFQNSAILTIQPK' | 'Complement C5' |
| 2580 | 'C5_No80' | 'ENSQYQPIK' | 'Complement C5' |
| 2581 | 'C5_No81' | 'QLPGGQNPVSYVYLEVVSK' | 'Complement C5' |
| 2582 | 'C5_No82' | 'IPLDLVPKTEIKR' | 'Complement C5' |
| 2583 | 'C5_No83' | 'DINYVNPVIK' | 'Complement C5' |
| 2584 | 'C5_No84' | 'KLKEGMLSIMSYR' | 'Complement C5' |
| 2585 | 'C5_No85' | 'DSLDQLVGGVPVTLNAQTIDVNQETSDLDPSK' | 'Complement C5' |
| 2586 | 'C5_No86' | 'TSGMQFCVK' | 'Complement C5' |
| 2587 | 'C5_No87' | 'FWKDNLQHKDSSVPNTGTAR' | 'Complement C5' |
| 2588 | 'C5_No88' | 'QCTMFYSTSNIK' | 'Complement C5' |
| 2589 | 'C5_No89' | 'LSMDIDVSYKHK' | 'Complement C5' |
| 2590 | 'C5_No90' | 'ESYSGVTLDPR' | 'Complement C5' |
| 2591 | 'C5_No91' | 'VFKDVFLEMNIPYSVVRGEQIQLK' | 'Complement C5' |
| 2592 | 'C5_No92' | 'ENSLYLTAFTVIGIR' | 'Complement C5' |
| 2593 | 'C5_No93' | 'KVTCTNAELVK' | 'Complement C5' |
| 2594 | 'C5_No94' | 'LNLVATPLFLK' | 'Complement C5' |
| 2595 | 'C5_No95' | 'DSEITFIKK' | 'Complement C5' |
| 2596 | 'C5_No96' | 'DSEITFIK' | 'Complement C5' |
| 2597 | 'C5_No97' | 'VVTEADVYITFGIREDLKDDQK' | 'Complement C5' |

| | A | B | C |
|---|---|---|---|
| 1 | **Peptide Name** | **Peptide Sequence** | **Peptide Description** |
| 2598 | 'C5_No98' | 'LKEGMLSIMSYR' | 'Complement C5' |
| 2599 | 'C5_No99' | 'YKEDFSTTGTAYFEVK' | 'Complement C5' |
| 2600 | 'C5_No100' | 'GTVYNYR' | 'Complement C5' |
| 2601 | 'C5_No101' | 'LSMDIDVSYK' | 'Complement C5' |
| 2602 | 'C5_No102' | 'EALVKGNPPIYR' | 'Complement C5' |
| 2603 | 'C5_No103' | 'ATLLDIYKTGEAVAEKDSEITFIK' | 'Complement C5' |
| 2604 | 'C5_No104' | 'FSDASYQSINIPVTQNMVPSSR' | 'Complement C5' |
| 2605 | 'C5_No105' | 'RESYSGVTLDPR' | 'Complement C5' |
| 2606 | 'C5_No106' | 'SDLGCGAGGGLNNANVFHLAGLTFLTNANADDSQENDEPCK' | 'Complement C5' |
| 2607 | 'C5_No107' | 'EFPYRIPLDLVPK' | 'Complement C5' |
| 2608 | 'C5_No108' | 'QLPGGQNPVSYVYLEVVSKHFSK' | 'Complement C5' |
| 2609 | 'C5_No109' | 'VVPEGVKR' | 'Complement C5' |
| 2610 | 'C5_No110' | 'EGMLSIMSYR' | 'Complement C5' |
| 2611 | 'C5_No111' | 'IIHFGTR' | 'Complement C5' |
| 2612 | 'C5_No112' | 'KEFPYRIPLDLVPK' | 'Complement C5' |
| 2613 | 'C5_No113' | 'EKFSDASYQSINIPVTQNMVPSSR' | 'Complement C5' |
| 2614 | 'C5_No114' | 'LSMDIDVSYKHK' | 'Complement C5' |
| 2615 | 'C5_No115' | 'AKVFKDVFLEMNIPYSVVR' | 'Complement C5' |
| 2616 | 'C5_No116' | 'KIEEIAAK' | 'Complement C5' |
| 2617 | 'C5_No117' | 'ENSQYQPIKLQGTLPVEAR' | 'Complement C5' |
| 2618 | 'C5_No118' | 'FSDASYQSINIPVTQNMVPSSR' | 'Complement C5' |
| 2619 | 'C5_No119' | 'QLPGGQNPVSYVYLEVVSKHFSK' | 'Complement C5' |
| 2620 | 'C5_No120' | 'NFKNFEITIK' | 'Complement C5' |
| 2621 | 'C5_No121' | 'EGMLSIMSYR' | 'Complement C5' |
| 2622 | 'C5_No122' | 'KFSYSSGHVHLSSENK' | 'Complement C5' |
| 2623 | 'C5_No123' | 'EMMQTAMQNTMLINGIAQVTFDSETAVK' | 'Complement C5' |
| 2624 | 'C5_No124' | 'NFKNFEITIKAR' | 'Complement C5' |
| 2625 | 'C5_No125' | 'CVEADCGQMQEELDLTISAETR' | 'Complement C5' |
| 2626 | 'C5_No126' | 'GEQIQLKGTVYNYR' | 'Complement C5' |
| 2627 | 'C5_No127' | 'YKATLLDIYKTGEAVAEK' | 'Complement C5' |
| 2628 | 'C5_No128' | 'GYGNSDYKR' | 'Complement C5' |
| 2629 | 'C5_No129' | 'IVACASYKPSREESSSGSSHAVMDISLPTGISANEEDLK' | 'Complement C5' |
| 2630 | 'C5_No130' | 'AKVFKDVFLEMNIPYSVVR' | 'Complement C5' |
| 2631 | 'C5_No131' | 'CVEADCGQMQEELDLTISAETRK' | 'Complement C5' |
| 2632 | 'C5_No132' | 'ATLLDIYKTGEAVAEK' | 'Complement C5' |
| 2633 | 'C5_No133' | 'EMMQTAMQNTMLINGIAQVTFDSETAVK' | 'Complement C5' |
| 2634 | 'C5_No134' | 'EMMQTAMQNTMLINGIAQVTFDSETAVK' | 'Complement C5' |
| 2635 | 'C5_No135' | 'GNPPIYR' | 'Complement C5' |
| 2636 | 'C5_No136' | 'VCEGAACK' | 'Complement C5' |
| 2637 | 'C5_No137' | 'EDFSTTGTAYFEVK' | 'Complement C5' |
| 2638 | 'C5_No138' | 'SPYIDKITHYNYLILSKGK' | 'Complement C5' |
| 2639 | 'C5_No139' | 'GALHNYK' | 'Complement C5' |
| 2640 | 'C5_No140' | 'EMMQTAMQNTMLINGIAQVTFDSETAVK' | 'Complement C5' |
| 2641 | 'C5_No141' | 'KLKEGMLSIMSYR' | 'Complement C5' |

| | A | B | C |
|---|---|---|---|
| 1 | **Peptide Name** | **Peptide Sequence** | **Peptide Description** |
| 2642 | 'C5_No142' | 'EESSSGSSHAVMDISLPTGISANEEDLK' | 'Complement C5' |
| 2643 | 'C5_No143' | 'AFDICPLVKIDTALIK' | 'Complement C5' |
| 2644 | 'C5_No144' | 'EDLKDDQK' | 'Complement C5' |
| 2645 | 'TIMP1_No1' | 'FVYTPAMESVCGYFHR' | 'Metalloproteinase inhibitor 1' |
| 2646 | 'TIMP1_No2' | 'GFQALGDAADIR' | 'Metalloproteinase inhibitor 1' |
| 2647 | 'TIMP1_No3' | 'LQSGTHCLWTDQLLQGSEK' | 'Metalloproteinase inhibitor 1' |
| 2648 | 'TIMP1_No4' | 'EPGLCTWQSLR' | 'Metalloproteinase inhibitor 1' |
| 2649 | 'TIMP1_No5' | 'FVYTPAMESVCGYFHR' | 'Metalloproteinase inhibitor 1' |
| 2650 | 'CST3_No1' | 'LVGGPMDASVEEEGVR' | 'Cystatin-C' |
| 2651 | 'CST3_No2' | 'LVGGPMDASVEEEGVRR' | 'Cystatin-C' |
| 2652 | 'CST3_No3' | 'TQPNLDNCPFHDQPHLK' | 'Cystatin-C' |
| 2653 | 'CST3_No4' | 'AFCSFQIYAVPWQGTMTLSK' | 'Cystatin-C' |
| 2654 | 'CST3_No5' | 'LVGGPMDASVEEEGVRR' | 'Cystatin-C' |
| 2655 | 'CST3_No6' | 'AFCSFQIYAVPWQGTMTLSK' | 'Cystatin-C' |
| 2656 | 'CST3_No7' | 'TQPNLDNCPFHDQPHLKR' | 'Cystatin-C' |
| 2657 | 'CST3_No8' | 'LVGGPMDASVEEEGVR' | 'Cystatin-C' |
| 2658 | 'CST3_No9' | 'KAFCSFQIYAVPWQGTMTLSK' | 'Cystatin-C' |
| 2659 | 'CST3_No10' | 'ALDFAVGEYNK' | 'Cystatin-C' |
| 2660 | 'CST3_No11' | 'RALDFAVGEYNK' | 'Cystatin-C' |
| 2661 | 'CST3_No12' | 'KQIVAGVNYFLDVELGR' | 'Cystatin-C' |
| 2662 | 'CST3_No13' | 'KAFCSFQIYAVPWQGTMTLSK' | 'Cystatin-C' |
| 2663 | 'KNG1_No1' | 'YFIDFVAR' | 'Kininogen-1' |
| 2664 | 'KNG1_No2' | 'DIPTNSPELEETLTHTITK' | 'Kininogen-1' |
| 2665 | 'KNG1_No3' | 'KYFIDFVAR' | 'Kininogen-1' |
| 2666 | 'KNG1_No4' | 'KIYPTVNCQPLGMISLMK' | 'Kininogen-1' |
| 2667 | 'KNG1_No5' | 'TVGSDTFYSFK' | 'Kininogen-1' |
| 2668 | 'KNG1_No6' | 'TVGSDTFYSFKYEIK' | 'Kininogen-1' |
| 2669 | 'KNG1_No7' | 'QVVAGLNFR' | 'Kininogen-1' |
| 2670 | 'KNG1_No8' | 'RPPGFSPFR' | 'Kininogen-1' |
| 2671 | 'KNG1_No9' | 'TWQDCEYKDAAK' | 'Kininogen-1' |
| 2672 | 'KNG1_No10' | 'ENFLFLTPDCK' | 'Kininogen-1' |
| 2673 | 'KNG1_No11' | 'KYNSQNQSNNQFVLYR' | 'Kininogen-1' |
| 2674 | 'KNG1_No12' | 'ICVGCPR' | 'Kininogen-1' |
| 2675 | 'KNG1_No13' | 'KLGQSLDCNAEVYVVPWEK' | 'Kininogen-1' |
| 2676 | 'KNG1_No14' | 'FSVATQTCQITPAEGPVVTAQYDCLGCVHPISTQSPDLEPILR' | 'Kininogen-1' |
| 2677 | 'KNG1_No15' | 'FKLDDDLEHQGGHVLDHGHK' | 'Kininogen-1' |
| 2678 | 'KNG1_No16' | 'QVVAGLNFR' | 'Kininogen-1' |
| 2679 | 'KNG1_No17' | 'YEIKEGDCPVQSGK' | 'Kininogen-1' |
| 2680 | 'KNG1_No18' | 'ESNEELTESCETK' | 'Kininogen-1' |
| 2681 | 'KNG1_No19' | 'AATGECTATVGK' | 'Kininogen-1' |
| 2682 | 'KNG1_No20' | 'DFVQPPTK' | 'Kininogen-1' |
| 2683 | 'KNG1_No21' | 'KIYPTVNCQPLGMISLMK' | 'Kininogen-1' |
| 2684 | 'KNG1_No22' | 'IYPTVNCQPLGMISLMK' | 'Kininogen-1' |
| 2685 | 'KNG1_No23' | 'TWQDCEYK' | 'Kininogen-1' |

| | A | B | C |
|---|---|---|---|
| 1 | **Peptide Name** | **Peptide Sequence** | **Peptide Description** |
| 2686 | 'KNG1_No24' | 'ETTCSKESNEELTESCETKK' | 'Kininogen-1' |
| 2687 | 'KNG1_No25' | 'LDDDLEHQGGHVLDHGHK' | 'Kininogen-1' |
| 2688 | 'KNG1_No26' | 'IYPTVNCQPLGMISLMK' | 'Kininogen-1' |
| 2689 | 'KNG1_No27' | 'KLGQSLDCNAEVYVVPWEKK' | 'Kininogen-1' |
| 2690 | 'KNG1_No28' | 'KIYPTVNCQPLGMISLMK' | 'Kininogen-1' |
| 2691 | 'KNG1_No29' | 'KIYPTVNCQPLGMISLMK' | 'Kininogen-1' |
| 2692 | 'KNG1_No30' | 'LGQSLDCNAEVYVVPWEK' | 'Kininogen-1' |
| 2693 | 'KNG1_No31' | 'IYPTVNCQPLGMISLMK' | 'Kininogen-1' |
| 2694 | 'KNG1_No32' | 'AATGECTATVGKR' | 'Kininogen-1' |
| 2695 | 'KNG1_No33' | 'EGDCPVQSGK' | 'Kininogen-1' |
| 2696 | 'KNG1_No34' | 'AVDAALKK' | 'Kininogen-1' |
| 2697 | 'KNG1_No35' | 'FKLDDDLEHQGGHVLDHGHKHK' | 'Kininogen-1' |
| 2698 | 'KNG1_No36' | 'YNSQNQSNNQFVLYR' | 'Kininogen-1' |
| 2699 | 'KNG1_No37' | 'IYPTVNCQPLGMISLMK' | 'Kininogen-1' |
| 2700 | 'KNG1_No38' | 'LGQSLDCNAEVYVVPWEKK' | 'Kininogen-1' |
| 2701 | 'KNG1_No39' | 'ETTCSKESNEELTESCETK' | 'Kininogen-1' |
| 2702 | 'KNG1_No40' | 'IASFSQNCDIYPGKDFVQPPTK' | 'Kininogen-1' |
| 2703 | 'KNG1_No41' | 'TVGSDTFYSFKYEIKEGDCPVQSGK' | 'Kininogen-1' |
| 2704 | 'KNG1_No42' | 'IASFSQNCDIYPGK' | 'Kininogen-1' |
| 2705 | 'KNG1_No43' | 'LDDDLEHQGGHVLDHGHKHK' | 'Kininogen-1' |
| 2706 | 'KNG1_No44' | 'ESNEELTESCETKK' | 'Kininogen-1' |
| 2707 | 'KNG1_No45' | 'SVSEINPTTQMK' | 'Kininogen-1' |
| 2708 | 'KNG1_No46' | 'EGDCPVQSGKTWQDCEYKDAAK' | 'Kininogen-1' |
| 2709 | 'KNG1_No47' | 'AVDAALKKYNSQNQSNNQFVLYR' | 'Kininogen-1' |
| 2710 | 'KNG1_No48' | 'CPGRPWK' | 'Kininogen-1' |
| 2711 | 'KNG1_No49' | 'ITEATKTVGSDTFYSFK' | 'Kininogen-1' |
| 2712 | 'KNG1_No50' | 'ITEATKTVGSDTFYSFKYEIK' | 'Kininogen-1' |
| 2713 | 'KNG1_No51' | 'AQRQVVAGLNFR' | 'Kininogen-1' |
| 2714 | 'KNG1_No52' | 'GHGLGHGHEQQHGLGHGHK' | 'Kininogen-1' |
| 2715 | 'KNG1_No53' | 'VQVVAGKKYFIDFVAR' | 'Kininogen-1' |
| 2716 | 'TGFB1_No1' | 'VAGESAEPEPEPEADYYAK' | 'Transforming growth factor beta-1 proprotein' |
| 2717 | 'TGFB1_No2' | 'VEQLSNMIVR' | 'Transforming growth factor beta-1 proprotein' |
| 2718 | 'TGFB1_No3' | 'ALDTNYCFSSTEK' | 'Transforming growth factor beta-1 proprotein' |
| 2719 | 'TGFB1_No4' | 'VLMVETHNEIYDKFK' | 'Transforming growth factor beta-1 proprotein' |
| 2720 | 'TGFB1_No5' | 'LKVEQHVELYQK' | 'Transforming growth factor beta-1 proprotein' |
| 2721 | 'TGFB1_No6' | 'DNTLQVDINGFTTGR' | 'Transforming growth factor beta-1 proprotein' |
| 2722 | 'TGFB1_No7' | 'LLAPSDSPEWLSFDVTGVVR' | 'Transforming growth factor beta-1 proprotein' |
| 2723 | 'TGFB1_No8' | 'VEQLSNMIVR' | 'Transforming growth factor beta-1 proprotein' |
| 2724 | 'TGFB1_No9' | 'VLMVETHNEIYDKFK' | 'Transforming growth factor beta-1 proprotein' |
| 2725 | 'IGF2_No1' | 'SCDLALLETYCATPAK' | 'Insulin-like growth factor II' |
| 2726 | 'IGF2_No2' | 'GFYFSRPASR' | 'Insulin-like growth factor II' |
| 2727 | 'IGF2_No3' | 'GIVEECCFR' | 'Insulin-like growth factor II' |
| 2728 | 'IGF2_No4' | 'SCDLALLETYCATPAKSER' | 'Insulin-like growth factor II' |
| 2729 | 'JCHAIN_No1' | 'SSEDPNEDIVER' | 'Immunoglobulin J chain' |

| | A | B | C |
|---|---|---|---|
| 1 | **Peptide Name** | **Peptide Sequence** | **Peptide Description** |
| 2730 | 'JCHAIN_No2' | 'FVYHLSDLCK' | 'Immunoglobulin J chain' |
| 2731 | 'JCHAIN_No3' | 'FVYHLSDLCKK' | 'Immunoglobulin J chain' |
| 2732 | 'JCHAIN_No4' | 'IIVPLNNR' | 'Immunoglobulin J chain' |
| 2733 | 'JCHAIN_No5' | 'TRFVYHLSDLCK' | 'Immunoglobulin J chain' |
| 2734 | 'JCHAIN_No6' | 'IVLVDNK' | 'Immunoglobulin J chain' |
| 2735 | 'JCHAIN_No7' | 'TRFVYHLSDLCKK' | 'Immunoglobulin J chain' |
| 2736 | 'JCHAIN_No8' | 'CYTAVVPLVYGGETK' | 'Immunoglobulin J chain' |
| 2737 | 'JCHAIN_No9' | 'NKCYTAVVPLVYGGETK' | 'Immunoglobulin J chain' |
| 2738 | 'JCHAIN_No10' | 'IIRSSEDPNEDIVER' | 'Immunoglobulin J chain' |
| 2739 | 'JCHAIN_No11' | 'KCDPTEVELDNQIVTATQSNICDEDSATETCYTYDRNK' | 'Immunoglobulin J chain' |
| 2740 | 'JCHAIN_No12' | 'IIVPLNNRENISDPTSPLR' | 'Immunoglobulin J chain' |
| 2741 | 'JCHAIN_No13' | 'SSEDPNEDIVERNIR' | 'Immunoglobulin J chain' |
| 2742 | 'JCHAIN_No14' | 'CDPTEVELDNQIVTATQSNICDEDSATETCYTYDRNK' | 'Immunoglobulin J chain' |
| 2743 | 'IGKV1D-33_No1' | 'LLIYDASNLETGVPSR' | 'Immunoglobulin kappa variable 1D-33' |
| 2744 | 'IGKV1D-33_No2' | 'VTITCQASQDISNYLNWYQQKPGK' | 'Immunoglobulin kappa variable 1D-33' |
| 2745 | 'IGKV1D-33_No3' | 'VTITCQASQDISNYLNWYQQKPGKAPK' | 'Immunoglobulin kappa variable 1D-33' |
| 2746 | 'IGKV1D-33_No4' | 'VTITCQASQDISNYLNWYQQK' | 'Immunoglobulin kappa variable 1D-33' |
| 2747 | 'IGKV1D-33_No5' | 'APKLLIYDASNLETGVPSR' | 'Immunoglobulin kappa variable 1D-33' |
| 2748 | 'IGKV1-39_No1' | 'ASQSISSYLNWYQQKPGK' | 'Immunoglobulin kappa variable 1-39' |
| 2749 | 'IGKV1-39_No2' | 'VTITCRASQSISSYLNWYQQKPGK' | 'Immunoglobulin kappa variable 1-39' |
| 2750 | 'IGKV1-39_No3' | 'ASQSISSYLNWYQQK' | 'Immunoglobulin kappa variable 1-39' |
| 2751 | 'IGKV1-39_No4' | 'ASQSISSYLNWYQQKPGKAPK' | 'Immunoglobulin kappa variable 1-39' |
| 2752 | 'IGKV1-17_No1' | 'LIYAASSLQSGVPSR' | 'Immunoglobulin kappa variable 1-17' |
| 2753 | 'IGKV1-17_No2' | 'RLIYAASSLQSGVPSR' | 'Immunoglobulin kappa variable 1-17' |
| 2754 | 'IGKV1D-16_No1' | 'ASQGISSWLAWYQQKPEK' | 'Immunoglobulin kappa variable 1D-16' |
| 2755 | 'IGKV1D-16_No2' | 'SLIYAASSLQSGVPSR' | 'Immunoglobulin kappa variable 1D-16' |
| 2756 | 'IGKV1D-16_No3' | 'APKSLIYAASSLQSGVPSR' | 'Immunoglobulin kappa variable 1D-16' |
| 2757 | 'IGKV1-5_No1' | 'ASQSISSWLAWYQQKPGK' | 'Immunoglobulin kappa variable 1-5' |
| 2758 | 'IGKV1-5_No2' | 'ASQSISSWLAWYQQK' | 'Immunoglobulin kappa variable 1-5' |
| 2759 | 'IGKV1-5_No3' | 'LLIYKASSLESGVPSR' | 'Immunoglobulin kappa variable 1-5' |
| 2760 | 'IGKV1-5_No4' | 'APKLLIYKASSLESGVPSR' | 'Immunoglobulin kappa variable 1-5' |
| 2761 | 'IGKV1-5_No5' | 'ASQSISSWLAWYQQKPGKAPK' | 'Immunoglobulin kappa variable 1-5' |
| 2762 | 'IGKV3-20_No1' | 'FSGSGSGTDFTLTISR' | 'Immunoglobulin kappa variable 3-20' |
| 2763 | 'IGKV3-20_No2' | 'ASQSVSSSYLAWYQQK' | 'Immunoglobulin kappa variable 3-20' |
| 2764 | 'IGKV3-20_No3' | 'ATGIPDRFSGSGSGTDFTLTISR' | 'Immunoglobulin kappa variable 3-20' |
| 2765 | 'IGKV3-20_No4' | 'LLIYGASSR' | 'Immunoglobulin kappa variable 3-20' |
| 2766 | 'IGKV3-20_No5' | 'ASQSVSSSYLAWYQQKPGQAPR' | 'Immunoglobulin kappa variable 3-20' |
| 2767 | 'IGKV3-15_No1' | 'ASQSVSSNLAWYQQK' | 'Immunoglobulin kappa variable 3-15' |
| 2768 | 'IGKV3-15_No2' | 'ASQSVSSNLAWYQQKPGQAPR' | 'Immunoglobulin kappa variable 3-15' |
| 2769 | 'IGKV3-15_No3' | 'LLIYGASTR' | 'Immunoglobulin kappa variable 3-15' |
| 2770 | 'IGLV1-44_No1' | 'LLIYSNNQR' | 'Immunoglobulin lambda variable 1-44' |
| 2771 | 'IGLV1-44_No2' | 'LLIYSNNQRPSGVPDR' | 'Immunoglobulin lambda variable 1-44' |
| 2772 | 'IGLV1-44_No3' | 'LLIYSNNQRPSGVPDRFSGSK' | 'Immunoglobulin lambda variable 1-44' |
| 2773 | 'IGLV1-44_No4' | 'VTISCSGSSSNIGSNTVNWYQQLPGTAPK' | 'Immunoglobulin lambda variable 1-44' |

| | A | B | C |
|---|---|---|---|
| 1 | **Peptide Name** | **Peptide Sequence** | **Peptide Description** |
| 2774 | 'IGLV1-47_No1' | 'VTISCSGSSSNIGSNYVYWYQQLPGTAPK' | 'Immunoglobulin lambda variable 1-47' |
| 2775 | 'IGLV1-47_No2' | 'SGTSASLAISGLR' | 'Immunoglobulin lambda variable 1-47' |
| 2776 | 'IGLV1-51_No1' | 'LLIYDNNKRPSGIPDR' | 'Immunoglobulin lambda variable 1-51' |
| 2777 | 'IGLV1-51_No2' | 'VTISCSGSSSNIGNNYVSWYQQLPGTAPK' | 'Immunoglobulin lambda variable 1-51' |
| 2778 | 'IGLV1-51_No3' | 'RPSGIPDRFSGSK' | 'Immunoglobulin lambda variable 1-51' |
| 2779 | 'IGLV1-51_No4' | 'LLIYDNNK' | 'Immunoglobulin lambda variable 1-51' |
| 2780 | 'IGLV1-51_No5' | 'LLIYDNNKR' | 'Immunoglobulin lambda variable 1-51' |
| 2781 | 'IGLV1-51_No6' | 'RPSGIPDR' | 'Immunoglobulin lambda variable 1-51' |
| 2782 | 'IGLV1-40_No1' | 'LLIYGNSNRPSGVPDR' | 'Immunoglobulin lambda variable 1-40' |
| 2783 | 'IGLV1-40_No2' | 'VTISCTGSSSNIGAGYDVHWYQQLPGTAPK' | 'Immunoglobulin lambda variable 1-40' |
| 2784 | 'IGLV1-40_No3' | 'LLIYGNSNRPSGVPDRFSGSK' | 'Immunoglobulin lambda variable 1-40' |
| 2785 | 'IGLV2-14_No1' | 'LMIYEVSNRPSGVSNR' | 'Immunoglobulin lambda variable 2-14' |
| 2786 | 'IGLV2-14_No2' | 'LMIYEVSNRPSGVSNR' | 'Immunoglobulin lambda variable 2-14' |
| 2787 | 'IGLV2-23_No1' | 'LMIYEGSK' | 'Immunoglobulin lambda variable 2-23' |
| 2788 | 'IGLV2-23_No2' | 'LMIYEGSKRPSGVSNR' | 'Immunoglobulin lambda variable 2-23' |
| 2789 | 'IGLV2-23_No3' | 'LMIYEGSKRPSGVSNR' | 'Immunoglobulin lambda variable 2-23' |
| 2790 | 'IGLV2-23_No4' | 'RPSGVSNR' | 'Immunoglobulin lambda variable 2-23' |
| 2791 | 'IGLV2-23_No5' | 'LMIYEGSK' | 'Immunoglobulin lambda variable 2-23' |
| 2792 | 'IGLV2-11_No1' | 'LMIYDVSK' | 'Immunoglobulin lambda variable 2-11' |
| 2793 | 'IGLV2-11_No2' | 'LMIYDVSK' | 'Immunoglobulin lambda variable 2-11' |
| 2794 | 'IGLV2-11_No3' | 'SVSGSPGQSVTISCTGTSSDVGGYNYVSWYQQHPGKAPK' | 'Immunoglobulin lambda variable 2-11' |
| 2795 | 'IGLV2-11_No4' | 'LMIYDVSKRPSGVPDR' | 'Immunoglobulin lambda variable 2-11' |
| 2796 | 'IGLV2-11_No5' | 'LMIYDVSKRPSGVPDR' | 'Immunoglobulin lambda variable 2-11' |
| 2797 | 'IGLV2-8_No1' | 'LMIYEVSK' | 'Immunoglobulin lambda variable 2-8' |
| 2798 | 'IGLV2-8_No2' | 'LMIYEVSKRPSGVPDR' | 'Immunoglobulin lambda variable 2-8' |
| 2799 | 'IGLV2-8_No3' | 'LMIYEVSKR' | 'Immunoglobulin lambda variable 2-8' |
| 2800 | 'IGLV2-8_No4' | 'LMIYEVSK' | 'Immunoglobulin lambda variable 2-8' |
| 2801 | 'IGLV2-8_No5' | 'LMIYEVSKRPSGVPDR' | 'Immunoglobulin lambda variable 2-8' |
| 2802 | 'IGLV2-8_No6' | 'LMIYEVSKR' | 'Immunoglobulin lambda variable 2-8' |
| 2803 | 'IGLV3-19_No1' | 'SYYASWYQQKPGQAPVLVIYGK' | 'Immunoglobulin lambda variable 3-19' |
| 2804 | 'IGLV3-19_No2' | 'ITCQGDSLR' | 'Immunoglobulin lambda variable 3-19' |
| 2805 | 'IGLV3-19_No3' | 'PGQAPVLVIYGK' | 'Immunoglobulin lambda variable 3-19' |
| 2806 | 'IGLV3-19_No4' | 'NNRPSGIPDR' | 'Immunoglobulin lambda variable 3-19' |
| 2807 | 'IGLV3-19_No5' | 'SYYASWYQQK' | 'Immunoglobulin lambda variable 3-19' |
| 2808 | 'IGLV3-19_No6' | 'FSGSSSGNTASLTITGAQAEDEADYYCNSR' | 'Immunoglobulin lambda variable 3-19' |
| 2809 | 'IGLV3-19_No7' | 'NNRPSGIPDRFSGSSSGNTASLTITGAQAEDEADYYCNSR' | 'Immunoglobulin lambda variable 3-19' |
| 2810 | 'IGLV3-19_No8' | 'PGQAPVLVIYGKNNRPSGIPDR' | 'Immunoglobulin lambda variable 3-19' |
| 2811 | 'IGLV3-1_No1' | 'YACWYQQKPGQSPVLVIYQDSK' | 'Immunoglobulin lambda variable 3-1' |
| 2812 | 'IGLV3-1_No2' | 'PGQSPVLVIYQDSK' | 'Immunoglobulin lambda variable 3-1' |
| 2813 | 'IGLV3-1_No3' | 'YACWYQQK' | 'Immunoglobulin lambda variable 3-1' |
| 2814 | 'IGLV3-1_No4' | 'LGDKYACWYQQKPGQSPVLVIYQDSK' | 'Immunoglobulin lambda variable 3-1' |
| 2815 | 'IGLV3-1_No5' | 'LGDKYACWYQQK' | 'Immunoglobulin lambda variable 3-1' |
| 2816 | 'IGLV3-25_No1' | 'QYAYWYQQKPGQAPVLVIYK' | 'Immunoglobulin lambda variable 3-25' |
| 2817 | 'IGLV3-25_No2' | 'ITCSGDALPK' | 'Immunoglobulin lambda variable 3-25' |

| | A | B | C |
|---|---|---|---|
| 1 | **Peptide Name** | **Peptide Sequence** | **Peptide Description** |
| 2818 | 'IGLV3-25_No3' | 'DSERPSGIPER' | 'Immunoglobulin lambda variable 3-25' |
| 2819 | 'IGLV3-25_No4' | 'QYAYWYQQKPGQAPVLVIYK' | 'Immunoglobulin lambda variable 3-25' |
| 2820 | 'IGLV3-25_No5' | 'QYAYWYQQK' | 'Immunoglobulin lambda variable 3-25' |
| 2821 | 'IGLV3-25_No6' | 'PGQAPVLVIYK' | 'Immunoglobulin lambda variable 3-25' |
| 2822 | 'IGLV3-25_No7' | 'ITCSGDALPKQYAYWYQQK' | 'Immunoglobulin lambda variable 3-25' |
| 2823 | 'IGLV3-27_No1' | 'WFQQKPGQAPVLVIYK' | 'Immunoglobulin lambda variable 3-27' |
| 2824 | 'IGLV3-27_No2' | 'ITCSGDVLAK' | 'Immunoglobulin lambda variable 3-27' |
| 2825 | 'IGLV6-57_No1' | 'FSGSIDSSSNSASLTISGLK' | 'Immunoglobulin lambda variable 6-57' |
| 2826 | 'IGLV6-57_No2' | 'PGSAPTTVIYEDNQRPSGVPDR' | 'Immunoglobulin lambda variable 6-57' |
| 2827 | 'IGLV6-57_No3' | 'PGSAPTTVIYEDNQR' | 'Immunoglobulin lambda variable 6-57' |
| 2828 | 'IGHV1-69_No1' | 'STSTAYMELSSLR' | 'Immunoglobulin heavy variable 1-69' |
| 2829 | 'IGHV1-69_No2' | 'STSTAYMELSSLR' | 'Immunoglobulin heavy variable 1-69' |
| 2830 | 'IGHV1-69_No3' | 'ASGGTFSSYAISWVR' | 'Immunoglobulin heavy variable 1-69' |
| 2831 | 'IGHV1-69_No4' | 'SEDTAVYYCAR' | 'Immunoglobulin heavy variable 1-69' |
| 2832 | 'IGHV1-69_No5' | 'VTITADKSTSTAYMELSSLR' | 'Immunoglobulin heavy variable 1-69' |
| 2833 | 'IGHV1-69_No6' | 'QAPGQGLEWMGGIIPIFGTANYAQK' | 'Immunoglobulin heavy variable 1-69' |
| 2834 | 'IGHV1-69_No7' | 'QAPGQGLEWMGGIIPIFGTANYAQK' | 'Immunoglobulin heavy variable 1-69' |
| 2835 | 'IGHV1-69_No8' | 'VTITADKSTSTAYMELSSLR' | 'Immunoglobulin heavy variable 1-69' |
| 2836 | 'IGHV1-69_No9' | 'STSTAYMELSSLRSEDTAVYYCAR' | 'Immunoglobulin heavy variable 1-69' |
| 2837 | 'IGHV1-46_No1' | 'DTSTSTVYMELSSLR' | 'Immunoglobulin heavy variable 1-46' |
| 2838 | 'IGHV1-46_No2' | 'DTSTSTVYMELSSLR' | 'Immunoglobulin heavy variable 1-46' |
| 2839 | 'IGHV1-46_No3' | 'ASGYTFTSYYMHWVR' | 'Immunoglobulin heavy variable 1-46' |
| 2840 | 'IGHV1-46_No4' | 'ASGYTFTSYYMHWVR' | 'Immunoglobulin heavy variable 1-46' |
| 2841 | 'IGHV1-46_No5' | 'QAPGQGLEWMGIINPSGGSTSYAQK' | 'Immunoglobulin heavy variable 1-46' |
| 2842 | 'IGHV3-11_No1' | 'LSCAASGFTFSDYYMSWIR' | 'Immunoglobulin heavy variable 3-11' |
| 2843 | 'IGHV3-11_No2' | 'LSCAASGFTFSDYYMSWIR' | 'Immunoglobulin heavy variable 3-11' |
| 2844 | 'IGHV3-48_No1' | 'LSCAASGFTFSSYEMNWVR' | 'Immunoglobulin heavy variable 3-48' |
| 2845 | 'IGHV3-48_No2' | 'LSCAASGFTFSSYEMNWVR' | 'Immunoglobulin heavy variable 3-48' |
| 2846 | 'IGHV3-23_No1' | 'AEDTAVYYCAK' | 'Immunoglobulin heavy variable 3-23' |
| 2847 | 'IGHV3-23_No2' | 'GLEWVSAISGSGGSTYYADSVK' | 'Immunoglobulin heavy variable 3-23' |
| 2848 | 'IGHV3-23_No3' | 'LSCAASGFTFSSYAMSWVR' | 'Immunoglobulin heavy variable 3-23' |
| 2849 | 'IGHV3-23_No4' | 'LSCAASGFTFSSYAMSWVR' | 'Immunoglobulin heavy variable 3-23' |
| 2850 | 'IGHV3-23_No5' | 'QAPGKGLEWVSAISGSGGSTYYADSVK' | 'Immunoglobulin heavy variable 3-23' |
| 2851 | 'IGHV3-23_No6' | 'DNSKNTLYLQMNSLRAEDTAVYYCAK' | 'Immunoglobulin heavy variable 3-23' |
| 2852 | 'IGHV3-23_No7' | 'NTLYLQMNSLRAEDTAVYYCAK' | 'Immunoglobulin heavy variable 3-23' |
| 2853 | 'IGHV3-23_No8' | 'DNSKNTLYLQMNSLRAEDTAVYYCAK' | 'Immunoglobulin heavy variable 3-23' |
| 2854 | 'IGHV3-23_No9' | 'NTLYLQMNSLRAEDTAVYYCAK' | 'Immunoglobulin heavy variable 3-23' |
| 2855 | 'IGHV3-23_No10' | 'GLEWVSAISGSGGSTYYADSVKGR' | 'Immunoglobulin heavy variable 3-23' |
| 2856 | 'IGHV3-13_No1' | 'AGDTAVYYCAR' | 'Immunoglobulin heavy variable 3-13' |
| 2857 | 'IGHV3-13_No2' | 'ENAKNSLYLQMNSLR' | 'Immunoglobulin heavy variable 3-13' |
| 2858 | 'IGHV3-13_No3' | 'LSCAASGFTFSSYDMHWVR' | 'Immunoglobulin heavy variable 3-13' |
| 2859 | 'IGHV3-13_No4' | 'LSCAASGFTFSSYDMHWVR' | 'Immunoglobulin heavy variable 3-13' |
| 2860 | 'IGHV3-13_No5' | 'ENAKNSLYLQMNSLR' | 'Immunoglobulin heavy variable 3-13' |
| 2861 | 'IGHV3-53_No1' | 'LSCAASGFTVSSNYMSWVR' | 'Immunoglobulin heavy variable 3-53' |

| | A | B | C |
|---|---|---|---|
| 1 | **Peptide Name** | **Peptide Sequence** | **Peptide Description** |
| 2862 | 'IGHV3-53_No2' | 'GLEWVSVIYSGGSTYYADSVK' | 'Immunoglobulin heavy variable 3-53' |
| 2863 | 'IGHV3-53_No3' | 'LSCAASGFTVSSNYMSWVR' | 'Immunoglobulin heavy variable 3-53' |
| 2864 | 'IGHV3-53_No4' | 'QAPGKGLEWVSVIYSGGSTYYADSVK' | 'Immunoglobulin heavy variable 3-53' |
| 2865 | 'IGHV3-30_No1' | 'LSCAASGFTFSSYGMHWVR' | 'Immunoglobulin heavy variable 3-30' |
| 2866 | 'IGHV3-30_No2' | 'LSCAASGFTFSSYGMHWVR' | 'Immunoglobulin heavy variable 3-30' |
| 2867 | 'IGHV3-7_No1' | 'GLEWVANIK' | 'Immunoglobulin heavy variable 3-7' |
| 2868 | 'IGHV3-7_No2' | 'NSLYLQMNSLRAEDTAVYYCAR' | 'Immunoglobulin heavy variable 3-7' |
| 2869 | 'IGHV3-7_No3' | 'YYVDSVK' | 'Immunoglobulin heavy variable 3-7' |
| 2870 | 'IGHV3-7_No4' | 'GLEWVANIKQDGSEK' | 'Immunoglobulin heavy variable 3-7' |
| 2871 | 'IGHV3-7_No5' | 'QDGSEKYYVDSVK' | 'Immunoglobulin heavy variable 3-7' |
| 2872 | 'IGHV3-7_No6' | 'LSCAASGFTFSSYWMSWVR' | 'Immunoglobulin heavy variable 3-7' |
| 2873 | 'IGHV3-7_No7' | 'NSLYLQMNSLRAEDTAVYYCAR' | 'Immunoglobulin heavy variable 3-7' |
| 2874 | 'IGHV3-7_No8' | 'QDGSEKYYVDSVK' | 'Immunoglobulin heavy variable 3-7' |
| 2875 | 'IGHV3-7_No9' | 'QAPGKGLEWVANIK' | 'Immunoglobulin heavy variable 3-7' |
| 2876 | 'IGHV3-7_No10' | 'QAPGKGLEWVANIK' | 'Immunoglobulin heavy variable 3-7' |
| 2877 | 'IGHV3-7_No11' | 'GLEWVANIKQDGSEKYYVDSVK' | 'Immunoglobulin heavy variable 3-7' |
| 2878 | 'IGHV3-7_No12' | 'LSCAASGFTFSSYWMSWVR' | 'Immunoglobulin heavy variable 3-7' |
| 2879 | 'IGHV3-7_No13' | 'QDGSEKYYVDSVKGR' | 'Immunoglobulin heavy variable 3-7' |
| 2880 | 'IGHV3-7_No14' | 'DNAKNSLYLQMNSLRAEDTAVYYCAR' | 'Immunoglobulin heavy variable 3-7' |
| 2881 | 'IGHV3-7_No15' | 'QAPGKGLEWVANIKQDGSEK' | 'Immunoglobulin heavy variable 3-7' |
| 2882 | 'IGHV3-7_No16' | 'DNAKNSLYLQMNSLRAEDTAVYYCAR' | 'Immunoglobulin heavy variable 3-7' |
| 2883 | 'IGHV3-7_No17' | 'QAPGKGLEWVANIKQDGSEK' | 'Immunoglobulin heavy variable 3-7' |
| 2884 | 'IGHV3-9_No1' | 'NSLYLQMNSLR' | 'Immunoglobulin heavy variable 3-9' |
| 2885 | 'IGHV3-9_No2' | 'AEDTALYYCAK' | 'Immunoglobulin heavy variable 3-9' |
| 2886 | 'IGHV3-9_No3' | 'LSCAASGFTFDDYAMHWVR' | 'Immunoglobulin heavy variable 3-9' |
| 2887 | 'IGHV3-9_No4' | 'LSCAASGFTFDDYAMHWVR' | 'Immunoglobulin heavy variable 3-9' |
| 2888 | 'IGHV3-9_No5' | 'GLEWVSGISWNSGSIGYADSVK' | 'Immunoglobulin heavy variable 3-9' |
| 2889 | 'IGHV3-9_No6' | 'DNAKNSLYLQMNSLR' | 'Immunoglobulin heavy variable 3-9' |
| 2890 | 'IGHV3-9_No7' | 'NSLYLQMNSLR' | 'Immunoglobulin heavy variable 3-9' |
| 2891 | 'IGHV3-9_No8' | 'NSLYLQMNSLRAEDTALYYCAK' | 'Immunoglobulin heavy variable 3-9' |
| 2892 | 'IGHV3-9_No9' | 'AEDTALYYCAKD' | 'Immunoglobulin heavy variable 3-9' |
| 2893 | 'IGHV3-9_No10' | 'NSLYLQMNSLRAEDTALYYCAK' | 'Immunoglobulin heavy variable 3-9' |
| 2894 | 'IGHV3-9_No11' | 'DNAKNSLYLQMNSLR' | 'Immunoglobulin heavy variable 3-9' |
| 2895 | 'IGHV3-9_No12' | 'FTISRDNAK' | 'Immunoglobulin heavy variable 3-9' |
| 2896 | 'IGHV3-9_No13' | 'QAPGKGLEWVSGISWNSGSIGYADSVK' | 'Immunoglobulin heavy variable 3-9' |
| 2897 | 'IGHV2-5_No1' | 'NQVVLTMTNMDPVDTATYYCAHR' | 'Immunoglobulin heavy variable 2-5' |
| 2898 | 'IGHV2-5_No2' | 'NQVVLTMTNMDPVDTATYYCAHR' | 'Immunoglobulin heavy variable 2-5' |
| 2899 | 'IGHV2-5_No3' | 'NQVVLTMTNMDPVDTATYYCAHR' | 'Immunoglobulin heavy variable 2-5' |
| 2900 | 'IGHV2-5_No4' | 'NQVVLTMTNMDPVDTATYYCAHR' | 'Immunoglobulin heavy variable 2-5' |
| 2901 | 'IGHV2-5_No5' | 'DTSKNQVVLTMTNMDPVDTATYYCAHR' | 'Immunoglobulin heavy variable 2-5' |
| 2902 | 'IGHV2-5_No6' | 'DTSKNQVVLTMTNMDPVDTATYYCAHR' | 'Immunoglobulin heavy variable 2-5' |
| 2903 | 'IGHV2-5_No7' | 'ESGPTLVK' | 'Immunoglobulin heavy variable 2-5' |
| 2904 | 'IGHV4-39' | 'GLEWIGSIYYSGSTYYNPSLK' | 'Immunoglobulin heavy variable 4-39' |
| 2905 | 'PIGR_No1' | 'QSSGENCDVVVNTLGK' | 'Polymeric immunoglobulin receptor' |

| | A | B | C |
|---|---|---|---|
| 1 | **Peptide Name** | **Peptide Sequence** | **Peptide Description** |
| 2906 | 'PIGR_No2' | 'ADEGWYWCGVK' | 'Polymeric immunoglobulin receptor' |
| 2907 | 'PIGR_No3' | 'YWCLWEGAQNGR' | 'Polymeric immunoglobulin receptor' |
| 2908 | 'PIGR_No4' | 'LSDAGQYLCQAGDDSNSNKK' | 'Polymeric immunoglobulin receptor' |
| 2909 | 'PIGR_No5' | 'NADLQVLKPEPELVYEDLR' | 'Polymeric immunoglobulin receptor' |
| 2910 | 'PIGR_No6' | 'ILLNPQDKDGSFSVVITGLR' | 'Polymeric immunoglobulin receptor' |
| 2911 | 'PIGR_No7' | 'WNNTGCQALPSQDEGPSK' | 'Polymeric immunoglobulin receptor' |
| 2912 | 'PIGR_No8' | 'LVSLTLNLVTR' | 'Polymeric immunoglobulin receptor' |
| 2913 | 'PIGR_No9' | 'GGCITLISSEGYVSSK' | 'Polymeric immunoglobulin receptor' |
| 2914 | 'PIGR_No10' | 'CPLLVDSEGWVK' | 'Polymeric immunoglobulin receptor' |
| 2915 | 'PIGR_No11' | 'GSVTFHCALGPEVANVAK' | 'Polymeric immunoglobulin receptor' |
| 2916 | 'PIGR_No12' | 'TVTINCPFK' | 'Polymeric immunoglobulin receptor' |
| 2917 | 'PIGR_No13' | 'GVAGGSVAVLCPYNRK' | 'Polymeric immunoglobulin receptor' |
| 2918 | 'PIGR_No14' | 'TVTINCPFKTENAQK' | 'Polymeric immunoglobulin receptor' |
| 2919 | 'PIGR_No15' | 'ILLNPQDKDGSFSVVITGLRK' | 'Polymeric immunoglobulin receptor' |
| 2920 | 'PIGR_No16' | 'VLDSGFREIENK' | 'Polymeric immunoglobulin receptor' |
| 2921 | 'PIGR_No17' | 'NADLQVLKPEPELVYEDLRGSVTFHCALGPEVANVAK' | 'Polymeric immunoglobulin receptor' |
| 2922 | 'PIGR_No18' | 'LSDAGQYLCQAGDDSNSNK' | 'Polymeric immunoglobulin receptor' |
| 2923 | 'PIGR_No19' | 'ADAAPDEKVLDSGFR' | 'Polymeric immunoglobulin receptor' |
| 2924 | 'PIGR_No20' | 'ADAAPDEKVLDSGFREIENK' | 'Polymeric immunoglobulin receptor' |
| 2925 | 'PIGR_No21' | 'QSSGENCDVVVNTLGK' | 'Polymeric immunoglobulin receptor' |
| 2926 | 'PIGR_No22' | 'SIKYWCLWEGAQNGR' | 'Polymeric immunoglobulin receptor' |
| 2927 | 'PIGR_No23' | 'QGHFYGETAAVYVAVEERK' | 'Polymeric immunoglobulin receptor' |
| 2928 | 'PIGR_No24' | 'QGHFYGETAAVYVAVEER' | 'Polymeric immunoglobulin receptor' |
| 2929 | 'PIGR_No25' | 'QGHFYGETAAVYVAVEER' | 'Polymeric immunoglobulin receptor' |
| 2930 | 'PIGR_No26' | 'TVTINCPFKTENAQKR' | 'Polymeric immunoglobulin receptor' |
| 2931 | 'PIGR_No27' | 'QSSGENCDVVVNTLGKR' | 'Polymeric immunoglobulin receptor' |
| 2932 | 'IGKC_No1' | 'TVAAPSVFIFPPSDEQLK' | 'Immunoglobulin kappa constant' |
| 2933 | 'IGKC_No2' | 'TVAAPSVFIFPPSDEQLKSGTASVVCLLNNFYPR' | 'Immunoglobulin kappa constant' |
| 2934 | 'IGKC_No3' | 'RTVAAPSVFIFPPSDEQLK' | 'Immunoglobulin kappa constant' |
| 2935 | 'IGKC_No4' | 'RTVAAPSVFIFPPSDEQLKSGTASVVCLLNNFYPR' | 'Immunoglobulin kappa constant' |
| 2936 | 'IGHE_No1' | 'TYTCQVTYQGHTFEDSTKK' | 'Immunoglobulin heavy constant epsilon' |
| 2937 | 'IGHE_No2' | 'TYTCQVTYQGHTFEDSTK' | 'Immunoglobulin heavy constant epsilon' |
| 2938 | 'IGHE_No3' | 'AEWEQKDEFICR' | 'Immunoglobulin heavy constant epsilon' |
| 2939 | 'IGHG1_No1' | 'ASTKGPSVFPLAPSSK' | 'Immunoglobulin heavy constant gamma 1' |
| 2940 | 'IGHG2_No1' | 'VVSVLTVVHQDWLNGKEYK' | 'Immunoglobulin heavy constant gamma 2' |
| 2941 | 'IGHG2_No2' | 'EPQVYTLPPSREEMTK' | 'Immunoglobulin heavy constant gamma 2' |
| 2942 | 'IGHG2_No3' | 'TTPPMLDSDGSFFLYSK' | 'Immunoglobulin heavy constant gamma 2' |
| 2943 | 'IGHG2_No4' | 'EPQVYTLPPSREEMTK' | 'Immunoglobulin heavy constant gamma 2' |
| 2944 | 'IGHG2_No5' | 'CCVECPPCPAPPVAGPSVFLFPPKPK' | 'Immunoglobulin heavy constant gamma 2' |
| 2945 | 'IGHG2_No6' | 'VVSVLTVVHQDWLNGK' | 'Immunoglobulin heavy constant gamma 2' |
| 2946 | 'IGHG2_No7' | 'EPQVYTLPPSREEMTKNQVSLTCLVK' | 'Immunoglobulin heavy constant gamma 2' |
| 2947 | 'IGHG2_No8' | 'STSESTAALGCLVK' | 'Immunoglobulin heavy constant gamma 2' |
| 2948 | 'IGHG2_No9' | 'TPEVTCVVVDVSHEDPEVQFNWYVDGVEVHNAK' | 'Immunoglobulin heavy constant gamma 2' |
| 2949 | 'IGHG2_No10' | 'GPSVFPLAPCSRSTSESTAALGCLVK' | 'Immunoglobulin heavy constant gamma 2' |

| | A | B | C |
|---|---|---|---|
| 1 | **Peptide Name** | **Peptide Sequence** | **Peptide Description** |
| 2950 | 'IGHG2_No11' | 'KCCVECPPCPAPPVAGPSVFLFPPKPK' | 'Immunoglobulin heavy constant gamma 2' |
| 2951 | 'IGHG2_No12' | 'EEMTKNQVSLTCLVK' | 'Immunoglobulin heavy constant gamma 2' |
| 2952 | 'IGHG2_No13' | 'VSNKGLPAPIEK' | 'Immunoglobulin heavy constant gamma 2' |
| 2953 | 'IGHG2_No14' | 'GQPREPQVYTLPPSREEMTK' | 'Immunoglobulin heavy constant gamma 2' |
| 2954 | 'IGHG2_No15' | 'GQPREPQVYTLPPSREEMTK' | 'Immunoglobulin heavy constant gamma 2' |
| 2955 | 'IGHG2_No16' | 'TKGQPREPQVYTLPPSR' | 'Immunoglobulin heavy constant gamma 2' |
| 2956 | 'IGHG2_No17' | 'GPSVFPLAPCSR' | 'Immunoglobulin heavy constant gamma 2' |
| 2957 | 'IGHG2_No18' | 'TTPPMLDSDGSFFLYSKLTVDKSR' | 'Immunoglobulin heavy constant gamma 2' |
| 2958 | 'IGHG2_No19' | 'EPQVYTLPPSREEMTKNQVSLTCLVK' | 'Immunoglobulin heavy constant gamma 2' |
| 2959 | 'IGHG2_No20' | 'GFYPSDISVEWESNGQPENNYK' | 'Immunoglobulin heavy constant gamma 2' |
| 2960 | 'IGHG2_No21' | 'TTPPMLDSDGSFFLYSKLTVDK' | 'Immunoglobulin heavy constant gamma 2' |
| 2961 | 'IGHG2_No22' | 'TTPPMLDSDGSFFLYSKLTVDKSR' | 'Immunoglobulin heavy constant gamma 2' |
| 2962 | 'IGHG2_No23' | 'GLPAPIEK' | 'Immunoglobulin heavy constant gamma 2' |
| 2963 | 'IGHG2_No24' | 'TKPREEQFNSTFR' | 'Immunoglobulin heavy constant gamma 2' |
| 2964 | 'IGHG2_No25' | 'TTPPMLDSDGSFFLYSK' | 'Immunoglobulin heavy constant gamma 2' |
| 2965 | 'IGHG2_No26' | 'GLPAPIEKTISK' | 'Immunoglobulin heavy constant gamma 2' |
| 2966 | 'IGHG2_No27' | 'EEMTKNQVSLTCLVK' | 'Immunoglobulin heavy constant gamma 2' |
| 2967 | 'IGHG2_No28' | 'TTPPMLDSDGSFFLYSKLTVDK' | 'Immunoglobulin heavy constant gamma 2' |
| 2968 | 'IGHG2_No29' | 'VSNKGLPAPIEKTISK' | 'Immunoglobulin heavy constant gamma 2' |
| 2969 | 'IGHG2_No30' | 'VDKTVER' | 'Immunoglobulin heavy constant gamma 2' |
| 2970 | 'IGHG2_No31' | 'CKVSNKGLPAPIEK' | 'Immunoglobulin heavy constant gamma 2' |
| 2971 | 'IGHG3_No1' | 'WYVDGVEVHNAK' | 'Immunoglobulin heavy constant gamma 3' |
| 2972 | 'IGHG3_No2' | 'TPEVTCVVVDVSHEDPEVQFK' | 'Immunoglobulin heavy constant gamma 3' |
| 2973 | 'IGHG3_No3' | 'SCDTPPPCPR' | 'Immunoglobulin heavy constant gamma 3' |
| 2974 | 'IGHG3_No4' | 'TPLGDTTHTCPR' | 'Immunoglobulin heavy constant gamma 3' |
| 2975 | 'IGHG3_No5' | 'WQQGNIFSCSVMHEALHNR' | 'Immunoglobulin heavy constant gamma 3' |
| 2976 | 'IGHG3_No6' | 'VELKTPLGDTTHTCPR' | 'Immunoglobulin heavy constant gamma 3' |
| 2977 | 'IGHG3_No7' | 'WQQGNIFSCSVMHEALHNR' | 'Immunoglobulin heavy constant gamma 3' |
| 2978 | 'IGHG3_No8' | 'CPAPELLGGPSVFLFPPKPK' | 'Immunoglobulin heavy constant gamma 3' |
| 2979 | 'IGHG3_No9' | 'TPEVTCVVVDVSHEDPEVQFKWYVDGVEVHNAK' | 'Immunoglobulin heavy constant gamma 3' |
| 2980 | 'IGHG3_No10' | 'SRWQQGNIFSCSVMHEALHNR' | 'Immunoglobulin heavy constant gamma 3' |
| 2981 | 'IGHG3_No11' | 'CPAPELLGGPSVFLFPPKPKDTLMISR' | 'Immunoglobulin heavy constant gamma 3' |
| 2982 | 'IGHG3_No12' | 'LTVDKSRWQQGNIFSCSVMHEALHNR' | 'Immunoglobulin heavy constant gamma 3' |
| 2983 | 'IGHG3_No13' | 'RVELKTPLGDTTHTCPR' | 'Immunoglobulin heavy constant gamma 3' |
| 2984 | 'IGHG3_No14' | 'WQQGNIFSCSVMHEALHNRFTQK' | 'Immunoglobulin heavy constant gamma 3' |
| 2985 | 'IGHG3_No15' | 'GPSVFPLAPCSRSTSGGTAALGCLVK' | 'Immunoglobulin heavy constant gamma 3' |
| 2986 | 'IGHG3_No16' | 'CPAPELLGGPSVFLFPPK' | 'Immunoglobulin heavy constant gamma 3' |
| 2987 | 'IGHG3_No17' | 'DTLMISRTPEVTCVVVDVSHEDPEVQFK' | 'Immunoglobulin heavy constant gamma 3' |
| 2988 | 'IGHG3_No18' | 'CPEPKSCDTPPPCPR' | 'Immunoglobulin heavy constant gamma 3' |
| 2989 | 'IGHG3_No19' | 'LTVDKSRWQQGNIFSCSVMHEALHNR' | 'Immunoglobulin heavy constant gamma 3' |
| 2990 | 'IGHG3_No20' | 'SRWQQGNIFSCSVMHEALHNR' | 'Immunoglobulin heavy constant gamma 3' |
| 2991 | 'IGHG3_No21' | 'DTLMISRTPEVTCVVVDVSHEDPEVQFK' | 'Immunoglobulin heavy constant gamma 3' |
| 2992 | 'IGHG3_No22' | 'TPLGDTTHTCPRCPEPK' | 'Immunoglobulin heavy constant gamma 3' |
| 2993 | 'IGHG3_No23' | 'WQQGNIFSCSVMHEALHNRFTQK' | 'Immunoglobulin heavy constant gamma 3' |

| | A | B | C |
|---|---|---|---|
| 1 | **Peptide Name** | **Peptide Sequence** | **Peptide Description** |
| 2994 | 'IGHG4_No1' | 'YGPPCPSCPAPEFLGGPSVFLFPPKPK' | 'Immunoglobulin heavy constant gamma 4' |
| 2995 | 'IGHG4_No2' | 'EPQVYTLPPSQEEMTK' | 'Immunoglobulin heavy constant gamma 4' |
| 2996 | 'IGHG4_No3' | 'WQEGNVFSCSVMHEALHNHYTQK' | 'Immunoglobulin heavy constant gamma 4' |
| 2997 | 'IGHG4_No4' | 'SRWQEGNVFSCSVMHEALHNHYTQK' | 'Immunoglobulin heavy constant gamma 4' |
| 2998 | 'IGHG4_No5' | 'WQEGNVFSCSVMHEALHNHYTQK' | 'Immunoglobulin heavy constant gamma 4' |
| 2999 | 'IGHG4_No6' | 'GLPSSIEK' | 'Immunoglobulin heavy constant gamma 4' |
| 3000 | 'IGHG4_No7' | 'TYTCNVDHKPSNTKVDK' | 'Immunoglobulin heavy constant gamma 4' |
| 3001 | 'IGHG4_No8' | 'GQPREPQVYTLPPSQEEMTK' | 'Immunoglobulin heavy constant gamma 4' |
| 3002 | 'IGHG4_No9' | 'TYTCNVDHKPSNTK' | 'Immunoglobulin heavy constant gamma 4' |
| 3003 | 'IGHG4_No10' | 'EPQVYTLPPSQEEMTKNQVSLTCLVK' | 'Immunoglobulin heavy constant gamma 4' |
| 3004 | 'IGHG4_No11' | 'VSNKGLPSSIEK' | 'Immunoglobulin heavy constant gamma 4' |
| 3005 | 'IGHG4_No12' | 'AKGQPREPQVYTLPPSQEEMTK' | 'Immunoglobulin heavy constant gamma 4' |
| 3006 | 'IGHG4_No13' | 'SRWQEGNVFSCSVMHEALHNHYTQK' | 'Immunoglobulin heavy constant gamma 4' |
| 3007 | 'IGHG4_No14' | 'LTVDKSRWQEGNVFSCSVMHEALHNHYTQK' | 'Immunoglobulin heavy constant gamma 4' |
| 3008 | 'IGHG4_No15' | 'EPQVYTLPPSQEEMTK' | 'Immunoglobulin heavy constant gamma 4' |
| 3009 | 'IGHG4_No16' | 'GLPSSIEKTISK' | 'Immunoglobulin heavy constant gamma 4' |
| 3010 | 'IGHG4_No17' | 'GQPREPQVYTLPPSQEEMTK' | 'Immunoglobulin heavy constant gamma 4' |
| 3011 | 'IGHG4_No18' | 'EPQVYTLPPSQEEMTKNQVSLTCLVK' | 'Immunoglobulin heavy constant gamma 4' |
| 3012 | 'IGHG4_No19' | 'TTPPVLDSDGSFFLYSR' | 'Immunoglobulin heavy constant gamma 4' |
| 3013 | 'IGHG4_No20' | 'GQPREPQVYTLPPSQEEMTKNQVSLTCLVK' | 'Immunoglobulin heavy constant gamma 4' |
| 3014 | 'IGHG4_No21' | 'AKGQPREPQVYTLPPSQEEMTK' | 'Immunoglobulin heavy constant gamma 4' |
| 3015 | 'IGHG4_No22' | 'LTVDKSRWQEGNVFSCSVMHEALHNHYTQK' | 'Immunoglobulin heavy constant gamma 4' |
| 3016 | 'IGHG4_No23' | 'GQPREPQVYTLPPSQEEMTKNQVSLTCLVK' | 'Immunoglobulin heavy constant gamma 4' |
| 3017 | 'IGHG4_No24' | 'TTPPVLDSDGSFFLYSRLTVDKSR' | 'Immunoglobulin heavy constant gamma 4' |
| 3018 | 'IGHG4_No25' | 'VSNKGLPSSIEKTISK' | 'Immunoglobulin heavy constant gamma 4' |
| 3019 | 'IGHM_No1' | 'YAATSQVLLPSKDVMQGTDEHVVCK' | 'Immunoglobulin heavy constant mu' |
| 3020 | 'IGHM_No2' | 'YAATSQVLLPSKDVMQGTDEHVVCK' | 'Immunoglobulin heavy constant mu' |
| 3021 | 'IGHM_No3' | 'DVMQGTDEHVVCK' | 'Immunoglobulin heavy constant mu' |
| 3022 | 'IGHM_No4' | 'DVMQGTDEHVVCK' | 'Immunoglobulin heavy constant mu' |
| 3023 | 'IGHM_No5' | 'EKNVPLPVIAELPPK' | 'Immunoglobulin heavy constant mu' |
| 3024 | 'IGHM_No6' | 'GQPLSPEK' | 'Immunoglobulin heavy constant mu' |
| 3025 | 'IGHM_No7' | 'YFAHSILTVSEEEWNTGETYTCVVAHEALPNRVTER' | 'Immunoglobulin heavy constant mu' |
| 3026 | 'IGHM_No8' | 'YFAHSILTVSEEEWNTGETYTCVVAHEALPNR' | 'Immunoglobulin heavy constant mu' |
| 3027 | 'IGHM_No9' | 'GQPLSPEKYVTSAPMPEPQAPGR' | 'Immunoglobulin heavy constant mu' |
| 3028 | 'IGHM_No10' | 'GGKYAATSQVLLPSKDVMQGTDEHVVCK' | 'Immunoglobulin heavy constant mu' |
| 3029 | 'IGHM_No11' | 'GGKYAATSQVLLPSKDVMQGTDEHVVCK' | 'Immunoglobulin heavy constant mu' |
| 3030 | 'IGHM_No12' | 'QIQVSWLR' | 'Immunoglobulin heavy constant mu' |
| 3031 | 'IGHM_No13' | 'YAATSQVLLPSKDVMQGTDEHVVCKVQHPNGNK' | 'Immunoglobulin heavy constant mu' |
| 3032 | 'IGHM_No14' | 'ESDWLGQSMFTCR' | 'Immunoglobulin heavy constant mu' |
| 3033 | 'IGHM_No15' | 'GQPLSPEKYVTSAPMPEPQAPGR' | 'Immunoglobulin heavy constant mu' |
| 3034 | 'IGHM_No16' | 'QIQVSWLR' | 'Immunoglobulin heavy constant mu' |
| 3035 | 'IGHM_No17' | 'LICQATGFSPRQIQVSWLR' | 'Immunoglobulin heavy constant mu' |
| 3036 | 'IGHM_No18' | 'ESDWLGQSMFTCR' | 'Immunoglobulin heavy constant mu' |
| 3037 | 'IGHM_No19' | 'VTSTLTIKESDWLGQSMFTCR' | 'Immunoglobulin heavy constant mu' |

| | A | B | C |
|---|---|---|---|
| 1 | **Peptide Name** | **Peptide Sequence** | **Peptide Description** |
| 3038 | 'IGHM_No20' | 'EKNVPLPVIAELPPKVSVFVPPR' | 'Immunoglobulin heavy constant mu' |
| 3039 | 'IGHM_No21' | 'DVMQGTDEHVVCKVQHPNGNKEK' | 'Immunoglobulin heavy constant mu' |
| 3040 | 'IGHM_No22' | 'YAATSQVLLPSKDVMQGTDEHVVCKVQHPNGNK' | 'Immunoglobulin heavy constant mu' |
| 3041 | 'IGHM_No23' | 'QIQVSWLREGKQVGSGVTTDQVQAEAK' | 'Immunoglobulin heavy constant mu' |
| 3042 | 'IGHM_No24' | 'DVMQGTDEHVVCKVQHPNGNKEK' | 'Immunoglobulin heavy constant mu' |
| 3043 | 'IGHM_No25' | 'QIQVSWLREGKQVGSGVTTDQVQAEAK' | 'Immunoglobulin heavy constant mu' |
| 3044 | 'IGHM_No26' | 'DVMQGTDEHVVCKVQHPNGNK' | 'Immunoglobulin heavy constant mu' |
| 3045 | 'IGHM_No27' | 'VTSTLTIKESDWLGQSMFTCR' | 'Immunoglobulin heavy constant mu' |
| 3046 | 'IGHA1_No1' | 'DASGVTFTWTPSSGK' | 'Immunoglobulin heavy constant alpha 1' |
| 3047 | 'IGHA1_No2' | 'TFTCTAAYPESK' | 'Immunoglobulin heavy constant alpha 1' |
| 3048 | 'IGHA1_No3' | 'QEPSQGTTTFAVTSILR' | 'Immunoglobulin heavy constant alpha 1' |
| 3049 | 'IGHA1_No4' | 'KGDTFSCMVGHEALPLAFTQK' | 'Immunoglobulin heavy constant alpha 1' |
| 3050 | 'IGHA1_No5' | 'VFPLSLCSTQPDGNVVIACLVQGFFPQEPLSVTWSESGQGVTAR' | 'Immunoglobulin heavy constant alpha 1' |
| 3051 | 'IGHA1_No6' | 'DLCGCYSVSSVLPGCAEPWNHGK' | 'Immunoglobulin heavy constant alpha 1' |
| 3052 | 'IGHA1_No7' | 'GDTFSCMVGHEALPLAFTQK' | 'Immunoglobulin heavy constant alpha 1' |
| 3053 | 'IGHA1_No8' | 'TFTCTAAYPESKTPLTATLSK' | 'Immunoglobulin heavy constant alpha 1' |
| 3054 | 'IGHA1_No9' | 'NFPPSQDASGDLYTTSSQLTLPATQCLAGK' | 'Immunoglobulin heavy constant alpha 1' |
| 3055 | 'IGHA1_No10' | 'GDTFSCMVGHEALPLAFTQK' | 'Immunoglobulin heavy constant alpha 1' |
| 3056 | 'IGHA1_No11' | 'LAGKPTHVNVSVVMAEVDGTCY' | 'Immunoglobulin heavy constant alpha 1' |
| 3057 | 'IGHA1_No12' | 'KGDTFSCMVGHEALPLAFTQK' | 'Immunoglobulin heavy constant alpha 1' |
| 3058 | 'IGHA1_No13' | 'VAAEDWKKGDTFSCMVGHEALPLAFTQK' | 'Immunoglobulin heavy constant alpha 1' |
| 3059 | 'IGHA1_No14' | 'TPLTATLSK' | 'Immunoglobulin heavy constant alpha 1' |
| 3060 | 'IGHA1_No15' | 'SAVQGPPERDLCGCYSVSSVLPGCAEPWNHGKTFTCTAAYPESK' | 'Immunoglobulin heavy constant alpha 1' |
| 3061 | 'IGHA1_No16' | 'VAAEDWKKGDTFSCMVGHEALPLAFTQK' | 'Immunoglobulin heavy constant alpha 1' |
| 3062 | 'IGHA1_No17' | 'YLTWASRQEPSQGTTTFAVTSILR' | 'Immunoglobulin heavy constant alpha 1' |
| 3063 | 'IGHA1_No18' | 'QEPSQGTTTFAVTSILR' | 'Immunoglobulin heavy constant alpha 1' |
| 3064 | 'IGHA1_No19' | 'SAVQGPPERDLCGCYSVSSVLPGCAEPWNHGK' | 'Immunoglobulin heavy constant alpha 1' |
| 3065 | 'IGHA1_No20' | 'LAGKPTHVNVSVVMAEVDGTCY' | 'Immunoglobulin heavy constant alpha 1' |
| 3066 | 'IGHA1_No21' | 'QEPSQGTTTFAVTSILRVAAEDWKK' | 'Immunoglobulin heavy constant alpha 1' |
| 3067 | 'IGHA1_No22' | 'EKYLTWASRQEPSQGTTTFAVTSILR' | 'Immunoglobulin heavy constant alpha 1' |
| 3068 | 'IGHA1_No23' | 'TIDRLAGKPTHVNVSVVMAEVDGTCY' | 'Immunoglobulin heavy constant alpha 1' |
| 3069 | 'IGHA1_No24' | 'PTHVNVSVVMAEVDGTCY' | 'Immunoglobulin heavy constant alpha 1' |
| 3070 | 'IGHA1_No25' | 'TIDRLAGKPTHVNVSVVMAEVDGTCY' | 'Immunoglobulin heavy constant alpha 1' |
| 3071 | 'IGHA1_No26' | 'KGDTFSCMVGHEALPLAFTQKTIDR' | 'Immunoglobulin heavy constant alpha 1' |
| 3072 | 'IGHA1_No27' | 'GDTFSCMVGHEALPLAFTQKTIDR' | 'Immunoglobulin heavy constant alpha 1' |
| 3073 | 'IGHA1_No28' | 'QEPSQGTTTFAVTSILRVAAEDWK' | 'Immunoglobulin heavy constant alpha 1' |
| 3074 | 'IGHA2_No1' | 'WLQGSQELPR' | 'Immunoglobulin heavy constant alpha 2' |
| 3075 | 'IGHA2_No2' | 'DASGATFTWTPSSGK' | 'Immunoglobulin heavy constant alpha 2' |
| 3076 | 'IGHA2_No3' | 'SAVQGPPER' | 'Immunoglobulin heavy constant alpha 2' |
| 3077 | 'IGHA2_No4' | 'VAAEDWK' | 'Immunoglobulin heavy constant alpha 2' |
| 3078 | 'IGHA2_No5' | 'WLQGSQELPREK' | 'Immunoglobulin heavy constant alpha 2' |
| 3079 | 'IGHA2_No6' | 'GFSPKDVLVR' | 'Immunoglobulin heavy constant alpha 2' |
| 3080 | 'IGHA2_No7' | 'SAVQGPPERDLCGCYSVSSVLPGCAQPWNHGETFTCTAAHPELK' | 'Immunoglobulin heavy constant alpha 2' |
| 3081 | 'IGHA2_No8' | 'GETFSCMVGHEALPLAFTQK' | 'Immunoglobulin heavy constant alpha 2' |

| | A | B | C |
|---|---|---|---|
| 1 | **Peptide Name** | **Peptide Sequence** | **Peptide Description** |
| 3082 | 'IGHA2_No9' | 'SGNTFRPEVHLLPPPSEELALNELVTLTCLAR' | 'Immunoglobulin heavy constant alpha 2' |
| 3083 | 'IGHA2_No10' | 'KGETFSCMVGHEALPLAFTQK' | 'Immunoglobulin heavy constant alpha 2' |
| 3084 | 'IGHA2_No11' | 'DLCGCYSVSSVLPGCAQPWNHGETFTCTAAHPELK' | 'Immunoglobulin heavy constant alpha 2' |
| 3085 | 'IGHA2_No12' | 'YLTWASR' | 'Immunoglobulin heavy constant alpha 2' |
| 3086 | 'IGHA2_No13' | 'EKYLTWASR' | 'Immunoglobulin heavy constant alpha 2' |
| 3087 | 'IGHA2_No14' | 'NFPPSQDASGDLYTTSSQLTLPATQCPDGK' | 'Immunoglobulin heavy constant alpha 2' |
| 3088 | 'IGHA2_No15' | 'VAAEDWKK' | 'Immunoglobulin heavy constant alpha 2' |
| 3089 | 'IGHA2_No16' | 'PEVHLLPPPSEELALNELVTLTCLAR' | 'Immunoglobulin heavy constant alpha 2' |
| 3090 | 'IGHA2_No17' | 'GETFSCMVGHEALPLAFTQK' | 'Immunoglobulin heavy constant alpha 2' |
| 3091 | 'IGHA2_No18' | 'QEPSQGTTTYAVTSILR' | 'Immunoglobulin heavy constant alpha 2' |
| 3092 | 'IGHA2_No19' | 'KGETFSCMVGHEALPLAFTQK' | 'Immunoglobulin heavy constant alpha 2' |
| 3093 | 'IGHA2_No20' | 'VPPPPPCCHPR' | 'Immunoglobulin heavy constant alpha 2' |
| 3094 | 'IGHA2_No21' | 'MAGKPTHINVSVVMAEADGTCY' | 'Immunoglobulin heavy constant alpha 2' |
| 3095 | 'IGHA2_No22' | 'SVTCHVK' | 'Immunoglobulin heavy constant alpha 2' |
| 3096 | 'IGHA2_No23' | 'QEPSQGTTTYAVTSILR' | 'Immunoglobulin heavy constant alpha 2' |
| 3097 | 'IGHA2_No24' | 'VAAEDWKKGETFSCMVGHEALPLAFTQK' | 'Immunoglobulin heavy constant alpha 2' |
| 3098 | 'IGHA2_No25' | 'HYTNSSQDVTVPCR' | 'Immunoglobulin heavy constant alpha 2' |
| 3099 | 'IGHA2_No26' | 'MAGKPTHINVSVVMAEADGTCY' | 'Immunoglobulin heavy constant alpha 2' |
| 3100 | 'IGHA2_No27' | 'GFSPKDVLVRWLQGSQELPR' | 'Immunoglobulin heavy constant alpha 2' |
| 3101 | 'IGHA2_No28' | 'MAGKPTHINVSVVMAEADGTCY' | 'Immunoglobulin heavy constant alpha 2' |
| 3102 | 'IGHA2_No29' | 'VAAEDWKKGETFSCMVGHEALPLAFTQK' | 'Immunoglobulin heavy constant alpha 2' |
| 3103 | 'IGHA2_No30' | 'NFPPSQDASGDLYTTSSQLTLPATQCPDGKSVTCHVK' | 'Immunoglobulin heavy constant alpha 2' |
| 3104 | 'IGHD_No1' | 'STTFWAWSVLR' | 'Immunoglobulin heavy constant delta' |
| 3105 | 'IGHD_No2' | 'SLEVSYVTDHGPMK' | 'Immunoglobulin heavy constant delta' |
| 3106 | 'IGHD_No3' | 'SLEVSYVTDHGPMK' | 'Immunoglobulin heavy constant delta' |
| 3107 | 'HLA-B_No1' | 'DGEDQTQDTELVETRPAGDR' | 'HLA class I histocompatibility antigen, B alpha chain' |
| 3108 | 'HLA-B_No2' | 'ADPPKTHVTHHPISDHEATLR' | 'HLA class I histocompatibility antigen, B alpha chain' |
| 3109 | 'HLA-B_No3' | 'AYLEGECVEWLR' | 'HLA class I histocompatibility antigen, B alpha chain' |
| 3110 | 'HLA-B_No4' | 'APWIEQEGPEYWDR' | 'HLA class I histocompatibility antigen, B alpha chain' |
| 3111 | 'HLA-B_No5' | 'SWTAADTAAQITQR' | 'HLA class I histocompatibility antigen, B alpha chain' |
| 3112 | 'HLA-B_No6' | 'DGEDQTQDTELVETRPAGDRTFQK' | 'HLA class I histocompatibility antigen, B alpha chain' |
| 3113 | 'HLA-H' | 'YTCHVQHEGLPEPLTLR' | 'Putative HLA class I histocompatibility antigen, alpha chain H' |
| 3114 | 'HBD_No1' | 'LLGNVLVCVLAR' | 'Hemoglobin subunit delta' |
| 3115 | 'HBD_No2' | 'TAVNALWGK' | 'Hemoglobin subunit delta' |
| 3116 | 'HBD_No3' | 'GTFSQLSELHCDKLHVDPENFR' | 'Hemoglobin subunit delta' |
| 3117 | 'HBD_No4' | 'EFTPQMQAAYQK' | 'Hemoglobin subunit delta' |
| 3118 | 'HBD_No5' | 'GTFSQLSELHCDK' | 'Hemoglobin subunit delta' |
| 3119 | 'HBD_No6' | 'EFTPQMQAAYQK' | 'Hemoglobin subunit delta' |
| 3120 | 'HBD_No7' | 'VLGAFSDGLAHLDNLKGTFSQLSELHCDKLHVDPENFR' | 'Hemoglobin subunit delta' |
| 3121 | 'HBD_No8' | 'FFESFGDLSSPDAVMGNPK' | 'Hemoglobin subunit delta' |
| 3122 | 'HBD_No9' | 'VNVDAVGGEALGR' | 'Hemoglobin subunit delta' |
| 3123 | 'HBD_No10' | 'FFESFGDLSSPDAVMGNPK' | 'Hemoglobin subunit delta' |
| 3124 | 'HBD_No11' | 'NFGKEFTPQMQAAYQK' | 'Hemoglobin subunit delta' |
| 3125 | 'HBD_No12' | 'VLGAFSDGLAHLDNLKGTFSQLSELHCDK' | 'Hemoglobin subunit delta' |

| | A | B | C |
|---|---|---|---|
| 1 | **Peptide Name** | **Peptide Sequence** | **Peptide Description** |
| 3126 | 'HBD_No13' | 'TAVNALWGKVNVDAVGGEALGR' | 'Hemoglobin subunit delta' |
| 3127 | 'HBD_No14' | 'VHLTPEEKTAVNALWGK' | 'Hemoglobin subunit delta' |
| 3128 | 'HBD_No15' | 'NFGKEFTPQMQAAYQK' | 'Hemoglobin subunit delta' |
| 3129 | 'HBE1' | 'LSELHCDK' | 'Hemoglobin subunit epsilon' |
| 3130 | 'MB_No1' | 'HGATVLTALGGILK' | 'Myoglobin' |
| 3131 | 'MB_No2' | 'VEADIPGHGQEVLIR' | 'Myoglobin' |
| 3132 | 'COL1A1_No1' | 'NSVAYMDQQTGNLKK' | 'Collagen alpha-1(I) chain' |
| 3133 | 'COL1A1_No2' | 'SGEYWIDPNQGCNLDAIK' | 'Collagen alpha-1(I) chain' |
| 3134 | 'KRT14_No1' | 'LASYLDKVR' | 'Keratin, type I cytoskeletal 14' |
| 3135 | 'KRT14_No2' | 'APSTYGGGLSVSSSR' | 'Keratin, type I cytoskeletal 14' |
| 3136 | 'KRT14_No3' | 'VTMQNLNDR' | 'Keratin, type I cytoskeletal 14' |
| 3137 | 'KRT14_No4' | 'EVATNSELVQSGK' | 'Keratin, type I cytoskeletal 14' |
| 3138 | 'KRT14_No5' | 'MSVEADINGLRR' | 'Keratin, type I cytoskeletal 14' |
| 3139 | 'KRT14_No6' | 'DAEEWFFTK' | 'Keratin, type I cytoskeletal 14' |
| 3140 | 'KRT14_No7' | 'ALEEANADLEVK' | 'Keratin, type I cytoskeletal 14' |
| 3141 | 'KRT14_No8' | 'VTMQNLNDRLASYLDKVR' | 'Keratin, type I cytoskeletal 14' |
| 3142 | 'KRT14_No9' | 'LEQEIATYR' | 'Keratin, type I cytoskeletal 14' |
| 3143 | 'KRT14_No10' | 'ILTATVDNANVLLQIDNAR' | 'Keratin, type I cytoskeletal 14' |
| 3144 | 'KRT14_No11' | 'VTMQNLNDR' | 'Keratin, type I cytoskeletal 14' |
| 3145 | 'KRT14_No12' | 'VLDELTLAR' | 'Keratin, type I cytoskeletal 14' |
| 3146 | 'KRT14_No13' | 'VTMQNLNDRLASYLDKVR' | 'Keratin, type I cytoskeletal 14' |
| 3147 | 'KRT14_No14' | 'ASLENSLEETKGR' | 'Keratin, type I cytoskeletal 14' |
| 3148 | 'KRT14_No15' | 'ILNEMRDQYEK' | 'Keratin, type I cytoskeletal 14' |
| 3149 | 'KRT14_No16' | 'ADLEMQIESLKEELAYLKK' | 'Keratin, type I cytoskeletal 14' |
| 3150 | 'KRT14_No17' | 'DAEEWFFTKTEELNR' | 'Keratin, type I cytoskeletal 14' |
| 3151 | 'KRT14_No18' | 'ASLENSLEETK' | 'Keratin, type I cytoskeletal 14' |
| 3152 | 'KRT14_No19' | 'MSVEADINGLRR' | 'Keratin, type I cytoskeletal 14' |
| 3153 | 'KRT14_No20' | 'GQVGGDVNVEMDAAPGVDLSR' | 'Keratin, type I cytoskeletal 14' |
| 3154 | 'KRT14_No21' | 'ADLEMQIESLKEELAYLKK' | 'Keratin, type I cytoskeletal 14' |
| 3155 | 'KRT14_No22' | 'GQVGGDVNVEMDAAPGVDLSR' | 'Keratin, type I cytoskeletal 14' |
| 3156 | 'KRT14_No23' | 'QRPAEIKDYSPYFK' | 'Keratin, type I cytoskeletal 14' |
| 3157 | 'KRT14_No24' | 'LLEGEDAHLSSSQFSSGSQSSR' | 'Keratin, type I cytoskeletal 14' |
| 3158 | 'KRT14_No25' | 'RVLDELTLAR' | 'Keratin, type I cytoskeletal 14' |
| 3159 | 'KRT14_No26' | 'TKYETELNLR' | 'Keratin, type I cytoskeletal 14' |
| 3160 | 'KRT14_No27' | 'NHEEEMNALR' | 'Keratin, type I cytoskeletal 14' |
| 3161 | 'KRT14_No28' | 'ILNEMRDQYEK' | 'Keratin, type I cytoskeletal 14' |
| 3162 | 'KRT14_No29' | 'TEELNREVATNSELVQSGK' | 'Keratin, type I cytoskeletal 14' |
| 3163 | 'KRT14_No30' | 'VTMQNLNDRLASYLDK' | 'Keratin, type I cytoskeletal 14' |
| 3164 | 'KRT14_No31' | 'MSVEADINGLR' | 'Keratin, type I cytoskeletal 14' |
| 3165 | 'KRT14_No32' | 'TRLEQEIATYRR' | 'Keratin, type I cytoskeletal 14' |
| 3166 | 'KRT14_No33' | 'TMQNLEIELQSQLSMK' | 'Keratin, type I cytoskeletal 14' |
| 3167 | 'KRT14_No34' | 'TRLEQEIATYR' | 'Keratin, type I cytoskeletal 14' |
| 3168 | 'KRT14_No35' | 'LASYLDK' | 'Keratin, type I cytoskeletal 14' |
| 3169 | 'KRT14_No36' | 'QRPAEIKDYSPYFK' | 'Keratin, type I cytoskeletal 14' |

| | A | B | C |
|---|---|---|---|
| 1 | **Peptide Name** | **Peptide Sequence** | **Peptide Description** |
| 3170 | 'KRT14_No37' | 'ADLEMQIESLKEELAYLK' | 'Keratin, type I cytoskeletal 14' |
| 3171 | 'KRT14_No38' | 'EVATNSELVQSGKSEISELRR' | 'Keratin, type I cytoskeletal 14' |
| 3172 | 'KRT14_No39' | 'KDAEEWFFTKTEELNR' | 'Keratin, type I cytoskeletal 14' |
| 3173 | 'KRT14_No40' | 'LEQEIATYRR' | 'Keratin, type I cytoskeletal 14' |
| 3174 | 'KRT14_No41' | 'TMQNLEIELQSQLSMK' | 'Keratin, type I cytoskeletal 14' |
| 3175 | 'KRT14_No42' | 'NHEEEMNALR' | 'Keratin, type I cytoskeletal 14' |
| 3176 | 'KRT6A_No1' | 'NLDLDSIIAEVK' | 'Keratin, type II cytoskeletal 6A' |
| 3177 | 'KRT6A_No2' | 'FASFIDKVR' | 'Keratin, type II cytoskeletal 6A' |
| 3178 | 'KRT6A_No3' | 'LALDVEIATYR' | 'Keratin, type II cytoskeletal 6A' |
| 3179 | 'KRT6A_No4' | 'NKYEDEINKR' | 'Keratin, type II cytoskeletal 6A' |
| 3180 | 'KRT6A_No5' | 'AQYEEIAQR' | 'Keratin, type II cytoskeletal 6A' |
| 3181 | 'KRT6A_No6' | 'ADTLTDEINFLR' | 'Keratin, type II cytoskeletal 6A' |
| 3182 | 'KRT6A_No7' | 'FASFIDK' | 'Keratin, type II cytoskeletal 6A' |
| 3183 | 'KRT6A_No8' | 'TAAENEFVTLKK' | 'Keratin, type II cytoskeletal 6A' |
| 3184 | 'KRT6A_No9' | 'LALDVEIATYRK' | 'Keratin, type II cytoskeletal 6A' |
| 3185 | 'KRT6A_No10' | 'QLDSIVGER' | 'Keratin, type II cytoskeletal 6A' |
| 3186 | 'KRT6A_No11' | 'WTLLQEQGTK' | 'Keratin, type II cytoskeletal 6A' |
| 3187 | 'KRT6A_No12' | 'YEELQVTAGR' | 'Keratin, type II cytoskeletal 6A' |
| 3188 | 'KRT6A_No13' | 'QNLEPLFEQYINNLRR' | 'Keratin, type II cytoskeletal 6A' |
| 3189 | 'KRT6A_No14' | 'AIGGGLSSVGGGSSTIK' | 'Keratin, type II cytoskeletal 6A' |
| 3190 | 'KRT6A_No15' | 'ALYDAELSQMQTHISDTSVVLSMDNNR' | 'Keratin, type II cytoskeletal 6A' |
| 3191 | 'KRT6A_No16' | 'DVDAAYMNKVELQAK' | 'Keratin, type II cytoskeletal 6A' |
| 3192 | 'KRT6A_No17' | 'LLKEYQELMNVK' | 'Keratin, type II cytoskeletal 6A' |
| 3193 | 'KRT6A_No18' | 'GSGGLGGACGGAGFGSR' | 'Keratin, type II cytoskeletal 6A' |
| 3194 | 'KRT6A_No19' | 'TLNNKFASFIDKVR' | 'Keratin, type II cytoskeletal 6A' |
| 3195 | 'KRT6A_No20' | 'LLEGEECR' | 'Keratin, type II cytoskeletal 6A' |
| 3196 | 'KRT6A_No21' | 'ISIGGGSCAISGGYGSR' | 'Keratin, type II cytoskeletal 6A' |
| 3197 | 'KRT6A_No22' | 'QLDSIVGER' | 'Keratin, type II cytoskeletal 6A' |
| 3198 | 'KRT6A_No23' | 'SGFSSVSVSR' | 'Keratin, type II cytoskeletal 6A' |
| 3199 | 'KRT6A_No24' | 'SRAEAESWYQTK' | 'Keratin, type II cytoskeletal 6A' |
| 3200 | 'KRT6A_No25' | 'NKLEGLEDALQK' | 'Keratin, type II cytoskeletal 6A' |
| 3201 | 'KRT6A_No26' | 'TLNNKFASFIDK' | 'Keratin, type II cytoskeletal 6A' |
| 3202 | 'KRT6A_No27' | 'DVDAAYMNKVELQAK' | 'Keratin, type II cytoskeletal 6A' |
| 3203 | 'KRT6A_No28' | 'YEDEINKR' | 'Keratin, type II cytoskeletal 6A' |
| 3204 | 'KRT6A_No29' | 'SRAEAESWYQTKYEELQVTAGR' | 'Keratin, type II cytoskeletal 6A' |
| 3205 | 'KRT6A_No30' | 'QNLEPLFEQYINNLR' | 'Keratin, type II cytoskeletal 6A' |
| 3206 | 'KRT6A_No31' | 'FLEQQNKVLETK' | 'Keratin, type II cytoskeletal 6A' |
| 3207 | 'KRT6A_No32' | 'TAAENEFVTLK' | 'Keratin, type II cytoskeletal 6A' |
| 3208 | 'KRT6A_No33' | 'RQLDSIVGER' | 'Keratin, type II cytoskeletal 6A' |
| 3209 | 'KRT6A_No34' | 'LLKEYQELMNVK' | 'Keratin, type II cytoskeletal 6A' |
| 3210 | 'KRT6A_No35' | 'AIGGGLSSVGGGSSTIKYTTTSSSSR' | 'Keratin, type II cytoskeletal 6A' |
| 3211 | 'KRT6A_No36' | 'AEAESWYQTKYEELQVTAGR' | 'Keratin, type II cytoskeletal 6A' |
| 3212 | 'KRT6A_No37' | 'LEGLEDALQK' | 'Keratin, type II cytoskeletal 6A' |
| 3213 | 'KRT6A_No38' | 'QCANLQAAIADAEQR' | 'Keratin, type II cytoskeletal 6A' |

| | A | B | C |
|---|---|---|---|
| 1 | **Peptide Name** | **Peptide Sequence** | **Peptide Description** |
| 3214 | 'SPTA1_No1' | 'QDTLDASLQSFQQER' | 'Spectrin alpha chain, erythrocytic 1' |
| 3215 | 'SPTA1_No2' | 'VLETAEEIQER' | 'Spectrin alpha chain, erythrocytic 1' |
| 3216 | 'SPTA1_No3' | 'MVEEGHFAAEDVASR' | 'Spectrin alpha chain, erythrocytic 1' |
| 3217 | 'SPTA1_No4' | 'VILENIASHEPR' | 'Spectrin alpha chain, erythrocytic 1' |
| 3218 | 'SPTA1_No5' | 'AYFLDGSLLK' | 'Spectrin alpha chain, erythrocytic 1' |
| 3219 | 'SPTA1_No6' | 'FYLFLSK' | 'Spectrin alpha chain, erythrocytic 1' |
| 3220 | 'SPTA1_No7' | 'LSESHPDATEDLQR' | 'Spectrin alpha chain, erythrocytic 1' |
| 3221 | 'SPTA1_No8' | 'HEALENDFAVHETR' | 'Spectrin alpha chain, erythrocytic 1' |
| 3222 | 'SPTA1_No9' | 'LYEDSDDLKNWINK' | 'Spectrin alpha chain, erythrocytic 1' |
| 3223 | 'SPTA1_No10' | 'HLSDIIEEREQELQKEEAR' | 'Spectrin alpha chain, erythrocytic 1' |
| 3224 | 'SPTA1_No11' | 'CQALSAADPGSDLFSVQALQR' | 'Spectrin alpha chain, erythrocytic 1' |
| 3225 | 'SPTA1_No12' | 'VQNVCAQGEDILNK' | 'Spectrin alpha chain, erythrocytic 1' |
| 3226 | 'SPTA1_No13' | 'KGDVLTLLSSINK' | 'Spectrin alpha chain, erythrocytic 1' |
| 3227 | 'SPTA1_No14' | 'FEALKEPLATR' | 'Spectrin alpha chain, erythrocytic 1' |
| 3228 | 'SPTA1_No15' | 'FSSDFDELSGWMNEK' | 'Spectrin alpha chain, erythrocytic 1' |
| 3229 | 'SPTA1_No16' | 'LEDSYHLQVFK' | 'Spectrin alpha chain, erythrocytic 1' |
| 3230 | 'SPTA1_No17' | 'FLNVQELAAAHHEK' | 'Spectrin alpha chain, erythrocytic 1' |
| 3231 | 'SPTA1_No18' | 'SSDEIENAFQALAEGK' | 'Spectrin alpha chain, erythrocytic 1' |
| 3232 | 'SPTA1_No19' | 'LTLSHPSDAPQIQEMKEDLVSSWEHIR' | 'Spectrin alpha chain, erythrocytic 1' |
| 3233 | 'APOA1_No1' | 'VKDLATVYVDVLK' | 'Apolipoprotein A-I' |
| 3234 | 'APOA1_No2' | 'VSFLSALEEYTK' | 'Apolipoprotein A-I' |
| 3235 | 'APOA1_No3' | 'QGLLPVLESFK' | 'Apolipoprotein A-I' |
| 3236 | 'APOA1_No4' | 'WQEEMELYR' | 'Apolipoprotein A-I' |
| 3237 | 'APOA1_No5' | 'LREQLGPVTQEFWDNLEK' | 'Apolipoprotein A-I' |
| 3238 | 'APOA1_No6' | 'DLATVYVDVLK' | 'Apolipoprotein A-I' |
| 3239 | 'APOA1_No7' | 'QGLLPVLESFK' | 'Apolipoprotein A-I' |
| 3240 | 'APOA1_No8' | 'AKPALEDLRQGLLPVLESFK' | 'Apolipoprotein A-I' |
| 3241 | 'APOA1_No9' | 'QKVEPLRAELQEGAR' | 'Apolipoprotein A-I' |
| 3242 | 'APOA1_No10' | 'DYVSQFEGSALGK' | 'Apolipoprotein A-I' |
| 3243 | 'APOA1_No11' | 'VSFLSALEEYTKK' | 'Apolipoprotein A-I' |
| 3244 | 'APOA1_No12' | 'VEPLRAELQEGAR' | 'Apolipoprotein A-I' |
| 3245 | 'APOA1_No13' | 'KWQEEMELYR' | 'Apolipoprotein A-I' |
| 3246 | 'APOA1_No14' | 'VQPYLDDFQKK' | 'Apolipoprotein A-I' |
| 3247 | 'APOA1_No15' | 'THLAPYSDELRQR' | 'Apolipoprotein A-I' |
| 3248 | 'APOA1_No16' | 'AKPALEDLR' | 'Apolipoprotein A-I' |
| 3249 | 'APOA1_No17' | 'LSPLGEEMRDR' | 'Apolipoprotein A-I' |
| 3250 | 'APOA1_No18' | 'LSPLGEEMRDR' | 'Apolipoprotein A-I' |
| 3251 | 'APOA1_No19' | 'DSGRDYVSQFEGSALGK' | 'Apolipoprotein A-I' |
| 3252 | 'APOA1_No20' | 'ATEHLSTLSEK' | 'Apolipoprotein A-I' |
| 3253 | 'APOA1_No21' | 'LREQLGPVTQEFWDNLEKETEGLR' | 'Apolipoprotein A-I' |
| 3254 | 'APOA1_No22' | 'KWQEEMELYR' | 'Apolipoprotein A-I' |
| 3255 | 'APOA1_No23' | 'LEALKENGGAR' | 'Apolipoprotein A-I' |
| 3256 | 'APOA1_No24' | 'QKVEPLR' | 'Apolipoprotein A-I' |
| 3257 | 'APOA1_No25' | 'VKDLATVYVDVLKDSGR' | 'Apolipoprotein A-I' |

| | A | B | C |
|---|---|---|---|
| 1 | **Peptide Name** | **Peptide Sequence** | **Peptide Description** |
| 3258 | 'APOA1_No26' | 'THLAPYSDELR' | 'Apolipoprotein A-I' |
| 3259 | 'APOA1_No27' | 'AELQEGAR' | 'Apolipoprotein A-I' |
| 3260 | 'APOA1_No28' | 'PALEDLR' | 'Apolipoprotein A-I' |
| 3261 | 'APOA1_No29' | 'QEMSKDLEEVK' | 'Apolipoprotein A-I' |
| 3262 | 'APOA1_No30' | 'AKVQPYLDDFQKK' | 'Apolipoprotein A-I' |
| 3263 | 'APOA1_No31' | 'DLATVYVDVLKDSGR' | 'Apolipoprotein A-I' |
| 3264 | 'APOA1_No32' | 'LHELQEK' | 'Apolipoprotein A-I' |
| 3265 | 'APOA1_No33' | 'VQPYLDDFQK' | 'Apolipoprotein A-I' |
| 3266 | 'APOA1_No34' | 'QKVEPLR' | 'Apolipoprotein A-I' |
| 3267 | 'APOA1_No35' | 'EQLGPVTQEFWDNLEKETEGLR' | 'Apolipoprotein A-I' |
| 3268 | 'APOA1_No36' | 'VQPYLDDFQKKWQEEMELYR' | 'Apolipoprotein A-I' |
| 3269 | 'APOA1_No37' | 'HFWQQDEPPQSPWDR' | 'Apolipoprotein A-I' |
| 3270 | 'APOA1_No38' | 'LAEYHAK' | 'Apolipoprotein A-I' |
| 3271 | 'APOA1_No39' | 'LLDNWDSVTSTFSK' | 'Apolipoprotein A-I' |
| 3272 | 'APOA1_No40' | 'ETEGLRQEMSK' | 'Apolipoprotein A-I' |
| 3273 | 'APOA1_No41' | 'VQPYLDDFQKKWQEEMELYR' | 'Apolipoprotein A-I' |
| 3274 | 'APOA1_No42' | 'DLATVYVDVLKDSGRDYVSQFEGSALGK' | 'Apolipoprotein A-I' |
| 3275 | 'APOA1_No43' | 'AKVQPYLDDFQK' | 'Apolipoprotein A-I' |
| 3276 | 'APOA1_No44' | 'EQLGPVTQEFWDNLEK' | 'Apolipoprotein A-I' |
| 3277 | 'APOA1_No45' | 'WQEEMELYRQK' | 'Apolipoprotein A-I' |
| 3278 | 'APOA1_No46' | 'LAARLEALKENGGAR' | 'Apolipoprotein A-I' |
| 3279 | 'APOA1_No47' | 'AHVDALRTHLAPYSDELRQR' | 'Apolipoprotein A-I' |
| 3280 | 'APOA1_No48' | 'ETEGLRQEMSKDLEEVK' | 'Apolipoprotein A-I' |
| 3281 | 'APOA1_No49' | 'QEMSKDLEEVK' | 'Apolipoprotein A-I' |
| 3282 | 'APOA1_No50' | 'QKLHELQEK' | 'Apolipoprotein A-I' |
| 3283 | 'APOA1_No51' | 'LAEYHAKATEHLSTLSEK' | 'Apolipoprotein A-I' |
| 3284 | 'APOA1_No52' | 'QKLHELQEK' | 'Apolipoprotein A-I' |
| 3285 | 'APOA1_No53' | 'EQLGPVTQEFWDNLEKETEGLRQEMSK' | 'Apolipoprotein A-I' |
| 3286 | 'APOA1_No54' | 'QLNLKLLDNWDSVTSTFSK' | 'Apolipoprotein A-I' |
| 3287 | 'APOA1_No55' | 'QEMSKDLEEVK' | 'Apolipoprotein A-I' |
| 3288 | 'APOA1_No56' | 'ETEGLRQEMSKDLEEVK' | 'Apolipoprotein A-I' |
| 3289 | 'APOA1_No57' | 'QEMSKDLEEVKAK' | 'Apolipoprotein A-I' |
| 3290 | 'APOA1_No58' | 'EQLGPVTQEFWDNLEKETEGLRQEMSK' | 'Apolipoprotein A-I' |
| 3291 | 'APOA1_No59' | 'AHVDALRTHLAPYSDELR' | 'Apolipoprotein A-I' |
| 3292 | 'APOA1_No60' | 'QKLHELQEKLSPLGEEMR' | 'Apolipoprotein A-I' |
| 3293 | 'APOA1_No61' | 'QKVEPLRAELQEGAR' | 'Apolipoprotein A-I' |
| 3294 | 'APOA1_No62' | 'LHELQEKLSPLGEEMRDR' | 'Apolipoprotein A-I' |
| 3295 | 'APOA1_No63' | 'VSFLSALEEYTKKLNTQ' | 'Apolipoprotein A-I' |
| 3296 | 'APOA1_No64' | 'LSPLGEEMR' | 'Apolipoprotein A-I' |
| 3297 | 'APOA1_No65' | 'QEMSKDLEEVKAK' | 'Apolipoprotein A-I' |
| 3298 | 'APOA1_No66' | 'LSPLGEEMR' | 'Apolipoprotein A-I' |
| 3299 | 'APOA1_No67' | 'ATEHLSTLSEKAKPALEDLR' | 'Apolipoprotein A-I' |
| 3300 | 'APOA1_No68' | 'THLAPYSDELRQRLAAR' | 'Apolipoprotein A-I' |
| 3301 | 'APOA1_No69' | 'KWQEEMELYRQK' | 'Apolipoprotein A-I' |

| | A | B | C |
|---|---|---|---|
| 1 | **Peptide Name** | **Peptide Sequence** | **Peptide Description** |
| 3302 | 'APOA1_No70' | 'WQEEMELYR' | 'Apolipoprotein A-I' |
| 3303 | 'APOA1_No71' | 'QEMSKDLEEVK' | 'Apolipoprotein A-I' |
| 3304 | 'APOA1_No72' | 'QEMSKDLEEVKAK' | 'Apolipoprotein A-I' |
| 3305 | 'APOA1_No73' | 'DSGRDYVSQFEGSALGKQLNLK' | 'Apolipoprotein A-I' |
| 3306 | 'APOA1_No74' | 'LHELQEKLSPLGEEMR' | 'Apolipoprotein A-I' |
| 3307 | 'APOA1_No75' | 'QKLHELQEKLSPLGEEMR' | 'Apolipoprotein A-I' |
| 3308 | 'APOA1_No76' | 'QKLHELQEKLSPLGEEMR' | 'Apolipoprotein A-I' |
| 3309 | 'APOA1_No77' | 'LHELQEKLSPLGEEMRDR' | 'Apolipoprotein A-I' |
| 3310 | 'APOA1_No78' | 'ETEGLRQEMSK' | 'Apolipoprotein A-I' |
| 3311 | 'APOA1_No79' | 'QEMSKDLEEVKAK' | 'Apolipoprotein A-I' |
| 3312 | 'APOA1_No80' | 'LHELQEKLSPLGEEMR' | 'Apolipoprotein A-I' |
| 3313 | 'APOA1_No81' | 'LSPLGEEMRDRAR' | 'Apolipoprotein A-I' |
| 3314 | 'APOA1_No82' | 'WQEEMELYRQK' | 'Apolipoprotein A-I' |
| 3315 | 'APOA1_No83' | 'QKLHELQEKLSPLGEEMR' | 'Apolipoprotein A-I' |
| 3316 | 'APOA1_No84' | 'DYVSQFEGSALGKQLNLK' | 'Apolipoprotein A-I' |
| 3317 | 'APOA1_No85' | 'VEPLRAELQEGARQK' | 'Apolipoprotein A-I' |
| 3318 | 'APOA1_No86' | 'QLNLKLLDNWDSVTSTFSKLR' | 'Apolipoprotein A-I' |
| 3319 | 'APOA1_No87' | 'KWQEEMELYRQK' | 'Apolipoprotein A-I' |
| 3320 | 'APOA1_No88' | 'LLDNWDSVTSTFSKLR' | 'Apolipoprotein A-I' |
| 3321 | 'APOA1_No89' | 'LSPLGEEMRDRAR' | 'Apolipoprotein A-I' |
| 3322 | 'APOA1_No90' | 'LEALKENGGARLAEYHAK' | 'Apolipoprotein A-I' |
| 3323 | 'APOA1_No91' | 'PALEDLRQGLLPVLESFK' | 'Apolipoprotein A-I' |
| 3324 | 'APOA1_No92' | 'QLNLKLLDNWDSVTSTFSK' | 'Apolipoprotein A-I' |
| 3325 | 'APOA1_No93' | 'ENGGARLAEYHAKATEHLSTLSEK' | 'Apolipoprotein A-I' |
| 3326 | 'APOA1_No94' | 'ARAHVDALR' | 'Apolipoprotein A-I' |
| 3327 | 'APOA1_No95' | 'AHVDALR' | 'Apolipoprotein A-I' |
| 3328 | 'APOE_No1' | 'WVQTLSEQVQEELLSSQVTQELR' | 'Apolipoprotein E' |
| 3329 | 'APOE_No2' | 'AATVGSLAGQPLQER' | 'Apolipoprotein E' |
| 3330 | 'APOE_No3' | 'SELEEQLTPVAEETR' | 'Apolipoprotein E' |
| 3331 | 'APOE_No4' | 'WELALGR' | 'Apolipoprotein E' |
| 3332 | 'APOE_No5' | 'AKLEEQAQQIR' | 'Apolipoprotein E' |
| 3333 | 'APOE_No6' | 'GEVQAMLGQSTEELR' | 'Apolipoprotein E' |
| 3334 | 'APOE_No7' | 'LDEVKEQVAEVR' | 'Apolipoprotein E' |
| 3335 | 'APOE_No8' | 'VEQAVETEPEPELR' | 'Apolipoprotein E' |
| 3336 | 'APOE_No9' | 'LGADMEDVCGR' | 'Apolipoprotein E' |
| 3337 | 'APOE_No10' | 'LGADMEDVCGR' | 'Apolipoprotein E' |
| 3338 | 'APOE_No11' | 'LQAEAFQAR' | 'Apolipoprotein E' |
| 3339 | 'APOE_No12' | 'LAVYQAGAR' | 'Apolipoprotein E' |
| 3340 | 'APOE_No13' | 'ALMDETMKELK' | 'Apolipoprotein E' |
| 3341 | 'APOE_No14' | 'DRLDEVKEQVAEVR' | 'Apolipoprotein E' |
| 3342 | 'APOE_No15' | 'AYKSELEEQLTPVAEETR' | 'Apolipoprotein E' |
| 3343 | 'APOE_No16' | 'ALMDETMKELK' | 'Apolipoprotein E' |
| 3344 | 'APOE_No17' | 'SWFEPLVEDMQR' | 'Apolipoprotein E' |
| 3345 | 'APOE_No18' | 'LLRDADDLQK' | 'Apolipoprotein E' |

| | A | B | C |
|---|---|---|---|
| 1 | **Peptide Name** | **Peptide Sequence** | **Peptide Description** |
| 3346 | 'APOE_No19' | 'SWFEPLVEDMQR' | 'Apolipoprotein E' |
| 3347 | 'APOE_No20' | 'LKSWFEPLVEDMQR' | 'Apolipoprotein E' |
| 3348 | 'APOE_No21' | 'ALMDETMKELK' | 'Apolipoprotein E' |
| 3349 | 'APOE_No22' | 'LEEQAQQIR' | 'Apolipoprotein E' |
| 3350 | 'APOE_No23' | 'QQTEWQSGQR' | 'Apolipoprotein E' |
| 3351 | 'APOE_No24' | 'ERLGPLVEQGR' | 'Apolipoprotein E' |
| 3352 | 'APOE_No25' | 'LSKELQAAQAR' | 'Apolipoprotein E' |
| 3353 | 'APOE_No26' | 'GEVQAMLGQSTEELRVR' | 'Apolipoprotein E' |
| 3354 | 'APOE_No27' | 'LKSWFEPLVEDMQR' | 'Apolipoprotein E' |
| 3355 | 'APOE_No28' | 'GEVQAMLGQSTEELR' | 'Apolipoprotein E' |
| 3356 | 'APOE_No29' | 'VQAAVGTSAAPVPSDNH' | 'Apolipoprotein E' |
| 3357 | 'APOE_No30' | 'LGPLVEQGR' | 'Apolipoprotein E' |
| 3358 | 'APOE_No31' | 'LLRDADDLQKR' | 'Apolipoprotein E' |
| 3359 | 'APOE_No32' | 'ALMDETMKELK' | 'Apolipoprotein E' |
| 3360 | 'APOE_No33' | 'ELQAAQAR' | 'Apolipoprotein E' |
| 3361 | 'APOE_No34' | 'VRAATVGSLAGQPLQER' | 'Apolipoprotein E' |
| 3362 | 'APOE_No35' | 'QQTEWQSGQRWELALGR' | 'Apolipoprotein E' |
| 3363 | 'APOE_No36' | 'GEVQAMLGQSTEELRVR' | 'Apolipoprotein E' |
| 3364 | 'APOE_No37' | 'QQTEWQSGQR' | 'Apolipoprotein E' |
| 3365 | 'APOE_No38' | 'MEEMGSR' | 'Apolipoprotein E' |
| 3366 | 'APOE_No39' | 'AQAWGER' | 'Apolipoprotein E' |
| 3367 | 'APOE_No40' | 'LVQYRGEVQAMLGQSTEELRVR' | 'Apolipoprotein E' |
| 3368 | 'APOE_No41' | 'LGPLVEQGRVR' | 'Apolipoprotein E' |
| 3369 | 'APOE_No42' | 'EQVAEVR' | 'Apolipoprotein E' |
| 3370 | 'APOE_No43' | 'AKLEEQAQQIRLQAEAFQAR' | 'Apolipoprotein E' |
| 3371 | 'APOE_No44' | 'RLAVYQAGAR' | 'Apolipoprotein E' |
| 3372 | 'APOE_No45' | 'AYKSELEEQLTPVAEETRAR' | 'Apolipoprotein E' |
| 3373 | 'APOE_No46' | 'DADDLQK' | 'Apolipoprotein E' |
| 3374 | 'APOE_No47' | 'QWAGLVEKVQAAVGTSAAPVPSDNH' | 'Apolipoprotein E' |
| 3375 | 'APOE_No48' | 'DADDLQKR' | 'Apolipoprotein E' |
| 3376 | 'APOE_No49' | 'LVQYRGEVQAMLGQSTEELR' | 'Apolipoprotein E' |
| 3377 | 'APOE_No50' | 'GLSAIRER' | 'Apolipoprotein E' |
| 3378 | 'APOE_No51' | 'LVQYRGEVQAMLGQSTEELRVR' | 'Apolipoprotein E' |
| 3379 | 'APOE_No52' | 'QWAGLVEKVQAAVGTSAAPVPSDNH' | 'Apolipoprotein E' |
| 3380 | 'APOE_No53' | 'QQTEWQSGQRWELALGR' | 'Apolipoprotein E' |
| 3381 | 'APOA2_No1' | 'AGTELVNFLSYFVELGTQPATQ' | 'Apolipoprotein A-II' |
| 3382 | 'APOA2_No2' | 'EPCVESLVSQYFQTVTDYGKDLMEK' | 'Apolipoprotein A-II' |
| 3383 | 'APOA2_No3' | 'SKEQLTPLIK' | 'Apolipoprotein A-II' |
| 3384 | 'APOA2_No4' | 'EQLTPLIKK' | 'Apolipoprotein A-II' |
| 3385 | 'APOA2_No5' | 'EPCVESLVSQYFQTVTDYGK' | 'Apolipoprotein A-II' |
| 3386 | 'APOA2_No6' | 'VKSPELQAEAK' | 'Apolipoprotein A-II' |
| 3387 | 'APOA2_No7' | 'SKEQLTPLIKK' | 'Apolipoprotein A-II' |
| 3388 | 'APOA2_No8' | 'SPELQAEAK' | 'Apolipoprotein A-II' |
| 3389 | 'APOA2_No9' | 'SYFEKSKEQLTPLIK' | 'Apolipoprotein A-II' |

| | A | B | C |
|---|---|---|---|
| 1 | **Peptide Name** | **Peptide Sequence** | **Peptide Description** |
| 3390 | 'APOA2_No10' | 'EPCVESLVSQYFQTVTDYGKDLMEK' | 'Apolipoprotein A-II' |
| 3391 | 'APOA2_No11' | 'QAKEPCVESLVSQYFQTVTDYGK' | 'Apolipoprotein A-II' |
| 3392 | 'APOA2_No12' | 'DLMEKVKSPELQAEAK' | 'Apolipoprotein A-II' |
| 3393 | 'APOA2_No13' | 'DLMEKVKSPELQAEAK' | 'Apolipoprotein A-II' |
| 3394 | 'APOA2_No14' | 'SPELQAEAKSYFEK' | 'Apolipoprotein A-II' |
| 3395 | 'APOA2_No15' | 'VKSPELQAEAKSYFEK' | 'Apolipoprotein A-II' |
| 3396 | 'APOC1_No1' | 'LKEFGNTLEDK' | 'Apolipoprotein C-I' |
| 3397 | 'APOC1_No2' | 'EFGNTLEDKAR' | 'Apolipoprotein C-I' |
| 3398 | 'APOC1_No3' | 'MREWFSETFQK' | 'Apolipoprotein C-I' |
| 3399 | 'APOC1_No4' | 'EWFSETFQK' | 'Apolipoprotein C-I' |
| 3400 | 'APOC1_No5' | 'LKEFGNTLEDKAR' | 'Apolipoprotein C-I' |
| 3401 | 'APOC1_No6' | 'MREWFSETFQK' | 'Apolipoprotein C-I' |
| 3402 | 'APOC1_No7' | 'EFGNTLEDK' | 'Apolipoprotein C-I' |
| 3403 | 'APOC1_No8' | 'IKQSELSAK' | 'Apolipoprotein C-I' |
| 3404 | 'APOC1_No9' | 'MREWFSETFQKVK' | 'Apolipoprotein C-I' |
| 3405 | 'APOC1_No10' | 'MREWFSETFQKVK' | 'Apolipoprotein C-I' |
| 3406 | 'APOC2_No1' | 'TYLPAVDEKLR' | 'Apolipoprotein C-II' |
| 3407 | 'APOC2_No2' | 'TYLPAVDEK' | 'Apolipoprotein C-II' |
| 3408 | 'APOC2_No3' | 'ESLSSYWESAK' | 'Apolipoprotein C-II' |
| 3409 | 'APOC2_No4' | 'TYLPAVDEKLRDLYSK' | 'Apolipoprotein C-II' |
| 3410 | 'APOC2_No5' | 'TAAQNLYEK' | 'Apolipoprotein C-II' |
| 3411 | 'APOC2_No6' | 'STAAMSTYTGIFTDQVLSVLKGEE' | 'Apolipoprotein C-II' |
| 3412 | 'APOC2_No7' | 'LRDLYSK' | 'Apolipoprotein C-II' |
| 3413 | 'APOC2_No8' | 'STAAMSTYTGIFTDQVLSVLK' | 'Apolipoprotein C-II' |
| 3414 | 'APOC2_No9' | 'STAAMSTYTGIFTDQVLSVLK' | 'Apolipoprotein C-II' |
| 3415 | 'APOC3_No1' | 'GWVTDGFSSLKDYWSTVK' | 'Apolipoprotein C-III' |
| 3416 | 'APOC3_No2' | 'DKFSEFWDLDPEVRPTSAVAA' | 'Apolipoprotein C-III' |
| 3417 | 'APOC3_No3' | 'FSEFWDLDPEVRPTSAVAA' | 'Apolipoprotein C-III' |
| 3418 | 'APOC3_No4' | 'GWVTDGFSSLK' | 'Apolipoprotein C-III' |
| 3419 | 'APOC3_No5' | 'DYWSTVK' | 'Apolipoprotein C-III' |
| 3420 | 'APOC3_No6' | 'GWVTDGFSSLKDYWSTVKDK' | 'Apolipoprotein C-III' |
| 3421 | 'APOC3_No7' | 'DALSSVQESQVAQQAR' | 'Apolipoprotein C-III' |
| 3422 | 'APOC3_No8' | 'TAKDALSSVQESQVAQQAR' | 'Apolipoprotein C-III' |
| 3423 | 'APOC3_No9' | 'DYWSTVKDK' | 'Apolipoprotein C-III' |
| 3424 | 'APOC3_No10' | 'HATKTAKDALSSVQESQVAQQAR' | 'Apolipoprotein C-III' |
| 3425 | 'FGA_No1' | 'DSHSLTTNIMEILR' | 'Fibrinogen alpha chain' |
| 3426 | 'FGA_No2' | 'MELERPGGNEITR' | 'Fibrinogen alpha chain' |
| 3427 | 'FGA_No3' | 'DSHSLTTNIMEILR' | 'Fibrinogen alpha chain' |
| 3428 | 'FGA_No4' | 'EVDLKDYEDQQK' | 'Fibrinogen alpha chain' |
| 3429 | 'FGA_No5' | 'GSAGHWTSESSVSGSTGQWHSESGSFRPDSPGSGNAR' | 'Fibrinogen alpha chain' |
| 3430 | 'FGA_No6' | 'MKPVPDLVPGNFK' | 'Fibrinogen alpha chain' |
| 3431 | 'FGA_No7' | 'ESSSHHPGIAEFPSR' | 'Fibrinogen alpha chain' |
| 3432 | 'FGA_No8' | 'MELERPGGNEITR' | 'Fibrinogen alpha chain' |
| 3433 | 'FGA_No9' | 'ALTDMPQMR' | 'Fibrinogen alpha chain' |

| | A | B | C |
|---|---|---|---|
| 1 | **Peptide Name** | **Peptide Sequence** | **Peptide Description** |
| 3434 | 'FGA_No10' | 'QHLPLIK' | 'Fibrinogen alpha chain' |
| 3435 | 'FGA_No11' | 'ALTDMPQMR' | 'Fibrinogen alpha chain' |
| 3436 | 'FGA_No12' | 'ALTDMPQMR' | 'Fibrinogen alpha chain' |
| 3437 | 'FGA_No13' | 'LKNSLFEYQK' | 'Fibrinogen alpha chain' |
| 3438 | 'FGA_No14' | 'PGSTGTWNPGSSER' | 'Fibrinogen alpha chain' |
| 3439 | 'FGA_No15' | 'DLLPSRDR' | 'Fibrinogen alpha chain' |
| 3440 | 'FGA_No16' | 'QFTSSTSYNR' | 'Fibrinogen alpha chain' |
| 3441 | 'FGA_No17' | 'HRHPDEAAFFDTASTGK' | 'Fibrinogen alpha chain' |
| 3442 | 'FGA_No18' | 'MKPVPDLVPGNFK' | 'Fibrinogen alpha chain' |
| 3443 | 'FGA_No19' | 'QFTSSTSYNRGDSTFESK' | 'Fibrinogen alpha chain' |
| 3444 | 'FGA_No20' | 'DRQHLPLIK' | 'Fibrinogen alpha chain' |
| 3445 | 'FGA_No21' | 'VIEKVQHIQLLQK' | 'Fibrinogen alpha chain' |
| 3446 | 'FGA_No22' | 'VQHIQLLQK' | 'Fibrinogen alpha chain' |
| 3447 | 'FGA_No23' | 'QLEQVIAKDLLPSR' | 'Fibrinogen alpha chain' |
| 3448 | 'FGA_No24' | 'GDFSSANNRDNTYNR' | 'Fibrinogen alpha chain' |
| 3449 | 'FGA_No25' | 'NPSSAGSWNSGSSGPGSTGNR' | 'Fibrinogen alpha chain' |
| 3450 | 'FGA_No26' | 'ALTDMPQMR' | 'Fibrinogen alpha chain' |
| 3451 | 'FGA_No27' | 'PNNPDWGTFEEVSGNVSPGTRR' | 'Fibrinogen alpha chain' |
| 3452 | 'FGA_No28' | 'GLIDEVNQDFTNR' | 'Fibrinogen alpha chain' |
| 3453 | 'FGA_No29' | 'NNKDSHSLTTNIMEILR' | 'Fibrinogen alpha chain' |
| 3454 | 'FGA_No30' | 'NSLFEYQK' | 'Fibrinogen alpha chain' |
| 3455 | 'FGA_No31' | 'NNKDSHSLTTNIMEILR' | 'Fibrinogen alpha chain' |
| 3456 | 'FGA_No32' | 'TVIGPDGHKEVTK' | 'Fibrinogen alpha chain' |
| 3457 | 'FGA_No33' | 'TFPGFFSPMLGEFVSETESR' | 'Fibrinogen alpha chain' |
| 3458 | 'FGA_No34' | 'NNSPYEIENGVVWVSFR' | 'Fibrinogen alpha chain' |
| 3459 | 'FGA_No35' | 'QLEQVIAK' | 'Fibrinogen alpha chain' |
| 3460 | 'FGA_No36' | 'LEVDIDIK' | 'Fibrinogen alpha chain' |
| 3461 | 'FGA_No37' | 'GDFSSANNR' | 'Fibrinogen alpha chain' |
| 3462 | 'FGA_No38' | 'PVPDLVPGNFK' | 'Fibrinogen alpha chain' |
| 3463 | 'FGA_No39' | 'MKGLIDEVNQDFTNR' | 'Fibrinogen alpha chain' |
| 3464 | 'FGA_No40' | 'QLEQVIAKDLLPSR' | 'Fibrinogen alpha chain' |
| 3465 | 'FGA_No41' | 'TVIGPDGHK' | 'Fibrinogen alpha chain' |
| 3466 | 'FGA_No42' | 'PNNPDWGTFEEVSGNVSPGTR' | 'Fibrinogen alpha chain' |
| 3467 | 'FGA_No43' | 'VELEDWAGNEAYAEYHFR' | 'Fibrinogen alpha chain' |
| 3468 | 'FGA_No44' | 'DCDDVLQTHPSGTQSGIFNIK' | 'Fibrinogen alpha chain' |
| 3469 | 'FGA_No45' | 'TFPGFFSPMLGEFVSETESR' | 'Fibrinogen alpha chain' |
| 3470 | 'FGA_No46' | 'EVDLKDYEDQQKQLEQVIAK' | 'Fibrinogen alpha chain' |
| 3471 | 'FGA_No47' | 'GLIDEVNQDFTNRINK' | 'Fibrinogen alpha chain' |
| 3472 | 'FGA_No48' | 'KVIEKVQHIQLLQK' | 'Fibrinogen alpha chain' |
| 3473 | 'FGA_No49' | 'PVPDLVPGNFKSQLQK' | 'Fibrinogen alpha chain' |
| 3474 | 'FGA_No50' | 'RLEVDIDIKIR' | 'Fibrinogen alpha chain' |
| 3475 | 'FGA_No51' | 'EVVTSEDGSDCPEAMDLGTLSGIGTLDGFR' | 'Fibrinogen alpha chain' |
| 3476 | 'FGA_No52' | 'LEVDIDIKIR' | 'Fibrinogen alpha chain' |
| 3477 | 'FGA_No53' | 'RLEVDIDIK' | 'Fibrinogen alpha chain' |

| | A | B | C |
|---|---|---|---|
| 1 | **Peptide Name** | **Peptide Sequence** | **Peptide Description** |
| 3478 | 'FGA_No54' | 'DSDWPFCSDEDWNYKCPSGCR' | 'Fibrinogen alpha chain' |
| 3479 | 'FGA_No55' | 'MKGLIDEVNQDFTNR' | 'Fibrinogen alpha chain' |
| 3480 | 'FGA_No56' | 'DYEDQQK' | 'Fibrinogen alpha chain' |
| 3481 | 'FGA_No57' | 'QHLPLIK' | 'Fibrinogen alpha chain' |
| 3482 | 'FGA_No58' | 'HPDEAAFFDTASTGK' | 'Fibrinogen alpha chain' |
| 3483 | 'FGA_No59' | 'GSESGIFTNTK' | 'Fibrinogen alpha chain' |
| 3484 | 'FGA_No60' | 'MKGLIDEVNQDFTNRINK' | 'Fibrinogen alpha chain' |
| 3485 | 'FGA_No61' | 'ESSSHHPGIAEFPSRGK' | 'Fibrinogen alpha chain' |
| 3486 | 'FGA_No62' | 'DSDWPFCSDEDWNYK' | 'Fibrinogen alpha chain' |
| 3487 | 'FGA_No63' | 'GSESGIFTNTKESSSHHPGIAEFPSR' | 'Fibrinogen alpha chain' |
| 3488 | 'FGA_No64' | 'MADEAGSEADHEGTHSTKR' | 'Fibrinogen alpha chain' |
| 3489 | 'FGA_No65' | 'SQLQKVPPEWK' | 'Fibrinogen alpha chain' |
| 3490 | 'FGA_No66' | 'EVTKEVVTSEDGSDCPEAMDLGTLSGIGTLDGFR' | 'Fibrinogen alpha chain' |
| 3491 | 'FGA_No67' | 'NPGSSGTGGTATWK' | 'Fibrinogen alpha chain' |
| 3492 | 'FGA_No68' | 'MADEAGSEADHEGTHSTK' | 'Fibrinogen alpha chain' |
| 3493 | 'FGA_No69' | 'DYEDQQKQLEQVIAK' | 'Fibrinogen alpha chain' |
| 3494 | 'FGA_No70' | 'AQLVDMK' | 'Fibrinogen alpha chain' |
| 3495 | 'FGA_No71' | 'LVTSKGDKELR' | 'Fibrinogen alpha chain' |
| 3496 | 'FGA_No72' | 'QLEQVIAKDLLPSRDR' | 'Fibrinogen alpha chain' |
| 3497 | 'FGA_No73' | 'TVIGPDGHKEVTKEVVTSEDGSDCPEAMDLGTLSGIGTLDGFR' | 'Fibrinogen alpha chain' |
| 3498 | 'FGA_No74' | 'SYKMADEAGSEADHEGTHSTKR' | 'Fibrinogen alpha chain' |
| 3499 | 'FGA_No75' | 'NPGSSGTGGTATWKPGSSGPGSTGSWNSGSSGTGSTGNQNPGSPR' | 'Fibrinogen alpha chain' |
| 3500 | 'FGA_No76' | 'PGSSGPGSTGSWNSGSSGTGSTGNQNPGSPRPGSTGTWNPGSSER' | 'Fibrinogen alpha chain' |
| 3501 | 'FGA_No77' | 'MKPVPDLVPGNFKSQLQK' | 'Fibrinogen alpha chain' |
| 3502 | 'FGA_No78' | 'GLIDEVNQDFTNRINKLK' | 'Fibrinogen alpha chain' |
| 3503 | 'FGA_No79' | 'GSESGIFTNTKESSSHHPGIAEFPSRGK' | 'Fibrinogen alpha chain' |
| 3504 | 'FGA_No80' | 'MADEAGSEADHEGTHSTK' | 'Fibrinogen alpha chain' |
| 3505 | 'FGA_No81' | 'EVVTSEDGSDCPEAMDLGTLSGIGTLDGFR' | 'Fibrinogen alpha chain' |
| 3506 | 'FGA_No82' | 'GGSTSYGTGSETESPR' | 'Fibrinogen alpha chain' |
| 3507 | 'FGA_No83' | 'QFTSSTSYNRGDSTFESK' | 'Fibrinogen alpha chain' |
| 3508 | 'FGA_No84' | 'MKGLIDEVNQDFTNRINK' | 'Fibrinogen alpha chain' |
| 3509 | 'FGA_No85' | 'MADEAGSEADHEGTHSTKR' | 'Fibrinogen alpha chain' |
| 3510 | 'FGA_No86' | 'GDFSSANNRDNTYNRVSEDLR' | 'Fibrinogen alpha chain' |
| 3511 | 'FGA_No87' | 'NSLFEYQKNNKDSHSLTTNIMEILR' | 'Fibrinogen alpha chain' |
| 3512 | 'FGA_No88' | 'HQSACKDSDWPFCSDEDWNYK' | 'Fibrinogen alpha chain' |
| 3513 | 'FGA_No89' | 'VSEDLRSRIEVLK' | 'Fibrinogen alpha chain' |
| 3514 | 'FGA_No90' | 'EVTKEVVTSEDGSDCPEAMDLGTLSGIGTLDGFR' | 'Fibrinogen alpha chain' |
| 3515 | 'FGA_No91' | 'GDSTFESK' | 'Fibrinogen alpha chain' |
| 3516 | 'FGA_No92' | 'GSAGHWTSESSVSGSTGQWHSESGSFR' | 'Fibrinogen alpha chain' |
| 3517 | 'FGA_No93' | 'SSSYSKQFTSSTSYNR' | 'Fibrinogen alpha chain' |
| 3518 | 'FGA_No94' | 'AQLVDMKR' | 'Fibrinogen alpha chain' |
| 3519 | 'FGA_No95' | 'QFTSSTSYNRGDSTFESKSYK' | 'Fibrinogen alpha chain' |
| 3520 | 'FGA_No96' | 'QLEQVIAK' | 'Fibrinogen alpha chain' |
| 3521 | 'FGA_No97' | 'EYHTEKLVTSK' | 'Fibrinogen alpha chain' |

| | A | B | C |
|---|---|---|---|
| 1 | **Peptide Name** | **Peptide Sequence** | **Peptide Description** |
| 3522 | 'FGA_No98' | 'SYKMADEAGSEADHEGTHSTK' | 'Fibrinogen alpha chain' |
| 3523 | 'FGA_No99' | 'HQSACKDSDWPFCSDEDWNYKCPSGCR' | 'Fibrinogen alpha chain' |
| 3524 | 'FGA_No100' | 'TVIGPDGHKEVTKEVVTSEDGSDCPEAMDLGTLSGIGTLDGFR' | 'Fibrinogen alpha chain' |
| 3525 | 'FGA_No101' | 'QLEQVIAKDLLPSRDR' | 'Fibrinogen alpha chain' |
| 3526 | 'FGA_No102' | 'VTSGSTTTTR' | 'Fibrinogen alpha chain' |
| 3527 | 'FGA_No103' | 'VQHIQLLQKNVR' | 'Fibrinogen alpha chain' |
| 3528 | 'FGA_No104' | 'PGGNEITR' | 'Fibrinogen alpha chain' |
| 3529 | 'FGA_No105' | 'DLLPSRDRQHLPLIK' | 'Fibrinogen alpha chain' |
| 3530 | 'FGA_No106' | 'MKPVPDLVPGNFKSQLQK' | 'Fibrinogen alpha chain' |
| 3531 | 'FGA_No107' | 'EYHTEKLVTSKGDK' | 'Fibrinogen alpha chain' |
| 3532 | 'FGA_No108' | 'SYKMADEAGSEADHEGTHSTKR' | 'Fibrinogen alpha chain' |
| 3533 | 'FGA_No109' | 'SYKMADEAGSEADHEGTHSTK' | 'Fibrinogen alpha chain' |
| 3534 | 'FGA_No110' | 'SRPVRDCDDVLQTHPSGTQSGIFNIK' | 'Fibrinogen alpha chain' |
| 3535 | 'FGA_No111' | 'PGSSGPGSTGSWNSGSSGTGSTGNQNPGSPR' | 'Fibrinogen alpha chain' |
| 3536 | 'FGA_No112' | 'VIEKVQHIQLLQKNVR' | 'Fibrinogen alpha chain' |
| 3537 | 'FGA_No113' | 'TVTKTVIGPDGHKEVTK' | 'Fibrinogen alpha chain' |
| 3538 | 'FGA_No114' | 'QFTSSTSYNRGDSTFESKSYK' | 'Fibrinogen alpha chain' |
| 3539 | 'FGA_No115' | 'DCDDVLQTHPSGTQSGIFNIKLPGSSK' | 'Fibrinogen alpha chain' |
| 3540 | 'FGA_No116' | 'VVERHQSACKDSDWPFCSDEDWNYK' | 'Fibrinogen alpha chain' |
| 3541 | 'FGA_No117' | 'MELERPGGNEITRGGSTSYGTGSETESPR' | 'Fibrinogen alpha chain' |
| 3542 | 'FGA_No118' | 'EVTKEVVTSEDGSDCPEAMDLGTLSGIGTLDGFRHR' | 'Fibrinogen alpha chain' |
| 3543 | 'FGA_No119' | 'NSLFEYQKNNKDSHSLTTNIMEILR' | 'Fibrinogen alpha chain' |
| 3544 | 'FGA_No120' | 'GGSTSYGTGSETESPRNPSSAGSWNSGSSGPGSTGNR' | 'Fibrinogen alpha chain' |
| 3545 | 'FGA_No121' | 'NSLFEYQKNNK' | 'Fibrinogen alpha chain' |
| 3546 | 'FGA_No122' | 'EVVTSEDGSDCPEAMDLGTLSGIGTLDGFRHR' | 'Fibrinogen alpha chain' |
| 3547 | 'FGA_No123' | 'VPPEWKALTDMPQMR' | 'Fibrinogen alpha chain' |
| 3548 | 'FGA_No124' | 'DYEDQQKQLEQVIAKDLLPSR' | 'Fibrinogen alpha chain' |
| 3549 | 'FGA_No125' | 'SSSYSKQFTSSTSYNRGDSTFESK' | 'Fibrinogen alpha chain' |
| 3550 | 'FGA_No126' | 'DNTYNRVSEDLR' | 'Fibrinogen alpha chain' |
| 3551 | 'FGA_No127' | 'AQLVDMKRLEVDIDIK' | 'Fibrinogen alpha chain' |
| 3552 | 'FGA_No128' | 'INKLKNSLFEYQK' | 'Fibrinogen alpha chain' |
| 3553 | 'FGA_No129' | 'EVVTSEDGSDCPEAMDLGTLSGIGTLDGFRHR' | 'Fibrinogen alpha chain' |
| 3554 | 'FGA_No130' | 'EVVTSEDGSDCPEAMDLGTLSGIGTLDGFRHRHPDEAAFFDTASTGK' | 'Fibrinogen alpha chain' |
| 3555 | 'FGA_No131' | 'TFPGFFSPMLGEFVSETESRGSESGIFTNTK' | 'Fibrinogen alpha chain' |
| 3556 | 'FGA_No132' | 'AQLVDMKRLEVDIDIK' | 'Fibrinogen alpha chain' |
| 3557 | 'FGA_No133' | 'NNKDSHSLTTNIMEILRGDFSSANNR' | 'Fibrinogen alpha chain' |
| 3558 | 'FGA_No134' | 'DSDWPFCSDEDWNYKCPSGCRMK' | 'Fibrinogen alpha chain' |
| 3559 | 'FGA_No135' | 'EKVTSGSTTTTR' | 'Fibrinogen alpha chain' |
| 3560 | 'FGA_No136' | 'TFPGFFSPMLGEFVSETESRGSESGIFTNTK' | 'Fibrinogen alpha chain' |
| 3561 | 'FGA_No137' | 'NPSSAGSWNSGSSGPGSTGNRNPGSSGTGGTATWK' | 'Fibrinogen alpha chain' |
| 3562 | 'FGA_No138' | 'REYHTEKLVTSK' | 'Fibrinogen alpha chain' |
| 3563 | 'FGA_No139' | 'NVRAQLVDMK' | 'Fibrinogen alpha chain' |
| 3564 | 'FGA_No140' | 'ESSSHHPGIAEFPSRGKSSSYSK' | 'Fibrinogen alpha chain' |
| 3565 | 'FGA_No141' | 'EVTKEVVTSEDGSDCPEAMDLGTLSGIGTLDGFRHR' | 'Fibrinogen alpha chain' |

| | A | B | C |
|---|---|---|---|
| 1 | **Peptide Name** | **Peptide Sequence** | **Peptide Description** |
| 3566 | 'FGA_No142' | 'TVTKTVIGPDGHK' | 'Fibrinogen alpha chain' |
| 3567 | 'FGA_No143' | 'EKVTSGSTTTTRR' | 'Fibrinogen alpha chain' |
| 3568 | 'FGA_No144' | 'NVRAQLVDMK' | 'Fibrinogen alpha chain' |
| 3569 | 'FGA_No145' | 'DNTYNRVSEDLRSR' | 'Fibrinogen alpha chain' |
| 3570 | 'FGA_No146' | 'NVRAQLVDMKR' | 'Fibrinogen alpha chain' |
| 3571 | 'FGA_No147' | 'PDSPGSGNARPNNPDWGTFEEVSGNVSPGTR' | 'Fibrinogen alpha chain' |
| 3572 | 'FGB_No1' | 'NYCGLPGEYWLGNDK' | 'Fibrinogen beta chain' |
| 3573 | 'FGB_No2' | 'KAPDAGGCLHADPDLGVLCPTGCQLQEALLQQERPIR' | 'Fibrinogen beta chain' |
| 3574 | 'FGB_No3' | 'NYCGLPGEYWLGNDKISQLTR' | 'Fibrinogen beta chain' |
| 3575 | 'FGB_No4' | 'MGPTELLIEMEDWKGDK' | 'Fibrinogen beta chain' |
| 3576 | 'FGB_No5' | 'AHYGGFTVQNEANK' | 'Fibrinogen beta chain' |
| 3577 | 'FGB_No6' | 'EDGGGWWYNR' | 'Fibrinogen beta chain' |
| 3578 | 'FGB_No7' | 'TPCTVSCNIPVVSGKECEEIIR' | 'Fibrinogen beta chain' |
| 3579 | 'FGB_No8' | 'YYWGGQYTWDMAK' | 'Fibrinogen beta chain' |
| 3580 | 'FGB_No9' | 'SILENLR' | 'Fibrinogen beta chain' |
| 3581 | 'FGB_No10' | 'EEAPSLRPAPPPISGGGYR' | 'Fibrinogen beta chain' |
| 3582 | 'FGB_No11' | 'TPCTVSCNIPVVSGKECEEIIRK' | 'Fibrinogen beta chain' |
| 3583 | 'FGB_No12' | 'MGPTELLIEMEDWKGDKVK' | 'Fibrinogen beta chain' |
| 3584 | 'FGB_No13' | 'MGPTELLIEMEDWKGDK' | 'Fibrinogen beta chain' |
| 3585 | 'FGB_No14' | 'IQKLESDVSAQMEYCR' | 'Fibrinogen beta chain' |
| 3586 | 'FGB_No15' | 'DNENVVNEYSSELEK' | 'Fibrinogen beta chain' |
| 3587 | 'FGB_No16' | 'AHYGGFTVQNEANKYQISVNKYR' | 'Fibrinogen beta chain' |
| 3588 | 'FGB_No17' | 'QGFGNVATNTDGK' | 'Fibrinogen beta chain' |
| 3589 | 'FGB_No18' | 'PAPPPISGGGYR' | 'Fibrinogen beta chain' |
| 3590 | 'FGB_No19' | 'REEAPSLRPAPPPISGGGYR' | 'Fibrinogen beta chain' |
| 3591 | 'FGB_No20' | 'MGPTELLIEMEDWKGDKVK' | 'Fibrinogen beta chain' |
| 3592 | 'FGB_No21' | 'KWDPYKQGFGNVATNTDGK' | 'Fibrinogen beta chain' |
| 3593 | 'FGB_No22' | 'GGETSEMYLIQPDSSVKPYR' | 'Fibrinogen beta chain' |
| 3594 | 'FGB_No23' | 'QDGSVDFGR' | 'Fibrinogen beta chain' |
| 3595 | 'FGB_No24' | 'MGPTELLIEMEDWKGDK' | 'Fibrinogen beta chain' |
| 3596 | 'FGB_No25' | 'ECEEIIR' | 'Fibrinogen beta chain' |
| 3597 | 'FGB_No26' | 'YQISVNK' | 'Fibrinogen beta chain' |
| 3598 | 'FGB_No27' | 'AHYGGFTVQNEANKYQISVNK' | 'Fibrinogen beta chain' |
| 3599 | 'FGB_No28' | 'HGTDDGVVWMNWK' | 'Fibrinogen beta chain' |
| 3600 | 'FGB_No29' | 'QCSKEDGGGWWYNR' | 'Fibrinogen beta chain' |
| 3601 | 'FGB_No30' | 'HGTDDGVVWMNWK' | 'Fibrinogen beta chain' |
| 3602 | 'FGB_No31' | 'WDPYKQGFGNVATNTDGK' | 'Fibrinogen beta chain' |
| 3603 | 'FGB_No32' | 'TMTIHNGMFFSTYDR' | 'Fibrinogen beta chain' |
| 3604 | 'FGB_No33' | 'QVKDNENVVNEYSSELEK' | 'Fibrinogen beta chain' |
| 3605 | 'FGB_No34' | 'QGFGNVATNTDGKNYCGLPGEYWLGNDKISQLTR' | 'Fibrinogen beta chain' |
| 3606 | 'FGB_No35' | 'DNDGWLTSDPR' | 'Fibrinogen beta chain' |
| 3607 | 'FGB_No36' | 'TMTIHNGMFFSTYDR' | 'Fibrinogen beta chain' |
| 3608 | 'FGB_No37' | 'NSVDELNNNVEAVSQTSSSSFQYMYLLK' | 'Fibrinogen beta chain' |
| 3609 | 'FGB_No38' | 'QGFGNVATNTDGK' | 'Fibrinogen beta chain' |

| | A | B | C |
|---|---|---|---|
| 1 | **Peptide Name** | **Peptide Sequence** | **Peptide Description** |
| 3610 | 'FGB_No39' | 'MGPTELLIEMEDWKGDKVK' | 'Fibrinogen beta chain' |
| 3611 | 'FGB_No40' | 'TMTIHNGMFFSTYDR' | 'Fibrinogen beta chain' |
| 3612 | 'FGB_No41' | 'KGGETSEMYLIQPDSSVKPYR' | 'Fibrinogen beta chain' |
| 3613 | 'FGB_No42' | 'MGPTELLIEMEDWKGDKVK' | 'Fibrinogen beta chain' |
| 3614 | 'FGB_No43' | 'TMTIHNGMFFSTYDR' | 'Fibrinogen beta chain' |
| 3615 | 'FGB_No44' | 'TMTIHNGMFFSTYDRDNDGWLTSDPRK' | 'Fibrinogen beta chain' |
| 3616 | 'FGB_No45' | 'MGPTELLIEMEDWK' | 'Fibrinogen beta chain' |
| 3617 | 'FGB_No46' | 'QCSKEDGGGWWYNR' | 'Fibrinogen beta chain' |
| 3618 | 'FGB_No47' | 'QVKDNENVVNEYSSELEK' | 'Fibrinogen beta chain' |
| 3619 | 'FGB_No48' | 'GSWYSMR' | 'Fibrinogen beta chain' |
| 3620 | 'FGB_No49' | 'QDGSVDFGRK' | 'Fibrinogen beta chain' |
| 3621 | 'FGB_No50' | 'NSVDELNNNVEAVSQTSSSSFQYMYLLKDLWQKR' | 'Fibrinogen beta chain' |
| 3622 | 'FGB_No51' | 'MGPTELLIEMEDWK' | 'Fibrinogen beta chain' |
| 3623 | 'FGB_No52' | 'SKIQKLESDVSAQMEYCR' | 'Fibrinogen beta chain' |
| 3624 | 'FGB_No53' | 'TMTIHNGMFFSTYDRDNDGWLTSDPR' | 'Fibrinogen beta chain' |
| 3625 | 'FGB_No54' | 'NSVDELNNNVEAVSQTSSSSFQYMYLLKDLWQKR' | 'Fibrinogen beta chain' |
| 3626 | 'FGB_No55' | 'DNDGWLTSDPRK' | 'Fibrinogen beta chain' |
| 3627 | 'FGB_No56' | 'DNENVVNEYSSELEKHQLYIDETVNSNIPTNLR' | 'Fibrinogen beta chain' |
| 3628 | 'FGB_No57' | 'MGPTELLIEMEDWK' | 'Fibrinogen beta chain' |
| 3629 | 'FGB_No58' | 'TMTIHNGMFFSTYDRDNDGWLTSDPRK' | 'Fibrinogen beta chain' |
| 3630 | 'FGB_No59' | 'KGGETSEMYLIQPDSSVK' | 'Fibrinogen beta chain' |
| 3631 | 'FGB_No60' | 'VYCDMNTENGGWTVIQNR' | 'Fibrinogen beta chain' |
| 3632 | 'FGB_No61' | 'GGETSEMYLIQPDSSVKPYRVYCDMNTENGGWTVIQNR' | 'Fibrinogen beta chain' |
| 3633 | 'FGB_No62' | 'TMTIHNGMFFSTYDRDNDGWLTSDPRK' | 'Fibrinogen beta chain' |
| 3634 | 'FGB_No63' | 'ECEEIIRK' | 'Fibrinogen beta chain' |
| 3635 | 'FGB_No64' | 'GGETSEMYLIQPDSSVKPYR' | 'Fibrinogen beta chain' |
| 3636 | 'FGB_No65' | 'QVKDNENVVNEYSSELEKHQLYIDETVNSNIPTNLR' | 'Fibrinogen beta chain' |
| 3637 | 'FGB_No66' | 'TMTIHNGMFFSTYDRDNDGWLTSDPRK' | 'Fibrinogen beta chain' |
| 3638 | 'FGB_No67' | 'IRPFFPQQ' | 'Fibrinogen beta chain' |
| 3639 | 'FGB_No68' | 'YQISVNKYR' | 'Fibrinogen beta chain' |
| 3640 | 'FGB_No69' | 'LESDVSAQMEYCR' | 'Fibrinogen beta chain' |
| 3641 | 'FGB_No70' | 'TMTIHNGMFFSTYDRDNDGWLTSDPR' | 'Fibrinogen beta chain' |
| 3642 | 'FGB_No71' | 'HQLYIDETVNSNIPTNLR' | 'Fibrinogen beta chain' |
| 3643 | 'FGB_No72' | 'KGGETSEMYLIQPDSSVKPYR' | 'Fibrinogen beta chain' |
| 3644 | 'FGB_No73' | 'QDGSVDFGRK' | 'Fibrinogen beta chain' |
| 3645 | 'FGB_No74' | 'VYCDMNTENGGWTVIQNR' | 'Fibrinogen beta chain' |
| 3646 | 'FGB_No75' | 'APDAGGCLHADPDLGVLCPTGCQLQEALLQQERPIR' | 'Fibrinogen beta chain' |
| 3647 | 'FGB_No76' | 'IQKLESDVSAQMEYCR' | 'Fibrinogen beta chain' |
| 3648 | 'FGB_No77' | 'LESDVSAQMEYCR' | 'Fibrinogen beta chain' |
| 3649 | 'FGB_No78' | 'MGPTELLIEMEDWKGDK' | 'Fibrinogen beta chain' |
| 3650 | 'FGB_No79' | 'SKIQKLESDVSAQMEYCR' | 'Fibrinogen beta chain' |
| 3651 | 'FGB_No80' | 'MGPTELLIEMEDWK' | 'Fibrinogen beta chain' |
| 3652 | 'FGB_No81' | 'KQCSKEDGGGWWYNR' | 'Fibrinogen beta chain' |
| 3653 | 'FGB_No82' | 'YYWGGQYTWDMAK' | 'Fibrinogen beta chain' |

| | A | B | C |
|---|---|---|---|
| 1 | **Peptide Name** | **Peptide Sequence** | **Peptide Description** |
| 3654 | 'FGB_No83' | 'TMTIHNGMFFSTYDRDNDGWLTSDPR' | 'Fibrinogen beta chain' |
| 3655 | 'FGB_No84' | 'TMTIHNGMFFSTYDRDNDGWLTSDPR' | 'Fibrinogen beta chain' |
| 3656 | 'FGB_No85' | 'TPCTVSCNIPVVSGK' | 'Fibrinogen beta chain' |
| 3657 | 'FGB_No86' | 'QVKDNENVVNEYSSELEKHQLYIDETVNSNIPTNLR' | 'Fibrinogen beta chain' |
| 3658 | 'FGB_No87' | 'VKAHYGGFTVQNEANK' | 'Fibrinogen beta chain' |
| 3659 | 'FGB_No88' | 'QKQVKDNENVVNEYSSELEK' | 'Fibrinogen beta chain' |
| 3660 | 'FGB_No89' | 'VKAHYGGFTVQNEANKYQISVNK' | 'Fibrinogen beta chain' |
| 3661 | 'FGB_No90' | 'QKQVKDNENVVNEYSSELEK' | 'Fibrinogen beta chain' |
| 3662 | 'FGB_No91' | 'HGTDDGVVWMNWKGSWYSMR' | 'Fibrinogen beta chain' |
| 3663 | 'FGB_No92' | 'GTAGNALMDGASQLMGENR' | 'Fibrinogen beta chain' |
| 3664 | 'FGB_No93' | 'GGETSEMYLIQPDSSVKPYRVYCDMNTENGGWTVIQNR' | 'Fibrinogen beta chain' |
| 3665 | 'FGB_No94' | 'MSMKIRPFFPQQ' | 'Fibrinogen beta chain' |
| 3666 | 'FGB_No95' | 'DNDGWLTSDPRKQCSK' | 'Fibrinogen beta chain' |
| 3667 | 'FGB_No96' | 'GGETSEMYLIQPDSSVK' | 'Fibrinogen beta chain' |
| 3668 | 'FGB_No97' | 'VLRSILENLR' | 'Fibrinogen beta chain' |
| 3669 | 'FGB_No98' | 'QGFGNVATNTDGKNYCGLPGEYWLGNDK' | 'Fibrinogen beta chain' |
| 3670 | 'FGB_No99' | 'HGTDDGVVWMNWKGSWYSMRK' | 'Fibrinogen beta chain' |
| 3671 | 'FGB_No100' | 'KAPDAGGCLHADPDLGVLCPTGCQLQEALLQQER' | 'Fibrinogen beta chain' |
| 3672 | 'FGB_No101' | 'QGFGNVATNTDGKNYCGLPGEYWLGNDK' | 'Fibrinogen beta chain' |
| 3673 | 'FGB_No102' | 'APDAGGCLHADPDLGVLCPTGCQLQEALLQQER' | 'Fibrinogen beta chain' |
| 3674 | 'FGB_No103' | 'HGTDDGVVWMNWKGSWYSMRK' | 'Fibrinogen beta chain' |
| 3675 | 'FGB_No104' | 'KGGETSEMYLIQPDSSVK' | 'Fibrinogen beta chain' |
| 3676 | 'FGB_No105' | 'WDPYKQGFGNVATNTDGKNYCGLPGEYWLGNDK' | 'Fibrinogen beta chain' |
| 3677 | 'FGB_No106' | 'NSVDELNNNVEAVSQTSSSSFQYMYLLK' | 'Fibrinogen beta chain' |
| 3678 | 'FGB_No107' | 'VYCDMNTENGGWTVIQNRQDGSVDFGRK' | 'Fibrinogen beta chain' |
| 3679 | 'FGB_No108' | 'GTAGNALMDGASQLMGENR' | 'Fibrinogen beta chain' |
| 3680 | 'FGB_No109' | 'HGTDDGVVWMNWKGSWYSMRK' | 'Fibrinogen beta chain' |
| 3681 | 'FGB_No110' | 'HGTDDGVVWMNWKGSWYSMR' | 'Fibrinogen beta chain' |
| 3682 | 'FGB_No111' | 'REEAPSLR' | 'Fibrinogen beta chain' |
| 3683 | 'FGB_No112' | 'HGTDDGVVWMNWKGSWYSMRK' | 'Fibrinogen beta chain' |
| 3684 | 'FGB_No113' | 'HGTDDGVVWMNWKGSWYSMR' | 'Fibrinogen beta chain' |
| 3685 | 'FGB_No114' | 'GGETSEMYLIQPDSSVK' | 'Fibrinogen beta chain' |
| 3686 | 'FGB_No115' | 'QDGSVDFGR' | 'Fibrinogen beta chain' |
| 3687 | 'FGB_No116' | 'MSMKIRPFFPQQ' | 'Fibrinogen beta chain' |
| 3688 | 'FGB_No117' | 'VYCDMNTENGGWTVIQNRQDGSVDFGR' | 'Fibrinogen beta chain' |
| 3689 | 'FGB_No118' | 'IQKLESDVSAQMEYCRTPCTVSCNIPVVSGK' | 'Fibrinogen beta chain' |
| 3690 | 'FGB_No119' | 'HGTDDGVVWMNWKGSWYSMR' | 'Fibrinogen beta chain' |
| 3691 | 'FGB_No120' | 'GHRPLDK' | 'Fibrinogen beta chain' |
| 3692 | 'FGB_No121' | 'DNENVVNEYSSELEKHQLYIDETVNSNIPTNLRVLR' | 'Fibrinogen beta chain' |
| 3693 | 'FGB_No122' | 'GTAGNALMDGASQLMGENR' | 'Fibrinogen beta chain' |
| 3694 | 'FGB_No123' | 'KREEAPSLR' | 'Fibrinogen beta chain' |
| 3695 | 'FGB_No124' | 'CHAANPNGR' | 'Fibrinogen beta chain' |
| 3696 | 'FGB_No125' | 'GTAGNALMDGASQLMGENR' | 'Fibrinogen beta chain' |
| 3697 | 'FGB_No126' | 'VYCDMNTENGGWTVIQNRQDGSVDFGR' | 'Fibrinogen beta chain' |

| | A | B | C |
|---|---|---|---|
| 1 | **Peptide Name** | **Peptide Sequence** | **Peptide Description** |
| 3698 | 'FGB_No127' | 'QDGSVDFGRKWDPYK' | 'Fibrinogen beta chain' |
| 3699 | 'FGB_No128' | 'QCSKEDGGGWWYNRCHAANPNGR' | 'Fibrinogen beta chain' |
| 3700 | 'FGG_No1' | 'QSGLYFIKPLK' | 'Fibrinogen gamma chain' |
| 3701 | 'FGG_No2' | 'YEASILTHDSSIR' | 'Fibrinogen gamma chain' |
| 3702 | 'FGG_No3' | 'FGSYCPTTCGIADFLSTYQTK' | 'Fibrinogen gamma chain' |
| 3703 | 'FGG_No4' | 'AIQLTYNPDESSKPNMIDAATLK' | 'Fibrinogen gamma chain' |
| 3704 | 'FGG_No5' | 'VAQLEAQCQEPCKDTVQIHDITGK' | 'Fibrinogen gamma chain' |
| 3705 | 'FGG_No6' | 'VAQLEAQCQEPCKDTVQIHDITGKDCQDIANK' | 'Fibrinogen gamma chain' |
| 3706 | 'FGG_No7' | 'ANQQFLVYCEIDGSGNGWTVFQK' | 'Fibrinogen gamma chain' |
| 3707 | 'FGG_No8' | 'IHLISTQSAIPYALRVELEDWNGR' | 'Fibrinogen gamma chain' |
| 3708 | 'FGG_No9' | 'VELEDWNGR' | 'Fibrinogen gamma chain' |
| 3709 | 'FGG_No10' | 'EGFGHLSPTGTTEFWLGNEKIHLISTQSAIPYALR' | 'Fibrinogen gamma chain' |
| 3710 | 'FGG_No11' | 'MLEEIMKYEASILTHDSSIR' | 'Fibrinogen gamma chain' |
| 3711 | 'FGG_No12' | 'VAQLEAQCQEPCK' | 'Fibrinogen gamma chain' |
| 3712 | 'FGG_No13' | 'MLEEIMK' | 'Fibrinogen gamma chain' |
| 3713 | 'FGG_No14' | 'VGPEADKYR' | 'Fibrinogen gamma chain' |
| 3714 | 'FGG_No15' | 'QSGLYFIK' | 'Fibrinogen gamma chain' |
| 3715 | 'FGG_No16' | 'RLDGSVDFKK' | 'Fibrinogen gamma chain' |
| 3716 | 'FGG_No17' | 'MLEEIMK' | 'Fibrinogen gamma chain' |
| 3717 | 'FGG_No18' | 'LTIGEGQQHHLGGAK' | 'Fibrinogen gamma chain' |
| 3718 | 'FGG_No19' | 'KMLEEIMKYEASILTHDSSIR' | 'Fibrinogen gamma chain' |
| 3719 | 'FGG_No20' | 'LDGSVDFKK' | 'Fibrinogen gamma chain' |
| 3720 | 'FGG_No21' | 'KMLEEIMKYEASILTHDSSIR' | 'Fibrinogen gamma chain' |
| 3721 | 'FGG_No22' | 'EGFGHLSPTGTTEFWLGNEK' | 'Fibrinogen gamma chain' |
| 3722 | 'FGG_No23' | 'ANQQFLVYCEIDGSGNGWTVFQKR' | 'Fibrinogen gamma chain' |
| 3723 | 'FGG_No24' | 'PNMIDAATLK' | 'Fibrinogen gamma chain' |
| 3724 | 'FGG_No25' | 'ASTPNGYDNGIIWATWK' | 'Fibrinogen gamma chain' |
| 3725 | 'FGG_No26' | 'TSTADYAMFK' | 'Fibrinogen gamma chain' |
| 3726 | 'FGG_No27' | 'QSGLYFIK' | 'Fibrinogen gamma chain' |
| 3727 | 'FGG_No28' | 'CHAGHLNGVYYQGGTYSK' | 'Fibrinogen gamma chain' |
| 3728 | 'FGG_No29' | 'NWIQYKEGFGHLSPTGTTEFWLGNEKIHLISTQSAIPYALR' | 'Fibrinogen gamma chain' |
| 3729 | 'FGG_No30' | 'FEGNCAEQDGSGWWMNK' | 'Fibrinogen gamma chain' |
| 3730 | 'FGG_No31' | 'DNCCILDER' | 'Fibrinogen gamma chain' |
| 3731 | 'FGG_No32' | 'RLDGSVDFK' | 'Fibrinogen gamma chain' |
| 3732 | 'FGG_No33' | 'TSTADYAMFK' | 'Fibrinogen gamma chain' |
| 3733 | 'FGG_No34' | 'MLEEIMK' | 'Fibrinogen gamma chain' |
| 3734 | 'FGG_No35' | 'KMLEEIMK' | 'Fibrinogen gamma chain' |
| 3735 | 'FGG_No36' | 'KMLEEIMK' | 'Fibrinogen gamma chain' |
| 3736 | 'FGG_No37' | 'KMLEEIMK' | 'Fibrinogen gamma chain' |
| 3737 | 'FGG_No38' | 'KMLEEIMKYEASILTHDSSIR' | 'Fibrinogen gamma chain' |
| 3738 | 'FGG_No39' | 'DCQDIANK' | 'Fibrinogen gamma chain' |
| 3739 | 'FGG_No40' | 'YLQEIYNSNNQK' | 'Fibrinogen gamma chain' |
| 3740 | 'FGG_No41' | 'IHLISTQSAIPYALR' | 'Fibrinogen gamma chain' |
| 3741 | 'FGG_No42' | 'KNWIQYK' | 'Fibrinogen gamma chain' |

| | A | B | C |
|---|---|---|---|
| 1 | **Peptide Name** | **Peptide Sequence** | **Peptide Description** |
| 3742 | 'FGG_No43' | 'KMLEEIMK' | 'Fibrinogen gamma chain' |
| 3743 | 'FGG_No44' | 'QVRPEHPAETEYDSLYPEDDL' | 'Fibrinogen gamma chain' |
| 3744 | 'FGG_No45' | 'MLEEIMKYEASILTHDSSIR' | 'Fibrinogen gamma chain' |
| 3745 | 'FGG_No46' | 'FEGNCAEQDGSGWWMNK' | 'Fibrinogen gamma chain' |
| 3746 | 'FGG_No47' | 'KMLEEIMKYEASILTHDSSIR' | 'Fibrinogen gamma chain' |
| 3747 | 'FGG_No48' | 'AIQLTYNPDESSK' | 'Fibrinogen gamma chain' |
| 3748 | 'FGG_No49' | 'NWIQYKEGFGHLSPTGTTEFWLGNEK' | 'Fibrinogen gamma chain' |
| 3749 | 'FGG_No50' | 'MLEEIMKYEASILTHDSSIR' | 'Fibrinogen gamma chain' |
| 3750 | 'FGG_No51' | 'DTVQIHDITGK' | 'Fibrinogen gamma chain' |
| 3751 | 'FGG_No52' | 'DTVQIHDITGKDCQDIANKGAK' | 'Fibrinogen gamma chain' |
| 3752 | 'FGG_No53' | 'PNMIDAATLK' | 'Fibrinogen gamma chain' |
| 3753 | 'FGG_No54' | 'TTMKIIPFNR' | 'Fibrinogen gamma chain' |
| 3754 | 'FGG_No55' | 'AIQLTYNPDESSKPNMIDAATLKSR' | 'Fibrinogen gamma chain' |
| 3755 | 'FGG_No56' | 'DTVQIHDITGKDCQDIANK' | 'Fibrinogen gamma chain' |
| 3756 | 'FGG_No57' | 'EKVAQLEAQCQEPCKDTVQIHDITGK' | 'Fibrinogen gamma chain' |
| 3757 | 'FGG_No58' | 'MLEEIMKYEASILTHDSSIR' | 'Fibrinogen gamma chain' |
| 3758 | 'FGG_No59' | 'QSGLYFIKPLK' | 'Fibrinogen gamma chain' |
| 3759 | 'FGG_No60' | 'LDGSVDFKKNWIQYK' | 'Fibrinogen gamma chain' |
| 3760 | 'FGG_No61' | 'IIPFNRLTIGEGQQHHLGGAK' | 'Fibrinogen gamma chain' |
| 3761 | 'FGG_No62' | 'AIQLTYNPDESSKPNMIDAATLK' | 'Fibrinogen gamma chain' |
| 3762 | 'FGG_No63' | 'EKVAQLEAQCQEPCK' | 'Fibrinogen gamma chain' |
| 3763 | 'FGG_No64' | 'FGSYCPTTCGIADFLSTYQTKVDK' | 'Fibrinogen gamma chain' |
| 3764 | 'FGG_No65' | 'VELEDWNGRTSTADYAMFK' | 'Fibrinogen gamma chain' |
| 3765 | 'FGG_No66' | 'ASTPNGYDNGIIWATWKTR' | 'Fibrinogen gamma chain' |
| 3766 | 'FGG_No67' | 'YLQEIYNSNNQKIVNLK' | 'Fibrinogen gamma chain' |
| 3767 | 'FGG_No68' | 'VDKDLQSLEDILHQVENK' | 'Fibrinogen gamma chain' |
| 3768 | 'FGG_No69' | 'YLQEIYNSNNQKIVNLKEK' | 'Fibrinogen gamma chain' |
| 3769 | 'FGG_No70' | 'TTMKIIPFNRLTIGEGQQHHLGGAK' | 'Fibrinogen gamma chain' |
| 3770 | 'FGG_No71' | 'MLEEIMK' | 'Fibrinogen gamma chain' |
| 3771 | 'FGG_No72' | 'TSTADYAMFKVGPEADKYR' | 'Fibrinogen gamma chain' |
| 3772 | 'FGG_No73' | 'KNWIQYKEGFGHLSPTGTTEFWLGNEK' | 'Fibrinogen gamma chain' |
| 3773 | 'FGG_No74' | 'TTMKIIPFNR' | 'Fibrinogen gamma chain' |
| 3774 | 'FGG_No75' | 'DNCCILDERFGSYCPTTCGIADFLSTYQTK' | 'Fibrinogen gamma chain' |
| 3775 | 'FGG_No76' | 'VELEDWNGRTSTADYAMFK' | 'Fibrinogen gamma chain' |
| 3776 | 'FGG_No77' | 'FFTSHNGMQFSTWDNDNDK' | 'Fibrinogen gamma chain' |
| 3777 | 'FGG_No78' | 'YEASILTHDSSIRYLQEIYNSNNQK' | 'Fibrinogen gamma chain' |
| 3778 | 'FGG_No79' | 'YEASILTHDSSIRYLQEIYNSNNQKIVNLK' | 'Fibrinogen gamma chain' |
| 3779 | 'FGG_No80' | 'TRWYSMK' | 'Fibrinogen gamma chain' |
| 3780 | 'FGG_No81' | 'TSTADYAMFKVGPEADKYR' | 'Fibrinogen gamma chain' |
| 3781 | 'FGG_No82' | 'VDKDLQSLEDILHQVENKTSEVK' | 'Fibrinogen gamma chain' |
| 3782 | 'FGG_No83' | 'LDGSVDFK' | 'Fibrinogen gamma chain' |
| 3783 | 'FGG_No84' | 'FFTSHNGMQFSTWDNDNDK' | 'Fibrinogen gamma chain' |
| 3784 | 'FGG_No85' | 'DCQDIANKGAK' | 'Fibrinogen gamma chain' |
| 3785 | 'FGG_No86' | 'TTMKIIPFNRLTIGEGQQHHLGGAK' | 'Fibrinogen gamma chain' |

| | A | B | C |
|---|---|---|---|
| 1 | **Peptide Name** | **Peptide Sequence** | **Peptide Description** |
| 3786 | 'FGG_No87' | 'AIQLTYNPDESSKPNMIDAATLKSR' | 'Fibrinogen gamma chain' |
| 3787 | 'FGG_No88' | 'QLIKAIQLTYNPDESSKPNMIDAATLK' | 'Fibrinogen gamma chain' |
| 3788 | 'FGG_No89' | 'DLQSLEDILHQVENK' | 'Fibrinogen gamma chain' |
| 3789 | 'FGG_No90' | 'FFTSHNGMQFSTWDNDNDKFEGNCAEQDGSGWWMNK' | 'Fibrinogen gamma chain' |
| 3790 | 'SLC4A1_No1' | 'VLLPLIFR' | 'Band 3 anion transport protein' |
| 3791 | 'SLC4A1_No2' | 'ADFLEQPVLGFVR' | 'Band 3 anion transport protein' |
| 3792 | 'SLC4A1_No3' | 'IPPDSEATLVLVGR' | 'Band 3 anion transport protein' |
| 3793 | 'SLC4A1_No4' | 'HSHAGELEALGGVKPAVLTR' | 'Band 3 anion transport protein' |
| 3794 | 'SLC4A1_No5' | 'LQEAAELEAVELPVPIR' | 'Band 3 anion transport protein' |
| 3795 | 'SLC4A1_No6' | 'RYQSSPAKPDSSFYK' | 'Band 3 anion transport protein' |
| 3796 | 'SLC4A1_No7' | 'NVELQCLDADDAK' | 'Band 3 anion transport protein' |
| 3797 | 'SLC4A1_No8' | 'ATFDEEEGRDEYDEVAMPV' | 'Band 3 anion transport protein' |
| 3798 | 'SLC4A1_No9' | 'WVQLEENLGENGAWGR' | 'Band 3 anion transport protein' |
| 3799 | 'SLC4A1_No10' | 'YHPDVPYVK' | 'Band 3 anion transport protein' |
| 3800 | 'SLC4A1_No11' | 'YQSSPAKPDSSFYK' | 'Band 3 anion transport protein' |
| 3801 | 'SLC4A1_No12' | 'GWVIHPLGLR' | 'Band 3 anion transport protein' |
| 3802 | 'SLC4A1_No13' | 'VYVELQELVMDEKNQELR' | 'Band 3 anion transport protein' |
| 3803 | 'SLC4A1_No14' | 'VYVELQELVMDEKNQELR' | 'Band 3 anion transport protein' |
| 3804 | 'SLC4A1_No15' | 'VYVELQELVMDEK' | 'Band 3 anion transport protein' |
| 3805 | 'SLC4A1_No16' | 'FIFEDQIRPQDR' | 'Band 3 anion transport protein' |
| 3806 | 'CRP_No1' | 'YEVQGEVFTKPQLWP' | 'C-reactive protein' |
| 3807 | 'CRP_No2' | 'RQDNEILIFWSK' | 'C-reactive protein' |
| 3808 | 'CRP_No3' | 'QDNEILIFWSK' | 'C-reactive protein' |
| 3809 | 'CRP_No4' | 'ESDTSYVSLK' | 'C-reactive protein' |
| 3810 | 'CRP_No5' | 'ALKYEVQGEVFTKPQLWP' | 'C-reactive protein' |
| 3811 | 'CRP_No6' | 'ESDTSYVSLKAPLTKPLK' | 'C-reactive protein' |
| 3812 | 'CRP_No7' | 'GYSIFSYATKR' | 'C-reactive protein' |
| 3813 | 'CRP_No8' | 'APLTKPLK' | 'C-reactive protein' |
| 3814 | 'CRP_No9' | 'GYSIFSYATK' | 'C-reactive protein' |
| 3815 | 'APCS_No1' | 'GYVIIKPLVWV' | 'Serum amyloid P-component' |
| 3816 | 'APCS_No2' | 'IVLGQEQDSYGGKFDR' | 'Serum amyloid P-component' |
| 3817 | 'APCS_No3' | 'IVLGQEQDSYGGK' | 'Serum amyloid P-component' |
| 3818 | 'APCS_No4' | 'DNELLVYKER' | 'Serum amyloid P-component' |
| 3819 | 'APCS_No5' | 'DNELLVYK' | 'Serum amyloid P-component' |
| 3820 | 'APCS_No6' | 'QGYFVEAQPK' | 'Serum amyloid P-component' |
| 3821 | 'APCS_No7' | 'AYSLFSYNTQGRDNELLVYK' | 'Serum amyloid P-component' |
| 3822 | 'APCS_No8' | 'AYSLFSYNTQGRDNELLVYKER' | 'Serum amyloid P-component' |
| 3823 | 'APCS_No9' | 'AYSDLSR' | 'Serum amyloid P-component' |
| 3824 | 'APCS_No10' | 'VGEYSLYIGR' | 'Serum amyloid P-component' |
| 3825 | 'APCS_No11' | 'ERVGEYSLYIGR' | 'Serum amyloid P-component' |
| 3826 | 'APCS_No12' | 'AYSLFSYNTQGR' | 'Serum amyloid P-component' |
| 3827 | 'APCS_No13' | 'VGEYSLYIGRHKVTSK' | 'Serum amyloid P-component' |
| 3828 | 'APCS_No14' | 'ESVTDHVNLITPLEK' | 'Serum amyloid P-component' |
| 3829 | 'APCS_No15' | 'QGYFVEAQPK' | 'Serum amyloid P-component' |

| | A | B | C |
|---|---|---|---|
| 1 | **Peptide Name** | **Peptide Sequence** | **Peptide Description** |
| 3830 | 'APCS_No16' | 'DNELLVYKERVGEYSLYIGR' | 'Serum amyloid P-component' |
| 3831 | 'C1QA_No1' | 'KGHIYQGSEADSVFSGFLIFPSA' | 'Complement C1q subcomponent subunit A' |
| 3832 | 'C1QA_No2' | 'GLFQVVSGGMVLQLQQGDQVWVEKDPK' | 'Complement C1q subcomponent subunit A' |
| 3833 | 'C1QA_No3' | 'GLFQVVSGGMVLQLQQGDQVWVEKDPKK' | 'Complement C1q subcomponent subunit A' |
| 3834 | 'C1QA_No4' | 'SLGFCDTTNK' | 'Complement C1q subcomponent subunit A' |
| 3835 | 'C1QA_No5' | 'GLFQVVSGGMVLQLQQGDQVWVEKDPKK' | 'Complement C1q subcomponent subunit A' |
| 3836 | 'C1QA_No6' | 'DQPRPAFSAIR' | 'Complement C1q subcomponent subunit A' |
| 3837 | 'C1QA_No7' | 'GLFQVVSGGMVLQLQQGDQVWVEKDPK' | 'Complement C1q subcomponent subunit A' |
| 3838 | 'C1QA_No8' | 'GHIYQGSEADSVFSGFLIFPSA' | 'Complement C1q subcomponent subunit A' |
| 3839 | 'C1QA_No9' | 'RSLGFCDTTNK' | 'Complement C1q subcomponent subunit A' |
| 3840 | 'C1QA_No10' | 'DQPRPAFSAIRR' | 'Complement C1q subcomponent subunit A' |
| 3841 | 'C1QA_No11' | 'GLFQVVSGGMVLQLQQGDQVWVEK' | 'Complement C1q subcomponent subunit A' |
| 3842 | 'C1QA_No12' | 'NPPMGGNVVIFDTVITNQEEPYQNHSGR' | 'Complement C1q subcomponent subunit A' |
| 3843 | 'C1QA_No13' | 'SLGFCDTTNKGLFQVVSGGMVLQLQQGDQVWVEKDPK' | 'Complement C1q subcomponent subunit A' |
| 3844 | 'C1QA_No14' | 'NPPMGGNVVIFDTVITNQEEPYQNHSGR' | 'Complement C1q subcomponent subunit A' |
| 3845 | 'C1QB_No1' | 'LEQGENVFLQATDK' | 'Complement C1q subcomponent subunit B' |
| 3846 | 'C1QB_No2' | 'VPGLYYFTYHASSR' | 'Complement C1q subcomponent subunit B' |
| 3847 | 'C1QB_No3' | 'VVTFCDYAYNTFQVTTGGMVLK' | 'Complement C1q subcomponent subunit B' |
| 3848 | 'C1QB_No4' | 'TINVPLRR' | 'Complement C1q subcomponent subunit B' |
| 3849 | 'C1QB_No5' | 'FDHVITNMNNNYEPR' | 'Complement C1q subcomponent subunit B' |
| 3850 | 'C1QB_No6' | 'FDHVITNMNNNYEPR' | 'Complement C1q subcomponent subunit B' |
| 3851 | 'C1QB_No7' | 'VVTFCDYAYNTFQVTTGGMVLK' | 'Complement C1q subcomponent subunit B' |
| 3852 | 'C1QB_No8' | 'GNLCVNLMR' | 'Complement C1q subcomponent subunit B' |
| 3853 | 'C1QB_No9' | 'GNLCVNLMR' | 'Complement C1q subcomponent subunit B' |
| 3854 | 'C1QB_No10' | 'DQTIRFDHVITNMNNNYEPR' | 'Complement C1q subcomponent subunit B' |
| 3855 | 'C1QB_No11' | 'DQTIRFDHVITNMNNNYEPR' | 'Complement C1q subcomponent subunit B' |
| 3856 | 'C1QB_No12' | 'FTCKVPGLYYFTYHASSR' | 'Complement C1q subcomponent subunit B' |
| 3857 | 'C1QC_No1' | 'FNAVLTNPQGDYDTSTGK' | 'Complement C1q subcomponent subunit C' |
| 3858 | 'C1QC_No2' | 'QTHQPPAPNSLIR' | 'Complement C1q subcomponent subunit C' |
| 3859 | 'C1QC_No3' | 'TNQVNSGGVLLR' | 'Complement C1q subcomponent subunit C' |
| 3860 | 'C1QC_No4' | 'QKFQSVFTVTR' | 'Complement C1q subcomponent subunit C' |
| 3861 | 'C1QC_No5' | 'QKFQSVFTVTR' | 'Complement C1q subcomponent subunit C' |
| 3862 | 'C1QC_No6' | 'VVTFCGHTSK' | 'Complement C1q subcomponent subunit C' |
| 3863 | 'C1QC_No7' | 'FQSVFTVTR' | 'Complement C1q subcomponent subunit C' |
| 3864 | 'C1QC_No8' | 'QTHQPPAPNSLIR' | 'Complement C1q subcomponent subunit C' |
| 3865 | 'C1QC_No9' | 'SGVKVVTFCGHTSK' | 'Complement C1q subcomponent subunit C' |
| 3866 | 'C1QC_No10' | 'VVTFCGHTSKTNQVNSGGVLLR' | 'Complement C1q subcomponent subunit C' |
| 3867 | 'C9_No1' | 'FTPTETNKAEQCCEETASSISLHGK' | 'Complement component C9' |
| 3868 | 'C9_No2' | 'AEQCCEETASSISLHGK' | 'Complement component C9' |
| 3869 | 'C9_No3' | 'DVVLTTTFVDDIK' | 'Complement component C9' |
| 3870 | 'C9_No4' | 'LSPIYNLVPVK' | 'Complement component C9' |
| 3871 | 'C9_No5' | 'TEHYEEQIEAFK' | 'Complement component C9' |
| 3872 | 'C9_No6' | 'RPWNVASLIYETK' | 'Complement component C9' |
| 3873 | 'C9_No7' | 'VVEESELAR' | 'Complement component C9' |

| | A | B | C |
|---|---|---|---|
| 1 | **Peptide Name** | **Peptide Sequence** | **Peptide Description** |
| 3874 | 'C9_No8' | 'DRVVEESELAR' | 'Complement component C9' |
| 3875 | 'C9_No9' | 'AIEDYINEFSVRK' | 'Complement component C9' |
| 3876 | 'C9_No10' | 'CLCACPFK' | 'Complement component C9' |
| 3877 | 'C9_No11' | 'ISEGLPALEFPNEK' | 'Complement component C9' |
| 3878 | 'C9_No12' | 'SIEVFGQFNGKR' | 'Complement component C9' |
| 3879 | 'C9_No13' | 'YAFELKEK' | 'Complement component C9' |
| 3880 | 'C9_No14' | 'FEGIACEISK' | 'Complement component C9' |
| 3881 | 'C9_No15' | 'DGNTLTYYR' | 'Complement component C9' |
| 3882 | 'C9_No16' | 'CHTCQNGGTVILMDGK' | 'Complement component C9' |
| 3883 | 'C9_No17' | 'DRDGNTLTYYR' | 'Complement component C9' |
| 3884 | 'C9_No18' | 'KCHTCQNGGTVILMDGK' | 'Complement component C9' |
| 3885 | 'C9_No19' | 'NFRTEHYEEQIEAFK' | 'Complement component C9' |
| 3886 | 'C9_No20' | 'TSNFNAAISLK' | 'Complement component C9' |
| 3887 | 'C9_No21' | 'QCVPTEPCEDAEDDCGNDFQCSTGR' | 'Complement component C9' |
| 3888 | 'C9_No22' | 'AIEDYINEFSVR' | 'Complement component C9' |
| 3889 | 'C9_No23' | 'LRCNGDNDCGDFSDEDDCESEPRPPCR' | 'Complement component C9' |
| 3890 | 'C9_No24' | 'CLCACPFKFEGIACEISK' | 'Complement component C9' |
| 3891 | 'C9_No25' | 'TEHYEEQIEAFKSIIQEK' | 'Complement component C9' |
| 3892 | 'C9_No26' | 'NRDVVLTTTFVDDIK' | 'Complement component C9' |
| 3893 | 'C9_No27' | 'CNGDNDCGDFSDEDDCESEPRPPCRDR' | 'Complement component C9' |
| 3894 | 'C9_No28' | 'GTVIDVTDFVNWASSINDAPVLISQK' | 'Complement component C9' |
| 3895 | 'C9_No29' | 'RPWNVASLIYETKGEK' | 'Complement component C9' |
| 3896 | 'C9_No30' | 'QCVPTEPCEDAEDDCGNDFQCSTGR' | 'Complement component C9' |
| 3897 | 'C9_No31' | 'SRSIEVFGQFNGKR' | 'Complement component C9' |
| 3898 | 'C9_No32' | 'KCHTCQNGGTVILMDGK' | 'Complement component C9' |
| 3899 | 'C9_No33' | 'GEKNFRTEHYEEQIEAFK' | 'Complement component C9' |
| 3900 | 'C9_No34' | 'NFRTEHYEEQIEAFKSIIQEK' | 'Complement component C9' |
| 3901 | 'C9_No35' | 'CLCACPFKFEGIACEISKQK' | 'Complement component C9' |
| 3902 | 'C9_No36' | 'CHTCQNGGTVILMDGK' | 'Complement component C9' |
| 3903 | 'C9_No37' | 'FTPTETNK' | 'Complement component C9' |
| 3904 | 'C9_No38' | 'SIEVFGQFNGK' | 'Complement component C9' |
| 3905 | 'C9_No39' | 'RQCVPTEPCEDAEDDCGNDFQCSTGR' | 'Complement component C9' |
| 3906 | 'C9_No40' | 'TAGYGINILGMDPLSTPFDNEFYNGLCNR' | 'Complement component C9' |
| 3907 | 'C9_No41' | 'TAGYGINILGMDPLSTPFDNEFYNGLCNRDR' | 'Complement component C9' |
| 3908 | 'C9_No42' | 'CNGDNDCGDFSDEDDCESEPRPPCR' | 'Complement component C9' |
| 3909 | 'C9_No43' | 'MSPWSEWSQCDPCLR' | 'Complement component C9' |
| 3910 | 'C9_No44' | 'QKISEGLPALEFPNEK' | 'Complement component C9' |
| 3911 | 'C9_No45' | 'FTPTETNKAEQCCEETASSISLHGKGSFR' | 'Complement component C9' |
| 3912 | 'C9_No46' | 'AEQCCEETASSISLHGKGSFR' | 'Complement component C9' |
| 3913 | 'C9_No47' | 'DVVLTTTFVDDIKALPTTYEK' | 'Complement component C9' |
| 3914 | 'C9_No48' | 'CTDAVGDRR' | 'Complement component C9' |
| 3915 | 'C9_No49' | 'SIIQEKTSNFNAAISLK' | 'Complement component C9' |
| 3916 | 'C9_No50' | 'TSNFNAAISLKFTPTETNKAEQCCEETASSISLHGK' | 'Complement component C9' |
| 3917 | 'APOH_No1' | 'VCPFAGILENGAVR' | 'Beta-2-glycoprotein 1' |

| | A | B | C |
|---|---|---|---|
| 1 | **Peptide Name** | **Peptide Sequence** | **Peptide Description** |
| 3918 | 'APOH_No2' | 'KFICPLTGLWPINTLK' | 'Beta-2-glycoprotein 1' |
| 3919 | 'APOH_No3' | 'ATFGCHDGYSLDGPEEIECTK' | 'Beta-2-glycoprotein 1' |
| 3920 | 'APOH_No4' | 'CPFPSRPDNGFVNYPAKPTLYYK' | 'Beta-2-glycoprotein 1' |
| 3921 | 'APOH_No5' | 'CSYTEDAQCIDGTIEVPK' | 'Beta-2-glycoprotein 1' |
| 3922 | 'APOH_No6' | 'TCPKPDDLPFSTVVPLK' | 'Beta-2-glycoprotein 1' |
| 3923 | 'APOH_No7' | 'TFYEPGEEITYSCK' | 'Beta-2-glycoprotein 1' |
| 3924 | 'APOH_No8' | 'ATVVYQGER' | 'Beta-2-glycoprotein 1' |
| 3925 | 'APOH_No9' | 'YTTFEYPNTISFSCNTGFYLNGADSAK' | 'Beta-2-glycoprotein 1' |
| 3926 | 'APOH_No10' | 'TFYEPGEEITYSCKPGYVSR' | 'Beta-2-glycoprotein 1' |
| 3927 | 'APOH_No11' | 'KCSYTEDAQCIDGTIEVPK' | 'Beta-2-glycoprotein 1' |
| 3928 | 'APOH_No12' | 'EHSSLAFWK' | 'Beta-2-glycoprotein 1' |
| 3929 | 'APOH_No13' | 'NGMLHGDKVSFFCK' | 'Beta-2-glycoprotein 1' |
| 3930 | 'APOH_No14' | 'TDASDVKPC' | 'Beta-2-glycoprotein 1' |
| 3931 | 'APOH_No15' | 'FKNGMLHGDKVSFFCK' | 'Beta-2-glycoprotein 1' |
| 3932 | 'APOH_No16' | 'CPFPSRPDNGFVNYPAK' | 'Beta-2-glycoprotein 1' |
| 3933 | 'APOH_No17' | 'PDDLPFSTVVPLK' | 'Beta-2-glycoprotein 1' |
| 3934 | 'APOH_No18' | 'CFKEHSSLAFWK' | 'Beta-2-glycoprotein 1' |
| 3935 | 'APOH_No19' | 'CTEEGKWSPELPVCAPIICPPPSIPTFATLR' | 'Beta-2-glycoprotein 1' |
| 3936 | 'APOH_No20' | 'NGMLHGDKVSFFCK' | 'Beta-2-glycoprotein 1' |
| 3937 | 'APOH_No21' | 'FICPLTGLWPINTLK' | 'Beta-2-glycoprotein 1' |
| 3938 | 'APOH_No22' | 'DKATFGCHDGYSLDGPEEIECTK' | 'Beta-2-glycoprotein 1' |
| 3939 | 'APOH_No23' | 'FKNGMLHGDKVSFFCK' | 'Beta-2-glycoprotein 1' |
| 3940 | 'APOH_No24' | 'LGNWSAMPSCK' | 'Beta-2-glycoprotein 1' |
| 3941 | 'APOH_No25' | 'KATVVYQGER' | 'Beta-2-glycoprotein 1' |
| 3942 | 'APOH_No26' | 'KCSYTEDAQCIDGTIEVPKCFK' | 'Beta-2-glycoprotein 1' |
| 3943 | 'APOH_No27' | 'LGNWSAMPSCK' | 'Beta-2-glycoprotein 1' |
| 3944 | 'APOH_No28' | 'PDNGFVNYPAKPTLYYK' | 'Beta-2-glycoprotein 1' |
| 3945 | 'APOH_No29' | 'VYKPSAGNNSLYR' | 'Beta-2-glycoprotein 1' |
| 3946 | 'APOH_No30' | 'CSYTEDAQCIDGTIEVPKCFKEHSSLAFWK' | 'Beta-2-glycoprotein 1' |
| 3947 | 'APOH_No31' | 'CSYTEDAQCIDGTIEVPKCFK' | 'Beta-2-glycoprotein 1' |
| 3948 | 'APOH_No32' | 'EHSSLAFWKTDASDVKPC' | 'Beta-2-glycoprotein 1' |
| 3949 | 'APOH_No33' | 'VPVKKATVVYQGER' | 'Beta-2-glycoprotein 1' |
| 3950 | 'APOH_No34' | 'TCPKPDDLPFSTVVPLKTFYEPGEEITYSCK' | 'Beta-2-glycoprotein 1' |
| 3951 | 'APOH_No35' | 'ATVVYQGERVKIQEK' | 'Beta-2-glycoprotein 1' |
| 3952 | 'APOH_No36' | 'NGMLHGDK' | 'Beta-2-glycoprotein 1' |
| 3953 | 'APOH_No37' | 'ASCKVPVK' | 'Beta-2-glycoprotein 1' |
| 3954 | 'LRG1_No1' | 'ENQLEVLEVSWLHGLK' | 'Leucine-rich alpha-2-glycoprotein' |
| 3955 | 'LRG1_No2' | 'TLDLGENQLETLPPDLLR' | 'Leucine-rich alpha-2-glycoprotein' |
| 3956 | 'LRG1_No3' | 'VAAGAFQGLR' | 'Leucine-rich alpha-2-glycoprotein' |
| 3957 | 'LRG1_No4' | 'YLFLNGNK' | 'Leucine-rich alpha-2-glycoprotein' |
| 3958 | 'LRG1_No5' | 'DGFDISGNPWICDQNLSDLYR' | 'Leucine-rich alpha-2-glycoprotein' |
| 3959 | 'LRG1_No6' | 'ALGHLDLSGNR' | 'Leucine-rich alpha-2-glycoprotein' |
| 3960 | 'LRG1_No7' | 'LHLEGNKLQVLGKDLLLPQPDLR' | 'Leucine-rich alpha-2-glycoprotein' |
| 3961 | 'LRG1_No8' | 'CAGPEAVKGQTLLAVAK' | 'Leucine-rich alpha-2-glycoprotein' |

| | A | B | C |
|---|---|---|---|
| 1 | **Peptide Name** | **Peptide Sequence** | **Peptide Description** |
| 3962 | 'LRG1_No9' | 'TLDLGENQLETLPPDLLRGPLQLER' | 'Leucine-rich alpha-2-glycoprotein' |
| 3963 | 'LRG1_No10' | 'LHLEGNKLQVLGK' | 'Leucine-rich alpha-2-glycoprotein' |
| 3964 | 'LRG1_No11' | 'NALTGLPPGLFQASATLDTLVLK' | 'Leucine-rich alpha-2-glycoprotein' |
| 3965 | 'LRG1_No12' | 'DLLLPQPDLR' | 'Leucine-rich alpha-2-glycoprotein' |
| 3966 | 'LRG1_No13' | 'LQELHLSSNGLESLSPEFLRPVPQLR' | 'Leucine-rich alpha-2-glycoprotein' |
| 3967 | 'LRG1_No14' | 'ALGHLDLSGNRLR' | 'Leucine-rich alpha-2-glycoprotein' |
| 3968 | 'LRG1_No15' | 'LQVLGKDLLLPQPDLR' | 'Leucine-rich alpha-2-glycoprotein' |
| 3969 | 'LRG1_No16' | 'LQELHLSSNGLESLSPEFLR' | 'Leucine-rich alpha-2-glycoprotein' |
| 3970 | 'LRG1_No17' | 'CAGPEAVK' | 'Leucine-rich alpha-2-glycoprotein' |
| 3971 | 'LRG1_No18' | 'LHLEGNK' | 'Leucine-rich alpha-2-glycoprotein' |
| 3972 | 'LRG1_No19' | 'GPLQLER' | 'Leucine-rich alpha-2-glycoprotein' |
| 3973 | 'FN1_No1' | 'FLATTPNSLLVSWQPPR' | 'Fibronectin' |
| 3974 | 'FN1_No2' | 'LLCQCLGFGSGHFR' | 'Fibronectin' |
| 3975 | 'FN1_No3' | 'EESPLLIGQQSTVSDVPR' | 'Fibronectin' |
| 3976 | 'FN1_No4' | 'TKTETITGFQVDAVPANGQTPIQR' | 'Fibronectin' |
| 3977 | 'FN1_No5' | 'PRPGVTEATITGLEPGTEYTIYVIALK' | 'Fibronectin' |
| 3978 | 'FN1_No6' | 'VPGTSTSATLTGLTR' | 'Fibronectin' |
| 3979 | 'FN1_No7' | 'EYLGAICSCTCFGGQR' | 'Fibronectin' |
| 3980 | 'FN1_No8' | 'DLQFVEVTDVK' | 'Fibronectin' |
| 3981 | 'FN1_No9' | 'TEIDKPSQMQVTDVQDNSISVK' | 'Fibronectin' |
| 3982 | 'FN1_No10' | 'TGLDSPTGIDFSDITANSFTVHWIAPR' | 'Fibronectin' |
| 3983 | 'FN1_No11' | 'NLQPASEYTVSLVAIK' | 'Fibronectin' |
| 3984 | 'FN1_No12' | 'GDSPASSKPISINYR' | 'Fibronectin' |
| 3985 | 'FN1_No13' | 'VDVIPVNLPGEHGQR' | 'Fibronectin' |
| 3986 | 'FN1_No14' | 'SYTITGLQPGTDYK' | 'Fibronectin' |
| 3987 | 'FN1_No15' | 'WSRPQAPITGYR' | 'Fibronectin' |
| 3988 | 'FN1_No16' | 'TFYSCTTEGR' | 'Fibronectin' |
| 3989 | 'FN1_No17' | 'TYLGNALVCTCYGGSR' | 'Fibronectin' |
| 3990 | 'FN1_No18' | 'GFNCESKPEAEETCFDKYTGNTYR' | 'Fibronectin' |
| 3991 | 'FN1_No19' | 'TEIDKPSQMQVTDVQDNSISVK' | 'Fibronectin' |
| 3992 | 'FN1_No20' | 'FGFCPMAAHEEICTTNEGVMYR' | 'Fibronectin' |
| 3993 | 'FN1_No21' | 'FGFCPMAAHEEICTTNEGVMYR' | 'Fibronectin' |
| 3994 | 'FN1_No22' | 'YQCYCYGR' | 'Fibronectin' |
| 3995 | 'FN1_No23' | 'FGFCPMAAHEEICTTNEGVMYR' | 'Fibronectin' |
| 3996 | 'FN1_No24' | 'TPFVTHPGYDTGNGIQLPGTSGQQPSVGQQMIFEEHGFRR' | 'Fibronectin' |
| 3997 | 'FN1_No25' | 'QAQQMVQPQSPVAVSQSKPGCYDNGK' | 'Fibronectin' |
| 3998 | 'FN1_No26' | 'HTSVQTTSSGSGPFTDVR' | 'Fibronectin' |
| 3999 | 'FN1_No27' | 'IYLYTLNDNAR' | 'Fibronectin' |
| 4000 | 'FN1_No28' | 'AAVYQPQPHPQPPPYGHCVTDSGVVYSVGMQWLK' | 'Fibronectin' |
| 4001 | 'FN1_No29' | 'LGVRPSQGGEAPR' | 'Fibronectin' |
| 4002 | 'FN1_No30' | 'QGENGQMMSCTCLGNGK' | 'Fibronectin' |
| 4003 | 'FN1_No31' | 'YSFCTDHTVLVQTR' | 'Fibronectin' |
| 4004 | 'FN1_No32' | 'QGENGQMMSCTCLGNGK' | 'Fibronectin' |
| 4005 | 'FN1_No33' | 'QGENGQMMSCTCLGNGK' | 'Fibronectin' |

| | A | B | C |
|---|---|---|---|
| 1 | **Peptide Name** | **Peptide Sequence** | **Peptide Description** |
| 4006 | 'FN1_No34' | 'QAQQMVQPQSPVAVSQSK' | 'Fibronectin' |
| 4007 | 'FN1_No35' | 'QYNVGPSVSK' | 'Fibronectin' |
| 4008 | 'FN1_No36' | 'EINLAPDSSSVVVSGLMVATK' | 'Fibronectin' |
| 4009 | 'FN1_No37' | 'EATIPGHLNSYTIK' | 'Fibronectin' |
| 4010 | 'FN1_No38' | 'GFNCESKPEAEETCFDK' | 'Fibronectin' |
| 4011 | 'FN1_No39' | 'QKTGLDSPTGIDFSDITANSFTVHWIAPR' | 'Fibronectin' |
| 4012 | 'FN1_No40' | 'QYNVGPSVSKYPLR' | 'Fibronectin' |
| 4013 | 'FN1_No41' | 'DSMIWDCTCIGAGR' | 'Fibronectin' |
| 4014 | 'FN1_No42' | 'FTNIGPDTMR' | 'Fibronectin' |
| 4015 | 'FN1_No43' | 'IGDQWDKQHDMGHMMR' | 'Fibronectin' |
| 4016 | 'FN1_No44' | 'IGDQWDKQHDMGHMMR' | 'Fibronectin' |
| 4017 | 'FN1_No45' | 'FGFCPMAAHEEICTTNEGVMYR' | 'Fibronectin' |
| 4018 | 'FN1_No46' | 'HYQINQQWER' | 'Fibronectin' |
| 4019 | 'FN1_No47' | 'YEVSVYALK' | 'Fibronectin' |
| 4020 | 'FN1_No48' | 'PAQGVVTTLENVSPPR' | 'Fibronectin' |
| 4021 | 'FN1_No49' | 'GEWTCIAYSQLR' | 'Fibronectin' |
| 4022 | 'FN1_No50' | 'QYNVGPSVSKYPLR' | 'Fibronectin' |
| 4023 | 'FN1_No51' | 'TYHVGEQWQK' | 'Fibronectin' |
| 4024 | 'FN1_No52' | 'VFAVSHGR' | 'Fibronectin' |
| 4025 | 'FN1_No53' | 'STATISGLKPGVDYTITVYAVTGR' | 'Fibronectin' |
| 4026 | 'FN1_No54' | 'QAQQMVQPQSPVAVSQSKPGCYDNGK' | 'Fibronectin' |
| 4027 | 'FN1_No55' | 'WCHDNGVNYK' | 'Fibronectin' |
| 4028 | 'FN1_No56' | 'PAQGVVTTLENVSPPRR' | 'Fibronectin' |
| 4029 | 'FN1_No57' | 'PISINYR' | 'Fibronectin' |
| 4030 | 'FN1_No58' | 'ITGYIIK' | 'Fibronectin' |
| 4031 | 'FN1_No59' | 'VTIMWTPPESAVTGYRVDVIPVNLPGEHGQR' | 'Fibronectin' |
| 4032 | 'FN1_No60' | 'GEWTCKPIAEK' | 'Fibronectin' |
| 4033 | 'FN1_No61' | 'GNLLQCICTGNGR' | 'Fibronectin' |
| 4034 | 'FN1_No62' | 'NTFAEVTGLSPGVTYYFKVFAVSHGR' | 'Fibronectin' |
| 4035 | 'FN1_No63' | 'YEVSVYALKDTLTSR' | 'Fibronectin' |
| 4036 | 'FN1_No64' | 'WCGTTQNYDADQK' | 'Fibronectin' |
| 4037 | 'FN1_No65' | 'QGENGQMMSCTCLGNGK' | 'Fibronectin' |
| 4038 | 'FN1_No66' | 'QDGHLWCSTTSNYEQDQKYSFCTDHTVLVQTR' | 'Fibronectin' |
| 4039 | 'FN1_No67' | 'SSPVVIDASTAIDAPSNLR' | 'Fibronectin' |
| 4040 | 'FN1_No68' | 'RPHETGGYMLECVCLGNGK' | 'Fibronectin' |
| 4041 | 'FN1_No69' | 'VGDTYERPKDSMIWDCTCIGAGR' | 'Fibronectin' |
| 4042 | 'FN1_No70' | 'NLQPASEYTVSLVAIKGNQESPK' | 'Fibronectin' |
| 4043 | 'FN1_No71' | 'IGDQWDKQHDMGHMMR' | 'Fibronectin' |
| 4044 | 'FN1_No72' | 'IGDQWDKQHDMGHMMR' | 'Fibronectin' |
| 4045 | 'FN1_No73' | 'CFDHAAGTSYVVGETWEKPYQGWMMVDCTCLGEGSGR' | 'Fibronectin' |
| 4046 | 'FN1_No74' | 'KKTDELPQLVTLPHPNLHGPEILDVPSTVQK' | 'Fibronectin' |
| 4047 | 'FN1_No75' | 'WLPSSSPVTGYR' | 'Fibronectin' |
| 4048 | 'FN1_No76' | 'VTIMWTPPESAVTGYR' | 'Fibronectin' |
| 4049 | 'FN1_No77' | 'IGDQWDKQHDMGHMMR' | 'Fibronectin' |

| | A | B | C |
|---|---|---|---|
| 1 | **Peptide Name** | **Peptide Sequence** | **Peptide Description** |
| 4050 | 'FN1_No78' | 'WKEATIPGHLNSYTIK' | 'Fibronectin' |
| 4051 | 'FN1_No79' | 'KTDELPQLVTLPHPNLHGPEILDVPSTVQK' | 'Fibronectin' |
| 4052 | 'FN1_No80' | 'QGENGQMMSCTCLGNGKGEFK' | 'Fibronectin' |
| 4053 | 'FN1_No81' | 'PEAEETCFDK' | 'Fibronectin' |
| 4054 | 'FN1_No82' | 'TDELPQLVTLPHPNLHGPEILDVPSTVQK' | 'Fibronectin' |
| 4055 | 'FN1_No83' | 'IGDQWDKQHDMGHMMR' | 'Fibronectin' |
| 4056 | 'FN1_No84' | 'VVTPLSPPTNLHLEANPDTGVLTVSWER' | 'Fibronectin' |
| 4057 | 'FN1_No85' | 'SYTITGLQPGTDYKIYLYTLNDNAR' | 'Fibronectin' |
| 4058 | 'FN1_No86' | 'DNMKWCGTTQNYDADQK' | 'Fibronectin' |
| 4059 | 'FN1_No87' | 'VGDTYERPK' | 'Fibronectin' |
| 4060 | 'FN1_No88' | 'VTWAPPPSIDLTNFLVR' | 'Fibronectin' |
| 4061 | 'FN1_No89' | 'GNLLQCICTGNGRGEWK' | 'Fibronectin' |
| 4062 | 'FN1_No90' | 'VTIMWTPPESAVTGYR' | 'Fibronectin' |
| 4063 | 'FN1_No91' | 'MSESGFKLLCQCLGFGSGHFR' | 'Fibronectin' |
| 4064 | 'FN1_No92' | 'IGDQWDKQHDMGHMMR' | 'Fibronectin' |
| 4065 | 'FN1_No93' | 'YEVSVYALKDTLTSRPAQGVVTTLENVSPPR' | 'Fibronectin' |
| 4066 | 'FN1_No94' | 'TIKPDVR' | 'Fibronectin' |
| 4067 | 'FN1_No95' | 'DSMIWDCTCIGAGR' | 'Fibronectin' |
| 4068 | 'FN1_No96' | 'RPHETGGYMLECVCLGNGK' | 'Fibronectin' |
| 4069 | 'FN1_No97' | 'QGENGQMMSCTCLGNGK' | 'Fibronectin' |
| 4070 | 'FN1_No98' | 'TFYSCTTEGRQDGHLWCSTTSNYEQDQKYSFCTDHTVLVQTR' | 'Fibronectin' |
| 4071 | 'FN1_No99' | 'IGDQWDKQHDMGHMMR' | 'Fibronectin' |
| 4072 | 'FN1_No100' | 'WKCDPVDQCQDSETGTFYQIGDSWEKYVHGVR' | 'Fibronectin' |
| 4073 | 'FN1_No101' | 'NTFAEVTGLSPGVTYYFK' | 'Fibronectin' |
| 4074 | 'FN1_No102' | 'GLKPGVVYEGQLISIQQYGHQEVTR' | 'Fibronectin' |
| 4075 | 'FN1_No103' | 'QGENGQMMSCTCLGNGKGEFK' | 'Fibronectin' |
| 4076 | 'FN1_No104' | 'TNTNVNCPIECFMPLDVQADR' | 'Fibronectin' |
| 4077 | 'FN1_No105' | 'FTNIGPDTMR' | 'Fibronectin' |
| 4078 | 'FN1_No106' | 'DTLTSRPAQGVVTTLENVSPPRR' | 'Fibronectin' |
| 4079 | 'FN1_No107' | 'ITYGETGGNSPVQEFTVPGSK' | 'Fibronectin' |
| 4080 | 'FN1_No108' | 'QAQQMVQPQSPVAVSQSK' | 'Fibronectin' |
| 4081 | 'FN1_No109' | 'QGENGQMMSCTCLGNGK' | 'Fibronectin' |
| 4082 | 'FN1_No110' | 'EINLAPDSSSVVVSGLMVATK' | 'Fibronectin' |
| 4083 | 'FN1_No111' | 'RPGGEPSPEGTTGQSYNQYSQR' | 'Fibronectin' |
| 4084 | 'FN1_No112' | 'GATYNVIVEALKDQQR' | 'Fibronectin' |
| 4085 | 'FN1_No113' | 'DTLTSRPAQGVVTTLENVSPPR' | 'Fibronectin' |
| 4086 | 'FN1_No114' | 'CFDHAAGTSYVVGETWEKPYQGWMMVDCTCLGEGSGR' | 'Fibronectin' |
| 4087 | 'FN1_No115' | 'ITGYIIKYEKPGSPPR' | 'Fibronectin' |
| 4088 | 'FN1_No116' | 'CFDHAAGTSYVVGETWEKPYQGWMMVDCTCLGEGSGR' | 'Fibronectin' |
| 4089 | 'FN1_No117' | 'TETITGFQVDAVPANGQTPIQR' | 'Fibronectin' |
| 4090 | 'FN1_No118' | 'QDGHLWCSTTSNYEQDQK' | 'Fibronectin' |
| 4091 | 'FN1_No119' | 'QAQQMVQPQSPVAVSQSK' | 'Fibronectin' |
| 4092 | 'FN1_No120' | 'ISCTIANR' | 'Fibronectin' |
| 4093 | 'FN1_No121' | 'AAVYQPQPHPQPPPYGHCVTDSGVVYSVGMQWLK' | 'Fibronectin' |

| | A | B | C |
|---|---|---|---|
| 1 | **Peptide Name** | **Peptide Sequence** | **Peptide Description** |
| 4094 | 'FN1_No122' | 'YEKPGSPPREVVPR' | 'Fibronectin' |
| 4095 | 'FN1_No123' | 'QGENGQMMSCTCLGNGK' | 'Fibronectin' |
| 4096 | 'FN1_No124' | 'IGDQWDK' | 'Fibronectin' |
| 4097 | 'FN1_No125' | 'CDPVDQCQDSETGTFYQIGDSWEK' | 'Fibronectin' |
| 4098 | 'FN1_No126' | 'PQAPITGYR' | 'Fibronectin' |
| 4099 | 'FN1_No127' | 'GLAFTDVDVDSIK' | 'Fibronectin' |
| 4100 | 'FN1_No128' | 'QGENGQMMSCTCLGNGKGEFK' | 'Fibronectin' |
| 4101 | 'FN1_No129' | 'DGQERDAPIVNK' | 'Fibronectin' |
| 4102 | 'FN1_No130' | 'TFYSCTTEGRQDGHLWCSTTSNYEQDQK' | 'Fibronectin' |
| 4103 | 'FN1_No131' | 'STTPDITGYR' | 'Fibronectin' |
| 4104 | 'FN1_No132' | 'AQITGYR' | 'Fibronectin' |
| 4105 | 'FN1_No133' | 'PGCYDNGKHYQINQQWER' | 'Fibronectin' |
| 4106 | 'FN1_No134' | 'CDPHEATCYDDGKTYHVGEQWQK' | 'Fibronectin' |
| 4107 | 'FN1_No135' | 'QDGHLWCSTTSNYEQDQK' | 'Fibronectin' |
| 4108 | 'FN1_No136' | 'VREEVVTVGNSVNEGLNQPTDDSCFDPYTVSHYAVGDEWER' | 'Fibronectin' |
| 4109 | 'FN1_No137' | 'GNLLQCICTGNGRGEWKCER' | 'Fibronectin' |
| 4110 | 'FN1_No138' | 'SDTVPSPR' | 'Fibronectin' |
| 4111 | 'FN1_No139' | 'VTDATETTITISWR' | 'Fibronectin' |
| 4112 | 'FN1_No140' | 'QGENGQMMSCTCLGNGKGEFK' | 'Fibronectin' |
| 4113 | 'FN1_No141' | 'NSITLTNLTPGTEYVVSIVALNGR' | 'Fibronectin' |
| 4114 | 'FN1_No142' | 'PEAEETCFDKYTGNTYR' | 'Fibronectin' |
| 4115 | 'FN1_No143' | 'GEFKCDPHEATCYDDGK' | 'Fibronectin' |
| 4116 | 'FN1_No144' | 'ESKPLTAQQTTK' | 'Fibronectin' |
| 4117 | 'FN1_No145' | 'GEFKCDPHEATCYDDGKTYHVGEQWQK' | 'Fibronectin' |
| 4118 | 'FN1_No146' | 'VGDTYERPKDSMIWDCTCIGAGR' | 'Fibronectin' |
| 4119 | 'FN1_No147' | 'FTQVTPTSLSAQWTPPNVQLTGYR' | 'Fibronectin' |
| 4120 | 'FN1_No148' | 'PGVVYEGQLISIQQYGHQEVTR' | 'Fibronectin' |
| 4121 | 'FN1_No149' | 'TGPMKEINLAPDSSSVVVSGLMVATK' | 'Fibronectin' |
| 4122 | 'FN1_No150' | 'TTPPTTATPIR' | 'Fibronectin' |
| 4123 | 'FN1_No151' | 'CDPHEATCYDDGK' | 'Fibronectin' |
| 4124 | 'FN1_No152' | 'QAQQMVQPQSPVAVSQSKPGCYDNGK' | 'Fibronectin' |
| 4125 | 'FN1_No153' | 'TGPMKEINLAPDSSSVVVSGLMVATK' | 'Fibronectin' |
| 4126 | 'FN1_No154' | 'VDVIPVNLPGEHGQRLPISR' | 'Fibronectin' |
| 4127 | 'FN1_No155' | 'IAWESPQGQVSR' | 'Fibronectin' |
| 4128 | 'FN1_No156' | 'DNMKWCGTTQNYDADQK' | 'Fibronectin' |
| 4129 | 'FN1_No157' | 'HKVREEVVTVGNSVNEGLNQPTDDSCFDPYTVSHYAVGDEWER' | 'Fibronectin' |
| 4130 | 'FN1_No158' | 'HEEGHMLNCTCFGQGR' | 'Fibronectin' |
| 4131 | 'FN1_No159' | 'EVVPRPR' | 'Fibronectin' |
| 4132 | 'FN1_No160' | 'QMLCTCLGNGVSCQETAVTQTYGGNSNGEPCVLPFTYNGR' | 'Fibronectin' |
| 4133 | 'FN1_No161' | 'WCHDNGVNYKIGEKWDR' | 'Fibronectin' |
| 4134 | 'FN1_No162' | 'TNTNVNCPIECFMPLDVQADREDSRE' | 'Fibronectin' |
| 4135 | 'FN1_No163' | 'PYPPNVGEEIQIGHIPR' | 'Fibronectin' |
| 4136 | 'FN1_No164' | 'TPFVTHPGYDTGNGIQLPGTSGQQPSVGQQMIFEEHGFR' | 'Fibronectin' |
| 4137 | 'FN1_No165' | 'CDPVDQCQDSETGTFYQIGDSWEKYVHGVR' | 'Fibronectin' |

| | A | B | C |
|---|---|---|---|
| 1 | **Peptide Name** | **Peptide Sequence** | **Peptide Description** |
| 4138 | 'FN1_No166' | 'TNTNVNCPIECFMPLDVQADREDSR' | 'Fibronectin' |
| 4139 | 'FN1_No167' | 'CDNCRRPGGEPSPEGTTGQSYNQYSQR' | 'Fibronectin' |
| 4140 | 'FN1_No168' | 'TGPMKEINLAPDSSSVVVSGLMVATK' | 'Fibronectin' |
| 4141 | 'FN1_No169' | 'WKCDPVDQCQDSETGTFYQIGDSWEK' | 'Fibronectin' |
| 4142 | 'FN1_No170' | 'MSESGFKLLCQCLGFGSGHFR' | 'Fibronectin' |
| 4143 | 'FN1_No171' | 'YTGNTYR' | 'Fibronectin' |
| 4144 | 'FN1_No172' | 'RHEEGHMLNCTCFGQGR' | 'Fibronectin' |
| 4145 | 'FN1_No173' | 'CHEGGQSYKIGDTWR' | 'Fibronectin' |
| 4146 | 'FN1_No174' | 'IGEKWDR' | 'Fibronectin' |
| 4147 | 'FN1_No175' | 'DQCIVDDITYNVNDTFHK' | 'Fibronectin' |
| 4148 | 'FN1_No176' | 'TNTNVNCPIECFMPLDVQADR' | 'Fibronectin' |
| 4149 | 'FN1_No177' | 'QGENGQMMSCTCLGNGKGEFK' | 'Fibronectin' |
| 4150 | 'FN1_No178' | 'WCGTTQNYDADQKFGFCPMAAHEEICTTNEGVMYR' | 'Fibronectin' |
| 4151 | 'FN1_No179' | 'CFDHAAGTSYVVGETWEK' | 'Fibronectin' |
| 4152 | 'FN1_No180' | 'TYHVGEQWQKEYLGAICSCTCFGGQR' | 'Fibronectin' |
| 4153 | 'FN1_No181' | 'VFAVSHGRESKPLTAQQTTK' | 'Fibronectin' |
| 4154 | 'FN1_No182' | 'QDGHLWCSTTSNYEQDQKYSFCTDHTVLVQTR' | 'Fibronectin' |
| 4155 | 'FN1_No183' | 'QGENGQMMSCTCLGNGK' | 'Fibronectin' |
| 4156 | 'FN1_No184' | 'HHPEHFSGRPR' | 'Fibronectin' |
| 4157 | 'FN1_No185' | 'PGGEPSPEGTTGQSYNQYSQR' | 'Fibronectin' |
| 4158 | 'FN1_No186' | 'MSESGFK' | 'Fibronectin' |
| 4159 | 'FN1_No187' | 'PSQMQVTDVQDNSISVK' | 'Fibronectin' |
| 4160 | 'FN1_No188' | 'STATISGLKPGVDYTITVYAVTGRGDSPASSK' | 'Fibronectin' |
| 4161 | 'FN1_No189' | 'HEEGHMLNCTCFGQGR' | 'Fibronectin' |
| 4162 | 'FN1_No190' | 'ARVTDATETTITISWR' | 'Fibronectin' |
| 4163 | 'FN1_No191' | 'QAQQMVQPQSPVAVSQSK' | 'Fibronectin' |
| 4164 | 'FN1_No192' | 'WCHDNGVNYKIGEK' | 'Fibronectin' |
| 4165 | 'FN1_No193' | 'QAQQMVQPQSPVAVSQSKPGCYDNGKHYQINQQWER' | 'Fibronectin' |
| 4166 | 'FN1_No194' | 'DQCIVDDITYNVNDTFHKR' | 'Fibronectin' |
| 4167 | 'FN1_No195' | 'WCGTTQNYDADQKFGFCPMAAHEEICTTNEGVMYR' | 'Fibronectin' |
| 4168 | 'RBP4_No1' | 'GNDDHWIVDTDYDTYAVQYSCR' | 'Retinol-binding protein 4' |
| 4169 | 'RBP4_No2' | 'LLNNWDVCADMVGTFTDTEDPAKFK' | 'Retinol-binding protein 4' |
| 4170 | 'RBP4_No3' | 'LIVHNGYCDGR' | 'Retinol-binding protein 4' |
| 4171 | 'RBP4_No4' | 'MKYWGVASFLQK' | 'Retinol-binding protein 4' |
| 4172 | 'RBP4_No5' | 'QRQEELCLAR' | 'Retinol-binding protein 4' |
| 4173 | 'RBP4_No6' | 'LLNLDGTCADSYSFVFSR' | 'Retinol-binding protein 4' |
| 4174 | 'RBP4_No7' | 'YWGVASFLQK' | 'Retinol-binding protein 4' |
| 4175 | 'RBP4_No8' | 'MKYWGVASFLQK' | 'Retinol-binding protein 4' |
| 4176 | 'RBP4_No9' | 'QRQEELCLAR' | 'Retinol-binding protein 4' |
| 4177 | 'RBP4_No10' | 'FSGTWYAMAK' | 'Retinol-binding protein 4' |
| 4178 | 'RBP4_No11' | 'LIVHNGYCDGRSER' | 'Retinol-binding protein 4' |
| 4179 | 'RBP4_No12' | 'LLNNWDVCADMVGTFTDTEDPAKFK' | 'Retinol-binding protein 4' |
| 4180 | 'RBP4_No13' | 'KDPEGLFLQDNIVAEFSVDETGQMSATAK' | 'Retinol-binding protein 4' |
| 4181 | 'RBP4_No14' | 'FSGTWYAMAK' | 'Retinol-binding protein 4' |

| | A | B | C |
|---|---|---|---|
| 1 | **Peptide Name** | **Peptide Sequence** | **Peptide Description** |
| 4182 | 'RBP4_No15' | 'DPNGLPPEAQK' | 'Retinol-binding protein 4' |
| 4183 | 'RBP4_No16' | 'KDPEGLFLQDNIVAEFSVDETGQMSATAK' | 'Retinol-binding protein 4' |
| 4184 | 'RBP4_No17' | 'LLNNWDVCADMVGTFTDTEDPAK' | 'Retinol-binding protein 4' |
| 4185 | 'RBP4_No18' | 'QEELCLAR' | 'Retinol-binding protein 4' |
| 4186 | 'RBP4_No19' | 'VKENFDK' | 'Retinol-binding protein 4' |
| 4187 | 'RBP4_No20' | 'LLNLDGTCADSYSFVFSRDPNGLPPEAQK' | 'Retinol-binding protein 4' |
| 4188 | 'RBP4_No21' | 'LLNNWDVCADMVGTFTDTEDPAK' | 'Retinol-binding protein 4' |
| 4189 | 'RBP4_No22' | 'VRLLNNWDVCADMVGTFTDTEDPAKFK' | 'Retinol-binding protein 4' |
| 4190 | 'RBP4_No23' | 'QEELCLAR' | 'Retinol-binding protein 4' |
| 4191 | 'RBP4_No24' | 'FSGTWYAMAKK' | 'Retinol-binding protein 4' |
| 4192 | 'RBP4_No25' | 'IVRQRQEELCLAR' | 'Retinol-binding protein 4' |
| 4193 | 'RBP4_No26' | 'FSGTWYAMAKK' | 'Retinol-binding protein 4' |
| 4194 | 'RBP4_No27' | 'VRLLNNWDVCADMVGTFTDTEDPAKFK' | 'Retinol-binding protein 4' |
| 4195 | 'RBP4_No28' | 'VKENFDKAR' | 'Retinol-binding protein 4' |
| 4196 | 'RBP4_No29' | 'QRQEELCLARQYR' | 'Retinol-binding protein 4' |
| 4197 | 'AMBP_No1' | 'AFIQLWAFDAVK' | 'Protein AMBP' |
| 4198 | 'AMBP_No2' | 'EYCGVPGDGDEELLR' | 'Protein AMBP' |
| 4199 | 'AMBP_No3' | 'EDSCQLGYSAGPCMGMTSR' | 'Protein AMBP' |
| 4200 | 'AMBP_No4' | 'EDSCQLGYSAGPCMGMTSR' | 'Protein AMBP' |
| 4201 | 'AMBP_No5' | 'TVAACNLPIVR' | 'Protein AMBP' |
| 4202 | 'AMBP_No6' | 'GVCEETSGAYEK' | 'Protein AMBP' |
| 4203 | 'AMBP_No7' | 'GVCEETSGAYEKTDTDGK' | 'Protein AMBP' |
| 4204 | 'AMBP_No8' | 'MTVSTLVLGEGATEAEISMTSTR' | 'Protein AMBP' |
| 4205 | 'AMBP_No9' | 'KEDSCQLGYSAGPCMGMTSR' | 'Protein AMBP' |
| 4206 | 'AMBP_No10' | 'VVAQGVGIPEDSIFTMADRGECVPGEQEPEPILIPR' | 'Protein AMBP' |
| 4207 | 'AMBP_No11' | 'KEDSCQLGYSAGPCMGMTSR' | 'Protein AMBP' |
| 4208 | 'AMBP_No12' | 'WYNLAIGSTCPWLK' | 'Protein AMBP' |
| 4209 | 'AMBP_No13' | 'WYNLAIGSTCPWLKK' | 'Protein AMBP' |
| 4210 | 'AMBP_No14' | 'VVAQGVGIPEDSIFTMADR' | 'Protein AMBP' |
| 4211 | 'AMBP_No15' | 'EDSCQLGYSAGPCMGMTSR' | 'Protein AMBP' |
| 4212 | 'AMBP_No16' | 'EDSCQLGYSAGPCMGMTSR' | 'Protein AMBP' |
| 4213 | 'AMBP_No17' | 'EYCGVPGDGDEELLRFSN' | 'Protein AMBP' |
| 4214 | 'AMBP_No18' | 'ECLQTCR' | 'Protein AMBP' |
| 4215 | 'AMBP_No19' | 'KGVCEETSGAYEKTDTDGK' | 'Protein AMBP' |
| 4216 | 'AMBP_No20' | 'KEDSCQLGYSAGPCMGMTSR' | 'Protein AMBP' |
| 4217 | 'AMBP_No21' | 'CVLFPYGGCQGNGNKFYSEKECR' | 'Protein AMBP' |
| 4218 | 'AMBP_No22' | 'VVAQGVGIPEDSIFTMADR' | 'Protein AMBP' |
| 4219 | 'AMBP_No23' | 'KEDSCQLGYSAGPCMGMTSR' | 'Protein AMBP' |
| 4220 | 'AMBP_No24' | 'CVLFPYGGCQGNGNK' | 'Protein AMBP' |
| 4221 | 'AMBP_No25' | 'ETLLQDFR' | 'Protein AMBP' |
| 4222 | 'AMBP_No26' | 'KGVCEETSGAYEK' | 'Protein AMBP' |
| 4223 | 'AMBP_No27' | 'GKCVLFPYGGCQGNGNK' | 'Protein AMBP' |
| 4224 | 'AMBP_No28' | 'GVCEETSGAYEKTDTDGKFLYHK' | 'Protein AMBP' |
| 4225 | 'AMBP_No29' | 'GECVPGEQEPEPILIPR' | 'Protein AMBP' |

| | A | B | C |
|---|---|---|---|
| 1 | **Peptide Name** | **Peptide Sequence** | **Peptide Description** |
| 4226 | 'AMBP_No30' | 'CVLFPYGGCQGNGNKFYSEK' | 'Protein AMBP' |
| 4227 | 'AMBP_No31' | 'MTVSTLVLGEGATEAEISMTSTR' | 'Protein AMBP' |
| 4228 | 'AMBP_No32' | 'AFIQLWAFDAVKGK' | 'Protein AMBP' |
| 4229 | 'AMBP_No33' | 'MTVSTLVLGEGATEAEISMTSTR' | 'Protein AMBP' |
| 4230 | 'AMBP_No34' | 'FYSEKECR' | 'Protein AMBP' |
| 4231 | 'AMBP_No35' | 'TDTDGKFLYHK' | 'Protein AMBP' |
| 4232 | 'AMBP_No36' | 'IYGKWYNLAIGSTCPWLK' | 'Protein AMBP' |
| 4233 | 'AMBP_No37' | 'VVAQGVGIPEDSIFTMADRGECVPGEQEPEPILIPR' | 'Protein AMBP' |
| 4234 | 'AMBP_No38' | 'MTVSTLVLGEGATEAEISMTSTR' | 'Protein AMBP' |
| 4235 | 'AMBP_No39' | 'APQLRETLLQDFR' | 'Protein AMBP' |
| 4236 | 'AMBP_No40' | 'ECREYCGVPGDGDEELLRFSN' | 'Protein AMBP' |
| 4237 | 'AMBP_No41' | 'FYSEKECREYCGVPGDGDEELLR' | 'Protein AMBP' |
| 4238 | 'AMBP_No42' | 'WRKGVCEETSGAYEK' | 'Protein AMBP' |
| 4239 | 'AMBP_No43' | 'TVAACNLPIVRGPCR' | 'Protein AMBP' |
| 4240 | 'AMBP_No44' | 'HHGPTITAK' | 'Protein AMBP' |
| 4241 | 'ORM1_No1' | 'YVGGQEHFAHLLILR' | 'Alpha-1-acid glycoprotein 1' |
| 4242 | 'ORM1_No2' | 'TYMLAFDVNDEKNWGLSVYADKPETTK' | 'Alpha-1-acid glycoprotein 1' |
| 4243 | 'ORM1_No3' | 'TEDTIFLR' | 'Alpha-1-acid glycoprotein 1' |
| 4244 | 'ORM1_No4' | 'TYMLAFDVNDEKNWGLSVYADKPETTK' | 'Alpha-1-acid glycoprotein 1' |
| 4245 | 'ORM1_No5' | 'SDVVYTDWKK' | 'Alpha-1-acid glycoprotein 1' |
| 4246 | 'ORM1_No6' | 'WFYIASAFRNEEYNK' | 'Alpha-1-acid glycoprotein 1' |
| 4247 | 'ORM1_No7' | 'TYMLAFDVNDEK' | 'Alpha-1-acid glycoprotein 1' |
| 4248 | 'ORM1_No8' | 'YVGGQEHFAHLLILRDTK' | 'Alpha-1-acid glycoprotein 1' |
| 4249 | 'ORM1_No9' | 'DKCEPLEK' | 'Alpha-1-acid glycoprotein 1' |
| 4250 | 'ORM1_No10' | 'SDVVYTDWKKDK' | 'Alpha-1-acid glycoprotein 1' |
| 4251 | 'ORM1_No11' | 'NWGLSVYADKPETTK' | 'Alpha-1-acid glycoprotein 1' |
| 4252 | 'ORM1_No12' | 'EQLGEFYEALDCLR' | 'Alpha-1-acid glycoprotein 1' |
| 4253 | 'ORM1_No13' | 'EQLGEFYEALDCLRIPK' | 'Alpha-1-acid glycoprotein 1' |
| 4254 | 'ORM1_No14' | 'NWGLSVYADKPETTKEQLGEFYEALDCLR' | 'Alpha-1-acid glycoprotein 1' |
| 4255 | 'ORM1_No15' | 'SVQEIQATFFYFTPNK' | 'Alpha-1-acid glycoprotein 1' |
| 4256 | 'ORM1_No16' | 'WFYIASAFR' | 'Alpha-1-acid glycoprotein 1' |
| 4257 | 'ORM1_No17' | 'SDVVYTDWK' | 'Alpha-1-acid glycoprotein 1' |
| 4258 | 'ORM1_No18' | 'QDQCIYNTTYLNVQR' | 'Alpha-1-acid glycoprotein 1' |
| 4259 | 'ORM1_No19' | 'IPKSDVVYTDWKK' | 'Alpha-1-acid glycoprotein 1' |
| 4260 | 'ORM1_No20' | 'IPKSDVVYTDWK' | 'Alpha-1-acid glycoprotein 1' |
| 4261 | 'ORM1_No21' | 'TYMLAFDVNDEK' | 'Alpha-1-acid glycoprotein 1' |
| 4262 | 'ORM1_No22' | 'DTKTYMLAFDVNDEK' | 'Alpha-1-acid glycoprotein 1' |
| 4263 | 'ORM1_No23' | 'TEDTIFLREYQTR' | 'Alpha-1-acid glycoprotein 1' |
| 4264 | 'ORM1_No24' | 'DKCEPLEKQHEK' | 'Alpha-1-acid glycoprotein 1' |
| 4265 | 'ORM1_No25' | 'CEPLEKQHEKER' | 'Alpha-1-acid glycoprotein 1' |
| 4266 | 'ORM1_No26' | 'QDQCIYNTTYLNVQR' | 'Alpha-1-acid glycoprotein 1' |
| 4267 | 'ORM1_No27' | 'KDKCEPLEK' | 'Alpha-1-acid glycoprotein 1' |
| 4268 | 'AHSG_No1' | 'AQLVPLPPSTYVEFTVSGTDCVAK' | 'Alpha-2-HS-glycoprotein' |
| 4269 | 'AHSG_No2' | 'QLKEHAVEGDCDFQLLK' | 'Alpha-2-HS-glycoprotein' |

| | A | B | C |
|---|---|---|---|
| 1 | **Peptide Name** | **Peptide Sequence** | **Peptide Description** |
| 4270 | 'AHSG_No3' | 'QLKEHAVEGDCDFQLLK' | 'Alpha-2-HS-glycoprotein' |
| 4271 | 'AHSG_No4' | 'HTFMGVVSLGSPSGEVSHPR' | 'Alpha-2-HS-glycoprotein' |
| 4272 | 'AHSG_No5' | 'FSVVYAK' | 'Alpha-2-HS-glycoprotein' |
| 4273 | 'AHSG_No6' | 'HTLNQIDEVK' | 'Alpha-2-HS-glycoprotein' |
| 4274 | 'AHSG_No7' | 'TVVQPSVGAAAGPVVPPCPGR' | 'Alpha-2-HS-glycoprotein' |
| 4275 | 'AHSG_No8' | 'EHAVEGDCDFQLLK' | 'Alpha-2-HS-glycoprotein' |
| 4276 | 'AHSG_No9' | 'EHAVEGDCDFQLLKLDGK' | 'Alpha-2-HS-glycoprotein' |
| 4277 | 'AHSG_No10' | 'QLKEHAVEGDCDFQLLKLDGK' | 'Alpha-2-HS-glycoprotein' |
| 4278 | 'AHSG_No11' | 'LDGKFSVVYAK' | 'Alpha-2-HS-glycoprotein' |
| 4279 | 'AHSG_No12' | 'HTFMGVVSLGSPSGEVSHPR' | 'Alpha-2-HS-glycoprotein' |
| 4280 | 'AHSG_No13' | 'QPNCDDPETEEAALVAIDYINQNLPWGYK' | 'Alpha-2-HS-glycoprotein' |
| 4281 | 'AHSG_No14' | 'AQLVPLPPSTYVEFTVSGTDCVAKEATEAAKCNLLAEK' | 'Alpha-2-HS-glycoprotein' |
| 4282 | 'AHSG_No15' | 'AQLVPLPPSTYVEFTVSGTDCVAKEATEAAK' | 'Alpha-2-HS-glycoprotein' |
| 4283 | 'AHSG_No16' | 'CNLLAEKQYGFCK' | 'Alpha-2-HS-glycoprotein' |
| 4284 | 'AHSG_No17' | 'QLKEHAVEGDCDFQLLKLDGK' | 'Alpha-2-HS-glycoprotein' |
| 4285 | 'AHSG_No18' | 'CDSSPDSAEDVR' | 'Alpha-2-HS-glycoprotein' |
| 4286 | 'AHSG_No19' | 'EATEAAKCNLLAEK' | 'Alpha-2-HS-glycoprotein' |
| 4287 | 'AHSG_No20' | 'EHAVEGDCDFQLLKLDGKFSVVYAK' | 'Alpha-2-HS-glycoprotein' |
| 4288 | 'AHSG_No21' | 'CNLLAEK' | 'Alpha-2-HS-glycoprotein' |
| 4289 | 'AHSG_No22' | 'EATEAAKCNLLAEKQYGFCK' | 'Alpha-2-HS-glycoprotein' |
| 4290 | 'AHSG_No23' | 'CDSSPDSAEDVRK' | 'Alpha-2-HS-glycoprotein' |
| 4291 | 'AHSG_No24' | 'AALAAFNAQNNGSNFQLEEISR' | 'Alpha-2-HS-glycoprotein' |
| 4292 | 'AHSG_No25' | 'VCQDCPLLAPLNDTR' | 'Alpha-2-HS-glycoprotein' |
| 4293 | 'AHSG_No26' | 'AHYDLRHTFMGVVSLGSPSGEVSHPR' | 'Alpha-2-HS-glycoprotein' |
| 4294 | 'AHSG_No27' | 'QYGFCKATLSEK' | 'Alpha-2-HS-glycoprotein' |
| 4295 | 'TTR_No1' | 'GSPAINVAVHVFR' | 'Transthyretin' |
| 4296 | 'TTR_No2' | 'TSESGELHGLTTEEEFVEGIYK' | 'Transthyretin' |
| 4297 | 'TTR_No3' | 'ALGISPFHEHAEVVFTANDSGPR' | 'Transthyretin' |
| 4298 | 'TTR_No4' | 'ALGISPFHEHAEVVFTANDSGPRR' | 'Transthyretin' |
| 4299 | 'TTR_No5' | 'YTIAALLSPYSYSTTAVVTNPKE' | 'Transthyretin' |
| 4300 | 'TTR_No6' | 'TSESGELHGLTTEEEFVEGIYKVEIDTK' | 'Transthyretin' |
| 4301 | 'TTR_No7' | 'GSPAINVAVHVFRK' | 'Transthyretin' |
| 4302 | 'TTR_No8' | 'AADDTWEPFASGK' | 'Transthyretin' |
| 4303 | 'TTR_No9' | 'RYTIAALLSPYSYSTTAVVTNPK' | 'Transthyretin' |
| 4304 | 'TTR_No10' | 'VLDAVRGSPAINVAVHVFRK' | 'Transthyretin' |
| 4305 | 'TTR_No11' | 'RYTIAALLSPYSYSTTAVVTNPKE' | 'Transthyretin' |
| 4306 | 'TTR_No12' | 'VLDAVRGSPAINVAVHVFR' | 'Transthyretin' |
| 4307 | 'TTR_No13' | 'GSPAINVAVHVFRKAADDTWEPFASGK' | 'Transthyretin' |
| 4308 | 'TTR_No14' | 'KAADDTWEPFASGK' | 'Transthyretin' |
| 4309 | 'TTR_No15' | 'CPLMVKVLDAVR' | 'Transthyretin' |
| 4310 | 'TTR_No16' | 'YTIAALLSPYSYSTTAVVTNPK' | 'Transthyretin' |
| 4311 | 'TTR_No17' | 'VEIDTKSYWK' | 'Transthyretin' |
| 4312 | 'TTR_No18' | 'CPLMVKVLDAVR' | 'Transthyretin' |
| 4313 | 'ALB_No1' | 'RHPDYSVVLLLR' | 'Serum albumin' |

| | A | B | C |
|---|---|---|---|
| 1 | **Peptide Name** | **Peptide Sequence** | **Peptide Description** |
| 4314 | 'ALB_No2' | 'QNCELFEQLGEYK' | 'Serum albumin' |
| 4315 | 'ALB_No3' | 'HPDYSVVLLLR' | 'Serum albumin' |
| 4316 | 'ALB_No4' | 'VPQVSTPTLVEVSR' | 'Serum albumin' |
| 4317 | 'ALB_No5' | 'QNCELFEQLGEYK' | 'Serum albumin' |
| 4318 | 'ALB_No6' | 'FKDLGEENFK' | 'Serum albumin' |
| 4319 | 'ALB_No7' | 'ETYGEMADCCAK' | 'Serum albumin' |
| 4320 | 'ALB_No8' | 'AEFAEVSKLVTDLTK' | 'Serum albumin' |
| 4321 | 'ALB_No9' | 'VHTECCHGDLLECADDR' | 'Serum albumin' |
| 4322 | 'ALB_No10' | 'SLHTLFGDKLCTVATLR' | 'Serum albumin' |
| 4323 | 'ALB_No11' | 'LVRPEVDVMCTAFHDNEETFLKK' | 'Serum albumin' |
| 4324 | 'ALB_No12' | 'LVRPEVDVMCTAFHDNEETFLK' | 'Serum albumin' |
| 4325 | 'ALB_No13' | 'TCVADESAENCDKSLHTLFGDK' | 'Serum albumin' |
| 4326 | 'ALB_No14' | 'RPCFSALEVDETYVPKEFNAETFTFHADICTLSEK' | 'Serum albumin' |
| 4327 | 'ALB_No15' | 'TCVADESAENCDK' | 'Serum albumin' |
| 4328 | 'ALB_No16' | 'YKAAFTECCQAADKAACLLPK' | 'Serum albumin' |
| 4329 | 'ALB_No17' | 'EQLKAVMDDFAAFVEK' | 'Serum albumin' |
| 4330 | 'ALB_No18' | 'PLVEEPQNLIK' | 'Serum albumin' |
| 4331 | 'ALB_No19' | 'SLHTLFGDKLCTVATLRETYGEMADCCAK' | 'Serum albumin' |
| 4332 | 'ALB_No20' | 'SHCIAEVENDEMPADLPSLAADFVESKDVCKNYAEAK' | 'Serum albumin' |
| 4333 | 'ALB_No21' | 'NYAEAKDVFLGMFLYEYAR' | 'Serum albumin' |
| 4334 | 'ALB_No22' | 'NYAEAKDVFLGMFLYEYAR' | 'Serum albumin' |
| 4335 | 'ALB_No23' | 'PEVDVMCTAFHDNEETFLKK' | 'Serum albumin' |
| 4336 | 'ALB_No24' | 'QNCELFEQLGEYKFQNALLVR' | 'Serum albumin' |
| 4337 | 'ALB_No25' | 'RHPDYSVVLLLRLAK' | 'Serum albumin' |
| 4338 | 'ALB_No26' | 'NYAEAKDVFLGMFLYEYARR' | 'Serum albumin' |
| 4339 | 'ALB_No27' | 'QEPERNECFLQHK' | 'Serum albumin' |
| 4340 | 'ALB_No28' | 'NECFLQHKDDNPNLPR' | 'Serum albumin' |
| 4341 | 'ALB_No29' | 'SLHTLFGDK' | 'Serum albumin' |
| 4342 | 'ALB_No30' | 'LKECCEKPLLEK' | 'Serum albumin' |
| 4343 | 'ALB_No31' | 'CCAAADPHECYAK' | 'Serum albumin' |
| 4344 | 'ALB_No32' | 'LDELRDEGK' | 'Serum albumin' |
| 4345 | 'ALB_No33' | 'ADDKETCFAEEGK' | 'Serum albumin' |
| 4346 | 'ALB_No34' | 'ECCEKPLLEK' | 'Serum albumin' |
| 4347 | 'ALB_No35' | 'FPKAEFAEVSK' | 'Serum albumin' |
| 4348 | 'ALB_No36' | 'KLVAASQAALGL' | 'Serum albumin' |
| 4349 | 'ALB_No37' | 'ALVLIAFAQYLQQCPFEDHVKLVNEVTEFAK' | 'Serum albumin' |
| 4350 | 'ALB_No38' | 'TPVSDRVTK' | 'Serum albumin' |
| 4351 | 'ALB_No39' | 'LSQRFPK' | 'Serum albumin' |
| 4352 | 'ALB_No40' | 'PEVDVMCTAFHDNEETFLKK' | 'Serum albumin' |
| 4353 | 'ALB_No41' | 'LKCASLQK' | 'Serum albumin' |
| 4354 | 'ALB_No42' | 'KQTALVELVK' | 'Serum albumin' |
| 4355 | 'ALB_No43' | 'QEPERNECFLQHK' | 'Serum albumin' |
| 4356 | 'ALB_No44' | 'ADDKETCFAEEGKK' | 'Serum albumin' |
| 4357 | 'ALB_No45' | 'QTALVELVK' | 'Serum albumin' |

| | A | B | C |
|---|---|---|---|
| 1 | **Peptide Name** | **Peptide Sequence** | **Peptide Description** |
| 4358 | 'ALB_No46' | 'QTALVELVK' | 'Serum albumin' |
| 4359 | 'ALB_No47' | 'SLHTLFGDKLCTVATLRETYGEMADCCAK' | 'Serum albumin' |
| 4360 | 'ALB_No48' | 'YLYEIARR' | 'Serum albumin' |
| 4361 | 'ALB_No49' | 'YICENQDSISSKLK' | 'Serum albumin' |
| 4362 | 'ALB_No50' | 'AVMDDFAAFVEKCCK' | 'Serum albumin' |
| 4363 | 'ALB_No51' | 'VFDEFKPLVEEPQNLIK' | 'Serum albumin' |
| 4364 | 'ALB_No52' | 'LDELRDEGKASSAK' | 'Serum albumin' |
| 4365 | 'ALB_No53' | 'DVCKNYAEAKDVFLGMFLYEYAR' | 'Serum albumin' |
| 4366 | 'ALB_No54' | 'PEVDVMCTAFHDNEETFLK' | 'Serum albumin' |
| 4367 | 'ALB_No55' | 'QIKKQTALVELVK' | 'Serum albumin' |
| 4368 | 'ALB_No56' | 'QEPERNECFLQHKDDNPNLPR' | 'Serum albumin' |
| 4369 | 'ALB_No57' | 'KYLYEIAR' | 'Serum albumin' |
| 4370 | 'ALB_No58' | 'KVPQVSTPTLVEVSR' | 'Serum albumin' |
| 4371 | 'ALB_No59' | 'MPCAEDYLSVVLNQLCVLHEKTPVSDRVTK' | 'Serum albumin' |
| 4372 | 'ALB_No60' | 'LCTVATLRETYGEMADCCAK' | 'Serum albumin' |
| 4373 | 'ALB_No61' | 'LAKTYETTLEK' | 'Serum albumin' |
| 4374 | 'ALB_No62' | 'QNCELFEQLGEYKFQNALLVRYTK' | 'Serum albumin' |
| 4375 | 'ALB_No63' | 'PCFSALEVDETYVPKEFNAETFTFHADICTLSEKER' | 'Serum albumin' |
| 4376 | 'ALB_No64' | 'LVAASQAALGL' | 'Serum albumin' |
| 4377 | 'ALB_No65' | 'QNCELFEQLGEYKFQNALLVR' | 'Serum albumin' |
| 4378 | 'ALB_No66' | 'SHCIAEVENDEMPADLPSLAADFVESKDVCKNYAEAK' | 'Serum albumin' |
| 4379 | 'ALB_No67' | 'TCVADESAENCDKSLHTLFGDKLCTVATLR' | 'Serum albumin' |
| 4380 | 'ALB_No68' | 'DVCKNYAEAKDVFLGMFLYEYAR' | 'Serum albumin' |
| 4381 | 'ALB_No69' | 'AACLLPKLDELRDEGK' | 'Serum albumin' |
| 4382 | 'ALB_No70' | 'RMPCAEDYLSVVLNQLCVLHEK' | 'Serum albumin' |
| 4383 | 'ALB_No71' | 'ATKEQLKAVMDDFAAFVEK' | 'Serum albumin' |
| 4384 | 'ALB_No72' | 'LAKTYETTLEKCCAAADPHECYAK' | 'Serum albumin' |
| 4385 | 'ALB_No73' | 'TYETTLEKCCAAADPHECYAK' | 'Serum albumin' |
| 4386 | 'ALB_No74' | 'RHPYFYAPELLFFAKR' | 'Serum albumin' |
| 4387 | 'ALB_No75' | 'LVRPEVDVMCTAFHDNEETFLKK' | 'Serum albumin' |
| 4388 | 'ALB_No76' | 'DVFLGMFLYEYAR' | 'Serum albumin' |
| 4389 | 'ALB_No77' | 'QTALVELVKHKPK' | 'Serum albumin' |
| 4390 | 'ALB_No78' | 'MPCAEDYLSVVLNQLCVLHEK' | 'Serum albumin' |
| 4391 | 'ALB_No79' | 'AWAVARLSQR' | 'Serum albumin' |
| 4392 | 'ALB_No80' | 'DLGEENFK' | 'Serum albumin' |
| 4393 | 'ALB_No81' | 'TPVSDRVTKCCTESLVNR' | 'Serum albumin' |
| 4394 | 'ALB_No82' | 'LVNEVTEFAK' | 'Serum albumin' |
| 4395 | 'ALB_No83' | 'DVCKNYAEAK' | 'Serum albumin' |
| 4396 | 'ALB_No84' | 'TYETTLEK' | 'Serum albumin' |
| 4397 | 'ALB_No85' | 'ALVLIAFAQYLQQCPFEDHVK' | 'Serum albumin' |
| 4398 | 'ALB_No86' | 'SHCIAEVENDEMPADLPSLAADFVESK' | 'Serum albumin' |
| 4399 | 'ALB_No87' | 'RHPYFYAPELLFFAK' | 'Serum albumin' |
| 4400 | 'ALB_No88' | 'VHTECCHGDLLECADDRADLAK' | 'Serum albumin' |
| 4401 | 'ALB_No89' | 'HPYFYAPELLFFAK' | 'Serum albumin' |

| | A | B | C |
|---|---|---|---|
| 1 | **Peptide Name** | **Peptide Sequence** | **Peptide Description** |
| 4402 | 'ALB_No90' | 'QEPERNECFLQHKDDNPNLPR' | 'Serum albumin' |
| 4403 | 'ALB_No91' | 'LVTDLTKVHTECCHGDLLECADDRADLAK' | 'Serum albumin' |
| 4404 | 'ALB_No92' | 'YLYEIAR' | 'Serum albumin' |
| 4405 | 'ALB_No93' | 'ETYGEMADCCAKQEPER' | 'Serum albumin' |
| 4406 | 'ALB_No94' | 'MPCAEDYLSVVLNQLCVLHEKTPVSDR' | 'Serum albumin' |
| 4407 | 'ALB_No95' | 'MPCAEDYLSVVLNQLCVLHEKTPVSDRVTK' | 'Serum albumin' |
| 4408 | 'ALB_No96' | 'HPDYSVVLLLRLAKTYETTLEK' | 'Serum albumin' |
| 4409 | 'ALB_No97' | 'EFNAETFTFHADICTLSEKER' | 'Serum albumin' |
| 4410 | 'ALB_No98' | 'LSQRFPKAEFAEVSK' | 'Serum albumin' |
| 4411 | 'ALB_No99' | 'QTALVELVKHKPK' | 'Serum albumin' |
| 4412 | 'ALB_No100' | 'RMPCAEDYLSVVLNQLCVLHEK' | 'Serum albumin' |
| 4413 | 'ALB_No101' | 'AVMDDFAAFVEK' | 'Serum albumin' |
| 4414 | 'ALB_No102' | 'ETYGEMADCCAKQEPERNECFLQHK' | 'Serum albumin' |
| 4415 | 'ALB_No103' | 'SHCIAEVENDEMPADLPSLAADFVESKDVCK' | 'Serum albumin' |
| 4416 | 'ALB_No104' | 'AAFTECCQAADKAACLLPKLDELR' | 'Serum albumin' |
| 4417 | 'ALB_No105' | 'AVMDDFAAFVEK' | 'Serum albumin' |
| 4418 | 'ALB_No106' | 'ETYGEMADCCAK' | 'Serum albumin' |
| 4419 | 'ALB_No107' | 'AAFTECCQAADKAACLLPK' | 'Serum albumin' |
| 4420 | 'ALB_No108' | 'VTKCCTESLVNR' | 'Serum albumin' |
| 4421 | 'ALB_No109' | 'PCFSALEVDETYVPK' | 'Serum albumin' |
| 4422 | 'ALB_No110' | 'KYLYEIARR' | 'Serum albumin' |
| 4423 | 'ALB_No111' | 'LVTDLTK' | 'Serum albumin' |
| 4424 | 'ALB_No112' | 'HPYFYAPELLFFAKR' | 'Serum albumin' |
| 4425 | 'ALB_No113' | 'FQNALLVR' | 'Serum albumin' |
| 4426 | 'ALB_No114' | 'LCTVATLRETYGEMADCCAK' | 'Serum albumin' |
| 4427 | 'ALB_No115' | 'RMPCAEDYLSVVLNQLCVLHEKTPVSDR' | 'Serum albumin' |
| 4428 | 'ALB_No116' | 'FPKAEFAEVSKLVTDLTK' | 'Serum albumin' |
| 4429 | 'ALB_No117' | 'CCTESLVNRR' | 'Serum albumin' |
| 4430 | 'ALB_No118' | 'RPCFSALEVDETYVPK' | 'Serum albumin' |
| 4431 | 'ALB_No119' | 'LVRPEVDVMCTAFHDNEETFLK' | 'Serum albumin' |
| 4432 | 'ALB_No120' | 'YTKKVPQVSTPTLVEVSR' | 'Serum albumin' |
| 4433 | 'ALB_No121' | 'SHCIAEVENDEMPADLPSLAADFVESK' | 'Serum albumin' |
| 4434 | 'ALB_No122' | 'CCTESLVNR' | 'Serum albumin' |
| 4435 | 'ALB_No123' | 'PEVDVMCTAFHDNEETFLK' | 'Serum albumin' |
| 4436 | 'ALB_No124' | 'AACLLPKLDELR' | 'Serum albumin' |
| 4437 | 'ALB_No125' | 'EFNAETFTFHADICTLSEK' | 'Serum albumin' |
| 4438 | 'ALB_No126' | 'RMPCAEDYLSVVLNQLCVLHEKTPVSDR' | 'Serum albumin' |
| 4439 | 'ALB_No127' | 'HPYFYAPELLFFAKRYK' | 'Serum albumin' |
| 4440 | 'ALB_No128' | 'VHTECCHGDLLECADDRADLAKYICENQDSISSK' | 'Serum albumin' |
| 4441 | 'ALB_No129' | 'QIKKQTALVELVK' | 'Serum albumin' |
| 4442 | 'ALB_No130' | 'AEFAEVSK' | 'Serum albumin' |
| 4443 | 'ALB_No131' | 'MPCAEDYLSVVLNQLCVLHEKTPVSDR' | 'Serum albumin' |
| 4444 | 'ALB_No132' | 'LKCASLQKFGER' | 'Serum albumin' |
| 4445 | 'ALB_No133' | 'MPCAEDYLSVVLNQLCVLHEK' | 'Serum albumin' |

| | A | B | C |
|---|---|---|---|
| 1 | **Peptide Name** | **Peptide Sequence** | **Peptide Description** |
| 4446 | 'ALB_No134' | 'HPDYSVVLLLRLAK' | 'Serum albumin' |
| 4447 | 'ALB_No135' | 'YICENQDSISSK' | 'Serum albumin' |
| 4448 | 'ALB_No136' | 'DVFLGMFLYEYAR' | 'Serum albumin' |
| 4449 | 'ALB_No137' | 'ETYGEMADCCAKQEPER' | 'Serum albumin' |
| 4450 | 'ALB_No138' | 'YKAAFTECCQAADK' | 'Serum albumin' |
| 4451 | 'ALB_No139' | 'CCTESLVNRRPCFSALEVDETYVPK' | 'Serum albumin' |
| 4452 | 'ALB_No140' | 'AAFTECCQAADK' | 'Serum albumin' |
| 4453 | 'ALB_No141' | 'AVMDDFAAFVEKCCK' | 'Serum albumin' |
| 4454 | 'ALB_No142' | 'EQLKAVMDDFAAFVEK' | 'Serum albumin' |
| 4455 | 'ALB_No143' | 'ADLAKYICENQDSISSK' | 'Serum albumin' |
| 4456 | 'ALB_No144' | 'ATKEQLKAVMDDFAAFVEK' | 'Serum albumin' |
| 4457 | 'ALB_No145' | 'AEFAEVSKLVTDLTKVHTECCHGDLLECADDR' | 'Serum albumin' |
| 4458 | 'ALB_No146' | 'ETCFAEEGKK' | 'Serum albumin' |
| 4459 | 'ALB_No147' | 'FQNALLVRYTK' | 'Serum albumin' |
| 4460 | 'ALB_No148' | 'CCAAADPHECYAKVFDEFKPLVEEPQNLIK' | 'Serum albumin' |
| 4461 | 'ALB_No149' | 'KVPQVSTPTLVEVSRNLGK' | 'Serum albumin' |
| 4462 | 'ALB_No150' | 'AFKAWAVAR' | 'Serum albumin' |
| 4463 | 'ALB_No151' | 'CCKADDKETCFAEEGK' | 'Serum albumin' |
| 4464 | 'ALB_No152' | 'LVTDLTKVHTECCHGDLLECADDR' | 'Serum albumin' |
| 4465 | 'ALB_No153' | 'DDNPNLPR' | 'Serum albumin' |
| 4466 | 'ALB_No154' | 'FGERAFKAWAVAR' | 'Serum albumin' |
| 4467 | 'ALB_No155' | 'QNCELFEQLGEYKFQNALLVRYTK' | 'Serum albumin' |
| 4468 | 'ALB_No156' | 'LCTVATLR' | 'Serum albumin' |
| 4469 | 'ALB_No157' | 'NECFLQHK' | 'Serum albumin' |
| 4470 | 'ALB_No158' | 'AACLLPK' | 'Serum albumin' |
| 4471 | 'ALB_No159' | 'SHCIAEVENDEMPADLPSLAADFVESKDVCK' | 'Serum albumin' |
| 4472 | 'ALB_No160' | 'EQLKAVMDDFAAFVEKCCK' | 'Serum albumin' |
| 4473 | 'ALB_No161' | 'ETYGEMADCCAKQEPERNECFLQHK' | 'Serum albumin' |
| 4474 | 'ALB_No162' | 'LVNEVTEFAKTCVADESAENCDK' | 'Serum albumin' |
| 4475 | 'ALB_No163' | 'LCTVATLRETYGEMADCCAKQEPER' | 'Serum albumin' |
| 4476 | 'ALB_No164' | 'NYAEAKDVFLGMFLYEYARR' | 'Serum albumin' |
| 4477 | 'ALB_No165' | 'CASLQKFGER' | 'Serum albumin' |
| 4478 | 'ALB_No166' | 'RYKAAFTECCQAADK' | 'Serum albumin' |
| 4479 | 'ALB_No167' | 'LVNEVTEFAKTCVADESAENCDKSLHTLFGDK' | 'Serum albumin' |
| 4480 | 'ALB_No168' | 'EFNAETFTFHADICTLSEKERQIK' | 'Serum albumin' |
| 4481 | 'ALB_No169' | 'QRLKCASLQK' | 'Serum albumin' |
| 4482 | 'ALB_No170' | 'ETCFAEEGK' | 'Serum albumin' |
| 4483 | 'ALB_No171' | 'DVFLGMFLYEYARR' | 'Serum albumin' |
| 4484 | 'ALB_No172' | 'VFDEFKPLVEEPQNLIKQNCELFEQLGEYK' | 'Serum albumin' |
| 4485 | 'ALB_No173' | 'SEVAHRFKDLGEENFK' | 'Serum albumin' |
| 4486 | 'ALB_No174' | 'VPQVSTPTLVEVSRNLGKVGSK' | 'Serum albumin' |
| 4487 | 'ALB_No175' | 'LKECCEK' | 'Serum albumin' |
| 4488 | 'ALB_No176' | 'VPQVSTPTLVEVSRNLGK' | 'Serum albumin' |
| 4489 | 'ALB_No177' | 'CCKHPEAK' | 'Serum albumin' |

| | A | B | C |
|---|---|---|---|
| 1 | **Peptide Name** | **Peptide Sequence** | **Peptide Description** |
| 4490 | 'ALB_No178' | 'YICENQDSISSKLKECCEK' | 'Serum albumin' |
| 4491 | 'ALB_No179' | 'ETCFAEEGKKLVAASQAALGL' | 'Serum albumin' |
| 4492 | 'ALB_No180' | 'EQLKAVMDDFAAFVEKCCK' | 'Serum albumin' |
| 4493 | 'ALB_No181' | 'NLGKVGSK' | 'Serum albumin' |
| 4494 | 'ALB_No182' | 'DAHKSEVAHRFK' | 'Serum albumin' |
| 4495 | 'ALB_No183' | 'CASLQKFGERAFK' | 'Serum albumin' |
| 4496 | 'ALB_No184' | 'LCTVATLRETYGEMADCCAKQEPER' | 'Serum albumin' |
| 4497 | 'ALB_No185' | 'ATKEQLK' | 'Serum albumin' |
| 4498 | 'ALB_No186' | 'SEVAHRFK' | 'Serum albumin' |
| 4499 | 'ALB_No187' | 'ADLAKYICENQDSISSKLK' | 'Serum albumin' |
| 4500 | 'ALB_No188' | 'ECCEKPLLEKSHCIAEVENDEMPADLPSLAADFVESK' | 'Serum albumin' |
| 4501 | 'ALB_No189' | 'PLVEEPQNLIKQNCELFEQLGEYK' | 'Serum albumin' |
| 4502 | 'ALB_No190' | 'DAHKSEVAHR' | 'Serum albumin' |
| 4503 | 'ALB_No191' | 'ECCEKPLLEKSHCIAEVENDEMPADLPSLAADFVESK' | 'Serum albumin' |
| 4504 | 'GC_No1' | 'HLSLLTTLSNR' | 'Vitamin D-binding protein' |
| 4505 | 'GC_No2' | 'VMDKYTFELSR' | 'Vitamin D-binding protein' |
| 4506 | 'GC_No3' | 'VMDKYTFELSR' | 'Vitamin D-binding protein' |
| 4507 | 'GC_No4' | 'EDFTSLSLVLYSR' | 'Vitamin D-binding protein' |
| 4508 | 'GC_No5' | 'GQELCADYSENTFTEYK' | 'Vitamin D-binding protein' |
| 4509 | 'GC_No6' | 'RTHLPEVFLSK' | 'Vitamin D-binding protein' |
| 4510 | 'GC_No7' | 'VCSQYAAYGEK' | 'Vitamin D-binding protein' |
| 4511 | 'GC_No8' | 'YTFELSR' | 'Vitamin D-binding protein' |
| 4512 | 'GC_No9' | 'SCESNSPFPVHPGTAECCTK' | 'Vitamin D-binding protein' |
| 4513 | 'GC_No10' | 'EFSHLGKEDFTSLSLVLYSR' | 'Vitamin D-binding protein' |
| 4514 | 'GC_No11' | 'KFPSGTFEQVSQLVK' | 'Vitamin D-binding protein' |
| 4515 | 'GC_No12' | 'VPTADLEDVLPLAEDITNILSK' | 'Vitamin D-binding protein' |
| 4516 | 'GC_No13' | 'FPSGTFEQVSQLVK' | 'Vitamin D-binding protein' |
| 4517 | 'GC_No14' | 'GQELCADYSENTFTEYKK' | 'Vitamin D-binding protein' |
| 4518 | 'GC_No15' | 'LCDNLSTK' | 'Vitamin D-binding protein' |
| 4519 | 'GC_No16' | 'VCKEFSHLGKEDFTSLSLVLYSR' | 'Vitamin D-binding protein' |
| 4520 | 'GC_No17' | 'FEDCCQEK' | 'Vitamin D-binding protein' |
| 4521 | 'GC_No18' | 'CCESASEDCMAK' | 'Vitamin D-binding protein' |
| 4522 | 'GC_No19' | 'ELSSFIDKGQELCADYSENTFTEYKK' | 'Vitamin D-binding protein' |
| 4523 | 'GC_No20' | 'ELPEHTVK' | 'Vitamin D-binding protein' |
| 4524 | 'GC_No21' | 'THLPEVFLSK' | 'Vitamin D-binding protein' |
| 4525 | 'GC_No22' | 'VMDKYTFELSRR' | 'Vitamin D-binding protein' |
| 4526 | 'GC_No23' | 'SCESNSPFPVHPGTAECCTKEGLER' | 'Vitamin D-binding protein' |
| 4527 | 'GC_No24' | 'CCESASEDCMAKELPEHTVK' | 'Vitamin D-binding protein' |
| 4528 | 'GC_No25' | 'DVCDPGNTK' | 'Vitamin D-binding protein' |
| 4529 | 'GC_No26' | 'LAQKVPTADLEDVLPLAEDITNILSK' | 'Vitamin D-binding protein' |
| 4530 | 'GC_No27' | 'ELPEHTVKLCDNLSTK' | 'Vitamin D-binding protein' |
| 4531 | 'GC_No28' | 'SYLSMVGSCCTSASPTVCFLK' | 'Vitamin D-binding protein' |
| 4532 | 'GC_No29' | 'KELSSFIDK' | 'Vitamin D-binding protein' |
| 4533 | 'GC_No30' | 'HQPQEFPTYVEPTNDEICEAFRK' | 'Vitamin D-binding protein' |

| | A | B | C |
|---|---|---|---|
| 1 | **Peptide Name** | **Peptide Sequence** | **Peptide Description** |
| 4534 | 'GC_No31' | 'NSKFEDCCQEK' | 'Vitamin D-binding protein' |
| 4535 | 'GC_No32' | 'LQLKHLSLLTTLSNR' | 'Vitamin D-binding protein' |
| 4536 | 'GC_No33' | 'EVVSLTEACCAEGADPDCYDTR' | 'Vitamin D-binding protein' |
| 4537 | 'GC_No34' | 'SLGECCDVEDSTTCFNAK' | 'Vitamin D-binding protein' |
| 4538 | 'GC_No35' | 'SYLSMVGSCCTSASPTVCFLK' | 'Vitamin D-binding protein' |
| 4539 | 'GC_No36' | 'VMDKYTFELSRR' | 'Vitamin D-binding protein' |
| 4540 | 'GC_No37' | 'CCESASEDCMAK' | 'Vitamin D-binding protein' |
| 4541 | 'GC_No38' | 'HQPQEFPTYVEPTNDEICEAFR' | 'Vitamin D-binding protein' |
| 4542 | 'GC_No39' | 'AKLPDATPTELAK' | 'Vitamin D-binding protein' |
| 4543 | 'GC_No40' | 'ELSSFIDKGQELCADYSENTFTEYK' | 'Vitamin D-binding protein' |
| 4544 | 'GC_No41' | 'LQLKHLSLLTTLSNRVCSQYAAYGEK' | 'Vitamin D-binding protein' |
| 4545 | 'GC_No42' | 'LPDATPTELAK' | 'Vitamin D-binding protein' |
| 4546 | 'GC_No43' | 'CCESASEDCMAKELPEHTVK' | 'Vitamin D-binding protein' |
| 4547 | 'GC_No44' | 'SCESNSPFPVHPGTAECCTKEGLERK' | 'Vitamin D-binding protein' |
| 4548 | 'GC_No45' | 'LCMAALK' | 'Vitamin D-binding protein' |
| 4549 | 'GC_No46' | 'SYLSMVGSCCTSASPTVCFLKER' | 'Vitamin D-binding protein' |
| 4550 | 'GC_No47' | 'SYLSMVGSCCTSASPTVCFLKER' | 'Vitamin D-binding protein' |
| 4551 | 'GC_No48' | 'GQELCADYSENTFTEYKKK' | 'Vitamin D-binding protein' |
| 4552 | 'GC_No49' | 'SLGECCDVEDSTTCFNAKGPLLKK' | 'Vitamin D-binding protein' |
| 4553 | 'GC_No50' | 'KELSSFIDKGQELCADYSENTFTEYK' | 'Vitamin D-binding protein' |
| 4554 | 'GC_No51' | 'SLGECCDVEDSTTCFNAKGPLLK' | 'Vitamin D-binding protein' |
| 4555 | 'GC_No52' | 'KLCMAALK' | 'Vitamin D-binding protein' |
| 4556 | 'GC_No53' | 'LCDNLSTKNSKFEDCCQEK' | 'Vitamin D-binding protein' |
| 4557 | 'GC_No54' | 'HQPQEFPTYVEPTNDEICEAFRKDPK' | 'Vitamin D-binding protein' |
| 4558 | 'GC_No55' | 'DVCDPGNTKVMDKYTFELSR' | 'Vitamin D-binding protein' |
| 4559 | 'GC_No56' | 'LKAKLPDATPTELAK' | 'Vitamin D-binding protein' |
| 4560 | 'GC_No57' | 'KLCMAALK' | 'Vitamin D-binding protein' |
| 4561 | 'GC_No58' | 'VLEPTLK' | 'Vitamin D-binding protein' |
| 4562 | 'GC_No59' | 'ERLQLKHLSLLTTLSNR' | 'Vitamin D-binding protein' |
| 4563 | 'GC_No60' | 'HLSLLTTLSNRVCSQYAAYGEKK' | 'Vitamin D-binding protein' |
| 4564 | 'GC_No61' | 'EVVSLTEACCAEGADPDCYDTRTSALSAK' | 'Vitamin D-binding protein' |
| 4565 | 'GC_No62' | 'VLEPTLKSLGECCDVEDSTTCFNAK' | 'Vitamin D-binding protein' |
| 4566 | 'GC_No63' | 'HLSLLTTLSNRVCSQYAAYGEK' | 'Vitamin D-binding protein' |
| 4567 | 'GC_No64' | 'THLPEVFLSKVLEPTLK' | 'Vitamin D-binding protein' |
| 4568 | 'GC_No65' | 'CCESASEDCMAKELPEHTVKLCDNLSTK' | 'Vitamin D-binding protein' |
| 4569 | 'GC_No66' | 'CCESASEDCMAKELPEHTVKLCDNLSTK' | 'Vitamin D-binding protein' |
| 4570 | 'GC_No67' | 'TSALSAKSCESNSPFPVHPGTAECCTK' | 'Vitamin D-binding protein' |
| 4571 | 'GC_No68' | 'YTFELSRR' | 'Vitamin D-binding protein' |
| 4572 | 'GC_No69' | 'VCKEFSHLGK' | 'Vitamin D-binding protein' |
| 4573 | 'GC_No70' | 'ELSSFIDK' | 'Vitamin D-binding protein' |
| 4574 | 'GC_No71' | 'ELPEHTVKLCDNLSTKNSK' | 'Vitamin D-binding protein' |
| 4575 | 'PPBP_No1' | 'GTHCNQVEVIATLK' | 'Platelet basic protein' |
| 4576 | 'PPBP_No2' | 'KICLDPDAPR' | 'Platelet basic protein' |
| 4577 | 'PPBP_No3' | 'ICLDPDAPR' | 'Platelet basic protein' |

| | A | B | C |
|---|---|---|---|
| 1 | **Peptide Name** | **Peptide Sequence** | **Peptide Description** |
| 4578 | 'PPBP_No4' | 'NIQSLEVIGK' | 'Platelet basic protein' |
| 4579 | 'PPBP_No5' | 'GTHCNQVEVIATLKDGR' | 'Platelet basic protein' |
| 4580 | 'PPBP_No6' | 'GKEESLDSDLYAELR' | 'Platelet basic protein' |
| 4581 | 'PPBP_No7' | 'GTHCNQVEVIATLKDGRK' | 'Platelet basic protein' |
| 4582 | 'PPBP_No8' | 'EESLDSDLYAELR' | 'Platelet basic protein' |
| 4583 | 'PPBP_No9' | 'NIQSLEVIGKGTHCNQVEVIATLKDGR' | 'Platelet basic protein' |
| 4584 | 'PF4_No1' | 'ICLDLQAPLYK' | 'Platelet factor 4' |
| 4585 | 'PF4_No2' | 'KICLDLQAPLYK' | 'Platelet factor 4' |
| 4586 | 'PF4_No3' | 'KICLDLQAPLYKK' | 'Platelet factor 4' |
| 4587 | 'PF4_No4' | 'ICLDLQAPLYKK' | 'Platelet factor 4' |
| 4588 | 'TFRC_No1' | 'VSASPLLYTLIEK' | 'Transferrin receptor protein 1' |
| 4589 | 'TFRC_No2' | 'ILNIFGVIK' | 'Transferrin receptor protein 1' |
| 4590 | 'TFRC_No3' | 'YNSQLLSFVR' | 'Transferrin receptor protein 1' |
| 4591 | 'TFRC_No4' | 'LFGNMEGDCPSDWK' | 'Transferrin receptor protein 1' |
| 4592 | 'TFRC_No5' | 'LVYLVENPGGYVAYSK' | 'Transferrin receptor protein 1' |
| 4593 | 'TFRC_No6' | 'LAQMFSDMVLK' | 'Transferrin receptor protein 1' |
| 4594 | 'TFRC_No7' | 'LLNENSYVPR' | 'Transferrin receptor protein 1' |
| 4595 | 'TFRC_No8' | 'RLYWDDLKR' | 'Transferrin receptor protein 1' |
| 4596 | 'TFRC_No9' | 'HVFWGSGSHTLPALLENLK' | 'Transferrin receptor protein 1' |
| 4597 | 'TFRC_No10' | 'LTHDVELNLDYER' | 'Transferrin receptor protein 1' |
| 4598 | 'TFRC_No11' | 'GFVEPDHYVVVGAQR' | 'Transferrin receptor protein 1' |
| 4599 | 'TFRC_No12' | 'HPVTGQFLYQDSNWASK' | 'Transferrin receptor protein 1' |
| 4600 | 'TFRC_No13' | 'VEYHFLSPYVSPK' | 'Transferrin receptor protein 1' |
| 4601 | 'TFRC_No14' | 'SSGLPNIPVQTISR' | 'Transferrin receptor protein 1' |
| 4602 | 'TFRC_No15' | 'LFGNMEGDCPSDWKTDSTCR' | 'Transferrin receptor protein 1' |
| 4603 | 'TFRC_No16' | 'LFGNMEGDCPSDWK' | 'Transferrin receptor protein 1' |
| 4604 | 'TFRC_No17' | 'EMGLSLQWLYSAR' | 'Transferrin receptor protein 1' |
| 4605 | 'TFRC_No18' | 'EAGSQKDENLALYVENQFR' | 'Transferrin receptor protein 1' |
| 4606 | 'TFRC_No19' | 'DSAQNSVIIVDKNGR' | 'Transferrin receptor protein 1' |
| 4607 | 'TFRC_No20' | 'LTVSNVLK' | 'Transferrin receptor protein 1' |
| 4608 | 'TFRC_No21' | 'RLYWDDLK' | 'Transferrin receptor protein 1' |
| 4609 | 'TFRC_No22' | 'TMQNVKHPVTGQFLYQDSNWASK' | 'Transferrin receptor protein 1' |
| 4610 | 'TFRC_No23' | 'LTTDFGNAEK' | 'Transferrin receptor protein 1' |
| 4611 | 'TFRC_No24' | 'LSEKLDSTDFTGTIK' | 'Transferrin receptor protein 1' |
| 4612 | 'TFRC_No25' | 'VANAESLNAIGVLIYMDQTK' | 'Transferrin receptor protein 1' |
| 4613 | 'TFRC_No26' | 'LYWDDLKR' | 'Transferrin receptor protein 1' |
| 4614 | 'TFRC_No27' | 'IQVKDSAQNSVIIVDK' | 'Transferrin receptor protein 1' |
| 4615 | 'TFRC_No28' | 'AFTYINLDKAVLGTSNFK' | 'Transferrin receptor protein 1' |
| 4616 | 'TFRC_No29' | 'KLSEKLDSTDFTGTIK' | 'Transferrin receptor protein 1' |
| 4617 | 'TFRC_No30' | 'SGVGTALLLK' | 'Transferrin receptor protein 1' |
| 4618 | 'TFRC_No31' | 'AFTYINLDK' | 'Transferrin receptor protein 1' |
| 4619 | 'TFRC_No32' | 'LAVDEEENADNNTK' | 'Transferrin receptor protein 1' |
| 4620 | 'TFRC_No33' | 'VANAESLNAIGVLIYMDQTK' | 'Transferrin receptor protein 1' |
| 4621 | 'TFRC_No34' | 'LFGNMEGDCPSDWKTDSTCR' | 'Transferrin receptor protein 1' |

| | A | B | C |
|---|---|---|---|
| 1 | **Peptide Name** | **Peptide Sequence** | **Peptide Description** |
| 4622 | 'TFRC_No35' | 'AAAEKLFGNMEGDCPSDWKTDSTCR' | 'Transferrin receptor protein 1' |
| 4623 | 'TFRC_No36' | 'LTTDFGNAEKTDR' | 'Transferrin receptor protein 1' |
| 4624 | 'TFRC_No37' | 'EAGSQKDENLALYVENQFREFK' | 'Transferrin receptor protein 1' |
| 4625 | 'TFRC_No38' | 'DENLALYVENQFR' | 'Transferrin receptor protein 1' |
| 4626 | 'TF_No1' | 'EFQLFSSPHGK' | 'Serotransferrin' |
| 4627 | 'TF_No2' | 'EGYYGYTGAFR' | 'Serotransferrin' |
| 4628 | 'TF_No3' | 'KPVDEYKDCHLAQVPSHTVVAR' | 'Serotransferrin' |
| 4629 | 'TF_No4' | 'LKCDEWSVNSVGK' | 'Serotransferrin' |
| 4630 | 'TF_No5' | 'DLLFRDDTVCLAK' | 'Serotransferrin' |
| 4631 | 'TF_No6' | 'YLGEEYVK' | 'Serotransferrin' |
| 4632 | 'TF_No7' | 'EDPQTFYYAVAVVK' | 'Serotransferrin' |
| 4633 | 'TF_No8' | 'SASDLTWDNLK' | 'Serotransferrin' |
| 4634 | 'TF_No9' | 'CLVEKGDVAFVK' | 'Serotransferrin' |
| 4635 | 'TF_No10' | 'FDEFFSEGCAPGSKK' | 'Serotransferrin' |
| 4636 | 'TF_No11' | 'SMGGKEDLIWELLNQAQEHFGK' | 'Serotransferrin' |
| 4637 | 'TF_No12' | 'TAGWNIPMGLLYNK' | 'Serotransferrin' |
| 4638 | 'TF_No13' | 'SKEFQLFSSPHGK' | 'Serotransferrin' |
| 4639 | 'TF_No14' | 'SMGGKEDLIWELLNQAQEHFGKDK' | 'Serotransferrin' |
| 4640 | 'TF_No15' | 'DCHLAQVPSHTVVAR' | 'Serotransferrin' |
| 4641 | 'TF_No16' | 'SETKDLLFRDDTVCLAK' | 'Serotransferrin' |
| 4642 | 'TF_No17' | 'PVDEYKDCHLAQVPSHTVVAR' | 'Serotransferrin' |
| 4643 | 'TF_No18' | 'WCAVSEHEATK' | 'Serotransferrin' |
| 4644 | 'TF_No19' | 'KSASDLTWDNLK' | 'Serotransferrin' |
| 4645 | 'TF_No20' | 'IMNGEADAMSLDGGFVYIAGK' | 'Serotransferrin' |
| 4646 | 'TF_No21' | 'HSTIFENLANKADR' | 'Serotransferrin' |
| 4647 | 'TF_No22' | 'DSGFQMNQLR' | 'Serotransferrin' |
| 4648 | 'TF_No23' | 'DSAHGFLK' | 'Serotransferrin' |
| 4649 | 'TF_No24' | 'MYLGYEYVTAIR' | 'Serotransferrin' |
| 4650 | 'TF_No25' | 'HQTVPQNTGGKNPDPWAK' | 'Serotransferrin' |
| 4651 | 'TF_No26' | 'CLKDGAGDVAFVK' | 'Serotransferrin' |
| 4652 | 'TF_No27' | 'DLLFKDSAHGFLK' | 'Serotransferrin' |
| 4653 | 'TF_No28' | 'KDSGFQMNQLR' | 'Serotransferrin' |
| 4654 | 'TF_No29' | 'SETKDLLFR' | 'Serotransferrin' |
| 4655 | 'TF_No30' | 'TAGWNIPMGLLYNK' | 'Serotransferrin' |
| 4656 | 'TF_No31' | 'HSTIFENLANK' | 'Serotransferrin' |
| 4657 | 'TF_No32' | 'HSTIFENLANKADRDQYELLCLDNTR' | 'Serotransferrin' |
| 4658 | 'TF_No33' | 'SMGGKEDLIWELLNQAQEHFGK' | 'Serotransferrin' |
| 4659 | 'TF_No34' | 'EDLIWELLNQAQEHFGK' | 'Serotransferrin' |
| 4660 | 'TF_No35' | 'DQYELLCLDNTR' | 'Serotransferrin' |
| 4661 | 'TF_No36' | 'QQQHLFGSNVTDCSGNFCLFR' | 'Serotransferrin' |
| 4662 | 'TF_No37' | 'EFQLFSSPHGKDLLFK' | 'Serotransferrin' |
| 4663 | 'TF_No38' | 'AIAANEADAVTLDAGLVYDAYLAPNNLKPVVAEFYGSK' | 'Serotransferrin' |
| 4664 | 'TF_No39' | 'CDEWSVNSVGK' | 'Serotransferrin' |
| 4665 | 'TF_No40' | 'AIAANEADAVTLDAGLVYDAYLAPNNLKPVVAEFYGSKEDPQTFYYAVAV | 'Serotransferrin' |

| | A | B | C |
|---|---|---|---|
| 1 | **Peptide Name** | **Peptide Sequence** | **Peptide Description** |
| 4666 | 'TF_No41' | 'WCALSHHER' | 'Serotransferrin' |
| 4667 | 'TF_No42' | 'IMNGEADAMSLDGGFVYIAGK' | 'Serotransferrin' |
| 4668 | 'TF_No43' | 'SASDLTWDNLKGK' | 'Serotransferrin' |
| 4669 | 'TF_No44' | 'EGTCPEAPTDECK' | 'Serotransferrin' |
| 4670 | 'TF_No45' | 'KPVEEYANCHLAR' | 'Serotransferrin' |
| 4671 | 'TF_No46' | 'KDSGFQMNQLR' | 'Serotransferrin' |
| 4672 | 'TF_No47' | 'SKEFQLFSSPHGKDLLFK' | 'Serotransferrin' |
| 4673 | 'TF_No48' | 'EGTCPEAPTDECKPVKWCALSHHER' | 'Serotransferrin' |
| 4674 | 'TF_No49' | 'APNHAVVTR' | 'Serotransferrin' |
| 4675 | 'TF_No50' | 'DSGFQMNQLR' | 'Serotransferrin' |
| 4676 | 'TF_No51' | 'EDLIWELLNQAQEHFGKDK' | 'Serotransferrin' |
| 4677 | 'TF_No52' | 'NPDPWAK' | 'Serotransferrin' |
| 4678 | 'TF_No53' | 'LCMGSGLNLCEPNNKEGYYGYTGAFR' | 'Serotransferrin' |
| 4679 | 'TF_No54' | 'SAGWNIPIGLLYCDLPEPR' | 'Serotransferrin' |
| 4680 | 'TF_No55' | 'DDTVCLAK' | 'Serotransferrin' |
| 4681 | 'TF_No56' | 'PVVAEFYGSK' | 'Serotransferrin' |
| 4682 | 'TF_No57' | 'NLREGTCPEAPTDECKPVK' | 'Serotransferrin' |
| 4683 | 'TF_No58' | 'HQTVPQNTGGK' | 'Serotransferrin' |
| 4684 | 'TF_No59' | 'TAGWNIPMGLLYNKINHCRFDEFFSEGCAPGSK' | 'Serotransferrin' |
| 4685 | 'TF_No60' | 'EFQLFSSPHGKDLLFKDSAHGFLK' | 'Serotransferrin' |
| 4686 | 'TF_No61' | 'IMNGEADAMSLDGGFVYIAGK' | 'Serotransferrin' |
| 4687 | 'TF_No62' | 'IMNGEADAMSLDGGFVYIAGK' | 'Serotransferrin' |
| 4688 | 'TF_No63' | 'DSAHGFLKVPPR' | 'Serotransferrin' |
| 4689 | 'TF_No64' | 'CLVEKGDVAFVKHQTVPQNTGGK' | 'Serotransferrin' |
| 4690 | 'TF_No65' | 'EGTCPEAPTDECKPVK' | 'Serotransferrin' |
| 4691 | 'TF_No66' | 'DYELLCLDGTR' | 'Serotransferrin' |
| 4692 | 'TF_No67' | 'EDPQTFYYAVAVVKK' | 'Serotransferrin' |
| 4693 | 'TF_No68' | 'DGAGDVAFVK' | 'Serotransferrin' |
| 4694 | 'TF_No69' | 'INHCRFDEFFSEGCAPGSK' | 'Serotransferrin' |
| 4695 | 'TF_No70' | 'KASYLDCIR' | 'Serotransferrin' |
| 4696 | 'TF_No71' | 'SVIPSDGPSVACVKK' | 'Serotransferrin' |
| 4697 | 'TF_No72' | 'MYLGYEYVTAIR' | 'Serotransferrin' |
| 4698 | 'TF_No73' | 'ADRDQYELLCLDNTR' | 'Serotransferrin' |
| 4699 | 'TF_No74' | 'SMGGKEDLIWELLNQAQEHFGKDK' | 'Serotransferrin' |
| 4700 | 'TF_No75' | 'TAGWNIPMGLLYNKINHCR' | 'Serotransferrin' |
| 4701 | 'TF_No76' | 'INHCRFDEFFSEGCAPGSKK' | 'Serotransferrin' |
| 4702 | 'TF_No77' | 'CSTSSLLEACTFRRP' | 'Serotransferrin' |
| 4703 | 'TF_No78' | 'WCAVSEHEATKCQSFRDHMK' | 'Serotransferrin' |
| 4704 | 'TF_No79' | 'LHDRNTYEKYLGEEYVK' | 'Serotransferrin' |
| 4705 | 'TF_No80' | 'GDVAFVKHQTVPQNTGGK' | 'Serotransferrin' |
| 4706 | 'TF_No81' | 'WCAVSEHEATKCQSFR' | 'Serotransferrin' |
| 4707 | 'TF_No82' | 'GDVAFVKHQTVPQNTGGKNPDPWAK' | 'Serotransferrin' |
| 4708 | 'TF_No83' | 'NTYEKYLGEEYVK' | 'Serotransferrin' |
| 4709 | 'TF_No84' | 'IECVSAETTEDCIAK' | 'Serotransferrin' |

| | A | B | C |
|---|---|---|---|
| 1 | **Peptide Name** | **Peptide Sequence** | **Peptide Description** |
| 4710 | 'TF_No85' | 'KCSTSSLLEACTFR' | 'Serotransferrin' |
| 4711 | 'TF_No86' | 'PVKWCALSHHER' | 'Serotransferrin' |
| 4712 | 'TF_No87' | 'DKSKEFQLFSSPHGK' | 'Serotransferrin' |
| 4713 | 'TF_No88' | 'SCHTGLGR' | 'Serotransferrin' |
| 4714 | 'TF_No89' | 'NLNEKDYELLCLDGTR' | 'Serotransferrin' |
| 4715 | 'TF_No90' | 'CDEWSVNSVGKIECVSAETTEDCIAK' | 'Serotransferrin' |
| 4716 | 'TF_No91' | 'ASYLDCIR' | 'Serotransferrin' |
| 4717 | 'TF_No92' | 'DLLFKDSAHGFLKVPPR' | 'Serotransferrin' |
| 4718 | 'TF_No93' | 'SVIPSDGPSVACVK' | 'Serotransferrin' |
| 4719 | 'TF_No94' | 'LCMGSGLNLCEPNNK' | 'Serotransferrin' |
| 4720 | 'TF_No95' | 'SASDLTWDNLKGKK' | 'Serotransferrin' |
| 4721 | 'TF_No96' | 'LCMGSGLNLCEPNNKEGYYGYTGAFR' | 'Serotransferrin' |
| 4722 | 'TF_No97' | 'LCMGSGLNLCEPNNK' | 'Serotransferrin' |
| 4723 | 'TF_No98' | 'CSTSSLLEACTFR' | 'Serotransferrin' |
| 4724 | 'TF_No99' | 'KSASDLTWDNLKGK' | 'Serotransferrin' |
| 4725 | 'TF_No100' | 'AIAANEADAVTLDAGLVYDAYLAPNNLK' | 'Serotransferrin' |
| 4726 | 'TF_No101' | 'FDEFFSEGCAPGSKKDSSLCK' | 'Serotransferrin' |
| 4727 | 'TF_No102' | 'YLGEEYVKAVGNLR' | 'Serotransferrin' |
| 4728 | 'TF_No103' | 'FDEFFSEGCAPGSK' | 'Serotransferrin' |
| 4729 | 'TF_No104' | 'QQQHLFGSNVTDCSGNFCLFR' | 'Serotransferrin' |
| 4730 | 'TF_No105' | 'DGAGDVAFVKHSTIFENLANK' | 'Serotransferrin' |
| 4731 | 'TF_No106' | 'NLNEKDYELLCLDGTRK' | 'Serotransferrin' |
| 4732 | 'TF_No107' | 'DYELLCLDGTRKPVEEYANCHLAR' | 'Serotransferrin' |
| 4733 | 'TF_No108' | 'LKCDEWSVNSVGKIECVSAETTEDCIAK' | 'Serotransferrin' |
| 4734 | 'TF_No109' | 'NPDPWAKNLNEKDYELLCLDGTR' | 'Serotransferrin' |
| 4735 | 'TF_No110' | 'CGLVPVLAENYNK' | 'Serotransferrin' |
| 4736 | 'TF_No111' | 'PVEEYANCHLAR' | 'Serotransferrin' |
| 4737 | 'TF_No112' | 'NLREGTCPEAPTDECK' | 'Serotransferrin' |
| 4738 | 'TF_No113' | 'ADRDQYELLCLDNTRK' | 'Serotransferrin' |
| 4739 | 'TF_No114' | 'DYELLCLDGTRK' | 'Serotransferrin' |
| 4740 | 'TF_No115' | 'WCAVSEHEATKCQSFRDHMK' | 'Serotransferrin' |
| 4741 | 'TF_No116' | 'EDPQTFYYAVAVVKKDSGFQMNQLR' | 'Serotransferrin' |
| 4742 | 'TF_No117' | 'YLGEEYVKAVGNLRK' | 'Serotransferrin' |
| 4743 | 'TF_No118' | 'DHMKSVIPSDGPSVACVK' | 'Serotransferrin' |
| 4744 | 'TF_No119' | 'DSAHGFLKVPPRMDAK' | 'Serotransferrin' |
| 4745 | 'TF_No120' | 'PVVAEFYGSKEDPQTFYYAVAVVKK' | 'Serotransferrin' |
| 4746 | 'TF_No121' | 'MYLGYEYVTAIRNLR' | 'Serotransferrin' |
| 4747 | 'TF_No122' | 'CQSFRDHMK' | 'Serotransferrin' |
| 4748 | 'TF_No123' | 'MYLGYEYVTAIRNLR' | 'Serotransferrin' |
| 4749 | 'TF_No124' | 'EDPQTFYYAVAVVKKDSGFQMNQLR' | 'Serotransferrin' |
| 4750 | 'TF_No125' | 'DSAHGFLKVPPRMDAK' | 'Serotransferrin' |
| 4751 | 'TF_No126' | 'AVGNLRKCSTSSLLEACTFR' | 'Serotransferrin' |
| 4752 | 'TF_No127' | 'DKEACVHKILR' | 'Serotransferrin' |
| 4753 | 'TF_No128' | 'SDNCEDTPEAGYFAIAVVK' | 'Serotransferrin' |

| | A | B | C |
|---|---|---|---|
| 1 | **Peptide Name** | **Peptide Sequence** | **Peptide Description** |
| 4754 | 'TF_No129' | 'SAGWNIPIGLLYCDLPEPRKPLEK' | 'Serotransferrin' |
| 4755 | 'TF_No130' | 'DLLFRDDTVCLAKLHDR' | 'Serotransferrin' |
| 4756 | 'TF_No131' | 'SVIPSDGPSVACVKKASYLDCIR' | 'Serotransferrin' |
| 4757 | 'TF_No132' | 'CLKDGAGDVAFVKHSTIFENLANK' | 'Serotransferrin' |
| 4758 | 'TF_No133' | 'DSGFQMNQLRGK' | 'Serotransferrin' |
| 4759 | 'TF_No134' | 'DSSLCKLCMGSGLNLCEPNNKEGYYGYTGAFR' | 'Serotransferrin' |
| 4760 | 'TF_No135' | 'DQYELLCLDNTRKPVDEYK' | 'Serotransferrin' |
| 4761 | 'TF_No136' | 'DHMKSVIPSDGPSVACVKK' | 'Serotransferrin' |
| 4762 | 'TF_No137' | 'MDAKMYLGYEYVTAIR' | 'Serotransferrin' |
| 4763 | 'TF_No138' | 'DGAGDVAFVKHSTIFENLANKADR' | 'Serotransferrin' |
| 4764 | 'TF_No139' | 'KPVDEYK' | 'Serotransferrin' |
| 4765 | 'TF_No140' | 'ILRQQQHLFGSNVTDCSGNFCLFR' | 'Serotransferrin' |
| 4766 | 'TF_No141' | 'KDSSLCKLCMGSGLNLCEPNNK' | 'Serotransferrin' |
| 4767 | 'TF_No142' | 'SDNCEDTPEAGYFAIAVVKK' | 'Serotransferrin' |
| 4768 | 'TF_No143' | 'DHMKSVIPSDGPSVACVK' | 'Serotransferrin' |
| 4769 | 'TF_No144' | 'KDSSLCK' | 'Serotransferrin' |
| 4770 | 'TF_No145' | 'PVVAEFYGSKEDPQTFYYAVAVVK' | 'Serotransferrin' |
| 4771 | 'TF_No146' | 'SCHTAVGR' | 'Serotransferrin' |
| 4772 | 'TF_No147' | 'HQTVPQNTGGKNPDPWAKNLNEK' | 'Serotransferrin' |
| 4773 | 'TF_No148' | 'KSCHTAVGR' | 'Serotransferrin' |
| 4774 | 'TF_No149' | 'APNHAVVTRK' | 'Serotransferrin' |
| 4775 | 'TF_No150' | 'LHDRNTYEK' | 'Serotransferrin' |
| 4776 | 'TF_No151' | 'TAGWNIPMGLLYNKINHCR' | 'Serotransferrin' |
| 4777 | 'TF_No152' | 'DSSLCKLCMGSGLNLCEPNNKEGYYGYTGAFR' | 'Serotransferrin' |
| 4778 | 'TF_No153' | 'KDSSLCKLCMGSGLNLCEPNNK' | 'Serotransferrin' |
| 4779 | 'TF_No154' | 'TVRWCAVSEHEATK' | 'Serotransferrin' |
| 4780 | 'TF_No155' | 'DKEACVHK' | 'Serotransferrin' |
| 4781 | 'TF_No156' | 'DSGFQMNQLRGK' | 'Serotransferrin' |
| 4782 | 'TF_No157' | 'KSCHTGLGR' | 'Serotransferrin' |
| 4783 | 'TF_No158' | 'CQSFRDHMK' | 'Serotransferrin' |
| 4784 | 'TF_No159' | 'GDVAFVK' | 'Serotransferrin' |
| 4785 | 'LTF_No1' | 'YYGYTGAFR' | 'Lactotransferrin' |
| 4786 | 'LTF_No2' | 'DVTVLQNTDGNNNEAWAK' | 'Lactotransferrin' |
| 4787 | 'LTF_No3' | 'SNLCALCIGDEQGENK' | 'Lactotransferrin' |
| 4788 | 'LTF_No4' | 'LADFALLCLDGK' | 'Lactotransferrin' |
| 4789 | 'LTF_No5' | 'FDEYFSQSCAPGSDPR' | 'Lactotransferrin' |
| 4790 | 'LTF_No6' | 'IDSGLYLGSGYFTAIQNLR' | 'Lactotransferrin' |
| 4791 | 'LTF_No7' | 'YLGPQYVAGITNLK' | 'Lactotransferrin' |
| 4792 | 'LTF_No8' | 'ADAVTLDGGFIYEAGLAPYK' | 'Lactotransferrin' |
| 4793 | 'LTF_No9' | 'SCHLAMAPNHAVVSR' | 'Lactotransferrin' |
| 4794 | 'LTF_No10' | 'CAFSSQEPYFSYSGAFK' | 'Lactotransferrin' |
| 4795 | 'LTF_No11' | 'YLGPQYVAGITNLKK' | 'Lactotransferrin' |
| 4796 | 'LTF_No12' | 'KGGSFQLNELQGLK' | 'Lactotransferrin' |
| 4797 | 'LTF_No13' | 'RSDTSLTWNSVK' | 'Lactotransferrin' |

| | A | B | C |
|---|---|---|---|
| 1 | **Peptide Name** | **Peptide Sequence** | **Peptide Description** |
| 4798 | 'LTF_No14' | 'NGSDCPDKFCLFQSETK' | 'Lactotransferrin' |
| 4799 | 'LTF_No15' | 'GEADAMSLDGGYVYTAGK' | 'Lactotransferrin' |
| 4800 | 'LTF_No16' | 'FFSASCVPGADKGQFPNLCR' | 'Lactotransferrin' |
| 4801 | 'LTF_No17' | 'SVNGKEDAIWNLLR' | 'Lactotransferrin' |
| 4802 | 'LTF_No18' | 'SCHLAMAPNHAVVSR' | 'Lactotransferrin' |
| 4803 | 'LTF_No19' | 'LRPVAAEVYGTER' | 'Lactotransferrin' |
| 4804 | 'LTF_No20' | 'DSPIQCIQAIAENR' | 'Lactotransferrin' |
| 4805 | 'LTF_No21' | 'SQQSSDPDPNCVDRPVEGYLAVAVVR' | 'Lactotransferrin' |
| 4806 | 'LTF_No22' | 'CGLVPVLAENYK' | 'Lactotransferrin' |
| 4807 | 'LTF_No23' | 'VVWCAVGEQELR' | 'Lactotransferrin' |
| 4808 | 'LTF_No24' | 'CLAENAGDVAFVK' | 'Lactotransferrin' |
| 4809 | 'LTF_No25' | 'GEADAMSLDGGYVYTAGK' | 'Lactotransferrin' |
| 4810 | 'LTF_No26' | 'FQLFGSPSGQK' | 'Lactotransferrin' |
| 4811 | 'LTF_No27' | 'VPPRIDSGLYLGSGYFTAIQNLR' | 'Lactotransferrin' |
| 4812 | 'LTF_No28' | 'NLLFNDNTECLAR' | 'Lactotransferrin' |
| 4813 | 'LTF_No29' | 'CSTSPLLEACEFLR' | 'Lactotransferrin' |
| 4814 | 'HPX_No1' | 'LYLVQGTQVYVFLTK' | 'Hemopexin' |
| 4815 | 'HPX_No2' | 'WKNFPSPVDAAFR' | 'Hemopexin' |
| 4816 | 'HPX_No3' | 'SLGPNSCSANGPGLYLIHGPNLYCYSDVEKLNAAK' | 'Hemopexin' |
| 4817 | 'HPX_No4' | 'SGAQATWTELPWPHEK' | 'Hemopexin' |
| 4818 | 'HPX_No5' | 'FDPVRGEVPPR' | 'Hemopexin' |
| 4819 | 'HPX_No6' | 'GGYTLVSGYPK' | 'Hemopexin' |
| 4820 | 'HPX_No7' | 'ALPQPQNVTSLLGCTH' | 'Hemopexin' |
| 4821 | 'HPX_No8' | 'GDKVWVYPPEK' | 'Hemopexin' |
| 4822 | 'HPX_No9' | 'SGAQATWTELPWPHEKVDGALCMEK' | 'Hemopexin' |
| 4823 | 'HPX_No10' | 'FDPVRGEVPPRYPR' | 'Hemopexin' |
| 4824 | 'HPX_No11' | 'SGAQATWTELPWPHEKVDGALCMEK' | 'Hemopexin' |
| 4825 | 'HPX_No12' | 'VDGALCMEK' | 'Hemopexin' |
| 4826 | 'HPX_No13' | 'GDKVWVYPPEKK' | 'Hemopexin' |
| 4827 | 'HPX_No14' | 'DVRDYFMPCPGR' | 'Hemopexin' |
| 4828 | 'HPX_No15' | 'QGHNSVFLIK' | 'Hemopexin' |
| 4829 | 'HPX_No16' | 'EWFWDLATGTMK' | 'Hemopexin' |
| 4830 | 'HPX_No17' | 'DYFMPCPGR' | 'Hemopexin' |
| 4831 | 'HPX_No18' | 'VWVYPPEKK' | 'Hemopexin' |
| 4832 | 'HPX_No19' | 'WDRELISER' | 'Hemopexin' |
| 4833 | 'HPX_No20' | 'LLQDEFPGIPSPLDAAVECHR' | 'Hemopexin' |
| 4834 | 'HPX_No21' | 'VWVYPPEK' | 'Hemopexin' |
| 4835 | 'HPX_No22' | 'EVGTPHGIILDSVDAAFICPGSSR' | 'Hemopexin' |
| 4836 | 'HPX_No23' | 'SHKWDRELISER' | 'Hemopexin' |
| 4837 | 'HPX_No24' | 'QGHNSVFLIK' | 'Hemopexin' |
| 4838 | 'HPX_No25' | 'LWWLDLK' | 'Hemopexin' |
| 4839 | 'HPX_No26' | 'EWFWDLATGTMKER' | 'Hemopexin' |
| 4840 | 'HPX_No27' | 'NFPSPVDAAFR' | 'Hemopexin' |
| 4841 | 'HPX_No28' | 'CSPHLVLSALTSDNHGATYAFSGTHYWR' | 'Hemopexin' |

| | A | B | C |
|---|---|---|---|
| 1 | **Peptide Name** | **Peptide Sequence** | **Peptide Description** |
| 4842 | 'HPX_No29' | 'DVRDYFMPCPGR' | 'Hemopexin' |
| 4843 | 'HPX_No30' | 'RLWWLDLK' | 'Hemopexin' |
| 4844 | 'HPX_No31' | 'EWFWDLATGTMK' | 'Hemopexin' |
| 4845 | 'HPX_No32' | 'GEVPPRYPR' | 'Hemopexin' |
| 4846 | 'HPX_No33' | 'QGHNSVFLIKGDK' | 'Hemopexin' |
| 4847 | 'HPX_No34' | 'VDGALCMEK' | 'Hemopexin' |
| 4848 | 'HPX_No35' | 'LLQDEFPGIPSPLDAAVECHRGECQAEGVLFFQGDR' | 'Hemopexin' |
| 4849 | 'HPX_No36' | 'GECQAEGVLFFQGDR' | 'Hemopexin' |
| 4850 | 'HPX_No37' | 'DYFMPCPGR' | 'Hemopexin' |
| 4851 | 'HPX_No38' | 'LEKEVGTPHGIILDSVDAAFICPGSSR' | 'Hemopexin' |
| 4852 | 'HPX_No39' | 'YYCFQGNQFLR' | 'Hemopexin' |
| 4853 | 'HPX_No40' | 'SLGPNSCSANGPGLYLIHGPNLYCYSDVEK' | 'Hemopexin' |
| 4854 | 'HPX_No41' | 'SWPAVGNCSSALR' | 'Hemopexin' |
| 4855 | 'HPX_No42' | 'DVRDYFMPCPGRGHGHR' | 'Hemopexin' |
| 4856 | 'HPX_No43' | 'DYFMPCPGRGHGHR' | 'Hemopexin' |
| 4857 | 'HPX_No44' | 'ELISERWK' | 'Hemopexin' |
| 4858 | 'HPX_No45' | 'WLGRYYCFQGNQFLR' | 'Hemopexin' |
| 4859 | 'HPX_No46' | 'RLEKEVGTPHGIILDSVDAAFICPGSSR' | 'Hemopexin' |
| 4860 | 'HPX_No47' | 'YYCFQGNQFLRFDPVR' | 'Hemopexin' |
| 4861 | 'HPX_No48' | 'GGYTLVSGYPKRLEK' | 'Hemopexin' |
| 4862 | 'HPX_No49' | 'DYFMPCPGRGHGHR' | 'Hemopexin' |
| 4863 | 'HPX_No50' | 'QGHNSVFLIKGDK' | 'Hemopexin' |
| 4864 | 'HPX_No51' | 'DVRDYFMPCPGRGHGHR' | 'Hemopexin' |
| 4865 | 'HPX_No52' | 'DGWHSWPIAHQWPQGPSAVDAAFSWEEK' | 'Hemopexin' |
| 4866 | 'HPX_No53' | 'PDPDVTER' | 'Hemopexin' |
| 4867 | 'HPX_No54' | 'NFPSPVDAAFRQGHNSVFLIK' | 'Hemopexin' |
| 4868 | 'HPX_No55' | 'ELISERWKNFPSPVDAAFR' | 'Hemopexin' |
| 4869 | 'HPX_No56' | 'QGHNSVFLIKGDKVWVYPPEK' | 'Hemopexin' |
| 4870 | 'HPX_No57' | 'QGHNSVFLIKGDKVWVYPPEK' | 'Hemopexin' |
| 4871 | 'HPX_No58' | 'EWFWDLATGTMKER' | 'Hemopexin' |
| 4872 | 'HPX_No59' | 'WDRELISERWK' | 'Hemopexin' |
| 4873 | 'FTL_No1' | 'KLNQALLDLHALGSAR' | 'Ferritin light chain' |
| 4874 | 'FTL_No2' | 'TDPHLCDFLETHFLDEEVK' | 'Ferritin light chain' |
| 4875 | 'FTL_No3' | 'LGGPEAGLGEYLFER' | 'Ferritin light chain' |
| 4876 | 'FTL_No4' | 'KPAEDEWGKTPDAMK' | 'Ferritin light chain' |
| 4877 | 'FTL_No5' | 'LNQALLDLHALGSAR' | 'Ferritin light chain' |
| 4878 | 'FTL_No6' | 'KPAEDEWGKTPDAMK' | 'Ferritin light chain' |
| 4879 | 'ANG_No1' | 'NVVVACENGLPVHLDQSIFR' | 'Angiogenin' |
| 4880 | 'ANG_No2' | 'YTHFLTQHYDAKPQGR' | 'Angiogenin' |
| 4881 | 'ANG_No3' | 'DDRYCESIMR' | 'Angiogenin' |
| 4882 | 'ANG_No4' | 'LHGGSPWPPCQYR' | 'Angiogenin' |
| 4883 | 'ANG_No5' | 'DINTFIHGNKR' | 'Angiogenin' |
| 4884 | 'ANG_No6' | 'NVVVACENGLPVHLDQSIFRRP' | 'Angiogenin' |
| 4885 | 'ANG_No7' | 'GLTSPCKDINTFIHGNKR' | 'Angiogenin' |

| | A | B | C |
|---|---|---|---|
| 1 | **Peptide Name** | **Peptide Sequence** | **Peptide Description** |
| 4886 | 'ANG_No8' | 'DDRYCESIMR' | 'Angiogenin' |
| 4887 | 'ANG_No9' | 'YTHFLTQHYDAKPQGRDDR' | 'Angiogenin' |
| 4888 | 'ANG_No10' | 'YTHFLTQHYDAK' | 'Angiogenin' |
| 4889 | 'F11_No1' | 'SCALSNLACIR' | 'Coagulation factor XI' |
| 4890 | 'F11_No2' | 'ICTHHPGCLFFTFFSQEWPK' | 'Coagulation factor XI' |
| 4891 | 'F11_No3' | 'YCQVVCTYHPR' | 'Coagulation factor XI' |
| 4892 | 'F11_No4' | 'HLCGGSIIGNQWILTAAHCFYGVESPK' | 'Coagulation factor XI' |
| 4893 | 'F11_No5' | 'IPLVTNEECQK' | 'Coagulation factor XI' |
| 4894 | 'F11_No6' | 'MAESGYDIALLK' | 'Coagulation factor XI' |
| 4895 | 'F11_No7' | 'ALSGFSLQSCR' | 'Coagulation factor XI' |
| 4896 | 'F11_No8' | 'DTCFEGGDITTVFTPSAK' | 'Coagulation factor XI' |
| 4897 | 'F11_No9' | 'TAAISGYSFK' | 'Coagulation factor XI' |
| 4898 | 'F11_No10' | 'NVIYTDCWVTGWGYR' | 'Coagulation factor XI' |
| 4899 | 'F11_No11' | 'GEWPWQVTLHTTSPTQR' | 'Coagulation factor XI' |
| 4900 | 'F11_No12' | 'HSIPVFCHSSFYHDTDFLGEELDIVAAK' | 'Coagulation factor XI' |
| 4901 | 'F11_No13' | 'AKIPLVTNEECQK' | 'Coagulation factor XI' |
| 4902 | 'F11_No14' | 'QFPSLEHR' | 'Coagulation factor XI' |
| 4903 | 'F11_No15' | 'QFPSLEHR' | 'Coagulation factor XI' |
| 4904 | 'F11_No16' | 'HNEVWHLVGITSWGEGCAQR' | 'Coagulation factor XI' |
| 4905 | 'F11_No17' | 'CTDDVHCHFFTYATR' | 'Coagulation factor XI' |
| 4906 | 'F11_No18' | 'AKIPLVTNEECQKR' | 'Coagulation factor XI' |
| 4907 | 'F11_No19' | 'IPLVTNEECQKR' | 'Coagulation factor XI' |
| 4908 | 'F11_No20' | 'MAESGYDIALLK' | 'Coagulation factor XI' |
| 4909 | 'F11_No21' | 'LDKVVSGFSLK' | 'Coagulation factor XI' |
| 4910 | 'F11_No22' | 'DIFPNTVFADSNIDSVMAPDAFVCGR' | 'Coagulation factor XI' |
| 4911 | 'F11_No23' | 'NVIYTDCWVTGWGYRK' | 'Coagulation factor XI' |
| 4912 | 'F11_No24' | 'ITKLDKVVSGFSLK' | 'Coagulation factor XI' |
| 4913 | 'F11_No25' | 'QCSHQISACNKDIYVDLDMK' | 'Coagulation factor XI' |
| 4914 | 'F11_No26' | 'QCSHQISACNKDIYVDLDMK' | 'Coagulation factor XI' |
| 4915 | 'F11_No27' | 'WFTCVLKDSVTETLPR' | 'Coagulation factor XI' |
| 4916 | 'F11_No28' | 'IVGGTASVR' | 'Coagulation factor XI' |
| 4917 | 'F11_No29' | 'WFTCVLK' | 'Coagulation factor XI' |
| 4918 | 'F11_No30' | 'LCTNAVR' | 'Coagulation factor XI' |
| 4919 | 'F11_No31' | 'SAQECQERCTDDVHCHFFTYATR' | 'Coagulation factor XI' |
| 4920 | 'F11_No32' | 'QCSHQISACNKDIYVDLDMK' | 'Coagulation factor XI' |
| 4921 | 'F11_No33' | 'DSVTETLPR' | 'Coagulation factor XI' |
| 4922 | 'F11_No34' | 'EDTSFFGVQEIIIHDQYK' | 'Coagulation factor XI' |
| 4923 | 'F11_No35' | 'SKALSGFSLQSCR' | 'Coagulation factor XI' |
| 4924 | 'F11_No36' | 'LETTVNYTDSQRPICLPSKGDR' | 'Coagulation factor XI' |
| 4925 | 'F11_No37' | 'LETTVNYTDSQRPICLPSK' | 'Coagulation factor XI' |
| 4926 | 'KLKB1_No1' | 'VNIPLVTNEECQK' | 'Plasma kallikrein' |
| 4927 | 'KLKB1_No2' | 'IAYGTQGSSGYSLR' | 'Plasma kallikrein' |
| 4928 | 'KLKB1_No3' | 'IYSGILNLSDITK' | 'Plasma kallikrein' |
| 4929 | 'KLKB1_No4' | 'VAEYMDWILEK' | 'Plasma kallikrein' |

| | A | B | C |
|---|---|---|---|
| 1 | **Peptide Name** | **Peptide Sequence** | **Peptide Description** |
| 4930 | 'KLKB1_No5' | 'VNIPLVTNEECQKR' | 'Plasma kallikrein' |
| 4931 | 'KLKB1_No6' | 'GGDVASMYTPNAQYCQMR' | 'Plasma kallikrein' |
| 4932 | 'KLKB1_No7' | 'GGDVASMYTPNAQYCQMR' | 'Plasma kallikrein' |
| 4933 | 'KLKB1_No8' | 'VLSNVESGFSLKPCALSEIGCHMNIFQHLAFSDVDVAR' | 'Plasma kallikrein' |
| 4934 | 'KLKB1_No9' | 'CQFFTYSLLPEDCKEEK' | 'Plasma kallikrein' |
| 4935 | 'KLKB1_No10' | 'CLLFSFLPASSINDMEKR' | 'Plasma kallikrein' |
| 4936 | 'KLKB1_No11' | 'EIIIHQNYK' | 'Plasma kallikrein' |
| 4937 | 'KLKB1_No12' | 'GGDVASMYTPNAQYCQMR' | 'Plasma kallikrein' |
| 4938 | 'KLKB1_No13' | 'GVNVCQETCTK' | 'Plasma kallikrein' |
| 4939 | 'KLKB1_No14' | 'GDTSTIYTNCWVTGWGFSK' | 'Plasma kallikrein' |
| 4940 | 'KLKB1_No15' | 'CLLFSFLPASSINDMEK' | 'Plasma kallikrein' |
| 4941 | 'KLKB1_No16' | 'TSESGTPSSSTPQENTISGYSLLTCKR' | 'Plasma kallikrein' |
| 4942 | 'KLKB1_No17' | 'VAEYMDWILEK' | 'Plasma kallikrein' |
| 4943 | 'KLKB1_No18' | 'LVGITSWGEGCAR' | 'Plasma kallikrein' |
| 4944 | 'KLKB1_No19' | 'CLLFSFLPASSINDMEKR' | 'Plasma kallikrein' |
| 4945 | 'KLKB1_No20' | 'PICLPSK' | 'Plasma kallikrein' |
| 4946 | 'KLKB1_No21' | 'LCNTGDNSVCTTK' | 'Plasma kallikrein' |
| 4947 | 'KLKB1_No22' | 'HLCGGSLIGHQWVLTAAHCFDGLPLQDVWR' | 'Plasma kallikrein' |
| 4948 | 'KLKB1_No23' | 'LSMDGSPTR' | 'Plasma kallikrein' |
| 4949 | 'KLKB1_No24' | 'DIYKGVDMR' | 'Plasma kallikrein' |
| 4950 | 'KLKB1_No25' | 'EKGEIQNILQK' | 'Plasma kallikrein' |
| 4951 | 'KLKB1_No26' | 'DTPFSQIK' | 'Plasma kallikrein' |
| 4952 | 'KLKB1_No27' | 'DSVTGTLPK' | 'Plasma kallikrein' |
| 4953 | 'KLKB1_No28' | 'VLTPDAFVCR' | 'Plasma kallikrein' |
| 4954 | 'KLKB1_No29' | 'LSMDGSPTR' | 'Plasma kallikrein' |
| 4955 | 'KLKB1_No30' | 'GGDVASMYTPNAQYCQMR' | 'Plasma kallikrein' |
| 4956 | 'KLKB1_No31' | 'CQFFSYATQTFHK' | 'Plasma kallikrein' |
| 4957 | 'KLKB1_No32' | 'VLSNVESGFSLKPCALSEIGCHMNIFQHLAFSDVDVAR' | 'Plasma kallikrein' |
| 4958 | 'KLKB1_No33' | 'VSEGNHDIALIK' | 'Plasma kallikrein' |
| 4959 | 'KLKB1_No34' | 'DACKGDSGGPLVCK' | 'Plasma kallikrein' |
| 4960 | 'KLKB1_No35' | 'YSPGGTPTAIK' | 'Plasma kallikrein' |
| 4961 | 'KLKB1_No36' | 'TSESGTPSSSTPQENTISGYSLLTCK' | 'Plasma kallikrein' |
| 4962 | 'KLKB1_No37' | 'VAEYMDWILEKTQSSDGK' | 'Plasma kallikrein' |
| 4963 | 'KLKB1_No38' | 'CLLFSFLPASSINDMEK' | 'Plasma kallikrein' |
| 4964 | 'KLKB1_No39' | 'DTPFSQIKEIIIHQNYK' | 'Plasma kallikrein' |
| 4965 | 'KLKB1_No40' | 'EGGKDACKGDSGGPLVCK' | 'Plasma kallikrein' |
| 4966 | 'KLKB1_No41' | 'IYSGILNLSDITKDTPFSQIK' | 'Plasma kallikrein' |
| 4967 | 'KLKB1_No42' | 'TGAVSGHSLKQCGHQISACHR' | 'Plasma kallikrein' |
| 4968 | 'KLKB1_No43' | 'FGCFLKDSVTGTLPK' | 'Plasma kallikrein' |
| 4969 | 'KLKB1_No44' | 'TLPEPCHSK' | 'Plasma kallikrein' |
| 4970 | 'KLKB1_No45' | 'QCGHQISACHRDIYKGVDMR' | 'Plasma kallikrein' |
| 4971 | 'KLKB1_No46' | 'QCGHQISACHRDIYK' | 'Plasma kallikrein' |
| 4972 | 'KLKB1_No47' | 'QCGHQISACHR' | 'Plasma kallikrein' |
| 4973 | 'KLKB1_No48' | 'MVCAGYK' | 'Plasma kallikrein' |

| | A | B | C |
|---|---|---|---|
| 1 | **Peptide Name** | **Peptide Sequence** | **Peptide Description** |
| 4974 | 'KLKB1_No49' | 'VAEYMDWILEKTQSSDGKAQMQSPA' | 'Plasma kallikrein' |
| 4975 | 'KLKB1_No50' | 'CQFFTYSLLPEDCKEEKCK' | 'Plasma kallikrein' |
| 4976 | 'KLKB1_No51' | 'TGAVSGHSLK' | 'Plasma kallikrein' |
| 4977 | 'KLKB1_No52' | 'DIYKGVDMR' | 'Plasma kallikrein' |
| 4978 | 'KLKB1_No53' | 'REQPGVYTK' | 'Plasma kallikrein' |
| 4979 | 'KLKB1_No54' | 'VSSVEECQKR' | 'Plasma kallikrein' |
| 4980 | 'KLKB1_No55' | 'CQFFSYATQTFHKAEYR' | 'Plasma kallikrein' |
| 4981 | 'KLKB1_No56' | 'VSSVEECQK' | 'Plasma kallikrein' |
| 4982 | 'KLKB1_No57' | 'GEIQNILQK' | 'Plasma kallikrein' |
| 4983 | 'KLKB1_No58' | 'QCGHQISACHRDIYKGVDMR' | 'Plasma kallikrein' |
| 4984 | 'KLKB1_No59' | 'GDSGGPLVCK' | 'Plasma kallikrein' |
| 4985 | 'KLKB1_No60' | 'QCGHQISACHRDIYK' | 'Plasma kallikrein' |
| 4986 | 'KLKB1_No61' | 'LVGITSWGEGCARR' | 'Plasma kallikrein' |
| 4987 | 'KLKB1_No62' | 'VAEYMDWILEKTQSSDGKAQMQSPA' | 'Plasma kallikrein' |
| 4988 | 'KLKB1_No63' | 'CQFFSYATQTFHKAEYRNNCLLK' | 'Plasma kallikrein' |
| 4989 | 'KLKB1_No64' | 'QCGHQISACHR' | 'Plasma kallikrein' |
| 4990 | 'KLKB1_No65' | 'MVCAGYK' | 'Plasma kallikrein' |
| 4991 | 'KLKB1_No66' | 'VAEYMDWILEKTQSSDGKAQMQSPA' | 'Plasma kallikrein' |
| 4992 | 'KLKB1_No67' | 'VAEYMDWILEKTQSSDGK' | 'Plasma kallikrein' |
| 4993 | 'C4BPA_No1' | 'LSLEIEQLELQR' | 'C4b-binding protein alpha chain' |
| 4994 | 'C4BPA_No2' | 'WTPYQGCEALCCPEPK' | 'C4b-binding protein alpha chain' |
| 4995 | 'C4BPA_No3' | 'SHSTQTLTCNSDGEWVYNTFCIYK' | 'C4b-binding protein alpha chain' |
| 4996 | 'C4BPA_No4' | 'LMQCLPNPEDVK' | 'C4b-binding protein alpha chain' |
| 4997 | 'C4BPA_No5' | 'LSCSYSHWSAPAPQCK' | 'C4b-binding protein alpha chain' |
| 4998 | 'C4BPA_No6' | 'FSAICQGDGTWSPR' | 'C4b-binding protein alpha chain' |
| 4999 | 'C4BPA_No7' | 'TPSCGDICNFPPK' | 'C4b-binding protein alpha chain' |
| 5000 | 'C4BPA_No8' | 'LMQCLPNPEDVK' | 'C4b-binding protein alpha chain' |
| 5001 | 'C4BPA_No9' | 'CEWETPEGCEQVLTGK' | 'C4b-binding protein alpha chain' |
| 5002 | 'C4BPA_No10' | 'TWYPEVPK' | 'C4b-binding protein alpha chain' |
| 5003 | 'C4BPA_No11' | 'PTKEDVYVVGTVLR' | 'C4b-binding protein alpha chain' |
| 5004 | 'C4BPA_No12' | 'QSSSYSFFKEEIIYECDKGYILVGQAK' | 'C4b-binding protein alpha chain' |
| 5005 | 'C4BPA_No13' | 'CHPGYKPTTDEPTTVICQK' | 'C4b-binding protein alpha chain' |
| 5006 | 'C4BPA_No14' | 'KPDVSHGEMVSGFGPIYNYKDTIVFK' | 'C4b-binding protein alpha chain' |
| 5007 | 'C4BPA_No15' | 'MALEVYK' | 'C4b-binding protein alpha chain' |
| 5008 | 'C4BPA_No16' | 'PTTDEPTTVICQK' | 'C4b-binding protein alpha chain' |
| 5009 | 'C4BPA_No17' | 'KPDVSHGEMVSGFGPIYNYK' | 'C4b-binding protein alpha chain' |
| 5010 | 'C4BPA_No18' | 'KPDVSHGEMVSGFGPIYNYKDTIVFK' | 'C4b-binding protein alpha chain' |
| 5011 | 'C4BPA_No19' | 'GVGWSHPLPQCEIVK' | 'C4b-binding protein alpha chain' |
| 5012 | 'C4BPA_No20' | 'SHSTQTLTCNSDGEWVYNTFCIYKR' | 'C4b-binding protein alpha chain' |
| 5013 | 'C4BPA_No21' | 'QSSSYSFFKEEIIYECDK' | 'C4b-binding protein alpha chain' |
| 5014 | 'C4BPA_No22' | 'LNNGEITQHR' | 'C4b-binding protein alpha chain' |
| 5015 | 'C4BPA_No23' | 'RLMQCLPNPEDVK' | 'C4b-binding protein alpha chain' |
| 5016 | 'C4BPA_No24' | 'WNPSPPACEPNSCINLPDIPHASWETYPRPTKEDVYVVGTVLR' | 'C4b-binding protein alpha chain' |
| 5017 | 'C4BPA_No25' | 'EDVYVVGTVLR' | 'C4b-binding protein alpha chain' |

| | A | B | C |
|---|---|---|---|
| 1 | **Peptide Name** | **Peptide Sequence** | **Peptide Description** |
| 5018 | 'C4BPA_No26' | 'QSTLDKEL' | 'C4b-binding protein alpha chain' |
| 5019 | 'C4BPA_No27' | 'PANHCVYFYGDEISFSCHETSR' | 'C4b-binding protein alpha chain' |
| 5020 | 'C4BPA_No28' | 'WNPSPPACEPNSCINLPDIPHASWETYPRPTK' | 'C4b-binding protein alpha chain' |
| 5021 | 'C4BPA_No29' | 'KPDVSHGEMVSGFGPIYNYK' | 'C4b-binding protein alpha chain' |
| 5022 | 'C4BPA_No30' | 'CEWETPEGCEQVLTGKR' | 'C4b-binding protein alpha chain' |
| 5023 | 'C4BPA_No31' | 'FSLLGHASISCTVENETIGVWRPSPPTCEK' | 'C4b-binding protein alpha chain' |
| 5024 | 'C4BPA_No32' | 'YTCLPGYVR' | 'C4b-binding protein alpha chain' |
| 5025 | 'C4BPA_No33' | 'QSSSYSFFK' | 'C4b-binding protein alpha chain' |
| 5026 | 'C4BPA_No34' | 'QSSSYSFFKEEIIYECDKGYILVGQAK' | 'C4b-binding protein alpha chain' |
| 5027 | 'C4BPA_No35' | 'SRPANHCVYFYGDEISFSCHETSR' | 'C4b-binding protein alpha chain' |
| 5028 | 'C4BPA_No36' | 'TWYPEVPKCEWETPEGCEQVLTGK' | 'C4b-binding protein alpha chain' |
| 5029 | 'C4BPA_No37' | 'TWYPEVPKCEWETPEGCEQVLTGKR' | 'C4b-binding protein alpha chain' |
| 5030 | 'C4BPA_No38' | 'GSSVIHCDADSK' | 'C4b-binding protein alpha chain' |
| 5031 | 'C4BPA_No39' | 'LMQCLPNPEDVKMALEVYK' | 'C4b-binding protein alpha chain' |
| 5032 | 'C4BPA_No40' | 'MALEVYKLSLEIEQLELQR' | 'C4b-binding protein alpha chain' |
| 5033 | 'C4BPA_No41' | 'MALEVYK' | 'C4b-binding protein alpha chain' |
| 5034 | 'C4BPA_No42' | 'KSRPANHCVYFYGDEISFSCHETSR' | 'C4b-binding protein alpha chain' |
| 5035 | 'C4BPA_No43' | 'TGTTLKYTCLPGYVR' | 'C4b-binding protein alpha chain' |
| 5036 | 'C4BPA_No44' | 'KPELVNGR' | 'C4b-binding protein alpha chain' |
| 5037 | 'C4BPA_No45' | 'EEIIYECDKGYILVGQAK' | 'C4b-binding protein alpha chain' |
| 5038 | 'C4BPA_No46' | 'PDVSHGEMVSGFGPIYNYK' | 'C4b-binding protein alpha chain' |
| 5039 | 'C4BPA_No47' | 'YRCHPGYKPTTDEPTTVICQK' | 'C4b-binding protein alpha chain' |
| 5040 | 'C4BPA_No48' | 'FKTGTTLKYTCLPGYVR' | 'C4b-binding protein alpha chain' |
| 5041 | 'C4BPA_No49' | 'CKPPPDIR' | 'C4b-binding protein alpha chain' |
| 5042 | 'C4BPA_No50' | 'FKTGTTLK' | 'C4b-binding protein alpha chain' |
| 5043 | 'C4BPA_No51' | 'GYILVGQAK' | 'C4b-binding protein alpha chain' |
| 5044 | 'C4BPA_No52' | 'RLMQCLPNPEDVK' | 'C4b-binding protein alpha chain' |
| 5045 | 'C4BPA_No53' | 'GFVLRGSSVIHCDADSK' | 'C4b-binding protein alpha chain' |
| 5046 | 'C4BPA_No54' | 'QSSSYSFFKEEIIYECDK' | 'C4b-binding protein alpha chain' |
| 5047 | 'C4BPA_No55' | 'MALEVYKLSLEIEQLELQRDSAR' | 'C4b-binding protein alpha chain' |
| 5048 | 'C4BPA_No56' | 'EEIIYECDK' | 'C4b-binding protein alpha chain' |
| 5049 | 'C4BPA_No57' | 'NLRWTPYQGCEALCCPEPK' | 'C4b-binding protein alpha chain' |
| 5050 | 'C4BPA_No58' | 'LNNGEITQHRK' | 'C4b-binding protein alpha chain' |
| 5051 | 'C4BPA_No59' | 'LMQCLPNPEDVKMALEVYK' | 'C4b-binding protein alpha chain' |
| 5052 | 'C4BPA_No60' | 'LSLEIEQLELQRDSAR' | 'C4b-binding protein alpha chain' |
| 5053 | 'C4BPA_No61' | 'LMQCLPNPEDVKMALEVYK' | 'C4b-binding protein alpha chain' |
| 5054 | 'C4BPA_No62' | 'PDVSHGEMVSGFGPIYNYK' | 'C4b-binding protein alpha chain' |
| 5055 | 'C4BPA_No63' | 'PSPPTCEK' | 'C4b-binding protein alpha chain' |
| 5056 | 'C4BPA_No64' | 'QSTLDKEL' | 'C4b-binding protein alpha chain' |
| 5057 | 'C4BPA_No65' | 'IAHGHYK' | 'C4b-binding protein alpha chain' |
| 5058 | 'C4BPA_No66' | 'WNPSPPACEPNSCINLPDIPHASWETYPR' | 'C4b-binding protein alpha chain' |
| 5059 | 'C4BPA_No67' | 'LMQCLPNPEDVKMALEVYK' | 'C4b-binding protein alpha chain' |
| 5060 | 'C4BPA_No68' | 'CEVQDRGVGWSHPLPQCEIVK' | 'C4b-binding protein alpha chain' |
| 5061 | 'VTN_No1' | 'GQYCYELDEK' | 'Vitronectin' |

| | A | B | C |
|---|---|---|---|
| 1 | **Peptide Name** | **Peptide Sequence** | **Peptide Description** |
| 5062 | 'VTN_No2' | 'FEDGVLDPDYPR' | 'Vitronectin' |
| 5063 | 'VTN_No3' | 'IYISGMAPR' | 'Vitronectin' |
| 5064 | 'VTN_No4' | 'IYISGMAPR' | 'Vitronectin' |
| 5065 | 'VTN_No5' | 'RVDTVDPPYPR' | 'Vitronectin' |
| 5066 | 'VTN_No6' | 'PQPPAEEELCSGKPFDAFTDLK' | 'Vitronectin' |
| 5067 | 'VTN_No7' | 'VDTVDPPYPR' | 'Vitronectin' |
| 5068 | 'VTN_No8' | 'ERVYFFK' | 'Vitronectin' |
| 5069 | 'VTN_No9' | 'DWHGVPGQVDAAMAGR' | 'Vitronectin' |
| 5070 | 'VTN_No10' | 'IYISGMAPRPSLAK' | 'Vitronectin' |
| 5071 | 'VTN_No11' | 'DWHGVPGQVDAAMAGR' | 'Vitronectin' |
| 5072 | 'VTN_No12' | 'SIAQYWLGCPAPGHL' | 'Vitronectin' |
| 5073 | 'VTN_No13' | 'LIRDVWGIEGPIDAAFTR' | 'Vitronectin' |
| 5074 | 'VTN_No14' | 'QPQFISR' | 'Vitronectin' |
| 5075 | 'VTN_No15' | 'KCQCDELCSYYQSCCTDYTAECKPQVTR' | 'Vitronectin' |
| 5076 | 'VTN_No16' | 'DVWGIEGPIDAAFTR' | 'Vitronectin' |
| 5077 | 'VTN_No17' | 'IYISGMAPRPSLAK' | 'Vitronectin' |
| 5078 | 'VTN_No18' | 'AVRPGYPK' | 'Vitronectin' |
| 5079 | 'VTN_No19' | 'GQYCYELDEKAVRPGYPK' | 'Vitronectin' |
| 5080 | 'VTN_No20' | 'TRRVDTVDPPYPR' | 'Vitronectin' |
| 5081 | 'VTN_No21' | 'TSAGTRQPQFISR' | 'Vitronectin' |
| 5082 | 'VTN_No22' | 'GDVFTMPEDEYTVYDDGEEK' | 'Vitronectin' |
| 5083 | 'VTN_No23' | 'CQCDELCSYYQSCCTDYTAECKPQVTR' | 'Vitronectin' |
| 5084 | 'VTN_No24' | 'TYLFKGSQYWR' | 'Vitronectin' |
| 5085 | 'VTN_No25' | 'CTEGFNVDKK' | 'Vitronectin' |
| 5086 | 'VTN_No26' | 'GNPEQTPVLKPEEEAPAPEVGASKPEGIDSR' | 'Vitronectin' |
| 5087 | 'VTN_No27' | 'MDWLVPATCEPIQSVFFFSGDK' | 'Vitronectin' |
| 5088 | 'VTN_No28' | 'NISDGFDGIPDNVDAALALPAHSYSGR' | 'Vitronectin' |
| 5089 | 'VTN_No29' | 'INCQGKTYLFK' | 'Vitronectin' |
| 5090 | 'VTN_No30' | 'NGSLFAFR' | 'Vitronectin' |
| 5091 | 'VTN_No31' | 'QPQFISRDWHGVPGQVDAAMAGR' | 'Vitronectin' |
| 5092 | 'VTN_No32' | 'GSQYWRFEDGVLDPDYPR' | 'Vitronectin' |
| 5093 | 'VTN_No33' | 'INCQGKTYLFKGSQYWR' | 'Vitronectin' |
| 5094 | 'CAT_No1' | 'GAGAFGYFEVTHDITK' | 'Catalase' |
| 5095 | 'CAT_No2' | 'ADVLTTGAGNPVGDK' | 'Catalase' |
| 5096 | 'CAT_No3' | 'FNTANDDNVTQVR' | 'Catalase' |
| 5097 | 'CAT_No4' | 'FSTVAGESGSADTVRDPR' | 'Catalase' |
| 5098 | 'CAT_No5' | 'DLFNAIATGK' | 'Catalase' |
| 5099 | 'CAT_No6' | 'GPLLVQDVVFTDEMAHFDRER' | 'Catalase' |
| 5100 | 'CAT_No7' | 'DYPLIPVGK' | 'Catalase' |
| 5101 | 'CAT_No8' | 'NAIHTFVQSGSHLAAR' | 'Catalase' |
| 5102 | 'CAT_No9' | 'LVNANGEAVYCK' | 'Catalase' |
| 5103 | 'CAT_No10' | 'NFTEVHPDYGSHIQALLDKYNAEKPK' | 'Catalase' |
| 5104 | 'CAT_No11' | 'DPILFPSFIHSQK' | 'Catalase' |
| 5105 | 'CAT_No12' | 'GPLLVQDVVFTDEMAHFDR' | 'Catalase' |

| | A | B | C |
|---|---|---|---|
| 1 | **Peptide Name** | **Peptide Sequence** | **Peptide Description** |
| 5106 | 'CAT_No13' | 'FSTVAGESGSADTVR' | 'Catalase' |
| 5107 | 'CAT_No14' | 'VWPHKDYPLIPVGK' | 'Catalase' |
| 5108 | 'CAT_No15' | 'GPLLVQDVVFTDEMAHFDRER' | 'Catalase' |
| 5109 | 'CAT_No16' | 'ADVLTTGAGNPVGDKLNVITVGPR' | 'Catalase' |
| 5110 | 'CAT_No17' | 'RFNTANDDNVTQVR' | 'Catalase' |
| 5111 | 'CAT_No18' | 'LGPNYLHIPVNCPYR' | 'Catalase' |
| 5112 | 'CAT_No19' | 'ADSRDPASDQMQHWKEQR' | 'Catalase' |
| 5113 | 'CAT_No20' | 'LCENIAGHLK' | 'Catalase' |
| 5114 | 'CAT_No21' | 'ADSRDPASDQMQHWK' | 'Catalase' |
| 5115 | 'CAT_No22' | 'AAQKADVLTTGAGNPVGDKLNVITVGPR' | 'Catalase' |
| 5116 | 'CAT_No23' | 'RLCENIAGHLK' | 'Catalase' |
| 5117 | 'CAT_No24' | 'ADSRDPASDQMQHWK' | 'Catalase' |
| 5118 | 'CAT_No25' | 'GPLLVQDVVFTDEMAHFDR' | 'Catalase' |
| 5119 | 'CAT_No26' | 'NFTEVHPDYGSHIQALLDK' | 'Catalase' |
| 5120 | 'CAT_No27' | 'GAGAFGYFEVTHDITKYSK' | 'Catalase' |
| 5121 | 'CAT_No28' | 'ADSRDPASDQMQHWKEQR' | 'Catalase' |
| 5122 | 'CAT_No29' | 'LCENIAGHLKDAQIFIQK' | 'Catalase' |
| 5123 | 'FUCA1_No1' | 'DLVGELGTALR' | 'Tissue alpha-L-fucosidase' |
| 5124 | 'FUCA1_No2' | 'FFHPEEWADLFQAAGAK' | 'Tissue alpha-L-fucosidase' |
| 5125 | 'FUCA1_No3' | 'DGLIVPIFQER' | 'Tissue alpha-L-fucosidase' |
| 5126 | 'FUCA1_No4' | 'DNYPPGFSYADFGPQFTAR' | 'Tissue alpha-L-fucosidase' |
| 5127 | 'PROC_No1' | 'TFVLNFIK' | 'Vitamin K-dependent protein C' |
| 5128 | 'PROC_No2' | 'LGDDLLQCHPAVK' | 'Vitamin K-dependent protein C' |
| 5129 | 'PROC_No3' | 'YLDWIHGHIR' | 'Vitamin K-dependent protein C' |
| 5130 | 'PROC_No4' | 'DTEDQEDQVDPR' | 'Vitamin K-dependent protein C' |
| 5131 | 'PROC_No5' | 'LACGAVLIHPSWVLTAAHCMDESKK' | 'Vitamin K-dependent protein C' |
| 5132 | 'PROC_No6' | 'LACGAVLIHPSWVLTAAHCMDESK' | 'Vitamin K-dependent protein C' |
| 5133 | 'PROC_No7' | 'RGDSPWQVVLLDSK' | 'Vitamin K-dependent protein C' |
| 5134 | 'PROC_No8' | 'ELNQAGQETLVTGWGYHSSR' | 'Vitamin K-dependent protein C' |
| 5135 | 'PROC_No9' | 'FPCGRPWK' | 'Vitamin K-dependent protein C' |
| 5136 | 'PROC_No10' | 'YLDWIHGHIRDK' | 'Vitamin K-dependent protein C' |
| 5137 | 'PROC_No11' | 'LACGAVLIHPSWVLTAAHCMDESKK' | 'Vitamin K-dependent protein C' |
| 5138 | 'PROC_No12' | 'RGDSPWQVVLLDSKK' | 'Vitamin K-dependent protein C' |
| 5139 | 'PROC_No13' | 'KLACGAVLIHPSWVLTAAHCMDESK' | 'Vitamin K-dependent protein C' |
| 5140 | 'PROC_No14' | 'GDSPWQVVLLDSK' | 'Vitamin K-dependent protein C' |
| 5141 | 'PROC_No15' | 'YLDWIHGHIRDKEAPQK' | 'Vitamin K-dependent protein C' |
| 5142 | 'PROC_No16' | 'DTEDQEDQVDPRLIDGK' | 'Vitamin K-dependent protein C' |
| 5143 | 'PROC_No17' | 'IPVVPHNECSEVMSNMVSENMLCAGILGDR' | 'Vitamin K-dependent protein C' |
| 5144 | 'PROC_No18' | 'STTDNDIALLHLAQPATLSQTIVPICLPDSGLAER' | 'Vitamin K-dependent protein C' |
| 5145 | 'PROC_No19' | 'LACGAVLIHPSWVLTAAHCMDESK' | 'Vitamin K-dependent protein C' |
| 5146 | 'ALDOA_No1' | 'TVPPAVTGITFLSGGQSEEEASINLNAINK' | 'Fructose-bisphosphate aldolase A' |
| 5147 | 'ALDOA_No2' | 'IGEHTPSALAIMENANVLAR' | 'Fructose-bisphosphate aldolase A' |
| 5148 | 'ALDOA_No3' | 'PYQYPALTPEQK' | 'Fructose-bisphosphate aldolase A' |
| 5149 | 'ALDOA_No4' | 'GILAADESTGSIAKR' | 'Fructose-bisphosphate aldolase A' |

| | A | B | C |
|---|---|---|---|
| 1 | **Peptide Name** | **Peptide Sequence** | **Peptide Description** |
| 5150 | 'ALDOA_No5' | 'GVVPLAGTNGETTTQGLDGLSER' | 'Fructose-bisphosphate aldolase A' |
| 5151 | 'ALDOA_No6' | 'LQSIGTENTEENRR' | 'Fructose-bisphosphate aldolase A' |
| 5152 | 'ALDOA_No7' | 'ADDGRPFPQVIK' | 'Fructose-bisphosphate aldolase A' |
| 5153 | 'ALDOA_No8' | 'RLQSIGTENTEENRR' | 'Fructose-bisphosphate aldolase A' |
| 5154 | 'ALDOA_No9' | 'ALANSLACQGK' | 'Fructose-bisphosphate aldolase A' |
| 5155 | 'ALDOA_No10' | 'YTPSGQAGAAASESLFVSNHAY' | 'Fructose-bisphosphate aldolase A' |
| 5156 | 'ALDOA_No11' | 'CPLLKPWALTFSYGR' | 'Fructose-bisphosphate aldolase A' |
| 5157 | 'ALDOA_No12' | 'VDKGVVPLAGTNGETTTQGLDGLSER' | 'Fructose-bisphosphate aldolase A' |
| 5158 | 'ALDOA_No13' | 'YASICQQNGIVPIVEPEILPDGDHDLKR' | 'Fructose-bisphosphate aldolase A' |
| 5159 | 'ALDOA_No14' | 'VNPCIGGVILFHETLYQK' | 'Fructose-bisphosphate aldolase A' |
| 5160 | 'ALDOA_No15' | 'FSHEEIAMATVTALR' | 'Fructose-bisphosphate aldolase A' |
| 5161 | 'ALDOA_No16' | 'FSHEEIAMATVTALRR' | 'Fructose-bisphosphate aldolase A' |
| 5162 | 'ALDOA_No17' | 'PYQYPALTPEQKK' | 'Fructose-bisphosphate aldolase A' |
| 5163 | 'ALDOA_No18' | 'IVAPGKGILAADESTGSIAK' | 'Fructose-bisphosphate aldolase A' |
| 5164 | 'ALDOA_No19' | 'FSHEEIAMATVTALR' | 'Fructose-bisphosphate aldolase A' |
| 5165 | 'ALDOA_No20' | 'ENLKAAQEEYVKR' | 'Fructose-bisphosphate aldolase A' |
| 5166 | 'ALDOA_No21' | 'IGEHTPSALAIMENANVLAR' | 'Fructose-bisphosphate aldolase A' |
| 5167 | 'ALDOA_No22' | 'FSHEEIAMATVTALRR' | 'Fructose-bisphosphate aldolase A' |
| 5168 | 'ALDOA_No23' | 'YASICQQNGIVPIVEPEILPDGDHDLK' | 'Fructose-bisphosphate aldolase A' |
| 5169 | 'CSTB' | 'VHVGDEDFVHLR' | 'Cystatin-B' |
| 5170 | 'ANXA1_No1' | 'AAYLQETGKPLDETLKK' | 'Annexin A1' |
| 5171 | 'ANXA1_No2' | 'DITSDTSGDFR' | 'Annexin A1' |
| 5172 | 'ANXA1_No3' | 'GVDEATIIDILTKR' | 'Annexin A1' |
| 5173 | 'ANXA1_No4' | 'TPAQFDADELR' | 'Annexin A1' |
| 5174 | 'ANXA1_No5' | 'SEDFGVNEDLADSDAR' | 'Annexin A1' |
| 5175 | 'ANXA1_No6' | 'GTDVNVFNTILTTR' | 'Annexin A1' |
| 5176 | 'APOB_No1' | 'QTIIVVLENVQR' | 'Apolipoprotein B-100' |
| 5177 | 'APOB_No2' | 'IAELSATAQEIIK' | 'Apolipoprotein B-100' |
| 5178 | 'APOB_No3' | 'IGQDGISTSATTNLK' | 'Apolipoprotein B-100' |
| 5179 | 'APOB_No4' | 'VIGNMGQTMEQLTPELK' | 'Apolipoprotein B-100' |
| 5180 | 'APOB_No5' | 'GFEPTLEALFGK' | 'Apolipoprotein B-100' |
| 5181 | 'APOB_No6' | 'QVFLYPEKDEPTYILNIK' | 'Apolipoprotein B-100' |
| 5182 | 'APOB_No7' | 'FPEVDVLTK' | 'Apolipoprotein B-100' |
| 5183 | 'APOB_No8' | 'IDDIWNLEVK' | 'Apolipoprotein B-100' |
| 5184 | 'APOB_No9' | 'YGMVAQVTQTLK' | 'Apolipoprotein B-100' |
| 5185 | 'APOB_No10' | 'INCKVELEVPQLCSFILK' | 'Apolipoprotein B-100' |
| 5186 | 'APOB_No11' | 'LALWGEHTGQLYSK' | 'Apolipoprotein B-100' |
| 5187 | 'APOB_No12' | 'DAVEKPQEFTIVAFVK' | 'Apolipoprotein B-100' |
| 5188 | 'APOB_No13' | 'TLADLTLLDSPIK' | 'Apolipoprotein B-100' |
| 5189 | 'APOB_No14' | 'HFVINLIGDFEVAEK' | 'Apolipoprotein B-100' |
| 5190 | 'APOB_No15' | 'NFVASHIANILNSEELDIQDLKK' | 'Apolipoprotein B-100' |
| 5191 | 'APOB_No16' | 'INNQLTLDSNTK' | 'Apolipoprotein B-100' |
| 5192 | 'APOB_No17' | 'SGSSTASWIQNVDTK' | 'Apolipoprotein B-100' |
| 5193 | 'APOB_No18' | 'LTISEQNIQR' | 'Apolipoprotein B-100' |

| | A | B | C |
|---|---|---|---|
| 1 | **Peptide Name** | **Peptide Sequence** | **Peptide Description** |
| 5194 | 'APOB_No19' | 'ITENDIQIALDDAK' | 'Apolipoprotein B-100' |
| 5195 | 'APOB_No20' | 'VHANPLLIDVVTYLVALIPEPSAQQLR' | 'Apolipoprotein B-100' |
| 5196 | 'APOB_No21' | 'TEHGSEMLFFGNAIEGK' | 'Apolipoprotein B-100' |
| 5197 | 'APOB_No22' | 'YTYNYEAESSSGVPGTADSR' | 'Apolipoprotein B-100' |
| 5198 | 'APOB_No23' | 'KYTYNYEAESSSGVPGTADSR' | 'Apolipoprotein B-100' |
| 5199 | 'APOB_No24' | 'LEIQSQVDSQHVGHSVLTAK' | 'Apolipoprotein B-100' |
| 5200 | 'APOB_No25' | 'FSDEGTHESQISFTIEGPLTSFGLSNK' | 'Apolipoprotein B-100' |
| 5201 | 'APOB_No26' | 'VNWEEEAASGLLTSLK' | 'Apolipoprotein B-100' |
| 5202 | 'APOB_No27' | 'TILGTMPAFEVSLQALQK' | 'Apolipoprotein B-100' |
| 5203 | 'APOB_No28' | 'SFDYHQFVDETNDK' | 'Apolipoprotein B-100' |
| 5204 | 'APOB_No29' | 'SGVQMNTNFFHESGLEAHVALK' | 'Apolipoprotein B-100' |
| 5205 | 'APOB_No30' | 'LIVAMSSWLQK' | 'Apolipoprotein B-100' |
| 5206 | 'APOB_No31' | 'DKIGVELTGR' | 'Apolipoprotein B-100' |
| 5207 | 'APOB_No32' | 'LTLDIQNK' | 'Apolipoprotein B-100' |
| 5208 | 'APOB_No33' | 'GESKLEVLNFDFQANAQLSNPK' | 'Apolipoprotein B-100' |
| 5209 | 'APOB_No34' | 'EVGTVLSQVYSK' | 'Apolipoprotein B-100' |
| 5210 | 'APOB_No35' | 'NTLELSNGVIVK' | 'Apolipoprotein B-100' |
| 5211 | 'APOB_No36' | 'SGVQMNTNFFHESGLEAHVALK' | 'Apolipoprotein B-100' |
| 5212 | 'APOB_No37' | 'VNWEEEAASGLLTSLKDNVPK' | 'Apolipoprotein B-100' |
| 5213 | 'APOB_No38' | 'MYQMDIQQELQR' | 'Apolipoprotein B-100' |
| 5214 | 'APOB_No39' | 'NFVASHIANILNSEELDIQDLK' | 'Apolipoprotein B-100' |
| 5215 | 'APOB_No40' | 'TGISPLALIK' | 'Apolipoprotein B-100' |
| 5216 | 'APOB_No41' | 'EQHLFLPFSYK' | 'Apolipoprotein B-100' |
| 5217 | 'APOB_No42' | 'VPQTDMTFR' | 'Apolipoprotein B-100' |
| 5218 | 'APOB_No43' | 'HIQNIDIQHLAGK' | 'Apolipoprotein B-100' |
| 5219 | 'APOB_No44' | 'ATLYALSHAVNNYHK' | 'Apolipoprotein B-100' |
| 5220 | 'APOB_No45' | 'SEILAHWSPAK' | 'Apolipoprotein B-100' |
| 5221 | 'APOB_No46' | 'FQFPGKPGIYTR' | 'Apolipoprotein B-100' |
| 5222 | 'APOB_No47' | 'IYSLWEHSTK' | 'Apolipoprotein B-100' |
| 5223 | 'APOB_No48' | 'AQIPILR' | 'Apolipoprotein B-100' |
| 5224 | 'APOB_No49' | 'LSLPDFK' | 'Apolipoprotein B-100' |
| 5225 | 'APOB_No50' | 'VLADKFIIPGLK' | 'Apolipoprotein B-100' |
| 5226 | 'APOB_No51' | 'SVGFHLPSR' | 'Apolipoprotein B-100' |
| 5227 | 'APOB_No52' | 'LDNIYSSDKFYK' | 'Apolipoprotein B-100' |
| 5228 | 'APOB_No53' | 'YHWEHTGLTLR' | 'Apolipoprotein B-100' |
| 5229 | 'APOB_No54' | 'IGVELTGR' | 'Apolipoprotein B-100' |
| 5230 | 'APOB_No55' | 'KLTISEQNIQR' | 'Apolipoprotein B-100' |
| 5231 | 'APOB_No56' | 'IISDYHQQFR' | 'Apolipoprotein B-100' |
| 5232 | 'APOB_No57' | 'YNALDLTNNGK' | 'Apolipoprotein B-100' |
| 5233 | 'APOB_No58' | 'FIIPSPK' | 'Apolipoprotein B-100' |
| 5234 | 'APOB_No59' | 'DNVFDGLVR' | 'Apolipoprotein B-100' |
| 5235 | 'APOB_No60' | 'LKQHIEAIDVR' | 'Apolipoprotein B-100' |
| 5236 | 'APOB_No61' | 'AQNLYQELLTQEGQASFQGLK' | 'Apolipoprotein B-100' |
| 5237 | 'APOB_No62' | 'SFDYHQFVDETNDKIR' | 'Apolipoprotein B-100' |

| | A | B | C |
|---|---|---|---|
| 1 | **Peptide Name** | **Peptide Sequence** | **Peptide Description** |
| 5238 | 'APOB_No63' | 'LVGFIDDAVK' | 'Apolipoprotein B-100' |
| 5239 | 'APOB_No64' | 'LNGEIQALELPQKAEALK' | 'Apolipoprotein B-100' |
| 5240 | 'APOB_No65' | 'YGMVAQVTQTLK' | 'Apolipoprotein B-100' |
| 5241 | 'APOB_No66' | 'VPQTDMTFR' | 'Apolipoprotein B-100' |
| 5242 | 'APOB_No67' | 'LDFSSQADLRNEIK' | 'Apolipoprotein B-100' |
| 5243 | 'APOB_No68' | 'LVELAHQYK' | 'Apolipoprotein B-100' |
| 5244 | 'APOB_No69' | 'QSFDLSVK' | 'Apolipoprotein B-100' |
| 5245 | 'APOB_No70' | 'QTIIVVLENVQR' | 'Apolipoprotein B-100' |
| 5246 | 'APOB_No71' | 'DFSLWEK' | 'Apolipoprotein B-100' |
| 5247 | 'APOB_No72' | 'LIVAMSSWLQK' | 'Apolipoprotein B-100' |
| 5248 | 'APOB_No73' | 'FRETLEDTR' | 'Apolipoprotein B-100' |
| 5249 | 'APOB_No74' | 'NMEVSVATTTK' | 'Apolipoprotein B-100' |
| 5250 | 'APOB_No75' | 'TQFNNNEYSQDLDAYNTKDK' | 'Apolipoprotein B-100' |
| 5251 | 'APOB_No76' | 'LSNVLQQVK' | 'Apolipoprotein B-100' |
| 5252 | 'APOB_No77' | 'LFLEETK' | 'Apolipoprotein B-100' |
| 5253 | 'APOB_No78' | 'VNQNLVYESGSLNFSKLEIQSQVDSQHVGHSVLTAK' | 'Apolipoprotein B-100' |
| 5254 | 'APOB_No79' | 'LNDLNSVLVMPTFHVPFTDLQVPSCK' | 'Apolipoprotein B-100' |
| 5255 | 'APOB_No80' | 'LKTQFNNNEYSQDLDAYNTK' | 'Apolipoprotein B-100' |
| 5256 | 'APOB_No81' | 'TEVIPPLIENR' | 'Apolipoprotein B-100' |
| 5257 | 'APOB_No82' | 'TIHDLHLFIENIDFNK' | 'Apolipoprotein B-100' |
| 5258 | 'APOB_No83' | 'LAAYLMLMR' | 'Apolipoprotein B-100' |
| 5259 | 'APOB_No84' | 'GLLIFDASSSWGPQMSASVHLDSK' | 'Apolipoprotein B-100' |
| 5260 | 'APOB_No85' | 'IHSGSFQSQVELSNDQEK' | 'Apolipoprotein B-100' |
| 5261 | 'APOB_No86' | 'ITLPDFR' | 'Apolipoprotein B-100' |
| 5262 | 'APOB_No87' | 'ATGVLYDYVNK' | 'Apolipoprotein B-100' |
| 5263 | 'APOB_No88' | 'YEVDQQIQVLMDK' | 'Apolipoprotein B-100' |
| 5264 | 'APOB_No89' | 'HSITNPLAVLCEFISQSIK' | 'Apolipoprotein B-100' |
| 5265 | 'APOB_No90' | 'ITEVALMGHLSCDTK' | 'Apolipoprotein B-100' |
| 5266 | 'APOB_No91' | 'ITEVALMGHLSCDTKEER' | 'Apolipoprotein B-100' |
| 5267 | 'APOB_No92' | 'VIGNMGQTMEQLTPELK' | 'Apolipoprotein B-100' |
| 5268 | 'APOB_No93' | 'LLLQMDSSATAYGSTVSKR' | 'Apolipoprotein B-100' |
| 5269 | 'APOB_No94' | 'IKGVISIPR' | 'Apolipoprotein B-100' |
| 5270 | 'APOB_No95' | 'LNDLNSVLVMPTFHVPFTDLQVPSCK' | 'Apolipoprotein B-100' |
| 5271 | 'APOB_No96' | 'KITEVALMGHLSCDTK' | 'Apolipoprotein B-100' |
| 5272 | 'APOB_No97' | 'FFSLLSGSLNSHGLELNADILGTDK' | 'Apolipoprotein B-100' |
| 5273 | 'APOB_No98' | 'GMTRPLSTLISSSQSCQYTLDAK' | 'Apolipoprotein B-100' |
| 5274 | 'APOB_No99' | 'LEVANMQAELVAK' | 'Apolipoprotein B-100' |
| 5275 | 'APOB_No100' | 'MTSNFPVDLSDYPK' | 'Apolipoprotein B-100' |
| 5276 | 'APOB_No101' | 'KIISDYHQQFR' | 'Apolipoprotein B-100' |
| 5277 | 'APOB_No102' | 'QVFLYPEKDEPTYILNIKR' | 'Apolipoprotein B-100' |
| 5278 | 'APOB_No103' | 'NPNGYSFSIPVK' | 'Apolipoprotein B-100' |
| 5279 | 'APOB_No104' | 'IPSVQINFK' | 'Apolipoprotein B-100' |
| 5280 | 'APOB_No105' | 'DSYDLHDLK' | 'Apolipoprotein B-100' |
| 5281 | 'APOB_No106' | 'YEVDQQIQVLMDK' | 'Apolipoprotein B-100' |

| | A | B | C |
|---|---|---|---|
| 1 | **Peptide Name** | **Peptide Sequence** | **Peptide Description** |
| 5282 | 'APOB_No107' | 'RHIQNIDIQHLAGK' | 'Apolipoprotein B-100' |
| 5283 | 'APOB_No108' | 'ESDEETQIKVNWEEEAASGLLTSLK' | 'Apolipoprotein B-100' |
| 5284 | 'APOB_No109' | 'LDVTTSIGRR' | 'Apolipoprotein B-100' |
| 5285 | 'APOB_No110' | 'LSNDMMGSYAEMK' | 'Apolipoprotein B-100' |
| 5286 | 'APOB_No111' | 'IEIPLPFGGK' | 'Apolipoprotein B-100' |
| 5287 | 'APOB_No112' | 'TLQGIPQMIGEVIR' | 'Apolipoprotein B-100' |
| 5288 | 'APOB_No113' | 'EFQVPTFTIPK' | 'Apolipoprotein B-100' |
| 5289 | 'APOB_No114' | 'MTSNFPVDLSDYPK' | 'Apolipoprotein B-100' |
| 5290 | 'APOB_No115' | 'FLDMLIK' | 'Apolipoprotein B-100' |
| 5291 | 'APOB_No116' | 'QHIEAIDVR' | 'Apolipoprotein B-100' |
| 5292 | 'APOB_No117' | 'LEPLKLHVAGNLK' | 'Apolipoprotein B-100' |
| 5293 | 'APOB_No118' | 'LAAYLMLMR' | 'Apolipoprotein B-100' |
| 5294 | 'APOB_No119' | 'LSNDMMGSYAEMK' | 'Apolipoprotein B-100' |
| 5295 | 'APOB_No120' | 'LSNDMMGSYAEMK' | 'Apolipoprotein B-100' |
| 5296 | 'APOB_No121' | 'LSNDMMGSYAEMK' | 'Apolipoprotein B-100' |
| 5297 | 'APOB_No122' | 'MYQMDIQQELQR' | 'Apolipoprotein B-100' |
| 5298 | 'APOB_No123' | 'ATGVLYDYVNKYHWEHTGLTLR' | 'Apolipoprotein B-100' |
| 5299 | 'APOB_No124' | 'CVQSTKPSLMIQK' | 'Apolipoprotein B-100' |
| 5300 | 'APOB_No125' | 'ATFQTPDFIVPLTDLR' | 'Apolipoprotein B-100' |
| 5301 | 'APOB_No126' | 'FFSLLSGSLNSHGLELNADILGTDKINSGAHK' | 'Apolipoprotein B-100' |
| 5302 | 'APOB_No127' | 'TLQGIPQMIGEVIRK' | 'Apolipoprotein B-100' |
| 5303 | 'APOB_No128' | 'LVGFIDDAVKK' | 'Apolipoprotein B-100' |
| 5304 | 'APOB_No129' | 'RGIISALLVPPETEEAK' | 'Apolipoprotein B-100' |
| 5305 | 'APOB_No130' | 'GMTRPLSTLISSSQSCQYTLDAK' | 'Apolipoprotein B-100' |
| 5306 | 'APOB_No131' | 'NIQEYLSILTDPDGKGKEK' | 'Apolipoprotein B-100' |
| 5307 | 'APOB_No132' | 'LPQQANDYLNSFNWER' | 'Apolipoprotein B-100' |
| 5308 | 'APOB_No133' | 'LDVTTSIGR' | 'Apolipoprotein B-100' |
| 5309 | 'APOB_No134' | 'DDKHEQDMVNGIMLSVEK' | 'Apolipoprotein B-100' |
| 5310 | 'APOB_No135' | 'QHIEAIDVR' | 'Apolipoprotein B-100' |
| 5311 | 'APOB_No136' | 'AASGTTGTYQEWKDK' | 'Apolipoprotein B-100' |
| 5312 | 'APOB_No137' | 'SNTVASLHTEKNTLELSNGVIVK' | 'Apolipoprotein B-100' |
| 5313 | 'APOB_No138' | 'VPLLLSEPINIIDALEMR' | 'Apolipoprotein B-100' |
| 5314 | 'APOB_No139' | 'TSSFALNLPTLPEVKFPEVDVLTK' | 'Apolipoprotein B-100' |
| 5315 | 'APOB_No140' | 'YKLQDFSDQLSDYYEK' | 'Apolipoprotein B-100' |
| 5316 | 'APOB_No141' | 'LDFSSQADLR' | 'Apolipoprotein B-100' |
| 5317 | 'APOB_No142' | 'TKNSEEFAAAMSR' | 'Apolipoprotein B-100' |
| 5318 | 'APOB_No143' | 'YENYELTLK' | 'Apolipoprotein B-100' |
| 5319 | 'APOB_No144' | 'LSLESLTSYFSIESSTKGDVK' | 'Apolipoprotein B-100' |
| 5320 | 'APOB_No145' | 'MNFKQELNGNTK' | 'Apolipoprotein B-100' |
| 5321 | 'APOB_No146' | 'LQSTTVMNPYMK' | 'Apolipoprotein B-100' |
| 5322 | 'APOB_No147' | 'LSNDMMGSYAEMK' | 'Apolipoprotein B-100' |
| 5323 | 'APOB_No148' | 'SKPTVSSSMEFK' | 'Apolipoprotein B-100' |
| 5324 | 'APOB_No149' | 'KITEVALMGHLSCDTKEER' | 'Apolipoprotein B-100' |
| 5325 | 'APOB_No150' | 'LTLDIQNKK' | 'Apolipoprotein B-100' |

| | A | B | C |
|---|---|---|---|
| 1 | **Peptide Name** | **Peptide Sequence** | **Peptide Description** |
| 5326 | 'APOB_No151' | 'IDDIWNLEVKENFAGEATLQR' | 'Apolipoprotein B-100' |
| 5327 | 'APOB_No152' | 'LSNDMMGSYAEMK' | 'Apolipoprotein B-100' |
| 5328 | 'APOB_No153' | 'HVAEAICKEQHLFLPFSYK' | 'Apolipoprotein B-100' |
| 5329 | 'APOB_No154' | 'SVSDGIAALDLNAVANK' | 'Apolipoprotein B-100' |
| 5330 | 'APOB_No155' | 'LSNDMMGSYAEMK' | 'Apolipoprotein B-100' |
| 5331 | 'APOB_No156' | 'LEVLNFDFQANAQLSNPK' | 'Apolipoprotein B-100' |
| 5332 | 'APOB_No157' | 'CVQSTKPSLMIQK' | 'Apolipoprotein B-100' |
| 5333 | 'APOB_No158' | 'INPLALKESVK' | 'Apolipoprotein B-100' |
| 5334 | 'APOB_No159' | 'AEPLAFTFSHDYKGSTSHHLVSR' | 'Apolipoprotein B-100' |
| 5335 | 'APOB_No160' | 'NHLQLEGLFFTNGEHTSK' | 'Apolipoprotein B-100' |
| 5336 | 'APOB_No161' | 'WNFYYSPQSSPDK' | 'Apolipoprotein B-100' |
| 5337 | 'APOB_No162' | 'ALYWVNGQVPDGVSK' | 'Apolipoprotein B-100' |
| 5338 | 'APOB_No163' | 'QTVNLQLQPYSLVTTLNSDLK' | 'Apolipoprotein B-100' |
| 5339 | 'APOB_No164' | 'VQGVEFSHR' | 'Apolipoprotein B-100' |
| 5340 | 'APOB_No165' | 'VELEVPQLCSFILK' | 'Apolipoprotein B-100' |
| 5341 | 'APOB_No166' | 'TILGTMPAFEVSLQALQK' | 'Apolipoprotein B-100' |
| 5342 | 'APOB_No167' | 'DKAQNLYQELLTQEGQASFQGLK' | 'Apolipoprotein B-100' |
| 5343 | 'APOB_No168' | 'SEYQADYESLR' | 'Apolipoprotein B-100' |
| 5344 | 'APOB_No169' | 'ASGSLPYTQTLQDHLNSLK' | 'Apolipoprotein B-100' |
| 5345 | 'APOB_No170' | 'SVMAPFTMTIDAHTNGNGK' | 'Apolipoprotein B-100' |
| 5346 | 'APOB_No171' | 'VSTAFVYTK' | 'Apolipoprotein B-100' |
| 5347 | 'APOB_No172' | 'GTYGLSCQRDPNTGR' | 'Apolipoprotein B-100' |
| 5348 | 'APOB_No173' | 'LQSTTVMNPYMK' | 'Apolipoprotein B-100' |
| 5349 | 'APOB_No174' | 'IVQILPWEQNEQVK' | 'Apolipoprotein B-100' |
| 5350 | 'APOB_No175' | 'NSLKIEIPLPFGGK' | 'Apolipoprotein B-100' |
| 5351 | 'APOB_No176' | 'EIFNMAR' | 'Apolipoprotein B-100' |
| 5352 | 'APOB_No177' | 'NNALDFVTK' | 'Apolipoprotein B-100' |
| 5353 | 'APOB_No178' | 'ITEVALMGHLSCDTK' | 'Apolipoprotein B-100' |
| 5354 | 'APOB_No179' | 'SNTVASLHTEK' | 'Apolipoprotein B-100' |
| 5355 | 'APOB_No180' | 'LNELSFK' | 'Apolipoprotein B-100' |
| 5356 | 'APOB_No181' | 'VPLLLSEPINIIDALEMR' | 'Apolipoprotein B-100' |
| 5357 | 'APOB_No182' | 'AALGKLPQQANDYLNSFNWER' | 'Apolipoprotein B-100' |
| 5358 | 'APOB_No183' | 'LTIFKTELR' | 'Apolipoprotein B-100' |
| 5359 | 'APOB_No184' | 'MGLAFESTK' | 'Apolipoprotein B-100' |
| 5360 | 'APOB_No185' | 'ESQLPTVMDFRK' | 'Apolipoprotein B-100' |
| 5361 | 'APOB_No186' | 'VAWHYDEEKIEFEWNTGTNVDTKK' | 'Apolipoprotein B-100' |
| 5362 | 'APOB_No187' | 'HLIDSLIDFLNFPR' | 'Apolipoprotein B-100' |
| 5363 | 'APOB_No188' | 'DLKVEDIPLAR' | 'Apolipoprotein B-100' |
| 5364 | 'APOB_No189' | 'AASGTTGTYQEWK' | 'Apolipoprotein B-100' |
| 5365 | 'APOB_No190' | 'LSNDMMGSYAEMK' | 'Apolipoprotein B-100' |
| 5366 | 'APOB_No191' | 'KGNVATEISTER' | 'Apolipoprotein B-100' |
| 5367 | 'APOB_No192' | 'WNFYYSPQSSPDKK' | 'Apolipoprotein B-100' |
| 5368 | 'APOB_No193' | 'IEDGTLASK' | 'Apolipoprotein B-100' |
| 5369 | 'APOB_No194' | 'AGVKLEVANMQAELVAK' | 'Apolipoprotein B-100' |

| | A | B | C |
|---|---|---|---|
| 1 | **Peptide Name** | **Peptide Sequence** | **Peptide Description** |
| 5370 | 'APOB_No195' | 'VHELIER' | 'Apolipoprotein B-100' |
| 5371 | 'APOB_No196' | 'HKLIDVISMYR' | 'Apolipoprotein B-100' |
| 5372 | 'APOB_No197' | 'QVFLYPEKDEPTYILNIK' | 'Apolipoprotein B-100' |
| 5373 | 'APOB_No198' | 'ATLELSPWQMSALVQVHASQPSSFHDFPDLGQEVALNANTK' | 'Apolipoprotein B-100' |
| 5374 | 'APOB_No199' | 'FDHTNSLNIAGLSLDFSSK' | 'Apolipoprotein B-100' |
| 5375 | 'APOB_No200' | 'SVMAPFTMTIDAHTNGNGK' | 'Apolipoprotein B-100' |
| 5376 | 'APOB_No201' | 'GMALFGEGK' | 'Apolipoprotein B-100' |
| 5377 | 'APOB_No202' | 'KLQSTTVMNPYMK' | 'Apolipoprotein B-100' |
| 5378 | 'APOB_No203' | 'ATLELSPWQMSALVQVHASQPSSFHDFPDLGQEVALNANTK' | 'Apolipoprotein B-100' |
| 5379 | 'APOB_No204' | 'FRETLEDTRDR' | 'Apolipoprotein B-100' |
| 5380 | 'APOB_No205' | 'FVTQAEGAK' | 'Apolipoprotein B-100' |
| 5381 | 'APOB_No206' | 'QVFLYPEKDEPTYILNIKR' | 'Apolipoprotein B-100' |
| 5382 | 'APOB_No207' | 'MDMTFSK' | 'Apolipoprotein B-100' |
| 5383 | 'APOB_No208' | 'LQDFSDQLSDYYEK' | 'Apolipoprotein B-100' |
| 5384 | 'APOB_No209' | 'LSQLQTYMIQFDQYIK' | 'Apolipoprotein B-100' |
| 5385 | 'APOB_No210' | 'TTLTAFGFASADLIEIGLEGK' | 'Apolipoprotein B-100' |
| 5386 | 'APOB_No211' | 'ELLKDLSK' | 'Apolipoprotein B-100' |
| 5387 | 'APOB_No212' | 'SISAALEHK' | 'Apolipoprotein B-100' |
| 5388 | 'APOB_No213' | 'LGNNPVSK' | 'Apolipoprotein B-100' |
| 5389 | 'APOB_No214' | 'LDFREIQIYKK' | 'Apolipoprotein B-100' |
| 5390 | 'APOB_No215' | 'GISTSAASPAVGTVGMDMDEDDDFSKWNFYYSPQSSPDKK' | 'Apolipoprotein B-100' |
| 5391 | 'APOB_No216' | 'LELELRPTGEIEQYSVSATYELQR' | 'Apolipoprotein B-100' |
| 5392 | 'APOB_No217' | 'EELCTMFIR' | 'Apolipoprotein B-100' |
| 5393 | 'APOB_No218' | 'IAIANIIDEIIEK' | 'Apolipoprotein B-100' |
| 5394 | 'APOB_No219' | 'GTYGLSCQR' | 'Apolipoprotein B-100' |
| 5395 | 'APOB_No220' | 'NSLFFSAQPFEITASTNNEGNLK' | 'Apolipoprotein B-100' |
| 5396 | 'APOB_No221' | 'ALVEQGFTVPEIK' | 'Apolipoprotein B-100' |
| 5397 | 'APOB_No222' | 'YSQPEDSLIPFFEITVPESQLTVSQFTLPK' | 'Apolipoprotein B-100' |
| 5398 | 'APOB_No223' | 'YEVDQQIQVLMDKLVELAHQYK' | 'Apolipoprotein B-100' |
| 5399 | 'APOB_No224' | 'GNVATEISTERDLGQCDR' | 'Apolipoprotein B-100' |
| 5400 | 'APOB_No225' | 'STSPPKQAEAVLK' | 'Apolipoprotein B-100' |
| 5401 | 'APOB_No226' | 'VPSYTLILPSLELPVLHVPR' | 'Apolipoprotein B-100' |
| 5402 | 'APOB_No227' | 'SVMAPFTMTIDAHTNGNGK' | 'Apolipoprotein B-100' |
| 5403 | 'APOB_No228' | 'LLSGGNTLHLVSTTK' | 'Apolipoprotein B-100' |
| 5404 | 'APOB_No229' | 'LQSTTVMNPYMK' | 'Apolipoprotein B-100' |
| 5405 | 'APOB_No230' | 'VNQNLVYESGSLNFSK' | 'Apolipoprotein B-100' |
| 5406 | 'APOB_No231' | 'LLDHRVPQTDMTFR' | 'Apolipoprotein B-100' |
| 5407 | 'APOB_No232' | 'DKDQEVLLQTFLDDASPGDK' | 'Apolipoprotein B-100' |
| 5408 | 'APOB_No233' | 'SVMAPFTMTIDAHTNGNGK' | 'Apolipoprotein B-100' |
| 5409 | 'APOB_No234' | 'TGLKEFLK' | 'Apolipoprotein B-100' |
| 5410 | 'APOB_No235' | 'YDKNQDVHSINLPFFETLQEYFER' | 'Apolipoprotein B-100' |
| 5411 | 'APOB_No236' | 'ITEVALMGHLSCDTKEER' | 'Apolipoprotein B-100' |
| 5412 | 'APOB_No237' | 'SLDEHYHIR' | 'Apolipoprotein B-100' |
| 5413 | 'APOB_No238' | 'NSEEFAAAMSR' | 'Apolipoprotein B-100' |

| | A | B | C |
|---|---|---|---|
| 1 | **Peptide Name** | **Peptide Sequence** | **Peptide Description** |
| 5414 | 'APOB_No239' | 'GISTSAASPAVGTVGMDMDEDDDFSKWNFYYSPQSSPDKK' | 'Apolipoprotein B-100' |
| 5415 | 'APOB_No240' | 'EFNLQNMGLPDFHIPENLFLK' | 'Apolipoprotein B-100' |
| 5416 | 'APOB_No241' | 'AEPLAFTFSHDYK' | 'Apolipoprotein B-100' |
| 5417 | 'APOB_No242' | 'YGMVAQVTQTLKLEDTPK' | 'Apolipoprotein B-100' |
| 5418 | 'APOB_No243' | 'LLLQMDSSATAYGSTVSKR' | 'Apolipoprotein B-100' |
| 5419 | 'APOB_No244' | 'LKTQFNNNEYSQDLDAYNTKDK' | 'Apolipoprotein B-100' |
| 5420 | 'APOB_No245' | 'GIISALLVPPETEEAK' | 'Apolipoprotein B-100' |
| 5421 | 'APOB_No246' | 'TLQGIPQMIGEVIR' | 'Apolipoprotein B-100' |
| 5422 | 'APOB_No247' | 'LLDHRVPQTDMTFR' | 'Apolipoprotein B-100' |
| 5423 | 'APOB_No248' | 'GAYQNNEIKHIYAISSAALSASYK' | 'Apolipoprotein B-100' |
| 5424 | 'APOB_No249' | 'LNGESNLR' | 'Apolipoprotein B-100' |
| 5425 | 'APOB_No250' | 'QGFFPDSVNK' | 'Apolipoprotein B-100' |
| 5426 | 'APOB_No251' | 'LDNIYSSDK' | 'Apolipoprotein B-100' |
| 5427 | 'APOB_No252' | 'EFNLQNMGLPDFHIPENLFLK' | 'Apolipoprotein B-100' |
| 5428 | 'APOB_No253' | 'AALTELSLGSAYQAMILGVDSK' | 'Apolipoprotein B-100' |
| 5429 | 'APOB_No254' | 'GAYQNNEIK' | 'Apolipoprotein B-100' |
| 5430 | 'APOB_No255' | 'NRNNALDFVTK' | 'Apolipoprotein B-100' |
| 5431 | 'APOB_No256' | 'FLLKAEPLAFTFSHDYK' | 'Apolipoprotein B-100' |
| 5432 | 'APOB_No257' | 'AHLDIAGSLEGHLR' | 'Apolipoprotein B-100' |
| 5433 | 'APOB_No258' | 'NMEVSVATTTK' | 'Apolipoprotein B-100' |
| 5434 | 'APOB_No259' | 'DEPTYILNIK' | 'Apolipoprotein B-100' |
| 5435 | 'APOB_No260' | 'AQNLYQELLTQEGQASFQGLKDNVFDGLVR' | 'Apolipoprotein B-100' |
| 5436 | 'APOB_No261' | 'IPSVQINFKDLK' | 'Apolipoprotein B-100' |
| 5437 | 'APOB_No262' | 'LIDVISMYR' | 'Apolipoprotein B-100' |
| 5438 | 'APOB_No263' | 'LNTDIAGLASAIDMSTNYNSDSLHFSNVFR' | 'Apolipoprotein B-100' |
| 5439 | 'APOB_No264' | 'DDKHEQDMVNGIMLSVEK' | 'Apolipoprotein B-100' |
| 5440 | 'APOB_No265' | 'EVYGFNPEGK' | 'Apolipoprotein B-100' |
| 5441 | 'APOB_No266' | 'ESQLPTVMDFR' | 'Apolipoprotein B-100' |
| 5442 | 'APOB_No267' | 'DKDQEVLLQTFLDDASPGDKR' | 'Apolipoprotein B-100' |
| 5443 | 'APOB_No268' | 'GAVDHKLSLESLTSYFSIESSTKGDVK' | 'Apolipoprotein B-100' |
| 5444 | 'APOB_No269' | 'GISTSAASPAVGTVGMDMDEDDDFSKWNFYYSPQSSPDKK' | 'Apolipoprotein B-100' |
| 5445 | 'APOB_No270' | 'NKYGMVAQVTQTLK' | 'Apolipoprotein B-100' |
| 5446 | 'APOB_No271' | 'LHVAGNLK' | 'Apolipoprotein B-100' |
| 5447 | 'APOB_No272' | 'VSALLTPAEQTGTWK' | 'Apolipoprotein B-100' |
| 5448 | 'APOB_No273' | 'NSEEFAAAMSR' | 'Apolipoprotein B-100' |
| 5449 | 'APOB_No274' | 'AVSMPSFSILGSDVR' | 'Apolipoprotein B-100' |
| 5450 | 'APOB_No275' | 'ALVDTLKFVTQAEGAK' | 'Apolipoprotein B-100' |
| 5451 | 'APOB_No276' | 'LPYTIITTPPLKDFSLWEK' | 'Apolipoprotein B-100' |
| 5452 | 'APOB_No277' | 'VLVDHFGYTK' | 'Apolipoprotein B-100' |
| 5453 | 'APOB_No278' | 'LATALSLSNK' | 'Apolipoprotein B-100' |
| 5454 | 'APOB_No279' | 'LNTDIAGLASAIDMSTNYNSDSLHFSNVFR' | 'Apolipoprotein B-100' |
| 5455 | 'APOB_No280' | 'ELLKDLSKEAQEVFK' | 'Apolipoprotein B-100' |
| 5456 | 'APOB_No281' | 'LEVANMQAELVAK' | 'Apolipoprotein B-100' |
| 5457 | 'APOB_No282' | 'FLLKAEPLAFTFSHDYKGSTSHHLVSR' | 'Apolipoprotein B-100' |

| | A | B | C |
|---|---|---|---|
| 1 | **Peptide Name** | **Peptide Sequence** | **Peptide Description** |
| 5458 | 'APOB_No283' | 'KLTIFKTELR' | 'Apolipoprotein B-100' |
| 5459 | 'APOB_No284' | 'SLHMYANR' | 'Apolipoprotein B-100' |
| 5460 | 'APOB_No285' | 'GAYQNNEIKHIYAISSAALSASYKADTVAK' | 'Apolipoprotein B-100' |
| 5461 | 'APOB_No286' | 'KITEVALMGHLSCDTKEER' | 'Apolipoprotein B-100' |
| 5462 | 'APOB_No287' | 'SKPTVSSSMEFK' | 'Apolipoprotein B-100' |
| 5463 | 'APOB_No288' | 'TLQGIPQMIGEVIRK' | 'Apolipoprotein B-100' |
| 5464 | 'APOB_No289' | 'ESQLPTVMDFR' | 'Apolipoprotein B-100' |
| 5465 | 'APOB_No290' | 'CSLLVLENELNAELGLSGASMK' | 'Apolipoprotein B-100' |
| 5466 | 'APOB_No291' | 'FSVPAGIVIPSFQALTAR' | 'Apolipoprotein B-100' |
| 5467 | 'APOB_No292' | 'FQFPGKPGIYTREELCTMFIR' | 'Apolipoprotein B-100' |
| 5468 | 'APOB_No293' | 'IAIANIIDEIIEKLK' | 'Apolipoprotein B-100' |
| 5469 | 'APOB_No294' | 'HIYAISSAALSASYK' | 'Apolipoprotein B-100' |
| 5470 | 'APOB_No295' | 'LLKENLCLNLHK' | 'Apolipoprotein B-100' |
| 5471 | 'APOB_No296' | 'IEGNLIFDPNNYLPK' | 'Apolipoprotein B-100' |
| 5472 | 'APOB_No297' | 'KHVAEAICKEQHLFLPFSYK' | 'Apolipoprotein B-100' |
| 5473 | 'APOB_No298' | 'VRESDEETQIKVNWEEEAASGLLTSLK' | 'Apolipoprotein B-100' |
| 5474 | 'APOB_No299' | 'TPALHFK' | 'Apolipoprotein B-100' |
| 5475 | 'APOB_No300' | 'NTASLKYENYELTLK' | 'Apolipoprotein B-100' |
| 5476 | 'APOB_No301' | 'HINIDQFVR' | 'Apolipoprotein B-100' |
| 5477 | 'APOB_No302' | 'LPYTIITTPPLK' | 'Apolipoprotein B-100' |
| 5478 | 'APOB_No303' | 'ADSVVDLLSYNVQGSGETTYDHK' | 'Apolipoprotein B-100' |
| 5479 | 'APOB_No304' | 'LELELRPTGEIEQYSVSATYELQREDR' | 'Apolipoprotein B-100' |
| 5480 | 'APOB_No305' | 'IHSGSFQSQVELSNDQEKAHLDIAGSLEGHLR' | 'Apolipoprotein B-100' |
| 5481 | 'APOB_No306' | 'FLDSNIKFSHVEK' | 'Apolipoprotein B-100' |
| 5482 | 'APOB_No307' | 'DAVEKPQEFTIVAFVKYDK' | 'Apolipoprotein B-100' |
| 5483 | 'APOB_No308' | 'VRESDEETQIK' | 'Apolipoprotein B-100' |
| 5484 | 'APOB_No309' | 'EALKESQLPTVMDFR' | 'Apolipoprotein B-100' |
| 5485 | 'APOB_No310' | 'IQIQEKLQQLKR' | 'Apolipoprotein B-100' |
| 5486 | 'APOB_No311' | 'LAAYLMLMR' | 'Apolipoprotein B-100' |
| 5487 | 'APOB_No312' | 'VLVDHFGYTKDDKHEQDMVNGIMLSVEK' | 'Apolipoprotein B-100' |
| 5488 | 'APOB_No313' | 'LLSGGNTLHLVSTTKTEVIPPLIENR' | 'Apolipoprotein B-100' |
| 5489 | 'APOB_No314' | 'SPAFTDLHLR' | 'Apolipoprotein B-100' |
| 5490 | 'APOB_No315' | 'EALKESQLPTVMDFRK' | 'Apolipoprotein B-100' |
| 5491 | 'APOB_No316' | 'KSISAALEHKVSALLTPAEQTGTWK' | 'Apolipoprotein B-100' |
| 5492 | 'APOB_No317' | 'DLSKEAQEVFK' | 'Apolipoprotein B-100' |
| 5493 | 'APOB_No318' | 'EVKIDGQFR' | 'Apolipoprotein B-100' |
| 5494 | 'APOB_No319' | 'VLVDHFGYTKDDKHEQDMVNGIMLSVEK' | 'Apolipoprotein B-100' |
| 5495 | 'APOB_No320' | 'YNALDLTNNGKLR' | 'Apolipoprotein B-100' |
| 5496 | 'APOB_No321' | 'QGFFPDSVNK' | 'Apolipoprotein B-100' |
| 5497 | 'APOB_No322' | 'ATVAVYLESLQDTK' | 'Apolipoprotein B-100' |
| 5498 | 'APOB_No323' | 'NKYGMVAQVTQTLKLEDTPK' | 'Apolipoprotein B-100' |
| 5499 | 'APOB_No324' | 'AVSMPSFSILGSDVR' | 'Apolipoprotein B-100' |
| 5500 | 'APOB_No325' | 'TQFNNNEYSQDLDAYNTK' | 'Apolipoprotein B-100' |
| 5501 | 'APOB_No326' | 'AHLNIKSPAFTDLHLR' | 'Apolipoprotein B-100' |

| | A | B | C |
|---|---|---|---|
| 1 | **Peptide Name** | **Peptide Sequence** | **Peptide Description** |
| 5502 | 'APOB_No327' | 'NIQEYLSILTDPDGK' | 'Apolipoprotein B-100' |
| 5503 | 'APOB_No328' | 'LLLQMDSSATAYGSTVSK' | 'Apolipoprotein B-100' |
| 5504 | 'APOB_No329' | 'ANLFNKLVTELR' | 'Apolipoprotein B-100' |
| 5505 | 'APOB_No330' | 'YEDGTLSLTSTSDLQSGIIK' | 'Apolipoprotein B-100' |
| 5506 | 'APOB_No331' | 'TSSFALNLPTLPEVK' | 'Apolipoprotein B-100' |
| 5507 | 'APOB_No332' | 'TSQCTLKEVYGFNPEGK' | 'Apolipoprotein B-100' |
| 5508 | 'APOB_No333' | 'SRATLYALSHAVNNYHK' | 'Apolipoprotein B-100' |
| 5509 | 'APOB_No334' | 'VIGNMGQTMEQLTPELK' | 'Apolipoprotein B-100' |
| 5510 | 'APOB_No335' | 'AALTELSLGSAYQAMILGVDSK' | 'Apolipoprotein B-100' |
| 5511 | 'APOB_No336' | 'VKHLIDSLIDFLNFPR' | 'Apolipoprotein B-100' |
| 5512 | 'APOB_No337' | 'NLTDFAEQYSIQDWAK' | 'Apolipoprotein B-100' |
| 5513 | 'APOB_No338' | 'KMTSNFPVDLSDYPK' | 'Apolipoprotein B-100' |
| 5514 | 'APOB_No339' | 'GMTRPLSTLISSSQSCQYTLDAKR' | 'Apolipoprotein B-100' |
| 5515 | 'APOB_No340' | 'EELCTMFIR' | 'Apolipoprotein B-100' |
| 5516 | 'APOB_No341' | 'ILGEELGFASLHDLQLLGK' | 'Apolipoprotein B-100' |
| 5517 | 'APOB_No342' | 'GNVATEISTER' | 'Apolipoprotein B-100' |
| 5518 | 'APOB_No343' | 'ENLCLNLHK' | 'Apolipoprotein B-100' |
| 5519 | 'APOB_No344' | 'KMTSNFPVDLSDYPK' | 'Apolipoprotein B-100' |
| 5520 | 'APOB_No345' | 'LQDFSDQLSDYYEKFIAESKR' | 'Apolipoprotein B-100' |
| 5521 | 'APOB_No346' | 'VLLDQLGTTISFER' | 'Apolipoprotein B-100' |
| 5522 | 'APOB_No347' | 'SLWDFLK' | 'Apolipoprotein B-100' |
| 5523 | 'APOB_No348' | 'LSQLQTYMIQFDQYIK' | 'Apolipoprotein B-100' |
| 5524 | 'APOB_No349' | 'VIGNMGQTMEQLTPELK' | 'Apolipoprotein B-100' |
| 5525 | 'APOB_No350' | 'NIQEYLSILTDPDGKGK' | 'Apolipoprotein B-100' |
| 5526 | 'APOB_No351' | 'LLLQMDSSATAYGSTVSK' | 'Apolipoprotein B-100' |
| 5527 | 'APOB_No352' | 'MYQMDIQQELQR' | 'Apolipoprotein B-100' |
| 5528 | 'APOB_No353' | 'SVGFHLPSREFQVPTFTIPK' | 'Apolipoprotein B-100' |
| 5529 | 'APOB_No354' | 'HINIDQFVRK' | 'Apolipoprotein B-100' |
| 5530 | 'APOB_No355' | 'KLNELSFK' | 'Apolipoprotein B-100' |
| 5531 | 'APOB_No356' | 'EEYFDPSIVGWTVK' | 'Apolipoprotein B-100' |
| 5532 | 'APOB_No357' | 'SISAALEHKVSALLTPAEQTGTWK' | 'Apolipoprotein B-100' |
| 5533 | 'APOB_No358' | 'KGNVATEISTERDLGQCDR' | 'Apolipoprotein B-100' |
| 5534 | 'APOB_No359' | 'DLGQCDR' | 'Apolipoprotein B-100' |
| 5535 | 'APOB_No360' | 'SPSQADINKIVQILPWEQNEQVK' | 'Apolipoprotein B-100' |
| 5536 | 'APOB_No361' | 'IKDYFEK' | 'Apolipoprotein B-100' |
| 5537 | 'APOB_No362' | 'DDKHEQDMVNGIMLSVEK' | 'Apolipoprotein B-100' |
| 5538 | 'APOB_No363' | 'NLQNNAEWVYQGAIR' | 'Apolipoprotein B-100' |
| 5539 | 'APOB_No364' | 'SGSSTASWIQNVDTKYQIR' | 'Apolipoprotein B-100' |
| 5540 | 'APOB_No365' | 'TKNSEEFAAAMSR' | 'Apolipoprotein B-100' |
| 5541 | 'APOB_No366' | 'LRLEPLKLHVAGNLK' | 'Apolipoprotein B-100' |
| 5542 | 'APOB_No367' | 'TQFNNNEYSQDLDAYNTKDKIGVELTGR' | 'Apolipoprotein B-100' |
| 5543 | 'APOB_No368' | 'QTEATMTFK' | 'Apolipoprotein B-100' |
| 5544 | 'APOB_No369' | 'MGLAFESTK' | 'Apolipoprotein B-100' |
| 5545 | 'APOB_No370' | 'CSLLVLENELNAELGLSGASMK' | 'Apolipoprotein B-100' |

| | A | B | C |
|---|---|---|---|
| 1 | **Peptide Name** | **Peptide Sequence** | **Peptide Description** |
| 5546 | 'APOB_No371' | 'KLQSTTVMNPYMK' | 'Apolipoprotein B-100' |
| 5547 | 'APOB_No372' | 'ESQLPTVMDFRK' | 'Apolipoprotein B-100' |
| 5548 | 'APOB_No373' | 'LDFREIQIYK' | 'Apolipoprotein B-100' |
| 5549 | 'APOB_No374' | 'DKAQNLYQELLTQEGQASFQGLKDNVFDGLVR' | 'Apolipoprotein B-100' |
| 5550 | 'APOB_No375' | 'SVSLPSLDPASAK' | 'Apolipoprotein B-100' |
| 5551 | 'APOB_No376' | 'KGISTSAASPAVGTVGMDMDEDDDFSK' | 'Apolipoprotein B-100' |
| 5552 | 'APOB_No377' | 'DDKHEQDMVNGIMLSVEK' | 'Apolipoprotein B-100' |
| 5553 | 'APOB_No378' | 'IQIQEKLQQLK' | 'Apolipoprotein B-100' |
| 5554 | 'APOB_No379' | 'RNLQNNAEWVYQGAIR' | 'Apolipoprotein B-100' |
| 5555 | 'APOB_No380' | 'TEHGSEMLFFGNAIEGK' | 'Apolipoprotein B-100' |
| 5556 | 'APOB_No381' | 'VSTAFVYTKNPNGYSFSIPVK' | 'Apolipoprotein B-100' |
| 5557 | 'APOB_No382' | 'SHDELPR' | 'Apolipoprotein B-100' |
| 5558 | 'APOB_No383' | 'ESDEETQIKVNWEEEAASGLLTSLKDNVPK' | 'Apolipoprotein B-100' |
| 5559 | 'APOB_No384' | 'YGMVAQVTQTLKLEDTPK' | 'Apolipoprotein B-100' |
| 5560 | 'APOB_No385' | 'HVAEAICK' | 'Apolipoprotein B-100' |
| 5561 | 'APOB_No386' | 'NLKHINIDQFVR' | 'Apolipoprotein B-100' |
| 5562 | 'APOB_No387' | 'EKIAELSATAQEIIK' | 'Apolipoprotein B-100' |
| 5563 | 'APOB_No388' | 'FNSSYLQGTNQITGR' | 'Apolipoprotein B-100' |
| 5564 | 'APOB_No389' | 'VEDIPLAR' | 'Apolipoprotein B-100' |
| 5565 | 'APOB_No390' | 'GLLIFDASSSWGPQMSASVHLDSK' | 'Apolipoprotein B-100' |
| 5566 | 'APOB_No391' | 'ENFAGEATLQR' | 'Apolipoprotein B-100' |
| 5567 | 'APOB_No392' | 'MNFKQELNGNTK' | 'Apolipoprotein B-100' |
| 5568 | 'APOB_No393' | 'IYSLWEHSTKNHLQLEGLFFTNGEHTSK' | 'Apolipoprotein B-100' |
| 5569 | 'APOB_No394' | 'QTVNLQLQPYSLVTTLNSDLK' | 'Apolipoprotein B-100' |
| 5570 | 'APOB_No395' | 'QTEATMTFKYNR' | 'Apolipoprotein B-100' |
| 5571 | 'APOB_No396' | 'KITEVALMGHLSCDTK' | 'Apolipoprotein B-100' |
| 5572 | 'APOB_No397' | 'PLSTLISSSQSCQYTLDAK' | 'Apolipoprotein B-100' |
| 5573 | 'APOB_No398' | 'YELKLAIPEGK' | 'Apolipoprotein B-100' |
| 5574 | 'APOB_No399' | 'GFEPTLEALFGKQGFFPDSVNK' | 'Apolipoprotein B-100' |
| 5575 | 'APOB_No400' | 'VLVDHFGYTKDDK' | 'Apolipoprotein B-100' |
| 5576 | 'APOB_No401' | 'ITEVALMGHLSCDTKEERK' | 'Apolipoprotein B-100' |
| 5577 | 'APOB_No402' | 'TIDQMLNSELQWPVPDIYLR' | 'Apolipoprotein B-100' |
| 5578 | 'APOB_No403' | 'HKLIDVISMYR' | 'Apolipoprotein B-100' |
| 5579 | 'APOB_No404' | 'HIYAISSAALSASYKADTVAK' | 'Apolipoprotein B-100' |
| 5580 | 'APOB_No405' | 'AGHIAWTSSGK' | 'Apolipoprotein B-100' |
| 5581 | 'APOB_No406' | 'GMALFGEGKAEFTGR' | 'Apolipoprotein B-100' |
| 5582 | 'APOB_No407' | 'LFLEETKATVAVYLESLQDTK' | 'Apolipoprotein B-100' |
| 5583 | 'APOB_No408' | 'IDFLNNYALFLSPSAQQASWQVSAR' | 'Apolipoprotein B-100' |
| 5584 | 'APOB_No409' | 'SPSQADINK' | 'Apolipoprotein B-100' |
| 5585 | 'APOB_No410' | 'INNQLTLDSNTKYFHK' | 'Apolipoprotein B-100' |
| 5586 | 'APOB_No411' | 'ASGSLPYTQTLQDHLNSLKEFNLQNMGLPDFHIPENLFLK' | 'Apolipoprotein B-100' |
| 5587 | 'APOB_No412' | 'AEPLAFTFSHDYKGSTSHHLVSRK' | 'Apolipoprotein B-100' |
| 5588 | 'APOB_No413' | 'ITENDIQIALDDAKINFNEK' | 'Apolipoprotein B-100' |
| 5589 | 'APOB_No414' | 'DFSAEYEEDGKYEGLQEWEGK' | 'Apolipoprotein B-100' |

| | A | B | C |
|---|---|---|---|
| 1 | **Peptide Name** | **Peptide Sequence** | **Peptide Description** |
| 5590 | 'APOB_No415' | 'IAELSATAQEIIKSQAIATKK' | 'Apolipoprotein B-100' |
| 5591 | 'APOB_No416' | 'GTFAHRDFSAEYEEDGKYEGLQEWEGK' | 'Apolipoprotein B-100' |
| 5592 | 'APOB_No417' | 'FLDMLIKK' | 'Apolipoprotein B-100' |
| 5593 | 'APOB_No418' | 'FLDMLIK' | 'Apolipoprotein B-100' |
| 5594 | 'APOB_No419' | 'VSSFYAK' | 'Apolipoprotein B-100' |
| 5595 | 'APOB_No420' | 'GAVDHKLSLESLTSYFSIESSTK' | 'Apolipoprotein B-100' |
| 5596 | 'APOB_No421' | 'VIGNMGQTMEQLTPELKSSILK' | 'Apolipoprotein B-100' |
| 5597 | 'APOB_No422' | 'GTYGLSCQRDPNTGRLNGESNLR' | 'Apolipoprotein B-100' |
| 5598 | 'APOB_No423' | 'LTGKIDFLNNYALFLSPSAQQASWQVSAR' | 'Apolipoprotein B-100' |
| 5599 | 'APOB_No424' | 'RLAAYLMLMR' | 'Apolipoprotein B-100' |
| 5600 | 'APOB_No425' | 'QTEATMTFK' | 'Apolipoprotein B-100' |
| 5601 | 'APOB_No426' | 'LKSFDYHQFVDETNDK' | 'Apolipoprotein B-100' |
| 5602 | 'APOB_No427' | 'IEGNLIFDPNNYLPKESMLK' | 'Apolipoprotein B-100' |
| 5603 | 'APOB_No428' | 'LNGEIQALELPQK' | 'Apolipoprotein B-100' |
| 5604 | 'APOB_No429' | 'FSHVEKLGNNPVSK' | 'Apolipoprotein B-100' |
| 5605 | 'APOB_No430' | 'LSLESLTSYFSIESSTK' | 'Apolipoprotein B-100' |
| 5606 | 'APOB_No431' | 'QIDDIDVR' | 'Apolipoprotein B-100' |
| 5607 | 'APOB_No432' | 'IEFEWNTGTNVDTKK' | 'Apolipoprotein B-100' |
| 5608 | 'APOB_No433' | 'TLQELKK' | 'Apolipoprotein B-100' |
| 5609 | 'APOB_No434' | 'KSISAALEHK' | 'Apolipoprotein B-100' |
| 5610 | 'APOB_No435' | 'LIDVISMYR' | 'Apolipoprotein B-100' |
| 5611 | 'APOB_No436' | 'YLRTEHGSEMLFFGNAIEGK' | 'Apolipoprotein B-100' |
| 5612 | 'APOB_No437' | 'AEALKLFLEETK' | 'Apolipoprotein B-100' |
| 5613 | 'APOB_No438' | 'QSWSVCK' | 'Apolipoprotein B-100' |
| 5614 | 'APOB_No439' | 'LRTSSFALNLPTLPEVKFPEVDVLTK' | 'Apolipoprotein B-100' |
| 5615 | 'APOB_No440' | 'VTQEFHMK' | 'Apolipoprotein B-100' |
| 5616 | 'APOB_No441' | 'YYELEEKIVSLIK' | 'Apolipoprotein B-100' |
| 5617 | 'APOB_No442' | 'LAAYLMLMR' | 'Apolipoprotein B-100' |
| 5618 | 'APOB_No443' | 'NKYGMVAQVTQTLKLEDTPK' | 'Apolipoprotein B-100' |
| 5619 | 'APOB_No444' | 'EIQIYKK' | 'Apolipoprotein B-100' |
| 5620 | 'APOB_No445' | 'QIDDIDVRFQK' | 'Apolipoprotein B-100' |
| 5621 | 'APOB_No446' | 'LKSFDYHQFVDETNDKIR' | 'Apolipoprotein B-100' |
| 5622 | 'APOB_No447' | 'TIDQMLNSELQWPVPDIYLR' | 'Apolipoprotein B-100' |
| 5623 | 'APOB_No448' | 'FLDMLIKK' | 'Apolipoprotein B-100' |
| 5624 | 'APOB_No449' | 'SVSLPSLDPASAKIEGNLIFDPNNYLPK' | 'Apolipoprotein B-100' |
| 5625 | 'APOB_No450' | 'EALKESQLPTVMDFR' | 'Apolipoprotein B-100' |
| 5626 | 'APOB_No451' | 'LNIPKLDFSSQADLR' | 'Apolipoprotein B-100' |
| 5627 | 'APOB_No452' | 'EALKESQLPTVMDFRK' | 'Apolipoprotein B-100' |
| 5628 | 'APOB_No453' | 'MDMTFSK' | 'Apolipoprotein B-100' |
| 5629 | 'APOB_No454' | 'MDMTFSKQNALLR' | 'Apolipoprotein B-100' |
| 5630 | 'APOB_No455' | 'SLHMYANR' | 'Apolipoprotein B-100' |
| 5631 | 'APOB_No456' | 'VSQEGLKLSNDMMGSYAEMK' | 'Apolipoprotein B-100' |
| 5632 | 'APOB_No457' | 'LQGTSKIDDIWNLEVKENFAGEATLQR' | 'Apolipoprotein B-100' |
| 5633 | 'APOB_No458' | 'AGVKLEVANMQAELVAK' | 'Apolipoprotein B-100' |

| | A | B | C |
|---|---|---|---|
| 1 | **Peptide Name** | **Peptide Sequence** | **Peptide Description** |
| 5634 | 'APOB_No459' | 'DFSAEYEEDGKYEGLQEWEGKAHLNIK' | 'Apolipoprotein B-100' |
| 5635 | 'APOB_No460' | 'KLQSTTVMNPYMK' | 'Apolipoprotein B-100' |
| 5636 | 'APOB_No461' | 'LVKEALKESQLPTVMDFR' | 'Apolipoprotein B-100' |
| 5637 | 'APOB_No462' | 'NFATSNKMDMTFSK' | 'Apolipoprotein B-100' |
| 5638 | 'APOB_No463' | 'INPLALKESVKFSSK' | 'Apolipoprotein B-100' |
| 5639 | 'APOB_No464' | 'LNIPKLDFSSQADLRNEIK' | 'Apolipoprotein B-100' |
| 5640 | 'APOB_No465' | 'ETLEDTRDR' | 'Apolipoprotein B-100' |
| 5641 | 'APOB_No466' | 'SEILAHWSPAKLLLQMDSSATAYGSTVSKR' | 'Apolipoprotein B-100' |
| 5642 | 'APOB_No467' | 'VNDESTEGKTSYR' | 'Apolipoprotein B-100' |
| 5643 | 'APOB_No468' | 'IEFEWNTGTNVDTK' | 'Apolipoprotein B-100' |
| 5644 | 'APOB_No469' | 'YEDGTLSLTSTSDLQSGIIKNTASLK' | 'Apolipoprotein B-100' |
| 5645 | 'APOB_No470' | 'NTASLKYENYELTLKSDTNGK' | 'Apolipoprotein B-100' |
| 5646 | 'APOB_No471' | 'MDMTFSK' | 'Apolipoprotein B-100' |
| 5647 | 'APOB_No472' | 'NFATSNKMDMTFSKQNALLR' | 'Apolipoprotein B-100' |
| 5648 | 'APOB_No473' | 'NLKHINIDQFVRK' | 'Apolipoprotein B-100' |
| 5649 | 'APOB_No474' | 'HKFLDSNIKFSHVEK' | 'Apolipoprotein B-100' |
| 5650 | 'APOB_No475' | 'IAELSATAQEIIKSQAIATK' | 'Apolipoprotein B-100' |
| 5651 | 'APOB_No476' | 'FFGEGTKK' | 'Apolipoprotein B-100' |
| 5652 | 'APOB_No477' | 'KGISTSAASPAVGTVGMDMDEDDDFSK' | 'Apolipoprotein B-100' |
| 5653 | 'APOB_No478' | 'ADTVAKVQGVEFSHR' | 'Apolipoprotein B-100' |
| 5654 | 'APOB_No479' | 'LRTSSFALNLPTLPEVK' | 'Apolipoprotein B-100' |
| 5655 | 'APOB_No480' | 'YTLNKNSLKIEIPLPFGGK' | 'Apolipoprotein B-100' |
| 5656 | 'APOB_No481' | 'NKYGMVAQVTQTLK' | 'Apolipoprotein B-100' |
| 5657 | 'APOB_No482' | 'LSLESLTSYFSIESSTKGDVKGSVLSR' | 'Apolipoprotein B-100' |
| 5658 | 'APOB_No483' | 'INNQLTLDSNTKYFHKLNIPK' | 'Apolipoprotein B-100' |
| 5659 | 'APOB_No484' | 'LKETIQK' | 'Apolipoprotein B-100' |
| 5660 | 'APOB_No485' | 'YLRTEHGSEMLFFGNAIEGK' | 'Apolipoprotein B-100' |
| 5661 | 'APOB_No486' | 'HIQNIDIQHLAGKLK' | 'Apolipoprotein B-100' |
| 5662 | 'APOB_No487' | 'GISTSAASPAVGTVGMDMDEDDDFSKWNFYYSPQSSPDKK' | 'Apolipoprotein B-100' |
| 5663 | 'APOB_No488' | 'IEGNLIFDPNNYLPKESMLK' | 'Apolipoprotein B-100' |
| 5664 | 'APOB_No489' | 'VHELIERYEVDQQIQVLMDK' | 'Apolipoprotein B-100' |
| 5665 | 'APOB_No490' | 'EDRALVDTLKFVTQAEGAK' | 'Apolipoprotein B-100' |
| 5666 | 'APOB_No491' | 'AKVHELIER' | 'Apolipoprotein B-100' |
| 5667 | 'APOB_No492' | 'AEFTGRHDAHLNGK' | 'Apolipoprotein B-100' |
| 5668 | 'APOB_No493' | 'YHMKADSVVDLLSYNVQGSGETTYDHK' | 'Apolipoprotein B-100' |
| 5669 | 'APOB_No494' | 'EYSGTIASEANTYLNSK' | 'Apolipoprotein B-100' |
| 5670 | 'APOB_No495' | 'GMALFGEGKAEFTGRHDAHLNGK' | 'Apolipoprotein B-100' |
| 5671 | 'APOB_No496' | 'FLDSNIKFSHVEKLGNNPVSK' | 'Apolipoprotein B-100' |
| 5672 | 'APOB_No497' | 'DQEVLLQTFLDDASPGDKR' | 'Apolipoprotein B-100' |
| 5673 | 'APOB_No498' | 'EKLTALTK' | 'Apolipoprotein B-100' |
| 5674 | 'APOB_No499' | 'GISTSAASPAVGTVGMDMDEDDDFSK' | 'Apolipoprotein B-100' |
| 5675 | 'APOB_No500' | 'MYQMDIQQELQR' | 'Apolipoprotein B-100' |
| 5676 | 'APOB_No501' | 'GMALFGEGKAEFTGR' | 'Apolipoprotein B-100' |
| 5677 | 'APOB_No502' | 'QVLFLDTVYGNCSTHFTVK' | 'Apolipoprotein B-100' |

| | A | B | C |
|---|---|---|---|
| 1 | **Peptide Name** | **Peptide Sequence** | **Peptide Description** |
| 5678 | 'APOB_No503' | 'HKFLDSNIK' | 'Apolipoprotein B-100' |
| 5679 | 'APOB_No504' | 'DLKMLETVR' | 'Apolipoprotein B-100' |
| 5680 | 'APOB_No505' | 'YGMVAQVTQTLKLEDTPKINSR' | 'Apolipoprotein B-100' |
| 5681 | 'APOB_No506' | 'QIDDIDVRFQK' | 'Apolipoprotein B-100' |
| 5682 | 'APOB_No507' | 'NFATSNKMDMTFSK' | 'Apolipoprotein B-100' |
| 5683 | 'APOB_No508' | 'FIAESKR' | 'Apolipoprotein B-100' |
| 5684 | 'APOB_No509' | 'YENYELTLKSDTNGK' | 'Apolipoprotein B-100' |
| 5685 | 'APOB_No510' | 'NLTDFAEQYSIQDWAKR' | 'Apolipoprotein B-100' |
| 5686 | 'APOB_No511' | 'ATGVLYDYVNKYHWEHTGLTLREVSSK' | 'Apolipoprotein B-100' |
| 5687 | 'APOB_No512' | 'SFDYHQFVDETNDKIREVTQR' | 'Apolipoprotein B-100' |
| 5688 | 'APOB_No513' | 'NFATSNKMDMTFSK' | 'Apolipoprotein B-100' |
| 5689 | 'APOB_No514' | 'PQEFTIVAFVK' | 'Apolipoprotein B-100' |
| 5690 | 'APOB_No515' | 'VIGNMGQTMEQLTPELKSSILK' | 'Apolipoprotein B-100' |
| 5691 | 'APOB_No516' | 'NFATSNKMDMTFSK' | 'Apolipoprotein B-100' |
| 5692 | 'APOB_No517' | 'TTKQSFDLSVK' | 'Apolipoprotein B-100' |
| 5693 | 'APOB_No518' | 'ITEVALMGHLSCDTKEERK' | 'Apolipoprotein B-100' |
| 5694 | 'APOB_No519' | 'EQHLFLPFSYKNK' | 'Apolipoprotein B-100' |
| 5695 | 'APOB_No520' | 'ETLEDTR' | 'Apolipoprotein B-100' |
| 5696 | 'APOB_No521' | 'QAEAVLKTLQELK' | 'Apolipoprotein B-100' |
| 5697 | 'APOB_No522' | 'VIGTLKNSLFFSAQPFEITASTNNEGNLK' | 'Apolipoprotein B-100' |
| 5698 | 'APOB_No523' | 'DLGQCDRFKPIR' | 'Apolipoprotein B-100' |
| 5699 | 'APOB_No524' | 'LDVTTSIGRRQHLR' | 'Apolipoprotein B-100' |
| 5700 | 'APOB_No525' | 'QSFDLSVKAQYKK' | 'Apolipoprotein B-100' |
| 5701 | 'APOB_No526' | 'PTGEIEQYSVSATYELQR' | 'Apolipoprotein B-100' |
| 5702 | 'APOB_No527' | 'QTVNLQLQPYSLVTTLNSDLKYNALDLTNNGK' | 'Apolipoprotein B-100' |
| 5703 | 'APOB_No528' | 'TLLKAGHIAWTSSGK' | 'Apolipoprotein B-100' |
| 5704 | 'APOB_No529' | 'NIFNFKVSQEGLK' | 'Apolipoprotein B-100' |
| 5705 | 'APOB_No530' | 'AKVHELIERYEVDQQIQVLMDK' | 'Apolipoprotein B-100' |
| 5706 | 'APOB_No531' | 'NMEVSVATTTKAQIPILR' | 'Apolipoprotein B-100' |
| 5707 | 'APOB_No532' | 'LQGTSKIDDIWNLEVK' | 'Apolipoprotein B-100' |
| 5708 | 'APOB_No533' | 'LQSTTVMNPYMK' | 'Apolipoprotein B-100' |
| 5709 | 'APOB_No534' | 'YRITENDIQIALDDAK' | 'Apolipoprotein B-100' |
| 5710 | 'APOB_No535' | 'NRQTIIVVLENVQR' | 'Apolipoprotein B-100' |
| 5711 | 'APOB_No536' | 'FLKNIILPVYDK' | 'Apolipoprotein B-100' |
| 5712 | 'APOB_No537' | 'NYQLYKSVSLPSLDPASAK' | 'Apolipoprotein B-100' |
| 5713 | 'APOB_No538' | 'NSEEFAAAMSRYELK' | 'Apolipoprotein B-100' |
| 5714 | 'APOB_No539' | 'STSPPKQAEAVLKTLQELK' | 'Apolipoprotein B-100' |
| 5715 | 'APOB_No540' | 'GMTRPLSTLISSSQSCQYTLDAKR' | 'Apolipoprotein B-100' |
| 5716 | 'APOB_No541' | 'ESDEETQIK' | 'Apolipoprotein B-100' |
| 5717 | 'APOB_No542' | 'LKFIIPSPK' | 'Apolipoprotein B-100' |
| 5718 | 'APOB_No543' | 'YGMVAQVTQTLKLEDTPKINSR' | 'Apolipoprotein B-100' |
| 5719 | 'APOB_No544' | 'LVGFIDDAVKKLNELSFK' | 'Apolipoprotein B-100' |
| 5720 | 'APOB_No545' | 'NSEEFAAAMSRYELKLAIPEGK' | 'Apolipoprotein B-100' |
| 5721 | 'APOB_No546' | 'FLDSNIK' | 'Apolipoprotein B-100' |

| | A | B | C |
|---|---|---|---|
| 1 | **Peptide Name** | **Peptide Sequence** | **Peptide Description** |
| 5722 | 'APOB_No547' | 'QAEAVLKTLQELKK' | 'Apolipoprotein B-100' |
| 5723 | 'APOB_No548' | 'TSQCTLK' | 'Apolipoprotein B-100' |
| 5724 | 'APOB_No549' | 'NIILPVYDK' | 'Apolipoprotein B-100' |
| 5725 | 'APOB_No550' | 'SEILAHWSPAKLLLQMDSSATAYGSTVSK' | 'Apolipoprotein B-100' |
| 5726 | 'APOB_No551' | 'MKALVEQGFTVPEIK' | 'Apolipoprotein B-100' |
| 5727 | 'APOB_No552' | 'YKNFATSNKMDMTFSK' | 'Apolipoprotein B-100' |
| 5728 | 'APOB_No553' | 'HEQDMVNGIMLSVEK' | 'Apolipoprotein B-100' |
| 5729 | 'APOB_No554' | 'DPNTGRLNGESNLR' | 'Apolipoprotein B-100' |
| 5730 | 'APOB_No555' | 'MKALVEQGFTVPEIK' | 'Apolipoprotein B-100' |
| 5731 | 'APOB_No556' | 'HEQDMVNGIMLSVEK' | 'Apolipoprotein B-100' |
| 5732 | 'APOB_No557' | 'IDGQFRVSSFYAK' | 'Apolipoprotein B-100' |
| 5733 | 'APOB_No558' | 'SSVKLQGTSKIDDIWNLEVK' | 'Apolipoprotein B-100' |
| 5734 | 'APOB_No559' | 'QTEATMTFKYNR' | 'Apolipoprotein B-100' |
| 5735 | 'APOB_No560' | 'SFDRHFEK' | 'Apolipoprotein B-100' |
| 5736 | 'APOB_No561' | 'VKYTLNK' | 'Apolipoprotein B-100' |
| 5737 | 'APOB_No562' | 'EYSGTIASEANTYLNSKSTR' | 'Apolipoprotein B-100' |
| 5738 | 'APOB_No563' | 'GMALFGEGK' | 'Apolipoprotein B-100' |
| 5739 | 'APOB_No564' | 'LDFSSQADLRNEIKTLLK' | 'Apolipoprotein B-100' |
| 5740 | 'APOB_No565' | 'FVTQAEGAKQTEATMTFK' | 'Apolipoprotein B-100' |
| 5741 | 'APOB_No566' | 'LKSLDEHYHIR' | 'Apolipoprotein B-100' |
| 5742 | 'APOB_No567' | 'SKEVPEAR' | 'Apolipoprotein B-100' |
| 5743 | 'APOB_No568' | 'IADFELPTIIVPEQTIEIPSIK' | 'Apolipoprotein B-100' |
| 5744 | 'APOB_No569' | 'YHWEHTGLTLREVSSK' | 'Apolipoprotein B-100' |
| 5745 | 'APOB_No570' | 'MDMTFSKQNALLR' | 'Apolipoprotein B-100' |
| 5746 | 'APOB_No571' | 'FVTQAEGAKQTEATMTFK' | 'Apolipoprotein B-100' |
| 5747 | 'APOB_No572' | 'TKNSEEFAAAMSRYELK' | 'Apolipoprotein B-100' |
| 5748 | 'APOB_No573' | 'YKNFATSNK' | 'Apolipoprotein B-100' |
| 5749 | 'APOB_No574' | 'ALVDTLKFVTQAEGAKQTEATMTFK' | 'Apolipoprotein B-100' |
| 5750 | 'APOB_No575' | 'LVKEALKESQLPTVMDFR' | 'Apolipoprotein B-100' |
| 5751 | 'APOB_No576' | 'HDAHLNGKVIGTLK' | 'Apolipoprotein B-100' |
| 5752 | 'APOB_No577' | 'LQDFSDQLSDYYEKFIAESK' | 'Apolipoprotein B-100' |
| 5753 | 'APOB_No578' | 'SLDEHYHIRVNLVK' | 'Apolipoprotein B-100' |
| 5754 | 'APOB_No579' | 'NSEEFAAAMSRYELK' | 'Apolipoprotein B-100' |
| 5755 | 'APOB_No580' | 'VSALLTPAEQTGTWKLK' | 'Apolipoprotein B-100' |
| 5756 | 'APOB_No581' | 'NMEVSVATTTKAQIPILR' | 'Apolipoprotein B-100' |
| 5757 | 'APOB_No582' | 'GSTSHHLVSR' | 'Apolipoprotein B-100' |
| 5758 | 'APOB_No583' | 'YHMKADSVVDLLSYNVQGSGETTYDHK' | 'Apolipoprotein B-100' |
| 5759 | 'APOB_No584' | 'QELNGNTK' | 'Apolipoprotein B-100' |
| 5760 | 'APOB_No585' | 'NSEEFAAAMSRYELKLAIPEGK' | 'Apolipoprotein B-100' |
| 5761 | 'APOB_No586' | 'SEILAHWSPAKLLLQMDSSATAYGSTVSKR' | 'Apolipoprotein B-100' |
| 5762 | 'APOB_No587' | 'QNALLRSEYQADYESLR' | 'Apolipoprotein B-100' |
| 5763 | 'APOB_No588' | 'QTEATMTFKYNR' | 'Apolipoprotein B-100' |
| 5764 | 'APOB_No589' | 'YRITENDIQIALDDAKINFNEK' | 'Apolipoprotein B-100' |
| 5765 | 'APOB_No590' | 'AALTELSLGSAYQAMILGVDSKNIFNFK' | 'Apolipoprotein B-100' |

| | A | B | C |
|---|---|---|---|
| 1 | **Peptide Name** | **Peptide Sequence** | **Peptide Description** |
| 5766 | 'APOB_No591' | 'KHVAEAICK' | 'Apolipoprotein B-100' |
| 5767 | 'APOB_No592' | 'TGLKEFLKTTK' | 'Apolipoprotein B-100' |
| 5768 | 'APOB_No593' | 'VSQEGLKLSNDMMGSYAEMK' | 'Apolipoprotein B-100' |
| 5769 | 'PRNP' | 'PIIHFGSDYEDR' | 'Major prion protein' |
| 5770 | 'SOD2_No1' | 'GELLEAIKR' | 'Superoxide dismutase [Mn], mitochondrial' |
| 5771 | 'SOD2_No2' | 'AIWNVINWENVTER' | 'Superoxide dismutase [Mn], mitochondrial' |
| 5772 | 'SOD2_No3' | 'HHAAYVNNLNVTEEK' | 'Superoxide dismutase [Mn], mitochondrial' |
| 5773 | 'SOD2_No4' | 'LTAASVGVQGSGWGWLGFNK' | 'Superoxide dismutase [Mn], mitochondrial' |
| 5774 | 'SOD2_No5' | 'HHAAYVNNLNVTEEKYQEALAK' | 'Superoxide dismutase [Mn], mitochondrial' |
| 5775 | 'SOD2_No6' | 'DFGSFDKFKEK' | 'Superoxide dismutase [Mn], mitochondrial' |
| 5776 | 'LCAT_No1' | 'LEPGQQEEYYR' | 'Phosphatidylcholine-sterol acyltransferase' |
| 5777 | 'LCAT_No2' | 'SSGLVSNAPGVQIR' | 'Phosphatidylcholine-sterol acyltransferase' |
| 5778 | 'LCAT_No3' | 'TYSVEYLDSSK' | 'Phosphatidylcholine-sterol acyltransferase' |
| 5779 | 'LCAT_No4' | 'LAGYLHTLVQNLVNNGYVR' | 'Phosphatidylcholine-sterol acyltransferase' |
| 5780 | 'LCAT_No5' | 'PVILVPGCLGNQLEAK' | 'Phosphatidylcholine-sterol acyltransferase' |
| 5781 | 'LCAT_No6' | 'ITTTSPWMFPSR' | 'Phosphatidylcholine-sterol acyltransferase' |
| 5782 | 'LCAT_No7' | 'LAGYLHTLVQNLVNNGYVRDETVR' | 'Phosphatidylcholine-sterol acyltransferase' |
| 5783 | 'LCAT_No8' | 'LEPGQQEEYYRK' | 'Phosphatidylcholine-sterol acyltransferase' |
| 5784 | 'LCAT_No9' | 'ITTTSPWMFPSR' | 'Phosphatidylcholine-sterol acyltransferase' |
| 5785 | 'LCAT_No10' | 'PMLVLASGDNQGIPIMSSIK' | 'Phosphatidylcholine-sterol acyltransferase' |
| 5786 | 'LCAT_No11' | 'LDKPDVVNWMCYR' | 'Phosphatidylcholine-sterol acyltransferase' |
| 5787 | 'LCAT_No12' | 'TYIYDHGFPYTDPVGVLYEDGDDTVATR' | 'Phosphatidylcholine-sterol acyltransferase' |
| 5788 | 'LCAT_No13' | 'STELCGLWQGR' | 'Phosphatidylcholine-sterol acyltransferase' |
| 5789 | 'LCAT_No14' | 'PMLVLASGDNQGIPIMSSIK' | 'Phosphatidylcholine-sterol acyltransferase' |
| 5790 | 'LCAT_No15' | 'LAGLVEEMHAAYGK' | 'Phosphatidylcholine-sterol acyltransferase' |
| 5791 | 'LCAT_No16' | 'LDKPDVVNWMCYR' | 'Phosphatidylcholine-sterol acyltransferase' |
| 5792 | 'LCAT_No17' | 'KLAGLVEEMHAAYGK' | 'Phosphatidylcholine-sterol acyltransferase' |
| 5793 | 'HRG_No1' | 'ADLFYDVEALDLESPK' | 'Histidine-rich glycoprotein' |
| 5794 | 'HRG_No2' | 'KGEVLPLPEANFPSFPLPHHK' | 'Histidine-rich glycoprotein' |
| 5795 | 'HRG_No3' | 'YKEENDDFASFR' | 'Histidine-rich glycoprotein' |
| 5796 | 'HRG_No4' | 'KYWNDCEPPDSR' | 'Histidine-rich glycoprotein' |
| 5797 | 'HRG_No5' | 'RPSEIVIGQCK' | 'Histidine-rich glycoprotein' |
| 5798 | 'HRG_No6' | 'ALDLINKR' | 'Histidine-rich glycoprotein' |
| 5799 | 'HRG_No7' | 'YWNDCEPPDSR' | 'Histidine-rich glycoprotein' |
| 5800 | 'HRG_No8' | 'RDGYLFQLLR' | 'Histidine-rich glycoprotein' |
| 5801 | 'HRG_No9' | 'SGFPQVSMFFTHTFPK' | 'Histidine-rich glycoprotein' |
| 5802 | 'HRG_No10' | 'GEVLPLPEANFPSFPLPHHK' | 'Histidine-rich glycoprotein' |
| 5803 | 'HRG_No11' | 'DSPVLIDFFEDTERYR' | 'Histidine-rich glycoprotein' |
| 5804 | 'HRG_No12' | 'SGFPQVSMFFTHTFPK' | 'Histidine-rich glycoprotein' |
| 5805 | 'HRG_No13' | 'YKEENDDFASFRVDR' | 'Histidine-rich glycoprotein' |
| 5806 | 'HRG_No14' | 'VIDFNCTTSSVSSALANTK' | 'Histidine-rich glycoprotein' |
| 5807 | 'HRG_No15' | 'DGYLFQLLR' | 'Histidine-rich glycoprotein' |
| 5808 | 'HRG_No16' | 'QIGSVYR' | 'Histidine-rich glycoprotein' |
| 5809 | 'HRG_No17' | 'VRGGEGTGYFVDFSVR' | 'Histidine-rich glycoprotein' |

| | A | B | C |
|---|---|---|---|
| 1 | **Peptide Name** | **Peptide Sequence** | **Peptide Description** |
| 5810 | 'HRG_No18' | 'ALEKYKEENDDFASFR' | 'Histidine-rich glycoprotein' |
| 5811 | 'HRG_No19' | 'HPLKPDNQPFPQSVSESCPGK' | 'Histidine-rich glycoprotein' |
| 5812 | 'HRG_No20' | 'GGEGTGYFVDFSVR' | 'Histidine-rich glycoprotein' |
| 5813 | 'HRG_No21' | 'RRDGYLFQLLR' | 'Histidine-rich glycoprotein' |
| 5814 | 'HRG_No22' | 'DSPVLIDFFEDTER' | 'Histidine-rich glycoprotein' |
| 5815 | 'HRG_No23' | 'NLVINCEVFDPQEHENINGVPPHLGHPFHWGGHER' | 'Histidine-rich glycoprotein' |
| 5816 | 'HRG_No24' | 'HPNVFGFCR' | 'Histidine-rich glycoprotein' |
| 5817 | 'HRG_No25' | 'EENDDFASFR' | 'Histidine-rich glycoprotein' |
| 5818 | 'HRG_No26' | 'DHHHPHKPHEHGPPPPPDER' | 'Histidine-rich glycoprotein' |
| 5819 | 'HRG_No27' | 'QIGSVYRLPPLR' | 'Histidine-rich glycoprotein' |
| 5820 | 'HRG_No28' | 'DSPVLIDFFEDTERYRK' | 'Histidine-rich glycoprotein' |
| 5821 | 'HRG_No29' | 'IADAHLDR' | 'Histidine-rich glycoprotein' |
| 5822 | 'HRG_No30' | 'QIGSVYRLPPLR' | 'Histidine-rich glycoprotein' |
| 5823 | 'HRG_No31' | 'FKSGFPQVSMFFTHTFPK' | 'Histidine-rich glycoprotein' |
| 5824 | 'HRG_No32' | 'EENDDFASFRVDRIER' | 'Histidine-rich glycoprotein' |
| 5825 | 'HRG_No33' | 'EENDDFASFRVDR' | 'Histidine-rich glycoprotein' |
| 5826 | 'HRG_No34' | 'FKSGFPQVSMFFTHTFPK' | 'Histidine-rich glycoprotein' |
| 5827 | 'HRG_No35' | 'HSHESQDLR' | 'Histidine-rich glycoprotein' |
| 5828 | 'IGLV7-43_No1' | 'FSGSLLGGK' | 'Immunoglobulin lambda variable 7-43' |
| 5829 | 'IGLV7-43_No2' | 'HSWTPAR' | 'Immunoglobulin lambda variable 7-43' |
| 5830 | 'IGLV7-43_No3' | 'ALIYSTSNKHSWTPAR' | 'Immunoglobulin lambda variable 7-43' |
| 5831 | 'THY1' | 'HVLFGTVGVPEHTYR' | 'Thy-1 membrane glycoprotein' |
| 5832 | 'A1BG_No1' | 'VTLTCVAPLSGVDFQLR' | 'Alpha-1B-glycoprotein' |
| 5833 | 'A1BG_No2' | 'IFFHLNAVALGDGGHYTCR' | 'Alpha-1B-glycoprotein' |
| 5834 | 'A1BG_No3' | 'GVTFLLR' | 'Alpha-1B-glycoprotein' |
| 5835 | 'A1BG_No4' | 'SGLSTGWTQLSK' | 'Alpha-1B-glycoprotein' |
| 5836 | 'A1BG_No5' | 'LELHVDGPPPRPQLR' | 'Alpha-1B-glycoprotein' |
| 5837 | 'A1BG_No6' | 'HQFLLTGDTQGR' | 'Alpha-1B-glycoprotein' |
| 5838 | 'A1BG_No7' | 'NGVAQEPVHLDSPAIK' | 'Alpha-1B-glycoprotein' |
| 5839 | 'A1BG_No8' | 'CEGPIPDVTFELLR' | 'Alpha-1B-glycoprotein' |
| 5840 | 'A1BG_No9' | 'TDGEGALSEPSATVTIEELAAPPPPVLMHHGESSQVLHPGNK' | 'Alpha-1B-glycoprotein' |
| 5841 | 'A1BG_No10' | 'SLPAPWLSMAPVSWITPGLK' | 'Alpha-1B-glycoprotein' |
| 5842 | 'A1BG_No11' | 'TPGAAANLELIFVGPQHAGNYR' | 'Alpha-1B-glycoprotein' |
| 5843 | 'A1BG_No12' | 'CLAPLEGAR' | 'Alpha-1B-glycoprotein' |
| 5844 | 'A1BG_No13' | 'VTLTCVAPLSGVDFQLRR' | 'Alpha-1B-glycoprotein' |
| 5845 | 'A1BG_No14' | 'FALVREDR' | 'Alpha-1B-glycoprotein' |
| 5846 | 'A1BG_No15' | 'SSTSPDRIFFHLNAVALGDGGHYTCR' | 'Alpha-1B-glycoprotein' |
| 5847 | 'A1BG_No16' | 'GVTFLLRR' | 'Alpha-1B-glycoprotein' |
| 5848 | 'A1BG_No17' | 'TDGEGALSEPSATVTIEELAAPPPPVLMHHGESSQVLHPGNK' | 'Alpha-1B-glycoprotein' |
| 5849 | 'A1BG_No18' | 'SLPAPWLSMAPVSWITPGLK' | 'Alpha-1B-glycoprotein' |
| 5850 | 'A1BG_No19' | 'NGVAQEPVHLDSPAIKHQFLLTGDTQGR' | 'Alpha-1B-glycoprotein' |
| 5851 | 'A1BG_No20' | 'CEGPIPDVTFELLREGETK' | 'Alpha-1B-glycoprotein' |
| 5852 | 'A1BG_No21' | 'YRSWVPHTFESELSDPVELLVAES' | 'Alpha-1B-glycoprotein' |
| 5853 | 'A1BG_No22' | 'ATWSGAVLAGR' | 'Alpha-1B-glycoprotein' |

| | A | B | C |
|---|---|---|---|
| 1 | **Peptide Name** | **Peptide Sequence** | **Peptide Description** |
| 5854 | 'A1BG_No23' | 'LLELTGPK' | 'Alpha-1B-glycoprotein' |
| 5855 | 'A1BG_No24' | 'DAVLRCEGPIPDVTFELLR' | 'Alpha-1B-glycoprotein' |
| 5856 | 'A1BG_No25' | 'RGEKELLVPR' | 'Alpha-1B-glycoprotein' |
| 5857 | 'A1BG_No26' | 'DAVLRCEGPIPDVTFELLREGETK' | 'Alpha-1B-glycoprotein' |
| 5858 | 'A1BG_No27' | 'GEKELLVPR' | 'Alpha-1B-glycoprotein' |
| 5859 | 'A1BG_No28' | 'ATWSGAVLAGRDAVLR' | 'Alpha-1B-glycoprotein' |
| 5860 | 'A1BG_No29' | 'TVRTPGAAANLELIFVGPQHAGNYR' | 'Alpha-1B-glycoprotein' |
| 5861 | 'A1BG_No30' | 'LRCLAPLEGAR' | 'Alpha-1B-glycoprotein' |
| 5862 | 'A1BG_No31' | 'LHDNQNGWSGDSAPVELILSDETLPAPEFSPEPESGR' | 'Alpha-1B-glycoprotein' |
| 5863 | 'A1BG_No32' | 'LELHVDGPPPR' | 'Alpha-1B-glycoprotein' |
| 5864 | 'A1BG_No33' | 'GVLRGVTFLLR' | 'Alpha-1B-glycoprotein' |
| 5865 | 'A1BG_No34' | 'HQFLLTGDTQGRYR' | 'Alpha-1B-glycoprotein' |
| 5866 | 'A1BG_No35' | 'CRSGLSTGWTQLSK' | 'Alpha-1B-glycoprotein' |
| 5867 | 'A1BG_No36' | 'SGLSTGWTQLSKLLELTGPK' | 'Alpha-1B-glycoprotein' |
| 5868 | 'A1BG_No37' | 'CEGPIPDVTFELLREGETKAVK' | 'Alpha-1B-glycoprotein' |
| 5869 | 'KRT6B_No1' | 'YEELQITAGR' | 'Keratin, type II cytoskeletal 6B' |
| 5870 | 'KRT6B_No2' | 'ATGGGLSSVGGGSSTIK' | 'Keratin, type II cytoskeletal 6B' |
| 5871 | 'KRT6B_No3' | 'SGFSSISVSR' | 'Keratin, type II cytoskeletal 6B' |
| 5872 | 'KRT6B_No4' | 'QLDNIVGER' | 'Keratin, type II cytoskeletal 6B' |
| 5873 | 'KRT6B_No5' | 'VLDTKWTLLQEQGTK' | 'Keratin, type II cytoskeletal 6B' |
| 5874 | 'KRT6B_No6' | 'FLEQQNKVLDTK' | 'Keratin, type II cytoskeletal 6B' |
| 5875 | 'KRT1_No1' | 'THNLEPYFESFINNLR' | 'Keratin, type II cytoskeletal 1' |
| 5876 | 'KRT1_No2' | 'SLDLDSIIAEVK' | 'Keratin, type II cytoskeletal 1' |
| 5877 | 'KRT1_No3' | 'WELLQQVDTSTR' | 'Keratin, type II cytoskeletal 1' |
| 5878 | 'KRT1_No4' | 'SLNNQFASFIDK' | 'Keratin, type II cytoskeletal 1' |
| 5879 | 'KRT1_No5' | 'TNAENEFVTIKK' | 'Keratin, type II cytoskeletal 1' |
| 5880 | 'KRT1_No6' | 'SLNNQFASFIDKVR' | 'Keratin, type II cytoskeletal 1' |
| 5881 | 'KRT1_No7' | 'FSSCGGGGGSFGAGGGFGSR' | 'Keratin, type II cytoskeletal 1' |
| 5882 | 'KRT1_No8' | 'QISNLQQSISDAEQR' | 'Keratin, type II cytoskeletal 1' |
| 5883 | 'KRT1_No9' | 'SGGGFSSGSAGIINYQR' | 'Keratin, type II cytoskeletal 1' |
| 5884 | 'KRT1_No10' | 'MSGECAPNVSVSVSTSHTTISGGGSR' | 'Keratin, type II cytoskeletal 1' |
| 5885 | 'KRT1_No11' | 'TNAENEFVTIK' | 'Keratin, type II cytoskeletal 1' |
| 5886 | 'KRT1_No12' | 'NMQDMVEDYR' | 'Keratin, type II cytoskeletal 1' |
| 5887 | 'KRT1_No13' | 'MSGECAPNVSVSVSTSHTTISGGGSR' | 'Keratin, type II cytoskeletal 1' |
| 5888 | 'KRT1_No14' | 'LLRDYQELMNTK' | 'Keratin, type II cytoskeletal 1' |
| 5889 | 'KRT1_No15' | 'QISNLQQSISDAEQRGENALKDAK' | 'Keratin, type II cytoskeletal 1' |
| 5890 | 'KRT1_No16' | 'NMQDMVEDYR' | 'Keratin, type II cytoskeletal 1' |
| 5891 | 'KRT1_No17' | 'THNLEPYFESFINNLRR' | 'Keratin, type II cytoskeletal 1' |
| 5892 | 'KRT1_No18' | 'QISNLQQSISDAEQRGENALKDAK' | 'Keratin, type II cytoskeletal 1' |
| 5893 | 'KRT1_No19' | 'GGGGGGYGSGGSSYGSGGGSYGSGGGGGGGR' | 'Keratin, type II cytoskeletal 1' |
| 5894 | 'KRT1_No20' | 'LNDLEDALQQAK' | 'Keratin, type II cytoskeletal 1' |
| 5895 | 'KRT1_No21' | 'DYQELMNTK' | 'Keratin, type II cytoskeletal 1' |
| 5896 | 'KRT1_No22' | 'TLLEGEESR' | 'Keratin, type II cytoskeletal 1' |
| 5897 | 'KRT1_No23' | 'SKAEAESLYQSKYEELQITAGR' | 'Keratin, type II cytoskeletal 1' |

| | A | B | C |
|---|---|---|---|
| 1 | **Peptide Name** | **Peptide Sequence** | **Peptide Description** |
| 5898 | 'KRT1_No24' | 'NKLNDLEDALQQAKEDLAR' | 'Keratin, type II cytoskeletal 1' |
| 5899 | 'KRT1_No25' | 'SKAEAESLYQSK' | 'Keratin, type II cytoskeletal 1' |
| 5900 | 'KRT1_No26' | 'LRSEIDNVKK' | 'Keratin, type II cytoskeletal 1' |
| 5901 | 'KRT1_No27' | 'QISNLQQSISDAEQR' | 'Keratin, type II cytoskeletal 1' |
| 5902 | 'KRT1_No28' | 'SISISVAR' | 'Keratin, type II cytoskeletal 1' |
| 5903 | 'KRT1_No29' | 'FLEQQNQVLQTKWELLQQVDTSTR' | 'Keratin, type II cytoskeletal 1' |
| 5904 | 'KRT1_No30' | 'SLVNLGGSK' | 'Keratin, type II cytoskeletal 1' |
| 5905 | 'KRT1_No31' | 'LNDLEDALQQAKEDLAR' | 'Keratin, type II cytoskeletal 1' |
| 5906 | 'KRT1_No32' | 'LDSELKNMQDMVEDYR' | 'Keratin, type II cytoskeletal 1' |
| 5907 | 'KRT1_No33' | 'KQISNLQQSISDAEQR' | 'Keratin, type II cytoskeletal 1' |
| 5908 | 'KRT1_No34' | 'LDSELKNMQDMVEDYR' | 'Keratin, type II cytoskeletal 1' |
| 5909 | 'KRT1_No35' | 'LDSELKNMQDMVEDYR' | 'Keratin, type II cytoskeletal 1' |
| 5910 | 'KRT1_No36' | 'NMQDMVEDYR' | 'Keratin, type II cytoskeletal 1' |
| 5911 | 'KRT1_No37' | 'NKLNDLEDALQQAK' | 'Keratin, type II cytoskeletal 1' |
| 5912 | 'KRT1_No38' | 'AEAESLYQSK' | 'Keratin, type II cytoskeletal 1' |
| 5913 | 'KRT1_No39' | 'RTNAENEFVTIKK' | 'Keratin, type II cytoskeletal 1' |
| 5914 | 'KRT1_No40' | 'DVDGAYMTK' | 'Keratin, type II cytoskeletal 1' |
| 5915 | 'KRT1_No41' | 'EQIKSLNNQFASFIDKVR' | 'Keratin, type II cytoskeletal 1' |
| 5916 | 'KRT1_No42' | 'DYQELMNTK' | 'Keratin, type II cytoskeletal 1' |
| 5917 | 'KRT1_No43' | 'NSKIEISELNR' | 'Keratin, type II cytoskeletal 1' |
| 5918 | 'KRT1_No44' | 'AEAESLYQSKYEELQITAGR' | 'Keratin, type II cytoskeletal 1' |
| 5919 | 'KRT1_No45' | 'NSKIEISELNRVIQR' | 'Keratin, type II cytoskeletal 1' |
| 5920 | 'KRT1_No46' | 'LDSELKNMQDMVEDYR' | 'Keratin, type II cytoskeletal 1' |
| 5921 | 'KRT1_No47' | 'DVDGAYMTK' | 'Keratin, type II cytoskeletal 1' |
| 5922 | 'KRT1_No48' | 'TNAENEFVTIKKDVDGAYMTK' | 'Keratin, type II cytoskeletal 1' |
| 5923 | 'KRT1_No49' | 'QISNLQQSISDAEQRGENALK' | 'Keratin, type II cytoskeletal 1' |
| 5924 | 'KRT1_No50' | 'QISNLQQSISDAEQRGENALK' | 'Keratin, type II cytoskeletal 1' |
| 5925 | 'KRT1_No51' | 'SDQSRLDSELK' | 'Keratin, type II cytoskeletal 1' |
| 5926 | 'KRT1_No52' | 'LLRDYQELMNTK' | 'Keratin, type II cytoskeletal 1' |
| 5927 | 'KRT1_No53' | 'TNAENEFVTIKKDVDGAYMTK' | 'Keratin, type II cytoskeletal 1' |
| 5928 | 'KRT1_No54' | 'DVDGAYMTKVDLQAK' | 'Keratin, type II cytoskeletal 1' |
| 5929 | 'KRT1_No55' | 'GSGGGSSGGSIGGR' | 'Keratin, type II cytoskeletal 1' |
| 5930 | 'KRT1_No56' | 'AQYEDIAQKSKAEAESLYQSK' | 'Keratin, type II cytoskeletal 1' |
| 5931 | 'KRT1_No57' | 'SDQSRLDSELKNMQDMVEDYR' | 'Keratin, type II cytoskeletal 1' |
| 5932 | 'KRT1_No58' | 'SGGGFSSGSAGIINYQRR' | 'Keratin, type II cytoskeletal 1' |
| 5933 | 'KRT1_No59' | 'DVDGAYMTKVDLQAK' | 'Keratin, type II cytoskeletal 1' |
| 5934 | 'KRT1_No60' | 'GSYGSGGSSYGSGGGSYGSGGGGGGHGSYGSGSSSGGYR' | 'Keratin, type II cytoskeletal 1' |
| 5935 | 'KRT1_No61' | 'SEIDNVKK' | 'Keratin, type II cytoskeletal 1' |
| 5936 | 'VWF_No1' | 'RLPGDIQVVPIGVGPNANVQELER' | 'von Willebrand factor' |
| 5937 | 'VWF_No2' | 'YTLFQIFSK' | 'von Willebrand factor' |
| 5938 | 'VWF_No3' | 'NSQWICSNEECPGECLVTGQSHFK' | 'von Willebrand factor' |
| 5939 | 'VWF_No4' | 'WTCPCVCTGSSTR' | 'von Willebrand factor' |
| 5940 | 'VWF_No5' | 'YAGSQVASTSEVLK' | 'von Willebrand factor' |
| 5941 | 'VWF_No6' | 'VLAPATFYAICQQDSCHQEQVCEVIASYAHLCR' | 'von Willebrand factor' |

| | A | B | C |
|---|---|---|---|
| 1 | **Peptide Name** | **Peptide Sequence** | **Peptide Description** |
| 5942 | 'VWF_No7' | 'LSEAEFEVLK' | 'von Willebrand factor' |
| 5943 | 'VWF_No8' | 'IEDLPTMVTLGNSFLHK' | 'von Willebrand factor' |
| 5944 | 'VWF_No9' | 'TYGLCGICDENGANDFMLR' | 'von Willebrand factor' |
| 5945 | 'VWF_No10' | 'ILTSDVFQDCNK' | 'von Willebrand factor' |
| 5946 | 'VWF_No11' | 'GLRPSCPNSQSPVK' | 'von Willebrand factor' |
| 5947 | 'VWF_No12' | 'HIVTFDGQNFK' | 'von Willebrand factor' |
| 5948 | 'VWF_No13' | 'AHLLSLVDVMQR' | 'von Willebrand factor' |
| 5949 | 'VWF_No14' | 'VKEEVFIQQR' | 'von Willebrand factor' |
| 5950 | 'VWF_No15' | 'TATLCPQSCEER' | 'von Willebrand factor' |
| 5951 | 'VWF_No16' | 'RPMKDETHFEVVESGR' | 'von Willebrand factor' |
| 5952 | 'VWF_No17' | 'YLTSEMHGARPGASK' | 'von Willebrand factor' |
| 5953 | 'VWF_No18' | 'FNHLGHIFTFTPQNNEFQLQLSPK' | 'von Willebrand factor' |
| 5954 | 'VWF_No19' | 'LVSVPYVGGNMEVNVYGAIMHEVR' | 'von Willebrand factor' |
| 5955 | 'VWF_No20' | 'VTVFPIGIGDR' | 'von Willebrand factor' |
| 5956 | 'VWF_No21' | 'TPDFCAMSCPPSLVYNHCEHGCPR' | 'von Willebrand factor' |
| 5957 | 'VWF_No22' | 'EGGPSQIGDALGFAVR' | 'von Willebrand factor' |
| 5958 | 'VWF_No23' | 'TCQSLHINEMCQER' | 'von Willebrand factor' |
| 5959 | 'VWF_No24' | 'YIILLLGK' | 'von Willebrand factor' |
| 5960 | 'VWF_No25' | 'VTVFPIGIGDRYDAAQLR' | 'von Willebrand factor' |
| 5961 | 'VWF_No26' | 'VAVVEYHDGSHAYIGLK' | 'von Willebrand factor' |
| 5962 | 'VWF_No27' | 'CLPSACEVVTGSPR' | 'von Willebrand factor' |
| 5963 | 'VWF_No28' | 'YDVCSCSDGR' | 'von Willebrand factor' |
| 5964 | 'VWF_No29' | 'KVPLDSSPATCHNNIMK' | 'von Willebrand factor' |
| 5965 | 'VWF_No30' | 'SLSCRPPMVK' | 'von Willebrand factor' |
| 5966 | 'VWF_No31' | 'FSEEACAVLTSPTFEACHR' | 'von Willebrand factor' |
| 5967 | 'VWF_No32' | 'DETLQDGCDTHFCK' | 'von Willebrand factor' |
| 5968 | 'VWF_No33' | 'LPGLHNSLVK' | 'von Willebrand factor' |
| 5969 | 'VWF_No34' | 'KVPLDSSPATCHNNIMK' | 'von Willebrand factor' |
| 5970 | 'VWF_No35' | 'HLSISVVLK' | 'von Willebrand factor' |
| 5971 | 'VWF_No36' | 'TTCNPCPLGYKEENNTGECCGR' | 'von Willebrand factor' |
| 5972 | 'VWF_No37' | 'SVGSQWASPENPCLINECVR' | 'von Willebrand factor' |
| 5973 | 'VWF_No38' | 'EQDLEVILHNGACSPGAR' | 'von Willebrand factor' |
| 5974 | 'VWF_No39' | 'RPMKDETHFEVVESGR' | 'von Willebrand factor' |
| 5975 | 'VWF_No40' | 'TNGVCVDWR' | 'von Willebrand factor' |
| 5976 | 'VWF_No41' | 'RDETLQDGCDTHFCK' | 'von Willebrand factor' |
| 5977 | 'VWF_No42' | 'GLWEQCQLLK' | 'von Willebrand factor' |
| 5978 | 'VWF_No43' | 'YLSDHSFLVSQGDREQAPNLVYMVTGNPASDEIKR' | 'von Willebrand factor' |
| 5979 | 'VWF_No44' | 'KTTCNPCPLGYKEENNTGECCGR' | 'von Willebrand factor' |
| 5980 | 'VWF_No45' | 'GQVYLQCGTPCNLTCR' | 'von Willebrand factor' |
| 5981 | 'VWF_No46' | 'IGWPNAPILIQDFETLPR' | 'von Willebrand factor' |
| 5982 | 'VWF_No47' | 'VAVVEYHDGSHAYIGLKDR' | 'von Willebrand factor' |
| 5983 | 'VWF_No48' | 'AVVILVTDVSVDSVDAAADAAR' | 'von Willebrand factor' |
| 5984 | 'VWF_No49' | 'GLRPSCPNSQSPVKVEETCGCR' | 'von Willebrand factor' |
| 5985 | 'VWF_No50' | 'ITLLLMASQEPQR' | 'von Willebrand factor' |

| | A | B | C |
|---|---|---|---|
| 1 | **Peptide Name** | **Peptide Sequence** | **Peptide Description** |
| 5986 | 'VWF_No51' | 'TYGLCGICDENGANDFMLR' | 'von Willebrand factor' |
| 5987 | 'VWF_No52' | 'LLDLVFLLDGSSR' | 'von Willebrand factor' |
| 5988 | 'VWF_No53' | 'ILDELLQTCVDPEDCPVCEVAGRR' | 'von Willebrand factor' |
| 5989 | 'VWF_No54' | 'SGFTYVLHEGECCGR' | 'von Willebrand factor' |
| 5990 | 'VWF_No55' | 'AHLLSLVDVMQR' | 'von Willebrand factor' |
| 5991 | 'VWF_No56' | 'IEDLPTMVTLGNSFLHK' | 'von Willebrand factor' |
| 5992 | 'VWF_No57' | 'IPGTCCDTCEEPECNDITAR' | 'von Willebrand factor' |
| 5993 | 'VWF_No58' | 'EAPDLVLQR' | 'von Willebrand factor' |
| 5994 | 'VWF_No59' | 'YLFPGECQYVLVQDYCGSNPGTFR' | 'von Willebrand factor' |
| 5995 | 'VWF_No60' | 'SEVEVDIHYCQGK' | 'von Willebrand factor' |
| 5996 | 'VWF_No61' | 'SKEFMEEVIQR' | 'von Willebrand factor' |
| 5997 | 'VWF_No62' | 'DCQDHSFSIVIETVQCADDRDAVCTR' | 'von Willebrand factor' |
| 5998 | 'VWF_No63' | 'CLPTACTIQLR' | 'von Willebrand factor' |
| 5999 | 'VWF_No64' | 'ILDELLQTCVDPEDCPVCEVAGR' | 'von Willebrand factor' |
| 6000 | 'VWF_No65' | 'ECLCGALASYAAACAGR' | 'von Willebrand factor' |
| 6001 | 'VWF_No66' | 'SFSIIGDFQNGK' | 'von Willebrand factor' |
| 6002 | 'VWF_No67' | 'ILAGPAGDSNVVK' | 'von Willebrand factor' |
| 6003 | 'VWF_No68' | 'YLSDHSFLVSQGDREQAPNLVYMVTGNPASDEIK' | 'von Willebrand factor' |
| 6004 | 'VWF_No69' | 'LTGSCSYVLFQNK' | 'von Willebrand factor' |
| 6005 | 'VWF_No70' | 'VNERGEYFWEK' | 'von Willebrand factor' |
| 6006 | 'VWF_No71' | 'ALSVVWDR' | 'von Willebrand factor' |
| 6007 | 'VWF_No72' | 'VAWREPGRCELNCPK' | 'von Willebrand factor' |
| 6008 | 'VWF_No73' | 'IGCNTCVCQDR' | 'von Willebrand factor' |
| 6009 | 'VWF_No74' | 'YLTSEMHGARPGASK' | 'von Willebrand factor' |
| 6010 | 'VWF_No75' | 'RDETLQDGCDTHFCKVNER' | 'von Willebrand factor' |
| 6011 | 'VWF_No76' | 'VNERGEYFWEKR' | 'von Willebrand factor' |
| 6012 | 'VWF_No77' | 'VPLDSSPATCHNNIMK' | 'von Willebrand factor' |
| 6013 | 'VWF_No78' | 'GEYFWEK' | 'von Willebrand factor' |
| 6014 | 'VWF_No79' | 'GLQPTLTNPGECRPNFTCACR' | 'von Willebrand factor' |
| 6015 | 'VWF_No80' | 'TCQSLHINEMCQER' | 'von Willebrand factor' |
| 6016 | 'VWF_No81' | 'CMVQVGVISGFK' | 'von Willebrand factor' |
| 6017 | 'VWF_No82' | 'DETHFEVVESGR' | 'von Willebrand factor' |
| 6018 | 'VWF_No83' | 'LSYGEDLQMDWDGR' | 'von Willebrand factor' |
| 6019 | 'VWF_No84' | 'LHGDCQDLQKQHSDPCALNPR' | 'von Willebrand factor' |
| 6020 | 'VWF_No85' | 'HGAGVAMDGQDVQLPLLK' | 'von Willebrand factor' |
| 6021 | 'VWF_No86' | 'AFVVDMMER' | 'von Willebrand factor' |
| 6022 | 'VWF_No87' | 'LVCPADNLR' | 'von Willebrand factor' |
| 6023 | 'VWF_No88' | 'ENGYECEWR' | 'von Willebrand factor' |
| 6024 | 'VWF_No89' | 'LQRIEDLPTMVTLGNSFLHK' | 'von Willebrand factor' |
| 6025 | 'VWF_No90' | 'IMKIPGTCCDTCEEPECNDITAR' | 'von Willebrand factor' |
| 6026 | 'VWF_No91' | 'SKEFMEEVIQR' | 'von Willebrand factor' |
| 6027 | 'VWF_No92' | 'VEDFGNAWK' | 'von Willebrand factor' |
| 6028 | 'VWF_No93' | 'LVCPADNLRAEGLECTK' | 'von Willebrand factor' |
| 6029 | 'VWF_No94' | 'IMKIPGTCCDTCEEPECNDITAR' | 'von Willebrand factor' |

| | A | B | C |
|---|---|---|---|
| 1 | **Peptide Name** | **Peptide Sequence** | **Peptide Description** |
| 6030 | 'VWF_No95' | 'YLSDHSFLVSQGDR' | 'von Willebrand factor' |
| 6031 | 'VWF_No96' | 'CPCFHQGKEYAPGETVK' | 'von Willebrand factor' |
| 6032 | 'VWF_No97' | 'AMYSIDINDVQDQCSCCSPTR' | 'von Willebrand factor' |
| 6033 | 'VWF_No98' | 'RPGDVWTLPDQCHTVTCQPDGQTLLK' | 'von Willebrand factor' |
| 6034 | 'VWF_No99' | 'SFSIIGDFQNGKR' | 'von Willebrand factor' |
| 6035 | 'VWF_No100' | 'LPGDIQVVPIGVGPNANVQELER' | 'von Willebrand factor' |
| 6036 | 'VWF_No101' | 'TTCNPCPLGYK' | 'von Willebrand factor' |
| 6037 | 'VWF_No102' | 'RVTGCPPFDEHKCLAEGGK' | 'von Willebrand factor' |
| 6038 | 'VWF_No103' | 'HSALSVELHSDMEVTVNGR' | 'von Willebrand factor' |
| 6039 | 'VWF_No104' | 'CVDGCSCPEGQLLDEGLCVESTECPCVHSGKR' | 'von Willebrand factor' |
| 6040 | 'VWF_No105' | 'STIYPVGQFWEEGCDVCTCTDMEDAVMGLR' | 'von Willebrand factor' |
| 6041 | 'VWF_No106' | 'EQAPNLVYMVTGNPASDEIK' | 'von Willebrand factor' |
| 6042 | 'VWF_No107' | 'VTGCPPFDEHKCLAEGGK' | 'von Willebrand factor' |
| 6043 | 'VWF_No108' | 'TCQNYDLECMSMGCVSGCLCPPGMVR' | 'von Willebrand factor' |
| 6044 | 'VWF_No109' | 'GLYLETEAGYYK' | 'von Willebrand factor' |
| 6045 | 'VWF_No110' | 'TLCECAGGLECACPALLEYAR' | 'von Willebrand factor' |
| 6046 | 'VWF_No111' | 'EYAPGETVK' | 'von Willebrand factor' |
| 6047 | 'VWF_No112' | 'EFMEEVIQR' | 'von Willebrand factor' |
| 6048 | 'VWF_No113' | 'TCAQEGMVLYGWTDHSACSPVCPAGMEYR' | 'von Willebrand factor' |
| 6049 | 'VWF_No114' | 'AFVLSSVDELEQQR' | 'von Willebrand factor' |
| 6050 | 'VWF_No115' | 'IQHTVTASVR' | 'von Willebrand factor' |
| 6051 | 'VWF_No116' | 'GEYFWEKR' | 'von Willebrand factor' |
| 6052 | 'VWF_No117' | 'AFVVDMMER' | 'von Willebrand factor' |
| 6053 | 'VWF_No118' | 'YDVCSCSDGRECLCGALASYAAACAGR' | 'von Willebrand factor' |
| 6054 | 'VWF_No119' | 'CMVQVGVISGFK' | 'von Willebrand factor' |
| 6055 | 'VWF_No120' | 'EQAPNLVYMVTGNPASDEIKR' | 'von Willebrand factor' |
| 6056 | 'VWF_No121' | 'VGSCKSEVEVDIHYCQGK' | 'von Willebrand factor' |
| 6057 | 'VWF_No122' | 'KTTCNPCPLGYK' | 'von Willebrand factor' |
| 6058 | 'VWF_No123' | 'AMYSIDINDVQDQCSCCSPTR' | 'von Willebrand factor' |
| 6059 | 'VWF_No124' | 'HCDGNVSSCGDHPSEGCFCPPDK' | 'von Willebrand factor' |
| 6060 | 'VWF_No125' | 'YFTFSGICQYLLAR' | 'von Willebrand factor' |
| 6061 | 'VWF_No126' | 'LSGEAYGFVAR' | 'von Willebrand factor' |
| 6062 | 'VWF_No127' | 'CHPLVDPEPFVALCEK' | 'von Willebrand factor' |
| 6063 | 'VWF_No128' | 'LSYGEDLQMDWDGR' | 'von Willebrand factor' |
| 6064 | 'VWF_No129' | 'VPLDSSPATCHNNIMK' | 'von Willebrand factor' |
| 6065 | 'VWF_No130' | 'VIVIPVGIGPHANLK' | 'von Willebrand factor' |
| 6066 | 'VWF_No131' | 'AVSPLPYLR' | 'von Willebrand factor' |
| 6067 | 'VWF_No132' | 'YLSDHSFLVSQGDREQAPNLVYMVTGNPASDEIKR' | 'von Willebrand factor' |
| 6068 | 'VWF_No133' | 'QNADQCCPEYECVCDPVSCDLPPVPHCER' | 'von Willebrand factor' |
| 6069 | 'VWF_No134' | 'CVDGCSCPEGQLLDEGLCVESTECPCVHSGK' | 'von Willebrand factor' |
| 6070 | 'SHBG_No1' | 'IALGGLLFPASNLR' | 'Sex hormone-binding globulin' |
| 6071 | 'SHBG_No2' | 'LPLVPALDGCLR' | 'Sex hormone-binding globulin' |
| 6072 | 'SHBG_No3' | 'QAEISASAPTSLR' | 'Sex hormone-binding globulin' |
| 6073 | 'SHBG_No4' | 'DIPQPHAEPWAFSLDLGLK' | 'Sex hormone-binding globulin' |

| | A | B | C |
|---|---|---|---|
| 1 | **Peptide Name** | **Peptide Sequence** | **Peptide Description** |
| 6074 | 'SHBG_No5' | 'DGRPEIQLHNHWAQLTVGAGPR' | 'Sex hormone-binding globulin' |
| 6075 | 'SHBG_No6' | 'DDWFMLGLR' | 'Sex hormone-binding globulin' |
| 6076 | 'SHBG_No7' | 'DSWLDKQAEISASAPTSLR' | 'Sex hormone-binding globulin' |
| 6077 | 'SHBG_No8' | 'ALALPPLGLAPLLNLWAK' | 'Sex hormone-binding globulin' |
| 6078 | 'SHBG_No9' | 'QAAGSGHLLALGTPENPSWLSLHLQDQK' | 'Sex hormone-binding globulin' |
| 6079 | 'SHBG_No10' | 'VVLSSGSGPGLDLPLVLGLPLQLK' | 'Sex hormone-binding globulin' |
| 6080 | 'SHBG_No11' | 'DDWFMLGLR' | 'Sex hormone-binding globulin' |
| 6081 | 'SHBG_No12' | 'QAEISASAPTSLR' | 'Sex hormone-binding globulin' |
| 6082 | 'SHBG_No13' | 'SCDVESNPGIFLPPGTQAEFNLR' | 'Sex hormone-binding globulin' |
| 6083 | 'SHBG_No14' | 'LDVDQALNR' | 'Sex hormone-binding globulin' |
| 6084 | 'SHBG_No15' | 'QAAGSGHLLALGTPENPSWLSLHLQDQK' | 'Sex hormone-binding globulin' |
| 6085 | 'SHBG_No16' | 'LPLVPALDGCLRR' | 'Sex hormone-binding globulin' |
| 6086 | 'SHBG_No17' | 'LDDGRWHQVEVK' | 'Sex hormone-binding globulin' |
| 6087 | 'SHBG_No18' | 'TWDPEGVIFYGDTNPK' | 'Sex hormone-binding globulin' |
| 6088 | 'SHBG_No19' | 'MEGDSVLLEVDGEEVLR' | 'Sex hormone-binding globulin' |
| 6089 | 'SHBG_No20' | 'RDSWLDKQAEISASAPTSLR' | 'Sex hormone-binding globulin' |
| 6090 | 'SHBG_No21' | 'MEGDSVLLEVDGEEVLR' | 'Sex hormone-binding globulin' |
| 6091 | 'SHBG_No22' | 'TSSSFEVR' | 'Sex hormone-binding globulin' |
| 6092 | 'GAPDH_No1' | 'IISNASCTTNCLAPLAK' | 'Glyceraldehyde-3-phosphate dehydrogenase' |
| 6093 | 'GAPDH_No2' | 'VPTANVSVVDLTCR' | 'Glyceraldehyde-3-phosphate dehydrogenase' |
| 6094 | 'GAPDH_No3' | 'LISWYDNEFGYSNR' | 'Glyceraldehyde-3-phosphate dehydrogenase' |
| 6095 | 'GAPDH_No4' | 'GALQNIIPASTGAAK' | 'Glyceraldehyde-3-phosphate dehydrogenase' |
| 6096 | 'GAPDH_No5' | 'VIISAPSADAPMFVMGVNHEK' | 'Glyceraldehyde-3-phosphate dehydrogenase' |
| 6097 | 'GAPDH_No6' | 'RVIISAPSADAPMFVMGVNHEK' | 'Glyceraldehyde-3-phosphate dehydrogenase' |
| 6098 | 'GAPDH_No7' | 'LVINGNPITIFQERDPSK' | 'Glyceraldehyde-3-phosphate dehydrogenase' |
| 6099 | 'GAPDH_No8' | 'WGDAGAEYVVESTGVFTTMEK' | 'Glyceraldehyde-3-phosphate dehydrogenase' |
| 6100 | 'GAPDH_No9' | 'GILGYTEHQVVSSDFNSDTHSSTFDAGAGIALNDHFVK' | 'Glyceraldehyde-3-phosphate dehydrogenase' |
| 6101 | 'GAPDH_No10' | 'VIISAPSADAPMFVMGVNHEKYDNSLK' | 'Glyceraldehyde-3-phosphate dehydrogenase' |
| 6102 | 'GAPDH_No11' | 'VVDLMAHMASKE' | 'Glyceraldehyde-3-phosphate dehydrogenase' |
| 6103 | 'GAPDH_No12' | 'VIHDNFGIVEGLMTTVHAITATQK' | 'Glyceraldehyde-3-phosphate dehydrogenase' |
| 6104 | 'GAPDH_No13' | 'VIISAPSADAPMFVMGVNHEK' | 'Glyceraldehyde-3-phosphate dehydrogenase' |
| 6105 | 'GAPDH_No14' | 'IKWGDAGAEYVVESTGVFTTMEK' | 'Glyceraldehyde-3-phosphate dehydrogenase' |
| 6106 | 'GAPDH_No15' | 'VIHDNFGIVEGLMTTVHAITATQK' | 'Glyceraldehyde-3-phosphate dehydrogenase' |
| 6107 | 'GAPDH_No16' | 'VVDLMAHMASKE' | 'Glyceraldehyde-3-phosphate dehydrogenase' |
| 6108 | 'GAPDH_No17' | 'VPTANVSVVDLTCRLEKPAK' | 'Glyceraldehyde-3-phosphate dehydrogenase' |
| 6109 | 'GAPDH_No18' | 'WGDAGAEYVVESTGVFTTMEK' | 'Glyceraldehyde-3-phosphate dehydrogenase' |
| 6110 | 'GAPDH_No19' | 'VIISAPSADAPMFVMGVNHEK' | 'Glyceraldehyde-3-phosphate dehydrogenase' |
| 6111 | 'GAPDH_No20' | 'VVDLMAHMASK' | 'Glyceraldehyde-3-phosphate dehydrogenase' |
| 6112 | 'GAPDH_No21' | 'VIISAPSADAPMFVMGVNHEKYDNSLK' | 'Glyceraldehyde-3-phosphate dehydrogenase' |
| 6113 | 'GAPDH_No22' | 'VVDLMAHMASK' | 'Glyceraldehyde-3-phosphate dehydrogenase' |
| 6114 | 'GAPDH_No23' | 'VVDLMAHMASK' | 'Glyceraldehyde-3-phosphate dehydrogenase' |
| 6115 | 'GAPDH_No24' | 'LVINGNPITIFQER' | 'Glyceraldehyde-3-phosphate dehydrogenase' |
| 6116 | 'IGKV1-16_No1' | 'ASQGISNYLAWFQQKPGK' | 'Immunoglobulin kappa variable 1-16' |
| 6117 | 'IGKV1-16_No2' | 'SLIYAASSLQSGVPSK' | 'Immunoglobulin kappa variable 1-16' |

| | A | B | C |
|---|---|---|---|
| 1 | **Peptide Name** | **Peptide Sequence** | **Peptide Description** |
| 6118 | 'IGKV1-16_No3' | 'ASQGISNYLAWFQQK' | 'Immunoglobulin kappa variable 1-16' |
| 6119 | 'IGKV1-16_No4' | 'ASQGISNYLAWFQQKPGKAPK' | 'Immunoglobulin kappa variable 1-16' |
| 6120 | 'HLA-A_No1' | 'DGEDQTQDTELVETRPAGDGTFQK' | 'HLA class I histocompatibility antigen, A alpha chain' |
| 6121 | 'HLA-A_No2' | 'WAAVVVPSGEEQR' | 'HLA class I histocompatibility antigen, A alpha chain' |
| 6122 | 'HLA-A_No3' | 'YLENGKETLQR' | 'HLA class I histocompatibility antigen, A alpha chain' |
| 6123 | 'HLA-A_No4' | 'FIAVGYVDDTQFVR' | 'HLA class I histocompatibility antigen, A alpha chain' |
| 6124 | 'HLA-A_No5' | 'YTCHVQHEGLPKPLTLR' | 'HLA class I histocompatibility antigen, A alpha chain' |
| 6125 | 'HLA-A_No6' | 'TDPPKTHMTHHPISDHEATLR' | 'HLA class I histocompatibility antigen, A alpha chain' |
| 6126 | 'HLA-A_No7' | 'AQSQTDRVDLGTLR' | 'HLA class I histocompatibility antigen, A alpha chain' |
| 6127 | 'HLA-A_No8' | 'TDPPKTHMTHHPISDHEATLR' | 'HLA class I histocompatibility antigen, A alpha chain' |
| 6128 | 'HLA-A_No9' | 'APWIEQEGPEYWDQETR' | 'HLA class I histocompatibility antigen, A alpha chain' |
| 6129 | 'HLA-A_No10' | 'THMTHHPISDHEATLR' | 'HLA class I histocompatibility antigen, A alpha chain' |
| 6130 | 'CAPNS1_No1' | 'THYSNIEANESEEVR' | 'Calpain small subunit 1' |
| 6131 | 'CAPNS1_No2' | 'MFLVNSFLK' | 'Calpain small subunit 1' |
| 6132 | 'CAPNS1_No3' | 'SMVAVMDSDTTGK' | 'Calpain small subunit 1' |
| 6133 | 'AMY1A_No1' | 'GHGAGGASILTFWDAR' | 'Alpha-amylase 1' |
| 6134 | 'AMY1A_No2' | 'TGSGDIENYNDATQVR' | 'Alpha-amylase 1' |
| 6135 | 'AMY1A_No3' | 'TSIVHLFEWR' | 'Alpha-amylase 1' |
| 6136 | 'AMY1A_No4' | 'LSGLLDLALGK' | 'Alpha-amylase 1' |
| 6137 | 'AMY1A_No5' | 'DFPAVPYSGWDFNDGK' | 'Alpha-amylase 1' |
| 6138 | 'AMY1A_No6' | 'IAEYMNHLIDIGVAGFR' | 'Alpha-amylase 1' |
| 6139 | 'AMY1A_No7' | 'EVTINPDTTCGNDWVCEHR' | 'Alpha-amylase 1' |
| 6140 | 'AMY1A_No8' | 'WVDIALECER' | 'Alpha-amylase 1' |
| 6141 | 'AMY1A_No9' | 'ALVFVDNHDNQR' | 'Alpha-amylase 1' |
| 6142 | 'AMY1A_No10' | 'LSGLLDLALGKDYVR' | 'Alpha-amylase 1' |
| 6143 | 'AMY1A_No11' | 'PFIYQEVIDLGGEPIK' | 'Alpha-amylase 1' |
| 6144 | 'AMY1A_No12' | 'IYVDAVINHMCGNAVSAGTSSTCGSYFNPGSR' | 'Alpha-amylase 1' |
| 6145 | 'AMY1A_No13' | 'NWGEGWGFMPSDR' | 'Alpha-amylase 1' |
| 6146 | 'AMY1A_No14' | 'MAVGFMLAHPYGFTR' | 'Alpha-amylase 1' |
| 6147 | 'AMY1A_No15' | 'AHFSISNSAEDPFIAIHAESK' | 'Alpha-amylase 1' |
| 6148 | 'AMY2A_No1' | 'LTGLLDLALEKDYVR' | 'Pancreatic alpha-amylase' |
| 6149 | 'AMY2A_No2' | 'LTGLLDLALEK' | 'Pancreatic alpha-amylase' |
| 6150 | 'AMY2A_No3' | 'NWGEGWGFVPSDR' | 'Pancreatic alpha-amylase' |
| 6151 | 'HSPB1_No1' | 'LFDQAFGLPR' | 'Heat shock protein beta-1' |
| 6152 | 'HSPB1_No2' | 'GPSWDPFRDWYPHSR' | 'Heat shock protein beta-1' |
| 6153 | 'HSPB1_No3' | 'LATQSNEITIPVTFESR' | 'Heat shock protein beta-1' |
| 6154 | 'HSPB1_No4' | 'VSLDVNHFAPDELTVK' | 'Heat shock protein beta-1' |
| 6155 | 'HSPB1_No5' | 'DGVVEITGKHEERQDEHGYISR' | 'Heat shock protein beta-1' |
| 6156 | 'RPN2' | 'LQVTNVLSQPLTQATVK' | 'Dolichyl-diphosphooligosaccharide--protein glycosyltransferase subunit 2' |
| 6157 | 'GNAI2_No1' | 'IAQSDYIPTQQDVLR' | 'Guanine nucleotide-binding protein G(i) subunit alpha-2' |
| 6158 | 'GNAI2_No2' | 'IIHEDGYSEEECR' | 'Guanine nucleotide-binding protein G(i) subunit alpha-2' |
| 6159 | 'GNAI2_No3' | 'TTGIVETHFTFK' | 'Guanine nucleotide-binding protein G(i) subunit alpha-2' |
| 6160 | 'GNAI2_No4' | 'RLWADHGVQACFGR' | 'Guanine nucleotide-binding protein G(i) subunit alpha-2' |
| 6161 | 'GNAI2_No5' | 'EYQLNDSAAYYLNDLER' | 'Guanine nucleotide-binding protein G(i) subunit alpha-2' |

| | A | B | C |
|---|---|---|---|
| 1 | **Peptide Name** | **Peptide Sequence** | **Peptide Description** |
| 6162 | 'GNAI2_No6' | 'SREYQLNDSAAYYLNDLER' | 'Guanine nucleotide-binding protein G(i) subunit alpha-2' |
| 6163 | 'GNAI2_No7' | 'AMGNLQIDFADPSR' | 'Guanine nucleotide-binding protein G(i) subunit alpha-2' |
| 6164 | 'GNAI2_No8' | 'EIYTHFTCATDTK' | 'Guanine nucleotide-binding protein G(i) subunit alpha-2' |
| 6165 | 'GNAI2_No9' | 'LWADHGVQACFGR' | 'Guanine nucleotide-binding protein G(i) subunit alpha-2' |
| 6166 | 'H2AC4_No1' | 'AGLQFPVGR' | 'Histone H2A type 1-B/E' |
| 6167 | 'H2AC4_No2' | 'VTIAQGGVLPNIQAVLLPK' | 'Histone H2A type 1-B/E' |
| 6168 | 'H2AC4_No3' | 'HLQLAIRNDEELNK' | 'Histone H2A type 1-B/E' |
| 6169 | 'H2AC4_No4' | 'NDEELNKLLGR' | 'Histone H2A type 1-B/E' |
| 6170 | 'H2AC4_No5' | 'AGLQFPVGRVHR' | 'Histone H2A type 1-B/E' |
| 6171 | 'IGF1_No1' | 'RLEMYCAPLKPAK' | 'Insulin-like growth factor I' |
| 6172 | 'IGF1_No2' | 'GFYFNKPTGYGSSSR' | 'Insulin-like growth factor I' |
| 6173 | 'IGF1_No3' | 'RLEMYCAPLKPAK' | 'Insulin-like growth factor I' |
| 6174 | 'IGF1_No4' | 'APQTGIVDECCFR' | 'Insulin-like growth factor I' |
| 6175 | 'IGF1_No5' | 'RAPQTGIVDECCFR' | 'Insulin-like growth factor I' |
| 6176 | 'ALDOB_No1' | 'IADQCPSSLAIQENANALAR' | 'Fructose-bisphosphate aldolase B' |
| 6177 | 'ALDOB_No2' | 'KYTPEQVAMATVTALHR' | 'Fructose-bisphosphate aldolase B' |
| 6178 | 'ALDOB_No3' | 'YASICQQNGLVPIVEPEVIPDGDHDLEHCQYVTEK' | 'Fructose-bisphosphate aldolase B' |
| 6179 | 'ALDOB_No4' | 'YTPEQVAMATVTALHR' | 'Fructose-bisphosphate aldolase B' |
| 6180 | 'ALDOB_No5' | 'FPALTQEQK' | 'Fructose-bisphosphate aldolase B' |
| 6181 | 'ALDOB_No6' | 'ALNDHHVYLEGTLLKPNMVTAGHACTK' | 'Fructose-bisphosphate aldolase B' |
| 6182 | 'ALDOB_No7' | 'KYTPEQVAMATVTALHR' | 'Fructose-bisphosphate aldolase B' |
| 6183 | 'ALDOB_No8' | 'GILAADESVGTMGNR' | 'Fructose-bisphosphate aldolase B' |
| 6184 | 'ALDOB_No9' | 'LDQGGAPLAGTNK' | 'Fructose-bisphosphate aldolase B' |
| 6185 | 'ALDOB_No10' | 'YTPEQVAMATVTALHR' | 'Fructose-bisphosphate aldolase B' |
| 6186 | 'ALDOB_No11' | 'DGVDFGKWR' | 'Fructose-bisphosphate aldolase B' |
| 6187 | 'APP_No1' | 'THPHFVIPYR' | 'Amyloid-beta precursor protein' |
| 6188 | 'APP_No2' | 'EVCSEQAETGPCR' | 'Amyloid-beta precursor protein' |
| 6189 | 'APP_No3' | 'STNLHDYGMLLPCGIDKFR' | 'Amyloid-beta precursor protein' |
| 6190 | 'APP_No4' | 'YLETPGDENEHAHFQK' | 'Amyloid-beta precursor protein' |
| 6191 | 'ARG1_No1' | 'VMEETLSYLLGR' | 'Arginase-1' |
| 6192 | 'ARG1_No2' | 'TIGIIGAPFSK' | 'Arginase-1' |
| 6193 | 'ARG1_No3' | 'ISLVLGGDHSLAIGSISGHAR' | 'Arginase-1' |
| 6194 | 'ARG1_No4' | 'VMEETLSYLLGR' | 'Arginase-1' |
| 6195 | 'ARG1_No5' | 'TGLLSGLDIMEVNPSLGK' | 'Arginase-1' |
| 6196 | 'ARG1_No6' | 'IPDVPGFSWVTPCISAK' | 'Arginase-1' |
| 6197 | 'ARG1_No7' | 'TGLLSGLDIMEVNPSLGK' | 'Arginase-1' |
| 6198 | 'ARG1_No8' | 'GKIPDVPGFSWVTPCISAK' | 'Arginase-1' |
| 6199 | 'APOD_No1' | 'NILTSNNIDVK' | 'Apolipoprotein D' |
| 6200 | 'APOD_No2' | 'IPTTFENGR' | 'Apolipoprotein D' |
| 6201 | 'APOD_No3' | 'MTVTDQVNCPK' | 'Apolipoprotein D' |
| 6202 | 'APOD_No4' | 'NPNLPPETVDSLK' | 'Apolipoprotein D' |
| 6203 | 'APOD_No5' | 'WYEIEKIPTTFENGR' | 'Apolipoprotein D' |
| 6204 | 'APOD_No6' | 'MTVTDQVNCPK' | 'Apolipoprotein D' |
| 6205 | 'APOD_No7' | 'NPNLPPETVDSLKNILTSNNIDVKK' | 'Apolipoprotein D' |

| | A | B | C |
|---|---|---|---|
| 1 | **Peptide Name** | **Peptide Sequence** | **Peptide Description** |
| 6206 | 'APOD_No8' | 'VLNQELR' | 'Apolipoprotein D' |
| 6207 | 'APOD_No9' | 'CPNPPVQENFDVNKYLGR' | 'Apolipoprotein D' |
| 6208 | 'APOD_No10' | 'KMTVTDQVNCPK' | 'Apolipoprotein D' |
| 6209 | 'APOD_No11' | 'NILTSNNIDVKK' | 'Apolipoprotein D' |
| 6210 | 'APOD_No12' | 'KMTVTDQVNCPK' | 'Apolipoprotein D' |
| 6211 | 'APOD_No13' | 'ADGTVNQIEGEATPVNLTEPAKLEVK' | 'Apolipoprotein D' |
| 6212 | 'APOD_No14' | 'MTVTDQVNCPKLS' | 'Apolipoprotein D' |
| 6213 | 'APOD_No15' | 'NPNLPPETVDSLKNILTSNNIDVK' | 'Apolipoprotein D' |
| 6214 | 'APOD_No16' | 'CPNPPVQENFDVNK' | 'Apolipoprotein D' |
| 6215 | 'APOD_No17' | 'KMTVTDQVNCPKLS' | 'Apolipoprotein D' |
| 6216 | 'APOD_No18' | 'IKVLNQELR' | 'Apolipoprotein D' |
| 6217 | 'APOD_No19' | 'CIQANYSLMENGK' | 'Apolipoprotein D' |
| 6218 | 'APOD_No20' | 'KMTVTDQVNCPKLS' | 'Apolipoprotein D' |
| 6219 | 'APOD_No21' | 'ADGTVNQIEGEATPVNLTEPAK' | 'Apolipoprotein D' |
| 6220 | 'APOD_No22' | 'CIQANYSLMENGK' | 'Apolipoprotein D' |
| 6221 | 'APOD_No23' | 'MTVTDQVNCPKLS' | 'Apolipoprotein D' |
| 6222 | 'ITGB3_No1' | 'YCECDDFSCVR' | 'Integrin beta-3' |
| 6223 | 'ITGB3_No2' | 'GSGDSSQVTQVSPQR' | 'Integrin beta-3' |
| 6224 | 'ITGB3_No3' | 'VLEDRPLSDKGSGDSSQVTQVSPQR' | 'Integrin beta-3' |
| 6225 | 'ITGB3_No4' | 'HVLTLTDQVTR' | 'Integrin beta-3' |
| 6226 | 'ITGB3_No5' | 'SILYVVEEPECPK' | 'Integrin beta-3' |
| 6227 | 'ITGB3_No6' | 'NDASHLLVFTTDAK' | 'Integrin beta-3' |
| 6228 | 'ITGB3_No7' | 'SFTIKPVGFK' | 'Integrin beta-3' |
| 6229 | 'ITGB3_No8' | 'YCRDEIESVK' | 'Integrin beta-3' |
| 6230 | 'ITGB3_No9' | 'CPTCPDACTFK' | 'Integrin beta-3' |
| 6231 | 'ITGB3_No10' | 'AKWDTANNPLYK' | 'Integrin beta-3' |
| 6232 | 'ITGB3_No11' | 'CPTCPDACTFKK' | 'Integrin beta-3' |
| 6233 | 'ITGB3_No12' | 'GECLCGQCVCHSSDFGK' | 'Integrin beta-3' |
| 6234 | 'ITGB3_No13' | 'LAGIVQPNDGQCHVGSDNHYSASTTMDYPSLGLMTEK' | 'Integrin beta-3' |
| 6235 | 'ITGB3_No14' | 'TTCLPMFGYK' | 'Integrin beta-3' |
| 6236 | 'ITGB3_No15' | 'TDTCMSSNGLLCSGR' | 'Integrin beta-3' |
| 6237 | 'ITGB3_No16' | 'GKCECGSCVCIQPGSYGDTCEK' | 'Integrin beta-3' |
| 6238 | 'ITGB3_No17' | 'CECGSCVCIQPGSYGDTCEK' | 'Integrin beta-3' |
| 6239 | 'ITGB3_No18' | 'GECLCGQCVCHSSDFGKITGK' | 'Integrin beta-3' |
| 6240 | 'ITGB3_No19' | 'DAPEGGFDAIMQATVCDEK' | 'Integrin beta-3' |
| 6241 | 'ITGB3_No20' | 'IGFGAFVDKPVSPYMYISPPEALENPCYDMK' | 'Integrin beta-3' |
| 6242 | 'ITGB3_No21' | 'LAGIVQPNDGQCHVGSDNHYSASTTMDYPSLGLMTEK' | 'Integrin beta-3' |
| 6243 | 'ITGB3_No22' | 'NRDAPEGGFDAIMQATVCDEK' | 'Integrin beta-3' |
| 6244 | 'ITGB3_No23' | 'CDLKENLLKDNCAPESIEFPVSEAR' | 'Integrin beta-3' |
| 6245 | 'ITGB3_No24' | 'CGPGWLGSQCECSEEDYRPSQQDECSPR' | 'Integrin beta-3' |
| 6246 | 'ITGB3_No25' | 'EATSTFTNITYRGT' | 'Integrin beta-3' |
| 6247 | 'ITGB3_No26' | 'DDLWSIQNLGTK' | 'Integrin beta-3' |
| 6248 | 'ITGB3_No27' | 'DAPEGGFDAIMQATVCDEK' | 'Integrin beta-3' |
| 6249 | 'S100A8_No1' | 'ALNSIIDVYHK' | 'Protein S100-A8' |

| | A | B | C |
|---|---|---|---|
| 1 | **Peptide Name** | **Peptide Sequence** | **Peptide Description** |
| 6250 | 'S100A8_No2' | 'GNFHAVYRDDLK' | 'Protein S100-A8' |
| 6251 | 'S100A8_No3' | 'KLLETECPQYIR' | 'Protein S100-A8' |
| 6252 | 'S100A8_No4' | 'LLETECPQYIR' | 'Protein S100-A8' |
| 6253 | 'S100A8_No5' | 'LLETECPQYIRK' | 'Protein S100-A8' |
| 6254 | 'S100A8_No6' | 'GNFHAVYRDDLKK' | 'Protein S100-A8' |
| 6255 | 'SERPINE1_No1' | 'LVQGFMPHFFR' | 'Plasminogen activator inhibitor 1' |
| 6256 | 'SERPINE1_No2' | 'QFQADFTSLSDQEPLHVAQALQK' | 'Plasminogen activator inhibitor 1' |
| 6257 | 'SERPINA5_No1' | 'AAAATGTIFTFR' | 'Plasma serine protease inhibitor' |
| 6258 | 'SERPINA5_No2' | 'AVVEVDESGTR' | 'Plasma serine protease inhibitor' |
| 6259 | 'SERPINA5_No3' | 'GTQEQDFYVTSETVVR' | 'Plasma serine protease inhibitor' |
| 6260 | 'SERPINA5_No4' | 'DFTFDLYR' | 'Plasma serine protease inhibitor' |
| 6261 | 'SERPINA5_No5' | 'GFQQLLQELNQPR' | 'Plasma serine protease inhibitor' |
| 6262 | 'SERPINA5_No6' | 'TLYLADTFPTNFR' | 'Plasma serine protease inhibitor' |
| 6263 | 'SERPINA5_No7' | 'MQQVENGLSEK' | 'Plasma serine protease inhibitor' |
| 6264 | 'SERPINA5_No8' | 'FSIEGSYQLEK' | 'Plasma serine protease inhibitor' |
| 6265 | 'SERPINA5_No9' | 'MQQVENGLSEK' | 'Plasma serine protease inhibitor' |
| 6266 | 'SERPINA5_No10' | 'MQILEGLGLNLQK' | 'Plasma serine protease inhibitor' |
| 6267 | 'SERPINA5_No11' | 'MQILEGLGLNLQK' | 'Plasma serine protease inhibitor' |
| 6268 | 'SERPINA5_No12' | 'RVEDLHVGATVAPSSR' | 'Plasma serine protease inhibitor' |
| 6269 | 'SERPINA5_No13' | 'EDQYHYLLDR' | 'Plasma serine protease inhibitor' |
| 6270 | 'SERPINA5_No14' | 'VEDLHVGATVAPSSR' | 'Plasma serine protease inhibitor' |
| 6271 | 'SERPINA5_No15' | 'GKIVDLLK' | 'Plasma serine protease inhibitor' |
| 6272 | 'SERPINA5_No16' | 'RDFTFDLYR' | 'Plasma serine protease inhibitor' |
| 6273 | 'SERPINA5_No17' | 'QINDYVAK' | 'Plasma serine protease inhibitor' |
| 6274 | 'SERPINA5_No18' | 'VVGVPYQGNATALFILPSEGK' | 'Plasma serine protease inhibitor' |
| 6275 | 'SERPINA5_No19' | 'AKWETSFNHK' | 'Plasma serine protease inhibitor' |
| 6276 | 'SERPINA5_No20' | 'TLYLADTFPTNFRDSAGAMK' | 'Plasma serine protease inhibitor' |
| 6277 | 'SERPINA5_No21' | 'TLYLADTFPTNFRDSAGAMK' | 'Plasma serine protease inhibitor' |
| 6278 | 'SERPING1_No1' | 'LVLLNAIYLSAK' | 'Plasma protease C1 inhibitor' |
| 6279 | 'SERPING1_No2' | 'VTTSQDMLSIMEK' | 'Plasma protease C1 inhibitor' |
| 6280 | 'SERPING1_No3' | 'TNLESILSYPKDFTCVHQALK' | 'Plasma protease C1 inhibitor' |
| 6281 | 'SERPING1_No4' | 'KYPVAHFIDQTLK' | 'Plasma protease C1 inhibitor' |
| 6282 | 'SERPING1_No5' | 'FPVFMGR' | 'Plasma protease C1 inhibitor' |
| 6283 | 'SERPING1_No6' | 'DFTCVHQALK' | 'Plasma protease C1 inhibitor' |
| 6284 | 'SERPING1_No7' | 'LEDMEQALSPSVFK' | 'Plasma protease C1 inhibitor' |
| 6285 | 'SERPING1_No8' | 'VTTSQDMLSIMEK' | 'Plasma protease C1 inhibitor' |
| 6286 | 'SERPING1_No9' | 'FPVFMGR' | 'Plasma protease C1 inhibitor' |
| 6287 | 'SERPING1_No10' | 'VTTSQDMLSIMEK' | 'Plasma protease C1 inhibitor' |
| 6288 | 'SERPING1_No11' | 'VTTSQDMLSIMEK' | 'Plasma protease C1 inhibitor' |
| 6289 | 'SERPING1_No12' | 'IKVTTSQDMLSIMEK' | 'Plasma protease C1 inhibitor' |
| 6290 | 'SERPING1_No13' | 'TLYSSSPR' | 'Plasma protease C1 inhibitor' |
| 6291 | 'SERPING1_No14' | 'MEPFHFK' | 'Plasma protease C1 inhibitor' |
| 6292 | 'SERPING1_No15' | 'AIMEKLEMSK' | 'Plasma protease C1 inhibitor' |
| 6293 | 'SERPING1_No16' | 'KVETNMAFSPFSIASLLTQVLLGAGENTK' | 'Plasma protease C1 inhibitor' |

| | A | B | C |
|---|---|---|---|
| 1 | **Peptide Name** | **Peptide Sequence** | **Peptide Description** |
| 6294 | 'SERPING1_No17' | 'LLDSLPSDTR' | 'Plasma protease C1 inhibitor' |
| 6295 | 'SERPING1_No18' | 'YPVAHFIDQTLK' | 'Plasma protease C1 inhibitor' |
| 6296 | 'SERPING1_No19' | 'GVTSVSQIFHSPDLAIR' | 'Plasma protease C1 inhibitor' |
| 6297 | 'SERPING1_No20' | 'LYHAFSAMK' | 'Plasma protease C1 inhibitor' |
| 6298 | 'SERPING1_No21' | 'HRLEDMEQALSPSVFK' | 'Plasma protease C1 inhibitor' |
| 6299 | 'SERPING1_No22' | 'HRLEDMEQALSPSVFK' | 'Plasma protease C1 inhibitor' |
| 6300 | 'SERPING1_No23' | 'NSVIKVPMMNSK' | 'Plasma protease C1 inhibitor' |
| 6301 | 'SERPING1_No24' | 'IKVTTSQDMLSIMEK' | 'Plasma protease C1 inhibitor' |
| 6302 | 'SERPING1_No25' | 'DFTCVHQALKGFTTK' | 'Plasma protease C1 inhibitor' |
| 6303 | 'SERPING1_No26' | 'AIMEKLEMSK' | 'Plasma protease C1 inhibitor' |
| 6304 | 'SERPING1_No27' | 'VPMMNSK' | 'Plasma protease C1 inhibitor' |
| 6305 | 'SERPING1_No28' | 'MEPFHFK' | 'Plasma protease C1 inhibitor' |
| 6306 | 'SERPING1_No29' | 'TNLESILSYPK' | 'Plasma protease C1 inhibitor' |
| 6307 | 'SERPING1_No30' | 'FQPTLLTLPR' | 'Plasma protease C1 inhibitor' |
| 6308 | 'SERPING1_No31' | 'LEDMEQALSPSVFK' | 'Plasma protease C1 inhibitor' |
| 6309 | 'SERPING1_No32' | 'LYHAFSAMK' | 'Plasma protease C1 inhibitor' |
| 6310 | 'SERPING1_No33' | 'TNLESILSYPKDFTCVHQALKGFTTK' | 'Plasma protease C1 inhibitor' |
| 6311 | 'SERPING1_No34' | 'AIMEKLEMSK' | 'Plasma protease C1 inhibitor' |
| 6312 | 'SERPING1_No35' | 'AIMEKLEMSKFQPTLLTLPR' | 'Plasma protease C1 inhibitor' |
| 6313 | 'SERPING1_No36' | 'IKVTTSQDMLSIMEK' | 'Plasma protease C1 inhibitor' |
| 6314 | 'SERPING1_No37' | 'NSVIKVPMMNSK' | 'Plasma protease C1 inhibitor' |
| 6315 | 'SERPING1_No38' | 'VLSNNSDANLELINTWVAK' | 'Plasma protease C1 inhibitor' |
| 6316 | 'SERPING1_No39' | 'NSVIKVPMMNSK' | 'Plasma protease C1 inhibitor' |
| 6317 | 'SERPING1_No40' | 'VGQLQLSHNLSLVILVPQNLK' | 'Plasma protease C1 inhibitor' |
| 6318 | 'SERPING1_No41' | 'LEMSKFQPTLLTLPR' | 'Plasma protease C1 inhibitor' |
| 6319 | 'SERPING1_No42' | 'KYPVAHFIDQTLKAK' | 'Plasma protease C1 inhibitor' |
| 6320 | 'SERPING1_No43' | 'IKVTTSQDMLSIMEK' | 'Plasma protease C1 inhibitor' |
| 6321 | 'SERPING1_No44' | 'LLDSLPSDTRLVLLNAIYLSAK' | 'Plasma protease C1 inhibitor' |
| 6322 | 'SERPING1_No45' | 'AIMEKLEMSK' | 'Plasma protease C1 inhibitor' |
| 6323 | 'SERPING1_No46' | 'TTFDPKK' | 'Plasma protease C1 inhibitor' |
| 6324 | 'SERPING1_No47' | 'MEPFHFKNSVIK' | 'Plasma protease C1 inhibitor' |
| 6325 | 'SERPING1_No48' | 'DFTCVHQALKGFTTKGVTSVSQIFHSPDLAIR' | 'Plasma protease C1 inhibitor' |
| 6326 | 'SERPING1_No49' | 'LEMSKFQPTLLTLPR' | 'Plasma protease C1 inhibitor' |
| 6327 | 'SERPING1_No50' | 'YPVAHFIDQTLKAK' | 'Plasma protease C1 inhibitor' |
| 6328 | 'SERPING1_No51' | 'MEPFHFKNSVIK' | 'Plasma protease C1 inhibitor' |
| 6329 | 'CFI_No1' | 'YQIWTTVVDWIHPDLKR' | 'Complement factor I' |
| 6330 | 'CFI_No2' | 'VFSLQWGEVK' | 'Complement factor I' |
| 6331 | 'CFI_No3' | 'VFCQPWQR' | 'Complement factor I' |
| 6332 | 'CFI_No4' | 'VANYFDWISYHVGR' | 'Complement factor I' |
| 6333 | 'CFI_No5' | 'GLETSLAECTFTK' | 'Complement factor I' |
| 6334 | 'CFI_No6' | 'TMGYQDFADVVCYTQK' | 'Complement factor I' |
| 6335 | 'CFI_No7' | 'LVDQDKTMFICK' | 'Complement factor I' |
| 6336 | 'CFI_No8' | 'IIFHENYNAGTYQNDIALIEMK' | 'Complement factor I' |
| 6337 | 'CFI_No9' | 'ADSPMDDFFQCVNGK' | 'Complement factor I' |

| | A | B | C |
|---|---|---|---|
| 1 | **Peptide Name** | **Peptide Sequence** | **Peptide Description** |
| 6338 | 'CFI_No10' | 'EMECAGTYDGSIDACK' | 'Complement factor I' |
| 6339 | 'CFI_No11' | 'ACDGINDCGDQSDELCCK' | 'Complement factor I' |
| 6340 | 'CFI_No12' | 'RAQLGDLPWQVAIK' | 'Complement factor I' |
| 6341 | 'CFI_No13' | 'RIVIEYVDR' | 'Complement factor I' |
| 6342 | 'CFI_No14' | 'SLECLHPGTK' | 'Complement factor I' |
| 6343 | 'CFI_No15' | 'VTYTSQEDLVEKK' | 'Complement factor I' |
| 6344 | 'CFI_No16' | 'VANYFDWISYHVGRPFISQYNV' | 'Complement factor I' |
| 6345 | 'CFI_No17' | 'LVDQDKTMFICK' | 'Complement factor I' |
| 6346 | 'CFI_No18' | 'LPYQCPK' | 'Complement factor I' |
| 6347 | 'CFI_No19' | 'KYTHLSCDKVFCQPWQR' | 'Complement factor I' |
| 6348 | 'CFI_No20' | 'ADSPMDDFFQCVNGK' | 'Complement factor I' |
| 6349 | 'CFI_No21' | 'IIFHENYNAGTYQNDIALIEMKK' | 'Complement factor I' |
| 6350 | 'CFI_No22' | 'YTHLSCDKVFCQPWQR' | 'Complement factor I' |
| 6351 | 'CFI_No23' | 'HGNTDSEGIVEVK' | 'Complement factor I' |
| 6352 | 'CFI_No24' | 'CIEGTCVCK' | 'Complement factor I' |
| 6353 | 'CFI_No25' | 'FYEKEMECAGTYDGSIDACK' | 'Complement factor I' |
| 6354 | 'CFI_No26' | 'YQIWTTVVDWIHPDLK' | 'Complement factor I' |
| 6355 | 'CFI_No27' | 'GLETSLAECTFTKR' | 'Complement factor I' |
| 6356 | 'CFI_No28' | 'TMGYQDFADVVCYTQK' | 'Complement factor I' |
| 6357 | 'CFI_No29' | 'AQLGDLPWQVAIK' | 'Complement factor I' |
| 6358 | 'CFI_No30' | 'SFPTYCQQK' | 'Complement factor I' |
| 6359 | 'CFI_No31' | 'IIFHENYNAGTYQNDIALIEMK' | 'Complement factor I' |
| 6360 | 'CFI_No32' | 'EANVACLDLGFQQGADTQR' | 'Complement factor I' |
| 6361 | 'CFI_No33' | 'FSVSLKHGNTDSEGIVEVK' | 'Complement factor I' |
| 6362 | 'CFI_No34' | 'IVIEYVDR' | 'Complement factor I' |
| 6363 | 'CFI_No35' | 'CIEGTCVCKLPYQCPK' | 'Complement factor I' |
| 6364 | 'CFI_No36' | 'VTYTSQEDLVEK' | 'Complement factor I' |
| 6365 | 'CFI_No37' | 'RSFPTYCQQK' | 'Complement factor I' |
| 6366 | 'CFI_No38' | 'EMECAGTYDGSIDACK' | 'Complement factor I' |
| 6367 | 'CFI_No39' | 'EANVACLDLGFQQGADTQRR' | 'Complement factor I' |
| 6368 | 'CFI_No40' | 'DASGITCGGIYIGGCWILTAAHCLR' | 'Complement factor I' |
| 6369 | 'CFI_No41' | 'HGNTDSEGIVEVKLVDQDKTMFICK' | 'Complement factor I' |
| 6370 | 'CFI_No42' | 'EKDNERVFSLQWGEVK' | 'Complement factor I' |
| 6371 | 'CFI_No43' | 'IIFHENYNAGTYQNDIALIEMKK' | 'Complement factor I' |
| 6372 | 'CFI_No44' | 'RTMGYQDFADVVCYTQK' | 'Complement factor I' |
| 6373 | 'CFI_No45' | 'RTMGYQDFADVVCYTQK' | 'Complement factor I' |
| 6374 | 'CFI_No46' | 'ACDGINDCGDQSDELCCKACQGK' | 'Complement factor I' |
| 6375 | 'CFI_No47' | 'FYEKEMECAGTYDGSIDACK' | 'Complement factor I' |
| 6376 | 'CFI_No48' | 'DGNKKDCELPR' | 'Complement factor I' |
| 6377 | 'CFI_No49' | 'HGNTDSEGIVEVKLVDQDKTMFICK' | 'Complement factor I' |
| 6378 | 'CFI_No50' | 'YTHLSCDK' | 'Complement factor I' |
| 6379 | 'CFI_No51' | 'KYTHLSCDK' | 'Complement factor I' |
| 6380 | 'CFI_No52' | 'GLETSLAECTFTKRR' | 'Complement factor I' |
| 6381 | 'F13B_No1' | 'LIENGYFHPVK' | 'Coagulation factor XIII B chain' |

| | A | B | C |
|---|---|---|---|
| 1 | **Peptide Name** | **Peptide Sequence** | **Peptide Description** |
| 6382 | 'F13B_No2' | 'LSFFCLAGYTTESGR' | 'Coagulation factor XIII B chain' |
| 6383 | 'F13B_No3' | 'VQYECATGYYTAGGK' | 'Coagulation factor XIII B chain' |
| 6384 | 'F13B_No4' | 'TTGGKDEEVVQCLSDGWSSQPTCR' | 'Coagulation factor XIII B chain' |
| 6385 | 'F13B_No5' | 'QGYDLSPLTPLSELSVQCNR' | 'Coagulation factor XIII B chain' |
| 6386 | 'F13B_No6' | 'HPPVVMNGAVADGILASYATGSSVEYR' | 'Coagulation factor XIII B chain' |
| 6387 | 'F13B_No7' | 'WSSPPVCLEPCTVNVDYMNR' | 'Coagulation factor XIII B chain' |
| 6388 | 'F13B_No8' | 'WDFDNRPHILHGEYIEFICR' | 'Coagulation factor XIII B chain' |
| 6389 | 'F13B_No9' | 'GMCTSPPLIK' | 'Coagulation factor XIII B chain' |
| 6390 | 'F13B_No10' | 'WTLPPECVENNENCK' | 'Coagulation factor XIII B chain' |
| 6391 | 'F13B_No11' | 'VACEEPPFIENGAANLHSK' | 'Coagulation factor XIII B chain' |
| 6392 | 'F13B_No12' | 'QSTLSYQEPLRT' | 'Coagulation factor XIII B chain' |
| 6393 | 'F13B_No13' | 'CNEYYLLR' | 'Coagulation factor XIII B chain' |
| 6394 | 'F13B_No14' | 'IYYNGDKVTYACK' | 'Coagulation factor XIII B chain' |
| 6395 | 'F13B_No15' | 'SFYFPMSIDKK' | 'Coagulation factor XIII B chain' |
| 6396 | 'F13B_No16' | 'GDTYPAELYITGSILR' | 'Coagulation factor XIII B chain' |
| 6397 | 'F13B_No17' | 'SFYFPMSIDK' | 'Coagulation factor XIII B chain' |
| 6398 | 'F13B_No18' | 'CPPPPLPINSK' | 'Coagulation factor XIII B chain' |
| 6399 | 'F13B_No19' | 'QEEQTTCTTEGWSPEPR' | 'Coagulation factor XIII B chain' |
| 6400 | 'F13B_No20' | 'CTKPDLSNGYISDVK' | 'Coagulation factor XIII B chain' |
| 6401 | 'F13B_No21' | 'TEEVECLTYGWSLTPK' | 'Coagulation factor XIII B chain' |
| 6402 | 'F13B_No22' | 'IAQYYYTFK' | 'Coagulation factor XIII B chain' |
| 6403 | 'F13B_No23' | 'HPPVVMNGAVADGILASYATGSSVEYR' | 'Coagulation factor XIII B chain' |
| 6404 | 'F13B_No24' | 'SGYLLHGSNEITCNR' | 'Coagulation factor XIII B chain' |
| 6405 | 'F13B_No25' | 'VKDKVQYECATGYYTAGGK' | 'Coagulation factor XIII B chain' |
| 6406 | 'F13B_No26' | 'HGEIVHIECELNFEIHGSAEIR' | 'Coagulation factor XIII B chain' |
| 6407 | 'F13B_No27' | 'CFDHHFLEGSR' | 'Coagulation factor XIII B chain' |
| 6408 | 'F13B_No28' | 'QEEQTTCTTEGWSPEPR' | 'Coagulation factor XIII B chain' |
| 6409 | 'F13B_No29' | 'VLHGDLIDFVCK' | 'Coagulation factor XIII B chain' |
| 6410 | 'F13B_No30' | 'SFYFPMSIDKK' | 'Coagulation factor XIII B chain' |
| 6411 | 'F13B_No31' | 'VQYECATGYYTAGGKK' | 'Coagulation factor XIII B chain' |
| 6412 | 'F13B_No32' | 'GEVKYPLCTR' | 'Coagulation factor XIII B chain' |
| 6413 | 'F13B_No33' | 'DKVQYECATGYYTAGGKK' | 'Coagulation factor XIII B chain' |
| 6414 | 'F13B_No34' | 'KCTKPDLSNGYISDVK' | 'Coagulation factor XIII B chain' |
| 6415 | 'F13B_No35' | 'GMCTSPPLIK' | 'Coagulation factor XIII B chain' |
| 6416 | 'F13B_No36' | 'KLSFFCLAGYTTESGR' | 'Coagulation factor XIII B chain' |
| 6417 | 'F13B_No37' | 'GKWTLPPECVENNENCK' | 'Coagulation factor XIII B chain' |
| 6418 | 'F13B_No38' | 'KTEEVECLTYGWSLTPK' | 'Coagulation factor XIII B chain' |
| 6419 | 'F13B_No39' | 'NRCPPPPLPINSK' | 'Coagulation factor XIII B chain' |
| 6420 | 'F13B_No40' | 'QGYDLSPLTPLSELSVQCNRGEVK' | 'Coagulation factor XIII B chain' |
| 6421 | 'F13B_No41' | 'DKVQYECATGYYTAGGK' | 'Coagulation factor XIII B chain' |
| 6422 | 'F13B_No42' | 'QGYDLSPLTPLSELSVQCNRGEVKYPLCTR' | 'Coagulation factor XIII B chain' |
| 6423 | 'F13B_No43' | 'DEEVVQCLSDGWSSQPTCR' | 'Coagulation factor XIII B chain' |
| 6424 | 'F13B_No44' | 'MQCDRGQLKYPR' | 'Coagulation factor XIII B chain' |
| 6425 | 'F13B_No45' | 'CEDGKWTEPPKCIEGQEK' | 'Coagulation factor XIII B chain' |

| | A | B | C |
|---|---|---|---|
| 1 | **Peptide Name** | **Peptide Sequence** | **Peptide Description** |
| 6426 | 'F13B_No46' | 'WTEPPKCIEGQEK' | 'Coagulation factor XIII B chain' |
| 6427 | 'F13B_No47' | 'SFYFPMSIDK' | 'Coagulation factor XIII B chain' |
| 6428 | 'F13B_No48' | 'IQTHSTTYR' | 'Coagulation factor XIII B chain' |
| 6429 | 'F13B_No49' | 'LSFFCLAGYTTESGRQEEQTTCTTEGWSPEPR' | 'Coagulation factor XIII B chain' |
| 6430 | 'F13B_No50' | 'TTGGKDEEVVQCLSDGWSSQPTCRK' | 'Coagulation factor XIII B chain' |
| 6431 | 'F13B_No51' | 'QGYDLSPLTPLSELSVQCNR' | 'Coagulation factor XIII B chain' |
| 6432 | 'F13B_No52' | 'CTKPDLSNGYISDVKLLYK' | 'Coagulation factor XIII B chain' |
| 6433 | 'F13B_No53' | 'CEQGKWSSPPVCLEPCTVNVDYMNR' | 'Coagulation factor XIII B chain' |
| 6434 | 'MPO_No1' | 'VVLEGGIDPILR' | 'Myeloperoxidase' |
| 6435 | 'MPO_No2' | 'IICDNTGITTVSK' | 'Myeloperoxidase' |
| 6436 | 'MPO_No3' | 'IGLDLPALNMQR' | 'Myeloperoxidase' |
| 6437 | 'MPO_No4' | 'VFFASWR' | 'Myeloperoxidase' |
| 6438 | 'MPO_No5' | 'FPTDQLTPDQER' | 'Myeloperoxidase' |
| 6439 | 'MPO_No6' | 'VGPLLACIIGTQFR' | 'Myeloperoxidase' |
| 6440 | 'MPO_No7' | 'QNQIAVDEIR' | 'Myeloperoxidase' |
| 6441 | 'MPO_No8' | 'DYLPLVLGPTAMR' | 'Myeloperoxidase' |
| 6442 | 'MPO_No9' | 'IANVFTNAFR' | 'Myeloperoxidase' |
| 6443 | 'MPO_No10' | 'QALAQISLPR' | 'Myeloperoxidase' |
| 6444 | 'MPO_No11' | 'IKNQADCIPFFR' | 'Myeloperoxidase' |
| 6445 | 'MPO_No12' | 'IGLDLPALNMQR' | 'Myeloperoxidase' |
| 6446 | 'MPO_No13' | 'SSEMPELTSMHTLLLR' | 'Myeloperoxidase' |
| 6447 | 'MPO_No14' | 'FWWENEGVFSMQQR' | 'Myeloperoxidase' |
| 6448 | 'MPO_No15' | 'DYLPLVLGPTAMR' | 'Myeloperoxidase' |
| 6449 | 'MPO_No16' | 'SRDHGLPGYNAWR' | 'Myeloperoxidase' |
| 6450 | 'ICAM1_No1' | 'VELAPLPSWQPVGK' | 'Intercellular adhesion molecule 1' |
| 6451 | 'ICAM1_No2' | 'LLGIETPLPK' | 'Intercellular adhesion molecule 1' |
| 6452 | 'ICAM1_No3' | 'GGSVLVTCSTSCDQPK' | 'Intercellular adhesion molecule 1' |
| 6453 | 'ICAM1_No4' | 'TFLTVYWTPER' | 'Intercellular adhesion molecule 1' |
| 6454 | 'ICAM1_No5' | 'ASVSVTAEDEGTQR' | 'Intercellular adhesion molecule 1' |
| 6455 | 'ICAM1_No6' | 'DGTFPLPIGESVTVTR' | 'Intercellular adhesion molecule 1' |
| 6456 | 'ICAM1_No7' | 'EPAVGEPAEVTTTVLVR' | 'Intercellular adhesion molecule 1' |
| 6457 | 'ICAM1_No8' | 'ELKREPAVGEPAEVTTTVLVR' | 'Intercellular adhesion molecule 1' |
| 6458 | 'ICAM1_No9' | 'SFSCSATLEVAGQLIHK' | 'Intercellular adhesion molecule 1' |
| 6459 | 'ICAM1_No10' | 'CLKDGTFPLPIGESVTVTR' | 'Intercellular adhesion molecule 1' |
| 6460 | 'ICAM1_No11' | 'VYELSNVQEDSQPMCYSNCPDGQSTAK' | 'Intercellular adhesion molecule 1' |
| 6461 | 'ICAM1_No12' | 'DLEGTYLCR' | 'Intercellular adhesion molecule 1' |
| 6462 | 'RPLP0' | 'AGAIAPCEVTVPAQNTGLGPEK' | '60S acidic ribosomal protein P0' |
| 6463 | 'REG1A_No1' | 'ISCPEGTNAYR' | 'Lithostathine-1-alpha' |
| 6464 | 'REG1A_No2' | 'ESGTDDFNVWIGLHDPKK' | 'Lithostathine-1-alpha' |
| 6465 | 'REG1A_No3' | 'WHWSSGSLVSYK' | 'Lithostathine-1-alpha' |
| 6466 | 'REG1A_No4' | 'DVPCEDKFSFVCK' | 'Lithostathine-1-alpha' |
| 6467 | 'REG1A_No5' | 'ESGTDDFNVWIGLHDPK' | 'Lithostathine-1-alpha' |
| 6468 | 'REG1A_No6' | 'WKDVPCEDKFSFVCK' | 'Lithostathine-1-alpha' |
| 6469 | 'CLEC3B_No1' | 'LDTLAQEVALLK' | 'Tetranectin' |

| | A | B | C |
|---|---|---|---|
| 1 | **Peptide Name** | **Peptide Sequence** | **Peptide Description** |
| 6470 | 'CLEC3B_No2' | 'SRLDTLAQEVALLK' | 'Tetranectin' |
| 6471 | 'CLEC3B_No3' | 'TFHEASEDCISR' | 'Tetranectin' |
| 6472 | 'CLEC3B_No4' | 'TENCAVLSGAANGK' | 'Tetranectin' |
| 6473 | 'CLEC3B_No5' | 'NWETEITAQPDGGKTENCAVLSGAANGK' | 'Tetranectin' |
| 6474 | 'CLEC3B_No6' | 'EQQALQTVCLK' | 'Tetranectin' |
| 6475 | 'CLEC3B_No7' | 'CFLAFTQTK' | 'Tetranectin' |
| 6476 | 'CLEC3B_No8' | 'GGTLGTPQTGSENDALYEYLR' | 'Tetranectin' |
| 6477 | 'CLEC3B_No9' | 'CRDQLPYICQFGIV' | 'Tetranectin' |
| 6478 | 'CLEC3B_No10' | 'SRLDTLAQEVALLKEQQALQTVCLK' | 'Tetranectin' |
| 6479 | 'CLEC3B_No11' | 'IAYKNWETEITAQPDGGK' | 'Tetranectin' |
| 6480 | 'CLEC3B_No12' | 'IAYKNWETEITAQPDGGKTENCAVLSGAANGK' | 'Tetranectin' |
| 6481 | 'CLEC3B_No13' | 'MFEELKSR' | 'Tetranectin' |
| 6482 | 'CLEC3B_No14' | 'LDTLAQEVALLKEQQALQTVCLKGTK' | 'Tetranectin' |
| 6483 | 'CLEC3B_No15' | 'LDTLAQEVALLKEQQALQTVCLK' | 'Tetranectin' |
| 6484 | 'CLEC3B_No16' | 'MFEELKSR' | 'Tetranectin' |
| 6485 | 'CLEC3B_No17' | 'EQQALQTVCLKGTK' | 'Tetranectin' |
| 6486 | 'SERPINA7_No1' | 'FSISATYDLGATLLK' | 'Thyroxine-binding globulin' |
| 6487 | 'SERPINA7_No2' | 'SFMLLILER' | 'Thyroxine-binding globulin' |
| 6488 | 'SERPINA7_No3' | 'KELELQIGNALFIGK' | 'Thyroxine-binding globulin' |
| 6489 | 'SERPINA7_No4' | 'AQWANPFDPSKTEDSSSFLIDK' | 'Thyroxine-binding globulin' |
| 6490 | 'SERPINA7_No5' | 'MGIQHAYSENADFSGLTEDNGLK' | 'Thyroxine-binding globulin' |
| 6491 | 'SERPINA7_No6' | 'EGQMESVEAAMSSK' | 'Thyroxine-binding globulin' |
| 6492 | 'SERPINA7_No7' | 'NALALFVLPK' | 'Thyroxine-binding globulin' |
| 6493 | 'SERPINA7_No8' | 'MSSINADFAFNLYR' | 'Thyroxine-binding globulin' |
| 6494 | 'SERPINA7_No9' | 'ELELQIGNALFIGK' | 'Thyroxine-binding globulin' |
| 6495 | 'SERPINA7_No10' | 'EGQMESVEAAMSSK' | 'Thyroxine-binding globulin' |
| 6496 | 'SERPINA7_No11' | 'GWVDLFVPK' | 'Thyroxine-binding globulin' |
| 6497 | 'SERPINA7_No12' | 'SFMLLILER' | 'Thyroxine-binding globulin' |
| 6498 | 'SERPINA7_No13' | 'GTEAAAVPEVELSDQPENTFLHPIIQIDR' | 'Thyroxine-binding globulin' |
| 6499 | 'SERPINA7_No14' | 'EGQMESVEAAMSSK' | 'Thyroxine-binding globulin' |
| 6500 | 'SERPINA7_No15' | 'MGIQHAYSENADFSGLTEDNGLK' | 'Thyroxine-binding globulin' |
| 6501 | 'SERPINA7_No16' | 'FTVETPDK' | 'Thyroxine-binding globulin' |
| 6502 | 'SERPINA7_No17' | 'QEINSHVEMQTK' | 'Thyroxine-binding globulin' |
| 6503 | 'SERPINA7_No18' | 'TEDSSSFLIDK' | 'Thyroxine-binding globulin' |
| 6504 | 'SERPINA7_No19' | 'QEINSHVEMQTK' | 'Thyroxine-binding globulin' |
| 6505 | 'SERPINA7_No20' | 'VVGLIQDLKPNTIMVLVNYIHFK' | 'Thyroxine-binding globulin' |
| 6506 | 'SERPINA7_No21' | 'MSSINADFAFNLYR' | 'Thyroxine-binding globulin' |
| 6507 | 'SERPINA7_No22' | 'MGIQHAYSENADFSGLTEDNGLKLSNAAHK' | 'Thyroxine-binding globulin' |
| 6508 | 'SERPINA7_No23' | 'MGIQHAYSENADFSGLTEDNGLKLSNAAHK' | 'Thyroxine-binding globulin' |
| 6509 | 'SERPINA7_No24' | 'QEINSHVEMQTK' | 'Thyroxine-binding globulin' |
| 6510 | 'SERPINA7_No25' | 'TLYETEVFSTDFSNISAAK' | 'Thyroxine-binding globulin' |
| 6511 | 'SERPINA7_No26' | 'QEINSHVEMQTK' | 'Thyroxine-binding globulin' |
| 6512 | 'SERPINA7_No27' | 'AVLHIGEKGTEAAAVPEVELSDQPENTFLHPIIQIDR' | 'Thyroxine-binding globulin' |
| 6513 | 'SERPINA7_No28' | 'AVLHIGEK' | 'Thyroxine-binding globulin' |

| | A | B | C |
|---|---|---|---|
| 1 | **Peptide Name** | **Peptide Sequence** | **Peptide Description** |
| 6514 | 'SERPINA7_No29' | 'EGQMESVEAAMSSK' | 'Thyroxine-binding globulin' |
| 6515 | 'SERPINA7_No30' | 'RFTVETPDK' | 'Thyroxine-binding globulin' |
| 6516 | 'SERPINA7_No31' | 'SILFLGKVVNPTEA' | 'Thyroxine-binding globulin' |
| 6517 | 'SERPINA7_No32' | 'LLQKGWVDLFVPK' | 'Thyroxine-binding globulin' |
| 6518 | 'SERPIND1_No1' | 'FAFNLYR' | 'Heparin cofactor 2' |
| 6519 | 'SERPIND1_No2' | 'IAIDLFK' | 'Heparin cofactor 2' |
| 6520 | 'SERPIND1_No3' | 'YEITTIHNLFR' | 'Heparin cofactor 2' |
| 6521 | 'SERPIND1_No4' | 'GPLDQLEK' | 'Heparin cofactor 2' |
| 6522 | 'SERPIND1_No5' | 'GGETAQSADPQWEQLNNK' | 'Heparin cofactor 2' |
| 6523 | 'SERPIND1_No6' | 'FPVEMTHNHNFR' | 'Heparin cofactor 2' |
| 6524 | 'SERPIND1_No7' | 'FTVDRPFLFLIYEHR' | 'Heparin cofactor 2' |
| 6525 | 'SERPIND1_No8' | 'GSWVNKFPVEMTHNHNFR' | 'Heparin cofactor 2' |
| 6526 | 'SERPIND1_No9' | 'NFGYTLR' | 'Heparin cofactor 2' |
| 6527 | 'SERPIND1_No10' | 'NYNLVESLK' | 'Heparin cofactor 2' |
| 6528 | 'SERPIND1_No11' | 'QFPILLDFK' | 'Heparin cofactor 2' |
| 6529 | 'SERPIND1_No12' | 'SVNDLYIQK' | 'Heparin cofactor 2' |
| 6530 | 'SERPIND1_No13' | 'GETHEQVHSILHFK' | 'Heparin cofactor 2' |
| 6531 | 'SERPIND1_No14' | 'VLKDQVNTFDNIFIAPVGISTAMGMISLGLK' | 'Heparin cofactor 2' |
| 6532 | 'SERPIND1_No15' | 'MLFDKNGNMAGISDQR' | 'Heparin cofactor 2' |
| 6533 | 'SERPIND1_No16' | 'TSCLLFMGR' | 'Heparin cofactor 2' |
| 6534 | 'SERPIND1_No17' | 'NGNMAGISDQR' | 'Heparin cofactor 2' |
| 6535 | 'SERPIND1_No18' | 'TSCLLFMGR' | 'Heparin cofactor 2' |
| 6536 | 'SERPIND1_No19' | 'FPVEMTHNHNFR' | 'Heparin cofactor 2' |
| 6537 | 'SERPIND1_No20' | 'GSWVNKFPVEMTHNHNFR' | 'Heparin cofactor 2' |
| 6538 | 'SERPIND1_No21' | 'RNFGYTLR' | 'Heparin cofactor 2' |
| 6539 | 'SERPIND1_No22' | 'NGNMAGISDQR' | 'Heparin cofactor 2' |
| 6540 | 'SERPIND1_No23' | 'TLEAQLTPR' | 'Heparin cofactor 2' |
| 6541 | 'SERPIND1_No24' | 'MLFDKNGNMAGISDQR' | 'Heparin cofactor 2' |
| 6542 | 'SERPIND1_No25' | 'HQGTITVNEEGTQATTVTTVGFMPLSTQVR' | 'Heparin cofactor 2' |
| 6543 | 'SERPIND1_No26' | 'QFPILLDFK' | 'Heparin cofactor 2' |
| 6544 | 'SERPIND1_No27' | 'QFPILLDFKTK' | 'Heparin cofactor 2' |
| 6545 | 'SERPIND1_No28' | 'MLFDKNGNMAGISDQR' | 'Heparin cofactor 2' |
| 6546 | 'SERPIND1_No29' | 'DALENIDPATQMMILNCIYFK' | 'Heparin cofactor 2' |
| 6547 | 'SERPIND1_No30' | 'VREYYFAEAQIADFSDPAFISK' | 'Heparin cofactor 2' |
| 6548 | 'SERPIND1_No31' | 'PFLFLIYEHR' | 'Heparin cofactor 2' |
| 6549 | 'SERPIND1_No32' | 'EVVKVSMMQTK' | 'Heparin cofactor 2' |
| 6550 | 'SERPIND1_No33' | 'YEITTIHNLFRK' | 'Heparin cofactor 2' |
| 6551 | 'SERPIND1_No34' | 'EYYFAEAQIADFSDPAFISK' | 'Heparin cofactor 2' |
| 6552 | 'SERPIND1_No35' | 'LNEREVVKVSMMQTK' | 'Heparin cofactor 2' |
| 6553 | 'SERPIND1_No36' | 'IQRLNILNAK' | 'Heparin cofactor 2' |
| 6554 | 'SERPIND1_No37' | 'LNEREVVKVSMMQTK' | 'Heparin cofactor 2' |
| 6555 | 'SERPIND1_No38' | 'VSMMQTK' | 'Heparin cofactor 2' |
| 6556 | 'SERPIND1_No39' | 'QFPILLDFKTK' | 'Heparin cofactor 2' |
| 6557 | 'SERPIND1_No40' | 'NYNLVESLKLMGIR' | 'Heparin cofactor 2' |

| | A | B | C |
|---|---|---|---|
| 1 | **Peptide Name** | **Peptide Sequence** | **Peptide Description** |
| 6558 | 'SERPIND1_No41' | 'FKLEKNYNLVESLK' | 'Heparin cofactor 2' |
| 6559 | 'SERPIND1_No42' | 'MLFDKNGNMAGISDQR' | 'Heparin cofactor 2' |
| 6560 | 'SERPIND1_No43' | 'SVNDLYIQKQFPILLDFKTK' | 'Heparin cofactor 2' |
| 6561 | 'SERPIND1_No44' | 'LEKNYNLVESLK' | 'Heparin cofactor 2' |
| 6562 | 'SERPIND1_No45' | 'SVNDLYIQKQFPILLDFK' | 'Heparin cofactor 2' |
| 6563 | 'SERPIND1_No46' | 'VLKDQVNTFDNIFIAPVGISTAMGMISLGLK' | 'Heparin cofactor 2' |
| 6564 | 'SERPIND1_No47' | 'HQGTITVNEEGTQATTVTTVGFMPLSTQVR' | 'Heparin cofactor 2' |
| 6565 | 'SERPIND1_No48' | 'NYNLVESLKLMGIR' | 'Heparin cofactor 2' |
| 6566 | 'ITGB1_No1' | 'LKPEDITQIQPQQLVLR' | 'Integrin beta-1' |
| 6567 | 'ITGB1_No2' | 'DKLPQPVQPDPVSHCK' | 'Integrin beta-1' |
| 6568 | 'ITGB1_No3' | 'HCECSTDEVNSEDMDAYCR' | 'Integrin beta-1' |
| 6569 | 'ITGB1_No4' | 'LLVFSTDAGFHFAGDGK' | 'Integrin beta-1' |
| 6570 | 'ITGB1_No5' | 'SLGTDLMNEMR' | 'Integrin beta-1' |
| 6571 | 'ITGB1_No6' | 'KGCPPDDIENPR' | 'Integrin beta-1' |
| 6572 | 'ITGB1_No7' | 'HCECSTDEVNSEDMDAYCR' | 'Integrin beta-1' |
| 6573 | 'ITGB1_No8' | 'NVLSLTNKGEVFNELVGK' | 'Integrin beta-1' |
| 6574 | 'ITGB1_No9' | 'FQGQTCEMCQTCLGVCAEHK' | 'Integrin beta-1' |
| 6575 | 'ITGB1_No10' | 'GCPPDDIENPR' | 'Integrin beta-1' |
| 6576 | 'ITGB1_No11' | 'DNTNEIYSGK' | 'Integrin beta-1' |
| 6577 | 'BCHE_No1' | 'YLTLNTESTR' | 'Cholinesterase' |
| 6578 | 'BCHE_No2' | 'IFFPGVSEFGK' | 'Cholinesterase' |
| 6579 | 'BCHE_No3' | 'SVTLFGESAGAASVSLHLLSPGSHSLFTR' | 'Cholinesterase' |
| 6580 | 'BCHE_No4' | 'TQILVGVNKDEGTAFLVYGAPGFSK' | 'Cholinesterase' |
| 6581 | 'BCHE_No5' | 'WNNYMMDWK' | 'Cholinesterase' |
| 6582 | 'BCHE_No6' | 'VLEMTGNIDEAEWEWK' | 'Cholinesterase' |
| 6583 | 'BCHE_No7' | 'EALGDVVGDYNFICPALEFTKK' | 'Cholinesterase' |
| 6584 | 'BCHE_No8' | 'ESILFHYTDWVDDQRPENYR' | 'Cholinesterase' |
| 6585 | 'BCHE_No9' | 'FSEWGNNAFFYYFEHR' | 'Cholinesterase' |
| 6586 | 'BCHE_No10' | 'VLEMTGNIDEAEWEWK' | 'Cholinesterase' |
| 6587 | 'BCHE_No11' | 'EALGDVVGDYNFICPALEFTK' | 'Cholinesterase' |
| 6588 | 'BCHE_No12' | 'DEGTAFLVYGAPGFSK' | 'Cholinesterase' |
| 6589 | 'BCHE_No13' | 'AILQSGSFNAPWAVTSLYEAR' | 'Cholinesterase' |
| 6590 | 'BCHE_No14' | 'FWTSFFPK' | 'Cholinesterase' |
| 6591 | 'BCHE_No15' | 'KTQILVGVNKDEGTAFLVYGAPGFSK' | 'Cholinesterase' |
| 6592 | 'BCHE_No16' | 'WNNYMMDWK' | 'Cholinesterase' |
| 6593 | 'BCHE_No17' | 'NQFNDYTSK' | 'Cholinesterase' |
| 6594 | 'BCHE_No18' | 'WNNYMMDWK' | 'Cholinesterase' |
| 6595 | 'BCHE_No19' | 'STEQKYLTLNTESTR' | 'Cholinesterase' |
| 6596 | 'IGKV2-30_No1' | 'DSGVPDRFSGSGSGTDFTLK' | 'Immunoglobulin kappa variable 2-30' |
| 6597 | 'IGKV2-30_No2' | 'SSQSLVYSDGNTYLNWFQQR' | 'Immunoglobulin kappa variable 2-30' |
| 6598 | 'IGKV2-30_No3' | 'SSQSLVYSDGNTYLNWFQQRPGQSPR' | 'Immunoglobulin kappa variable 2-30' |
| 6599 | 'IGKV2-30_No4' | 'VSNRDSGVPDRFSGSGSGTDFTLK' | 'Immunoglobulin kappa variable 2-30' |
| 6600 | 'IGKV2-30_No5' | 'VSNRDSGVPDR' | 'Immunoglobulin kappa variable 2-30' |
| 6601 | 'IGKV4-1_No1' | 'LLIYWASTR' | 'Immunoglobulin kappa variable 4-1' |

| | A | B | C |
|---|---|---|---|
| 1 | **Peptide Name** | **Peptide Sequence** | **Peptide Description** |
| 6602 | 'IGKV4-1_No2' | 'NYLAWYQQKPGQPPK' | 'Immunoglobulin kappa variable 4-1' |
| 6603 | 'IGKV4-1_No3' | 'SSQSVLYSSNNK' | 'Immunoglobulin kappa variable 4-1' |
| 6604 | 'IGKV4-1_No4' | 'SSQSVLYSSNNKNYLAWYQQKPGQPPK' | 'Immunoglobulin kappa variable 4-1' |
| 6605 | 'IGKV4-1_No5' | 'NYLAWYQQK' | 'Immunoglobulin kappa variable 4-1' |
| 6606 | 'IGKV4-1_No6' | 'PGQPPKLLIYWASTR' | 'Immunoglobulin kappa variable 4-1' |
| 6607 | 'IGKV4-1_No7' | 'ATINCKSSQSVLYSSNNK' | 'Immunoglobulin kappa variable 4-1' |
| 6608 | 'IGHV4-34_No1' | 'SRVTISVDTSK' | 'Immunoglobulin heavy variable 4-34' |
| 6609 | 'IGHV4-34_No2' | 'GLEWIGEINHSGSTNYNPSLK' | 'Immunoglobulin heavy variable 4-34' |
| 6610 | 'IGHV4-34_No3' | 'VTISVDTSKNQFSLK' | 'Immunoglobulin heavy variable 4-34' |
| 6611 | 'IGHV4-34_No4' | 'LSSVTAADTAVYYCAR' | 'Immunoglobulin heavy variable 4-34' |
| 6612 | 'IGHV4-34_No5' | 'VTISVDTSK' | 'Immunoglobulin heavy variable 4-34' |
| 6613 | 'IGHV4-34_No6' | 'SRVTISVDTSKNQFSLK' | 'Immunoglobulin heavy variable 4-34' |
| 6614 | 'GSN_No1' | 'AGALNSNDAFVLK' | 'Gelsolin' |
| 6615 | 'GSN_No2' | 'VPVDPATYGQFYGGDSYIILYNYR' | 'Gelsolin' |
| 6616 | 'GSN_No3' | 'HVVPNEVVVQR' | 'Gelsolin' |
| 6617 | 'GSN_No4' | 'AGKEPGLQIWR' | 'Gelsolin' |
| 6618 | 'GSN_No5' | 'SEDCFILDHGK' | 'Gelsolin' |
| 6619 | 'GSN_No6' | 'TGAQELLR' | 'Gelsolin' |
| 6620 | 'GSN_No7' | 'DSQEEEKTEALTSAK' | 'Gelsolin' |
| 6621 | 'GSN_No8' | 'PALPAGTEDTAK' | 'Gelsolin' |
| 6622 | 'GSN_No9' | 'NWRDPDQTDGLGLSYLSSHIANVER' | 'Gelsolin' |
| 6623 | 'GSN_No10' | 'SEDCFILDHGKDGK' | 'Gelsolin' |
| 6624 | 'GSN_No11' | 'EPAHLMSLFGGKPMIIYK' | 'Gelsolin' |
| 6625 | 'GSN_No12' | 'VPFDAATLHTSTAMAAQHGMDDDGTGQK' | 'Gelsolin' |
| 6626 | 'GSN_No13' | 'VPFDAATLHTSTAMAAQHGMDDDGTGQK' | 'Gelsolin' |
| 6627 | 'GSN_No14' | 'VPFDAATLHTSTAMAAQHGMDDDGTGQK' | 'Gelsolin' |
| 6628 | 'GSN_No15' | 'PNSMVVEHPEFLK' | 'Gelsolin' |
| 6629 | 'GSN_No16' | 'EVQGFESATFLGYFK' | 'Gelsolin' |
| 6630 | 'GSN_No17' | 'VPEARPNSMVVEHPEFLK' | 'Gelsolin' |
| 6631 | 'GSN_No18' | 'QTQVSVLPEGGETPLFK' | 'Gelsolin' |
| 6632 | 'GSN_No19' | 'DPDQTDGLGLSYLSSHIANVER' | 'Gelsolin' |
| 6633 | 'GSN_No20' | 'PNSMVVEHPEFLK' | 'Gelsolin' |
| 6634 | 'GSN_No21' | 'EPAHLMSLFGGKPMIIYK' | 'Gelsolin' |
| 6635 | 'GSN_No22' | 'YIETDPANRDR' | 'Gelsolin' |
| 6636 | 'GSN_No23' | 'PALPAGTEDTAKEDAANR' | 'Gelsolin' |
| 6637 | 'GSN_No24' | 'EPAHLMSLFGGK' | 'Gelsolin' |
| 6638 | 'GSN_No25' | 'VVQGKEPAHLMSLFGGKPMIIYK' | 'Gelsolin' |
| 6639 | 'GSN_No26' | 'QTQVSVLPEGGETPLFK' | 'Gelsolin' |
| 6640 | 'GSN_No27' | 'VHVSEEGTEPEAMLQVLGPKPALPAGTEDTAKEDAANR' | 'Gelsolin' |
| 6641 | 'GSN_No28' | 'EPAHLMSLFGGKPMIIYK' | 'Gelsolin' |
| 6642 | 'GSN_No29' | 'VHVSEEGTEPEAMLQVLGPKPALPAGTEDTAKEDAANR' | 'Gelsolin' |
| 6643 | 'GSN_No30' | 'VPFDAATLHTSTAMAAQHGMDDDGTGQK' | 'Gelsolin' |
| 6644 | 'GSN_No31' | 'RYIETDPANRDR' | 'Gelsolin' |
| 6645 | 'GSN_No32' | 'YIETDPANR' | 'Gelsolin' |

| | A | B | C |
|---|---|---|---|
| 1 | **Peptide Name** | **Peptide Sequence** | **Peptide Description** |
| 6646 | 'GSN_No33' | 'VPEARPNSMVVEHPEFLK' | 'Gelsolin' |
| 6647 | 'GSN_No34' | 'RYIETDPANR' | 'Gelsolin' |
| 6648 | 'GSN_No35' | 'TPSAAYLWVGTGASEAEKTGAQELLR' | 'Gelsolin' |
| 6649 | 'GSN_No36' | 'YIETDPANRDRR' | 'Gelsolin' |
| 6650 | 'GSN_No37' | 'VHVSEEGTEPEAMLQVLGPK' | 'Gelsolin' |
| 6651 | 'GSN_No38' | 'EPAHLMSLFGGKPMIIYK' | 'Gelsolin' |
| 6652 | 'GSN_No39' | 'PALPAGTEDTAKEDAANRK' | 'Gelsolin' |
| 6653 | 'GSN_No40' | 'VHVSEEGTEPEAMLQVLGPK' | 'Gelsolin' |
| 6654 | 'GSN_No41' | 'VSNGAGTMSVSLVADENPFAQGALK' | 'Gelsolin' |
| 6655 | 'GSN_No42' | 'EPGLQIWR' | 'Gelsolin' |
| 6656 | 'GSN_No43' | 'TPSAAYLWVGTGASEAEK' | 'Gelsolin' |
| 6657 | 'GSN_No44' | 'DSQEEEKTEALTSAKR' | 'Gelsolin' |
| 6658 | 'GSN_No45' | 'AQPVQVAEGSEPDGFWEALGGK' | 'Gelsolin' |
| 6659 | 'GSN_No46' | 'GGVASGFKHVVPNEVVVQR' | 'Gelsolin' |
| 6660 | 'GSN_No47' | 'VSNGAGTMSVSLVADENPFAQGALKSEDCFILDHGK' | 'Gelsolin' |
| 6661 | 'GSN_No48' | 'VSNGAGTMSVSLVADENPFAQGALKSEDCFILDHGKDGK' | 'Gelsolin' |
| 6662 | 'GSN_No49' | 'TASDFITK' | 'Gelsolin' |
| 6663 | 'GSN_No50' | 'QFFKNWRDPDQTDGLGLSYLSSHIANVER' | 'Gelsolin' |
| 6664 | 'GSN_No51' | 'QTQVSVLPEGGETPLFKQFFK' | 'Gelsolin' |
| 6665 | 'GSN_No52' | 'VHVSEEGTEPEAMLQVLGPKPALPAGTEDTAK' | 'Gelsolin' |
| 6666 | 'GSN_No53' | 'VHVSEEGTEPEAMLQVLGPKPALPAGTEDTAK' | 'Gelsolin' |
| 6667 | 'GSN_No54' | 'GASQAGAPQGRVPEARPNSMVVEHPEFLK' | 'Gelsolin' |
| 6668 | 'GSN_No55' | 'AGALNSNDAFVLKTPSAAYLWVGTGASEAEKTGAQELLR' | 'Gelsolin' |
| 6669 | 'GSN_No56' | 'VPFDAATLHTSTAMAAQHGMDDDGTGQKQIWR' | 'Gelsolin' |
| 6670 | 'GSN_No57' | 'VEKFDLVPVPTNLYGDFFTGDAYVILK' | 'Gelsolin' |
| 6671 | 'GSN_No58' | 'VVQGKEPAHLMSLFGGKPMIIYK' | 'Gelsolin' |
| 6672 | 'GSN_No59' | 'VVQGKEPAHLMSLFGGKPMIIYK' | 'Gelsolin' |
| 6673 | 'GSN_No60' | 'AALKTASDFITK' | 'Gelsolin' |
| 6674 | 'GSN_No61' | 'VSNGAGTMSVSLVADENPFAQGALK' | 'Gelsolin' |
| 6675 | 'GSN_No62' | 'LYKVSNGAGTMSVSLVADENPFAQGALK' | 'Gelsolin' |
| 6676 | 'GSN_No63' | 'VSNGAGTMSVSLVADENPFAQGALKSEDCFILDHGKDGK' | 'Gelsolin' |
| 6677 | 'GSN_No64' | 'SEDCFILDHGKDGKIFVWK' | 'Gelsolin' |
| 6678 | 'GSN_No65' | 'MDYPKQTQVSVLPEGGETPLFK' | 'Gelsolin' |
| 6679 | 'GSN_No66' | 'LFACSNKIGR' | 'Gelsolin' |
| 6680 | 'GSN_No67' | 'QTQVSVLPEGGETPLFKQFFK' | 'Gelsolin' |
| 6681 | 'GSN_No68' | 'QFFKNWRDPDQTDGLGLSYLSSHIANVER' | 'Gelsolin' |
| 6682 | 'GSN_No69' | 'IEGSNKVPVDPATYGQFYGGDSYIILYNYR' | 'Gelsolin' |
| 6683 | 'GSN_No70' | 'GASQAGAPQGRVPEARPNSMVVEHPEFLK' | 'Gelsolin' |
| 6684 | 'GSN_No71' | 'TASDFITKMDYPK' | 'Gelsolin' |
| 6685 | 'GSN_No72' | 'EGGQTAPASTR' | 'Gelsolin' |
| 6686 | 'GSN_No73' | 'EPAHLMSLFGGK' | 'Gelsolin' |
| 6687 | 'GSN_No74' | 'GASQAGAPQGR' | 'Gelsolin' |
| 6688 | 'GSN_No75' | 'MDAHPPR' | 'Gelsolin' |
| 6689 | 'GSN_No76' | 'AGKEPGLQIWRVEK' | 'Gelsolin' |

| | A | B | C |
|---|---|---|---|
| 1 | **Peptide Name** | **Peptide Sequence** | **Peptide Description** |
| 6690 | 'GSN_No77' | 'VVQGKEPAHLMSLFGGK' | 'Gelsolin' |
| 6691 | 'GSN_No78' | 'VVQGKEPAHLMSLFGGK' | 'Gelsolin' |
| 6692 | 'GSN_No79' | 'EVQGFESATFLGYFKSGLK' | 'Gelsolin' |
| 6693 | 'GSN_No80' | 'VSNGAGTMSVSLVADENPFAQGALKSEDCFILDHGK' | 'Gelsolin' |
| 6694 | 'GSN_No81' | 'HVVPNEVVVQRLFQVKGR' | 'Gelsolin' |
| 6695 | 'GSN_No82' | 'TASDFITKMDYPK' | 'Gelsolin' |
| 6696 | 'GSN_No83' | 'LFACSNK' | 'Gelsolin' |
| 6697 | 'GSN_No84' | 'VPFDAATLHTSTAMAAQHGMDDDGTGQKQIWR' | 'Gelsolin' |
| 6698 | 'GSN_No85' | 'KGGVASGFKHVVPNEVVVQR' | 'Gelsolin' |
| 6699 | 'GSN_No86' | 'MDAHPPRLFACSNK' | 'Gelsolin' |
| 6700 | 'GSN_No87' | 'KGGVASGFK' | 'Gelsolin' |
| 6701 | 'GSN_No88' | 'MDYPKQTQVSVLPEGGETPLFKQFFK' | 'Gelsolin' |
| 6702 | 'GSN_No89' | 'LKATQVSK' | 'Gelsolin' |
| 6703 | 'GSN_No90' | 'AVQHREVQGFESATFLGYFK' | 'Gelsolin' |
| 6704 | 'GSN_No91' | 'KAALKTASDFITK' | 'Gelsolin' |
| 6705 | 'ATP5F1B_No1' | 'VVDLLAPYAK' | 'ATP synthase subunit beta, mitochondrial' |
| 6706 | 'ATP5F1B_No2' | 'FTQAGSEVSALLGR' | 'ATP synthase subunit beta, mitochondrial' |
| 6707 | 'ATP5F1B_No3' | 'VLDSGAPIKIPVGPETLGR' | 'ATP synthase subunit beta, mitochondrial' |
| 6708 | 'ATP5F1B_No4' | 'LVLEVAQHLGESTVR' | 'ATP synthase subunit beta, mitochondrial' |
| 6709 | 'ATP5F1B_No5' | 'FLSQPFQVAEVFTGHMGK' | 'ATP synthase subunit beta, mitochondrial' |
| 6710 | 'ATP5F1B_No6' | 'VALTGLTVAEYFR' | 'ATP synthase subunit beta, mitochondrial' |
| 6711 | 'ATP5F1B_No7' | 'FLSQPFQVAEVFTGHMGK' | 'ATP synthase subunit beta, mitochondrial' |
| 6712 | 'ATP5F1B_No8' | 'IPSAVGYQPTLATDMGTMQER' | 'ATP synthase subunit beta, mitochondrial' |
| 6713 | 'ATP5F1B_No9' | 'AIAELGIYPAVDPLDSTSR' | 'ATP synthase subunit beta, mitochondrial' |
| 6714 | 'ATP5F1B_No10' | 'IPSAVGYQPTLATDMGTMQER' | 'ATP synthase subunit beta, mitochondrial' |
| 6715 | 'ATP5F1B_No11' | 'TIAMDGTEGLVR' | 'ATP synthase subunit beta, mitochondrial' |
| 6716 | 'C2_No1' | 'DFHINLFR' | 'Complement C2' |
| 6717 | 'C2_No2' | 'AVISPGFDVFAK' | 'Complement C2' |
| 6718 | 'C2_No3' | 'HAIILLTDGK' | 'Complement C2' |
| 6719 | 'C2_No4' | 'RNDYLDIYAIGVGK' | 'Complement C2' |
| 6720 | 'C2_No5' | 'PICLPCTMEANLALR' | 'Complement C2' |
| 6721 | 'C2_No6' | 'EVVTDQFLCSGTQEDESPCK' | 'Complement C2' |
| 6722 | 'C2_No7' | 'ALHQVFEHMLDVSK' | 'Complement C2' |
| 6723 | 'C2_No8' | 'VLMSVLNDNSR' | 'Complement C2' |
| 6724 | 'C2_No9' | 'LLGMETMAWQEIR' | 'Complement C2' |
| 6725 | 'C2_No10' | 'SSGQWQTPGATR' | 'Complement C2' |
| 6726 | 'C2_No11' | 'TPWHVTIKPK' | 'Complement C2' |
| 6727 | 'C2_No12' | 'QHLGDVLNFLPL' | 'Complement C2' |
| 6728 | 'C2_No13' | 'LLGMETMAWQEIR' | 'Complement C2' |
| 6729 | 'C2_No14' | 'GALISDQWVLTAAHCFR' | 'Complement C2' |
| 6730 | 'C2_No15' | 'LLGMETMAWQEIR' | 'Complement C2' |
| 6731 | 'C2_No16' | 'SKVPPPRDFHINLFR' | 'Complement C2' |
| 6732 | 'C2_No17' | 'CPAPVSFENGIYTPR' | 'Complement C2' |
| 6733 | 'C2_No18' | 'ECQGNGVWSGTEPICR' | 'Complement C2' |

| | A | B | C |
|---|---|---|---|
| 1 | **Peptide Name** | **Peptide Sequence** | **Peptide Description** |
| 6734 | 'C2_No19' | 'VLMSVLNDNSR' | 'Complement C2' |
| 6735 | 'C2_No20' | 'SQWGKEFLIEK' | 'Complement C2' |
| 6736 | 'C2_No21' | 'QPYSYDFPEDVAPALGTSFSHMLGATNPTQK' | 'Complement C2' |
| 6737 | 'C2_No22' | 'LLGMETMAWQEIR' | 'Complement C2' |
| 6738 | 'C2_No23' | 'ALHQVFEHMLDVSK' | 'Complement C2' |
| 6739 | 'C2_No24' | 'NDYLDIYAIGVGK' | 'Complement C2' |
| 6740 | 'C2_No25' | 'KNQGILEFYGDDIALLK' | 'Complement C2' |
| 6741 | 'C2_No26' | 'PICLPCTMEANLALR' | 'Complement C2' |
| 6742 | 'C2_No27' | 'QCRPNGMWDGETAVCDNGAGHCPNPGISLGAVR' | 'Complement C2' |
| 6743 | 'C2_No28' | 'LDVDWRELNELGSK' | 'Complement C2' |
| 6744 | 'C2_No29' | 'VPPPRDFHINLFR' | 'Complement C2' |
| 6745 | 'C2_No30' | 'CSSNLVLTGSSER' | 'Complement C2' |
| 6746 | 'C2_No31' | 'QHLGDVLNFLPL' | 'Complement C2' |
| 6747 | 'C2_No32' | 'AVISPGFDVFAKK' | 'Complement C2' |
| 6748 | 'C2_No33' | 'RPQGSTCRDHENELLNK' | 'Complement C2' |
| 6749 | 'C2_No34' | 'MGVEWTSCAEVVSQEK' | 'Complement C2' |
| 6750 | 'C2_No35' | 'EILNINQK' | 'Complement C2' |
| 6751 | 'C2_No36' | 'ESASLMVDR' | 'Complement C2' |
| 6752 | 'C2_No37' | 'GALISDQWVLTAAHCFRDGNDHSLWR' | 'Complement C2' |
| 6753 | 'C2_No38' | 'DHENELLNK' | 'Complement C2' |
| 6754 | 'C2_No39' | 'NQGILEFYGDDIALLK' | 'Complement C2' |
| 6755 | 'C2_No40' | 'QPYSYDFPEDVAPALGTSFSHMLGATNPTQK' | 'Complement C2' |
| 6756 | 'C2_No41' | 'MGVEWTSCAEVVSQEK' | 'Complement C2' |
| 6757 | 'C2_No42' | 'LDVDWRELNELGSKK' | 'Complement C2' |
| 6758 | 'C2_No43' | 'HAIILLTDGKSNMGGSPK' | 'Complement C2' |
| 6759 | 'C2_No44' | 'DMTEVISSLENANYK' | 'Complement C2' |
| 6760 | 'C2_No45' | 'DMTEVISSLENANYK' | 'Complement C2' |
| 6761 | 'C2_No46' | 'TAVDHIREILNINQKR' | 'Complement C2' |
| 6762 | 'C2_No47' | 'TAVDHIREILNINQK' | 'Complement C2' |
| 6763 | 'C2_No48' | 'TAVDHIR' | 'Complement C2' |
| 6764 | 'C2_No49' | 'GESGGAVFLER' | 'Complement C2' |
| 6765 | 'C2_No50' | 'MSTHARPICLPCTMEANLALR' | 'Complement C2' |
| 6766 | 'C2_No51' | 'ESASLMVDR' | 'Complement C2' |
| 6767 | 'C2_No52' | 'EVVTDQFLCSGTQEDESPCKGESGGAVFLER' | 'Complement C2' |
| 6768 | 'C2_No53' | 'HAIILLTDGKSNMGGSPK' | 'Complement C2' |
| 6769 | 'C2_No54' | 'QPYSYDFPEDVAPALGTSFSHMLGATNPTQK' | 'Complement C2' |
| 6770 | 'C2_No55' | 'QPYSYDFPEDVAPALGTSFSHMLGATNPTQK' | 'Complement C2' |
| 6771 | 'C2_No56' | 'AVCKPVR' | 'Complement C2' |
| 6772 | 'C2_No57' | 'CSSNLVLTGSSERECQGNGVWSGTEPICR' | 'Complement C2' |
| 6773 | 'S100A9_No1' | 'LGHPDTLNQGEFK' | 'Protein S100-A9' |
| 6774 | 'S100A9_No2' | 'NIETIINTFHQYSVK' | 'Protein S100-A9' |
| 6775 | 'S100A9_No3' | 'LGHPDTLNQGEFKELVR' | 'Protein S100-A9' |
| 6776 | 'S100A9_No4' | 'LGHPDTLNQGEFKELVRK' | 'Protein S100-A9' |
| 6777 | 'S100A9_No5' | 'VIEHIMEDLDTNADK' | 'Protein S100-A9' |

| | A | B | C |
|---|---|---|---|
| 1 | **Peptide Name** | **Peptide Sequence** | **Peptide Description** |
| 6778 | 'S100A9_No6' | 'VIEHIMEDLDTNADK' | 'Protein S100-A9' |
| 6779 | 'APOA4_No1' | 'SELTQQLNALFQDK' | 'Apolipoprotein A-IV' |
| 6780 | 'APOA4_No2' | 'LGPHAGDVEGHLSFLEK' | 'Apolipoprotein A-IV' |
| 6781 | 'APOA4_No3' | 'LNHQLEGLTFQMK' | 'Apolipoprotein A-IV' |
| 6782 | 'APOA4_No4' | 'SLAELGGHLDQQVEEFR' | 'Apolipoprotein A-IV' |
| 6783 | 'APOA4_No5' | 'LGEVNTYAGDLQK' | 'Apolipoprotein A-IV' |
| 6784 | 'APOA4_No6' | 'SLAPYAQDTQEK' | 'Apolipoprotein A-IV' |
| 6785 | 'APOA4_No7' | 'IDQTVEELRR' | 'Apolipoprotein A-IV' |
| 6786 | 'APOA4_No8' | 'KLVPFATELHER' | 'Apolipoprotein A-IV' |
| 6787 | 'APOA4_No9' | 'ENADSLQASLRPHADELK' | 'Apolipoprotein A-IV' |
| 6788 | 'APOA4_No10' | 'RVEPYGENFNK' | 'Apolipoprotein A-IV' |
| 6789 | 'APOA4_No11' | 'IDQNVEELKGR' | 'Apolipoprotein A-IV' |
| 6790 | 'APOA4_No12' | 'LLPHANEVSQK' | 'Apolipoprotein A-IV' |
| 6791 | 'APOA4_No13' | 'IDQNVEELK' | 'Apolipoprotein A-IV' |
| 6792 | 'APOA4_No14' | 'ALVQQMEQLR' | 'Apolipoprotein A-IV' |
| 6793 | 'APOA4_No15' | 'LTPYADEFKVK' | 'Apolipoprotein A-IV' |
| 6794 | 'APOA4_No16' | 'ISASAEELR' | 'Apolipoprotein A-IV' |
| 6795 | 'APOA4_No17' | 'VNSFFSTFK' | 'Apolipoprotein A-IV' |
| 6796 | 'APOA4_No18' | 'ALVQQMEQLR' | 'Apolipoprotein A-IV' |
| 6797 | 'APOA4_No19' | 'SLAELGGHLDQQVEEFRR' | 'Apolipoprotein A-IV' |
| 6798 | 'APOA4_No20' | 'LKEEIGKELEELR' | 'Apolipoprotein A-IV' |
| 6799 | 'APOA4_No21' | 'LNHQLEGLTFQMK' | 'Apolipoprotein A-IV' |
| 6800 | 'APOA4_No22' | 'ISASAEELRQR' | 'Apolipoprotein A-IV' |
| 6801 | 'APOA4_No23' | 'AKIDQNVEELKGR' | 'Apolipoprotein A-IV' |
| 6802 | 'APOA4_No24' | 'LVPFATELHER' | 'Apolipoprotein A-IV' |
| 6803 | 'APOA4_No25' | 'VKIDQTVEELRR' | 'Apolipoprotein A-IV' |
| 6804 | 'APOA4_No26' | 'TQVNTQAEQLR' | 'Apolipoprotein A-IV' |
| 6805 | 'APOA4_No27' | 'QLTPYAQR' | 'Apolipoprotein A-IV' |
| 6806 | 'APOA4_No28' | 'TQVNTQAEQLRR' | 'Apolipoprotein A-IV' |
| 6807 | 'APOA4_No29' | 'SLAPYAQDTQEKLNHQLEGLTFQMK' | 'Apolipoprotein A-IV' |
| 6808 | 'APOA4_No30' | 'VEPYGENFNK' | 'Apolipoprotein A-IV' |
| 6809 | 'APOA4_No31' | 'IGDNLRELQQR' | 'Apolipoprotein A-IV' |
| 6810 | 'APOA4_No32' | 'VKIDQTVEELR' | 'Apolipoprotein A-IV' |
| 6811 | 'APOA4_No33' | 'RRVEPYGENFNK' | 'Apolipoprotein A-IV' |
| 6812 | 'APOA4_No34' | 'AKIDQNVEELK' | 'Apolipoprotein A-IV' |
| 6813 | 'APOA4_No35' | 'LAPLAEDVRGNLR' | 'Apolipoprotein A-IV' |
| 6814 | 'APOA4_No36' | 'LGPHAGDVEGHLSFLEKDLR' | 'Apolipoprotein A-IV' |
| 6815 | 'APOA4_No37' | 'SLAPYAQDTQEKLNHQLEGLTFQMK' | 'Apolipoprotein A-IV' |
| 6816 | 'APOA4_No38' | 'LGPHAGDVEGHLSFLEKDLRDK' | 'Apolipoprotein A-IV' |
| 6817 | 'APOA4_No39' | 'SELTQQLNALFQDKLGEVNTYAGDLQK' | 'Apolipoprotein A-IV' |
| 6818 | 'APOA4_No40' | 'VLRENADSLQASLRPHADELK' | 'Apolipoprotein A-IV' |
| 6819 | 'APOA4_No41' | 'LEPYADQLR' | 'Apolipoprotein A-IV' |
| 6820 | 'APOA4_No42' | 'DKVNSFFSTFKEK' | 'Apolipoprotein A-IV' |
| 6821 | 'APOA4_No43' | 'LAPLAEDVRGNLRGNTEGLQK' | 'Apolipoprotein A-IV' |

| | A | B | C |
|---|---|---|---|
| 1 | **Peptide Name** | **Peptide Sequence** | **Peptide Description** |
| 6822 | 'APOA4_No44' | 'EEIGKELEELR' | 'Apolipoprotein A-IV' |
| 6823 | 'APOA4_No45' | 'DLRDKVNSFFSTFK' | 'Apolipoprotein A-IV' |
| 6824 | 'APOA4_No46' | 'LTPYADEFK' | 'Apolipoprotein A-IV' |
| 6825 | 'APOA4_No47' | 'DKVNSFFSTFK' | 'Apolipoprotein A-IV' |
| 6826 | 'APOA4_No48' | 'QKLGPHAGDVEGHLSFLEKDLR' | 'Apolipoprotein A-IV' |
| 6827 | 'APOA4_No49' | 'QKLGPHAGDVEGHLSFLEK' | 'Apolipoprotein A-IV' |
| 6828 | 'APOA4_No50' | 'RSLAPYAQDTQEK' | 'Apolipoprotein A-IV' |
| 6829 | 'APOA4_No51' | 'IGDNLRELQQRLEPYADQLR' | 'Apolipoprotein A-IV' |
| 6830 | 'APOA4_No52' | 'SLAELGGHLDQQVEEFRRR' | 'Apolipoprotein A-IV' |
| 6831 | 'APOA4_No53' | 'SLAPYAQDTQEKLNHQLEGLTFQMKK' | 'Apolipoprotein A-IV' |
| 6832 | 'APOA4_No54' | 'ENADSLQASLRPHADELKAK' | 'Apolipoprotein A-IV' |
| 6833 | 'APOA4_No55' | 'VNSFFSTFKEK' | 'Apolipoprotein A-IV' |
| 6834 | 'APOA4_No56' | 'ALVQQMEQLRQK' | 'Apolipoprotein A-IV' |
| 6835 | 'APOA4_No57' | 'SELTQQLNALFQDKLGEVNTYAGDLQKK' | 'Apolipoprotein A-IV' |
| 6836 | 'APOA4_No58' | 'LEPYADQLRTQVNTQAEQLRR' | 'Apolipoprotein A-IV' |
| 6837 | 'APOA4_No59' | 'EAVEHLQK' | 'Apolipoprotein A-IV' |
| 6838 | 'APOA4_No60' | 'SLAPYAQDTQEKLNHQLEGLTFQMKK' | 'Apolipoprotein A-IV' |
| 6839 | 'APOA4_No61' | 'RVEPYGENFNKALVQQMEQLR' | 'Apolipoprotein A-IV' |
| 6840 | 'APOA4_No62' | 'QKLGPHAGDVEGHLSFLEK' | 'Apolipoprotein A-IV' |
| 6841 | 'APOA4_No63' | 'LLPHANEVSQKIGDNLR' | 'Apolipoprotein A-IV' |
| 6842 | 'APOA4_No64' | 'GRLTPYADEFKVK' | 'Apolipoprotein A-IV' |
| 6843 | 'APOA4_No65' | 'RQLTPYAQR' | 'Apolipoprotein A-IV' |
| 6844 | 'APOA4_No66' | 'EAVEHLQKSELTQQLNALFQDK' | 'Apolipoprotein A-IV' |
| 6845 | 'APOA4_No67' | 'LNHQLEGLTFQMKK' | 'Apolipoprotein A-IV' |
| 6846 | 'APOA4_No68' | 'ELQQRLEPYADQLR' | 'Apolipoprotein A-IV' |
| 6847 | 'APOA4_No69' | 'EEIGKELEELRAR' | 'Apolipoprotein A-IV' |
| 6848 | 'APOA4_No70' | 'IDQTVEELR' | 'Apolipoprotein A-IV' |
| 6849 | 'APOA4_No71' | 'VNSFFSTFKEKESQDK' | 'Apolipoprotein A-IV' |
| 6850 | 'APOA4_No72' | 'LAPLAEDVR' | 'Apolipoprotein A-IV' |
| 6851 | 'APOA4_No73' | 'TLSLPELEQQQEQQQEQQQEQVQMLAPLES' | 'Apolipoprotein A-IV' |
| 6852 | 'APOA4_No74' | 'VEPYGENFNKALVQQMEQLR' | 'Apolipoprotein A-IV' |
| 6853 | 'APOA4_No75' | 'EAVEHLQKSELTQQLNALFQDKLGEVNTYAGDLQK' | 'Apolipoprotein A-IV' |
| 6854 | 'APOA4_No76' | 'QKLGPHAGDVEGHLSFLEKDLR' | 'Apolipoprotein A-IV' |
| 6855 | 'APOA4_No77' | 'LGEVNTYAGDLQKK' | 'Apolipoprotein A-IV' |
| 6856 | 'APOA4_No78' | 'LTPYADEFKVKIDQTVEELR' | 'Apolipoprotein A-IV' |
| 6857 | 'APOA4_No79' | 'ARISASAEELRQR' | 'Apolipoprotein A-IV' |
| 6858 | 'APOA4_No80' | 'ALVQQMEQLRQK' | 'Apolipoprotein A-IV' |
| 6859 | 'APOA4_No81' | 'GRLTPYADEFK' | 'Apolipoprotein A-IV' |
| 6860 | 'APOA4_No82' | 'RSLAPYAQDTQEKLNHQLEGLTFQMK' | 'Apolipoprotein A-IV' |
| 6861 | 'APOA4_No83' | 'EKESQDKTLSLPELEQQQEQQQEQQQEQVQMLAPLES' | 'Apolipoprotein A-IV' |
| 6862 | 'APOA4_No84' | 'VEPYGENFNKALVQQMEQLRQK' | 'Apolipoprotein A-IV' |
| 6863 | 'APOA4_No85' | 'QRLAPLAEDVR' | 'Apolipoprotein A-IV' |
| 6864 | 'APOA4_No86' | 'LNHQLEGLTFQMKK' | 'Apolipoprotein A-IV' |
| 6865 | 'APOA4_No87' | 'RVEPYGENFNKALVQQMEQLR' | 'Apolipoprotein A-IV' |

| | A | B | C |
|---|---|---|---|
| 1 | **Peptide Name** | **Peptide Sequence** | **Peptide Description** |
| 6866 | 'APOA4_No88' | 'TLSLPELEQQQEQQQEQQQEQVQMLAPLES' | 'Apolipoprotein A-IV' |
| 6867 | 'APOA4_No89' | 'EKESQDKTLSLPELEQQQEQQQEQQQEQVQMLAPLES' | 'Apolipoprotein A-IV' |
| 6868 | 'APOA4_No90' | 'NAEELKAR' | 'Apolipoprotein A-IV' |
| 6869 | 'CKM_No1' | 'LNYKPEEEYPDLSK' | 'Creatine kinase M-type' |
| 6870 | 'CKM_No2' | 'LGSSEVEQVQLVVDGVK' | 'Creatine kinase M-type' |
| 6871 | 'CKM_No3' | 'LSVEALNSLTGEFK' | 'Creatine kinase M-type' |
| 6872 | 'CKM_No4' | 'GTGGVDTAAVGSVFDVSNADR' | 'Creatine kinase M-type' |
| 6873 | 'CKM_No5' | 'SFLVWVNEEDHLR' | 'Creatine kinase M-type' |
| 6874 | 'ENO1_No1' | 'DATNVGDEGGFAPNILENKEGLELLK' | 'Alpha-enolase' |
| 6875 | 'ENO1_No2' | 'AAVPSGASTGIYEALELR' | 'Alpha-enolase' |
| 6876 | 'ENO1_No3' | 'YISPDQLADLYK' | 'Alpha-enolase' |
| 6877 | 'ENO1_No4' | 'IGAEVYHNLK' | 'Alpha-enolase' |
| 6878 | 'ENO1_No5' | 'DATNVGDEGGFAPNILENK' | 'Alpha-enolase' |
| 6879 | 'ENO1_No6' | 'SGKYDLDFKSPDDPSR' | 'Alpha-enolase' |
| 6880 | 'ENO1_No7' | 'HIADLAGNSEVILPVPAFNVINGGSHAGNK' | 'Alpha-enolase' |
| 6881 | 'ENO1_No8' | 'AGYTDKVVIGMDVAASEFFR' | 'Alpha-enolase' |
| 6882 | 'ENO1_No9' | 'LAQANGWGVMVSHR' | 'Alpha-enolase' |
| 6883 | 'ENO1_No10' | 'FTASAGIQVVGDDLTVTNPKR' | 'Alpha-enolase' |
| 6884 | 'ENO1_No11' | 'FGANAILGVSLAVCK' | 'Alpha-enolase' |
| 6885 | 'ENO1_No12' | 'GNPTVEVDLFTSK' | 'Alpha-enolase' |
| 6886 | 'ENO1_No13' | 'VVIGMDVAASEFFR' | 'Alpha-enolase' |
| 6887 | 'ENO1_No14' | 'YDLDFKSPDDPSR' | 'Alpha-enolase' |
| 6888 | 'ENO1_No15' | 'VVIGMDVAASEFFR' | 'Alpha-enolase' |
| 6889 | 'ENO1_No16' | 'LAQANGWGVMVSHR' | 'Alpha-enolase' |
| 6890 | 'ENO1_No17' | 'LAMQEFMILPVGAANFR' | 'Alpha-enolase' |
| 6891 | 'ENO1_No18' | 'LAMQEFMILPVGAANFR' | 'Alpha-enolase' |
| 6892 | 'ENO1_No19' | 'IDKLMIEMDGTENK' | 'Alpha-enolase' |
| 6893 | 'PYGL_No1' | 'LHSFLGDDVFLR' | 'Glycogen phosphorylase, liver form' |
| 6894 | 'PYGL_No2' | 'WPVDLVEK' | 'Glycogen phosphorylase, liver form' |
| 6895 | 'PYGL_No3' | 'GIVGVENVAELKK' | 'Glycogen phosphorylase, liver form' |
| 6896 | 'PYGL_No4' | 'IFVDIEKLPWSK' | 'Glycogen phosphorylase, liver form' |
| 6897 | 'GPI_No1' | 'TLAQLNPESSLFIIASK' | 'Glucose-6-phosphate isomerase' |
| 6898 | 'GPI_No2' | 'VFEGNRPTNSIVFTK' | 'Glucose-6-phosphate isomerase' |
| 6899 | 'GPI_No3' | 'VWYVSNIDGTHIAK' | 'Glucose-6-phosphate isomerase' |
| 6900 | 'GPI_No4' | 'TFTTQETITNAETAKEWFLQAAK' | 'Glucose-6-phosphate isomerase' |
| 6901 | 'GPI_No5' | 'SNTPILVDGKDVMPEVNK' | 'Glucose-6-phosphate isomerase' |
| 6902 | 'GPI_No6' | 'TFTTQETITNAETAK' | 'Glucose-6-phosphate isomerase' |
| 6903 | 'GPI_No7' | 'KIEPELDGSAQVTSHDASTNGLINFIK' | 'Glucose-6-phosphate isomerase' |
| 6904 | 'TPM3_No1' | 'KLVIIEGDLERTEER' | 'Tropomyosin alpha-3 chain' |
| 6905 | 'TPM3_No2' | 'AADAEAEVASLNRR' | 'Tropomyosin alpha-3 chain' |
| 6906 | 'TPM3_No3' | 'ILTDKLKEAETR' | 'Tropomyosin alpha-3 chain' |
| 6907 | 'TPM3_No4' | 'ALKDEEKMELQEIQLK' | 'Tropomyosin alpha-3 chain' |
| 6908 | 'TPM3_No5' | 'KLVIIEGDLER' | 'Tropomyosin alpha-3 chain' |
| 6909 | 'TPM3_No6' | 'YSQKEDKYEEEIK' | 'Tropomyosin alpha-3 chain' |

| | A | B | C |
|---|---|---|---|
| 1 | **Peptide Name** | **Peptide Sequence** | **Peptide Description** |
| 6910 | 'TPM3_No7' | 'ALKDEEKMELQEIQLK' | 'Tropomyosin alpha-3 chain' |
| 6911 | 'FABP1' | 'AIGLPEELIQK' | 'Fatty acid-binding protein, liver' |
| 6912 | 'LDHB_No1' | 'FIIPQIVK' | 'L-lactate dehydrogenase B chain' |
| 6913 | 'LDHB_No2' | 'IVVVTAGVR' | 'L-lactate dehydrogenase B chain' |
| 6914 | 'LDHB_No3' | 'IVADKDYSVTANSK' | 'L-lactate dehydrogenase B chain' |
| 6915 | 'LDHB_No4' | 'LKDDEVAQLKK' | 'L-lactate dehydrogenase B chain' |
| 6916 | 'LDHB_No5' | 'LIAPVAEEEATVPNNK' | 'L-lactate dehydrogenase B chain' |
| 6917 | 'LDHB_No6' | 'EKLIAPVAEEEATVPNNK' | 'L-lactate dehydrogenase B chain' |
| 6918 | 'LDHB_No7' | 'SADTLWDIQKDLKDL' | 'L-lactate dehydrogenase B chain' |
| 6919 | 'LDHB_No8' | 'GEMMDLQHGSLFLQTPK' | 'L-lactate dehydrogenase B chain' |
| 6920 | 'LDHB_No9' | 'MVVESAYEVIK' | 'L-lactate dehydrogenase B chain' |
| 6921 | 'LDHB_No10' | 'ITVVGVGQVGMACAISILGK' | 'L-lactate dehydrogenase B chain' |
| 6922 | 'LDHB_No11' | 'SADTLWDIQK' | 'L-lactate dehydrogenase B chain' |
| 6923 | 'LDHB_No12' | 'SLADELALVDVLEDKLK' | 'L-lactate dehydrogenase B chain' |
| 6924 | 'GPX1_No1' | 'EALPAPSDDATALMTDPK' | 'Glutathione peroxidase 1' |
| 6925 | 'GPX1_No2' | 'YVRPGGGFEPNFMLFEK' | 'Glutathione peroxidase 1' |
| 6926 | 'GPX1_No3' | 'YVRPGGGFEPNFMLFEK' | 'Glutathione peroxidase 1' |
| 6927 | 'GPX1_No4' | 'PLAGGEPVSLGSLR' | 'Glutathione peroxidase 1' |
| 6928 | 'GPX1_No5' | 'EALPAPSDDATALMTDPK' | 'Glutathione peroxidase 1' |
| 6929 | 'GPX1_No6' | 'GLVVLGFPCNQFGHQENAK' | 'Glutathione peroxidase 1' |
| 6930 | 'GPX1_No7' | 'DYTQMNELQR' | 'Glutathione peroxidase 1' |
| 6931 | 'GPX1_No8' | 'CEVNGAGAHPLFAFLR' | 'Glutathione peroxidase 1' |
| 6932 | 'PROS1_No1' | 'FSAEFDFR' | 'Vitamin K-dependent protein S' |
| 6933 | 'PROS1_No2' | 'KVESELIKPINPR' | 'Vitamin K-dependent protein S' |
| 6934 | 'PROS1_No3' | 'FRLPEISR' | 'Vitamin K-dependent protein S' |
| 6935 | 'PROS1_No4' | 'IETISHEDLQR' | 'Vitamin K-dependent protein S' |
| 6936 | 'PROS1_No5' | 'VYFAGFPR' | 'Vitamin K-dependent protein S' |
| 6937 | 'PROS1_No6' | 'IQALSLCSDQQSHLEFR' | 'Vitamin K-dependent protein S' |
| 6938 | 'PROS1_No7' | 'EAVMDINKPGPLFKPENGLLETK' | 'Vitamin K-dependent protein S' |
| 6939 | 'PROS1_No8' | 'QSTNAYPDLR' | 'Vitamin K-dependent protein S' |
| 6940 | 'PROS1_No9' | 'DCKDVDECSLKPSICGTAVCK' | 'Vitamin K-dependent protein S' |
| 6941 | 'PROS1_No10' | 'EAVMDINKPGPLFKPENGLLETK' | 'Vitamin K-dependent protein S' |
| 6942 | 'PROS1_No11' | 'QGASGIKEIIQEK' | 'Vitamin K-dependent protein S' |
| 6943 | 'PROS1_No12' | 'ASFTCTCKPGWQGEK' | 'Vitamin K-dependent protein S' |
| 6944 | 'PROS1_No13' | 'SCEVVSVCLPLNLDTK' | 'Vitamin K-dependent protein S' |
| 6945 | 'PROS1_No14' | 'PSICGTAVCK' | 'Vitamin K-dependent protein S' |
| 6946 | 'PROS1_No15' | 'VESELIKPINPR' | 'Vitamin K-dependent protein S' |
| 6947 | 'PROS1_No16' | 'NIPGDFECECPEGYRYNLK' | 'Vitamin K-dependent protein S' |
| 6948 | 'PROS1_No17' | 'ITTGGDVINNGLWNMVSVEELEHSISIK' | 'Vitamin K-dependent protein S' |
| 6949 | 'PROS1_No18' | 'SQDILLSVENTVIYR' | 'Vitamin K-dependent protein S' |
| 6950 | 'PROS1_No19' | 'NNLELSTPLKIETISHEDLQR' | 'Vitamin K-dependent protein S' |
| 6951 | 'PROS1_No20' | 'VNRNNLELSTPLKIETISHEDLQR' | 'Vitamin K-dependent protein S' |
| 6952 | 'PROS1_No21' | 'QGASGIKEIIQEK' | 'Vitamin K-dependent protein S' |
| 6953 | 'PROS1_No22' | 'SCVNAIPDQCSPLPCNEDGYMSCK' | 'Vitamin K-dependent protein S' |

| | A | B | C |
|---|---|---|---|
| 1 | **Peptide Name** | **Peptide Sequence** | **Peptide Description** |
| 6954 | 'PROS1_No23' | 'QLAVLDKAMK' | 'Vitamin K-dependent protein S' |
| 6955 | 'PROS1_No24' | 'IEVQLKNEHTSK' | 'Vitamin K-dependent protein S' |
| 6956 | 'PROS1_No25' | 'SCVNAIPDQCSPLPCNEDGYMSCK' | 'Vitamin K-dependent protein S' |
| 6957 | 'PROS1_No26' | 'IQALSLCSDQQSHLEFRVNR' | 'Vitamin K-dependent protein S' |
| 6958 | 'PROS1_No27' | 'VNRNNLELSTPLK' | 'Vitamin K-dependent protein S' |
| 6959 | 'PROS1_No28' | 'SFQTGLFTAAR' | 'Vitamin K-dependent protein S' |
| 6960 | 'PROS1_No29' | 'QGASGIKEIIQEKQNK' | 'Vitamin K-dependent protein S' |
| 6961 | 'PROS1_No30' | 'ITTGGDVINNGLWNMVSVEELEHSISIK' | 'Vitamin K-dependent protein S' |
| 6962 | 'PROS1_No31' | 'SCVNAIPDQCSPLPCNEDGYMSCKDGK' | 'Vitamin K-dependent protein S' |
| 6963 | 'PROS1_No32' | 'QSTNAYPDLR' | 'Vitamin K-dependent protein S' |
| 6964 | 'PROS1_No33' | 'AHSCPSVWK' | 'Vitamin K-dependent protein S' |
| 6965 | 'PROS1_No34' | 'DVDECSLKPSICGTAVCK' | 'Vitamin K-dependent protein S' |
| 6966 | 'PROS1_No35' | 'QGASGIKEIIQEKQNK' | 'Vitamin K-dependent protein S' |
| 6967 | 'PROS1_No36' | 'SCVNAIPDQCSPLPCNEDGYMSCKDGK' | 'Vitamin K-dependent protein S' |
| 6968 | 'PROS1_No37' | 'GGKIEVQLKNEHTSK' | 'Vitamin K-dependent protein S' |
| 6969 | 'PROS1_No38' | 'NGFVMLSNK' | 'Vitamin K-dependent protein S' |
| 6970 | 'PROS1_No39' | 'PGPLFKPENGLLETK' | 'Vitamin K-dependent protein S' |
| 6971 | 'P4HB_No1' | 'THILLFLPK' | 'Protein disulfide-isomerase' |
| 6972 | 'P4HB_No2' | 'VDATEESDLAQQYGVR' | 'Protein disulfide-isomerase' |
| 6973 | 'P4HB_No3' | 'ILEFFGLK' | 'Protein disulfide-isomerase' |
| 6974 | 'P4HB_No4' | 'MDSTANEVEAVK' | 'Protein disulfide-isomerase' |
| 6975 | 'P4HB_No5' | 'LGETYKDHENIVIAK' | 'Protein disulfide-isomerase' |
| 6976 | 'P4HB_No6' | 'NNFEGEVTKENLLDFIK' | 'Protein disulfide-isomerase' |
| 6977 | 'P4HB_No7' | 'ENLLDFIK' | 'Protein disulfide-isomerase' |
| 6978 | 'P4HB_No8' | 'NFEDVAFDEKK' | 'Protein disulfide-isomerase' |
| 6979 | 'P4HB_No9' | 'LAKVDATEESDLAQQYGVR' | 'Protein disulfide-isomerase' |
| 6980 | 'P4HB_No10' | 'HNQLPLVIEFTEQTAPK' | 'Protein disulfide-isomerase' |
| 6981 | 'P4HB_No11' | 'YQLDKDGVVLFK' | 'Protein disulfide-isomerase' |
| 6982 | 'P4HB_No12' | 'IKPHLMSQELPEDWDKQPVK' | 'Protein disulfide-isomerase' |
| 6983 | 'P4HB_No13' | 'QLAPIWDKLGETYKDHENIVIAK' | 'Protein disulfide-isomerase' |
| 6984 | 'P4HB_No14' | 'EADDIVNWLK' | 'Protein disulfide-isomerase' |
| 6985 | 'P4HB_No15' | 'SVSDYDGKLSNFK' | 'Protein disulfide-isomerase' |
| 6986 | 'P4HB_No16' | 'QLAPIWDKLGETYKDHENIVIAK' | 'Protein disulfide-isomerase' |
| 6987 | 'P4HB_No17' | 'YQLDKDGVVLFKK' | 'Protein disulfide-isomerase' |
| 6988 | 'ASGR2_No1' | 'FIVQHTNPFNTWIGLTDSDGSWK' | 'Asialoglycoprotein receptor 2' |
| 6989 | 'ASGR2_No2' | 'ADHDALLFHLK' | 'Asialoglycoprotein receptor 2' |
| 6990 | 'ASGR2_No3' | 'TCCPVNWVEHQGSCYWFSHSGK' | 'Asialoglycoprotein receptor 2' |
| 6991 | 'ASGR2_No4' | 'NWAVTQPDNWHGHELGGSEDCVEVQPDGR' | 'Asialoglycoprotein receptor 2' |
| 6992 | 'CSF1R_No1' | 'ALTFELTLR' | 'Macrophage colony-stimulating factor 1 receptor' |
| 6993 | 'CSF1R_No2' | 'FIQSQDYQCSALMGGR' | 'Macrophage colony-stimulating factor 1 receptor' |
| 6994 | 'CSF1R_No3' | 'HTFTLSLPR' | 'Macrophage colony-stimulating factor 1 receptor' |
| 6995 | 'CSF1R_No4' | 'VIPGPPALTLVPAELVR' | 'Macrophage colony-stimulating factor 1 receptor' |
| 6996 | 'CSF1R_No5' | 'LAIPQQSDFHNNR' | 'Macrophage colony-stimulating factor 1 receptor' |
| 6997 | 'CSF1R_No6' | 'CTEPGDPLGGSAAIHLYVK' | 'Macrophage colony-stimulating factor 1 receptor' |

| | A | B | C |
|---|---|---|---|
| 1 | **Peptide Name** | **Peptide Sequence** | **Peptide Description** |
| 6998 | 'CSF1R_No7' | 'FIQSQDYQCSALMGGR' | 'Macrophage colony-stimulating factor 1 receptor' |
| 6999 | 'CSF1R_No8' | 'CDEAQVLQVWDDPYPEVLSQEPFHK' | 'Macrophage colony-stimulating factor 1 receptor' |
| 7000 | 'CTSD_No1' | 'FDGILGMAYPR' | 'Cathepsin D' |
| 7001 | 'CTSD_No2' | 'AIGAVPLIQGEYMIPCEK' | 'Cathepsin D' |
| 7002 | 'CTSD_No3' | 'LLDIACWIHHK' | 'Cathepsin D' |
| 7003 | 'CTSD_No4' | 'YSQAVPAVTEGPIPEVLK' | 'Cathepsin D' |
| 7004 | 'CTSD_No5' | 'DPDAQPGGELMLGGTDSK' | 'Cathepsin D' |
| 7005 | 'CTSD_No6' | 'AIGAVPLIQGEYMIPCEK' | 'Cathepsin D' |
| 7006 | 'CTSD_No7' | 'DPDAQPGGELMLGGTDSK' | 'Cathepsin D' |
| 7007 | 'CTSD_No8' | 'ISVNNVLPVFDNLMQQK' | 'Cathepsin D' |
| 7008 | 'CTSD_No9' | 'EGCEAIVDTGTSLMVGPVDEVR' | 'Cathepsin D' |
| 7009 | 'CTSD_No10' | 'YYTVFDRDNNR' | 'Cathepsin D' |
| 7010 | 'CTSD_No11' | 'FDGILGMAYPR' | 'Cathepsin D' |
| 7011 | 'CTSD_No12' | 'TMSEVGGSVEDLIAK' | 'Cathepsin D' |
| 7012 | 'CTSD_No13' | 'ISVNNVLPVFDNLMQQK' | 'Cathepsin D' |
| 7013 | 'CTSD_No14' | 'QVFGEATKQPGITFIAAK' | 'Cathepsin D' |
| 7014 | 'CTSD_No15' | 'EGCEAIVDTGTSLMVGPVDEVR' | 'Cathepsin D' |
| 7015 | 'ANXA2_No1' | 'GVDEVTIVNILTNR' | 'Annexin A2' |
| 7016 | 'ANXA2_No2' | 'TNQELQEINR' | 'Annexin A2' |
| 7017 | 'ANXA2_No3' | 'TDLEKDIISDTSGDFRK' | 'Annexin A2' |
| 7018 | 'ANXA2_No4' | 'TPAQYDASELK' | 'Annexin A2' |
| 7019 | 'ANXA2_No5' | 'AYTNFDAERDALNIETAIK' | 'Annexin A2' |
| 7020 | 'ANXA2_No6' | 'SYSPYDMLESIRK' | 'Annexin A2' |
| 7021 | 'ANXA2_No7' | 'SALSGHLETVILGLLK' | 'Annexin A2' |
| 7022 | 'ANXA2_No8' | 'SYSPYDMLESIR' | 'Annexin A2' |
| 7023 | 'ANXA2_No9' | 'QDIAFAYQR' | 'Annexin A2' |
| 7024 | 'C8A_No1' | 'YHFEALADTGISSEFYDNANDLLSK' | 'Complement component C8 alpha chain' |
| 7025 | 'C8A_No2' | 'AIDEDCSQYEPIPGSQK' | 'Complement component C8 alpha chain' |
| 7026 | 'C8A_No3' | 'CGPCFNNGVPILEGTSCR' | 'Complement component C8 alpha chain' |
| 7027 | 'C8A_No4' | 'KAMAVEDIISR' | 'Complement component C8 alpha chain' |
| 7028 | 'C8A_No5' | 'ALDQYLMEFNACR' | 'Complement component C8 alpha chain' |
| 7029 | 'C8A_No6' | 'LYYGDDEKYFR' | 'Complement component C8 alpha chain' |
| 7030 | 'C8A_No7' | 'AMAVEDIISR' | 'Complement component C8 alpha chain' |
| 7031 | 'C8A_No8' | 'YNPVVIDFEMQPIHEVLR' | 'Complement component C8 alpha chain' |
| 7032 | 'C8A_No9' | 'FINDYGTHYITSGSMGGIYEYILVIDKAK' | 'Complement component C8 alpha chain' |
| 7033 | 'C8A_No10' | 'DITTCFGGSLGIQYEDKINVGGGLSGDHCKK' | 'Complement component C8 alpha chain' |
| 7034 | 'C8A_No11' | 'HLVCNGDQDCLDGSDEDDCEDVR' | 'Complement component C8 alpha chain' |
| 7035 | 'C8A_No12' | 'INVGGGLSGDHCK' | 'Complement component C8 alpha chain' |
| 7036 | 'C8A_No13' | 'HTSLGPLEAK' | 'Complement component C8 alpha chain' |
| 7037 | 'C8A_No14' | 'ECDNPAPQNGGASCPGR' | 'Complement component C8 alpha chain' |
| 7038 | 'C8A_No15' | 'DITTCFGGSLGIQYEDKINVGGGLSGDHCK' | 'Complement component C8 alpha chain' |
| 7039 | 'C8A_No16' | 'YNPVVIDFEMQPIHEVLR' | 'Complement component C8 alpha chain' |
| 7040 | 'C8A_No17' | 'ALDQYLMEFNACR' | 'Complement component C8 alpha chain' |
| 7041 | 'C8A_No18' | 'SLKYNPVVIDFEMQPIHEVLR' | 'Complement component C8 alpha chain' |

| | A | B | C |
|---|---|---|---|
| 1 | **Peptide Name** | **Peptide Sequence** | **Peptide Description** |
| 7042 | 'C8A_No19' | 'RHLVCNGDQDCLDGSDEDDCEDVR' | 'Complement component C8 alpha chain' |
| 7043 | 'C8A_No20' | 'RECDNPAPQNGGASCPGR' | 'Complement component C8 alpha chain' |
| 7044 | 'C8A_No21' | 'LGSLGAACEQTQTEGAK' | 'Complement component C8 alpha chain' |
| 7045 | 'C8A_No22' | 'MESLGITSR' | 'Complement component C8 alpha chain' |
| 7046 | 'C8A_No23' | 'ECDNPAPQNGGASCPGRK' | 'Complement component C8 alpha chain' |
| 7047 | 'C8A_No24' | 'FGGTICSGDIWDQASCSSSTTCVR' | 'Complement component C8 alpha chain' |
| 7048 | 'C8A_No25' | 'RRECDNPAPQNGGASCPGR' | 'Complement component C8 alpha chain' |
| 7049 | 'C8A_No26' | 'KAMAVEDIISR' | 'Complement component C8 alpha chain' |
| 7050 | 'C8A_No27' | 'RALDQYLMEFNACR' | 'Complement component C8 alpha chain' |
| 7051 | 'C8A_No28' | 'AALGYNILTQEDAQSVYDASYYGGQCETVYNGEWR' | 'Complement component C8 alpha chain' |
| 7052 | 'C8A_No29' | 'QAQCGQDFQCK' | 'Complement component C8 alpha chain' |
| 7053 | 'C8A_No30' | 'QAQCGQDFQCKETGR' | 'Complement component C8 alpha chain' |
| 7054 | 'C8A_No31' | 'KPYNFLK' | 'Complement component C8 alpha chain' |
| 7055 | 'C8A_No32' | 'AMAVEDIISR' | 'Complement component C8 alpha chain' |
| 7056 | 'C8A_No33' | 'QAQCGQDFQCKETGR' | 'Complement component C8 alpha chain' |
| 7057 | 'C8A_No34' | 'INVGGGLSGDHCKK' | 'Complement component C8 alpha chain' |
| 7058 | 'C8A_No35' | 'MESLGITSR' | 'Complement component C8 alpha chain' |
| 7059 | 'C8A_No36' | 'SLLQPNKFGGTICSGDIWDQASCSSSTTCVR' | 'Complement component C8 alpha chain' |
| 7060 | 'C8A_No37' | 'IFTKVQTAHFK' | 'Complement component C8 alpha chain' |
| 7061 | 'C8A_No38' | 'SLKYNPVVIDFEMQPIHEVLR' | 'Complement component C8 alpha chain' |
| 7062 | 'C8A_No39' | 'FINDYGTHYITSGSMGGIYEYILVIDKAK' | 'Complement component C8 alpha chain' |
| 7063 | 'C8A_No40' | 'HTSLGPLEAKR' | 'Complement component C8 alpha chain' |
| 7064 | 'C8A_No41' | 'DITTCFGGSLGIQYEDK' | 'Complement component C8 alpha chain' |
| 7065 | 'C8A_No42' | 'QAQCGQDFQCK' | 'Complement component C8 alpha chain' |
| 7066 | 'C8A_No43' | 'YDSTCER' | 'Complement component C8 alpha chain' |
| 7067 | 'C8A_No44' | 'RALDQYLMEFNACR' | 'Complement component C8 alpha chain' |
| 7068 | 'C8A_No45' | 'VQTAHFK' | 'Complement component C8 alpha chain' |
| 7069 | 'C8B_No1' | 'GILNEIKDR' | 'Complement component C8 beta chain' |
| 7070 | 'C8B_No2' | 'LPLEYSYGEYR' | 'Complement component C8 beta chain' |
| 7071 | 'C8B_No3' | 'GDYTLNNVHACAK' | 'Complement component C8 beta chain' |
| 7072 | 'C8B_No4' | 'KPYNVESYTPQTQGK' | 'Complement component C8 beta chain' |
| 7073 | 'C8B_No5' | 'SGFSFGFK' | 'Complement component C8 beta chain' |
| 7074 | 'C8B_No6' | 'EVSSCHCAPCQGNGVPVLK' | 'Complement component C8 beta chain' |
| 7075 | 'C8B_No7' | 'SLMLHYEFLQR' | 'Complement component C8 beta chain' |
| 7076 | 'C8B_No8' | 'CDCICPVGSQGLACEVSYR' | 'Complement component C8 beta chain' |
| 7077 | 'C8B_No9' | 'RLPLEYSYGEYR' | 'Complement component C8 beta chain' |
| 7078 | 'C8B_No10' | 'LLCNGDNDCGDQSDEANCR' | 'Complement component C8 beta chain' |
| 7079 | 'C8B_No11' | 'YAYLLQPSQFHGEPCNFSDK' | 'Complement component C8 beta chain' |
| 7080 | 'C8B_No12' | 'DTMVEDLVVLVR' | 'Complement component C8 beta chain' |
| 7081 | 'C8B_No13' | 'YYAGGCSPHYILNTR' | 'Complement component C8 beta chain' |
| 7082 | 'C8B_No14' | 'LLCNGDNDCGDQSDEANCRR' | 'Complement component C8 beta chain' |
| 7083 | 'C8B_No15' | 'QCNNPPPQNGGSPCSGPASETLDCS' | 'Complement component C8 beta chain' |
| 7084 | 'C8B_No16' | 'CDCICPVGSQGLACEVSYRK' | 'Complement component C8 beta chain' |
| 7085 | 'C8B_No17' | 'RLLCNGDNDCGDQSDEANCRR' | 'Complement component C8 beta chain' |

| | A | B | C |
|---|---|---|---|
| 1 | **Peptide Name** | **Peptide Sequence** | **Peptide Description** |
| 7086 | 'C8B_No18' | 'RLLCNGDNDCGDQSDEANCR' | 'Complement component C8 beta chain' |
| 7087 | 'C8B_No19' | 'QALEEFQK' | 'Complement component C8 beta chain' |
| 7088 | 'C8B_No20' | 'SLMLHYEFLQR' | 'Complement component C8 beta chain' |
| 7089 | 'C8B_No21' | 'QALEEFQKEVSSCHCAPCQGNGVPVLK' | 'Complement component C8 beta chain' |
| 7090 | 'C8B_No22' | 'IGGAIEEVYVSLGVSVGK' | 'Complement component C8 beta chain' |
| 7091 | 'C8B_No23' | 'SDLEVAHYK' | 'Complement component C8 beta chain' |
| 7092 | 'C8B_No24' | 'IPGIFELGISSQSDR' | 'Complement component C8 beta chain' |
| 7093 | 'C8B_No25' | 'YAYLLQPSQFHGEPCNFSDKEVEDCVTNRPCR' | 'Complement component C8 beta chain' |
| 7094 | 'C8B_No26' | 'DTMVEDLVVLVR' | 'Complement component C8 beta chain' |
| 7095 | 'C8B_No27' | 'LPLEYSYGEYRDLFR' | 'Complement component C8 beta chain' |
| 7096 | 'C8B_No28' | 'VKVEPLYELVTATDFAYSSTVR' | 'Complement component C8 beta chain' |
| 7097 | 'C8B_No29' | 'RDTMVEDLVVLVR' | 'Complement component C8 beta chain' |
| 7098 | 'C8B_No30' | 'CEGFVCAQTGR' | 'Complement component C8 beta chain' |
| 7099 | 'C8B_No31' | 'QALEEFQKEVSSCHCAPCQGNGVPVLK' | 'Complement component C8 beta chain' |
| 7100 | 'C8B_No32' | 'FRKPYNVESYTPQTQGK' | 'Complement component C8 beta chain' |
| 7101 | 'C8B_No33' | 'SVFLHAR' | 'Complement component C8 beta chain' |
| 7102 | 'C8B_No34' | 'RLPLEYSYGEYRDLFR' | 'Complement component C8 beta chain' |
| 7103 | 'C8B_No35' | 'EVEDCVTNRPCR' | 'Complement component C8 beta chain' |
| 7104 | 'C8B_No36' | 'QRQCNNPPPQNGGSPCSGPASETLDCS' | 'Complement component C8 beta chain' |
| 7105 | 'C8B_No37' | 'GSRCDCICPVGSQGLACEVSYR' | 'Complement component C8 beta chain' |
| 7106 | 'C8B_No38' | 'NKRDTMVEDLVVLVR' | 'Complement component C8 beta chain' |
| 7107 | 'C8B_No39' | 'EYESYSDFER' | 'Complement component C8 beta chain' |
| 7108 | 'C8B_No40' | 'GSRCDCICPVGSQGLACEVSYRK' | 'Complement component C8 beta chain' |
| 7109 | 'C8B_No41' | 'EAMERGDYTLNNVHACAK' | 'Complement component C8 beta chain' |
| 7110 | 'C8B_No42' | 'IPGIFELGISSQSDRGK' | 'Complement component C8 beta chain' |
| 7111 | 'C8B_No43' | 'RDTMVEDLVVLVR' | 'Complement component C8 beta chain' |
| 7112 | 'C8B_No44' | 'QCNNPPPQNGGSPCSGPASETLDCS' | 'Complement component C8 beta chain' |
| 7113 | 'C8B_No45' | 'NKRDTMVEDLVVLVR' | 'Complement component C8 beta chain' |
| 7114 | 'C8B_No46' | 'VKRLPLEYSYGEYR' | 'Complement component C8 beta chain' |
| 7115 | 'C8B_No47' | 'GGASEHITTLAYQELPTADLMQEWGDAVQYNPAIIK' | 'Complement component C8 beta chain' |
| 7116 | 'C8B_No48' | 'SDLEVAHYKLKPR' | 'Complement component C8 beta chain' |
| 7117 | 'C8B_No49' | 'EAMERGDYTLNNVHACAK' | 'Complement component C8 beta chain' |
| 7118 | 'C8B_No50' | 'KPYNVESYTPQTQGKYEFILK' | 'Complement component C8 beta chain' |
| 7119 | 'C8B_No51' | 'PYNVESYTPQTQGK' | 'Complement component C8 beta chain' |
| 7120 | 'GP1BA_No1' | 'LTSLPLGALR' | 'Platelet glycoprotein Ib alpha chain' |
| 7121 | 'GP1BA_No2' | 'GLGELQELYLK' | 'Platelet glycoprotein Ib alpha chain' |
| 7122 | 'GP1BA_No3' | 'LTQLNLDR' | 'Platelet glycoprotein Ib alpha chain' |
| 7123 | 'GP1BA_No4' | 'AWLLFLR' | 'Platelet glycoprotein Ib alpha chain' |
| 7124 | 'GP1BA_No5' | 'LTQLNLDRCELTK' | 'Platelet glycoprotein Ib alpha chain' |
| 7125 | 'GP1BA_No6' | 'GLGELQELYLKGNELK' | 'Platelet glycoprotein Ib alpha chain' |
| 7126 | 'GP1BA_No7' | 'GVLQGHLESSR' | 'Platelet glycoprotein Ib alpha chain' |
| 7127 | 'GP1BA_No8' | 'AMTSNVASVQCDNSDKFPVYK' | 'Platelet glycoprotein Ib alpha chain' |
| 7128 | 'GP1BA_No9' | 'WLQDNAENVYVWK' | 'Platelet glycoprotein Ib alpha chain' |
| 7129 | 'GP1BA_No10' | 'AMTSNVASVQCDNSDKFPVYK' | 'Platelet glycoprotein Ib alpha chain' |

| | A | B | C |
|---|---|---|---|
| 1 | **Peptide Name** | **Peptide Sequence** | **Peptide Description** |
| 7130 | 'GP1BA_No11' | 'AMTSNVASVQCDNSDKFPVYKYPGK' | 'Platelet glycoprotein Ib alpha chain' |
| 7131 | 'C8G_No1' | 'AEATTLHVAPQGTAMAVSTFR' | 'Complement component C8 gamma chain' |
| 7132 | 'C8G_No2' | 'RPASPISTIQPK' | 'Complement component C8 gamma chain' |
| 7133 | 'C8G_No3' | 'KLDGICWQVR' | 'Complement component C8 gamma chain' |
| 7134 | 'C8G_No4' | 'YGFCEAADQFHVLDEVR' | 'Complement component C8 gamma chain' |
| 7135 | 'C8G_No5' | 'YGFCEAADQFHVLDEVRR' | 'Complement component C8 gamma chain' |
| 7136 | 'C8G_No6' | 'VQEAHLTEDQIFYFPK' | 'Complement component C8 gamma chain' |
| 7137 | 'C8G_No7' | 'QLYGDTGVLGR' | 'Complement component C8 gamma chain' |
| 7138 | 'C8G_No8' | 'SLPVSDSVLSGFEQR' | 'Complement component C8 gamma chain' |
| 7139 | 'C8G_No9' | 'FLQEQGHRAEATTLHVAPQGTAMAVSTFR' | 'Complement component C8 gamma chain' |
| 7140 | 'C8G_No10' | 'QLYGDTGVLGR' | 'Complement component C8 gamma chain' |
| 7141 | 'C8G_No11' | 'AEATTLHVAPQGTAMAVSTFR' | 'Complement component C8 gamma chain' |
| 7142 | 'C8G_No12' | 'ANFDAQQFAGTWLLVAVGSACR' | 'Complement component C8 gamma chain' |
| 7143 | 'C8G_No13' | 'FLQEQGHR' | 'Complement component C8 gamma chain' |
| 7144 | 'C8G_No14' | 'FLQEQGHRAEATTLHVAPQGTAMAVSTFR' | 'Complement component C8 gamma chain' |
| 7145 | 'C8G_No15' | 'AEATTLHVAPQGTAMAVSTFRK' | 'Complement component C8 gamma chain' |
| 7146 | 'C8G_No16' | 'AEATTLHVAPQGTAMAVSTFRK' | 'Complement component C8 gamma chain' |
| 7147 | 'CAPN1_No1' | 'LVFVHSAEGNEFWSALLEK' | 'Calpain-1 catalytic subunit' |
| 7148 | 'CAPN1_No2' | 'KLYELIITR' | 'Calpain-1 catalytic subunit' |
| 7149 | 'CAPN1_No3' | 'LEICNLTPDALK' | 'Calpain-1 catalytic subunit' |
| 7150 | 'CAPN1_No4' | 'NYLSIFR' | 'Calpain-1 catalytic subunit' |
| 7151 | 'CAPN1_No5' | 'KAPSDLYQIILK' | 'Calpain-1 catalytic subunit' |
| 7152 | 'CAPN1_No6' | 'APSDLYQIILK' | 'Calpain-1 catalytic subunit' |
| 7153 | 'CAPN1_No7' | 'NYPATFWVNPQFK' | 'Calpain-1 catalytic subunit' |
| 7154 | 'CAPN1_No8' | 'SEEIITPVYCTGVSAQVQK' | 'Calpain-1 catalytic subunit' |
| 7155 | 'CAPN1_No9' | 'LYELIITR' | 'Calpain-1 catalytic subunit' |
| 7156 | 'CAPN1_No10' | 'LPPGEYVVVPSTFEPNKEGDFVLR' | 'Calpain-1 catalytic subunit' |
| 7157 | 'CAPN1_No11' | 'DFFLANASR' | 'Calpain-1 catalytic subunit' |
| 7158 | 'CAPN1_No12' | 'YLGQDYEQLR' | 'Calpain-1 catalytic subunit' |
| 7159 | 'TUBB_No1' | 'ISVYYNEATGGK' | 'Tubulin beta chain' |
| 7160 | 'TUBB_No2' | 'MAVTFIGNSTAIQELFKR' | 'Tubulin beta chain' |
| 7161 | 'TUBB_No3' | 'FWEVISDEHGIDPTGTYHGDSDLQLDR' | 'Tubulin beta chain' |
| 7162 | 'TUBB_No4' | 'ALTVPELTQQVFDAK' | 'Tubulin beta chain' |
| 7163 | 'TUBB_No5' | 'MAVTFIGNSTAIQELFK' | 'Tubulin beta chain' |
| 7164 | 'TUBB_No6' | 'MAVTFIGNSTAIQELFK' | 'Tubulin beta chain' |
| 7165 | 'CA3_No1' | 'FDPSCLFPACR' | 'Carbonic anhydrase 3' |
| 7166 | 'CA3_No2' | 'IGHENGEFQIFLDALDK' | 'Carbonic anhydrase 3' |
| 7167 | 'CA3_No3' | 'YAAELHLVHWNPK' | 'Carbonic anhydrase 3' |
| 7168 | 'PRSS1_No1' | 'LGEHNIEVLEGNEQFINAAK' | 'Trypsin-1' |
| 7169 | 'PRSS1_No2' | 'VSTISLPTAPPATGTK' | 'Trypsin-1' |
| 7170 | 'PRSS1_No3' | 'TLNNDIMLIK' | 'Trypsin-1' |
| 7171 | 'PRSS2' | 'TLDNDILLIK' | 'Trypsin-2' |
| 7172 | 'PSAP' | 'IGACPSAHKPLLGTEK' | 'Prosaposin' |
| 7173 | 'HEXB_No1' | 'PGPALWPLPLSVK' | 'Beta-hexosaminidase subunit beta' |

| | A | B | C |
|---|---|---|---|
| 1 | **Peptide Name** | **Peptide Sequence** | **Peptide Description** |
| 7174 | 'HEXB_No2' | 'VLPEFDTPGHTLSWGK' | 'Beta-hexosaminidase subunit beta' |
| 7175 | 'HEXB_No3' | 'VLDIIATINK' | 'Beta-hexosaminidase subunit beta' |
| 7176 | 'HEXB_No4' | 'GSIVWQEVFDDKAK' | 'Beta-hexosaminidase subunit beta' |
| 7177 | 'PFN1_No1' | 'TLVLLMGK' | 'Profilin-1' |
| 7178 | 'PFN1_No2' | 'STGGAPTFNVTVTK' | 'Profilin-1' |
| 7179 | 'PFN1_No3' | 'TFVNITPAEVGVLVGK' | 'Profilin-1' |
| 7180 | 'PFN1_No4' | 'DSLLQDGEFSMDLR' | 'Profilin-1' |
| 7181 | 'PFN1_No5' | 'DSLLQDGEFSMDLR' | 'Profilin-1' |
| 7182 | 'PFN1_No6' | 'TDKTLVLLMGK' | 'Profilin-1' |
| 7183 | 'PFN1_No7' | 'TFVNITPAEVGVLVGKDR' | 'Profilin-1' |
| 7184 | 'PFN1_No8' | 'SSFYVNGLTLGGQK' | 'Profilin-1' |
| 7185 | 'PFN1_No9' | 'TDKTLVLLMGK' | 'Profilin-1' |
| 7186 | 'PFN1_No10' | 'TKSTGGAPTFNVTVTK' | 'Profilin-1' |
| 7187 | 'PFN1_No11' | 'CSVIRDSLLQDGEFSMDLR' | 'Profilin-1' |
| 7188 | 'BPGM_No1' | 'VCDVPLDQLPR' | 'Bisphosphoglycerate mutase' |
| 7189 | 'BPGM_No2' | 'YKVCDVPLDQLPR' | 'Bisphosphoglycerate mutase' |
| 7190 | 'BPGM_No3' | 'SESLKDVLER' | 'Bisphosphoglycerate mutase' |
| 7191 | 'BPGM_No4' | 'EQMALNHGEEQVR' | 'Bisphosphoglycerate mutase' |
| 7192 | 'BPGM_No5' | 'HYGALIGLNR' | 'Bisphosphoglycerate mutase' |
| 7193 | 'APRT' | 'DISPVLKDPASFR' | 'Adenine phosphoribosyltransferase' |
| 7194 | 'CTSB_No1' | 'VMFTEDLKLPASFDAR' | 'Cathepsin B' |
| 7195 | 'CTSB_No2' | 'GQDHCGIESEVVAGIPR' | 'Cathepsin B' |
| 7196 | 'CTSB_No3' | 'DQGSCGSCWAFGAVEAISDR' | 'Cathepsin B' |
| 7197 | 'CTSB_No4' | 'SGVYQHVTGEMMGGHAIR' | 'Cathepsin B' |
| 7198 | 'CTSB_No5' | 'ICEPGYSPTYKQDK' | 'Cathepsin B' |
| 7199 | 'CTSB_No6' | 'ICEPGYSPTYK' | 'Cathepsin B' |
| 7200 | 'CTSB_No7' | 'TDQYWEKI' | 'Cathepsin B' |
| 7201 | 'CTSB_No8' | 'VMFTEDLKLPASFDAR' | 'Cathepsin B' |
| 7202 | 'HSP90AA1_No1' | 'HLEINPDHSIIETLR' | 'Heat shock protein HSP 90-alpha' |
| 7203 | 'HSP90AA1_No2' | 'KHLEINPDHSIIETLR' | 'Heat shock protein HSP 90-alpha' |
| 7204 | 'HSP90AA1_No3' | 'RAPFDLFENR' | 'Heat shock protein HSP 90-alpha' |
| 7205 | 'HSP90AA1_No4' | 'DQVANSAFVER' | 'Heat shock protein HSP 90-alpha' |
| 7206 | 'HSP90AA1_No5' | 'ELISNSSDALDKIR' | 'Heat shock protein HSP 90-alpha' |
| 7207 | 'HSP90AA1_No6' | 'RAPFDLFENRK' | 'Heat shock protein HSP 90-alpha' |
| 7208 | 'UMOD_No1' | 'TALQPMVSALNIR' | 'Uromodulin' |
| 7209 | 'UMOD_No2' | 'FSVQMFR' | 'Uromodulin' |
| 7210 | 'LAMB1_No1' | 'IPSWTGAGFVR' | 'Laminin subunit beta-1' |
| 7211 | 'LAMB1_No2' | 'EGFYDLSSEDPFGCK' | 'Laminin subunit beta-1' |
| 7212 | 'LAMB1_No3' | 'DIRDPNFCER' | 'Laminin subunit beta-1' |
| 7213 | 'LAMB1_No4' | 'KVDDIICDSR' | 'Laminin subunit beta-1' |
| 7214 | 'LAMB1_No5' | 'CGNTIPDDDNQVVSLSPGSR' | 'Laminin subunit beta-1' |
| 7215 | 'LAMB1_No6' | 'SCACNPLGTIPGGNPCDSETGHCYCK' | 'Laminin subunit beta-1' |
| 7216 | 'LAMB1_No7' | 'TCSECQELFWGDPDVECR' | 'Laminin subunit beta-1' |
| 7217 | 'LAMB1_No8' | 'LNVEGEHCDVCK' | 'Laminin subunit beta-1' |

| | A | B | C |
|---|---|---|---|
| 1 | **Peptide Name** | **Peptide Sequence** | **Peptide Description** |
| 7218 | 'LAMB1_No9' | 'NCEQCKPFYYQHPER' | 'Laminin subunit beta-1' |
| 7219 | 'LAMB1_No10' | 'ALDPAFKIEDPYSPR' | 'Laminin subunit beta-1' |
| 7220 | 'LAMB1_No11' | 'VESLSQVEVILQHSAADIAR' | 'Laminin subunit beta-1' |
| 7221 | 'LAMB1_No12' | 'YVVLPRPVCFEK' | 'Laminin subunit beta-1' |
| 7222 | 'LAMB1_No13' | 'YSDIEPSTEGEVIFR' | 'Laminin subunit beta-1' |
| 7223 | 'LAMB1_No14' | 'LSVTSTCGLHKPEPYCIVSHLQEDKK' | 'Laminin subunit beta-1' |
| 7224 | 'LAMB1_No15' | 'YFAYDCEASFPGISTGPMK' | 'Laminin subunit beta-1' |
| 7225 | 'LAMB1_No16' | 'CLYHTEGEHCQFCR' | 'Laminin subunit beta-1' |
| 7226 | 'LAMB1_No17' | 'SCYQDPVTLQLACVCDPGYIGSR' | 'Laminin subunit beta-1' |
| 7227 | 'LYN_No1' | 'GSLLDFLK' | 'Tyrosine-protein kinase Lyn' |
| 7228 | 'LYN_No2' | 'LNTLETEEWFFK' | 'Tyrosine-protein kinase Lyn' |
| 7229 | 'LYN_No3' | 'QLLAPGNSAGAFLIR' | 'Tyrosine-protein kinase Lyn' |
| 7230 | 'LYN_No4' | 'WTAPEAINFGCFTIK' | 'Tyrosine-protein kinase Lyn' |
| 7231 | 'TPM2_No1' | 'HIAEDSDRKYEEVAR' | 'Tropomyosin beta chain' |
| 7232 | 'TPM2_No2' | 'MELQEMQLK' | 'Tropomyosin beta chain' |
| 7233 | 'TPM2_No3' | 'MELQEMQLK' | 'Tropomyosin beta chain' |
| 7234 | 'SFTPB_No1' | 'LVLPVLPGALQAR' | 'Pulmonary surfactant-associated protein B' |
| 7235 | 'SFTPB_No2' | 'GALAVAVAQVCR' | 'Pulmonary surfactant-associated protein B' |
| 7236 | 'SFTPB_No3' | 'QFVEQHTPQLLTLVPR' | 'Pulmonary surfactant-associated protein B' |
| 7237 | 'SFTPB_No4' | 'VVPLVAGGICQCLAER' | 'Pulmonary surfactant-associated protein B' |
| 7238 | 'SFTPB_No5' | 'QFVEQHTPQLLTLVPR' | 'Pulmonary surfactant-associated protein B' |
| 7239 | 'SFTPB_No6' | 'YSVILLDTLLGR' | 'Pulmonary surfactant-associated protein B' |
| 7240 | 'SFTPB_No7' | 'ALGHCLQEVWGHVGADDLCQECEDIVHILNK' | 'Pulmonary surfactant-associated protein B' |
| 7241 | 'SFTPB_No8' | 'SPTGEWLPR' | 'Pulmonary surfactant-associated protein B' |
| 7242 | 'SFTPB_No9' | 'KFLEQECNVLPLK' | 'Pulmonary surfactant-associated protein B' |
| 7243 | 'SFTPB_No10' | 'MLPQLVCR' | 'Pulmonary surfactant-associated protein B' |
| 7244 | 'SFTPB_No11' | 'GWDAHTTCQALGVCGTMSSPLQCIHSPDL' | 'Pulmonary surfactant-associated protein B' |
| 7245 | 'SFTPB_No12' | 'EAIFQDTMR' | 'Pulmonary surfactant-associated protein B' |
| 7246 | 'THBS1_No1' | 'FVFGTTPEDILR' | 'Thrombospondin-1' |
| 7247 | 'THBS1_No2' | 'GFLLLASLR' | 'Thrombospondin-1' |
| 7248 | 'THBS1_No3' | 'DNCQYVYNVDQR' | 'Thrombospondin-1' |
| 7249 | 'THBS1_No4' | 'TKDLQAICGISCDELSSMVLELR' | 'Thrombospondin-1' |
| 7250 | 'THBS1_No5' | 'FYVVMWK' | 'Thrombospondin-1' |
| 7251 | 'THBS1_No6' | 'GPDPSSPAFR' | 'Thrombospondin-1' |
| 7252 | 'THBS1_No7' | 'FQMIPLDPK' | 'Thrombospondin-1' |
| 7253 | 'THBS1_No8' | 'CENTDPGYNCLPCPPR' | 'Thrombospondin-1' |
| 7254 | 'THBS1_No9' | 'AQLYIDCEKMENAELDVPIQSVFTR' | 'Thrombospondin-1' |
| 7255 | 'THBS1_No10' | 'LCNNPTPQFGGK' | 'Thrombospondin-1' |
| 7256 | 'THBS1_No11' | 'CNYLGHYSDPMYR' | 'Thrombospondin-1' |
| 7257 | 'THBS1_No12' | 'QHVVSVEEALLATGQWK' | 'Thrombospondin-1' |
| 7258 | 'THBS1_No13' | 'LCNSPSPQMNGKPCEGEAR' | 'Thrombospondin-1' |
| 7259 | 'THBS1_No14' | 'KDHSGQVFSVVSNGK' | 'Thrombospondin-1' |
| 7260 | 'THBS1_No15' | 'NALWHTGNTPGQVR' | 'Thrombospondin-1' |
| 7261 | 'THBS1_No16' | 'QVTQSYWDTNPTR' | 'Thrombospondin-1' |

| | A | B | C |
|---|---|---|---|
| 1 | **Peptide Name** | **Peptide Sequence** | **Peptide Description** |
| 7262 | 'THBS1_No17' | 'RPPLCYHNGVQYR' | 'Thrombospondin-1' |
| 7263 | 'THBS1_No18' | 'KVTEENKELANELR' | 'Thrombospondin-1' |
| 7264 | 'THBS1_No19' | 'SCDSLNNRCEGSSVQTR' | 'Thrombospondin-1' |
| 7265 | 'THBS1_No20' | 'CTSYPDGSWK' | 'Thrombospondin-1' |
| 7266 | 'THBS1_No21' | 'NNEEWTVDSCTECHCQNSVTICK' | 'Thrombospondin-1' |
| 7267 | 'THBS1_No22' | 'AGTLDLSLTVQGK' | 'Thrombospondin-1' |
| 7268 | 'THBS1_No23' | 'LCNSPSPQMNGKPCEGEAR' | 'Thrombospondin-1' |
| 7269 | 'THBS1_No24' | 'IMADSGPIYDK' | 'Thrombospondin-1' |
| 7270 | 'THBS1_No25' | 'IEDANLIPPVPDDKFQDLVDAVR' | 'Thrombospondin-1' |
| 7271 | 'THBS1_No26' | 'IMADSGPIYDK' | 'Thrombospondin-1' |
| 7272 | 'THBS1_No27' | 'GTSQNDPNWVVR' | 'Thrombospondin-1' |
| 7273 | 'THBS1_No28' | 'QHVVSVEEALLATGQWK' | 'Thrombospondin-1' |
| 7274 | 'THBS1_No29' | 'RFQMIPLDPK' | 'Thrombospondin-1' |
| 7275 | 'THBS1_No30' | 'DHSGQVFSVVSNGK' | 'Thrombospondin-1' |
| 7276 | 'THBS1_No31' | 'GDACDHDDDNDGIPDDKDNCR' | 'Thrombospondin-1' |
| 7277 | 'THBS1_No32' | 'TIVTTLQDSIRK' | 'Thrombospondin-1' |
| 7278 | 'THBS1_No33' | 'FTGSQPFGQGVEHATANK' | 'Thrombospondin-1' |
| 7279 | 'THBS1_No34' | 'CNYLGHYSDPMYR' | 'Thrombospondin-1' |
| 7280 | 'THBS1_No35' | 'IMADSGPIYDKTYAGGR' | 'Thrombospondin-1' |
| 7281 | 'THBS1_No36' | 'EVPDACFNHNGEHR' | 'Thrombospondin-1' |
| 7282 | 'THBS1_No37' | 'KIMADSGPIYDK' | 'Thrombospondin-1' |
| 7283 | 'THBS1_No38' | 'LVPNPDQKDSDGDGR' | 'Thrombospondin-1' |
| 7284 | 'THBS1_No39' | 'LCNNPTPQFGGKDCVGDVTENQICNK' | 'Thrombospondin-1' |
| 7285 | 'THBS1_No40' | 'SRLCNNPTPQFGGK' | 'Thrombospondin-1' |
| 7286 | 'THBS1_No41' | 'DCVGDVTENQICNK' | 'Thrombospondin-1' |
| 7287 | 'THBS1_No42' | 'GGVNDNFQGVLQNVR' | 'Thrombospondin-1' |
| 7288 | 'THBS1_No43' | 'SITLFVQEDR' | 'Thrombospondin-1' |
| 7289 | 'THBS1_No44' | 'AEKGFLLLASLR' | 'Thrombospondin-1' |
| 7290 | 'THBS1_No45' | 'VTEENKELANELR' | 'Thrombospondin-1' |
| 7291 | 'THBS1_No46' | 'MENAELDVPIQSVFTR' | 'Thrombospondin-1' |
| 7292 | 'THBS1_No47' | 'IEDANLIPPVPDDKFQDLVDAVRAEK' | 'Thrombospondin-1' |
| 7293 | 'THBS1_No48' | 'DGKGDACDHDDDNDGIPDDKDNCR' | 'Thrombospondin-1' |
| 7294 | 'THBS1_No49' | 'HIGWKDFTAYR' | 'Thrombospondin-1' |
| 7295 | 'THBS1_No50' | 'IRLCNSPSPQMNGKPCEGEAR' | 'Thrombospondin-1' |
| 7296 | 'THBS1_No51' | 'FYVVMWK' | 'Thrombospondin-1' |
| 7297 | 'THBS1_No52' | 'LVKGPDPSSPAFR' | 'Thrombospondin-1' |
| 7298 | 'THBS1_No53' | 'FQMIPLDPKGTSQNDPNWVVR' | 'Thrombospondin-1' |
| 7299 | 'THBS1_No54' | 'NNEEWTVDSCTECHCQNSVTICKK' | 'Thrombospondin-1' |
| 7300 | 'THBS1_No55' | 'QDCPIDGCLSNPCFAGVK' | 'Thrombospondin-1' |
| 7301 | 'THBS1_No56' | 'MENAELDVPIQSVFTR' | 'Thrombospondin-1' |
| 7302 | 'THBS1_No57' | 'GTLLALER' | 'Thrombospondin-1' |
| 7303 | 'THBS1_No58' | 'TNYIGHK' | 'Thrombospondin-1' |
| 7304 | 'THBS1_No59' | 'IPESGGDNSVFDIFELTGAARK' | 'Thrombospondin-1' |
| 7305 | 'THBS1_No60' | 'AQLYIDCEKMENAELDVPIQSVFTR' | 'Thrombospondin-1' |

| | A | B | C |
|---|---|---|---|
| 1 | **Peptide Name** | **Peptide Sequence** | **Peptide Description** |
| 7306 | 'THBS1_No61' | 'DTDMDGVGDQCDNCPLEHNPDQLDSDSDR' | 'Thrombospondin-1' |
| 7307 | 'THBS1_No62' | 'IPESGGDNSVFDIFELTGAAR' | 'Thrombospondin-1' |
| 7308 | 'THBS1_No63' | 'IMADSGPIYDKTYAGGR' | 'Thrombospondin-1' |
| 7309 | 'THBS1_No64' | 'FQMIPLDPK' | 'Thrombospondin-1' |
| 7310 | 'THBS1_No65' | 'IGDTCDNNQDIDEDGHQNNLDNCPYVPNANQADHDKDGK' | 'Thrombospondin-1' |
| 7311 | 'RNASE1_No1' | 'QHMDSDSSPSSSSTYCNQMMR' | 'Ribonuclease pancreatic' |
| 7312 | 'RNASE1_No2' | 'QHMDSDSSPSSSSTYCNQMMR' | 'Ribonuclease pancreatic' |
| 7313 | 'RNASE1_No3' | 'QHMDSDSSPSSSSTYCNQMMR' | 'Ribonuclease pancreatic' |
| 7314 | 'RNASE1_No4' | 'HIIVACEGSPYVPVHFDASVEDST' | 'Ribonuclease pancreatic' |
| 7315 | 'RNASE1_No5' | 'QHMDSDSSPSSSSTYCNQMMR' | 'Ribonuclease pancreatic' |
| 7316 | 'RNASE1_No6' | 'QHMDSDSSPSSSSTYCNQMMR' | 'Ribonuclease pancreatic' |
| 7317 | 'RNASE1_No7' | 'QHMDSDSSPSSSSTYCNQMMR' | 'Ribonuclease pancreatic' |
| 7318 | 'RNASE1_No8' | 'QHMDSDSSPSSSSTYCNQMMR' | 'Ribonuclease pancreatic' |
| 7319 | 'RNASE1_No9' | 'QHMDSDSSPSSSSTYCNQMMR' | 'Ribonuclease pancreatic' |
| 7320 | 'CD55_No1' | 'QPYITQNYFPVGTVVEYECRPGYR' | 'Complement decay-accelerating factor' |
| 7321 | 'CD55_No2' | 'IPGEKDSVICLK' | 'Complement decay-accelerating factor' |
| 7322 | 'SERPINA6_No1' | 'EENFYVDETTVVK' | 'Corticosteroid-binding globulin' |
| 7323 | 'SERPINA6_No2' | 'GLASANVDFAFSLYK' | 'Corticosteroid-binding globulin' |
| 7324 | 'SERPINA6_No3' | 'GTWTQPFDLASTR' | 'Corticosteroid-binding globulin' |
| 7325 | 'SERPINA6_No4' | 'MNTVIAALSR' | 'Corticosteroid-binding globulin' |
| 7326 | 'SERPINA6_No5' | 'HYYESEVLAMNFQDWATASR' | 'Corticosteroid-binding globulin' |
| 7327 | 'SERPINA6_No6' | 'SETEIHQGFQHLHQLFAK' | 'Corticosteroid-binding globulin' |
| 7328 | 'SERPINA6_No7' | 'HLVALSPK' | 'Corticosteroid-binding globulin' |
| 7329 | 'SERPINA6_No8' | 'ITQDAQLK' | 'Corticosteroid-binding globulin' |
| 7330 | 'SERPINA6_No9' | 'AVLQLNEEGVDTAGSTGVTLNLTSKPIILR' | 'Corticosteroid-binding globulin' |
| 7331 | 'SERPINA6_No10' | 'WSAGLTSSQVDLYIPK' | 'Corticosteroid-binding globulin' |
| 7332 | 'SERPINA6_No11' | 'GTWTQPFDLASTREENFYVDETTVVK' | 'Corticosteroid-binding globulin' |
| 7333 | 'SERPINA6_No12' | 'HYYESEVLAMNFQDWATASR' | 'Corticosteroid-binding globulin' |
| 7334 | 'SERPINA6_No13' | 'MNTVIAALSR' | 'Corticosteroid-binding globulin' |
| 7335 | 'SERPINA6_No14' | 'AQLLQGLGFNLTER' | 'Corticosteroid-binding globulin' |
| 7336 | 'SERPINA6_No15' | 'AVLQLNEEGVDTAGSTGVTLNLTSK' | 'Corticosteroid-binding globulin' |
| 7337 | 'SERPINA6_No16' | 'MNTVIAALSRDTINR' | 'Corticosteroid-binding globulin' |
| 7338 | 'SERPINA6_No17' | 'GKMNTVIAALSR' | 'Corticosteroid-binding globulin' |
| 7339 | 'SERPINA6_No18' | 'DTINRWSAGLTSSQVDLYIPK' | 'Corticosteroid-binding globulin' |
| 7340 | 'SLC3A2_No1' | 'LLTSFLPAQLLR' | '4F2 cell-surface antigen heavy chain' |
| 7341 | 'SLC3A2_No2' | 'VILDLTPNYR' | '4F2 cell-surface antigen heavy chain' |
| 7342 | 'SLC3A2_No3' | 'IGDLQAFQGHGAGNLAGLK' | '4F2 cell-surface antigen heavy chain' |
| 7343 | 'SLC3A2_No4' | 'NQKDDVAQTDLLQIDPNFGSK' | '4F2 cell-surface antigen heavy chain' |
| 7344 | 'SLC3A2_No5' | 'ADLLLSTQPGREEGSPLELER' | '4F2 cell-surface antigen heavy chain' |
| 7345 | 'SLC3A2_No6' | 'GQSEDPGSLLSLFR' | '4F2 cell-surface antigen heavy chain' |
| 7346 | 'SLC3A2_No7' | 'DDVAQTDLLQIDPNFGSK' | '4F2 cell-surface antigen heavy chain' |
| 7347 | 'SLC3A2_No8' | 'SLLHGDFHAFSAGPGLFSYIR' | '4F2 cell-surface antigen heavy chain' |
| 7348 | 'SLC3A2_No9' | 'LKLEPHEGLLLR' | '4F2 cell-surface antigen heavy chain' |
| 7349 | 'SLC3A2_No10' | 'NQKDDVAQTDLLQIDPNFGSKEDFDSLLQSAK' | '4F2 cell-surface antigen heavy chain' |

| | A | B | C |
|---|---|---|---|
| 1 | **Peptide Name** | **Peptide Sequence** | **Peptide Description** |
| 7350 | 'CELA2A' | 'LGCNYYHKPSVFTR' | 'Chymotrypsin-like elastase family member 2A' |
| 7351 | 'HSP90AB1_No1' | 'ELISNASDALDKIR' | 'Heat shock protein HSP 90-beta' |
| 7352 | 'HSP90AB1_No2' | 'ALLFIPR' | 'Heat shock protein HSP 90-beta' |
| 7353 | 'HSP90AB1_No3' | 'SLTNDWEDHLAVK' | 'Heat shock protein HSP 90-beta' |
| 7354 | 'HSP90AB1_No4' | 'VILHLKEDQTEYLEER' | 'Heat shock protein HSP 90-beta' |
| 7355 | 'HSP90AB1_No5' | 'HLEINPDHPIVETLR' | 'Heat shock protein HSP 90-beta' |
| 7356 | 'HSP90AB1_No6' | 'HFSVEGQLEFR' | 'Heat shock protein HSP 90-beta' |
| 7357 | 'HSP90AB1_No7' | 'ADLINNLGTIAK' | 'Heat shock protein HSP 90-beta' |
| 7358 | 'HSP90AB1_No8' | 'VILHLKEDQTEYLEERR' | 'Heat shock protein HSP 90-beta' |
| 7359 | 'HSP90AB1_No9' | 'YHTSQSGDEMTSLSEYVSR' | 'Heat shock protein HSP 90-beta' |
| 7360 | 'HSP90AB1_No10' | 'HNDDEQYAWESSAGGSFTVR' | 'Heat shock protein HSP 90-beta' |
| 7361 | 'HSP90AB1_No11' | 'ELISNASDALDK' | 'Heat shock protein HSP 90-beta' |
| 7362 | 'HSP90AB1_No12' | 'EGLELPEDEEEKK' | 'Heat shock protein HSP 90-beta' |
| 7363 | 'ELANE_No1' | 'QAGVCFGDSGSPLVCNGLIHGIASFVR' | 'Neutrophil elastase' |
| 7364 | 'ELANE_No2' | 'SEDNPCPHPRDPDPASR' | 'Neutrophil elastase' |
| 7365 | 'MMP2_No1' | 'AFQVWSDVTPLR' | '72 kDa type IV collagenase' |
| 7366 | 'MMP2_No2' | 'FQGTSYDSCTTEGR' | '72 kDa type IV collagenase' |
| 7367 | 'MMP2_No3' | 'FFGLPQTGDLDQNTIETMR' | '72 kDa type IV collagenase' |
| 7368 | 'MMP2_No4' | 'IIGYTPDLDPETVDDAFAR' | '72 kDa type IV collagenase' |
| 7369 | 'MMP2_No5' | 'MWCATTANYDDDRK' | '72 kDa type IV collagenase' |
| 7370 | 'MMP2_No6' | 'DGLLAHAFAPGTGVGGDSHFDDDELWTLGEGQVVR' | '72 kDa type IV collagenase' |
| 7371 | 'MMP2_No7' | 'ESCNLFVLK' | '72 kDa type IV collagenase' |
| 7372 | 'MMP2_No8' | 'TYIFAGDKFWR' | '72 kDa type IV collagenase' |
| 7373 | 'MMP2_No9' | 'ESCNLFVLKDTLKK' | '72 kDa type IV collagenase' |
| 7374 | 'MMP2_No10' | 'TDKELAVQYLNTFYGCPK' | '72 kDa type IV collagenase' |
| 7375 | 'MMP2_No11' | 'YGFCPHEALFTMGGNAEGQPCK' | '72 kDa type IV collagenase' |
| 7376 | 'MMP2_No12' | 'WEHGDGYPFDGK' | '72 kDa type IV collagenase' |
| 7377 | 'MMP2_No13' | 'GYPKPLTSLGLPPDVQR' | '72 kDa type IV collagenase' |
| 7378 | 'MMP2_No14' | 'WCGTTEDYDR' | '72 kDa type IV collagenase' |
| 7379 | 'MMP2_No15' | 'MWCATTANYDDDRK' | '72 kDa type IV collagenase' |
| 7380 | 'MMP2_No16' | 'FFGLPQTGDLDQNTIETMR' | '72 kDa type IV collagenase' |
| 7381 | 'MMP2_No17' | 'WDKNQITYR' | '72 kDa type IV collagenase' |
| 7382 | 'MMP2_No18' | 'IHDGEADIMINFGR' | '72 kDa type IV collagenase' |
| 7383 | 'MMP2_No19' | 'DKPMGPLLVATFWPELPEK' | '72 kDa type IV collagenase' |
| 7384 | 'MMP2_No20' | 'WCGTTEDYDRDK' | '72 kDa type IV collagenase' |
| 7385 | 'SOD3_No1' | 'AVVVHAGEDDLGR' | 'Extracellular superoxide dismutase [Cu-Zn]' |
| 7386 | 'SOD3_No2' | 'VTEIWQEVMQR' | 'Extracellular superoxide dismutase [Cu-Zn]' |
| 7387 | 'SOD3_No3' | 'AGLAASLAGPHSIVGR' | 'Extracellular superoxide dismutase [Cu-Zn]' |
| 7388 | 'SOD3_No4' | 'LACCVVGVCGPGLWER' | 'Extracellular superoxide dismutase [Cu-Zn]' |
| 7389 | 'SOD3_No5' | 'RDDDGALHAACQVQPSATLDAAQPR' | 'Extracellular superoxide dismutase [Cu-Zn]' |
| 7390 | 'SOD3_No6' | 'AVVVHAGEDDLGRGGNQASVENGNAGRR' | 'Extracellular superoxide dismutase [Cu-Zn]' |
| 7391 | 'SOD3_No7' | 'AVVVHAGEDDLGRGGNQASVENGNAGR' | 'Extracellular superoxide dismutase [Cu-Zn]' |
| 7392 | 'SOD3_No8' | 'VTEIWQEVMQR' | 'Extracellular superoxide dismutase [Cu-Zn]' |
| 7393 | 'CTSG_No1' | 'VSSFLPWIR' | 'Cathepsin G' |

| | A | B | C |
|---|---|---|---|
| 1 | **Peptide Name** | **Peptide Sequence** | **Peptide Description** |
| 7394 | 'CTSG_No2' | 'TIQNDIMLLQLSR' | 'Cathepsin G' |
| 7395 | 'CTSG_No3' | 'TIQNDIMLLQLSR' | 'Cathepsin G' |
| 7396 | 'CTSG_No4' | 'AQEGLRPGTLCTVAGWGR' | 'Cathepsin G' |
| 7397 | 'CTSG_No5' | 'GDSGGPLLCNNVAHGIVSYGK' | 'Cathepsin G' |
| 7398 | 'ADH4_No1' | 'VTPGSTCAVFGLGGVGLSAVMGCK' | 'All-trans-retinol dehydrogenase [NAD(+)] ADH4' |
| 7399 | 'ADH4_No2' | 'VCLLGCGFSTGYGAAINNAK' | 'All-trans-retinol dehydrogenase [NAD(+)] ADH4' |
| 7400 | 'ADH4_No3' | 'FCLSPLTNLCGK' | 'All-trans-retinol dehydrogenase [NAD(+)] ADH4' |
| 7401 | 'MGP_No1' | 'NANTFISPQQR' | 'Matrix Gla protein' |
| 7402 | 'MGP_No2' | 'YAMVYGYNAAYNR' | 'Matrix Gla protein' |
| 7403 | 'MGP_No3' | 'YAMVYGYNAAYNR' | 'Matrix Gla protein' |
| 7404 | 'ITGA2B_No1' | 'VYLFLQPR' | 'Integrin alpha-IIb' |
| 7405 | 'ITGA2B_No2' | 'GPHALGAPSLLLTGTQLYGR' | 'Integrin alpha-IIb' |
| 7406 | 'ITGA2B_No3' | 'IVLLDVPVR' | 'Integrin alpha-IIb' |
| 7407 | 'ITGA2B_No4' | 'GQVLVFLGQSEGLR' | 'Integrin alpha-IIb' |
| 7408 | 'ITGA2B_No5' | 'NRPPLEEDDEEGE' | 'Integrin alpha-IIb' |
| 7409 | 'ITGA2B_No6' | 'VVLCELGNPMKK' | 'Integrin alpha-IIb' |
| 7410 | 'ITGA2B_No7' | 'TPVGSCFLAQPESGR' | 'Integrin alpha-IIb' |
| 7411 | 'ITGA2B_No8' | 'DGYNDIAVAAPYGGPSGR' | 'Integrin alpha-IIb' |
| 7412 | 'ITGA2B_No9' | 'IYVENDFSWDKR' | 'Integrin alpha-IIb' |
| 7413 | 'ITGA2B_No10' | 'VAIVVGAPR' | 'Integrin alpha-IIb' |
| 7414 | 'ITGA2B_No11' | 'IVLLDVPVRAEAQVELR' | 'Integrin alpha-IIb' |
| 7415 | 'ITGA2B_No12' | 'VLLLGSQQAGTTLNLDLGGK' | 'Integrin alpha-IIb' |
| 7416 | 'ITGA2B_No13' | 'TLGPSQEETGGVFLCPWR' | 'Integrin alpha-IIb' |
| 7417 | 'ITGA2B_No14' | 'SRPSQVLDSPFPTGSAFGFSLR' | 'Integrin alpha-IIb' |
| 7418 | 'ITGA2B_No15' | 'LQDPVLVSCDSAPCTVVQCDLQEMAR' | 'Integrin alpha-IIb' |
| 7419 | 'ITGA2B_No16' | 'AEGGQCPSLLFDLRDETR' | 'Integrin alpha-IIb' |
| 7420 | 'ITGA2B_No17' | 'HSPICHTTMAFLR' | 'Integrin alpha-IIb' |
| 7421 | 'ITGA2B_No18' | 'GNSFPASLVVAAEEGEREQNSLDSWGPK' | 'Integrin alpha-IIb' |
| 7422 | 'ITGA2B_No19' | 'AEGGQCPSLLFDLR' | 'Integrin alpha-IIb' |
| 7423 | 'ITGA2B_No20' | 'VVLCELGNPMK' | 'Integrin alpha-IIb' |
| 7424 | 'ITGA2B_No21' | 'RVLLLGSQQAGTTLNLDLGGK' | 'Integrin alpha-IIb' |
| 7425 | 'ITGA2B_No22' | 'TEEAEKTPVGSCFLAQPESGR' | 'Integrin alpha-IIb' |
| 7426 | 'ITGA2B_No23' | 'GEAQVWTQLLR' | 'Integrin alpha-IIb' |
| 7427 | 'ITGA2B_No24' | 'TEEAEKTPVGSCFLAQPESGRR' | 'Integrin alpha-IIb' |
| 7428 | 'ITGA2B_No25' | 'FGSAIAPLGDLDR' | 'Integrin alpha-IIb' |
| 7429 | 'ITGA2B_No26' | 'GAVDIDDNGYPDLIVGAYGANQVAVYR' | 'Integrin alpha-IIb' |
| 7430 | 'ITGA2B_No27' | 'HDLLVGAPLYMESR' | 'Integrin alpha-IIb' |
| 7431 | 'ITGA2B_No28' | 'TPVGSCFLAQPESGRR' | 'Integrin alpha-IIb' |
| 7432 | 'ITGA2B_No29' | 'ALSNVEGFER' | 'Integrin alpha-IIb' |
| 7433 | 'ITGA2B_No30' | 'FGSAIAPLGDLDRDGYNDIAVAAPYGGPSGR' | 'Integrin alpha-IIb' |
| 7434 | 'ITGA2B_No31' | 'HSPICHTTMAFLRDEADFRDK' | 'Integrin alpha-IIb' |
| 7435 | 'ITGA2B_No32' | 'TPVSCFNIQMCVGATGHNIPQK' | 'Integrin alpha-IIb' |
| 7436 | 'ITGA2B_No33' | 'LSLNAELQLDR' | 'Integrin alpha-IIb' |
| 7437 | 'ITGA2B_No34' | 'GNSFPASLVVAAEEGER' | 'Integrin alpha-IIb' |

| | A | B | C |
|---|---|---|---|
| 1 | **Peptide Name** | **Peptide Sequence** | **Peptide Description** |
| 7438 | 'ITGA2B_No35' | 'LQDPVLVSCDSAPCTVVQCDLQEMAR' | 'Integrin alpha-IIb' |
| 7439 | 'LPA_No1' | 'NPDAVAAPYCYTR' | 'Apolipoprotein(a)' |
| 7440 | 'LPA_No2' | 'TCQAWSSMTPHSHSR' | 'Apolipoprotein(a)' |
| 7441 | 'LPA_No3' | 'TPEYYPNAGLIMNYCR' | 'Apolipoprotein(a)' |
| 7442 | 'LPA_No4' | 'TPENYPNAGLTR' | 'Apolipoprotein(a)' |
| 7443 | 'LPA_No5' | 'GTDSCQGDSGGPLVCFEK' | 'Apolipoprotein(a)' |
| 7444 | 'LPA_No6' | 'NPDPVAAPYCYTR' | 'Apolipoprotein(a)' |
| 7445 | 'LPA_No7' | 'ATTVTGTPCQEWAAQEPHR' | 'Apolipoprotein(a)' |
| 7446 | 'LPA_No8' | 'FVTWIEGMMR' | 'Apolipoprotein(a)' |
| 7447 | 'LPA_No9' | 'TPEYYPNAGLIMNYCR' | 'Apolipoprotein(a)' |
| 7448 | 'LPA_No10' | 'TTEYYPNGGLTR' | 'Apolipoprotein(a)' |
| 7449 | 'LPA_No11' | 'TPAYYPNAGLIK' | 'Apolipoprotein(a)' |
| 7450 | 'LPA_No12' | 'IPLYYPNAGLTR' | 'Apolipoprotein(a)' |
| 7451 | 'LPA_No13' | 'GTYSTTVTGR' | 'Apolipoprotein(a)' |
| 7452 | 'LPA_No14' | 'KATTVTGTPCQEWAAQEPHR' | 'Apolipoprotein(a)' |
| 7453 | 'LPA_No15' | 'VMPACLPSPDYMVTAR' | 'Apolipoprotein(a)' |
| 7454 | 'LPA_No16' | 'TPENYPNDGLTMNYCR' | 'Apolipoprotein(a)' |
| 7455 | 'LPA_No17' | 'NPDGDINGPWCYTMNPR' | 'Apolipoprotein(a)' |
| 7456 | 'LPA_No18' | 'QPWCYTTDPCVR' | 'Apolipoprotein(a)' |
| 7457 | 'LPA_No19' | 'NPDAEISPWCYTMDPNVR' | 'Apolipoprotein(a)' |
| 7458 | 'LPA_No20' | 'NPDAEISPWCYTMDPNVR' | 'Apolipoprotein(a)' |
| 7459 | 'LPA_No21' | 'NPDAEIRPWCYTMDPSVR' | 'Apolipoprotein(a)' |
| 7460 | 'LPA_No22' | 'NPDSGKQPWCYTTDPCVR' | 'Apolipoprotein(a)' |
| 7461 | 'LPA_No23' | 'TCQSWSSMTPHR' | 'Apolipoprotein(a)' |
| 7462 | 'LPA_No24' | 'TCQSWSSMTPHWHR' | 'Apolipoprotein(a)' |
| 7463 | 'LPA_No25' | 'HFCGGTLISPEWVLTAAHCLK' | 'Apolipoprotein(a)' |
| 7464 | 'LPA_No26' | 'GTFSTTVTGR' | 'Apolipoprotein(a)' |
| 7465 | 'LPA_No27' | 'LFDYCDIPLCASSSFDCGKPQVEPK' | 'Apolipoprotein(a)' |
| 7466 | 'LPA_No28' | 'LSRPAVITDK' | 'Apolipoprotein(a)' |
| 7467 | 'LPA_No29' | 'NPDPVAAPWCYTTDPSVR' | 'Apolipoprotein(a)' |
| 7468 | 'LPA_No30' | 'LFLEPTQADIALLK' | 'Apolipoprotein(a)' |
| 7469 | 'LPA_No31' | 'TCQSWSSMTPHWHQR' | 'Apolipoprotein(a)' |
| 7470 | 'LPA_No32' | 'TPENYPNDGLTMNYCR' | 'Apolipoprotein(a)' |
| 7471 | 'LPA_No33' | 'TCQAWSSMTPHQHSR' | 'Apolipoprotein(a)' |
| 7472 | 'LPA_No34' | 'EAQLLVIENEVCNHYK' | 'Apolipoprotein(a)' |
| 7473 | 'LPA_No35' | 'TCQAWSSMTPHSHSR' | 'Apolipoprotein(a)' |
| 7474 | 'LPA_No36' | 'VILGAHQEVNLESHVQEIEVSR' | 'Apolipoprotein(a)' |
| 7475 | 'LPA_No37' | 'YICAEHLAR' | 'Apolipoprotein(a)' |
| 7476 | 'LPA_No38' | 'SPVVQDCYHGDGR' | 'Apolipoprotein(a)' |
| 7477 | 'LPA_No39' | 'NPDAEIRPWCYTMDPSVR' | 'Apolipoprotein(a)' |
| 7478 | 'LPA_No40' | 'NPDAVAAPYCYTRDPGVR' | 'Apolipoprotein(a)' |
| 7479 | 'LPA_No41' | 'TCQSWSSMIPHWHQR' | 'Apolipoprotein(a)' |
| 7480 | 'LPA_No42' | 'VMPACLPSPDYMVTAR' | 'Apolipoprotein(a)' |
| 7481 | 'LPA_No43' | 'NPDGDINGPWCYTMNPR' | 'Apolipoprotein(a)' |

| | A | B | C |
|---|---|---|---|
| 1 | **Peptide Name** | **Peptide Sequence** | **Peptide Description** |
| 7482 | 'LPA_No44' | 'RIPLYYPNAGLTR' | 'Apolipoprotein(a)' |
| 7483 | 'LPA_No45' | 'TPENYPNAGLTENYCR' | 'Apolipoprotein(a)' |
| 7484 | 'LPA_No46' | 'HFCGGTLISPEWVLTAAHCLKK' | 'Apolipoprotein(a)' |
| 7485 | 'LPA_No47' | 'NPDADTGPWCFTMDPSIR' | 'Apolipoprotein(a)' |
| 7486 | 'LPA_No48' | 'VMPACLPSPDYMVTAR' | 'Apolipoprotein(a)' |
| 7487 | 'LPA_No49' | 'GTLSTTITGR' | 'Apolipoprotein(a)' |
| 7488 | 'LPA_No50' | 'TTENYPNAGLIMNYCR' | 'Apolipoprotein(a)' |
| 7489 | 'LPA_No51' | 'KCPGSIVGGCVAHPHSWPWQVSLR' | 'Apolipoprotein(a)' |
| 7490 | 'LPA_No52' | 'TECYITGWGETQGTFGTGLLK' | 'Apolipoprotein(a)' |
| 7491 | 'LPA_No53' | 'TCQSWSSMTPHWHQR' | 'Apolipoprotein(a)' |
| 7492 | 'LPA_No54' | 'TCQSWSSMTPHWHR' | 'Apolipoprotein(a)' |
| 7493 | 'LPA_No55' | 'HSTFIPGTNKWAGLEK' | 'Apolipoprotein(a)' |
| 7494 | 'LPA_No56' | 'NPDADTGPWCFTMDPSIR' | 'Apolipoprotein(a)' |
| 7495 | 'LPA_No57' | 'TCQSWSSMIPHWHQR' | 'Apolipoprotein(a)' |
| 7496 | 'LPA_No58' | 'TCQSWSSMTPHR' | 'Apolipoprotein(a)' |
| 7497 | 'LPA_No59' | 'KLFDYCDIPLCASSSFDCGKPQVEPK' | 'Apolipoprotein(a)' |
| 7498 | 'LPA_No60' | 'VMPACLPSPDYMVTAR' | 'Apolipoprotein(a)' |
| 7499 | 'LPA_No61' | 'TCQAWSSMTPHQHSR' | 'Apolipoprotein(a)' |
| 7500 | 'LPA_No62' | 'NPDPVAAPYCYTRDPSVR' | 'Apolipoprotein(a)' |
| 7501 | 'LPA_No63' | 'NYCRNPDAEISPWCYTMDPNVR' | 'Apolipoprotein(a)' |
| 7502 | 'LPA_No64' | 'TPENYPNAGLTENYCRNPDSGKQPWCYTTDPCVR' | 'Apolipoprotein(a)' |
| 7503 | 'LPA_No65' | 'GTDSCQGDSGGPLVCFEKDK' | 'Apolipoprotein(a)' |
| 7504 | 'LPA_No66' | 'TPENYPNAGLTENYCRNPDSGK' | 'Apolipoprotein(a)' |
| 7505 | 'LPA_No67' | 'PWCYTMDPSVR' | 'Apolipoprotein(a)' |
| 7506 | 'LPA_No68' | 'TTENYPNAGLIMNYCRNPDAVAAPYCYTR' | 'Apolipoprotein(a)' |
| 7507 | 'LPA_No69' | 'TPEYYPNAGLIMNYCRNPDAVAAPYCYTR' | 'Apolipoprotein(a)' |
| 7508 | 'LPA_No70' | 'NYCRNPDAEISPWCYTMDPNVR' | 'Apolipoprotein(a)' |
| 7509 | 'LPA_No71' | 'PWCYTMDPSVR' | 'Apolipoprotein(a)' |
| 7510 | 'LPA_No72' | 'TTENYPNAGLIMNYCRNPDAVAAPYCYTR' | 'Apolipoprotein(a)' |
| 7511 | 'LPA_No73' | 'TPEYYPNAGLIMNYCRNPDAVAAPYCYTR' | 'Apolipoprotein(a)' |
| 7512 | 'PLEK_No1' | 'EDPAYLHYYDPAGAEDPLGAIHLR' | 'Pleckstrin' |
| 7513 | 'PLEK_No2' | 'IFNHCFTGNCVIDWLVSNQSVR' | 'Pleckstrin' |
| 7514 | 'PLEK_No3' | 'QQDHFFQAAFLEER' | 'Pleckstrin' |
| 7515 | 'PLEK_No4' | 'FILREDPAYLHYYDPAGAEDPLGAIHLR' | 'Pleckstrin' |
| 7516 | 'PLEK_No5' | 'GSTLTSPCQDFGKR' | 'Pleckstrin' |
| 7517 | 'PLEK_No6' | 'NRQEGLMIASSLLNEGYLQPAGDMSK' | 'Pleckstrin' |
| 7518 | 'PLEK_No7' | 'NRQEGLMIASSLLNEGYLQPAGDMSK' | 'Pleckstrin' |
| 7519 | 'PLEK_No8' | 'KFILREDPAYLHYYDPAGAEDPLGAIHLR' | 'Pleckstrin' |
| 7520 | 'PLEK_No9' | 'LPETIDLGALYLSMKDTEK' | 'Pleckstrin' |
| 7521 | 'PLEK_No10' | 'LPETIDLGALYLSMK' | 'Pleckstrin' |
| 7522 | 'PLEK_No11' | 'NRQEGLMIASSLLNEGYLQPAGDMSK' | 'Pleckstrin' |
| 7523 | 'PLEK_No12' | 'LPETIDLGALYLSMKDTEK' | 'Pleckstrin' |
| 7524 | 'PLEK_No13' | 'LPETIDLGALYLSMK' | 'Pleckstrin' |
| 7525 | 'PLEK_No14' | 'QQDHFFQAAFLEER' | 'Pleckstrin' |

| | A | B | C |
|---|---|---|---|
| 1 | **Peptide Name** | **Peptide Sequence** | **Peptide Description** |
| 7526 | 'PLEK_No15' | 'KIFNHCFTGNCVIDWLVSNQSVR' | 'Pleckstrin' |
| 7527 | 'PLEK_No16' | 'NRQEGLMIASSLLNEGYLQPAGDMSK' | 'Pleckstrin' |
| 7528 | 'CD14_No1' | 'LTVGAAQVPAQLLVGALR' | 'Monocyte differentiation antigen CD14' |
| 7529 | 'CD14_No2' | 'FPAIQNLALR' | 'Monocyte differentiation antigen CD14' |
| 7530 | 'CD14_No3' | 'VLSIAQAHSPAFSCEQVR' | 'Monocyte differentiation antigen CD14' |
| 7531 | 'CD14_No4' | 'LKELTLEDLK' | 'Monocyte differentiation antigen CD14' |
| 7532 | 'CD14_No5' | 'GLMAALCPHK' | 'Monocyte differentiation antigen CD14' |
| 7533 | 'CD14_No6' | 'AFPALTSLDLSDNPGLGER' | 'Monocyte differentiation antigen CD14' |
| 7534 | 'CD14_No7' | 'NTGMETPTGVCAALAAAGVQPHSLDLSHNSLR' | 'Monocyte differentiation antigen CD14' |
| 7535 | 'CD14_No8' | 'RLTVGAAQVPAQLLVGALR' | 'Monocyte differentiation antigen CD14' |
| 7536 | 'CD14_No9' | 'GLMAALCPHKFPAIQNLALR' | 'Monocyte differentiation antigen CD14' |
| 7537 | 'CD14_No10' | 'ITGTMPPLPLEATGLALSSLR' | 'Monocyte differentiation antigen CD14' |
| 7538 | 'CD14_No11' | 'NTGMETPTGVCAALAAAGVQPHSLDLSHNSLR' | 'Monocyte differentiation antigen CD14' |
| 7539 | 'CD14_No12' | 'SWLAELQQWLKPGLK' | 'Monocyte differentiation antigen CD14' |
| 7540 | 'CD14_No13' | 'ITGTMPPLPLEATGLALSSLR' | 'Monocyte differentiation antigen CD14' |
| 7541 | 'CD14_No14' | 'RVDADADPRQYADTVK' | 'Monocyte differentiation antigen CD14' |
| 7542 | 'CD14_No15' | 'GLMAALCPHK' | 'Monocyte differentiation antigen CD14' |
| 7543 | 'CD14_No16' | 'VDADADPRQYADTVK' | 'Monocyte differentiation antigen CD14' |
| 7544 | 'CD14_No17' | 'GLMAALCPHKFPAIQNLALR' | 'Monocyte differentiation antigen CD14' |
| 7545 | 'CD14_No18' | 'VLDLSCNRLNR' | 'Monocyte differentiation antigen CD14' |
| 7546 | 'CD14_No19' | 'ATVNPSAPR' | 'Monocyte differentiation antigen CD14' |
| 7547 | 'CFH_No1' | 'SSNLIILEEHLK' | 'Complement factor H' |
| 7548 | 'CFH_No2' | 'DTSCVNPPTVQNAYIVSR' | 'Complement factor H' |
| 7549 | 'CFH_No3' | 'RPCGHPGDTPFGTFTLTGGNVFEYGVK' | 'Complement factor H' |
| 7550 | 'CFH_No4' | 'LGYVTADGETSGSITCGK' | 'Complement factor H' |
| 7551 | 'CFH_No5' | 'AGEQVTYTCATYYK' | 'Complement factor H' |
| 7552 | 'CFH_No6' | 'AQTTVTCMENGWSPTPR' | 'Complement factor H' |
| 7553 | 'CFH_No7' | 'VSVLCQENYLIQEGEEITCK' | 'Complement factor H' |
| 7554 | 'CFH_No8' | 'SITCIHGVWTQLPQCVAIDKLK' | 'Complement factor H' |
| 7555 | 'CFH_No9' | 'SSIDIENGFISESQYTYALKEK' | 'Complement factor H' |
| 7556 | 'CFH_No10' | 'WSSPPQCEGLPCK' | 'Complement factor H' |
| 7557 | 'CFH_No11' | 'SPDVINGSPISQK' | 'Complement factor H' |
| 7558 | 'CFH_No12' | 'EGWIHTVCINGR' | 'Complement factor H' |
| 7559 | 'CFH_No13' | 'GDAVCTESGWRPLPSCEEK' | 'Complement factor H' |
| 7560 | 'CFH_No14' | 'KGEWVALNPLRK' | 'Complement factor H' |
| 7561 | 'CFH_No15' | 'AQTTVTCMENGWSPTPR' | 'Complement factor H' |
| 7562 | 'CFH_No16' | 'WQSIPLCVEK' | 'Complement factor H' |
| 7563 | 'CFH_No17' | 'TGESVEFVCKR' | 'Complement factor H' |
| 7564 | 'CFH_No18' | 'NDFTWFK' | 'Complement factor H' |
| 7565 | 'CFH_No19' | 'GEWVALNPLR' | 'Complement factor H' |
| 7566 | 'CFH_No20' | 'IDVHLVPDRK' | 'Complement factor H' |
| 7567 | 'CFH_No21' | 'SIDVACHPGYALPK' | 'Complement factor H' |
| 7568 | 'CFH_No22' | 'SCDIPVFMNAR' | 'Complement factor H' |
| 7569 | 'CFH_No23' | 'GKEGWIHTVCINGR' | 'Complement factor H' |

| | A | B | C |
|---|---|---|---|
| 1 | **Peptide Name** | **Peptide Sequence** | **Peptide Description** |
| 7570 | 'CFH_No24' | 'RNTEILTGSWSDQTYPEGTQAIYK' | 'Complement factor H' |
| 7571 | 'CFH_No25' | 'SCDNPYIPNGDYSPLR' | 'Complement factor H' |
| 7572 | 'CFH_No26' | 'IVSSAMEPDREYHFGQAVR' | 'Complement factor H' |
| 7573 | 'CFH_No27' | 'TKEEYGHSEVVEYYCNPR' | 'Complement factor H' |
| 7574 | 'CFH_No28' | 'GEWVALNPLRK' | 'Complement factor H' |
| 7575 | 'CFH_No29' | 'IEGDEEMHCSDDGFWSK' | 'Complement factor H' |
| 7576 | 'CFH_No30' | 'TDCLSLPSFENAIPMGEKK' | 'Complement factor H' |
| 7577 | 'CFH_No31' | 'PGFTIVGPNSVQCYHFGLSPDLPICK' | 'Complement factor H' |
| 7578 | 'CFH_No32' | 'IEGDEEMHCSDDGFWSK' | 'Complement factor H' |
| 7579 | 'CFH_No33' | 'CNMGYEYSER' | 'Complement factor H' |
| 7580 | 'CFH_No34' | 'CTLKPCDYPDIK' | 'Complement factor H' |
| 7581 | 'CFH_No35' | 'CFEGFGIDGPAIAK' | 'Complement factor H' |
| 7582 | 'CFH_No36' | 'FVCNSGYKIEGDEEMHCSDDGFWSK' | 'Complement factor H' |
| 7583 | 'CFH_No37' | 'TGDEITYQCR' | 'Complement factor H' |
| 7584 | 'CFH_No38' | 'WSHPPSCIK' | 'Complement factor H' |
| 7585 | 'CFH_No39' | 'KGEWVALNPLR' | 'Complement factor H' |
| 7586 | 'CFH_No40' | 'CTSTGWIPAPR' | 'Complement factor H' |
| 7587 | 'CFH_No41' | 'AVYTCNEGYQLLGEINYRECDTDGWTNDIPICEVVK' | 'Complement factor H' |
| 7588 | 'CFH_No42' | 'IVSSAMEPDR' | 'Complement factor H' |
| 7589 | 'CFH_No43' | 'EFDHNSNIR' | 'Complement factor H' |
| 7590 | 'CFH_No44' | 'EQVQSCGPPPELLNGNVK' | 'Complement factor H' |
| 7591 | 'CFH_No45' | 'SITCIHGVWTQLPQCVAIDKLKK' | 'Complement factor H' |
| 7592 | 'CFH_No46' | 'CNMGYEYSER' | 'Complement factor H' |
| 7593 | 'CFH_No47' | 'TDCLSLPSFENAIPMGEK' | 'Complement factor H' |
| 7594 | 'CFH_No48' | 'SCDIPVFMNAR' | 'Complement factor H' |
| 7595 | 'CFH_No49' | 'ECELPKIDVHLVPDR' | 'Complement factor H' |
| 7596 | 'CFH_No50' | 'VSVLCQENYLIQEGEEITCKDGR' | 'Complement factor H' |
| 7597 | 'CFH_No51' | 'PLPSCEEK' | 'Complement factor H' |
| 7598 | 'CFH_No52' | 'TDCLSLPSFENAIPMGEKK' | 'Complement factor H' |
| 7599 | 'CFH_No53' | 'NTEILTGSWSDQTYPEGTQAIYK' | 'Complement factor H' |
| 7600 | 'CFH_No54' | 'SPPEISHGVVAHMSDSYQYGEEVTYK' | 'Complement factor H' |
| 7601 | 'CFH_No55' | 'ISEENETTCYMGK' | 'Complement factor H' |
| 7602 | 'CFH_No56' | 'IDVHLVPDR' | 'Complement factor H' |
| 7603 | 'CFH_No57' | 'IIYKENERFQYK' | 'Complement factor H' |
| 7604 | 'CFH_No58' | 'SITCIHGVWTQLPQCVAIDK' | 'Complement factor H' |
| 7605 | 'CFH_No59' | 'TGESVEFVCK' | 'Complement factor H' |
| 7606 | 'CFH_No60' | 'IVSSAMEPDREYHFGQAVR' | 'Complement factor H' |
| 7607 | 'CFH_No61' | 'TDCLSLPSFENAIPMGEKKDVYK' | 'Complement factor H' |
| 7608 | 'CFH_No62' | 'IIYKENER' | 'Complement factor H' |
| 7609 | 'CFH_No63' | 'TDCLSLPSFENAIPMGEK' | 'Complement factor H' |
| 7610 | 'CFH_No64' | 'DGWSAQPTCIK' | 'Complement factor H' |
| 7611 | 'CFH_No65' | 'HRTGDEITYQCR' | 'Complement factor H' |
| 7612 | 'CFH_No66' | 'CLPVTAPENGK' | 'Complement factor H' |
| 7613 | 'CFH_No67' | 'NGFYPATR' | 'Complement factor H' |

| | A | B | C |
|---|---|---|---|
| 1 | **Peptide Name** | **Peptide Sequence** | **Peptide Description** |
| 7614 | 'CFH_No68' | 'EKTKEEYGHSEVVEYYCNPR' | 'Complement factor H' |
| 7615 | 'CFH_No69' | 'AVYTCNEGYQLLGEINYR' | 'Complement factor H' |
| 7616 | 'CFH_No70' | 'LSYTCEGGFR' | 'Complement factor H' |
| 7617 | 'CFH_No71' | 'EQVQSCGPPPELLNGNVKEK' | 'Complement factor H' |
| 7618 | 'CFH_No72' | 'YYSYYCDEHFETPSGSYWDHIHCTQDGWSPAVPCLR' | 'Complement factor H' |
| 7619 | 'CFH_No73' | 'TKNDFTWFK' | 'Complement factor H' |
| 7620 | 'CFH_No74' | 'FVCNSGYKIEGDEEMHCSDDGFWSK' | 'Complement factor H' |
| 7621 | 'CFH_No75' | 'VSVLCQENYLIQEGEEITCKDGRWQSIPLCVEK' | 'Complement factor H' |
| 7622 | 'CFH_No76' | 'FSCKPGFTIVGPNSVQCYHFGLSPDLPICK' | 'Complement factor H' |
| 7623 | 'CFH_No77' | 'DVYKAGEQVTYTCATYYK' | 'Complement factor H' |
| 7624 | 'CFH_No78' | 'IVSSAMEPDR' | 'Complement factor H' |
| 7625 | 'CFH_No79' | 'HGGLYHENMR' | 'Complement factor H' |
| 7626 | 'CFH_No80' | 'ECDTDGWTNDIPICEVVK' | 'Complement factor H' |
| 7627 | 'CFH_No81' | 'CNMGYEYSERGDAVCTESGWRPLPSCEEK' | 'Complement factor H' |
| 7628 | 'CFH_No82' | 'DQYKVGEVLK' | 'Complement factor H' |
| 7629 | 'CFH_No83' | 'SSIDIENGFISESQYTYALK' | 'Complement factor H' |
| 7630 | 'CFH_No84' | 'KDVYKAGEQVTYTCATYYK' | 'Complement factor H' |
| 7631 | 'CFH_No85' | 'DGEKVSVLCQENYLIQEGEEITCKDGR' | 'Complement factor H' |
| 7632 | 'CFH_No86' | 'CTLKPCDYPDIKHGGLYHENMR' | 'Complement factor H' |
| 7633 | 'CFH_No87' | 'FVCNSGYK' | 'Complement factor H' |
| 7634 | 'CFH_No88' | 'SLGNVIMVCR' | 'Complement factor H' |
| 7635 | 'CFH_No89' | 'CVEISCKSPDVINGSPISQK' | 'Complement factor H' |
| 7636 | 'CFH_No90' | 'SPPEISHGVVAHMSDSYQYGEEVTYK' | 'Complement factor H' |
| 7637 | 'CFH_No91' | 'EYHFGQAVR' | 'Complement factor H' |
| 7638 | 'CFH_No92' | 'CKSSNLIILEEHLK' | 'Complement factor H' |
| 7639 | 'CFH_No93' | 'CNMGYEYSERGDAVCTESGWRPLPSCEEK' | 'Complement factor H' |
| 7640 | 'CFH_No94' | 'ECELPKIDVHLVPDRK' | 'Complement factor H' |
| 7641 | 'CFH_No95' | 'SSQESYAHGTKLSYTCEGGFR' | 'Complement factor H' |
| 7642 | 'CFH_No96' | 'SSNLIILEEHLKNKK' | 'Complement factor H' |
| 7643 | 'CFH_No97' | 'SLGNVIMVCR' | 'Complement factor H' |
| 7644 | 'CFH_No98' | 'IPCSQPPQIEHGTINSSR' | 'Complement factor H' |
| 7645 | 'CFH_No99' | 'DGEKVSVLCQENYLIQEGEEITCK' | 'Complement factor H' |
| 7646 | 'CFH_No100' | 'EEYGHSEVVEYYCNPR' | 'Complement factor H' |
| 7647 | 'CFH_No101' | 'CLPVTAPENGKIVSSAMEPDREYHFGQAVR' | 'Complement factor H' |
| 7648 | 'CFH_No102' | 'WTGRPTCR' | 'Complement factor H' |
| 7649 | 'CFH_No103' | 'WSHPPSCIKTDCLSLPSFENAIPMGEK' | 'Complement factor H' |
| 7650 | 'CFH_No104' | 'LGYVTADGETSGSITCGKDGWSAQPTCIK' | 'Complement factor H' |
| 7651 | 'CFH_No105' | 'CVEISCK' | 'Complement factor H' |
| 7652 | 'CFH_No106' | 'CLGEKWSHPPSCIK' | 'Complement factor H' |
| 7653 | 'CFH_No107' | 'ISEENETTCYMGK' | 'Complement factor H' |
| 7654 | 'CFH_No108' | 'SPPEISHGVVAHMSDSYQYGEEVTYKCFEGFGIDGPAIAK' | 'Complement factor H' |
| 7655 | 'CFH_No109' | 'HGGLYHENMR' | 'Complement factor H' |
| 7656 | 'CFH_No110' | 'CLPVTAPENGKIVSSAMEPDREYHFGQAVR' | 'Complement factor H' |
| 7657 | 'CFH_No111' | 'DGRWQSIPLCVEK' | 'Complement factor H' |

| | A | B | C |
|---|---|---|---|
| 1 | **Peptide Name** | **Peptide Sequence** | **Peptide Description** |
| 7658 | 'CFH_No112' | 'FLMKGPNK' | 'Complement factor H' |
| 7659 | 'CFH_No113' | 'TGESVEFVCKRGYR' | 'Complement factor H' |
| 7660 | 'CFH_No114' | 'SSIDIENGFISESQYTYALKEKAK' | 'Complement factor H' |
| 7661 | 'CFH_No115' | 'SPDVINGSPISQKIIYKENER' | 'Complement factor H' |
| 7662 | 'CFH_No116' | 'CTLKPCDYPDIKHGGLYHENMR' | 'Complement factor H' |
| 7663 | 'CFH_No117' | 'IEGDEEMHCSDDGFWSKEKPK' | 'Complement factor H' |
| 7664 | 'CFH_No118' | 'TDCLSLPSFENAIPMGEKKDVYK' | 'Complement factor H' |
| 7665 | 'CFH_No119' | 'WSSPPQCEGLPCKSPPEISHGVVAHMSDSYQYGEEVTYK' | 'Complement factor H' |
| 7666 | 'CFH_No120' | 'SSQESYAHGTK' | 'Complement factor H' |
| 7667 | 'CFH_No121' | 'NKKEFDHNSNIR' | 'Complement factor H' |
| 7668 | 'CFH_No122' | 'WSHPPSCIKTDCLSLPSFENAIPMGEK' | 'Complement factor H' |
| 7669 | 'CFH_No123' | 'IEGDEEMHCSDDGFWSKEKPK' | 'Complement factor H' |
| 7670 | 'CFH_No124' | 'GDAVCTESGWRPLPSCEEKSCDNPYIPNGDYSPLR' | 'Complement factor H' |
| 7671 | 'CFH_No125' | 'KEFDHNSNIR' | 'Complement factor H' |
| 7672 | 'FCGR3A_No1' | 'WVFKEEDPIHLR' | 'Low affinity immunoglobulin gamma Fc region receptor III-A' |
| 7673 | 'FCGR3A_No2' | 'AVVFLEPQWYR' | 'Low affinity immunoglobulin gamma Fc region receptor III-A' |
| 7674 | 'FCGR3A_No3' | 'VLEKDSVTLK' | 'Low affinity immunoglobulin gamma Fc region receptor III-A' |
| 7675 | 'FCGR3A_No4' | 'KYFHHNSDFYIPK' | 'Low affinity immunoglobulin gamma Fc region receptor III-A' |
| 7676 | 'FCGR3A_No5' | 'ATLKDSGSYFCR' | 'Low affinity immunoglobulin gamma Fc region receptor III-A' |
| 7677 | 'FCGR3A_No6' | 'YFHHNSDFYIPK' | 'Low affinity immunoglobulin gamma Fc region receptor III-A' |
| 7678 | 'FCGR3A_No7' | 'EEDPIHLR' | 'Low affinity immunoglobulin gamma Fc region receptor III-A' |
| 7679 | 'VIM_No1' | 'ISLPLPNFSSLNLR' | 'Vimentin' |
| 7680 | 'VIM_No2' | 'DGQVINETSQHHDDLE' | 'Vimentin' |
| 7681 | 'VIM_No3' | 'KVESLQEEIAFLK' | 'Vimentin' |
| 7682 | 'VIM_No4' | 'TNEKVELQELNDR' | 'Vimentin' |
| 7683 | 'SERPINF2_No1' | 'LQQVLHAGSGPCLPHLLSR' | 'Alpha-2-antiplasmin' |
| 7684 | 'SERPINF2_No2' | 'WFLLEQPEIQVAHFPFK' | 'Alpha-2-antiplasmin' |
| 7685 | 'SERPINF2_No3' | 'GFPIKEDFLEQSEQLFGAKPVSLTGK' | 'Alpha-2-antiplasmin' |
| 7686 | 'SERPINF2_No4' | 'GFPIKEDFLEQSEQLFGAK' | 'Alpha-2-antiplasmin' |
| 7687 | 'SERPINF2_No5' | 'LGNQEPGGQTALK' | 'Alpha-2-antiplasmin' |
| 7688 | 'SERPINF2_No6' | 'LCQDLGPGAFR' | 'Alpha-2-antiplasmin' |
| 7689 | 'SERPINF2_No7' | 'GDKLFGPDLK' | 'Alpha-2-antiplasmin' |
| 7690 | 'SERPINF2_No8' | 'DFLQSLK' | 'Alpha-2-antiplasmin' |
| 7691 | 'SERPINF2_No9' | 'NKFDPSLTQR' | 'Alpha-2-antiplasmin' |
| 7692 | 'SERPINF2_No10' | 'QEDDLANINQWVK' | 'Alpha-2-antiplasmin' |
| 7693 | 'SERPINF2_No11' | 'DSFHLDEQFTVPVEMMQAR' | 'Alpha-2-antiplasmin' |
| 7694 | 'SERPINF2_No12' | 'DSFHLDEQFTVPVEMMQAR' | 'Alpha-2-antiplasmin' |
| 7695 | 'SERPINF2_No13' | 'GISEQSLVVSGVQHQSTLELSEVGVEAAAATSIAMSR' | 'Alpha-2-antiplasmin' |
| 7696 | 'SERPINF2_No14' | 'PVSLTGKQEDDLANINQWVK' | 'Alpha-2-antiplasmin' |
| 7697 | 'SERPINF2_No15' | 'QLTSGPNQEQVSPLTLLK' | 'Alpha-2-antiplasmin' |
| 7698 | 'SERPINF2_No16' | 'ELKEQQDSPGNKDFLQSLK' | 'Alpha-2-antiplasmin' |
| 7699 | 'SERPINF2_No17' | 'HQMDLVATLSQLGLQELFQAPDLR' | 'Alpha-2-antiplasmin' |
| 7700 | 'SERPINF2_No18' | 'LGNQEPGGQTALKSPPGVCSRDPTPEQTHR' | 'Alpha-2-antiplasmin' |
| 7701 | 'SERPINF2_No19' | 'LGNQEPGGQTALKSPPGVCSR' | 'Alpha-2-antiplasmin' |

| | A | B | C |
|---|---|---|---|
| 1 | **Peptide Name** | **Peptide Sequence** | **Peptide Description** |
| 7702 | 'SERPINF2_No20' | 'SPPGVCSRDPTPEQTHR' | 'Alpha-2-antiplasmin' |
| 7703 | 'SERPINF2_No21' | 'DSFHLDEQFTVPVEMMQAR' | 'Alpha-2-antiplasmin' |
| 7704 | 'SERPINF2_No22' | 'SPPGVCSR' | 'Alpha-2-antiplasmin' |
| 7705 | 'SERPINF2_No23' | 'MSLSSFSVNR' | 'Alpha-2-antiplasmin' |
| 7706 | 'SERPINF2_No24' | 'EDFLEQSEQLFGAKPVSLTGK' | 'Alpha-2-antiplasmin' |
| 7707 | 'SERPINF2_No25' | 'DSFHLDEQFTVPVEMMQAR' | 'Alpha-2-antiplasmin' |
| 7708 | 'SERPINF2_No26' | 'QEDDLANINQWVK' | 'Alpha-2-antiplasmin' |
| 7709 | 'SERPINF2_No27' | 'QLTSGPNQEQVSPLTLLK' | 'Alpha-2-antiplasmin' |
| 7710 | 'SERPINF2_No28' | 'NPNPSAPR' | 'Alpha-2-antiplasmin' |
| 7711 | 'SERPINF2_No29' | 'EDFLEQSEQLFGAK' | 'Alpha-2-antiplasmin' |
| 7712 | 'SERPINF2_No30' | 'EQQDSPGNKDFLQSLK' | 'Alpha-2-antiplasmin' |
| 7713 | 'SERPINF2_No31' | 'GFPRGDKLFGPDLK' | 'Alpha-2-antiplasmin' |
| 7714 | 'SERPINF2_No32' | 'NKFDPSLTQRDSFHLDEQFTVPVEMMQAR' | 'Alpha-2-antiplasmin' |
| 7715 | 'SERPINF2_No33' | 'GISEQSLVVSGVQHQSTLELSEVGVEAAAATSIAMSR' | 'Alpha-2-antiplasmin' |
| 7716 | 'SERPINF2_No34' | 'DPTPEQTHRLAR' | 'Alpha-2-antiplasmin' |
| 7717 | 'SERPINF2_No35' | 'HQMDLVATLSQLGLQELFQAPDLR' | 'Alpha-2-antiplasmin' |
| 7718 | 'SERPINF2_No36' | 'ELKEQQDSPGNK' | 'Alpha-2-antiplasmin' |
| 7719 | 'SERPINF2_No37' | 'FDPSLTQR' | 'Alpha-2-antiplasmin' |
| 7720 | 'SERPINF2_No38' | 'LVPPMEEDYPQFGSPK' | 'Alpha-2-antiplasmin' |
| 7721 | 'SERPINF2_No39' | 'DPTPEQTHR' | 'Alpha-2-antiplasmin' |
| 7722 | 'SERPINF2_No40' | 'EQQDSPGNKDFLQSLKGFPR' | 'Alpha-2-antiplasmin' |
| 7723 | 'SERPINF2_No41' | 'DFLQSLKGFPR' | 'Alpha-2-antiplasmin' |
| 7724 | 'SERPINF2_No42' | 'GDKLFGPDLKLVPPMEEDYPQFGSPK' | 'Alpha-2-antiplasmin' |
| 7725 | 'F7_No1' | 'VAQVIIPSTYVPGTTNHDIALLR' | 'Coagulation factor VII' |
| 7726 | 'F7_No2' | 'FSLVSGWGQLLDR' | 'Coagulation factor VII' |
| 7727 | 'F7_No3' | 'NLIAVLGEHDLSEHDGDEQSRR' | 'Coagulation factor VII' |
| 7728 | 'F7_No4' | 'SEPRPGVLLR' | 'Coagulation factor VII' |
| 7729 | 'F7_No5' | 'LMTQDCLQQSR' | 'Coagulation factor VII' |
| 7730 | 'F7_No6' | 'LMTQDCLQQSR' | 'Coagulation factor VII' |
| 7731 | 'F7_No7' | 'NLIAVLGEHDLSEHDGDEQSR' | 'Coagulation factor VII' |
| 7732 | 'F7_No8' | 'NCETHKDDQLICVNENGGCEQYCSDHTGTKR' | 'Coagulation factor VII' |
| 7733 | 'F7_No9' | 'GATALELMVLNVPR' | 'Coagulation factor VII' |
| 7734 | 'F7_No10' | 'CHEGYSLLADGVSCTPTVEYPCGK' | 'Coagulation factor VII' |
| 7735 | 'F7_No11' | 'VSQYIEWLQK' | 'Coagulation factor VII' |
| 7736 | 'F7_No12' | 'GATALELMVLNVPR' | 'Coagulation factor VII' |
| 7737 | 'F7_No13' | 'NCETHKDDQLICVNENGGCEQYCSDHTGTK' | 'Coagulation factor VII' |
| 7738 | 'F7_No14' | 'LHQPVVLTDHVVPLCLPER' | 'Coagulation factor VII' |
| 7739 | 'KRT16_No1' | 'IIAATIENAQPILQIDNAR' | 'Keratin, type I cytoskeletal 16' |
| 7740 | 'KRT16_No2' | 'APSTYGGGLSVSSR' | 'Keratin, type I cytoskeletal 16' |
| 7741 | 'KRT16_No3' | 'TKYEHELALR' | 'Keratin, type I cytoskeletal 16' |
| 7742 | 'KRT16_No4' | 'GQTGGDVNVEMDAAPGVDLSR' | 'Keratin, type I cytoskeletal 16' |
| 7743 | 'KRT16_No5' | 'EVASNSELVQSSR' | 'Keratin, type I cytoskeletal 16' |
| 7744 | 'KRT16_No6' | 'GQTGGDVNVEMDAAPGVDLSR' | 'Keratin, type I cytoskeletal 16' |
| 7745 | 'KRT16_No7' | 'QRPSEIKDYSPYFK' | 'Keratin, type I cytoskeletal 16' |

| | A | B | C |
|---|---|---|---|
| 1 | **Peptide Name** | **Peptide Sequence** | **Peptide Description** |
| 7746 | 'KRT16_No8' | 'KNHEEEMLALR' | 'Keratin, type I cytoskeletal 16' |
| 7747 | 'KRT16_No9' | 'LLEGEDAHLSSQQASGQSYSSR' | 'Keratin, type I cytoskeletal 16' |
| 7748 | 'KRT16_No10' | 'TEELNKEVASNSELVQSSR' | 'Keratin, type I cytoskeletal 16' |
| 7749 | 'KRT16_No11' | 'DAETWFLSK' | 'Keratin, type I cytoskeletal 16' |
| 7750 | 'KRT16_No12' | 'QTVEADVNGLRR' | 'Keratin, type I cytoskeletal 16' |
| 7751 | 'KRT16_No13' | 'TDLEMQIEGLKEELAYLRK' | 'Keratin, type I cytoskeletal 16' |
| 7752 | 'KRT16_No14' | 'QTVEADVNGLRR' | 'Keratin, type I cytoskeletal 16' |
| 7753 | 'KRT16_No15' | 'ILNEMRDQYEQMAEK' | 'Keratin, type I cytoskeletal 16' |
| 7754 | 'KRT16_No16' | 'TDLEMQIEGLKEELAYLRK' | 'Keratin, type I cytoskeletal 16' |
| 7755 | 'KRT16_No17' | 'ILNEMRDQYEQMAEK' | 'Keratin, type I cytoskeletal 16' |
| 7756 | 'KRT16_No18' | 'QRPSEIKDYSPYFK' | 'Keratin, type I cytoskeletal 16' |
| 7757 | 'KRT16_No19' | 'ILNEMRDQYEQMAEK' | 'Keratin, type I cytoskeletal 16' |
| 7758 | 'KRT16_No20' | 'ILNEMRDQYEQMAEK' | 'Keratin, type I cytoskeletal 16' |
| 7759 | 'KRT16_No21' | 'VLQGLEIELQSQLSMK' | 'Keratin, type I cytoskeletal 16' |
| 7760 | 'KRT16_No22' | 'RLLEGEDAHLSSQQASGQSYSSR' | 'Keratin, type I cytoskeletal 16' |
| 7761 | 'IGFBP1_No1' | 'ALPGEQQPLHALTR' | 'Insulin-like growth factor-binding protein 1' |
| 7762 | 'IGFBP1_No2' | 'FYLPNCNK' | 'Insulin-like growth factor-binding protein 1' |
| 7763 | 'ACAA1_No1' | 'IAQFLSDIPETVPLSTVNR' | '3-ketoacyl-CoA thiolase, peroxisomal' |
| 7764 | 'ACAA1_No2' | 'DGSTTAGNSSQVSDGAAAILLAR' | '3-ketoacyl-CoA thiolase, peroxisomal' |
| 7765 | 'DBH_No1' | 'TPEGLTLLFK' | 'Dopamine beta-hydroxylase' |
| 7766 | 'DBH_No2' | 'VISTLEEPTPQCPTSQGR' | 'Dopamine beta-hydroxylase' |
| 7767 | 'DBH_No3' | 'SPAGPTVVSIGGGK' | 'Dopamine beta-hydroxylase' |
| 7768 | 'DBH_No4' | 'YFHLINR' | 'Dopamine beta-hydroxylase' |
| 7769 | 'DBH_No5' | 'AFYYPEEAGLAFGGPGSSR' | 'Dopamine beta-hydroxylase' |
| 7770 | 'DBH_No6' | 'HVLAAWALGAK' | 'Dopamine beta-hydroxylase' |
| 7771 | 'DBH_No7' | 'FQGEWNLQPLPK' | 'Dopamine beta-hydroxylase' |
| 7772 | 'DBH_No8' | 'LEVHYHNPLVIEGR' | 'Dopamine beta-hydroxylase' |
| 7773 | 'DBH_No9' | 'GQIHLDPQQDYQLLQVQR' | 'Dopamine beta-hydroxylase' |
| 7774 | 'DBH_No10' | 'SAVDAGFLQK' | 'Dopamine beta-hydroxylase' |
| 7775 | 'DBH_No11' | 'SPAGPTVVSIGGGKG' | 'Dopamine beta-hydroxylase' |
| 7776 | 'DBH_No12' | 'ETAFILTGYCTDK' | 'Dopamine beta-hydroxylase' |
| 7777 | 'DBH_No13' | 'EWEIVNQDNHYSPHFQEIR' | 'Dopamine beta-hydroxylase' |
| 7778 | 'DBH_No14' | 'DGREWEIVNQDNHYSPHFQEIR' | 'Dopamine beta-hydroxylase' |
| 7779 | 'DBH_No15' | 'VVSVHPGDVLITSCTYNTEDR' | 'Dopamine beta-hydroxylase' |
| 7780 | 'DBH_No16' | 'FNAGIMELGLVYTPVMAIPPR' | 'Dopamine beta-hydroxylase' |
| 7781 | 'DBH_No17' | 'KVVSVHPGDVLITSCTYNTEDR' | 'Dopamine beta-hydroxylase' |
| 7782 | 'GSTP1_No1' | 'ALPGQLKPFETLLSQNQGGK' | 'Glutathione S-transferase P' |
| 7783 | 'GSTP1_No2' | 'PPYTVVYFPVR' | 'Glutathione S-transferase P' |
| 7784 | 'GSTP1_No3' | 'FQDGDLTLYQSNTILR' | 'Glutathione S-transferase P' |
| 7785 | 'GSTP1_No4' | 'YISLIYTNYEAGKDDYVK' | 'Glutathione S-transferase P' |
| 7786 | 'GSTP1_No5' | 'MLLADQGQSWKEEVVTVETWQEGSLK' | 'Glutathione S-transferase P' |
| 7787 | 'GSTP1_No6' | 'AFLASPEYVNLPINGNGK' | 'Glutathione S-transferase P' |
| 7788 | 'GSTP1_No7' | 'MLLADQGQSWKEEVVTVETWQEGSLK' | 'Glutathione S-transferase P' |
| 7789 | 'LGALS1_No1' | 'FNAHGDANTIVCNSK' | 'Galectin-1' |

| | A | B | C |
|---|---|---|---|
| 1 | **Peptide Name** | **Peptide Sequence** | **Peptide Description** |
| 7790 | 'LGALS1_No2' | 'ACGLVASNLNLKPGECLR' | 'Galectin-1' |
| 7791 | 'LGALS1_No3' | 'LPDGYEFK' | 'Galectin-1' |
| 7792 | 'LGALS1_No4' | 'LNLEAINYMAADGDFK' | 'Galectin-1' |
| 7793 | 'LGALS1_No5' | 'SFVLNLGK' | 'Galectin-1' |
| 7794 | 'LGALS1_No6' | 'LPDGYEFKFPNR' | 'Galectin-1' |
| 7795 | 'LGALS1_No7' | 'DSNNLCLHFNPR' | 'Galectin-1' |
| 7796 | 'LGALS1_No8' | 'LNLEAINYMAADGDFK' | 'Galectin-1' |
| 7797 | 'SPARC_No1' | 'LHLDYIGPCK' | 'SPARC' |
| 7798 | 'SPARC_No2' | 'RLEAGDHPVELLAR' | 'SPARC' |
| 7799 | 'SPARC_No3' | 'NVLVTLYERDEDNNLLTEK' | 'SPARC' |
| 7800 | 'SPARC_No4' | 'APLIPMEHCTTR' | 'SPARC' |
| 7801 | 'SPARC_No5' | 'YIPPCLDSELTEFPLR' | 'SPARC' |
| 7802 | 'SPARC_No6' | 'TFDSSCHFFATK' | 'SPARC' |
| 7803 | 'SPARC_No7' | 'APLIPMEHCTTR' | 'SPARC' |
| 7804 | 'SPARC_No8' | 'NVLVTLYERDEDNNLLTEKQK' | 'SPARC' |
| 7805 | 'GSTM1' | 'LGLDFPNLPYLIDGAHK' | 'Glutathione S-transferase Mu 1' |
| 7806 | 'TPM1_No1' | 'HIAEDADRKYEEVAR' | 'Tropomyosin alpha-1 chain' |
| 7807 | 'TPM1_No2' | 'KLVIIESDLERAEER' | 'Tropomyosin alpha-1 chain' |
| 7808 | 'TPM1_No3' | 'KLVIIESDLER' | 'Tropomyosin alpha-1 chain' |
| 7809 | 'TPM1_No4' | 'AQKDEEKMEIQEIQLK' | 'Tropomyosin alpha-1 chain' |
| 7810 | 'TPM1_No5' | 'KATDAEADVASLNRR' | 'Tropomyosin alpha-1 chain' |
| 7811 | 'TPM1_No6' | 'ATDAEADVASLNRR' | 'Tropomyosin alpha-1 chain' |
| 7812 | 'TPM1_No7' | 'KATDAEADVASLNR' | 'Tropomyosin alpha-1 chain' |
| 7813 | 'PDGFRB_No1' | 'LLGEVGTLQFAELHR' | 'Platelet-derived growth factor receptor beta' |
| 7814 | 'PDGFRB_No2' | 'VTDPQLVVTLHEK' | 'Platelet-derived growth factor receptor beta' |
| 7815 | 'PDGFRB_No3' | 'LVEPVTDFLLDMPYHIR' | 'Platelet-derived growth factor receptor beta' |
| 7816 | 'PDGFRB_No4' | 'VLELSESHPDSGEQTVR' | 'Platelet-derived growth factor receptor beta' |
| 7817 | 'C1S_No1' | 'LLEVPEGR' | 'Complement C1s subcomponent' |
| 7818 | 'C1S_No2' | 'SSNNPHSPIVEEFQVPYNK' | 'Complement C1s subcomponent' |
| 7819 | 'C1S_No3' | 'QFGPYCGHGFPGPLNIETK' | 'Complement C1s subcomponent' |
| 7820 | 'C1S_No4' | 'VGATSFYSTCQSNGK' | 'Complement C1s subcomponent' |
| 7821 | 'C1S_No5' | 'GDSGGAFAVQDPNDKTK' | 'Complement C1s subcomponent' |
| 7822 | 'C1S_No6' | 'MLTPEHVFIHPGWK' | 'Complement C1s subcomponent' |
| 7823 | 'C1S_No7' | 'LPVAPLRK' | 'Complement C1s subcomponent' |
| 7824 | 'C1S_No8' | 'FYAAGLVSWGPQCGTYGLYTR' | 'Complement C1s subcomponent' |
| 7825 | 'C1S_No9' | 'VEKPTADAEAYVFTPNMICAGGEK' | 'Complement C1s subcomponent' |
| 7826 | 'C1S_No10' | 'SNALDIIFQTDLTGQK' | 'Complement C1s subcomponent' |
| 7827 | 'C1S_No11' | 'NYVDWIMK' | 'Complement C1s subcomponent' |
| 7828 | 'C1S_No12' | 'TNFDNDIALVR' | 'Complement C1s subcomponent' |
| 7829 | 'C1S_No13' | 'GDSGGAFAVQDPNDK' | 'Complement C1s subcomponent' |
| 7830 | 'C1S_No14' | 'GMDSCKGDSGGAFAVQDPNDKTK' | 'Complement C1s subcomponent' |
| 7831 | 'C1S_No15' | 'EPTMYVGSTSVQTSR' | 'Complement C1s subcomponent' |
| 7832 | 'C1S_No16' | 'VKNYVDWIMK' | 'Complement C1s subcomponent' |
| 7833 | 'C1S_No17' | 'SNALDIIFQTDLTGQKK' | 'Complement C1s subcomponent' |

| | A | B | C |
|---|---|---|---|
| 1 | **Peptide Name** | **Peptide Sequence** | **Peptide Description** |
| 7834 | 'C1S_No18' | 'IIGGSDADIK' | 'Complement C1s subcomponent' |
| 7835 | 'C1S_No19' | 'DVVQITCLDGFEVVEGR' | 'Complement C1s subcomponent' |
| 7836 | 'C1S_No20' | 'EVKVEKPTADAEAYVFTPNMICAGGEK' | 'Complement C1s subcomponent' |
| 7837 | 'C1S_No21' | 'YHGDPMPCPKEDTPNSVWEPAK' | 'Complement C1s subcomponent' |
| 7838 | 'C1S_No22' | 'GMDSCKGDSGGAFAVQDPNDKTK' | 'Complement C1s subcomponent' |
| 7839 | 'C1S_No23' | 'MLTPEHVFIHPGWK' | 'Complement C1s subcomponent' |
| 7840 | 'C1S_No24' | 'CVPVCGVPR' | 'Complement C1s subcomponent' |
| 7841 | 'C1S_No25' | 'GFQVVVTLR' | 'Complement C1s subcomponent' |
| 7842 | 'C1S_No26' | 'VEDPESTLFGSVIR' | 'Complement C1s subcomponent' |
| 7843 | 'C1S_No27' | 'VEKPTADAEAYVFTPNMICAGGEK' | 'Complement C1s subcomponent' |
| 7844 | 'C1S_No28' | 'TMQENSTPRED' | 'Complement C1s subcomponent' |
| 7845 | 'C1S_No29' | 'EDTPNSVWEPAK' | 'Complement C1s subcomponent' |
| 7846 | 'C1S_No30' | 'YHGDPMPCPK' | 'Complement C1s subcomponent' |
| 7847 | 'C1S_No31' | 'VKNYVDWIMK' | 'Complement C1s subcomponent' |
| 7848 | 'C1S_No32' | 'GMDSCKGDSGGAFAVQDPNDK' | 'Complement C1s subcomponent' |
| 7849 | 'C1S_No33' | 'CVPVCGVPREPFEEK' | 'Complement C1s subcomponent' |
| 7850 | 'C1S_No34' | 'QFGPYCGHGFPGPLNIETK' | 'Complement C1s subcomponent' |
| 7851 | 'C1S_No35' | 'CQPVDCGIPESIENGK' | 'Complement C1s subcomponent' |
| 7852 | 'C1S_No36' | 'LKCQPVDCGIPESIENGKVEDPESTLFGSVIR' | 'Complement C1s subcomponent' |
| 7853 | 'C1S_No37' | 'YHGDPMPCPKEDTPNSVWEPAKAK' | 'Complement C1s subcomponent' |
| 7854 | 'C1S_No38' | 'CVPVCGVPREPFEEKQR' | 'Complement C1s subcomponent' |
| 7855 | 'C1S_No39' | 'EVKVEKPTADAEAYVFTPNMICAGGEK' | 'Complement C1s subcomponent' |
| 7856 | 'C1S_No40' | 'LRYHGDPMPCPK' | 'Complement C1s subcomponent' |
| 7857 | 'C1S_No41' | 'YHGDPMPCPK' | 'Complement C1s subcomponent' |
| 7858 | 'C1S_No42' | 'EPTMYVGSTSVQTSR' | 'Complement C1s subcomponent' |
| 7859 | 'C1S_No43' | 'NYVDWIMK' | 'Complement C1s subcomponent' |
| 7860 | 'C1S_No44' | 'EDTPNSVWEPAKAK' | 'Complement C1s subcomponent' |
| 7861 | 'C1S_No45' | 'SKMLTPEHVFIHPGWK' | 'Complement C1s subcomponent' |
| 7862 | 'C1S_No46' | 'REDFDVEAADSAGNCLDSLVFVAGDR' | 'Complement C1s subcomponent' |
| 7863 | 'C1S_No47' | 'LLEVPEGRTNFDNDIALVR' | 'Complement C1s subcomponent' |
| 7864 | 'C1S_No48' | 'YHGDPMPCPKEDTPNSVWEPAK' | 'Complement C1s subcomponent' |
| 7865 | 'C1S_No49' | 'CQPVDCGIPESIENGKVEDPESTLFGSVIR' | 'Complement C1s subcomponent' |
| 7866 | 'C1S_No50' | 'TMQENSTPRED' | 'Complement C1s subcomponent' |
| 7867 | 'C1S_No51' | 'GMDSCKGDSGGAFAVQDPNDK' | 'Complement C1s subcomponent' |
| 7868 | 'C1S_No52' | 'PTADAEAYVFTPNMICAGGEK' | 'Complement C1s subcomponent' |
| 7869 | 'C1S_No53' | 'SSNNPHSPIVEEFQVPYNKLQVIFK' | 'Complement C1s subcomponent' |
| 7870 | 'C1S_No54' | 'SNALDIIFQTDLTGQKKGWK' | 'Complement C1s subcomponent' |
| 7871 | 'C1S_No55' | 'YHGDPMPCPKEDTPNSVWEPAKAK' | 'Complement C1s subcomponent' |
| 7872 | 'C1S_No56' | 'CEYQIRLEK' | 'Complement C1s subcomponent' |
| 7873 | 'LTA4H_No1' | 'LVVDLTDIDPDVAYSSVPYEK' | 'Leukotriene A-4 hydrolase' |
| 7874 | 'LTA4H_No2' | 'MQEVYNFNAINNSEIR' | 'Leukotriene A-4 hydrolase' |
| 7875 | 'LTA4H_No3' | 'WEDAIPLALK' | 'Leukotriene A-4 hydrolase' |
| 7876 | 'LTA4H_No4' | 'DGETPDPEDPSR' | 'Leukotriene A-4 hydrolase' |
| 7877 | 'LTA4H_No5' | 'SLSNVIAHEISHSWTGNLVTNK' | 'Leukotriene A-4 hydrolase' |

| | A | B | C |
|---|---|---|---|
| 1 | **Peptide Name** | **Peptide Sequence** | **Peptide Description** |
| 7878 | 'LTA4H_No6' | 'TLTGTAALTVQSQEDNLR' | 'Leukotriene A-4 hydrolase' |
| 7879 | 'LTA4H_No7' | 'EHPYLFSQCQAIHCR' | 'Leukotriene A-4 hydrolase' |
| 7880 | 'LTA4H_No8' | 'GSPMEISLPIALSK' | 'Leukotriene A-4 hydrolase' |
| 7881 | 'LTA4H_No9' | 'AILPCQDTPSVK' | 'Leukotriene A-4 hydrolase' |
| 7882 | 'LTA4H_No10' | 'SAYEFSETESMLK' | 'Leukotriene A-4 hydrolase' |
| 7883 | 'LTA4H_No11' | 'LTYTAEVSVPK' | 'Leukotriene A-4 hydrolase' |
| 7884 | 'LTA4H_No12' | 'PEIVDTCSLASPASVCR' | 'Leukotriene A-4 hydrolase' |
| 7885 | 'LTA4H_No13' | 'HFNALGGWGELQNSVK' | 'Leukotriene A-4 hydrolase' |
| 7886 | 'LTA4H_No14' | 'SAYEFSETESMLK' | 'Leukotriene A-4 hydrolase' |
| 7887 | 'ALDOC_No1' | 'GVVPLAGTDGETTTQGLDGLSER' | 'Fructose-bisphosphate aldolase C' |
| 7888 | 'ALDOC_No2' | 'VDKGVVPLAGTDGETTTQGLDGLSER' | 'Fructose-bisphosphate aldolase C' |
| 7889 | 'ALDOC_No3' | 'TPSALAILENANVLAR' | 'Fructose-bisphosphate aldolase C' |
| 7890 | 'C4A_No1' | 'HLVPGAPFLLQALVR' | 'Complement C4-A' |
| 7891 | 'C4A_No2' | 'ASAGLLGAHAAAITAYALTLTK' | 'Complement C4-A' |
| 7892 | 'C4A_No3' | 'VHYTVCIWR' | 'Complement C4-A' |
| 7893 | 'C4A_No4' | 'PVAFSVVPTAAAAVSLK' | 'Complement C4-A' |
| 7894 | 'C4A_No5' | 'GPEVQLVAHSPWLK' | 'Complement C4-A' |
| 7895 | 'C4A_No6' | 'LTVAAPPSGGPGFLSIERPDSRPPR' | 'Complement C4-A' |
| 7896 | 'C4A_No7' | 'GLEEELQFSLGSK' | 'Complement C4-A' |
| 7897 | 'C4A_No8' | 'LNMGITDLQGLR' | 'Complement C4-A' |
| 7898 | 'C4A_No9' | 'EPFLSCCQFAESLR' | 'Complement C4-A' |
| 7899 | 'C4A_No10' | 'LNMGITDLQGLR' | 'Complement C4-A' |
| 7900 | 'C4A_No11' | 'SFFPENWLWR' | 'Complement C4-A' |
| 7901 | 'C4A_No12' | 'LHLETDSLALVALGALDTALYAAGSK' | 'Complement C4-A' |
| 7902 | 'C4A_No13' | 'VDVQAGACEGKLELSVDGAK' | 'Complement C4-A' |
| 7903 | 'C4A_No14' | 'ECVGFEAVQEVPVGLVQPASATLYDYYNPERR' | 'Complement C4-A' |
| 7904 | 'C4A_No15' | 'YIYGKPVQGVAYVR' | 'Complement C4-A' |
| 7905 | 'C4A_No16' | 'EGAIHREELVYELNPLDHR' | 'Complement C4-A' |
| 7906 | 'C4A_No17' | 'VLSLAQEQVGGSPEK' | 'Complement C4-A' |
| 7907 | 'C4A_No18' | 'YVSHFETEGPHVLLYFDSVPTSR' | 'Complement C4-A' |
| 7908 | 'C4A_No19' | 'VDFTLSSERDFALLSLQVPLKDAK' | 'Complement C4-A' |
| 7909 | 'C4A_No20' | 'FACYYPR' | 'Complement C4-A' |
| 7910 | 'C4A_No21' | 'SCGLHQLLR' | 'Complement C4-A' |
| 7911 | 'C4A_No22' | 'LVNGQSHISLSK' | 'Complement C4-A' |
| 7912 | 'C4A_No23' | 'EELVYELNPLDHR' | 'Complement C4-A' |
| 7913 | 'C4A_No24' | 'DHAVDLIQK' | 'Complement C4-A' |
| 7914 | 'C4A_No25' | 'KADGSYAAWLSR' | 'Complement C4-A' |
| 7915 | 'C4A_No26' | 'LTVAAPPSGGPGFLSIER' | 'Complement C4-A' |
| 7916 | 'C4A_No27' | 'GLQDEDGYR' | 'Complement C4-A' |
| 7917 | 'C4A_No28' | 'PVQGVAYVR' | 'Complement C4-A' |
| 7918 | 'C4A_No29' | 'FGLLDEDGKK' | 'Complement C4-A' |
| 7919 | 'C4A_No30' | 'DFALLSLQVPLK' | 'Complement C4-A' |
| 7920 | 'C4A_No31' | 'KYVLPNFEVK' | 'Complement C4-A' |
| 7921 | 'C4A_No32' | 'AACAQLNDFLQEYGTQGCQV' | 'Complement C4-A' |

| | A | B | C |
|---|---|---|---|
| 1 | **Peptide Name** | **Peptide Sequence** | **Peptide Description** |
| 7922 | 'C4A_No33' | 'AVGSGATFSHYYYMILSR' | 'Complement C4-A' |
| 7923 | 'C4A_No34' | 'VQQPDCREPFLSCCQFAESLRK' | 'Complement C4-A' |
| 7924 | 'C4A_No35' | 'EPFLSCCQFAESLRK' | 'Complement C4-A' |
| 7925 | 'C4A_No36' | 'YVLPNFEVK' | 'Complement C4-A' |
| 7926 | 'C4A_No37' | 'QGSFQGGFR' | 'Complement C4-A' |
| 7927 | 'C4A_No38' | 'GLCVATPVQLR' | 'Complement C4-A' |
| 7928 | 'C4A_No39' | 'DSSTWLTAFVLK' | 'Complement C4-A' |
| 7929 | 'C4A_No40' | 'VDFTLSSERDFALLSLQVPLK' | 'Complement C4-A' |
| 7930 | 'C4A_No41' | 'SMQGGLVGNDETVALTAFVTIALHHGLAVFQDEGAEPLKQR' | 'Complement C4-A' |
| 7931 | 'C4A_No42' | 'GQIVFMNR' | 'Complement C4-A' |
| 7932 | 'C4A_No43' | 'TEQWSTLPPETK' | 'Complement C4-A' |
| 7933 | 'C4A_No44' | 'SMQGGLVGNDETVALTAFVTIALHHGLAVFQDEGAEPLK' | 'Complement C4-A' |
| 7934 | 'C4A_No45' | 'EMSGSPASGIPVK' | 'Complement C4-A' |
| 7935 | 'C4A_No46' | 'DFALLSLQVPLKDAK' | 'Complement C4-A' |
| 7936 | 'C4A_No47' | 'GQIVFMNR' | 'Complement C4-A' |
| 7937 | 'C4A_No48' | 'SMQGGLVGNDETVALTAFVTIALHHGLAVFQDEGAEPLK' | 'Complement C4-A' |
| 7938 | 'C4A_No49' | 'GQIVFMNREPK' | 'Complement C4-A' |
| 7939 | 'C4A_No50' | 'VDVQAGACEGK' | 'Complement C4-A' |
| 7940 | 'C4A_No51' | 'LGQYASPTAK' | 'Complement C4-A' |
| 7941 | 'C4A_No52' | 'ITPGKPYILTVPGHLDEMQLDIQAR' | 'Complement C4-A' |
| 7942 | 'C4A_No53' | 'AVGSGATFSHYYYMILSR' | 'Complement C4-A' |
| 7943 | 'C4A_No54' | 'AEFQDALEKLNMGITDLQGLR' | 'Complement C4-A' |
| 7944 | 'C4A_No55' | 'RHLVPGAPFLLQALVR' | 'Complement C4-A' |
| 7945 | 'C4A_No56' | 'QGSFQGGFR' | 'Complement C4-A' |
| 7946 | 'C4A_No57' | 'YRVFALDQK' | 'Complement C4-A' |
| 7947 | 'C4A_No58' | 'TTNIQGINLLFSSR' | 'Complement C4-A' |
| 7948 | 'C4A_No59' | 'PVAFSVVPTAAAAVSLKVVAR' | 'Complement C4-A' |
| 7949 | 'C4A_No60' | 'ALEILQEEDLIDEDDIPVR' | 'Complement C4-A' |
| 7950 | 'C4A_No61' | 'LLLFSPSVVHLGVPLSVGVQLQDVPR' | 'Complement C4-A' |
| 7951 | 'C4A_No62' | 'STQDTVIALDALSAYWIASHTTEER' | 'Complement C4-A' |
| 7952 | 'C4A_No63' | 'YLDKTEQWSTLPPETKDHAVDLIQK' | 'Complement C4-A' |
| 7953 | 'C4A_No64' | 'GCGEQTMIYLAPTLAASR' | 'Complement C4-A' |
| 7954 | 'C4A_No65' | 'PYILTVPGHLDEMQLDIQAR' | 'Complement C4-A' |
| 7955 | 'C4A_No66' | 'NGKVGLSGMAIADVTLLSGFHALR' | 'Complement C4-A' |
| 7956 | 'C4A_No67' | 'GRTLEIPGNSDPNMIPDGDFNSYVR' | 'Complement C4-A' |
| 7957 | 'C4A_No68' | 'TLEIPGNSDPNMIPDGDFNSYVR' | 'Complement C4-A' |
| 7958 | 'C4A_No69' | 'VQQPDCREPFLSCCQFAESLR' | 'Complement C4-A' |
| 7959 | 'C4A_No70' | 'VGLSGMAIADVTLLSGFHALR' | 'Complement C4-A' |
| 7960 | 'C4A_No71' | 'CSVFYGAPSK' | 'Complement C4-A' |
| 7961 | 'C4A_No72' | 'AEFQDALEKLNMGITDLQGLR' | 'Complement C4-A' |
| 7962 | 'C4A_No73' | 'MRPSTDTITVMVENSHGLR' | 'Complement C4-A' |
| 7963 | 'C4A_No74' | 'MRPSTDTITVMVENSHGLR' | 'Complement C4-A' |
| 7964 | 'C4A_No75' | 'VLQIEKEGAIHREELVYELNPLDHR' | 'Complement C4-A' |
| 7965 | 'C4A_No76' | 'YLDKTEQWSTLPPETK' | 'Complement C4-A' |

| | A | B | C |
|---|---|---|---|
| 1 | **Peptide Name** | **Peptide Sequence** | **Peptide Description** |
| 7966 | 'C4A_No77' | 'FGLLDEDGK' | 'Complement C4-A' |
| 7967 | 'C4A_No78' | 'TYNVLDMK' | 'Complement C4-A' |
| 7968 | 'C4A_No79' | 'ITQVLHFTK' | 'Complement C4-A' |
| 7969 | 'C4A_No80' | 'GHLFLQTDQPIYNPGQR' | 'Complement C4-A' |
| 7970 | 'C4A_No81' | 'AEFQDALEK' | 'Complement C4-A' |
| 7971 | 'C4A_No82' | 'TKGLCVATPVQLR' | 'Complement C4-A' |
| 7972 | 'C4A_No83' | 'SMQGGLVGNDETVALTAFVTIALHHGLAVFQDEGAEPLKQR' | 'Complement C4-A' |
| 7973 | 'C4A_No84' | 'EAPKVVEEQESR' | 'Complement C4-A' |
| 7974 | 'C4A_No85' | 'EFHLHLR' | 'Complement C4-A' |
| 7975 | 'C4A_No86' | 'RGHLFLQTDQPIYNPGQR' | 'Complement C4-A' |
| 7976 | 'C4A_No87' | 'NNVPCSPK' | 'Complement C4-A' |
| 7977 | 'C4A_No88' | 'RCSVFYGAPSK' | 'Complement C4-A' |
| 7978 | 'C4A_No89' | 'LLLFSPSVVHLGVPLSVGVQLQDVPRGQVVK' | 'Complement C4-A' |
| 7979 | 'C4A_No90' | 'LTSLSDRYVSHFETEGPHVLLYFDSVPTSR' | 'Complement C4-A' |
| 7980 | 'C4A_No91' | 'AINEKLGQYASPTAK' | 'Complement C4-A' |
| 7981 | 'C4A_No92' | 'ITPGKPYILTVPGHLDEMQLDIQAR' | 'Complement C4-A' |
| 7982 | 'C4A_No93' | 'VEYGFQVK' | 'Complement C4-A' |
| 7983 | 'C4A_No94' | 'LLATLCSAEVCQCAEGK' | 'Complement C4-A' |
| 7984 | 'C4A_No95' | 'VFREFHLHLR' | 'Complement C4-A' |
| 7985 | 'C4A_No96' | 'RCCQDGVTRLPMMR' | 'Complement C4-A' |
| 7986 | 'C4A_No97' | 'KKEVYMPSSIFQDDFVIPDISEPGTWK' | 'Complement C4-A' |
| 7987 | 'C4A_No98' | 'TYNVLDMK' | 'Complement C4-A' |
| 7988 | 'C4A_No99' | 'MRPSTDTITVMVENSHGLR' | 'Complement C4-A' |
| 7989 | 'C4A_No100' | 'RFEQLELR' | 'Complement C4-A' |
| 7990 | 'C4A_No101' | 'VGDTLNLNLR' | 'Complement C4-A' |
| 7991 | 'C4A_No102' | 'AINEKLGQYASPTAKR' | 'Complement C4-A' |
| 7992 | 'C4A_No103' | 'LGQYASPTAKR' | 'Complement C4-A' |
| 7993 | 'C4A_No104' | 'CCQDGVTRLPMMR' | 'Complement C4-A' |
| 7994 | 'C4A_No105' | 'VFALDQK' | 'Complement C4-A' |
| 7995 | 'C4A_No106' | 'VDFTLSSER' | 'Complement C4-A' |
| 7996 | 'C4A_No107' | 'EMSGSPASGIPVK' | 'Complement C4-A' |
| 7997 | 'C4A_No108' | 'GSVFLRNPSR' | 'Complement C4-A' |
| 7998 | 'C4A_No109' | 'GQIVFMNREPKR' | 'Complement C4-A' |
| 7999 | 'C4A_No110' | 'SCGLHQLLRGPEVQLVAHSPWLK' | 'Complement C4-A' |
| 8000 | 'C4A_No111' | 'GRTLEIPGNSDPNMIPDGDFNSYVR' | 'Complement C4-A' |
| 8001 | 'C4A_No112' | 'TEQWSTLPPETKDHAVDLIQK' | 'Complement C4-A' |
| 8002 | 'C4A_No113' | 'MRPSTDTITVMVENSHGLR' | 'Complement C4-A' |
| 8003 | 'C4A_No114' | 'ADGSYAAWLSR' | 'Complement C4-A' |
| 8004 | 'C4A_No115' | 'RFEQLELRPVLYNYLDK' | 'Complement C4-A' |
| 8005 | 'C4A_No116' | 'ANSFLGEK' | 'Complement C4-A' |
| 8006 | 'C4A_No117' | 'VTASDPLDTLGSEGALSPGGVASLLR' | 'Complement C4-A' |
| 8007 | 'C4A_No118' | 'VLSLAQEQVGGSPEKLQETSNWLLSQQQADGSFQDPCPVLDR' | 'Complement C4-A' |
| 8008 | 'C4A_No119' | 'GQIVFMNREPK' | 'Complement C4-A' |
| 8009 | 'C4A_No120' | 'FEQLELRPVLYNYLDK' | 'Complement C4-A' |

| | A | B | C |
|---|---|---|---|
| 1 | **Peptide Name** | **Peptide Sequence** | **Peptide Description** |
| 8010 | 'C4A_No121' | 'GPEVQLVAHSPWLKDSLSR' | 'Complement C4-A' |
| 8011 | 'C4A_No122' | 'VGLSGMAIADVTLLSGFHALR' | 'Complement C4-A' |
| 8012 | 'C4A_No123' | 'PSTDTITVMVENSHGLR' | 'Complement C4-A' |
| 8013 | 'C4A_No124' | 'LLATLCSAEVCQCAEGKCPR' | 'Complement C4-A' |
| 8014 | 'C4A_No125' | 'VLQIEKEGAIHR' | 'Complement C4-A' |
| 8015 | 'C4A_No126' | 'GCGEQTMIYLAPTLAASR' | 'Complement C4-A' |
| 8016 | 'C4A_No127' | 'DHAVDLIQKGYMR' | 'Complement C4-A' |
| 8017 | 'C4A_No128' | 'SHALQLNNR' | 'Complement C4-A' |
| 8018 | 'C4A_No129' | 'KKEVYMPSSIFQDDFVIPDISEPGTWK' | 'Complement C4-A' |
| 8019 | 'C4A_No130' | 'TTNIQGINLLFSSRR' | 'Complement C4-A' |
| 8020 | 'C4A_No131' | 'ADLEKLTSLSDR' | 'Complement C4-A' |
| 8021 | 'C4A_No132' | 'GLEEELQFSLGSKINVK' | 'Complement C4-A' |
| 8022 | 'C4A_No133' | 'FGLLDEDGKKTFFR' | 'Complement C4-A' |
| 8023 | 'C4A_No134' | 'RCCQDGVTRLPMMR' | 'Complement C4-A' |
| 8024 | 'C4A_No135' | 'EAPKVVEEQESRVHYTVCIWR' | 'Complement C4-A' |
| 8025 | 'C4A_No136' | 'LELSVDGAK' | 'Complement C4-A' |
| 8026 | 'C4A_No137' | 'QRAACAQLNDFLQEYGTQGCQV' | 'Complement C4-A' |
| 8027 | 'C4A_No138' | 'GLQDEDGYRMK' | 'Complement C4-A' |
| 8028 | 'C4A_No139' | 'ANSFLGEKASAGLLGAHAAAITAYALTLTK' | 'Complement C4-A' |
| 8029 | 'C4A_No140' | 'VVEEQESR' | 'Complement C4-A' |
| 8030 | 'C4A_No141' | 'LQETSNWLLSQQQADGSFQDPCPVLDR' | 'Complement C4-A' |
| 8031 | 'C4A_No142' | 'RCCQDGVTRLPMMR' | 'Complement C4-A' |
| 8032 | 'C4A_No143' | 'MKFACYYPR' | 'Complement C4-A' |
| 8033 | 'C4A_No144' | 'NGFKSHALQLNNR' | 'Complement C4-A' |
| 8034 | 'C4A_No145' | 'CCQDGVTRLPMMR' | 'Complement C4-A' |
| 8035 | 'C4A_No146' | 'CCQDGVTRLPMMR' | 'Complement C4-A' |
| 8036 | 'C4A_No147' | 'ITQVLHFTKDVK' | 'Complement C4-A' |
| 8037 | 'C4A_No148' | 'VEASISKANSFLGEK' | 'Complement C4-A' |
| 8038 | 'C4A_No149' | 'SCGLHQLLRGPEVQLVAHSPWLKDSLSR' | 'Complement C4-A' |
| 8039 | 'C4A_No150' | 'TLEIPGNSDPNMIPDGDFNSYVR' | 'Complement C4-A' |
| 8040 | 'C4A_No151' | 'GSFEFPVGDAVSK' | 'Complement C4-A' |
| 8041 | 'C4A_No152' | 'PYILTVPGHLDEMQLDIQAR' | 'Complement C4-A' |
| 8042 | 'C4A_No153' | 'LVNGQSHISLSKAEFQDALEK' | 'Complement C4-A' |
| 8043 | 'C4A_No154' | 'RCCQDGVTRLPMMR' | 'Complement C4-A' |
| 8044 | 'C4A_No155' | 'VVEEQESRVHYTVCIWR' | 'Complement C4-A' |
| 8045 | 'C4A_No156' | 'GLQDEDGYRMK' | 'Complement C4-A' |
| 8046 | 'C4A_No157' | 'KEVYMPSSIFQDDFVIPDISEPGTWK' | 'Complement C4-A' |
| 8047 | 'C4A_No158' | 'IQQFRKADGSYAAWLSR' | 'Complement C4-A' |
| 8048 | 'C4A_No159' | 'AEMADQASAWLTR' | 'Complement C4-A' |
| 8049 | 'C4A_No160' | 'GQIVFMNREPKR' | 'Complement C4-A' |
| 8050 | 'C4A_No161' | 'DKGQAGLQR' | 'Complement C4-A' |
| 8051 | 'C4A_No162' | 'CCQDGVTRLPMMR' | 'Complement C4-A' |
| 8052 | 'C4A_No163' | 'LFETKITQVLHFTKDVK' | 'Complement C4-A' |
| 8053 | 'C4A_No164' | 'DDPDAPLQPVTPLQLFEGRR' | 'Complement C4-A' |

| | A | B | C |
|---|---|---|---|
| 1 | **Peptide Name** | **Peptide Sequence** | **Peptide Description** |
| 8054 | 'C4A_No165' | 'FACYYPRVEYGFQVK' | 'Complement C4-A' |
| 8055 | 'C4A_No166' | 'VQQPDCR' | 'Complement C4-A' |
| 8056 | 'C4A_No167' | 'LFETKITQVLHFTK' | 'Complement C4-A' |
| 8057 | 'C4A_No168' | 'AAFRLFETK' | 'Complement C4-A' |
| 8058 | 'C4A_No169' | 'NNVPCSPKVDFTLSSER' | 'Complement C4-A' |
| 8059 | 'C4A_No170' | 'GSFEFPVGDAVSKVLQIEK' | 'Complement C4-A' |
| 8060 | 'C4A_No171' | 'KADGSYAAWLSRDSSTWLTAFVLK' | 'Complement C4-A' |
| 8061 | 'C4A_No172' | 'DHAVDLIQKGYMR' | 'Complement C4-A' |
| 8062 | 'C4A_No173' | 'KEVYMPSSIFQDDFVIPDISEPGTWK' | 'Complement C4-A' |
| 8063 | 'C4A_No174' | 'SRLLATLCSAEVCQCAEGK' | 'Complement C4-A' |
| 8064 | 'C4A_No175' | 'AEMADQASAWLTR' | 'Complement C4-A' |
| 8065 | 'C4A_No176' | 'NGKVGLSGMAIADVTLLSGFHALR' | 'Complement C4-A' |
| 8066 | 'C4A_No177' | 'VLREDSR' | 'Complement C4-A' |
| 8067 | 'C4A_No178' | 'GLESQTKLVNGQSHISLSK' | 'Complement C4-A' |
| 8068 | 'C4A_No179' | 'CCQDGVTR' | 'Complement C4-A' |
| 8069 | 'C4A_No180' | 'VTASDPLDTLGSEGALSPGGVASLLRLPR' | 'Complement C4-A' |
| 8070 | 'C4A_No181' | 'VEASISKANSFLGEKASAGLLGAHAAAITAYALTLTK' | 'Complement C4-A' |
| 8071 | 'C4A_No182' | 'RCCQDGVTR' | 'Complement C4-A' |
| 8072 | 'C4A_No183' | 'FEQLELR' | 'Complement C4-A' |
| 8073 | 'C4A_No184' | 'TEQWSTLPPETKDHAVDLIQKGYMR' | 'Complement C4-A' |
| 8074 | 'C4A_No185' | 'DDPDAPLQPVTPLQLFEGR' | 'Complement C4-A' |
| 8075 | 'C4A_No186' | 'VEYGFQVKVLREDSR' | 'Complement C4-A' |
| 8076 | 'C4A_No187' | 'MKFACYYPR' | 'Complement C4-A' |
| 8077 | 'C4A_No188' | 'SHKPLNMGK' | 'Complement C4-A' |
| 8078 | 'C4A_No189' | 'ECVGFEAVQEVPVGLVQPASATLYDYYNPER' | 'Complement C4-A' |
| 8079 | 'C4A_No190' | 'FSDGLESNSSTQFEVK' | 'Complement C4-A' |
| 8080 | 'C4A_No191' | 'QIRGLEEELQFSLGSKINVK' | 'Complement C4-A' |
| 8081 | 'C4A_No192' | 'QIRGLEEELQFSLGSK' | 'Complement C4-A' |
| 8082 | 'C4A_No193' | 'DSLSRTTNIQGINLLFSSR' | 'Complement C4-A' |
| 8083 | 'C4A_No194' | 'PSTDTITVMVENSHGLR' | 'Complement C4-A' |
| 8084 | 'C4A_No195' | 'ITQVLHFTKDVKAAANQMR' | 'Complement C4-A' |
| 8085 | 'C4A_No196' | 'SRLLATLCSAEVCQCAEGKCPR' | 'Complement C4-A' |
| 8086 | 'C4A_No197' | 'VDVQAGACEGKLELSVDGAKQYR' | 'Complement C4-A' |
| 8087 | 'C4B_No1' | 'GSSTWLTAFVLK' | 'Complement C4-B' |
| 8088 | 'C4B_No2' | 'AEMADQAAAWLTR' | 'Complement C4-B' |
| 8089 | 'C4B_No3' | 'ASAGLLGAHAAAITAYALTLTKAPADLR' | 'Complement C4-B' |
| 8090 | 'C4B_No4' | 'VLSLAQEQVGGSPEKLQETSNWLLSQQQADGSFQDLSPVIHR' | 'Complement C4-B' |
| 8091 | 'C4B_No5' | 'AEMADQAAAWLTR' | 'Complement C4-B' |
| 8092 | 'C4B_No6' | 'LQETSNWLLSQQQADGSFQDLSPVIHR' | 'Complement C4-B' |
| 8093 | 'C4B_No7' | 'PVAFSVVPTAATAVSLK' | 'Complement C4-B' |
| 8094 | 'C4B_No8' | 'ASSFLGEKASAGLLGAHAAAITAYALTLTK' | 'Complement C4-B' |
| 8095 | 'C4B_No9' | 'ASSFLGEKASAGLLGAHAAAITAYALTLTKAPADLR' | 'Complement C4-B' |
| 8096 | 'C4B_No10' | 'VEASISKASSFLGEK' | 'Complement C4-B' |
| 8097 | 'C4B_No11' | 'GVAHNNLMAMAQETGDNLYWGSVTGSQSNAVSPTPAPR' | 'Complement C4-B' |

| | A | B | C |
|---|---|---|---|
| 1 | **Peptide Name** | **Peptide Sequence** | **Peptide Description** |
| 8098 | 'H2AC11_No1' | 'NDEELNKLLGK' | 'Histone H2A type 1' |
| 8099 | 'H2AC11_No2' | 'HLQLAIRNDEELNKLLGK' | 'Histone H2A type 1' |
| 8100 | 'UBB_No1' | 'ESTLHLVLR' | 'Polyubiquitin-B' |
| 8101 | 'UBB_No2' | 'TITLEVEPSDTIENVK' | 'Polyubiquitin-B' |
| 8102 | 'UBB_No3' | 'TLSDYNIQKESTLHLVLR' | 'Polyubiquitin-B' |
| 8103 | 'UBB_No4' | 'TITLEVEPSDTIENVKAK' | 'Polyubiquitin-B' |
| 8104 | 'SAA1_No1' | 'FFGHGAEDSLADQAANEWGR' | 'Serum amyloid A-1 protein' |
| 8105 | 'SAA1_No2' | 'DPNHFRPAGLPEKY' | 'Serum amyloid A-1 protein' |
| 8106 | 'SAA1_No3' | 'SFFSFLGEAFDGAR' | 'Serum amyloid A-1 protein' |
| 8107 | 'SAA1_No4' | 'EANYIGSDKYFHAR' | 'Serum amyloid A-1 protein' |
| 8108 | 'SAA1_No5' | 'GPGGVWAAEAISDAR' | 'Serum amyloid A-1 protein' |
| 8109 | 'SAA1_No6' | 'SGKDPNHFRPAGLPEK' | 'Serum amyloid A-1 protein' |
| 8110 | 'SAA1_No7' | 'PAGLPEKY' | 'Serum amyloid A-1 protein' |
| 8111 | 'SAA1_No8' | 'RGPGGVWAAEAISDAR' | 'Serum amyloid A-1 protein' |
| 8112 | 'SAA1_No9' | 'RGPGGVWAAEAISDARENIQR' | 'Serum amyloid A-1 protein' |
| 8113 | 'SAA1_No10' | 'DPNHFRPAGLPEK' | 'Serum amyloid A-1 protein' |
| 8114 | 'SAA1_No11' | 'GPGGVWAAEAISDARENIQR' | 'Serum amyloid A-1 protein' |
| 8115 | 'SAA1_No12' | 'AYSDMREANYIGSDKYFHAR' | 'Serum amyloid A-1 protein' |
| 8116 | 'SAA1_No13' | 'AYSDMREANYIGSDKYFHAR' | 'Serum amyloid A-1 protein' |
| 8117 | 'SAA2_No1' | 'RGPGGAWAAEVISNAR' | 'Serum amyloid A-2 protein' |
| 8118 | 'SAA2_No2' | 'SGRDPNHFRPAGLPEK' | 'Serum amyloid A-2 protein' |
| 8119 | 'SAA2_No3' | 'GAEDSLADQAANKWGR' | 'Serum amyloid A-2 protein' |
| 8120 | 'SAA2_No4' | 'GPGGAWAAEVISNAR' | 'Serum amyloid A-2 protein' |
| 8121 | 'SAA2_No5' | 'GAEDSLADQAANK' | 'Serum amyloid A-2 protein' |
| 8122 | 'SULT1A3_No1' | 'DVAVSYYHFHR' | 'Sulfotransferase 1A3' |
| 8123 | 'SULT1A3_No2' | 'SLPEETMDFMVQHTSFK' | 'Sulfotransferase 1A3' |
| 8124 | 'SULT1A3_No3' | 'SHLPLALLPQTLLDQK' | 'Sulfotransferase 1A3' |
| 8125 | 'SULT1A3_No4' | 'SLPEETMDFMVQHTSFK' | 'Sulfotransferase 1A3' |
| 8126 | 'HSPA1A_No1' | 'ATAGDTHLGGEDFDNR' | 'Heat shock 70 kDa protein 1A' |
| 8127 | 'HSPA1A_No2' | 'NQVALNPQNTVFDAK' | 'Heat shock 70 kDa protein 1A' |
| 8128 | 'HSPA1A_No3' | 'LVNHFVEEFKR' | 'Heat shock 70 kDa protein 1A' |
| 8129 | 'HSPA1A_No4' | 'DAGVIAGLNVLR' | 'Heat shock 70 kDa protein 1A' |
| 8130 | 'HSPA1A_No5' | 'TTPSYVAFTDTER' | 'Heat shock 70 kDa protein 1A' |
| 8131 | 'HSPA1A_No6' | 'LLQDFFNGR' | 'Heat shock 70 kDa protein 1A' |
| 8132 | 'HSPA1A_No7' | 'IINEPTAAAIAYGLDR' | 'Heat shock 70 kDa protein 1A' |
| 8133 | 'HSPA1A_No8' | 'AFYPEEISSMVLTK' | 'Heat shock 70 kDa protein 1A' |
| 8134 | 'HSPA1A_No9' | 'NALESYAFNMK' | 'Heat shock 70 kDa protein 1A' |
| 8135 | 'HSPA1A_No10' | 'AAAIGIDLGTTYSCVGVFQHGK' | 'Heat shock 70 kDa protein 1A' |
| 8136 | 'HSPA1A_No11' | 'HWPFQVINDGDKPK' | 'Heat shock 70 kDa protein 1A' |
| 8137 | 'HSPA1A_No12' | 'TTPSYVAFTDTERLIGDAAK' | 'Heat shock 70 kDa protein 1A' |
| 8138 | 'HSPA1A_No13' | 'FEELCSDLFR' | 'Heat shock 70 kDa protein 1A' |
| 8139 | 'IGHA1_No29' | 'NTVYLQMNSLR' | 'Immunoglobulin alpha-2 heavy chain' |
| 8140 | 'IGHA1_No30' | 'NTVYLQMNSLR' | 'Immunoglobulin alpha-2 heavy chain' |
| 8141 | 'IGHA1_No31' | 'EVQLVETGGGLIQPGGSLR' | 'Immunoglobulin alpha-2 heavy chain' |

| | A | B | C |
|---|---|---|---|
| 1 | **Peptide Name** | **Peptide Sequence** | **Peptide Description** |
| 8142 | 'IGHA1_No32' | 'GTTVTVSSASPTSPK' | 'Immunoglobulin alpha-2 heavy chain' |
| 8143 | 'IGHA1_No33' | 'LSCAASGFTVSNHSMSWVR' | 'Immunoglobulin alpha-2 heavy chain' |
| 8144 | 'IGHA1_No34' | 'NTVYLQMNSLRAEDTAVYYCAR' | 'Immunoglobulin alpha-2 heavy chain' |
| 8145 | 'IGHA1_No35' | 'LSCAASGFTVSNHSMSWVR' | 'Immunoglobulin alpha-2 heavy chain' |
| 8146 | 'IGHD_No4' | 'NQFSLNLR' | 'Immunoglobulin delta heavy chain' |
| 8147 | 'IGHD_No5' | 'VTISVDTSR' | 'Immunoglobulin delta heavy chain' |
| 8148 | 'IGHD_No6' | 'LMALREPAAQAPVK' | 'Immunoglobulin delta heavy chain' |
| 8149 | 'IGHD_No7' | 'SLEVSYVTDHGPM' | 'Immunoglobulin delta heavy chain' |
| 8150 | 'IGHD_No8' | 'VPAPPSPQPATYTCVVSHEDSR' | 'Immunoglobulin delta heavy chain' |
| 8151 | 'IGHD_No9' | 'DAHLTWEVAGK' | 'Immunoglobulin delta heavy chain' |
| 8152 | 'IGHD_No10' | 'TGYYWGWIR' | 'Immunoglobulin delta heavy chain' |
| 8153 | 'IGHD_No11' | 'ATFTCFVVGSDLK' | 'Immunoglobulin delta heavy chain' |
| 8154 | 'IGHD_No12' | 'APDVFPIISGCR' | 'Immunoglobulin delta heavy chain' |
| 8155 | 'IGHD_No13' | 'TPECPSHTQPLGVYLLTPAVQDLWLR' | 'Immunoglobulin delta heavy chain' |
| 8156 | 'IGHD_No14' | 'ATFTCFVVGSDLKDAHLTWEVAGK' | 'Immunoglobulin delta heavy chain' |
| 8157 | 'IGHD_No15' | 'EVNTSGFAPARPPPQPGSTTFWAWSVLR' | 'Immunoglobulin delta heavy chain' |
| 8158 | 'IGHD_No16' | 'DSYYMTSSQLSTPLQQWR' | 'Immunoglobulin delta heavy chain' |
| 8159 | 'IGHD_No17' | 'EIFRWPESPK' | 'Immunoglobulin delta heavy chain' |
| 8160 | 'IGHD_No18' | 'DKATFTCFVVGSDLKDAHLTWEVAGK' | 'Immunoglobulin delta heavy chain' |
| 8161 | 'IGHD_No19' | 'SLWNAGTSVTCTLNHPSLPPQR' | 'Immunoglobulin delta heavy chain' |
| 8162 | 'IGHD_No20' | 'TFPEIQR' | 'Immunoglobulin delta heavy chain' |
| 8163 | 'IGHD_No21' | 'LMALREPAAQAPVK' | 'Immunoglobulin delta heavy chain' |
| 8164 | 'IGHD_No22' | 'DSYYMTSSQLSTPLQQWR' | 'Immunoglobulin delta heavy chain' |
| 8165 | 'IGHD_No23' | 'DAHLTWEVAGKVPTGGVEEGLLER' | 'Immunoglobulin delta heavy chain' |
| 8166 | 'IGHD_No24' | 'VPTGGVEEGLLER' | 'Immunoglobulin delta heavy chain' |
| 8167 | 'IGHD_No25' | 'RLQLQESGPGLVK' | 'Immunoglobulin delta heavy chain' |
| 8168 | 'IGHD_No26' | 'GLEWIGGVYYTGSIYYNPSLRGR' | 'Immunoglobulin delta heavy chain' |
| 8169 | 'IGHD_No27' | 'DKATFTCFVVGSDLK' | 'Immunoglobulin delta heavy chain' |
| 8170 | 'IGHD_No28' | 'SLEVSYVTDHGPM' | 'Immunoglobulin delta heavy chain' |
| 8171 | 'IGHD_No29' | 'RDSYYMTSSQLSTPLQQWR' | 'Immunoglobulin delta heavy chain' |
| 8172 | 'IGHD_No30' | 'RDSYYMTSSQLSTPLQQWR' | 'Immunoglobulin delta heavy chain' |
| 8173 | 'IGHE_No4' | 'QVQLVQSGAEVRK' | 'Immunoglobulin epsilon heavy chain' |
| 8174 | 'IGHE_No5' | 'PSPFDLFIR' | 'Immunoglobulin epsilon heavy chain' |
| 8175 | 'IGHE_No6' | 'GSGFFVFSR' | 'Immunoglobulin epsilon heavy chain' |
| 8176 | 'IGHE_No7' | 'QVQLVQSGAEVRKPGASVR' | 'Immunoglobulin epsilon heavy chain' |
| 8177 | 'IGHE_No8' | 'AVHEAASPSQTVQR' | 'Immunoglobulin epsilon heavy chain' |
| 8178 | 'IGHE_No9' | 'KSPTITCLVVDLAPSK' | 'Immunoglobulin epsilon heavy chain' |
| 8179 | 'IGHE_No10' | 'SDDSAVFYCAK' | 'Immunoglobulin epsilon heavy chain' |
| 8180 | 'IGHE_No11' | 'GVSAYLSRPSPFDLFIR' | 'Immunoglobulin epsilon heavy chain' |
| 8181 | 'IGHE_No12' | 'ASGYTFIDSYVGWIR' | 'Immunoglobulin epsilon heavy chain' |
| 8182 | 'IGHE_No13' | 'QVQLVQSGAEVR' | 'Immunoglobulin epsilon heavy chain' |
| 8183 | 'IGHE_No14' | 'QVQLVQSGAEVRKPGASVR' | 'Immunoglobulin epsilon heavy chain' |
| 8184 | 'IGHE_No15' | 'SPTITCLVVDLAPSK' | 'Immunoglobulin epsilon heavy chain' |
| 8185 | 'IGHE_No16' | 'AAPEVYAFATPEWPGSR' | 'Immunoglobulin epsilon heavy chain' |

| | A | B | C |
|---|---|---|---|
| 1 | **Peptide Name** | **Peptide Sequence** | **Peptide Description** |
| 8186 | 'IGHE_No17' | 'QVQLVQSGAEVR' | 'Immunoglobulin epsilon heavy chain' |
| 8187 | 'IGHE_No18' | 'QVQLVQSGAEVRKPGASVR' | 'Immunoglobulin epsilon heavy chain' |
| 8188 | 'IGHG1_No2' | 'GPSVFPLAPSSK' | 'Immunoglobulin gamma-1 heavy chain' |
| 8189 | 'IGHG1_No3' | 'FNWYVDGVEVHNAK' | 'Immunoglobulin gamma-1 heavy chain' |
| 8190 | 'IGHG1_No4' | 'VVSVLTVLHQDWLNGKEYK' | 'Immunoglobulin gamma-1 heavy chain' |
| 8191 | 'IGHG1_No5' | 'NQVSLTCLVK' | 'Immunoglobulin gamma-1 heavy chain' |
| 8192 | 'IGHG1_No6' | 'EPQVYTLPPSR' | 'Immunoglobulin gamma-1 heavy chain' |
| 8193 | 'IGHG1_No7' | 'WQQGNVFSCSVMHEALHNHYTQK' | 'Immunoglobulin gamma-1 heavy chain' |
| 8194 | 'IGHG1_No8' | 'TTPPVLDSDGSFFLYSKLTVDKSR' | 'Immunoglobulin gamma-1 heavy chain' |
| 8195 | 'IGHG1_No9' | 'VVSVLTVLHQDWLNGK' | 'Immunoglobulin gamma-1 heavy chain' |
| 8196 | 'IGHG1_No10' | 'THTCPPCPAPELLGGPSVFLFPPKPK' | 'Immunoglobulin gamma-1 heavy chain' |
| 8197 | 'IGHG1_No11' | 'WQQGNVFSCSVMHEALHNHYTQK' | 'Immunoglobulin gamma-1 heavy chain' |
| 8198 | 'IGHG1_No12' | 'THTCPPCPAPELLGGPSVFLFPPK' | 'Immunoglobulin gamma-1 heavy chain' |
| 8199 | 'IGHG1_No13' | 'TTPPVLDSDGSFFLYSK' | 'Immunoglobulin gamma-1 heavy chain' |
| 8200 | 'IGHG1_No14' | 'LSCAASGFTFSR' | 'Immunoglobulin gamma-1 heavy chain' |
| 8201 | 'IGHG1_No15' | 'DTLMISR' | 'Immunoglobulin gamma-1 heavy chain' |
| 8202 | 'IGHG1_No16' | 'SRWQQGNVFSCSVMHEALHNHYTQK' | 'Immunoglobulin gamma-1 heavy chain' |
| 8203 | 'IGHG1_No17' | 'GFYPSDIAVEWESNGQPENNYK' | 'Immunoglobulin gamma-1 heavy chain' |
| 8204 | 'IGHG1_No18' | 'DTLMISR' | 'Immunoglobulin gamma-1 heavy chain' |
| 8205 | 'IGHG1_No19' | 'VSNKALPAPIEK' | 'Immunoglobulin gamma-1 heavy chain' |
| 8206 | 'IGHG1_No20' | 'NTLYLNMNSLR' | 'Immunoglobulin gamma-1 heavy chain' |
| 8207 | 'IGHG1_No21' | 'STSGGTAALGCLVK' | 'Immunoglobulin gamma-1 heavy chain' |
| 8208 | 'IGHG1_No22' | 'TPEVTCVVVDVSHEDPEVKFNWYVDGVEVHNAK' | 'Immunoglobulin gamma-1 heavy chain' |
| 8209 | 'IGHG1_No23' | 'ALPAPIEK' | 'Immunoglobulin gamma-1 heavy chain' |
| 8210 | 'IGHG1_No24' | 'NTLYLNMNSLR' | 'Immunoglobulin gamma-1 heavy chain' |
| 8211 | 'IGHG1_No25' | 'SCDKTHTCPPCPAPELLGGPSVFLFPPKPK' | 'Immunoglobulin gamma-1 heavy chain' |
| 8212 | 'IGHG1_No26' | 'GPSVFPLAPSSKSTSGGTAALGCLVK' | 'Immunoglobulin gamma-1 heavy chain' |
| 8213 | 'IGHG1_No27' | 'QVQLVQSGGGVVQPGR' | 'Immunoglobulin gamma-1 heavy chain' |
| 8214 | 'IGHG1_No28' | 'VVSVLTVLHQDWLNGKEYKCK' | 'Immunoglobulin gamma-1 heavy chain' |
| 8215 | 'IGHG1_No29' | 'IRDTAMFFAHWGQGTLVTVSSASTK' | 'Immunoglobulin gamma-1 heavy chain' |
| 8216 | 'IGHG1_No30' | 'LTVDKSR' | 'Immunoglobulin gamma-1 heavy chain' |
| 8217 | 'IGHG1_No31' | 'LTVDKSRWQQGNVFSCSVMHEALHNHYTQK' | 'Immunoglobulin gamma-1 heavy chain' |
| 8218 | 'IGHG1_No32' | 'AKGQPREPQVYTLPPSR' | 'Immunoglobulin gamma-1 heavy chain' |
| 8219 | 'IGHG1_No33' | 'SRWQQGNVFSCSVMHEALHNHYTQK' | 'Immunoglobulin gamma-1 heavy chain' |
| 8220 | 'IGHG1_No34' | 'EPQVYTLPPSRDELTKNQVSLTCLVK' | 'Immunoglobulin gamma-1 heavy chain' |
| 8221 | 'IGHG1_No35' | 'TPEVTCVVVDVSHEDPEVK' | 'Immunoglobulin gamma-1 heavy chain' |
| 8222 | 'IGHG1_No36' | 'DTLMISRTPEVTCVVVDVSHEDPEVK' | 'Immunoglobulin gamma-1 heavy chain' |
| 8223 | 'IGHG1_No37' | 'GQPREPQVYTLPPSRDELTK' | 'Immunoglobulin gamma-1 heavy chain' |
| 8224 | 'IGHG1_No38' | 'EPQVYTLPPSRDELTK' | 'Immunoglobulin gamma-1 heavy chain' |
| 8225 | 'IGHG1_No39' | 'GQPREPQVYTLPPSR' | 'Immunoglobulin gamma-1 heavy chain' |
| 8226 | 'IGHG1_No40' | 'QVQLVQSGGGVVQPGR' | 'Immunoglobulin gamma-1 heavy chain' |
| 8227 | 'IGHG1_No41' | 'IRDTAMFFAHWGQGTLVTVSSASTK' | 'Immunoglobulin gamma-1 heavy chain' |
| 8228 | 'IGHG1_No42' | 'ALPAPIEKTISK' | 'Immunoglobulin gamma-1 heavy chain' |
| 8229 | 'IGHG1_No43' | 'DTLMISRTPEVTCVVVDVSHEDPEVK' | 'Immunoglobulin gamma-1 heavy chain' |

| | A | B | C |
|---|---|---|---|
| 1 | **Peptide Name** | **Peptide Sequence** | **Peptide Description** |
| 8230 | 'IGHG1_No44' | 'PKDTLMISR' | 'Immunoglobulin gamma-1 heavy chain' |
| 8231 | 'IGHG1_No45' | 'YTIHWVR' | 'Immunoglobulin gamma-1 heavy chain' |
| 8232 | 'IGHG1_No46' | 'TTPPVLDSDGSFFLYSKLTVDK' | 'Immunoglobulin gamma-1 heavy chain' |
| 8233 | 'IGHG1_No47' | 'NTLYLNMNSLRPEDTAVYYCAR' | 'Immunoglobulin gamma-1 heavy chain' |
| 8234 | 'IGHG1_No48' | 'FNWYVDGVEVHNAKTKPR' | 'Immunoglobulin gamma-1 heavy chain' |
| 8235 | 'IGHG1_No49' | 'DELTKNQVSLTCLVK' | 'Immunoglobulin gamma-1 heavy chain' |
| 8236 | 'IGHG1_No50' | 'QVQLVQSGGGVVQPGR' | 'Immunoglobulin gamma-1 heavy chain' |
| 8237 | 'IGHG1_No51' | 'QVQLVQSGGGVVQPGRSLR' | 'Immunoglobulin gamma-1 heavy chain' |
| 8238 | 'IGHG1_No52' | 'EEQYNSTYR' | 'Immunoglobulin gamma-1 heavy chain' |
| 8239 | 'IGHG1_No53' | 'QVQLVQSGGGVVQPGR' | 'Immunoglobulin gamma-1 heavy chain' |
| 8240 | 'IGHG1_No54' | 'LTVDKSRWQQGNVFSCSVMHEALHNHYTQK' | 'Immunoglobulin gamma-1 heavy chain' |
| 8241 | 'IGHG1_No55' | 'PEDTAVYYCAR' | 'Immunoglobulin gamma-1 heavy chain' |
| 8242 | 'IGHG1_No56' | 'DTAMFFAHWGQGTLVTVSSASTK' | 'Immunoglobulin gamma-1 heavy chain' |
| 8243 | 'IGHG1_No57' | 'NDSKNTLYLNMNSLR' | 'Immunoglobulin gamma-1 heavy chain' |
| 8244 | 'IGHG1_No58' | 'PKDTLMISR' | 'Immunoglobulin gamma-1 heavy chain' |
| 8245 | 'IGHG1_No59' | 'CKVSNKALPAPIEK' | 'Immunoglobulin gamma-1 heavy chain' |
| 8246 | 'IGHG1_No60' | 'NTLYLNMNSLRPEDTAVYYCAR' | 'Immunoglobulin gamma-1 heavy chain' |
| 8247 | 'IGHG1_No61' | 'NDSKNTLYLNMNSLR' | 'Immunoglobulin gamma-1 heavy chain' |
| 8248 | 'IGHM_No28' | 'GVALHRPDVYLLPPAR' | 'Immunoglobulin mu heavy chain' |
| 8249 | 'IGHM_No29' | 'FTCTVTHTDLPSPLK' | 'Immunoglobulin mu heavy chain' |
| 8250 | 'IGHM_No30' | 'LICQATGFSPR' | 'Immunoglobulin mu heavy chain' |
| 8251 | 'IGHM_No31' | 'YAATSQVLLPSK' | 'Immunoglobulin mu heavy chain' |
| 8252 | 'IGHM_No32' | 'STGKPTLYNVSLVMSDTAGTCY' | 'Immunoglobulin mu heavy chain' |
| 8253 | 'IGHM_No33' | 'QVGSGVTTDQVQAEAK' | 'Immunoglobulin mu heavy chain' |
| 8254 | 'IGHM_No34' | 'DGFFGNPR' | 'Immunoglobulin mu heavy chain' |
| 8255 | 'IGHM_No35' | 'YFAHSILTVSEEEWNTGQTYTCVVAHEALPNRVTER' | 'Immunoglobulin mu heavy chain' |
| 8256 | 'IGHM_No36' | 'STGKPTLYNVSLVMSDTAGTCY' | 'Immunoglobulin mu heavy chain' |
| 8257 | 'IGHM_No37' | 'LTCLVTDLTTYDSVTISWTR' | 'Immunoglobulin mu heavy chain' |
| 8258 | 'IGHM_No38' | 'VTSTLTIK' | 'Immunoglobulin mu heavy chain' |
| 8259 | 'IGHM_No39' | 'QKNVPLPVIAELPPK' | 'Immunoglobulin mu heavy chain' |
| 8260 | 'IGHM_No40' | 'EGKQVGSGVTTDQVQAEAK' | 'Immunoglobulin mu heavy chain' |
| 8261 | 'IGHM_No41' | 'VSVFVPPRDGFFGNPR' | 'Immunoglobulin mu heavy chain' |
| 8262 | 'IGHM_No42' | 'TVDKSTGKPTLYNVSLVMSDTAGTCY' | 'Immunoglobulin mu heavy chain' |
| 8263 | 'IGHM_No43' | 'NVPLPVIAELPPK' | 'Immunoglobulin mu heavy chain' |
| 8264 | 'IGHM_No44' | 'QVGSGVTTDQVQAEAKESGPTTYK' | 'Immunoglobulin mu heavy chain' |
| 8265 | 'IGHM_No45' | 'ESGPTTYK' | 'Immunoglobulin mu heavy chain' |
| 8266 | 'IGHM_No46' | 'GLTFQQNASSMCVPDQDTAIR' | 'Immunoglobulin mu heavy chain' |
| 8267 | 'IGHM_No47' | 'NVPLPVIAELPPKVSVFVPPR' | 'Immunoglobulin mu heavy chain' |
| 8268 | 'IGHM_No48' | 'NVPLPVIAELPPKVSVFVPPRDGFFGNPR' | 'Immunoglobulin mu heavy chain' |
| 8269 | 'IGHM_No49' | 'YVTSAPMPEPQAPGR' | 'Immunoglobulin mu heavy chain' |
| 8270 | 'IGHM_No50' | 'GEPLSPEKYVTSAPMPEPQAPGR' | 'Immunoglobulin mu heavy chain' |
| 8271 | 'IGHM_No51' | 'ESATITCLVTGFSPADVFVQWMQR' | 'Immunoglobulin mu heavy chain' |
| 8272 | 'IGHM_No52' | 'GVALHRPDVYLLPPAREQLNLR' | 'Immunoglobulin mu heavy chain' |
| 8273 | 'IGHM_No53' | 'SKLICQATGFSPR' | 'Immunoglobulin mu heavy chain' |

| | A | B | C |
|---|---|---|---|
| 1 | **Peptide Name** | **Peptide Sequence** | **Peptide Description** |
| 8274 | 'IGHM_No54' | 'FTCTVTHTDLPSPLKQTISRPK' | 'Immunoglobulin mu heavy chain' |
| 8275 | 'IGHM_No55' | 'QVGSGVTTDQVQAEAKESGPTTYK' | 'Immunoglobulin mu heavy chain' |
| 8276 | 'IGHM_No56' | 'VFAIPPSFASIFLTK' | 'Immunoglobulin mu heavy chain' |
| 8277 | 'IGHM_No57' | 'YFAHSILTVSEEEWNTGQTYTCVVAHEALPNR' | 'Immunoglobulin mu heavy chain' |
| 8278 | 'IGHM_No58' | 'YVTSAPMPEPQAPGR' | 'Immunoglobulin mu heavy chain' |
| 8279 | 'IGHM_No59' | 'QVGSGVTTDQVQAEAK' | 'Immunoglobulin mu heavy chain' |
| 8280 | 'IGHM_No60' | 'EGKQVGSGVTTDQVQAEAKESGPTTYK' | 'Immunoglobulin mu heavy chain' |
| 8281 | 'IGHM_No61' | 'QTISRPK' | 'Immunoglobulin mu heavy chain' |
| 8282 | 'IGHM_No62' | 'QKNVPLPVIAELPPK' | 'Immunoglobulin mu heavy chain' |
| 8283 | 'IGHM_No63' | 'VSVFVPPR' | 'Immunoglobulin mu heavy chain' |
| 8284 | 'IGHM_No64' | 'ESGPTTYKVTSTLTIK' | 'Immunoglobulin mu heavy chain' |
| 8285 | 'IGHM_No65' | 'QTISRPK' | 'Immunoglobulin mu heavy chain' |
| 8286 | 'IGHM_No66' | 'DGFFGNPRK' | 'Immunoglobulin mu heavy chain' |
| 8287 | 'IGHM_No67' | 'GLTFQQNASSMCVPDQDTAIR' | 'Immunoglobulin mu heavy chain' |
| 8288 | 'IGHM_No68' | 'PTLYNVSLVMSDTAGTCY' | 'Immunoglobulin mu heavy chain' |
| 8289 | 'IGHM_No69' | 'QVGSGVTTDQVQAEAKESGPTTYKVTSTLTIK' | 'Immunoglobulin mu heavy chain' |
| 8290 | 'IGHM_No70' | 'GGKYAATSQVLLPSK' | 'Immunoglobulin mu heavy chain' |
| 8291 | 'IGHM_No71' | 'TVDKSTGKPTLYNVSLVMSDTAGTCY' | 'Immunoglobulin mu heavy chain' |
| 8292 | 'IGHM_No72' | 'GFPSVLRGGK' | 'Immunoglobulin mu heavy chain' |
| 8293 | 'IGHM_No73' | 'PDVYLLPPAR' | 'Immunoglobulin mu heavy chain' |
| 8294 | 'IGHM_No74' | 'VSVFVPPRDGFFGNPRK' | 'Immunoglobulin mu heavy chain' |
| 8295 | 'IGHM_No75' | 'FTCTVTHTDLPSPLKQTISR' | 'Immunoglobulin mu heavy chain' |
| 8296 | 'IGHM_No76' | 'VFAIPPSFASIFLTKSTK' | 'Immunoglobulin mu heavy chain' |
| 8297 | 'IGHM_No77' | 'QVGSGVTTDQVQAEAKESGPTTYKVTSTLTIK' | 'Immunoglobulin mu heavy chain' |
| 8298 | 'IGHM_No78' | 'ESDWLGESMFTCR' | 'Immunoglobulin mu heavy chain' |
| 8299 | 'IGHM_No79' | 'ESATITCLVTGFSPADVFVQWMQR' | 'Immunoglobulin mu heavy chain' |
| 8300 | 'IGHM_No80' | 'QVTLTESGPALVKPK' | 'Immunoglobulin mu heavy chain' |
| 8301 | 'IGK_No1' | 'SGTASVVCLLNNFYPR' | 'Immunoglobulin kappa light chain' |
| 8302 | 'IGK_No2' | 'VDNALQSGNSQESVTEQDSK' | 'Immunoglobulin kappa light chain' |
| 8303 | 'IGK_No3' | 'DSTYSLSSTLTLSK' | 'Immunoglobulin kappa light chain' |
| 8304 | 'IGK_No4' | 'DIQMTQSPSTLSASVGDR' | 'Immunoglobulin kappa light chain' |
| 8305 | 'IGK_No5' | 'VQWKVDNALQSGNSQESVTEQDSK' | 'Immunoglobulin kappa light chain' |
| 8306 | 'IGK_No6' | 'VDNALQSGNSQESVTEQDSKDSTYSLSSTLTLSKADYEK' | 'Immunoglobulin kappa light chain' |
| 8307 | 'IGK_No7' | 'ASSLESGVPSR' | 'Immunoglobulin kappa light chain' |
| 8308 | 'IGK_No8' | 'SFNRGEC' | 'Immunoglobulin kappa light chain' |
| 8309 | 'IGK_No9' | 'HKVYACEVTHQGLSSPVTK' | 'Immunoglobulin kappa light chain' |
| 8310 | 'IGK_No10' | 'LLMYKASSLESGVPSR' | 'Immunoglobulin kappa light chain' |
| 8311 | 'IGK_No11' | 'GTVAAPSVFIFPPSDEQLK' | 'Immunoglobulin kappa light chain' |
| 8312 | 'IGK_No12' | 'SGTASVVCLLNNFYPREAK' | 'Immunoglobulin kappa light chain' |
| 8313 | 'IGK_No13' | 'VQWKVDNALQSGNSQESVTEQDSKDSTYSLSSTLTLSK' | 'Immunoglobulin kappa light chain' |
| 8314 | 'IGK_No14' | 'ADYEKHKVYACEVTHQGLSSPVTK' | 'Immunoglobulin kappa light chain' |
| 8315 | 'IGK_No15' | 'DIQMTQSPSTLSASVGDRVTITCR' | 'Immunoglobulin kappa light chain' |
| 8316 | 'IGK_No16' | 'ASQSINTWLAWYQQK' | 'Immunoglobulin kappa light chain' |
| 8317 | 'IGK_No17' | 'VYACEVTHQGLSSPVTK' | 'Immunoglobulin kappa light chain' |

| | A | B | C |
|---|---|---|---|
| 1 | **Peptide Name** | **Peptide Sequence** | **Peptide Description** |
| 8318 | 'IGK_No18' | 'ASQSINTWLAWYQQKPGK' | 'Immunoglobulin kappa light chain' |
| 8319 | 'IGK_No19' | 'DIQMTQSPSTLSASVGDR' | 'Immunoglobulin kappa light chain' |
| 8320 | 'IGK_No20' | 'VDNALQSGNSQESVTEQDSKDSTYSLSSTLTLSK' | 'Immunoglobulin kappa light chain' |
| 8321 | 'IGK_No21' | 'DSTYSLSSTLTLSKADYEK' | 'Immunoglobulin kappa light chain' |
| 8322 | 'IGK_No22' | 'VYACEVTHQGLSSPVTKSFNR' | 'Immunoglobulin kappa light chain' |
| 8323 | 'IGK_No23' | 'VEVKGTVAAPSVFIFPPSDEQLK' | 'Immunoglobulin kappa light chain' |
| 8324 | 'IGK_No24' | 'DIQMTQSPSTLSASVGDRVTITCR' | 'Immunoglobulin kappa light chain' |
| 8325 | 'IGK_No25' | 'DSTYSLSSTLTLSKADYEKHK' | 'Immunoglobulin kappa light chain' |
| 8326 | 'IGK_No26' | 'LLMYKASSLESGVPSR' | 'Immunoglobulin kappa light chain' |
| 8327 | 'IGK_No27' | 'VYACEVTHQGLSSPVTKSFNRGEC' | 'Immunoglobulin kappa light chain' |
| 8328 | 'IGK_No28' | 'DIQMTQSPSTLSASVGDR' | 'Immunoglobulin kappa light chain' |
| 8329 | 'IGK_No29' | 'DIQMTQSPSTLSASVGDR' | 'Immunoglobulin kappa light chain' |
| 8330 | 'IGK_No30' | 'SGTASVVCLLNNFYPREAKVQWK' | 'Immunoglobulin kappa light chain' |
| 8331 | 'IGK_No31' | 'ADYEKHK' | 'Immunoglobulin kappa light chain' |
| 8332 | 'IGL1' | 'YAASSYLSLTPEQWK' | 'Immunoglobulin lambda-1 light chain' |
| 8333 | 'IGL2' | 'SYSCQVTHEGSTVEK' | 'Immunoglobulin lambda-1 light chain' |
| 8334 | 'IGL3' | 'ATLVCLISDFYPGAVTVAWK' | 'Immunoglobulin lambda-1 light chain' |
| 8335 | 'IGL4' | 'VTVLGQPK' | 'Immunoglobulin lambda-1 light chain' |
| 8336 | 'IGL5' | 'QSNNKYAASSYLSLTPEQWK' | 'Immunoglobulin lambda-1 light chain' |
| 8337 | 'IGL6' | 'QSNNKYAASSYLSLTPEQWK' | 'Immunoglobulin lambda-1 light chain' |
| 8338 | 'IGL7' | 'SYSCQVTHEGSTVEKTVAPTECS' | 'Immunoglobulin lambda-1 light chain' |
| 8339 | 'IGL8' | 'ANPTVTLFPPSSEELQANK' | 'Immunoglobulin lambda-1 light chain' |
| 8340 | 'IGL9' | 'ADGSPVKAGVETTKPSK' | 'Immunoglobulin lambda-1 light chain' |
| 8341 | 'IGL10' | 'RPSGVPDR' | 'Immunoglobulin lambda-1 light chain' |
| 8342 | 'IGL11' | 'AGVETTKPSK' | 'Immunoglobulin lambda-1 light chain' |
| 8343 | 'IGL12' | 'RPSGVPDRFSGSK' | 'Immunoglobulin lambda-1 light chain' |
| 8344 | 'IGL13' | 'SHRSYSCQVTHEGSTVEK' | 'Immunoglobulin lambda-1 light chain' |
| 8345 | 'IGL14' | 'ATLVCLISDFYPGAVTVAWKADGSPVK' | 'Immunoglobulin lambda-1 light chain' |
| 8346 | 'IGL15' | 'YAASSYLSLTPEQWKSHR' | 'Immunoglobulin lambda-1 light chain' |
| 8347 | 'IGL16' | 'VIIYEVNKRPSGVPDR' | 'Immunoglobulin lambda-1 light chain' |
| 8348 | 'IGL17' | 'VIIYEVNK' | 'Immunoglobulin lambda-1 light chain' |
| 8349 | 'IGL18' | 'TVAPTECS' | 'Immunoglobulin lambda-1 light chain' |
| 8350 | 'IGL19' | 'SHRSYSCQVTHEGSTVEKTVAPTECS' | 'Immunoglobulin lambda-1 light chain' |
| 8351 | 'IGL20' | 'PSGVPDRFSGSK' | 'Immunoglobulin lambda-1 light chain' |
| 8352 | 'IGL21' | 'QSNNKYAASSYLSLTPEQWKSHR' | 'Immunoglobulin lambda-1 light chain' |
| 8353 | 'IGL22' | 'YAASSYLSLTPEQWKSHRSYSCQVTHEGSTVEK' | 'Immunoglobulin lambda-1 light chain' |
| 8354 | 'IGLC3_No1' | 'AAPSVTLFPPSSEELQANK' | 'Immunoglobulin lambda constant 3' |
| 8355 | 'IGLC3_No2' | 'AGVETTTPSK' | 'Immunoglobulin lambda constant 3' |
| 8356 | 'IGLC3_No3' | 'ADSSPVKAGVETTTPSK' | 'Immunoglobulin lambda constant 3' |
| 8357 | 'IGLC3_No4' | 'ATLVCLISDFYPGAVTVAWKADSSPVK' | 'Immunoglobulin lambda constant 3' |
| 8358 | 'IGLC3_No5' | 'SHKSYSCQVTHEGSTVEK' | 'Immunoglobulin lambda constant 3' |
| 8359 | 'IGLC3_No6' | 'ADSSPVKAGVETTTPSKQSNNK' | 'Immunoglobulin lambda constant 3' |
| 8360 | 'IGLC3_No7' | 'SHKSYSCQVTHEGSTVEKTVAPTECS' | 'Immunoglobulin lambda constant 3' |
| 8361 | 'IGLC3_No8' | 'ATLVCLISDFYPGAVTVAWKADSSPVKAGVETTTPSK' | 'Immunoglobulin lambda constant 3' |

| | A | B | C |
|---|---|---|---|
| 1 | **Peptide Name** | **Peptide Sequence** | **Peptide Description** |
| 8362 | 'IGLC3_No9' | 'AGVETTTPSKQSNNKYAASSYLSLTPEQWK' | 'Immunoglobulin lambda constant 3' |
| 8363 | 'IGLC3_No10' | 'YAASSYLSLTPEQWKSHK' | 'Immunoglobulin lambda constant 3' |
| 8364 | 'IGLC3_No11' | 'AGVETTTPSKQSNNK' | 'Immunoglobulin lambda constant 3' |
| 8365 | 'IGLC3_No12' | 'YAASSYLSLTPEQWKSHKSYSCQVTHEGSTVEK' | 'Immunoglobulin lambda constant 3' |
| 8366 | 'IGHV1-8_No1' | 'NTSISTAYMELSSLR' | 'Immunoglobulin heavy variable 1-8' |
| 8367 | 'IGHV1-8_No2' | 'ASGYTFTSYDINWVR' | 'Immunoglobulin heavy variable 1-8' |
| 8368 | 'IGHV1-8_No3' | 'NTSISTAYMELSSLR' | 'Immunoglobulin heavy variable 1-8' |
| 8369 | 'IGHV1-8_No4' | 'QATGQGLEWMGWMNPNSGNTGYAQK' | 'Immunoglobulin heavy variable 1-8' |
| 8370 | 'IGHV1-8_No5' | 'QATGQGLEWMGWMNPNSGNTGYAQK' | 'Immunoglobulin heavy variable 1-8' |
| 8371 | 'IGHV1-8_No6' | 'QATGQGLEWMGWMNPNSGNTGYAQK' | 'Immunoglobulin heavy variable 1-8' |
| 8372 | 'IGHV3-30-3_No1' | 'NTLYLQMNSLR' | 'Immunoglobulin heavy variable 3-30-3' |
| 8373 | 'IGHV3-30-3_No2' | 'NTLYLQMNSLR' | 'Immunoglobulin heavy variable 3-30-3' |
| 8374 | 'IGHV3-30-3_No3' | 'LSCAASGFTFSSYAMHWVR' | 'Immunoglobulin heavy variable 3-30-3' |
| 8375 | 'IGHV3-30-3_No4' | 'LSCAASGFTFSSYAMHWVR' | 'Immunoglobulin heavy variable 3-30-3' |
| 8376 | 'IGHV3-30-3_No5' | 'GLEWVAVISYDGSNK' | 'Immunoglobulin heavy variable 3-30-3' |
| 8377 | 'IGHV3-30-3_No6' | 'DNSKNTLYLQMNSLR' | 'Immunoglobulin heavy variable 3-30-3' |
| 8378 | 'IGHV3-30-3_No7' | 'YYADSVK' | 'Immunoglobulin heavy variable 3-30-3' |
| 8379 | 'IGHV3-30-3_No8' | 'AEDTAVYYCAR' | 'Immunoglobulin heavy variable 3-30-3' |
| 8380 | 'IGHV3-30-3_No9' | 'DNSKNTLYLQMNSLR' | 'Immunoglobulin heavy variable 3-30-3' |
| 8381 | 'IGHV3-30-3_No10' | 'GRFTISR' | 'Immunoglobulin heavy variable 3-30-3' |
| 8382 | 'IGHV3-30-3_No11' | 'NTLYLQMNSLRAEDTAVYYCAR' | 'Immunoglobulin heavy variable 3-30-3' |
| 8383 | 'IGHV3-30-3_No12' | 'DNSKNTLYLQMNSLRAEDTAVYYCAR' | 'Immunoglobulin heavy variable 3-30-3' |
| 8384 | 'IGHV3-30-3_No13' | 'DNSKNTLYLQMNSLRAEDTAVYYCAR' | 'Immunoglobulin heavy variable 3-30-3' |
| 8385 | 'IGHV3-30-3_No14' | 'YYADSVKGR' | 'Immunoglobulin heavy variable 3-30-3' |
| 8386 | 'IGHV3-30-3_No15' | 'NTLYLQMNSLRAEDTAVYYCAR' | 'Immunoglobulin heavy variable 3-30-3' |
| 8387 | 'IGHV4-38-2' | 'GLEWIGSIYHSGSTYYNPSLK' | 'Immunoglobulin heavy variable 4-38-2' |
| 8388 | 'RNASE2_No1' | 'RDPPQYPVVPVHLDR' | 'Non-secretory ribonuclease' |
| 8389 | 'RNASE2_No2' | 'YAQTPANMFYIVACDNR' | 'Non-secretory ribonuclease' |
| 8390 | 'HLA-C_No1' | 'AYLEGTCVEWLR' | 'HLA class I histocompatibility antigen, C alpha chain' |
| 8391 | 'HLA-C_No2' | 'APWVEQEGPEYWDRETQK' | 'HLA class I histocompatibility antigen, C alpha chain' |
| 8392 | 'HLA-C_No3' | 'AYLEGTCVEWLRR' | 'HLA class I histocompatibility antigen, C alpha chain' |
| 8393 | 'H1-4_No1' | 'KASGPPVSELITK' | 'Histone H1.4' |
| 8394 | 'H1-4_No2' | 'SETAPAAPAAPAPAEKTPVK' | 'Histone H1.4' |
| 8395 | 'SPP1_No1' | 'GKDSYETSQLDDQSAETHSHK' | 'Osteopontin' |
| 8396 | 'SPP1_No2' | 'ISHELDSASSEVN' | 'Osteopontin' |
| 8397 | 'SPP1_No3' | 'KANDESNEHSDVIDSQELSK' | 'Osteopontin' |
| 8398 | 'PTPRF_No1' | 'VLAFTAVGDGPPSPTIQVK' | 'Receptor-type tyrosine-protein phosphatase F' |
| 8399 | 'PTPRF_No2' | 'AAGTEGPFQEVDGVATTR' | 'Receptor-type tyrosine-protein phosphatase F' |
| 8400 | 'PTPRF_No3' | 'TDEDVPSGPPR' | 'Receptor-type tyrosine-protein phosphatase F' |
| 8401 | 'PTPRF_No4' | 'IQLSWLLPPQER' | 'Receptor-type tyrosine-protein phosphatase F' |
| 8402 | 'PTPRF_No5' | 'TQQGVPAQPADFQAEVESDTR' | 'Receptor-type tyrosine-protein phosphatase F' |
| 8403 | 'PTPRF_No6' | 'FEVIEFDDGAGSVLR' | 'Receptor-type tyrosine-protein phosphatase F' |
| 8404 | 'PTPRF_No7' | 'GSGPLSPSIQSR' | 'Receptor-type tyrosine-protein phosphatase F' |
| 8405 | 'PTPRF_No8' | 'IISYTVVFR' | 'Receptor-type tyrosine-protein phosphatase F' |

| | A | B | C |
|---|---|---|---|
| 1 | **Peptide Name** | **Peptide Sequence** | **Peptide Description** |
| 8406 | 'PTPRF_No9' | 'ADEARPNTIDFGKDDQHFTVTGLHK' | 'Receptor-type tyrosine-protein phosphatase F' |
| 8407 | 'PTPRF_No10' | 'FTLTGLKPDTTYDIK' | 'Receptor-type tyrosine-protein phosphatase F' |
| 8408 | 'PTPRF_No11' | 'GSSAGGLQHLVSIR' | 'Receptor-type tyrosine-protein phosphatase F' |
| 8409 | 'PTPRF_No12' | 'NGVITQYSVAYEAVDGEDR' | 'Receptor-type tyrosine-protein phosphatase F' |
| 8410 | 'PTPRF_No13' | 'VQRDEAIYECTATNSLGEINTSAK' | 'Receptor-type tyrosine-protein phosphatase F' |
| 8411 | 'PTPRF_No14' | 'AGLGEEFEK' | 'Receptor-type tyrosine-protein phosphatase F' |
| 8412 | 'PTPRF_No15' | 'DEAIYECTATNSLGEINTSAK' | 'Receptor-type tyrosine-protein phosphatase F' |
| 8413 | 'PTPRF_No16' | 'SDMGVGVFTPTIEAR' | 'Receptor-type tyrosine-protein phosphatase F' |
| 8414 | 'TXN_No1' | 'VGEFSGANKEKLEATINELV' | 'Thioredoxin' |
| 8415 | 'TXN_No2' | 'CMPTFQFFK' | 'Thioredoxin' |
| 8416 | 'TXN_No3' | 'CMPTFQFFK' | 'Thioredoxin' |
| 8417 | 'TXN_No4' | 'MIKPFFHSLSEK' | 'Thioredoxin' |
| 8418 | 'TXN_No5' | 'TAFQEALDAAGDK' | 'Thioredoxin' |
| 8419 | 'C7_No1' | 'LSGNVLSYTFQVK' | 'Complement component C7' |
| 8420 | 'C7_No2' | 'SCVGETTESTQCEDEELEHLR' | 'Complement component C7' |
| 8421 | 'C7_No3' | 'LLEPHCFPLSLVPTEFCPSPPALK' | 'Complement component C7' |
| 8422 | 'C7_No4' | 'LTPLYELVK' | 'Complement component C7' |
| 8423 | 'C7_No5' | 'EQTMSECEAGALR' | 'Complement component C7' |
| 8424 | 'C7_No6' | 'VLFYVDSEKLK' | 'Complement component C7' |
| 8425 | 'C7_No7' | 'MPYECGPSLDVCAQDER' | 'Complement component C7' |
| 8426 | 'C7_No8' | 'GQSISVTSIRPCAAETQ' | 'Complement component C7' |
| 8427 | 'C7_No9' | 'NVVYTCNEGYSLIGNPVAR' | 'Complement component C7' |
| 8428 | 'C7_No10' | 'ILPLTVCK' | 'Complement component C7' |
| 8429 | 'C7_No11' | 'RILPLTVCK' | 'Complement component C7' |
| 8430 | 'C7_No12' | 'RYSAWAESVTNLPQVIK' | 'Complement component C7' |
| 8431 | 'C7_No13' | 'ELSHLPSLYDYSAYR' | 'Complement component C7' |
| 8432 | 'C7_No14' | 'SSGWHFVVK' | 'Complement component C7' |
| 8433 | 'C7_No15' | 'SLVCNGDSDCDEDSADEDR' | 'Complement component C7' |
| 8434 | 'C7_No16' | 'ACGACPLWGK' | 'Complement component C7' |
| 8435 | 'C7_No17' | 'LKQNDFNSVEEK' | 'Complement component C7' |
| 8436 | 'C7_No18' | 'DGFVQDEGTMFPVGK' | 'Complement component C7' |
| 8437 | 'C7_No19' | 'EQTMSECEAGALR' | 'Complement component C7' |
| 8438 | 'C7_No20' | 'IACVLPVLMDGIQSHPQKPFYTVGEK' | 'Complement component C7' |
| 8439 | 'C7_No21' | 'DSCTLPASAEK' | 'Complement component C7' |
| 8440 | 'C7_No22' | 'DGFVQDEGTMFPVGK' | 'Complement component C7' |
| 8441 | 'C7_No23' | 'WLVGEMHCQK' | 'Complement component C7' |
| 8442 | 'C7_No24' | 'VLFYVDSEKLKQNDFNSVEEK' | 'Complement component C7' |
| 8443 | 'C7_No25' | 'KVFSGDGKDFYR' | 'Complement component C7' |
| 8444 | 'C7_No26' | 'VTVSCSGGMSLEGPSAFLCGSSLK' | 'Complement component C7' |
| 8445 | 'C7_No27' | 'ELSHLPSLYDYSAYRR' | 'Complement component C7' |
| 8446 | 'C7_No28' | 'FSSHGCKELENALK' | 'Complement component C7' |
| 8447 | 'C7_No29' | 'GGGAGFISGLSYLELDNPAGNK' | 'Complement component C7' |
| 8448 | 'C7_No30' | 'LIDQYGTHYLQSGSLGGEYR' | 'Complement component C7' |
| 8449 | 'C7_No31' | 'IACVLPVLMDGIQSHPQKPFYTVGEK' | 'Complement component C7' |

| | A | B | C |
|---|---|---|---|
| 1 | **Peptide Name** | **Peptide Sequence** | **Peptide Description** |
| 8450 | 'C7_No32' | 'GGGAGFISGLSYLELDNPAGNKR' | 'Complement component C7' |
| 8451 | 'C7_No33' | 'ACGACPLWGKCDAESSK' | 'Complement component C7' |
| 8452 | 'C7_No34' | 'QKLTPLYELVK' | 'Complement component C7' |
| 8453 | 'C7_No35' | 'YSAWAESVTNLPQVIK' | 'Complement component C7' |
| 8454 | 'C7_No36' | 'VFSGDGKDFYR' | 'Complement component C7' |
| 8455 | 'C7_No37' | 'CRGQSISVTSIRPCAAETQ' | 'Complement component C7' |
| 8456 | 'C7_No38' | 'WLVGEMHCQK' | 'Complement component C7' |
| 8457 | 'C7_No39' | 'RPSCDIDKPPPNIELTGNGYNELTGQFR' | 'Complement component C7' |
| 8458 | 'C7_No40' | 'RLIDQYGTHYLQSGSLGGEYR' | 'Complement component C7' |
| 8459 | 'C7_No41' | 'MPYECGPSLDVCAQDER' | 'Complement component C7' |
| 8460 | 'C7_No42' | 'CFSGQCISK' | 'Complement component C7' |
| 8461 | 'C7_No43' | 'AASGTQNNVLR' | 'Complement component C7' |
| 8462 | 'C7_No44' | 'IACVLPVLMDGIQSHPQK' | 'Complement component C7' |
| 8463 | 'C7_No45' | 'LKQNDFNSVEEKK' | 'Complement component C7' |
| 8464 | 'C7_No46' | 'GGGAGFISGLSYLELDNPAGNKRR' | 'Complement component C7' |
| 8465 | 'C7_No47' | 'QKLTPLYELVK' | 'Complement component C7' |
| 8466 | 'C7_No48' | 'QNDFNSVEEK' | 'Complement component C7' |
| 8467 | 'C7_No49' | 'VTVSCSGGMSLEGPSAFLCGSSLK' | 'Complement component C7' |
| 8468 | 'C7_No50' | 'SYTSHTNEIHK' | 'Complement component C7' |
| 8469 | 'C7_No51' | 'YSAWAESVTNLPQVIKQK' | 'Complement component C7' |
| 8470 | 'C7_No52' | 'EVPCASVK' | 'Complement component C7' |
| 8471 | 'C7_No53' | 'GCPTEEGCGERFR' | 'Complement component C7' |
| 8472 | 'C7_No54' | 'VLFYVDSEK' | 'Complement component C7' |
| 8473 | 'C7_No55' | 'SVAVYGQYGGQPCVGNAFETQSCEPTRGCPTEEGCGER' | 'Complement component C7' |
| 8474 | 'C7_No56' | 'SLVCNGDSDCDEDSADEDRCEDSER' | 'Complement component C7' |
| 8475 | 'C7_No57' | 'QNDFNSVEEKK' | 'Complement component C7' |
| 8476 | 'C7_No58' | 'LTPLYELVKEVPCASVK' | 'Complement component C7' |
| 8477 | 'C7_No59' | 'CVQKENPLTQAVPK' | 'Complement component C7' |
| 8478 | 'C7_No60' | 'RSVAVYGQYGGQPCVGNAFETQSCEPTR' | 'Complement component C7' |
| 8479 | 'C7_No61' | 'SRECNNPPPSGGGR' | 'Complement component C7' |
| 8480 | 'C7_No62' | 'ELENALKAASGTQNNVLR' | 'Complement component C7' |
| 8481 | 'PRKAR1A_No1' | 'VSILESLDKWER' | 'cAMP-dependent protein kinase type I-alpha regulatory subunit' |
| 8482 | 'PRKAR1A_No2' | 'LTVADALEPVQFEDGQK' | 'cAMP-dependent protein kinase type I-alpha regulatory subunit' |
| 8483 | 'PRKAR1A_No3' | 'GAISAEVYTEEDAASYVR' | 'cAMP-dependent protein kinase type I-alpha regulatory subunit' |
| 8484 | 'PRKAR1A_No4' | 'MESGSTAASEEAR' | 'cAMP-dependent protein kinase type I-alpha regulatory subunit' |
| 8485 | 'CHGA_No1' | 'RLEGQEEEEDNRDSSMK' | 'Chromogranin-A' |
| 8486 | 'CHGA_No2' | 'SGEATDGARPQALPEPMQESK' | 'Chromogranin-A' |
| 8487 | 'CHGA_No3' | 'LEGQEEEEDNRDSSMK' | 'Chromogranin-A' |
| 8488 | 'CHGA_No4' | 'YPGPQAEGDSEGLSQGLVDREK' | 'Chromogranin-A' |
| 8489 | 'CHGA_No5' | 'RLEGQEEEEDNRDSSMK' | 'Chromogranin-A' |
| 8490 | 'CHGA_No6' | 'SEALAVDGAGKPGAEEAQDPEGK' | 'Chromogranin-A' |
| 8491 | 'CHGA_No7' | 'RPEDQELESLSAIEAELEK' | 'Chromogranin-A' |
| 8492 | 'CHGA_No8' | 'YPGPQAEGDSEGLSQGLVDR' | 'Chromogranin-A' |
| 8493 | 'CHGA_No9' | 'AEGNNQAPGEEEEEEEEATNTHPPASLPSQK' | 'Chromogranin-A' |

| | A | B | C |
|---|---|---|---|
| 1 | **Peptide Name** | **Peptide Sequence** | **Peptide Description** |
| 8494 | 'TFPI_No1' | 'FFFNIFTR' | 'Tissue factor pathway inhibitor' |
| 8495 | 'TFPI_No2' | 'YGGCLGNMNNFETLEECK' | 'Tissue factor pathway inhibitor' |
| 8496 | 'TFPI_No3' | 'TTLQQEKPDFCFLEEDPGICR' | 'Tissue factor pathway inhibitor' |
| 8497 | 'TFPI_No4' | 'QCEEFIYGGCEGNQNRFESLEECKK' | 'Tissue factor pathway inhibitor' |
| 8498 | 'TFPI_No5' | 'YGGCLGNMNNFETLEECK' | 'Tissue factor pathway inhibitor' |
| 8499 | 'PF4V1_No1' | 'HITSLEVIK' | 'Platelet factor 4 variant' |
| 8500 | 'PF4V1_No2' | 'AGPHCPTAQLIATLK' | 'Platelet factor 4 variant' |
| 8501 | 'PF4V1_No3' | 'KICLDLQALLYK' | 'Platelet factor 4 variant' |
| 8502 | 'PF4V1_No4' | 'AEAEEDGDLQCLCVK' | 'Platelet factor 4 variant' |
| 8503 | 'KIT_No1' | 'CPLTDPEVTNYSLK' | 'Mast/stem cell growth factor receptor Kit' |
| 8504 | 'KIT_No2' | 'LCLHCSVDQEGK' | 'Mast/stem cell growth factor receptor Kit' |
| 8505 | 'KIT_No3' | 'HGLSNSIYVFVRDPAK' | 'Mast/stem cell growth factor receptor Kit' |
| 8506 | 'KIT_No4' | 'YNSWHHGDFNYER' | 'Mast/stem cell growth factor receptor Kit' |
| 8507 | 'ESD' | 'SVSAFAPICNPVLCPWGK' | 'S-formylglutathione hydrolase' |
| 8508 | 'CLU_No1' | 'LFDSDPITVTVPVEVSR' | 'Clusterin' |
| 8509 | 'CLU_No2' | 'YVNKEIQNAVNGVK' | 'Clusterin' |
| 8510 | 'CLU_No3' | 'TLLSNLEEAKK' | 'Clusterin' |
| 8511 | 'CLU_No4' | 'KTLLSNLEEAKK' | 'Clusterin' |
| 8512 | 'CLU_No5' | 'ELDESLQVAER' | 'Clusterin' |
| 8513 | 'CLU_No6' | 'FFTREPQDTYHYLPFSLPHR' | 'Clusterin' |
| 8514 | 'CLU_No7' | 'VTTVASHTSDSDVPSGVTEVVVK' | 'Clusterin' |
| 8515 | 'CLU_No8' | 'RPHFFFPK' | 'Clusterin' |
| 8516 | 'CLU_No9' | 'ASSIIDELFQDRFFTREPQDTYHYLPFSLPHR' | 'Clusterin' |
| 8517 | 'CLU_No10' | 'QQTHMLDVMQDHFSR' | 'Clusterin' |
| 8518 | 'CLU_No11' | 'IDSLLENDRQQTHMLDVMQDHFSR' | 'Clusterin' |
| 8519 | 'CLU_No12' | 'KTLLSNLEEAK' | 'Clusterin' |
| 8520 | 'CLU_No13' | 'EIQNAVNGVK' | 'Clusterin' |
| 8521 | 'CLU_No14' | 'KYNELLK' | 'Clusterin' |
| 8522 | 'CLU_No15' | 'QQTHMLDVMQDHFSR' | 'Clusterin' |
| 8523 | 'CLU_No16' | 'QQTHMLDVMQDHFSR' | 'Clusterin' |
| 8524 | 'CLU_No17' | 'IDSLLENDRQQTHMLDVMQDHFSR' | 'Clusterin' |
| 8525 | 'CLU_No18' | 'EPQDTYHYLPFSLPHR' | 'Clusterin' |
| 8526 | 'CLU_No19' | 'QQTHMLDVMQDHFSR' | 'Clusterin' |
| 8527 | 'CLU_No20' | 'ASSIIDELFQDR' | 'Clusterin' |
| 8528 | 'CLU_No21' | 'IDSLLENDRQQTHMLDVMQDHFSR' | 'Clusterin' |
| 8529 | 'CLU_No22' | 'IDSLLENDRQQTHMLDVMQDHFSR' | 'Clusterin' |
| 8530 | 'CLU_No23' | 'TLLSNLEEAK' | 'Clusterin' |
| 8531 | 'CLU_No24' | 'QQTHMLDVMQDHFSR' | 'Clusterin' |
| 8532 | 'CLU_No25' | 'QQTHMLDVMQDHFSR' | 'Clusterin' |
| 8533 | 'CLU_No26' | 'ASSIIDELFQDRFFTR' | 'Clusterin' |
| 8534 | 'CLU_No27' | 'LFDSDPITVTVPVEVSRK' | 'Clusterin' |
| 8535 | 'CLU_No28' | 'EILSVDCSTNNPSQAK' | 'Clusterin' |
| 8536 | 'CLU_No29' | 'QQTHMLDVMQDHFSR' | 'Clusterin' |
| 8537 | 'CLU_No30' | 'QQTHMLDVMQDHFSR' | 'Clusterin' |

| | A | B | C |
|---|---|---|---|
| 1 | **Peptide Name** | **Peptide Sequence** | **Peptide Description** |
| 8538 | 'CLU_No31' | 'RELDESLQVAER' | 'Clusterin' |
| 8539 | 'CLU_No32' | 'IDSLLENDR' | 'Clusterin' |
| 8540 | 'CLU_No33' | 'CREILSVDCSTNNPSQAK' | 'Clusterin' |
| 8541 | 'CLU_No34' | 'LANLTQGEDQYYLR' | 'Clusterin' |
| 8542 | 'CLU_No35' | 'FMETVAEKALQEYR' | 'Clusterin' |
| 8543 | 'CLU_No36' | 'TLIEKTNEER' | 'Clusterin' |
| 8544 | 'CLU_No37' | 'TLLSNLEEAKKK' | 'Clusterin' |
| 8545 | 'CLU_No38' | 'FMETVAEK' | 'Clusterin' |
| 8546 | 'CLU_No39' | 'FMETVAEKALQEYRK' | 'Clusterin' |
| 8547 | 'CLU_No40' | 'TLIEKTNEERK' | 'Clusterin' |
| 8548 | 'CLU_No41' | 'FMETVAEKALQEYR' | 'Clusterin' |
| 8549 | 'CLU_No42' | 'EGDDDRTVCR' | 'Clusterin' |
| 8550 | 'CLU_No43' | 'LRRELDESLQVAER' | 'Clusterin' |
| 8551 | 'CLU_No44' | 'EILSVDCSTNNPSQAKLRR' | 'Clusterin' |
| 8552 | 'CLU_No45' | 'KYNELLKSYQWK' | 'Clusterin' |
| 8553 | 'CLU_No46' | 'ELDESLQVAERLTR' | 'Clusterin' |
| 8554 | 'CLU_No47' | 'QIKTLIEKTNEER' | 'Clusterin' |
| 8555 | 'CLU_No48' | 'TNEERKTLLSNLEEAK' | 'Clusterin' |
| 8556 | 'HSPA5_No1' | 'SQIFSTASDNQPTVTIK' | 'Endoplasmic reticulum chaperone BiP' |
| 8557 | 'HSPA5_No2' | 'IINEPTAAAIAYGLDKR' | 'Endoplasmic reticulum chaperone BiP' |
| 8558 | 'HSPA5_No3' | 'NQLTSNPENTVFDAK' | 'Endoplasmic reticulum chaperone BiP' |
| 8559 | 'HSPA5_No4' | 'AKFEELNMDLFR' | 'Endoplasmic reticulum chaperone BiP' |
| 8560 | 'HSPA5_No5' | 'TKPYIQVDIGGGQTK' | 'Endoplasmic reticulum chaperone BiP' |
| 8561 | 'HSPA5_No6' | 'MKETAEAYLGK' | 'Endoplasmic reticulum chaperone BiP' |
| 8562 | 'HSPA5_No7' | 'DAGTIAGLNVMR' | 'Endoplasmic reticulum chaperone BiP' |
| 8563 | 'HSPA5_No8' | 'NGRVEIIANDQGNR' | 'Endoplasmic reticulum chaperone BiP' |
| 8564 | 'HSPA5_No9' | 'VTHAVVTVPAYFNDAQR' | 'Endoplasmic reticulum chaperone BiP' |
| 8565 | 'HSPA5_No10' | 'KSDIDEIVLVGGSTR' | 'Endoplasmic reticulum chaperone BiP' |
| 8566 | 'HSPA5_No11' | 'TWNDPSVQQDIK' | 'Endoplasmic reticulum chaperone BiP' |
| 8567 | 'HSPA5_No12' | 'EFFNGKEPSR' | 'Endoplasmic reticulum chaperone BiP' |
| 8568 | 'HSPA5_No13' | 'VYEGERPLTK' | 'Endoplasmic reticulum chaperone BiP' |
| 8569 | 'HSPA5_No14' | 'TFAPEEISAMVLTK' | 'Endoplasmic reticulum chaperone BiP' |
| 8570 | 'HSPA5_No15' | 'ELEEIVQPIISK' | 'Endoplasmic reticulum chaperone BiP' |
| 8571 | 'HSPA5_No16' | 'ITPSYVAFTPEGER' | 'Endoplasmic reticulum chaperone BiP' |
| 8572 | 'HSPA5_No17' | 'NQLTSNPENTVFDAKR' | 'Endoplasmic reticulum chaperone BiP' |
| 8573 | 'HSPA5_No18' | 'IDTRNELESYAYSLK' | 'Endoplasmic reticulum chaperone BiP' |
| 8574 | 'HSPA5_No19' | 'KTKPYIQVDIGGGQTK' | 'Endoplasmic reticulum chaperone BiP' |
| 8575 | 'HSPA5_No20' | 'KVTHAVVTVPAYFNDAQR' | 'Endoplasmic reticulum chaperone BiP' |
| 8576 | 'HSPA5_No21' | 'ITPSYVAFTPEGERLIGDAAK' | 'Endoplasmic reticulum chaperone BiP' |
| 8577 | 'HSPA5_No22' | 'IEWLESHQDADIEDFKAK' | 'Endoplasmic reticulum chaperone BiP' |
| 8578 | 'HSPA5_No23' | 'AVEEKIEWLESHQDADIEDFKAK' | 'Endoplasmic reticulum chaperone BiP' |
| 8579 | 'HSPA5_No24' | 'LYGSAGPPPTGEEDTAEKDEL' | 'Endoplasmic reticulum chaperone BiP' |
| 8580 | 'HSPA5_No25' | 'VLEDSDLKKSDIDEIVLVGGSTR' | 'Endoplasmic reticulum chaperone BiP' |
| 8581 | 'HSPA5_No26' | 'IEWLESHQDADIEDFK' | 'Endoplasmic reticulum chaperone BiP' |

| | A | B | C |
|---|---|---|---|
| 1 | **Peptide Name** | **Peptide Sequence** | **Peptide Description** |
| 8582 | 'HSPA5_No27' | 'DAGTIAGLNVMR' | 'Endoplasmic reticulum chaperone BiP' |
| 8583 | 'HSPA5_No28' | 'AKFEELNMDLFR' | 'Endoplasmic reticulum chaperone BiP' |
| 8584 | 'HSPA5_No29' | 'MVNDAEKFAEEDKK' | 'Endoplasmic reticulum chaperone BiP' |
| 8585 | 'HSPA5_No30' | 'TFAPEEISAMVLTK' | 'Endoplasmic reticulum chaperone BiP' |
| 8586 | 'HSPA5_No31' | 'KSQIFSTASDNQPTVTIK' | 'Endoplasmic reticulum chaperone BiP' |
| 8587 | 'LAMC1_No1' | 'LSAEDLVLEGAGLR' | 'Laminin subunit gamma-1' |
| 8588 | 'LAMC1_No2' | 'LCQCSDNIDPNAVGNCNR' | 'Laminin subunit gamma-1' |
| 8589 | 'LAMC1_No3' | 'EAQQALGSAAADATEAK' | 'Laminin subunit gamma-1' |
| 8590 | 'LAMC1_No4' | 'LNTFGDEVFNDPK' | 'Laminin subunit gamma-1' |
| 8591 | 'LAMC1_No5' | 'ATAESASECLPCDCNGR' | 'Laminin subunit gamma-1' |
| 8592 | 'LAMC1_No6' | 'TGQCECQPGITGQHCER' | 'Laminin subunit gamma-1' |
| 8593 | 'HSPA8_No1' | 'IINEPTAAAIAYGLDKK' | 'Heat shock cognate 71 kDa protein' |
| 8594 | 'HSPA8_No2' | 'NQVAMNPTNTVFDAK' | 'Heat shock cognate 71 kDa protein' |
| 8595 | 'HSPA8_No3' | 'STAGDTHLGGEDFDNR' | 'Heat shock cognate 71 kDa protein' |
| 8596 | 'HSPA8_No4' | 'LLQDFFNGK' | 'Heat shock cognate 71 kDa protein' |
| 8597 | 'HSPA8_No5' | 'NQVAMNPTNTVFDAK' | 'Heat shock cognate 71 kDa protein' |
| 8598 | 'HSPA8_No6' | 'DAGTIAGLNVLR' | 'Heat shock cognate 71 kDa protein' |
| 8599 | 'HSPA8_No7' | 'SQIHDIVLVGGSTR' | 'Heat shock cognate 71 kDa protein' |
| 8600 | 'HSPA8_No8' | 'FEELNADLFR' | 'Heat shock cognate 71 kDa protein' |
| 8601 | 'HSPA8_No9' | 'MKEIAEAYLGK' | 'Heat shock cognate 71 kDa protein' |
| 8602 | 'HSPA8_No10' | 'MVNHFIAEFK' | 'Heat shock cognate 71 kDa protein' |
| 8603 | 'HSPA8_No11' | 'GPAVGIDLGTTYSCVGVFQHGK' | 'Heat shock cognate 71 kDa protein' |
| 8604 | 'HSPA8_No12' | 'NQVAMNPTNTVFDAKR' | 'Heat shock cognate 71 kDa protein' |
| 8605 | 'HSPA8_No13' | 'MVNHFIAEFKR' | 'Heat shock cognate 71 kDa protein' |
| 8606 | 'HSPA8_No14' | 'LDKSQIHDIVLVGGSTR' | 'Heat shock cognate 71 kDa protein' |
| 8607 | 'HSPA8_No15' | 'RFDDAVVQSDMK' | 'Heat shock cognate 71 kDa protein' |
| 8608 | 'HSPA8_No16' | 'IINEPTAAAIAYGLDK' | 'Heat shock cognate 71 kDa protein' |
| 8609 | 'HSPA8_No17' | 'ARFEELNADLFR' | 'Heat shock cognate 71 kDa protein' |
| 8610 | 'HSPA8_No18' | 'TVTNAVVTVPAYFNDSQR' | 'Heat shock cognate 71 kDa protein' |
| 8611 | 'HSPA8_No19' | 'NSLESYAFNMK' | 'Heat shock cognate 71 kDa protein' |
| 8612 | 'HSPA8_No20' | 'NQVAMNPTNTVFDAKR' | 'Heat shock cognate 71 kDa protein' |
| 8613 | 'HSPA8_No21' | 'MVNHFIAEFKR' | 'Heat shock cognate 71 kDa protein' |
| 8614 | 'HSPA8_No22' | 'QTQTFTTYSDNQPGVLIQVYEGER' | 'Heat shock cognate 71 kDa protein' |
| 8615 | 'HSPA8_No23' | 'LLQDFFNGKELNK' | 'Heat shock cognate 71 kDa protein' |
| 8616 | 'HSPA8_No24' | 'MVNHFIAEFK' | 'Heat shock cognate 71 kDa protein' |
| 8617 | 'HSPA8_No25' | 'NQTAEKEEFEHQQKELEK' | 'Heat shock cognate 71 kDa protein' |
| 8618 | 'SLC2A1_No1' | 'VTILELFR' | 'Solute carrier family 2, facilitated glucose transporter member 1' |
| 8619 | 'SLC2A1_No2' | 'QGGASQSDKTPEELFHPLGADSQV' | 'Solute carrier family 2, facilitated glucose transporter member 1' |
| 8620 | 'SLC2A1_No3' | 'GTADVTHDLQEMKEESR' | 'Solute carrier family 2, facilitated glucose transporter member 1' |
| 8621 | 'SLC2A3_No1' | 'LWGTQDVSQDIQEMKDESAR' | 'Solute carrier family 2, facilitated glucose transporter member 3' |
| 8622 | 'SLC2A3_No2' | 'QVTVLELFR' | 'Solute carrier family 2, facilitated glucose transporter member 3' |
| 8623 | 'SLC2A3_No3' | 'SGKDGVMEMNSIEPAKETTTNV' | 'Solute carrier family 2, facilitated glucose transporter member 3' |
| 8624 | 'SLC2A3_No4' | 'LWGTQDVSQDIQEMKDESAR' | 'Solute carrier family 2, facilitated glucose transporter member 3' |
| 8625 | 'SLC2A3_No5' | 'DGVMEMNSIEPAKETTTNV' | 'Solute carrier family 2, facilitated glucose transporter member 3' |

| | A | B | C |
|---|---|---|---|
| 1 | **Peptide Name** | **Peptide Sequence** | **Peptide Description** |
| 8626 | 'EPB41_No1' | 'HSNLMLEDLDKSQEEIKK' | 'Protein 4.1' |
| 8627 | 'EPB41_No2' | 'VSLLDDTVYECVVEK' | 'Protein 4.1' |
| 8628 | 'EPB41_No3' | 'SMTPAQADLEFLENAK' | 'Protein 4.1' |
| 8629 | 'PYGB_No1' | 'IGEEFLTDLSQLKK' | 'Glycogen phosphorylase, brain form' |
| 8630 | 'PYGB_No2' | 'LLPLVSDEVFIR' | 'Glycogen phosphorylase, brain form' |
| 8631 | 'PYGB_No3' | 'IGEEFLTDLSQLK' | 'Glycogen phosphorylase, brain form' |
| 8632 | 'PYGB_No4' | 'WLLLCNPGLADTIVEK' | 'Glycogen phosphorylase, brain form' |
| 8633 | 'PYGB_No5' | 'QLLNCLHVVTLYNR' | 'Glycogen phosphorylase, brain form' |
| 8634 | 'PYGB_No6' | 'IVNGWQVEEADDWLR' | 'Glycogen phosphorylase, brain form' |
| 8635 | 'MBL2_No1' | 'FFLTNGEIMTFEK' | 'Mannose-binding protein C' |
| 8636 | 'MBL2_No2' | 'WLTFSLGK' | 'Mannose-binding protein C' |
| 8637 | 'MBL2_No3' | 'FQASVATPR' | 'Mannose-binding protein C' |
| 8638 | 'MBL2_No4' | 'NAAENGAIQNLIKEEAFLGITDEKTEGQFVDLTGNR' | 'Mannose-binding protein C' |
| 8639 | 'MBL2_No5' | 'SPDGDSSLAASER' | 'Mannose-binding protein C' |
| 8640 | 'MBL2_No6' | 'LTYTNWNEGEPNNAGSDEDCVLLLK' | 'Mannose-binding protein C' |
| 8641 | 'MBL2_No7' | 'NAAENGAIQNLIK' | 'Mannose-binding protein C' |
| 8642 | 'MBL2_No8' | 'EEAFLGITDEKTEGQFVDLTGNR' | 'Mannose-binding protein C' |
| 8643 | 'MBL2_No9' | 'FFLTNGEIMTFEK' | 'Mannose-binding protein C' |
| 8644 | 'MBL2_No10' | 'QVGNKFFLTNGEIMTFEK' | 'Mannose-binding protein C' |
| 8645 | 'MBL2_No11' | 'NAAENGAIQNLIKEEAFLGITDEK' | 'Mannose-binding protein C' |
| 8646 | 'MBL2_No12' | 'TEGQFVDLTGNR' | 'Mannose-binding protein C' |
| 8647 | 'SPTB_No1' | 'FLDLLEPLGR' | 'Spectrin beta chain, erythrocytic' |
| 8648 | 'SPTB_No2' | 'FAALEKPTTLELK' | 'Spectrin beta chain, erythrocytic' |
| 8649 | 'SPTB_No3' | 'VIQGQTDPEYLLLGQR' | 'Spectrin beta chain, erythrocytic' |
| 8650 | 'SPTB_No4' | 'ALEDLAQELEKENYHDQKR' | 'Spectrin beta chain, erythrocytic' |
| 8651 | 'SPTB_No5' | 'HLLEVEDLLQK' | 'Spectrin beta chain, erythrocytic' |
| 8652 | 'SPTB_No6' | 'LLSGEDVGQDEGATR' | 'Spectrin beta chain, erythrocytic' |
| 8653 | 'SPTB_No7' | 'VHLENMGSHDIVDGNHR' | 'Spectrin beta chain, erythrocytic' |
| 8654 | 'SPTB_No8' | 'KHEAIETDTAAYEER' | 'Spectrin beta chain, erythrocytic' |
| 8655 | 'SPTB_No9' | 'ELHLLGVQVQQFQDVATR' | 'Spectrin beta chain, erythrocytic' |
| 8656 | 'SPTB_No10' | 'HNLEHAFNVAER' | 'Spectrin beta chain, erythrocytic' |
| 8657 | 'SPTB_No11' | 'SDDPGKDLTSVNR' | 'Spectrin beta chain, erythrocytic' |
| 8658 | 'SPTB_No12' | 'DGLAFNALIHK' | 'Spectrin beta chain, erythrocytic' |
| 8659 | 'SPTB_No13' | 'FANSLTGVQQQLQAFSTYR' | 'Spectrin beta chain, erythrocytic' |
| 8660 | 'SPTB_No14' | 'DLTSVLILQR' | 'Spectrin beta chain, erythrocytic' |
| 8661 | 'SPTB_No15' | 'LPLAQSADYGTNLQTVQLFMK' | 'Spectrin beta chain, erythrocytic' |
| 8662 | 'SPTB_No16' | 'YHQGINAEIETR' | 'Spectrin beta chain, erythrocytic' |
| 8663 | 'SPTB_No17' | 'TQLVDTADKFR' | 'Spectrin beta chain, erythrocytic' |
| 8664 | 'SPTB_No18' | 'FQIQDIVVQTQEGR' | 'Spectrin beta chain, erythrocytic' |
| 8665 | 'SPTB_No19' | 'ELYQQVVAQADLR' | 'Spectrin beta chain, erythrocytic' |
| 8666 | 'SPTB_No20' | 'LLEVLSGEMLPKPTK' | 'Spectrin beta chain, erythrocytic' |
| 8667 | 'SPTB_No21' | 'GQQLVEAAEIDCQDLEER' | 'Spectrin beta chain, erythrocytic' |
| 8668 | 'SPTB_No22' | 'AQGLLSAGHPEGEQIIR' | 'Spectrin beta chain, erythrocytic' |
| 8669 | 'SPTB_No23' | 'TSATEFENVGNQPPYSR' | 'Spectrin beta chain, erythrocytic' |

| | A | B | C |
|---|---|---|---|
| 1 | **Peptide Name** | **Peptide Sequence** | **Peptide Description** |
| 8670 | 'SPTB_No24' | 'HRELPEDVGLDASTAESFHR' | 'Spectrin beta chain, erythrocytic' |
| 8671 | 'SPTB_No25' | 'LVAQDNFGYDLAAVEAAK' | 'Spectrin beta chain, erythrocytic' |
| 8672 | 'SPTB_No26' | 'QIAERPAEETGPQEEEGETAGEAPVSHHAATER' | 'Spectrin beta chain, erythrocytic' |
| 8673 | 'SPTB_No27' | 'ETDDLEQWISEK' | 'Spectrin beta chain, erythrocytic' |
| 8674 | 'SPTB_No28' | 'LIDAGHSEAATIAEWK' | 'Spectrin beta chain, erythrocytic' |
| 8675 | 'SPTB_No29' | 'HQAFVAELASHEGWLENIDAEGK' | 'Spectrin beta chain, erythrocytic' |
| 8676 | 'SPTB_No30' | 'VHLENMGSHDIVDGNHR' | 'Spectrin beta chain, erythrocytic' |
| 8677 | 'SPTB_No31' | 'ELVASSPEMGQDFDHVTLLR' | 'Spectrin beta chain, erythrocytic' |
| 8678 | 'SPTB_No32' | 'LQGQVDKHYAGLKDVAEER' | 'Spectrin beta chain, erythrocytic' |
| 8679 | 'LAMP1_No1' | 'AFSVNIFK' | 'Lysosome-associated membrane glycoprotein 1' |
| 8680 | 'LAMP1_No2' | 'FFLQGIQLNTILPDARDPAFK' | 'Lysosome-associated membrane glycoprotein 1' |
| 8681 | 'LAMP1_No3' | 'TVESITDIRADIDKK' | 'Lysosome-associated membrane glycoprotein 1' |
| 8682 | 'FGFR1_No1' | 'IGPDNLPYVQILK' | 'Fibroblast growth factor receptor 1' |
| 8683 | 'FGFR1_No2' | 'VYSDPQPHIQWLK' | 'Fibroblast growth factor receptor 1' |
| 8684 | 'G6PD_No1' | 'GGYFDEFGIIR' | 'Glucose-6-phosphate 1-dehydrogenase' |
| 8685 | 'G6PD_No2' | 'LILDVFCGSQMHFVR' | 'Glucose-6-phosphate 1-dehydrogenase' |
| 8686 | 'G6PD_No3' | 'NSYVAGQYDDAASYQR' | 'Glucose-6-phosphate 1-dehydrogenase' |
| 8687 | 'G6PD_No4' | 'DGLLPENTFIVGYAR' | 'Glucose-6-phosphate 1-dehydrogenase' |
| 8688 | 'G6PD_No5' | 'LQFHDVAGDIFHQQCK' | 'Glucose-6-phosphate 1-dehydrogenase' |
| 8689 | 'G6PD_No6' | 'LKLEDFFAR' | 'Glucose-6-phosphate 1-dehydrogenase' |
| 8690 | 'G6PD_No7' | 'LSNHISSLFR' | 'Glucose-6-phosphate 1-dehydrogenase' |
| 8691 | 'G6PD_No8' | 'WDGVPFILR' | 'Glucose-6-phosphate 1-dehydrogenase' |
| 8692 | 'MTHFD1_No1' | 'GVPTGFILPIR' | 'C-1-tetrahydrofolate synthase, cytoplasmic' |
| 8693 | 'MTHFD1_No2' | 'LDIDPETITWQR' | 'C-1-tetrahydrofolate synthase, cytoplasmic' |
| 8694 | 'MTHFD1_No3' | 'TDPTTLTDEEINR' | 'C-1-tetrahydrofolate synthase, cytoplasmic' |
| 8695 | 'CETP_No1' | 'YGLHNIQISHLSIASSQVELVEAK' | 'Cholesteryl ester transfer protein' |
| 8696 | 'CETP_No2' | 'ITKPALLVLNHETAK' | 'Cholesteryl ester transfer protein' |
| 8697 | 'CETP_No3' | 'LMLSLMGDEFK' | 'Cholesteryl ester transfer protein' |
| 8698 | 'CETP_No4' | 'TDAPDCYLSFHK' | 'Cholesteryl ester transfer protein' |
| 8699 | 'CETP_No5' | 'LLLHLQGEREPGWIK' | 'Cholesteryl ester transfer protein' |
| 8700 | 'CETP_No6' | 'KLFLSLLDFQITPK' | 'Cholesteryl ester transfer protein' |
| 8701 | 'CETP_No7' | 'LEVVFTALMNSK' | 'Cholesteryl ester transfer protein' |
| 8702 | 'CETP_No8' | 'VIQTAFQR' | 'Cholesteryl ester transfer protein' |
| 8703 | 'CETP_No9' | 'EINVISNIMADFVQTR' | 'Cholesteryl ester transfer protein' |
| 8704 | 'CETP_No10' | 'PALLVLNHETAK' | 'Cholesteryl ester transfer protein' |
| 8705 | 'CETP_No11' | 'EINVISNIMADFVQTR' | 'Cholesteryl ester transfer protein' |
| 8706 | 'CETP_No12' | 'LLLHLQGER' | 'Cholesteryl ester transfer protein' |
| 8707 | 'CETP_No13' | 'LMLSLMGDEFK' | 'Cholesteryl ester transfer protein' |
| 8708 | 'CETP_No14' | 'AASILSDGDIGVDISLTGDPVITASYLESHHK' | 'Cholesteryl ester transfer protein' |
| 8709 | 'CETP_No15' | 'GVVVNSSVMVK' | 'Cholesteryl ester transfer protein' |
| 8710 | 'CETP_No16' | 'GVSLFDIINPEIITR' | 'Cholesteryl ester transfer protein' |
| 8711 | 'CETP_No17' | 'VRTDAPDCYLSFHK' | 'Cholesteryl ester transfer protein' |
| 8712 | 'CETP_No18' | 'LMLSLMGDEFK' | 'Cholesteryl ester transfer protein' |
| 8713 | 'CETP_No19' | 'LEVVFTALMNSK' | 'Cholesteryl ester transfer protein' |

| | A | B | C |
|---|---|---|---|
| 1 | **Peptide Name** | **Peptide Sequence** | **Peptide Description** |
| 8714 | 'CETP_No20' | 'AMMLLGQVK' | 'Cholesteryl ester transfer protein' |
| 8715 | 'CETP_No21' | 'GTSHEAGIVCR' | 'Cholesteryl ester transfer protein' |
| 8716 | 'IGF2R_No1' | 'ATLITFLCDR' | 'Cation-independent mannose-6-phosphate receptor' |
| 8717 | 'IGF2R_No2' | 'YDLSALVR' | 'Cation-independent mannose-6-phosphate receptor' |
| 8718 | 'IGF2R_No3' | 'FLHQDIDSGQGIR' | 'Cation-independent mannose-6-phosphate receptor' |
| 8719 | 'IGF2R_No4' | 'PLNPVPGCNR' | 'Cation-independent mannose-6-phosphate receptor' |
| 8720 | 'IGF2R_No5' | 'FVCNDDVYSGPLK' | 'Cation-independent mannose-6-phosphate receptor' |
| 8721 | 'IGF2R_No6' | 'GCPEDAAVCAVDK' | 'Cation-independent mannose-6-phosphate receptor' |
| 8722 | 'IGF2R_No7' | 'TLGTPEFVTATECVHYFEWR' | 'Cation-independent mannose-6-phosphate receptor' |
| 8723 | 'IGF2R_No8' | 'YVDGDLCPDGIR' | 'Cation-independent mannose-6-phosphate receptor' |
| 8724 | 'IGF2R_No9' | 'TYHSVGDSVLR' | 'Cation-independent mannose-6-phosphate receptor' |
| 8725 | 'IGF2R_No10' | 'VEGDNCEVKDPR' | 'Cation-independent mannose-6-phosphate receptor' |
| 8726 | 'IGF2R_No11' | 'RYDLSALVR' | 'Cation-independent mannose-6-phosphate receptor' |
| 8727 | 'IGF2R_No12' | 'KYDFYINVCGPVSVSPCQPDSGACQVAK' | 'Cation-independent mannose-6-phosphate receptor' |
| 8728 | 'IGF2R_No13' | 'LDFCDGHSPAVTITFVCPSER' | 'Cation-independent mannose-6-phosphate receptor' |
| 8729 | 'IGF2R_No14' | 'LSYYDGMIQLNYR' | 'Cation-independent mannose-6-phosphate receptor' |
| 8730 | 'IGF2R_No15' | 'DIDTLRDPGSQLR' | 'Cation-independent mannose-6-phosphate receptor' |
| 8731 | 'IGF2R_No16' | 'HFFINICHR' | 'Cation-independent mannose-6-phosphate receptor' |
| 8732 | 'IGF2R_No17' | 'GNIQLSYSDGDDCGHGK' | 'Cation-independent mannose-6-phosphate receptor' |
| 8733 | 'IGF2R_No18' | 'SFSLGDIYFK' | 'Cation-independent mannose-6-phosphate receptor' |
| 8734 | 'IGF2R_No19' | 'TYSVGVCTFAVGPEQGGCK' | 'Cation-independent mannose-6-phosphate receptor' |
| 8735 | 'IGF2R_No20' | 'HAEPEQNWEAVDGSQTETEK' | 'Cation-independent mannose-6-phosphate receptor' |
| 8736 | 'IGF2R_No21' | 'VPIDGPPIDIGR' | 'Cation-independent mannose-6-phosphate receptor' |
| 8737 | 'IGF2R_No22' | 'DGAGNSFDLSSLSR' | 'Cation-independent mannose-6-phosphate receptor' |
| 8738 | 'IGF2R_No23' | 'HAEPEQNWEAVDGSQTETEKK' | 'Cation-independent mannose-6-phosphate receptor' |
| 8739 | 'IGF2R_No24' | 'PASGCEAETQTEELK' | 'Cation-independent mannose-6-phosphate receptor' |
| 8740 | 'IGF2R_No25' | 'GNIQLSYSDGDDCGHGKK' | 'Cation-independent mannose-6-phosphate receptor' |
| 8741 | 'IGF2R_No26' | 'EKEDLLCGATDGKK' | 'Cation-independent mannose-6-phosphate receptor' |
| 8742 | 'ADH5_No1' | 'AGDTVIPLYIPQCGECK' | 'Alcohol dehydrogenase class-3' |
| 8743 | 'ADH5_No2' | 'VCLLGCGISTGYGAAVNTAK' | 'Alcohol dehydrogenase class-3' |
| 8744 | 'ADH5_No3' | 'AAVAWEAGKPLSIEEIEVAPPK' | 'Alcohol dehydrogenase class-3' |
| 8745 | 'ADH5_No4' | 'IKVDEFVTHNLSFDEINK' | 'Alcohol dehydrogenase class-3' |
| 8746 | 'ADH5_No5' | 'VDEFVTHNLSFDEINK' | 'Alcohol dehydrogenase class-3' |
| 8747 | 'COL6A1_No1' | 'LLLFSDGNSQGATPAAIEK' | 'Collagen alpha-1(VI) chain' |
| 8748 | 'COL6A1_No2' | 'VPSYQALLR' | 'Collagen alpha-1(VI) chain' |
| 8749 | 'COL6A1_No3' | 'TAEYDVAYGESHLFR' | 'Collagen alpha-1(VI) chain' |
| 8750 | 'COL6A1_No4' | 'VAVVQYSGTGQQRPER' | 'Collagen alpha-1(VI) chain' |
| 8751 | 'COL6A1_No5' | 'DTTPLNVLCSPGIQVVSVGIK' | 'Collagen alpha-1(VI) chain' |
| 8752 | 'COL6A1_No6' | 'VFSVAITPDHLEPR' | 'Collagen alpha-1(VI) chain' |
| 8753 | 'COL6A1_No7' | 'LSIIATDHTYR' | 'Collagen alpha-1(VI) chain' |
| 8754 | 'COL6A1_No8' | 'FEPGQSYAGVVQYSHSQMQEHVSLR' | 'Collagen alpha-1(VI) chain' |
| 8755 | 'COL6A1_No9' | 'FEPGQSYAGVVQYSHSQMQEHVSLR' | 'Collagen alpha-1(VI) chain' |
| 8756 | 'COL6A1_No10' | 'SLQWMAGGTFTGEALQYTR' | 'Collagen alpha-1(VI) chain' |
| 8757 | 'COL6A3_No1' | 'LLTPITTLTSEQIQK' | 'Collagen alpha-3(VI) chain' |

| | A | B | C |
|---|---|---|---|
| 1 | **Peptide Name** | **Peptide Sequence** | **Peptide Description** |
| 8758 | 'COL6A3_No2' | 'QFGVAPFTIAR' | 'Collagen alpha-3(VI) chain' |
| 8759 | 'COL6A3_No3' | 'LLVLITGGK' | 'Collagen alpha-3(VI) chain' |
| 8760 | 'COL6A3_No4' | 'LQPVLQPLPSPGVGGK' | 'Collagen alpha-3(VI) chain' |
| 8761 | 'COL6A3_No5' | 'IVVLMLTGEVPEQQLEEAQR' | 'Collagen alpha-3(VI) chain' |
| 8762 | 'COL6A3_No6' | 'IAVAQYSDDVKVESR' | 'Collagen alpha-3(VI) chain' |
| 8763 | 'COL6A3_No7' | 'LLPSFVSSENAFYLSPDIR' | 'Collagen alpha-3(VI) chain' |
| 8764 | 'COL6A3_No8' | 'AAPLQGMLPGLLAPLR' | 'Collagen alpha-3(VI) chain' |
| 8765 | 'COL6A3_No9' | 'MKPLDGSALYTGSALDFVR' | 'Collagen alpha-3(VI) chain' |
| 8766 | 'COL6A3_No10' | 'SLDEISQPAQELKR' | 'Collagen alpha-3(VI) chain' |
| 8767 | 'COL6A3_No11' | 'QINVGNALEYVSR' | 'Collagen alpha-3(VI) chain' |
| 8768 | 'COL6A3_No12' | 'IIDELNVKPEGTR' | 'Collagen alpha-3(VI) chain' |
| 8769 | 'COL6A3_No13' | 'IGDLHPQIVNLLK' | 'Collagen alpha-3(VI) chain' |
| 8770 | 'COL6A3_No14' | 'ISLSPEYVFSVSTFR' | 'Collagen alpha-3(VI) chain' |
| 8771 | 'COL6A3_No15' | 'GLEYIMQSHLTK' | 'Collagen alpha-3(VI) chain' |
| 8772 | 'COL6A3_No16' | 'VAVIQFSDDPKVEFLLNAHSSKDEVQNAVQR' | 'Collagen alpha-3(VI) chain' |
| 8773 | 'COL6A3_No17' | 'QSGVVPFIFQAK' | 'Collagen alpha-3(VI) chain' |
| 8774 | 'COL6A3_No18' | 'VAVVQYSDR' | 'Collagen alpha-3(VI) chain' |
| 8775 | 'COL6A3_No19' | 'LVDFLSR' | 'Collagen alpha-3(VI) chain' |
| 8776 | 'COL6A3_No20' | 'VGVVQFSDEPR' | 'Collagen alpha-3(VI) chain' |
| 8777 | 'COL6A3_No21' | 'ALNLGYALDYAQR' | 'Collagen alpha-3(VI) chain' |
| 8778 | 'COL6A3_No22' | 'VGVVQFSNDVFPEFYLK' | 'Collagen alpha-3(VI) chain' |
| 8779 | 'COL6A3_No23' | 'QINVGNALEYVSR' | 'Collagen alpha-3(VI) chain' |
| 8780 | 'COL6A3_No24' | 'EVQVFEITENSAK' | 'Collagen alpha-3(VI) chain' |
| 8781 | 'COL6A3_No25' | 'VVESLDVGQDRVR' | 'Collagen alpha-3(VI) chain' |
| 8782 | 'COL6A3_No26' | 'SSIMAFAIGNK' | 'Collagen alpha-3(VI) chain' |
| 8783 | 'COL6A3_No27' | 'SDDEVDDPAVELKQFGVAPFTIAR' | 'Collagen alpha-3(VI) chain' |
| 8784 | 'COL6A3_No28' | 'FDEHQSKPEILNLVK' | 'Collagen alpha-3(VI) chain' |
| 8785 | 'COL6A3_No29' | 'LVDKSTELNEEPLMR' | 'Collagen alpha-3(VI) chain' |
| 8786 | 'COL6A3_No30' | 'IEDGVPQHLVLVLGGK' | 'Collagen alpha-3(VI) chain' |
| 8787 | 'COL6A3_No31' | 'IVVLMLTGEVPEQQLEEAQR' | 'Collagen alpha-3(VI) chain' |
| 8788 | 'COL6A3_No32' | 'VPQIAFVITGGK' | 'Collagen alpha-3(VI) chain' |
| 8789 | 'COL6A3_No33' | 'SADVNVFAIGVEDADEGALK' | 'Collagen alpha-3(VI) chain' |
| 8790 | 'COL6A3_No34' | 'TRPEFYLNSYMNK' | 'Collagen alpha-3(VI) chain' |
| 8791 | 'COL6A3_No35' | 'DVVFLLDGSEGVR' | 'Collagen alpha-3(VI) chain' |
| 8792 | 'COL6A3_No36' | 'QASVFSFGLGAQAASR' | 'Collagen alpha-3(VI) chain' |
| 8793 | 'COL6A3_No37' | 'SLDEISQPAQELK' | 'Collagen alpha-3(VI) chain' |
| 8794 | 'COL6A3_No38' | 'AAPLQGMLPGLLAPLR' | 'Collagen alpha-3(VI) chain' |
| 8795 | 'COL6A3_No39' | 'EFRELPNIEER' | 'Collagen alpha-3(VI) chain' |
| 8796 | 'COL6A3_No40' | 'EVYTFASEPNDVFFK' | 'Collagen alpha-3(VI) chain' |
| 8797 | 'COL6A3_No41' | 'QLGTVQQVISER' | 'Collagen alpha-3(VI) chain' |
| 8798 | 'COL6A3_No42' | 'VEFLLNAHSSKDEVQNAVQR' | 'Collagen alpha-3(VI) chain' |
| 8799 | 'COL6A3_No43' | 'ELPNIEER' | 'Collagen alpha-3(VI) chain' |
| 8800 | 'COL6A3_No44' | 'SGDDVRNPSVVVK' | 'Collagen alpha-3(VI) chain' |
| 8801 | 'COL6A3_No45' | 'LVDYLDVGFDTTR' | 'Collagen alpha-3(VI) chain' |

| | A | B | C |
|---|---|---|---|
| 1 | **Peptide Name** | **Peptide Sequence** | **Peptide Description** |
| 8802 | 'COL6A3_No46' | 'VCAPVLAKPGVISVMGT' | 'Collagen alpha-3(VI) chain' |
| 8803 | 'COL6A3_No47' | 'VVIHFTDGADGDLADLHR' | 'Collagen alpha-3(VI) chain' |
| 8804 | 'COL6A3_No48' | 'MKPLDGSALYTGSALDFVR' | 'Collagen alpha-3(VI) chain' |
| 8805 | 'COL6A3_No49' | 'VIGGLLAGQTYHVAVVCYLR' | 'Collagen alpha-3(VI) chain' |
| 8806 | 'COL6A3_No50' | 'SVEDAQDVSLALTQR' | 'Collagen alpha-3(VI) chain' |
| 8807 | 'COL6A3_No51' | 'FDEHQSKPEILNLVKR' | 'Collagen alpha-3(VI) chain' |
| 8808 | 'COL6A3_No52' | 'QCDWFQGDQPTK' | 'Collagen alpha-3(VI) chain' |
| 8809 | 'COL6A3_No53' | 'AELQHIATDDNLVFTVPEFR' | 'Collagen alpha-3(VI) chain' |
| 8810 | 'COL6A3_No54' | 'SSIMAFAIGNK' | 'Collagen alpha-3(VI) chain' |
| 8811 | 'COL6A3_No55' | 'LLPSFVSSENAFYLSPDIRK' | 'Collagen alpha-3(VI) chain' |
| 8812 | 'COL6A3_No56' | 'VVESLDVGQDR' | 'Collagen alpha-3(VI) chain' |
| 8813 | 'COL6A3_No57' | 'TRPEFYLNSYMNK' | 'Collagen alpha-3(VI) chain' |
| 8814 | 'SLC25A6_No1' | 'YFPTQALNFAFK' | 'ADP/ATP translocase 3' |
| 8815 | 'SLC25A6_No2' | 'EQGVLSFWR' | 'ADP/ATP translocase 3' |
| 8816 | 'SLC25A6_No3' | 'GADIMYTGTVDCWR' | 'ADP/ATP translocase 3' |
| 8817 | 'SLC25A6_No4' | 'DFLAGGIAAAISK' | 'ADP/ATP translocase 3' |
| 8818 | 'F5_No1' | 'EFNPLVIVGLSK' | 'Coagulation factor V' |
| 8819 | 'F5_No2' | 'ETDIEDSDDIPEDTTYK' | 'Coagulation factor V' |
| 8820 | 'F5_No3' | 'SEAYNTFSER' | 'Coagulation factor V' |
| 8821 | 'F5_No4' | 'NFFNPPIISR' | 'Coagulation factor V' |
| 8822 | 'F5_No5' | 'SQHLDNFSNQIGK' | 'Coagulation factor V' |
| 8823 | 'F5_No6' | 'DIHSGLIGPLLICQK' | 'Coagulation factor V' |
| 8824 | 'F5_No7' | 'QWLEIDLLK' | 'Coagulation factor V' |
| 8825 | 'F5_No8' | 'VMYTQYEDESFTK' | 'Coagulation factor V' |
| 8826 | 'F5_No9' | 'WIISSLTPK' | 'Coagulation factor V' |
| 8827 | 'F5_No10' | 'LNNGGSYNAWSVEK' | 'Coagulation factor V' |
| 8828 | 'F5_No11' | 'QHQLGVWPLLPGSFK' | 'Coagulation factor V' |
| 8829 | 'F5_No12' | 'ETDIEDSDDIPEDTTYKK' | 'Coagulation factor V' |
| 8830 | 'F5_No13' | 'LELQGCEVNGCSTPLGMENGK' | 'Coagulation factor V' |
| 8831 | 'F5_No14' | 'AWGESTPLANKPGK' | 'Coagulation factor V' |
| 8832 | 'F5_No15' | 'LSEGASYLDHTFPAEK' | 'Coagulation factor V' |
| 8833 | 'F5_No16' | 'MYEQEWVR' | 'Coagulation factor V' |
| 8834 | 'F5_No17' | 'DPPSDLLLLK' | 'Coagulation factor V' |
| 8835 | 'F5_No18' | 'FCENPDEVKR' | 'Coagulation factor V' |
| 8836 | 'F5_No19' | 'QHQLGVWPLLPGSFK' | 'Coagulation factor V' |
| 8837 | 'F5_No20' | 'AADIEQQAVFAVFDENK' | 'Coagulation factor V' |
| 8838 | 'F5_No21' | 'NKADKPLSIHPQGIR' | 'Coagulation factor V' |
| 8839 | 'F5_No22' | 'MPMGLSTGIISDSQIK' | 'Coagulation factor V' |
| 8840 | 'F5_No23' | 'AVQPGETYTYK' | 'Coagulation factor V' |
| 8841 | 'F5_No24' | 'LAAEFASKPWIQVDMQK' | 'Coagulation factor V' |
| 8842 | 'F5_No25' | 'EDGILGPIIR' | 'Coagulation factor V' |
| 8843 | 'F5_No26' | 'DPDNIAAWYLR' | 'Coagulation factor V' |
| 8844 | 'F5_No27' | 'SYTIHYSEQGVEWKPYR' | 'Coagulation factor V' |
| 8845 | 'F5_No28' | 'ASKPGWWLLNTEVGENQR' | 'Coagulation factor V' |

| | A | B | C |
|---|---|---|---|
| 1 | **Peptide Name** | **Peptide Sequence** | **Peptide Description** |
| 8846 | 'F5_No29' | 'KITAIITQGCK' | 'Coagulation factor V' |
| 8847 | 'F5_No30' | 'MRPWKDPPSDLLLLK' | 'Coagulation factor V' |
| 8848 | 'F5_No31' | 'VMYTQYEDESFTK' | 'Coagulation factor V' |
| 8849 | 'F5_No32' | 'HLQAGMQAYIDIK' | 'Coagulation factor V' |
| 8850 | 'F5_No33' | 'LELQGCEVNGCSTPLGMENGKIENK' | 'Coagulation factor V' |
| 8851 | 'F5_No34' | 'SWWGDYWEPFR' | 'Coagulation factor V' |
| 8852 | 'F5_No35' | 'GEYEEHLGILGPIIR' | 'Coagulation factor V' |
| 8853 | 'F5_No36' | 'HTVNPNMKEDGILGPIIR' | 'Coagulation factor V' |
| 8854 | 'F5_No37' | 'SHEFHAINGMIYSLPGLK' | 'Coagulation factor V' |
| 8855 | 'F5_No38' | 'PYSLHAHGLSYEK' | 'Coagulation factor V' |
| 8856 | 'F5_No39' | 'YLDSTFTK' | 'Coagulation factor V' |
| 8857 | 'F5_No40' | 'NVMYFNGNSDASTIKENQFDPPIVAR' | 'Coagulation factor V' |
| 8858 | 'F5_No41' | 'RHEDTLTLFPMR' | 'Coagulation factor V' |
| 8859 | 'F5_No42' | 'FTVNNLAEPQK' | 'Coagulation factor V' |
| 8860 | 'F5_No43' | 'TFHPLRSEAYNTFSER' | 'Coagulation factor V' |
| 8861 | 'F5_No44' | 'SWYLEDNINKFCENPDEVKR' | 'Coagulation factor V' |
| 8862 | 'F5_No45' | 'ASEFLGYWEPR' | 'Coagulation factor V' |
| 8863 | 'F5_No46' | 'AWAYYSAVNPEKDIHSGLIGPLLICQK' | 'Coagulation factor V' |
| 8864 | 'F5_No47' | 'LELQGCEVNGCSTPLGMENGK' | 'Coagulation factor V' |
| 8865 | 'F5_No48' | 'ADKPLSIHPQGIR' | 'Coagulation factor V' |
| 8866 | 'F5_No49' | 'LLSLGAGEFK' | 'Coagulation factor V' |
| 8867 | 'F5_No50' | 'NLASRPYSLHAHGLSYEK' | 'Coagulation factor V' |
| 8868 | 'F5_No51' | 'ENQFDPPIVAR' | 'Coagulation factor V' |
| 8869 | 'F5_No52' | 'WNILEFDEPTENDAQCLTR' | 'Coagulation factor V' |
| 8870 | 'F5_No53' | 'IVYREYEPYFKK' | 'Coagulation factor V' |
| 8871 | 'F5_No54' | 'MPMGLSTGIISDSQIK' | 'Coagulation factor V' |
| 8872 | 'F5_No55' | 'HTVNPNMKEDGILGPIIR' | 'Coagulation factor V' |
| 8873 | 'F5_No56' | 'FCENPDEVKRDDPK' | 'Coagulation factor V' |
| 8874 | 'F5_No57' | 'AGMQTPFLIMDRDCR' | 'Coagulation factor V' |
| 8875 | 'F5_No58' | 'DPRGEYEEHLGILGPIIR' | 'Coagulation factor V' |
| 8876 | 'F5_No59' | 'NVMYFNGNSDASTIK' | 'Coagulation factor V' |
| 8877 | 'F5_No60' | 'MRPWKDPPSDLLLLK' | 'Coagulation factor V' |
| 8878 | 'F5_No61' | 'SHEFHAINGMIYSLPGLK' | 'Coagulation factor V' |
| 8879 | 'F5_No62' | 'AGMQTPFLIMDR' | 'Coagulation factor V' |
| 8880 | 'F5_No63' | 'SSSPELSEMLEYDR' | 'Coagulation factor V' |
| 8881 | 'F5_No64' | 'PGWWLLNTEVGENQR' | 'Coagulation factor V' |
| 8882 | 'F5_No65' | 'DIASGLIGLLLICK' | 'Coagulation factor V' |
| 8883 | 'F5_No66' | 'QWLEIDLLK' | 'Coagulation factor V' |
| 8884 | 'F5_No67' | 'SSSPELSEMLEYDR' | 'Coagulation factor V' |
| 8885 | 'F5_No68' | 'TWNQSIALR' | 'Coagulation factor V' |
| 8886 | 'F5_No69' | 'SSMVDKIFEGNTNTK' | 'Coagulation factor V' |
| 8887 | 'F5_No70' | 'AEVDDVIQVR' | 'Coagulation factor V' |
| 8888 | 'F5_No71' | 'HLSQDTGSPSGMRPWEDLPSQDTGSPSR' | 'Coagulation factor V' |
| 8889 | 'F5_No72' | 'RDPRGEYEEHLGILGPIIR' | 'Coagulation factor V' |

| | A | B | C |
|---|---|---|---|
| 1 | **Peptide Name** | **Peptide Sequence** | **Peptide Description** |
| 8890 | 'F5_No73' | 'HLSQDTGSPSGMRPWEDLPSQDTGSPSR' | 'Coagulation factor V' |
| 8891 | 'F5_No74' | 'AGMQTPFLIMDRDCR' | 'Coagulation factor V' |
| 8892 | 'F5_No75' | 'KSHEFHAINGMIYSLPGLK' | 'Coagulation factor V' |
| 8893 | 'F5_No76' | 'AADIEQQAVFAVFDENKSWYLEDNINK' | 'Coagulation factor V' |
| 8894 | 'F5_No77' | 'FTVNNLAEPQKAPSHQQATTAGSPLR' | 'Coagulation factor V' |
| 8895 | 'F5_No78' | 'APSHQQATTAGSPLR' | 'Coagulation factor V' |
| 8896 | 'F5_No79' | 'LKSSMVDKIFEGNTNTK' | 'Coagulation factor V' |
| 8897 | 'F5_No80' | 'AGMQTPFLIMDRDCR' | 'Coagulation factor V' |
| 8898 | 'F5_No81' | 'FKNLASRPYSLHAHGLSYEK' | 'Coagulation factor V' |
| 8899 | 'F5_No82' | 'SWYLEDNINK' | 'Coagulation factor V' |
| 8900 | 'F5_No83' | 'RHEDTLTLFPMR' | 'Coagulation factor V' |
| 8901 | 'F5_No84' | 'LELQGCEVNGCSTPLGMENGKIENK' | 'Coagulation factor V' |
| 8902 | 'F5_No85' | 'EKPQSTISGLLGPTLYAEVGDIIK' | 'Coagulation factor V' |
| 8903 | 'F5_No86' | 'KVMYTQYEDESFTK' | 'Coagulation factor V' |
| 8904 | 'F5_No87' | 'SSMVDKIFEGNTNTK' | 'Coagulation factor V' |
| 8905 | 'F5_No88' | 'SQHLDNFSNQIGKHYKK' | 'Coagulation factor V' |
| 8906 | 'F5_No89' | 'LAAEFASKPWIQVDMQK' | 'Coagulation factor V' |
| 8907 | 'F5_No90' | 'KYLDSTFTK' | 'Coagulation factor V' |
| 8908 | 'F5_No91' | 'MYEQEWVR' | 'Coagulation factor V' |
| 8909 | 'F5_No92' | 'PYYSDVDIMR' | 'Coagulation factor V' |
| 8910 | 'F5_No93' | 'AGMQTPFLIMDRDCR' | 'Coagulation factor V' |
| 8911 | 'F5_No94' | 'HLQAGMQAYIDIK' | 'Coagulation factor V' |
| 8912 | 'PIP_No1' | 'TVQIAAVVDVIR' | 'Prolactin-inducible protein' |
| 8913 | 'PIP_No2' | 'YTACLCDDNPK' | 'Prolactin-inducible protein' |
| 8914 | 'PIP_No3' | 'TYLISSIPLQGAFNYK' | 'Prolactin-inducible protein' |
| 8915 | 'PIP_No4' | 'ELGICPDDAAVIPIK' | 'Prolactin-inducible protein' |
| 8916 | 'ANXA3_No1' | 'MLISILTER' | 'Annexin A3' |
| 8917 | 'ANXA3_No2' | 'GAGTNEDALIEILTTR' | 'Annexin A3' |
| 8918 | 'ANXA3_No3' | 'SEIDLLDIR' | 'Annexin A3' |
| 8919 | 'ANXA3_No4' | 'SLGDDISSETSGDFRK' | 'Annexin A3' |
| 8920 | 'ACTN1_No1' | 'LASDLLEWIR' | 'Alpha-actinin-1' |
| 8921 | 'ACTN1_No2' | 'FAIQDISVEETSAK' | 'Alpha-actinin-1' |
| 8922 | 'ACTN1_No3' | 'TINEVENQILTR' | 'Alpha-actinin-1' |
| 8923 | 'ACTN1_No4' | 'DGLGFCALIHR' | 'Alpha-actinin-1' |
| 8924 | 'ACTN1_No5' | 'MVSDINNAWGCLEQVEK' | 'Alpha-actinin-1' |
| 8925 | 'ACTN1_No6' | 'EGLLLWCQR' | 'Alpha-actinin-1' |
| 8926 | 'ACTN1_No7' | 'LAILGIHNEVSK' | 'Alpha-actinin-1' |
| 8927 | 'ACTN1_No8' | 'QFGAQANVIGPWIQTK' | 'Alpha-actinin-1' |
| 8928 | 'ACTN1_No9' | 'TFTAWCNSHLR' | 'Alpha-actinin-1' |
| 8929 | 'ACTN1_No10' | 'AIMTYVSSFYHAFSGAQK' | 'Alpha-actinin-1' |
| 8930 | 'ACTN1_No11' | 'AGTQIENIEEDFRDGLK' | 'Alpha-actinin-1' |
| 8931 | 'ACTN1_No12' | 'GYEEWLLNEIR' | 'Alpha-actinin-1' |
| 8932 | 'ACTN1_No13' | 'LVSIGAEEIVDGNVK' | 'Alpha-actinin-1' |
| 8933 | 'ACTN1_No14' | 'ILAGDKNYITMDELRR' | 'Alpha-actinin-1' |

| | A | B | C |
|---|---|---|---|
| 1 | **Peptide Name** | **Peptide Sequence** | **Peptide Description** |
| 8934 | 'ACTN1_No15' | 'ISIEMHGTLEDQLSHLR' | 'Alpha-actinin-1' |
| 8935 | 'ACTN1_No16' | 'QKDYETATLSEIK' | 'Alpha-actinin-1' |
| 8936 | 'ACTN1_No17' | 'MLDAEDIVGTARPDEK' | 'Alpha-actinin-1' |
| 8937 | 'ACTN1_No18' | 'LLETIDQLYLEYAK' | 'Alpha-actinin-1' |
| 8938 | 'ACTN1_No19' | 'HRPELIDYGK' | 'Alpha-actinin-1' |
| 8939 | 'ACTN1_No20' | 'ELPPDQAEYCIAR' | 'Alpha-actinin-1' |
| 8940 | 'ACTN1_No21' | 'KTFTAWCNSHLR' | 'Alpha-actinin-1' |
| 8941 | 'ACTN1_No22' | 'LEDFRDYR' | 'Alpha-actinin-1' |
| 8942 | 'ACTN1_No23' | 'NVNIQNFHISWK' | 'Alpha-actinin-1' |
| 8943 | 'ACTN1_No24' | 'VGWEQLLTTIAR' | 'Alpha-actinin-1' |
| 8944 | 'ACTN1_No25' | 'ALDFIASK' | 'Alpha-actinin-1' |
| 8945 | 'ACTN1_No26' | 'KAGTQIENIEEDFRDGLK' | 'Alpha-actinin-1' |
| 8946 | 'ACTN1_No27' | 'GISQEQMNEFR' | 'Alpha-actinin-1' |
| 8947 | 'ACTN1_No28' | 'ISIEMHGTLEDQLSHLR' | 'Alpha-actinin-1' |
| 8948 | 'ACTN1_No29' | 'AIMTYVSSFYHAFSGAQK' | 'Alpha-actinin-1' |
| 8949 | 'ACTN1_No30' | 'QKDYETATLSEIK' | 'Alpha-actinin-1' |
| 8950 | 'ACTN1_No31' | 'ASIHEAWTDGKEAMLR' | 'Alpha-actinin-1' |
| 8951 | 'ACTN1_No32' | 'LLETIDQLYLEYAKR' | 'Alpha-actinin-1' |
| 8952 | 'ACTN1_No33' | 'LRKDDPLTNLNTAFDVAEK' | 'Alpha-actinin-1' |
| 8953 | 'ACTN1_No34' | 'ILAGDKNYITMDELR' | 'Alpha-actinin-1' |
| 8954 | 'ACTN1_No35' | 'ATLPDADKER' | 'Alpha-actinin-1' |
| 8955 | 'ACTN1_No36' | 'LSNRPAFMPSEGR' | 'Alpha-actinin-1' |
| 8956 | 'ACTN1_No37' | 'CQLEINFNTLQTK' | 'Alpha-actinin-1' |
| 8957 | 'ACTN1_No38' | 'GISQEQMNEFR' | 'Alpha-actinin-1' |
| 8958 | 'ACTN1_No39' | 'VLAVNQENEQLMEDYEK' | 'Alpha-actinin-1' |
| 8959 | 'ACTN1_No40' | 'ICDQWDNLGALTQK' | 'Alpha-actinin-1' |
| 8960 | 'ACTN1_No41' | 'MVSDINNAWGCLEQVEK' | 'Alpha-actinin-1' |
| 8961 | 'ACTN1_No42' | 'IDQLEGDHQLIQEALIFDNK' | 'Alpha-actinin-1' |
| 8962 | 'ACTN1_No43' | 'VLAVNQENEQLMEDYEK' | 'Alpha-actinin-1' |
| 8963 | 'ACTN1_No44' | 'MLDAEDIVGTARPDEK' | 'Alpha-actinin-1' |
| 8964 | 'ACTN1_No45' | 'KQFGAQANVIGPWIQTK' | 'Alpha-actinin-1' |
| 8965 | 'ACTN1_No46' | 'IVQTYHVNMAGTNPYTTITPQEINGK' | 'Alpha-actinin-1' |
| 8966 | 'ACTN1_No47' | 'ETADTDTADQVMASFK' | 'Alpha-actinin-1' |
| 8967 | 'ACTN1_No48' | 'ASFNHFDRDHSGTLGPEEFK' | 'Alpha-actinin-1' |
| 8968 | 'ACTN1_No49' | 'ACLISLGYDIGNDPQGEAEFAR' | 'Alpha-actinin-1' |
| 8969 | 'ACTN1_No50' | 'ILAGDKNYITMDELR' | 'Alpha-actinin-1' |
| 8970 | 'ACTN1_No51' | 'DDPLTNLNTAFDVAEK' | 'Alpha-actinin-1' |
| 8971 | 'ACTN1_No52' | 'KDDPLTNLNTAFDVAEK' | 'Alpha-actinin-1' |
| 8972 | 'ACTN1_No53' | 'IVQTYHVNMAGTNPYTTITPQEINGK' | 'Alpha-actinin-1' |
| 8973 | 'ACTN1_No54' | 'LSNRPAFMPSEGR' | 'Alpha-actinin-1' |
| 8974 | 'ACTN1_No55' | 'GYEEWLLNEIRR' | 'Alpha-actinin-1' |
| 8975 | 'ACTN1_No56' | 'DLLLDPAWEK' | 'Alpha-actinin-1' |
| 8976 | 'ACTN1_No57' | 'NYITMDELR' | 'Alpha-actinin-1' |
| 8977 | 'ACTN1_No58' | 'HTNYTMEHIR' | 'Alpha-actinin-1' |

| | A | B | C |
|---|---|---|---|
| 1 | **Peptide Name** | **Peptide Sequence** | **Peptide Description** |
| 8978 | 'ACTN1_No59' | 'ASIHEAWTDGKEAMLR' | 'Alpha-actinin-1' |
| 8979 | 'ACTN1_No60' | 'RDQALTEEHAR' | 'Alpha-actinin-1' |
| 8980 | 'ACE_No1' | 'LFQELQPLYLNLHAYVR' | 'Angiotensin-converting enzyme' |
| 8981 | 'ACE_No2' | 'TATCWSLDPDLTNILASSR' | 'Angiotensin-converting enzyme' |
| 8982 | 'ACE_No3' | 'ENYNQEWWSLR' | 'Angiotensin-converting enzyme' |
| 8983 | 'ACE_No4' | 'GANPGFHEAIGDVLALSVSTPEHLHK' | 'Angiotensin-converting enzyme' |
| 8984 | 'ACE_No5' | 'SMYETPSLEQDLER' | 'Angiotensin-converting enzyme' |
| 8985 | 'ACE_No6' | 'EGANPGFHEAIGDVLALSVSTPK' | 'Angiotensin-converting enzyme' |
| 8986 | 'ACE_No7' | 'DMVGLDALDAQPLLK' | 'Angiotensin-converting enzyme' |
| 8987 | 'ACE_No8' | 'RQEEAALLSQEFAEAWGQK' | 'Angiotensin-converting enzyme' |
| 8988 | 'ACE_No9' | 'PLLDWLR' | 'Angiotensin-converting enzyme' |
| 8989 | 'ACE_No10' | 'EAGYEGPLHQCDIYR' | 'Angiotensin-converting enzyme' |
| 8990 | 'ACE_No11' | 'AALPAQELEEYNK' | 'Angiotensin-converting enzyme' |
| 8991 | 'ACE_No12' | 'DMVGLDALDAQPLLK' | 'Angiotensin-converting enzyme' |
| 8992 | 'CDH1_No1' | 'DTANWLEINPDTGAISTR' | 'Cadherin-1' |
| 8993 | 'CDH1_No2' | 'VFYSITGQGADTPPVGVFIIER' | 'Cadherin-1' |
| 8994 | 'CDH1_No3' | 'DWVIPPISCPENEKGPFPK' | 'Cadherin-1' |
| 8995 | 'CDH1_No4' | 'IWRDTANWLEINPDTGAISTR' | 'Cadherin-1' |
| 8996 | 'CDH1_No5' | 'GQVPENEANVVITTLK' | 'Cadherin-1' |
| 8997 | 'CDH1_No6' | 'VGTDGVITVK' | 'Cadherin-1' |
| 8998 | 'CDH1_No7' | 'AELDREDFEHVK' | 'Cadherin-1' |
| 8999 | 'SRC_No1' | 'LTTVCPTSKPQTQGLAK' | 'Proto-oncogene tyrosine-protein kinase Src' |
| 9000 | 'SRC_No2' | 'LFGGFNSSDTVTSPQR' | 'Proto-oncogene tyrosine-protein kinase Src' |
| 9001 | 'SRC_No3' | 'TQFNSLQQLVAYYSK' | 'Proto-oncogene tyrosine-protein kinase Src' |
| 9002 | 'SRC_No4' | 'KLDSGGFYITSR' | 'Proto-oncogene tyrosine-protein kinase Src' |
| 9003 | 'PEPD_No1' | 'VPLALFALNR' | 'Xaa-Pro dipeptidase' |
| 9004 | 'PEPD_No2' | 'AAATGPSFWLGNETLK' | 'Xaa-Pro dipeptidase' |
| 9005 | 'PEPD_No3' | 'IEEDVVVTDSGIELLTCVPR' | 'Xaa-Pro dipeptidase' |
| 9006 | 'PEPD_No4' | 'HSSYTCICGSGENSAVLHYGHAGAPNDR' | 'Xaa-Pro dipeptidase' |
| 9007 | 'PEPD_No5' | 'VGMKEYELESLFEHYCYSR' | 'Xaa-Pro dipeptidase' |
| 9008 | 'PEPD_No6' | 'FEVNNTILHPEIVECR' | 'Xaa-Pro dipeptidase' |
| 9009 | 'PEPD_No7' | 'YCTDTGVLFR' | 'Xaa-Pro dipeptidase' |
| 9010 | 'PEPD_No8' | 'NPAVQAGSIVVLQGGEETQR' | 'Xaa-Pro dipeptidase' |
| 9011 | 'PEPD_No9' | 'PGVWWPDMHR' | 'Xaa-Pro dipeptidase' |
| 9012 | 'PEPD_No10' | 'EYELESLFEHYCYSR' | 'Xaa-Pro dipeptidase' |
| 9013 | 'PEPD_No11' | 'TDMELEVLR' | 'Xaa-Pro dipeptidase' |
| 9014 | 'PEPD_No12' | 'KNPAVQAGSIVVLQGGEETQR' | 'Xaa-Pro dipeptidase' |
| 9015 | 'PEPD_No13' | 'AFTPFSGPK' | 'Xaa-Pro dipeptidase' |
| 9016 | 'PEPD_No14' | 'EASFDGISKFEVNNTILHPEIVECR' | 'Xaa-Pro dipeptidase' |
| 9017 | 'PEPD_No15' | 'TVEEIEACMAGCDK' | 'Xaa-Pro dipeptidase' |
| 9018 | 'PEPD_No16' | 'TDMELEVLR' | 'Xaa-Pro dipeptidase' |
| 9019 | 'GP1BB_No1' | 'LLPYLAEDELR' | 'Platelet glycoprotein Ib beta chain' |
| 9020 | 'GP1BB_No2' | 'LSLTDPLVAER' | 'Platelet glycoprotein Ib beta chain' |
| 9021 | 'GP1BB_No3' | 'TAHLGANPWR' | 'Platelet glycoprotein Ib beta chain' |

| | A | B | C |
|---|---|---|---|
| 1 | **Peptide Name** | **Peptide Sequence** | **Peptide Description** |
| 9022 | 'LAMP2_No1' | 'IPLNDLFR' | 'Lysosome-associated membrane glycoprotein 2' |
| 9023 | 'LAMP2_No2' | 'GILTVDELLAIR' | 'Lysosome-associated membrane glycoprotein 2' |
| 9024 | 'LAMP2_No3' | 'YLDFVFAVK' | 'Lysosome-associated membrane glycoprotein 2' |
| 9025 | 'RNH1_No1' | 'VLWLADCDVSDSSCSSLAATLLANHSLR' | 'Ribonuclease inhibitor' |
| 9026 | 'RNH1_No2' | 'LSLQNCCLTGAGCGVLSSTLR' | 'Ribonuclease inhibitor' |
| 9027 | 'RNH1_No3' | 'ELTVSNNDINEAGVR' | 'Ribonuclease inhibitor' |
| 9028 | 'RNH1_No4' | 'LQLEYCSLSAASCEPLASVLR' | 'Ribonuclease inhibitor' |
| 9029 | 'RNH1_No5' | 'ELSLAGNELGDEGAR' | 'Ribonuclease inhibitor' |
| 9030 | 'NCAM1_No1' | 'FFLCQVAGDAK' | 'Neural cell adhesion molecule 1' |
| 9031 | 'NCAM1_No2' | 'EASMEGIVTIVGLKPETTYAVR' | 'Neural cell adhesion molecule 1' |
| 9032 | 'NCAM1_No3' | 'CVVTGEDGSESEATVNVK' | 'Neural cell adhesion molecule 1' |
| 9033 | 'NCAM1_No4' | 'KVDKNDEAEYICIAENK' | 'Neural cell adhesion molecule 1' |
| 9034 | 'NCAM1_No5' | 'LEGQMGEDGNSIK' | 'Neural cell adhesion molecule 1' |
| 9035 | 'NCAM1_No6' | 'FIVLSNNYLQIR' | 'Neural cell adhesion molecule 1' |
| 9036 | 'NCAM1_No7' | 'DKDISWFSPNGEK' | 'Neural cell adhesion molecule 1' |
| 9037 | 'NCAM1_No8' | 'EASMEGIVTIVGLKPETTYAVR' | 'Neural cell adhesion molecule 1' |
| 9038 | 'NCAM1_No9' | 'GLGEISAASEFK' | 'Neural cell adhesion molecule 1' |
| 9039 | 'NCAM1_No10' | 'ALSSEWKPEIR' | 'Neural cell adhesion molecule 1' |
| 9040 | 'NCAM1_No11' | 'NAPTPQEFR' | 'Neural cell adhesion molecule 1' |
| 9041 | 'NCAM1_No12' | 'FFLCQVAGDAKDKDISWFSPNGEK' | 'Neural cell adhesion molecule 1' |
| 9042 | 'NCAM1_No13' | 'LPSGSDHVMLK' | 'Neural cell adhesion molecule 1' |
| 9043 | 'ICAM2_No1' | 'QVILTLQPTLVAVGK' | 'Intercellular adhesion molecule 2' |
| 9044 | 'ICAM2_No2' | 'ILLDEQAQWK' | 'Intercellular adhesion molecule 2' |
| 9045 | 'EEF2_No1' | 'TFCQLILDPIFK' | 'Elongation factor 2' |
| 9046 | 'EEF2_No2' | 'KIWCFGPDGTGPNILTDITK' | 'Elongation factor 2' |
| 9047 | 'EEF2_No3' | 'EDLYLKPIQR' | 'Elongation factor 2' |
| 9048 | 'KRT10_No1' | 'LKYENEVALR' | 'Keratin, type I cytoskeletal 10' |
| 9049 | 'KRT10_No2' | 'ALEESNYELEGK' | 'Keratin, type I cytoskeletal 10' |
| 9050 | 'KRT10_No3' | 'SQYEQLAEQNRK' | 'Keratin, type I cytoskeletal 10' |
| 9051 | 'KRT10_No4' | 'QSVEADINGLRR' | 'Keratin, type I cytoskeletal 10' |
| 9052 | 'KRT10_No5' | 'ELTTEIDNNIEQISSYK' | 'Keratin, type I cytoskeletal 10' |
| 9053 | 'KRT10_No6' | 'NQILNLTTDNANILLQIDNAR' | 'Keratin, type I cytoskeletal 10' |
| 9054 | 'KRT10_No7' | 'SKELTTEIDNNIEQISSYK' | 'Keratin, type I cytoskeletal 10' |
| 9055 | 'KRT10_No8' | 'GSLGGGFSSGGFSGGSFSR' | 'Keratin, type I cytoskeletal 10' |
| 9056 | 'KRT10_No9' | 'QSVEADINGLRR' | 'Keratin, type I cytoskeletal 10' |
| 9057 | 'KRT10_No10' | 'VLDELTLTK' | 'Keratin, type I cytoskeletal 10' |
| 9058 | 'KRT10_No11' | 'SQYEQLAEQNR' | 'Keratin, type I cytoskeletal 10' |
| 9059 | 'KRT10_No12' | 'AETECQNTEYQQLLDIK' | 'Keratin, type I cytoskeletal 10' |
| 9060 | 'KRT10_No13' | 'LENEIQTYR' | 'Keratin, type I cytoskeletal 10' |
| 9061 | 'KRT10_No14' | 'SLLEGEGSSGGGGR' | 'Keratin, type I cytoskeletal 10' |
| 9062 | 'KRT10_No15' | 'IRLENEIQTYR' | 'Keratin, type I cytoskeletal 10' |
| 9063 | 'KRT10_No16' | 'GSSGGGCFGGSSGGYGGLGGFGGGSFR' | 'Keratin, type I cytoskeletal 10' |
| 9064 | 'KRT10_No17' | 'NVQALEIELQSQLALK' | 'Keratin, type I cytoskeletal 10' |
| 9065 | 'KRT10_No18' | 'RVLDELTLTK' | 'Keratin, type I cytoskeletal 10' |

| | A | B | C |
|---|---|---|---|
| 1 | **Peptide Name** | **Peptide Sequence** | **Peptide Description** |
| 9066 | 'KRT10_No19' | 'ADLEMQIESLTEELAYLK' | 'Keratin, type I cytoskeletal 10' |
| 9067 | 'KRT10_No20' | 'ELTTEIDNNIEQISSYKSEITELRR' | 'Keratin, type I cytoskeletal 10' |
| 9068 | 'KRT10_No21' | 'SQYEQLAEQNRKDAEAWFNEK' | 'Keratin, type I cytoskeletal 10' |
| 9069 | 'KRT10_No22' | 'SEITELRR' | 'Keratin, type I cytoskeletal 10' |
| 9070 | 'KRT10_No23' | 'QSVEADINGLR' | 'Keratin, type I cytoskeletal 10' |
| 9071 | 'KRT10_No24' | 'DAEAWFNEK' | 'Keratin, type I cytoskeletal 10' |
| 9072 | 'KRT10_No25' | 'NVSTGDVNVEMNAAPGVDLTQLLNNMR' | 'Keratin, type I cytoskeletal 10' |
| 9073 | 'KRT10_No26' | 'NHEEEMKDLR' | 'Keratin, type I cytoskeletal 10' |
| 9074 | 'KRT10_No27' | 'ALEESNYELEGKIKEWYEK' | 'Keratin, type I cytoskeletal 10' |
| 9075 | 'KRT10_No28' | 'QSLEASLAETEGR' | 'Keratin, type I cytoskeletal 10' |
| 9076 | 'KRT10_No29' | 'NVSTGDVNVEMNAAPGVDLTQLLNNMR' | 'Keratin, type I cytoskeletal 10' |
| 9077 | 'KRT10_No30' | 'ADLEMQIESLTEELAYLKK' | 'Keratin, type I cytoskeletal 10' |
| 9078 | 'KRT10_No31' | 'ADLEMQIESLTEELAYLKK' | 'Keratin, type I cytoskeletal 10' |
| 9079 | 'KRT10_No32' | 'LAADDFRLKYENEVALR' | 'Keratin, type I cytoskeletal 10' |
| 9080 | 'KRT10_No33' | 'KDAEAWFNEK' | 'Keratin, type I cytoskeletal 10' |
| 9081 | 'KRT10_No34' | 'NVSTGDVNVEMNAAPGVDLTQLLNNMR' | 'Keratin, type I cytoskeletal 10' |
| 9082 | 'KRT10_No35' | 'IKEWYEKHGNSHQGEPR' | 'Keratin, type I cytoskeletal 10' |
| 9083 | 'KRT10_No36' | 'KNHEEEMKDLR' | 'Keratin, type I cytoskeletal 10' |
| 9084 | 'KRT10_No37' | 'NHEEEMKDLR' | 'Keratin, type I cytoskeletal 10' |
| 9085 | 'KRT10_No38' | 'YENEVALR' | 'Keratin, type I cytoskeletal 10' |
| 9086 | 'KRT10_No39' | 'SKELTTEIDNNIEQISSYKSEITELR' | 'Keratin, type I cytoskeletal 10' |
| 9087 | 'KRT10_No40' | 'KNHEEEMKDLR' | 'Keratin, type I cytoskeletal 10' |
| 9088 | 'KRT5_No1' | 'YEELQQTAGR' | 'Keratin, type II cytoskeletal 5' |
| 9089 | 'KRT5_No2' | 'AQYEEIANR' | 'Keratin, type II cytoskeletal 5' |
| 9090 | 'KRT5_No3' | 'ISISTSGGSFR' | 'Keratin, type II cytoskeletal 5' |
| 9091 | 'KRT5_No4' | 'DVDAAYMNKVELEAK' | 'Keratin, type II cytoskeletal 5' |
| 9092 | 'KRT5_No5' | 'NMQDLVEDFK' | 'Keratin, type II cytoskeletal 5' |
| 9093 | 'KRT5_No6' | 'TTAENEFVMLKK' | 'Keratin, type II cytoskeletal 5' |
| 9094 | 'KRT5_No7' | 'TTAENEFVMLKK' | 'Keratin, type II cytoskeletal 5' |
| 9095 | 'KRT5_No8' | 'VSLAGACGVGGYGSR' | 'Keratin, type II cytoskeletal 5' |
| 9096 | 'KRT5_No9' | 'SRTEAESWYQTK' | 'Keratin, type II cytoskeletal 5' |
| 9097 | 'KRT5_No10' | 'LAELEEALQK' | 'Keratin, type II cytoskeletal 5' |
| 9098 | 'KRT5_No11' | 'SFSTASAITPSVSR' | 'Keratin, type II cytoskeletal 5' |
| 9099 | 'KRT5_No12' | 'QCANLQNAIADAEQRGELALK' | 'Keratin, type II cytoskeletal 5' |
| 9100 | 'KRT5_No13' | 'TTAENEFVMLK' | 'Keratin, type II cytoskeletal 5' |
| 9101 | 'KRT5_No14' | 'TTAENEFVMLK' | 'Keratin, type II cytoskeletal 5' |
| 9102 | 'KRT5_No15' | 'DVDAAYMNKVELEAK' | 'Keratin, type II cytoskeletal 5' |
| 9103 | 'KRT5_No16' | 'QCANLQNAIADAEQR' | 'Keratin, type II cytoskeletal 5' |
| 9104 | 'KRT5_No17' | 'GLGVGFGSGGGSSSSVK' | 'Keratin, type II cytoskeletal 5' |
| 9105 | 'KRT5_No18' | 'LLREYQELMNTK' | 'Keratin, type II cytoskeletal 5' |
| 9106 | 'KRT5_No19' | 'SRTEAESWYQTKYEELQQTAGR' | 'Keratin, type II cytoskeletal 5' |
| 9107 | 'KRT5_No20' | 'VDALMDEINFMK' | 'Keratin, type II cytoskeletal 5' |
| 9108 | 'KRT5_No21' | 'QCANLQNAIADAEQRGELALK' | 'Keratin, type II cytoskeletal 5' |
| 9109 | 'KRT5_No22' | 'QCANLQNAIADAEQRGELALKDAR' | 'Keratin, type II cytoskeletal 5' |

| | A | B | C |
|---|---|---|---|
| 1 | **Peptide Name** | **Peptide Sequence** | **Peptide Description** |
| 9110 | 'KRT5_No23' | 'VDALMDEINFMK' | 'Keratin, type II cytoskeletal 5' |
| 9111 | 'KRT5_No24' | 'VDALMDEINFMK' | 'Keratin, type II cytoskeletal 5' |
| 9112 | 'KRT5_No25' | 'VDALMDEINFMK' | 'Keratin, type II cytoskeletal 5' |
| 9113 | 'KRT5_No26' | 'TEAESWYQTKYEELQQTAGR' | 'Keratin, type II cytoskeletal 5' |
| 9114 | 'KRT5_No27' | 'NKLAELEEALQK' | 'Keratin, type II cytoskeletal 5' |
| 9115 | 'KRT5_No28' | 'NMQDLVEDFK' | 'Keratin, type II cytoskeletal 5' |
| 9116 | 'KRT5_No29' | 'LLREYQELMNTK' | 'Keratin, type II cytoskeletal 5' |
| 9117 | 'KRT5_No30' | 'RISISTSGGSFR' | 'Keratin, type II cytoskeletal 5' |
| 9118 | 'PDIA4_No1' | 'VSQGQLVVMQPEK' | 'Protein disulfide-isomerase A4' |
| 9119 | 'PDIA4_No2' | 'MDATANDVPSDR' | 'Protein disulfide-isomerase A4' |
| 9120 | 'PDIA4_No3' | 'EVSQPDWTPPPEVTLVLTK' | 'Protein disulfide-isomerase A4' |
| 9121 | 'PDIA4_No4' | 'YGIVDYMIEQSGPPSK' | 'Protein disulfide-isomerase A4' |
| 9122 | 'PDIA4_No5' | 'IDATSASVLASR' | 'Protein disulfide-isomerase A4' |
| 9123 | 'PDIA4_No6' | 'DLGLSESGEDVNAAILDESGKK' | 'Protein disulfide-isomerase A4' |
| 9124 | 'C6_No1' | 'IGESIELTCPK' | 'Complement component C6' |
| 9125 | 'C6_No2' | 'GEVLDNSFTGGICK' | 'Complement component C6' |
| 9126 | 'C6_No3' | 'DLHLSDVFLK' | 'Complement component C6' |
| 9127 | 'C6_No4' | 'TFSEWLESVK' | 'Complement component C6' |
| 9128 | 'C6_No5' | 'AKDLHLSDVFLK' | 'Complement component C6' |
| 9129 | 'C6_No6' | 'YTCQGNSWTPPISNSLTCEKDTLTK' | 'Complement component C6' |
| 9130 | 'C6_No7' | 'ESCGYDTCYDWEK' | 'Complement component C6' |
| 9131 | 'C6_No8' | 'YNPIPSVQLMGNGFHFLAGEPR' | 'Complement component C6' |
| 9132 | 'C6_No9' | 'YTCQGNSWTPPISNSLTCEK' | 'Complement component C6' |
| 9133 | 'C6_No10' | 'QLEWGLER' | 'Complement component C6' |
| 9134 | 'C6_No11' | 'VPANLENVGFEVQTAEDDLKTDFYK' | 'Complement component C6' |
| 9135 | 'C6_No12' | 'KYNPIPSVQLMGNGFHFLAGEPR' | 'Complement component C6' |
| 9136 | 'C6_No13' | 'YYQENFCEQICSK' | 'Complement component C6' |
| 9137 | 'C6_No14' | 'ENPAVIDFELAPIVDLVR' | 'Complement component C6' |
| 9138 | 'C6_No15' | 'TLNICEVGTIR' | 'Complement component C6' |
| 9139 | 'C6_No16' | 'CVCLLPPQCFK' | 'Complement component C6' |
| 9140 | 'C6_No17' | 'KESCGYDTCYDWEK' | 'Complement component C6' |
| 9141 | 'C6_No18' | 'CPINCLLGDFGPWSDCDPCIEK' | 'Complement component C6' |
| 9142 | 'C6_No19' | 'LSEKHEGSFIQGAEK' | 'Complement component C6' |
| 9143 | 'C6_No20' | 'SEYGAALAWEK' | 'Complement component C6' |
| 9144 | 'C6_No21' | 'SVLRPSQFGGQPCTAPLVAFQPCIPSK' | 'Complement component C6' |
| 9145 | 'C6_No22' | 'CLNNQQLHFLHIGSCQDGR' | 'Complement component C6' |
| 9146 | 'C6_No23' | 'HRQIVVDKYYQENFCEQICSK' | 'Complement component C6' |
| 9147 | 'C6_No24' | 'TECIKPVVQEVLTITPFQR' | 'Complement component C6' |
| 9148 | 'C6_No25' | 'NSGLTEEEAKHCVR' | 'Complement component C6' |
| 9149 | 'C6_No26' | 'KLECNGENDCGDNSDERDCGR' | 'Complement component C6' |
| 9150 | 'C6_No27' | 'ALNHLPLEYNSALYSR' | 'Complement component C6' |
| 9151 | 'C6_No28' | 'TSNPYRVPANLENVGFEVQTAEDDLKTDFYK' | 'Complement component C6' |
| 9152 | 'C6_No29' | 'SEYGAALAWEKGSSGLEEK' | 'Complement component C6' |
| 9153 | 'C6_No30' | 'FDPCQCAPCPNNGR' | 'Complement component C6' |

| | A | B | C |
|---|---|---|---|
| 1 | **Peptide Name** | **Peptide Sequence** | **Peptide Description** |
| 9154 | 'C6_No31' | 'KLECNGENDCGDNSDER' | 'Complement component C6' |
| 9155 | 'C6_No32' | 'ESCGYDTCYDWEKCSASTSK' | 'Complement component C6' |
| 9156 | 'C6_No33' | 'KYNPIPSVQLMGNGFHFLAGEPR' | 'Complement component C6' |
| 9157 | 'C6_No34' | 'QGDVECQRTECIKPVVQEVLTITPFQR' | 'Complement component C6' |
| 9158 | 'C6_No35' | 'HEGSFIQGAEK' | 'Complement component C6' |
| 9159 | 'C6_No36' | 'VPANLENVGFEVQTAEDDLK' | 'Complement component C6' |
| 9160 | 'C6_No37' | 'LECNGENDCGDNSDERDCGR' | 'Complement component C6' |
| 9161 | 'C6_No38' | 'QIVVDKYYQENFCEQICSK' | 'Complement component C6' |
| 9162 | 'C6_No39' | 'CLPDGTWR' | 'Complement component C6' |
| 9163 | 'C6_No40' | 'QIVVDKYYQENFCEQICSK' | 'Complement component C6' |
| 9164 | 'C6_No41' | 'CTTNKLSEKHEGSFIQGAEK' | 'Complement component C6' |
| 9165 | 'C6_No42' | 'GFVVAGPSR' | 'Complement component C6' |
| 9166 | 'C6_No43' | 'NIPCAVTKR' | 'Complement component C6' |
| 9167 | 'C6_No44' | 'SNAVDGQWGCWSSWSTCDATYKR' | 'Complement component C6' |
| 9168 | 'C6_No45' | 'SENINHNSAFK' | 'Complement component C6' |
| 9169 | 'C6_No46' | 'CPINCLLGDFGPWSDCDPCIEKQSK' | 'Complement component C6' |
| 9170 | 'C6_No47' | 'MGSSTSEKTLNICEVGTIR' | 'Complement component C6' |
| 9171 | 'C6_No48' | 'MEILHPGK' | 'Complement component C6' |
| 9172 | 'C6_No49' | 'QGDVECQRTECIKPVVQEVLTITPFQR' | 'Complement component C6' |
| 9173 | 'C6_No50' | 'NSGLTEEEAK' | 'Complement component C6' |
| 9174 | 'C6_No51' | 'RSENINHNSAFK' | 'Complement component C6' |
| 9175 | 'C6_No52' | 'DLTSLGHNENQQGSFSSQGGSSFSVPIFYSSK' | 'Complement component C6' |
| 9176 | 'C6_No53' | 'KALQEYAAK' | 'Complement component C6' |
| 9177 | 'C6_No54' | 'QIVVDKYYQENFCEQICSKQETR' | 'Complement component C6' |
| 9178 | 'C6_No55' | 'MEILHPGK' | 'Complement component C6' |
| 9179 | 'C6_No56' | 'GGRSEYGAALAWEK' | 'Complement component C6' |
| 9180 | 'C6_No57' | 'IEEADCK' | 'Complement component C6' |
| 9181 | 'C6_No58' | 'TSNPYRVPANLENVGFEVQTAEDDLK' | 'Complement component C6' |
| 9182 | 'C6_No59' | 'IEEADCKNKFR' | 'Complement component C6' |
| 9183 | 'C6_No60' | 'NIPCAVTK' | 'Complement component C6' |
| 9184 | 'C6_No61' | 'ECNNPAPQR' | 'Complement component C6' |
| 9185 | 'C6_No62' | 'EVDLPEIEADSGCPQPVPPENGFIR' | 'Complement component C6' |
| 9186 | 'C6_No63' | 'YNPIPSVQLMGNGFHFLAGEPR' | 'Complement component C6' |
| 9187 | 'C6_No64' | 'GGRSEYGAALAWEKGSSGLEEK' | 'Complement component C6' |
| 9188 | 'C6_No65' | 'TRECNNPAPQR' | 'Complement component C6' |
| 9189 | 'C6_No66' | 'MGSSTSEKTLNICEVGTIR' | 'Complement component C6' |
| 9190 | 'C6_No67' | 'GSSGLEEKTFSEWLESVK' | 'Complement component C6' |
| 9191 | 'C6_No68' | 'QIVVDKYYQENFCEQICSKQETR' | 'Complement component C6' |
| 9192 | 'C6_No69' | 'DLTSLGHNENQQGSFSSQGGSSFSVPIFYSSKR' | 'Complement component C6' |
| 9193 | 'C6_No70' | 'YTCQGNSWTPPISNSLTCEKDTLTKLK' | 'Complement component C6' |
| 9194 | 'C6_No71' | 'SENINHNSAFKQAIQASHKK' | 'Complement component C6' |
| 9195 | 'C6_No72' | 'QSGSECICMSPEEDCSHHSEDLCVFDTDSNDYFTSPACK' | 'Complement component C6' |
| 9196 | 'C6_No73' | 'ALQEYAAK' | 'Complement component C6' |
| 9197 | 'CEACAM1' | 'QIVGYAIGTQQATPGPANSGR' | 'Carcinoembryonic antigen-related cell adhesion molecule 1' |

| | A | B | C |
|---|---|---|---|
| 1 | **Peptide Name** | **Peptide Sequence** | **Peptide Description** |
| 9198 | 'TPT1' | 'DLISHDEMFSDIYK' | 'Translationally-controlled tumor protein' |
| 9199 | 'ALAD_No1' | 'MQPQSVLHSGYFHPLLR' | 'Delta-aminolevulinic acid dehydratase' |
| 9200 | 'ALAD_No2' | 'GSAADSEESPAIEAIHLLR' | 'Delta-aminolevulinic acid dehydratase' |
| 9201 | 'ALAD_No3' | 'AAVLEAMTAFR' | 'Delta-aminolevulinic acid dehydratase' |
| 9202 | 'ALAD_No4' | 'CVLIFGVPSR' | 'Delta-aminolevulinic acid dehydratase' |
| 9203 | 'ALAD_No5' | 'RLEEMLRPLVEEGLR' | 'Delta-aminolevulinic acid dehydratase' |
| 9204 | 'PRG2_No1' | 'VVGIPGCQTCR' | 'Bone marrow proteoglycan' |
| 9205 | 'PRG2_No2' | 'SLQTFSQAWFTCR' | 'Bone marrow proteoglycan' |
| 9206 | 'PRG2_No3' | 'WNFAYWAAHQPWSR' | 'Bone marrow proteoglycan' |
| 9207 | 'PRG2_No4' | 'GNLVSIHNFNINYR' | 'Bone marrow proteoglycan' |
| 9208 | 'PRG2_No5' | 'TLPEDEETPEQEMEETPCR' | 'Bone marrow proteoglycan' |
| 9209 | 'PRG2_No6' | 'RFQWVDGSR' | 'Bone marrow proteoglycan' |
| 9210 | 'PRG2_No7' | 'FQWVDGSR' | 'Bone marrow proteoglycan' |
| 9211 | 'PRG2_No8' | 'TLPEDEETPEQEMEETPCR' | 'Bone marrow proteoglycan' |
| 9212 | 'LCP1_No1' | 'LSPEELLLR' | 'Plastin-2' |
| 9213 | 'LCP1_No2' | 'ISFDEFIK' | 'Plastin-2' |
| 9214 | 'LCP1_No3' | 'NEALIALLR' | 'Plastin-2' |
| 9215 | 'LCP1_No4' | 'VYALPEDLVEVNPK' | 'Plastin-2' |
| 9216 | 'LCP1_No5' | 'IKVPVDWNR' | 'Plastin-2' |
| 9217 | 'LCP1_No6' | 'AYYHLLEQVAPKGDEEGVPAVVIDMSGLR' | 'Plastin-2' |
| 9218 | 'LCP1_No7' | 'IGNFSTDIKDSK' | 'Plastin-2' |
| 9219 | 'LCP1_No8' | 'AECMLQQAER' | 'Plastin-2' |
| 9220 | 'LCP1_No9' | 'YPALHKPENQDIDWGALEGETREER' | 'Plastin-2' |
| 9221 | 'LCP1_No10' | 'YPALHKPENQDIDWGALEGETR' | 'Plastin-2' |
| 9222 | 'LCP1_No11' | 'AYYHLLEQVAPK' | 'Plastin-2' |
| 9223 | 'LCP1_No12' | 'TENLNDDEKLNNAK' | 'Plastin-2' |
| 9224 | 'LCP1_No13' | 'MINLSVPDTIDER' | 'Plastin-2' |
| 9225 | 'LCP1_No14' | 'HVIPMNPNTNDLFNAVGDGIVLCK' | 'Plastin-2' |
| 9226 | 'LCP1_No15' | 'YAFVNWINK' | 'Plastin-2' |
| 9227 | 'LCP1_No16' | 'WANYHLENAGCNK' | 'Plastin-2' |
| 9228 | 'LCP1_No17' | 'MVMTVFACLMGK' | 'Plastin-2' |
| 9229 | 'LCP1_No18' | 'GSVSDEEMMELR' | 'Plastin-2' |
| 9230 | 'LCP1_No19' | 'GDEEGVPAVVIDMSGLR' | 'Plastin-2' |
| 9231 | 'LCP1_No20' | 'YTLNILEEIGGGQK' | 'Plastin-2' |
| 9232 | 'LCP1_No21' | 'QFVTATDVVR' | 'Plastin-2' |
| 9233 | 'LCP1_No22' | 'MINLSVPDTIDER' | 'Plastin-2' |
| 9234 | 'LCP1_No23' | 'RYTLNILEEIGGGQK' | 'Plastin-2' |
| 9235 | 'LCP1_No24' | 'FSLVGIGGQDLNEGNR' | 'Plastin-2' |
| 9236 | 'LCP1_No25' | 'EGICAIGGTSEQSSVGTQHSYSEEEK' | 'Plastin-2' |
| 9237 | 'LCP1_No26' | 'AACLPLPGYR' | 'Plastin-2' |
| 9238 | 'LCP1_No27' | 'HVIPMNPNTNDLFNAVGDGIVLCK' | 'Plastin-2' |
| 9239 | 'LCP1_No28' | 'QFVTATDVVR' | 'Plastin-2' |
| 9240 | 'LCP1_No29' | 'EITENLMATGDLDQDGR' | 'Plastin-2' |
| 9241 | 'LCP1_No30' | 'GDEEGVPAVVIDMSGLR' | 'Plastin-2' |

| | A | B | C |
|---|---|---|---|
| 1 | **Peptide Name** | **Peptide Sequence** | **Peptide Description** |
| 9242 | 'LCP1_No31' | 'VREITENLMATGDLDQDGR' | 'Plastin-2' |
| 9243 | 'LCP1_No32' | 'VPVDWNR' | 'Plastin-2' |
| 9244 | 'LCP1_No33' | 'NWMNSLGVNPR' | 'Plastin-2' |
| 9245 | 'LCP1_No34' | 'NWMNSLGVNPR' | 'Plastin-2' |
| 9246 | 'CD59_No1' | 'FEHCNFNDVTTR' | 'CD59 glycoprotein' |
| 9247 | 'CD59_No2' | 'LRENELTYYCCK' | 'CD59 glycoprotein' |
| 9248 | 'SELL_No1' | 'SYYWIGIR' | 'L-selectin' |
| 9249 | 'SELL_No2' | 'NKEDCVEIYIK' | 'L-selectin' |
| 9250 | 'SELL_No3' | 'SLTEEAENWGDGEPNNKK' | 'L-selectin' |
| 9251 | 'SELL_No4' | 'NKEDCVEIYIKR' | 'L-selectin' |
| 9252 | 'SELL_No5' | 'SYYWIGIRK' | 'L-selectin' |
| 9253 | 'SELL_No6' | 'KTICESSGIWSNPSPICQK' | 'L-selectin' |
| 9254 | 'SELL_No7' | 'SLTEEAENWGDGEPNNK' | 'L-selectin' |
| 9255 | 'MIF_No1' | 'PMFIVNTNVPR' | 'Macrophage migration inhibitory factor' |
| 9256 | 'MIF_No2' | 'LLCGLLAER' | 'Macrophage migration inhibitory factor' |
| 9257 | 'FDPS_No1' | 'QDFVQHFSQIVR' | 'Farnesyl pyrophosphate synthase' |
| 9258 | 'FDPS_No2' | 'QDFVQHFSQIVR' | 'Farnesyl pyrophosphate synthase' |
| 9259 | 'NID1_No1' | 'QAEVTFVGHPGNLVIK' | 'Nidogen-1' |
| 9260 | 'NID1_No2' | 'ASLHGGEPTTIIR' | 'Nidogen-1' |
| 9261 | 'NID1_No3' | 'KALEGLQYPFAVTSYGK' | 'Nidogen-1' |
| 9262 | 'NID1_No4' | 'QAEVTFVGHPGNLVIK' | 'Nidogen-1' |
| 9263 | 'NID1_No5' | 'NIFWTDSNLDRIEVAK' | 'Nidogen-1' |
| 9264 | 'NID1_No6' | 'VLFETDLVNPR' | 'Nidogen-1' |
| 9265 | 'NID1_No7' | 'EYTVTEPERDGASPSR' | 'Nidogen-1' |
| 9266 | 'NID1_No8' | 'ALEGLQYPFAVTSYGK' | 'Nidogen-1' |
| 9267 | 'NID1_No9' | 'QDLGSPEGIAVDHLGR' | 'Nidogen-1' |
| 9268 | 'NID1_No10' | 'SNGAYNIFANDRESVENLAK' | 'Nidogen-1' |
| 9269 | 'NID1_No11' | 'VIIGLAFDCVDK' | 'Nidogen-1' |
| 9270 | 'NID1_No12' | 'GNLYWTDWNRDNPK' | 'Nidogen-1' |
| 9271 | 'NID1_No13' | 'IFVGSSQVPIVFENTDLHSYVVMNHGR' | 'Nidogen-1' |
| 9272 | 'NID1_No14' | 'EGSPDALQNPCYIGTHGCDTNAACR' | 'Nidogen-1' |
| 9273 | 'NID1_No15' | 'TCYDIDECSEQPSVCGSHTICNNHPGTFR' | 'Nidogen-1' |
| 9274 | 'NID1_No16' | 'VYYREDLSPSITQR' | 'Nidogen-1' |
| 9275 | 'NID1_No17' | 'QDLGSPEGIAVDHLGR' | 'Nidogen-1' |
| 9276 | 'NID1_No18' | 'ASLHGGEPTTIIRQDLGSPEGIAVDHLGR' | 'Nidogen-1' |
| 9277 | 'PKM_No1' | 'GIFPVLCKDPVQEAWAEDVDLR' | 'Pyruvate kinase PKM' |
| 9278 | 'PKM_No2' | 'DPVQEAWAEDVDLR' | 'Pyruvate kinase PKM' |
| 9279 | 'PKM_No3' | 'IYVDDGLISLQVK' | 'Pyruvate kinase PKM' |
| 9280 | 'PKM_No4' | 'EAEAAIYHLQLFEELRR' | 'Pyruvate kinase PKM' |
| 9281 | 'PKM_No5' | 'GADFLVTEVENGGSLGSK' | 'Pyruvate kinase PKM' |
| 9282 | 'PKM_No6' | 'GVNLPGAAVDLPAVSEKDIQDLK' | 'Pyruvate kinase PKM' |
| 9283 | 'PKM_No7' | 'LNFSHGTHEYHAETIK' | 'Pyruvate kinase PKM' |
| 9284 | 'PKM_No8' | 'LAPITSDPTEATAVGAVEASFK' | 'Pyruvate kinase PKM' |
| 9285 | 'PKM_No9' | 'SVETLKEMIK' | 'Pyruvate kinase PKM' |

| | A | B | C |
|---|---|---|---|
| 1 | **Peptide Name** | **Peptide Sequence** | **Peptide Description** |
| 9286 | 'PKM_No10' | 'GIFPVLCK' | 'Pyruvate kinase PKM' |
| 9287 | 'PKM_No11' | 'RFDEILEASDGIMVAR' | 'Pyruvate kinase PKM' |
| 9288 | 'PKM_No12' | 'AEGSDVANAVLDGADCIMLSGETAKGDYPLEAVR' | 'Pyruvate kinase PKM' |
| 9289 | 'PKM_No13' | 'LDIDSPPITAR' | 'Pyruvate kinase PKM' |
| 9290 | 'PKM_No14' | 'AGKPVICATQMLESMIK' | 'Pyruvate kinase PKM' |
| 9291 | 'PKM_No15' | 'IISKIENHEGVR' | 'Pyruvate kinase PKM' |
| 9292 | 'PKM_No16' | 'KGVNLPGAAVDLPAVSEKDIQDLK' | 'Pyruvate kinase PKM' |
| 9293 | 'PKM_No17' | 'LNFSHGTHEYHAETIKNVR' | 'Pyruvate kinase PKM' |
| 9294 | 'PKM_No18' | 'ITLDNAYMEK' | 'Pyruvate kinase PKM' |
| 9295 | 'PKM_No19' | 'TATESFASDPILYRPVAVALDTKGPEIR' | 'Pyruvate kinase PKM' |
| 9296 | 'PKM_No20' | 'RFDEILEASDGIMVAR' | 'Pyruvate kinase PKM' |
| 9297 | 'PKM_No21' | 'CDENILWLDYK' | 'Pyruvate kinase PKM' |
| 9298 | 'PKM_No22' | 'GADFLVTEVENGGSLGSKK' | 'Pyruvate kinase PKM' |
| 9299 | 'PKM_No23' | 'KGDVVIVLTGWRPGSGFTNTMR' | 'Pyruvate kinase PKM' |
| 9300 | 'PKM_No24' | 'FDEILEASDGIMVAR' | 'Pyruvate kinase PKM' |
| 9301 | 'PKM_No25' | 'AGKPVICATQMLESMIK' | 'Pyruvate kinase PKM' |
| 9302 | 'PKM_No26' | 'GVNLPGAAVDLPAVSEK' | 'Pyruvate kinase PKM' |
| 9303 | 'PKM_No27' | 'YRPRAPIIAVTR' | 'Pyruvate kinase PKM' |
| 9304 | 'HSP90B1_No1' | 'LISLTDENALSGNEELTVK' | 'Endoplasmin' |
| 9305 | 'HSP90B1_No2' | 'NLLHVTDTGVGMTREELVK' | 'Endoplasmin' |
| 9306 | 'HSP90B1_No3' | 'KYSQFINFPIYVWSSK' | 'Endoplasmin' |
| 9307 | 'HSP90B1_No4' | 'FQSSHHPTDITSLDQYVER' | 'Endoplasmin' |
| 9308 | 'HSP90B1_No5' | 'SILFVPTSAPR' | 'Endoplasmin' |
| 9309 | 'HSP90B1_No6' | 'RVFITDDFHDMMPK' | 'Endoplasmin' |
| 9310 | 'HSP90B1_No7' | 'DISTNYYASQK' | 'Endoplasmin' |
| 9311 | 'HSP90B1_No8' | 'LTESPCALVASQYGWSGNMER' | 'Endoplasmin' |
| 9312 | 'HSP90B1_No9' | 'VFITDDFHDMMPK' | 'Endoplasmin' |
| 9313 | 'HSP90B1_No10' | 'EVEEDEYKAFYK' | 'Endoplasmin' |
| 9314 | 'HSP90B1_No11' | 'EAVEKEFEPLLNWMK' | 'Endoplasmin' |
| 9315 | 'HSP90B1_No12' | 'IADDKYNDTFWK' | 'Endoplasmin' |
| 9316 | 'HSP90B1_No13' | 'NLLHVTDTGVGMTR' | 'Endoplasmin' |
| 9317 | 'HSP90B1_No14' | 'KGYEVIYLTEPVDEYCIQALPEFDGKR' | 'Endoplasmin' |
| 9318 | 'HSP90B1_No15' | 'EEEAIQLDGLNASQIR' | 'Endoplasmin' |
| 9319 | 'HSP90B1_No16' | 'EAVEKEFEPLLNWMKDK' | 'Endoplasmin' |
| 9320 | 'HSP90B1_No17' | 'TETVEEPMEEEEAAKEEK' | 'Endoplasmin' |
| 9321 | 'HSP90B1_No18' | 'IADDKYNDTFWKEFGTNIK' | 'Endoplasmin' |
| 9322 | 'HSP90B1_No19' | 'GVVDSDDLPLNVSR' | 'Endoplasmin' |
| 9323 | 'HSP90B1_No20' | 'TETVEEPMEEEEAAKEEK' | 'Endoplasmin' |
| 9324 | 'HSP90B1_No21' | 'LTESPCALVASQYGWSGNMER' | 'Endoplasmin' |
| 9325 | 'HSP90B1_No22' | 'NLLHVTDTGVGMTREELVK' | 'Endoplasmin' |
| 9326 | 'GP9_No1' | 'TPEALLQVR' | 'Platelet glycoprotein IX' |
| 9327 | 'GP9_No2' | 'GHGLTALPALPAR' | 'Platelet glycoprotein IX' |
| 9328 | 'GP9_No3' | 'LWLEDRTPEALLQVR' | 'Platelet glycoprotein IX' |
| 9329 | 'GP9_No4' | 'CASPSLAAHGPLGR' | 'Platelet glycoprotein IX' |

| | A | B | C |
|---|---|---|---|
| 1 | **Peptide Name** | **Peptide Sequence** | **Peptide Description** |
| 9330 | 'GP9_No5' | 'ALETMGLWVDCR' | 'Platelet glycoprotein IX' |
| 9331 | 'MMP9_No1' | 'QLSLPETGELDSATLK' | 'Matrix metalloproteinase-9' |
| 9332 | 'MMP9_No2' | 'LGLGADVAQVTGALR' | 'Matrix metalloproteinase-9' |
| 9333 | 'MMP9_No3' | 'MFPGVPLDTHDVFQYR' | 'Matrix metalloproteinase-9' |
| 9334 | 'MMP9_No4' | 'LWCATTSNFDSDKK' | 'Matrix metalloproteinase-9' |
| 9335 | 'MMP9_No5' | 'FTEGPPLHKDDVNGIR' | 'Matrix metalloproteinase-9' |
| 9336 | 'MMP9_No6' | 'SLGPALLLLQK' | 'Matrix metalloproteinase-9' |
| 9337 | 'MMP9_No7' | 'LFGFCPTR' | 'Matrix metalloproteinase-9' |
| 9338 | 'MMP9_No8' | 'QSTLVLFPGDLR' | 'Matrix metalloproteinase-9' |
| 9339 | 'MMP9_No9' | 'QVWVYTGASVLGPR' | 'Matrix metalloproteinase-9' |
| 9340 | 'MMP9_No10' | 'FGNADGAACHFPFIFEGR' | 'Matrix metalloproteinase-9' |
| 9341 | 'MMP9_No11' | 'SDGLPWCSTTANYDTDDRFGFCPSER' | 'Matrix metalloproteinase-9' |
| 9342 | 'MMP9_No12' | 'QSTLVLFPGDLR' | 'Matrix metalloproteinase-9' |
| 9343 | 'DARS' | 'IGSCTQQDVELHVQK' | 'Aspartate--tRNA ligase, cytoplasmic' |
| 9344 | 'JUP_No1' | 'ALMGSPQLVAAVVR' | 'Junction plakoglobin' |
| 9345 | 'JUP_No2' | 'VSVELTNSLFK' | 'Junction plakoglobin' |
| 9346 | 'JUP_No3' | 'EGLLAIFK' | 'Junction plakoglobin' |
| 9347 | 'JUP_No4' | 'LIILANGGPQALVQIMR' | 'Junction plakoglobin' |
| 9348 | 'JUP_No5' | 'LLNQPNQWPLVK' | 'Junction plakoglobin' |
| 9349 | 'JUP_No6' | 'TLVTQNSGVEALIHAILR' | 'Junction plakoglobin' |
| 9350 | 'JUP_No7' | 'LIILANGGPQALVQIMR' | 'Junction plakoglobin' |
| 9351 | 'JUP_No8' | 'ISEDKNPDYR' | 'Junction plakoglobin' |
| 9352 | 'JUP_No9' | 'SAIVHLINYQDDAELATR' | 'Junction plakoglobin' |
| 9353 | 'JUP_No10' | 'NEGTATYAAAVLFR' | 'Junction plakoglobin' |
| 9354 | 'JUP_No11' | 'NLALCPANHAPLQEAAVIPR' | 'Junction plakoglobin' |
| 9355 | 'JUP_No12' | 'MEEIVEGCTGALHILAR' | 'Junction plakoglobin' |
| 9356 | 'JUP_No13' | 'NLSDVATKQEGLESVLK' | 'Junction plakoglobin' |
| 9357 | 'JUP_No14' | 'ALMGSPQLVAAVVR' | 'Junction plakoglobin' |
| 9358 | 'JUP_No15' | 'LAEPSQLLK' | 'Junction plakoglobin' |
| 9359 | 'JUP_No16' | 'LNYGIPAIVK' | 'Junction plakoglobin' |
| 9360 | 'CPA1' | 'ISVADEAQVQK' | 'Carboxypeptidase A1' |
| 9361 | 'FABP4_No1' | 'LVSSENFDDYMK' | 'Fatty acid-binding protein, adipocyte' |
| 9362 | 'FABP4_No2' | 'STITLDGGVLVHVQK' | 'Fatty acid-binding protein, adipocyte' |
| 9363 | 'FABP4_No3' | 'CDAFVGTWK' | 'Fatty acid-binding protein, adipocyte' |
| 9364 | 'ANPEP_No1' | 'YLSYTLNPDLIR' | 'Aminopeptidase N' |
| 9365 | 'ANPEP_No2' | 'ELWILNR' | 'Aminopeptidase N' |
| 9366 | 'ANPEP_No3' | 'MLSSFLSEDVFK' | 'Aminopeptidase N' |
| 9367 | 'ANPEP_No4' | 'QVTPLFIHFR' | 'Aminopeptidase N' |
| 9368 | 'ANPEP_No5' | 'VVATTQMQAADAR' | 'Aminopeptidase N' |
| 9369 | 'ANPEP_No6' | 'AQIINDAFNLASAHK' | 'Aminopeptidase N' |
| 9370 | 'ANPEP_No7' | 'GVGGSQPPDIDKTELVEPTEYLVVHLK' | 'Aminopeptidase N' |
| 9371 | 'ANPEP_No8' | 'QVTPLFIHFR' | 'Aminopeptidase N' |
| 9372 | 'ANPEP_No9' | 'YRLPNTLKPDSYR' | 'Aminopeptidase N' |
| 9373 | 'ANPEP_No10' | 'RFSTEYELQQLEQFKK' | 'Aminopeptidase N' |

| | A | B | C |
|---|---|---|---|
| 1 | **Peptide Name** | **Peptide Sequence** | **Peptide Description** |
| 9374 | 'ANPEP_No11' | 'QQQDYWLIDVR' | 'Aminopeptidase N' |
| 9375 | 'ANPEP_No12' | 'QQQDYWLIDVR' | 'Aminopeptidase N' |
| 9376 | 'ANPEP_No13' | 'EATDVIIIHSK' | 'Aminopeptidase N' |
| 9377 | 'ANPEP_No14' | 'FSTEYELQQLEQFKK' | 'Aminopeptidase N' |
| 9378 | 'ANPEP_No15' | 'VTLRPYLTPNDR' | 'Aminopeptidase N' |
| 9379 | 'ANPEP_No16' | 'DHSAIPVINR' | 'Aminopeptidase N' |
| 9380 | 'ANPEP_No17' | 'MLSSFLSEDVFK' | 'Aminopeptidase N' |
| 9381 | 'ANPEP_No18' | 'LPNTLKPDSYR' | 'Aminopeptidase N' |
| 9382 | 'ANPEP_No19' | 'KVVATTQMQAADAR' | 'Aminopeptidase N' |
| 9383 | 'ANPEP_No20' | 'ENSLLFDPLSSSSSNK' | 'Aminopeptidase N' |
| 9384 | 'ANPEP_No21' | 'ENKEVVLQWFTENSK' | 'Aminopeptidase N' |
| 9385 | 'ANPEP_No22' | 'VVATTQMQAADAR' | 'Aminopeptidase N' |
| 9386 | 'ANPEP_No23' | 'QWMENPNNNPIHPNLR' | 'Aminopeptidase N' |
| 9387 | 'ANPEP_No24' | 'DLTALSNMLPK' | 'Aminopeptidase N' |
| 9388 | 'ANPEP_No25' | 'ENSLLFDPLSSSSSNKER' | 'Aminopeptidase N' |
| 9389 | 'ANPEP_No26' | 'KVVATTQMQAADAR' | 'Aminopeptidase N' |
| 9390 | 'ANPEP_No27' | 'LMFDRSEVYGPMK' | 'Aminopeptidase N' |
| 9391 | 'ANPEP_No28' | 'EVVLQWFTENSK' | 'Aminopeptidase N' |
| 9392 | 'ANPEP_No29' | 'FSTEYELQQLEQFK' | 'Aminopeptidase N' |
| 9393 | 'ANPEP_No30' | 'DLTALSNMLPK' | 'Aminopeptidase N' |
| 9394 | 'ANPEP_No31' | 'QWMENPNNNPIHPNLR' | 'Aminopeptidase N' |
| 9395 | 'PVR_No1' | 'HGESGSMAVFHQTQGPSYSESK' | 'Poliovirus receptor' |
| 9396 | 'PVR_No2' | 'VQLTGEPVPMAR' | 'Poliovirus receptor' |
| 9397 | 'PVR_No3' | 'HGESGSMAVFHQTQGPSYSESK' | 'Poliovirus receptor' |
| 9398 | 'RAC2_No1' | 'HHCPSTPIILVGTK' | 'Ras-related C3 botulinum toxin substrate 2' |
| 9399 | 'RAC2_No2' | 'YLECSALTQR' | 'Ras-related C3 botulinum toxin substrate 2' |
| 9400 | 'RAC2_No3' | 'LAPITYPQGLALAK' | 'Ras-related C3 botulinum toxin substrate 2' |
| 9401 | 'RAC2_No4' | 'KLAPITYPQGLALAK' | 'Ras-related C3 botulinum toxin substrate 2' |
| 9402 | 'RAC2_No5' | 'AVLCPQPTR' | 'Ras-related C3 botulinum toxin substrate 2' |
| 9403 | 'RAC2_No6' | 'LDLRDDKDTIEK' | 'Ras-related C3 botulinum toxin substrate 2' |
| 9404 | 'CPN1_No1' | 'EALIQFLEQVHQGIK' | 'Carboxypeptidase N catalytic chain' |
| 9405 | 'CPN1_No2' | 'NFPDLNTYIYYNEK' | 'Carboxypeptidase N catalytic chain' |
| 9406 | 'CPN1_No3' | 'GMVLDENYNNLANAVISVSGINHDVTSGDHGDYFR' | 'Carboxypeptidase N catalytic chain' |
| 9407 | 'CPN1_No4' | 'YVGNMHGNEALGR' | 'Carboxypeptidase N catalytic chain' |
| 9408 | 'CPN1_No5' | 'YGGPNHHLPLPDNWK' | 'Carboxypeptidase N catalytic chain' |
| 9409 | 'CPN1_No6' | 'GMVLDENYNNLANAVISVSGINHDVTSGDHGDYFR' | 'Carboxypeptidase N catalytic chain' |
| 9410 | 'CPN1_No7' | 'IVQLIQDTR' | 'Carboxypeptidase N catalytic chain' |
| 9411 | 'CPN1_No8' | 'IHILPSMNPDGYEVAAAQGPNKPGYLVGR' | 'Carboxypeptidase N catalytic chain' |
| 9412 | 'CPN1_No9' | 'TASTPTPDDKLFQK' | 'Carboxypeptidase N catalytic chain' |
| 9413 | 'CPN1_No10' | 'VQNECPGITR' | 'Carboxypeptidase N catalytic chain' |
| 9414 | 'CPN1_No11' | 'IHILPSMNPDGYEVAAAQGPNKPGYLVGR' | 'Carboxypeptidase N catalytic chain' |
| 9415 | 'CPN1_No12' | 'HLYVLEFSDHPGIHEPLEPEVK' | 'Carboxypeptidase N catalytic chain' |
| 9416 | 'CPN1_No13' | 'YVGNMHGNEALGR' | 'Carboxypeptidase N catalytic chain' |
| 9417 | 'CPN1_No14' | 'SIPQVSPVRR' | 'Carboxypeptidase N catalytic chain' |

| | A | B | C |
|---|---|---|---|
| 1 | **Peptide Name** | **Peptide Sequence** | **Peptide Description** |
| 9418 | 'CPN1_No15' | 'RTASTPTPDDKLFQK' | 'Carboxypeptidase N catalytic chain' |
| 9419 | 'CPN1_No16' | 'TLYKVQNECPGITR' | 'Carboxypeptidase N catalytic chain' |
| 9420 | 'CPN1_No17' | 'RSIPQVSPVR' | 'Carboxypeptidase N catalytic chain' |
| 9421 | 'CPN1_No18' | 'NNANGVDLNR' | 'Carboxypeptidase N catalytic chain' |
| 9422 | 'CPN1_No19' | 'SQVEPETR' | 'Carboxypeptidase N catalytic chain' |
| 9423 | 'CPN1_No20' | 'TASTPTPDDKLFQKLAK' | 'Carboxypeptidase N catalytic chain' |
| 9424 | 'B4GALT1_No1' | 'YWLYYLHPVLQR' | 'Beta-1,4-galactosyltransferase 1' |
| 9425 | 'B4GALT1_No2' | 'QQLDYGIYVINQAGDTIFNR' | 'Beta-1,4-galactosyltransferase 1' |
| 9426 | 'B4GALT1_No3' | 'LPQLVGVSTPLQGGSNSAAAIGQSSGELR' | 'Beta-1,4-galactosyltransferase 1' |
| 9427 | 'NME1_No1' | 'YMHSGPVVAMVWEGLNVVK' | 'Nucleoside diphosphate kinase A' |
| 9428 | 'NME1_No2' | 'NIIHGSDSVESAEK' | 'Nucleoside diphosphate kinase A' |
| 9429 | 'IGLL1_No1' | 'ATPSVTLFPPSSEELQANK' | 'Immunoglobulin lambda-like polypeptide 1' |
| 9430 | 'IGLL1_No2' | 'YAASSYLSLTPEQWR' | 'Immunoglobulin lambda-like polypeptide 1' |
| 9431 | 'IGLL1_No3' | 'SYSCQVMHEGSTVEK' | 'Immunoglobulin lambda-like polypeptide 1' |
| 9432 | 'IGLL1_No4' | 'QSNNKYAASSYLSLTPEQWR' | 'Immunoglobulin lambda-like polypeptide 1' |
| 9433 | 'IGLL1_No5' | 'SYSCQVMHEGSTVEK' | 'Immunoglobulin lambda-like polypeptide 1' |
| 9434 | 'IGLL1_No6' | 'QSNNKYAASSYLSLTPEQWR' | 'Immunoglobulin lambda-like polypeptide 1' |
| 9435 | 'DSP_No1' | 'ALLQAILQTEDMLK' | 'Desmoplakin' |
| 9436 | 'DSP_No2' | 'GFFDPNTEENLTYLQLK' | 'Desmoplakin' |
| 9437 | 'DSP_No3' | 'LLEAQIATGGIIDPK' | 'Desmoplakin' |
| 9438 | 'DSP_No4' | 'TLELQGLINDLQR' | 'Desmoplakin' |
| 9439 | 'DSP_No5' | 'YGDGIQLTR' | 'Desmoplakin' |
| 9440 | 'DSP_No6' | 'AQQIHSQTSQQYPLYDLDLGK' | 'Desmoplakin' |
| 9441 | 'DSP_No7' | 'SVQNDSQAIAEVLNQLK' | 'Desmoplakin' |
| 9442 | 'DSP_No8' | 'NLPLADQGSSHHITVK' | 'Desmoplakin' |
| 9443 | 'DSP_No9' | 'GYFNEELSEILSDPSDDTK' | 'Desmoplakin' |
| 9444 | 'DSP_No10' | 'LASLEELKR' | 'Desmoplakin' |
| 9445 | 'DSP_No11' | 'SVEEVASEIQPFLR' | 'Desmoplakin' |
| 9446 | 'DSP_No12' | 'DLKDEIVR' | 'Desmoplakin' |
| 9447 | 'DSP_No13' | 'TIADLELHYQEFIR' | 'Desmoplakin' |
| 9448 | 'DSP_No14' | 'LNDSILQATEQR' | 'Desmoplakin' |
| 9449 | 'DSP_No15' | 'ESHRLPVDIAYKR' | 'Desmoplakin' |
| 9450 | 'DSP_No16' | 'YIELLTR' | 'Desmoplakin' |
| 9451 | 'DSP_No17' | 'FLSEMLK' | 'Desmoplakin' |
| 9452 | 'DSP_No18' | 'LLEAQIASGGVVDPVNSVFLPK' | 'Desmoplakin' |
| 9453 | 'DSP_No19' | 'LENINGVTDGYLNSLCTVR' | 'Desmoplakin' |
| 9454 | 'DSP_No20' | 'ITNLTQQLEQASIVK' | 'Desmoplakin' |
| 9455 | 'DSP_No21' | 'TMIQSPSGVILQEAADVHAR' | 'Desmoplakin' |
| 9456 | 'DSP_No22' | 'LQRLEDELNR' | 'Desmoplakin' |
| 9457 | 'DSP_No23' | 'AVTGYNDPETGNIISLFQAMNK' | 'Desmoplakin' |
| 9458 | 'DSP_No24' | 'LPVDIAYK' | 'Desmoplakin' |
| 9459 | 'DSP_No25' | 'TMIQSPSGVILQEAADVHAR' | 'Desmoplakin' |
| 9460 | 'DSP_No26' | 'ESHRLPVDIAYK' | 'Desmoplakin' |
| 9461 | 'DSP_No27' | 'AVTGYNDPETGNIISLFQAMNK' | 'Desmoplakin' |

| | A | B | C |
|---|---|---|---|
| 1 | **Peptide Name** | **Peptide Sequence** | **Peptide Description** |
| 9462 | 'DSP_No28' | 'GGGGYTCQSGSGWDEFTK' | 'Desmoplakin' |
| 9463 | 'DSP_No29' | 'YCYLQNEVFGLFQK' | 'Desmoplakin' |
| 9464 | 'DSP_No30' | 'RLSSEVEALRR' | 'Desmoplakin' |
| 9465 | 'DSP_No31' | 'LKNTLTQTTENLR' | 'Desmoplakin' |
| 9466 | 'DSP_No32' | 'GVITDQNSDGYCQTGTMSR' | 'Desmoplakin' |
| 9467 | 'DSP_No33' | 'LTYEIEDEK' | 'Desmoplakin' |
| 9468 | 'DSP_No34' | 'AEMDMVAWGVDLASVEQHINSHR' | 'Desmoplakin' |
| 9469 | 'DSP_No35' | 'QLQNIIQATSR' | 'Desmoplakin' |
| 9470 | 'DSP_No36' | 'NQFETEINITK' | 'Desmoplakin' |
| 9471 | 'DSP_No37' | 'LTYEIEDEKR' | 'Desmoplakin' |
| 9472 | 'TIMP2_No1' | 'AVSEKEVDSGNDIYGNPIKR' | 'Metalloproteinase inhibitor 2' |
| 9473 | 'TIMP2_No2' | 'EVDSGNDIYGNPIKR' | 'Metalloproteinase inhibitor 2' |
| 9474 | 'TIMP2_No3' | 'MHITLCDFIVPWDTLSTTQKK' | 'Metalloproteinase inhibitor 2' |
| 9475 | 'TIMP2_No4' | 'DIEFIYTAPSSAVCGVSLDVGGK' | 'Metalloproteinase inhibitor 2' |
| 9476 | 'TIMP2_No5' | 'GPEKDIEFIYTAPSSAVCGVSLDVGGKK' | 'Metalloproteinase inhibitor 2' |
| 9477 | 'CD44_No1' | 'YGFIEGHVVIPR' | 'CD44 antigen' |
| 9478 | 'CD44_No2' | 'FAGVFHVEK' | 'CD44 antigen' |
| 9479 | 'CD44_No3' | 'ALSIGFETCR' | 'CD44 antigen' |
| 9480 | 'SELP_No1' | 'LEGPNNVECTTSGR' | 'P-selectin' |
| 9481 | 'SELP_No2' | 'YTDLVAIQNK' | 'P-selectin' |
| 9482 | 'SELP_No3' | 'HALCYTASCQDMSCSK' | 'P-selectin' |
| 9483 | 'SELP_No4' | 'YTDLVAIQNKNEIDYLNK' | 'P-selectin' |
| 9484 | 'SELP_No5' | 'STCQFICDEGYSLSGPER' | 'P-selectin' |
| 9485 | 'SELP_No6' | 'ALTNEAENWADNEPNNKR' | 'P-selectin' |
| 9486 | 'SELP_No7' | 'AVQCQHLEAPSEGTMDCVHPLTAFAYGSSCK' | 'P-selectin' |
| 9487 | 'SELP_No8' | 'WTDSPPMCEAIK' | 'P-selectin' |
| 9488 | 'SELP_No9' | 'HALCYTASCQDMSCSK' | 'P-selectin' |
| 9489 | 'SELP_No10' | 'SPSAPGKWNDEHCLK' | 'P-selectin' |
| 9490 | 'SELP_No11' | 'KALTNEAENWADNEPNNKR' | 'P-selectin' |
| 9491 | 'ANK1_No1' | 'SGLTPLHLVAQEGHVPVADVLIK' | 'Ankyrin-1' |
| 9492 | 'ANK1_No2' | 'LGYISVTDVLK' | 'Ankyrin-1' |
| 9493 | 'ANK1_No3' | 'TGFTPLHIAAHYENLNVAQLLLNR' | 'Ankyrin-1' |
| 9494 | 'ANK1_No4' | 'LLCSVIGGTDQAQWEDITGTTK' | 'Ankyrin-1' |
| 9495 | 'ANK1_No5' | 'ITHSPTVSQVTER' | 'Ankyrin-1' |
| 9496 | 'ANK1_No6' | 'TGASIDAVTESGLTPLHVASFMGHLPIVK' | 'Ankyrin-1' |
| 9497 | 'ANK1_No7' | 'LLGNQATFSPIVTVEPR' | 'Ankyrin-1' |
| 9498 | 'ANK1_No8' | 'GFTPLYMAAQENHLEVVK' | 'Ankyrin-1' |
| 9499 | 'ANK1_No9' | 'TPTPLALR' | 'Ankyrin-1' |
| 9500 | 'ANK1_No10' | 'NGASPNEVSSDGTTPLAIAK' | 'Ankyrin-1' |
| 9501 | 'ANK1_No11' | 'EGHVETVLALLEK' | 'Ankyrin-1' |
| 9502 | 'ANK1_No12' | 'TAEAVNFATLLYK' | 'Ankyrin-1' |
| 9503 | 'ANK1_No13' | 'TKDELTPLHCAAR' | 'Ankyrin-1' |
| 9504 | 'ANK1_No14' | 'LVPLVQATFPENAVTK' | 'Ankyrin-1' |
| 9505 | 'ANK1_No15' | 'LALQAQPVPDELVTK' | 'Ankyrin-1' |

| | A | B | C |
|---|---|---|---|
| 1 | **Peptide Name** | **Peptide Sequence** | **Peptide Description** |
| 9506 | 'ANK1_No16' | 'TLEQHENFVEVAR' | 'Ankyrin-1' |
| 9507 | 'ANK1_No17' | 'YSILSESTPGSLSGTEQAEMK' | 'Ankyrin-1' |
| 9508 | 'ANK1_No18' | 'LGYSPLHQAAQQGHTDIVTLLLK' | 'Ankyrin-1' |
| 9509 | 'ANK1_No19' | 'LCQDYDTIGPEGGSLK' | 'Ankyrin-1' |
| 9510 | 'ANK1_No20' | 'RQDDATGAGQDSENEVSLVSGHQR' | 'Ankyrin-1' |
| 9511 | 'PNLIP' | 'ITGLDPAEPCFQGTPELVR' | 'Pancreatic triacylglycerol lipase' |
| 9512 | 'PECAM1_No1' | 'SELVTVTESFSTPK' | 'Platelet endothelial cell adhesion molecule' |
| 9513 | 'PECAM1_No2' | 'CESISGTLPISYQLLK' | 'Platelet endothelial cell adhesion molecule' |
| 9514 | 'PECAM1_No3' | 'STESYFIPEVR' | 'Platelet endothelial cell adhesion molecule' |
| 9515 | 'PECAM1_No4' | 'VIAPVDEVQISILSSK' | 'Platelet endothelial cell adhesion molecule' |
| 9516 | 'PECAM1_No5' | 'IISGIHMQTSESTK' | 'Platelet endothelial cell adhesion molecule' |
| 9517 | 'PECAM1_No6' | 'DNPTEDVEYQCVADNCHSHAK' | 'Platelet endothelial cell adhesion molecule' |
| 9518 | 'H1-5' | 'SETAPAETATPAPVEK' | 'Histone H1.5' |
| 9519 | 'EPB42_No1' | 'VLPTPQTQATQEGALLNK' | 'Erythrocyte membrane protein band 4.2' |
| 9520 | 'EPB42_No2' | 'FQFTPTHVGLQR' | 'Erythrocyte membrane protein band 4.2' |
| 9521 | 'EPB42_No3' | 'GDAQISVTLVNHSEQEK' | 'Erythrocyte membrane protein band 4.2' |
| 9522 | 'ATP2A2_No1' | 'VGEATETALTCLVEK' | 'Sarcoplasmic/endoplasmic reticulum calcium ATPase 2' |
| 9523 | 'ATP2A2_No2' | 'TGTLTTNQMSVCR' | 'Sarcoplasmic/endoplasmic reticulum calcium ATPase 2' |
| 9524 | 'ATP2A2_No3' | 'NYLEPGKECVQPATK' | 'Sarcoplasmic/endoplasmic reticulum calcium ATPase 2' |
| 9525 | 'ATP2A2_No4' | 'SMSVYCTPNKPSR' | 'Sarcoplasmic/endoplasmic reticulum calcium ATPase 2' |
| 9526 | 'ATP2A2_No5' | 'VDQSILTGESVSVIK' | 'Sarcoplasmic/endoplasmic reticulum calcium ATPase 2' |
| 9527 | 'ATP2A2_No6' | 'IGIFGQDEDVTSK' | 'Sarcoplasmic/endoplasmic reticulum calcium ATPase 2' |
| 9528 | 'ATP2A2_No7' | 'TGTLTTNQMSVCR' | 'Sarcoplasmic/endoplasmic reticulum calcium ATPase 2' |
| 9529 | 'ATP2A2_No8' | 'RIGIFGQDEDVTSK' | 'Sarcoplasmic/endoplasmic reticulum calcium ATPase 2' |
| 9530 | 'CD36_No1' | 'RLQVNLLVKPSEK' | 'Platelet glycoprotein 4' |
| 9531 | 'CD36_No2' | 'LQVNLLVKPSEK' | 'Platelet glycoprotein 4' |
| 9532 | 'CD36_No3' | 'QVVLEEGTIAFK' | 'Platelet glycoprotein 4' |
| 9533 | 'CD36_No4' | 'VAIIDTYK' | 'Platelet glycoprotein 4' |
| 9534 | 'CD36_No5' | 'SQVLQFFSSDICR' | 'Platelet glycoprotein 4' |
| 9535 | 'CD36_No6' | 'KQVVLEEGTIAFK' | 'Platelet glycoprotein 4' |
| 9536 | 'CD36_No7' | 'AFASPVENPDNYCFCTEK' | 'Platelet glycoprotein 4' |
| 9537 | 'FAH_No1' | 'VFLQNLLSVSQAR' | 'Fumarylacetoacetase' |
| 9538 | 'FAH_No2' | 'DKENALMPNWLHLPVGYHGR' | 'Fumarylacetoacetase' |
| 9539 | 'FAH_No3' | 'WEYVPLGPFLGK' | 'Fumarylacetoacetase' |
| 9540 | 'FAH_No4' | 'HLFTGPVLSK' | 'Fumarylacetoacetase' |
| 9541 | 'FAH_No5' | 'HQDVFNQPTLNSFMGLGQAAWK' | 'Fumarylacetoacetase' |
| 9542 | 'FAH_No6' | 'DIQKWEYVPLGPFLGK' | 'Fumarylacetoacetase' |
| 9543 | 'FAH_No7' | 'HQDVFNQPTLNSFMGLGQAAWK' | 'Fumarylacetoacetase' |
| 9544 | 'FAH_No8' | 'FLLDGDEVIITGYCQGDGYR' | 'Fumarylacetoacetase' |
| 9545 | 'GOT1_No1' | 'APPSVFAEVPQAQPVLVFK' | 'Aspartate aminotransferase, cytoplasmic' |
| 9546 | 'GOT1_No2' | 'NTPVYVSSPTWENHNAVFSAAGFK' | 'Aspartate aminotransferase, cytoplasmic' |
| 9547 | 'GOT1_No3' | 'NLDYVATSIHEAVTK' | 'Aspartate aminotransferase, cytoplasmic' |
| 9548 | 'GOT1_No4' | 'IANDNSLNHEYLPILGLAEFR' | 'Aspartate aminotransferase, cytoplasmic' |
| 9549 | 'GOT1_No5' | 'IGADFLAR' | 'Aspartate aminotransferase, cytoplasmic' |

| | A | B | C |
|---|---|---|---|
| 1 | **Peptide Name** | **Peptide Sequence** | **Peptide Description** |
| 9550 | 'GOT1_No6' | 'IVASTLSNPELFEEWTGNVK' | 'Aspartate aminotransferase, cytoplasmic' |
| 9551 | 'BPI' | 'GLDYASQQGTAALQK' | 'Bactericidal permeability-increasing protein' |
| 9552 | 'ITGA2_No1' | 'VDISLENPGTSPALEAYSETAK' | 'Integrin alpha-2' |
| 9553 | 'ITGA2_No2' | 'FGIAVLGYLNR' | 'Integrin alpha-2' |
| 9554 | 'ITGA2_No3' | 'VYLFTIK' | 'Integrin alpha-2' |
| 9555 | 'ITGA2_No4' | 'FVQGLDIGPTK' | 'Integrin alpha-2' |
| 9556 | 'ITGA2_No5' | 'SVACDVGYPALKR' | 'Integrin alpha-2' |
| 9557 | 'ITGA2_No6' | 'DCGEDGLCISDLVLDVR' | 'Integrin alpha-2' |
| 9558 | 'ITGA2_No7' | 'IPLLYDAEIHLTR' | 'Integrin alpha-2' |
| 9559 | 'ITGA2_No8' | 'STNINFYEISSDGNVPSIVHSFEDVGPK' | 'Integrin alpha-2' |
| 9560 | 'ITGA2_No9' | 'AGDISCNADINPLK' | 'Integrin alpha-2' |
| 9561 | 'ITGA2_No10' | 'NMVVNQAQSCPEHIIYIQEPSDVVNSLDLR' | 'Integrin alpha-2' |
| 9562 | 'ITGA2_No11' | 'VMVVVTDGESHDGSMLK' | 'Integrin alpha-2' |
| 9563 | 'ITGA2_No12' | 'VTTGSVPVSMATVIIHIPQYTK' | 'Integrin alpha-2' |
| 9564 | 'ITGA2_No13' | 'IFSGPSSEQFGYAVQQFINPK' | 'Integrin alpha-2' |
| 9565 | 'CTRB1_No1' | 'FSILTVNNDITLLK' | 'Chymotrypsinogen B' |
| 9566 | 'CTRB1_No2' | 'IVNGEDAVPGSWPWQVSLQDK' | 'Chymotrypsinogen B' |
| 9567 | 'PRKACA_No1' | 'FPSHFSSDLKDLLR' | 'cAMP-dependent protein kinase catalytic subunit alpha' |
| 9568 | 'PRKACA_No2' | 'ILQAVNFPFLVK' | 'cAMP-dependent protein kinase catalytic subunit alpha' |
| 9569 | 'ENG_No1' | 'GPITSAAELNDPQSILLR' | 'Endoglin' |
| 9570 | 'ENG_No2' | 'GFKLPDTPQGLLGEAR' | 'Endoglin' |
| 9571 | 'ENG_No3' | 'TQILEWAAER' | 'Endoglin' |
| 9572 | 'PFKL_No1' | 'IMEVIDAITTTAQSHQR' | 'ATP-dependent 6-phosphofructokinase, liver type' |
| 9573 | 'PFKL_No2' | 'ISMAAYVSGELEHVTR' | 'ATP-dependent 6-phosphofructokinase, liver type' |
| 9574 | 'PFKL_No3' | 'EQWWLSLR' | 'ATP-dependent 6-phosphofructokinase, liver type' |
| 9575 | 'PFKL_No4' | 'AIGVLTSGGDAQGMNAAVR' | 'ATP-dependent 6-phosphofructokinase, liver type' |
| 9576 | 'PFKL_No5' | 'AVAFSPVTELKK' | 'ATP-dependent 6-phosphofructokinase, liver type' |
| 9577 | 'PFKL_No6' | 'GQVQEVGWHDVAGWLGR' | 'ATP-dependent 6-phosphofructokinase, liver type' |
| 9578 | 'PFKL_No7' | 'VFANAPDSACVIGLK' | 'ATP-dependent 6-phosphofructokinase, liver type' |
| 9579 | 'PFKL_No8' | 'GGSFENNWNIYK' | 'ATP-dependent 6-phosphofructokinase, liver type' |
| 9580 | 'LGALS3_No1' | 'MLITILGTVKPNANR' | 'Galectin-3' |
| 9581 | 'LGALS3_No2' | 'GNDVAFHFNPR' | 'Galectin-3' |
| 9582 | 'LGALS3_No3' | 'IQVLVEPDHFK' | 'Galectin-3' |
| 9583 | 'LGALS3_No4' | 'VAVNDAHLLQYNHR' | 'Galectin-3' |
| 9584 | 'LGALS3_No5' | 'MLITILGTVKPNANR' | 'Galectin-3' |
| 9585 | 'LGALS3_No6' | 'QSVFPFESGKPFK' | 'Galectin-3' |
| 9586 | 'IGFBP3_No1' | 'ALAQCAPPPAVCAELVR' | 'Insulin-like growth factor-binding protein 3' |
| 9587 | 'IGFBP3_No2' | 'GFCWCVDK' | 'Insulin-like growth factor-binding protein 3' |
| 9588 | 'IGFBP3_No3' | 'YGQPLPGYTTK' | 'Insulin-like growth factor-binding protein 3' |
| 9589 | 'IGFBP3_No4' | 'GFCWCVDKYGQPLPGYTTK' | 'Insulin-like growth factor-binding protein 3' |
| 9590 | 'IGFBP3_No5' | 'ALAQCAPPPAVCAELVREPGCGCCLTCALSEGQPCGIYTER' | 'Insulin-like growth factor-binding protein 3' |
| 9591 | 'IGFBP3_No6' | 'EMEDTLNHLK' | 'Insulin-like growth factor-binding protein 3' |
| 9592 | 'IGFBP3_No7' | 'YKVDYESQSTDTQNFSSESKR' | 'Insulin-like growth factor-binding protein 3' |
| 9593 | 'IGFBP3_No8' | 'REMEDTLNHLK' | 'Insulin-like growth factor-binding protein 3' |

| | A | B | C |
|---|---|---|---|
| 1 | **Peptide Name** | **Peptide Sequence** | **Peptide Description** |
| 9594 | 'IGFBP3_No9' | 'CQPSPDEARPLQALLDGR' | 'Insulin-like growth factor-binding protein 3' |
| 9595 | 'IGFBP3_No10' | 'EMEDTLNHLK' | 'Insulin-like growth factor-binding protein 3' |
| 9596 | 'IGFBP3_No11' | 'RGFCWCVDK' | 'Insulin-like growth factor-binding protein 3' |
| 9597 | 'IGFBP3_No12' | 'VDYESQSTDTQNFSSESK' | 'Insulin-like growth factor-binding protein 3' |
| 9598 | 'IGFBP3_No13' | 'FLNVLSPR' | 'Insulin-like growth factor-binding protein 3' |
| 9599 | 'PSMC3' | 'EKAPSIIFIDELDAIGTK' | '26S proteasome regulatory subunit 6A' |
| 9600 | 'TCP1_No1' | 'EGPLSVFGDR' | 'T-complex protein 1 subunit alpha' |
| 9601 | 'TCP1_No2' | 'SLLVIPNTLAVNAAQDSTDLVAK' | 'T-complex protein 1 subunit alpha' |
| 9602 | 'TCP1_No3' | 'YINENLIVNTDELGR' | 'T-complex protein 1 subunit alpha' |
| 9603 | 'TCP1_No4' | 'ILATGANVILTTGGIDDMCLK' | 'T-complex protein 1 subunit alpha' |
| 9604 | 'TCP1_No5' | 'DDKHGSYEDAVHSGALND' | 'T-complex protein 1 subunit alpha' |
| 9605 | 'TCP1_No6' | 'LGVQVVITDPEKLDQIR' | 'T-complex protein 1 subunit alpha' |
| 9606 | 'IGFBP2_No1' | 'LIQGAPTIR' | 'Insulin-like growth factor-binding protein 2' |
| 9607 | 'IGFBP2_No2' | 'LEGEACGVYTPR' | 'Insulin-like growth factor-binding protein 2' |
| 9608 | 'IGFBP2_No3' | 'GECWCVNPNTGK' | 'Insulin-like growth factor-binding protein 2' |
| 9609 | 'IGFBP2_No4' | 'MPCAELVR' | 'Insulin-like growth factor-binding protein 2' |
| 9610 | 'IGFBP2_No5' | 'LAACGPPPVAPPAAVAAVAGGAR' | 'Insulin-like growth factor-binding protein 2' |
| 9611 | 'IGFBP2_No6' | 'MPCAELVREPGCGCCSVCAR' | 'Insulin-like growth factor-binding protein 2' |
| 9612 | 'IGFBP2_No7' | 'TPCQQELDQVLER' | 'Insulin-like growth factor-binding protein 2' |
| 9613 | 'IGFBP2_No8' | 'EPGCGCCSVCAR' | 'Insulin-like growth factor-binding protein 2' |
| 9614 | 'IGFBP2_No9' | 'GDPECHLFYNEQQEAR' | 'Insulin-like growth factor-binding protein 2' |
| 9615 | 'IGFBP2_No10' | 'GPLEHLYSLHIPNCDK' | 'Insulin-like growth factor-binding protein 2' |
| 9616 | 'IGFBP2_No11' | 'CYPHPGSELPLQALVMGEGTCEK' | 'Insulin-like growth factor-binding protein 2' |
| 9617 | 'VCL_No1' | 'AQQVSQGLDVLTAK' | 'Vinculin' |
| 9618 | 'VCL_No2' | 'DPSASPGDAGEQAIR' | 'Vinculin' |
| 9619 | 'VCL_No3' | 'ELLPVLISAMK' | 'Vinculin' |
| 9620 | 'VCL_No4' | 'IPTISTQLK' | 'Vinculin' |
| 9621 | 'VCL_No5' | 'MALLMAEMSR' | 'Vinculin' |
| 9622 | 'VCL_No6' | 'AGEVINQPMMMAAR' | 'Vinculin' |
| 9623 | 'VCL_No7' | 'AVAGNISDPGLQK' | 'Vinculin' |
| 9624 | 'VCL_No8' | 'GILSGTSDLLLTFDEAEVRK' | 'Vinculin' |
| 9625 | 'VCL_No9' | 'VAMANIQPQMLVAGATSIAR' | 'Vinculin' |
| 9626 | 'VCL_No10' | 'ALASQLQDSLK' | 'Vinculin' |
| 9627 | 'VCL_No11' | 'AIPDLTAPVAAVQAAVSNLVR' | 'Vinculin' |
| 9628 | 'VCL_No12' | 'VLQLTSWDEDAWASK' | 'Vinculin' |
| 9629 | 'VCL_No13' | 'ETVQTTEDQILKR' | 'Vinculin' |
| 9630 | 'VCL_No14' | 'GWLRDPSASPGDAGEQAIR' | 'Vinculin' |
| 9631 | 'VCL_No15' | 'VAMANIQPQMLVAGATSIAR' | 'Vinculin' |
| 9632 | 'VCL_No16' | 'VGKETVQTTEDQILKR' | 'Vinculin' |
| 9633 | 'VCL_No17' | 'MIDERQQELTHQEHR' | 'Vinculin' |
| 9634 | 'VCL_No18' | 'ELLPVLISAMK' | 'Vinculin' |
| 9635 | 'VCL_No19' | 'SLLDASEEAIKK' | 'Vinculin' |
| 9636 | 'VCL_No20' | 'MLGQMTDQVADLR' | 'Vinculin' |
| 9637 | 'VCL_No21' | 'VDQLTAQLADLAAR' | 'Vinculin' |

| | A | B | C |
|---|---|---|---|
| 1 | **Peptide Name** | **Peptide Sequence** | **Peptide Description** |
| 9638 | 'VCL_No22' | 'KIDAAQNWLADPNGGPEGEEQIR' | 'Vinculin' |
| 9639 | 'VCL_No23' | 'MQEAMTQEVSDVFSDTTTPIK' | 'Vinculin' |
| 9640 | 'VCL_No24' | 'LVQAAQMLQSDPYSVPAR' | 'Vinculin' |
| 9641 | 'VCL_No25' | 'QVATALQNLQTK' | 'Vinculin' |
| 9642 | 'VCL_No26' | 'AGEVINQPMMMAAR' | 'Vinculin' |
| 9643 | 'VCL_No27' | 'WIDNPTVDDR' | 'Vinculin' |
| 9644 | 'VCL_No28' | 'MLGQMTDQVADLR' | 'Vinculin' |
| 9645 | 'VCL_No29' | 'EVENSEDPKFR' | 'Vinculin' |
| 9646 | 'VCL_No30' | 'VMLVNSMNTVK' | 'Vinculin' |
| 9647 | 'VCL_No31' | 'MLGQMTDQVADLR' | 'Vinculin' |
| 9648 | 'VCL_No32' | 'LANVMMGPYR' | 'Vinculin' |
| 9649 | 'VCL_No33' | 'MSAEINEIIR' | 'Vinculin' |
| 9650 | 'VCL_No34' | 'LVQAAQMLQSDPYSVPARDYLIDGSR' | 'Vinculin' |
| 9651 | 'VCL_No35' | 'TNISDEESEQATEMLVHNAQNLMQSVK' | 'Vinculin' |
| 9652 | 'VCL_No36' | 'LVQAAQMLQSDPYSVPAR' | 'Vinculin' |
| 9653 | 'VCL_No37' | 'AGEVINQPMMMAAR' | 'Vinculin' |
| 9654 | 'VCL_No38' | 'CDRVDQLTAQLADLAARGEGESPQAR' | 'Vinculin' |
| 9655 | 'VCL_No39' | 'AGEVINQPMMMAAR' | 'Vinculin' |
| 9656 | 'VCL_No40' | 'LANVMMGPYRQDLLAK' | 'Vinculin' |
| 9657 | 'VCL_No41' | 'ALASQLQDSLKDLK' | 'Vinculin' |
| 9658 | 'VCL_No42' | 'ETVQTTEDQILK' | 'Vinculin' |
| 9659 | 'VCL_No43' | 'QILDEAGKVGELCAGK' | 'Vinculin' |
| 9660 | 'VCL_No44' | 'TNISDEESEQATEMLVHNAQNLMQSVK' | 'Vinculin' |
| 9661 | 'VCL_No45' | 'MLGQMTDQVADLR' | 'Vinculin' |
| 9662 | 'VCL_No46' | 'NPGNQAAYEHFETMK' | 'Vinculin' |
| 9663 | 'VCL_No47' | 'MIDERQQELTHQEHR' | 'Vinculin' |
| 9664 | 'VCL_No48' | 'MQEAMTQEVSDVFSDTTTPIK' | 'Vinculin' |
| 9665 | 'VCL_No49' | 'WIDNPTVDDRGVGQAAIR' | 'Vinculin' |
| 9666 | 'VCL_No50' | 'AAVHLEGKIEQAQR' | 'Vinculin' |
| 9667 | 'VCL_No51' | 'QILDEAGKVGELCAGK' | 'Vinculin' |
| 9668 | 'VCL_No52' | 'LLAVAATAPPDAPNREEVFDER' | 'Vinculin' |
| 9669 | 'VCL_No53' | 'CDRVDQLTAQLADLAAR' | 'Vinculin' |
| 9670 | 'VCL_No54' | 'LVQAAQMLQSDPYSVPARDYLIDGSR' | 'Vinculin' |
| 9671 | 'VCL_No55' | 'IDAAQNWLADPNGGPEGEEQIR' | 'Vinculin' |
| 9672 | 'VCL_No56' | 'EVENSEDPKFREAVK' | 'Vinculin' |
| 9673 | 'VCL_No57' | 'VMLVNSMNTVK' | 'Vinculin' |
| 9674 | 'VCL_No58' | 'MSAEINEIIR' | 'Vinculin' |
| 9675 | 'VCL_No59' | 'NPGNQAAYEHFETMK' | 'Vinculin' |
| 9676 | 'VCL_No60' | 'REVENSEDPKFR' | 'Vinculin' |
| 9677 | 'VCL_No61' | 'TNISDEESEQATEMLVHNAQNLMQSVK' | 'Vinculin' |
| 9678 | 'VCL_No62' | 'VGKETVQTTEDQILK' | 'Vinculin' |
| 9679 | 'VCL_No63' | 'TISPMVMDAK' | 'Vinculin' |
| 9680 | 'VCL_No64' | 'MTGLVDEAIDTK' | 'Vinculin' |
| 9681 | 'LBP_No1' | 'SFRPFVPR' | 'Lipopolysaccharide-binding protein' |

| | A | B | C |
|---|---|---|---|
| 1 | **Peptide Name** | **Peptide Sequence** | **Peptide Description** |
| 9682 | 'LBP_No2' | 'ITLPDFTGDLR' | 'Lipopolysaccharide-binding protein' |
| 9683 | 'LBP_No3' | 'FNDKLAEGFPLPLLK' | 'Lipopolysaccharide-binding protein' |
| 9684 | 'LBP_No4' | 'VQLYDLGLQIHK' | 'Lipopolysaccharide-binding protein' |
| 9685 | 'LBP_No5' | 'SPVTLLAAVMSLPEEHNK' | 'Lipopolysaccharide-binding protein' |
| 9686 | 'LBP_No6' | 'GLQYAAQEGLLALQSELLR' | 'Lipopolysaccharide-binding protein' |
| 9687 | 'LBP_No7' | 'LAEGFPLPLLKR' | 'Lipopolysaccharide-binding protein' |
| 9688 | 'LBP_No8' | 'FNDKLAEGFPLPLLKR' | 'Lipopolysaccharide-binding protein' |
| 9689 | 'LBP_No9' | 'ATAQMLEVMFK' | 'Lipopolysaccharide-binding protein' |
| 9690 | 'LBP_No10' | 'RVQLYDLGLQIHK' | 'Lipopolysaccharide-binding protein' |
| 9691 | 'LBP_No11' | 'ITLPDFTGDLRIPHVGR' | 'Lipopolysaccharide-binding protein' |
| 9692 | 'LBP_No12' | 'ITGFLKPGK' | 'Lipopolysaccharide-binding protein' |
| 9693 | 'LBP_No13' | 'SPVTLLAAVMSLPEEHNK' | 'Lipopolysaccharide-binding protein' |
| 9694 | 'LBP_No14' | 'LAEGFPLPLLK' | 'Lipopolysaccharide-binding protein' |
| 9695 | 'LBP_No15' | 'ICEMIQK' | 'Lipopolysaccharide-binding protein' |
| 9696 | 'LBP_No16' | 'ATAQMLEVMFKGEIFHR' | 'Lipopolysaccharide-binding protein' |
| 9697 | 'LBP_No17' | 'DFLFLGANVQYMR' | 'Lipopolysaccharide-binding protein' |
| 9698 | 'LBP_No18' | 'ICEMIQK' | 'Lipopolysaccharide-binding protein' |
| 9699 | 'LBP_No19' | 'LQGSFDVSVK' | 'Lipopolysaccharide-binding protein' |
| 9700 | 'LBP_No20' | 'DFLFLGANVQYMR' | 'Lipopolysaccharide-binding protein' |
| 9701 | 'LBP_No21' | 'ITDKGLQYAAQEGLLALQSELLR' | 'Lipopolysaccharide-binding protein' |
| 9702 | 'LBP_No22' | 'ATAQMLEVMFK' | 'Lipopolysaccharide-binding protein' |
| 9703 | 'PGAM1_No1' | 'ALPFWNEEIVPQIK' | 'Phosphoglycerate mutase 1' |
| 9704 | 'PGAM1_No2' | 'FSGWYDADLSPAGHEEAKR' | 'Phosphoglycerate mutase 1' |
| 9705 | 'KRT4_No1' | 'NLDLDSIIAEVR' | 'Keratin, type II cytoskeletal 4' |
| 9706 | 'KRT4_No2' | 'FASFIDKVQFLEQQNK' | 'Keratin, type II cytoskeletal 4' |
| 9707 | 'KRT4_No3' | 'WNLLQQQTTTTSSK' | 'Keratin, type II cytoskeletal 4' |
| 9708 | 'PAM_No1' | 'IPVDEEAFVIDFKPR' | 'Peptidyl-glycine alpha-amidating monooxygenase' |
| 9709 | 'PAM_No2' | 'REEEEVLDQGDFYSLLSK' | 'Peptidyl-glycine alpha-amidating monooxygenase' |
| 9710 | 'PAM_No3' | 'DNNKDCSGVSLHLTR' | 'Peptidyl-glycine alpha-amidating monooxygenase' |
| 9711 | 'PAM_No4' | 'LLGEREDVVHVHK' | 'Peptidyl-glycine alpha-amidating monooxygenase' |
| 9712 | 'PAM_No5' | 'DCSGVSLHLTR' | 'Peptidyl-glycine alpha-amidating monooxygenase' |
| 9713 | 'CDH2_No1' | 'FLEAGIYEVPIIITDSGNPPK' | 'Cadherin-2' |
| 9714 | 'CDH2_No2' | 'VKVCQCDSNGDCTDVDR' | 'Cadherin-2' |
| 9715 | 'CDH2_No3' | 'VCQCDSNGDCTDVDR' | 'Cadherin-2' |
| 9716 | 'CDH2_No4' | 'DWVIPPINLPENSR' | 'Cadherin-2' |
| 9717 | 'CDH2_No5' | 'QEEGLHAGTMLTTFTAQDPDR' | 'Cadherin-2' |
| 9718 | 'CDH2_No6' | 'IDPVNGQITTIAVLDR' | 'Cadherin-2' |
| 9719 | 'CDH2_No7' | 'LSDPANWLK' | 'Cadherin-2' |
| 9720 | 'VCAM1_No1' | 'NTVISVNPSTK' | 'Vascular cell adhesion protein 1' |
| 9721 | 'VCAM1_No2' | 'LHIDEMDSVPTVR' | 'Vascular cell adhesion protein 1' |
| 9722 | 'VCAM1_No3' | 'MEDSGVYLCEGINQAGR' | 'Vascular cell adhesion protein 1' |
| 9723 | 'VCAM1_No4' | 'SLEVTFTPVIEDIGK' | 'Vascular cell adhesion protein 1' |
| 9724 | 'VCAM1_No5' | 'EVELIVQEKPFTVEISPGPR' | 'Vascular cell adhesion protein 1' |
| 9725 | 'VCAM1_No6' | 'SQEFLEDADRK' | 'Vascular cell adhesion protein 1' |

| | A | B | C |
|---|---|---|---|
| 1 | **Peptide Name** | **Peptide Sequence** | **Peptide Description** |
| 9726 | 'VCAM1_No7' | 'MEDSGIYVCEGVNLIGK' | 'Vascular cell adhesion protein 1' |
| 9727 | 'VCAM1_No8' | 'QSTQTLYVNVAPR' | 'Vascular cell adhesion protein 1' |
| 9728 | 'VCAM1_No9' | 'MEDSGVYLCEGINQAGR' | 'Vascular cell adhesion protein 1' |
| 9729 | 'VCAM1_No10' | 'LHIDDMEFEPK' | 'Vascular cell adhesion protein 1' |
| 9730 | 'VCAM1_No11' | 'SLEMTFIPTIEDTGK' | 'Vascular cell adhesion protein 1' |
| 9731 | 'VCAM1_No12' | 'MEDSGIYVCEGVNLIGK' | 'Vascular cell adhesion protein 1' |
| 9732 | 'VCAM1_No13' | 'QAVKELQVYISPK' | 'Vascular cell adhesion protein 1' |
| 9733 | 'VCAM1_No14' | 'VPSVYPLDRLEIELLK' | 'Vascular cell adhesion protein 1' |
| 9734 | 'VCAM1_No15' | 'LHIDEMDSVPTVR' | 'Vascular cell adhesion protein 1' |
| 9735 | 'VCAM1_No16' | 'VTNEGTTSTLTMNPVSFGNEHSYLCTATCESR' | 'Vascular cell adhesion protein 1' |
| 9736 | 'VCAM1_No17' | 'LEIDLLK' | 'Vascular cell adhesion protein 1' |
| 9737 | 'VCAM1_No18' | 'LHIDDMEFEPK' | 'Vascular cell adhesion protein 1' |
| 9738 | 'VCAM1_No19' | 'DPEIHLSGPLEAGKPITVK' | 'Vascular cell adhesion protein 1' |
| 9739 | 'VCAM1_No20' | 'SLEMTFIPTIEDTGK' | 'Vascular cell adhesion protein 1' |
| 9740 | 'VCAM1_No21' | 'LQEGGSVTMTCSSEGLPAPEIFWSK' | 'Vascular cell adhesion protein 1' |
| 9741 | 'VCAM1_No22' | 'SIDGAYTIR' | 'Vascular cell adhesion protein 1' |
| 9742 | 'VCAM1_No23' | 'AKLHIDEMDSVPTVR' | 'Vascular cell adhesion protein 1' |
| 9743 | 'VCAM1_No24' | 'AKLHIDEMDSVPTVR' | 'Vascular cell adhesion protein 1' |
| 9744 | 'VCAM1_No25' | 'GETILENIEFLEDTDMK' | 'Vascular cell adhesion protein 1' |
| 9745 | 'VCAM1_No26' | 'AQLKDAGVYECESK' | 'Vascular cell adhesion protein 1' |
| 9746 | 'VCAM1_No27' | 'VTNEGTTSTLTMNPVSFGNEHSYLCTATCESR' | 'Vascular cell adhesion protein 1' |
| 9747 | 'VCAM1_No28' | 'VPSVYPLDR' | 'Vascular cell adhesion protein 1' |
| 9748 | 'VCAM1_No29' | 'QAVKELQVYISPK' | 'Vascular cell adhesion protein 1' |
| 9749 | 'VCAM1_No30' | 'SQEFLEDADRKSLETK' | 'Vascular cell adhesion protein 1' |
| 9750 | 'VCAM1_No31' | 'AQLKDAGVYECESKNK' | 'Vascular cell adhesion protein 1' |
| 9751 | 'HK1_No1' | 'MVSGMYLGELVR' | 'Hexokinase-1' |
| 9752 | 'HK1_No2' | 'LSDETLIDIMTR' | 'Hexokinase-1' |
| 9753 | 'HK1_No3' | 'AILQQLGLNSTCDDSILVK' | 'Hexokinase-1' |
| 9754 | 'HK1_No4' | 'MISGMYLGEIVR' | 'Hexokinase-1' |
| 9755 | 'HK1_No5' | 'QIEETLAHFHLTK' | 'Hexokinase-1' |
| 9756 | 'HK1_No6' | 'ATDCVGHDVVTLLR' | 'Hexokinase-1' |
| 9757 | 'HK1_No7' | 'FLSQIESDR' | 'Hexokinase-1' |
| 9758 | 'ORM2_No1' | 'EHVAHLLFLR' | 'Alpha-1-acid glycoprotein 2' |
| 9759 | 'ORM2_No2' | 'TLMFGSYLDDEKNWGLSFYADKPETTK' | 'Alpha-1-acid glycoprotein 2' |
| 9760 | 'ORM2_No3' | 'YEGGREHVAHLLFLR' | 'Alpha-1-acid glycoprotein 2' |
| 9761 | 'ORM2_No4' | 'TLMFGSYLDDEKNWGLSFYADKPETTK' | 'Alpha-1-acid glycoprotein 2' |
| 9762 | 'ORM2_No5' | 'SDVMYTDWKK' | 'Alpha-1-acid glycoprotein 2' |
| 9763 | 'ORM2_No6' | 'TLMFGSYLDDEK' | 'Alpha-1-acid glycoprotein 2' |
| 9764 | 'ORM2_No7' | 'SDVMYTDWKK' | 'Alpha-1-acid glycoprotein 2' |
| 9765 | 'ORM2_No8' | 'SDVMYTDWK' | 'Alpha-1-acid glycoprotein 2' |
| 9766 | 'ORM2_No9' | 'EQLGEFYEALDCLCIPR' | 'Alpha-1-acid glycoprotein 2' |
| 9767 | 'ORM2_No10' | 'EHVAHLLFLRDTK' | 'Alpha-1-acid glycoprotein 2' |
| 9768 | 'ORM2_No11' | 'SDVMYTDWKKDK' | 'Alpha-1-acid glycoprotein 2' |
| 9769 | 'ORM2_No12' | 'YEGGREHVAHLLFLRDTK' | 'Alpha-1-acid glycoprotein 2' |

| | A | B | C |
|---|---|---|---|
| 1 | **Peptide Name** | **Peptide Sequence** | **Peptide Description** |
| 9770 | 'ORM2_No13' | 'ITGKWFYIASAFR' | 'Alpha-1-acid glycoprotein 2' |
| 9771 | 'ORM2_No14' | 'SDVMYTDWK' | 'Alpha-1-acid glycoprotein 2' |
| 9772 | 'ORM2_No15' | 'QNQCFYNSSYLNVQR' | 'Alpha-1-acid glycoprotein 2' |
| 9773 | 'ORM2_No16' | 'NWGLSFYADKPETTK' | 'Alpha-1-acid glycoprotein 2' |
| 9774 | 'ORM2_No17' | 'SDVMYTDWKKDK' | 'Alpha-1-acid glycoprotein 2' |
| 9775 | 'ORM2_No18' | 'TLMFGSYLDDEK' | 'Alpha-1-acid glycoprotein 2' |
| 9776 | 'ORM2_No19' | 'ITGKWFYIASAFRNEEYNK' | 'Alpha-1-acid glycoprotein 2' |
| 9777 | 'ITIH2_No1' | 'FLHVPDTFEGHFDGVPVISK' | 'Inter-alpha-trypsin inhibitor heavy chain H2' |
| 9778 | 'ITIH2_No2' | 'VVNNSPQPQNVVFDVQIPK' | 'Inter-alpha-trypsin inhibitor heavy chain H2' |
| 9779 | 'ITIH2_No3' | 'IQPSGGTNINEALLR' | 'Inter-alpha-trypsin inhibitor heavy chain H2' |
| 9780 | 'ITIH2_No4' | 'SILQMSLDHHIVTPLTSLVIENEAGDER' | 'Inter-alpha-trypsin inhibitor heavy chain H2' |
| 9781 | 'ITIH2_No5' | 'KLWAYLTINQLLAER' | 'Inter-alpha-trypsin inhibitor heavy chain H2' |
| 9782 | 'ITIH2_No6' | 'TILDDLRAEDHFSVIDFNQNIR' | 'Inter-alpha-trypsin inhibitor heavy chain H2' |
| 9783 | 'ITIH2_No7' | 'NVQFNYPHTSVTDVTQNNFHNYFGGSEIVVAGK' | 'Inter-alpha-trypsin inhibitor heavy chain H2' |
| 9784 | 'ITIH2_No8' | 'VQFELHYQEVK' | 'Inter-alpha-trypsin inhibitor heavy chain H2' |
| 9785 | 'ITIH2_No9' | 'FYNQVSTPLLR' | 'Inter-alpha-trypsin inhibitor heavy chain H2' |
| 9786 | 'ITIH2_No10' | 'IYLQPGR' | 'Inter-alpha-trypsin inhibitor heavy chain H2' |
| 9787 | 'ITIH2_No11' | 'AHVSFKPTVAQQR' | 'Inter-alpha-trypsin inhibitor heavy chain H2' |
| 9788 | 'ITIH2_No12' | 'IYGNQDTSSQLKK' | 'Inter-alpha-trypsin inhibitor heavy chain H2' |
| 9789 | 'ITIH2_No13' | 'IYGNQDTSSQLK' | 'Inter-alpha-trypsin inhibitor heavy chain H2' |
| 9790 | 'ITIH2_No14' | 'HLEVDVWVIEPQGLR' | 'Inter-alpha-trypsin inhibitor heavy chain H2' |
| 9791 | 'ITIH2_No15' | 'SSALDMENFR' | 'Inter-alpha-trypsin inhibitor heavy chain H2' |
| 9792 | 'ITIH2_No16' | 'ETAVDGELVVLYDVKR' | 'Inter-alpha-trypsin inhibitor heavy chain H2' |
| 9793 | 'ITIH2_No17' | 'TEVNVLPGAK' | 'Inter-alpha-trypsin inhibitor heavy chain H2' |
| 9794 | 'ITIH2_No18' | 'VQSTITSR' | 'Inter-alpha-trypsin inhibitor heavy chain H2' |
| 9795 | 'ITIH2_No19' | 'MLADAPPQDPSCCSGALYYGSK' | 'Inter-alpha-trypsin inhibitor heavy chain H2' |
| 9796 | 'ITIH2_No20' | 'TWRNDLISATK' | 'Inter-alpha-trypsin inhibitor heavy chain H2' |
| 9797 | 'ITIH2_No21' | 'AEDHFSVIDFNQNIR' | 'Inter-alpha-trypsin inhibitor heavy chain H2' |
| 9798 | 'ITIH2_No22' | 'SSALDMENFRTEVNVLPGAK' | 'Inter-alpha-trypsin inhibitor heavy chain H2' |
| 9799 | 'ITIH2_No23' | 'MATTMIQSK' | 'Inter-alpha-trypsin inhibitor heavy chain H2' |
| 9800 | 'ITIH2_No24' | 'ETAVDGELVVLYDVK' | 'Inter-alpha-trypsin inhibitor heavy chain H2' |
| 9801 | 'ITIH2_No25' | 'LSNENHGIAQR' | 'Inter-alpha-trypsin inhibitor heavy chain H2' |
| 9802 | 'ITIH2_No26' | 'KFYNQVSTPLLR' | 'Inter-alpha-trypsin inhibitor heavy chain H2' |
| 9803 | 'ITIH2_No27' | 'HADPDFTR' | 'Inter-alpha-trypsin inhibitor heavy chain H2' |
| 9804 | 'ITIH2_No28' | 'TILDDLR' | 'Inter-alpha-trypsin inhibitor heavy chain H2' |
| 9805 | 'ITIH2_No29' | 'SILQMSLDHHIVTPLTSLVIENEAGDER' | 'Inter-alpha-trypsin inhibitor heavy chain H2' |
| 9806 | 'ITIH2_No30' | 'PTVAQQR' | 'Inter-alpha-trypsin inhibitor heavy chain H2' |
| 9807 | 'ITIH2_No31' | 'SSALDMENFR' | 'Inter-alpha-trypsin inhibitor heavy chain H2' |
| 9808 | 'ITIH2_No32' | 'ETAVDGELVVLYDVKREEK' | 'Inter-alpha-trypsin inhibitor heavy chain H2' |
| 9809 | 'ITIH2_No33' | 'VQFELHYQEVKWR' | 'Inter-alpha-trypsin inhibitor heavy chain H2' |
| 9810 | 'ITIH2_No34' | 'SLAPTAAAK' | 'Inter-alpha-trypsin inhibitor heavy chain H2' |
| 9811 | 'ITIH2_No35' | 'LWAYLTINQLLAER' | 'Inter-alpha-trypsin inhibitor heavy chain H2' |
| 9812 | 'ITIH2_No36' | 'YIEKIQPSGGTNINEALLR' | 'Inter-alpha-trypsin inhibitor heavy chain H2' |
| 9813 | 'ITIH2_No37' | 'FLHVPDTFEGHFDGVPVISKGQQK' | 'Inter-alpha-trypsin inhibitor heavy chain H2' |

| | A | B | C |
|---|---|---|---|
| 1 | **Peptide Name** | **Peptide Sequence** | **Peptide Description** |
| 9814 | 'ITIH2_No38' | 'MLADAPPQDPSCCSGALYYGSK' | 'Inter-alpha-trypsin inhibitor heavy chain H2' |
| 9815 | 'ITIH2_No39' | 'NVQFNYPHTSVTDVTQNNFHNYFGGSEIVVAGKFDPAK' | 'Inter-alpha-trypsin inhibitor heavy chain H2' |
| 9816 | 'ITIH2_No40' | 'MATTMIQSK' | 'Inter-alpha-trypsin inhibitor heavy chain H2' |
| 9817 | 'ITIH2_No41' | 'NDLISATK' | 'Inter-alpha-trypsin inhibitor heavy chain H2' |
| 9818 | 'ITIH2_No42' | 'RLSNENHGIAQR' | 'Inter-alpha-trypsin inhibitor heavy chain H2' |
| 9819 | 'ITIH2_No43' | 'MATTMIQSK' | 'Inter-alpha-trypsin inhibitor heavy chain H2' |
| 9820 | 'ITIH2_No44' | 'SSALDMENFRTEVNVLPGAK' | 'Inter-alpha-trypsin inhibitor heavy chain H2' |
| 9821 | 'ITIH2_No45' | 'DKHADPDFTR' | 'Inter-alpha-trypsin inhibitor heavy chain H2' |
| 9822 | 'ITIH2_No46' | 'MATTMIQSK' | 'Inter-alpha-trypsin inhibitor heavy chain H2' |
| 9823 | 'ITIH2_No47' | 'LGSYEHR' | 'Inter-alpha-trypsin inhibitor heavy chain H2' |
| 9824 | 'ITIH2_No48' | 'KLGSYEHRIYLQPGR' | 'Inter-alpha-trypsin inhibitor heavy chain H2' |
| 9825 | 'ITIH2_No49' | 'KLGSYEHR' | 'Inter-alpha-trypsin inhibitor heavy chain H2' |
| 9826 | 'ITIH2_No50' | 'MKQTVEAMK' | 'Inter-alpha-trypsin inhibitor heavy chain H2' |
| 9827 | 'ITIH2_No51' | 'LGSYEHRIYLQPGR' | 'Inter-alpha-trypsin inhibitor heavy chain H2' |
| 9828 | 'ITIH2_No52' | 'VQFELHYQEVKWRK' | 'Inter-alpha-trypsin inhibitor heavy chain H2' |
| 9829 | 'ITIH2_No53' | 'MATTMIQSKVVNNSPQPQNVVFDVQIPK' | 'Inter-alpha-trypsin inhibitor heavy chain H2' |
| 9830 | 'ITIH2_No54' | 'GKTAGLVR' | 'Inter-alpha-trypsin inhibitor heavy chain H2' |
| 9831 | 'ITIH2_No55' | 'QTVEAMK' | 'Inter-alpha-trypsin inhibitor heavy chain H2' |
| 9832 | 'ITIH2_No56' | 'TEVNVLPGAKVQFELHYQEVK' | 'Inter-alpha-trypsin inhibitor heavy chain H2' |
| 9833 | 'ITIH2_No57' | 'NDLISATKTQVADAKR' | 'Inter-alpha-trypsin inhibitor heavy chain H2' |
| 9834 | 'ITIH2_No58' | 'SLPGESEEMMEEVDQVTLYSYK' | 'Inter-alpha-trypsin inhibitor heavy chain H2' |
| 9835 | 'ITIH2_No59' | 'IYLQPGRLAK' | 'Inter-alpha-trypsin inhibitor heavy chain H2' |
| 9836 | 'ITIH2_No60' | 'ILNLVSDPESGIVVNGQLVGAK' | 'Inter-alpha-trypsin inhibitor heavy chain H2' |
| 9837 | 'ITIH2_No61' | 'TWRNDLISATKTQVADAK' | 'Inter-alpha-trypsin inhibitor heavy chain H2' |
| 9838 | 'ITIH2_No62' | 'RYIEKIQPSGGTNINEALLR' | 'Inter-alpha-trypsin inhibitor heavy chain H2' |
| 9839 | 'ITIH2_No63' | 'DKHADPDFTRK' | 'Inter-alpha-trypsin inhibitor heavy chain H2' |
| 9840 | 'ITIH2_No64' | 'NDLISATKTQVADAK' | 'Inter-alpha-trypsin inhibitor heavy chain H2' |
| 9841 | 'ITIH1_No1' | 'VTFQLTYEEVLKR' | 'Inter-alpha-trypsin inhibitor heavy chain H1' |
| 9842 | 'ITIH1_No2' | 'QYYEGSEIVVAGR' | 'Inter-alpha-trypsin inhibitor heavy chain H1' |
| 9843 | 'ITIH1_No3' | 'QAVDTAVDGVFIR' | 'Inter-alpha-trypsin inhibitor heavy chain H1' |
| 9844 | 'ITIH1_No4' | 'TMEQFTIHLTVNPQSK' | 'Inter-alpha-trypsin inhibitor heavy chain H1' |
| 9845 | 'ITIH1_No5' | 'PTVSQQQSCPTCSTSLLNGHFK' | 'Inter-alpha-trypsin inhibitor heavy chain H1' |
| 9846 | 'ITIH1_No6' | 'KGHVLFRPTVSQQQSCPTCSTSLLNGHFK' | 'Inter-alpha-trypsin inhibitor heavy chain H1' |
| 9847 | 'ITIH1_No7' | 'EVAFDLEIPK' | 'Inter-alpha-trypsin inhibitor heavy chain H1' |
| 9848 | 'ITIH1_No8' | 'FAHYVVTSQVVNTANEAR' | 'Inter-alpha-trypsin inhibitor heavy chain H1' |
| 9849 | 'ITIH1_No9' | 'QLVHHFEIDVDIFEPQGISK' | 'Inter-alpha-trypsin inhibitor heavy chain H1' |
| 9850 | 'ITIH1_No10' | 'KAAISGENAGLVR' | 'Inter-alpha-trypsin inhibitor heavy chain H1' |
| 9851 | 'ITIH1_No11' | 'TAFISDFAVTADGNAFIGDIKDK' | 'Inter-alpha-trypsin inhibitor heavy chain H1' |
| 9852 | 'ITIH1_No12' | 'ADVQAHGEGQEFSITCLVDEEEMKK' | 'Inter-alpha-trypsin inhibitor heavy chain H1' |
| 9853 | 'ITIH1_No13' | 'LDAQASFLPK' | 'Inter-alpha-trypsin inhibitor heavy chain H1' |
| 9854 | 'ITIH1_No14' | 'TAFISDFAVTADGNAFIGDIKDKVTAWK' | 'Inter-alpha-trypsin inhibitor heavy chain H1' |
| 9855 | 'ITIH1_No15' | 'GMADQDGLKPTIDKPSEDSPPLEMLGPR' | 'Inter-alpha-trypsin inhibitor heavy chain H1' |
| 9856 | 'ITIH1_No16' | 'TMEQFTIHLTVNPQSK' | 'Inter-alpha-trypsin inhibitor heavy chain H1' |
| 9857 | 'ITIH1_No17' | 'QLVHHFEIDVDIFEPQGISK' | 'Inter-alpha-trypsin inhibitor heavy chain H1' |

| | A | B | C |
|---|---|---|---|
| 1 | **Peptide Name** | **Peptide Sequence** | **Peptide Description** |
| 9858 | 'ITIH1_No18' | 'NVVFVIDISGSMR' | 'Inter-alpha-trypsin inhibitor heavy chain H1' |
| 9859 | 'ITIH1_No19' | 'PLLVDVDLQYPQDAVLALTQNHHK' | 'Inter-alpha-trypsin inhibitor heavy chain H1' |
| 9860 | 'ITIH1_No20' | 'ADVQAHGEGQEFSITCLVDEEEMK' | 'Inter-alpha-trypsin inhibitor heavy chain H1' |
| 9861 | 'ITIH1_No21' | 'LWAYLTIQELLAKR' | 'Inter-alpha-trypsin inhibitor heavy chain H1' |
| 9862 | 'ITIH1_No22' | 'RNHMQYEIVIK' | 'Inter-alpha-trypsin inhibitor heavy chain H1' |
| 9863 | 'ITIH1_No23' | 'ADVQAHGEGQEFSITCLVDEEEMK' | 'Inter-alpha-trypsin inhibitor heavy chain H1' |
| 9864 | 'ITIH1_No24' | 'GSLVQASEANLQAAQDFVR' | 'Inter-alpha-trypsin inhibitor heavy chain H1' |
| 9865 | 'ITIH1_No25' | 'RTFVLSALQPSPTHSSSNTQR' | 'Inter-alpha-trypsin inhibitor heavy chain H1' |
| 9866 | 'ITIH1_No26' | 'TAFISDFAVTADGNAFIGDIK' | 'Inter-alpha-trypsin inhibitor heavy chain H1' |
| 9867 | 'ITIH1_No27' | 'VTFQLTYEEVLK' | 'Inter-alpha-trypsin inhibitor heavy chain H1' |
| 9868 | 'ITIH1_No28' | 'GHVLFRPTVSQQQSCPTCSTSLLNGHFK' | 'Inter-alpha-trypsin inhibitor heavy chain H1' |
| 9869 | 'ITIH1_No29' | 'ILGDMQPGDYFDLVLFGTR' | 'Inter-alpha-trypsin inhibitor heavy chain H1' |
| 9870 | 'ITIH1_No30' | 'NVVFVIDISGSMR' | 'Inter-alpha-trypsin inhibitor heavy chain H1' |
| 9871 | 'ITIH1_No31' | 'NHMQYEIVIK' | 'Inter-alpha-trypsin inhibitor heavy chain H1' |
| 9872 | 'ITIH1_No32' | 'IYEDHDATQQLQGFYSQVAK' | 'Inter-alpha-trypsin inhibitor heavy chain H1' |
| 9873 | 'ITIH1_No33' | 'VTYDVSR' | 'Inter-alpha-trypsin inhibitor heavy chain H1' |
| 9874 | 'ITIH1_No34' | 'QAVDTAVDGVFIR' | 'Inter-alpha-trypsin inhibitor heavy chain H1' |
| 9875 | 'ITIH1_No35' | 'GHMLENHVER' | 'Inter-alpha-trypsin inhibitor heavy chain H1' |
| 9876 | 'ITIH1_No36' | 'ILGDMQPGDYFDLVLFGTR' | 'Inter-alpha-trypsin inhibitor heavy chain H1' |
| 9877 | 'ITIH1_No37' | 'ELAAQTIK' | 'Inter-alpha-trypsin inhibitor heavy chain H1' |
| 9878 | 'ITIH1_No38' | 'ADVQAHGEGQEFSITCLVDEEEMKK' | 'Inter-alpha-trypsin inhibitor heavy chain H1' |
| 9879 | 'ITIH1_No39' | 'GMADQDGLKPTIDKPSEDSPPLEMLGPR' | 'Inter-alpha-trypsin inhibitor heavy chain H1' |
| 9880 | 'ITIH1_No40' | 'NHMQYEIVIK' | 'Inter-alpha-trypsin inhibitor heavy chain H1' |
| 9881 | 'ITIH1_No41' | 'RNHMQYEIVIK' | 'Inter-alpha-trypsin inhibitor heavy chain H1' |
| 9882 | 'ITIH1_No42' | 'AAISGENAGLVR' | 'Inter-alpha-trypsin inhibitor heavy chain H1' |
| 9883 | 'ITIH1_No43' | 'GFSLDEATNLNGGLLR' | 'Inter-alpha-trypsin inhibitor heavy chain H1' |
| 9884 | 'ITIH1_No44' | 'GMADQDGLKPTIDKPSEDSPPLEMLGPR' | 'Inter-alpha-trypsin inhibitor heavy chain H1' |
| 9885 | 'ITIH1_No45' | 'ELAAQTIKK' | 'Inter-alpha-trypsin inhibitor heavy chain H1' |
| 9886 | 'ITIH1_No46' | 'GSSVHQDFLGFYVLDSHR' | 'Inter-alpha-trypsin inhibitor heavy chain H1' |
| 9887 | 'ITIH1_No47' | 'TFVLSALQPSPTHSSSNTQR' | 'Inter-alpha-trypsin inhibitor heavy chain H1' |
| 9888 | 'ITIH1_No48' | 'RQAVDTAVDGVFIR' | 'Inter-alpha-trypsin inhibitor heavy chain H1' |
| 9889 | 'ITIH1_No49' | 'QYYEGSEIVVAGR' | 'Inter-alpha-trypsin inhibitor heavy chain H1' |
| 9890 | 'ITIH1_No50' | 'LWAYLTIQELLAK' | 'Inter-alpha-trypsin inhibitor heavy chain H1' |
| 9891 | 'ITIH1_No51' | 'ADVQAHGEGQEFSITCLVDEEEMKKLLR' | 'Inter-alpha-trypsin inhibitor heavy chain H1' |
| 9892 | 'ITIH1_No52' | 'DKICDLLVANNHFAHFFAPQNLTNMNK' | 'Inter-alpha-trypsin inhibitor heavy chain H1' |
| 9893 | 'ITIH1_No53' | 'VQSWKGSLVQASEANLQAAQDFVR' | 'Inter-alpha-trypsin inhibitor heavy chain H1' |
| 9894 | 'ITIH1_No54' | 'DKICDLLVANNHFAHFFAPQNLTNMNK' | 'Inter-alpha-trypsin inhibitor heavy chain H1' |
| 9895 | 'ITIH1_No55' | 'GHMLENHVER' | 'Inter-alpha-trypsin inhibitor heavy chain H1' |
| 9896 | 'ITIH1_No56' | 'ERGHMLENHVER' | 'Inter-alpha-trypsin inhibitor heavy chain H1' |
| 9897 | 'ITIH1_No57' | 'DKVTAWK' | 'Inter-alpha-trypsin inhibitor heavy chain H1' |
| 9898 | 'ITIH1_No58' | 'ICDLLVANNHFAHFFAPQNLTNMNK' | 'Inter-alpha-trypsin inhibitor heavy chain H1' |
| 9899 | 'ITIH1_No59' | 'LDAQASFLPKELAAQTIKK' | 'Inter-alpha-trypsin inhibitor heavy chain H1' |
| 9900 | 'ITIH1_No60' | 'QSSFKADVQAHGEGQEFSITCLVDEEEMKK' | 'Inter-alpha-trypsin inhibitor heavy chain H1' |
| 9901 | 'ITIH1_No61' | 'ERGHMLENHVER' | 'Inter-alpha-trypsin inhibitor heavy chain H1' |

| | A | B | C |
|---|---|---|---|
| 1 | **Peptide Name** | **Peptide Sequence** | **Peptide Description** |
| 9902 | 'ITIH1_No62' | 'VTYDVSRDK' | 'Inter-alpha-trypsin inhibitor heavy chain H1' |
| 9903 | 'ITIH1_No63' | 'GMADQDGLKPTIDKPSEDSPPLEMLGPR' | 'Inter-alpha-trypsin inhibitor heavy chain H1' |
| 9904 | 'ITIH1_No64' | 'LDAQASFLPKELAAQTIK' | 'Inter-alpha-trypsin inhibitor heavy chain H1' |
| 9905 | 'ITIH1_No65' | 'VTSRFAHYVVTSQVVNTANEAR' | 'Inter-alpha-trypsin inhibitor heavy chain H1' |
| 9906 | 'ITIH1_No66' | 'KGHVLFR' | 'Inter-alpha-trypsin inhibitor heavy chain H1' |
| 9907 | 'ITIH1_No67' | 'ADVQAHGEGQEFSITCLVDEEEMKKLLR' | 'Inter-alpha-trypsin inhibitor heavy chain H1' |
| 9908 | 'ITIH1_No68' | 'DQAVLRQDGVVVTINK' | 'Inter-alpha-trypsin inhibitor heavy chain H1' |
| 9909 | 'ITIH1_No69' | 'QAVDTAVDGVFIRSLKVNCK' | 'Inter-alpha-trypsin inhibitor heavy chain H1' |
| 9910 | 'ITIH1_No70' | 'IADNKQSSFK' | 'Inter-alpha-trypsin inhibitor heavy chain H1' |
| 9911 | 'ITIH1_No71' | 'QSSFKADVQAHGEGQEFSITCLVDEEEMKK' | 'Inter-alpha-trypsin inhibitor heavy chain H1' |
| 9912 | 'ITIH1_No72' | 'QAVDTAVDGVFIRSLKVNCK' | 'Inter-alpha-trypsin inhibitor heavy chain H1' |
| 9913 | 'ITIH1_No73' | 'ANLSSQALQMSLDYGFVTPLTSMSIR' | 'Inter-alpha-trypsin inhibitor heavy chain H1' |
| 9914 | 'ITIH1_No74' | 'SLKVNCK' | 'Inter-alpha-trypsin inhibitor heavy chain H1' |
| 9915 | 'ANXA7_No1' | 'GFGTDEQAIVDVVANR' | 'Annexin A7' |
| 9916 | 'ANXA7_No2' | 'LGTDESCFNMILATR' | 'Annexin A7' |
| 9917 | 'ANXA7_No3' | 'VLIEILCTR' | 'Annexin A7' |
| 9918 | 'AZU1_No1' | 'QFPFLASIQNQGR' | 'Azurocidin' |
| 9919 | 'AZU1_No2' | 'DWIDGVLNNPGPGPA' | 'Azurocidin' |
| 9920 | 'PSMB1_No1' | 'NMQNVEHVPLSLDR' | 'Proteasome subunit beta type-1' |
| 9921 | 'PSMB1_No2' | 'AGGSASAMLQPLLDNQVGFK' | 'Proteasome subunit beta type-1' |
| 9922 | 'PSMB1_No3' | 'EGIREETVSLR' | 'Proteasome subunit beta type-1' |
| 9923 | 'PSMB1_No4' | 'LSEGFSIHTR' | 'Proteasome subunit beta type-1' |
| 9924 | 'PSMB1_No5' | 'AGGSASAMLQPLLDNQVGFK' | 'Proteasome subunit beta type-1' |
| 9925 | 'PZP_No1' | 'GSFALSFPVESDVAPIAR' | 'Pregnancy zone protein' |
| 9926 | 'PZP_No2' | 'QQNAQGGFSSTQDTVVALHALSR' | 'Pregnancy zone protein' |
| 9927 | 'PZP_No3' | 'SGTHTLPVESGDMK' | 'Pregnancy zone protein' |
| 9928 | 'PZP_No4' | 'YNILPEKEDSPFALK' | 'Pregnancy zone protein' |
| 9929 | 'PZP_No5' | 'AVGYLITGYQR' | 'Pregnancy zone protein' |
| 9930 | 'PZP_No6' | 'MLQITNTGFEMK' | 'Pregnancy zone protein' |
| 9931 | 'PZP_No7' | 'ASPAFLASQNTK' | 'Pregnancy zone protein' |
| 9932 | 'PZP_No8' | 'NELIPLIYLENPRR' | 'Pregnancy zone protein' |
| 9933 | 'PZP_No9' | 'QAMGELSELSFHYLIMAK' | 'Pregnancy zone protein' |
| 9934 | 'PZP_No10' | 'SLFTDLVAEK' | 'Pregnancy zone protein' |
| 9935 | 'PZP_No11' | 'CVYLQTSMK' | 'Pregnancy zone protein' |
| 9936 | 'PZP_No12' | 'NALFCLESAWNVAK' | 'Pregnancy zone protein' |
| 9937 | 'PZP_No13' | 'TLLVEAEGIEQEK' | 'Pregnancy zone protein' |
| 9938 | 'PZP_No14' | 'DLFHCVSFTLPR' | 'Pregnancy zone protein' |
| 9939 | 'PZP_No15' | 'GEESYCICGNER' | 'Pregnancy zone protein' |
| 9940 | 'PZP_No16' | 'VFSLSGSYIHLEPVAGTLPCGHTETITAHYTLNR' | 'Pregnancy zone protein' |
| 9941 | 'PZP_No17' | 'EILNSLDKEAVK' | 'Pregnancy zone protein' |
| 9942 | 'PZP_No18' | 'ISEITNIVSK' | 'Pregnancy zone protein' |
| 9943 | 'PZP_No19' | 'QAMGELSELSFHYLIMAK' | 'Pregnancy zone protein' |
| 9944 | 'PZP_No20' | 'NELIPLIYLENPR' | 'Pregnancy zone protein' |
| 9945 | 'PZP_No21' | 'IAQWQSLK' | 'Pregnancy zone protein' |

| | A | B | C |
|---|---|---|---|
| 1 | **Peptide Name** | **Peptide Sequence** | **Peptide Description** |
| 9946 | 'PZP_No22' | 'SGTHTLPVESGDMK' | 'Pregnancy zone protein' |
| 9947 | 'PZP_No23' | 'ALLAYAFSLLGK' | 'Pregnancy zone protein' |
| 9948 | 'PZP_No24' | 'VFTVHPNLCFHYSWVAEDHQGAQHTANR' | 'Pregnancy zone protein' |
| 9949 | 'PZP_No25' | 'IQHPFTVEEFVLPK' | 'Pregnancy zone protein' |
| 9950 | 'PZP_No26' | 'EDNLVHWERPQRPK' | 'Pregnancy zone protein' |
| 9951 | 'PZP_No27' | 'NTVLVLNTQSLVFVQTDKPMYKPGQTVR' | 'Pregnancy zone protein' |
| 9952 | 'PZP_No28' | 'VLNCDKQEVCEEFSQQLNSNGCITQQVHTK' | 'Pregnancy zone protein' |
| 9953 | 'PZP_No29' | 'NTVLVLNTQSLVFVQTDKPMYKPGQTVR' | 'Pregnancy zone protein' |
| 9954 | 'PZP_No30' | 'IREEGTDLEVTANR' | 'Pregnancy zone protein' |
| 9955 | 'PZP_No31' | 'AVDQSVLLMKPEAELSVSSVYNLLTVK' | 'Pregnancy zone protein' |
| 9956 | 'PZP_No32' | 'QAMGELSELSFHYLIMAK' | 'Pregnancy zone protein' |
| 9957 | 'PZP_No33' | 'IQHPFTVEEFVLPKFEVK' | 'Pregnancy zone protein' |
| 9958 | 'PZP_No34' | 'VVSVDENFRPR' | 'Pregnancy zone protein' |
| 9959 | 'PZP_No35' | 'SLFTDLVAEKDLFHCVSFTLPR' | 'Pregnancy zone protein' |
| 9960 | 'PZP_No36' | 'QQNAQGGFSSTQDTVVALHALSR' | 'Pregnancy zone protein' |
| 9961 | 'PZP_No37' | 'AVDQSVLLMKPEAELSVSSVYNLLTVK' | 'Pregnancy zone protein' |
| 9962 | 'PZP_No38' | 'EILNSLDKEAVKEDNLVHWER' | 'Pregnancy zone protein' |
| 9963 | 'PZP_No39' | 'QGIPFFAQVLLVDGK' | 'Pregnancy zone protein' |
| 9964 | 'PZP_No40' | 'QAMGELSELSFHYLIMAK' | 'Pregnancy zone protein' |
| 9965 | 'PZP_No41' | 'QAMGELSELSFHYLIMAK' | 'Pregnancy zone protein' |
| 9966 | 'PZP_No42' | 'EILNSLDK' | 'Pregnancy zone protein' |
| 9967 | 'PZP_No43' | 'FRVVSVDENFRPR' | 'Pregnancy zone protein' |
| 9968 | 'PZP_No44' | 'QAMGELSELSFHYLIMAK' | 'Pregnancy zone protein' |
| 9969 | 'PZP_No45' | 'IAQWQSLKLEAGINQLSFPLSSEPIQGSYR' | 'Pregnancy zone protein' |
| 9970 | 'PZP_No46' | 'VVVQTESGGR' | 'Pregnancy zone protein' |
| 9971 | 'PZP_No47' | 'MLQITNTGFEMK' | 'Pregnancy zone protein' |
| 9972 | 'PZP_No48' | 'QLNYKHQDGSYSTFGER' | 'Pregnancy zone protein' |
| 9973 | 'PZP_No49' | 'MLQITNTGFEMK' | 'Pregnancy zone protein' |
| 9974 | 'PZP_No50' | 'EEGTDLEVTANR' | 'Pregnancy zone protein' |
| 9975 | 'PZP_No51' | 'HQDGSYSTFGER' | 'Pregnancy zone protein' |
| 9976 | 'PZP_No52' | 'IISIMDEK' | 'Pregnancy zone protein' |
| 9977 | 'PZP_No53' | 'PVPGLATVSLCR' | 'Pregnancy zone protein' |
| 9978 | 'PZP_No54' | 'FEIENCLANK' | 'Pregnancy zone protein' |
| 9979 | 'PZP_No55' | 'ISASSEVAFLSIQIK' | 'Pregnancy zone protein' |
| 9980 | 'PZP_No56' | 'VQTVPQTCDGHK' | 'Pregnancy zone protein' |
| 9981 | 'PZP_No57' | 'QAMGELSELSFHYLIMAK' | 'Pregnancy zone protein' |
| 9982 | 'PZP_No58' | 'QLNYKHQDGSYSTFGER' | 'Pregnancy zone protein' |
| 9983 | 'PZP_No59' | 'QEVCEEFSQQLNSNGCITQQVHTK' | 'Pregnancy zone protein' |
| 9984 | 'PZP_No60' | 'PYVPQLGTYNVIPLNNEQSSGPVPETVR' | 'Pregnancy zone protein' |
| 9985 | 'PZP_No61' | 'SYIFIDEAHITQSLTWLSQMQK' | 'Pregnancy zone protein' |
| 9986 | 'PZP_No62' | 'VDLSFSPAQSPPASHAHLQVAAAPQSLCALR' | 'Pregnancy zone protein' |
| 9987 | 'PZP_No63' | 'IREEGTDLEVTANRISEITNIVSK' | 'Pregnancy zone protein' |
| 9988 | 'PZP_No64' | 'LEAGINQLSFPLSSEPIQGSYR' | 'Pregnancy zone protein' |
| 9989 | 'PZP_No65' | 'QGIPFFAQVLLVDGK' | 'Pregnancy zone protein' |

| | A | B | C |
|---|---|---|---|
| 1 | **Peptide Name** | **Peptide Sequence** | **Peptide Description** |
| 9990 | 'PZP_No66' | 'GPTQDFR' | 'Pregnancy zone protein' |
| 9991 | 'PZP_No67' | 'EAVKEDNLVHWER' | 'Pregnancy zone protein' |
| 9992 | 'C4BPB_No1' | 'LIQEAPKPECEK' | 'C4b-binding protein beta chain' |
| 9993 | 'C4BPB_No2' | 'SQCLEDHTWAPPFPICK' | 'C4b-binding protein beta chain' |
| 9994 | 'C4BPB_No3' | 'ITFMCNDHYILK' | 'C4b-binding protein beta chain' |
| 9995 | 'C4BPB_No4' | 'EVEGQILGTYVCIK' | 'C4b-binding protein beta chain' |
| 9996 | 'C4BPB_No5' | 'NLCEAMENFMQQLK' | 'C4b-binding protein beta chain' |
| 9997 | 'C4BPB_No6' | 'ALLAFQESK' | 'C4b-binding protein beta chain' |
| 9998 | 'C4BPB_No7' | 'ESGMTMEELK' | 'C4b-binding protein beta chain' |
| 9999 | 'C4BPB_No8' | 'ITFMCNDHYILK' | 'C4b-binding protein beta chain' |
| 10000 | 'C4BPB_No9' | 'ESGMTMEELKYSLELK' | 'C4b-binding protein beta chain' |
| 10001 | 'C4BPB_No10' | 'ESGMTMEELK' | 'C4b-binding protein beta chain' |
| 10002 | 'C4BPB_No11' | 'YYLVGVQEQQCVDGEWSSALPVCK' | 'C4b-binding protein beta chain' |
| 10003 | 'C4BPB_No12' | 'ESGMTMEELKYSLELKK' | 'C4b-binding protein beta chain' |
| 10004 | 'C4BPB_No13' | 'NLCEAMENFMQQLK' | 'C4b-binding protein beta chain' |
| 10005 | 'C4BPB_No14' | 'LIQEAPKPECEKALLAFQESK' | 'C4b-binding protein beta chain' |
| 10006 | 'C4BPB_No15' | 'EWDNTTTECR' | 'C4b-binding protein beta chain' |
| 10007 | 'C4BPB_No16' | 'YSLELKKAELK' | 'C4b-binding protein beta chain' |
| 10008 | 'COL5A1' | 'VLDFHNLPDGITK' | 'Collagen alpha-1(V) chain' |
| 10009 | 'CSRP1_No1' | 'GYGYGQGAGTLSTDKGESLGIK' | 'Cysteine and glycine-rich protein 1' |
| 10010 | 'CSRP1_No2' | 'NLDSTTVAVHGEEIYCK' | 'Cysteine and glycine-rich protein 1' |
| 10011 | 'CSRP1_No3' | 'TVYFAEEVQCEGNSFHK' | 'Cysteine and glycine-rich protein 1' |
| 10012 | 'CSRP1_No4' | 'SCFLCMVCK' | 'Cysteine and glycine-rich protein 1' |
| 10013 | 'FLNA_No1' | 'VANPSGNLTETYVQDR' | 'Filamin-A' |
| 10014 | 'FLNA_No2' | 'AFGPGLQGGSAGSPAR' | 'Filamin-A' |
| 10015 | 'FLNA_No3' | 'LPQLPITNFSR' | 'Filamin-A' |
| 10016 | 'FLNA_No4' | 'SPFSVAVSPSLDLSK' | 'Filamin-A' |
| 10017 | 'FLNA_No5' | 'YTPVQQGPVGVNVTYGGDPIPK' | 'Filamin-A' |
| 10018 | 'FLNA_No6' | 'IVGPSGAAVPCKVEPGLGADNSVVR' | 'Filamin-A' |
| 10019 | 'FLNA_No7' | 'FGGEHVPNSPFQVTALAGDQPSVQPPLR' | 'Filamin-A' |
| 10020 | 'FLNA_No8' | 'LYSVSYLLK' | 'Filamin-A' |
| 10021 | 'FLNA_No9' | 'LIALLEVLSQK' | 'Filamin-A' |
| 10022 | 'FLNA_No10' | 'VTAQGPGLEPSGNIANK' | 'Filamin-A' |
| 10023 | 'FLNA_No11' | 'TGVAVNKPAEFTVDAK' | 'Filamin-A' |
| 10024 | 'FLNA_No12' | 'TFSVWYVPEVTGTHK' | 'Filamin-A' |
| 10025 | 'FLNA_No13' | 'VQVQDNEGCPVEALVK' | 'Filamin-A' |
| 10026 | 'FLNA_No14' | 'EGPYSISVLYGDEEVPR' | 'Filamin-A' |
| 10027 | 'FLNA_No15' | 'WGDEHIPGSPYR' | 'Filamin-A' |
| 10028 | 'FLNA_No16' | 'MDCQECPEGYR' | 'Filamin-A' |
| 10029 | 'FLNA_No17' | 'VTVLFAGQHIAK' | 'Filamin-A' |
| 10030 | 'FLNA_No18' | 'AWGPGLEGGVVGK' | 'Filamin-A' |
| 10031 | 'FLNA_No19' | 'FNEEHIPDSPFVVPVASPSGDAR' | 'Filamin-A' |
| 10032 | 'FLNA_No20' | 'VTYTPMAPGSYLISIK' | 'Filamin-A' |
| 10033 | 'FLNA_No21' | 'VEPGLGADNSVVR' | 'Filamin-A' |

| | A | B | C |
|---|---|---|---|
| 1 | **Peptide Name** | **Peptide Sequence** | **Peptide Description** |
| 10034 | 'FLNA_No22' | 'FADQHVPGSPFSVK' | 'Filamin-A' |
| 10035 | 'FLNA_No23' | 'VAQPTITDNKDGTVTVR' | 'Filamin-A' |
| 10036 | 'FLNA_No24' | 'YGGPYHIGGSPFK' | 'Filamin-A' |
| 10037 | 'FLNA_No25' | 'VNQPASFAVSLNGAK' | 'Filamin-A' |
| 10038 | 'FLNA_No26' | 'SAGQGEVLVYVEDPAGHQEEAK' | 'Filamin-A' |
| 10039 | 'FLNA_No27' | 'IANLQTDLSDGLR' | 'Filamin-A' |
| 10040 | 'FLNA_No28' | 'ATCAPQHGAPGPGPADASK' | 'Filamin-A' |
| 10041 | 'FLNA_No29' | 'KTHIQDNHDGTYTVAYVPDVTGR' | 'Filamin-A' |
| 10042 | 'FLNA_No30' | 'YWPQEAGEYAVHVLCNSEDIR' | 'Filamin-A' |
| 10043 | 'FLNA_No31' | 'CSGPGLSPGMVR' | 'Filamin-A' |
| 10044 | 'FLNA_No32' | 'AGNNMLLVGVHGPR' | 'Filamin-A' |
| 10045 | 'FLNA_No33' | 'SPFEVYVDK' | 'Filamin-A' |
| 10046 | 'FLNA_No34' | 'LVSNHSLHETSSVFVDSLTK' | 'Filamin-A' |
| 10047 | 'FLNA_No35' | 'AGQSAAGAAPGGGVDTR' | 'Filamin-A' |
| 10048 | 'FLNA_No36' | 'YNEQHVPGSPFTAR' | 'Filamin-A' |
| 10049 | 'FLNA_No37' | 'TGVELGKPTHFTVNAK' | 'Filamin-A' |
| 10050 | 'FLNA_No38' | 'SPYTVTVGQACNPSACR' | 'Filamin-A' |
| 10051 | 'FLNA_No39' | 'YAPSEAGLHEMDIR' | 'Filamin-A' |
| 10052 | 'FLNA_No40' | 'EAGAGGLAIAVEGPSK' | 'Filamin-A' |
| 10053 | 'FLNA_No41' | 'AHVVPCFDASK' | 'Filamin-A' |
| 10054 | 'FLNA_No42' | 'RLTVSSLQESGLK' | 'Filamin-A' |
| 10055 | 'FLNA_No43' | 'VTYCPTEPGNYIINIK' | 'Filamin-A' |
| 10056 | 'FLNA_No44' | 'GTVEPQLEAR' | 'Filamin-A' |
| 10057 | 'FLNA_No45' | 'DVDIIDHHDNTYTVK' | 'Filamin-A' |
| 10058 | 'FLNA_No46' | 'ASGPGLNTTGVPASLPVEFTIDAK' | 'Filamin-A' |
| 10059 | 'FLNA_No47' | 'YGGDEIPFSPYR' | 'Filamin-A' |
| 10060 | 'FLNA_No48' | 'VHSPSGALEECYVTEIDQDKYAVR' | 'Filamin-A' |
| 10061 | 'FLNA_No49' | 'TPCEEILVK' | 'Filamin-A' |
| 10062 | 'FLNA_No50' | 'AYGPGIEPTGNMVK' | 'Filamin-A' |
| 10063 | 'FLNA_No51' | 'LIALLEVLSQKK' | 'Filamin-A' |
| 10064 | 'FLNA_No52' | 'MDCQECPEGYR' | 'Filamin-A' |
| 10065 | 'FLNA_No53' | 'VHGPGIQSGTTNKPNKFTVETR' | 'Filamin-A' |
| 10066 | 'FLNA_No54' | 'FVPAEMGTHTVSVK' | 'Filamin-A' |
| 10067 | 'FLNA_No55' | 'YGGQPVPNFPSK' | 'Filamin-A' |
| 10068 | 'FLNA_No56' | 'FNGTHIPGSPFK' | 'Filamin-A' |
| 10069 | 'FLNA_No57' | 'GKLDVQFSGLTK' | 'Filamin-A' |
| 10070 | 'FLNA_No58' | 'VDVGKDQEFTVK' | 'Filamin-A' |
| 10071 | 'FLNA_No59' | 'DAGEGLLAVQITDPEGKPK' | 'Filamin-A' |
| 10072 | 'FLNA_No60' | 'AYGPGIEPTGNMVK' | 'Filamin-A' |
| 10073 | 'FLNA_No61' | 'VDINTEDLEDGTCR' | 'Filamin-A' |
| 10074 | 'FLNA_No62' | 'DGSCGVAYVVQEPGDYEVSVK' | 'Filamin-A' |
| 10075 | 'FLNA_No63' | 'GAGTGGLGLAVEGPSEAK' | 'Filamin-A' |
| 10076 | 'FLNA_No64' | 'LQVEPAVDTSGVQCYGPGIEGQGVFR' | 'Filamin-A' |
| 10077 | 'FLNA_No65' | 'DAPQDFHPDRVK' | 'Filamin-A' |

| | A | B | C |
|---|---|---|---|
| 1 | **Peptide Name** | **Peptide Sequence** | **Peptide Description** |
| 10078 | 'FLNA_No66' | 'THEAEIVEGENHTYCIR' | 'Filamin-A' |
| 10079 | 'FLNA_No67' | 'VTYTPMAPGSYLISIK' | 'Filamin-A' |
| 10080 | 'FLNA_No68' | 'SPFEVYVDKSQGDASK' | 'Filamin-A' |
| 10081 | 'FLNA_No69' | 'QMQLENVSVALEFLDRESIK' | 'Filamin-A' |
| 10082 | 'FLNA_No70' | 'DAEMPATEKDLAEDAPWKK' | 'Filamin-A' |
| 10083 | 'FLNA_No71' | 'DAPQDFHPDR' | 'Filamin-A' |
| 10084 | 'FLNA_No72' | 'EGPYSISVLYGDEEVPRSPFK' | 'Filamin-A' |
| 10085 | 'FLNA_No73' | 'EAGAGGLAIAVEGPSKAEISFEDRK' | 'Filamin-A' |
| 10086 | 'FLNA_No74' | 'AHEPTYFTVDCAEAGQGDVSIGIK' | 'Filamin-A' |
| 10087 | 'FLNA_No75' | 'HTAMVSWGGVSIPNSPFR' | 'Filamin-A' |
| 10088 | 'FLNA_No76' | 'VGSAADIPINISETDLSLLTATVVPPSGREEPCLLKR' | 'Filamin-A' |
| 10089 | 'FLNA_No77' | 'AEAGVPAEFSIWTR' | 'Filamin-A' |
| 10090 | 'FLNA_No78' | 'EATTEFSVDAR' | 'Filamin-A' |
| 10091 | 'FLNA_No79' | 'VGSAADIPINISETDLSLLTATVVPPSGR' | 'Filamin-A' |
| 10092 | 'FLNA_No80' | 'YAPSEAGLHEMDIR' | 'Filamin-A' |
| 10093 | 'FLNA_No81' | 'AGGPGLERAEAGVPAEFSIWTR' | 'Filamin-A' |
| 10094 | 'FLNA_No82' | 'DAGEGGLSLAIEGPSK' | 'Filamin-A' |
| 10095 | 'FLNA_No83' | 'VSGLGEKVDVGKDQEFTVK' | 'Filamin-A' |
| 10096 | 'FLNA_No84' | 'FVPAEMGTHTVSVK' | 'Filamin-A' |
| 10097 | 'FLNA_No85' | 'LSPFMADIR' | 'Filamin-A' |
| 10098 | 'FLNA_No86' | 'RIANLQTDLSDGLR' | 'Filamin-A' |
| 10099 | 'FLNA_No87' | 'HTAMVSWGGVSIPNSPFR' | 'Filamin-A' |
| 10100 | 'FLNA_No88' | 'GDAVRDVDIIDHHDNTYTVK' | 'Filamin-A' |
| 10101 | 'FLNA_No89' | 'DNGNGTYSCSYVPR' | 'Filamin-A' |
| 10102 | 'FLNA_No90' | 'CAPGVVGPAEADIDFDIIR' | 'Filamin-A' |
| 10103 | 'FLNA_No91' | 'DAEMPATEKDLAEDAPWKK' | 'Filamin-A' |
| 10104 | 'FLNA_No92' | 'ALGALVDSCAPGLCPDWDSWDASKPVTNAR' | 'Filamin-A' |
| 10105 | 'FLNA_No93' | 'AHVVPCFDASKVK' | 'Filamin-A' |
| 10106 | 'FLNA_No94' | 'APSVANVGSHCDLSLK' | 'Filamin-A' |
| 10107 | 'FLNA_No95' | 'DLAEDAPWKK' | 'Filamin-A' |
| 10108 | 'FLNA_No96' | 'CSGPGLSPGMVR' | 'Filamin-A' |
| 10109 | 'FLNA_No97' | 'IVGPSGAAVPCK' | 'Filamin-A' |
| 10110 | 'FLNA_No98' | 'NGHVGISFVPK' | 'Filamin-A' |
| 10111 | 'FLNA_No99' | 'LSPFMADIR' | 'Filamin-A' |
| 10112 | 'FLNA_No100' | 'SPFEVKVGTECGNQK' | 'Filamin-A' |
| 10113 | 'FLNA_No101' | 'VHSPSGALEECYVTEIDQDK' | 'Filamin-A' |
| 10114 | 'FLNA_No102' | 'SAGQGEVLVYVEDPAGHQEEAKVTANNDKNR' | 'Filamin-A' |
| 10115 | 'FLNA_No103' | 'ANLPQSFQVDTSKAGVAPLQVK' | 'Filamin-A' |
| 10116 | 'FLNA_No104' | 'ALTQTGGPHVK' | 'Filamin-A' |
| 10117 | 'FLNA_No105' | 'IPEISIQDMTAQVTSPSGK' | 'Filamin-A' |
| 10118 | 'FLNA_No106' | 'EAGAGGLAIAVEGPSKAEISFEDR' | 'Filamin-A' |
| 10119 | 'FLNA_No107' | 'QMQLENVSVALEFLDRESIK' | 'Filamin-A' |
| 10120 | 'FLNA_No108' | 'THIQDNHDGTYTVAYVPDVTGR' | 'Filamin-A' |
| 10121 | 'FLNA_No109' | 'VKETADFKVYTK' | 'Filamin-A' |

| | A | B | C |
|---|---|---|---|
| 1 | **Peptide Name** | **Peptide Sequence** | **Peptide Description** |
| 10122 | 'FLNA_No110' | 'AGNNMLLVGVHGPR' | 'Filamin-A' |
| 10123 | 'FLNA_No111' | 'VSGQGLHEGHTFEPAEFIIDTR' | 'Filamin-A' |
| 10124 | 'FLNA_No112' | 'VPVHDVTDASKVK' | 'Filamin-A' |
| 10125 | 'FLNA_No113' | 'ENGVYLIDVK' | 'Filamin-A' |
| 10126 | 'FLNA_No114' | 'VNVGAGSHPNK' | 'Filamin-A' |
| 10127 | 'FLNA_No115' | 'EEGPYEVEVTYDGVPVPGSPFPLEAVAPTKPSK' | 'Filamin-A' |
| 10128 | 'ACO1_No1' | 'YQQAGLPLIVLAGK' | 'Cytoplasmic aconitate hydratase' |
| 10129 | 'ACO1_No2' | 'GPFLLGIK' | 'Cytoplasmic aconitate hydratase' |
| 10130 | 'ACO1_No3' | 'QAPQTIHLPSGEILDVFDAAER' | 'Cytoplasmic aconitate hydratase' |
| 10131 | 'CD9_No1' | 'KDVLETFTVK' | 'CD9 antigen' |
| 10132 | 'CD9_No2' | 'EVQEFYKDTYNK' | 'CD9 antigen' |
| 10133 | 'PCMT1' | 'LILPVGPAGGNQMLEQYDKLQDGSIK' | 'Protein-L-isoaspartate(D-aspartate) O-methyltransferase' |
| 10134 | 'TNXB_No1' | 'FLLYGLLGGK' | 'Tenascin-X' |
| 10135 | 'TNXB_No2' | 'TLSPVLESPR' | 'Tenascin-X' |
| 10136 | 'TNXB_No3' | 'SGTLYSLTLYGLR' | 'Tenascin-X' |
| 10137 | 'TNXB_No4' | 'VSYQLADGGEPQSVQVDGQAR' | 'Tenascin-X' |
| 10138 | 'TNXB_No5' | 'FLLYGLSGR' | 'Tenascin-X' |
| 10139 | 'TNXB_No6' | 'LGPLSAEGTTGLAPAGQTSEESRPR' | 'Tenascin-X' |
| 10140 | 'TNXB_No7' | 'LGPISADSTTAPLEK' | 'Tenascin-X' |
| 10141 | 'TNXB_No8' | 'GFEFSVPFTEMK' | 'Tenascin-X' |
| 10142 | 'TNXB_No9' | 'NCHYANLNGLYGSTVDHQGVSWYHWK' | 'Tenascin-X' |
| 10143 | 'TNXB_No10' | 'FLLYGLHEGKR' | 'Tenascin-X' |
| 10144 | 'TNXB_No11' | 'VRGEESEVTVGGLEPGRK' | 'Tenascin-X' |
| 10145 | 'TNXB_No12' | 'GFEESEPLTGFLTTVPDGPTQLR' | 'Tenascin-X' |
| 10146 | 'TNXB_No13' | 'LGVLTVTDTTPDSMR' | 'Tenascin-X' |
| 10147 | 'TNXB_No14' | 'LGVLTVTDTTPDSMR' | 'Tenascin-X' |
| 10148 | 'TNXB_No15' | 'FLLFGIQDGKR' | 'Tenascin-X' |
| 10149 | 'TNXB_No16' | 'EVTVQGLEPSRK' | 'Tenascin-X' |
| 10150 | 'TNXB_No17' | 'LSQLSVTDVTTSSLR' | 'Tenascin-X' |
| 10151 | 'TNXB_No18' | 'AGDEAVFAQYDSFHVDSAAEYYR' | 'Tenascin-X' |
| 10152 | 'TNXB_No19' | 'ILISGLEPSTPYR' | 'Tenascin-X' |
| 10153 | 'TNXB_No20' | 'YEVTVVSVR' | 'Tenascin-X' |
| 10154 | 'TNXB_No21' | 'LHLEGYHGTAGDSMSYHSGSVFSAR' | 'Tenascin-X' |
| 10155 | 'TNXB_No22' | 'MHLYGLHEGQR' | 'Tenascin-X' |
| 10156 | 'TNXB_No23' | 'LQGLIPGAR' | 'Tenascin-X' |
| 10157 | 'TNXB_No24' | 'AVAVSGLDPAR' | 'Tenascin-X' |
| 10158 | 'TNXB_No25' | 'DLQFSEIR' | 'Tenascin-X' |
| 10159 | 'TNXB_No26' | 'TLCSLHGVFDLSR' | 'Tenascin-X' |
| 10160 | 'TNXB_No27' | 'FLLYGLLGGKR' | 'Tenascin-X' |
| 10161 | 'TNXB_No28' | 'FLLFGIQDGK' | 'Tenascin-X' |
| 10162 | 'TNXB_No29' | 'ELPPHLGELTVAEETSSSLR' | 'Tenascin-X' |
| 10163 | 'TNXB_No30' | 'AVRVPGHEDGVTISGLEPDHKYK' | 'Tenascin-X' |
| 10164 | 'TNXB_No31' | 'VGPVSAVGVTAPEEESPDAPLAK' | 'Tenascin-X' |
| 10165 | 'TNXB_No32' | 'FLLYGLHEGK' | 'Tenascin-X' |

| | A | B | C |
|---|---|---|---|
| 1 | **Peptide Name** | **Peptide Sequence** | **Peptide Description** |
| 10166 | 'TNXB_No33' | 'GEESEVTVGGLEPGRK' | 'Tenascin-X' |
| 10167 | 'TNXB_No34' | 'LQAMWGQSLLPPVSTSFTTGGLR' | 'Tenascin-X' |
| 10168 | 'TNXB_No35' | 'DRDPNSLLISCAVSYR' | 'Tenascin-X' |
| 10169 | 'TNXB_No36' | 'GPNLTSPASITFTTGLEAPR' | 'Tenascin-X' |
| 10170 | 'TNXB_No37' | 'FLLYGLLGK' | 'Tenascin-X' |
| 10171 | 'TNXB_No38' | 'LHLEGYHGTAGDSMSYHSGSVFSAR' | 'Tenascin-X' |
| 10172 | 'TNXB_No39' | 'MNLYGFHGGQR' | 'Tenascin-X' |
| 10173 | 'TNXB_No40' | 'GFEFSVPFTEMK' | 'Tenascin-X' |
| 10174 | 'TNXB_No41' | 'DCGEEMQNGAGASR' | 'Tenascin-X' |
| 10175 | 'TNXB_No42' | 'DGYAGEDCGEEVPTIEGMR' | 'Tenascin-X' |
| 10176 | 'UBA1_No1' | 'YFLVGAGAIGCELLK' | 'Ubiquitin-like modifier-activating enzyme 1' |
| 10177 | 'UBA1_No2' | 'AAVATFLQSVQVPEFTPK' | 'Ubiquitin-like modifier-activating enzyme 1' |
| 10178 | 'UBA1_No3' | 'NEEDAAELVALAQAVNAR' | 'Ubiquitin-like modifier-activating enzyme 1' |
| 10179 | 'UBA1_No4' | 'ATLPSPDKLPGFK' | 'Ubiquitin-like modifier-activating enzyme 1' |
| 10180 | 'UBA1_No5' | 'IHVSDQELQSANASVDDSRLEELK' | 'Ubiquitin-like modifier-activating enzyme 1' |
| 10181 | 'UBA1_No6' | 'AENYDIPSADR' | 'Ubiquitin-like modifier-activating enzyme 1' |
| 10182 | 'UBA1_No7' | 'DEFEGLFK' | 'Ubiquitin-like modifier-activating enzyme 1' |
| 10183 | 'UBA1_No8' | 'ALPAVQQNNLDEDLIRK' | 'Ubiquitin-like modifier-activating enzyme 1' |
| 10184 | 'GPX3_No1' | 'YVRPGGGFVPNFQLFEK' | 'Glutathione peroxidase 3' |
| 10185 | 'GPX3_No2' | 'QEPGENSEILPTLK' | 'Glutathione peroxidase 3' |
| 10186 | 'GPX3_No3' | 'NSCPPTSELLGTSDR' | 'Glutathione peroxidase 3' |
| 10187 | 'GPX3_No4' | 'PGGGFVPNFQLFEK' | 'Glutathione peroxidase 3' |
| 10188 | 'GPX3_No5' | 'MDILSYMR' | 'Glutathione peroxidase 3' |
| 10189 | 'GPX3_No6' | 'FLVGPDGIPIMR' | 'Glutathione peroxidase 3' |
| 10190 | 'GPX3_No7' | 'LFWEPMK' | 'Glutathione peroxidase 3' |
| 10191 | 'GPX3_No8' | 'FLVGPDGIPIMR' | 'Glutathione peroxidase 3' |
| 10192 | 'GPX3_No9' | 'MDILSYMR' | 'Glutathione peroxidase 3' |
| 10193 | 'GPX3_No10' | 'LFWEPMKVHDIR' | 'Glutathione peroxidase 3' |
| 10194 | 'GPX3_No11' | 'QEPGENSEILPTLK' | 'Glutathione peroxidase 3' |
| 10195 | 'GPX3_No12' | 'MDILSYMR' | 'Glutathione peroxidase 3' |
| 10196 | 'GPX3_No13' | 'LFWEPMKVHDIR' | 'Glutathione peroxidase 3' |
| 10197 | 'NME2_No1' | 'TFIAIKPDGVQR' | 'Nucleoside diphosphate kinase B' |
| 10198 | 'NME2_No2' | 'VMLGETNPADSKPGTIR' | 'Nucleoside diphosphate kinase B' |
| 10199 | 'NME2_No3' | 'GLVGEIIKR' | 'Nucleoside diphosphate kinase B' |
| 10200 | 'NME2_No4' | 'VMLGETNPADSKPGTIR' | 'Nucleoside diphosphate kinase B' |
| 10201 | 'NME2_No5' | 'DRPFFPGLVK' | 'Nucleoside diphosphate kinase B' |
| 10202 | 'NME2_No6' | 'GDFCIQVGR' | 'Nucleoside diphosphate kinase B' |
| 10203 | 'IGFBP4_No1' | 'EDARPVPQGSCQSELHR' | 'Insulin-like growth factor-binding protein 4' |
| 10204 | 'IGFBP4_No2' | 'TGVKLPGGLEPK' | 'Insulin-like growth factor-binding protein 4' |
| 10205 | 'IGFBP4_No3' | 'GELDCHQLADSFRE' | 'Insulin-like growth factor-binding protein 4' |
| 10206 | 'IGFBP4_No4' | 'THEDLYIIPIPNCDR' | 'Insulin-like growth factor-binding protein 4' |
| 10207 | 'IGFBP4_No5' | 'LPGGLEPK' | 'Insulin-like growth factor-binding protein 4' |
| 10208 | 'IGFBP4_No6' | 'CRPPVGCEELVR' | 'Insulin-like growth factor-binding protein 4' |
| 10209 | 'PRKACB' | 'ILQAVNFPFLVR' | 'cAMP-dependent protein kinase catalytic subunit beta' |

| | A | B | C |
|---|---|---|---|
| 1 | **Peptide Name** | **Peptide Sequence** | **Peptide Description** |
| 10210 | 'CPN2_No1' | 'AGGSWDLAVQER' | 'Carboxypeptidase N subunit 2' |
| 10211 | 'CPN2_No2' | 'LLNIQTYCAGPAYLK' | 'Carboxypeptidase N subunit 2' |
| 10212 | 'CPN2_No3' | 'NIIFVETSFTTLETR' | 'Carboxypeptidase N subunit 2' |
| 10213 | 'CPN2_No4' | 'RLFQPLTHLK' | 'Carboxypeptidase N subunit 2' |
| 10214 | 'CPN2_No5' | 'SLMLSYNAITHLPAGIFR' | 'Carboxypeptidase N subunit 2' |
| 10215 | 'CPN2_No6' | 'TLNLAQNLLAQLPEELFHPLTSLQTLK' | 'Carboxypeptidase N subunit 2' |
| 10216 | 'CPN2_No7' | 'QLVCPVTR' | 'Carboxypeptidase N subunit 2' |
| 10217 | 'CPN2_No8' | 'DHLGFQVTWPDESK' | 'Carboxypeptidase N subunit 2' |
| 10218 | 'CPN2_No9' | 'VLPAGLFAHTPCLVGLSLTHNQLETVAEGTFAHLSNLR' | 'Carboxypeptidase N subunit 2' |
| 10219 | 'CPN2_No10' | 'SLMLSYNAITHLPAGIFR' | 'Carboxypeptidase N subunit 2' |
| 10220 | 'CPN2_No11' | 'LTVSIEAR' | 'Carboxypeptidase N subunit 2' |
| 10221 | 'CPN2_No12' | 'VVFLNTQLCQFRPDAFGGLPR' | 'Carboxypeptidase N subunit 2' |
| 10222 | 'CPN2_No13' | 'LFQPLTHLK' | 'Carboxypeptidase N subunit 2' |
| 10223 | 'CPN2_No14' | 'LSNNALSGLPQGVFGK' | 'Carboxypeptidase N subunit 2' |
| 10224 | 'CPN2_No15' | 'VVFLNTQLCQFR' | 'Carboxypeptidase N subunit 2' |
| 10225 | 'CPN2_No16' | 'SQCTYSNPEGTVVLACDQAQCR' | 'Carboxypeptidase N subunit 2' |
| 10226 | 'CPN2_No17' | 'LELLSLSK' | 'Carboxypeptidase N subunit 2' |
| 10227 | 'PROZ_No1' | 'APDLQDLPWQVK' | 'Vitamin K-dependent protein Z' |
| 10228 | 'PROZ_No2' | 'TDGCQHFCLPGQESYTCSCAQGYR' | 'Vitamin K-dependent protein Z' |
| 10229 | 'PROZ_No3' | 'SSVAAMHWMDGSVVTR' | 'Vitamin K-dependent protein Z' |
| 10230 | 'PROZ_No4' | 'LGEDHKQCVPHDQCACGVLTSEKR' | 'Vitamin K-dependent protein Z' |
| 10231 | 'PROZ_No5' | 'LTNSEGKDFCGGVIIR' | 'Vitamin K-dependent protein Z' |
| 10232 | 'PROZ_No6' | 'QCVPHDQCACGVLTSEK' | 'Vitamin K-dependent protein Z' |
| 10233 | 'PROZ_No7' | 'RAPDLQDLPWQVK' | 'Vitamin K-dependent protein Z' |
| 10234 | 'PROZ_No8' | 'LGEDHKQCVPHDQCACGVLTSEK' | 'Vitamin K-dependent protein Z' |
| 10235 | 'PROZ_No9' | 'QCVPHDQCACGVLTSEK' | 'Vitamin K-dependent protein Z' |
| 10236 | 'PROZ_No10' | 'DFAEHLLIPR' | 'Vitamin K-dependent protein Z' |
| 10237 | 'PROZ_No11' | 'DFCGGVIIR' | 'Vitamin K-dependent protein Z' |
| 10238 | 'PROZ_No12' | 'GLLSGWAR' | 'Vitamin K-dependent protein Z' |
| 10239 | 'PROZ_No13' | 'GSWFLTGVLGSQPVGGQAHMVLVTK' | 'Vitamin K-dependent protein Z' |
| 10240 | 'PROZ_No14' | 'SSVAAMHWMDGSVVTR' | 'Vitamin K-dependent protein Z' |
| 10241 | 'PROZ_No15' | 'SSVAAMHWMDGSVVTR' | 'Vitamin K-dependent protein Z' |
| 10242 | 'PROZ_No16' | 'QCVPHDQCACGVLTSEKR' | 'Vitamin K-dependent protein Z' |
| 10243 | 'PROZ_No17' | 'TSQDPLMIK' | 'Vitamin K-dependent protein Z' |
| 10244 | 'MRC1_No1' | 'ACIGFGGNLVSIQNEK' | 'Macrophage mannose receptor 1' |
| 10245 | 'MRC1_No2' | 'TGIAGGLWDVLK' | 'Macrophage mannose receptor 1' |
| 10246 | 'MRC1_No3' | 'WYADCTSAGR' | 'Macrophage mannose receptor 1' |
| 10247 | 'MRC1_No4' | 'FEGSESLWNKDPLTSVSYQINSK' | 'Macrophage mannose receptor 1' |
| 10248 | 'MRC1_No5' | 'IYGTTDNLCSR' | 'Macrophage mannose receptor 1' |
| 10249 | 'MRC1_No6' | 'MGSSLVSIESAAESSFLSYR' | 'Macrophage mannose receptor 1' |
| 10250 | 'MRC1_No7' | 'TNFWIGLFR' | 'Macrophage mannose receptor 1' |
| 10251 | 'MRC1_No8' | 'NFGDLVSIQSESEKK' | 'Macrophage mannose receptor 1' |
| 10252 | 'MRC1_No9' | 'DSTFSAWTGLNDVNSEHTFLWTDGR' | 'Macrophage mannose receptor 1' |
| 10253 | 'MRC1_No10' | 'GYEAMYTLLGNANGATCAFPFK' | 'Macrophage mannose receptor 1' |

| | A | B | C |
|---|---|---|---|
| 1 | **Peptide Name** | **Peptide Sequence** | **Peptide Description** |
| 10254 | 'MRC1_No11' | 'GEPSHENNRQEDCVVMK' | 'Macrophage mannose receptor 1' |
| 10255 | 'MRC1_No12' | 'QKFQWHEAETYCK' | 'Macrophage mannose receptor 1' |
| 10256 | 'MRC1_No13' | 'YEQAFLTSFVGLRPEK' | 'Macrophage mannose receptor 1' |
| 10257 | 'MRC1_No14' | 'NWGQASLECLR' | 'Macrophage mannose receptor 1' |
| 10258 | 'MRC1_No15' | 'FQWHEAETYCK' | 'Macrophage mannose receptor 1' |
| 10259 | 'MRC1_No16' | 'YFWTGLSDIQTK' | 'Macrophage mannose receptor 1' |
| 10260 | 'MRC1_No17' | 'SQGPEIVEVEK' | 'Macrophage mannose receptor 1' |
| 10261 | 'MRC1_No18' | 'ALGGDLASINNKEEQQTIWR' | 'Macrophage mannose receptor 1' |
| 10262 | 'MRC1_No19' | 'FTHWNSDMPGR' | 'Macrophage mannose receptor 1' |
| 10263 | 'MRC1_No20' | 'SSLSYEDADCVVIIGGASNEAGK' | 'Macrophage mannose receptor 1' |
| 10264 | 'MRC1_No21' | 'IFGFMEEERK' | 'Macrophage mannose receptor 1' |
| 10265 | 'MRC1_No22' | 'WVSESQIMSVAFK' | 'Macrophage mannose receptor 1' |
| 10266 | 'MRC1_No23' | 'GEPSHENNRQEDCVVMK' | 'Macrophage mannose receptor 1' |
| 10267 | 'IGHV1-2_No1' | 'ASGYTFTGYYMHWVR' | 'Immunoglobulin heavy variable 1-2' |
| 10268 | 'IGHV1-2_No2' | 'SDDTAVYYCAR' | 'Immunoglobulin heavy variable 1-2' |
| 10269 | 'IGHV1-2_No3' | 'LRSDDTAVYYCAR' | 'Immunoglobulin heavy variable 1-2' |
| 10270 | 'IGHV1-2_No4' | 'DTSISTAYMELSR' | 'Immunoglobulin heavy variable 1-2' |
| 10271 | 'IGHV1-2_No5' | 'QAPGQGLEWMGWINPNSGGTNYAQK' | 'Immunoglobulin heavy variable 1-2' |
| 10272 | 'IGHV1-2_No6' | 'FQGWVTMTR' | 'Immunoglobulin heavy variable 1-2' |
| 10273 | 'IGHV1-2_No7' | 'QAPGQGLEWMGWINPNSGGTNYAQK' | 'Immunoglobulin heavy variable 1-2' |
| 10274 | 'IGHV1-2_No8' | 'DTSISTAYMELSR' | 'Immunoglobulin heavy variable 1-2' |
| 10275 | 'IGHV1-2_No9' | 'FQGWVTMTR' | 'Immunoglobulin heavy variable 1-2' |
| 10276 | 'IGHV1-2_No10' | 'ASGYTFTGYYMHWVR' | 'Immunoglobulin heavy variable 1-2' |
| 10277 | 'IGHV1-2_No11' | 'ILFLVAAATGAHSQVQLVQSGAEVK' | 'Immunoglobulin heavy variable 1-2' |
| 10278 | 'CES1_No1' | 'LKDKEVAFWTNLFAK' | 'Liver carboxylesterase 1' |
| 10279 | 'CES1_No2' | 'TTTSAVMVHCLR' | 'Liver carboxylesterase 1' |
| 10280 | 'CES1_No3' | 'EGYLQIGANTQAAQK' | 'Liver carboxylesterase 1' |
| 10281 | 'CES1_No4' | 'LSEDCLYLNIYTPADLTK' | 'Liver carboxylesterase 1' |
| 10282 | 'CES1_No5' | 'QKTEEELLETTLK' | 'Liver carboxylesterase 1' |
| 10283 | 'CES1_No6' | 'NFHTVPYMVGINK' | 'Liver carboxylesterase 1' |
| 10284 | 'CES1_No7' | 'GNWGHLDQVAALR' | 'Liver carboxylesterase 1' |
| 10285 | 'FBLN1_No1' | 'AITPPHPASQANIIFDITEGNLR' | 'Fibulin-1' |
| 10286 | 'FBLN1_No2' | 'TGYYFDGISR' | 'Fibulin-1' |
| 10287 | 'FBLN1_No3' | 'DSFDIIKR' | 'Fibulin-1' |
| 10288 | 'FBLN1_No4' | 'IIEVEEEQEDPYLNDR' | 'Fibulin-1' |
| 10289 | 'FBLN1_No5' | 'SQETGDLDVGGLQETDK' | 'Fibulin-1' |
| 10290 | 'FBLN1_No6' | 'EFTRPEEIIFLR' | 'Fibulin-1' |
| 10291 | 'FBLN1_No7' | 'MCVDVNECQR' | 'Fibulin-1' |
| 10292 | 'FBLN1_No8' | 'DSSCGTGYELTEDNSCK' | 'Fibulin-1' |
| 10293 | 'FBLN1_No9' | 'YMDGMTVGVVR' | 'Fibulin-1' |
| 10294 | 'FBLN1_No10' | 'DCSLPYATESK' | 'Fibulin-1' |
| 10295 | 'FBLN1_No11' | 'LGESCINTVGSFR' | 'Fibulin-1' |
| 10296 | 'FBLN1_No12' | 'CVDVDECAPPAEPCGK' | 'Fibulin-1' |
| 10297 | 'FBLN1_No13' | 'LEMNYVVGGVVSHR' | 'Fibulin-1' |

| | A | B | C |
|---|---|---|---|
| 1 | **Peptide Name** | **Peptide Sequence** | **Peptide Description** |
| 10298 | 'FBLN1_No14' | 'QVRPIVGPFHAVLK' | 'Fibulin-1' |
| 10299 | 'FBLN1_No15' | 'DCSLPYATESKECR' | 'Fibulin-1' |
| 10300 | 'FBLN1_No16' | 'DSFDIIK' | 'Fibulin-1' |
| 10301 | 'FBLN1_No17' | 'YMDGMTVGVVR' | 'Fibulin-1' |
| 10302 | 'FBLN1_No18' | 'LEMNYVVGGVVSHR' | 'Fibulin-1' |
| 10303 | 'FBLN1_No19' | 'YMDGMTVGVVR' | 'Fibulin-1' |
| 10304 | 'FBLN1_No20' | 'YMDGMTVGVVR' | 'Fibulin-1' |
| 10305 | 'FBLN1_No21' | 'SCRPNDVTCVFDPVHTISHTVISLPTFR' | 'Fibulin-1' |
| 10306 | 'FBLN1_No22' | 'QVRPIVGPFHAVLK' | 'Fibulin-1' |
| 10307 | 'FBLN1_No23' | 'PEEIIFLR' | 'Fibulin-1' |
| 10308 | 'FBLN1_No24' | 'CLAFECPENYRR' | 'Fibulin-1' |
| 10309 | 'FBLN1_No25' | 'RGYQLSDVDGVTCEDIDECALPTGGHICSYR' | 'Fibulin-1' |
| 10310 | 'FBLN1_No26' | 'MVQEQCCHSQLEELHCATGISLANEQDR' | 'Fibulin-1' |
| 10311 | 'FBLN1_No27' | 'SQETGDLDVGGLQETDKIIEVEEEQEDPYLNDRCR' | 'Fibulin-1' |
| 10312 | 'FBLN1_No28' | 'GYHLNEEGTRCVDVDECAPPAEPCGK' | 'Fibulin-1' |
| 10313 | 'FBLN1_No29' | 'SAATLQQEKTDTVR' | 'Fibulin-1' |
| 10314 | 'FBLN1_No30' | 'CCHCCLLGR' | 'Fibulin-1' |
| 10315 | 'FBLN1_No31' | 'MVQEQCCHSQLEELHCATGISLANEQDR' | 'Fibulin-1' |
| 10316 | 'FBLN1_No32' | 'MCVDVNECQRYPGR' | 'Fibulin-1' |
| 10317 | 'FBLN1_No33' | 'GYHLNEEGTR' | 'Fibulin-1' |
| 10318 | 'FBLN1_No34' | 'MATHQKDCSLPYATESKECR' | 'Fibulin-1' |
| 10319 | 'FBLN1_No35' | 'MCVDVNECQR' | 'Fibulin-1' |
| 10320 | 'FBLN1_No36' | 'MCVDVNECQRYPGR' | 'Fibulin-1' |
| 10321 | 'FBLN1_No37' | 'MATHQKDCSLPYATESK' | 'Fibulin-1' |
| 10322 | 'FBLN1_No38' | 'MATHQKDCSLPYATESKECR' | 'Fibulin-1' |
| 10323 | 'FBLN1_No39' | 'CVNSPGSFR' | 'Fibulin-1' |
| 10324 | 'FBLN1_No40' | 'DIDECESGIHNCLPDFICQNTLGSFR' | 'Fibulin-1' |
| 10325 | 'FBLN1_No41' | 'MATHQKDCSLPYATESK' | 'Fibulin-1' |
| 10326 | 'FBLN1_No42' | 'SQETGDLDVGGLQETDKIIEVEEEQEDPYLNDR' | 'Fibulin-1' |
| 10327 | 'FBLN1_No43' | 'NVPNCGR' | 'Fibulin-1' |
| 10328 | 'FBLN1_No44' | 'IIEVEEEQEDPYLNDRCR' | 'Fibulin-1' |
| 10329 | 'FBLN1_No45' | 'PIVGPFHAVLK' | 'Fibulin-1' |
| 10330 | 'FBLN1_No46' | 'CECKTGYYFDGISR' | 'Fibulin-1' |
| 10331 | 'ITGA6_No1' | 'LIATFPDTLTYSAYR' | 'Integrin alpha-6' |
| 10332 | 'ITGA6_No2' | 'LNYLDILMR' | 'Integrin alpha-6' |
| 10333 | 'ITGA6_No3' | 'FGSCQQGVAATFTK' | 'Integrin alpha-6' |
| 10334 | 'ITGA6_No4' | 'EGCGDDNVCNSNLK' | 'Integrin alpha-6' |
| 10335 | 'ITGA6_No5' | 'NSYPDVAVGSLSDSVTIFR' | 'Integrin alpha-6' |
| 10336 | 'ITGA6_No6' | 'LETTSNQDNLAPITAK' | 'Integrin alpha-6' |
| 10337 | 'PPIB_No1' | 'VIFGLFGK' | 'Peptidyl-prolyl cis-trans isomerase B' |
| 10338 | 'PPIB_No2' | 'TVDNFVALATGEK' | 'Peptidyl-prolyl cis-trans isomerase B' |
| 10339 | 'PPIB_No3' | 'HYGPGWVSMANAGK' | 'Peptidyl-prolyl cis-trans isomerase B' |
| 10340 | 'PPIB_No4' | 'DKPLKDVIIADCGK' | 'Peptidyl-prolyl cis-trans isomerase B' |
| 10341 | 'PPIB_No5' | 'VIKDFMIQGGDFTR' | 'Peptidyl-prolyl cis-trans isomerase B' |

| | A | B | C |
|---|---|---|---|
| 1 | **Peptide Name** | **Peptide Sequence** | **Peptide Description** |
| 10342 | 'PPIB_No6' | 'VIKDFMIQGGDFTR' | 'Peptidyl-prolyl cis-trans isomerase B' |
| 10343 | 'PPIB_No7' | 'HYGPGWVSMANAGK' | 'Peptidyl-prolyl cis-trans isomerase B' |
| 10344 | 'PPIB_No8' | 'IEVEKPFAIAKE' | 'Peptidyl-prolyl cis-trans isomerase B' |
| 10345 | 'PTPRG_No1' | 'ETFLDPFVLR' | 'Receptor-type tyrosine-protein phosphatase gamma' |
| 10346 | 'PTPRG_No2' | 'DLLPASLGSYYR' | 'Receptor-type tyrosine-protein phosphatase gamma' |
| 10347 | 'PTPRG_No3' | 'DNSALDPIIHGLK' | 'Receptor-type tyrosine-protein phosphatase gamma' |
| 10348 | 'PTPRG_No4' | 'ATISHVSPDSLYLFR' | 'Receptor-type tyrosine-protein phosphatase gamma' |
| 10349 | 'PTPRG_No5' | 'TVAILLKDDYFVSGAGLPGR' | 'Receptor-type tyrosine-protein phosphatase gamma' |
| 10350 | 'PTPRG_No6' | 'ASGDPYWAYSGAYGPEHWVTSSVSCGGR' | 'Receptor-type tyrosine-protein phosphatase gamma' |
| 10351 | 'PTPRG_No7' | 'HQSPIDILDQYAR' | 'Receptor-type tyrosine-protein phosphatase gamma' |
| 10352 | 'PTPRG_No8' | 'GVVHHEKETFLDPFVLR' | 'Receptor-type tyrosine-protein phosphatase gamma' |
| 10353 | 'CFL1_No1' | 'LGGSAVISLEGKPL' | 'Cofilin-1' |
| 10354 | 'CFL1_No2' | 'NIILEEGKEILVGDVGQTVDDPYATFVK' | 'Cofilin-1' |
| 10355 | 'CFL1_No3' | 'HELQANCYEEVKDR' | 'Cofilin-1' |
| 10356 | 'CFL1_No4' | 'ESKKEDLVFIFWAPESAPLK' | 'Cofilin-1' |
| 10357 | 'CFL1_No5' | 'LTGIKHELQANCYEEVKDR' | 'Cofilin-1' |
| 10358 | 'CFL1_No6' | 'AVLFCLSEDKK' | 'Cofilin-1' |
| 10359 | 'CFL1_No7' | 'EILVGDVGQTVDDPYATFVK' | 'Cofilin-1' |
| 10360 | 'CFL1_No8' | 'ASGVAVSDGVIK' | 'Cofilin-1' |
| 10361 | 'CFL1_No9' | 'YALYDATYETK' | 'Cofilin-1' |
| 10362 | 'CFL1_No10' | 'ASGVAVSDGVIKVFNDMK' | 'Cofilin-1' |
| 10363 | 'CFL1_No11' | 'ASGVAVSDGVIKVFNDMK' | 'Cofilin-1' |
| 10364 | 'CFL1_No12' | 'LTGIKHELQANCYEEVK' | 'Cofilin-1' |
| 10365 | 'CFL1_No13' | 'HELQANCYEEVK' | 'Cofilin-1' |
| 10366 | 'LAMA2_No1' | 'LVEHVPGQPVR' | 'Laminin subunit alpha-2' |
| 10367 | 'LAMA2_No2' | 'AEECYYDENVAR' | 'Laminin subunit alpha-2' |
| 10368 | 'LAMA2_No3' | 'VAPQQDDLDSPQQISISNAEAR' | 'Laminin subunit alpha-2' |
| 10369 | 'LAMA2_No4' | 'PGNWILER' | 'Laminin subunit alpha-2' |
| 10370 | 'LAMA2_No5' | 'AEQTILPLVDEALQHTTTK' | 'Laminin subunit alpha-2' |
| 10371 | 'LAMA2_No6' | 'LVEHVPGQPVRNPQCR' | 'Laminin subunit alpha-2' |
| 10372 | 'PRTN3_No1' | 'VALYVDWIR' | 'Myeloblastin' |
| 10373 | 'PRTN3_No2' | 'LVNVVLGAHNVR' | 'Myeloblastin' |
| 10374 | 'PRTN3_No3' | 'LFPDFFTR' | 'Myeloblastin' |
| 10375 | 'PRTN3_No4' | 'TQEPTQQHFSVAQVFLNNYDAENK' | 'Myeloblastin' |
| 10376 | 'GPT' | 'LFLLADEVYQDNVYAAGSQFHSFKK' | 'Alanine aminotransferase 1' |
| 10377 | 'TBXAS1_No1' | 'DVFSSTGCKPNPSR' | 'Thromboxane-A synthase' |
| 10378 | 'TBXAS1_No2' | 'SVADSVLFLR' | 'Thromboxane-A synthase' |
| 10379 | 'TBXAS1_No3' | 'HSASPMGVQDFDIVR' | 'Thromboxane-A synthase' |
| 10380 | 'TBXAS1_No4' | 'YAESGDAFDIQR' | 'Thromboxane-A synthase' |
| 10381 | 'TBXAS1_No5' | 'QGFWESQMELR' | 'Thromboxane-A synthase' |
| 10382 | 'IGFBP6_No1' | 'HLDSVLQQLQTEVYR' | 'Insulin-like growth factor-binding protein 6' |
| 10383 | 'IGFBP6_No2' | 'GAQTLYVPNCDHR' | 'Insulin-like growth factor-binding protein 6' |
| 10384 | 'IGFBP6_No3' | 'EGQECGVYTPNCAPGLQCHPPKDDEAPLR' | 'Insulin-like growth factor-binding protein 6' |
| 10385 | 'IGFBP6_No4' | 'RGPCWCVDR' | 'Insulin-like growth factor-binding protein 6' |

| | A | B | C |
|---|---|---|---|
| 1 | **Peptide Name** | **Peptide Sequence** | **Peptide Description** |
| 10386 | 'IGFBP5_No1' | 'HMEASLQELK' | 'Insulin-like growth factor-binding protein 5' |
| 10387 | 'IGFBP5_No2' | 'AVYLPNCDR' | 'Insulin-like growth factor-binding protein 5' |
| 10388 | 'IGFBP5_No3' | 'ALSMCPPSPLGCELVK' | 'Insulin-like growth factor-binding protein 5' |
| 10389 | 'IGFBP5_No4' | 'QDEEKPLHALLHGR' | 'Insulin-like growth factor-binding protein 5' |
| 10390 | 'IGFBP5_No5' | 'HMEASLQELK' | 'Insulin-like growth factor-binding protein 5' |
| 10391 | 'ACP1_No1' | 'SPIAEAVFR' | 'Low molecular weight phosphotyrosine protein phosphatase' |
| 10392 | 'ACP1_No2' | 'SVLFVCLGNICR' | 'Low molecular weight phosphotyrosine protein phosphatase' |
| 10393 | 'TNC_No1' | 'WQPAIATVDSYVISYTGEK' | 'Tenascin' |
| 10394 | 'TNC_No2' | 'YAPISGGDHAEVDVPK' | 'Tenascin' |
| 10395 | 'TNC_No3' | 'WQPAIATVDSYVISYTGEKVPEITR' | 'Tenascin' |
| 10396 | 'TNC_No4' | 'EQCTAGAGCCLQPATGR' | 'Tenascin' |
| 10397 | 'TNC_No5' | 'LDAPSQIEVK' | 'Tenascin' |
| 10398 | 'TNC_No6' | 'REEFWLGLDNLNK' | 'Tenascin' |
| 10399 | 'TNC_No7' | 'ITAQGQYELR' | 'Tenascin' |
| 10400 | 'TNC_No8' | 'GYDCSDMSCPNDCHQHGR' | 'Tenascin' |
| 10401 | 'TNC_No9' | 'EVIVGPDTTSYSLADLSPSTHYTAK' | 'Tenascin' |
| 10402 | 'TNC_No10' | 'GLEPGQEYNVLLTAEK' | 'Tenascin' |
| 10403 | 'TNC_No11' | 'ITYVPITGGTPSMVTVDGTK' | 'Tenascin' |
| 10404 | 'TNC_No12' | 'DLTATEVQSETALLTWRPPR' | 'Tenascin' |
| 10405 | 'TNC_No13' | 'LEELENLVSSLR' | 'Tenascin' |
| 10406 | 'TNC_No14' | 'SFSTFDKDTDSAITNCALSYK' | 'Tenascin' |
| 10407 | 'TNC_No15' | 'AGTPYTVTLHGEVR' | 'Tenascin' |
| 10408 | 'TNC_No16' | 'CEEGQCVCDEGFAGVDCSEKR' | 'Tenascin' |
| 10409 | 'TNC_No17' | 'CICNEGYSGEDCSEVSPPK' | 'Tenascin' |
| 10410 | 'TNC_No18' | 'AATPYTVSIYGVIQGYR' | 'Tenascin' |
| 10411 | 'TNC_No19' | 'KQSEPLEITLLAPER' | 'Tenascin' |
| 10412 | 'TNC_No20' | 'GYDCSDMSCPNDCHQHGR' | 'Tenascin' |
| 10413 | 'TNC_No21' | 'DHGETAFAVYDK' | 'Tenascin' |
| 10414 | 'TNC_No22' | 'CECDDGFTGADCGELK' | 'Tenascin' |
| 10415 | 'TNC_No23' | 'EEFWLGLDNLNK' | 'Tenascin' |
| 10416 | 'TNC_No24' | 'LSWTADEGVFDNFVLK' | 'Tenascin' |
| 10417 | 'TNC_No25' | 'VATYLPAPEGLK' | 'Tenascin' |
| 10418 | 'TNC_No26' | 'ETFTTGLDAPR' | 'Tenascin' |
| 10419 | 'TNC_No27' | 'TVSGNTVEYALTDLEPATEYTLR' | 'Tenascin' |
| 10420 | 'TNC_No28' | 'AVDIPGLEAATPYR' | 'Tenascin' |
| 10421 | 'TNC_No29' | 'ITYVPITGGTPSMVTVDGTK' | 'Tenascin' |
| 10422 | 'TNC_No30' | 'ENFYQNWK' | 'Tenascin' |
| 10423 | 'TNC_No31' | 'LKVEGYSGTAGDSMAYHNGR' | 'Tenascin' |
| 10424 | 'TNC_No32' | 'TTLTGLRPGTEYGIGVSAVK' | 'Tenascin' |
| 10425 | 'TNC_No33' | 'YAPISGGDHAEVDVPKSQQATTK' | 'Tenascin' |
| 10426 | 'MYL9_No1' | 'ELLTTMGDRFTDEEVDEMYR' | 'Myosin regulatory light polypeptide 9' |
| 10427 | 'MYL9_No2' | 'NAFACFDEEASGFIHEDHLR' | 'Myosin regulatory light polypeptide 9' |
| 10428 | 'MYL9_No3' | 'FTDEEVDEMYR' | 'Myosin regulatory light polypeptide 9' |
| 10429 | 'MYL9_No4' | 'ELLTTMGDRFTDEEVDEMYR' | 'Myosin regulatory light polypeptide 9' |

| | A | B | C |
|---|---|---|---|
| 1 | **Peptide Name** | **Peptide Sequence** | **Peptide Description** |
| 10430 | 'AZGP1_No1' | 'YSLTYIYTGLSK' | 'Zinc-alpha-2-glycoprotein' |
| 10431 | 'AZGP1_No2' | 'QVEGMEDWKQDSQLQK' | 'Zinc-alpha-2-glycoprotein' |
| 10432 | 'AZGP1_No3' | 'AYLEEECPATLR' | 'Zinc-alpha-2-glycoprotein' |
| 10433 | 'AZGP1_No4' | 'QDPPSVVVTSHQAPGEK' | 'Zinc-alpha-2-glycoprotein' |
| 10434 | 'AZGP1_No5' | 'AREDIFMETLK' | 'Zinc-alpha-2-glycoprotein' |
| 10435 | 'AZGP1_No6' | 'YYYDGKDYIEFNK' | 'Zinc-alpha-2-glycoprotein' |
| 10436 | 'AZGP1_No7' | 'QKWEAEPVYVQR' | 'Zinc-alpha-2-glycoprotein' |
| 10437 | 'AZGP1_No8' | 'AGEVQEPELR' | 'Zinc-alpha-2-glycoprotein' |
| 10438 | 'AZGP1_No9' | 'EIPAWVPFDPAAQITK' | 'Zinc-alpha-2-glycoprotein' |
| 10439 | 'AZGP1_No10' | 'NILDRQDPPSVVVTSHQAPGEKK' | 'Zinc-alpha-2-glycoprotein' |
| 10440 | 'AZGP1_No11' | 'AYLEEECPATLRK' | 'Zinc-alpha-2-glycoprotein' |
| 10441 | 'AZGP1_No12' | 'CLAYDFYPGK' | 'Zinc-alpha-2-glycoprotein' |
| 10442 | 'AZGP1_No13' | 'AKAYLEEECPATLR' | 'Zinc-alpha-2-glycoprotein' |
| 10443 | 'AZGP1_No14' | 'SQPMGLWR' | 'Zinc-alpha-2-glycoprotein' |
| 10444 | 'AZGP1_No15' | 'IDVHWTR' | 'Zinc-alpha-2-glycoprotein' |
| 10445 | 'AZGP1_No16' | 'WEAEPVYVQR' | 'Zinc-alpha-2-glycoprotein' |
| 10446 | 'AZGP1_No17' | 'SQPMGLWR' | 'Zinc-alpha-2-glycoprotein' |
| 10447 | 'AZGP1_No18' | 'QDPPSVVVTSHQAPGEKK' | 'Zinc-alpha-2-glycoprotein' |
| 10448 | 'AZGP1_No19' | 'AKAYLEEECPATLRK' | 'Zinc-alpha-2-glycoprotein' |
| 10449 | 'AZGP1_No20' | 'NILDRQDPPSVVVTSHQAPGEK' | 'Zinc-alpha-2-glycoprotein' |
| 10450 | 'AZGP1_No21' | 'QDPPSVVVTSHQAPGEK' | 'Zinc-alpha-2-glycoprotein' |
| 10451 | 'AZGP1_No22' | 'AREDIFMETLK' | 'Zinc-alpha-2-glycoprotein' |
| 10452 | 'AZGP1_No23' | 'QVEGMEDWKQDSQLQK' | 'Zinc-alpha-2-glycoprotein' |
| 10453 | 'AZGP1_No24' | 'CLAYDFYPGKIDVHWTR' | 'Zinc-alpha-2-glycoprotein' |
| 10454 | 'AZGP1_No25' | 'QKWEAEPVYVQR' | 'Zinc-alpha-2-glycoprotein' |
| 10455 | 'AZGP1_No26' | 'QVEGMEDWKQDSQLQK' | 'Zinc-alpha-2-glycoprotein' |
| 10456 | 'AZGP1_No27' | 'QVEGMEDWKQDSQLQK' | 'Zinc-alpha-2-glycoprotein' |
| 10457 | 'AZGP1_No28' | 'LKCLAYDFYPGK' | 'Zinc-alpha-2-glycoprotein' |
| 10458 | 'AZGP1_No29' | 'EDIFMETLK' | 'Zinc-alpha-2-glycoprotein' |
| 10459 | 'AZGP1_No30' | 'DYIEFNKEIPAWVPFDPAAQITK' | 'Zinc-alpha-2-glycoprotein' |
| 10460 | 'AZGP1_No31' | 'HVEDVPAFQALGSLNDLQFFR' | 'Zinc-alpha-2-glycoprotein' |
| 10461 | 'AZGP1_No32' | 'DYIEFNK' | 'Zinc-alpha-2-glycoprotein' |
| 10462 | 'AZGP1_No33' | 'QDPPSVVVTSHQAPGEKK' | 'Zinc-alpha-2-glycoprotein' |
| 10463 | 'AZGP1_No34' | 'YYYDGKDYIEFNKEIPAWVPFDPAAQITK' | 'Zinc-alpha-2-glycoprotein' |
| 10464 | 'AZGP1_No35' | 'LKCLAYDFYPGKIDVHWTR' | 'Zinc-alpha-2-glycoprotein' |
| 10465 | 'AZGP1_No36' | 'YSKNILDRQDPPSVVVTSHQAPGEK' | 'Zinc-alpha-2-glycoprotein' |
| 10466 | 'AZGP1_No37' | 'DIVEYYNDSNGSHVLQGR' | 'Zinc-alpha-2-glycoprotein' |
| 10467 | 'AZGP1_No38' | 'QDPPSVVVTSHQAPGEKKK' | 'Zinc-alpha-2-glycoprotein' |
| 10468 | 'AZGP1_No39' | 'EDIFMETLK' | 'Zinc-alpha-2-glycoprotein' |
| 10469 | 'ATP5F1A_No1' | 'TGAIVDVPVGEELLGR' | 'ATP synthase subunit alpha, mitochondrial' |
| 10470 | 'ATP5F1A_No2' | 'FENAFLSHVVSQHQALLGTIR' | 'ATP synthase subunit alpha, mitochondrial' |
| 10471 | 'ATP5F1A_No3' | 'TGTAEMSSILEER' | 'ATP synthase subunit alpha, mitochondrial' |
| 10472 | 'ATP5F1A_No4' | 'LELAQYR' | 'ATP synthase subunit alpha, mitochondrial' |
| 10473 | 'ATP5F1A_No5' | 'HALIIYDDLSK' | 'ATP synthase subunit alpha, mitochondrial' |

| | A | B | C |
|---|---|---|---|
| 1 | **Peptide Name** | **Peptide Sequence** | **Peptide Description** |
| 10474 | 'ATP5F1A_No6' | 'GMSLNLEPDNVGVVVFGNDK' | 'ATP synthase subunit alpha, mitochondrial' |
| 10475 | 'ATP5F1A_No7' | 'ILGADTSVDLEETGR' | 'ATP synthase subunit alpha, mitochondrial' |
| 10476 | 'ATP5F1A_No8' | 'LKEIVTNFLAGFEA' | 'ATP synthase subunit alpha, mitochondrial' |
| 10477 | 'ATP5F1A_No9' | 'AVDSLVPIGR' | 'ATP synthase subunit alpha, mitochondrial' |
| 10478 | 'PSMA1_No1' | 'FVFDRPLPVSR' | 'Proteasome subunit alpha type-1' |
| 10479 | 'PSMA1_No2' | 'IHQIEYAMEAVK' | 'Proteasome subunit alpha type-1' |
| 10480 | 'PSMA1_No3' | 'ALRETLPAEQDLTTK' | 'Proteasome subunit alpha type-1' |
| 10481 | 'PSMA1_No4' | 'AQPAQPADEPAEKADEPMEH' | 'Proteasome subunit alpha type-1' |
| 10482 | 'PSMA2_No1' | 'KLAQQYYLVYQEPIPTAQLVQR' | 'Proteasome subunit alpha type-2' |
| 10483 | 'PSMA2_No2' | 'LVQIEYALAAVAGGAPSVGIK' | 'Proteasome subunit alpha type-2' |
| 10484 | 'PSMA2_No3' | 'GYSFSLTTFSPSGK' | 'Proteasome subunit alpha type-2' |
| 10485 | 'PSMA2_No4' | 'LAQQYYLVYQEPIPTAQLVQR' | 'Proteasome subunit alpha type-2' |
| 10486 | 'PSMA3_No1' | 'IIYIVHDEVKDK' | 'Proteasome subunit alpha type-3' |
| 10487 | 'PSMA3_No2' | 'SSIGTGYDLSASTFSPDGR' | 'Proteasome subunit alpha type-3' |
| 10488 | 'PSMA3_No3' | 'HVGMAVAGLLADAR' | 'Proteasome subunit alpha type-3' |
| 10489 | 'PSMA3_No4' | 'HVGMAVAGLLADAR' | 'Proteasome subunit alpha type-3' |
| 10490 | 'PSMA4_No1' | 'LLDEVFFSEK' | 'Proteasome subunit alpha type-4' |
| 10491 | 'PSMA4_No2' | 'HYGFQLYQSDPSGNYGGWK' | 'Proteasome subunit alpha type-4' |
| 10492 | 'MSN_No1' | 'APDFVFYAPR' | 'Moesin' |
| 10493 | 'MSN_No2' | 'LFFLQVK' | 'Moesin' |
| 10494 | 'MSN_No3' | 'GMLREDAVLEYLK' | 'Moesin' |
| 10495 | 'MSN_No4' | 'AQMVQEDLEK' | 'Moesin' |
| 10496 | 'MSN_No5' | 'VTTMDAELEFAIQPNTTGK' | 'Moesin' |
| 10497 | 'MSN_No6' | 'AKFYPEDVSEELIQDITQR' | 'Moesin' |
| 10498 | 'MSN_No7' | 'ERQEAEEAKEALLQASR' | 'Moesin' |
| 10499 | 'MSN_No8' | 'ALTSELANARDESKK' | 'Moesin' |
| 10500 | 'MSN_No9' | 'VTTMDAELEFAIQPNTTGK' | 'Moesin' |
| 10501 | 'MSN_No10' | 'VLEQHKLNKDQWEER' | 'Moesin' |
| 10502 | 'MSN_No11' | 'TQEQLALEMAELTAR' | 'Moesin' |
| 10503 | 'MSN_No12' | 'KAPDFVFYAPR' | 'Moesin' |
| 10504 | 'MSN_No13' | 'AQMVQEDLEK' | 'Moesin' |
| 10505 | 'MSN_No14' | 'IAQDLEMYGVNYFSIK' | 'Moesin' |
| 10506 | 'MSN_No15' | 'KTQEQLALEMAELTAR' | 'Moesin' |
| 10507 | 'MSN_No16' | 'QRIDEFESM' | 'Moesin' |
| 10508 | 'MSN_No17' | 'IGFPWSEIR' | 'Moesin' |
| 10509 | 'MSN_No18' | 'TAMSTPHVAEPAENEQDEQDENGAEASADLR' | 'Moesin' |
| 10510 | 'MSN_No19' | 'DQKKTQEQLALEMAELTAR' | 'Moesin' |
| 10511 | 'S100A4_No1' | 'ALDVMVSTFHK' | 'Protein S100-A4' |
| 10512 | 'S100A4_No2' | 'SELKELLTR' | 'Protein S100-A4' |
| 10513 | 'EEF1G_No1' | 'STFVLDEFKR' | 'Elongation factor 1-gamma' |
| 10514 | 'EEF1G_No2' | 'ILGLLDAYLK' | 'Elongation factor 1-gamma' |
| 10515 | 'EEF1G_No3' | 'KLDPGSEETQTLVR' | 'Elongation factor 1-gamma' |
| 10516 | 'MST1_No1' | 'SPLNDFQVLR' | 'Hepatocyte growth factor-like protein' |
| 10517 | 'MST1_No2' | 'FLDQGLDDNYCR' | 'Hepatocyte growth factor-like protein' |

| | A | B | C |
|---|---|---|---|
| 1 | **Peptide Name** | **Peptide Sequence** | **Peptide Description** |
| 10518 | 'MST1_No3' | 'TPFDYCALR' | 'Hepatocyte growth factor-like protein' |
| 10519 | 'MST1_No4' | 'VVGGHPGNSPWTVSLR' | 'Hepatocyte growth factor-like protein' |
| 10520 | 'MST1_No5' | 'GTMATTVGGLPCQAWSHK' | 'Hepatocyte growth factor-like protein' |
| 10521 | 'MST1_No6' | 'NPDGSERPWCYTTDPQIER' | 'Hepatocyte growth factor-like protein' |
| 10522 | 'MST1_No7' | 'VSVFVDWIHK' | 'Hepatocyte growth factor-like protein' |
| 10523 | 'MST1_No8' | 'VALICLPPEWYVVPPGTK' | 'Hepatocyte growth factor-like protein' |
| 10524 | 'MST1_No9' | 'WDLQHPHQHPFEPGK' | 'Hepatocyte growth factor-like protein' |
| 10525 | 'MST1_No10' | 'EFCDLPR' | 'Hepatocyte growth factor-like protein' |
| 10526 | 'MST1_No11' | 'WDAQIPHQHR' | 'Hepatocyte growth factor-like protein' |
| 10527 | 'MST1_No12' | 'AAFCYQIR' | 'Hepatocyte growth factor-like protein' |
| 10528 | 'MST1_No13' | 'NPDGDPGGPWCYTTDPAVR' | 'Hepatocyte growth factor-like protein' |
| 10529 | 'MST1_No14' | 'GTMATTVGGLPCQAWSHKFPNDHK' | 'Hepatocyte growth factor-like protein' |
| 10530 | 'MST1_No15' | 'GTELQHLLHAVVPGPWQEDVADAEECAGR' | 'Hepatocyte growth factor-like protein' |
| 10531 | 'MST1_No16' | 'TPFDYCALRR' | 'Hepatocyte growth factor-like protein' |
| 10532 | 'MST1_No17' | 'NPDGSEAPWCFTLRPGMR' | 'Hepatocyte growth factor-like protein' |
| 10533 | 'MST1_No18' | 'CADDQPPSILDPPDQVQFEK' | 'Hepatocyte growth factor-like protein' |
| 10534 | 'MST1_No19' | 'NPDGDSHGPWCYTMDPR' | 'Hepatocyte growth factor-like protein' |
| 10535 | 'MST1_No20' | 'WSAETPHKPQFTFTSEPHAQLEENFCR' | 'Hepatocyte growth factor-like protein' |
| 10536 | 'MST1_No21' | 'MVCGPSGSQLVLLKLER' | 'Hepatocyte growth factor-like protein' |
| 10537 | 'MST1_No22' | 'GTMATTVGGLPCQAWSHK' | 'Hepatocyte growth factor-like protein' |
| 10538 | 'MST1_No23' | 'RCTDDVRPQDCYHGAGEQYR' | 'Hepatocyte growth factor-like protein' |
| 10539 | 'MST1_No24' | 'CTDDVRPQDCYHGAGEQYR' | 'Hepatocyte growth factor-like protein' |
| 10540 | 'MST1_No25' | 'EAACVWCNGEEYRGAVDR' | 'Hepatocyte growth factor-like protein' |
| 10541 | 'MST1_No26' | 'MVCGPSGSQLVLLK' | 'Hepatocyte growth factor-like protein' |
| 10542 | 'MST1_No27' | 'TCIMNNGVGYR' | 'Hepatocyte growth factor-like protein' |
| 10543 | 'MST1_No28' | 'SRWPAVFTR' | 'Hepatocyte growth factor-like protein' |
| 10544 | 'MST1_No29' | 'MVCGPSGSQLVLLK' | 'Hepatocyte growth factor-like protein' |
| 10545 | 'MST1_No30' | 'QGQHFCGGSLVK' | 'Hepatocyte growth factor-like protein' |
| 10546 | 'MST1_No31' | 'NPDGDSHGPWCYTMDPR' | 'Hepatocyte growth factor-like protein' |
| 10547 | 'MST1_No32' | 'FLDQGLDDNYCRNPDGSERPWCYTTDPQIER' | 'Hepatocyte growth factor-like protein' |
| 10548 | 'MST1_No33' | 'RCADDQPPSILDPPDQVQFEK' | 'Hepatocyte growth factor-like protein' |
| 10549 | 'MST1_No34' | 'NPDGSEAPWCFTLRPGMR' | 'Hepatocyte growth factor-like protein' |
| 10550 | 'MST1_No35' | 'CEIAGWGETK' | 'Hepatocyte growth factor-like protein' |
| 10551 | 'MST1_No36' | 'SVTLNQR' | 'Hepatocyte growth factor-like protein' |
| 10552 | 'MST1_No37' | 'MVCGPSGSQLVLLKLER' | 'Hepatocyte growth factor-like protein' |
| 10553 | 'MST1_No38' | 'QGQHFCGGSLVK' | 'Hepatocyte growth factor-like protein' |
| 10554 | 'MST1_No39' | 'GTMATTVGGLPCQAWSHKFPNDHK' | 'Hepatocyte growth factor-like protein' |
| 10555 | 'MST1_No40' | 'FTPEKYACKDLR' | 'Hepatocyte growth factor-like protein' |
| 10556 | 'MST1_No41' | 'NPDGSERPWCYTTDPQIEREFCDLPR' | 'Hepatocyte growth factor-like protein' |
| 10557 | 'MST1_No42' | 'CTDDVRPQDCYHGAGEQYRGTVSK' | 'Hepatocyte growth factor-like protein' |
| 10558 | 'STOM_No1' | 'VQNATLAVANITNADSATR' | 'Erythrocyte band 7 integral membrane protein' |
| 10559 | 'STOM_No2' | 'VIAAEGEMNASR' | 'Erythrocyte band 7 integral membrane protein' |
| 10560 | 'STOM_No3' | 'NSTIVFPLPIDMLQGIIGAK' | 'Erythrocyte band 7 integral membrane protein' |
| 10561 | 'STOM_No4' | 'YLQTLTTIAAEK' | 'Erythrocyte band 7 integral membrane protein' |

| | A | B | C |
|---|---|---|---|
| 1 | **Peptide Name** | **Peptide Sequence** | **Peptide Description** |
| 10562 | 'STOM_No5' | 'ALKEASMVITESPAALQLR' | 'Erythrocyte band 7 integral membrane protein' |
| 10563 | 'STOM_No6' | 'EASMVITESPAALQLR' | 'Erythrocyte band 7 integral membrane protein' |
| 10564 | 'STOM_No7' | 'TISFDIPPQEILTK' | 'Erythrocyte band 7 integral membrane protein' |
| 10565 | 'STOM_No8' | 'NLSQILSDREEIAHNMQSTLDDATDAWGIK' | 'Erythrocyte band 7 integral membrane protein' |
| 10566 | 'STOM_No9' | 'VEIKDVKLPVQLQR' | 'Erythrocyte band 7 integral membrane protein' |
| 10567 | 'STOM_No10' | 'AKVIAAEGEMNASR' | 'Erythrocyte band 7 integral membrane protein' |
| 10568 | 'STOM_No11' | 'ALKEASMVITESPAALQLR' | 'Erythrocyte band 7 integral membrane protein' |
| 10569 | 'STOM_No12' | 'LLAQTTLR' | 'Erythrocyte band 7 integral membrane protein' |
| 10570 | 'STOM_No13' | 'GPGLFFILPCTDSFIK' | 'Erythrocyte band 7 integral membrane protein' |
| 10571 | 'PON1_No1' | 'YVYIAELLAHK' | 'Serum paraoxonase/arylesterase 1' |
| 10572 | 'PON1_No2' | 'EVQPVELPNCNLVK' | 'Serum paraoxonase/arylesterase 1' |
| 10573 | 'PON1_No3' | 'FDVSSFNPHGISTFTDEDNAMYLLVVNHPDAK' | 'Serum paraoxonase/arylesterase 1' |
| 10574 | 'PON1_No4' | 'VTQVYAENGTVLQGSTVASVYK' | 'Serum paraoxonase/arylesterase 1' |
| 10575 | 'PON1_No5' | 'IFFYDSENPPASEVLR' | 'Serum paraoxonase/arylesterase 1' |
| 10576 | 'PON1_No6' | 'IQNILTEEPK' | 'Serum paraoxonase/arylesterase 1' |
| 10577 | 'PON1_No7' | 'ILLMDLNEEDPTVLELGITGSK' | 'Serum paraoxonase/arylesterase 1' |
| 10578 | 'PON1_No8' | 'FQEEEKSLLHLK' | 'Serum paraoxonase/arylesterase 1' |
| 10579 | 'PON1_No9' | 'LNALREVQPVELPNCNLVK' | 'Serum paraoxonase/arylesterase 1' |
| 10580 | 'PON1_No10' | 'GKLLIGTVFHK' | 'Serum paraoxonase/arylesterase 1' |
| 10581 | 'PON1_No11' | 'STVELFKFQEEEK' | 'Serum paraoxonase/arylesterase 1' |
| 10582 | 'PON1_No12' | 'STVELFKFQEEEKSLLHLK' | 'Serum paraoxonase/arylesterase 1' |
| 10583 | 'PON1_No13' | 'SFNPNSPGK' | 'Serum paraoxonase/arylesterase 1' |
| 10584 | 'PON1_No14' | 'VVAEGFDFANGINISPDGK' | 'Serum paraoxonase/arylesterase 1' |
| 10585 | 'PON1_No15' | 'FDVSSFNPHGISTFTDEDNAMYLLVVNHPDAK' | 'Serum paraoxonase/arylesterase 1' |
| 10586 | 'PON1_No16' | 'ILLMDLNEEDPTVLELGITGSK' | 'Serum paraoxonase/arylesterase 1' |
| 10587 | 'PON1_No17' | 'VTQVYAENGTVLQGSTVASVYKGK' | 'Serum paraoxonase/arylesterase 1' |
| 10588 | 'PON1_No18' | 'IQNILTEEPKVTQVYAENGTVLQGSTVASVYKGK' | 'Serum paraoxonase/arylesterase 1' |
| 10589 | 'PON1_No19' | 'YVYIAELLAHKIHVYEK' | 'Serum paraoxonase/arylesterase 1' |
| 10590 | 'PON1_No20' | 'SLDFNTLVDNISVDPETGDLWVGCHPNGMK' | 'Serum paraoxonase/arylesterase 1' |
| 10591 | 'PON1_No21' | 'SLDFNTLVDNISVDPETGDLWVGCHPNGMK' | 'Serum paraoxonase/arylesterase 1' |
| 10592 | 'PON1_No22' | 'GIETGSEDLEILPNGLAFISSGLKYPGIK' | 'Serum paraoxonase/arylesterase 1' |
| 10593 | 'YWHAQ_No1' | 'AVTEQGAELSNEER' | '14-3-3 protein theta' |
| 10594 | 'YWHAQ_No2' | 'YLIANATNPESK' | '14-3-3 protein theta' |
| 10595 | 'YWHAQ_No3' | 'YLAEVACGDDRK' | '14-3-3 protein theta' |
| 10596 | 'DPP4_No1' | 'ISLQWLR' | 'Dipeptidyl peptidase 4' |
| 10597 | 'DPP4_No2' | 'LAYVWNNDIYVK' | 'Dipeptidyl peptidase 4' |
| 10598 | 'DPP4_No3' | 'FRPSEPHFTLDGNSFYK' | 'Dipeptidyl peptidase 4' |
| 10599 | 'DPP4_No4' | 'HSYTASYDIYDLNKR' | 'Dipeptidyl peptidase 4' |
| 10600 | 'DPP4_No5' | 'LGTFEVEDQIEAAR' | 'Dipeptidyl peptidase 4' |
| 10601 | 'DPP4_No6' | 'VTCLSCELNPER' | 'Dipeptidyl peptidase 4' |
| 10602 | 'DPP4_No7' | 'TYTLTDYLK' | 'Dipeptidyl peptidase 4' |
| 10603 | 'DPP4_No8' | 'WEYYDSVYTER' | 'Dipeptidyl peptidase 4' |
| 10604 | 'DPP4_No9' | 'HICYFQIDK' | 'Dipeptidyl peptidase 4' |
| 10605 | 'DPP4_No10' | 'QVEYLLIHGTADDNVHFQQSAQISK' | 'Dipeptidyl peptidase 4' |

| | A | B | C |
|---|---|---|---|
| 1 | **Peptide Name** | **Peptide Sequence** | **Peptide Description** |
| 10606 | 'DPP4_No11' | 'KTYTLTDYLK' | 'Dipeptidyl peptidase 4' |
| 10607 | 'DPP4_No12' | 'CSGPGLPLYTLHSSVNDK' | 'Dipeptidyl peptidase 4' |
| 10608 | 'DPP4_No13' | 'WISDHEYLYK' | 'Dipeptidyl peptidase 4' |
| 10609 | 'DPP4_No14' | 'HSYTASYDIYDLNK' | 'Dipeptidyl peptidase 4' |
| 10610 | 'DPP4_No15' | 'KYPLLLDVYAGPCSQK' | 'Dipeptidyl peptidase 4' |
| 10611 | 'DPP4_No16' | 'YPLLLDVYAGPCSQK' | 'Dipeptidyl peptidase 4' |
| 10612 | 'DPP4_No17' | 'QHIEMSTTGWVGR' | 'Dipeptidyl peptidase 4' |
| 10613 | 'DPP4_No18' | 'RLGTFEVEDQIEAAR' | 'Dipeptidyl peptidase 4' |
| 10614 | 'DPP4_No19' | 'AENFKQVEYLLIHGTADDNVHFQQSAQISK' | 'Dipeptidyl peptidase 4' |
| 10615 | 'CALR_No1' | 'FYALSASFEPFSNK' | 'Calreticulin' |
| 10616 | 'CALR_No2' | 'IKDPDASKPEDWDER' | 'Calreticulin' |
| 10617 | 'CALR_No3' | 'CKDDEFTHLYTLIVRPDNTYEVK' | 'Calreticulin' |
| 10618 | 'CALR_No4' | 'GQTLVVQFTVK' | 'Calreticulin' |
| 10619 | 'CALR_No5' | 'IDDPTDSKPEDWDKPEHIPDPDAK' | 'Calreticulin' |
| 10620 | 'CALR_No6' | 'IDNSQVESGSLEDDWDFLPPKK' | 'Calreticulin' |
| 10621 | 'CALR_No7' | 'FYGDEEKDKGLQTSQDAR' | 'Calreticulin' |
| 10622 | 'CALR_No8' | 'IDNSQVESGSLEDDWDFLPPK' | 'Calreticulin' |
| 10623 | 'CANX_No1' | 'AEEDEILNR' | 'Calnexin' |
| 10624 | 'CANX_No2' | 'GTLSGWILSK' | 'Calnexin' |
| 10625 | 'CANX_No3' | 'KIPNPDFFEDLEPFR' | 'Calnexin' |
| 10626 | 'CANX_No4' | 'APVPTGEVYFADSFDR' | 'Calnexin' |
| 10627 | 'CFP_No1' | 'TCNHPVPQHGGPFCAGDATR' | 'Properdin' |
| 10628 | 'CFP_No2' | 'RPCLHVPACKDPEEEEL' | 'Properdin' |
| 10629 | 'CFP_No3' | 'LCTPLLPK' | 'Properdin' |
| 10630 | 'CFP_No4' | 'CSAPEPSQKPPGKPCPGLAYEQR' | 'Properdin' |
| 10631 | 'CFP_No5' | 'GLLGGGVSVEDCCLNTAFAYQK' | 'Properdin' |
| 10632 | 'CFP_No6' | 'HCYSIQHCPLK' | 'Properdin' |
| 10633 | 'CFP_No7' | 'CEELQGQKLVVEEK' | 'Properdin' |
| 10634 | 'CFP_No8' | 'YPPTVSMVEGQGEK' | 'Properdin' |
| 10635 | 'CFP_No9' | 'QRLCTPLLPK' | 'Properdin' |
| 10636 | 'CFP_No10' | 'SGGLCQPCR' | 'Properdin' |
| 10637 | 'CFP_No11' | 'SISCQEIPGQQSR' | 'Properdin' |
| 10638 | 'CFP_No12' | 'YPPTVSMVEGQGEK' | 'Properdin' |
| 10639 | 'CFP_No13' | 'CAGQQQDIR' | 'Properdin' |
| 10640 | 'CFP_No14' | 'GLLGGGVSVEDCCLNTAFAYQKR' | 'Properdin' |
| 10641 | 'PSMB8_No1' | 'FQHGVIAAVDSR' | 'Proteasome subunit beta type-8' |
| 10642 | 'PSMB8_No2' | 'KGPGLYYVDEHGTR' | 'Proteasome subunit beta type-8' |
| 10643 | 'PSMA5_No1' | 'LFQVEYAIEAIK' | 'Proteasome subunit alpha type-5' |
| 10644 | 'PSMA5_No2' | 'AIGSASEGAQSSLQEVYHK' | 'Proteasome subunit alpha type-5' |
| 10645 | 'PSMA5_No3' | 'ITSPLMEPSSIEK' | 'Proteasome subunit alpha type-5' |
| 10646 | 'GRN_No1' | 'CDMEVSCPDGYTCCR' | 'Progranulin' |
| 10647 | 'GRN_No2' | 'CDMEVSCPDGYTCCR' | 'Progranulin' |
| 10648 | 'GRN_No3' | 'DVECGEGHFCHDNQTCCR' | 'Progranulin' |
| 10649 | 'GRN_No4' | 'SRCPDGSTCCELPSGK' | 'Progranulin' |

| | A | B | C |
|---|---|---|---|
| 1 | **Peptide Name** | **Peptide Sequence** | **Peptide Description** |
| 10650 | 'GRN_No5' | 'AVALSSSVMCPDAR' | 'Progranulin' |
| 10651 | 'PCSK6_No1' | 'NVVVTILDDGIER' | 'Proprotein convertase subtilisin/kexin type 6' |
| 10652 | 'PCSK6_No2' | 'SIPLVQVLR' | 'Proprotein convertase subtilisin/kexin type 6' |
| 10653 | 'PCSK6_No3' | 'GSCIPDCEPGTYFDSELIR' | 'Proprotein convertase subtilisin/kexin type 6' |
| 10654 | 'PTPN6_No1' | 'GEPWTFLVR' | 'Tyrosine-protein phosphatase non-receptor type 6' |
| 10655 | 'PTPN6_No2' | 'AGFWEEFESLQK' | 'Tyrosine-protein phosphatase non-receptor type 6' |
| 10656 | 'PTPN6_No3' | 'AYGPYSVTNCGEHDTTEYKLR' | 'Tyrosine-protein phosphatase non-receptor type 6' |
| 10657 | 'TKT_No1' | 'ILATPPQEDAPSVDIANIR' | 'Transketolase' |
| 10658 | 'TKT_No2' | 'LDNLVAILDINR' | 'Transketolase' |
| 10659 | 'TKT_No3' | 'VLDPFTIKPLDRK' | 'Transketolase' |
| 10660 | 'TKT_No4' | 'LGQSDPAPLQHQMDIYQK' | 'Transketolase' |
| 10661 | 'TKT_No5' | 'GITGVEDKESWHGKPLPK' | 'Transketolase' |
| 10662 | 'TKT_No6' | 'ISSDLDGHPVPK' | 'Transketolase' |
| 10663 | 'TKT_No7' | 'SKDDQVTVIGAGVTLHEALAAAELLKK' | 'Transketolase' |
| 10664 | 'TKT_No8' | 'NSTFSEIFKK' | 'Transketolase' |
| 10665 | 'SERPINB3_No1' | 'GLVLSGVLHK' | 'Serpin B3' |
| 10666 | 'SERPINB3_No2' | 'TYLFLQEYLDAIKK' | 'Serpin B3' |
| 10667 | 'SERPINB3_No3' | 'FKVEESYDLKDTLR' | 'Serpin B3' |
| 10668 | 'SERPINB3_No4' | 'VLHFDQVTENTTGK' | 'Serpin B3' |
| 10669 | 'SERPINA4_No1' | 'LFHTNFYDTVGTIQLINDHVK' | 'Kallistatin' |
| 10670 | 'SERPINA4_No2' | 'VGSALFLSHNLK' | 'Kallistatin' |
| 10671 | 'SERPINA4_No3' | 'WADLSGITK' | 'Kallistatin' |
| 10672 | 'SERPINA4_No4' | 'IVDLVSELKK' | 'Kallistatin' |
| 10673 | 'SERPINA4_No5' | 'ALWEKPFISSR' | 'Kallistatin' |
| 10674 | 'SERPINA4_No6' | 'GFQHLLHTLNLPGHGLETR' | 'Kallistatin' |
| 10675 | 'SERPINA4_No7' | 'FYYLIASETPGK' | 'Kallistatin' |
| 10676 | 'SERPINA4_No8' | 'LFHTNFYDTVGTIQLINDHVKK' | 'Kallistatin' |
| 10677 | 'SERPINA4_No9' | 'IAPANADFAFR' | 'Kallistatin' |
| 10678 | 'SERPINA4_No10' | 'LGFTDLFSK' | 'Kallistatin' |
| 10679 | 'SERPINA4_No11' | 'VPMMLQDQEHHWYLHDR' | 'Kallistatin' |
| 10680 | 'SERPINA4_No12' | 'VPMMLQDQEHHWYLHDR' | 'Kallistatin' |
| 10681 | 'SERPINA4_No13' | 'IVDLVSELK' | 'Kallistatin' |
| 10682 | 'SERPINA4_No14' | 'VPMMLQDQEHHWYLHDR' | 'Kallistatin' |
| 10683 | 'SERPINA4_No15' | 'GKIVDLVSELKK' | 'Kallistatin' |
| 10684 | 'SERPINA4_No16' | 'KLELHLPK' | 'Kallistatin' |
| 10685 | 'SERPINA4_No17' | 'FSISGSYVLDQILPR' | 'Kallistatin' |
| 10686 | 'SERPINA4_No18' | 'ATLDVDEAGTEAAAATSFAIK' | 'Kallistatin' |
| 10687 | 'SERPINA4_No19' | 'EIEEVLTPEMLMR' | 'Kallistatin' |
| 10688 | 'SERPINA4_No20' | 'TTPKDFYVDENTTVR' | 'Kallistatin' |
| 10689 | 'SERPINA4_No21' | 'VPMMLQDQEHHWYLHDR' | 'Kallistatin' |
| 10690 | 'SERPINA4_No22' | 'MREIEEVLTPEMLMR' | 'Kallistatin' |
| 10691 | 'SERPINA4_No23' | 'FFSAQTNR' | 'Kallistatin' |
| 10692 | 'SERPINA4_No24' | 'EIEEVLTPEMLMR' | 'Kallistatin' |
| 10693 | 'SERPINA4_No25' | 'YLPCSVLR' | 'Kallistatin' |

| | A | B | C |
|---|---|---|---|
| 1 | **Peptide Name** | **Peptide Sequence** | **Peptide Description** |
| 10694 | 'SERPINA4_No26' | 'GDATVFFILPNQGK' | 'Kallistatin' |
| 10695 | 'SERPINA4_No27' | 'EIEEVLTPEMLMR' | 'Kallistatin' |
| 10696 | 'SERPINA4_No28' | 'MDYKGDATVFFILPNQGK' | 'Kallistatin' |
| 10697 | 'SERPINA4_No29' | 'MDYKGDATVFFILPNQGK' | 'Kallistatin' |
| 10698 | 'SERPINA4_No30' | 'MREIEEVLTPEMLMR' | 'Kallistatin' |
| 10699 | 'SERPINA4_No31' | 'DFYVDENTTVR' | 'Kallistatin' |
| 10700 | 'SERPINA4_No32' | 'MREIEEVLTPEMLMR' | 'Kallistatin' |
| 10701 | 'SERPINA4_No33' | 'MREIEEVLTPEMLMR' | 'Kallistatin' |
| 10702 | 'SERPINA4_No34' | 'VGSALFLSHNLKFLAK' | 'Kallistatin' |
| 10703 | 'SERPINA4_No35' | 'EIEEVLTPEMLMR' | 'Kallistatin' |
| 10704 | 'SERPINA4_No36' | 'LFHTNFYDTVGTIQLINDHVKKETR' | 'Kallistatin' |
| 10705 | 'SERPINA4_No37' | 'MREIEEVLTPEMLMR' | 'Kallistatin' |
| 10706 | 'SERPINA4_No38' | 'WADLSGITKQQKLEASK' | 'Kallistatin' |
| 10707 | 'PRDX6_No1' | 'FHDFLGDSWGILFSHPR' | 'Peroxiredoxin-6' |
| 10708 | 'PRDX6_No2' | 'LPFPIIDDR' | 'Peroxiredoxin-6' |
| 10709 | 'PRDX6_No3' | 'PGGLLLGDVAPNFEANTTVGR' | 'Peroxiredoxin-6' |
| 10710 | 'PRDX6_No4' | 'VVFVFGPDKK' | 'Peroxiredoxin-6' |
| 10711 | 'PRDX6_No5' | 'VATPVDWKDGDSVMVLPTIPEEEAK' | 'Peroxiredoxin-6' |
| 10712 | 'PRDX6_No6' | 'DINAYNCEEPTEK' | 'Peroxiredoxin-6' |
| 10713 | 'PRDX6_No7' | 'DFTPVCTTELGR' | 'Peroxiredoxin-6' |
| 10714 | 'PRDX6_No8' | 'DINAYNCEEPTEKLPFPIIDDR' | 'Peroxiredoxin-6' |
| 10715 | 'PRDX6_No9' | 'VATPVDWKDGDSVMVLPTIPEEEAK' | 'Peroxiredoxin-6' |
| 10716 | 'PRDX6_No10' | 'VVFVFGPDK' | 'Peroxiredoxin-6' |
| 10717 | 'PRDX6_No11' | 'LPFPIIDDRNR' | 'Peroxiredoxin-6' |
| 10718 | 'PRDX6_No12' | 'LSILYPATTGR' | 'Peroxiredoxin-6' |
| 10719 | 'BLVRB_No1' | 'YVAVMPPHIGDQPLTGAYTVTLDGR' | 'Flavin reductase (NADPH)' |
| 10720 | 'BLVRB_No2' | 'LPSEGPRPAHVVVGDVLQAADVDK' | 'Flavin reductase (NADPH)' |
| 10721 | 'BLVRB_No3' | 'CLTTDEYDGHSTYPSHQYQ' | 'Flavin reductase (NADPH)' |
| 10722 | 'BLVRB_No4' | 'NDLSPTTVMSEGAR' | 'Flavin reductase (NADPH)' |
| 10723 | 'BLVRB_No5' | 'LQAVTDDHIR' | 'Flavin reductase (NADPH)' |
| 10724 | 'BLVRB_No6' | 'TVAGQDAVIVLLGTR' | 'Flavin reductase (NADPH)' |
| 10725 | 'BLVRB_No7' | 'YVAVMPPHIGDQPLTGAYTVTLDGR' | 'Flavin reductase (NADPH)' |
| 10726 | 'BLVRB_No8' | 'VVACTSAFLLWDPTK' | 'Flavin reductase (NADPH)' |
| 10727 | 'BLVRB_No9' | 'HDLGHFMLR' | 'Flavin reductase (NADPH)' |
| 10728 | 'BLVRB_No10' | 'PAHVVVGDVLQAADVDK' | 'Flavin reductase (NADPH)' |
| 10729 | 'BLVRB_No11' | 'VVACTSAFLLWDPTKVPPR' | 'Flavin reductase (NADPH)' |
| 10730 | 'PRDX5_No1' | 'THLPGFVEQAEALK' | 'Peroxiredoxin-5, mitochondrial' |
| 10731 | 'PRDX5_No2' | 'VGDAIPAVEVFEGEPGNK' | 'Peroxiredoxin-5, mitochondrial' |
| 10732 | 'PRDX5_No3' | 'VNLAELFK' | 'Peroxiredoxin-5, mitochondrial' |
| 10733 | 'PRDX5_No4' | 'LLADPTGAFGK' | 'Peroxiredoxin-5, mitochondrial' |
| 10734 | 'DDT_No1' | 'FFPLESWQIGK' | 'D-dopachrome decarboxylase' |
| 10735 | 'DDT_No2' | 'LCAAAASILGKPADR' | 'D-dopachrome decarboxylase' |
| 10736 | 'DDT_No3' | 'PFLELDTNLPANR' | 'D-dopachrome decarboxylase' |
| 10737 | 'DDT_No4' | 'SHSAHFFEFLTK' | 'D-dopachrome decarboxylase' |

| | A | B | C |
|---|---|---|---|
| 1 | **Peptide Name** | **Peptide Sequence** | **Peptide Description** |
| 10738 | 'DDT_No5' | 'PFLELDTNLPANRVPAGLEK' | 'D-dopachrome decarboxylase' |
| 10739 | 'PRDX3_No1' | 'HLSVNDLPVGR' | 'Thioredoxin-dependent peroxide reductase, mitochondrial' |
| 10740 | 'PRDX3_No2' | 'GLFIIDPNGVIK' | 'Thioredoxin-dependent peroxide reductase, mitochondrial' |
| 10741 | 'CMPK1_No1' | 'FLIDGFPR' | 'UMP-CMP kinase' |
| 10742 | 'CMPK1_No2' | 'MKPLVVFVLGGPGAGK' | 'UMP-CMP kinase' |
| 10743 | 'CMPK1_No3' | 'IQTYLQSTKPIIDLYEEMGK' | 'UMP-CMP kinase' |
| 10744 | 'PEBP1_No1' | 'YVWLVYEQDRPLK' | 'Phosphatidylethanolamine-binding protein 1' |
| 10745 | 'PEBP1_No2' | 'GNDISSGTVLSDYVGSGPPK' | 'Phosphatidylethanolamine-binding protein 1' |
| 10746 | 'PEBP1_No3' | 'APVAGTCYQAEWDDYVPK' | 'Phosphatidylethanolamine-binding protein 1' |
| 10747 | 'PDIA3_No1' | 'LAPEYEAAATR' | 'Protein disulfide-isomerase A3' |
| 10748 | 'PDIA3_No2' | 'ELSDFISYLQR' | 'Protein disulfide-isomerase A3' |
| 10749 | 'PDIA3_No3' | 'EATNPPVIQEEKPK' | 'Protein disulfide-isomerase A3' |
| 10750 | 'PDIA3_No4' | 'TFSHELSDFGLESTAGEIPVVAIR' | 'Protein disulfide-isomerase A3' |
| 10751 | 'PDIA3_No5' | 'KTFSHELSDFGLESTAGEIPVVAIR' | 'Protein disulfide-isomerase A3' |
| 10752 | 'PDIA3_No6' | 'FISDKDASIVGFFDDSFSEAHSEFLK' | 'Protein disulfide-isomerase A3' |
| 10753 | 'PDIA3_No7' | 'FIQENIFGICPHMTEDNKDLIQGK' | 'Protein disulfide-isomerase A3' |
| 10754 | 'PDIA3_No8' | 'MDATANDVPSPYEVR' | 'Protein disulfide-isomerase A3' |
| 10755 | 'PDIA3_No9' | 'KFIQENIFGICPHMTEDNKDLIQGK' | 'Protein disulfide-isomerase A3' |
| 10756 | 'PDIA3_No10' | 'FVMQEEFSR' | 'Protein disulfide-isomerase A3' |
| 10757 | 'PDIA3_No11' | 'LSKDPNIVIAK' | 'Protein disulfide-isomerase A3' |
| 10758 | 'PDIA3_No12' | 'IFRDGEEAGAYDGPR' | 'Protein disulfide-isomerase A3' |
| 10759 | 'PDIA3_No13' | 'TADGIVSHLK' | 'Protein disulfide-isomerase A3' |
| 10760 | 'PDIA3_No14' | 'QAGPASVPLRTEEEFKK' | 'Protein disulfide-isomerase A3' |
| 10761 | 'PDIA3_No15' | 'RLAPEYEAAATR' | 'Protein disulfide-isomerase A3' |
| 10762 | 'PDIA3_No16' | 'FLQDYFDGNLKR' | 'Protein disulfide-isomerase A3' |
| 10763 | 'PDIA3_No17' | 'GFPTIYFSPANK' | 'Protein disulfide-isomerase A3' |
| 10764 | 'PDIA3_No18' | 'MDATANDVPSPYEVR' | 'Protein disulfide-isomerase A3' |
| 10765 | 'PDIA3_No19' | 'FIQENIFGICPHMTEDNKDLIQGK' | 'Protein disulfide-isomerase A3' |
| 10766 | 'PDIA3_No20' | 'YLKSEPIPESNDGPVK' | 'Protein disulfide-isomerase A3' |
| 10767 | 'PPP2R1A' | 'IGPILDNSTLQSEVKPILEK' | 'Serine/threonine-protein phosphatase 2A 65 kDa regulatory subunit A alpha isoform' |
| 10768 | 'SERPINB1_No1' | 'TYNFLPEFLVSTQK' | 'Leukocyte elastase inhibitor' |
| 10769 | 'SERPINB1_No2' | 'LVLVNAIYFK' | 'Leukocyte elastase inhibitor' |
| 10770 | 'SERPINB1_No3' | 'FQSLNADINKR' | 'Leukocyte elastase inhibitor' |
| 10771 | 'SERPINB1_No4' | 'FAYGYIEDLK' | 'Leukocyte elastase inhibitor' |
| 10772 | 'SERPINB1_No5' | 'LGVQDLFNSSK' | 'Leukocyte elastase inhibitor' |
| 10773 | 'SERPINB1_No6' | 'TFHFNTVEEVHSR' | 'Leukocyte elastase inhibitor' |
| 10774 | 'SERPINB1_No7' | 'KIEEQLTLEK' | 'Leukocyte elastase inhibitor' |
| 10775 | 'SERPINB1_No8' | 'TYGADLASVDFQHASEDAR' | 'Leukocyte elastase inhibitor' |
| 10776 | 'SERPINB1_No9' | 'HNSSGSILFLGR' | 'Leukocyte elastase inhibitor' |
| 10777 | 'SERPINB1_No10' | 'GQTEGKIPELLASGMVDNMTK' | 'Leukocyte elastase inhibitor' |
| 10778 | 'SERPINB1_No11' | 'TYGADLASVDFQHASEDARK' | 'Leukocyte elastase inhibitor' |
| 10779 | 'SERPINB1_No12' | 'FKLEESYTLNSDLAR' | 'Leukocyte elastase inhibitor' |
| 10780 | 'CORO1A_No1' | 'AAPEASGTPSSDAVSR' | 'Coronin-1A' |
| 10781 | 'CORO1A_No2' | 'KSDLFQEDLYPPTAGPDPALTAEEWLGGR' | 'Coronin-1A' |

| | A | B | C |
|---|---|---|---|
| 1 | **Peptide Name** | **Peptide Sequence** | **Peptide Description** |
| 10782 | 'CORO1A_No3' | 'VSQTTWDSGFCAVNPK' | 'Coronin-1A' |
| 10783 | 'CORO1A_No4' | 'KLQATVQELQK' | 'Coronin-1A' |
| 10784 | 'CORO1A_No5' | 'RAAPEASGTPSSDAVSR' | 'Coronin-1A' |
| 10785 | 'CORO1A_No6' | 'DAGPLLISLKDGYVPPK' | 'Coronin-1A' |
| 10786 | 'CORO1A_No7' | 'RCEPIAMTVPR' | 'Coronin-1A' |
| 10787 | 'CORO1A_No8' | 'DGGLICTSCR' | 'Coronin-1A' |
| 10788 | 'GDI1_No1' | 'TDDYLDQPCLETVNR' | 'Rab GDP dissociation inhibitor alpha' |
| 10789 | 'GDI1_No2' | 'NPYYGGESSSITPLEELYK' | 'Rab GDP dissociation inhibitor alpha' |
| 10790 | 'GDI1_No3' | 'KSDIYVCMISYAHNVAAQGK' | 'Rab GDP dissociation inhibitor alpha' |
| 10791 | 'S100A7_No1' | 'SIIGMIDMFHK' | 'Protein S100-A7' |
| 10792 | 'S100A7_No2' | 'SIIGMIDMFHK' | 'Protein S100-A7' |
| 10793 | 'CASP14_No1' | 'GHILELLTEVTR' | 'Caspase-14' |
| 10794 | 'CASP14_No2' | 'SLEEEKYDMSGAR' | 'Caspase-14' |
| 10795 | 'CASP14_No3' | 'LALILCVTK' | 'Caspase-14' |
| 10796 | 'CASP14_No4' | 'LENLFEALNNK' | 'Caspase-14' |
| 10797 | 'CASP14_No5' | 'GHILELLTEVTRR' | 'Caspase-14' |
| 10798 | 'CASP14_No6' | 'MAEAELVQEGK' | 'Caspase-14' |
| 10799 | 'CASP14_No7' | 'AREGSEEDLDALEHMFR' | 'Caspase-14' |
| 10800 | 'CASP14_No8' | 'EDPVSCAFVVLMAHGR' | 'Caspase-14' |
| 10801 | 'CASP14_No9' | 'SLEEEKYDMSGAR' | 'Caspase-14' |
| 10802 | 'YWHAB_No1' | 'DSTLIMQLLR' | '14-3-3 protein beta/alpha' |
| 10803 | 'YWHAB_No2' | 'AVTEQGHELSNEER' | '14-3-3 protein beta/alpha' |
| 10804 | 'YWHAB_No3' | 'QTTVSNSQQAYQEAFEISK' | '14-3-3 protein beta/alpha' |
| 10805 | 'YWHAB_No4' | 'DSTLIMQLLR' | '14-3-3 protein beta/alpha' |
| 10806 | 'YWHAB_No5' | 'LAEQAERYDDMAAAMK' | '14-3-3 protein beta/alpha' |
| 10807 | 'YWHAB_No6' | 'NLLSVAYK' | '14-3-3 protein beta/alpha' |
| 10808 | 'YWHAB_No7' | 'QTTVSNSQQAYQEAFEISKK' | '14-3-3 protein beta/alpha' |
| 10809 | 'STIP1_No1' | 'ALSVGNIDDALQCYSEAIK' | 'Stress-induced-phosphoprotein 1' |
| 10810 | 'STIP1_No2' | 'LAYINPDLALEEK' | 'Stress-induced-phosphoprotein 1' |
| 10811 | 'STIP1_No3' | 'TYEEGLKHEANNPQLK' | 'Stress-induced-phosphoprotein 1' |
| 10812 | 'PRDX2_No1' | 'ATAVVDGAFKEVK' | 'Peroxiredoxin-2' |
| 10813 | 'PRDX2_No2' | 'GLFIIDGK' | 'Peroxiredoxin-2' |
| 10814 | 'PRDX2_No3' | 'KEGGLGPLNIPLLADVTR' | 'Peroxiredoxin-2' |
| 10815 | 'PRDX2_No4' | 'RLSEDYGVLK' | 'Peroxiredoxin-2' |
| 10816 | 'PRDX2_No5' | 'GLFIIDGKGVLR' | 'Peroxiredoxin-2' |
| 10817 | 'PRDX2_No6' | 'EGGLGPLNIPLLADVTRR' | 'Peroxiredoxin-2' |
| 10818 | 'PRDX2_No7' | 'LVQAFQYTDEHGEVCPAGWKPGSDTIKPNVDDSK' | 'Peroxiredoxin-2' |
| 10819 | 'PRDX2_No8' | 'LSEDYGVLKTDEGIAYR' | 'Peroxiredoxin-2' |
| 10820 | 'PRDX2_No9' | 'KEGGLGPLNIPLLADVTRR' | 'Peroxiredoxin-2' |
| 10821 | 'PRDX2_No10' | 'EGGLGPLNIPLLADVTR' | 'Peroxiredoxin-2' |
| 10822 | 'PRDX2_No11' | 'TDEGIAYR' | 'Peroxiredoxin-2' |
| 10823 | 'PRDX2_No12' | 'RLSEDYGVLKTDEGIAYR' | 'Peroxiredoxin-2' |
| 10824 | 'HPD' | 'FAVLQTYGDTTHTLVEK' | '4-hydroxyphenylpyruvate dioxygenase' |
| 10825 | 'ICAM3_No1' | 'TFVLPVTPPR' | 'Intercellular adhesion molecule 3' |

| | A | B | C |
|---|---|---|---|
| 1 | **Peptide Name** | **Peptide Sequence** | **Peptide Description** |
| 10826 | 'ICAM3_No2' | 'IALETSLSK' | 'Intercellular adhesion molecule 3' |
| 10827 | 'ICAM3_No3' | 'QPAVEEPAEVTATVLASR' | 'Intercellular adhesion molecule 3' |
| 10828 | 'ICAM3_No4' | 'WEEELSRQPAVEEPAEVTATVLASR' | 'Intercellular adhesion molecule 3' |
| 10829 | 'ICAM3_No5' | 'QPAVEEPAEVTATVLASR' | 'Intercellular adhesion molecule 3' |
| 10830 | 'CDH5_No1' | 'YLLKGEYVGK' | 'Cadherin-5' |
| 10831 | 'CDH5_No2' | 'VHFLPVVISDNGMPSR' | 'Cadherin-5' |
| 10832 | 'CDH5_No3' | 'KPLIGTVLAMDPDAAR' | 'Cadherin-5' |
| 10833 | 'CDH5_No4' | 'ESIVQVHIEVLDENDNAPEFAKPYQPK' | 'Cadherin-5' |
| 10834 | 'CDH5_No5' | 'VGTSVGSLFVEDPDEPQNR' | 'Cadherin-5' |
| 10835 | 'CDH5_No6' | 'VHDVNDNWPVFTHR' | 'Cadherin-5' |
| 10836 | 'CDH5_No7' | 'VCENAVHGQLVLQISAIDKDITPR' | 'Cadherin-5' |
| 10837 | 'CDH5_No8' | 'KPLIGTVLAMDPDAAR' | 'Cadherin-5' |
| 10838 | 'CDH5_No9' | 'YEIVVEAR' | 'Cadherin-5' |
| 10839 | 'CDH5_No10' | 'HSIGYSIR' | 'Cadherin-5' |
| 10840 | 'CDH5_No11' | 'YTFVVPEDTR' | 'Cadherin-5' |
| 10841 | 'CDH5_No12' | 'VHFLPVVISDNGMPSR' | 'Cadherin-5' |
| 10842 | 'CDH5_No13' | 'LDRENISEYHLTAVIVDKDTGENLETPSSFTIK' | 'Cadherin-5' |
| 10843 | 'CDH5_No14' | 'GKEYFAIDNSGR' | 'Cadherin-5' |
| 10844 | 'CDH5_No15' | 'ELDSTGTPTGK' | 'Cadherin-5' |
| 10845 | 'CDH5_No16' | 'EYFAIDNSGR' | 'Cadherin-5' |
| 10846 | 'CDH5_No17' | 'DTGENLETPSSFTIK' | 'Cadherin-5' |
| 10847 | 'CDH5_No18' | 'VDAETGDVFAIER' | 'Cadherin-5' |
| 10848 | 'CDH5_No19' | 'VFRVDAETGDVFAIER' | 'Cadherin-5' |
| 10849 | 'CDH5_No20' | 'GDYQDAFTIETNPAHNEGIIK' | 'Cadherin-5' |
| 10850 | 'MAN1A1_No1' | 'WAWEAVEALENHCR' | 'Mannosyl-oligosaccharide 1,2-alpha-mannosidase IA' |
| 10851 | 'MAN1A1_No2' | 'MYFDAVQAIETHLIR' | 'Mannosyl-oligosaccharide 1,2-alpha-mannosidase IA' |
| 10852 | 'MAN1A1_No3' | 'GLPPVDFVPPIGVESR' | 'Mannosyl-oligosaccharide 1,2-alpha-mannosidase IA' |
| 10853 | 'MAN1A1_No4' | 'KMYFDAVQAIETHLIR' | 'Mannosyl-oligosaccharide 1,2-alpha-mannosidase IA' |
| 10854 | 'MAN1A1_No5' | 'LLPAFHTPSGIPWALLNMK' | 'Mannosyl-oligosaccharide 1,2-alpha-mannosidase IA' |
| 10855 | 'MAN1A1_No6' | 'LLPAFHTPSGIPWALLNMK' | 'Mannosyl-oligosaccharide 1,2-alpha-mannosidase IA' |
| 10856 | 'MAN1A1_No7' | 'MYFDAVQAIETHLIR' | 'Mannosyl-oligosaccharide 1,2-alpha-mannosidase IA' |
| 10857 | 'MAN1A1_No8' | 'KWAWEAVEALENHCR' | 'Mannosyl-oligosaccharide 1,2-alpha-mannosidase IA' |
| 10858 | 'MAN1A1_No9' | 'KMYFDAVQAIETHLIR' | 'Mannosyl-oligosaccharide 1,2-alpha-mannosidase IA' |
| 10859 | 'MAN1A1_No10' | 'GGHSSSLFGNIK' | 'Mannosyl-oligosaccharide 1,2-alpha-mannosidase IA' |
| 10860 | 'MAN1A1_No11' | 'AWLMSDKTDLEAK' | 'Mannosyl-oligosaccharide 1,2-alpha-mannosidase IA' |
| 10861 | 'MAN1A1_No12' | 'KSSSGLTYIAEWK' | 'Mannosyl-oligosaccharide 1,2-alpha-mannosidase IA' |
| 10862 | 'MAN1A1_No13' | 'GYAWGLNELKPISK' | 'Mannosyl-oligosaccharide 1,2-alpha-mannosidase IA' |
| 10863 | 'MAN1A1_No14' | 'AWLMSDKTDLEAK' | 'Mannosyl-oligosaccharide 1,2-alpha-mannosidase IA' |
| 10864 | 'MAN1A1_No15' | 'FVGGLLSAYYLSGEEIFRK' | 'Mannosyl-oligosaccharide 1,2-alpha-mannosidase IA' |
| 10865 | 'MAN1A1_No16' | 'MYFDAVQAIETHLIRK' | 'Mannosyl-oligosaccharide 1,2-alpha-mannosidase IA' |
| 10866 | 'MAN1A1_No17' | 'MYFDAVQAIETHLIRK' | 'Mannosyl-oligosaccharide 1,2-alpha-mannosidase IA' |
| 10867 | 'MAN1A1_No18' | 'ETLQKLPEEIQR' | 'Mannosyl-oligosaccharide 1,2-alpha-mannosidase IA' |
| 10868 | 'MAN1A1_No19' | 'AWLMSDKTDLEAKK' | 'Mannosyl-oligosaccharide 1,2-alpha-mannosidase IA' |
| 10869 | 'RNASE4_No1' | 'SICSTTNIQCK' | 'Ribonuclease 4' |

| | A | B | C |
|---|---|---|---|
| 1 | **Peptide Name** | **Peptide Sequence** | **Peptide Description** |
| 10870 | 'RNASE4_No2' | 'FNTFIHEDIWNIR' | 'Ribonuclease 4' |
| 10871 | 'RNASE4_No3' | 'RVVIACEGNPQVPVHFDG' | 'Ribonuclease 4' |
| 10872 | 'RNASE4_No4' | 'RFNTFIHEDIWNIR' | 'Ribonuclease 4' |
| 10873 | 'RNASE4_No5' | 'QHVHPEETGGSDRYCNLMMQR' | 'Ribonuclease 4' |
| 10874 | 'RNASE4_No6' | 'VVIACEGNPQVPVHFDG' | 'Ribonuclease 4' |
| 10875 | 'PRSS3' | 'VLEGNEQFINAAK' | 'Trypsin-3' |
| 10876 | 'SERPINB6_No1' | 'SCDFLSSFR' | 'Serpin B6' |
| 10877 | 'SERPINB6_No2' | 'GNTAAQMAQILSFNK' | 'Serpin B6' |
| 10878 | 'SERPINB6_No3' | 'FCADHPFLFFIQHSK' | 'Serpin B6' |
| 10879 | 'SERPINB6_No4' | 'IAELLSPGSVDPLTR' | 'Serpin B6' |
| 10880 | 'SERPINB6_No5' | 'SGGGGDIHQGFQSLLTEVNK' | 'Serpin B6' |
| 10881 | 'SERPINB6_No6' | 'GNWDEQFDKENTEER' | 'Serpin B6' |
| 10882 | 'SFTPD_No1' | 'QQVEALQGQVQHLQAAFSQYK' | 'Pulmonary surfactant-associated protein D' |
| 10883 | 'SFTPD_No2' | 'TAGFVKPFTEAQLLCTQAGGQLASPR' | 'Pulmonary surfactant-associated protein D' |
| 10884 | 'SFTPD_No3' | 'SAAENAALQQLVVAK' | 'Pulmonary surfactant-associated protein D' |
| 10885 | 'THBS4_No1' | 'WFLQHRPQVGYIR' | 'Thrombospondin-4' |
| 10886 | 'THBS4_No2' | 'NPELNPCSVNAQCIEER' | 'Thrombospondin-4' |
| 10887 | 'THBS4_No3' | 'GAGSLELYLDCIQVDSVHNLPR' | 'Thrombospondin-4' |
| 10888 | 'THBS4_No4' | 'CGPCKPGYTGDQIR' | 'Thrombospondin-4' |
| 10889 | 'THBS4_No5' | 'NSDKDIFGDACDNCLSVLNNDQK' | 'Thrombospondin-4' |
| 10890 | 'THBS4_No6' | 'IDVCPENAEVTLTDFR' | 'Thrombospondin-4' |
| 10891 | 'THBS4_No7' | 'QVCTDIDECR' | 'Thrombospondin-4' |
| 10892 | 'THBS4_No8' | 'NCRNPELNPCSVNAQCIEER' | 'Thrombospondin-4' |
| 10893 | 'THBS4_No9' | 'YHPCYPGVHCINLSPGFR' | 'Thrombospondin-4' |
| 10894 | 'THBS4_No10' | 'QFLGQMTQLNQLLGEVK' | 'Thrombospondin-4' |
| 10895 | 'THBS4_No11' | 'KPQDFLEELK' | 'Thrombospondin-4' |
| 10896 | 'THBS4_No12' | 'DVDIDSYPDEELPCSAR' | 'Thrombospondin-4' |
| 10897 | 'THBS4_No13' | 'CDACPVGFTGPMVQGVGISFAK' | 'Thrombospondin-4' |
| 10898 | 'THBS4_No14' | 'AFAGPSQKPETIELR' | 'Thrombospondin-4' |
| 10899 | 'THBS4_No15' | 'LGVFCFSQENIIWSNLK' | 'Thrombospondin-4' |
| 10900 | 'THBS4_No16' | 'QFLGQMTQLNQLLGEVK' | 'Thrombospondin-4' |
| 10901 | 'THBS4_No17' | 'CDACPVGFTGPMVQGVGISFAK' | 'Thrombospondin-4' |
| 10902 | 'THBS4_No18' | 'SSATIFGLYSSTDNSK' | 'Thrombospondin-4' |
| 10903 | 'THBS4_No19' | 'QFLGQMTQLNQLLGEVK' | 'Thrombospondin-4' |
| 10904 | 'THBS4_No20' | 'NSLWHTGDTSDQVR' | 'Thrombospondin-4' |
| 10905 | 'THBS4_No21' | 'QGDVTCVCGVGWAGDGYICGK' | 'Thrombospondin-4' |
| 10906 | 'KRT9_No1' | 'MTLDDFR' | 'Keratin, type I cytoskeletal 9' |
| 10907 | 'KRT9_No2' | 'TLNDMRQEYEQLIAK' | 'Keratin, type I cytoskeletal 9' |
| 10908 | 'KRT9_No3' | 'TLLDIDNTR' | 'Keratin, type I cytoskeletal 9' |
| 10909 | 'KRT9_No4' | 'IKFEMEQNLR' | 'Keratin, type I cytoskeletal 9' |
| 10910 | 'KRT9_No5' | 'IKFEMEQNLR' | 'Keratin, type I cytoskeletal 9' |
| 10911 | 'KRT9_No6' | 'TLNDMRQEYEQLIAK' | 'Keratin, type I cytoskeletal 9' |
| 10912 | 'KRT9_No7' | 'QFSSSYLSR' | 'Keratin, type I cytoskeletal 9' |
| 10913 | 'KRT9_No8' | 'HGVQELEIELQSQLSK' | 'Keratin, type I cytoskeletal 9' |

| | A | B | C |
|---|---|---|---|
| 1 | **Peptide Name** | **Peptide Sequence** | **Peptide Description** |
| 10914 | 'KRT9_No9' | 'VQALEEANNDLENKIQDWYDK' | 'Keratin, type I cytoskeletal 9' |
| 10915 | 'KRT9_No10' | 'QVLDNLTMEK' | 'Keratin, type I cytoskeletal 9' |
| 10916 | 'KRT9_No11' | 'QGVDADINGLR' | 'Keratin, type I cytoskeletal 9' |
| 10917 | 'KRT9_No12' | 'VQALEEANNDLENKIQDWYDKK' | 'Keratin, type I cytoskeletal 9' |
| 10918 | 'KRT9_No13' | 'MTLDDFR' | 'Keratin, type I cytoskeletal 9' |
| 10919 | 'KRT9_No14' | 'FSSSSGYGGGSSR' | 'Keratin, type I cytoskeletal 9' |
| 10920 | 'KRT9_No15' | 'GGSGGSYGGGGSGGGYGGGSGSR' | 'Keratin, type I cytoskeletal 9' |
| 10921 | 'KRT9_No16' | 'HGVQELEIELQSQLSKK' | 'Keratin, type I cytoskeletal 9' |
| 10922 | 'KRT9_No17' | 'EIETYHNLLEGGQEDFESSGAGK' | 'Keratin, type I cytoskeletal 9' |
| 10923 | 'KRT9_No18' | 'SGGGGGGGLGSGGSIR' | 'Keratin, type I cytoskeletal 9' |
| 10924 | 'KRT9_No19' | 'SDLEMQYETLQEELMALKK' | 'Keratin, type I cytoskeletal 9' |
| 10925 | 'KRT9_No20' | 'GGGGSFGYSYGGGSGGGFSASSLGGGFGGGSR' | 'Keratin, type I cytoskeletal 9' |
| 10926 | 'KRT9_No21' | 'NHKEEMSQLTGQNSGDVNVEINVAPGK' | 'Keratin, type I cytoskeletal 9' |
| 10927 | 'KRT9_No22' | 'SDLEMQYETLQEELMALKK' | 'Keratin, type I cytoskeletal 9' |
| 10928 | 'KRT9_No23' | 'LASYLDKVQALEEANNDLENK' | 'Keratin, type I cytoskeletal 9' |
| 10929 | 'KRT9_No24' | 'SDLEMQYETLQEELMALKK' | 'Keratin, type I cytoskeletal 9' |
| 10930 | 'KRT9_No25' | 'LASYLDKVQALEEANNDLENKIQDWYDK' | 'Keratin, type I cytoskeletal 9' |
| 10931 | 'KRT9_No26' | 'FEMEQNLR' | 'Keratin, type I cytoskeletal 9' |
| 10932 | 'KRT9_No27' | 'NHKEEMSQLTGQNSGDVNVEINVAPGK' | 'Keratin, type I cytoskeletal 9' |
| 10933 | 'KRT9_No28' | 'VQALEEANNDLENK' | 'Keratin, type I cytoskeletal 9' |
| 10934 | 'KRT9_No29' | 'SDLEMQYETLQEELMALK' | 'Keratin, type I cytoskeletal 9' |
| 10935 | 'KRT9_No30' | 'NYSPYYNTIDDLKDQIVDLTVGNNK' | 'Keratin, type I cytoskeletal 9' |
| 10936 | 'KRT9_No31' | 'QEYEQLIAK' | 'Keratin, type I cytoskeletal 9' |
| 10937 | 'KRT9_No32' | 'SDLEMQYETLQEELMALKK' | 'Keratin, type I cytoskeletal 9' |
| 10938 | 'KRT9_No33' | 'STMQELNSR' | 'Keratin, type I cytoskeletal 9' |
| 10939 | 'KRT9_No34' | 'DIENQYETQITQIEHEVSSSGQEVQSSAK' | 'Keratin, type I cytoskeletal 9' |
| 10940 | 'KRT9_No35' | 'QVLDNLTMEK' | 'Keratin, type I cytoskeletal 9' |
| 10941 | 'KRT9_No36' | 'STMQELNSR' | 'Keratin, type I cytoskeletal 9' |
| 10942 | 'KRT9_No37' | 'QEIECQNQEYSLLLSIK' | 'Keratin, type I cytoskeletal 9' |
| 10943 | 'KRT9_No38' | 'QVLDNLTMEKSDLEMQYETLQEELMALKK' | 'Keratin, type I cytoskeletal 9' |
| 10944 | 'KRT9_No39' | 'KDIENQYETQITQIEHEVSSSGQEVQSSAK' | 'Keratin, type I cytoskeletal 9' |
| 10945 | 'KRT9_No40' | 'LEKEIETYHNLLEGGQEDFESSGAGK' | 'Keratin, type I cytoskeletal 9' |
| 10946 | 'KRT9_No41' | 'IGLGGRGGSGGSYGR' | 'Keratin, type I cytoskeletal 9' |
| 10947 | 'KRT9_No42' | 'NHKEEMSQLTGQNSGDVNVEINVAPGKDLTK' | 'Keratin, type I cytoskeletal 9' |
| 10948 | 'KRT9_No43' | 'NHKEEMSQLTGQNSGDVNVEINVAPGKDLTK' | 'Keratin, type I cytoskeletal 9' |
| 10949 | 'KRT9_No44' | 'IQDWYDKKGPAAIQK' | 'Keratin, type I cytoskeletal 9' |
| 10950 | 'KRT9_No45' | 'QVLDNLTMEK' | 'Keratin, type I cytoskeletal 9' |
| 10951 | 'KRT9_No46' | 'NRKDIENQYETQITQIEHEVSSSGQEVQSSAK' | 'Keratin, type I cytoskeletal 9' |
| 10952 | 'KRT9_No47' | 'DIENQYETQITQIEHEVSSSGQEVQSSAKEVTQLR' | 'Keratin, type I cytoskeletal 9' |
| 10953 | 'KRT9_No48' | 'FEMEQNLR' | 'Keratin, type I cytoskeletal 9' |
| 10954 | 'KRT9_No49' | 'QGVDADINGLR' | 'Keratin, type I cytoskeletal 9' |
| 10955 | 'KRT9_No50' | 'SDLEMQYETLQEELMALK' | 'Keratin, type I cytoskeletal 9' |
| 10956 | 'KRT9_No51' | 'SDLEMQYETLQEELMALK' | 'Keratin, type I cytoskeletal 9' |
| 10957 | 'KRT9_No52' | 'MRLEKEIETYHNLLEGGQEDFESSGAGK' | 'Keratin, type I cytoskeletal 9' |

| | A | B | C |
|---|---|---|---|
| 1 | **Peptide Name** | **Peptide Sequence** | **Peptide Description** |
| 10958 | 'KRT9_No53' | 'GGSGGSHGGGSGFGGESGGSYGGGEEASGSGGGYGGGSGKSSHS' | 'Keratin, type I cytoskeletal 9' |
| 10959 | 'KRT9_No54' | 'MRLEKEIETYHNLLEGGQEDFESSGAGK' | 'Keratin, type I cytoskeletal 9' |
| 10960 | 'KRT9_No55' | 'TLLDIDNTRMTLDDFR' | 'Keratin, type I cytoskeletal 9' |
| 10961 | 'KRT9_No56' | 'HGVQELEIELQSQLSKKAALEK' | 'Keratin, type I cytoskeletal 9' |
| 10962 | 'KRT9_No57' | 'QGVDADINGLRQVLDNLTMEK' | 'Keratin, type I cytoskeletal 9' |
| 10963 | 'KRT9_No58' | 'FSSSGGGGGGGRFSSSSGYGGGSSR' | 'Keratin, type I cytoskeletal 9' |
| 10964 | 'KRT9_No59' | 'GGSGGSHGGGSGFGGESGGSYGGGEEASGSGGGYGGGSGK' | 'Keratin, type I cytoskeletal 9' |
| 10965 | 'KRT9_No60' | 'GSRGGSGGSYGGGGSGGGYGGGSGSR' | 'Keratin, type I cytoskeletal 9' |
| 10966 | 'KRT9_No61' | 'MTLDDFRIK' | 'Keratin, type I cytoskeletal 9' |
| 10967 | 'KRT9_No62' | 'NYSPYYNTIDDLK' | 'Keratin, type I cytoskeletal 9' |
| 10968 | 'KRT9_No63' | 'FSSSGGGGGGGR' | 'Keratin, type I cytoskeletal 9' |
| 10969 | 'KRT9_No64' | 'SGGGGGGGLGSGGSIRSSYSR' | 'Keratin, type I cytoskeletal 9' |
| 10970 | 'SAA4_No1' | 'AYWDIMISNHQNSNR' | 'Serum amyloid A-4 protein' |
| 10971 | 'SAA4_No2' | 'FRPDGLPK' | 'Serum amyloid A-4 protein' |
| 10972 | 'SAA4_No3' | 'EALQGVGDMGR' | 'Serum amyloid A-4 protein' |
| 10973 | 'SAA4_No4' | 'AYWDIMISNHQNSNR' | 'Serum amyloid A-4 protein' |
| 10974 | 'SAA4_No5' | 'EALQGVGDMGR' | 'Serum amyloid A-4 protein' |
| 10975 | 'SAA4_No6' | 'SFFKEALQGVGDMGR' | 'Serum amyloid A-4 protein' |
| 10976 | 'SAA4_No7' | 'SFFKEALQGVGDMGR' | 'Serum amyloid A-4 protein' |
| 10977 | 'SAA4_No8' | 'GPGGVWAAK' | 'Serum amyloid A-4 protein' |
| 10978 | 'SAA4_No9' | 'GNYDAAQR' | 'Serum amyloid A-4 protein' |
| 10979 | 'SAA4_No10' | 'SNEKAEEWGR' | 'Serum amyloid A-4 protein' |
| 10980 | 'SAA4_No11' | 'GPGGVWAAKLISR' | 'Serum amyloid A-4 protein' |
| 10981 | 'SAA4_No12' | 'GNYDAAQRGPGGVWAAK' | 'Serum amyloid A-4 protein' |
| 10982 | 'SAA4_No13' | 'GNYDAAQRGPGGVWAAKLISR' | 'Serum amyloid A-4 protein' |
| 10983 | 'SAA4_No14' | 'DPDRFRPDGLPK' | 'Serum amyloid A-4 protein' |
| 10984 | 'FBN1_No1' | 'TGCTDINECEIGAHNCGK' | 'Fibrillin-1' |
| 10985 | 'FBN1_No2' | 'YQCACNPGYHSTPDR' | 'Fibrillin-1' |
| 10986 | 'FBN1_No3' | 'GYIGTHCGQPVCESGCLNGGR' | 'Fibrillin-1' |
| 10987 | 'FBN1_No4' | 'NCMDIDECQRDPLLCR' | 'Fibrillin-1' |
| 10988 | 'FBN1_No5' | 'SGNCYLDIRPR' | 'Fibrillin-1' |
| 10989 | 'FBN1_No6' | 'GDNGDTACSNEIGVGVSK' | 'Fibrillin-1' |
| 10990 | 'FBN1_No7' | 'RPDGEGCVDENECQTKPGICENGR' | 'Fibrillin-1' |
| 10991 | 'FBN1_No8' | 'TLPGGNQCIVPICR' | 'Fibrillin-1' |
| 10992 | 'FBN1_No9' | 'EIPGVCENGVCINMVGSFR' | 'Fibrillin-1' |
| 10993 | 'FBN1_No10' | 'IKGTQCEDIDECEVFPGVCK' | 'Fibrillin-1' |
| 10994 | 'FBN1_No11' | 'MCCCSYNIGR' | 'Fibrillin-1' |
| 10995 | 'FBN1_No12' | 'EGYCFTEVLQNMCQIGSSNR' | 'Fibrillin-1' |
| 10996 | 'FBN1_No13' | 'TNDDQTMCLDINECER' | 'Fibrillin-1' |
| 10997 | 'FBN1_No14' | 'TECRDIDECLQNGR' | 'Fibrillin-1' |
| 10998 | 'FBN1_No15' | 'CPTGYYLNEDTR' | 'Fibrillin-1' |
| 10999 | 'FBN1_No16' | 'GQCIKPLFGAVTK' | 'Fibrillin-1' |
| 11000 | 'FBN1_No17' | 'CECFPGLAVGLDGR' | 'Fibrillin-1' |
| 11001 | 'FBN1_No18' | 'TCVDINECLLEPR' | 'Fibrillin-1' |

| | A | B | C |
|---|---|---|---|
| 1 | **Peptide Name** | **Peptide Sequence** | **Peptide Description** |
| 11002 | 'FBN1_No19' | 'CACTYGFTGPQCER' | 'Fibrillin-1' |
| 11003 | 'FBN1_No20' | 'CICNSGYEVDSTGK' | 'Fibrillin-1' |
| 11004 | 'FBN1_No21' | 'CQCPSGMTLDATGR' | 'Fibrillin-1' |
| 11005 | 'FBN1_No22' | 'CECPVGFFYNDK' | 'Fibrillin-1' |
| 11006 | 'FBN1_No23' | 'YEDEECTLPIAGR' | 'Fibrillin-1' |
| 11007 | 'FBN1_No24' | 'NGECVNDRGSYHCICK' | 'Fibrillin-1' |
| 11008 | 'FBN1_No25' | 'QICPYGSGIIVGPDDSAVDMDECKEPDVCK' | 'Fibrillin-1' |
| 11009 | 'FBN1_No26' | 'ILELLPALTTLTNHNR' | 'Fibrillin-1' |
| 11010 | 'FBN1_No27' | 'CSCSPGWIGDGIK' | 'Fibrillin-1' |
| 11011 | 'FBN1_No28' | 'MCCCSYNIGR' | 'Fibrillin-1' |
| 11012 | 'MYH9_No1' | 'QLLQANPILEAFGNAK' | 'Myosin-9' |
| 11013 | 'MYH9_No2' | 'VISGVLQLGNIVFK' | 'Myosin-9' |
| 11014 | 'MYH9_No3' | 'VSHLLGINVTDFTR' | 'Myosin-9' |
| 11015 | 'MYH9_No4' | 'ADFCIIHYAGK' | 'Myosin-9' |
| 11016 | 'MYH9_No5' | 'LDPHLVLDQLR' | 'Myosin-9' |
| 11017 | 'MYH9_No6' | 'NTDQASMPDNTAAQK' | 'Myosin-9' |
| 11018 | 'MYH9_No7' | 'KFDQLLAEEK' | 'Myosin-9' |
| 11019 | 'MYH9_No8' | 'VIQYLAYVASSHK' | 'Myosin-9' |
| 11020 | 'MYH9_No9' | 'TFHIFYYLLSGAGEHLK' | 'Myosin-9' |
| 11021 | 'MYH9_No10' | 'IRELESQISELQEDLESER' | 'Myosin-9' |
| 11022 | 'MYH9_No11' | 'RGDLPFVVPR' | 'Myosin-9' |
| 11023 | 'MYH9_No12' | 'QEEEMMAKEEELVK' | 'Myosin-9' |
| 11024 | 'MYH9_No13' | 'NKHEAMITDLEER' | 'Myosin-9' |
| 11025 | 'MYH9_No14' | 'NFINNPLAQADWAAK' | 'Myosin-9' |
| 11026 | 'MYH9_No15' | 'IAQLEEQLDNETKER' | 'Myosin-9' |
| 11027 | 'MYH9_No16' | 'VEDMAELTCLNEASVLHNLK' | 'Myosin-9' |
| 11028 | 'MYH9_No17' | 'LQQELDDLLVDLDHQR' | 'Myosin-9' |
| 11029 | 'MYH9_No18' | 'IMGIPEEEQMGLLR' | 'Myosin-9' |
| 11030 | 'MYH9_No19' | 'TEMEDLMSSKDDVGK' | 'Myosin-9' |
| 11031 | 'MYH9_No20' | 'IAEFTTNLTEEEEK' | 'Myosin-9' |
| 11032 | 'MYH9_No21' | 'KKVEAQLQELQVK' | 'Myosin-9' |
| 11033 | 'MYH9_No22' | 'KEEELQAALAR' | 'Myosin-9' |
| 11034 | 'MYH9_No23' | 'ALEEAMEQKAELER' | 'Myosin-9' |
| 11035 | 'MYH9_No24' | 'KVEAQLQELQVK' | 'Myosin-9' |
| 11036 | 'MYH9_No25' | 'DFSALESQLQDTQELLQEENRQK' | 'Myosin-9' |
| 11037 | 'MYH9_No26' | 'LTEMETLQSQLMAEK' | 'Myosin-9' |
| 11038 | 'MYH9_No27' | 'LEVNLQAMK' | 'Myosin-9' |
| 11039 | 'MYH9_No28' | 'EMEAELEDER' | 'Myosin-9' |
| 11040 | 'MYH9_No29' | 'VKPLLQVSR' | 'Myosin-9' |
| 11041 | 'MYH9_No30' | 'KVIQYLAYVASSHK' | 'Myosin-9' |
| 11042 | 'MYH9_No31' | 'ALELDSNLYR' | 'Myosin-9' |
| 11043 | 'MYH9_No32' | 'HSQAVEELAEQLEQTKR' | 'Myosin-9' |
| 11044 | 'MYH9_No33' | 'AGVLAHLEEERDLK' | 'Myosin-9' |
| 11045 | 'MYH9_No34' | 'EQLEEEEEAKHNLEK' | 'Myosin-9' |

| | A | B | C |
|---|---|---|---|
| 1 | **Peptide Name** | **Peptide Sequence** | **Peptide Description** |
| 11046 | 'MYH9_No35' | 'QIATLHAQVADMK' | 'Myosin-9' |
| 11047 | 'MYH9_No36' | 'TQLEELEDELQATEDAK' | 'Myosin-9' |
| 11048 | 'MYH9_No37' | 'RKLEGDSTDLSDQIAELQAQIAELK' | 'Myosin-9' |
| 11049 | 'MYH9_No38' | 'AGVLAHLEEER' | 'Myosin-9' |
| 11050 | 'MYH9_No39' | 'EMEAELEDER' | 'Myosin-9' |
| 11051 | 'MYH9_No40' | 'TVGQLYKEQLAK' | 'Myosin-9' |
| 11052 | 'MYH9_No41' | 'IIGLDQVAGMSETALPGAFK' | 'Myosin-9' |
| 11053 | 'MYH9_No42' | 'AQQAADKYLYVDKNFINNPLAQADWAAK' | 'Myosin-9' |
| 11054 | 'MYH9_No43' | 'SMEAEMIQLQEELAAAER' | 'Myosin-9' |
| 11055 | 'MYH9_No44' | 'MQQNIQELEEQLEEEESAR' | 'Myosin-9' |
| 11056 | 'MYH9_No45' | 'KLEEEQIILEDQNCK' | 'Myosin-9' |
| 11057 | 'MYH9_No46' | 'LQQLFNHTMFILEQEEYQR' | 'Myosin-9' |
| 11058 | 'MYH9_No47' | 'TLEEEAKTHEAQIQEMR' | 'Myosin-9' |
| 11059 | 'MYH9_No48' | 'HEAMITDLEER' | 'Myosin-9' |
| 11060 | 'MYH9_No49' | 'ASREEILAQAKENEK' | 'Myosin-9' |
| 11061 | 'MYH9_No50' | 'VISGVLQLGNIVFKK' | 'Myosin-9' |
| 11062 | 'MYH9_No51' | 'VEEEEERCQHLQAEK' | 'Myosin-9' |
| 11063 | 'MYH9_No52' | 'NMDPLNDNIATLLHQSSDK' | 'Myosin-9' |
| 11064 | 'MYH9_No53' | 'ELEDATETADAMNR' | 'Myosin-9' |
| 11065 | 'MYH9_No54' | 'NLPIYSEEIVEMYK' | 'Myosin-9' |
| 11066 | 'MYH9_No55' | 'LKDVLLQVDDERR' | 'Myosin-9' |
| 11067 | 'MYH9_No56' | 'IMGIPEEEQMGLLR' | 'Myosin-9' |
| 11068 | 'MYH9_No57' | 'VDYKADEWLMK' | 'Myosin-9' |
| 11069 | 'MYH9_No58' | 'IAEFTTNLTEEEEKSK' | 'Myosin-9' |
| 11070 | 'MYH9_No59' | 'QEEEMMAKEEELVK' | 'Myosin-9' |
| 11071 | 'MYH9_No60' | 'EMEAELEDERKQR' | 'Myosin-9' |
| 11072 | 'MYH9_No61' | 'FSKVEDMAELTCLNEASVLHNLKER' | 'Myosin-9' |
| 11073 | 'MYH9_No62' | 'QTLENERGELANEVK' | 'Myosin-9' |
| 11074 | 'MYH9_No63' | 'QIATLHAQVADMK' | 'Myosin-9' |
| 11075 | 'MYH9_No64' | 'RQLEEAEEEAQR' | 'Myosin-9' |
| 11076 | 'MYH9_No65' | 'QLLQANPILEAFGNAK' | 'Myosin-9' |
| 11077 | 'MYH9_No66' | 'DKADFCIIHYAGK' | 'Myosin-9' |
| 11078 | 'MYH9_No67' | 'TDLLLEPYNKYR' | 'Myosin-9' |
| 11079 | 'MYH9_No68' | 'LEEEQIILEDQNCK' | 'Myosin-9' |
| 11080 | 'MYH9_No69' | 'IAQLEEELEEEQGNTELINDRLK' | 'Myosin-9' |
| 11081 | 'MYH9_No70' | 'EMEAELEDERKQR' | 'Myosin-9' |
| 11082 | 'MYH9_No71' | 'QLEEAEEEAQR' | 'Myosin-9' |
| 11083 | 'MYH9_No72' | 'SMMQDREDQSILCTGESGAGK' | 'Myosin-9' |
| 11084 | 'MYH9_No73' | 'IIGLDQVAGMSETALPGAFK' | 'Myosin-9' |
| 11085 | 'MYH9_No74' | 'MQQNIQELEEQLEEEESAR' | 'Myosin-9' |
| 11086 | 'MYH9_No75' | 'TRLQQELDDLLVDLDHQR' | 'Myosin-9' |
| 11087 | 'MYH9_No76' | 'ANLQIDQINTDLNLER' | 'Myosin-9' |
| 11088 | 'MYH9_No77' | 'SGFEPASLKEEVGEEAIVELVENGKK' | 'Myosin-9' |
| 11089 | 'MYH9_No78' | 'KQELEEICHDLEAR' | 'Myosin-9' |

| | A | B | C |
|---|---|---|---|
| 1 | **Peptide Name** | **Peptide Sequence** | **Peptide Description** |
| 11090 | 'MYH9_No79' | 'SMMQDREDQSILCTGESGAGK' | 'Myosin-9' |
| 11091 | 'MYH9_No80' | 'LKNKHEAMITDLEER' | 'Myosin-9' |
| 11092 | 'MYH9_No81' | 'DVLLQVDDERR' | 'Myosin-9' |
| 11093 | 'MYH9_No82' | 'YAEERDRAEAEAR' | 'Myosin-9' |
| 11094 | 'MYH9_No83' | 'FVSELWK' | 'Myosin-9' |
| 11095 | 'MYH9_No84' | 'VKLQEMEGTVK' | 'Myosin-9' |
| 11096 | 'MYH9_No85' | 'TEMEDLMSSKDDVGK' | 'Myosin-9' |
| 11097 | 'MYH9_No86' | 'NKHEAMITDLEER' | 'Myosin-9' |
| 11098 | 'MYH9_No87' | 'MEDSVGCLETAEEVKR' | 'Myosin-9' |
| 11099 | 'MYH9_No88' | 'TLEEEAKTHEAQIQEMR' | 'Myosin-9' |
| 11100 | 'MYH9_No89' | 'ALEQQVEEMK' | 'Myosin-9' |
| 11101 | 'MYH9_No90' | 'EDQSILCTGESGAGKTENTKK' | 'Myosin-9' |
| 11102 | 'MYH9_No91' | 'NSFREQLEEEEEAKHNLEK' | 'Myosin-9' |
| 11103 | 'MYH9_No92' | 'KAGKLDPHLVLDQLR' | 'Myosin-9' |
| 11104 | 'MYH9_No93' | 'IAQLEEELEEEQGNTELINDRLKK' | 'Myosin-9' |
| 11105 | 'MYH9_No94' | 'NTDQASMPDNTAAQK' | 'Myosin-9' |
| 11106 | 'MYH9_No95' | 'FLSNGHVTIPGQQDKDMFQETMEAMR' | 'Myosin-9' |
| 11107 | 'MYH9_No96' | 'LKKLEEEQIILEDQNCK' | 'Myosin-9' |
| 11108 | 'MYH9_No97' | 'NLPIYSEEIVEMYK' | 'Myosin-9' |
| 11109 | 'MYH9_No98' | 'AQQAADKYLYVDK' | 'Myosin-9' |
| 11110 | 'MYH9_No99' | 'LTKDFSALESQLQDTQELLQEENR' | 'Myosin-9' |
| 11111 | 'MYH9_No100' | 'LQVELDNVTGLLSQSDSK' | 'Myosin-9' |
| 11112 | 'MYH9_No101' | 'QKHSQAVEELAEQLEQTKR' | 'Myosin-9' |
| 11113 | 'MYH9_No102' | 'LTEMETLQSQLMAEK' | 'Myosin-9' |
| 11114 | 'MYH9_No103' | 'TEMEDLMSSKDDVGK' | 'Myosin-9' |
| 11115 | 'MYH9_No104' | 'DLEAHIDSANKNRDEAIK' | 'Myosin-9' |
| 11116 | 'MYH9_No105' | 'QAQQERDELADEIANSSGK' | 'Myosin-9' |
| 11117 | 'MYH9_No106' | 'MEDSVGCLETAEEVK' | 'Myosin-9' |
| 11118 | 'MYH9_No107' | 'QKHSQAVEELAEQLEQTKR' | 'Myosin-9' |
| 11119 | 'MYH9_No108' | 'VEDMAELTCLNEASVLHNLKER' | 'Myosin-9' |
| 11120 | 'MYH9_No109' | 'ERNTDQASMPDNTAAQK' | 'Myosin-9' |
| 11121 | 'MYH9_No110' | 'IAQLEEELEEEQGNTELINDR' | 'Myosin-9' |
| 11122 | 'MYH9_No111' | 'LTEMETLQSQLMAEK' | 'Myosin-9' |
| 11123 | 'MYH9_No112' | 'QIATLHAQVADMK' | 'Myosin-9' |
| 11124 | 'MYH9_No113' | 'TEMEDLMSSKDDVGK' | 'Myosin-9' |
| 11125 | 'MYH9_No114' | 'QRYEILTPNSIPK' | 'Myosin-9' |
| 11126 | 'MYH9_No115' | 'VEDMAELTCLNEASVLHNLK' | 'Myosin-9' |
| 11127 | 'MYH9_No116' | 'ALEEAMEQKAELER' | 'Myosin-9' |
| 11128 | 'MYH9_No117' | 'AKQTLENERGELANEVK' | 'Myosin-9' |
| 11129 | 'MYH9_No118' | 'IMGIPEEEQMGLLR' | 'Myosin-9' |
| 11130 | 'MYH9_No119' | 'QLEEAEEEAQRANASR' | 'Myosin-9' |
| 11131 | 'MYH9_No120' | 'QAQQERDELADEIANSSGK' | 'Myosin-9' |
| 11132 | 'MYH9_No121' | 'HEMPPHIYAITDTAYR' | 'Myosin-9' |
| 11133 | 'MYH9_No122' | 'VDYKADEWLMK' | 'Myosin-9' |

| | A | B | C |
|---|---|---|---|
| 1 | **Peptide Name** | **Peptide Sequence** | **Peptide Description** |
| 11134 | 'MYH9_No123' | 'VEDMAELTCLNEASVLHNLKER' | 'Myosin-9' |
| 11135 | 'MYH9_No124' | 'DFSALESQLQDTQELLQEENR' | 'Myosin-9' |
| 11136 | 'MYH9_No125' | 'HEMPPHIYAITDTAYR' | 'Myosin-9' |
| 11137 | 'MYH9_No126' | 'FLSNGHVTIPGQQDKDMFQETMEAMR' | 'Myosin-9' |
| 11138 | 'MYH9_No127' | 'KEEELQAALARVEEEAAQK' | 'Myosin-9' |
| 11139 | 'MYH9_No128' | 'LVWVPSDK' | 'Myosin-9' |
| 11140 | 'MYH9_No129' | 'LKDVLLQVDDER' | 'Myosin-9' |
| 11141 | 'MYH9_No130' | 'ELEDATETADAMNR' | 'Myosin-9' |
| 11142 | 'MYH9_No131' | 'NMDPLNDNIATLLHQSSDK' | 'Myosin-9' |
| 11143 | 'MYH9_No132' | 'ELESQISELQEDLESER' | 'Myosin-9' |
| 11144 | 'MYH9_No133' | 'EMEAELEDERK' | 'Myosin-9' |
| 11145 | 'MYH9_No134' | 'LLEDRIAEFTTNLTEEEEKSK' | 'Myosin-9' |
| 11146 | 'MYH9_No135' | 'HSQAVEELAEQLEQTK' | 'Myosin-9' |
| 11147 | 'MYH9_No136' | 'KMEDSVGCLETAEEVKR' | 'Myosin-9' |
| 11148 | 'MYH9_No137' | 'ELEDATETADAMNREVSSLK' | 'Myosin-9' |
| 11149 | 'MYH9_No138' | 'ITDVIIGFQACCR' | 'Myosin-9' |
| 11150 | 'MYH9_No139' | 'ALEEAMEQKAELERLNK' | 'Myosin-9' |
| 11151 | 'MYH9_No140' | 'HEAMITDLEER' | 'Myosin-9' |
| 11152 | 'MYH9_No141' | 'RQAQQERDELADEIANSSGK' | 'Myosin-9' |
| 11153 | 'MYH9_No142' | 'MEDSVGCLETAEEVK' | 'Myosin-9' |
| 11154 | 'MYH9_No143' | 'MEDSVGCLETAEEVKR' | 'Myosin-9' |
| 11155 | 'MYH9_No144' | 'LTKDFSALESQLQDTQELLQEENRQK' | 'Myosin-9' |
| 11156 | 'MYH9_No145' | 'AGKLDPHLVLDQLR' | 'Myosin-9' |
| 11157 | 'MYH9_No146' | 'RQLEEAEEEAQRANASR' | 'Myosin-9' |
| 11158 | 'MYH9_No147' | 'MQQNIQELEEQLEEEESARQK' | 'Myosin-9' |
| 11159 | 'MYH9_No148' | 'KLEEEQIILEDQNCKLAK' | 'Myosin-9' |
| 11160 | 'MYH9_No149' | 'QTLENERGELANEVK' | 'Myosin-9' |
| 11161 | 'MYH9_No150' | 'YEILTPNSIPK' | 'Myosin-9' |
| 11162 | 'MYH9_No151' | 'KANLQIDQINTDLNLER' | 'Myosin-9' |
| 11163 | 'MYH9_No152' | 'LKNKHEAMITDLEER' | 'Myosin-9' |
| 11164 | 'MYH9_No153' | 'SMEAEMIQLQEELAAAER' | 'Myosin-9' |
| 11165 | 'MYH9_No154' | 'SGFEPASLKEEVGEEAIVELVENGK' | 'Myosin-9' |
| 11166 | 'MYH9_No155' | 'SMEAEMIQLQEELAAAER' | 'Myosin-9' |
| 11167 | 'MYH9_No156' | 'KMEDSVGCLETAEEVKR' | 'Myosin-9' |
| 11168 | 'MYH9_No157' | 'QIATLHAQVADMK' | 'Myosin-9' |
| 11169 | 'MYH9_No158' | 'KQELEEICHDLEARVEEEEER' | 'Myosin-9' |
| 11170 | 'MYH9_No159' | 'FLSNGHVTIPGQQDK' | 'Myosin-9' |
| 11171 | 'MYH9_No160' | 'KLEMDLKDLEAHIDSANK' | 'Myosin-9' |
| 11172 | 'MYH9_No161' | 'EEILAQAKENEK' | 'Myosin-9' |
| 11173 | 'MYH9_No162' | 'VKLQEMEGTVK' | 'Myosin-9' |
| 11174 | 'MYH9_No163' | 'TDLLLEPYNK' | 'Myosin-9' |
| 11175 | 'MYH9_No164' | 'LEMDLKDLEAHIDSANK' | 'Myosin-9' |
| 11176 | 'MYH9_No165' | 'LTEMETLQSQLMAEK' | 'Myosin-9' |
| 11177 | 'MYH9_No166' | 'KMEDSVGCLETAEEVK' | 'Myosin-9' |

| | A | B | C |
|---|---|---|---|
| 1 | **Peptide Name** | **Peptide Sequence** | **Peptide Description** |
| 11178 | 'MYH9_No167' | 'ERNTDQASMPDNTAAQK' | 'Myosin-9' |
| 11179 | 'MYH9_No168' | 'QEEEMMAKEEELVK' | 'Myosin-9' |
| 11180 | 'IGFALS_No1' | 'SFEGLGQLEVLTLDHNQLQEVK' | 'Insulin-like growth factor-binding protein complex acid labile subunit' |
| 11181 | 'IGFALS_No2' | 'LEYLLLSR' | 'Insulin-like growth factor-binding protein complex acid labile subunit' |
| 11182 | 'IGFALS_No3' | 'AFWLDVSHNR' | 'Insulin-like growth factor-binding protein complex acid labile subunit' |
| 11183 | 'IGFALS_No4' | 'SLALGTFAHTPALASLGLSNNR' | 'Insulin-like growth factor-binding protein complex acid labile subunit' |
| 11184 | 'IGFALS_No5' | 'LWLEGNPWDCGCPLK' | 'Insulin-like growth factor-binding protein complex acid labile subunit' |
| 11185 | 'IGFALS_No6' | 'LHSLHLEGSCLGR' | 'Insulin-like growth factor-binding protein complex acid labile subunit' |
| 11186 | 'IGFALS_No7' | 'TFTPQPPGLER' | 'Insulin-like growth factor-binding protein complex acid labile subunit' |
| 11187 | 'IGFALS_No8' | 'TFKDLHFLEELQLGHNR' | 'Insulin-like growth factor-binding protein complex acid labile subunit' |
| 11188 | 'IGFALS_No9' | 'DLHFLEELQLGHNR' | 'Insulin-like growth factor-binding protein complex acid labile subunit' |
| 11189 | 'IGFALS_No10' | 'LSHNAIASLRPR' | 'Insulin-like growth factor-binding protein complex acid labile subunit' |
| 11190 | 'IGFALS_No11' | 'WLDLSHNR' | 'Insulin-like growth factor-binding protein complex acid labile subunit' |
| 11191 | 'IGFALS_No12' | 'IRPHTFTGLSGLR' | 'Insulin-like growth factor-binding protein complex acid labile subunit' |
| 11192 | 'IGFALS_No13' | 'DFALQNPSAVPR' | 'Insulin-like growth factor-binding protein complex acid labile subunit' |
| 11193 | 'IGFALS_No14' | 'NLIAAVAPGAFLGLK' | 'Insulin-like growth factor-binding protein complex acid labile subunit' |
| 11194 | 'IGFALS_No15' | 'AFWLDVSHNRLEALPNSLLAPLGR' | 'Insulin-like growth factor-binding protein complex acid labile subunit' |
| 11195 | 'IGFALS_No16' | 'LAYLQPALFSGLAELR' | 'Insulin-like growth factor-binding protein complex acid labile subunit' |
| 11196 | 'IGFALS_No17' | 'ALRDFALQNPSAVPR' | 'Insulin-like growth factor-binding protein complex acid labile subunit' |
| 11197 | 'IGFALS_No18' | 'LEALPNSLLAPLGR' | 'Insulin-like growth factor-binding protein complex acid labile subunit' |
| 11198 | 'IGFALS_No19' | 'LAELPADALGPLQR' | 'Insulin-like growth factor-binding protein complex acid labile subunit' |
| 11199 | 'IGFALS_No20' | 'LFQGLGKLEYLLLSR' | 'Insulin-like growth factor-binding protein complex acid labile subunit' |
| 11200 | 'IGFALS_No21' | 'GLGKLHSLHLEGSCLGR' | 'Insulin-like growth factor-binding protein complex acid labile subunit' |
| 11201 | 'IGFALS_No22' | 'ANVFVQLPR' | 'Insulin-like growth factor-binding protein complex acid labile subunit' |
| 11202 | 'IGFALS_No23' | 'LYLDRNLIAAVAPGAFLGLK' | 'Insulin-like growth factor-binding protein complex acid labile subunit' |
| 11203 | 'IGFALS_No24' | 'VAGLLEDTFPGLLGLR' | 'Insulin-like growth factor-binding protein complex acid labile subunit' |
| 11204 | 'IGFALS_No25' | 'AGAFLGLTNVAVMNLSGNCLR' | 'Insulin-like growth factor-binding protein complex acid labile subunit' |
| 11205 | 'IGFALS_No26' | 'AGAFLGLTNVAVMNLSGNCLR' | 'Insulin-like growth factor-binding protein complex acid labile subunit' |
| 11206 | 'IGFALS_No27' | 'NLPEQVFR' | 'Insulin-like growth factor-binding protein complex acid labile subunit' |
| 11207 | 'IGFALS_No28' | 'DLSEAHFAPC' | 'Insulin-like growth factor-binding protein complex acid labile subunit' |
| 11208 | 'KRT2_No1' | 'FLEQQNQVLQTK' | 'Keratin, type II cytoskeletal 2 epidermal' |
| 11209 | 'KRT2_No2' | 'VDLLNQEIEFLK' | 'Keratin, type II cytoskeletal 2 epidermal' |
| 11210 | 'KRT2_No3' | 'IEISELNR' | 'Keratin, type II cytoskeletal 2 epidermal' |
| 11211 | 'KRT2_No4' | 'GFSSGSAVVSGGSR' | 'Keratin, type II cytoskeletal 2 epidermal' |
| 11212 | 'KRT2_No5' | 'YLDGLTAER' | 'Keratin, type II cytoskeletal 2 epidermal' |
| 11213 | 'KRT2_No6' | 'TAAENDFVTLKK' | 'Keratin, type II cytoskeletal 2 epidermal' |
| 11214 | 'KRT2_No7' | 'NVQDAIADAEQR' | 'Keratin, type II cytoskeletal 2 epidermal' |
| 11215 | 'KRT2_No8' | 'YEELQVTVGR' | 'Keratin, type II cytoskeletal 2 epidermal' |
| 11216 | 'KRT2_No9' | 'GGSGGGGSISGGGYGSGGGSGGR' | 'Keratin, type II cytoskeletal 2 epidermal' |
| 11217 | 'KRT2_No10' | 'VDPEIQNVK' | 'Keratin, type II cytoskeletal 2 epidermal' |
| 11218 | 'KRT2_No11' | 'NVQDAIADAEQRGEHALK' | 'Keratin, type II cytoskeletal 2 epidermal' |
| 11219 | 'KRT2_No12' | 'SKEEAEALYHSKYEELQVTVGR' | 'Keratin, type II cytoskeletal 2 epidermal' |
| 11220 | 'KRT2_No13' | 'NVQDAIADAEQRGEHALKDAR' | 'Keratin, type II cytoskeletal 2 epidermal' |
| 11221 | 'KRT2_No14' | 'LNDLEEALQQAKEDLAR' | 'Keratin, type II cytoskeletal 2 epidermal' |

| | A | B | C |
|---|---|---|---|
| 1 | **Peptide Name** | **Peptide Sequence** | **Peptide Description** |
| 11222 | 'KRT2_No15' | 'GGGFGGGSSFGGGSGFSGGGFGGGGFGGGR' | 'Keratin, type II cytoskeletal 2 epidermal' |
| 11223 | 'KRT2_No16' | 'EIKIEISELNR' | 'Keratin, type II cytoskeletal 2 epidermal' |
| 11224 | 'KRT2_No17' | 'LLRDYQELMNVK' | 'Keratin, type II cytoskeletal 2 epidermal' |
| 11225 | 'KRT2_No18' | 'VLYDAEISQIHQSVTDTNVILSMDNSR' | 'Keratin, type II cytoskeletal 2 epidermal' |
| 11226 | 'KRT2_No19' | 'TAAENDFVTLK' | 'Keratin, type II cytoskeletal 2 epidermal' |
| 11227 | 'KRT2_No20' | 'SKEEAEALYHSK' | 'Keratin, type II cytoskeletal 2 epidermal' |
| 11228 | 'KRT2_No21' | 'LNDLEEALQQAK' | 'Keratin, type II cytoskeletal 2 epidermal' |
| 11229 | 'KRT2_No22' | 'GSSSGGGYSSGSSSYGSGGR' | 'Keratin, type II cytoskeletal 2 epidermal' |
| 11230 | 'KRT2_No23' | 'TSQNSELNNMQDLVEDYKK' | 'Keratin, type II cytoskeletal 2 epidermal' |
| 11231 | 'KRT2_No24' | 'SISISVAGGGGGFGAAGGFGGR' | 'Keratin, type II cytoskeletal 2 epidermal' |
| 11232 | 'KRT2_No25' | 'NKLNDLEEALQQAKEDLAR' | 'Keratin, type II cytoskeletal 2 epidermal' |
| 11233 | 'KRT2_No26' | 'WELLQQMNVGTRPINLEPIFQGYIDSLKR' | 'Keratin, type II cytoskeletal 2 epidermal' |
| 11234 | 'KRT2_No27' | 'TSQNSELNNMQDLVEDYK' | 'Keratin, type II cytoskeletal 2 epidermal' |
| 11235 | 'KRT2_No28' | 'STSSFSCLSR' | 'Keratin, type II cytoskeletal 2 epidermal' |
| 11236 | 'KRT2_No29' | 'DVDNAYMIK' | 'Keratin, type II cytoskeletal 2 epidermal' |
| 11237 | 'KRT2_No30' | 'TSQNSELNNMQDLVEDYKK' | 'Keratin, type II cytoskeletal 2 epidermal' |
| 11238 | 'KRT2_No31' | 'VLYDAEISQIHQSVTDTNVILSMDNSR' | 'Keratin, type II cytoskeletal 2 epidermal' |
| 11239 | 'KRT2_No32' | 'HGGGGGGFGGGGFGSR' | 'Keratin, type II cytoskeletal 2 epidermal' |
| 11240 | 'KRT2_No33' | 'TSQNSELNNMQDLVEDYK' | 'Keratin, type II cytoskeletal 2 epidermal' |
| 11241 | 'KRT2_No34' | 'VELQSKVDLLNQEIEFLK' | 'Keratin, type II cytoskeletal 2 epidermal' |
| 11242 | 'KRT2_No35' | 'HGDSLKEIKIEISELNR' | 'Keratin, type II cytoskeletal 2 epidermal' |
| 11243 | 'KRT2_No36' | 'PINLEPIFQGYIDSLKR' | 'Keratin, type II cytoskeletal 2 epidermal' |
| 11244 | 'KRT2_No37' | 'TSQNSELNNMQDLVEDYKKK' | 'Keratin, type II cytoskeletal 2 epidermal' |
| 11245 | 'KRT2_No38' | 'NKLNDLEEALQQAK' | 'Keratin, type II cytoskeletal 2 epidermal' |
| 11246 | 'KRT2_No39' | 'WELLQQMNVGTR' | 'Keratin, type II cytoskeletal 2 epidermal' |
| 11247 | 'KRT2_No40' | 'LLRDYQELMNVK' | 'Keratin, type II cytoskeletal 2 epidermal' |
| 11248 | 'KRT2_No41' | 'GFSSGSAVVSGGSRR' | 'Keratin, type II cytoskeletal 2 epidermal' |
| 11249 | 'KRT2_No42' | 'WELLQQMNVGTR' | 'Keratin, type II cytoskeletal 2 epidermal' |
| 11250 | 'KRT2_No43' | 'IEISELNRVIQR' | 'Keratin, type II cytoskeletal 2 epidermal' |
| 11251 | 'KRT2_No44' | 'GGSISGGGYGSGGGK' | 'Keratin, type II cytoskeletal 2 epidermal' |
| 11252 | 'KRT2_No45' | 'DVDNAYMIKVELQSK' | 'Keratin, type II cytoskeletal 2 epidermal' |
| 11253 | 'KRT2_No46' | 'DYQELMNVK' | 'Keratin, type II cytoskeletal 2 epidermal' |
| 11254 | 'KRT2_No47' | 'RYLDGLTAER' | 'Keratin, type II cytoskeletal 2 epidermal' |
| 11255 | 'KRT2_No48' | 'MSGDLSSNVTVSVTSSTISSNVASK' | 'Keratin, type II cytoskeletal 2 epidermal' |
| 11256 | 'KRT2_No49' | 'TSQNSELNNMQDLVEDYKKK' | 'Keratin, type II cytoskeletal 2 epidermal' |
| 11257 | 'KRT2_No50' | 'AAFGGSGGRGSSSGGGYSSGSSSYGSGGR' | 'Keratin, type II cytoskeletal 2 epidermal' |
| 11258 | 'FLT4_No1' | 'VAPEHEGHYVCEVQDRR' | 'Vascular endothelial growth factor receptor 3' |
| 11259 | 'FLT4_No2' | 'DFEQPFINKPDTLLVNR' | 'Vascular endothelial growth factor receptor 3' |
| 11260 | 'FLT4_No3' | 'VAPEHEGHYVCEVQDR' | 'Vascular endothelial growth factor receptor 3' |
| 11261 | 'FLT4_No4' | 'ESTEVIVHENPFISVEWLK' | 'Vascular endothelial growth factor receptor 3' |
| 11262 | 'FLT4_No5' | 'DFEQPFINKPDTLLVNRK' | 'Vascular endothelial growth factor receptor 3' |
| 11263 | 'FLT4_No6' | 'GPILEATAGDELVKLPVK' | 'Vascular endothelial growth factor receptor 3' |
| 11264 | 'FLT4_No7' | 'EVTEASTGTYTLALWNSAAGLR' | 'Vascular endothelial growth factor receptor 3' |
| 11265 | 'FLT4_No8' | 'NVHLFATPLAASLEEVAPGAR' | 'Vascular endothelial growth factor receptor 3' |

| | A | B | C |
|---|---|---|---|
| 1 | **Peptide Name** | **Peptide Sequence** | **Peptide Description** |
| 11266 | 'FLT4_No9' | 'HATLSLSIPR' | 'Vascular endothelial growth factor receptor 3' |
| 11267 | 'CHI3L1_No1' | 'GNQWVGYDDQESVK' | 'Chitinase-3-like protein 1' |
| 11268 | 'CHI3L1_No2' | 'SFTLASSETGVGAPISGPGIPGR' | 'Chitinase-3-like protein 1' |
| 11269 | 'CHI3L1_No3' | 'LVMGIPTFGR' | 'Chitinase-3-like protein 1' |
| 11270 | 'CHI3L1_No4' | 'TLLSVGGWNFGSQR' | 'Chitinase-3-like protein 1' |
| 11271 | 'CHI3L1_No5' | 'ILGQQVPYATK' | 'Chitinase-3-like protein 1' |
| 11272 | 'PGM1_No1' | 'TQAYQDQKPGTSGLR' | 'Phosphoglucomutase-1' |
| 11273 | 'PGM1_No2' | 'SIFDFSALK' | 'Phosphoglucomutase-1' |
| 11274 | 'PGM1_No3' | 'LVIGQNGILSTPAVSCIIR' | 'Phosphoglucomutase-1' |
| 11275 | 'PPP1CC_No1' | 'IKYPENFFLLR' | 'Serine/threonine-protein phosphatase PP1-gamma catalytic subunit' |
| 11276 | 'PPP1CC_No2' | 'ADLDKLNIDSIIQR' | 'Serine/threonine-protein phosphatase PP1-gamma catalytic subunit' |
| 11277 | 'PPP1CC_No3' | 'HDLDLICR' | 'Serine/threonine-protein phosphatase PP1-gamma catalytic subunit' |
| 11278 | 'PPP1CC_No4' | 'AHQVVEDGYEFFAK' | 'Serine/threonine-protein phosphatase PP1-gamma catalytic subunit' |
| 11279 | 'SERPINF1_No1' | 'LQSLFDSPDFSK' | 'Pigment epithelium-derived factor' |
| 11280 | 'SERPINF1_No2' | 'TSLEDFYLDEER' | 'Pigment epithelium-derived factor' |
| 11281 | 'SERPINF1_No3' | 'KTSLEDFYLDEER' | 'Pigment epithelium-derived factor' |
| 11282 | 'SERPINF1_No4' | 'LAAAVSNFGYDLYR' | 'Pigment epithelium-derived factor' |
| 11283 | 'SERPINF1_No5' | 'ALYYDLISSPDIHGTYK' | 'Pigment epithelium-derived factor' |
| 11284 | 'SERPINF1_No6' | 'ELLDTVTAPQK' | 'Pigment epithelium-derived factor' |
| 11285 | 'SERPINF1_No7' | 'LSYEGEVTK' | 'Pigment epithelium-derived factor' |
| 11286 | 'SERPINF1_No8' | 'DTDTGALLFIGK' | 'Pigment epithelium-derived factor' |
| 11287 | 'SERPINF1_No9' | 'ALYYDLISSPDIHGTYKELLDTVTAPQK' | 'Pigment epithelium-derived factor' |
| 11288 | 'SERPINF1_No10' | 'LDLQEINNWVQAQMK' | 'Pigment epithelium-derived factor' |
| 11289 | 'SERPINF1_No11' | 'VPMMSDPK' | 'Pigment epithelium-derived factor' |
| 11290 | 'SERPINF1_No12' | 'SSTSPTTNVLLSPLSVATALSALSLGAEQR' | 'Pigment epithelium-derived factor' |
| 11291 | 'SERPINF1_No13' | 'ITGKPIKLTQVEHR' | 'Pigment epithelium-derived factor' |
| 11292 | 'SERPINF1_No14' | 'EIPDEISILLLGVAHFK' | 'Pigment epithelium-derived factor' |
| 11293 | 'SERPINF1_No15' | 'IKSSFVAPLEK' | 'Pigment epithelium-derived factor' |
| 11294 | 'SERPINF1_No16' | 'IAQLPLTGSMSIIFFLPLK' | 'Pigment epithelium-derived factor' |
| 11295 | 'SERPINF1_No17' | 'YGLDSDLSCK' | 'Pigment epithelium-derived factor' |
| 11296 | 'SERPINF1_No18' | 'SSFVAPLEK' | 'Pigment epithelium-derived factor' |
| 11297 | 'SERPINF1_No19' | 'LKLSYEGEVTK' | 'Pigment epithelium-derived factor' |
| 11298 | 'SERPINF1_No20' | 'TVRVPMMSDPK' | 'Pigment epithelium-derived factor' |
| 11299 | 'SERPINF1_No21' | 'LQSLFDSPDFSKITGKPIK' | 'Pigment epithelium-derived factor' |
| 11300 | 'SERPINF1_No22' | 'SSTSPTTNVLLSPLSVATALSALSLGAEQRTESIIHR' | 'Pigment epithelium-derived factor' |
| 11301 | 'SERPINF1_No23' | 'LDLQEINNWVQAQMK' | 'Pigment epithelium-derived factor' |
| 11302 | 'SERPINF1_No24' | 'TESIIHR' | 'Pigment epithelium-derived factor' |
| 11303 | 'SERPINF1_No25' | 'KTSLEDFYLDEERTVR' | 'Pigment epithelium-derived factor' |
| 11304 | 'SERPINF1_No26' | 'STKEIPDEISILLLGVAHFK' | 'Pigment epithelium-derived factor' |
| 11305 | 'SERPINF1_No27' | 'TVQAVLTVPK' | 'Pigment epithelium-derived factor' |
| 11306 | 'SERPINF1_No28' | 'ELLDTVTAPQKNLK' | 'Pigment epithelium-derived factor' |
| 11307 | 'SERPINF1_No29' | 'ALYYDLISSPDIHGTYKELLDTVTAPQKNLK' | 'Pigment epithelium-derived factor' |
| 11308 | 'SERPINF1_No30' | 'SLQEMKLQSLFDSPDFSK' | 'Pigment epithelium-derived factor' |
| 11309 | 'SERPINF1_No31' | 'LTQVEHR' | 'Pigment epithelium-derived factor' |

| | A | B | C |
|---|---|---|---|
| 1 | **Peptide Name** | **Peptide Sequence** | **Peptide Description** |
| 11310 | 'GMPR' | 'GHIISDGGCTCPGDVAK' | 'GMP reductase 1' |
| 11311 | 'CFHR2_No1' | 'ITCAEEGWSPTPK' | 'Complement factor H-related protein 2' |
| 11312 | 'CFHR2_No2' | 'LVYPSCEEK' | 'Complement factor H-related protein 2' |
| 11313 | 'CFHR2_No3' | 'CLDPCVISQEIMEK' | 'Complement factor H-related protein 2' |
| 11314 | 'CFHR2_No4' | 'TGDIVEFVCK' | 'Complement factor H-related protein 2' |
| 11315 | 'CFHR2_No5' | 'CLDPCVISQEIMEK' | 'Complement factor H-related protein 2' |
| 11316 | 'CFHR2_No6' | 'NGQWSEPPKCLDPCVISQEIMEK' | 'Complement factor H-related protein 2' |
| 11317 | 'CFHR2_No7' | 'CLDPCVISQEIMEKYNIK' | 'Complement factor H-related protein 2' |
| 11318 | 'CFHR2_No8' | 'AMCQNGKLVYPSCEEK' | 'Complement factor H-related protein 2' |
| 11319 | 'CFHR2_No9' | 'CLDPCVISQEIMEKYNIK' | 'Complement factor H-related protein 2' |
| 11320 | 'TAGLN2_No1' | 'TLMNLGGLAVAR' | 'Transgelin-2' |
| 11321 | 'TAGLN2_No2' | 'DDGLFSGDPNWFPK' | 'Transgelin-2' |
| 11322 | 'TAGLN2_No3' | 'ENFQNWLK' | 'Transgelin-2' |
| 11323 | 'TAGLN2_No4' | 'GASQAGMTGYGMPR' | 'Transgelin-2' |
| 11324 | 'TAGLN2_No5' | 'NFSDNQLQEGK' | 'Transgelin-2' |
| 11325 | 'TAGLN2_No6' | 'TLMNLGGLAVAR' | 'Transgelin-2' |
| 11326 | 'TAGLN2_No7' | 'DDGLFSGDPNWFPKK' | 'Transgelin-2' |
| 11327 | 'TAGLN2_No8' | 'DGTVLCELINALYPEGQAPVK' | 'Transgelin-2' |
| 11328 | 'TAGLN2_No9' | 'IQASTMAFK' | 'Transgelin-2' |
| 11329 | 'TAGLN2_No10' | 'QMEQISQFLQAAER' | 'Transgelin-2' |
| 11330 | 'TAGLN2_No11' | 'DGTVLCELINALYPEGQAPVKK' | 'Transgelin-2' |
| 11331 | 'TAGLN2_No12' | 'QMEQISQFLQAAER' | 'Transgelin-2' |
| 11332 | 'TAGLN2_No13' | 'DVGRPQPGRENFQNWLK' | 'Transgelin-2' |
| 11333 | 'TAGLN2_No14' | 'GASQAGMTGYGMPR' | 'Transgelin-2' |
| 11334 | 'TAGLN2_No15' | 'NVIGLQMGTNR' | 'Transgelin-2' |
| 11335 | 'TAGLN2_No16' | 'ANRGPAYGLSR' | 'Transgelin-2' |
| 11336 | 'TAGLN2_No17' | 'GASQAGMTGYGMPR' | 'Transgelin-2' |
| 11337 | 'TAGLN2_No18' | 'QMEQISQFLQAAER' | 'Transgelin-2' |
| 11338 | 'TAGLN2_No19' | 'YGINTTDIFQTVDLWEGK' | 'Transgelin-2' |
| 11339 | 'TAGLN2_No20' | 'NVIGLQMGTNR' | 'Transgelin-2' |
| 11340 | 'TAGLN2_No21' | 'QMEQISQFLQAAER' | 'Transgelin-2' |
| 11341 | 'TALDO1_No1' | 'ALAGCDFLTISPK' | 'Transaldolase' |
| 11342 | 'TALDO1_No2' | 'ILDWHVANTDKK' | 'Transaldolase' |
| 11343 | 'TALDO1_No3' | 'LLGELLQDNAK' | 'Transaldolase' |
| 11344 | 'TALDO1_No4' | 'LSFDKDAMVAR' | 'Transaldolase' |
| 11345 | 'SNCA_No1' | 'TKEQVTNVGGAVVTGVTAVAQK' | 'Alpha-synuclein' |
| 11346 | 'SNCA_No2' | 'TKEGVVHGVATVAEK' | 'Alpha-synuclein' |
| 11347 | 'SNCA_No3' | 'EGVVHGVATVAEK' | 'Alpha-synuclein' |
| 11348 | 'HSPA9_No1' | 'AQFEGIVTDLIR' | 'Stress-70 protein, mitochondrial' |
| 11349 | 'HSPA9_No2' | 'QAVTNPNNTFYATK' | 'Stress-70 protein, mitochondrial' |
| 11350 | 'COL18A1_No1' | 'TEAPSATGQASSLLGGR' | 'Collagen alpha-1(XVIII) chain' |
| 11351 | 'COL18A1_No2' | 'DFQPVLHLVALNSPLSGGMR' | 'Collagen alpha-1(XVIII) chain' |
| 11352 | 'COL18A1_No3' | 'LQDLYSIVR' | 'Collagen alpha-1(XVIII) chain' |
| 11353 | 'COL18A1_No4' | 'YHFPSLFFR' | 'Collagen alpha-1(XVIII) chain' |

| | A | B | C |
|---|---|---|---|
| 1 | **Peptide Name** | **Peptide Sequence** | **Peptide Description** |
| 11354 | 'COL18A1_No5' | 'GLELEPGAGLFVAQAGGADPDKFQGVIAELK' | 'Collagen alpha-1(XVIII) chain' |
| 11355 | 'COL18A1_No6' | 'ELLREETGAALKPR' | 'Collagen alpha-1(XVIII) chain' |
| 11356 | 'COL18A1_No7' | 'IFSFDGKDVLR' | 'Collagen alpha-1(XVIII) chain' |
| 11357 | 'COL18A1_No8' | 'DFQPVLHLVALNSPLSGGMR' | 'Collagen alpha-1(XVIII) chain' |
| 11358 | 'COL18A1_No9' | 'GADFQCFQQAR' | 'Collagen alpha-1(XVIII) chain' |
| 11359 | 'COL18A1_No10' | 'LSGVQDGHQDISLLYTEPGAGQTHTAASFR' | 'Collagen alpha-1(XVIII) chain' |
| 11360 | 'COL18A1_No11' | 'RLTESYCETWR' | 'Collagen alpha-1(XVIII) chain' |
| 11361 | 'IL6ST_No1' | 'SSFTVQDLKPFTEYVFR' | 'Interleukin-6 receptor subunit beta' |
| 11362 | 'IL6ST_No2' | 'CYLITVTPVYADGPGSPESIK' | 'Interleukin-6 receptor subunit beta' |
| 11363 | 'IL6ST_No3' | 'APCITDWQQEDGTVHR' | 'Interleukin-6 receptor subunit beta' |
| 11364 | 'IL6ST_No4' | 'APSFWYK' | 'Interleukin-6 receptor subunit beta' |
| 11365 | 'GP5_No1' | 'LPNLSSLTLSR' | 'Platelet glycoprotein V' |
| 11366 | 'GP5_No2' | 'LTEVLLGHNSWR' | 'Platelet glycoprotein V' |
| 11367 | 'GP5_No3' | 'LMISDSHISAVAPGTFSDLIK' | 'Platelet glycoprotein V' |
| 11368 | 'GP5_No4' | 'MVLLEQLFLDHNALR' | 'Platelet glycoprotein V' |
| 11369 | 'GP5_No5' | 'GVLQSQSFSGMTVLQR' | 'Platelet glycoprotein V' |
| 11370 | 'GP5_No6' | 'SIAPGAFDR' | 'Platelet glycoprotein V' |
| 11371 | 'GP5_No7' | 'LMISDSHISAVAPGTFSDLIK' | 'Platelet glycoprotein V' |
| 11372 | 'GP5_No8' | 'GIDQNMFQK' | 'Platelet glycoprotein V' |
| 11373 | 'CCT6A' | 'GIDPFSLDALSK' | 'T-complex protein 1 subunit zeta' |
| 11374 | 'MDH1_No1' | 'NVIIWGNHSSTQYPDVNHAK' | 'Malate dehydrogenase, cytoplasmic' |
| 11375 | 'MDH1_No2' | 'FVEGLPINDFSR' | 'Malate dehydrogenase, cytoplasmic' |
| 11376 | 'MDH1_No3' | 'EVGVYEALKDDSWLK' | 'Malate dehydrogenase, cytoplasmic' |
| 11377 | 'MDH1_No4' | 'VIVVGNPANTNCLTASK' | 'Malate dehydrogenase, cytoplasmic' |
| 11378 | 'MDH2_No1' | 'IFGVTTLDIVR' | 'Malate dehydrogenase, mitochondrial' |
| 11379 | 'MDH2_No2' | 'LTLYDIAHTPGVAADLSHIETK' | 'Malate dehydrogenase, mitochondrial' |
| 11380 | 'MDH2_No3' | 'VDFPQDQLTALTGR' | 'Malate dehydrogenase, mitochondrial' |
| 11381 | 'MDH2_No4' | 'FVFSLVDAMNGK' | 'Malate dehydrogenase, mitochondrial' |
| 11382 | 'MDH2_No5' | 'GYLGPEQLPDCLK' | 'Malate dehydrogenase, mitochondrial' |
| 11383 | 'HADHA_No1' | 'TVLGTPEVLLGALPGAGGTQR' | 'Trifunctional enzyme subunit alpha, mitochondrial' |
| 11384 | 'HADHA_No2' | 'TGIEQGSDAGYLCESQK' | 'Trifunctional enzyme subunit alpha, mitochondrial' |
| 11385 | 'MNDA' | 'SLLAYDLGLTTK' | 'Myeloid cell nuclear differentiation antigen' |
| 11386 | 'PTGDS_No1' | 'AQGFTEDTIVFLPQTDK' | 'Prostaglandin-H2 D-isomerase' |
| 11387 | 'PTGDS_No2' | 'TMLLQPAGSLGSYSYR' | 'Prostaglandin-H2 D-isomerase' |
| 11388 | 'PTGDS_No3' | 'TMLLQPAGSLGSYSYR' | 'Prostaglandin-H2 D-isomerase' |
| 11389 | 'PTGDS_No4' | 'AQGFTEDTIVFLPQTDKCMTEQ' | 'Prostaglandin-H2 D-isomerase' |
| 11390 | 'CSK_No1' | 'LLYPPETGLFLVR' | 'Tyrosine-protein kinase CSK' |
| 11391 | 'CSK_No2' | 'VMEGTVAAQDEFYR' | 'Tyrosine-protein kinase CSK' |
| 11392 | 'ECI1' | 'DADVQNFVSFISK' | 'Enoyl-CoA delta isomerase 1, mitochondrial' |
| 11393 | 'EPS15' | 'AAAAQLSLTQLSSGNPVYEK' | 'Epidermal growth factor receptor substrate 15' |
| 11394 | 'PRCP_No1' | 'PSWITTMYGGK' | 'Lysosomal Pro-X carboxypeptidase' |
| 11395 | 'PRCP_No2' | 'DITDTLVAVTISEGAHHLDLR' | 'Lysosomal Pro-X carboxypeptidase' |
| 11396 | 'PRCP_No3' | 'YYGESLPFGDNSFK' | 'Lysosomal Pro-X carboxypeptidase' |
| 11397 | 'MCAM_No1' | 'GATLALTQVTPQDER' | 'Cell surface glycoprotein MUC18' |

| | A | B | C |
|---|---|---|---|
| 1 | **Peptide Name** | **Peptide Sequence** | **Peptide Description** |
| 11398 | 'MCAM_No2' | 'VWLEVEPVGMLK' | 'Cell surface glycoprotein MUC18' |
| 11399 | 'MCAM_No3' | 'EAEEETTNDNGVLVLEPAR' | 'Cell surface glycoprotein MUC18' |
| 11400 | 'MCAM_No4' | 'GPVLQLHDLKR' | 'Cell surface glycoprotein MUC18' |
| 11401 | 'MCAM_No5' | 'EPEEVATCVGR' | 'Cell surface glycoprotein MUC18' |
| 11402 | 'MCAM_No6' | 'NGYPIPQVIWYK' | 'Cell surface glycoprotein MUC18' |
| 11403 | 'MCAM_No7' | 'VYKAPEEPNIQVNPLGIPVNSKEPEEVATCVGR' | 'Cell surface glycoprotein MUC18' |
| 11404 | 'MCAM_No8' | 'EDKDAQFYCELNYR' | 'Cell surface glycoprotein MUC18' |
| 11405 | 'MCAM_No9' | 'GPVLQLHDLK' | 'Cell surface glycoprotein MUC18' |
| 11406 | 'MCAM_No10' | 'VWLEVEPVGMLK' | 'Cell surface glycoprotein MUC18' |
| 11407 | 'MCAM_No11' | 'EVTVPVFYPTEK' | 'Cell surface glycoprotein MUC18' |
| 11408 | 'MCAM_No12' | 'VHIQSSQTVESSGLYTLQSILK' | 'Cell surface glycoprotein MUC18' |
| 11409 | 'MCAM_No13' | 'AQLVKEDKDAQFYCELNYR' | 'Cell surface glycoprotein MUC18' |
| 11410 | 'MCAM_No14' | 'EAEEETTNDNGVLVLEPARK' | 'Cell surface glycoprotein MUC18' |
| 11411 | 'MCAM_No15' | 'APEEPNIQVNPLGIPVNSKEPEEVATCVGR' | 'Cell surface glycoprotein MUC18' |
| 11412 | 'MCAM_No16' | 'VRQGQGQSEPGEYEQR' | 'Cell surface glycoprotein MUC18' |
| 11413 | 'MCAM_No17' | 'CLADGNPPPHFSISK' | 'Cell surface glycoprotein MUC18' |
| 11414 | 'BTD_No1' | 'LSSGLVTAALYGR' | 'Biotinidase' |
| 11415 | 'BTD_No2' | 'VDLITFDTPFAGR' | 'Biotinidase' |
| 11416 | 'BTD_No3' | 'EGYLHVCSNGLCCYLLYERPTLSK' | 'Biotinidase' |
| 11417 | 'BTD_No4' | 'SHLIIAQVAK' | 'Biotinidase' |
| 11418 | 'BTD_No5' | 'GDMFLVANLGTK' | 'Biotinidase' |
| 11419 | 'BTD_No6' | 'QEALELMNQNLDIYEQQVMTAAQK' | 'Biotinidase' |
| 11420 | 'BTD_No7' | 'HNLYFEAAFDVPLK' | 'Biotinidase' |
| 11421 | 'BTD_No8' | 'TSIYPFLDFMPSPQVVR' | 'Biotinidase' |
| 11422 | 'BTD_No9' | 'GDMFLVANLGTKEPCHSSDPR' | 'Biotinidase' |
| 11423 | 'BTD_No10' | 'ILSGDPYCEKDAQEVHCDEATK' | 'Biotinidase' |
| 11424 | 'BTD_No11' | 'TSIYPFLDFMPSPQVVR' | 'Biotinidase' |
| 11425 | 'BTD_No12' | 'WNPCLEPHR' | 'Biotinidase' |
| 11426 | 'BTD_No13' | 'GDMFLVANLGTKEPCHSSDPR' | 'Biotinidase' |
| 11427 | 'BTD_No14' | 'GDMFLVANLGTK' | 'Biotinidase' |
| 11428 | 'BTD_No15' | 'QEALELMNQNLDIYEQQVMTAAQK' | 'Biotinidase' |
| 11429 | 'BTD_No16' | 'LSCMAIR' | 'Biotinidase' |
| 11430 | 'BTD_No17' | 'SRLSSGLVTAALYGR' | 'Biotinidase' |
| 11431 | 'BTD_No18' | 'DAQEVHCDEATK' | 'Biotinidase' |
| 11432 | 'GPD2_No1' | 'LVSEFPYIEAEVK' | 'Glycerol-3-phosphate dehydrogenase, mitochondrial' |
| 11433 | 'GPD2_No2' | 'VIFFLPWQK' | 'Glycerol-3-phosphate dehydrogenase, mitochondrial' |
| 11434 | 'GPD2_No3' | 'RGDVLAAWSGIRPLVTDPK' | 'Glycerol-3-phosphate dehydrogenase, mitochondrial' |
| 11435 | 'AFM_No1' | 'RNPFVFAPTLLTVAVHFEEVAK' | 'Afamin' |
| 11436 | 'AFM_No2' | 'ESLLNHFLYEVAR' | 'Afamin' |
| 11437 | 'AFM_No3' | 'AESPEVCFNEESPK' | 'Afamin' |
| 11438 | 'AFM_No4' | 'FTDSENVCQER' | 'Afamin' |
| 11439 | 'AFM_No5' | 'FTFEYSR' | 'Afamin' |
| 11440 | 'AFM_No6' | 'ICAMEGLPQK' | 'Afamin' |
| 11441 | 'AFM_No7' | 'RLCFFYNK' | 'Afamin' |

| | A | B | C |
|---|---|---|---|
| 1 | **Peptide Name** | **Peptide Sequence** | **Peptide Description** |
| 11442 | 'AFM_No8' | 'LCFFYNKK' | 'Afamin' |
| 11443 | 'AFM_No9' | 'SCCEEQNKVNCLQTR' | 'Afamin' |
| 11444 | 'AFM_No10' | 'FLVNLVK' | 'Afamin' |
| 11445 | 'AFM_No11' | 'LKHELTDEELQSLFTNFANVVDK' | 'Afamin' |
| 11446 | 'AFM_No12' | 'FTDSENVCQERDADPDTFFAK' | 'Afamin' |
| 11447 | 'AFM_No13' | 'TDRFLVNLVK' | 'Afamin' |
| 11448 | 'AFM_No14' | 'IAPQLSTEELVSLGEK' | 'Afamin' |
| 11449 | 'AFM_No15' | 'KSDVGFLPPFPTLDPEEK' | 'Afamin' |
| 11450 | 'AFM_No16' | 'LVKDMVEYKDR' | 'Afamin' |
| 11451 | 'AFM_No17' | 'RHPDLSIPELLR' | 'Afamin' |
| 11452 | 'AFM_No18' | 'TINPAVDHCCK' | 'Afamin' |
| 11453 | 'AFM_No19' | 'RPCFESLK' | 'Afamin' |
| 11454 | 'AFM_No20' | 'EGKFTDSENVCQER' | 'Afamin' |
| 11455 | 'AFM_No21' | 'AESPEVCFNEESPKIGN' | 'Afamin' |
| 11456 | 'AFM_No22' | 'NCCNTENPPGCYR' | 'Afamin' |
| 11457 | 'AFM_No23' | 'VVHFIYIAILSQK' | 'Afamin' |
| 11458 | 'AFM_No24' | 'ICAMEGLPQK' | 'Afamin' |
| 11459 | 'AFM_No25' | 'KTDRFLVNLVK' | 'Afamin' |
| 11460 | 'AFM_No26' | 'GQCIINSNKDDRPK' | 'Afamin' |
| 11461 | 'AFM_No27' | 'RLCFFYNKK' | 'Afamin' |
| 11462 | 'AFM_No28' | 'CMADKTLPECSKLPNNVLQEK' | 'Afamin' |
| 11463 | 'AFM_No29' | 'EGKFTDSENVCQERDADPDTFFAK' | 'Afamin' |
| 11464 | 'AFM_No30' | 'AIPVTQYLK' | 'Afamin' |
| 11465 | 'AFM_No31' | 'NPFVFAPTLLTVAVHFEEVAK' | 'Afamin' |
| 11466 | 'AFM_No32' | 'IVQIYKDLLR' | 'Afamin' |
| 11467 | 'AFM_No33' | 'DGLKYHYLIR' | 'Afamin' |
| 11468 | 'AFM_No34' | 'CQAYESNRESLLNHFLYEVAR' | 'Afamin' |
| 11469 | 'AFM_No35' | 'DMVEYKDR' | 'Afamin' |
| 11470 | 'AFM_No36' | 'LPNNVLQEK' | 'Afamin' |
| 11471 | 'AFM_No37' | 'TYVPPPFSQDLFTFHADMCQSQNEELQR' | 'Afamin' |
| 11472 | 'AFM_No38' | 'CCKAESPEVCFNEESPK' | 'Afamin' |
| 11473 | 'AFM_No39' | 'HPDLSIPELLR' | 'Afamin' |
| 11474 | 'AFM_No40' | 'RPCFESLKADK' | 'Afamin' |
| 11475 | 'AFM_No41' | 'TLPECSKLPNNVLQEK' | 'Afamin' |
| 11476 | 'AFM_No42' | 'DADPDTFFAK' | 'Afamin' |
| 11477 | 'AFM_No43' | 'LVKDMVEYKDR' | 'Afamin' |
| 11478 | 'AFM_No44' | 'HFQNLGKDGLKYHYLIR' | 'Afamin' |
| 11479 | 'AFM_No45' | 'MVQQECKHFQNLGK' | 'Afamin' |
| 11480 | 'AFM_No46' | 'CCKAESPEVCFNEESPKIGN' | 'Afamin' |
| 11481 | 'AFM_No47' | 'ESLLNHFLYEVARR' | 'Afamin' |
| 11482 | 'AFM_No48' | 'VNCLQTR' | 'Afamin' |
| 11483 | 'AFM_No49' | 'SDVGFLPPFPTLDPEEKCQAYESNR' | 'Afamin' |
| 11484 | 'AFM_No50' | 'DMVEYKDR' | 'Afamin' |
| 11485 | 'AFM_No51' | 'HFQNLGK' | 'Afamin' |

| | A | B | C |
|---|---|---|---|
| 1 | **Peptide Name** | **Peptide Sequence** | **Peptide Description** |
| 11486 | 'AFM_No52' | 'ELISLVEDVSSNYDGCCEGDVVQCIR' | 'Afamin' |
| 11487 | 'AFM_No53' | 'MVQQECKHFQNLGKDGLK' | 'Afamin' |
| 11488 | 'AFM_No54' | 'CMADKTLPECSKLPNNVLQEK' | 'Afamin' |
| 11489 | 'AFM_No55' | 'HVCGALLK' | 'Afamin' |
| 11490 | 'AFM_No56' | 'HELTDEELQSLFTNFANVVDK' | 'Afamin' |
| 11491 | 'AFM_No57' | 'DLSLREGKFTDSENVCQER' | 'Afamin' |
| 11492 | 'AFM_No58' | 'QDSISSK' | 'Afamin' |
| 11493 | 'AFM_No59' | 'AFSSYQK' | 'Afamin' |
| 11494 | 'AFM_No60' | 'ELISLVEDVSSNYDGCCEGDVVQCIRDTSK' | 'Afamin' |
| 11495 | 'AFM_No61' | 'KSDVGFLPPFPTLDPEEKCQAYESNR' | 'Afamin' |
| 11496 | 'AFM_No62' | 'MVQQECKHFQNLGK' | 'Afamin' |
| 11497 | 'AFM_No63' | 'VMNHICSK' | 'Afamin' |
| 11498 | 'AFM_No64' | 'GQCIINSNK' | 'Afamin' |
| 11499 | 'AFM_No65' | 'LCFFYNK' | 'Afamin' |
| 11500 | 'AFM_No66' | 'TYVPPPFSQDLFTFHADMCQSQNEELQR' | 'Afamin' |
| 11501 | 'AFM_No67' | 'LCFFYNKKSDVGFLPPFPTLDPEEK' | 'Afamin' |
| 11502 | 'AFM_No68' | 'CQAYESNR' | 'Afamin' |
| 11503 | 'AFM_No69' | 'SDVGFLPPFPTLDPEEK' | 'Afamin' |
| 11504 | 'AFM_No70' | 'ADKTYVPPPFSQDLFTFHADMCQSQNEELQR' | 'Afamin' |
| 11505 | 'AFM_No71' | 'MVQQECK' | 'Afamin' |
| 11506 | 'AFM_No72' | 'MVQQECKHFQNLGKDGLK' | 'Afamin' |
| 11507 | 'AFM_No73' | 'YAEDKFNETTEK' | 'Afamin' |
| 11508 | 'AFM_No74' | 'CMADKTLPECSK' | 'Afamin' |
| 11509 | 'AFM_No75' | 'DTSKVMNHICSKQDSISSK' | 'Afamin' |
| 11510 | 'AFM_No76' | 'AFSSYQKHVCGALLK' | 'Afamin' |
| 11511 | 'AFM_No77' | 'DTSKVMNHICSKQDSISSK' | 'Afamin' |
| 11512 | 'AFM_No78' | 'DTSKVMNHICSK' | 'Afamin' |
| 11513 | 'PPIC_No1' | 'IVIGLFGK' | 'Peptidyl-prolyl cis-trans isomerase C' |
| 11514 | 'PPIC_No2' | 'TVENFVALATGEK' | 'Peptidyl-prolyl cis-trans isomerase C' |
| 11515 | 'VDAC2_No1' | 'VNNSSLIGVGYTQTLRPGVK' | 'Voltage-dependent anion-selective channel protein 2' |
| 11516 | 'VDAC2_No2' | 'LTLSALVDGK' | 'Voltage-dependent anion-selective channel protein 2' |
| 11517 | 'VDAC2_No3' | 'GFGFGLVK' | 'Voltage-dependent anion-selective channel protein 2' |
| 11518 | 'VDAC2_No4' | 'SCSGVEFSTSGSSNTDTGK' | 'Voltage-dependent anion-selective channel protein 2' |
| 11519 | 'CRKL' | 'IGDQEFDHLPALLEFYK' | 'Crk-like protein' |
| 11520 | 'CAPZA2_No1' | 'FTITPSTTQVVGILK' | 'F-actin-capping protein subunit alpha-2' |
| 11521 | 'CAPZA2_No2' | 'FIIHAPPGEFNEVFNDVR' | 'F-actin-capping protein subunit alpha-2' |
| 11522 | 'CAPZA2_No3' | 'ADLEEQLSDEEKVR' | 'F-actin-capping protein subunit alpha-2' |
| 11523 | 'CAPZA2_No4' | 'DIQDSLTVSNEVQTAK' | 'F-actin-capping protein subunit alpha-2' |
| 11524 | 'CAPZB_No1' | 'LVEDMENKIR' | 'F-actin-capping protein subunit beta' |
| 11525 | 'CAPZB_No2' | 'QMEKDETVSDCSPHIANIGR' | 'F-actin-capping protein subunit beta' |
| 11526 | 'CAPZB_No3' | 'GCWDSIHVVEVQEK' | 'F-actin-capping protein subunit beta' |
| 11527 | 'CAPZB_No4' | 'SPWSNKYDPPLEDGAMPSAR' | 'F-actin-capping protein subunit beta' |
| 11528 | 'CAPZB_No5' | 'KLEVEANNAFDQYR' | 'F-actin-capping protein subunit beta' |
| 11529 | 'CAPZB_No6' | 'SGSGTMNLGGSLTR' | 'F-actin-capping protein subunit beta' |

| | A | B | C |
|---|---|---|---|
| 1 | **Peptide Name** | **Peptide Sequence** | **Peptide Description** |
| 11530 | 'CAPZB_No7' | 'SGSGTMNLGGSLTR' | 'F-actin-capping protein subunit beta' |
| 11531 | 'CAPZB_No8' | 'DKVVGKDYLLCDYNR' | 'F-actin-capping protein subunit beta' |
| 11532 | 'CAPZB_No9' | 'VVGKDYLLCDYNR' | 'F-actin-capping protein subunit beta' |
| 11533 | 'CAPZB_No10' | 'STLNEIYFGK' | 'F-actin-capping protein subunit beta' |
| 11534 | 'CAPZB_No11' | 'LTSTVMLWLQTNK' | 'F-actin-capping protein subunit beta' |
| 11535 | 'CAPZB_No12' | 'SDQQLDCALDLMR' | 'F-actin-capping protein subunit beta' |
| 11536 | 'CAPZB_No13' | 'QMEKDETVSDCSPHIANIGR' | 'F-actin-capping protein subunit beta' |
| 11537 | 'CAPZB_No14' | 'LVEDMENKIR' | 'F-actin-capping protein subunit beta' |
| 11538 | 'LGALS7' | 'AVVGDAQYHHFR' | 'Galectin-7' |
| 11539 | 'LIMS1_No1' | 'MGVPICGACR' | 'LIM and senescent cell antigen-like-containing domain protein 1' |
| 11540 | 'LIMS1_No2' | 'AWCVNCFACSTCNTK' | 'LIM and senescent cell antigen-like-containing domain protein 1' |
| 11541 | 'LIMS1_No3' | 'NDPYHPDHFNCANCGK' | 'LIM and senescent cell antigen-like-containing domain protein 1' |
| 11542 | 'LIMS1_No4' | 'VIEGDVVSALNK' | 'LIM and senescent cell antigen-like-containing domain protein 1' |
| 11543 | 'LIMS1_No5' | 'AMNNSWHPECFR' | 'LIM and senescent cell antigen-like-containing domain protein 1' |
| 11544 | 'LIMS1_No6' | 'ELKGELYCLPCHDK' | 'LIM and senescent cell antigen-like-containing domain protein 1' |
| 11545 | 'LIMS1_No7' | 'FVEFDMKPVCK' | 'LIM and senescent cell antigen-like-containing domain protein 1' |
| 11546 | 'LIMS1_No8' | 'KGLAYCETHYNQLFGDVCFHCNR' | 'LIM and senescent cell antigen-like-containing domain protein 1' |
| 11547 | 'LIMS1_No9' | 'GLAYCETHYNQLFGDVCFHCNR' | 'LIM and senescent cell antigen-like-containing domain protein 1' |
| 11548 | 'LIMS1_No10' | 'GELYCLPCHDK' | 'LIM and senescent cell antigen-like-containing domain protein 1' |
| 11549 | 'LIMS1_No11' | 'FVEFDMKPVCK' | 'LIM and senescent cell antigen-like-containing domain protein 1' |
| 11550 | 'LIMS1_No12' | 'AMNNSWHPECFR' | 'LIM and senescent cell antigen-like-containing domain protein 1' |
| 11551 | 'PIP4K2A_No1' | 'FGIDDQDFQNSLTR' | 'Phosphatidylinositol 5-phosphate 4-kinase type-2 alpha' |
| 11552 | 'PIP4K2A_No2' | 'HGAGAEISTVNPEQYSK' | 'Phosphatidylinositol 5-phosphate 4-kinase type-2 alpha' |
| 11553 | 'GCLM' | 'EFPDVLECTVSHAVEK' | 'Glutamate--cysteine ligase regulatory subunit' |
| 11554 | 'GSS_No1' | 'EGIAQTVFLGLNR' | 'Glutathione synthetase' |
| 11555 | 'GSS_No2' | 'ALAEGVLLR' | 'Glutathione synthetase' |
| 11556 | 'GSS_No3' | 'AIEHADGGVAAGVAVLDNPYPV' | 'Glutathione synthetase' |
| 11557 | 'IDH2_No1' | 'LVPGWTKPITIGR' | 'Isocitrate dehydrogenase [NADP], mitochondrial' |
| 11558 | 'IDH2_No2' | 'IIWQFIK' | 'Isocitrate dehydrogenase [NADP], mitochondrial' |
| 11559 | 'IDH2_No3' | 'NILGGTVFREPIICK' | 'Isocitrate dehydrogenase [NADP], mitochondrial' |
| 11560 | 'IDH2_No4' | 'LIDDMVAQVLK' | 'Isocitrate dehydrogenase [NADP], mitochondrial' |
| 11561 | 'IDH2_No5' | 'GRPTSTNPIASIFAWTR' | 'Isocitrate dehydrogenase [NADP], mitochondrial' |
| 11562 | 'IDH2_No6' | 'VAKPVVEMDGDEMTR' | 'Isocitrate dehydrogenase [NADP], mitochondrial' |
| 11563 | 'IDH2_No7' | 'FKDIFQEIFDK' | 'Isocitrate dehydrogenase [NADP], mitochondrial' |
| 11564 | 'IDH2_No8' | 'VCVETVESGAMTK' | 'Isocitrate dehydrogenase [NADP], mitochondrial' |
| 11565 | 'IDH2_No9' | 'LILPHVDIQLK' | 'Isocitrate dehydrogenase [NADP], mitochondrial' |
| 11566 | 'IDH2_No10' | 'YFDLGLPNR' | 'Isocitrate dehydrogenase [NADP], mitochondrial' |
| 11567 | 'MASP1_No1' | 'VECSDNLFTQR' | 'Mannan-binding lectin serine protease 1' |
| 11568 | 'MASP1_No2' | 'SLPTCLPVCGLPK' | 'Mannan-binding lectin serine protease 1' |
| 11569 | 'MASP1_No3' | 'TGVITSPDFPNPYPK' | 'Mannan-binding lectin serine protease 1' |
| 11570 | 'MASP1_No4' | 'AAGNECPELQPPVHGK' | 'Mannan-binding lectin serine protease 1' |
| 11571 | 'MASP1_No5' | 'APEPISTQSHSVLILFHSDNSGENR' | 'Mannan-binding lectin serine protease 1' |
| 11572 | 'MASP1_No6' | 'APGELEHGLITFSTR' | 'Mannan-binding lectin serine protease 1' |
| 11573 | 'MASP1_No7' | 'VLGPFCGEK' | 'Mannan-binding lectin serine protease 1' |

| | A | B | C |
|---|---|---|---|
| 1 | **Peptide Name** | **Peptide Sequence** | **Peptide Description** |
| 11574 | 'MASP1_No8' | 'YSCQEPYYK' | 'Mannan-binding lectin serine protease 1' |
| 11575 | 'MASP1_No9' | 'DACAGDSGGPMVTLNR' | 'Mannan-binding lectin serine protease 1' |
| 11576 | 'MASP1_No10' | 'DACAGDSGGPMVTLNR' | 'Mannan-binding lectin serine protease 1' |
| 11577 | 'MASP1_No11' | 'DSDLLSPSDFK' | 'Mannan-binding lectin serine protease 1' |
| 11578 | 'MASP1_No12' | 'EGGKDACAGDSGGPMVTLNR' | 'Mannan-binding lectin serine protease 1' |
| 11579 | 'MASP1_No13' | 'FPETLMEIEIPIVDHSTCQK' | 'Mannan-binding lectin serine protease 1' |
| 11580 | 'MASP1_No14' | 'VLKDNVEMDTFQIECLK' | 'Mannan-binding lectin serine protease 1' |
| 11581 | 'MASP1_No15' | 'YGVYSYIHHNKDWIQR' | 'Mannan-binding lectin serine protease 1' |
| 11582 | 'MASP1_No16' | 'YGVYSYIHHNK' | 'Mannan-binding lectin serine protease 1' |
| 11583 | 'MASP1_No17' | 'YFFKDQVLVSCDTGYK' | 'Mannan-binding lectin serine protease 1' |
| 11584 | 'MASP1_No18' | 'DNVEMDTFQIECLK' | 'Mannan-binding lectin serine protease 1' |
| 11585 | 'MASP1_No19' | 'AAGNECPELQPPVHGKIEPSQAK' | 'Mannan-binding lectin serine protease 1' |
| 11586 | 'MASP1_No20' | 'DRYGVYSYIHHNKDWIQR' | 'Mannan-binding lectin serine protease 1' |
| 11587 | 'MASP1_No21' | 'VLKDNVEMDTFQIECLK' | 'Mannan-binding lectin serine protease 1' |
| 11588 | 'MASP1_No22' | 'EGGKDACAGDSGGPMVTLNR' | 'Mannan-binding lectin serine protease 1' |
| 11589 | 'MASP1_No23' | 'LRSDENEQHLGVK' | 'Mannan-binding lectin serine protease 1' |
| 11590 | 'MASP1_No24' | 'DNVEMDTFQIECLK' | 'Mannan-binding lectin serine protease 1' |
| 11591 | 'MASP1_No25' | 'FPETLMEIEIPIVDHSTCQK' | 'Mannan-binding lectin serine protease 1' |
| 11592 | 'MASP1_No26' | 'TCRVECSDNLFTQR' | 'Mannan-binding lectin serine protease 1' |
| 11593 | 'MASP1_No27' | 'DMICAGEKEGGKDACAGDSGGPMVTLNR' | 'Mannan-binding lectin serine protease 1' |
| 11594 | 'MASP1_No28' | 'FTGFDAHYMAVDVDECKER' | 'Mannan-binding lectin serine protease 1' |
| 11595 | 'MASP1_No29' | 'SDFSNEER' | 'Mannan-binding lectin serine protease 1' |
| 11596 | 'MASP1_No30' | 'FTGFDAHYMAVDVDECK' | 'Mannan-binding lectin serine protease 1' |
| 11597 | 'MASP1_No31' | 'ETTDTEQTPGQEVVLSPGSFMSITFR' | 'Mannan-binding lectin serine protease 1' |
| 11598 | 'MASP1_No32' | 'IVDCRAPGELEHGLITFSTR' | 'Mannan-binding lectin serine protease 1' |
| 11599 | 'MASP1_No33' | 'DRYGVYSYIHHNK' | 'Mannan-binding lectin serine protease 1' |
| 11600 | 'MASP1_No34' | 'DGTWSNKIPTCK' | 'Mannan-binding lectin serine protease 1' |
| 11601 | 'MASP1_No35' | 'FTGFDAHYMAVDVDECKER' | 'Mannan-binding lectin serine protease 1' |
| 11602 | 'MASP1_No36' | 'FGYILHTDNR' | 'Mannan-binding lectin serine protease 1' |
| 11603 | 'LMAN1' | 'YVSSLTEEISKR' | 'Protein ERGIC-53' |
| 11604 | 'CCT3_No1' | 'NVLLDPQLVPGGGASEMAVAHALTEK' | 'T-complex protein 1 subunit gamma' |
| 11605 | 'CCT3_No2' | 'TAVETAVLLLR' | 'T-complex protein 1 subunit gamma' |
| 11606 | 'CCT3_No3' | 'IPGGIIEDSCVLR' | 'T-complex protein 1 subunit gamma' |
| 11607 | 'CCT3_No4' | 'IVLLDSSLEYK' | 'T-complex protein 1 subunit gamma' |
| 11608 | 'CCT3_No5' | 'AVAQALEVIPR' | 'T-complex protein 1 subunit gamma' |
| 11609 | 'CCT3_No6' | 'WSSLACNIALDAVK' | 'T-complex protein 1 subunit gamma' |
| 11610 | 'ARRB1_No1' | 'VQYAPERPGPQPTAETTR' | 'Beta-arrestin-1' |
| 11611 | 'ARRB1_No2' | 'CPVAMEEADDTVAPSSTFCK' | 'Beta-arrestin-1' |
| 11612 | 'ARRB1_No3' | 'KVQYAPERPGPQPTAETTR' | 'Beta-arrestin-1' |
| 11613 | 'TUFM_No1' | 'DLEKPFLLPVEAVYSVPGR' | 'Elongation factor Tu, mitochondrial' |
| 11614 | 'TUFM_No2' | 'LLDAVDTYIPVPAR' | 'Elongation factor Tu, mitochondrial' |
| 11615 | 'TUFM_No3' | 'TIGTGLVTNTLAMTEEEK' | 'Elongation factor Tu, mitochondrial' |
| 11616 | 'PSMB2' | 'FILNLPTFSVR' | 'Proteasome subunit beta type-2' |
| 11617 | 'COMP_No1' | 'DSDGDGIGDACDNCPQK' | 'Cartilage oligomeric matrix protein' |

| | A | B | C |
|---|---|---|---|
| 1 | **Peptide Name** | **Peptide Sequence** | **Peptide Description** |
| 11618 | 'COMP_No2' | 'FYEGPELVADSNVVLDTTMR' | 'Cartilage oligomeric matrix protein' |
| 11619 | 'COMP_No3' | 'NTDEDKWGDACDNCR' | 'Cartilage oligomeric matrix protein' |
| 11620 | 'COMP_No4' | 'AFQTVVLDPEGDAQIDPNWVVLNQGR' | 'Cartilage oligomeric matrix protein' |
| 11621 | 'COMP_No5' | 'NTVMECDACGMQQSVR' | 'Cartilage oligomeric matrix protein' |
| 11622 | 'COMP_No6' | 'NALWHTGDTESQVR' | 'Cartilage oligomeric matrix protein' |
| 11623 | 'COMP_No7' | 'DTDLDGFPDEKLR' | 'Cartilage oligomeric matrix protein' |
| 11624 | 'COMP_No8' | 'FCPDGSPSECHEHADCVLER' | 'Cartilage oligomeric matrix protein' |
| 11625 | 'COMP_No9' | 'VPNSDQKDSDGDGIGDACDNCPQK' | 'Cartilage oligomeric matrix protein' |
| 11626 | 'COMP_No10' | 'NDDQKDTDQDGRGDACDDDIDGDR' | 'Cartilage oligomeric matrix protein' |
| 11627 | 'COMP_No11' | 'QMEQTYWQANPFR' | 'Cartilage oligomeric matrix protein' |
| 11628 | 'COMP_No12' | 'SCVCAVGWAGNGILCGR' | 'Cartilage oligomeric matrix protein' |
| 11629 | 'COMP_No13' | 'SNPDQADVDHDFVGDACDSDQDQDGDGHQDSR' | 'Cartilage oligomeric matrix protein' |
| 11630 | 'COMP_No14' | 'ELQETNAALQDVR' | 'Cartilage oligomeric matrix protein' |
| 11631 | 'COMP_No15' | 'DNCVTVPNSGQEDVDR' | 'Cartilage oligomeric matrix protein' |
| 11632 | 'COMP_No16' | 'DGIGDACDPDADGDGVPNEK' | 'Cartilage oligomeric matrix protein' |
| 11633 | 'COMP_No17' | 'LVPNPGQEDADRDGVGDVCQDDFDADK' | 'Cartilage oligomeric matrix protein' |
| 11634 | 'COMP_No18' | 'QVCTDINECETGQHNCVPNSVCINTR' | 'Cartilage oligomeric matrix protein' |
| 11635 | 'COMP_No19' | 'QMEQTYWQANPFR' | 'Cartilage oligomeric matrix protein' |
| 11636 | 'COMP_No20' | 'NTVMECDACGMQQSVR' | 'Cartilage oligomeric matrix protein' |
| 11637 | 'COMP_No21' | 'QMEQTYWQANPFR' | 'Cartilage oligomeric matrix protein' |
| 11638 | 'COMP_No22' | 'DTDQDGRGDACDDDIDGDR' | 'Cartilage oligomeric matrix protein' |
| 11639 | 'COMP_No23' | 'CEACPPGYSGPTHQGVGLAFAK' | 'Cartilage oligomeric matrix protein' |
| 11640 | 'COMP_No24' | 'DTDQDGRGDACDDDIDGDRIR' | 'Cartilage oligomeric matrix protein' |
| 11641 | 'COMP_No25' | 'ANKQVCTDINECETGQHNCVPNSVCINTR' | 'Cartilage oligomeric matrix protein' |
| 11642 | 'COMP_No26' | 'VVDKIDVCPENAEVTLTDFR' | 'Cartilage oligomeric matrix protein' |
| 11643 | 'COMP_No27' | 'QMEQTYWQANPFR' | 'Cartilage oligomeric matrix protein' |
| 11644 | 'COMP_No28' | 'FCPDGSPSECHEHADCVLERDGSR' | 'Cartilage oligomeric matrix protein' |
| 11645 | 'COMP_No29' | 'FYEGPELVADSNVVLDTTMR' | 'Cartilage oligomeric matrix protein' |
| 11646 | 'COMP_No30' | 'NTVMECDACGMQQSVR' | 'Cartilage oligomeric matrix protein' |
| 11647 | 'COMP_No31' | 'GDACDDDIDGDRIR' | 'Cartilage oligomeric matrix protein' |
| 11648 | 'COMP_No32' | 'DTDLDGFPDEKLRCPER' | 'Cartilage oligomeric matrix protein' |
| 11649 | 'COMP_No33' | 'NTVMECDACGMQQSVR' | 'Cartilage oligomeric matrix protein' |
| 11650 | 'COMP_No34' | 'GSFQCGPCQPGFVGDQASGCQRR' | 'Cartilage oligomeric matrix protein' |
| 11651 | 'SELENOP_No1' | 'LPTDSELAPR' | 'Selenoprotein P' |
| 11652 | 'SELENOP_No2' | 'DMPASEDLQDLQKK' | 'Selenoprotein P' |
| 11653 | 'SELENOP_No3' | 'CINQLLCK' | 'Selenoprotein P' |
| 11654 | 'SELENOP_No4' | 'DMPASEDLQDLQK' | 'Selenoprotein P' |
| 11655 | 'SELENOP_No5' | 'DMPASEDLQDLQKK' | 'Selenoprotein P' |
| 11656 | 'SELENOP_No6' | 'QPPAWSIR' | 'Selenoprotein P' |
| 11657 | 'SELENOP_No7' | 'EGYSNISYIVVNHQGISSR' | 'Selenoprotein P' |
| 11658 | 'SELENOP_No8' | 'QPPAWSIR' | 'Selenoprotein P' |
| 11659 | 'SELENOP_No9' | 'CINQLLCKLPTDSELAPR' | 'Selenoprotein P' |
| 11660 | 'SELENOP_No10' | 'QGHPENRDMPASEDLQDLQK' | 'Selenoprotein P' |
| 11661 | 'CAMP_No1' | 'FALLGDFFR' | 'Cathelicidin antimicrobial peptide' |

| | A | B | C |
|---|---|---|---|
| 1 | **Peptide Name** | **Peptide Sequence** | **Peptide Description** |
| 11662 | 'CAMP_No2' | 'FALLGDFFRK' | 'Cathelicidin antimicrobial peptide' |
| 11663 | 'CAMP_No3' | 'CMGTVTLNQAR' | 'Cathelicidin antimicrobial peptide' |
| 11664 | 'CAMP_No4' | 'TTQQSPEDCDFKK' | 'Cathelicidin antimicrobial peptide' |
| 11665 | 'CAMP_No5' | 'CMGTVTLNQAR' | 'Cathelicidin antimicrobial peptide' |
| 11666 | 'CAMP_No6' | 'TTQQSPEDCDFKKDGLVK' | 'Cathelicidin antimicrobial peptide' |
| 11667 | 'SULT1A1_No1' | 'THLPLALLPQTLLDQK' | 'Sulfotransferase 1A1' |
| 11668 | 'SULT1A1_No2' | 'MELIQDTSRPPLEYVK' | 'Sulfotransferase 1A1' |
| 11669 | 'SULT1A1_No3' | 'VHPEPGTWDSFLEK' | 'Sulfotransferase 1A1' |
| 11670 | 'GDI2_No1' | 'NTNDANSCQIIIPQNQVNR' | 'Rab GDP dissociation inhibitor beta' |
| 11671 | 'GDI2_No2' | 'FVSISDLLVPK' | 'Rab GDP dissociation inhibitor beta' |
| 11672 | 'GDI2_No3' | 'TYDATTHFETTCDDIK' | 'Rab GDP dissociation inhibitor beta' |
| 11673 | 'GDI2_No4' | 'LSAIYGGTYMLNKPIEEIIVQNGK' | 'Rab GDP dissociation inhibitor beta' |
| 11674 | 'GDI2_No5' | 'FKIPGSPPESMGR' | 'Rab GDP dissociation inhibitor beta' |
| 11675 | 'GDI2_No6' | 'FLVYVANFDEKDPR' | 'Rab GDP dissociation inhibitor beta' |
| 11676 | 'GDI2_No7' | 'SDIYVCMISFAHNVAAQGK' | 'Rab GDP dissociation inhibitor beta' |
| 11677 | 'GDI2_No8' | 'FLMANGQLVK' | 'Rab GDP dissociation inhibitor beta' |
| 11678 | 'GDI2_No9' | 'NPYYGGESASITPLEDLYKR' | 'Rab GDP dissociation inhibitor beta' |
| 11679 | 'GDI2_No10' | 'NPYYGGESASITPLEDLYK' | 'Rab GDP dissociation inhibitor beta' |
| 11680 | 'GDI2_No11' | 'SPYLYPLYGLGELPQGFAR' | 'Rab GDP dissociation inhibitor beta' |
| 11681 | 'GDI2_No12' | 'DLGTESQIFISR' | 'Rab GDP dissociation inhibitor beta' |
| 11682 | 'ST13_No1' | 'VAAIEALNDGELQK' | 'Hsc70-interacting protein' |
| 11683 | 'ST13_No2' | 'AIDLFTDAIK' | 'Hsc70-interacting protein' |
| 11684 | 'ST13_No3' | 'LLGHWEEAAHDLALACK' | 'Hsc70-interacting protein' |
| 11685 | 'ST13_No4' | 'QDPSVLHTEEMR' | 'Hsc70-interacting protein' |
| 11686 | 'ST13_No5' | 'AIDLFTDAIKLNPR' | 'Hsc70-interacting protein' |
| 11687 | 'VASP_No1' | 'ATVMLYDDGNKR' | 'Vasodilator-stimulated phosphoprotein' |
| 11688 | 'VASP_No2' | 'VKEEIIEAFVQELR' | 'Vasodilator-stimulated phosphoprotein' |
| 11689 | 'VASP_No3' | 'YNQATPNFHQWR' | 'Vasodilator-stimulated phosphoprotein' |
| 11690 | 'VASP_No4' | 'VKQELLEEVKK' | 'Vasodilator-stimulated phosphoprotein' |
| 11691 | 'VASP_No5' | 'MQPDQQVVINCAIVR' | 'Vasodilator-stimulated phosphoprotein' |
| 11692 | 'VASP_No6' | 'SGGGGLMEEMNAMLAR' | 'Vasodilator-stimulated phosphoprotein' |
| 11693 | 'VASP_No7' | 'ATVMLYDDGNKR' | 'Vasodilator-stimulated phosphoprotein' |
| 11694 | 'VASP_No8' | 'KMQPDQQVVINCAIVR' | 'Vasodilator-stimulated phosphoprotein' |
| 11695 | 'VASP_No9' | 'QVWGLNFGSK' | 'Vasodilator-stimulated phosphoprotein' |
| 11696 | 'VASP_No10' | 'ATQVGEKTPKDESANQEEPEAR' | 'Vasodilator-stimulated phosphoprotein' |
| 11697 | 'VASP_No11' | 'KMQPDQQVVINCAIVR' | 'Vasodilator-stimulated phosphoprotein' |
| 11698 | 'DNM2_No1' | 'VYSPHVLNLTLIDLPGITK' | 'Dynamin-2' |
| 11699 | 'DNM2_No2' | 'ESSLILAVTPANMDLANSDALK' | 'Dynamin-2' |
| 11700 | 'CCT4_No1' | 'AYILNLVK' | 'T-complex protein 1 subunit delta' |
| 11701 | 'CCT4_No2' | 'LGGTIDDCELVEGLVLTQK' | 'T-complex protein 1 subunit delta' |
| 11702 | 'CCT4_No3' | 'DALSDLALHFLNK' | 'T-complex protein 1 subunit delta' |
| 11703 | 'RAB5C_No1' | 'TAMNVNEIFMAIAK' | 'Ras-related protein Rab-5C' |
| 11704 | 'RAB5C_No2' | 'GAQAAIVVYDITNTDTFAR' | 'Ras-related protein Rab-5C' |
| 11705 | 'RAB5C_No3' | 'GVDLQENNPASR' | 'Ras-related protein Rab-5C' |

| | A | B | C |
|---|---|---|---|
| 1 | **Peptide Name** | **Peptide Sequence** | **Peptide Description** |
| 11706 | 'RAB7A_No1' | 'FQSLGVAFYR' | 'Ras-related protein Rab-7a' |
| 11707 | 'RAB7A_No2' | 'DPENFPFVVLGNK' | 'Ras-related protein Rab-7a' |
| 11708 | 'HSD17B4' | 'LGLLGLANSLAIEGR' | 'Peroxisomal multifunctional enzyme type 2' |
| 11709 | 'PSMD7' | 'VVVHPLVLLSVVDHFNR' | '26S proteasome non-ATPase regulatory subunit 7' |
| 11710 | 'LUM_No1' | 'NIPTVNENLENYYLEVNQLEK' | 'Lumican' |
| 11711 | 'LUM_No2' | 'FNALQYLR' | 'Lumican' |
| 11712 | 'LUM_No3' | 'SLEDLQLTHNK' | 'Lumican' |
| 11713 | 'LUM_No4' | 'ISETSLPPDMYECLR' | 'Lumican' |
| 11714 | 'LUM_No5' | 'ILGPLSYSK' | 'Lumican' |
| 11715 | 'LUM_No6' | 'RFNALQYLR' | 'Lumican' |
| 11716 | 'LUM_No7' | 'ISETSLPPDMYECLR' | 'Lumican' |
| 11717 | 'LUM_No8' | 'ISNIPDEYFKR' | 'Lumican' |
| 11718 | 'LUM_No9' | 'ISNIPDEYFK' | 'Lumican' |
| 11719 | 'LUM_No10' | 'NIPTVNENLENYYLEVNQLEKFDIK' | 'Lumican' |
| 11720 | 'LUM_No11' | 'SLEYLDLSFNQIAR' | 'Lumican' |
| 11721 | 'LUM_No12' | 'LPSGLPVSLLTLYLDNNK' | 'Lumican' |
| 11722 | 'LUM_No13' | 'LKEDAVSAAFK' | 'Lumican' |
| 11723 | 'LUM_No14' | 'SLEDLQLTHNKITK' | 'Lumican' |
| 11724 | 'LUM_No15' | 'NNQIDHIDEK' | 'Lumican' |
| 11725 | 'LUM_No16' | 'LDGNRISETSLPPDMYECLR' | 'Lumican' |
| 11726 | 'LUM_No17' | 'LKEDAVSAAFKGLK' | 'Lumican' |
| 11727 | 'LUM_No18' | 'SVPMVPPGIK' | 'Lumican' |
| 11728 | 'LUM_No19' | 'LKNIPTVNENLENYYLEVNQLEKFDIK' | 'Lumican' |
| 11729 | 'LUM_No20' | 'LPSGLPVSLLTLYLDNNKISNIPDEYFK' | 'Lumican' |
| 11730 | 'LUM_No21' | 'SVPMVPPGIK' | 'Lumican' |
| 11731 | 'LUM_No22' | 'LKNIPTVNENLENYYLEVNQLEK' | 'Lumican' |
| 11732 | 'PGD_No1' | 'VGTGEPCCDWVGDEGAGHFVK' | '6-phosphogluconate dehydrogenase, decarboxylating' |
| 11733 | 'PGD_No2' | 'SFLEDIRK' | '6-phosphogluconate dehydrogenase, decarboxylating' |
| 11734 | 'PGD_No3' | 'DYFGAHTYELLAK' | '6-phosphogluconate dehydrogenase, decarboxylating' |
| 11735 | 'PGD_No4' | 'IISYAQGFMLLR' | '6-phosphogluconate dehydrogenase, decarboxylating' |
| 11736 | 'PGD_No5' | 'TVSKVDDFLANEAK' | '6-phosphogluconate dehydrogenase, decarboxylating' |
| 11737 | 'PGD_No6' | 'IISYAQGFMLLR' | '6-phosphogluconate dehydrogenase, decarboxylating' |
| 11738 | 'ARHGDIA_No1' | 'SIQEIQELDKDDESLRK' | 'Rho GDP-dissociation inhibitor 1' |
| 11739 | 'ARHGDIA_No2' | 'AEEYEFLTPVEEAPK' | 'Rho GDP-dissociation inhibitor 1' |
| 11740 | 'ARHGDIA_No3' | 'AEQEPTAEQLAQIAAENEEDEHSVNYKPPAQK' | 'Rho GDP-dissociation inhibitor 1' |
| 11741 | 'ARHGDIA_No4' | 'SIQEIQELDKDDESLR' | 'Rho GDP-dissociation inhibitor 1' |
| 11742 | 'ARHGDIB_No1' | 'LTLVCESAPGPITMDLTGDLEALKK' | 'Rho GDP-dissociation inhibitor 2' |
| 11743 | 'ARHGDIB_No2' | 'ATFMVGSYGPRPEEYEFLTPVEEAPK' | 'Rho GDP-dissociation inhibitor 2' |
| 11744 | 'ARHGDIB_No3' | 'ELQEMDKDDESLIK' | 'Rho GDP-dissociation inhibitor 2' |
| 11745 | 'ARHGDIB_No4' | 'ATFMVGSYGPRPEEYEFLTPVEEAPK' | 'Rho GDP-dissociation inhibitor 2' |
| 11746 | 'ARHGDIB_No5' | 'LTLVCESAPGPITMDLTGDLEALKK' | 'Rho GDP-dissociation inhibitor 2' |
| 11747 | 'ARHGDIB_No6' | 'SLKELQEMDKDDESLIK' | 'Rho GDP-dissociation inhibitor 2' |
| 11748 | 'ARHGDIB_No7' | 'APEPHVEEDDDDELDSK' | 'Rho GDP-dissociation inhibitor 2' |
| 11749 | 'ARHGDIB_No8' | 'APEPHVEEDDDDELDSKLNYKPPPQK' | 'Rho GDP-dissociation inhibitor 2' |

| | A | B | C |
|---|---|---|---|
| 1 | **Peptide Name** | **Peptide Sequence** | **Peptide Description** |
| 11750 | 'ARHGDIB_No9' | 'TLLGDGPVVTDPK' | 'Rho GDP-dissociation inhibitor 2' |
| 11751 | 'ARHGDIB_No10' | 'TLLGDGPVVTDPKAPNVVVTR' | 'Rho GDP-dissociation inhibitor 2' |
| 11752 | 'ARHGDIB_No11' | 'ELQEMDKDDESLIK' | 'Rho GDP-dissociation inhibitor 2' |
| 11753 | 'CAPZA1_No1' | 'EASDPQPEEADGGLK' | 'F-actin-capping protein subunit alpha-1' |
| 11754 | 'CAPZA1_No2' | 'FTITPPTAQVVGVLK' | 'F-actin-capping protein subunit alpha-1' |
| 11755 | 'CAPZA1_No3' | 'TIDGQQTIIACIESHQFQPK' | 'F-actin-capping protein subunit alpha-1' |
| 11756 | 'CAPZA1_No4' | 'ADFDDRVSDEEKVR' | 'F-actin-capping protein subunit alpha-1' |
| 11757 | 'CAPZA1_No5' | 'LLLNNDNLLR' | 'F-actin-capping protein subunit alpha-1' |
| 11758 | 'CAPZA1_No6' | 'IQVHYYEDGNVQLVSHK' | 'F-actin-capping protein subunit alpha-1' |
| 11759 | 'BLVRA' | 'NPHPSSAFLNLIGFVSR' | 'Biliverdin reductase A' |
| 11760 | 'CTSC_No1' | 'ILHLPTSWDWR' | 'Dipeptidyl peptidase 1' |
| 11761 | 'CTSC_No2' | 'GTDECAIESIAVAATPIPK' | 'Dipeptidyl peptidase 1' |
| 11762 | 'PTTG1IP' | 'ACLDYPVTSVLPPASLCK' | 'Pituitary tumor-transforming gene 1 protein-interacting protein' |
| 11763 | 'CRISP3_No1' | 'YYYVCQYCPAGNWANR' | 'Cysteine-rich secretory protein 3' |
| 11764 | 'CRISP3_No2' | 'TPNAVVGHYTQVVWYSSYLVGCGNAYCPNQK' | 'Cysteine-rich secretory protein 3' |
| 11765 | 'CRISP3_No3' | 'YEDLYSNCK' | 'Cysteine-rich secretory protein 3' |
| 11766 | 'CRISP3_No4' | 'MEWNKEAAANAQK' | 'Cysteine-rich secretory protein 3' |
| 11767 | 'CRISP3_No5' | 'LYVPYEQGAPCASCPDNCDDGLCTNGCK' | 'Cysteine-rich secretory protein 3' |
| 11768 | 'CACNA2D1_No1' | 'PIGVGIPTINLR' | 'Voltage-dependent calcium channel subunit alpha-2/delta-1' |
| 11769 | 'CACNA2D1_No2' | 'YQDLYTVEPNNAR' | 'Voltage-dependent calcium channel subunit alpha-2/delta-1' |
| 11770 | 'CACNA2D1_No3' | 'KTPNNPSCNADLINR' | 'Voltage-dependent calcium channel subunit alpha-2/delta-1' |
| 11771 | 'CACNA2D1_No4' | 'GYYYEIPSIGAIR' | 'Voltage-dependent calcium channel subunit alpha-2/delta-1' |
| 11772 | 'CACNA2D1_No5' | 'TPNNPSCNADLINR' | 'Voltage-dependent calcium channel subunit alpha-2/delta-1' |
| 11773 | 'CACNA2D1_No6' | 'TASGVNQLVDIYEKYQDLYTVEPNNAR' | 'Voltage-dependent calcium channel subunit alpha-2/delta-1' |
| 11774 | 'CACNA2D1_No7' | 'INTQEYLDVLGRPMVLAGDK' | 'Voltage-dependent calcium channel subunit alpha-2/delta-1' |
| 11775 | 'CACNA2D1_No8' | 'IIMLFTDGGEER' | 'Voltage-dependent calcium channel subunit alpha-2/delta-1' |
| 11776 | 'CACNA2D1_No9' | 'VLLDAGFTNELVQNYWSK' | 'Voltage-dependent calcium channel subunit alpha-2/delta-1' |
| 11777 | 'CACNA2D1_No10' | 'TASGVNQLVDIYEK' | 'Voltage-dependent calcium channel subunit alpha-2/delta-1' |
| 11778 | 'CACNA2D1_No11' | 'AKLEETITQAR' | 'Voltage-dependent calcium channel subunit alpha-2/delta-1' |
| 11779 | 'CACNA2D1_No12' | 'IKPVFIEDANFGR' | 'Voltage-dependent calcium channel subunit alpha-2/delta-1' |
| 11780 | 'CACNA2D1_No13' | 'TSIRDPCAGPVCDCK' | 'Voltage-dependent calcium channel subunit alpha-2/delta-1' |
| 11781 | 'RAD23B_No1' | 'QEKPAEKPAETPVATSPTATDSTSGDSSR' | 'UV excision repair protein RAD23 homolog B' |
| 11782 | 'RAD23B_No2' | 'ILNDDTALKEYK' | 'UV excision repair protein RAD23 homolog B' |
| 11783 | 'NAGLU_No1' | 'FLLGSWLEQAR' | 'Alpha-N-acetylglucosaminidase' |
| 11784 | 'NAGLU_No2' | 'YDLLDLTR' | 'Alpha-N-acetylglucosaminidase' |
| 11785 | 'NAGLU_No3' | 'LLLTSAPSLATSPAFR' | 'Alpha-N-acetylglucosaminidase' |
| 11786 | 'NAGLU_No4' | 'GSTGVAAAAGLHR' | 'Alpha-N-acetylglucosaminidase' |
| 11787 | 'NAGLU_No5' | 'DFCGCHVAWSGSQLR' | 'Alpha-N-acetylglucosaminidase' |
| 11788 | 'NAGLU_No6' | 'QAVQELVSLYYEEAR' | 'Alpha-N-acetylglucosaminidase' |
| 11789 | 'NAGLU_No7' | 'YGVSHPDAGAAWR' | 'Alpha-N-acetylglucosaminidase' |
| 11790 | 'NAGLU_No8' | 'SFGMTPVLPAFAGHVPEAVTR' | 'Alpha-N-acetylglucosaminidase' |
| 11791 | 'NAGLU_No9' | 'PGLDTYSLGGGGAAR' | 'Alpha-N-acetylglucosaminidase' |
| 11792 | 'NAGLU_No10' | 'KDPVPDLAAWVTSFAAR' | 'Alpha-N-acetylglucosaminidase' |
| 11793 | 'NAPA_No1' | 'TIQGDEEDLR' | 'Alpha-soluble NSF attachment protein' |

| | A | B | C |
|---|---|---|---|
| 1 | **Peptide Name** | **Peptide Sequence** | **Peptide Description** |
| 11794 | 'NAPA_No2' | 'HDAATCFVDAGNAFK' | 'Alpha-soluble NSF attachment protein' |
| 11795 | 'NAPA_No3' | 'YEELFPAFSDSR' | 'Alpha-soluble NSF attachment protein' |
| 11796 | 'NAPA_No4' | 'AIAHYEQSADYYKGEESNSSANK' | 'Alpha-soluble NSF attachment protein' |
| 11797 | 'APOC4_No1' | 'GFMQTYYDDHLR' | 'Apolipoprotein C-IV' |
| 11798 | 'APOC4_No2' | 'MKELLETVVNR' | 'Apolipoprotein C-IV' |
| 11799 | 'APOC4_No3' | 'GFMQTYYDDHLR' | 'Apolipoprotein C-IV' |
| 11800 | 'APOC4_No4' | 'AWFLESK' | 'Apolipoprotein C-IV' |
| 11801 | 'APOC4_No5' | 'ELLETVVNR' | 'Apolipoprotein C-IV' |
| 11802 | 'APOC4_No6' | 'MKELLETVVNR' | 'Apolipoprotein C-IV' |
| 11803 | 'APOC4_No7' | 'GFMQTYYDDHLRDLGPLTK' | 'Apolipoprotein C-IV' |
| 11804 | 'APOC4_No8' | 'AWFLESKDSLLKK' | 'Apolipoprotein C-IV' |
| 11805 | 'APOC4_No9' | 'TRDGWQWFWSPSTFR' | 'Apolipoprotein C-IV' |
| 11806 | 'APOC4_No10' | 'GFMQTYYDDHLRDLGPLTK' | 'Apolipoprotein C-IV' |
| 11807 | 'APOC4_No11' | 'AWFLESKDSLLK' | 'Apolipoprotein C-IV' |
| 11808 | 'APOC4_No12' | 'LVCGDKDQG' | 'Apolipoprotein C-IV' |
| 11809 | 'APOC4_No13' | 'DGWQWFWSPSTFR' | 'Apolipoprotein C-IV' |
| 11810 | 'PLTP_No1' | 'AVEPQLQEEER' | 'Phospholipid transfer protein' |
| 11811 | 'PLTP_No2' | 'AGALQLLLVGDK' | 'Phospholipid transfer protein' |
| 11812 | 'PLTP_No3' | 'TMLQIGVMPMLNER' | 'Phospholipid transfer protein' |
| 11813 | 'PLTP_No4' | 'TGLELSRDPAGR' | 'Phospholipid transfer protein' |
| 11814 | 'PLTP_No5' | 'GAFFPLTER' | 'Phospholipid transfer protein' |
| 11815 | 'PLTP_No6' | 'AGALQLLLVGDKVPHDLDMLLR' | 'Phospholipid transfer protein' |
| 11816 | 'PLTP_No7' | 'VPHDLDMLLR' | 'Phospholipid transfer protein' |
| 11817 | 'PLTP_No8' | 'MHAAFGGTFK' | 'Phospholipid transfer protein' |
| 11818 | 'PLTP_No9' | 'FLEQELETITIPDLR' | 'Phospholipid transfer protein' |
| 11819 | 'PLTP_No10' | 'VYDFLSTFITSGMR' | 'Phospholipid transfer protein' |
| 11820 | 'PLTP_No11' | 'TMLQIGVMPMLNER' | 'Phospholipid transfer protein' |
| 11821 | 'PLTP_No12' | 'TMLQIGVMPMLNER' | 'Phospholipid transfer protein' |
| 11822 | 'PLTP_No13' | 'ALELVKQEGLR' | 'Phospholipid transfer protein' |
| 11823 | 'PLTP_No14' | 'KVYDFLSTFITSGMR' | 'Phospholipid transfer protein' |
| 11824 | 'PLTP_No15' | 'AGALQLLLVGDKVPHDLDMLLR' | 'Phospholipid transfer protein' |
| 11825 | 'PLTP_No16' | 'FLEQELETITIPDLRGK' | 'Phospholipid transfer protein' |
| 11826 | 'PLTP_No17' | 'DPVASTSNLDMDFR' | 'Phospholipid transfer protein' |
| 11827 | 'PLTP_No18' | 'KVYDFLSTFITSGMR' | 'Phospholipid transfer protein' |
| 11828 | 'PLTP_No19' | 'IYSNHSALESLALIPLQAPLK' | 'Phospholipid transfer protein' |
| 11829 | 'PLTP_No20' | 'VPHDLDMLLR' | 'Phospholipid transfer protein' |
| 11830 | 'PLTP_No21' | 'TMLQIGVMPMLNER' | 'Phospholipid transfer protein' |
| 11831 | 'PLTP_No22' | 'SSVDELVGIDYSLMKDPVASTSNLDMDFR' | 'Phospholipid transfer protein' |
| 11832 | 'PLTP_No23' | 'TMLQIGVMPMLNER' | 'Phospholipid transfer protein' |
| 11833 | 'PLTP_No24' | 'TMLQIGVMPMLNER' | 'Phospholipid transfer protein' |
| 11834 | 'PLTP_No25' | 'ATYFGSIVLLSPAVIDSPLK' | 'Phospholipid transfer protein' |
| 11835 | 'PLTP_No26' | 'TMLQIGVMPMLNER' | 'Phospholipid transfer protein' |
| 11836 | 'PLTP_No27' | 'VYDFLSTFITSGMR' | 'Phospholipid transfer protein' |
| 11837 | 'VCP_No1' | 'LDQLIYIPLPDEK' | 'Transitional endoplasmic reticulum ATPase' |

| | A | B | C |
|---|---|---|---|
| 1 | **Peptide Name** | **Peptide Sequence** | **Peptide Description** |
| 11838 | 'VCP_No2' | 'LIVDEAINEDNSVVSLSQPK' | 'Transitional endoplasmic reticulum ATPase' |
| 11839 | 'VCP_No3' | 'LADDVDLEQVANETHGHVGADLAALCSEAALQAIR' | 'Transitional endoplasmic reticulum ATPase' |
| 11840 | 'VCP_No4' | 'AIANECQANFISIK' | 'Transitional endoplasmic reticulum ATPase' |
| 11841 | 'VCP_No5' | 'MDELQLFR' | 'Transitional endoplasmic reticulum ATPase' |
| 11842 | 'VCP_No6' | 'ASGADSKGDDLSTAILK' | 'Transitional endoplasmic reticulum ATPase' |
| 11843 | 'VCP_No7' | 'ETVVEVPQVTWEDIGGLEDVKR' | 'Transitional endoplasmic reticulum ATPase' |
| 11844 | 'VCP_No8' | 'IVSQLLTLMDGLK' | 'Transitional endoplasmic reticulum ATPase' |
| 11845 | 'VCP_No9' | 'ELQELVQYPVEHPDK' | 'Transitional endoplasmic reticulum ATPase' |
| 11846 | 'VCP_No10' | 'QAAPCVLFFDELDSIAK' | 'Transitional endoplasmic reticulum ATPase' |
| 11847 | 'VCP_No11' | 'KYEMFAQTLQQSR' | 'Transitional endoplasmic reticulum ATPase' |
| 11848 | 'VCP_No12' | 'KYEMFAQTLQQSR' | 'Transitional endoplasmic reticulum ATPase' |
| 11849 | 'VCP_No13' | 'WALSQSNPSALR' | 'Transitional endoplasmic reticulum ATPase' |
| 11850 | 'VCP_No14' | 'MDELQLFR' | 'Transitional endoplasmic reticulum ATPase' |
| 11851 | 'VCP_No15' | 'ESIESEIRR' | 'Transitional endoplasmic reticulum ATPase' |
| 11852 | 'VCP_No16' | 'LGDVISIQPCPDVK' | 'Transitional endoplasmic reticulum ATPase' |
| 11853 | 'VCP_No17' | 'DVDLEFLAK' | 'Transitional endoplasmic reticulum ATPase' |
| 11854 | 'MFAP4_No1' | 'ADGEYWLGLQNMHLLTLK' | 'Microfibril-associated glycoprotein 4' |
| 11855 | 'MFAP4_No2' | 'ADGEYWLGLQNMHLLTLK' | 'Microfibril-associated glycoprotein 4' |
| 11856 | 'INHBC_No1' | 'AGGQCPACGGPTLELESQR' | 'Inhibin beta C chain' |
| 11857 | 'INHBC_No2' | 'ANTAAGTTGGGSCCVPTAR' | 'Inhibin beta C chain' |
| 11858 | 'INHBC_No3' | 'ELLLDLAKR' | 'Inhibin beta C chain' |
| 11859 | 'INHBC_No4' | 'ELLLDLAK' | 'Inhibin beta C chain' |
| 11860 | 'INHBC_No5' | 'QEFFVDFR' | 'Inhibin beta C chain' |
| 11861 | 'INHBC_No6' | 'LDFHFSSDR' | 'Inhibin beta C chain' |
| 11862 | 'INHBC_No7' | 'RPLSLLYYDR' | 'Inhibin beta C chain' |
| 11863 | 'INHBC_No8' | 'RPLSLLYYDRDSNIVK' | 'Inhibin beta C chain' |
| 11864 | 'INHBC_No9' | 'AGGQCPACGGPTLELESQRELLLDLAKR' | 'Inhibin beta C chain' |
| 11865 | 'MANF_No1' | 'IINEVSKPLAHHIPVEK' | 'Mesencephalic astrocyte-derived neurotrophic factor' |
| 11866 | 'MANF_No2' | 'QIDLSTVDLKK' | 'Mesencephalic astrocyte-derived neurotrophic factor' |
| 11867 | 'MANF_No3' | 'DVTFSPATIENELIK' | 'Mesencephalic astrocyte-derived neurotrophic factor' |
| 11868 | 'NAP1L1_No1' | 'FYEEVHDLER' | 'Nucleosome assembly protein 1-like 1' |
| 11869 | 'NAP1L1_No2' | 'QLTVQMMQNPQILAALQER' | 'Nucleosome assembly protein 1-like 1' |
| 11870 | 'NAP1L1_No3' | 'FYEEVHDLERK' | 'Nucleosome assembly protein 1-like 1' |
| 11871 | 'NAP1L1_No4' | 'NVDLLSDMVQEHDEPILK' | 'Nucleosome assembly protein 1-like 1' |
| 11872 | 'NAP1L1_No5' | 'YAVLYQPLFDKR' | 'Nucleosome assembly protein 1-like 1' |
| 11873 | 'NAP1L1_No6' | 'KYAVLYQPLFDK' | 'Nucleosome assembly protein 1-like 1' |
| 11874 | 'NAP1L1_No7' | 'YAVLYQPLFDK' | 'Nucleosome assembly protein 1-like 1' |
| 11875 | 'NAP1L1_No8' | 'LDGLVETPTGYIESLPR' | 'Nucleosome assembly protein 1-like 1' |
| 11876 | 'NAP1L1_No9' | 'NVDLLSDMVQEHDEPILK' | 'Nucleosome assembly protein 1-like 1' |
| 11877 | 'CDH6_No1' | 'IVVEDVDEPPVFSK' | 'Cadherin-6' |
| 11878 | 'CDH6_No2' | 'FLYLGPFK' | 'Cadherin-6' |
| 11879 | 'CDH6_No3' | 'TPESSPPGTPIGR' | 'Cadherin-6' |
| 11880 | 'CDH13_No1' | 'SIVVSPILIPENQR' | 'Cadherin-13' |
| 11881 | 'CDH13_No2' | 'TLEGPVPLEVIVIDQNDNRPIFR' | 'Cadherin-13' |

| | A | B | C |
|---|---|---|---|
| 1 | **Peptide Name** | **Peptide Sequence** | **Peptide Description** |
| 11882 | 'CDH13_No3' | 'MTAFDADDPATDNALLR' | 'Cadherin-13' |
| 11883 | 'CDH13_No4' | 'DIQGSLQDIFK' | 'Cadherin-13' |
| 11884 | 'CDH13_No5' | 'MTAFDADDPATDNALLR' | 'Cadherin-13' |
| 11885 | 'CCL18_No1' | 'FIVDYSETSPQCPKPGVILLTK' | 'C-C motif chemokine 18' |
| 11886 | 'CCL18_No2' | 'PGVILLTK' | 'C-C motif chemokine 18' |
| 11887 | 'CCL18_No3' | 'FIVDYSETSPQCPK' | 'C-C motif chemokine 18' |
| 11888 | 'CCL18_No4' | 'ELCCLVYTSWQIPQK' | 'C-C motif chemokine 18' |
| 11889 | 'CCL18_No5' | 'FIVDYSETSPQCPKPGVILLTKR' | 'C-C motif chemokine 18' |
| 11890 | 'NPEPPS_No1' | 'VLGATLLPDLIQK' | 'Puromycin-sensitive aminopeptidase' |
| 11891 | 'NPEPPS_No2' | 'VLTFALSEEVRPQDTVSVIGGVAGGSK' | 'Puromycin-sensitive aminopeptidase' |
| 11892 | 'INHBE_No1' | 'ITHPPPQAALTR' | 'Inhibin beta E chain' |
| 11893 | 'INHBE_No2' | 'RLLDTAGHQQPFLELK' | 'Inhibin beta E chain' |
| 11894 | 'INHBE_No3' | 'LWLHVLPTLPGTLCLR' | 'Inhibin beta E chain' |
| 11895 | 'INHBE_No4' | 'RPLSLLYLDHNGNVVK' | 'Inhibin beta E chain' |
| 11896 | 'INHBE_No5' | 'QQILDGLHLTSRPR' | 'Inhibin beta E chain' |
| 11897 | 'INHBE_No6' | 'DHYVDFQELGWR' | 'Inhibin beta E chain' |
| 11898 | 'ANTXR2_No1' | 'LDALWALLR' | 'Anthrax toxin receptor 2' |
| 11899 | 'ANTXR2_No2' | 'VSPVGETYIHEGLK' | 'Anthrax toxin receptor 2' |
| 11900 | 'ANTXR2_No3' | 'WPTVDASYYGGR' | 'Anthrax toxin receptor 2' |
| 11901 | 'ANTXR2_No4' | 'TSSIIIALTDGKLDGLVPSYAEK' | 'Anthrax toxin receptor 2' |
| 11902 | 'ANTXR2_No5' | 'IADSKEQVFPVK' | 'Anthrax toxin receptor 2' |
| 11903 | 'ANTXR2_No6' | 'SLGASVYCVGVLDFEQAQLER' | 'Anthrax toxin receptor 2' |
| 11904 | 'ANTXR2_No7' | 'FVSPEMR' | 'Anthrax toxin receptor 2' |
| 11905 | 'ANTXR2_No8' | 'WGDKGSTEEGARLEK' | 'Anthrax toxin receptor 2' |
| 11906 | 'MTPN_No1' | 'CDKEFMWALK' | 'Myotrophin' |
| 11907 | 'MTPN_No2' | 'NGDLDEVKDYVAK' | 'Myotrophin' |
| 11908 | 'MTPN_No3' | 'GADINAPDKHHITPLLSAVYEGHVSCVK' | 'Myotrophin' |
| 11909 | 'MTPN_No4' | 'CDKEFMWALK' | 'Myotrophin' |
| 11910 | 'MTPN_No5' | 'GPDGLTAFEATDNQAIK' | 'Myotrophin' |
| 11911 | 'HIST1H2BD_No1' | 'KESYSVYVYK' | 'Histone H2B type 1-D' |
| 11912 | 'HIST1H2BD_No2' | 'LLLPGELAK' | 'Histone H2B type 1-D' |
| 11913 | 'HIST1H2BD_No3' | 'AMGIMNSFVNDIFER' | 'Histone H2B type 1-D' |
| 11914 | 'HIST1H2BD_No4' | 'AMGIMNSFVNDIFER' | 'Histone H2B type 1-D' |
| 11915 | 'DEFA1_No1' | 'IPACIAGER' | 'Neutrophil defensin 1' |
| 11916 | 'DEFA1_No2' | 'YGTCIYQGR' | 'Neutrophil defensin 1' |
| 11917 | 'DEFA1_No3' | 'RYGTCIYQGR' | 'Neutrophil defensin 1' |
| 11918 | 'ARPC4_No1' | 'ATLQAALCLENFSSQVVER' | 'Actin-related protein 2/3 complex subunit 4' |
| 11919 | 'ARPC4_No2' | 'TATLRPYLSAVR' | 'Actin-related protein 2/3 complex subunit 4' |
| 11920 | 'ARPC4_No3' | 'IVAEEFLK' | 'Actin-related protein 2/3 complex subunit 4' |
| 11921 | 'ARPC4_No4' | 'VLIEGSINSVR' | 'Actin-related protein 2/3 complex subunit 4' |
| 11922 | 'ARPC4_No5' | 'ELLLQPVTISR' | 'Actin-related protein 2/3 complex subunit 4' |
| 11923 | 'TPI1_No1' | 'ELASQPDVDGFLVGGASLKPEFVDIINAK' | 'Triosephosphate isomerase' |
| 11924 | 'TPI1_No2' | 'DCGATWVVLGHSER' | 'Triosephosphate isomerase' |
| 11925 | 'TPI1_No3' | 'VAHALAEGLGVIACIGEK' | 'Triosephosphate isomerase' |

| | A | B | C |
|---|---|---|---|
| 1 | **Peptide Name** | **Peptide Sequence** | **Peptide Description** |
| 11926 | 'TPI1_No4' | 'IAVAAQNCYK' | 'Triosephosphate isomerase' |
| 11927 | 'TPI1_No5' | 'RHVFGESDELIGQK' | 'Triosephosphate isomerase' |
| 11928 | 'TPI1_No6' | 'QSLGELIGTLNAAK' | 'Triosephosphate isomerase' |
| 11929 | 'TPI1_No7' | 'SNVSDAVAQSTR' | 'Triosephosphate isomerase' |
| 11930 | 'TPI1_No8' | 'VVLAYEPVWAIGTGK' | 'Triosephosphate isomerase' |
| 11931 | 'TPI1_No9' | 'VPADTEVVCAPPTAYIDFAR' | 'Triosephosphate isomerase' |
| 11932 | 'TPI1_No10' | 'ELASQPDVDGFLVGGASLKPEFVDIINAKQ' | 'Triosephosphate isomerase' |
| 11933 | 'TPI1_No11' | 'HVFGESDELIGQK' | 'Triosephosphate isomerase' |
| 11934 | 'TPI1_No12' | 'QSLGELIGTLNAAK' | 'Triosephosphate isomerase' |
| 11935 | 'TPI1_No13' | 'TATPQQAQEVHEK' | 'Triosephosphate isomerase' |
| 11936 | 'TPI1_No14' | 'KQSLGELIGTLNAAK' | 'Triosephosphate isomerase' |
| 11937 | 'MYL6_No1' | 'ALGQNPTNAEVLK' | 'Myosin light polypeptide 6' |
| 11938 | 'MYL6_No2' | 'NKDQGTYEDYVEGLR' | 'Myosin light polypeptide 6' |
| 11939 | 'MYL6_No3' | 'ILYSQCGDVMR' | 'Myosin light polypeptide 6' |
| 11940 | 'MYL6_No4' | 'EAFQLFDR' | 'Myosin light polypeptide 6' |
| 11941 | 'MYL6_No5' | 'ILYSQCGDVMR' | 'Myosin light polypeptide 6' |
| 11942 | 'MYL6_No6' | 'HVLVTLGEK' | 'Myosin light polypeptide 6' |
| 11943 | 'MYL6_No7' | 'VLDFEHFLPMLQTVAK' | 'Myosin light polypeptide 6' |
| 11944 | 'MYL6_No8' | 'DQGTYEDYVEGLR' | 'Myosin light polypeptide 6' |
| 11945 | 'MYL6_No9' | 'VLDFEHFLPMLQTVAK' | 'Myosin light polypeptide 6' |
| 11946 | 'MYL6_No10' | 'TGDGKILYSQCGDVMR' | 'Myosin light polypeptide 6' |
| 11947 | 'MYL6_No11' | 'VLGNPKSDEMNVK' | 'Myosin light polypeptide 6' |
| 11948 | 'ACTB_No1' | 'IWHHTFYNELR' | 'Actin, cytoplasmic 1' |
| 11949 | 'ACTB_No2' | 'QEYDESGPSIVHR' | 'Actin, cytoplasmic 1' |
| 11950 | 'ACTB_No3' | 'DDDIAALVVDNGSGMCK' | 'Actin, cytoplasmic 1' |
| 11951 | 'ACTB_No4' | 'DDDIAALVVDNGSGMCK' | 'Actin, cytoplasmic 1' |
| 11952 | 'ACTB_No5' | 'AVFPSIVGRPR' | 'Actin, cytoplasmic 1' |
| 11953 | 'ACTB_No6' | 'CPEALFQPSFLGMESCGIHETTFNSIMK' | 'Actin, cytoplasmic 1' |
| 11954 | 'ACTB_No7' | 'VAPEEHPVLLTEAPLNPK' | 'Actin, cytoplasmic 1' |
| 11955 | 'ACTB_No8' | 'LCYVALDFEQEMATAASSSSLEK' | 'Actin, cytoplasmic 1' |
| 11956 | 'ACTB_No9' | 'DLYANTVLSGGTTMYPGIADR' | 'Actin, cytoplasmic 1' |
| 11957 | 'ACTB_No10' | 'TTGIVMDSGDGVTHTVPIYEGYALPHAILR' | 'Actin, cytoplasmic 1' |
| 11958 | 'ACTB_No11' | 'DLTDYLMK' | 'Actin, cytoplasmic 1' |
| 11959 | 'ACTB_No12' | 'LDLAGRDLTDYLMK' | 'Actin, cytoplasmic 1' |
| 11960 | 'ACTB_No13' | 'EKLCYVALDFEQEMATAASSSSLEK' | 'Actin, cytoplasmic 1' |
| 11961 | 'ACTB_No14' | 'LDLAGRDLTDYLMK' | 'Actin, cytoplasmic 1' |
| 11962 | 'ACTB_No15' | 'AGFAGDDAPR' | 'Actin, cytoplasmic 1' |
| 11963 | 'ACTB_No16' | 'DSYVGDEAQSK' | 'Actin, cytoplasmic 1' |
| 11964 | 'ACTB_No17' | 'QEYDESGPSIVHR' | 'Actin, cytoplasmic 1' |
| 11965 | 'ACTB_No18' | 'FRCPEALFQPSFLGMESCGIHETTFNSIMK' | 'Actin, cytoplasmic 1' |
| 11966 | 'ACTB_No19' | 'DSYVGDEAQSKR' | 'Actin, cytoplasmic 1' |
| 11967 | 'ACTB_No20' | 'SYELPDGQVITIGNER' | 'Actin, cytoplasmic 1' |
| 11968 | 'ACTB_No21' | 'TTGIVMDSGDGVTHTVPIYEGYALPHAILR' | 'Actin, cytoplasmic 1' |
| 11969 | 'ACTB_No22' | 'DLYANTVLSGGTTMYPGIADR' | 'Actin, cytoplasmic 1' |

| | A | B | C |
|---|---|---|---|
| 1 | **Peptide Name** | **Peptide Sequence** | **Peptide Description** |
| 11970 | 'ACTB_No23' | 'HQGVMVGMGQKDSYVGDEAQSKR' | 'Actin, cytoplasmic 1' |
| 11971 | 'ACTB_No24' | 'CPEALFQPSFLGMESCGIHETTFNSIMK' | 'Actin, cytoplasmic 1' |
| 11972 | 'ACTB_No25' | 'MQKEITALAPSTMK' | 'Actin, cytoplasmic 1' |
| 11973 | 'ACTB_No26' | 'KDLYANTVLSGGTTMYPGIADR' | 'Actin, cytoplasmic 1' |
| 11974 | 'ACTB_No27' | 'GYSFTTTAEREIVR' | 'Actin, cytoplasmic 1' |
| 11975 | 'ACTB_No28' | 'HQGVMVGMGQK' | 'Actin, cytoplasmic 1' |
| 11976 | 'ACTB_No29' | 'QEYDESGPSIVHRK' | 'Actin, cytoplasmic 1' |
| 11977 | 'ACTB_No30' | 'QEYDESGPSIVHRK' | 'Actin, cytoplasmic 1' |
| 11978 | 'ACTB_No31' | 'EITALAPSTMK' | 'Actin, cytoplasmic 1' |
| 11979 | 'ACTB_No32' | 'HQGVMVGMGQKDSYVGDEAQSKR' | 'Actin, cytoplasmic 1' |
| 11980 | 'ACTB_No33' | 'KDLYANTVLSGGTTMYPGIADR' | 'Actin, cytoplasmic 1' |
| 11981 | 'ACTB_No34' | 'MQKEITALAPSTMK' | 'Actin, cytoplasmic 1' |
| 11982 | 'ACTB_No35' | 'HQGVMVGMGQKDSYVGDEAQSKR' | 'Actin, cytoplasmic 1' |
| 11983 | 'ACTB_No36' | 'LCYVALDFEQEMATAASSSSLEK' | 'Actin, cytoplasmic 1' |
| 11984 | 'ACTB_No37' | 'HQGVMVGMGQK' | 'Actin, cytoplasmic 1' |
| 11985 | 'ACTB_No38' | 'DLTDYLMK' | 'Actin, cytoplasmic 1' |
| 11986 | 'ACTB_No39' | 'HQGVMVGMGQKDSYVGDEAQSKR' | 'Actin, cytoplasmic 1' |
| 11987 | 'ACTB_No40' | 'HQGVMVGMGQKDSYVGDEAQSK' | 'Actin, cytoplasmic 1' |
| 11988 | 'ACTB_No41' | 'HQGVMVGMGQK' | 'Actin, cytoplasmic 1' |
| 11989 | 'ACTB_No42' | 'HQGVMVGMGQKDSYVGDEAQSK' | 'Actin, cytoplasmic 1' |
| 11990 | 'ACTB_No43' | 'CDVDIRK' | 'Actin, cytoplasmic 1' |
| 11991 | 'ACTB_No44' | 'EKLCYVALDFEQEMATAASSSSLEK' | 'Actin, cytoplasmic 1' |
| 11992 | 'ACTB_No45' | 'HQGVMVGMGQKDSYVGDEAQSK' | 'Actin, cytoplasmic 1' |
| 11993 | 'ACTB_No46' | 'AVFPSIVGR' | 'Actin, cytoplasmic 1' |
| 11994 | 'ACTB_No47' | 'HQGVMVGMGQK' | 'Actin, cytoplasmic 1' |
| 11995 | 'ACTB_No48' | 'CPEALFQPSFLGMESCGIHETTFNSIMK' | 'Actin, cytoplasmic 1' |
| 11996 | 'ACTB_No49' | 'GYSFTTTAEREIVRDIK' | 'Actin, cytoplasmic 1' |
| 11997 | 'ACTB_No50' | 'CPEALFQPSFLGMESCGIHETTFNSIMK' | 'Actin, cytoplasmic 1' |
| 11998 | 'ACTB_No51' | 'CDVDIRKDLYANTVLSGGTTMYPGIADR' | 'Actin, cytoplasmic 1' |
| 11999 | 'ACTB_No52' | 'CDVDIRKDLYANTVLSGGTTMYPGIADR' | 'Actin, cytoplasmic 1' |
| 12000 | 'ACTB_No53' | 'MQKEITALAPSTMK' | 'Actin, cytoplasmic 1' |
| 12001 | 'EIF4A1_No1' | 'VLITTDLLAR' | 'Eukaryotic initiation factor 4A-I' |
| 12002 | 'EIF4A1_No2' | 'GIYAYGFEKPSAIQQR' | 'Eukaryotic initiation factor 4A-I' |
| 12003 | 'EIF4A1_No3' | 'MFVLDEADEMLSR' | 'Eukaryotic initiation factor 4A-I' |
| 12004 | 'EIF4A1_No4' | 'GFKDQIYDIFQK' | 'Eukaryotic initiation factor 4A-I' |
| 12005 | 'EIF4A1_No5' | 'GVAINMVTEEDKR' | 'Eukaryotic initiation factor 4A-I' |
| 12006 | 'PRPS1_No1' | 'VYAILTHGIFSGPAISR' | 'Ribose-phosphate pyrophosphokinase 1' |
| 12007 | 'PRPS1_No2' | 'VTAVIPCFPYAR' | 'Ribose-phosphate pyrophosphokinase 1' |
| 12008 | 'PRPS1_No3' | 'INNACFEAVVVTNTIPQEDK' | 'Ribose-phosphate pyrophosphokinase 1' |
| 12009 | 'PRPS1_No4' | 'VAILVDDMADTCGTICHAADK' | 'Ribose-phosphate pyrophosphokinase 1' |
| 12010 | 'CDC42_No1' | 'YVECSALTQK' | 'Cell division control protein 42 homolog' |
| 12011 | 'CDC42_No2' | 'TPFLLVGTQIDLR' | 'Cell division control protein 42 homolog' |
| 12012 | 'CDC42_No3' | 'WVPEITHHCPK' | 'Cell division control protein 42 homolog' |
| 12013 | 'CDC42_No4' | 'TPFLLVGTQIDLRDDPSTIEK' | 'Cell division control protein 42 homolog' |

| | A | B | C |
|---|---|---|---|
| 1 | **Peptide Name** | **Peptide Sequence** | **Peptide Description** |
| 12014 | 'DSTN_No1' | 'HFVGMLPEKDCR' | 'Destrin' |
| 12015 | 'DSTN_No2' | 'AVIFCLSADKK' | 'Destrin' |
| 12016 | 'DSTN_No3' | 'FQGIKHECQANGPEDLNR' | 'Destrin' |
| 12017 | 'RAB8A_No1' | 'ANINVENAFFTLAR' | 'Ras-related protein Rab-8A' |
| 12018 | 'RAB8A_No2' | 'NIEEHASADVEK' | 'Ras-related protein Rab-8A' |
| 12019 | 'RAB8A_No3' | 'TYDYLFK' | 'Ras-related protein Rab-8A' |
| 12020 | 'RAB10_No1' | 'AFLTLAEDILR' | 'Ras-related protein Rab-10' |
| 12021 | 'RAB10_No2' | 'KTPVKEPNSENVDISSGGGVTGWK' | 'Ras-related protein Rab-10' |
| 12022 | 'RAB14_No1' | 'NLTNPNTVIILIGNK' | 'Ras-related protein Rab-14' |
| 12023 | 'RAB14_No2' | 'SCLLHQFTEK' | 'Ras-related protein Rab-14' |
| 12024 | 'RAB14_No3' | 'STYNHLSSWLTDAR' | 'Ras-related protein Rab-14' |
| 12025 | 'RAB14_No4' | 'QFAEENGLLFLEASAK' | 'Ras-related protein Rab-14' |
| 12026 | 'RAB14_No5' | 'ATAPYNYSYIFK' | 'Ras-related protein Rab-14' |
| 12027 | 'ACTR3_No1' | 'TLTGTVIDSGDGVTHVIPVAEGYVIGSCIK' | 'Actin-related protein 3' |
| 12028 | 'ACTR3_No2' | 'LPACVVDCGTGYTK' | 'Actin-related protein 3' |
| 12029 | 'ACTR3_No3' | 'LGYAGNTEPQFIIPSCIAIK' | 'Actin-related protein 3' |
| 12030 | 'ACTR3_No4' | 'AEPEDHYFLLTEPPLNTPENR' | 'Actin-related protein 3' |
| 12031 | 'ACTR3_No5' | 'LKLSEELSGGR' | 'Actin-related protein 3' |
| 12032 | 'ACTR3_No6' | 'YAVWFGGSMLASTPEFYQVCHTK' | 'Actin-related protein 3' |
| 12033 | 'ACTR3_No7' | 'GVDDLDFFIGDEAIEKPTYATK' | 'Actin-related protein 3' |
| 12034 | 'ACTR3_No8' | 'AGRLPACVVDCGTGYTK' | 'Actin-related protein 3' |
| 12035 | 'ACTR3_No9' | 'FMEQVIFK' | 'Actin-related protein 3' |
| 12036 | 'ACTR3_No10' | 'HGIVEDWDLMER' | 'Actin-related protein 3' |
| 12037 | 'ACTR3_No11' | 'DREVGIPPEQSLETAK' | 'Actin-related protein 3' |
| 12038 | 'ACTR3_No12' | 'NIVLSGGSTMFR' | 'Actin-related protein 3' |
| 12039 | 'ACTR3_No13' | 'EFSIDVGYER' | 'Actin-related protein 3' |
| 12040 | 'ACTR3_No14' | 'FMEQVIFK' | 'Actin-related protein 3' |
| 12041 | 'ACTR3_No15' | 'QYTGINAISK' | 'Actin-related protein 3' |
| 12042 | 'ACTR2_No1' | 'LCYVGYNIEQEQK' | 'Actin-related protein 2' |
| 12043 | 'ACTR2_No2' | 'HLWDYTFGPEK' | 'Actin-related protein 2' |
| 12044 | 'ACTR2_No3' | 'HIVLSGGSTMYPGLPSR' | 'Actin-related protein 2' |
| 12045 | 'ACTR2_No4' | 'GYAFNHSADFETVR' | 'Actin-related protein 2' |
| 12046 | 'ACTR2_No5' | 'HMVFLGGAVLADIMK' | 'Actin-related protein 2' |
| 12047 | 'ACTR2_No6' | 'KVVVCDNGTGFVK' | 'Actin-related protein 2' |
| 12048 | 'ACTR2_No7' | 'LALETTVLVESYTLPDGR' | 'Actin-related protein 2' |
| 12049 | 'ACTR2_No8' | 'HIVLSGGSTMYPGLPSR' | 'Actin-related protein 2' |
| 12050 | 'ACTR2_No9' | 'VGNIEIKDLMVGDEASELR' | 'Actin-related protein 2' |
| 12051 | 'ACTR2_No10' | 'VGNIEIKDLMVGDEASELR' | 'Actin-related protein 2' |
| 12052 | 'ACTR1A_No1' | 'MESYDVIANQPVVIDNGSGVIK' | 'Alpha-centractin' |
| 12053 | 'ACTR1A_No2' | 'YCFPNYVGRPK' | 'Alpha-centractin' |
| 12054 | 'ARF3_No1' | 'LGEIVTTIPTIGFNVETVEYK' | 'ADP-ribosylation factor 3' |
| 12055 | 'ARF3_No2' | 'HYFQNTQGLIFVVDSNDRER' | 'ADP-ribosylation factor 3' |
| 12056 | 'ARF3_No3' | 'QDLPNAMNAAEITDK' | 'ADP-ribosylation factor 3' |
| 12057 | 'ARF3_No4' | 'MLAEDELR' | 'ADP-ribosylation factor 3' |

| | A | B | C |
|---|---|---|---|
| 1 | **Peptide Name** | **Peptide Sequence** | **Peptide Description** |
| 12058 | 'ARF3_No5' | 'DAVLLVFANK' | 'ADP-ribosylation factor 3' |
| 12059 | 'ARF3_No6' | 'QDLPNAMNAAEITDK' | 'ADP-ribosylation factor 3' |
| 12060 | 'ARF3_No7' | 'NISFTVWDVGGQDK' | 'ADP-ribosylation factor 3' |
| 12061 | 'ARF3_No8' | 'MLAEDELR' | 'ADP-ribosylation factor 3' |
| 12062 | 'RAP1B_No1' | 'QWNNCAFLESSAK' | 'Ras-related protein Rap-1b' |
| 12063 | 'RAP1B_No2' | 'VKDTDDVPMILVGNK' | 'Ras-related protein Rap-1b' |
| 12064 | 'RAP1B_No3' | 'QWNNCAFLESSAK' | 'Ras-related protein Rap-1b' |
| 12065 | 'RAP1B_No4' | 'VKDTDDVPMILVGNK' | 'Ras-related protein Rap-1b' |
| 12066 | 'RAP1B_No5' | 'VKDTDDVPMILVGNKCDLEDER' | 'Ras-related protein Rap-1b' |
| 12067 | 'RHOA_No1' | 'HFCPNVPIILVGNKK' | 'Transforming protein RhoA' |
| 12068 | 'RHOA_No2' | 'HFCPNVPIILVGNK' | 'Transforming protein RhoA' |
| 12069 | 'RHOA_No3' | 'TCLLIVFSK' | 'Transforming protein RhoA' |
| 12070 | 'RHOA_No4' | 'IGAFGYMECSAK' | 'Transforming protein RhoA' |
| 12071 | 'RHOA_No5' | 'LVIVGDGACGK' | 'Transforming protein RhoA' |
| 12072 | 'RHOA_No6' | 'IGAFGYMECSAK' | 'Transforming protein RhoA' |
| 12073 | 'HSPE1_No1' | 'FLPLFDR' | '10 kDa heat shock protein, mitochondrial' |
| 12074 | 'HSPE1_No2' | 'VVLDDKDYFLFR' | '10 kDa heat shock protein, mitochondrial' |
| 12075 | 'LYZ_No1' | 'GISLANWMCLAK' | 'Lysozyme C' |
| 12076 | 'LYZ_No2' | 'GISLANWMCLAK' | 'Lysozyme C' |
| 12077 | 'LYZ_No3' | 'YWCNDGKTPGAVNACHLSCSALLQDNIADAVACAK' | 'Lysozyme C' |
| 12078 | 'LYZ_No4' | 'LGMDGYR' | 'Lysozyme C' |
| 12079 | 'LYZ_No5' | 'TPGAVNACHLSCSALLQDNIADAVACAK' | 'Lysozyme C' |
| 12080 | 'LYZ_No6' | 'LGMDGYR' | 'Lysozyme C' |
| 12081 | 'LYZ_No7' | 'YWCNDGKTPGAVNACHLSCSALLQDNIADAVACAKR' | 'Lysozyme C' |
| 12082 | 'LYZ_No8' | 'ATNYNAGDRSTDYGIFQINSR' | 'Lysozyme C' |
| 12083 | 'LYZ_No9' | 'TPGAVNACHLSCSALLQDNIADAVACAKR' | 'Lysozyme C' |
| 12084 | 'B2M_No1' | 'SNFLNCYVSGFHPSDIEVDLLK' | 'Beta-2-microglobulin' |
| 12085 | 'B2M_No2' | 'IEKVEHSDLSFSK' | 'Beta-2-microglobulin' |
| 12086 | 'B2M_No3' | 'VEHSDLSFSK' | 'Beta-2-microglobulin' |
| 12087 | 'B2M_No4' | 'SNFLNCYVSGFHPSDIEVDLLKNGER' | 'Beta-2-microglobulin' |
| 12088 | 'B2M_No5' | 'IVKWDRDM' | 'Beta-2-microglobulin' |
| 12089 | 'NPC2' | 'EVNVSPCPTQPCQLSK' | 'NPC intracellular cholesterol transporter 2' |
| 12090 | 'YWHAG_No1' | 'NVTELNEPLSNEER' | '14-3-3 protein gamma' |
| 12091 | 'YWHAG_No2' | 'VDREQLVQK' | '14-3-3 protein gamma' |
| 12092 | 'YWHAG_No3' | 'YLAEVATGEKR' | '14-3-3 protein gamma' |
| 12093 | 'YWHAE_No1' | 'MDDREDLVYQAK' | '14-3-3 protein epsilon' |
| 12094 | 'YWHAE_No2' | 'LICCDILDVLDKHLIPAANTGESK' | '14-3-3 protein epsilon' |
| 12095 | 'YWHAE_No3' | 'YLAEFATGNDRK' | '14-3-3 protein epsilon' |
| 12096 | 'YWHAE_No4' | 'MDDREDLVYQAK' | '14-3-3 protein epsilon' |
| 12097 | 'YWHAE_No5' | 'AASDIAMTELPPTHPIR' | '14-3-3 protein epsilon' |
| 12098 | 'YWHAE_No6' | 'LICCDILDVLDK' | '14-3-3 protein epsilon' |
| 12099 | 'YWHAE_No7' | 'LAEQAERYDEMVESMKK' | '14-3-3 protein epsilon' |
| 12100 | 'YWHAE_No8' | 'IISSIEQKEENKGGEDK' | '14-3-3 protein epsilon' |
| 12101 | 'YWHAE_No9' | 'VAGMDVELTVEER' | '14-3-3 protein epsilon' |

| | A | B | C |
|---|---|---|---|
| 1 | **Peptide Name** | **Peptide Sequence** | **Peptide Description** |
| 12102 | 'YWHAE_No10' | 'AASDIAMTELPPTHPIR' | '14-3-3 protein epsilon' |
| 12103 | 'YWHAE_No11' | 'LAEQAERYDEMVESMKK' | '14-3-3 protein epsilon' |
| 12104 | 'YWHAE_No12' | 'LAEQAERYDEMVESMK' | '14-3-3 protein epsilon' |
| 12105 | 'TMSB4X_No1' | 'NPLPSKETIEQEK' | 'Thymosin beta-4' |
| 12106 | 'TMSB4X_No2' | 'SDKPDMAEIEK' | 'Thymosin beta-4' |
| 12107 | 'TMSB4X_No3' | 'SDKPDMAEIEKFDK' | 'Thymosin beta-4' |
| 12108 | 'TMSB4X_No4' | 'TETQEKNPLPSKETIEQEK' | 'Thymosin beta-4' |
| 12109 | 'TMSB4X_No5' | 'SDKPDMAEIEKFDK' | 'Thymosin beta-4' |
| 12110 | 'TMSB4X_No6' | 'SDKPDMAEIEK' | 'Thymosin beta-4' |
| 12111 | 'TMSB4X_No7' | 'NPLPSKETIEQEKQAGES' | 'Thymosin beta-4' |
| 12112 | 'PSMC6' | 'ALQSVGQIVGEVLK' | '26S proteasome regulatory subunit 10B' |
| 12113 | 'H4C1_No1' | 'TVTAMDVVYALK' | 'Histone H4' |
| 12114 | 'H4C1_No2' | 'TVTAMDVVYALK' | 'Histone H4' |
| 12115 | 'H4C1_No3' | 'DNIQGITKPAIR' | 'Histone H4' |
| 12116 | 'H4C1_No4' | 'TVTAMDVVYALKR' | 'Histone H4' |
| 12117 | 'H4C1_No5' | 'VFLENVIRDAVTYTEHAK' | 'Histone H4' |
| 12118 | 'H4C1_No6' | 'TVTAMDVVYALKR' | 'Histone H4' |
| 12119 | 'H4C1_No7' | 'RISGLIYEETR' | 'Histone H4' |
| 12120 | 'H4C1_No8' | 'VFLENVIR' | 'Histone H4' |
| 12121 | 'RAN_No1' | 'SNYNFEKPFLWLAR' | 'GTP-binding nuclear protein Ran' |
| 12122 | 'RAN_No2' | 'FNVWDTAGQEK' | 'GTP-binding nuclear protein Ran' |
| 12123 | 'RAP1A_No1' | 'LVVLGSGGVGK' | 'Ras-related protein Rap-1A' |
| 12124 | 'RAP1A_No2' | 'SKINVNEIFYDLVR' | 'Ras-related protein Rap-1A' |
| 12125 | 'RAP1A_No3' | 'YDPTIEDSYRK' | 'Ras-related protein Rap-1A' |
| 12126 | 'RAP1A_No4' | 'YDPTIEDSYR' | 'Ras-related protein Rap-1A' |
| 12127 | 'RAP1A_No5' | 'SALTVQFVQGIFVEK' | 'Ras-related protein Rap-1A' |
| 12128 | 'RAP1A_No6' | 'INVNEIFYDLVR' | 'Ras-related protein Rap-1A' |
| 12129 | 'RAP1A_No7' | 'QWCNCAFLESSAK' | 'Ras-related protein Rap-1A' |
| 12130 | 'GNB1_No1' | 'SELDQLRQEAEQLKNQIR' | 'Guanine nucleotide-binding protein G(I)/G(S)/G(T) subunit beta-1' |
| 12131 | 'GNB1_No2' | 'SELDQLRQEAEQLK' | 'Guanine nucleotide-binding protein G(I)/G(S)/G(T) subunit beta-1' |
| 12132 | 'GNB1_No3' | 'KACADATLSQITNNIDPVGR' | 'Guanine nucleotide-binding protein G(I)/G(S)/G(T) subunit beta-1' |
| 12133 | 'GNB1_No4' | 'ACADATLSQITNNIDPVGR' | 'Guanine nucleotide-binding protein G(I)/G(S)/G(T) subunit beta-1' |
| 12134 | 'GNB1_No5' | 'LFVSGACDASAK' | 'Guanine nucleotide-binding protein G(I)/G(S)/G(T) subunit beta-1' |
| 12135 | 'GNB2_No1' | 'ELPGHTGYLSCCR' | 'Guanine nucleotide-binding protein G(I)/G(S)/G(T) subunit beta-2' |
| 12136 | 'GNB2_No2' | 'ACGDSTLTQITAGLDPVGR' | 'Guanine nucleotide-binding protein G(I)/G(S)/G(T) subunit beta-2' |
| 12137 | 'PPIA_No1' | 'HTGPGILSMANAGPNTNGSQFFICTAK' | 'Peptidyl-prolyl cis-trans isomerase A' |
| 12138 | 'PPIA_No2' | 'VSFELFADKVPK' | 'Peptidyl-prolyl cis-trans isomerase A' |
| 12139 | 'PPIA_No3' | 'KITIADCGQLE' | 'Peptidyl-prolyl cis-trans isomerase A' |
| 12140 | 'PPIA_No4' | 'FEDENFILK' | 'Peptidyl-prolyl cis-trans isomerase A' |
| 12141 | 'PPIA_No5' | 'VKEGMNIVEAMER' | 'Peptidyl-prolyl cis-trans isomerase A' |
| 12142 | 'PPIA_No6' | 'VSFELFADK' | 'Peptidyl-prolyl cis-trans isomerase A' |
| 12143 | 'PPIA_No7' | 'HTGPGILSMANAGPNTNGSQFFICTAK' | 'Peptidyl-prolyl cis-trans isomerase A' |
| 12144 | 'PPIA_No8' | 'IIPGFMCQGGDFTR' | 'Peptidyl-prolyl cis-trans isomerase A' |
| 12145 | 'PPIA_No9' | 'EGMNIVEAMER' | 'Peptidyl-prolyl cis-trans isomerase A' |

| | A | B | C |
|---|---|---|---|
| 1 | **Peptide Name** | **Peptide Sequence** | **Peptide Description** |
| 12146 | 'PPIA_No10' | 'VNPTVFFDIAVDGEPLGR' | 'Peptidyl-prolyl cis-trans isomerase A' |
| 12147 | 'PPIA_No11' | 'SIYGEKFEDENFILK' | 'Peptidyl-prolyl cis-trans isomerase A' |
| 12148 | 'PPIA_No12' | 'VKEGMNIVEAMER' | 'Peptidyl-prolyl cis-trans isomerase A' |
| 12149 | 'PPIA_No13' | 'TEWLDGKHVVFGK' | 'Peptidyl-prolyl cis-trans isomerase A' |
| 12150 | 'PPIA_No14' | 'VKEGMNIVEAMER' | 'Peptidyl-prolyl cis-trans isomerase A' |
| 12151 | 'FKBP1A_No1' | 'RGQTCVVHYTGMLEDGKK' | 'Peptidyl-prolyl cis-trans isomerase FKBP1A' |
| 12152 | 'FKBP1A_No2' | 'RGQTCVVHYTGMLEDGKK' | 'Peptidyl-prolyl cis-trans isomerase FKBP1A' |
| 12153 | 'FKBP1A_No3' | 'RGQTCVVHYTGMLEDGK' | 'Peptidyl-prolyl cis-trans isomerase FKBP1A' |
| 12154 | 'YWHAZ_No1' | 'SVTEQGAELSNEER' | '14-3-3 protein zeta/delta' |
| 12155 | 'YWHAZ_No2' | 'YLAEVAAGDDKK' | '14-3-3 protein zeta/delta' |
| 12156 | 'YWHAZ_No3' | 'MDKNELVQK' | '14-3-3 protein zeta/delta' |
| 12157 | 'YWHAZ_No4' | 'FLIPNASQAESK' | '14-3-3 protein zeta/delta' |
| 12158 | 'YWHAZ_No5' | 'GIVDQSQQAYQEAFEISK' | '14-3-3 protein zeta/delta' |
| 12159 | 'YWHAZ_No6' | 'VVSSIEQKTEGAEK' | '14-3-3 protein zeta/delta' |
| 12160 | 'YWHAZ_No7' | 'MDKNELVQK' | '14-3-3 protein zeta/delta' |
| 12161 | 'YWHAZ_No8' | 'GIVDQSQQAYQEAFEISKK' | '14-3-3 protein zeta/delta' |
| 12162 | 'YWHAZ_No9' | 'SVTEQGAELSNEERNLLSVAYKNVVGAR' | '14-3-3 protein zeta/delta' |
| 12163 | 'YWHAZ_No10' | 'SVTEQGAELSNEERNLLSVAYK' | '14-3-3 protein zeta/delta' |
| 12164 | 'YWHAZ_No11' | 'AKLAEQAERYDDMAACMK' | '14-3-3 protein zeta/delta' |
| 12165 | 'YWHAZ_No12' | 'LAEQAERYDDMAACMK' | '14-3-3 protein zeta/delta' |
| 12166 | 'YWHAZ_No13' | 'KGIVDQSQQAYQEAFEISK' | '14-3-3 protein zeta/delta' |
| 12167 | 'YWHAZ_No14' | 'VVSSIEQKTEGAEKK' | '14-3-3 protein zeta/delta' |
| 12168 | 'DYNLL1_No1' | 'YNPTWHCIVGR' | 'Dynein light chain 1, cytoplasmic' |
| 12169 | 'DYNLL1_No2' | 'NFGSYVTHETK' | 'Dynein light chain 1, cytoplasmic' |
| 12170 | 'EIF5A_No1' | 'VHLVGIDIFTGK' | 'Eukaryotic translation initiation factor 5A-1' |
| 12171 | 'EIF5A_No2' | 'KYEDICPSTHNMDVPNIKR' | 'Eukaryotic translation initiation factor 5A-1' |
| 12172 | 'EIF5A_No3' | 'KYEDICPSTHNMDVPNIK' | 'Eukaryotic translation initiation factor 5A-1' |
| 12173 | 'EIF5A_No4' | 'KYEDICPSTHNMDVPNIK' | 'Eukaryotic translation initiation factor 5A-1' |
| 12174 | 'EIF5A_No5' | 'EDLRLPEGDLGKEIEQK' | 'Eukaryotic translation initiation factor 5A-1' |
| 12175 | 'EIF5A_No6' | 'EDLRLPEGDLGK' | 'Eukaryotic translation initiation factor 5A-1' |
| 12176 | 'ACTG1_No1' | 'EEEIAALVIDNGSGMCK' | 'Actin, cytoplasmic 2' |
| 12177 | 'ACTG1_No2' | 'EEEIAALVIDNGSGMCK' | 'Actin, cytoplasmic 2' |
| 12178 | 'TPM4_No1' | 'AGLNSLEAVKR' | 'Tropomyosin alpha-4 chain' |
| 12179 | 'TPM4_No2' | 'KIQALQQQADEAEDR' | 'Tropomyosin alpha-4 chain' |
| 12180 | 'TPM4_No3' | 'IQLVEEELDRAQER' | 'Tropomyosin alpha-4 chain' |
| 12181 | 'TPM4_No4' | 'IQALQQQADEAEDR' | 'Tropomyosin alpha-4 chain' |
| 12182 | 'TPM4_No5' | 'HIAEEADRKYEEVAR' | 'Tropomyosin alpha-4 chain' |
| 12183 | 'TPM4_No6' | 'KLVILEGELER' | 'Tropomyosin alpha-4 chain' |
| 12184 | 'TPM4_No7' | 'RIQLVEEELDRAQER' | 'Tropomyosin alpha-4 chain' |
| 12185 | 'TPM4_No8' | 'LEEAEKAADESER' | 'Tropomyosin alpha-4 chain' |
| 12186 | 'TPM4_No9' | 'EKAEGDVAALNRR' | 'Tropomyosin alpha-4 chain' |
| 12187 | 'TPM4_No10' | 'KLVILEGELERAEER' | 'Tropomyosin alpha-4 chain' |
| 12188 | 'TPM4_No11' | 'IQALQQQADEAEDRAQGLQR' | 'Tropomyosin alpha-4 chain' |
| 12189 | 'TPM4_No12' | 'LATALQKLEEAEKAADESER' | 'Tropomyosin alpha-4 chain' |

| | A | B | C |
|---|---|---|---|
| 1 | **Peptide Name** | **Peptide Sequence** | **Peptide Description** |
| 12190 | 'TPM4_No13' | 'IQLVEEELDR' | 'Tropomyosin alpha-4 chain' |
| 12191 | 'TPM4_No14' | 'AEVSELKCGDLEEELKNVTNNLK' | 'Tropomyosin alpha-4 chain' |
| 12192 | 'TPM4_No15' | 'EENVGLHQTLDQTLNELNCI' | 'Tropomyosin alpha-4 chain' |
| 12193 | 'TPM4_No16' | 'LAQAKEENVGLHQTLDQTLNELNCI' | 'Tropomyosin alpha-4 chain' |
| 12194 | 'TPM4_No17' | 'KIQALQQQADEAEDRAQGLQR' | 'Tropomyosin alpha-4 chain' |
| 12195 | 'TPM4_No18' | 'LATALQKLEEAEK' | 'Tropomyosin alpha-4 chain' |
| 12196 | 'TPM4_No19' | 'TIDDLEEKLAQAKEENVGLHQTLDQTLNELNCI' | 'Tropomyosin alpha-4 chain' |
| 12197 | 'TPM4_No20' | 'LVILEGELERAEER' | 'Tropomyosin alpha-4 chain' |
| 12198 | 'TPM4_No21' | 'TIDDLEEKLAQAK' | 'Tropomyosin alpha-4 chain' |
| 12199 | 'TPM4_No22' | 'YSEKEDKYEEEIK' | 'Tropomyosin alpha-4 chain' |
| 12200 | 'TPM4_No23' | 'AGLNSLEAVKRK' | 'Tropomyosin alpha-4 chain' |
| 12201 | 'TPM4_No24' | 'CGDLEEELKNVTNNLK' | 'Tropomyosin alpha-4 chain' |
| 12202 | 'TPM4_No25' | 'RIQLVEEELDR' | 'Tropomyosin alpha-4 chain' |
| 12203 | 'TPM4_No26' | 'LVILEGELER' | 'Tropomyosin alpha-4 chain' |
| 12204 | 'TPM4_No27' | 'AGLNSLEAVK' | 'Tropomyosin alpha-4 chain' |
| 12205 | 'ACTC1_No1' | 'YPIEHGIITNWDDMEK' | 'Actin, alpha cardiac muscle 1' |
| 12206 | 'ACTC1_No2' | 'YPIEHGIITNWDDMEK' | 'Actin, alpha cardiac muscle 1' |
| 12207 | 'ACTC1_No3' | 'YPIEHGIITNWDDMEKIWHHTFYNELR' | 'Actin, alpha cardiac muscle 1' |
| 12208 | 'ACTC1_No4' | 'YPIEHGIITNWDDMEKIWHHTFYNELR' | 'Actin, alpha cardiac muscle 1' |
| 12209 | 'ACTC1_No5' | 'LCYVALDFENEMATAASSSSLEK' | 'Actin, alpha cardiac muscle 1' |
| 12210 | 'UBE2L3_No1' | 'GQVCLPVISAENWKPATK' | 'Ubiquitin-conjugating enzyme E2 L3' |
| 12211 | 'UBE2L3_No2' | 'LMKELEEIRK' | 'Ubiquitin-conjugating enzyme E2 L3' |
| 12212 | 'UBE2L3_No3' | 'IEINFPAEYPFKPPK' | 'Ubiquitin-conjugating enzyme E2 L3' |
| 12213 | 'EEF1A1_No1' | 'YYVTIIDAPGHR' | 'Elongation factor 1-alpha 1' |
| 12214 | 'EEF1A1_No2' | 'IGGIGTVPVGR' | 'Elongation factor 1-alpha 1' |
| 12215 | 'EEF1A1_No3' | 'VETGVLKPGMVVTFAPVNVTTEVK' | 'Elongation factor 1-alpha 1' |
| 12216 | 'TUBA4A_No1' | 'AVFVDLEPTVIDEIR' | 'Tubulin alpha-4A chain' |
| 12217 | 'TUBA4A_No2' | 'TIGGGDDSFTTFFCETGAGK' | 'Tubulin alpha-4A chain' |
| 12218 | 'TUBA4A_No3' | 'SIQFVDWCPTGFK' | 'Tubulin alpha-4A chain' |
| 12219 | 'TUBA4A_No4' | 'AVFVDLEPTVIDEIRNGPYR' | 'Tubulin alpha-4A chain' |
| 12220 | 'TUBA4A_No5' | 'LISQIVSSITASLR' | 'Tubulin alpha-4A chain' |
| 12221 | 'TUBA4A_No6' | 'RSIQFVDWCPTGFK' | 'Tubulin alpha-4A chain' |
| 12222 | 'TUBB4B_No1' | 'LAVNMVPFPR' | 'Tubulin beta-4B chain' |
| 12223 | 'TUBB4B_No2' | 'ISEQFTAMFR' | 'Tubulin beta-4B chain' |
| 12224 | 'TUBB4B_No3' | 'LHFFMPGFAPLTSR' | 'Tubulin beta-4B chain' |
| 12225 | 'TUBB4B_No4' | 'GHYTEGAELVDSVLDVVR' | 'Tubulin beta-4B chain' |
| 12226 | 'TUBB4B_No5' | 'LTTPTYGDLNHLVSATMSGVTTCLR' | 'Tubulin beta-4B chain' |
| 12227 | 'TUBB4B_No6' | 'NSSYFVEWIPNNVK' | 'Tubulin beta-4B chain' |
| 12228 | 'TUBB4B_No7' | 'SGPFGQIFRPDNFVFGQSGAGNNWAK' | 'Tubulin beta-4B chain' |
| 12229 | 'TUBB4B_No8' | 'FPGQLNADLR' | 'Tubulin beta-4B chain' |
| 12230 | 'TUBB4B_No9' | 'LHFFMPGFAPLTSR' | 'Tubulin beta-4B chain' |
| 12231 | 'TUBB4B_No10' | 'KLAVNMVPFPR' | 'Tubulin beta-4B chain' |
| 12232 | 'TUBB4B_No11' | 'FPGQLNADLRK' | 'Tubulin beta-4B chain' |
| 12233 | 'TUBB4B_No12' | 'ALTVPELTQQMFDAK' | 'Tubulin beta-4B chain' |

| | A | B | C |
|---|---|---|---|
| 1 | **Peptide Name** | **Peptide Sequence** | **Peptide Description** |
| 12234 | 'TUBB4B_No13' | 'YLTVAAVFR' | 'Tubulin beta-4B chain' |
| 12235 | 'TUBB4B_No14' | 'EVDEQMLNVQNK' | 'Tubulin beta-4B chain' |
| 12236 | 'TUBB4B_No15' | 'INVYYNEATGGK' | 'Tubulin beta-4B chain' |
| 12237 | 'TUBB4B_No16' | 'GHYTEGAELVDSVLDVVRK' | 'Tubulin beta-4B chain' |
| 12238 | 'TUBB4B_No17' | 'IMNTFSVVPSPK' | 'Tubulin beta-4B chain' |
| 12239 | 'TUBB4B_No18' | 'MSATFIGNSTAIQELFK' | 'Tubulin beta-4B chain' |
| 12240 | 'TUBB4B_No19' | 'MSATFIGNSTAIQELFKR' | 'Tubulin beta-4B chain' |
| 12241 | 'TUBB4B_No20' | 'MSMKEVDEQMLNVQNK' | 'Tubulin beta-4B chain' |
| 12242 | 'TUBB4B_No21' | 'EVDEQMLNVQNK' | 'Tubulin beta-4B chain' |
| 12243 | 'TUBB4B_No22' | 'LTTPTYGDLNHLVSATMSGVTTCLR' | 'Tubulin beta-4B chain' |
| 12244 | 'TUBB4B_No23' | 'IREEYPDR' | 'Tubulin beta-4B chain' |
| 12245 | 'TUBB4B_No24' | 'LAVNMVPFPR' | 'Tubulin beta-4B chain' |
| 12246 | 'TUBB4B_No25' | 'AVLVDLEPGTMDSVR' | 'Tubulin beta-4B chain' |
| 12247 | 'PAFAH1B2' | 'IIVLGLLPR' | 'Platelet-activating factor acetylhydrolase IB subunit beta' |
| 12248 | 'H3C1' | 'EIAQDFKTDLR' | 'Histone H3.1' |
| 12249 | 'HBB_No1' | 'LLGNVLVCVLAHHFGK' | 'Hemoglobin subunit beta' |
| 12250 | 'HBB_No2' | 'VNVDEVGGEALGR' | 'Hemoglobin subunit beta' |
| 12251 | 'HBB_No3' | 'VLGAFSDGLAHLDNLK' | 'Hemoglobin subunit beta' |
| 12252 | 'HBB_No4' | 'LHVDPENFR' | 'Hemoglobin subunit beta' |
| 12253 | 'HBB_No5' | 'GTFATLSELHCDK' | 'Hemoglobin subunit beta' |
| 12254 | 'HBB_No6' | 'EFTPPVQAAYQK' | 'Hemoglobin subunit beta' |
| 12255 | 'HBB_No7' | 'GTFATLSELHCDKLHVDPENFR' | 'Hemoglobin subunit beta' |
| 12256 | 'HBB_No8' | 'VLGAFSDGLAHLDNLKGTFATLSELHCDKLHVDPENFR' | 'Hemoglobin subunit beta' |
| 12257 | 'HBB_No9' | 'VHLTPEEK' | 'Hemoglobin subunit beta' |
| 12258 | 'HBB_No10' | 'KVLGAFSDGLAHLDNLK' | 'Hemoglobin subunit beta' |
| 12259 | 'HBB_No11' | 'LLGNVLVCVLAHHFGKEFTPPVQAAYQK' | 'Hemoglobin subunit beta' |
| 12260 | 'HBB_No12' | 'VVAGVANALAHKYH' | 'Hemoglobin subunit beta' |
| 12261 | 'HBB_No13' | 'SAVTALWGK' | 'Hemoglobin subunit beta' |
| 12262 | 'HBB_No14' | 'FFESFGDLSTPDAVMGNPK' | 'Hemoglobin subunit beta' |
| 12263 | 'HBB_No15' | 'VVAGVANALAHK' | 'Hemoglobin subunit beta' |
| 12264 | 'HBB_No16' | 'VHLTPEEKSAVTALWGK' | 'Hemoglobin subunit beta' |
| 12265 | 'HBB_No17' | 'FFESFGDLSTPDAVMGNPK' | 'Hemoglobin subunit beta' |
| 12266 | 'HBB_No18' | 'VLGAFSDGLAHLDNLKGTFATLSELHCDK' | 'Hemoglobin subunit beta' |
| 12267 | 'HBB_No19' | 'SAVTALWGKVNVDEVGGEALGR' | 'Hemoglobin subunit beta' |
| 12268 | 'HBB_No20' | 'VHLTPEEKSAVTALWGKVNVDEVGGEALGR' | 'Hemoglobin subunit beta' |
| 12269 | 'HBB_No21' | 'KVLGAFSDGLAHLDNLKGTFATLSELHCDK' | 'Hemoglobin subunit beta' |
| 12270 | 'HBB_No22' | 'FFESFGDLSTPDAVMGNPKVK' | 'Hemoglobin subunit beta' |
| 12271 | 'HBB_No23' | 'EFTPPVQAAYQKVVAGVANALAHKYH' | 'Hemoglobin subunit beta' |
| 12272 | 'HBB_No24' | 'FFESFGDLSTPDAVMGNPKVK' | 'Hemoglobin subunit beta' |
| 12273 | 'HBG1_No1' | 'LLVVYPWTQR' | 'Hemoglobin subunit gamma-1' |
| 12274 | 'HBG1_No2' | 'GTFAQLSELHCDK' | 'Hemoglobin subunit gamma-1' |
| 12275 | 'HBG1_No3' | 'FFDSFGNLSSASAIMGNPK' | 'Hemoglobin subunit gamma-1' |
| 12276 | 'HBG1_No4' | 'FFDSFGNLSSASAIMGNPK' | 'Hemoglobin subunit gamma-1' |
| 12277 | 'HBG1_No5' | 'VLTSLGDAIK' | 'Hemoglobin subunit gamma-1' |

| | A | B | C |
|---|---|---|---|
| 1 | **Peptide Name** | **Peptide Sequence** | **Peptide Description** |
| 12278 | 'HBG1_No6' | 'GHFTEEDKATITSLWGK' | 'Hemoglobin subunit gamma-1' |
| 12279 | 'HBG1_No7' | 'KVLTSLGDAIK' | 'Hemoglobin subunit gamma-1' |
| 12280 | 'HBG1_No8' | 'LHVDPENFK' | 'Hemoglobin subunit gamma-1' |
| 12281 | 'HBG1_No9' | 'HLDDLKGTFAQLSELHCDKLHVDPENFK' | 'Hemoglobin subunit gamma-1' |
| 12282 | 'HBG1_No10' | 'MVTAVASALSSR' | 'Hemoglobin subunit gamma-1' |
| 12283 | 'HBG1_No11' | 'HLDDLKGTFAQLSELHCDK' | 'Hemoglobin subunit gamma-1' |
| 12284 | 'HBG1_No12' | 'GTFAQLSELHCDKLHVDPENFK' | 'Hemoglobin subunit gamma-1' |
| 12285 | 'HBG1_No13' | 'VNVEDAGGETLGR' | 'Hemoglobin subunit gamma-1' |
| 12286 | 'HBG1_No14' | 'MVTAVASALSSR' | 'Hemoglobin subunit gamma-1' |
| 12287 | 'HBA1_No1' | 'VGAHAGEYGAEALER' | 'Hemoglobin subunit alpha' |
| 12288 | 'HBA1_No2' | 'VADALTNAVAHVDDMPNALSALSDLHAHK' | 'Hemoglobin subunit alpha' |
| 12289 | 'HBA1_No3' | 'FLASVSTVLTSK' | 'Hemoglobin subunit alpha' |
| 12290 | 'HBA1_No4' | 'LLSHCLLVTLAAHLPAEFTPAVHASLDK' | 'Hemoglobin subunit alpha' |
| 12291 | 'HBA1_No5' | 'TYFPHFDLSHGSAQVK' | 'Hemoglobin subunit alpha' |
| 12292 | 'HBA1_No6' | 'KVADALTNAVAHVDDMPNALSALSDLHAHK' | 'Hemoglobin subunit alpha' |
| 12293 | 'HBA1_No7' | 'VLSPADKTNVK' | 'Hemoglobin subunit alpha' |
| 12294 | 'HBA1_No8' | 'MFLSFPTTK' | 'Hemoglobin subunit alpha' |
| 12295 | 'HBA1_No9' | 'LRVDPVNFK' | 'Hemoglobin subunit alpha' |
| 12296 | 'HBA1_No10' | 'MFLSFPTTK' | 'Hemoglobin subunit alpha' |
| 12297 | 'HBA1_No11' | 'VADALTNAVAHVDDMPNALSALSDLHAHK' | 'Hemoglobin subunit alpha' |
| 12298 | 'HBA1_No12' | 'KVADALTNAVAHVDDMPNALSALSDLHAHK' | 'Hemoglobin subunit alpha' |
| 12299 | 'HBA1_No13' | 'VLSPADKTNVK' | 'Hemoglobin subunit alpha' |
| 12300 | 'HBA1_No14' | 'KVADALTNAVAHVDDMPNALSALSDLHAHKLR' | 'Hemoglobin subunit alpha' |
| 12301 | 'HBA1_No15' | 'VADALTNAVAHVDDMPNALSALSDLHAHKLR' | 'Hemoglobin subunit alpha' |
| 12302 | 'HBA1_No16' | 'VADALTNAVAHVDDMPNALSALSDLHAHKLR' | 'Hemoglobin subunit alpha' |
| 12303 | 'HBA1_No17' | 'MFLSFPTTKTYFPHFDLSHGSAQVK' | 'Hemoglobin subunit alpha' |
| 12304 | 'HBA1_No18' | 'KVADALTNAVAHVDDMPNALSALSDLHAHKLR' | 'Hemoglobin subunit alpha' |
| 12305 | 'HBA1_No19' | 'MFLSFPTTKTYFPHFDLSHGSAQVK' | 'Hemoglobin subunit alpha' |
| 12306 | 'HBA1_No20' | 'VGAHAGEYGAEALERMFLSFPTTK' | 'Hemoglobin subunit alpha' |
| 12307 | 'HBA1_No21' | 'TYFPHFDLSHGSAQVKGHGKK' | 'Hemoglobin subunit alpha' |
| 12308 | 'HBA1_No22' | 'TYFPHFDLSHGSAQVKGHGK' | 'Hemoglobin subunit alpha' |
| 12309 | 'HBA1_No23' | 'AAWGKVGAHAGEYGAEALER' | 'Hemoglobin subunit alpha' |
| 12310 | 'HBA1_No24' | 'VDPVNFK' | 'Hemoglobin subunit alpha' |
| 12311 | 'KRT85' | 'VLQAHISDTSVIVK' | 'Keratin, type II cuticular Hb5' |
| 12312 | 'GSTO1_No1' | 'MILELFSK' | 'Glutathione S-transferase omega-1' |
| 12313 | 'GSTO1_No2' | 'HEVININLK' | 'Glutathione S-transferase omega-1' |
| 12314 | 'GSTO1_No3' | 'GSAPPGPVPEGSIR' | 'Glutathione S-transferase omega-1' |
| 12315 | 'GSTO1_No4' | 'NKPEWFFK' | 'Glutathione S-transferase omega-1' |
| 12316 | 'GSTO1_No5' | 'VPSLVGSFIR' | 'Glutathione S-transferase omega-1' |
| 12317 | 'GSTO1_No6' | 'MILELFSK' | 'Glutathione S-transferase omega-1' |
| 12318 | 'RELN_No1' | 'YIALEIPLK' | 'Reelin' |
| 12319 | 'RELN_No2' | 'ITIPLPNAALTR' | 'Reelin' |
| 12320 | 'RELN_No3' | 'FLQFTLR' | 'Reelin' |
| 12321 | 'RELN_No4' | 'DCLPTNVECSR' | 'Reelin' |

| | A | B | C |
|---|---|---|---|
| 1 | **Peptide Name** | **Peptide Sequence** | **Peptide Description** |
| 12322 | 'RELN_No5' | 'ITLPLPPYTR' | 'Reelin' |
| 12323 | 'RELN_No6' | 'KCGILSSGNNLFFNEDGLR' | 'Reelin' |
| 12324 | 'RELN_No7' | 'ITGAQVGTGCGTLNDGK' | 'Reelin' |
| 12325 | 'RELN_No8' | 'FVYLELPAAAK' | 'Reelin' |
| 12326 | 'RELN_No9' | 'DLTLKPGYVLQFK' | 'Reelin' |
| 12327 | 'RELN_No10' | 'TVMLDTFSSAPVPQHER' | 'Reelin' |
| 12328 | 'RELN_No11' | 'LSSYHNFYSIR' | 'Reelin' |
| 12329 | 'RELN_No12' | 'LLEHYSYLSYHEPR' | 'Reelin' |
| 12330 | 'RELN_No13' | 'QLITSFLDSSQSR' | 'Reelin' |
| 12331 | 'RELN_No14' | 'ITVYLPLSTISPR' | 'Reelin' |
| 12332 | 'RELN_No15' | 'TVMLDTFSSAPVPQHER' | 'Reelin' |
| 12333 | 'GPLD1_No1' | 'TLLLVGSPTWK' | 'Phosphatidylinositol-glycan-specific phospholipase D' |
| 12334 | 'GPLD1_No2' | 'FGSSLITVR' | 'Phosphatidylinositol-glycan-specific phospholipase D' |
| 12335 | 'GPLD1_No3' | 'GIVAAFYSGPSLSDKEK' | 'Phosphatidylinositol-glycan-specific phospholipase D' |
| 12336 | 'GPLD1_No4' | 'QVLLVGAPTYDDVSK' | 'Phosphatidylinositol-glycan-specific phospholipase D' |
| 12337 | 'GPLD1_No5' | 'TMFIGGSQLSQK' | 'Phosphatidylinositol-glycan-specific phospholipase D' |
| 12338 | 'GPLD1_No6' | 'LSGALHVYSLGSD' | 'Phosphatidylinositol-glycan-specific phospholipase D' |
| 12339 | 'GPLD1_No7' | 'IADVTSGLIGGEDGR' | 'Phosphatidylinositol-glycan-specific phospholipase D' |
| 12340 | 'GPLD1_No8' | 'AQYVLISPEASSR' | 'Phosphatidylinositol-glycan-specific phospholipase D' |
| 12341 | 'GPLD1_No9' | 'VAFLTVTLHQGGATR' | 'Phosphatidylinositol-glycan-specific phospholipase D' |
| 12342 | 'GPLD1_No10' | 'VYLIYGNDLGLPPVDLDLDKEAHR' | 'Phosphatidylinositol-glycan-specific phospholipase D' |
| 12343 | 'GPLD1_No11' | 'VYGYFPPNGQSWFTISGDK' | 'Phosphatidylinositol-glycan-specific phospholipase D' |
| 12344 | 'GPLD1_No12' | 'LGHLLHIRDEK' | 'Phosphatidylinositol-glycan-specific phospholipase D' |
| 12345 | 'GPLD1_No13' | 'SWITPCPEEK' | 'Phosphatidylinositol-glycan-specific phospholipase D' |
| 12346 | 'GPLD1_No14' | 'HVSSPLASYFLSFPYAR' | 'Phosphatidylinositol-glycan-specific phospholipase D' |
| 12347 | 'GPLD1_No15' | 'LGWAMTSADLNQDGHGDLVVGAPGYSR' | 'Phosphatidylinositol-glycan-specific phospholipase D' |
| 12348 | 'GPLD1_No16' | 'TMFIGGSQLSQK' | 'Phosphatidylinositol-glycan-specific phospholipase D' |
| 12349 | 'GPLD1_No17' | 'QVLLVGAPTYDDVSK' | 'Phosphatidylinositol-glycan-specific phospholipase D' |
| 12350 | 'GPLD1_No18' | 'QVLLVGAPTYDDVSKVAFLTVTLHQGGATR' | 'Phosphatidylinositol-glycan-specific phospholipase D' |
| 12351 | 'GPLD1_No19' | 'ALEFLQLHNGR' | 'Phosphatidylinositol-glycan-specific phospholipase D' |
| 12352 | 'GPLD1_No20' | 'GVFFSVNSWTPDSMSFIYK' | 'Phosphatidylinositol-glycan-specific phospholipase D' |
| 12353 | 'GPLD1_No21' | 'WYVPVKDLLGIYEK' | 'Phosphatidylinositol-glycan-specific phospholipase D' |
| 12354 | 'GPLD1_No22' | 'ILEGFQPSGR' | 'Phosphatidylinositol-glycan-specific phospholipase D' |
| 12355 | 'GPLD1_No23' | 'RWYVPVKDLLGIYEK' | 'Phosphatidylinositol-glycan-specific phospholipase D' |
| 12356 | 'GPLD1_No24' | 'LGHLLHIRDEKK' | 'Phosphatidylinositol-glycan-specific phospholipase D' |
| 12357 | 'GPLD1_No25' | 'ENYPLPWEK' | 'Phosphatidylinositol-glycan-specific phospholipase D' |
| 12358 | 'GPLD1_No26' | 'NQVVIAAGR' | 'Phosphatidylinositol-glycan-specific phospholipase D' |
| 12359 | 'GPLD1_No27' | 'ELLLEHQDAYQAGIVFPDCFYPSICK' | 'Phosphatidylinositol-glycan-specific phospholipase D' |
| 12360 | 'GPLD1_No28' | 'ENYPLPWEKDTEK' | 'Phosphatidylinositol-glycan-specific phospholipase D' |
| 12361 | 'GPLD1_No29' | 'GAVYVYFGSK' | 'Phosphatidylinositol-glycan-specific phospholipase D' |
| 12362 | 'GPLD1_No30' | 'VYVYNGKETTLGDMTGK' | 'Phosphatidylinositol-glycan-specific phospholipase D' |
| 12363 | 'GPLD1_No31' | 'DLLGIYEK' | 'Phosphatidylinositol-glycan-specific phospholipase D' |
| 12364 | 'GPLD1_No32' | 'VYVYNGKETTLGDMTGK' | 'Phosphatidylinositol-glycan-specific phospholipase D' |
| 12365 | 'GPLD1_No33' | 'QKGIVAAFYSGPSLSDKEK' | 'Phosphatidylinositol-glycan-specific phospholipase D' |

| | A | B | C |
|---|---|---|---|
| 1 | **Peptide Name** | **Peptide Sequence** | **Peptide Description** |
| 12366 | 'GPLD1_No34' | 'IADVTSGLIGGEDGRVYVYNGKETTLGDMTGK' | 'Phosphatidylinositol-glycan-specific phospholipase D' |
| 12367 | 'GPLD1_No35' | 'GIVAAFYSGPSLSDK' | 'Phosphatidylinositol-glycan-specific phospholipase D' |
| 12368 | 'GPLD1_No36' | 'IADVTSGLIGGEDGRVYVYNGK' | 'Phosphatidylinositol-glycan-specific phospholipase D' |
| 12369 | 'GPLD1_No37' | 'VYLIYGNDLGLPPVDLDLDK' | 'Phosphatidylinositol-glycan-specific phospholipase D' |
| 12370 | 'LCN2_No1' | 'TFVPGCQPGEFTLGNIK' | 'Neutrophil gelatinase-associated lipocalin' |
| 12371 | 'LCN2_No2' | 'SLGLPENHIVFPVPIDQCIDG' | 'Neutrophil gelatinase-associated lipocalin' |
| 12372 | 'LCN2_No3' | 'WYVVGLAGNAILR' | 'Neutrophil gelatinase-associated lipocalin' |
| 12373 | 'LCN2_No4' | 'ELTSELKENFIR' | 'Neutrophil gelatinase-associated lipocalin' |
| 12374 | 'LCN2_No5' | 'TKELTSELKENFIR' | 'Neutrophil gelatinase-associated lipocalin' |
| 12375 | 'LCN2_No6' | 'VVSTNYNQHAMVFFK' | 'Neutrophil gelatinase-associated lipocalin' |
| 12376 | 'LCN2_No7' | 'SYPGLTSYLVR' | 'Neutrophil gelatinase-associated lipocalin' |
| 12377 | 'LCN2_No8' | 'VPLQQNFQDNQFQGK' | 'Neutrophil gelatinase-associated lipocalin' |
| 12378 | 'LCN2_No9' | 'MYATIYELKEDK' | 'Neutrophil gelatinase-associated lipocalin' |
| 12379 | 'LCN2_No10' | 'VVSTNYNQHAMVFFK' | 'Neutrophil gelatinase-associated lipocalin' |
| 12380 | 'S100A12' | 'TKLEEHLEGIVNIFHQYSVR' | 'Protein S100-A12' |
| 12381 | 'BASP1_No1' | 'AEPPKAPEQEQAAPGPAAGGEAPK' | 'Brain acid soluble protein 1' |
| 12382 | 'BASP1_No2' | 'SDGAPASDSKPGSSEAAPSSK' | 'Brain acid soluble protein 1' |
| 12383 | 'BASP1_No3' | 'ETPAATEAPSSTPK' | 'Brain acid soluble protein 1' |
| 12384 | 'BASP1_No4' | 'AAEAAAAPAESAAPAAGEEPSKEEGEPK' | 'Brain acid soluble protein 1' |
| 12385 | 'BASP1_No5' | 'AEGAATEEEGTPKESEPQAAAEPAEAK' | 'Brain acid soluble protein 1' |
| 12386 | 'BASP1_No6' | 'AQGPAASAEEPKPVEAPAANSDQTVTVKE' | 'Brain acid soluble protein 1' |
| 12387 | 'IGLV3-21_No1' | 'FSGSNSGNTATLTISR' | 'Immunoglobulin lambda variable 3-21' |
| 12388 | 'IGLV3-21_No2' | 'PGQAPVLVVYDDSDRPSGIPER' | 'Immunoglobulin lambda variable 3-21' |
| 12389 | 'IGLV3-21_No3' | 'SVHWYQQKPGQAPVLVVYDDSDRPSGIPER' | 'Immunoglobulin lambda variable 3-21' |
| 12390 | 'IGLV3-21_No4' | 'ITCGGNNIGSK' | 'Immunoglobulin lambda variable 3-21' |
| 12391 | 'IGLV3-21_No5' | 'PGQAPVLVVYDDSDR' | 'Immunoglobulin lambda variable 3-21' |
| 12392 | 'IGLV3-21_No6' | 'SVHWYQQK' | 'Immunoglobulin lambda variable 3-21' |
| 12393 | 'IGLV3-21_No7' | 'SVHWYQQKPGQAPVLVVYDDSDR' | 'Immunoglobulin lambda variable 3-21' |
| 12394 | 'HAMP' | 'DTHFPICIFCCGCCHR' | 'Hepcidin' |
| 12395 | 'DCD_No1' | 'LGKDAVEDLESVGK' | 'Dermcidin' |
| 12396 | 'DCD_No2' | 'DAVEDLESVGK' | 'Dermcidin' |
| 12397 | 'DCD_No3' | 'GAVHDVKDVLDSVL' | 'Dermcidin' |
| 12398 | 'HSPG2_No1' | 'LLSGPYFWSLPSR' | 'Basement membrane-specific heparan sulfate proteoglycan core protein' |
| 12399 | 'HSPG2_No2' | 'LYIFQASPADAGQYVCR' | 'Basement membrane-specific heparan sulfate proteoglycan core protein' |
| 12400 | 'HSPG2_No3' | 'LPAVEPTDQAQYLCR' | 'Basement membrane-specific heparan sulfate proteoglycan core protein' |
| 12401 | 'HSPG2_No4' | 'HLISTHFAPGDFQGFALVNPQR' | 'Basement membrane-specific heparan sulfate proteoglycan core protein' |
| 12402 | 'HSPG2_No5' | 'SIVPQGGSHSLR' | 'Basement membrane-specific heparan sulfate proteoglycan core protein' |
| 12403 | 'HSPG2_No6' | 'TPSGLYLGTCER' | 'Basement membrane-specific heparan sulfate proteoglycan core protein' |
| 12404 | 'HSPG2_No7' | 'LVSEDPINDGEWHR' | 'Basement membrane-specific heparan sulfate proteoglycan core protein' |
| 12405 | 'HSPG2_No8' | 'CSCHGHSEACEPETGACQGCQHHTEGPR' | 'Basement membrane-specific heparan sulfate proteoglycan core protein' |
| 12406 | 'HSPG2_No9' | 'CAPGYYGNPSQGQPCQR' | 'Basement membrane-specific heparan sulfate proteoglycan core protein' |
| 12407 | 'HSPG2_No10' | 'IQVVVLSASDASPPPVK' | 'Basement membrane-specific heparan sulfate proteoglycan core protein' |
| 12408 | 'HSPG2_No11' | 'SPAYTLVWTR' | 'Basement membrane-specific heparan sulfate proteoglycan core protein' |
| 12409 | 'HSPG2_No12' | 'ATATSCRPCPCPYIDASR' | 'Basement membrane-specific heparan sulfate proteoglycan core protein' |

| | A | B | C |
|---|---|---|---|
| 1 | **Peptide Name** | **Peptide Sequence** | **Peptide Description** |
| 12410 | 'HSPG2_No13' | 'LLQVTPADSGEYVCR' | 'Basement membrane-specific heparan sulfate proteoglycan core protein' |
| 12411 | 'HSPG2_No14' | 'LPQVSPADSGEYVCR' | 'Basement membrane-specific heparan sulfate proteoglycan core protein' |
| 12412 | 'HSPG2_No15' | 'AMDFNGILTIR' | 'Basement membrane-specific heparan sulfate proteoglycan core protein' |
| 12413 | 'HSPG2_No16' | 'LHHVSPADSGEYVCR' | 'Basement membrane-specific heparan sulfate proteoglycan core protein' |
| 12414 | 'HSPG2_No17' | 'LCNECADGSFHLSTR' | 'Basement membrane-specific heparan sulfate proteoglycan core protein' |
| 12415 | 'HSPG2_No18' | 'CSATGSPTPTLEWTGGPGGQLPAK' | 'Basement membrane-specific heparan sulfate proteoglycan core protein' |
| 12416 | 'HSPG2_No19' | 'SLPEVPETIELEVR' | 'Basement membrane-specific heparan sulfate proteoglycan core protein' |
| 12417 | 'HSPG2_No20' | 'AELLVTEAPSKPITVTVEEQR' | 'Basement membrane-specific heparan sulfate proteoglycan core protein' |
| 12418 | 'HSPG2_No21' | 'GSVYIGGAPDVATLTGGR' | 'Basement membrane-specific heparan sulfate proteoglycan core protein' |
| 12419 | 'HSPG2_No22' | 'AMDFNGILTIR' | 'Basement membrane-specific heparan sulfate proteoglycan core protein' |
| 12420 | 'HSPG2_No23' | 'DFISLGLQDGHLVFR' | 'Basement membrane-specific heparan sulfate proteoglycan core protein' |
| 12421 | 'HSPG2_No24' | 'ATFSSVPLAASISAVSLEVAQPGPSNRPR' | 'Basement membrane-specific heparan sulfate proteoglycan core protein' |
| 12422 | 'HSPG2_No25' | 'SKSPAYTLVWTR' | 'Basement membrane-specific heparan sulfate proteoglycan core protein' |
| 12423 | 'HSPG2_No26' | 'CEQCQPGYYGDAQR' | 'Basement membrane-specific heparan sulfate proteoglycan core protein' |
| 12424 | 'HSPG2_No27' | 'FLGDKVTSYGGELR' | 'Basement membrane-specific heparan sulfate proteoglycan core protein' |
| 12425 | 'HSPG2_No28' | 'IEPSSSHVAEGQTLDLNCVVPGQAHAQVTWHK' | 'Basement membrane-specific heparan sulfate proteoglycan core protein' |
| 12426 | 'HSPG2_No29' | 'LDVEFKPLAPDGVLLFSGGK' | 'Basement membrane-specific heparan sulfate proteoglycan core protein' |
| 12427 | 'HSPG2_No30' | 'CQVSGSPPHYFYWSR' | 'Basement membrane-specific heparan sulfate proteoglycan core protein' |
| 12428 | 'HSPG2_No31' | 'ESDQGAYTCEAMNAR' | 'Basement membrane-specific heparan sulfate proteoglycan core protein' |
| 12429 | 'HSPG2_No32' | 'RGSIQVDGEELVSGR' | 'Basement membrane-specific heparan sulfate proteoglycan core protein' |
| 12430 | 'HSPG2_No33' | 'FSSGITGCVK' | 'Basement membrane-specific heparan sulfate proteoglycan core protein' |
| 12431 | 'HSPG2_No34' | 'SQSVRPGADVTFICTAK' | 'Basement membrane-specific heparan sulfate proteoglycan core protein' |
| 12432 | 'HSPG2_No35' | 'CPPGYIGLSCQDCAPGYTR' | 'Basement membrane-specific heparan sulfate proteoglycan core protein' |
| 12433 | 'HSPG2_No36' | 'AAGVPSATITWR' | 'Basement membrane-specific heparan sulfate proteoglycan core protein' |
| 12434 | 'HSPG2_No37' | 'QPQAIITWYK' | 'Basement membrane-specific heparan sulfate proteoglycan core protein' |
| 12435 | 'HSPG2_No38' | 'EQAWQRPDGQPATR' | 'Basement membrane-specific heparan sulfate proteoglycan core protein' |
| 12436 | 'HSPG2_No39' | 'LHQMSVADSGEYVCR' | 'Basement membrane-specific heparan sulfate proteoglycan core protein' |
| 12437 | 'HSPG2_No40' | 'GPSCQDCDTGYTR' | 'Basement membrane-specific heparan sulfate proteoglycan core protein' |
| 12438 | 'HSPG2_No41' | 'LHQMSVADSGEYVCR' | 'Basement membrane-specific heparan sulfate proteoglycan core protein' |
| 12439 | 'HSPG2_No42' | 'PSDAGTYVCLAQNALGTAQK' | 'Basement membrane-specific heparan sulfate proteoglycan core protein' |
| 12440 | 'HSPG2_No43' | 'ESDQGAYTCEAMNAR' | 'Basement membrane-specific heparan sulfate proteoglycan core protein' |
| 12441 | 'HSPG2_No44' | 'LYQASPADSGEYVCR' | 'Basement membrane-specific heparan sulfate proteoglycan core protein' |
| 12442 | 'HSPG2_No45' | 'LSGSHSQGVAYPVR' | 'Basement membrane-specific heparan sulfate proteoglycan core protein' |
| 12443 | 'HSPG2_No46' | 'IESSSSQVAEGQTLDLNCLVAGQAHAQVTWHK' | 'Basement membrane-specific heparan sulfate proteoglycan core protein' |
| 12444 | 'HSPG2_No47' | 'CPIGYSGLSCESCDAHFTR' | 'Basement membrane-specific heparan sulfate proteoglycan core protein' |
| 12445 | 'HSPG2_No48' | 'QPQAIITWYK' | 'Basement membrane-specific heparan sulfate proteoglycan core protein' |
| 12446 | 'HSPG2_No49' | 'EVSEAVVDTLESEYLK' | 'Basement membrane-specific heparan sulfate proteoglycan core protein' |
| 12447 | 'HSPG2_No50' | 'IAHVELADAGQYR' | 'Basement membrane-specific heparan sulfate proteoglycan core protein' |
| 12448 | 'HSPG2_No51' | 'RCESCAPGYEGNPIQPGGK' | 'Basement membrane-specific heparan sulfate proteoglycan core protein' |
| 12449 | 'CYCS_No1' | 'TGPNLHGLFGR' | 'Cytochrome c' |
| 12450 | 'CYCS_No2' | 'GIIWGEDTLMEYLENPKK' | 'Cytochrome c' |
| 12451 | 'CYCS_No3' | 'ADLIAYLK' | 'Cytochrome c' |
| 12452 | 'MPP1_No1' | 'VASMAQSAPSEAPSCSPFGK' | '55 kDa erythrocyte membrane protein' |
| 12453 | 'MPP1_No2' | 'IAILDIEPQTLK' | '55 kDa erythrocyte membrane protein' |

| | A | B | C |
|---|---|---|---|
| 1 | **Peptide Name** | **Peptide Sequence** | **Peptide Description** |
| 12454 | 'MPP1_No3' | 'FATGDIIQIINK' | '55 kDa erythrocyte membrane protein' |
| 12455 | 'MPP1_No4' | 'VASMAQSAPSEAPSCSPFGK' | '55 kDa erythrocyte membrane protein' |
| 12456 | 'SLC25A3_No1' | 'IQTQPGYANTLR' | 'Phosphate carrier protein, mitochondrial' |
| 12457 | 'SLC25A3_No2' | 'FGFYEVFK' | 'Phosphate carrier protein, mitochondrial' |
| 12458 | 'SLC25A3_No3' | 'GVAPLWMR' | 'Phosphate carrier protein, mitochondrial' |
| 12459 | 'SLC25A3_No4' | 'GWAPTFLGYSMQGLCK' | 'Phosphate carrier protein, mitochondrial' |
| 12460 | 'CLTC_No1' | 'LLLPWLEAR' | 'Clathrin heavy chain 1' |
| 12461 | 'CLTC_No2' | 'KVGYTPDWIFLLR' | 'Clathrin heavy chain 1' |
| 12462 | 'CLTC_No3' | 'NLQNLLILTAIK' | 'Clathrin heavy chain 1' |
| 12463 | 'CLTC_No4' | 'ALEHFTDLYDIKR' | 'Clathrin heavy chain 1' |
| 12464 | 'CLTC_No5' | 'SVNESLNNLFITEEDYQALR' | 'Clathrin heavy chain 1' |
| 12465 | 'CLTC_No6' | 'QLPLVKPYLR' | 'Clathrin heavy chain 1' |
| 12466 | 'CLTC_No7' | 'SVDPTLALSVYLR' | 'Clathrin heavy chain 1' |
| 12467 | 'CLTC_No8' | 'GQFSTDELVAEVEKR' | 'Clathrin heavy chain 1' |
| 12468 | 'CLTC_No9' | 'TSIDAYDNFDNISLAQR' | 'Clathrin heavy chain 1' |
| 12469 | 'CLTC_No10' | 'KFDVNTSAVQVLIEHIGNLDR' | 'Clathrin heavy chain 1' |
| 12470 | 'CLTC_No11' | 'WLLLTGISAQQNR' | 'Clathrin heavy chain 1' |
| 12471 | 'CLTC_No12' | 'IVLDNSVFSEHR' | 'Clathrin heavy chain 1' |
| 12472 | 'CLTC_No13' | 'TLQIFNIEMK' | 'Clathrin heavy chain 1' |
| 12473 | 'CLTC_No14' | 'ISGETIFVTAPHEATAGIIGVNR' | 'Clathrin heavy chain 1' |
| 12474 | 'CLTC_No15' | 'NNLAGAEELFAR' | 'Clathrin heavy chain 1' |
| 12475 | 'CLTC_No16' | 'VDKLDASESLR' | 'Clathrin heavy chain 1' |
| 12476 | 'CLTC_No17' | 'LEKHELIEFR' | 'Clathrin heavy chain 1' |
| 12477 | 'CLTC_No18' | 'VSQPIEGHAASFAQFK' | 'Clathrin heavy chain 1' |
| 12478 | 'CLTC_No19' | 'LASTLVHLGEYQAAVDGAR' | 'Clathrin heavy chain 1' |
| 12479 | 'CLTC_No20' | 'LLYNNVSNFGR' | 'Clathrin heavy chain 1' |
| 12480 | 'CLTC_No21' | 'KFNALFAQGNYSEAAK' | 'Clathrin heavy chain 1' |
| 12481 | 'CTBS_No1' | 'SYDWSQITTVATFGK' | 'Di-N-acetylchitobiase' |
| 12482 | 'CTBS_No2' | 'QQTEEMWEVLKPK' | 'Di-N-acetylchitobiase' |
| 12483 | 'CTBS_No3' | 'YDSELMCYAHSK' | 'Di-N-acetylchitobiase' |
| 12484 | 'CTBS_No4' | 'GDVSLKDIIDPAFR' | 'Di-N-acetylchitobiase' |
| 12485 | 'CTBS_No5' | 'QQTEEMWEVLKPK' | 'Di-N-acetylchitobiase' |
| 12486 | 'CTBS_No6' | 'QQTEEMWEVLKPK' | 'Di-N-acetylchitobiase' |
| 12487 | 'CTBS_No7' | 'ATYIQNYR' | 'Di-N-acetylchitobiase' |
| 12488 | 'CTBS_No8' | 'DPAGHFHQVWYDNPQSISLK' | 'Di-N-acetylchitobiase' |
| 12489 | 'CTBS_No9' | 'QQTEEMWEVLKPK' | 'Di-N-acetylchitobiase' |
| 12490 | 'CTBS_No10' | 'EIEGSQVTFDVAWSPK' | 'Di-N-acetylchitobiase' |
| 12491 | 'CTBS_No11' | 'VVLKGDVSLKDIIDPAFR' | 'Di-N-acetylchitobiase' |
| 12492 | 'CTBS_No12' | 'YDSELMCYAHSK' | 'Di-N-acetylchitobiase' |
| 12493 | 'CTBS_No13' | 'APYYNYKDPAGHFHQVWYDNPQSISLK' | 'Di-N-acetylchitobiase' |
| 12494 | 'FABP5_No1' | 'ELGVGIALR' | 'Fatty acid-binding protein 5' |
| 12495 | 'FABP5_No2' | 'TTQFSCTLGEKFEETTADGRK' | 'Fatty acid-binding protein 5' |
| 12496 | 'FABP5_No3' | 'LVVECVMNNVTCTR' | 'Fatty acid-binding protein 5' |
| 12497 | 'FABP5_No4' | 'LVVECVMNNVTCTR' | 'Fatty acid-binding protein 5' |

| | A | B | C |
|---|---|---|---|
| 1 | **Peptide Name** | **Peptide Sequence** | **Peptide Description** |
| 12498 | 'FABP5_No5' | 'TQTVCNFTDGALVQHQEWDGKESTITR' | 'Fatty acid-binding protein 5' |
| 12499 | 'FABP5_No6' | 'GFDEYMKELGVGIALR' | 'Fatty acid-binding protein 5' |
| 12500 | 'CAP1_No1' | 'ALLVTASQCQQPAENK' | 'Adenylyl cyclase-associated protein 1' |
| 12501 | 'CAP1_No2' | 'LSDLLAPISEQIK' | 'Adenylyl cyclase-associated protein 1' |
| 12502 | 'CAP1_No3' | 'NSLDCEIVSAK' | 'Adenylyl cyclase-associated protein 1' |
| 12503 | 'CAP1_No4' | 'VPTISINK' | 'Adenylyl cyclase-associated protein 1' |
| 12504 | 'CAP1_No5' | 'VENQENVSNLVIEDTELK' | 'Adenylyl cyclase-associated protein 1' |
| 12505 | 'CAP1_No6' | 'AGAAPYVQAFDSLLAGPVAEYLK' | 'Adenylyl cyclase-associated protein 1' |
| 12506 | 'CAP1_No7' | 'AVGRLEAVSHTSDMHR' | 'Adenylyl cyclase-associated protein 1' |
| 12507 | 'CAP1_No8' | 'EFHTTGLAWSK' | 'Adenylyl cyclase-associated protein 1' |
| 12508 | 'CAP1_No9' | 'LFNHLSAVSESIQALGWVAMAPK' | 'Adenylyl cyclase-associated protein 1' |
| 12509 | 'CAP1_No10' | 'LFNHLSAVSESIQALGWVAMAPKPGPYVK' | 'Adenylyl cyclase-associated protein 1' |
| 12510 | 'CAP1_No11' | 'ALLVTASQCQQPAENKLSDLLAPISEQIK' | 'Adenylyl cyclase-associated protein 1' |
| 12511 | 'CAP1_No12' | 'SSEMNVLIPTEGGDFNEFPVPEQFK' | 'Adenylyl cyclase-associated protein 1' |
| 12512 | 'CAP1_No13' | 'WRVENQENVSNLVIEDTELK' | 'Adenylyl cyclase-associated protein 1' |
| 12513 | 'CAP1_No14' | 'SSEMNVLIPTEGGDFNEFPVPEQFK' | 'Adenylyl cyclase-associated protein 1' |
| 12514 | 'CAP1_No15' | 'VPTISINKTDGCHAYLSK' | 'Adenylyl cyclase-associated protein 1' |
| 12515 | 'CAP1_No16' | 'VENQENVSNLVIEDTELKQVAYIYK' | 'Adenylyl cyclase-associated protein 1' |
| 12516 | 'CAP1_No17' | 'LFNHLSAVSESIQALGWVAMAPK' | 'Adenylyl cyclase-associated protein 1' |
| 12517 | 'CAP1_No18' | 'AVGRLEAVSHTSDMHR' | 'Adenylyl cyclase-associated protein 1' |
| 12518 | 'CAP1_No19' | 'ADMQNLVER' | 'Adenylyl cyclase-associated protein 1' |
| 12519 | 'PFKP_No1' | 'EWSGLLEELAR' | 'ATP-dependent 6-phosphofructokinase, platelet type' |
| 12520 | 'PFKP_No2' | 'NVIFQPVAELKK' | 'ATP-dependent 6-phosphofructokinase, platelet type' |
| 12521 | 'PFKP_No3' | 'EIGWTDVGGWTGQGGSILGTK' | 'ATP-dependent 6-phosphofructokinase, platelet type' |
| 12522 | 'PFKP_No4' | 'IIEVVDAIMTTAQSHQR' | 'ATP-dependent 6-phosphofructokinase, platelet type' |
| 12523 | 'PFKP_No5' | 'ASYDVSDSGQLEHVQPWSV' | 'ATP-dependent 6-phosphofructokinase, platelet type' |
| 12524 | 'PFKP_No6' | 'NVLGHMQQGGAPSPFDR' | 'ATP-dependent 6-phosphofructokinase, platelet type' |
| 12525 | 'PFKP_No7' | 'KEWSGLLEELAR' | 'ATP-dependent 6-phosphofructokinase, platelet type' |
| 12526 | 'DSG1_No1' | 'VGDFVATDLDTGRPSTTVR' | 'Desmoglein-1' |
| 12527 | 'DSG1_No2' | 'IHSDCAANQQVTYR' | 'Desmoglein-1' |
| 12528 | 'DSG1_No3' | 'TMNNFLDREQYGQYALAVR' | 'Desmoglein-1' |
| 12529 | 'DSG1_No4' | 'ASAISVTVLNVIEGPVFRPGSK' | 'Desmoglein-1' |
| 12530 | 'DSG1_No5' | 'ISGVGIDQPPYGIFVINQK' | 'Desmoglein-1' |
| 12531 | 'DSG1_No6' | 'YVMGNNPADLLAVDSR' | 'Desmoglein-1' |
| 12532 | 'DSG1_No7' | 'YQGTILSIDDNLQR' | 'Desmoglein-1' |
| 12533 | 'DSG1_No8' | 'YVMGNNPADLLAVDSR' | 'Desmoglein-1' |
| 12534 | 'DSG1_No9' | 'GSDRDGGADGMSAECECNIK' | 'Desmoglein-1' |
| 12535 | 'DSG1_No10' | 'SSSDHHFNQTIGSASPSTAR' | 'Desmoglein-1' |
| 12536 | 'DSG1_No11' | 'ALNSMGQDLERPLELR' | 'Desmoglein-1' |
| 12537 | 'DSG1_No12' | 'QEPSDSPMFIINR' | 'Desmoglein-1' |
| 12538 | 'DSG1_No13' | 'TYVVTGNMGSNDKVGDFVATDLDTGRPSTTVR' | 'Desmoglein-1' |
| 12539 | 'DSG1_No14' | 'ALNSMGQDLERPLELR' | 'Desmoglein-1' |
| 12540 | 'DSG1_No15' | 'QEPSDSPMFIINR' | 'Desmoglein-1' |
| 12541 | 'DSG1_No16' | 'IIRQEPSDSPMFIINR' | 'Desmoglein-1' |

| | A | B | C |
|---|---|---|---|
| 1 | **Peptide Name** | **Peptide Sequence** | **Peptide Description** |
| 12542 | 'SLC25A11' | 'AATASAGAGGIDGKPR' | 'Mitochondrial 2-oxoglutarate/malate carrier protein' |
| 12543 | 'CFHR3_No1' | 'RPYFPVAVGK' | 'Complement factor H-related protein 3' |
| 12544 | 'CFHR3_No2' | 'EGIVEYPRCE' | 'Complement factor H-related protein 3' |
| 12545 | 'CFHR3_No3' | 'CYFPYLENGYNQNYGR' | 'Complement factor H-related protein 3' |
| 12546 | 'CFHR3_No4' | 'SDIEIENGFISESSSIYILNK' | 'Complement factor H-related protein 3' |
| 12547 | 'CFHR3_No5' | 'TGDTIEFMCK' | 'Complement factor H-related protein 3' |
| 12548 | 'CFHR3_No6' | 'KCYFPYLENGYNQNYGR' | 'Complement factor H-related protein 3' |
| 12549 | 'CFHR3_No7' | 'CIHPCIITEENMNK' | 'Complement factor H-related protein 3' |
| 12550 | 'CFHR3_No8' | 'HGGLFHENMR' | 'Complement factor H-related protein 3' |
| 12551 | 'CFHR3_No9' | 'CGPPPPISNGDTTSFLLK' | 'Complement factor H-related protein 3' |
| 12552 | 'CFHR3_No10' | 'KCYFPYLENGYNQNYGRK' | 'Complement factor H-related protein 3' |
| 12553 | 'CFHR3_No11' | 'CIHPCIITEENMNKNNIK' | 'Complement factor H-related protein 3' |
| 12554 | 'CFHR3_No12' | 'CYFPYLENGYNQNYGRK' | 'Complement factor H-related protein 3' |
| 12555 | 'CFHR3_No13' | 'VYVPQSR' | 'Complement factor H-related protein 3' |
| 12556 | 'CFHR3_No14' | 'AQTTVTCTEKGWSPTPR' | 'Complement factor H-related protein 3' |
| 12557 | 'CFHR3_No15' | 'CIHPCIITEENMNK' | 'Complement factor H-related protein 3' |
| 12558 | 'CFHR3_No16' | 'PYFPVAVGK' | 'Complement factor H-related protein 3' |
| 12559 | 'CFHR3_No17' | 'HGGLFHENMR' | 'Complement factor H-related protein 3' |
| 12560 | 'CFHR3_No18' | 'CIHPCIITEENMNKNNIK' | 'Complement factor H-related protein 3' |
| 12561 | 'ACY1_No1' | 'LLPALASVPALPSDS' | 'Aminoacylase-1' |
| 12562 | 'ACY1_No2' | 'TVQPKPDYGAAVAFFEETAR' | 'Aminoacylase-1' |
| 12563 | 'CFHR1_No1' | 'INHGILYDEEK' | 'Complement factor H-related protein 1' |
| 12564 | 'CFHR1_No2' | 'TTCWDGKLEYPTCAK' | 'Complement factor H-related protein 1' |
| 12565 | 'CFHR1_No3' | 'LQNNENNISCVER' | 'Complement factor H-related protein 1' |
| 12566 | 'CFHR1_No4' | 'EIMENYNIALR' | 'Complement factor H-related protein 1' |
| 12567 | 'CFHR1_No5' | 'ITCTEEGWSPTPK' | 'Complement factor H-related protein 1' |
| 12568 | 'CFHR1_No6' | 'LCFFPFVENGHSESSGQTHLEGDTVQIICNTGYR' | 'Complement factor H-related protein 1' |
| 12569 | 'CFHR1_No7' | 'CLHPCVISR' | 'Complement factor H-related protein 1' |
| 12570 | 'CFHR1_No8' | 'TGESAEFVCK' | 'Complement factor H-related protein 1' |
| 12571 | 'CFHR1_No9' | 'NGQWSEPPKCLHPCVISR' | 'Complement factor H-related protein 1' |
| 12572 | 'CFHR1_No10' | 'YKPFSQVPTGEVFYYSCEYNFVSPSK' | 'Complement factor H-related protein 1' |
| 12573 | 'CFHR1_No11' | 'LEYPTCAK' | 'Complement factor H-related protein 1' |
| 12574 | 'CFHR1_No12' | 'STDTSCVNPPTVQNAHILSR' | 'Complement factor H-related protein 1' |
| 12575 | 'CFHR1_No13' | 'EIMENYNIALR' | 'Complement factor H-related protein 1' |
| 12576 | 'CFHR1_No14' | 'NGQWSEPPK' | 'Complement factor H-related protein 1' |
| 12577 | 'CFHR1_No15' | 'TGESAEFVCKR' | 'Complement factor H-related protein 1' |
| 12578 | 'CFHR1_No16' | 'EIMENYNIALRWTAKQK' | 'Complement factor H-related protein 1' |
| 12579 | 'CFHR1_No17' | 'QMSKYPSGER' | 'Complement factor H-related protein 1' |
| 12580 | 'CFHR1_No18' | 'TTCWDGK' | 'Complement factor H-related protein 1' |
| 12581 | 'CFHR1_No19' | 'TTCWDGKLEYPTCAKR' | 'Complement factor H-related protein 1' |
| 12582 | 'CFHR1_No20' | 'QMSKYPSGER' | 'Complement factor H-related protein 1' |
| 12583 | 'CFHR1_No21' | 'EIMENYNIALRWTAK' | 'Complement factor H-related protein 1' |
| 12584 | 'GBE1_No1' | 'IVLDSDAAEYGGHQR' | '1,4-alpha-glucan-branching enzyme' |
| 12585 | 'GBE1_No2' | 'IYESHVGISSHEGK' | '1,4-alpha-glucan-branching enzyme' |

| | A | B | C |
|---|---|---|---|
| 1 | **Peptide Name** | **Peptide Sequence** | **Peptide Description** |
| 12586 | 'KRT17_No1' | 'LLEGEDAHLTQYK' | 'Keratin, type I cytoskeletal 17' |
| 12587 | 'KRT17_No2' | 'GQVGGEINVEMDAAPGVDLSR' | 'Keratin, type I cytoskeletal 17' |
| 12588 | 'KRT17_No3' | 'DAEDWFFSK' | 'Keratin, type I cytoskeletal 17' |
| 12589 | 'KRT17_No4' | 'RLLEGEDAHLTQYK' | 'Keratin, type I cytoskeletal 17' |
| 12590 | 'KRT17_No5' | 'GQVGGEINVEMDAAPGVDLSR' | 'Keratin, type I cytoskeletal 17' |
| 12591 | 'KRT17_No6' | 'LSVEADINGLRR' | 'Keratin, type I cytoskeletal 17' |
| 12592 | 'KRT17_No7' | 'ADLEMQIENLKEELAYLKK' | 'Keratin, type I cytoskeletal 17' |
| 12593 | 'KRT17_No8' | 'DAEDWFFSKTEELNR' | 'Keratin, type I cytoskeletal 17' |
| 12594 | 'KRT17_No9' | 'ADLEMQIENLKEELAYLKK' | 'Keratin, type I cytoskeletal 17' |
| 12595 | 'NOTCH2_No1' | 'NCQTVLAPCSPNPCENAAVCK' | 'Neurogenic locus notch homolog protein 2' |
| 12596 | 'NOTCH2_No2' | 'VASFSCMCPEGK' | 'Neurogenic locus notch homolog protein 2' |
| 12597 | 'NOTCH2_No3' | 'QPPYYSCQCAPPFSGSR' | 'Neurogenic locus notch homolog protein 2' |
| 12598 | 'NOTCH2_No4' | 'GADCTEDVDECAMANSNPCEHAGK' | 'Neurogenic locus notch homolog protein 2' |
| 12599 | 'NOTCH2_No5' | 'CQNGGVCVDGVNTYNCR' | 'Neurogenic locus notch homolog protein 2' |
| 12600 | 'NOTCH2_No6' | 'DTYECTCQVGFTGK' | 'Neurogenic locus notch homolog protein 2' |
| 12601 | 'NOTCH2_No7' | 'IGGFTCLCMPGFK' | 'Neurogenic locus notch homolog protein 2' |
| 12602 | 'NOTCH2_No8' | 'VASFSCMCPEGK' | 'Neurogenic locus notch homolog protein 2' |
| 12603 | 'NOTCH2_No9' | 'GLLCEENIDDCAR' | 'Neurogenic locus notch homolog protein 2' |
| 12604 | 'NOTCH2_No10' | 'GADCTEDVDECAMANSNPCEHAGK' | 'Neurogenic locus notch homolog protein 2' |
| 12605 | 'HGFAC_No1' | 'LCNIEPDER' | 'Hepatocyte growth factor activator' |
| 12606 | 'HGFAC_No2' | 'VANYVDWINDR' | 'Hepatocyte growth factor activator' |
| 12607 | 'HGFAC_No3' | 'SQFVQPICLPEPGSTFPAGHK' | 'Hepatocyte growth factor activator' |
| 12608 | 'HGFAC_No4' | 'LEACESLTR' | 'Hepatocyte growth factor activator' |
| 12609 | 'HGFAC_No5' | 'SDACQGDSGGPLACEK' | 'Hepatocyte growth factor activator' |
| 12610 | 'HGFAC_No6' | 'EALVPLVADHK' | 'Hepatocyte growth factor activator' |
| 12611 | 'HGFAC_No7' | 'WCATTHNYDRDR' | 'Hepatocyte growth factor activator' |
| 12612 | 'HGFAC_No8' | 'YEYLEGGDR' | 'Hepatocyte growth factor activator' |
| 12613 | 'HGFAC_No9' | 'NPDNDERPWCYVVK' | 'Hepatocyte growth factor activator' |
| 12614 | 'HGFAC_No10' | 'CQIAGWGHLDENVSGYSSSLR' | 'Hepatocyte growth factor activator' |
| 12615 | 'HGFAC_No11' | 'AVPSSSSPQAQALTEDGRPCRFPFR' | 'Hepatocyte growth factor activator' |
| 12616 | 'HGFAC_No12' | 'QGHVEQCECFGGR' | 'Hepatocyte growth factor activator' |
| 12617 | 'HGFAC_No13' | 'VQLSPDLLATLPEPASPGR' | 'Hepatocyte growth factor activator' |
| 12618 | 'HGFAC_No14' | 'QGHVEQCECFGGR' | 'Hepatocyte growth factor activator' |
| 12619 | 'HGFAC_No15' | 'AVPSSSSPQAQALTEDGRPCR' | 'Hepatocyte growth factor activator' |
| 12620 | 'HGFAC_No16' | 'DSALSWEYCR' | 'Hepatocyte growth factor activator' |
| 12621 | 'HGFAC_No17' | 'TTDVTQTFGIEK' | 'Hepatocyte growth factor activator' |
| 12622 | 'HGFAC_No18' | 'MLHACTSEGSAHR' | 'Hepatocyte growth factor activator' |
| 12623 | 'HGFAC_No19' | 'TWCEGTR' | 'Hepatocyte growth factor activator' |
| 12624 | 'HGFAC_No20' | 'YIPYTLYSVFNPSDHDLVLIR' | 'Hepatocyte growth factor activator' |
| 12625 | 'HGFAC_No21' | 'DSALSWEYCRLEACESLTR' | 'Hepatocyte growth factor activator' |
| 12626 | 'HGFAC_No22' | 'CSSPEVYGADISPNMLCAGYFDCK' | 'Hepatocyte growth factor activator' |
| 12627 | 'HGFAC_No23' | 'MLHACTSEGSAHR' | 'Hepatocyte growth factor activator' |
| 12628 | 'HGFAC_No24' | 'CFDETRYEYLEGGDRWAR' | 'Hepatocyte growth factor activator' |
| 12629 | 'YWHAH_No1' | 'AVTELNEPLSNEDR' | '14-3-3 protein eta' |

| | A | B | C |
|---|---|---|---|
| 1 | **Peptide Name** | **Peptide Sequence** | **Peptide Description** |
| 12630 | 'YWHAH_No2' | 'AVTELNEPLSNEDRNLLSVAYK' | '14-3-3 protein eta' |
| 12631 | 'YWHAH_No3' | 'NSVVEASEAAYKEAFEISK' | '14-3-3 protein eta' |
| 12632 | 'YWHAH_No4' | 'GDREQLLQR' | '14-3-3 protein eta' |
| 12633 | 'YWHAH_No5' | 'NSVVEASEAAYK' | '14-3-3 protein eta' |
| 12634 | 'YWHAH_No6' | 'YLAEVASGEK' | '14-3-3 protein eta' |
| 12635 | 'CALD1_No1' | 'QQEAALELEELKK' | 'Caldesmon' |
| 12636 | 'CALD1_No2' | 'SAKPTKPAASDLPVPAEGVR' | 'Caldesmon' |
| 12637 | 'CALD1_No3' | 'ASVDTKEAEGAPQVEAGK' | 'Caldesmon' |
| 12638 | 'CALD1_No4' | 'RGETESEEFEKLK' | 'Caldesmon' |
| 12639 | 'CALD1_No5' | 'ASVDTKEAEGAPQVEAGKR' | 'Caldesmon' |
| 12640 | 'CALD1_No6' | 'GNVFSSPTAAGTPNKETAGLK' | 'Caldesmon' |
| 12641 | 'CALD1_No7' | 'QQEAALELEELKK' | 'Caldesmon' |
| 12642 | 'CALD1_No8' | 'QERYEIEETETVTK' | 'Caldesmon' |
| 12643 | 'CALD1_No9' | 'SPAPKPSDLRPGDVSSK' | 'Caldesmon' |
| 12644 | 'CALD1_No10' | 'LEQYTSAIEGTK' | 'Caldesmon' |
| 12645 | 'CALD1_No11' | 'QKEFDPTITDASLSLPSR' | 'Caldesmon' |
| 12646 | 'CALD1_No12' | 'YEIEETETVTK' | 'Caldesmon' |
| 12647 | 'CALD1_No13' | 'GNVFSSPTAAGTPNK' | 'Caldesmon' |
| 12648 | 'ITIH3_No1' | 'VTFELTYEELLKR' | 'Inter-alpha-trypsin inhibitor heavy chain H3' |
| 12649 | 'ITIH3_No2' | 'DYIFGNYIER' | 'Inter-alpha-trypsin inhibitor heavy chain H3' |
| 12650 | 'ITIH3_No3' | 'GHVSFKPSLDQQR' | 'Inter-alpha-trypsin inhibitor heavy chain H3' |
| 12651 | 'ITIH3_No4' | 'VTFELTYEELLK' | 'Inter-alpha-trypsin inhibitor heavy chain H3' |
| 12652 | 'ITIH3_No5' | 'LVDEDMNSFK' | 'Inter-alpha-trypsin inhibitor heavy chain H3' |
| 12653 | 'ITIH3_No6' | 'EHLVQATPENLQEAR' | 'Inter-alpha-trypsin inhibitor heavy chain H3' |
| 12654 | 'ITIH3_No7' | 'LVDEDMNSFK' | 'Inter-alpha-trypsin inhibitor heavy chain H3' |
| 12655 | 'ITIH3_No8' | 'YHFVTPLTSMVVTKPEDNEDER' | 'Inter-alpha-trypsin inhibitor heavy chain H3' |
| 12656 | 'ITIH3_No9' | 'GMTNINDGLLR' | 'Inter-alpha-trypsin inhibitor heavy chain H3' |
| 12657 | 'ITIH3_No10' | 'SLPEGVANGIEVYSTK' | 'Inter-alpha-trypsin inhibitor heavy chain H3' |
| 12658 | 'ITIH3_No11' | 'GMTNINDGLLR' | 'Inter-alpha-trypsin inhibitor heavy chain H3' |
| 12659 | 'ITIH3_No12' | 'LWAYLTIEQLLEK' | 'Inter-alpha-trypsin inhibitor heavy chain H3' |
| 12660 | 'ITIH3_No13' | 'FAHNVVTMR' | 'Inter-alpha-trypsin inhibitor heavy chain H3' |
| 12661 | 'ITIH3_No14' | 'KGHVSFKPSLDQQR' | 'Inter-alpha-trypsin inhibitor heavy chain H3' |
| 12662 | 'ITIH3_No15' | 'DFLGFYVVDSHR' | 'Inter-alpha-trypsin inhibitor heavy chain H3' |
| 12663 | 'ITIH3_No16' | 'SMEDKGMTNINDGLLR' | 'Inter-alpha-trypsin inhibitor heavy chain H3' |
| 12664 | 'ITIH3_No17' | 'EVSFDVELPK' | 'Inter-alpha-trypsin inhibitor heavy chain H3' |
| 12665 | 'ITIH3_No18' | 'STSIVIMLTDGDANVGESRPEK' | 'Inter-alpha-trypsin inhibitor heavy chain H3' |
| 12666 | 'ITIH3_No19' | 'FPLYNLGFGNNLNYNFLENMALENHGFAR' | 'Inter-alpha-trypsin inhibitor heavy chain H3' |
| 12667 | 'ITIH3_No20' | 'FTVSVNVAAGSK' | 'Inter-alpha-trypsin inhibitor heavy chain H3' |
| 12668 | 'ITIH3_No21' | 'YHFVTPLTSMVVTKPEDNEDER' | 'Inter-alpha-trypsin inhibitor heavy chain H3' |
| 12669 | 'ITIH3_No22' | 'STSIVIMLTDGDANVGESRPEK' | 'Inter-alpha-trypsin inhibitor heavy chain H3' |
| 12670 | 'ITIH3_No23' | 'GHGATNDLTFTEEVDMKEMEK' | 'Inter-alpha-trypsin inhibitor heavy chain H3' |
| 12671 | 'ITIH3_No24' | 'LVDEDMNSFKADVK' | 'Inter-alpha-trypsin inhibitor heavy chain H3' |
| 12672 | 'ITIH3_No25' | 'SMEDKGMTNINDGLLR' | 'Inter-alpha-trypsin inhibitor heavy chain H3' |
| 12673 | 'ITIH3_No26' | 'ALDLSLKYHFVTPLTSMVVTKPEDNEDER' | 'Inter-alpha-trypsin inhibitor heavy chain H3' |

| | A | B | C |
|---|---|---|---|
| 1 | **Peptide Name** | **Peptide Sequence** | **Peptide Description** |
| 12674 | 'ITIH3_No27' | 'GHGATNDLTFTEEVDMKEMEK' | 'Inter-alpha-trypsin inhibitor heavy chain H3' |
| 12675 | 'ITIH3_No28' | 'NAIGGKFPLYNLGFGNNLNYNFLENMALENHGFAR' | 'Inter-alpha-trypsin inhibitor heavy chain H3' |
| 12676 | 'ITIH3_No29' | 'VSDIRPGSDPTKPDATLVVK' | 'Inter-alpha-trypsin inhibitor heavy chain H3' |
| 12677 | 'ITIH3_No30' | 'LWAYLTIEQLLEKRK' | 'Inter-alpha-trypsin inhibitor heavy chain H3' |
| 12678 | 'ITIH3_No31' | 'KLEQTKEALLR' | 'Inter-alpha-trypsin inhibitor heavy chain H3' |
| 12679 | 'ITIH3_No32' | 'FAHNVVTMR' | 'Inter-alpha-trypsin inhibitor heavy chain H3' |
| 12680 | 'ITIH3_No33' | 'GHGATNDLTFTEEVDMKEMEK' | 'Inter-alpha-trypsin inhibitor heavy chain H3' |
| 12681 | 'ITIH3_No34' | 'ADTAKEVSFDVELPK' | 'Inter-alpha-trypsin inhibitor heavy chain H3' |
| 12682 | 'ITIH3_No35' | 'SLPEGVANGIEVYSTKINSK' | 'Inter-alpha-trypsin inhibitor heavy chain H3' |
| 12683 | 'ITIH3_No36' | 'GHGATNDLTFTEEVDMK' | 'Inter-alpha-trypsin inhibitor heavy chain H3' |
| 12684 | 'ITIH3_No37' | 'LIQDAVTGLTVNGQITGDKR' | 'Inter-alpha-trypsin inhibitor heavy chain H3' |
| 12685 | 'ITIH3_No38' | 'MSAQTHGLLGQFFQPFDFK' | 'Inter-alpha-trypsin inhibitor heavy chain H3' |
| 12686 | 'ITIH3_No39' | 'AVNRADTAKEVSFDVELPK' | 'Inter-alpha-trypsin inhibitor heavy chain H3' |
| 12687 | 'ITIH3_No40' | 'SMEDKGMTNINDGLLR' | 'Inter-alpha-trypsin inhibitor heavy chain H3' |
| 12688 | 'ITIH3_No41' | 'GHGATNDLTFTEEVDMK' | 'Inter-alpha-trypsin inhibitor heavy chain H3' |
| 12689 | 'ITIH3_No42' | 'NVAFVIDISGSMAGR' | 'Inter-alpha-trypsin inhibitor heavy chain H3' |
| 12690 | 'ITIH3_No43' | 'GKYEMYLK' | 'Inter-alpha-trypsin inhibitor heavy chain H3' |
| 12691 | 'ITIH3_No44' | 'GKYEMYLK' | 'Inter-alpha-trypsin inhibitor heavy chain H3' |
| 12692 | 'ITIH3_No45' | 'LVDEDMNSFKADVK' | 'Inter-alpha-trypsin inhibitor heavy chain H3' |
| 12693 | 'ITIH3_No46' | 'SMEDKGMTNINDGLLR' | 'Inter-alpha-trypsin inhibitor heavy chain H3' |
| 12694 | 'ITIH3_No47' | 'FPLYNLGFGNNLNYNFLENMALENHGFAR' | 'Inter-alpha-trypsin inhibitor heavy chain H3' |
| 12695 | 'ITIH3_No48' | 'ALQERDYIFGNYIER' | 'Inter-alpha-trypsin inhibitor heavy chain H3' |
| 12696 | 'ITIH3_No49' | 'MSAQTHGLLGQFFQPFDFK' | 'Inter-alpha-trypsin inhibitor heavy chain H3' |
| 12697 | 'ITIH3_No50' | 'STSIVIMLTDGDANVGESRPEKIQENVR' | 'Inter-alpha-trypsin inhibitor heavy chain H3' |
| 12698 | 'ITIH3_No51' | 'STSIVIMLTDGDANVGESRPEKIQENVR' | 'Inter-alpha-trypsin inhibitor heavy chain H3' |
| 12699 | 'ITIH3_No52' | 'SCPTCTDSLLNGDFTITYDVNR' | 'Inter-alpha-trypsin inhibitor heavy chain H3' |
| 12700 | 'ITIH3_No53' | 'ADVKGHGATNDLTFTEEVDMKEMEK' | 'Inter-alpha-trypsin inhibitor heavy chain H3' |
| 12701 | 'ITIH3_No54' | 'NVAFVIDISGSMAGR' | 'Inter-alpha-trypsin inhibitor heavy chain H3' |
| 12702 | 'ITIH3_No55' | 'NHQLIVTR' | 'Inter-alpha-trypsin inhibitor heavy chain H3' |
| 12703 | 'ITIH3_No56' | 'ALDLSLKYHFVTPLTSMVVTKPEDNEDER' | 'Inter-alpha-trypsin inhibitor heavy chain H3' |
| 12704 | 'ITIH3_No57' | 'SLPEGVANGIEVYSTKINSKVTSR' | 'Inter-alpha-trypsin inhibitor heavy chain H3' |
| 12705 | 'ITIH3_No58' | 'PEDNEDER' | 'Inter-alpha-trypsin inhibitor heavy chain H3' |
| 12706 | 'ITIH3_No59' | 'NAHGEEKENLTAR' | 'Inter-alpha-trypsin inhibitor heavy chain H3' |
| 12707 | 'REG3A_No1' | 'NPSTISSPGHCASLSR' | 'Regenerating islet-derived protein 3-alpha' |
| 12708 | 'REG3A_No2' | 'AYGSHCYALFLSPK' | 'Regenerating islet-derived protein 3-alpha' |
| 12709 | 'BTK_No1' | 'FTNSETAEHIAQGLR' | 'Tyrosine-protein kinase BTK' |
| 12710 | 'BTK_No2' | 'NAPSTAGLGYGSWEIDPK' | 'Tyrosine-protein kinase BTK' |
| 12711 | 'BTK_No3' | 'AAVILESIFLK' | 'Tyrosine-protein kinase BTK' |
| 12712 | 'PRDX1_No1' | 'GLFIIDDKGILR' | 'Peroxiredoxin-1' |
| 12713 | 'PRDX1_No2' | 'HGEVCPAGWKPGSDTIKPDVQK' | 'Peroxiredoxin-1' |
| 12714 | 'PRDX1_No3' | 'QITVNDLPVGR' | 'Peroxiredoxin-1' |
| 12715 | 'PRDX1_No4' | 'QITVNDLPVGR' | 'Peroxiredoxin-1' |
| 12716 | 'PRDX1_No5' | 'LNCQVIGASVDSHFCHLAWVNTPK' | 'Peroxiredoxin-1' |
| 12717 | 'PRDX1_No6' | 'KQGGLGPMNIPLVSDPK' | 'Peroxiredoxin-1' |

| | A | B | C |
|---|---|---|---|
| 1 | **Peptide Name** | **Peptide Sequence** | **Peptide Description** |
| 12718 | 'PRDX1_No7' | 'TIAQDYGVLKADEGISFR' | 'Peroxiredoxin-1' |
| 12719 | 'DPT_No1' | 'YFESVLDREWQFYCCR' | 'Dermatopontin' |
| 12720 | 'DPT_No2' | 'QGFSYQCPQGQVIVAVR' | 'Dermatopontin' |
| 12721 | 'DPT_No3' | 'GATTTFSAVER' | 'Dermatopontin' |
| 12722 | 'LRP1_No1' | 'NVIALAFDYR' | 'Prolow-density lipoprotein receptor-related protein 1' |
| 12723 | 'LRP1_No2' | 'LDGLCIPLR' | 'Prolow-density lipoprotein receptor-related protein 1' |
| 12724 | 'LRP1_No3' | 'TVLWPNGLSLDIPAGR' | 'Prolow-density lipoprotein receptor-related protein 1' |
| 12725 | 'LRP1_No4' | 'IVFPHGITLDLVSR' | 'Prolow-density lipoprotein receptor-related protein 1' |
| 12726 | 'LRP1_No5' | 'SLDPFKPFIIFSNR' | 'Prolow-density lipoprotein receptor-related protein 1' |
| 12727 | 'LRP1_No6' | 'DCPDGSDEAPEICPQSK' | 'Prolow-density lipoprotein receptor-related protein 1' |
| 12728 | 'LRP1_No7' | 'TCADVDECSTTFPCSQR' | 'Prolow-density lipoprotein receptor-related protein 1' |
| 12729 | 'LRP1_No8' | 'LYWVDAFYDR' | 'Prolow-density lipoprotein receptor-related protein 1' |
| 12730 | 'LRP1_No9' | 'ILWIDAR' | 'Prolow-density lipoprotein receptor-related protein 1' |
| 12731 | 'LRP1_No10' | 'NLFWTSYDTNKK' | 'Prolow-density lipoprotein receptor-related protein 1' |
| 12732 | 'LRP1_No11' | 'WKCDGDHDCADGSDEKDCTPR' | 'Prolow-density lipoprotein receptor-related protein 1' |
| 12733 | 'LRP1_No12' | 'AALSGANVLTLIEK' | 'Prolow-density lipoprotein receptor-related protein 1' |
| 12734 | 'LRP1_No13' | 'SHACENDQYGKPGGCSDICLLANSHK' | 'Prolow-density lipoprotein receptor-related protein 1' |
| 12735 | 'LRP1_No14' | 'AVTDEEPFLIFANR' | 'Prolow-density lipoprotein receptor-related protein 1' |
| 12736 | 'LRP1_No15' | 'KPEHELFLVYGK' | 'Prolow-density lipoprotein receptor-related protein 1' |
| 12737 | 'LRP1_No16' | 'ACACAHGMLAEDGASCR' | 'Prolow-density lipoprotein receptor-related protein 1' |
| 12738 | 'LRP1_No17' | 'NGDTCVTLLDLELYNPK' | 'Prolow-density lipoprotein receptor-related protein 1' |
| 12739 | 'LRP1_No18' | 'SERPPIFEIR' | 'Prolow-density lipoprotein receptor-related protein 1' |
| 12740 | 'LRP1_No19' | 'TVLVSSGLREPR' | 'Prolow-density lipoprotein receptor-related protein 1' |
| 12741 | 'LRP1_No20' | 'ETVITMSGDDHPR' | 'Prolow-density lipoprotein receptor-related protein 1' |
| 12742 | 'LRP1_No21' | 'SGQQACEGVGSFLLYSVHEGIR' | 'Prolow-density lipoprotein receptor-related protein 1' |
| 12743 | 'LRP1_No22' | 'ITWPNGLTLDYVTER' | 'Prolow-density lipoprotein receptor-related protein 1' |
| 12744 | 'LRP1_No23' | 'IFFSDIHFGNIQQINDDGSR' | 'Prolow-density lipoprotein receptor-related protein 1' |
| 12745 | 'LRP1_No24' | 'CVAEALLCNGQDDCGDSSDER' | 'Prolow-density lipoprotein receptor-related protein 1' |
| 12746 | 'LRP1_No25' | 'VNNGGCSNLCLLSPGGGHK' | 'Prolow-density lipoprotein receptor-related protein 1' |
| 12747 | 'LRP1_No26' | 'LWWADQVSEK' | 'Prolow-density lipoprotein receptor-related protein 1' |
| 12748 | 'LRP1_No27' | 'SGFHTVPGQPGCQDINECLR' | 'Prolow-density lipoprotein receptor-related protein 1' |
| 12749 | 'LRP1_No28' | 'CDNDNDCGDNSDEAGCSHSCSSTQFK' | 'Prolow-density lipoprotein receptor-related protein 1' |
| 12750 | 'LRP1_No29' | 'SCMCTAGYSLR' | 'Prolow-density lipoprotein receptor-related protein 1' |
| 12751 | 'LRP1_No30' | 'QGLNNAVALDFDYR' | 'Prolow-density lipoprotein receptor-related protein 1' |
| 12752 | 'LRP1_No31' | 'LYFSDATLDKIER' | 'Prolow-density lipoprotein receptor-related protein 1' |
| 12753 | 'LRP1_No32' | 'ACACAHGMLAEDGASCR' | 'Prolow-density lipoprotein receptor-related protein 1' |
| 12754 | 'LRP1_No33' | 'VYDESIQLDHK' | 'Prolow-density lipoprotein receptor-related protein 1' |
| 12755 | 'LRP1_No34' | 'ETVITMSGDDHPR' | 'Prolow-density lipoprotein receptor-related protein 1' |
| 12756 | 'ARHGAP1_No1' | 'FLLDHQGELFPSPDPSGL' | 'Rho GTPase-activating protein 1' |
| 12757 | 'ARHGAP1_No2' | 'NPEQEPIPIVLR' | 'Rho GTPase-activating protein 1' |
| 12758 | 'ARHGAP1_No3' | 'IFYVNYLSELSEHVK' | 'Rho GTPase-activating protein 1' |
| 12759 | 'ARHGAP1_No4' | 'VPATLQVLQTLPEENYQVLR' | 'Rho GTPase-activating protein 1' |
| 12760 | 'ARHGAP1_No5' | 'HQIVEVAGDDKYGRK' | 'Rho GTPase-activating protein 1' |
| 12761 | 'ARHGAP1_No6' | 'HQIVEVAGDDKYGR' | 'Rho GTPase-activating protein 1' |

| | A | B | C |
|---|---|---|---|
| 1 | **Peptide Name** | **Peptide Sequence** | **Peptide Description** |
| 12762 | 'TGM3_No1' | 'SDLGPSYGGWQVLDATPQER' | 'Protein-glutamine gamma-glutamyltransferase E' |
| 12763 | 'TGM3_No2' | 'SQGVFQCGPASVIGVR' | 'Protein-glutamine gamma-glutamyltransferase E' |
| 12764 | 'TGM3_No3' | 'VRFDILPSR' | 'Protein-glutamine gamma-glutamyltransferase E' |
| 12765 | 'TGM3_No4' | 'VITNFNSAHDTDR' | 'Protein-glutamine gamma-glutamyltransferase E' |
| 12766 | 'TGM3_No5' | 'QAHHTDKFSSQELILR' | 'Protein-glutamine gamma-glutamyltransferase E' |
| 12767 | 'TGM3_No6' | 'GQNFQVLMIMNK' | 'Protein-glutamine gamma-glutamyltransferase E' |
| 12768 | 'TGM3_No7' | 'ITWLYDNTTGK' | 'Protein-glutamine gamma-glutamyltransferase E' |
| 12769 | 'TGM3_No8' | 'VPDESEVVVER' | 'Protein-glutamine gamma-glutamyltransferase E' |
| 12770 | 'TGM3_No9' | 'EEYVQEDAGIIFVGSTNR' | 'Protein-glutamine gamma-glutamyltransferase E' |
| 12771 | 'TGM3_No10' | 'MDVTDKYKYPEGSDQER' | 'Protein-glutamine gamma-glutamyltransferase E' |
| 12772 | 'LGALS3BP_No1' | 'SDLAVPSELALLK' | 'Galectin-3-binding protein' |
| 12773 | 'LGALS3BP_No2' | 'TLQALEFHTVPFQLLAR' | 'Galectin-3-binding protein' |
| 12774 | 'LGALS3BP_No3' | 'YYPYQSFQTPQHPSFLFQDK' | 'Galectin-3-binding protein' |
| 12775 | 'LGALS3BP_No4' | 'RIDITLSSVK' | 'Galectin-3-binding protein' |
| 12776 | 'LGALS3BP_No5' | 'ASHEEVEGLVEK' | 'Galectin-3-binding protein' |
| 12777 | 'LGALS3BP_No6' | 'YSSDYFQAPSDYR' | 'Galectin-3-binding protein' |
| 12778 | 'LGALS3BP_No7' | 'GQWGTVCDNLWDLTDASVVCR' | 'Galectin-3-binding protein' |
| 12779 | 'LGALS3BP_No8' | 'STSSFPCPAGHFNGFR' | 'Galectin-3-binding protein' |
| 12780 | 'LGALS3BP_No9' | 'KTLQALEFHTVPFQLLAR' | 'Galectin-3-binding protein' |
| 12781 | 'LGALS3BP_No10' | 'IDITLSSVK' | 'Galectin-3-binding protein' |
| 12782 | 'LGALS3BP_No11' | 'IYTSPTWSAFVTDSSWSAR' | 'Galectin-3-binding protein' |
| 12783 | 'LGALS3BP_No12' | 'ELSEALGQIFDSQR' | 'Galectin-3-binding protein' |
| 12784 | 'LGALS3BP_No13' | 'YYPYQSFQTPQHPSFLFQDKR' | 'Galectin-3-binding protein' |
| 12785 | 'LGALS3BP_No14' | 'LADGGATNQGR' | 'Galectin-3-binding protein' |
| 12786 | 'LGALS3BP_No15' | 'IYTSPTWSAFVTDSSWSARK' | 'Galectin-3-binding protein' |
| 12787 | 'LGALS3BP_No16' | 'AAFGQGSGPIMLDEVQCTGTEASLADCK' | 'Galectin-3-binding protein' |
| 12788 | 'LGALS3BP_No17' | 'GLNLTEDTYKPR' | 'Galectin-3-binding protein' |
| 12789 | 'LGALS3BP_No18' | 'ASHEEVEGLVEKIR' | 'Galectin-3-binding protein' |
| 12790 | 'LGALS3BP_No19' | 'AVDTWSWGERASHEEVEGLVEK' | 'Galectin-3-binding protein' |
| 12791 | 'LGALS3BP_No20' | 'LADGGATNQGRVEIFYR' | 'Galectin-3-binding protein' |
| 12792 | 'LGALS3BP_No21' | 'AVDTWSWGER' | 'Galectin-3-binding protein' |
| 12793 | 'LGALS3BP_No22' | 'KSQLVYQSR' | 'Galectin-3-binding protein' |
| 12794 | 'LGALS3BP_No23' | 'AAFGQGSGPIMLDEVQCTGTEASLADCK' | 'Galectin-3-binding protein' |
| 12795 | 'LGALS3BP_No24' | 'LASAYGAR' | 'Galectin-3-binding protein' |
| 12796 | 'LGALS3BP_No25' | 'DLLRYFYSR' | 'Galectin-3-binding protein' |
| 12797 | 'DMTN' | 'SPGIISQASAPR' | 'Dematin' |
| 12798 | 'DSC1_No1' | 'ILEDGSIYTTHDLILSSER' | 'Desmocollin-1' |
| 12799 | 'DSC1_No2' | 'MKVQDQDLPNTPHSK' | 'Desmocollin-1' |
| 12800 | 'DSC1_No3' | 'MKVQDQDLPNTPHSK' | 'Desmocollin-1' |
| 12801 | 'DSC1_No4' | 'HFSIHPDTGVITTTTPFLDREK' | 'Desmocollin-1' |
| 12802 | 'DSC1_No5' | 'VTATDLDEPDTLHTR' | 'Desmocollin-1' |
| 12803 | 'DSC1_No6' | 'VPSHLQAETLVGK' | 'Desmocollin-1' |
| 12804 | 'FGL1_No1' | 'NLHFLTTQEDYTLK' | 'Fibrinogen-like protein 1' |
| 12805 | 'FGL1_No2' | 'IDLADFEK' | 'Fibrinogen-like protein 1' |

| | A | B | C |
|---|---|---|---|
| 1 | **Peptide Name** | **Peptide Sequence** | **Peptide Description** |
| 12806 | 'FGL1_No3' | 'QYADCSEIFNDGYK' | 'Fibrinogen-like protein 1' |
| 12807 | 'FGL1_No4' | 'IRPNDFIPNVI' | 'Fibrinogen-like protein 1' |
| 12808 | 'FGL1_No5' | 'GWKDYENGFGNFVQK' | 'Fibrinogen-like protein 1' |
| 12809 | 'FGL1_No6' | 'QLLQENEVQFLDKGDENTVIDLGSKR' | 'Fibrinogen-like protein 1' |
| 12810 | 'FGL1_No7' | 'CHSANLNGVYYSGPYTAK' | 'Fibrinogen-like protein 1' |
| 12811 | 'FGL1_No8' | 'QLLQENEVQFLDKGDENTVIDLGSK' | 'Fibrinogen-like protein 1' |
| 12812 | 'FGL1_No9' | 'IKPLQSPAEFSVYCDMSDGGGWTVIQR' | 'Fibrinogen-like protein 1' |
| 12813 | 'FGL1_No10' | 'QLLQENEVQFLDK' | 'Fibrinogen-like protein 1' |
| 12814 | 'SIGLEC14_No1' | 'ALNPSQTSMSGTLELPNIGAR' | 'Sialic acid-binding Ig-like lectin 14' |
| 12815 | 'SIGLEC14_No2' | 'ALNPSQTSMSGTLELPNIGAR' | 'Sialic acid-binding Ig-like lectin 14' |
| 12816 | 'SIGLEC14_No3' | 'DGEIPYYAEVVATNNPDRR' | 'Sialic acid-binding Ig-like lectin 14' |
| 12817 | 'BST1_No1' | 'VADFLSWCR' | 'ADP-ribosyl cyclase/cyclic ADP-ribose hydrolase 2' |
| 12818 | 'BST1_No2' | 'GFFADYEIPNLQK' | 'ADP-ribosyl cyclase/cyclic ADP-ribose hydrolase 2' |
| 12819 | 'BST1_No3' | 'LLQCVDHSTHPDCALK' | 'ADP-ribosyl cyclase/cyclic ADP-ribose hydrolase 2' |
| 12820 | 'BST1_No4' | 'SLFWENSHLLVNSFADNTRR' | 'ADP-ribosyl cyclase/cyclic ADP-ribose hydrolase 2' |
| 12821 | 'BST1_No5' | 'IEIWVMHEIGGPNVESCGEGSMK' | 'ADP-ribosyl cyclase/cyclic ADP-ribose hydrolase 2' |
| 12822 | 'BST1_No6' | 'DMGFQYSCINDYRPVK' | 'ADP-ribosyl cyclase/cyclic ADP-ribose hydrolase 2' |
| 12823 | 'BST1_No7' | 'FMPLSDVLYGR' | 'ADP-ribosyl cyclase/cyclic ADP-ribose hydrolase 2' |
| 12824 | 'BST1_No8' | 'LKDMGFQYSCINDYRPVK' | 'ADP-ribosyl cyclase/cyclic ADP-ribose hydrolase 2' |
| 12825 | 'BST1_No9' | 'LKDMGFQYSCINDYRPVK' | 'ADP-ribosyl cyclase/cyclic ADP-ribose hydrolase 2' |
| 12826 | 'BST1_No10' | 'FMPLSDVLYGR' | 'ADP-ribosyl cyclase/cyclic ADP-ribose hydrolase 2' |
| 12827 | 'BST1_No11' | 'DMGFQYSCINDYRPVK' | 'ADP-ribosyl cyclase/cyclic ADP-ribose hydrolase 2' |
| 12828 | 'BST1_No12' | 'SLFWENSHLLVNSFADNTR' | 'ADP-ribosyl cyclase/cyclic ADP-ribose hydrolase 2' |
| 12829 | 'HYAL1_No1' | 'ALYPSIYMPAVLEGTGK' | 'Hyaluronidase-1' |
| 12830 | 'HYAL1_No2' | 'GALSLEDQAQMAVEFK' | 'Hyaluronidase-1' |
| 12831 | 'HYAL1_No3' | 'ALYPSIYMPAVLEGTGK' | 'Hyaluronidase-1' |
| 12832 | 'HYAL1_No4' | 'AQNDQLGWLWGQSR' | 'Hyaluronidase-1' |
| 12833 | 'HYAL1_No5' | 'AWMAGTLQLGR' | 'Hyaluronidase-1' |
| 12834 | 'HYAL1_No6' | 'WAFNWDTK' | 'Hyaluronidase-1' |
| 12835 | 'EFEMP1_No1' | 'NPCQDPYILTPENR' | 'EGF-containing fibulin-like extracellular matrix protein 1' |
| 12836 | 'EFEMP1_No2' | 'FSCMCPQGYQVVR' | 'EGF-containing fibulin-like extracellular matrix protein 1' |
| 12837 | 'EFEMP1_No3' | 'RGEQCVDIDECTIPPYCHQR' | 'EGF-containing fibulin-like extracellular matrix protein 1' |
| 12838 | 'EFEMP1_No4' | 'ADQVCINLR' | 'EGF-containing fibulin-like extracellular matrix protein 1' |
| 12839 | 'EFEMP1_No5' | 'TCQDINECETTNECREDEMCWNYHGGFR' | 'EGF-containing fibulin-like extracellular matrix protein 1' |
| 12840 | 'EFEMP1_No6' | 'CVCPVSNAMCR' | 'EGF-containing fibulin-like extracellular matrix protein 1' |
| 12841 | 'EFEMP1_No7' | 'LNCEDIDECR' | 'EGF-containing fibulin-like extracellular matrix protein 1' |
| 12842 | 'EFEMP1_No8' | 'FSCMCPQGYQVVR' | 'EGF-containing fibulin-like extracellular matrix protein 1' |
| 12843 | 'EFEMP1_No9' | 'IQCAAGYEQSEHNVCQDIDECTAGTHNCR' | 'EGF-containing fibulin-like extracellular matrix protein 1' |
| 12844 | 'EFEMP1_No10' | 'TCQDINECETTNECREDEMCWNYHGGFR' | 'EGF-containing fibulin-like extracellular matrix protein 1' |
| 12845 | 'EFEMP1_No11' | 'RNPADPQRIPSNPSHR' | 'EGF-containing fibulin-like extracellular matrix protein 1' |
| 12846 | 'EFEMP1_No12' | 'TCQDINECETTNECR' | 'EGF-containing fibulin-like extracellular matrix protein 1' |
| 12847 | 'EFEMP1_No13' | 'DIDECDIVPDACK' | 'EGF-containing fibulin-like extracellular matrix protein 1' |
| 12848 | 'EFEMP1_No14' | 'EHIVDLEMLTVSSIGTFR' | 'EGF-containing fibulin-like extracellular matrix protein 1' |
| 12849 | 'EFEMP1_No15' | 'GSFACQCPPGYQK' | 'EGF-containing fibulin-like extracellular matrix protein 1' |

| | A | B | C |
|---|---|---|---|
| 1 | **Peptide Name** | **Peptide Sequence** | **Peptide Description** |
| 12850 | 'EFEMP1_No16' | 'SGNENGEFYLR' | 'EGF-containing fibulin-like extracellular matrix protein 1' |
| 12851 | 'EFEMP1_No17' | 'CVNHYGGYLCLPK' | 'EGF-containing fibulin-like extracellular matrix protein 1' |
| 12852 | 'EFEMP1_No18' | 'CVCPVSNAMCR' | 'EGF-containing fibulin-like extracellular matrix protein 1' |
| 12853 | 'EFEMP1_No19' | 'QTSPVSAMLVLVK' | 'EGF-containing fibulin-like extracellular matrix protein 1' |
| 12854 | 'EFEMP1_No20' | 'EHIVDLEMLTVSSIGTFR' | 'EGF-containing fibulin-like extracellular matrix protein 1' |
| 12855 | 'EFEMP1_No21' | 'NPADPQRIPSNPSHR' | 'EGF-containing fibulin-like extracellular matrix protein 1' |
| 12856 | 'EFEMP1_No22' | 'QQCKDIDECDIVPDACK' | 'EGF-containing fibulin-like extracellular matrix protein 1' |
| 12857 | 'EFEMP1_No23' | 'IKSGNENGEFYLR' | 'EGF-containing fibulin-like extracellular matrix protein 1' |
| 12858 | 'EFEMP1_No24' | 'GEQCVDIDECTIPPYCHQR' | 'EGF-containing fibulin-like extracellular matrix protein 1' |
| 12859 | 'EFEMP1_No25' | 'GSFACQCPPGYQKR' | 'EGF-containing fibulin-like extracellular matrix protein 1' |
| 12860 | 'EFEMP1_No26' | 'IQCAAGYEQSEHNVCQDIDECTAGTHNCRADQVCINLR' | 'EGF-containing fibulin-like extracellular matrix protein 1' |
| 12861 | 'EFEMP1_No27' | 'QQCKDIDECDIVPDACK' | 'EGF-containing fibulin-like extracellular matrix protein 1' |
| 12862 | 'EFEMP1_No28' | 'QTSPVSAMLVLVK' | 'EGF-containing fibulin-like extracellular matrix protein 1' |
| 12863 | 'EFEMP1_No29' | 'CYPRNPCQDPYILTPENR' | 'EGF-containing fibulin-like extracellular matrix protein 1' |
| 12864 | 'EFEMP1_No30' | 'QTSPVSAMLVLVK' | 'EGF-containing fibulin-like extracellular matrix protein 1' |
| 12865 | 'EFEMP1_No31' | 'EDEMCWNYHGGFR' | 'EGF-containing fibulin-like extracellular matrix protein 1' |
| 12866 | 'EFEMP1_No32' | 'TSSYLCQYQCVNEPGKFSCMCPQGYQVVR' | 'EGF-containing fibulin-like extracellular matrix protein 1' |
| 12867 | 'EFEMP1_No33' | 'DIDECDIVPDACKGGMK' | 'EGF-containing fibulin-like extracellular matrix protein 1' |
| 12868 | 'EFEMP1_No34' | 'QTSPVSAMLVLVK' | 'EGF-containing fibulin-like extracellular matrix protein 1' |
| 12869 | 'EFEMP1_No35' | 'SRTCQDINECETTNECREDEMCWNYHGGFR' | 'EGF-containing fibulin-like extracellular matrix protein 1' |
| 12870 | 'CNTN1_No1' | 'FIPLIPIPER' | 'Contactin-1' |
| 12871 | 'CNTN1_No2' | 'IKTDGAAPNVAPSDVGGGGGR' | 'Contactin-1' |
| 12872 | 'CNTN1_No3' | 'MNNGDVDLTSDR' | 'Contactin-1' |
| 12873 | 'CNTN1_No4' | 'TDGAAPNVAPSDVGGGGGR' | 'Contactin-1' |
| 12874 | 'CNTN1_No5' | 'VLEPMPSTAEISTSGAVLK' | 'Contactin-1' |
| 12875 | 'CNTN1_No6' | 'TTKPYPADIVVQFK' | 'Contactin-1' |
| 12876 | 'CNTN1_No7' | 'HSIEVPIPR' | 'Contactin-1' |
| 12877 | 'CNTN1_No8' | 'VLSSSEISVHWEHVLEK' | 'Contactin-1' |
| 12878 | 'CNTN1_No9' | 'DGEYVVEVR' | 'Contactin-1' |
| 12879 | 'CNTN1_No10' | 'DAGIYYCLASNNYGMVR' | 'Contactin-1' |
| 12880 | 'CNTN1_No11' | 'GDGPYSLVAVINSAQDAPSEAPTEVGVK' | 'Contactin-1' |
| 12881 | 'CNTN1_No12' | 'TILSDDWK' | 'Contactin-1' |
| 12882 | 'CNTN1_No13' | 'ILALAPTFEMNPMK' | 'Contactin-1' |
| 12883 | 'CNTN1_No14' | 'NFMLDSNGELLIR' | 'Contactin-1' |
| 12884 | 'CNTN1_No15' | 'ELTITWAPLSR' | 'Contactin-1' |
| 12885 | 'CNTN1_No16' | 'YVHKDETMSPSTAFQVK' | 'Contactin-1' |
| 12886 | 'CNTN1_No17' | 'YGHGVSEEDKGFGPIFEEQPINTIYPEESLEGK' | 'Contactin-1' |
| 12887 | 'CNTN1_No18' | 'YTCTAQTIVDNSSASADLVVR' | 'Contactin-1' |
| 12888 | 'FAP_No1' | 'GNELPRPIQYLCWSPVGSK' | 'Prolyl endopeptidase FAP' |
| 12889 | 'FAP_No2' | 'YALWWSPNGK' | 'Prolyl endopeptidase FAP' |
| 12890 | 'FAP_No3' | 'KLEVDEITLWYK' | 'Prolyl endopeptidase FAP' |
| 12891 | 'FAP_No4' | 'GTAFQGDKLLYAVYR' | 'Prolyl endopeptidase FAP' |
| 12892 | 'FAP_No5' | 'YYALVCYGPGIPISTLHDGR' | 'Prolyl endopeptidase FAP' |
| 12893 | 'FAP_No6' | 'NVDYLLIHGTADDNVHFQNSAQIAK' | 'Prolyl endopeptidase FAP' |

| | A | B | C |
|---|---|---|---|
| 1 | **Peptide Name** | **Peptide Sequence** | **Peptide Description** |
| 12894 | 'FAP_No7' | 'MILPPQFDR' | 'Prolyl endopeptidase FAP' |
| 12895 | 'LMAN2_No1' | 'DNFHGLAIFLDTYPNDETTER' | 'Vesicular integral-membrane protein VIP36' |
| 12896 | 'LMAN2_No2' | 'LTVMTDLEDKNEWK' | 'Vesicular integral-membrane protein VIP36' |
| 12897 | 'LMAN2_No3' | 'WTELAGCTADFR' | 'Vesicular integral-membrane protein VIP36' |
| 12898 | 'LMAN2_No4' | 'LPTGYYFGASAGTGDLSDNHDIISMK' | 'Vesicular integral-membrane protein VIP36' |
| 12899 | 'LMAN2_No5' | 'NLHGDGIALWYTR' | 'Vesicular integral-membrane protein VIP36' |
| 12900 | 'PTPRJ_No1' | 'AVSISPTNVILTWK' | 'Receptor-type tyrosine-protein phosphatase eta' |
| 12901 | 'PTPRJ_No2' | 'AGSPTAPVHDESLVGPVDPSSGQQSR' | 'Receptor-type tyrosine-protein phosphatase eta' |
| 12902 | 'PTPRJ_No3' | 'VITEPIPVSDLR' | 'Receptor-type tyrosine-protein phosphatase eta' |
| 12903 | 'PTPRJ_No4' | 'TKGDPLGTEGGLDASNTER' | 'Receptor-type tyrosine-protein phosphatase eta' |
| 12904 | 'PTPRJ_No5' | 'DTEVLLVGLEPGTR' | 'Receptor-type tyrosine-protein phosphatase eta' |
| 12905 | 'PTPRJ_No6' | 'TPSSTGPSPVFDIK' | 'Receptor-type tyrosine-protein phosphatase eta' |
| 12906 | 'PTPRJ_No7' | 'VLLESIGSHEELTQDSR' | 'Receptor-type tyrosine-protein phosphatase eta' |
| 12907 | 'PTPRJ_No8' | 'AYAVILTTGEAGHPSADVLK' | 'Receptor-type tyrosine-protein phosphatase eta' |
| 12908 | 'PTPRJ_No9' | 'VEITTNQSIIIGGLFPGTK' | 'Receptor-type tyrosine-protein phosphatase eta' |
| 12909 | 'PTPRJ_No10' | 'GDPLGTEGGLDASNTER' | 'Receptor-type tyrosine-protein phosphatase eta' |
| 12910 | 'PTPRJ_No11' | 'SPDGASEYVYHLVIESK' | 'Receptor-type tyrosine-protein phosphatase eta' |
| 12911 | 'ECH1' | 'VIGNQSLVNELAFTAR' | 'Delta(3,5)-Delta(2,4)-dienoyl-CoA isomerase, mitochondrial' |
| 12912 | 'PLA2G7_No1' | 'YPLVVFSHGLGAFR' | 'Platelet-activating factor acetylhydrolase' |
| 12913 | 'PLA2G7_No2' | 'TLKQEEETHIRNEQVR' | 'Platelet-activating factor acetylhydrolase' |
| 12914 | 'PLA2G7_No3' | 'ASLAFLQK' | 'Platelet-activating factor acetylhydrolase' |
| 12915 | 'PLA2G7_No4' | 'IAVIGHSFGGATVIQTLSEDQR' | 'Platelet-activating factor acetylhydrolase' |
| 12916 | 'PLA2G7_No5' | 'LLFGSMTTPANWNSPLRPGEK' | 'Platelet-activating factor acetylhydrolase' |
| 12917 | 'PLA2G7_No6' | 'LYYPSQDNDRLDTLWIPNK' | 'Platelet-activating factor acetylhydrolase' |
| 12918 | 'PLA2G7_No7' | 'GSVHQNFADFTFATGK' | 'Platelet-activating factor acetylhydrolase' |
| 12919 | 'PLA2G7_No8' | 'FDMEQLKDSIDREK' | 'Platelet-activating factor acetylhydrolase' |
| 12920 | 'PLA2G7_No9' | 'AKECSQALSLILDIDHGKPVK' | 'Platelet-activating factor acetylhydrolase' |
| 12921 | 'PLA2G7_No10' | 'ECSQALSLILDIDHGKPVK' | 'Platelet-activating factor acetylhydrolase' |
| 12922 | 'PLA2G7_No11' | 'IQVLMAAASFGQTK' | 'Platelet-activating factor acetylhydrolase' |
| 12923 | 'PLA2G7_No12' | 'GNGPYSVGCTDLMFDHTNK' | 'Platelet-activating factor acetylhydrolase' |
| 12924 | 'PLA2G7_No13' | 'FLGTHWLMGNILR' | 'Platelet-activating factor acetylhydrolase' |
| 12925 | 'PLA2G7_No14' | 'FDMEQLKDSIDR' | 'Platelet-activating factor acetylhydrolase' |
| 12926 | 'PLA2G7_No15' | 'DQSAAEIGDKSWLYLR' | 'Platelet-activating factor acetylhydrolase' |
| 12927 | 'PLA2G7_No16' | 'LLFGSMTTPANWNSPLRPGEK' | 'Platelet-activating factor acetylhydrolase' |
| 12928 | 'PLA2G7_No17' | 'NALDLKFDMEQLKDSIDR' | 'Platelet-activating factor acetylhydrolase' |
| 12929 | 'PLA2G7_No18' | 'LKGDIDSNVAIDLSNK' | 'Platelet-activating factor acetylhydrolase' |
| 12930 | 'PLA2G7_No19' | 'FLGTHWLMGNILR' | 'Platelet-activating factor acetylhydrolase' |
| 12931 | 'PLA2G7_No20' | 'FDMEQLKDSIDREK' | 'Platelet-activating factor acetylhydrolase' |
| 12932 | 'IFT88' | 'ISSTAVTR' | 'Intraflagellar transport protein 88 homolog' |
| 12933 | 'SPP2_No1' | 'VNSQSLSPYLFR' | 'Secreted phosphoprotein 24' |
| 12934 | 'SPP2_No2' | 'DSGEDPATCAFQR' | 'Secreted phosphoprotein 24' |
| 12935 | 'SPP2_No3' | 'VSAQQVQGVHAR' | 'Secreted phosphoprotein 24' |
| 12936 | 'SPP2_No4' | 'KDSGEDPATCAFQR' | 'Secreted phosphoprotein 24' |
| 12937 | 'SPP2_No5' | 'STVKVSAQQVQGVHAR' | 'Secreted phosphoprotein 24' |

| | A | B | C |
|---|---|---|---|
| 1 | **Peptide Name** | **Peptide Sequence** | **Peptide Description** |
| 12938 | 'MTAP' | 'TWGADVINMTTVPEVVLAK' | 'S-methyl-5''-thioadenosine phosphorylase' |
| 12939 | 'PRDX4_No1' | 'DYGVYLEDSGHTLR' | 'Peroxiredoxin-4' |
| 12940 | 'PRDX4_No2' | 'VSVADHSLHLSK' | 'Peroxiredoxin-4' |
| 12941 | 'MMRN1_No1' | 'NTDNIIYPEEYSSCSR' | 'Multimerin-1' |
| 12942 | 'MMRN1_No2' | 'LVEENALAPDFSK' | 'Multimerin-1' |
| 12943 | 'MMRN1_No3' | 'GLTEFVEPIIQIK' | 'Multimerin-1' |
| 12944 | 'MMRN1_No4' | 'FPPVTTFSGYLLYR' | 'Multimerin-1' |
| 12945 | 'MMRN1_No5' | 'AQEQQSLIHTNQAESHTAVGR' | 'Multimerin-1' |
| 12946 | 'MMRN1_No6' | 'QTHLEGALEQEHSR' | 'Multimerin-1' |
| 12947 | 'MMRN1_No7' | 'LSPTVILDNQVTYVPGGK' | 'Multimerin-1' |
| 12948 | 'MMRN1_No8' | 'GPCGWTGGSCPQR' | 'Multimerin-1' |
| 12949 | 'MMRN1_No9' | 'GVAEQQQQQGCGDPEVMQK' | 'Multimerin-1' |
| 12950 | 'MMRN1_No10' | 'YNFVLQVAK' | 'Multimerin-1' |
| 12951 | 'MMRN1_No11' | 'IENLTSAVNSLNFIIK' | 'Multimerin-1' |
| 12952 | 'MMRN1_No12' | 'GTIPAKFPPVTTFSGYLLYR' | 'Multimerin-1' |
| 12953 | 'MMRN1_No13' | 'DQALQLQVLNSR' | 'Multimerin-1' |
| 12954 | 'MMRN1_No14' | 'QEMTLTCEKPIK' | 'Multimerin-1' |
| 12955 | 'MMRN1_No15' | 'LKEVHEQLLSTEQVSDQK' | 'Multimerin-1' |
| 12956 | 'MMRN1_No16' | 'NWCAYVHTR' | 'Multimerin-1' |
| 12957 | 'MMRN1_No17' | 'IPYLGVYVFK' | 'Multimerin-1' |
| 12958 | 'MMRN1_No18' | 'KKIENLTSAVNSLNFIIK' | 'Multimerin-1' |
| 12959 | 'MMRN1_No19' | 'FVLVQENRPTLTDIVELR' | 'Multimerin-1' |
| 12960 | 'MMRN1_No20' | 'GVAEQQQQQGCGDPEVMQK' | 'Multimerin-1' |
| 12961 | 'MMRN1_No21' | 'ESLRGECEDMLSK' | 'Multimerin-1' |
| 12962 | 'MMRN1_No22' | 'HSLPDIQLLQK' | 'Multimerin-1' |
| 12963 | 'MMRN1_No23' | 'HSWTIPEDGNSQK' | 'Multimerin-1' |
| 12964 | 'MMRN1_No24' | 'QTHLEGALEQEHSR' | 'Multimerin-1' |
| 12965 | 'MMRN1_No25' | 'LAFESENINSEIHCDR' | 'Multimerin-1' |
| 12966 | 'MMRN1_No26' | 'IFQNDMQETVAQLFK' | 'Multimerin-1' |
| 12967 | 'MMRN1_No27' | 'KIENLTSAVNSLNFIIK' | 'Multimerin-1' |
| 12968 | 'MMRN1_No28' | 'VTQTLIPYYISVK' | 'Multimerin-1' |
| 12969 | 'MMRN1_No29' | 'KSNEQATSLNTVGGTGGIGGVGGTGGVGNR' | 'Multimerin-1' |
| 12970 | 'MMRN1_No30' | 'QEMTLTCEKPIKELEVK' | 'Multimerin-1' |
| 12971 | 'MMRN1_No31' | 'TVSSLSEDLESTR' | 'Multimerin-1' |
| 12972 | 'MMRN1_No32' | 'ESLRGECEDMLSK' | 'Multimerin-1' |
| 12973 | 'SELENBP1_No1' | 'LVLPSLISSR' | 'Methanethiol oxidase' |
| 12974 | 'SELENBP1_No2' | 'SPQYCQVIHR' | 'Methanethiol oxidase' |
| 12975 | 'SELENBP1_No3' | 'NTGTEAPDYLATVDVDPK' | 'Methanethiol oxidase' |
| 12976 | 'SELENBP1_No4' | 'FLHNPDAAQGFVGCALSSTIQR' | 'Methanethiol oxidase' |
| 12977 | 'SELENBP1_No5' | 'QYDISDPQRPR' | 'Methanethiol oxidase' |
| 12978 | 'SELENBP1_No6' | 'LNPNFLVDFGK' | 'Methanethiol oxidase' |
| 12979 | 'SELENBP1_No7' | 'DGLIPLEIR' | 'Methanethiol oxidase' |
| 12980 | 'SELENBP1_No8' | 'GGPVQVLEDEELK' | 'Methanethiol oxidase' |
| 12981 | 'SELENBP1_No9' | 'HEIVQTLSLKDGLIPLEIR' | 'Methanethiol oxidase' |

| | A | B | C |
|---|---|---|---|
| 1 | **Peptide Name** | **Peptide Sequence** | **Peptide Description** |
| 12982 | 'SELENBP1_No10' | 'GGFVLLDGETFEVK' | 'Methanethiol oxidase' |
| 12983 | 'SELENBP1_No11' | 'GGPVQVLEDEELKSQPEPLVVK' | 'Methanethiol oxidase' |
| 12984 | 'CHIT1_No1' | 'VGAPATGSGTPGPFTK' | 'Chitotriosidase-1' |
| 12985 | 'CHIT1_No2' | 'TLLAIGGWNFGTQK' | 'Chitotriosidase-1' |
| 12986 | 'CHIT1_No3' | 'LFQQSCPTGLVFSNSCK' | 'Chitotriosidase-1' |
| 12987 | 'CHIT1_No4' | 'YPLIQTLR' | 'Chitotriosidase-1' |
| 12988 | 'CHIT1_No5' | 'LVCYFTNWAQYR' | 'Chitotriosidase-1' |
| 12989 | 'PTPRS_No1' | 'SPQGLGAFTPVVR' | 'Receptor-type tyrosine-protein phosphatase S' |
| 12990 | 'PTPRS_No2' | 'YSSPANLYVR' | 'Receptor-type tyrosine-protein phosphatase S' |
| 12991 | 'PTPRS_No3' | 'ITTVAHTEVGPGPESSPVVVR' | 'Receptor-type tyrosine-protein phosphatase S' |
| 12992 | 'PTPRS_No4' | 'DENVYECVAQNSVGEITVHAK' | 'Receptor-type tyrosine-protein phosphatase S' |
| 12993 | 'PTPRS_No5' | 'DQIGVSGGVASFVCQATGDPKPR' | 'Receptor-type tyrosine-protein phosphatase S' |
| 12994 | 'PTPRS_No6' | 'ETELPAAAEPGAENALTLQGLKPDTAYDLQVR' | 'Receptor-type tyrosine-protein phosphatase S' |
| 12995 | 'PTPRS_No7' | 'TFDPTTSYVVEDLKPNTEYAFR' | 'Receptor-type tyrosine-protein phosphatase S' |
| 12996 | 'PTPRS_No8' | 'GGLGEEAAEVLSIPEDTPR' | 'Receptor-type tyrosine-protein phosphatase S' |
| 12997 | 'MFAP5_No1' | 'LVCKEHEAMKDELCR' | 'Microfibrillar-associated protein 5' |
| 12998 | 'MFAP5_No2' | 'QCIHQLCFTSLR' | 'Microfibrillar-associated protein 5' |
| 12999 | 'MFAP5_No3' | 'QCIHQLCFTSLR' | 'Microfibrillar-associated protein 5' |
| 13000 | 'MFAP5_No4' | 'LVCKEHEAMKDELCR' | 'Microfibrillar-associated protein 5' |
| 13001 | 'ILK_No1' | 'FSFQCPGR' | 'Integrin-linked protein kinase' |
| 13002 | 'ILK_No2' | 'GMAFLHTLEPLIPR' | 'Integrin-linked protein kinase' |
| 13003 | 'ILK_No3' | 'MYAPAWVAPEALQK' | 'Integrin-linked protein kinase' |
| 13004 | 'ILK_No4' | 'FALDMAR' | 'Integrin-linked protein kinase' |
| 13005 | 'ILK_No5' | 'VALEGLRPTIPPGISPHVCK' | 'Integrin-linked protein kinase' |
| 13006 | 'ILK_No6' | 'MDDIFTQCR' | 'Integrin-linked protein kinase' |
| 13007 | 'ILK_No7' | 'FDMIVPILEK' | 'Integrin-linked protein kinase' |
| 13008 | 'ILK_No8' | 'EVPFADLSNMEIGMK' | 'Integrin-linked protein kinase' |
| 13009 | 'ILK_No9' | 'FDMIVPILEK' | 'Integrin-linked protein kinase' |
| 13010 | 'ILK_No10' | 'GMAFLHTLEPLIPR' | 'Integrin-linked protein kinase' |
| 13011 | 'ILK_No11' | 'LWLDNTENDLNQGDDHGFSPLHWACR' | 'Integrin-linked protein kinase' |
| 13012 | 'ILK_No12' | 'MYAPAWVAPEALQK' | 'Integrin-linked protein kinase' |
| 13013 | 'ILK_No13' | 'SAVVEMLIMR' | 'Integrin-linked protein kinase' |
| 13014 | 'ILK_No14' | 'GDDTPLHLAASHGHR' | 'Integrin-linked protein kinase' |
| 13015 | 'ILK_No15' | 'SAVVEMLIMR' | 'Integrin-linked protein kinase' |
| 13016 | 'MADCAM1' | 'GLDTSLGAVQSDTGR' | 'Mucosal addressin cell adhesion molecule 1' |
| 13017 | 'TUBB3_No1' | 'AILVDLEPGTMDSVR' | 'Tubulin beta-3 chain' |
| 13018 | 'TUBB3_No2' | 'MSSTFIGNSTAIQELFK' | 'Tubulin beta-3 chain' |
| 13019 | 'TUBB3_No3' | 'VAVCDIPPR' | 'Tubulin beta-3 chain' |
| 13020 | 'TUBB3_No4' | 'MSSTFIGNSTAIQELFK' | 'Tubulin beta-3 chain' |
| 13021 | 'TUBB3_No5' | 'AILVDLEPGTMDSVR' | 'Tubulin beta-3 chain' |
| 13022 | 'DCTN2_No1' | 'LLGPDAAINLTDPDGALAK' | 'Dynactin subunit 2' |
| 13023 | 'DCTN2_No2' | 'WSPIASTLPELVQR' | 'Dynactin subunit 2' |
| 13024 | 'IQGAP2_No1' | 'NPNAVLTLVDDNLAPEYQK' | 'Ras GTPase-activating-like protein IQGAP2' |
| 13025 | 'IQGAP2_No2' | 'AWVNQLETQTGEASK' | 'Ras GTPase-activating-like protein IQGAP2' |

| | A | B | C |
|---|---|---|---|
| 1 | **Peptide Name** | **Peptide Sequence** | **Peptide Description** |
| 13026 | 'IQGAP2_No3' | 'IFYPETTDVYDRK' | 'Ras GTPase-activating-like protein IQGAP2' |
| 13027 | 'STIM1_No1' | 'SHSPSSPDPDTPSPVGDSR' | 'Stromal interaction molecule 1' |
| 13028 | 'STIM1_No2' | 'ALDTVLFGPPLLTR' | 'Stromal interaction molecule 1' |
| 13029 | 'STIM1_No3' | 'DLTHSDSESSLHMSDR' | 'Stromal interaction molecule 1' |
| 13030 | 'STIM1_No4' | 'DLTHSDSESSLHMSDR' | 'Stromal interaction molecule 1' |
| 13031 | 'RAB32_No1' | 'DSSQSPSQVDQFCK' | 'Ras-related protein Rab-32' |
| 13032 | 'RAB32_No2' | 'VLVIGELGVGK' | 'Ras-related protein Rab-32' |
| 13033 | 'RAB32_No3' | 'YVHQLFSQHYR' | 'Ras-related protein Rab-32' |
| 13034 | 'FHL1_No1' | 'AIVAGDQNVEYK' | 'Four and a half LIM domains protein 1' |
| 13035 | 'FHL1_No2' | 'FTAVEDQYYCVDCYK' | 'Four and a half LIM domains protein 1' |
| 13036 | 'FHL1_No3' | 'AITSGGITYQDQPWHADCFVCVTCSK' | 'Four and a half LIM domains protein 1' |
| 13037 | 'FHL1_No4' | 'CLHPLANETFVAK' | 'Four and a half LIM domains protein 1' |
| 13038 | 'FHL1_No5' | 'FWHDTCFR' | 'Four and a half LIM domains protein 1' |
| 13039 | 'ALCAM_No1' | 'ESLTLIVEGKPQIK' | 'CD166 antigen' |
| 13040 | 'ALCAM_No2' | 'ADIQMPFTCSVTYYGPSGQK' | 'CD166 antigen' |
| 13041 | 'ALCAM_No3' | 'FVCMLVTEDNVFEAPTIVK' | 'CD166 antigen' |
| 13042 | 'ALCAM_No4' | 'SMIASTAITVHYLDLSLNPSGEVTR' | 'CD166 antigen' |
| 13043 | 'ALCAM_No5' | 'VLHPLEGAVVIIFK' | 'CD166 antigen' |
| 13044 | 'ALCAM_No6' | 'VFKQPSKPEIVSK' | 'CD166 antigen' |
| 13045 | 'ALCAM_No7' | 'KSVQYDDVPEYKDR' | 'CD166 antigen' |
| 13046 | 'ALCAM_No8' | 'SSPSFSSLHYQDAGNYVCETALQEVEGLK' | 'CD166 antigen' |
| 13047 | 'ALCAM_No9' | 'SVQYDDVPEYKDR' | 'CD166 antigen' |
| 13048 | 'ALCAM_No10' | 'WKYEKPDGSPVFIAFR' | 'CD166 antigen' |
| 13049 | 'ALCAM_No11' | 'FVCMLVTEDNVFEAPTIVK' | 'CD166 antigen' |
| 13050 | 'ALCAM_No12' | 'ALFLETEQLKK' | 'CD166 antigen' |
| 13051 | 'ALCAM_No13' | 'ALFLETEQLK' | 'CD166 antigen' |
| 13052 | 'ALCAM_No14' | 'ADIQMPFTCSVTYYGPSGQK' | 'CD166 antigen' |
| 13053 | 'ALCAM_No15' | 'LDVPQNLMFGK' | 'CD166 antigen' |
| 13054 | 'ALCAM_No16' | 'SSNTYTLTDVRR' | 'CD166 antigen' |
| 13055 | 'APOF_No1' | 'SGVQQLIQYYQDQK' | 'Apolipoprotein F' |
| 13056 | 'APOF_No2' | 'FLVSLALR' | 'Apolipoprotein F' |
| 13057 | 'APOF_No3' | 'SYDLDPGAGSLEI' | 'Apolipoprotein F' |
| 13058 | 'APOF_No4' | 'DANISQPETTKEGLR' | 'Apolipoprotein F' |
| 13059 | 'ENPP2_No1' | 'WWGGQPLWITATK' | 'Ectonucleotide pyrophosphatase/phosphodiesterase family member 2' |
| 13060 | 'ENPP2_No2' | 'NGVNVISGPIFDYDYDGLHDTEDKIK' | 'Ectonucleotide pyrophosphatase/phosphodiesterase family member 2' |
| 13061 | 'ENPP2_No3' | 'YGPFGPEMTNPLR' | 'Ectonucleotide pyrophosphatase/phosphodiesterase family member 2' |
| 13062 | 'ENPP2_No4' | 'VRDIEHLTSLDFFR' | 'Ectonucleotide pyrophosphatase/phosphodiesterase family member 2' |
| 13063 | 'ENPP2_No5' | 'NEENACHCSEDCLAR' | 'Ectonucleotide pyrophosphatase/phosphodiesterase family member 2' |
| 13064 | 'ENPP2_No6' | 'QAEVSSVPDHLTSCVRPDVR' | 'Ectonucleotide pyrophosphatase/phosphodiesterase family member 2' |
| 13065 | 'ENPP2_No7' | 'DIEHLTSLDFFR' | 'Ectonucleotide pyrophosphatase/phosphodiesterase family member 2' |
| 13066 | 'ENPP2_No8' | 'SYTSCCHDFDELCLK' | 'Ectonucleotide pyrophosphatase/phosphodiesterase family member 2' |
| 13067 | 'ENPP2_No9' | 'RIEDIHLLVER' | 'Ectonucleotide pyrophosphatase/phosphodiesterase family member 2' |
| 13068 | 'ENPP2_No10' | 'GESHWVDDDCEEIK' | 'Ectonucleotide pyrophosphatase/phosphodiesterase family member 2' |
| 13069 | 'ENPP2_No11' | 'IEDIHLLVER' | 'Ectonucleotide pyrophosphatase/phosphodiesterase family member 2' |

| | A | B | C |
|---|---|---|---|
| 1 | **Peptide Name** | **Peptide Sequence** | **Peptide Description** |
| 13070 | 'ENPP2_No12' | 'GDCCTNYQVVCK' | 'Ectonucleotide pyrophosphatase/phosphodiesterase family member 2' |
| 13071 | 'ENPP2_No13' | 'VSPSFSQNCLAYK' | 'Ectonucleotide pyrophosphatase/phosphodiesterase family member 2' |
| 13072 | 'ENPP2_No14' | 'QMSYGFLFPPYLSSSPEAK' | 'Ectonucleotide pyrophosphatase/phosphodiesterase family member 2' |
| 13073 | 'ENPP2_No15' | 'YDAFLVTNMVPMYPAFK' | 'Ectonucleotide pyrophosphatase/phosphodiesterase family member 2' |
| 13074 | 'ENPP2_No16' | 'EIDKIVGQLMDGLK' | 'Ectonucleotide pyrophosphatase/phosphodiesterase family member 2' |
| 13075 | 'ENPP2_No17' | 'QMSYGFLFPPYLSSSPEAK' | 'Ectonucleotide pyrophosphatase/phosphodiesterase family member 2' |
| 13076 | 'ENPP2_No18' | 'HLLYGRPAVLYR' | 'Ectonucleotide pyrophosphatase/phosphodiesterase family member 2' |
| 13077 | 'ENPP2_No19' | 'VWNYFQR' | 'Ectonucleotide pyrophosphatase/phosphodiesterase family member 2' |
| 13078 | 'ENPP2_No20' | 'AGTFFWSVVIPHER' | 'Ectonucleotide pyrophosphatase/phosphodiesterase family member 2' |
| 13079 | 'ENPP2_No21' | 'QAEVSSVPDHLTSCVRPDVR' | 'Ectonucleotide pyrophosphatase/phosphodiesterase family member 2' |
| 13080 | 'ENPP2_No22' | 'CFFQGDHGFDNK' | 'Ectonucleotide pyrophosphatase/phosphodiesterase family member 2' |
| 13081 | 'ENPP2_No23' | 'VRDIEHLTSLDFFRK' | 'Ectonucleotide pyrophosphatase/phosphodiesterase family member 2' |
| 13082 | 'ENPP2_No24' | 'YGPFGPEMTNPLR' | 'Ectonucleotide pyrophosphatase/phosphodiesterase family member 2' |
| 13083 | 'ENPP2_No25' | 'YDAFLVTNMVPMYPAFK' | 'Ectonucleotide pyrophosphatase/phosphodiesterase family member 2' |
| 13084 | 'ENPP2_No26' | 'EIDKIVGQLMDGLK' | 'Ectonucleotide pyrophosphatase/phosphodiesterase family member 2' |
| 13085 | 'ENPP2_No27' | 'IVGQLMDGLK' | 'Ectonucleotide pyrophosphatase/phosphodiesterase family member 2' |
| 13086 | 'ENPP2_No28' | 'RVWNYFQR' | 'Ectonucleotide pyrophosphatase/phosphodiesterase family member 2' |
| 13087 | 'ENPP2_No29' | 'VSPSFSQNCLAYKNDK' | 'Ectonucleotide pyrophosphatase/phosphodiesterase family member 2' |
| 13088 | 'PKP1_No1' | 'SPNQNVQQAAAGALR' | 'Plakophilin-1' |
| 13089 | 'PKP1_No2' | 'LLLSDMWSSK' | 'Plakophilin-1' |
| 13090 | 'BLMH_No1' | 'IGPITPLEFYR' | 'Bleomycin hydrolase' |
| 13091 | 'BLMH_No2' | 'AQHVFQHAVPQEGKPITNQK' | 'Bleomycin hydrolase' |
| 13092 | 'BLMH_No3' | 'LYTVEYLSNMVGGR' | 'Bleomycin hydrolase' |
| 13093 | 'BLMH_No4' | 'LYTVEYLSNMVGGR' | 'Bleomycin hydrolase' |
| 13094 | 'BLMH_No5' | 'TLYNNQPIDFLK' | 'Bleomycin hydrolase' |
| 13095 | 'BLMH_No6' | 'TLYNNQPIDFLKK' | 'Bleomycin hydrolase' |
| 13096 | 'BLMH_No7' | 'ICLVNDPRPQHK' | 'Bleomycin hydrolase' |
| 13097 | 'BLMH_No8' | 'GEISATQDVMMEEIFR' | 'Bleomycin hydrolase' |
| 13098 | 'BLMH_No9' | 'LNSDPQFVLAQNVGTTHDLLDICLKR' | 'Bleomycin hydrolase' |
| 13099 | 'BLMH_No10' | 'KTLYNNQPIDFLK' | 'Bleomycin hydrolase' |
| 13100 | 'DAG1_No1' | 'VTIPTDLIASSGDIIK' | 'Dystroglycan' |
| 13101 | 'DAG1_No2' | 'LGCSLNQNSVPDIHGVEAPAR' | 'Dystroglycan' |
| 13102 | 'DAG1_No3' | 'EGAMSAQLGYPVVGWHIANK' | 'Dystroglycan' |
| 13103 | 'DAG1_No4' | 'SFSEVELHNMK' | 'Dystroglycan' |
| 13104 | 'DAG1_No5' | 'VVENGALLSWK' | 'Dystroglycan' |
| 13105 | 'DAG1_No6' | 'LFDMSAFMAGPGNAK' | 'Dystroglycan' |
| 13106 | 'DAG1_No7' | 'LFDMSAFMAGPGNAK' | 'Dystroglycan' |
| 13107 | 'DAG1_No8' | 'EGAMSAQLGYPVVGWHIANK' | 'Dystroglycan' |
| 13108 | 'DAG1_No9' | 'SFSEVELHNMK' | 'Dystroglycan' |
| 13109 | 'DSG2_No1' | 'IVSLEPAYPPVFYLNK' | 'Desmoglein-2' |
| 13110 | 'DSG2_No2' | 'AWITAPVALR' | 'Desmoglein-2' |
| 13111 | 'DSG2_No3' | 'GNNVEKPLELR' | 'Desmoglein-2' |
| 13112 | 'DSG2_No4' | 'IVAISEDYPR' | 'Desmoglein-2' |
| 13113 | 'DSG2_No5' | 'LEDRDNWISVDSVTSEIK' | 'Desmoglein-2' |

| | A | B | C |
|---|---|---|---|
| 1 | **Peptide Name** | **Peptide Sequence** | **Peptide Description** |
| 13114 | 'DSG2_No6' | 'SEIQFLISDNQGFSCPEK' | 'Desmoglein-2' |
| 13115 | 'DSG2_No7' | 'QVLTLTVCECLHGSGCR' | 'Desmoglein-2' |
| 13116 | 'DSG2_No8' | 'QESTSVLLQQSEKK' | 'Desmoglein-2' |
| 13117 | 'DSG2_No9' | 'GQIIGNFQAFDEDTGLPAHAR' | 'Desmoglein-2' |
| 13118 | 'DSG2_No10' | 'GITEPPFGIFVFNK' | 'Desmoglein-2' |
| 13119 | 'SEPTIN6_No1' | 'TAAELLQSQGSQAGGSQTLK' | 'Septin-6' |
| 13120 | 'SEPTIN6_No2' | 'TVPLAGHVGFDSLPDQLVNK' | 'Septin-6' |
| 13121 | 'GIT2' | 'LLSLGAQANFFHPEK' | 'ARF GTPase-activating protein GIT2' |
| 13122 | 'MLEC' | 'KEEEEEEEEYDEGSNLKK' | 'Malectin' |
| 13123 | 'FLNC_No1' | 'LKPGAPVR' | 'Filamin-C' |
| 13124 | 'FLNC_No2' | 'SPFEVQVSPEAGVQK' | 'Filamin-C' |
| 13125 | 'FLNC_No3' | 'EAGDGVFECEYYPVVPGK' | 'Filamin-C' |
| 13126 | 'FLNC_No4' | 'AKLDVQFAGTAK' | 'Filamin-C' |
| 13127 | 'SLBP' | 'LLINDFGR' | 'Histone RNA hairpin-binding protein' |
| 13128 | 'WFDC2' | 'DQCQVDSQCPGQMK' | 'WAP four-disulfide core domain protein 2' |
| 13129 | 'SPARCL1_No1' | 'VHAVDSCMSFQCK' | 'SPARC-like protein 1' |
| 13130 | 'SPARCL1_No2' | 'SIPTCTDFEVIQFPLR' | 'SPARC-like protein 1' |
| 13131 | 'SPARCL1_No3' | 'TGLEAISNHKETEEK' | 'SPARC-like protein 1' |
| 13132 | 'SPARCL1_No4' | 'DQGNQEQDPNISNGEEEEEKEPGEVGTHNDNQER' | 'SPARC-like protein 1' |
| 13133 | 'SPARCL1_No5' | 'ASLVPMEHCITR' | 'SPARC-like protein 1' |
| 13134 | 'SPARCL1_No6' | 'KGHQLQLDYFGACK' | 'SPARC-like protein 1' |
| 13135 | 'SPARCL1_No7' | 'VHENENIGTTEPGEHQEAK' | 'SPARC-like protein 1' |
| 13136 | 'SPARCL1_No8' | 'HSASDDYFIPSQAFLEAER' | 'SPARC-like protein 1' |
| 13137 | 'SPARCL1_No9' | 'HSQGLRDQGNQEQDPNISNGEEEEEKEPGEVGTHNDNQER' | 'SPARC-like protein 1' |
| 13138 | 'SPARCL1_No10' | 'VHAVDSCMSFQCK' | 'SPARC-like protein 1' |
| 13139 | 'SPARCL1_No11' | 'AQSIAYHLK' | 'SPARC-like protein 1' |
| 13140 | 'SPARCL1_No12' | 'NILMQLYEANSEHAGYLNEK' | 'SPARC-like protein 1' |
| 13141 | 'SPARCL1_No13' | 'TVSEALLMEPTDDGNTTPR' | 'SPARC-like protein 1' |
| 13142 | 'SPARCL1_No14' | 'LLAGDHPIDLLLR' | 'SPARC-like protein 1' |
| 13143 | 'SPARCL1_No15' | 'HIQETEWQSQEGK' | 'SPARC-like protein 1' |
| 13144 | 'SPARCL1_No16' | 'NILMQLYEANSEHAGYLNEK' | 'SPARC-like protein 1' |
| 13145 | 'SPARCL1_No17' | 'TVSEALLMEPTDDGNTTPR' | 'SPARC-like protein 1' |
| 13146 | 'SPARCL1_No18' | 'AEAEENEKETAVSTEDDSHHKAEK' | 'SPARC-like protein 1' |
| 13147 | 'SPARCL1_No19' | 'NHGVDDDGDDDGDDGGTDGPR' | 'SPARC-like protein 1' |
| 13148 | 'SPARCL1_No20' | 'ASLVPMEHCITR' | 'SPARC-like protein 1' |
| 13149 | 'SPARCL1_No21' | 'FFEECDPNKDKHITLK' | 'SPARC-like protein 1' |
| 13150 | 'SPARCL1_No22' | 'FFEECDPNKDK' | 'SPARC-like protein 1' |
| 13151 | 'SPARCL1_No23' | 'EKVHENENIGTTEPGEHQEAK' | 'SPARC-like protein 1' |
| 13152 | 'HABP2_No1' | 'FCEIGSDDCYVGDGYSYR' | 'Hyaluronan-binding protein 2' |
| 13153 | 'HABP2_No2' | 'LIANTLCNSR' | 'Hyaluronan-binding protein 2' |
| 13154 | 'HABP2_No3' | 'LPGFDSCGK' | 'Hyaluronan-binding protein 2' |
| 13155 | 'HABP2_No4' | 'GQCLITQSPPYYR' | 'Hyaluronan-binding protein 2' |
| 13156 | 'HABP2_No5' | 'TVCLPDGSFPSGSECHISGWGVTETGK' | 'Hyaluronan-binding protein 2' |
| 13157 | 'HABP2_No6' | 'PGQDTCQGDSGGPLTCEK' | 'Hyaluronan-binding protein 2' |

| | A | B | C |
|---|---|---|---|
| 1 | **Peptide Name** | **Peptide Sequence** | **Peptide Description** |
| 13158 | 'HABP2_No7' | 'YSHYNERDEIPHNDIALLK' | 'Hyaluronan-binding protein 2' |
| 13159 | 'HABP2_No8' | 'LKPVDGHCALESK' | 'Hyaluronan-binding protein 2' |
| 13160 | 'HABP2_No9' | 'VVLGDQDLK' | 'Hyaluronan-binding protein 2' |
| 13161 | 'HABP2_No10' | 'FTCACPDQFKGK' | 'Hyaluronan-binding protein 2' |
| 13162 | 'HABP2_No11' | 'GKFCEIGSDDCYVGDGYSYR' | 'Hyaluronan-binding protein 2' |
| 13163 | 'HABP2_No12' | 'VVLGDQDLKKEEFHEQSFR' | 'Hyaluronan-binding protein 2' |
| 13164 | 'HABP2_No13' | 'QLYDHMIDDSMICAGNLQKPGQDTCQGDSGGPLTCEK' | 'Hyaluronan-binding protein 2' |
| 13165 | 'HABP2_No14' | 'HPYTGPSCSQVVPVCR' | 'Hyaluronan-binding protein 2' |
| 13166 | 'HABP2_No15' | 'HPYTGPSCSQVVPVCRPNPCQNGATCSR' | 'Hyaluronan-binding protein 2' |
| 13167 | 'HABP2_No16' | 'NPDADEKPWCFIK' | 'Hyaluronan-binding protein 2' |
| 13168 | 'HABP2_No17' | 'DEIPHNDIALLK' | 'Hyaluronan-binding protein 2' |
| 13169 | 'HABP2_No18' | 'VVLGDQDLKK' | 'Hyaluronan-binding protein 2' |
| 13170 | 'HABP2_No19' | 'SKFTCACPDQFK' | 'Hyaluronan-binding protein 2' |
| 13171 | 'HABP2_No20' | 'VKLIANTLCNSR' | 'Hyaluronan-binding protein 2' |
| 13172 | 'HABP2_No21' | 'QLYDHMIDDSMICAGNLQK' | 'Hyaluronan-binding protein 2' |
| 13173 | 'HABP2_No22' | 'EEFHEQSFR' | 'Hyaluronan-binding protein 2' |
| 13174 | 'HABP2_No23' | 'SKFTCACPDQFKGK' | 'Hyaluronan-binding protein 2' |
| 13175 | 'HABP2_No24' | 'KEEFHEQSFR' | 'Hyaluronan-binding protein 2' |
| 13176 | 'HABP2_No25' | 'KEEFHEQSFRVEK' | 'Hyaluronan-binding protein 2' |
| 13177 | 'HABP2_No26' | 'QLYDHMIDDSMICAGNLQK' | 'Hyaluronan-binding protein 2' |
| 13178 | 'HABP2_No27' | 'QLYDHMIDDSMICAGNLQKPGQDTCQGDSGGPLTCEK' | 'Hyaluronan-binding protein 2' |
| 13179 | 'HABP2_No28' | 'QLYDHMIDDSMICAGNLQK' | 'Hyaluronan-binding protein 2' |
| 13180 | 'HABP2_No29' | 'QLYDHMIDDSMICAGNLQK' | 'Hyaluronan-binding protein 2' |
| 13181 | 'HABP2_No30' | 'QLYDHMIDDSMICAGNLQK' | 'Hyaluronan-binding protein 2' |
| 13182 | 'HABP2_No31' | 'TVCLPDGSFPSGSECHISGWGVTETGKGSR' | 'Hyaluronan-binding protein 2' |
| 13183 | 'HABP2_No32' | 'VQNTCKDNPCGR' | 'Hyaluronan-binding protein 2' |
| 13184 | 'HABP2_No33' | 'QLYDHMIDDSMICAGNLQKPGQDTCQGDSGGPLTCEK' | 'Hyaluronan-binding protein 2' |
| 13185 | 'HABP2_No34' | 'FTCACPDQFK' | 'Hyaluronan-binding protein 2' |
| 13186 | 'HABP2_No35' | 'FCEIGSDDCYVGDGYSYRGK' | 'Hyaluronan-binding protein 2' |
| 13187 | 'HABP2_No36' | 'EEFHEQSFRVEK' | 'Hyaluronan-binding protein 2' |
| 13188 | 'HABP2_No37' | 'LKPVDGHCALESKYVK' | 'Hyaluronan-binding protein 2' |
| 13189 | 'HABP2_No38' | 'QLYDHMIDDSMICAGNLQKPGQDTCQGDSGGPLTCEK' | 'Hyaluronan-binding protein 2' |
| 13190 | 'HABP2_No39' | 'IFKYSHYNERDEIPHNDIALLK' | 'Hyaluronan-binding protein 2' |
| 13191 | 'DSC3_No1' | 'VLNDGSVYTAR' | 'Desmocollin-3' |
| 13192 | 'DSC3_No2' | 'EVTVLLEHQK' | 'Desmocollin-3' |
| 13193 | 'DSC3_No3' | 'VILNVPSKLEADKIIGR' | 'Desmocollin-3' |
| 13194 | 'IHH_No1' | 'VAAVSTHVALGAYAPLTK' | 'Indian hedgehog protein' |
| 13195 | 'IHH_No2' | 'AFQVIETQDPPRR' | 'Indian hedgehog protein' |
| 13196 | 'IHH_No3' | 'LLLEEGSFHPLGMSGAGS' | 'Indian hedgehog protein' |
| 13197 | 'ITIH4_No1' | 'YIFHNFMER' | 'Inter-alpha-trypsin inhibitor heavy chain H4' |
| 13198 | 'ITIH4_No2' | 'AGFSWIEVTFK' | 'Inter-alpha-trypsin inhibitor heavy chain H4' |
| 13199 | 'ITIH4_No3' | 'WKETLFSVMPGLK' | 'Inter-alpha-trypsin inhibitor heavy chain H4' |
| 13200 | 'ITIH4_No4' | 'SPEQQETVLDGNLIIR' | 'Inter-alpha-trypsin inhibitor heavy chain H4' |
| 13201 | 'ITIH4_No5' | 'LQDRGPDVLTATVSGK' | 'Inter-alpha-trypsin inhibitor heavy chain H4' |

| | A | B | C |
|---|---|---|---|
| 1 | **Peptide Name** | **Peptide Sequence** | **Peptide Description** |
| 13202 | 'ITIH4_No6' | 'NMEQFQVSVSVAPNAK' | 'Inter-alpha-trypsin inhibitor heavy chain H4' |
| 13203 | 'ITIH4_No7' | 'LGVYELLLK' | 'Inter-alpha-trypsin inhibitor heavy chain H4' |
| 13204 | 'ITIH4_No8' | 'ANTVQEATFQMELPK' | 'Inter-alpha-trypsin inhibitor heavy chain H4' |
| 13205 | 'ITIH4_No9' | 'NGIDIYSLTVDSR' | 'Inter-alpha-trypsin inhibitor heavy chain H4' |
| 13206 | 'ITIH4_No10' | 'ITFELVYEELLKR' | 'Inter-alpha-trypsin inhibitor heavy chain H4' |
| 13207 | 'ITIH4_No11' | 'QGPVNLLSDPEQGVEVTGQYEREK' | 'Inter-alpha-trypsin inhibitor heavy chain H4' |
| 13208 | 'ITIH4_No12' | 'ETLFSVMPGLK' | 'Inter-alpha-trypsin inhibitor heavy chain H4' |
| 13209 | 'ITIH4_No13' | 'IPKPEASFSPR' | 'Inter-alpha-trypsin inhibitor heavy chain H4' |
| 13210 | 'ITIH4_No14' | 'ILDDLSPR' | 'Inter-alpha-trypsin inhibitor heavy chain H4' |
| 13211 | 'ITIH4_No15' | 'MNFRPGVLSSR' | 'Inter-alpha-trypsin inhibitor heavy chain H4' |
| 13212 | 'ITIH4_No16' | 'YIFHNFMER' | 'Inter-alpha-trypsin inhibitor heavy chain H4' |
| 13213 | 'ITIH4_No17' | 'RLDYQEGPPGVEISCWSVEL' | 'Inter-alpha-trypsin inhibitor heavy chain H4' |
| 13214 | 'ITIH4_No18' | 'QGPVNLLSDPEQGVEVTGQYER' | 'Inter-alpha-trypsin inhibitor heavy chain H4' |
| 13215 | 'ITIH4_No19' | 'PSLVPASAENVNK' | 'Inter-alpha-trypsin inhibitor heavy chain H4' |
| 13216 | 'ITIH4_No20' | 'MNFRPGVLSSR' | 'Inter-alpha-trypsin inhibitor heavy chain H4' |
| 13217 | 'ITIH4_No21' | 'TGLLLLSDPDK' | 'Inter-alpha-trypsin inhibitor heavy chain H4' |
| 13218 | 'ITIH4_No22' | 'NPLVWVHASPEHVVVTR' | 'Inter-alpha-trypsin inhibitor heavy chain H4' |
| 13219 | 'ITIH4_No23' | 'WKETLFSVMPGLK' | 'Inter-alpha-trypsin inhibitor heavy chain H4' |
| 13220 | 'ITIH4_No24' | 'QLGLPGPPDVPDHAAYHPFR' | 'Inter-alpha-trypsin inhibitor heavy chain H4' |
| 13221 | 'ITIH4_No25' | 'VTIGLLFWDGR' | 'Inter-alpha-trypsin inhibitor heavy chain H4' |
| 13222 | 'ITIH4_No26' | 'QLGLPGPPDVPDHAAYHPFRR' | 'Inter-alpha-trypsin inhibitor heavy chain H4' |
| 13223 | 'ITIH4_No27' | 'DTDRFSSHVGGTLGQFYQEVLWGSPAASDDGRR' | 'Inter-alpha-trypsin inhibitor heavy chain H4' |
| 13224 | 'ITIH4_No28' | 'FAHTVVTSR' | 'Inter-alpha-trypsin inhibitor heavy chain H4' |
| 13225 | 'ITIH4_No29' | 'EKAGFSWIEVTFK' | 'Inter-alpha-trypsin inhibitor heavy chain H4' |
| 13226 | 'ITIH4_No30' | 'LWAYLTIQQLLEQTVSASDADQQALR' | 'Inter-alpha-trypsin inhibitor heavy chain H4' |
| 13227 | 'ITIH4_No31' | 'FSSHVGGTLGQFYQEVLWGSPAASDDGR' | 'Inter-alpha-trypsin inhibitor heavy chain H4' |
| 13228 | 'ITIH4_No32' | 'VRPQQLVK' | 'Inter-alpha-trypsin inhibitor heavy chain H4' |
| 13229 | 'ITIH4_No33' | 'EKAEAQAQYSAAVAK' | 'Inter-alpha-trypsin inhibitor heavy chain H4' |
| 13230 | 'ITIH4_No34' | 'ITFELVYEELLK' | 'Inter-alpha-trypsin inhibitor heavy chain H4' |
| 13231 | 'ITIH4_No35' | 'ANTVQEATFQMELPKK' | 'Inter-alpha-trypsin inhibitor heavy chain H4' |
| 13232 | 'ITIH4_No36' | 'AGFSWIEVTFKNPLVWVHASPEHVVVTR' | 'Inter-alpha-trypsin inhibitor heavy chain H4' |
| 13233 | 'ITIH4_No37' | 'TGLLLLSDPDKVTIGLLFWDGR' | 'Inter-alpha-trypsin inhibitor heavy chain H4' |
| 13234 | 'ITIH4_No38' | 'QLGLPGPPDVPDHAAYHPFRR' | 'Inter-alpha-trypsin inhibitor heavy chain H4' |
| 13235 | 'ITIH4_No39' | 'GPDVLTATVSGK' | 'Inter-alpha-trypsin inhibitor heavy chain H4' |
| 13236 | 'ITIH4_No40' | 'ANTVQEATFQMELPKK' | 'Inter-alpha-trypsin inhibitor heavy chain H4' |
| 13237 | 'ITIH4_No41' | 'QGPVNLLSDPEQGVEVTGQYER' | 'Inter-alpha-trypsin inhibitor heavy chain H4' |
| 13238 | 'ITIH4_No42' | 'NMEQFQVSVSVAPNAK' | 'Inter-alpha-trypsin inhibitor heavy chain H4' |
| 13239 | 'ITIH4_No43' | 'SIQNNVR' | 'Inter-alpha-trypsin inhibitor heavy chain H4' |
| 13240 | 'ITIH4_No44' | 'NVVFVIDK' | 'Inter-alpha-trypsin inhibitor heavy chain H4' |
| 13241 | 'ITIH4_No45' | 'FSSHVGGTLGQFYQEVLWGSPAASDDGRR' | 'Inter-alpha-trypsin inhibitor heavy chain H4' |
| 13242 | 'ITIH4_No46' | 'EKAGFSWIEVTFKNPLVWVHASPEHVVVTR' | 'Inter-alpha-trypsin inhibitor heavy chain H4' |
| 13243 | 'ITIH4_No47' | 'LLFKGSEMVVAGK' | 'Inter-alpha-trypsin inhibitor heavy chain H4' |
| 13244 | 'ITIH4_No48' | 'FKPTLSQQQK' | 'Inter-alpha-trypsin inhibitor heavy chain H4' |
| 13245 | 'ITIH4_No49' | 'AISGGSIQIENGYFVHYFAPEGLTTMPK' | 'Inter-alpha-trypsin inhibitor heavy chain H4' |

| | A | B | C |
|---|---|---|---|
| 1 | **Peptide Name** | **Peptide Sequence** | **Peptide Description** |
| 13246 | 'ITIH4_No50' | 'SSAYKWKETLFSVMPGLK' | 'Inter-alpha-trypsin inhibitor heavy chain H4' |
| 13247 | 'ITIH4_No51' | 'HRQGPVNLLSDPEQGVEVTGQYEREK' | 'Inter-alpha-trypsin inhibitor heavy chain H4' |
| 13248 | 'ITIH4_No52' | 'QLGLPGPPDVPDHAAYHPFR' | 'Inter-alpha-trypsin inhibitor heavy chain H4' |
| 13249 | 'ITIH4_No53' | 'HRQGPVNLLSDPEQGVEVTGQYER' | 'Inter-alpha-trypsin inhibitor heavy chain H4' |
| 13250 | 'ITIH4_No54' | 'AISGGSIQIENGYFVHYFAPEGLTTMPK' | 'Inter-alpha-trypsin inhibitor heavy chain H4' |
| 13251 | 'ITIH4_No55' | 'RLGVYELLLK' | 'Inter-alpha-trypsin inhibitor heavy chain H4' |
| 13252 | 'ITIH4_No56' | 'QGPVNLLSDPEQGVEVTGQYEREK' | 'Inter-alpha-trypsin inhibitor heavy chain H4' |
| 13253 | 'ITIH4_No57' | 'LALDNGGLAR' | 'Inter-alpha-trypsin inhibitor heavy chain H4' |
| 13254 | 'ITIH4_No58' | 'AEAQAQYSAAVAK' | 'Inter-alpha-trypsin inhibitor heavy chain H4' |
| 13255 | 'ITIH4_No59' | 'ANTVQEATFQMELPK' | 'Inter-alpha-trypsin inhibitor heavy chain H4' |
| 13256 | 'ITIH4_No60' | 'LLFKGSEMVVAGK' | 'Inter-alpha-trypsin inhibitor heavy chain H4' |
| 13257 | 'ITIH4_No61' | 'NVVFVIDKSGSMSGR' | 'Inter-alpha-trypsin inhibitor heavy chain H4' |
| 13258 | 'ITIH4_No62' | 'DQFNLIVFSTEATQWRPSLVPASAENVNK' | 'Inter-alpha-trypsin inhibitor heavy chain H4' |
| 13259 | 'ITIH4_No63' | 'NVVFVIDKSGSMSGRK' | 'Inter-alpha-trypsin inhibitor heavy chain H4' |
| 13260 | 'ITIH4_No64' | 'ITFELVYEELLKRR' | 'Inter-alpha-trypsin inhibitor heavy chain H4' |
| 13261 | 'ITIH4_No65' | 'ERRLDYQEGPPGVEISCWSVEL' | 'Inter-alpha-trypsin inhibitor heavy chain H4' |
| 13262 | 'ITIH4_No66' | 'NVVFVIDKSGSMSGR' | 'Inter-alpha-trypsin inhibitor heavy chain H4' |
| 13263 | 'ITIH4_No67' | 'GSEMVVAGKLQDR' | 'Inter-alpha-trypsin inhibitor heavy chain H4' |
| 13264 | 'ITIH4_No68' | 'NVHSGSTFFK' | 'Inter-alpha-trypsin inhibitor heavy chain H4' |
| 13265 | 'ITIH4_No69' | 'YSLFCLGFGFDVSYAFLEK' | 'Inter-alpha-trypsin inhibitor heavy chain H4' |
| 13266 | 'ITIH4_No70' | 'LDYQEGPPGVEISCWSVEL' | 'Inter-alpha-trypsin inhibitor heavy chain H4' |
| 13267 | 'ITIH4_No71' | 'SSAYKWKETLFSVMPGLK' | 'Inter-alpha-trypsin inhibitor heavy chain H4' |
| 13268 | 'ITIH4_No72' | 'GSEMVVAGK' | 'Inter-alpha-trypsin inhibitor heavy chain H4' |
| 13269 | 'ITIH4_No73' | 'LAILPASAPPATSNPDPAVSR' | 'Inter-alpha-trypsin inhibitor heavy chain H4' |
| 13270 | 'ITIH4_No74' | 'AFITNFSMIIDGMTYPGIIK' | 'Inter-alpha-trypsin inhibitor heavy chain H4' |
| 13271 | 'ITIH4_No75' | 'RRLGVYELLLK' | 'Inter-alpha-trypsin inhibitor heavy chain H4' |
| 13272 | 'ITIH4_No76' | 'ETLFSVMPGLK' | 'Inter-alpha-trypsin inhibitor heavy chain H4' |
| 13273 | 'ITIH4_No77' | 'SFAAGIQALGGTNINDAMLMAVQLLDSSNQEER' | 'Inter-alpha-trypsin inhibitor heavy chain H4' |
| 13274 | 'ITIH4_No78' | 'NVVFVIDKSGSMSGRK' | 'Inter-alpha-trypsin inhibitor heavy chain H4' |
| 13275 | 'ITIH4_No79' | 'KAFITNFSMIIDGMTYPGIIK' | 'Inter-alpha-trypsin inhibitor heavy chain H4' |
| 13276 | 'ITIH4_No80' | 'VVNRANTVQEATFQMELPKK' | 'Inter-alpha-trypsin inhibitor heavy chain H4' |
| 13277 | 'ITIH4_No81' | 'FKPTLSQQQKSPEQQETVLDGNLIIR' | 'Inter-alpha-trypsin inhibitor heavy chain H4' |
| 13278 | 'ITIH4_No82' | 'VVNRANTVQEATFQMELPK' | 'Inter-alpha-trypsin inhibitor heavy chain H4' |
| 13279 | 'ITIH4_No83' | 'TLRVQGNDHSATR' | 'Inter-alpha-trypsin inhibitor heavy chain H4' |
| 13280 | 'ITIH4_No84' | 'LLFKGSEMVVAGKLQDR' | 'Inter-alpha-trypsin inhibitor heavy chain H4' |
| 13281 | 'ITIH4_No85' | 'PEASFSPR' | 'Inter-alpha-trypsin inhibitor heavy chain H4' |
| 13282 | 'ITIH4_No86' | 'FAHTVVTSRVVNR' | 'Inter-alpha-trypsin inhibitor heavy chain H4' |
| 13283 | 'ITIH4_No87' | 'NVHSGSTFFKYYLQGAK' | 'Inter-alpha-trypsin inhibitor heavy chain H4' |
| 13284 | 'ITIH4_No88' | 'VVNRANTVQEATFQMELPK' | 'Inter-alpha-trypsin inhibitor heavy chain H4' |
| 13285 | 'ITIH4_No89' | 'SIQNNVREAVSGR' | 'Inter-alpha-trypsin inhibitor heavy chain H4' |
| 13286 | 'ITIH4_No90' | 'GSEMVVAGK' | 'Inter-alpha-trypsin inhibitor heavy chain H4' |
| 13287 | 'ITIH4_No91' | 'ILDDLSPRDQFNLIVFSTEATQWRPSLVPASAENVNK' | 'Inter-alpha-trypsin inhibitor heavy chain H4' |
| 13288 | 'ITIH4_No92' | 'GSEMVVAGKLQDR' | 'Inter-alpha-trypsin inhibitor heavy chain H4' |
| 13289 | 'ITIH4_No93' | 'EALIKILDDLSPR' | 'Inter-alpha-trypsin inhibitor heavy chain H4' |

| | A | B | C |
|---|---|---|---|
| 1 | **Peptide Name** | **Peptide Sequence** | **Peptide Description** |
| 13290 | 'ITIH4_No94' | 'GSEMVVAGKLQDRGPDVLTATVSGK' | 'Inter-alpha-trypsin inhibitor heavy chain H4' |
| 13291 | 'ITIH4_No95' | 'PDDQEQSQVAEKPMEGESR' | 'Inter-alpha-trypsin inhibitor heavy chain H4' |
| 13292 | 'ITIH4_No96' | 'SPEQQETVLDGNLIIRYDVDR' | 'Inter-alpha-trypsin inhibitor heavy chain H4' |
| 13293 | 'ITIH4_No97' | 'PDDQEQSQVAEKPMEGESR' | 'Inter-alpha-trypsin inhibitor heavy chain H4' |
| 13294 | 'RASA3_No1' | 'LSEVITDTGVVCHK' | 'Ras GTPase-activating protein 3' |
| 13295 | 'RASA3_No2' | 'FGDEFLGELR' | 'Ras GTPase-activating protein 3' |
| 13296 | 'KIAA0100' | 'ILLYSSTLR' | 'Protein KIAA0100' |
| 13297 | 'CLINT1_No1' | 'HIHITQATETTTTR' | 'Clathrin interactor 1' |
| 13298 | 'CLINT1_No2' | 'EATNDDPWGPSGQLMGEIAK' | 'Clathrin interactor 1' |
| 13299 | 'GANAB_No1' | 'VLLVLELQGLQK' | 'Neutral alpha-glucosidase AB' |
| 13300 | 'GANAB_No2' | 'VSQGSKDPAEGDGAQPEETPR' | 'Neutral alpha-glucosidase AB' |
| 13301 | 'GANAB_No3' | 'LVAIVDPHIK' | 'Neutral alpha-glucosidase AB' |
| 13302 | 'GANAB_No4' | 'VVIIGAGKPAAVVLQTK' | 'Neutral alpha-glucosidase AB' |
| 13303 | 'GANAB_No5' | 'GLLEFEHQR' | 'Neutral alpha-glucosidase AB' |
| 13304 | 'GANAB_No6' | 'MMDYLQGSGETPQTDVR' | 'Neutral alpha-glucosidase AB' |
| 13305 | 'GANAB_No7' | 'LSFQHDPETSVLVLR' | 'Neutral alpha-glucosidase AB' |
| 13306 | 'GANAB_No8' | 'LDLLEDR' | 'Neutral alpha-glucosidase AB' |
| 13307 | 'GANAB_No9' | 'MMDYLQGSGETPQTDVR' | 'Neutral alpha-glucosidase AB' |
| 13308 | 'GANAB_No10' | 'FRIDELEPR' | 'Neutral alpha-glucosidase AB' |
| 13309 | 'GANAB_No11' | 'FSFSGNTLVSSSADPEGHFETPIWIER' | 'Neutral alpha-glucosidase AB' |
| 13310 | 'GANAB_No12' | 'DVHNIYGLYVHMATADGLR' | 'Neutral alpha-glucosidase AB' |
| 13311 | 'GANAB_No13' | 'FGAVWTGDNTAEWDHLK' | 'Neutral alpha-glucosidase AB' |
| 13312 | 'GANAB_No14' | 'KLVAIVDPHIK' | 'Neutral alpha-glucosidase AB' |
| 13313 | 'GANAB_No15' | 'REPWLLPSQHNDIIR' | 'Neutral alpha-glucosidase AB' |
| 13314 | 'GANAB_No16' | 'YFTWDPSRFPQPR' | 'Neutral alpha-glucosidase AB' |
| 13315 | 'GANAB_No17' | 'QYASLTGTQALPPLFSLGYHQSR' | 'Neutral alpha-glucosidase AB' |
| 13316 | 'GANAB_No18' | 'NPEPELLVR' | 'Neutral alpha-glucosidase AB' |
| 13317 | 'LTBP1_No1' | 'LCQIPVHGASVPK' | 'Latent-transforming growth factor beta-binding protein 1' |
| 13318 | 'LTBP1_No2' | 'HPPEASVQIHQVSR' | 'Latent-transforming growth factor beta-binding protein 1' |
| 13319 | 'LTBP1_No3' | 'VQEGYTCDCFDGYHLDTAK' | 'Latent-transforming growth factor beta-binding protein 1' |
| 13320 | 'LTBP1_No4' | 'MTCVDVNECDELNNR' | 'Latent-transforming growth factor beta-binding protein 1' |
| 13321 | 'LTBP1_No5' | 'DIDECQQGNLCVNGQCK' | 'Latent-transforming growth factor beta-binding protein 1' |
| 13322 | 'LTBP1_No6' | 'ASGLGDHCEDINECLEDK' | 'Latent-transforming growth factor beta-binding protein 1' |
| 13323 | 'LTBP1_No7' | 'CTCGQGYQLSAAKDQCEDIDECQHR' | 'Latent-transforming growth factor beta-binding protein 1' |
| 13324 | 'LTBP1_No8' | 'QEDCCGTVGTSWGFNK' | 'Latent-transforming growth factor beta-binding protein 1' |
| 13325 | 'LTBP1_No9' | 'TQTIHSTYSHQQVIPHVYPVAAK' | 'Latent-transforming growth factor beta-binding protein 1' |
| 13326 | 'LTBP1_No10' | 'KCVDIDECTQVQHLCSQGR' | 'Latent-transforming growth factor beta-binding protein 1' |
| 13327 | 'LTBP1_No11' | 'EICPGGMGYTVSGVHR' | 'Latent-transforming growth factor beta-binding protein 1' |
| 13328 | 'LTBP1_No12' | 'NGFCLNTRPGYECYCK' | 'Latent-transforming growth factor beta-binding protein 1' |
| 13329 | 'LTBP1_No13' | 'TSTDLDVDVDQPKEEK' | 'Latent-transforming growth factor beta-binding protein 1' |
| 13330 | 'LTBP1_No14' | 'MTCVDVNECDELNNR' | 'Latent-transforming growth factor beta-binding protein 1' |
| 13331 | 'LTBP1_No15' | 'ASGLGDHCEDINECLEDKSVCQR' | 'Latent-transforming growth factor beta-binding protein 1' |
| 13332 | 'LTBP1_No16' | 'DQCEDIDECQHR' | 'Latent-transforming growth factor beta-binding protein 1' |
| 13333 | 'LTBP1_No17' | 'TKEAQPGQSQVSYQGLPVQK' | 'Latent-transforming growth factor beta-binding protein 1' |

| | A | B | C |
|---|---|---|---|
| 1 | **Peptide Name** | **Peptide Sequence** | **Peptide Description** |
| 13334 | 'LTBP1_No18' | 'TSTDLDVDVDQPK' | 'Latent-transforming growth factor beta-binding protein 1' |
| 13335 | 'LTBP1_No19' | 'GFVPAGESSSEAGGENYKDADECLLFGQEICK' | 'Latent-transforming growth factor beta-binding protein 1' |
| 13336 | 'LTBP1_No20' | 'EICPGGMGYTVSGVHR' | 'Latent-transforming growth factor beta-binding protein 1' |
| 13337 | 'LTBP1_No21' | 'GISGEQSTEGSFPLR' | 'Latent-transforming growth factor beta-binding protein 1' |
| 13338 | 'LASP1_No1' | 'LKQQSELQSQVR' | 'LIM and SH3 domain protein 1' |
| 13339 | 'LASP1_No2' | 'QSFTMVADTPENLR' | 'LIM and SH3 domain protein 1' |
| 13340 | 'LASP1_No3' | 'QSFTMVADTPENLR' | 'LIM and SH3 domain protein 1' |
| 13341 | 'LASP1_No4' | 'VNCLDKFWHK' | 'LIM and SH3 domain protein 1' |
| 13342 | 'LASP1_No5' | 'QSFTMVADTPENLR' | 'LIM and SH3 domain protein 1' |
| 13343 | 'KPNB1_No1' | 'LAATNALLNSLEFTK' | 'Importin subunit beta-1' |
| 13344 | 'KPNB1_No2' | 'SSAYESLMEIVK' | 'Importin subunit beta-1' |
| 13345 | 'KPNB1_No3' | 'SDYDMVDYLNELR' | 'Importin subunit beta-1' |
| 13346 | 'SEPTIN2_No1' | 'LTVVDTPGYGDAINCR' | 'Septin-2' |
| 13347 | 'SEPTIN2_No2' | 'ASIPFSVVGSNQLIEAK' | 'Septin-2' |
| 13348 | 'SEPTIN2_No3' | 'LYPWGVVEVENPEHNDFLK' | 'Septin-2' |
| 13349 | 'SEPTIN2_No4' | 'ILDEIEEHNIK' | 'Septin-2' |
| 13350 | 'SEPTIN2_No5' | 'TIISYIDEQFER' | 'Septin-2' |
| 13351 | 'POSTN_No1' | 'AAAITSDILEALGR' | 'Periostin' |
| 13352 | 'POSTN_No2' | 'TTVLYECCPGYMR' | 'Periostin' |
| 13353 | 'POSTN_No3' | 'GCPAVLPIDHVYGTLGIVGATTTQR' | 'Periostin' |
| 13354 | 'POSTN_No4' | 'LILQNHILK' | 'Periostin' |
| 13355 | 'POSTN_No5' | 'VGLNELYNGQILETIGGK' | 'Periostin' |
| 13356 | 'POSTN_No6' | 'IIHGNQIATNGVVHVIDR' | 'Periostin' |
| 13357 | 'POSTN_No7' | 'GLESNVNVELLNALHSHMINKR' | 'Periostin' |
| 13358 | 'POSTN_No8' | 'GSFTYFAPSNEAWDNLDSDIRR' | 'Periostin' |
| 13359 | 'POSTN_No9' | 'LLYPADTPVGNDQLLEILNK' | 'Periostin' |
| 13360 | 'POSTN_No10' | 'DKNALQNIILYHLTPGVFIGK' | 'Periostin' |
| 13361 | 'POSTN_No11' | 'DGHFTLFAPTNEAFEK' | 'Periostin' |
| 13362 | 'POSTN_No12' | 'EGETITEVIHGEPIIK' | 'Periostin' |
| 13363 | 'POSTN_No13' | 'ISTGGGETEETLK' | 'Periostin' |
| 13364 | 'POSTN_No14' | 'IIDGVPVEITEKETR' | 'Periostin' |
| 13365 | 'POSTN_No15' | 'ELLTQPGDWTLFVPTNDAFK' | 'Periostin' |
| 13366 | 'POSTN_No16' | 'FIEGGDGHLFEDEEIKR' | 'Periostin' |
| 13367 | 'POSTN_No17' | 'GLESNVNVELLNALHSHMINK' | 'Periostin' |
| 13368 | 'PDIA6_No1' | 'VKLAAVDATVNQVLASR' | 'Protein disulfide-isomerase A6' |
| 13369 | 'PDIA6_No2' | 'GESPVDYDGGR' | 'Protein disulfide-isomerase A6' |
| 13370 | 'PDIA6_No3' | 'NSYLEVLLK' | 'Protein disulfide-isomerase A6' |
| 13371 | 'PDIA6_No4' | 'TGEAIVDAALSALR' | 'Protein disulfide-isomerase A6' |
| 13372 | 'PDIA6_No5' | 'IFQKGESPVDYDGGR' | 'Protein disulfide-isomerase A6' |
| 13373 | 'PDIA6_No6' | 'GSFSEQGINEFLR' | 'Protein disulfide-isomerase A6' |
| 13374 | 'PDIA6_No7' | 'NLEPEWAAAASEVKEQTK' | 'Protein disulfide-isomerase A6' |
| 13375 | 'PDIA6_No8' | 'GSTAPVGGGAFPTIVER' | 'Protein disulfide-isomerase A6' |
| 13376 | 'PCOLCE_No1' | 'TEESPSAPDAPTCPK' | 'Procollagen C-endopeptidase enhancer 1' |
| 13377 | 'PCOLCE_No2' | 'VFDLELHPACR' | 'Procollagen C-endopeptidase enhancer 1' |

| | A | B | C |
|---|---|---|---|
| 1 | **Peptide Name** | **Peptide Sequence** | **Peptide Description** |
| 13378 | 'PCOLCE_No3' | 'TGGLDLPSPPTGASLK' | 'Procollagen C-endopeptidase enhancer 1' |
| 13379 | 'PCOLCE_No4' | 'TGTLQSNFCASSLVVTATVK' | 'Procollagen C-endopeptidase enhancer 1' |
| 13380 | 'PCOLCE_No5' | 'GESGYVASEGFPNLYPPNK' | 'Procollagen C-endopeptidase enhancer 1' |
| 13381 | 'PCOLCE_No6' | 'SQPPEKTEESPSAPDAPTCPK' | 'Procollagen C-endopeptidase enhancer 1' |
| 13382 | 'PCOLCE_No7' | 'EPGEGLAVTVSLIGAYK' | 'Procollagen C-endopeptidase enhancer 1' |
| 13383 | 'PCOLCE_No8' | 'YDALEVFAGSGTSGQR' | 'Procollagen C-endopeptidase enhancer 1' |
| 13384 | 'PCOLCE_No9' | 'FDLEPDTYCR' | 'Procollagen C-endopeptidase enhancer 1' |
| 13385 | 'PCOLCE_No10' | 'YDSVSVFNGAVSDDSR' | 'Procollagen C-endopeptidase enhancer 1' |
| 13386 | 'PCOLCE_No11' | 'SMVREPGEGLAVTVSLIGAYK' | 'Procollagen C-endopeptidase enhancer 1' |
| 13387 | 'PCOLCE_No12' | 'ATSGTEHQFCGGR' | 'Procollagen C-endopeptidase enhancer 1' |
| 13388 | 'PCOLCE_No13' | 'GVSYLLMGQVEENR' | 'Procollagen C-endopeptidase enhancer 1' |
| 13389 | 'PCOLCE_No14' | 'GFLLWYSGR' | 'Procollagen C-endopeptidase enhancer 1' |
| 13390 | 'PLEC_No1' | 'SSIAGLLLK' | 'Plectin' |
| 13391 | 'PLEC_No2' | 'APVPASELLASGVLSR' | 'Plectin' |
| 13392 | 'PLEC_No3' | 'AYSDPSTGEPATYGELQQR' | 'Plectin' |
| 13393 | 'PLEC_No4' | 'LGFHLPLEVAYQR' | 'Plectin' |
| 13394 | 'PON3_No1' | 'LLNYNPEDPPGSEVLR' | 'Serum paraoxonase/lactonase 3' |
| 13395 | 'PON3_No2' | 'IFLMDLNEQNPR' | 'Serum paraoxonase/lactonase 3' |
| 13396 | 'PON3_No3' | 'IQNVLSEKPR' | 'Serum paraoxonase/lactonase 3' |
| 13397 | 'PON3_No4' | 'GFCSANGITVSADQK' | 'Serum paraoxonase/lactonase 3' |
| 13398 | 'PON3_No5' | 'STVEIFKFEEQQR' | 'Serum paraoxonase/lactonase 3' |
| 13399 | 'PON3_No6' | 'HDNWDLTQLK' | 'Serum paraoxonase/lactonase 3' |
| 13400 | 'PON3_No7' | 'YPGMPNFAPDEPGK' | 'Serum paraoxonase/lactonase 3' |
| 13401 | 'PON3_No8' | 'YPGMPNFAPDEPGK' | 'Serum paraoxonase/lactonase 3' |
| 13402 | 'PON3_No9' | 'NIHIMEKHDNWDLTQLK' | 'Serum paraoxonase/lactonase 3' |
| 13403 | 'PON3_No10' | 'VIQLGTLVDNLTVDPATGDILAGCHPNPMK' | 'Serum paraoxonase/lactonase 3' |
| 13404 | 'PON3_No11' | 'ELFNPHGISIFIDK' | 'Serum paraoxonase/lactonase 3' |
| 13405 | 'PON3_No12' | 'VIQLGTLVDNLTVDPATGDILAGCHPNPMK' | 'Serum paraoxonase/lactonase 3' |
| 13406 | 'PON3_No13' | 'IFLMDLNEQNPR' | 'Serum paraoxonase/lactonase 3' |
| 13407 | 'NECTIN1_No1' | 'VEFLRPSFTDGTIR' | 'Nectin-1' |
| 13408 | 'NECTIN1_No2' | 'LKGEAEYQEIR' | 'Nectin-1' |
| 13409 | 'KRT31_No1' | 'QNQEYQVLLDVR' | 'Keratin, type I cuticular Ha1' |
| 13410 | 'KRT31_No2' | 'QVVSSSEQLQSYQAEIIELRR' | 'Keratin, type I cuticular Ha1' |
| 13411 | 'KRT31_No3' | 'QLVESDINGLRR' | 'Keratin, type I cuticular Ha1' |
| 13412 | 'KRT31_No4' | 'SQQQEPLLCPSYQSYFK' | 'Keratin, type I cuticular Ha1' |
| 13413 | 'KRT31_No5' | 'SQYEALVETNRR' | 'Keratin, type I cuticular Ha1' |
| 13414 | 'KRT31_No6' | 'TVNALEIELQAQHNLR' | 'Keratin, type I cuticular Ha1' |
| 13415 | 'PCBP1_No1' | 'IITLTGPTNAIFK' | 'Poly(rC)-binding protein 1' |
| 13416 | 'PCBP1_No2' | 'LVVPATQCGSLIGK' | 'Poly(rC)-binding protein 1' |
| 13417 | 'PCBP2_No1' | 'INISEGNCPER' | 'Poly(rC)-binding protein 2' |
| 13418 | 'PCBP2_No2' | 'IITLAGPTNAIFK' | 'Poly(rC)-binding protein 2' |
| 13419 | 'PCBP2_No3' | 'ESTGAQVQVAGDMLPNSTER' | 'Poly(rC)-binding protein 2' |
| 13420 | 'PCBP2_No4' | 'ESTGAQVQVAGDMLPNSTER' | 'Poly(rC)-binding protein 2' |
| 13421 | 'PCBP2_No5' | 'AITIAGIPQSIIECVK' | 'Poly(rC)-binding protein 2' |

| | A | B | C |
|---|---|---|---|
| 1 | **Peptide Name** | **Peptide Sequence** | **Peptide Description** |
| 13422 | 'PCBP2_No6' | 'LVVPASQCGSLIGK' | 'Poly(rC)-binding protein 2' |
| 13423 | 'RSU1_No1' | 'EIGELTQLKELHIQGNR' | 'Ras suppressor protein 1' |
| 13424 | 'RSU1_No2' | 'LQILSLR' | 'Ras suppressor protein 1' |
| 13425 | 'RSU1_No3' | 'LTVLPPELGNLDLTGQK' | 'Ras suppressor protein 1' |
| 13426 | 'RSU1_No4' | 'ALYLSDNDFEILPPDIGK' | 'Ras suppressor protein 1' |
| 13427 | 'RSU1_No5' | 'LTMVPPNIAELK' | 'Ras suppressor protein 1' |
| 13428 | 'RSU1_No6' | 'LQILSLRDNDLISLPK' | 'Ras suppressor protein 1' |
| 13429 | 'RSU1_No7' | 'NQPEVDMSDR' | 'Ras suppressor protein 1' |
| 13430 | 'RSU1_No8' | 'DNDLISLPK' | 'Ras suppressor protein 1' |
| 13431 | 'FCN2_No1' | 'LGEFWLGNDNIHALTAQGTSELR' | 'Ficolin-2' |
| 13432 | 'FCN2_No2' | 'GTHGSFANGINWK' | 'Ficolin-2' |
| 13433 | 'FCN2_No3' | 'MVGLEGSDKLTILR' | 'Ficolin-2' |
| 13434 | 'FCN2_No4' | 'VDLVDFEDNYQFAK' | 'Ficolin-2' |
| 13435 | 'FCN2_No5' | 'RVDGSVDFYRDWATYK' | 'Ficolin-2' |
| 13436 | 'FCN2_No6' | 'MVGLEGSDKLTILR' | 'Ficolin-2' |
| 13437 | 'FCN2_No7' | 'VDGSVDFYRDWATYK' | 'Ficolin-2' |
| 13438 | 'FCN2_No8' | 'VDGSVDFYR' | 'Ficolin-2' |
| 13439 | 'FCN2_No9' | 'GHFLSGWHTIYLPDCR' | 'Ficolin-2' |
| 13440 | 'CDSN' | 'GSPGVPSFAAGPPISEGK' | 'Corneodesmosin' |
| 13441 | 'MAPRE2_No1' | 'FYDANYDGKEYDPVEAR' | 'Microtubule-associated protein RP/EB family member 2' |
| 13442 | 'MAPRE2_No2' | 'LEHEYIHNFK' | 'Microtubule-associated protein RP/EB family member 2' |
| 13443 | 'MAPRE2_No3' | 'SDKDLETQVIQLNEQVHSLK' | 'Microtubule-associated protein RP/EB family member 2' |
| 13444 | 'MAPRE2_No4' | 'LALEGVEKERDFYFGK' | 'Microtubule-associated protein RP/EB family member 2' |
| 13445 | 'TGFBI_No1' | 'LTLLAPLNSVFK' | 'Transforming growth factor-beta-induced protein ig-h3' |
| 13446 | 'TGFBI_No2' | 'SPYQLVLQHSR' | 'Transforming growth factor-beta-induced protein ig-h3' |
| 13447 | 'TGFBI_No3' | 'EGVYTVFAPTNEAFR' | 'Transforming growth factor-beta-induced protein ig-h3' |
| 13448 | 'TGFBI_No4' | 'STVISYECCPGYEK' | 'Transforming growth factor-beta-induced protein ig-h3' |
| 13449 | 'TGFBI_No5' | 'YLYHGQTLETLGGK' | 'Transforming growth factor-beta-induced protein ig-h3' |
| 13450 | 'TGFBI_No6' | 'ILGDPEALR' | 'Transforming growth factor-beta-induced protein ig-h3' |
| 13451 | 'TGFBI_No7' | 'ILGDPEALRDLLNNHILK' | 'Transforming growth factor-beta-induced protein ig-h3' |
| 13452 | 'TGFBI_No8' | 'ADHHATNGVVHLIDK' | 'Transforming growth factor-beta-induced protein ig-h3' |
| 13453 | 'TGFBI_No9' | 'NSLCIENSCIAAHDKR' | 'Transforming growth factor-beta-induced protein ig-h3' |
| 13454 | 'TGFBI_No10' | 'GDELADSALEIFK' | 'Transforming growth factor-beta-induced protein ig-h3' |
| 13455 | 'TGFBI_No11' | 'YHIGDEILVSGGIGALVR' | 'Transforming growth factor-beta-induced protein ig-h3' |
| 13456 | 'TGFBI_No12' | 'VLTPPMGTVMDVLK' | 'Transforming growth factor-beta-induced protein ig-h3' |
| 13457 | 'TGFBI_No13' | 'VLTPPMGTVMDVLK' | 'Transforming growth factor-beta-induced protein ig-h3' |
| 13458 | 'TGFBI_No14' | 'YLYHGQTLETLGGKK' | 'Transforming growth factor-beta-induced protein ig-h3' |
| 13459 | 'TGFBI_No15' | 'VLTPPMGTVMDVLK' | 'Transforming growth factor-beta-induced protein ig-h3' |
| 13460 | 'TGFBI_No16' | 'SLQGDKLEVSLK' | 'Transforming growth factor-beta-induced protein ig-h3' |
| 13461 | 'TGFBI_No17' | 'STVISYECCPGYEKVPGEK' | 'Transforming growth factor-beta-induced protein ig-h3' |
| 13462 | 'TGFBI_No18' | 'YGTLFTMDR' | 'Transforming growth factor-beta-induced protein ig-h3' |
| 13463 | 'TGFBI_No19' | 'VLTPPMGTVMDVLK' | 'Transforming growth factor-beta-induced protein ig-h3' |
| 13464 | 'TGFBI_No20' | 'LLGDAKELANILK' | 'Transforming growth factor-beta-induced protein ig-h3' |
| 13465 | 'TGFBI_No21' | 'FSMLVAAIQSAGLTETLNR' | 'Transforming growth factor-beta-induced protein ig-h3' |

| | A | B | C |
|---|---|---|---|
| 1 | **Peptide Name** | **Peptide Sequence** | **Peptide Description** |
| 13466 | 'TGFBI_No22' | 'LTLLAPLNSVFKDGTPPIDAHTR' | 'Transforming growth factor-beta-induced protein ig-h3' |
| 13467 | 'TGFBI_No23' | 'LKSLQGDKLEVSLK' | 'Transforming growth factor-beta-induced protein ig-h3' |
| 13468 | 'TGFBI_No24' | 'GCPAALPLSNLYETLGVVGSTTTQLYTDR' | 'Transforming growth factor-beta-induced protein ig-h3' |
| 13469 | 'TGFBI_No25' | 'NSLCIENSCIAAHDK' | 'Transforming growth factor-beta-induced protein ig-h3' |
| 13470 | 'TGFBI_No26' | 'DLLNNHILK' | 'Transforming growth factor-beta-induced protein ig-h3' |
| 13471 | 'TGFBI_No27' | 'AAVAASGLNTMLEGNGQYTLLAPTNEAFEKIPSETLNR' | 'Transforming growth factor-beta-induced protein ig-h3' |
| 13472 | 'TGFBI_No28' | 'SRLLGDAKELANILK' | 'Transforming growth factor-beta-induced protein ig-h3' |
| 13473 | 'TGFBI_No29' | 'QHGPNVCAVQK' | 'Transforming growth factor-beta-induced protein ig-h3' |
| 13474 | 'TGFBI_No30' | 'NHIIKDQLASK' | 'Transforming growth factor-beta-induced protein ig-h3' |
| 13475 | 'TGFBI_No31' | 'GRYGTLFTMDR' | 'Transforming growth factor-beta-induced protein ig-h3' |
| 13476 | 'TGFBI_No32' | 'GCPAALPLSNLYETLGVVGSTTTQLYTDRTEK' | 'Transforming growth factor-beta-induced protein ig-h3' |
| 13477 | 'TGFBI_No33' | 'FSMLVAAIQSAGLTETLNR' | 'Transforming growth factor-beta-induced protein ig-h3' |
| 13478 | 'TGFBI_No34' | 'QAGLGNHLSGSER' | 'Transforming growth factor-beta-induced protein ig-h3' |
| 13479 | 'MAPRE1_No1' | 'QGQETAVAPSLVAPALNKPK' | 'Microtubule-associated protein RP/EB family member 1' |
| 13480 | 'MAPRE1_No2' | 'FFDANYDGKDYDPVAAR' | 'Microtubule-associated protein RP/EB family member 1' |
| 13481 | 'MAPRE1_No3' | 'AVNVYSTSVTSDNLSR' | 'Microtubule-associated protein RP/EB family member 1' |
| 13482 | 'CD226_No1' | 'QIDLLTYCNLVHGR' | 'CD226 antigen' |
| 13483 | 'CD226_No2' | 'QIDLLTYCNLVHGR' | 'CD226 antigen' |
| 13484 | 'CD226_No3' | 'IGTQQDSIAIFSPTHGMVIR' | 'CD226 antigen' |
| 13485 | 'CST6_No1' | 'DLSPDDPQVQK' | 'Cystatin-M' |
| 13486 | 'CST6_No2' | 'AAQAAVASYNMGSNSIYYFR' | 'Cystatin-M' |
| 13487 | 'CST6_No3' | 'AAQAAVASYNMGSNSIYYFR' | 'Cystatin-M' |
| 13488 | 'CST6_No4' | 'VTGDHVDLTTCPLAAGAQQEK' | 'Cystatin-M' |
| 13489 | 'STXBP2_No1' | 'ADTPSLGEGPEK' | 'Syntaxin-binding protein 2' |
| 13490 | 'STXBP2_No2' | 'AAHIFFTDTCPEPLFSELGR' | 'Syntaxin-binding protein 2' |
| 13491 | 'STXBP2_No3' | 'LIQHANVQAHSSLIR' | 'Syntaxin-binding protein 2' |
| 13492 | 'STXBP2_No4' | 'KGPEDTAQLAHAVLAK' | 'Syntaxin-binding protein 2' |
| 13493 | 'STXBP2_No5' | 'LIVPVLLDAAVPAYDKIR' | 'Syntaxin-binding protein 2' |
| 13494 | 'STXBP2_No6' | 'YSTHLHLADDCMK' | 'Syntaxin-binding protein 2' |
| 13495 | 'STXBP2_No7' | 'ATEGKWEVLIGSSHILTPTR' | 'Syntaxin-binding protein 2' |
| 13496 | 'STXBP2_No8' | 'REPIPSLEAIYLLSPTEK' | 'Syntaxin-binding protein 2' |
| 13497 | 'STXBP2_No9' | 'NLWPFVSDPAPTASSQAAVSAR' | 'Syntaxin-binding protein 2' |
| 13498 | 'ADIPOQ_No1' | 'IFYNQQNHYDGSTGK' | 'Adiponectin' |
| 13499 | 'ADIPOQ_No2' | 'SAFSVGLETYVTIPNMPIR' | 'Adiponectin' |
| 13500 | 'ADIPOQ_No3' | 'GDIGETGVPGAEGPR' | 'Adiponectin' |
| 13501 | 'ADIPOQ_No4' | 'SAFSVGLETYVTIPNMPIR' | 'Adiponectin' |
| 13502 | 'ADIPOQ_No5' | 'NGLYADNDNDSTFTGFLLYHDTN' | 'Adiponectin' |
| 13503 | 'RAB11B_No1' | 'GAVGALLVYDIAK' | 'Ras-related protein Rab-11B' |
| 13504 | 'RAB11B_No2' | 'GTRDDEYDYLFK' | 'Ras-related protein Rab-11B' |
| 13505 | 'RAB11B_No3' | 'FTRNEFNLESK' | 'Ras-related protein Rab-11B' |
| 13506 | 'RAB11B_No4' | 'NNLSFIETSALDSTNVEEAFK' | 'Ras-related protein Rab-11B' |
| 13507 | 'ZYX_No1' | 'LGHPEALSAGTGSPQPPSFTYAQQR' | 'Zyxin' |
| 13508 | 'ZYX_No2' | 'SPGAPGPLTLK' | 'Zyxin' |
| 13509 | 'ZYX_No3' | 'AYHPHCFTCVVCAR' | 'Zyxin' |

| | A | B | C |
|---|---|---|---|
| 1 | **Peptide Name** | **Peptide Sequence** | **Peptide Description** |
| 13510 | 'ZYX_No4' | 'FSPGAPGGSGSQPNQK' | 'Zyxin' |
| 13511 | 'ZYX_No5' | 'VSSGYVPPPVATPFSSK' | 'Zyxin' |
| 13512 | 'ZYX_No6' | 'PLEGTSFIVDQANRPHCVPDYHK' | 'Zyxin' |
| 13513 | 'ZYX_No7' | 'PSPAISVSVSAPAFYAPQK' | 'Zyxin' |
| 13514 | 'ZYX_No8' | 'CEDCGKPLSIEADDNGCFPLDGHVLCR' | 'Zyxin' |
| 13515 | 'ZYX_No9' | 'CNTCGEPITDR' | 'Zyxin' |
| 13516 | 'ZYX_No10' | 'VVALDKNFHMK' | 'Zyxin' |
| 13517 | 'ZYX_No11' | 'PSPAISVSVSAPAFYAPQKK' | 'Zyxin' |
| 13518 | 'ZYX_No12' | 'SSTKPAAGGTAPLPPWK' | 'Zyxin' |
| 13519 | 'ZYX_No13' | 'QNVAVNELCGR' | 'Zyxin' |
| 13520 | 'SEPTIN7_No1' | 'NLEGYVGFANLPNQVYR' | 'Septin-7' |
| 13521 | 'SEPTIN7_No2' | 'DVTNNVHYENYR' | 'Septin-7' |
| 13522 | 'SEPTIN7_No3' | 'STLINSLFLTDLYSPEYPGPSHR' | 'Septin-7' |
| 13523 | 'SEPTIN7_No4' | 'ADTLTPEECQQFKK' | 'Septin-7' |
| 13524 | 'SEPTIN7_No5' | 'THMQDLKDVTNNVHYENYR' | 'Septin-7' |
| 13525 | 'IGFBP7_No1' | 'AGAAAGGPGVSGVCVCK' | 'Insulin-like growth factor-binding protein 7' |
| 13526 | 'IGFBP7_No2' | 'SRYPVCGSDGTTYPSGCQLR' | 'Insulin-like growth factor-binding protein 7' |
| 13527 | 'IGFBP7_No3' | 'ITVVDALHEIPVK' | 'Insulin-like growth factor-binding protein 7' |
| 13528 | 'IGFBP7_No4' | 'EDAGEYECHASNSQGQASASAK' | 'Insulin-like growth factor-binding protein 7' |
| 13529 | 'IGFBP7_No5' | 'TELLPGDRDNLAIQTR' | 'Insulin-like growth factor-binding protein 7' |
| 13530 | 'IGFBP7_No6' | 'ITVVDALHEIPVKKGEGAEL' | 'Insulin-like growth factor-binding protein 7' |
| 13531 | 'IGFBP7_No7' | 'ITVVDALHEIPVKK' | 'Insulin-like growth factor-binding protein 7' |
| 13532 | 'IGFBP7_No8' | 'YPVCGSDGTTYPSGCQLR' | 'Insulin-like growth factor-binding protein 7' |
| 13533 | 'IGFBP7_No9' | 'GGPEKHEVTGWVLVSPLSK' | 'Insulin-like growth factor-binding protein 7' |
| 13534 | 'EXT1_No1' | 'LLLQIPSTIR' | 'Exostosin-1' |
| 13535 | 'EXT1_No2' | 'IAESYQNILAAIEGSR' | 'Exostosin-1' |
| 13536 | 'EXT1_No3' | 'ASISTENFRPNFDVSIPLFSK' | 'Exostosin-1' |
| 13537 | 'EXT1_No4' | 'LDPVLFKDQVSILR' | 'Exostosin-1' |
| 13538 | 'EXT1_No5' | 'IVLTTLEIIQDR' | 'Exostosin-1' |
| 13539 | 'EXT1_No6' | 'NALYHVHNGEDVVLLTTCK' | 'Exostosin-1' |
| 13540 | 'SYPL1' | 'APNIYLVR' | 'Synaptophysin-like protein 1' |
| 13541 | 'ECM1_No1' | 'ACPSHQPDISSGLELPFPPGVPTLDNIK' | 'Extracellular matrix protein 1' |
| 13542 | 'ECM1_No2' | 'ELLALIQLER' | 'Extracellular matrix protein 1' |
| 13543 | 'ECM1_No3' | 'EVGPPLPQEAVPLQK' | 'Extracellular matrix protein 1' |
| 13544 | 'ECM1_No4' | 'LDGFPPGRPSPDNLNQICLPNR' | 'Extracellular matrix protein 1' |
| 13545 | 'ECM1_No5' | 'AWEDTLDKYCDR' | 'Extracellular matrix protein 1' |
| 13546 | 'ECM1_No6' | 'HPPSPTRDECFAR' | 'Extracellular matrix protein 1' |
| 13547 | 'ECM1_No7' | 'DPALCCYLSPGDEQVNCFNINYLR' | 'Extracellular matrix protein 1' |
| 13548 | 'ECM1_No8' | 'NVALVSGDTENAK' | 'Extracellular matrix protein 1' |
| 13549 | 'ECM1_No9' | 'LTFINDLCGPR' | 'Extracellular matrix protein 1' |
| 13550 | 'ECM1_No10' | 'QGETLNFLEIGYSR' | 'Extracellular matrix protein 1' |
| 13551 | 'ECM1_No11' | 'LLPAQLPAEKEVGPPLPQEAVPLQK' | 'Extracellular matrix protein 1' |
| 13552 | 'ECM1_No12' | 'QHVVYGPWNLPQSSYSHLTR' | 'Extracellular matrix protein 1' |
| 13553 | 'ECM1_No13' | 'VTPNLMGHLCGNQR' | 'Extracellular matrix protein 1' |

| | A | B | C |
|---|---|---|---|
| 1 | **Peptide Name** | **Peptide Sequence** | **Peptide Description** |
| 13554 | 'ECM1_No14' | 'LVWEEAMSR' | 'Extracellular matrix protein 1' |
| 13555 | 'ECM1_No15' | 'RAPYPNYDRDILTIDIGR' | 'Extracellular matrix protein 1' |
| 13556 | 'ECM1_No16' | 'AWEDTLDKYCDREYAVK' | 'Extracellular matrix protein 1' |
| 13557 | 'ECM1_No17' | 'ELPSLQHPNEQK' | 'Extracellular matrix protein 1' |
| 13558 | 'ECM1_No18' | 'VTPNLMGHLCGNQR' | 'Extracellular matrix protein 1' |
| 13559 | 'ECM1_No19' | 'CCDLPFPEQACCAEEEK' | 'Extracellular matrix protein 1' |
| 13560 | 'ECM1_No20' | 'FCEAEFSVK' | 'Extracellular matrix protein 1' |
| 13561 | 'ECM1_No21' | 'FSCFQEEAPQPHYQLR' | 'Extracellular matrix protein 1' |
| 13562 | 'ECM1_No22' | 'QHVVYGPWNLPQSSYSHLTR' | 'Extracellular matrix protein 1' |
| 13563 | 'ECM1_No23' | 'APYPNYDRDILTIDIGR' | 'Extracellular matrix protein 1' |
| 13564 | 'ECM1_No24' | 'DILTIDIGR' | 'Extracellular matrix protein 1' |
| 13565 | 'ECM1_No25' | 'ELLALIQLEREFQR' | 'Extracellular matrix protein 1' |
| 13566 | 'ECM1_No26' | 'CCDLPFPEQACCAEEEKLTFINDLCGPRR' | 'Extracellular matrix protein 1' |
| 13567 | 'ECM1_No27' | 'LVWEEAMSR' | 'Extracellular matrix protein 1' |
| 13568 | 'ECM1_No28' | 'CCDLPFPEQACCAEEEKLTFINDLCGPR' | 'Extracellular matrix protein 1' |
| 13569 | 'ECM1_No29' | 'NIWRDPALCCYLSPGDEQVNCFNINYLR' | 'Extracellular matrix protein 1' |
| 13570 | 'ECM1_No30' | 'TRPHWCCTR' | 'Extracellular matrix protein 1' |
| 13571 | 'ECM1_No31' | 'PSPDNLNQICLPNR' | 'Extracellular matrix protein 1' |
| 13572 | 'ECM1_No32' | 'GQGEQGSTGGTNISSTSEPKEE' | 'Extracellular matrix protein 1' |
| 13573 | 'CCL14_No1' | 'GPYHPSECCFTYTTYK' | 'C-C motif chemokine 14' |
| 13574 | 'CCL14_No2' | 'IMDYYETNSQCSKPGIVFITK' | 'C-C motif chemokine 14' |
| 13575 | 'CCL14_No3' | 'GHSVCTNPSDKWVQDYIK' | 'C-C motif chemokine 14' |
| 13576 | 'CCL14_No4' | 'IMDYYETNSQCSKPGIVFITK' | 'C-C motif chemokine 14' |
| 13577 | 'CCL14_No5' | 'PGIVFITK' | 'C-C motif chemokine 14' |
| 13578 | 'DBN1_No1' | 'LREDENAEPVGTTYQK' | 'Drebrin' |
| 13579 | 'DBN1_No2' | 'SPSDSSTASTPVAEQIER' | 'Drebrin' |
| 13580 | 'DBN1_No3' | 'KSESEVEEAAAIIAQRPDNPR' | 'Drebrin' |
| 13581 | 'DBN1_No4' | 'LAASGEGGLQELSGHFENQK' | 'Drebrin' |
| 13582 | 'MAN2A1_No1' | 'VLLAPLGDDFR' | 'Alpha-mannosidase 2' |
| 13583 | 'MAN2A1_No2' | 'VGNGHSNEAALILHR' | 'Alpha-mannosidase 2' |
| 13584 | 'MAN2A1_No3' | 'IYSEVTCFFDHVTHR' | 'Alpha-mannosidase 2' |
| 13585 | 'MAN2A1_No4' | 'SGWAIDPFGHSPTMAYLLNR' | 'Alpha-mannosidase 2' |
| 13586 | 'MAN2A1_No5' | 'WWDIIDIQK' | 'Alpha-mannosidase 2' |
| 13587 | 'MAN2A1_No6' | 'IIGNSAFLLILK' | 'Alpha-mannosidase 2' |
| 13588 | 'MAN2A1_No7' | 'SGAYLFLPDGNAKPYVYTTPPFVR' | 'Alpha-mannosidase 2' |
| 13589 | 'MAN2A1_No8' | 'FLSSSLYTALTEAR' | 'Alpha-mannosidase 2' |
| 13590 | 'MAN2A1_No9' | 'LPLQANVYPMTTMAYIQDAK' | 'Alpha-mannosidase 2' |
| 13591 | 'MAN2A1_No10' | 'FIWSEISYLSK' | 'Alpha-mannosidase 2' |
| 13592 | 'MAN2A1_No11' | 'DWVVVDYGTR' | 'Alpha-mannosidase 2' |
| 13593 | 'MAN2A1_No12' | 'TLEFFWR' | 'Alpha-mannosidase 2' |
| 13594 | 'MAN2A1_No13' | 'ILESASSNSHLADYVLYK' | 'Alpha-mannosidase 2' |
| 13595 | 'MAN2A1_No14' | 'FYTDLNGYQIQPR' | 'Alpha-mannosidase 2' |
| 13596 | 'MAN2A1_No15' | 'NLGLFQHHDAITGTAK' | 'Alpha-mannosidase 2' |
| 13597 | 'MAN2A1_No16' | 'LYHIQGIEGQSVEVSNIVDIR' | 'Alpha-mannosidase 2' |

| | A | B | C |
|---|---|---|---|
| 1 | **Peptide Name** | **Peptide Sequence** | **Peptide Description** |
| 13598 | 'MAN2A1_No17' | 'FGCPWGVPPETIHPGNVQSR' | 'Alpha-mannosidase 2' |
| 13599 | 'MAN2A1_No18' | 'LLAENNEIISNIR' | 'Alpha-mannosidase 2' |
| 13600 | 'MAN2A1_No19' | 'MLLDQYR' | 'Alpha-mannosidase 2' |
| 13601 | 'MAN2A1_No20' | 'DKTQYIFNNMVLK' | 'Alpha-mannosidase 2' |
| 13602 | 'MAN2A1_No21' | 'LPLQANVYPMTTMAYIQDAK' | 'Alpha-mannosidase 2' |
| 13603 | 'MAN2A1_No22' | 'DKTQYIFNNMVLK' | 'Alpha-mannosidase 2' |
| 13604 | 'MAN2A1_No23' | 'ICCQFDFK' | 'Alpha-mannosidase 2' |
| 13605 | 'MAN2A1_No24' | 'WWDIIDIQKK' | 'Alpha-mannosidase 2' |
| 13606 | 'MAN2A1_No25' | 'NYQQLFDYMNSQSK' | 'Alpha-mannosidase 2' |
| 13607 | 'MAN2A1_No26' | 'GLEQGIQDNKITANLFR' | 'Alpha-mannosidase 2' |
| 13608 | 'MAN2A1_No27' | 'ITANLFR' | 'Alpha-mannosidase 2' |
| 13609 | 'MAN2A1_No28' | 'RNLGLFQHHDAITGTAK' | 'Alpha-mannosidase 2' |
| 13610 | 'MAN2A1_No29' | 'SGWAIDPFGHSPTMAYLLNR' | 'Alpha-mannosidase 2' |
| 13611 | 'QPCT_No1' | 'SFSNIISTLNPTAK' | 'Glutaminyl-peptide cyclotransferase' |
| 13612 | 'QPCT_No2' | 'YPGSPGSYAAR' | 'Glutaminyl-peptide cyclotransferase' |
| 13613 | 'QPCT_No3' | 'VFVGATDSAVPCAMMLELAR' | 'Glutaminyl-peptide cyclotransferase' |
| 13614 | 'QPCT_No4' | 'LQAIEHELHELGLLK' | 'Glutaminyl-peptide cyclotransferase' |
| 13615 | 'UGP2_No1' | 'ILTTASSHEFEHTKK' | 'UTP--glucose-1-phosphate uridylyltransferase' |
| 13616 | 'UGP2_No2' | 'NENTFLDLTVQQIEHLNK' | 'UTP--glucose-1-phosphate uridylyltransferase' |
| 13617 | 'AOC3_No1' | 'LGPGLVDAAQAR' | 'Membrane primary amine oxidase' |
| 13618 | 'AOC3_No2' | 'ELPQASGLLHHCCFYK' | 'Membrane primary amine oxidase' |
| 13619 | 'AOC3_No3' | 'YQLAVTQR' | 'Membrane primary amine oxidase' |
| 13620 | 'AOC3_No4' | 'DAFCVFEQNQGLPLRR' | 'Membrane primary amine oxidase' |
| 13621 | 'AOC3_No5' | 'EALAIVFFGR' | 'Membrane primary amine oxidase' |
| 13622 | 'AOC3_No6' | 'YGNQVSEHTLGTVHTHSAHFK' | 'Membrane primary amine oxidase' |
| 13623 | 'AOC3_No7' | 'PSDNCVFSVELQLPPK' | 'Membrane primary amine oxidase' |
| 13624 | 'AOC3_No8' | 'DAFCVFEQNQGLPLR' | 'Membrane primary amine oxidase' |
| 13625 | 'AOC3_No9' | 'KLLEMEEQAAFLVGSATPR' | 'Membrane primary amine oxidase' |
| 13626 | 'INF2_No1' | 'ATEPVATSNPAGDPVGSTR' | 'Inverted formin-2' |
| 13627 | 'INF2_No2' | 'ALDELFEAIEQK' | 'Inverted formin-2' |
| 13628 | 'INF2_No3' | 'LLPEKHEIENLR' | 'Inverted formin-2' |
| 13629 | 'INF2_No4' | 'AVLLASDAQECTLEEVVER' | 'Inverted formin-2' |
| 13630 | 'INF2_No5' | 'ISDALLQLTCVSCVR' | 'Inverted formin-2' |
| 13631 | 'INF2_No6' | 'QLPIFCQLILR' | 'Inverted formin-2' |
| 13632 | 'INF2_No7' | 'DLEQPSQAAGINLEIIR' | 'Inverted formin-2' |
| 13633 | 'INF2_No8' | 'CPASEPGLDATTASESR' | 'Inverted formin-2' |
| 13634 | 'TKFC_No1' | 'VAPAEPQEAPDSTAAGGSASK' | 'Triokinase/FMN cyclase' |
| 13635 | 'TKFC_No2' | 'VAGALAEAGVGLEEIAK' | 'Triokinase/FMN cyclase' |
| 13636 | 'TKFC_No3' | 'SPGADLLQVLTK' | 'Triokinase/FMN cyclase' |
| 13637 | 'TKFC_No4' | 'VAPAEPQEAPDSTAAGGSASKR' | 'Triokinase/FMN cyclase' |
| 13638 | 'TKFC_No5' | 'LSVLLLEK' | 'Triokinase/FMN cyclase' |
| 13639 | 'KRT71_No1' | 'FLEQQNQVLETK' | 'Keratin, type II cytoskeletal 71' |
| 13640 | 'KRT71_No2' | 'NLDLDSIIDEVR' | 'Keratin, type II cytoskeletal 71' |
| 13641 | 'LGALSL_No1' | 'IQTLSAIDTIK' | 'Galectin-related protein' |

| | A | B | C |
|---|---|---|---|
| 1 | **Peptide Name** | **Peptide Sequence** | **Peptide Description** |
| 13642 | 'LGALSL_No2' | 'VFVDGHQLFDFYHR' | 'Galectin-related protein' |
| 13643 | 'LGALSL_No3' | 'AGSVADSDAVVKLDDGHLNNSLSSPVQADVYFPR' | 'Galectin-related protein' |
| 13644 | 'SVEP1_No1' | 'AFCLACPFYGTTPFAGSR' | 'Sushi, von Willebrand factor type A, EGF and pentraxin domain-containing protein 1' |
| 13645 | 'SVEP1_No2' | 'ISCGPPAHVENAIAR' | 'Sushi, von Willebrand factor type A, EGF and pentraxin domain-containing protein 1' |
| 13646 | 'SVEP1_No3' | 'EVTFHCHEGYILHGAPK' | 'Sushi, von Willebrand factor type A, EGF and pentraxin domain-containing protein 1' |
| 13647 | 'SVEP1_No4' | 'GGQWNPSPMSIQCIPVR' | 'Sushi, von Willebrand factor type A, EGF and pentraxin domain-containing protein 1' |
| 13648 | 'SVEP1_No5' | 'GAFQQAAQILLHAR' | 'Sushi, von Willebrand factor type A, EGF and pentraxin domain-containing protein 1' |
| 13649 | 'SVEP1_No6' | 'ITCLESGEWNHLIPYCK' | 'Sushi, von Willebrand factor type A, EGF and pentraxin domain-containing protein 1' |
| 13650 | 'SVEP1_No7' | 'SGYVIQGSSDLICTEK' | 'Sushi, von Willebrand factor type A, EGF and pentraxin domain-containing protein 1' |
| 13651 | 'SVEP1_No8' | 'QGFILSGVK' | 'Sushi, von Willebrand factor type A, EGF and pentraxin domain-containing protein 1' |
| 13652 | 'SVEP1_No9' | 'YSCVGGFFLR' | 'Sushi, von Willebrand factor type A, EGF and pentraxin domain-containing protein 1' |
| 13653 | 'SVEP1_No10' | 'EIEYTCNEGFLLEGAR' | 'Sushi, von Willebrand factor type A, EGF and pentraxin domain-containing protein 1' |
| 13654 | 'SVEP1_No11' | 'TTGPNVVYSCNR' | 'Sushi, von Willebrand factor type A, EGF and pentraxin domain-containing protein 1' |
| 13655 | 'SVEP1_No12' | 'SVGSPVFVCQANR' | 'Sushi, von Willebrand factor type A, EGF and pentraxin domain-containing protein 1' |
| 13656 | 'SVEP1_No13' | 'MVPSFCSDAEDIDCR' | 'Sushi, von Willebrand factor type A, EGF and pentraxin domain-containing protein 1' |
| 13657 | 'SVEP1_No14' | 'EGYTLAGLDTIECLADGK' | 'Sushi, von Willebrand factor type A, EGF and pentraxin domain-containing protein 1' |
| 13658 | 'SVEP1_No15' | 'IGSYQDEEGQLECK' | 'Sushi, von Willebrand factor type A, EGF and pentraxin domain-containing protein 1' |
| 13659 | 'SVEP1_No16' | 'GGQWNPSPMSIQCIPVR' | 'Sushi, von Willebrand factor type A, EGF and pentraxin domain-containing protein 1' |
| 13660 | 'SVEP1_No17' | 'FGTICYVSCR' | 'Sushi, von Willebrand factor type A, EGF and pentraxin domain-containing protein 1' |
| 13661 | 'SVEP1_No18' | 'CIAHFCEKPPSVSYSILESVSK' | 'Sushi, von Willebrand factor type A, EGF and pentraxin domain-containing protein 1' |
| 13662 | 'SVEP1_No19' | 'VIDAEPPVIDWCR' | 'Sushi, von Willebrand factor type A, EGF and pentraxin domain-containing protein 1' |
| 13663 | 'ACTBL2' | 'TTGIVMDSGDGVTHIVPIYEGYALPHAILR' | 'Beta-actin-like protein 2' |
| 13664 | 'FLG2_No1' | 'SVVTVIDVFYK' | 'Filaggrin-2' |
| 13665 | 'FLG2_No2' | 'HQEEESETEEDEEDTPGHK' | 'Filaggrin-2' |
| 13666 | 'FLG2_No3' | 'QSSYGQHGSGSSQSSGYGQYGSR' | 'Filaggrin-2' |
| 13667 | 'FLG2_No4' | 'ESGEEYESGSGSNSWER' | 'Filaggrin-2' |
| 13668 | 'FLG2_No5' | 'SSQSNYGQHGSGSSQSSGYGQHGSSSGQTTGFGQHR' | 'Filaggrin-2' |
| 13669 | 'FLG2_No6' | 'SGSGQSSGFGQHGSGSGQSSGFGQHESGSGK' | 'Filaggrin-2' |
| 13670 | 'FLG2_No7' | 'SHQSSYGQHGSGSSQSSGYGQHGSSSGQTFGFGQHR' | 'Filaggrin-2' |
| 13671 | 'FLG2_No8' | 'SGQSSYGQHSSGSSQSSGYGQHGSR' | 'Filaggrin-2' |
| 13672 | 'FLG2_No9' | 'SHQSSYGQHGSGSSQSSGYGQHGSSSGQTSGFGQHR' | 'Filaggrin-2' |
| 13673 | 'FLG2_No10' | 'HGSYGPAEYDYGHTGYGPSGGSR' | 'Filaggrin-2' |
| 13674 | 'WDR44_No1' | 'LLASAGQDNVVR' | 'WD repeat-containing protein 44' |
| 13675 | 'WDR44_No2' | 'DFAAVEEVAPAKPPR' | 'WD repeat-containing protein 44' |
| 13676 | 'WDR44_No3' | 'HLTPEPDIVASTK' | 'WD repeat-containing protein 44' |
| 13677 | 'LY6G6F' | 'DASIPQFKPEIQVYENIHLAR' | 'Lymphocyte antigen 6 complex locus protein G6f' |
| 13678 | 'KPRP_No1' | 'RLDQCPESPLQR' | 'Keratinocyte proline-rich protein' |
| 13679 | 'KPRP_No2' | 'IEISSPCCPR' | 'Keratinocyte proline-rich protein' |
| 13680 | 'PEAR1_No1' | 'FGQDCAETCDCAPDAR' | 'Platelet endothelial aggregation receptor 1' |
| 13681 | 'PEAR1_No2' | 'LQCCHGFYESR' | 'Platelet endothelial aggregation receptor 1' |
| 13682 | 'PEAR1_No3' | 'CSCENAIACSPIDGECVCK' | 'Platelet endothelial aggregation receptor 1' |
| 13683 | 'DMKN' | 'VGEAAHALGNTGHEIGR' | 'Dermokine' |
| 13684 | 'VASN_No1' | 'LAGLGLQQLDEGLFSR' | 'Vasorin' |
| 13685 | 'VASN_No2' | 'HIQPGAFDTLDR' | 'Vasorin' |

| | A | B | C |
|---|---|---|---|
| 1 | **Peptide Name** | **Peptide Sequence** | **Peptide Description** |
| 13686 | 'VASN_No3' | 'NLHDLDVSDNQLER' | 'Vasorin' |
| 13687 | 'VASN_No4' | 'SLTLGIEPVSPTSLR' | 'Vasorin' |
| 13688 | 'VASN_No5' | 'ESHVTLASPEETR' | 'Vasorin' |
| 13689 | 'VASN_No6' | 'YLQGSSVQLR' | 'Vasorin' |
| 13690 | 'VASN_No7' | 'IRHIQPGAFDTLDR' | 'Vasorin' |
| 13691 | 'VASN_No8' | 'LRNLHDLDVSDNQLER' | 'Vasorin' |
| 13692 | 'PM20D1' | 'NPLTNAIIR' | 'N-fatty-acyl-amino acid synthase/hydrolase PM20D1' |
| 13693 | 'TWF2_No1' | 'AVLPLLDAQQPCYLLYR' | 'Twinfilin-2' |
| 13694 | 'TWF2_No2' | 'KIEIGDGAELTAEFLYDEVHPK' | 'Twinfilin-2' |
| 13695 | 'TWF2_No3' | 'HLSSCAAPAPLTSAER' | 'Twinfilin-2' |
| 13696 | 'TWF2_No4' | 'LLDSVEQDFHLEIAK' | 'Twinfilin-2' |
| 13697 | 'TWF2_No5' | 'VVIEDEQLVLGASQEPVGR' | 'Twinfilin-2' |
| 13698 | 'TWF2_No6' | 'ETIELVHTEPTDVAQLPSR' | 'Twinfilin-2' |
| 13699 | 'ERAP2_No1' | 'PAETPTQIQEMFDEVSYNK' | 'Endoplasmic reticulum aminopeptidase 2' |
| 13700 | 'ERAP2_No2' | 'MAFPCFDEPLFK' | 'Endoplasmic reticulum aminopeptidase 2' |
| 13701 | 'ERAP2_No3' | 'TQNLAALLHAIAR' | 'Endoplasmic reticulum aminopeptidase 2' |
| 13702 | 'ERAP2_No4' | 'VGLIHDVFQLVGAGR' | 'Endoplasmic reticulum aminopeptidase 2' |
| 13703 | 'ERAP2_No5' | 'KFDLGSYDIR' | 'Endoplasmic reticulum aminopeptidase 2' |
| 13704 | 'ERAP2_No6' | 'VLSYPAHEQIALLVPEK' | 'Endoplasmic reticulum aminopeptidase 2' |
| 13705 | 'ERAP2_No7' | 'LACDLNHAPCIQK' | 'Endoplasmic reticulum aminopeptidase 2' |
| 13706 | 'APOA5_No1' | 'QDTYLQIAAFTR' | 'Apolipoprotein A-V' |
| 13707 | 'APOA5_No2' | 'AFAGTGTEEGAGPDPQMLSEEVR' | 'Apolipoprotein A-V' |
| 13708 | 'APOA5_No3' | 'IQQNLDQLREELSR' | 'Apolipoprotein A-V' |
| 13709 | 'APOA5_No4' | 'LPQDPVGMR' | 'Apolipoprotein A-V' |
| 13710 | 'APOA5_No5' | 'AFAGTGTEEGAGPDPQMLSEEVR' | 'Apolipoprotein A-V' |
| 13711 | 'APOA5_No6' | 'ELFHPYAESLVSGIGR' | 'Apolipoprotein A-V' |
| 13712 | 'APOA5_No7' | 'FKELFHPYAESLVSGIGR' | 'Apolipoprotein A-V' |
| 13713 | 'APOA5_No8' | 'KGFWDYFSQTSGDK' | 'Apolipoprotein A-V' |
| 13714 | 'APOA5_No9' | 'RQLQEELEEVK' | 'Apolipoprotein A-V' |
| 13715 | 'APOA5_No10' | 'EPATLKDSLEQDLNNMNK' | 'Apolipoprotein A-V' |
| 13716 | 'APOA5_No11' | 'QQLKPYTMDLMEQVALR' | 'Apolipoprotein A-V' |
| 13717 | 'APOA5_No12' | 'QDTYLQIAAFTR' | 'Apolipoprotein A-V' |
| 13718 | 'APOA5_No13' | 'GFWDYFSQTSGDK' | 'Apolipoprotein A-V' |
| 13719 | 'CSPG4_No1' | 'VAIQPVNDHAPVQTISR' | 'Chondroitin sulfate proteoglycan 4' |
| 13720 | 'CSPG4_No2' | 'ITVAALDASNLLASVPSPQR' | 'Chondroitin sulfate proteoglycan 4' |
| 13721 | 'SBSN_No1' | 'FGQGVHHAAGQAGNEAGR' | 'Suprabasin' |
| 13722 | 'SBSN_No2' | 'FGQGVDNAAGQAGNEAGR' | 'Suprabasin' |
| 13723 | 'SBSN_No3' | 'ALDGINSGITHAGR' | 'Suprabasin' |
| 13724 | 'SBSN_No4' | 'LGQGVNHAADQAGKEVEK' | 'Suprabasin' |
| 13725 | 'SBSN_No5' | 'EAEKFGQGAHHAAGQAGNEAGR' | 'Suprabasin' |
| 13726 | 'SBSN_No6' | 'FGQGAHHGLSEGWKETEK' | 'Suprabasin' |
| 13727 | 'SBSN_No7' | 'EADKLIHHGVHHGANQAGSEAGK' | 'Suprabasin' |
| 13728 | 'SBSN_No8' | 'EAGQFGQGVHHTLEQAGK' | 'Suprabasin' |
| 13729 | 'SBSN_No9' | 'FGQGVHHAASQVGKEEDR' | 'Suprabasin' |

| | A | B | C |
|---|---|---|---|
| 1 | **Peptide Name** | **Peptide Sequence** | **Peptide Description** |
| 13730 | 'SBSN_No10' | 'VAHEINHGIGQAGKEAEK' | 'Suprabasin' |
| 13731 | 'SBSN_No11' | 'FGQGVHHGLSEGWKETEK' | 'Suprabasin' |
| 13732 | 'PLXDC2_No1' | 'VGLSDAFVVVHR' | 'Plexin domain-containing protein 2' |
| 13733 | 'PLXDC2_No2' | 'HRQDWVDSGCPEESKEK' | 'Plexin domain-containing protein 2' |
| 13734 | 'PLXDC2_No3' | 'EIPVLVTQISSTNHPVK' | 'Plexin domain-containing protein 2' |
| 13735 | 'PLXDC2_No4' | 'DLWVNIDQMEKDK' | 'Plexin domain-containing protein 2' |
| 13736 | 'PLXDC2_No5' | 'MLTATQYIAPLMANFDPSVSR' | 'Plexin domain-containing protein 2' |
| 13737 | 'PLXDC2_No6' | 'DLWVNIDQMEKDKVK' | 'Plexin domain-containing protein 2' |
| 13738 | 'PLXDC2_No7' | 'HRQDWVDSGCPEESK' | 'Plexin domain-containing protein 2' |
| 13739 | 'PLXDC2_No8' | 'DLWVNIDQMEKDK' | 'Plexin domain-containing protein 2' |
| 13740 | 'PLXDC2_No9' | 'DLWVNIDQMEK' | 'Plexin domain-containing protein 2' |
| 13741 | 'PLXDC2_No10' | 'DLWVNIDQMEKDKVK' | 'Plexin domain-containing protein 2' |
| 13742 | 'PLXDC2_No11' | 'ASVGQDSPEPR' | 'Plexin domain-containing protein 2' |
| 13743 | 'PI16_No1' | 'RPYQEGTPCSQCPSGYHCK' | 'Peptidase inhibitor 16' |
| 13744 | 'PI16_No2' | 'LMVELHNLYR' | 'Peptidase inhibitor 16' |
| 13745 | 'PI16_No3' | 'AQVSPTASDMLHMR' | 'Peptidase inhibitor 16' |
| 13746 | 'PI16_No4' | 'GKRPYQEGTPCSQCPSGYHCK' | 'Peptidase inhibitor 16' |
| 13747 | 'PI16_No5' | 'LQGVEETNIELLVCNYEPPGNVK' | 'Peptidase inhibitor 16' |
| 13748 | 'PI16_No6' | 'GENLFAITDEGMDVPLAMEEWHHER' | 'Peptidase inhibitor 16' |
| 13749 | 'PI16_No7' | 'LMVELHNLYR' | 'Peptidase inhibitor 16' |
| 13750 | 'PI16_No8' | 'AQVSPTASDMLHMR' | 'Peptidase inhibitor 16' |
| 13751 | 'PI16_No9' | 'AQVSPTASDMLHMR' | 'Peptidase inhibitor 16' |
| 13752 | 'PI16_No10' | 'AQVSPTASDMLHMR' | 'Peptidase inhibitor 16' |
| 13753 | 'PI16_No11' | 'WDEELAAFAK' | 'Peptidase inhibitor 16' |
| 13754 | 'ADAMTSL4_No1' | 'LQIAQLRPSSNYLALR' | 'ADAMTS-like protein 4' |
| 13755 | 'ADAMTSL4_No2' | 'ILWIPAGALR' | 'ADAMTS-like protein 4' |
| 13756 | 'ADAMTSL4_No3' | 'VQDGTLCQPGAPDICVAGR' | 'ADAMTS-like protein 4' |
| 13757 | 'ADAMTSL4_No4' | 'RPDGCGVCGGDDSTCR' | 'ADAMTS-like protein 4' |
| 13758 | 'ADAMTSL4_No5' | 'RPSSQGWASPQVAGR' | 'ADAMTS-like protein 4' |
| 13759 | 'ADAMTSL4_No6' | 'ACSQAPCPPEQPDPR' | 'ADAMTS-like protein 4' |
| 13760 | 'ADAMTSL4_No7' | 'HTEKVQDGTLCQPGAPDICVAGR' | 'ADAMTS-like protein 4' |
| 13761 | 'ADAMTSL4_No8' | 'GVWRPIFLCISR' | 'ADAMTS-like protein 4' |
| 13762 | 'ADAMTSL4_No9' | 'RPDPFPSVPR' | 'ADAMTS-like protein 4' |
| 13763 | 'ADAMTSL4_No10' | 'DSSPHCPLVVQAR' | 'ADAMTS-like protein 4' |
| 13764 | 'CD109_No1' | 'TLTLPSLPLNSADEIYELR' | 'CD109 antigen' |
| 13765 | 'CD109_No2' | 'IPVQLVFK' | 'CD109 antigen' |
| 13766 | 'CD109_No3' | 'ISVTQPDSIVGIVAVDK' | 'CD109 antigen' |
| 13767 | 'CD109_No4' | 'TNIQVTVTGPSSPSPVK' | 'CD109 antigen' |
| 13768 | 'CD109_No5' | 'NSLGGFASTQDTTVALK' | 'CD109 antigen' |
| 13769 | 'CD109_No6' | 'IVTLFSDFKPYK' | 'CD109 antigen' |
| 13770 | 'CD109_No7' | 'VGSPFELVVSGNK' | 'CD109 antigen' |
| 13771 | 'CD109_No8' | 'TSLNILIKDPK' | 'CD109 antigen' |
| 13772 | 'CD109_No9' | 'SYSQSILLDLTDNR' | 'CD109 antigen' |
| 13773 | 'CD109_No10' | 'IEFPILEDSSELQLK' | 'CD109 antigen' |

| | A | B | C |
|---|---|---|---|
| 1 | **Peptide Name** | **Peptide Sequence** | **Peptide Description** |
| 13774 | 'CD109_No11' | 'FMEENEGHIVDIHDFSLGSSPHVR' | 'CD109 antigen' |
| 13775 | 'CD109_No12' | 'TLSFSFPPNTVTGSER' | 'CD109 antigen' |
| 13776 | 'CD109_No13' | 'VGSPFELVVSGNKR' | 'CD109 antigen' |
| 13777 | 'CD109_No14' | 'LKELSYMVVSR' | 'CD109 antigen' |
| 13778 | 'CD109_No15' | 'FMEENEGHIVDIHDFSLGSSPHVR' | 'CD109 antigen' |
| 13779 | 'CD109_No16' | 'LKELSYMVVSR' | 'CD109 antigen' |
| 13780 | 'CD109_No17' | 'SNLIQQWLSQQSDLGVISK' | 'CD109 antigen' |
| 13781 | 'CD109_No18' | 'AEQEGGMQFWVSSESK' | 'CD109 antigen' |
| 13782 | 'CD109_No19' | 'CFLEADPYIDIDQNVLHR' | 'CD109 antigen' |
| 13783 | 'CD109_No20' | 'VSNTQDASVSIVDYYEPR' | 'CD109 antigen' |
| 13784 | 'CD109_No21' | 'ALSEFAALMNTER' | 'CD109 antigen' |
| 13785 | 'UNC13D_No1' | 'ALLYEDALYTVLHR' | 'Protein unc-13 homolog D' |
| 13786 | 'UNC13D_No2' | 'AVQMDELVPLGELTK' | 'Protein unc-13 homolog D' |
| 13787 | 'UNC13D_No3' | 'WFQPAIPSWLQK' | 'Protein unc-13 homolog D' |
| 13788 | 'UNC13D_No4' | 'HSTSAVDLSTCFAQISHTAR' | 'Protein unc-13 homolog D' |
| 13789 | 'UNC13D_No5' | 'IQQQAETTSEELGAVTVK' | 'Protein unc-13 homolog D' |
| 13790 | 'TUBA1A_No1' | 'VGINYQPPTVVPGGDLAK' | 'Tubulin alpha-1A chain' |
| 13791 | 'TUBA1A_No2' | 'AYHEQLSVAEITNACFEPANQMVK' | 'Tubulin alpha-1A chain' |
| 13792 | 'TUBA1A_No3' | 'EIIDLVLDR' | 'Tubulin alpha-1A chain' |
| 13793 | 'TUBA1A_No4' | 'NLDIERPTYTNLNR' | 'Tubulin alpha-1A chain' |
| 13794 | 'TUBA1A_No5' | 'TIGGGDDSFNTFFSETGAGK' | 'Tubulin alpha-1A chain' |
| 13795 | 'TUBA1A_No6' | 'IHFPLATYAPVISAEK' | 'Tubulin alpha-1A chain' |
| 13796 | 'TUBA1A_No7' | 'AVFVDLEPTVIDEVR' | 'Tubulin alpha-1A chain' |
| 13797 | 'TUBA1A_No8' | 'LDHKFDLMYAK' | 'Tubulin alpha-1A chain' |
| 13798 | 'TUBA1A_No9' | 'QLFHPEQLITGKEDAANNYAR' | 'Tubulin alpha-1A chain' |
| 13799 | 'TUBA1A_No10' | 'YMACCLLYR' | 'Tubulin alpha-1A chain' |
| 13800 | 'TUBA1A_No11' | 'AVCMLSNTTAIAEAWAR' | 'Tubulin alpha-1A chain' |
| 13801 | 'TUBA1A_No12' | 'FDLMYAK' | 'Tubulin alpha-1A chain' |
| 13802 | 'TUBA1A_No13' | 'YMACCLLYR' | 'Tubulin alpha-1A chain' |
| 13803 | 'TUBA1A_No14' | 'AVCMLSNTTAIAEAWAR' | 'Tubulin alpha-1A chain' |
| 13804 | 'TUBA1A_No15' | 'QLFHPEQLITGKEDAANNYAR' | 'Tubulin alpha-1A chain' |
| 13805 | 'TUBA1A_No16' | 'TIQFVDWCPTGFK' | 'Tubulin alpha-1A chain' |
| 13806 | 'TUBA1A_No17' | 'FDLMYAK' | 'Tubulin alpha-1A chain' |
| 13807 | 'TUBA1A_No18' | 'LIGQIVSSITASLR' | 'Tubulin alpha-1A chain' |
| 13808 | 'TUBA1A_No19' | 'RNLDIERPTYTNLNR' | 'Tubulin alpha-1A chain' |
| 13809 | 'TUBA1A_No20' | 'LDHKFDLMYAK' | 'Tubulin alpha-1A chain' |
| 13810 | 'TUBA1A_No21' | 'AVFVDLEPTVIDEVRTGTYR' | 'Tubulin alpha-1A chain' |
| 13811 | 'TUBA1A_No22' | 'LDHKFDLMYAKR' | 'Tubulin alpha-1A chain' |
| 13812 | 'ADAMTS13_No1' | 'EVCQAVPCPAR' | 'A disintegrin and metalloproteinase with thrombospondin motifs 13' |
| 13813 | 'ADAMTS13_No2' | 'AGAQQPAVALETCNPQPCPAR' | 'A disintegrin and metalloproteinase with thrombospondin motifs 13' |
| 13814 | 'ADAMTS13_No3' | 'QAWVWAAVR' | 'A disintegrin and metalloproteinase with thrombospondin motifs 13' |
| 13815 | 'ADAMTS13_No4' | 'DTCLGPQAQAPVPADFCQHLPKPVTVR' | 'A disintegrin and metalloproteinase with thrombospondin motifs 13' |
| 13816 | 'ADAMTS13_No5' | 'YVLTNLNIGAELLR' | 'A disintegrin and metalloproteinase with thrombospondin motifs 13' |
| 13817 | 'ADAMTS13_No6' | 'SSPGGASFYHWGAAVPHSQGDALCR' | 'A disintegrin and metalloproteinase with thrombospondin motifs 13' |

| | A | B | C |
|---|---|---|---|
| 1 | **Peptide Name** | **Peptide Sequence** | **Peptide Description** |
| 13818 | 'ADAMTS13_No7' | 'LLVPLLDGTECGVEK' | 'A disintegrin and metalloproteinase with thrombospondin motifs 13' |
| 13819 | 'ADAMTS13_No8' | 'ACVGADLQAEMCNTQACEK' | 'A disintegrin and metalloproteinase with thrombospondin motifs 13' |
| 13820 | 'ADAMTS13_No9' | 'GPGQADCAVAIGRPLGEVVTLR' | 'A disintegrin and metalloproteinase with thrombospondin motifs 13' |
| 13821 | 'ADAMTS13_No10' | 'IWGPLQEDADIQVYR' | 'A disintegrin and metalloproteinase with thrombospondin motifs 13' |
| 13822 | 'ADAMTS13_No11' | 'VPVQEELCGLASKPGSR' | 'A disintegrin and metalloproteinase with thrombospondin motifs 13' |
| 13823 | 'ADAMTS13_No12' | 'LFINVAPHAR' | 'A disintegrin and metalloproteinase with thrombospondin motifs 13' |
| 13824 | 'ADAMTS13_No13' | 'YGSQLAPETFYR' | 'A disintegrin and metalloproteinase with thrombospondin motifs 13' |
| 13825 | 'ADAMTS13_No14' | 'GPGQADCAVAIGR' | 'A disintegrin and metalloproteinase with thrombospondin motifs 13' |
| 13826 | 'ADAMTS13_No15' | 'RQLLSLLSAGR' | 'A disintegrin and metalloproteinase with thrombospondin motifs 13' |
| 13827 | 'ADAMTS13_No16' | 'SLVELTPIAAVHGR' | 'A disintegrin and metalloproteinase with thrombospondin motifs 13' |
| 13828 | 'ADAMTS13_No17' | 'EHLDMCQALSCHTDPLDQSSCSR' | 'A disintegrin and metalloproteinase with thrombospondin motifs 13' |
| 13829 | 'ADAMTS13_No18' | 'VALTEDRLPR' | 'A disintegrin and metalloproteinase with thrombospondin motifs 13' |
| 13830 | 'ADAMTS13_No19' | 'LLVPLLDGTECGVEKWCSK' | 'A disintegrin and metalloproteinase with thrombospondin motifs 13' |
| 13831 | 'ADAMTS13_No20' | 'KLLDMTFSSK' | 'A disintegrin and metalloproteinase with thrombospondin motifs 13' |
| 13832 | 'ADAMTS13_No21' | 'VPVQEELCGLASK' | 'A disintegrin and metalloproteinase with thrombospondin motifs 13' |
| 13833 | 'ADAMTS13_No22' | 'FLCMDSALR' | 'A disintegrin and metalloproteinase with thrombospondin motifs 13' |
| 13834 | 'ADAMTS13_No23' | 'TGAQAAHVWTPAAGSCSVSCGR' | 'A disintegrin and metalloproteinase with thrombospondin motifs 13' |
| 13835 | 'ADAMTS13_No24' | 'WHVGTWMECSVSCGDGIQR' | 'A disintegrin and metalloproteinase with thrombospondin motifs 13' |
| 13836 | 'ADAMTS13_No25' | 'IWGPLQEDADIQVYRR' | 'A disintegrin and metalloproteinase with thrombospondin motifs 13' |
| 13837 | 'ADAMTS13_No26' | 'ACVGADLQAEMCNTQACEK' | 'A disintegrin and metalloproteinase with thrombospondin motifs 13' |
| 13838 | 'SND1_No1' | 'NLPGLVQEGEPFSEEATLFTK' | 'Staphylococcal nuclease domain-containing protein 1' |
| 13839 | 'SND1_No2' | 'ASSAQSGGSSGGPAVPTVQR' | 'Staphylococcal nuclease domain-containing protein 1' |
| 13840 | 'SND1_No3' | 'DTNGENIAESLVAEGLATR' | 'Staphylococcal nuclease domain-containing protein 1' |
| 13841 | 'DOK3' | 'GPALLVLGPDAIQLR' | 'Docking protein 3' |
| 13842 | 'MOB1B' | 'ELAPLQELIEK' | 'MOB kinase activator 1B' |
| 13843 | 'MAVS' | 'DTLWHLFNTLQR' | 'Mitochondrial antiviral-signaling protein' |
| 13844 | 'C6orf120' | 'VYYDGTVEQHPFGEAAYPADGADAGQK' | 'UPF0669 protein C6orf120' |
| 13845 | 'DCXR' | 'ECPGIEPVCVDLGDWEATER' | 'L-xylulose reductase' |
| 13846 | 'KRT77' | 'VDTLTGEVNFLK' | 'Keratin, type II cytoskeletal 1b' |
| 13847 | 'ABI3BP_No1' | 'YIQKPDNSPCSITDSVK' | 'Target of Nesh-SH3' |
| 13848 | 'ABI3BP_No2' | 'NPLGEGPVSNTVAFSTESADPR' | 'Target of Nesh-SH3' |
| 13849 | 'ABI3BP_No3' | 'GHGEDHCQFVDSFLDGR' | 'Target of Nesh-SH3' |
| 13850 | 'ABI3BP_No4' | 'YIQKPDNSPCSITDSVKR' | 'Target of Nesh-SH3' |
| 13851 | 'ABI3BP_No5' | 'KFVGVQLCNSLR' | 'Target of Nesh-SH3' |
| 13852 | 'ABI3BP_No6' | 'DAIWTERPFNSDSYSECK' | 'Target of Nesh-SH3' |
| 13853 | 'ABI3BP_No7' | 'TGQQLTSDQLPIK' | 'Target of Nesh-SH3' |
| 13854 | 'ABI3BP_No8' | 'FVGVQLCNSLR' | 'Target of Nesh-SH3' |
| 13855 | 'MEGF8_No1' | 'TPHDLFSSGLFR' | 'Multiple epidermal growth factor-like domains protein 8' |
| 13856 | 'MEGF8_No2' | 'TLQPGDGEASTPR' | 'Multiple epidermal growth factor-like domains protein 8' |
| 13857 | 'MEGF8_No3' | 'GPDTENMEEVGR' | 'Multiple epidermal growth factor-like domains protein 8' |
| 13858 | 'MEGF8_No4' | 'WCTNCPEGACIGR' | 'Multiple epidermal growth factor-like domains protein 8' |
| 13859 | 'MEGF8_No5' | 'LFHASALLGDTMVVLGGR' | 'Multiple epidermal growth factor-like domains protein 8' |
| 13860 | 'MEGF8_No6' | 'STTITLTPSAETDVSLVYR' | 'Multiple epidermal growth factor-like domains protein 8' |
| 13861 | 'MEGF8_No7' | 'APQTVELPAVAGHTLTAR' | 'Multiple epidermal growth factor-like domains protein 8' |

| | A | B | C |
|---|---|---|---|
| 1 | **Peptide Name** | **Peptide Sequence** | **Peptide Description** |
| 13862 | 'MEGF8_No8' | 'GPESCSLGCAQATQCALCLR' | 'Multiple epidermal growth factor-like domains protein 8' |
| 13863 | 'MEGF8_No9' | 'GAMYLLGGLTAGGVTR' | 'Multiple epidermal growth factor-like domains protein 8' |
| 13864 | 'MEGF8_No10' | 'GPGFCDECQDWTWGEHCER' | 'Multiple epidermal growth factor-like domains protein 8' |
| 13865 | 'MEGF8_No11' | 'LLGDCQACLAFSSPTAPPR' | 'Multiple epidermal growth factor-like domains protein 8' |
| 13866 | 'MEGF8_No12' | 'GWDDSVHSEPR' | 'Multiple epidermal growth factor-like domains protein 8' |
| 13867 | 'MEGF8_No13' | 'SPEECPPLCSQR' | 'Multiple epidermal growth factor-like domains protein 8' |
| 13868 | 'MEGF8_No14' | 'FLDTGVVQSDR' | 'Multiple epidermal growth factor-like domains protein 8' |
| 13869 | 'MEGF8_No15' | 'SASVGPPMEESVAHAVAAVGSR' | 'Multiple epidermal growth factor-like domains protein 8' |
| 13870 | 'MEGF8_No16' | 'LLALTLPPDPCR' | 'Multiple epidermal growth factor-like domains protein 8' |
| 13871 | 'MEGF8_No17' | 'LGCGGSPCSPMPR' | 'Multiple epidermal growth factor-like domains protein 8' |
| 13872 | 'MEGF8_No18' | 'WTQMLAGAEDGGPGPSPR' | 'Multiple epidermal growth factor-like domains protein 8' |
| 13873 | 'ADGRG6_No1' | 'VILPQTSDAYQVSVAK' | 'Adhesion G-protein coupled receptor G6' |
| 13874 | 'ADGRG6_No2' | 'ISVVIQNILR' | 'Adhesion G-protein coupled receptor G6' |
| 13875 | 'ADGRG6_No3' | 'NYETVPCDSTISK' | 'Adhesion G-protein coupled receptor G6' |
| 13876 | 'ADGRG6_No4' | 'LLLGSNQNEIVSLK' | 'Adhesion G-protein coupled receptor G6' |
| 13877 | 'ADGRG6_No5' | 'VVLSNPSGTFTSPCYPNDYPNSQACMWTLR' | 'Adhesion G-protein coupled receptor G6' |
| 13878 | 'ADAMTSL2_No1' | 'VVPESQCQMETKPLAIHPCGDK' | 'ADAMTS-like protein 2' |
| 13879 | 'ADAMTSL2_No2' | 'KPPEESTCFERPCFK' | 'ADAMTS-like protein 2' |
| 13880 | 'ADAMTSL2_No3' | 'MLSPGFDSSVYSDLCEAAEAVRPEER' | 'ADAMTS-like protein 2' |
| 13881 | 'ADAMTSL2_No4' | 'MLSPGFDSSVYSDLCEAAEAVRPEER' | 'ADAMTS-like protein 2' |
| 13882 | 'ADAMTSL2_No5' | 'VNLCGHWYYSK' | 'ADAMTS-like protein 2' |
| 13883 | 'ADAMTSL2_No6' | 'NPACGPQWEMSEWSECTAK' | 'ADAMTS-like protein 2' |
| 13884 | 'ADAMTSL2_No7' | 'NCPAHWLAQDWER' | 'ADAMTS-like protein 2' |
| 13885 | 'ADAMTSL2_No8' | 'SFREEQCVSFNSHVYNGR' | 'ADAMTS-like protein 2' |
| 13886 | 'ADAMTSL2_No9' | 'GNAHLGYSLVTHIPAGAR' | 'ADAMTS-like protein 2' |
| 13887 | 'ADAMTSL2_No10' | 'CGICQGDGSSCTHVTGNYR' | 'ADAMTS-like protein 2' |
| 13888 | 'ADAMTSL2_No11' | 'YDGVEVDDSYCDALTRPEPVHEFCAGR' | 'ADAMTS-like protein 2' |
| 13889 | 'ADAMTSL2_No12' | 'NFNIAGTVVK' | 'ADAMTS-like protein 2' |
| 13890 | 'ADAMTSL2_No13' | 'LVLCMELANGKPQTR' | 'ADAMTS-like protein 2' |
| 13891 | 'SERPINA11_No1' | 'HQLSEPAPAYHR' | 'Serpin A11' |
| 13892 | 'SERPINA11_No2' | 'QINDYLR' | 'Serpin A11' |
| 13893 | 'SERPINA11_No3' | 'GNALALLVLPDPGK' | 'Serpin A11' |
| 13894 | 'SERPINA11_No4' | 'ITPTITNFALR' | 'Serpin A11' |
| 13895 | 'SERPINA11_No5' | 'SLLHTLALPSPK' | 'Serpin A11' |
| 13896 | 'OAF_No1' | 'SYSFDFYVPQR' | 'Out at first protein homolog' |
| 13897 | 'OAF_No2' | 'ALILGELEK' | 'Out at first protein homolog' |
| 13898 | 'OAF_No3' | 'FWLEQGVDSSVFEALPK' | 'Out at first protein homolog' |
| 13899 | 'OAF_No4' | 'KPDGTLVSFTADFKK' | 'Out at first protein homolog' |
| 13900 | 'OAF_No5' | 'GQSQFQALCFVTQLQHNEIIPSEAMAK' | 'Out at first protein homolog' |
| 13901 | 'OAF_No6' | 'YGLSLAWYPCMLK' | 'Out at first protein homolog' |
| 13902 | 'RTN4RL2_No1' | 'VSSSDVTCATPPER' | 'Reticulon-4 receptor-like 2' |
| 13903 | 'RTN4RL2_No2' | 'ARPLWAWFQR' | 'Reticulon-4 receptor-like 2' |
| 13904 | 'RTN4RL2_No3' | 'CQLSSLPGNIFR' | 'Reticulon-4 receptor-like 2' |
| 13905 | 'FERMT3_No1' | 'VVLAGGVAPALFR' | 'Fermitin family homolog 3' |

| | A | B | C |
|---|---|---|---|
| 1 | **Peptide Name** | **Peptide Sequence** | **Peptide Description** |
| 13906 | 'FERMT3_No2' | 'LFFGPQHRPVILR' | 'Fermitin family homolog 3' |
| 13907 | 'FERMT3_No3' | 'VFVGEEDPEAESVTLR' | 'Fermitin family homolog 3' |
| 13908 | 'FERMT3_No4' | 'LLVPSPEGMSEIYLR' | 'Fermitin family homolog 3' |
| 13909 | 'FERMT3_No5' | 'HPEELSLLR' | 'Fermitin family homolog 3' |
| 13910 | 'FERMT3_No6' | 'ILEAHQNVAQLSLAEAQLR' | 'Fermitin family homolog 3' |
| 13911 | 'FERMT3_No7' | 'LEGSAPTDVLDSLTTIPELKDHLR' | 'Fermitin family homolog 3' |
| 13912 | 'FERMT3_No8' | 'VTGESHIGGVLLK' | 'Fermitin family homolog 3' |
| 13913 | 'FERMT3_No9' | 'LTQLYEQAR' | 'Fermitin family homolog 3' |
| 13914 | 'FERMT3_No10' | 'KDEILGIANNR' | 'Fermitin family homolog 3' |
| 13915 | 'FERMT3_No11' | 'GCEVVPDVNVSGQK' | 'Fermitin family homolog 3' |
| 13916 | 'FERMT3_No12' | 'EKEPEEELYDLSK' | 'Fermitin family homolog 3' |
| 13917 | 'FERMT3_No13' | 'ASFSQPLFQAVAAICR' | 'Fermitin family homolog 3' |
| 13918 | 'FERMT3_No14' | 'LEGSAPTDVLDSLTTIPELK' | 'Fermitin family homolog 3' |
| 13919 | 'FERMT3_No15' | 'AGDALWLR' | 'Fermitin family homolog 3' |
| 13920 | 'FERMT3_No16' | 'QWLLQTHWTLDK' | 'Fermitin family homolog 3' |
| 13921 | 'FERMT3_No17' | 'KQDWSDHAIWWEQK' | 'Fermitin family homolog 3' |
| 13922 | 'FERMT3_No18' | 'QDWSDHAIWWEQK' | 'Fermitin family homolog 3' |
| 13923 | 'FERMT3_No19' | 'LLVPSPEGMSEIYLR' | 'Fermitin family homolog 3' |
| 13924 | 'FERMT3_No20' | 'ARGEELDEDLFLQLTGGHEAF' | 'Fermitin family homolog 3' |
| 13925 | 'FERMT3_No21' | 'QWNVNWDIR' | 'Fermitin family homolog 3' |
| 13926 | 'FERMT3_No22' | 'GMPAHFSDSAQTEACYHMLSR' | 'Fermitin family homolog 3' |
| 13927 | 'FERMT3_No23' | 'LLSIRHPEELSLLR' | 'Fermitin family homolog 3' |
| 13928 | 'FERMT3_No24' | 'IVHEYIGGYIFLSTR' | 'Fermitin family homolog 3' |
| 13929 | 'FERMT3_No25' | 'QWLLQTHWTLDK' | 'Fermitin family homolog 3' |
| 13930 | 'FERMT3_No26' | 'YYSFFDLDPK' | 'Fermitin family homolog 3' |
| 13931 | 'FERMT3_No27' | 'SQDEAPGDPIQQLNLK' | 'Fermitin family homolog 3' |
| 13932 | 'FERMT3_No28' | 'QDWSDHAIWWEQK' | 'Fermitin family homolog 3' |
| 13933 | 'FERMT3_No29' | 'QWNVNWDIR' | 'Fermitin family homolog 3' |
| 13934 | 'FERMT3_No30' | 'TASGDYIDSSWELR' | 'Fermitin family homolog 3' |
| 13935 | 'CD163_No1' | 'SSMSETTVGVVCR' | 'Scavenger receptor cysteine-rich type 1 protein M130' |
| 13936 | 'CD163_No2' | 'HYCNHNEDAGVTCSDGSDLELR' | 'Scavenger receptor cysteine-rich type 1 protein M130' |
| 13937 | 'CD163_No3' | 'LVDGVTECSGR' | 'Scavenger receptor cysteine-rich type 1 protein M130' |
| 13938 | 'CD163_No4' | 'LASPSEETWITCDNK' | 'Scavenger receptor cysteine-rich type 1 protein M130' |
| 13939 | 'CD163_No5' | 'CGVALSTPGGAR' | 'Scavenger receptor cysteine-rich type 1 protein M130' |
| 13940 | 'CD163_No6' | 'WGTVCDDNFNIDHASVICR' | 'Scavenger receptor cysteine-rich type 1 protein M130' |
| 13941 | 'CD163_No7' | 'QLGCPTAVTAIGR' | 'Scavenger receptor cysteine-rich type 1 protein M130' |
| 13942 | 'CD163_No8' | 'HGDTWGSICDSDFSLEAASVLCR' | 'Scavenger receptor cysteine-rich type 1 protein M130' |
| 13943 | 'CD163_No9' | 'LEVFYNGAWGTVGK' | 'Scavenger receptor cysteine-rich type 1 protein M130' |
| 13944 | 'CD163_No10' | 'IWQCHSHGWGQQNCR' | 'Scavenger receptor cysteine-rich type 1 protein M130' |
| 13945 | 'CD163_No11' | 'AFKEAEFGQGTGPIWLNEVK' | 'Scavenger receptor cysteine-rich type 1 protein M130' |
| 13946 | 'CD163_No12' | 'AMSIPMWVDNVQCPK' | 'Scavenger receptor cysteine-rich type 1 protein M130' |
| 13947 | 'CD163_No13' | 'LVGGDIPCSGRVEVK' | 'Scavenger receptor cysteine-rich type 1 protein M130' |
| 13948 | 'CD163_No14' | 'GPDTLWQCPSSPWEK' | 'Scavenger receptor cysteine-rich type 1 protein M130' |
| 13949 | 'CD163_No15' | 'LVGGDIPCSGR' | 'Scavenger receptor cysteine-rich type 1 protein M130' |

| | A | B | C |
|---|---|---|---|
| 1 | **Peptide Name** | **Peptide Sequence** | **Peptide Description** |
| 13950 | 'CD163_No16' | 'EAEFGQGTGPIWLNEVK' | 'Scavenger receptor cysteine-rich type 1 protein M130' |
| 13951 | 'CD163_No17' | 'NWQWGGLTCDHYEEAK' | 'Scavenger receptor cysteine-rich type 1 protein M130' |
| 13952 | 'CD163_No18' | 'HKEDAGVICSEFMSLR' | 'Scavenger receptor cysteine-rich type 1 protein M130' |
| 13953 | 'CD163_No19' | 'SSMSETTVGVVCR' | 'Scavenger receptor cysteine-rich type 1 protein M130' |
| 13954 | 'CD163_No20' | 'AMSIPMWVDNVQCPK' | 'Scavenger receptor cysteine-rich type 1 protein M130' |
| 13955 | 'CD163_No21' | 'RLASPSEETWITCDNKIR' | 'Scavenger receptor cysteine-rich type 1 protein M130' |
| 13956 | 'CD163_No22' | 'TLGAWGSLCNSHWDIEDAHVLCQQLK' | 'Scavenger receptor cysteine-rich type 1 protein M130' |
| 13957 | 'CAND1_No1' | 'LGTLSALDILIK' | 'Cullin-associated NEDD8-dissociated protein 1' |
| 13958 | 'CAND1_No2' | 'EGPAVVGQFIQDVK' | 'Cullin-associated NEDD8-dissociated protein 1' |
| 13959 | 'CAND1_No3' | 'ITSEALLVTQQLVK' | 'Cullin-associated NEDD8-dissociated protein 1' |
| 13960 | 'CAND1_No4' | 'SVILEAFSSPSEEVK' | 'Cullin-associated NEDD8-dissociated protein 1' |
| 13961 | 'TREML1_No1' | 'VSLNILPPEEEEETHK' | 'Trem-like transcript 1 protein' |
| 13962 | 'TREML1_No2' | 'GGGTSCGPAQNPPNNQTPSS' | 'Trem-like transcript 1 protein' |
| 13963 | 'TREML1_No3' | 'FLPEGCQPLVSSAVDR' | 'Trem-like transcript 1 protein' |
| 13964 | 'HRNR_No1' | 'GPYESGSGHSSGLGHQESR' | 'Hornerin' |
| 13965 | 'HRNR_No2' | 'SSSSGQHGSGLGESSGFGHHESSSGQSSSYSQHGSGSGHSSGYGQH | 'Hornerin' |
| 13966 | 'HRNR_No3' | 'HGSGSGQSSSYGPYGSGSGWSSSR' | 'Hornerin' |
| 13967 | 'HRNR_No4' | 'HGSGSGQSSSYSPYGSGSGWSSSR' | 'Hornerin' |
| 13968 | 'HRNR_No5' | 'GPYESGSGHSSGLGHR' | 'Hornerin' |
| 13969 | 'HRNR_No6' | 'GSGSGQSPSSGQHGTGFGR' | 'Hornerin' |
| 13970 | 'HRNR_No7' | 'QSLGHGQHGSGSGQSPSPSR' | 'Hornerin' |
| 13971 | 'HRNR_No8' | 'HGSGSGQSSGFGHHESSSWQSSGYTQHGSGSGHSSSYEQHGSR' | 'Hornerin' |
| 13972 | 'HRNR_No9' | 'GHYESGSGQTSGFGQHESGSGQSSGYSK' | 'Hornerin' |
| 13973 | 'HRNR_No10' | 'SSSRGPYESGSGHSSGLGHQESR' | 'Hornerin' |
| 13974 | 'HRNR_No11' | 'SSSGQSSGYTQHGSGSGHSSSYEQHGSR' | 'Hornerin' |
| 13975 | 'HRNR_No12' | 'QSLGHGQHGSGSGQSPSPSR' | 'Hornerin' |
| 13976 | 'HRNR_No13' | 'GEQHGSSSGSSSSYGQHGSGSR' | 'Hornerin' |
| 13977 | 'FAM20C_No1' | 'LSLLMAESLR' | 'Extracellular serine/threonine protein kinase FAM20C' |
| 13978 | 'FAM20C_No2' | 'HNAEIAAFHLDR' | 'Extracellular serine/threonine protein kinase FAM20C' |
| 13979 | 'FAM20C_No3' | 'NGLHSVVDDDLDTEHR' | 'Extracellular serine/threonine protein kinase FAM20C' |
| 13980 | 'FAM20C_No4' | 'HNPAIEALLHDLSSQR' | 'Extracellular serine/threonine protein kinase FAM20C' |
| 13981 | 'FAM20C_No5' | 'GDQVAPVLYQPHLEALDR' | 'Extracellular serine/threonine protein kinase FAM20C' |
| 13982 | 'FAM20C_No6' | 'LIMTFQNYGQALFKPMK' | 'Extracellular serine/threonine protein kinase FAM20C' |
| 13983 | 'OSCAR_No1' | 'FGLFKPGEIAPLLFR' | 'Osteoclast-associated immunoglobulin-like receptor' |
| 13984 | 'OSCAR_No2' | 'APGTYSCYYHTPSAPYVLSQR' | 'Osteoclast-associated immunoglobulin-like receptor' |
| 13985 | 'OSCAR_No3' | 'HSAQPWADFTLLGAR' | 'Osteoclast-associated immunoglobulin-like receptor' |
| 13986 | 'ADGRF5_No1' | 'INIPGTPETDIDSSCSR' | 'Adhesion G protein-coupled receptor F5' |
| 13987 | 'ADGRF5_No2' | 'FSNVPSSPESPIGGTITYK' | 'Adhesion G protein-coupled receptor F5' |
| 13988 | 'ADGRF5_No3' | 'CVFWNFR' | 'Adhesion G protein-coupled receptor F5' |
| 13989 | 'ADGRF5_No4' | 'LILDIFEYECKK' | 'Adhesion G protein-coupled receptor F5' |
| 13990 | 'ADGRF5_No5' | 'SPSQDEMLPTYLK' | 'Adhesion G protein-coupled receptor F5' |
| 13991 | 'ADGRF5_No6' | 'SPSQDEMLPTYLK' | 'Adhesion G protein-coupled receptor F5' |
| 13992 | 'KRT78_No1' | 'TLNNQFASFIDK' | 'Keratin, type II cytoskeletal 78' |
| 13993 | 'KRT78_No2' | 'YLDFSSIITEVR' | 'Keratin, type II cytoskeletal 78' |

| | A | B | C |
|---|---|---|---|
| 1 | **Peptide Name** | **Peptide Sequence** | **Peptide Description** |
| 13994 | 'KRT78_No3' | 'IEIDPQFQVVR' | 'Keratin, type II cytoskeletal 78' |
| 13995 | 'KRT78_No4' | 'ATLENDFVVLKK' | 'Keratin, type II cytoskeletal 78' |
| 13996 | 'KRT78_No5' | 'VQISQLHQEIQR' | 'Keratin, type II cytoskeletal 78' |
| 13997 | 'KRT78_No6' | 'QNASLQAAITDAEQR' | 'Keratin, type II cytoskeletal 78' |
| 13998 | 'KRT78_No7' | 'TLNNQFASFIDKVR' | 'Keratin, type II cytoskeletal 78' |
| 13999 | 'KRT78_No8' | 'YQELQVSAQLHGDR' | 'Keratin, type II cytoskeletal 78' |
| 14000 | 'ARHGAP18_No1' | 'IPLIFQK' | 'Rho GTPase-activating protein 18' |
| 14001 | 'ARHGAP18_No2' | 'ETAPGGTESQSLR' | 'Rho GTPase-activating protein 18' |
| 14002 | 'ARHGAP18_No3' | 'FLSQESGVAQTLK' | 'Rho GTPase-activating protein 18' |
| 14003 | 'ARHGAP18_No4' | 'YGQYTMNQESTTIK' | 'Rho GTPase-activating protein 18' |
| 14004 | 'TMEM132C_No1' | 'EVGMTTIQVLSPLSDSILAEK' | 'Transmembrane protein 132C' |
| 14005 | 'TMEM132C_No2' | 'LPLQIEVSDTELSQIK' | 'Transmembrane protein 132C' |
| 14006 | 'LILRA3_No1' | 'KPSLSVQPGPVVAPGEK' | 'Leukocyte immunoglobulin-like receptor subfamily A member 3' |
| 14007 | 'LILRA3_No2' | 'YQAEFPMSPVTSAHAGTYR' | 'Leukocyte immunoglobulin-like receptor subfamily A member 3' |
| 14008 | 'LILRA3_No3' | 'TALWITR' | 'Leukocyte immunoglobulin-like receptor subfamily A member 3' |
| 14009 | 'LILRA3_No4' | 'PTLWAEPGSVITQGSPVTLR' | 'Leukocyte immunoglobulin-like receptor subfamily A member 3' |
| 14010 | 'LILRA3_No5' | 'CQGSLETQEYHLYR' | 'Leukocyte immunoglobulin-like receptor subfamily A member 3' |
| 14011 | 'LILRA3_No6' | 'EGEDEHPQCLNSHSHAR' | 'Leukocyte immunoglobulin-like receptor subfamily A member 3' |
| 14012 | 'LILRA3_No7' | 'DFLQRPGR' | 'Leukocyte immunoglobulin-like receptor subfamily A member 3' |
| 14013 | 'LILRA3_No8' | 'YQAEFPMSPVTSAHAGTYR' | 'Leukocyte immunoglobulin-like receptor subfamily A member 3' |
| 14014 | 'LILRA3_No9' | 'EGAADSPLR' | 'Leukocyte immunoglobulin-like receptor subfamily A member 3' |
| 14015 | 'GOLM1_No1' | 'IQSSHNFQLESVNK' | 'Golgi membrane protein 1' |
| 14016 | 'GOLM1_No2' | 'NIDVFNVEDQKR' | 'Golgi membrane protein 1' |
| 14017 | 'GOLM1_No3' | 'DQLVIPDGQEEEQEAAGEGR' | 'Golgi membrane protein 1' |
| 14018 | 'GOLM1_No4' | 'KFSYDLSQCINQMK' | 'Golgi membrane protein 1' |
| 14019 | 'GOLM1_No5' | 'LPQEPGREQVVEDRPVGGR' | 'Golgi membrane protein 1' |
| 14020 | 'GOLM1_No6' | 'KFSYDLSQCINQMK' | 'Golgi membrane protein 1' |
| 14021 | 'GOLM1_No7' | 'QVEKEETNEIQVVNEEPQRDR' | 'Golgi membrane protein 1' |
| 14022 | 'PCSK9_No1' | 'LPGTYVVVLK' | 'Proprotein convertase subtilisin/kexin type 9' |
| 14023 | 'PCSK9_No2' | 'GTVSGTLIGLEFIR' | 'Proprotein convertase subtilisin/kexin type 9' |
| 14024 | 'PCSK9_No3' | 'EASIHASCCHAPGLECK' | 'Proprotein convertase subtilisin/kexin type 9' |
| 14025 | 'PCSK9_No4' | 'VLTPNLVAALPPSTHGAGWQLFCR' | 'Proprotein convertase subtilisin/kexin type 9' |
| 14026 | 'PCSK9_No5' | 'DVINEAWFPEDQR' | 'Proprotein convertase subtilisin/kexin type 9' |
| 14027 | 'PCSK9_No6' | 'MSGDLLELALK' | 'Proprotein convertase subtilisin/kexin type 9' |
| 14028 | 'PCSK9_No7' | 'SEEDGLAEAPEHGTTATFHR' | 'Proprotein convertase subtilisin/kexin type 9' |
| 14029 | 'PCSK9_No8' | 'AHNAFGGEGVYAIAR' | 'Proprotein convertase subtilisin/kexin type 9' |
| 14030 | 'PCSK9_No9' | 'CDSHGTHLAGVVSGR' | 'Proprotein convertase subtilisin/kexin type 9' |
| 14031 | 'PCSK9_No10' | 'VMVTDFENVPEEDGTR' | 'Proprotein convertase subtilisin/kexin type 9' |
| 14032 | 'PCSK9_No11' | 'CAPDEELLSCSSFSR' | 'Proprotein convertase subtilisin/kexin type 9' |
| 14033 | 'PCSK9_No12' | 'SQLVQPVGPLVVLLPLAGGYSR' | 'Proprotein convertase subtilisin/kexin type 9' |
| 14034 | 'PCSK9_No13' | 'MSGDLLELALK' | 'Proprotein convertase subtilisin/kexin type 9' |
| 14035 | 'PCSK9_No14' | 'LPGTYVVVLKEETHLSQSER' | 'Proprotein convertase subtilisin/kexin type 9' |
| 14036 | 'TXNDC5_No1' | 'GYPTLLLFR' | 'Thioredoxin domain-containing protein 5' |
| 14037 | 'TXNDC5_No2' | 'VDCTAHSDVCSAQGVR' | 'Thioredoxin domain-containing protein 5' |

| | A | B | C |
|---|---|---|---|
| 1 | **Peptide Name** | **Peptide Sequence** | **Peptide Description** |
| 14038 | 'TXNDC5_No3' | 'GYPTLLWFR' | 'Thioredoxin domain-containing protein 5' |
| 14039 | 'TXNDC5_No4' | 'ALAPTWEQLALGLEHSETVK' | 'Thioredoxin domain-containing protein 5' |
| 14040 | 'TXNDC5_No5' | 'VDCTQHYELCSGNQVR' | 'Thioredoxin domain-containing protein 5' |
| 14041 | 'TXNDC5_No6' | 'EYVESQLQR' | 'Thioredoxin domain-containing protein 5' |
| 14042 | 'ANGPTL6_No1' | 'LAAADGAVAGEVR' | 'Angiopoietin-related protein 6' |
| 14043 | 'ANGPTL6_No2' | 'DSYSGNCALYQR' | 'Angiopoietin-related protein 6' |
| 14044 | 'ANGPTL6_No3' | 'ELAQLVTQQSSLIAR' | 'Angiopoietin-related protein 6' |
| 14045 | 'ANGPTL6_No4' | 'AHYDGFSLEPESDHYR' | 'Angiopoietin-related protein 6' |
| 14046 | 'ANGPTL6_No5' | 'MLDPAPEPQRDQTQR' | 'Angiopoietin-related protein 6' |
| 14047 | 'ANGPTL6_No6' | 'GDHELLVLLEDWGGR' | 'Angiopoietin-related protein 6' |
| 14048 | 'ANGPTL6_No7' | 'MLDPAPEPQR' | 'Angiopoietin-related protein 6' |
| 14049 | 'ANGPTL6_No8' | 'QDGSVNFFTTWQHYK' | 'Angiopoietin-related protein 6' |
| 14050 | 'ANGPTL6_No9' | 'FTGAVCWSGPASTR' | 'Angiopoietin-related protein 6' |
| 14051 | 'ANGPTL6_No10' | 'QDGSVNFFTTWQHYK' | 'Angiopoietin-related protein 6' |
| 14052 | 'ANGPTL6_No11' | 'RQDGSVNFFTTWQHYK' | 'Angiopoietin-related protein 6' |
| 14053 | 'ANGPTL6_No12' | 'ATPEAANASELAALR' | 'Angiopoietin-related protein 6' |
| 14054 | 'ANGPTL6_No13' | 'LGQYHGDAGDSLSWHNDKPFSTVDRDR' | 'Angiopoietin-related protein 6' |
| 14055 | 'RDH11_No1' | 'GSGVTTYSVHPGTVQSELVR' | 'Retinol dehydrogenase 11' |
| 14056 | 'RDH11_No2' | 'LANILFTQELAR' | 'Retinol dehydrogenase 11' |
| 14057 | 'BPIFB1_No1' | 'LSFLVNALAK' | 'BPI fold-containing family B member 1' |
| 14058 | 'BPIFB1_No2' | 'VITANILQLQVKPSANDQELLVK' | 'BPI fold-containing family B member 1' |
| 14059 | 'BPIFB1_No3' | 'QVMNLLVPSLPNLVK' | 'BPI fold-containing family B member 1' |
| 14060 | 'BPIFB1_No4' | 'IQLMNSGIGWFQPDVLK' | 'BPI fold-containing family B member 1' |
| 14061 | 'BPIFB1_No5' | 'DALVLTPASLWKPSSPVSQ' | 'BPI fold-containing family B member 1' |
| 14062 | 'BPIFB1_No6' | 'IPLDMVAGFNTPLVK' | 'BPI fold-containing family B member 1' |
| 14063 | 'BPIFB1_No7' | 'IPLDMVAGFNTPLVK' | 'BPI fold-containing family B member 1' |
| 14064 | 'BPIFB1_No8' | 'LVLSDCATSHGSLR' | 'BPI fold-containing family B member 1' |
| 14065 | 'BPIFB1_No9' | 'EKPAGGIPVLGSLVNTVLK' | 'BPI fold-containing family B member 1' |
| 14066 | 'BPIFB1_No10' | 'QVMNLLVPSLPNLVK' | 'BPI fold-containing family B member 1' |
| 14067 | 'BPIFB1_No11' | 'DALVLTPASLWK' | 'BPI fold-containing family B member 1' |
| 14068 | 'BPIFB1_No12' | 'GDQLILNLNNISSDR' | 'BPI fold-containing family B member 1' |
| 14069 | 'BPIFB1_No13' | 'SGVPVSLVK' | 'BPI fold-containing family B member 1' |
| 14070 | 'BPIFB1_No14' | 'IQLMNSGIGWFQPDVLK' | 'BPI fold-containing family B member 1' |
| 14071 | 'BPIFB1_No15' | 'ILTQDTPEFFIDQGHAK' | 'BPI fold-containing family B member 1' |
| 14072 | 'BPIFB1_No16' | 'GDTIQLYLGAK' | 'BPI fold-containing family B member 1' |
| 14073 | 'DTD1' | 'SASSGAEGDVSSEREP' | 'D-aminoacyl-tRNA deacylase 1' |
| 14074 | 'SNED1_No1' | 'EVSQFTPVAFPIAK' | 'Sushi, nidogen and EGF-like domain-containing protein 1' |
| 14075 | 'SNED1_No2' | 'RCQAEVDCGPPEEVK' | 'Sushi, nidogen and EGF-like domain-containing protein 1' |
| 14076 | 'SNED1_No3' | 'GYSLSAPSR' | 'Sushi, nidogen and EGF-like domain-containing protein 1' |
| 14077 | 'TSKU_No1' | 'AGLPAPTIQSLNLAWNR' | 'Tsukushin' |
| 14078 | 'TSKU_No2' | 'ALHVDLSHNLIHR' | 'Tsukushin' |
| 14079 | 'TSKU_No3' | 'EVSVSAFTTHSQGR' | 'Tsukushin' |
| 14080 | 'SECTM1' | 'AHGQESAIFNEVAPGYFSR' | 'Secreted and transmembrane protein 1' |
| 14081 | 'POF1B_No1' | 'YFGELLADLSR' | 'Protein POF1B' |

| | A | B | C |
|---|---|---|---|
| 1 | **Peptide Name** | **Peptide Sequence** | **Peptide Description** |
| 14082 | 'POF1B_No2' | 'NLEQENQNLR' | 'Protein POF1B' |
| 14083 | 'ITLN1_No1' | 'EFTAGFVQFR' | 'Intelectin-1' |
| 14084 | 'ITLN1_No2' | 'CWTDNGPVIPVVYDFGDAQK' | 'Intelectin-1' |
| 14085 | 'ITLN1_No3' | 'TASYYSPYGQR' | 'Intelectin-1' |
| 14086 | 'ITLN1_No4' | 'EITEAAVLLFYR' | 'Intelectin-1' |
| 14087 | 'ITLN1_No5' | 'EIKDECPSAFDGLYFLR' | 'Intelectin-1' |
| 14088 | 'ITLN1_No6' | 'YRTDTGFLQTLGHNLFGIYQK' | 'Intelectin-1' |
| 14089 | 'ITLN1_No7' | 'DLGIWHVPNK' | 'Intelectin-1' |
| 14090 | 'OIT3_No1' | 'LLIPVTCEFPR' | 'Oncoprotein-induced transcript 3 protein' |
| 14091 | 'OIT3_No2' | 'QTPGSSGDFIIR' | 'Oncoprotein-induced transcript 3 protein' |
| 14092 | 'OIT3_No3' | 'HFQVPVFK' | 'Oncoprotein-induced transcript 3 protein' |
| 14093 | 'OIT3_No4' | 'GLVLSEDNHTCQVPVLCK' | 'Oncoprotein-induced transcript 3 protein' |
| 14094 | 'OIT3_No5' | 'SNAIEVNIPR' | 'Oncoprotein-induced transcript 3 protein' |
| 14095 | 'OIT3_No6' | 'SDGKTCEDVEGCHNNNGGCSHSCLGSEK' | 'Oncoprotein-induced transcript 3 protein' |
| 14096 | 'OIT3_No7' | 'TCGTVVDVVNDKIVASNLVTGLPK' | 'Oncoprotein-induced transcript 3 protein' |
| 14097 | 'ROBO4_No1' | 'SDEGTYMCVATNSAGHR' | 'Roundabout homolog 4' |
| 14098 | 'ROBO4_No2' | 'GPDSNVLLLR' | 'Roundabout homolog 4' |
| 14099 | 'ROBO4_No3' | 'MSCQASGQPPPTIR' | 'Roundabout homolog 4' |
| 14100 | 'ROBO4_No4' | 'LSVAVLREDFQIQPR' | 'Roundabout homolog 4' |
| 14101 | 'ROBO4_No5' | 'VSIQEPQDYTEPVELLAVR' | 'Roundabout homolog 4' |
| 14102 | 'ROBO4_No6' | 'PAVWLSWK' | 'Roundabout homolog 4' |
| 14103 | 'ROBO4_No7' | 'MSCQASGQPPPTIR' | 'Roundabout homolog 4' |
| 14104 | 'ROBO4_No8' | 'AEKSDEGTYMCVATNSAGHR' | 'Roundabout homolog 4' |
| 14105 | 'ROBO4_No9' | 'HTVSGGSLLMAR' | 'Roundabout homolog 4' |
| 14106 | 'ROBO4_No10' | 'AEKSDEGTYMCVATNSAGHR' | 'Roundabout homolog 4' |
| 14107 | 'SMPDL3A' | 'LIDIFQK' | 'Acid sphingomyelinase-like phosphodiesterase 3a' |
| 14108 | 'CFHR4_No1' | 'VEYQCQSYYELQGSK' | 'Complement factor H-related protein 4' |
| 14109 | 'CFHR4_No2' | 'YVTCSNGDWSEPPR' | 'Complement factor H-related protein 4' |
| 14110 | 'CFHR4_No3' | 'FCDMPVFENSR' | 'Complement factor H-related protein 4' |
| 14111 | 'CFHR4_No4' | 'VYLPWSR' | 'Complement factor H-related protein 4' |
| 14112 | 'CFHR4_No5' | 'FCDMPVFENSR' | 'Complement factor H-related protein 4' |
| 14113 | 'CFHR4_No6' | 'LGYNANTSVLSFQAVCR' | 'Complement factor H-related protein 4' |
| 14114 | 'CFHR4_No7' | 'CIHPCIITEENMNKNNIQLK' | 'Complement factor H-related protein 4' |
| 14115 | 'FAM3C_No1' | 'LIADLGSTSITNLGFR' | 'Protein FAM3C' |
| 14116 | 'FAM3C_No2' | 'ICLEDNVLMSGVK' | 'Protein FAM3C' |
| 14117 | 'FAM3C_No3' | 'AIQDGTIVLMGTYDDGATK' | 'Protein FAM3C' |
| 14118 | 'PSMF1_No1' | 'AGLEVLFASAAPAITCR' | 'Proteasome inhibitor PI31 subunit' |
| 14119 | 'PSMF1_No2' | 'IVSGIITPIHEQWEK' | 'Proteasome inhibitor PI31 subunit' |
| 14120 | 'PXDN_No1' | 'VPSQLLNTELTER' | 'Peroxidasin homolog' |
| 14121 | 'PXDN_No2' | 'SPNDLLALFR' | 'Peroxidasin homolog' |
| 14122 | 'PXDN_No3' | 'LSTTECVDAGGESHANNTK' | 'Peroxidasin homolog' |
| 14123 | 'GGH_No1' | 'LDLTEKDYEILFK' | 'Gamma-glutamyl hydrolase' |
| 14124 | 'GGH_No2' | 'KPIIGILMQK' | 'Gamma-glutamyl hydrolase' |
| 14125 | 'GGH_No3' | 'NLDGISHAPNAVK' | 'Gamma-glutamyl hydrolase' |

| | A | B | C |
|---|---|---|---|
| 1 | **Peptide Name** | **Peptide Sequence** | **Peptide Description** |
| 14126 | 'GGH_No4' | 'FFNVLTTNTDGK' | 'Gamma-glutamyl hydrolase' |
| 14127 | 'GGH_No5' | 'VVPVRLDLTEKDYEILFK' | 'Gamma-glutamyl hydrolase' |
| 14128 | 'GGH_No6' | 'SINGILFPGGSVDLR' | 'Gamma-glutamyl hydrolase' |
| 14129 | 'GGH_No7' | 'TAFYLAEFFVNEAR' | 'Gamma-glutamyl hydrolase' |
| 14130 | 'GGH_No8' | 'YPVYGVQWHPEK' | 'Gamma-glutamyl hydrolase' |
| 14131 | 'GGH_No9' | 'SINGILFPGGSVDLRR' | 'Gamma-glutamyl hydrolase' |
| 14132 | 'GGH_No10' | 'IEFISTMEGYK' | 'Gamma-glutamyl hydrolase' |
| 14133 | 'NRCAM_No1' | 'NALGAIHHTISVR' | 'Neuronal cell adhesion molecule' |
| 14134 | 'NRCAM_No2' | 'ISWLTNGVPIEIAPDDPSR' | 'Neuronal cell adhesion molecule' |
| 14135 | 'NEO1_No1' | 'VIGQDVVLPCVASGLPTPTIK' | 'Neogenin' |
| 14136 | 'NEO1_No2' | 'KSDVTETLVSGTQLSQLIEGLDR' | 'Neogenin' |
| 14137 | 'NEO1_No3' | 'GYAIGYGIGSPHAQTIK' | 'Neogenin' |
| 14138 | 'NEO1_No4' | 'HNKPDEGYYQCVATVESLGTIISR' | 'Neogenin' |
| 14139 | 'NEO1_No5' | 'QLLPDGSLFISNVVHSK' | 'Neogenin' |
| 14140 | 'NEO1_No6' | 'DGTFLNLVSDDRR' | 'Neogenin' |
| 14141 | 'NEO1_No7' | 'KDGTFLNLVSDDRR' | 'Neogenin' |
| 14142 | 'NEO1_No8' | 'LTHQIQELTLDTPYYFK' | 'Neogenin' |
| 14143 | 'NEO1_No9' | 'WEQNRQPLLLDDR' | 'Neogenin' |
| 14144 | 'NEO1_No10' | 'YSDEVELK' | 'Neogenin' |
| 14145 | 'NEO1_No11' | 'VLPDPEVISDLVFLK' | 'Neogenin' |
| 14146 | 'NEO1_No12' | 'HGPGVSTPDVAVR' | 'Neogenin' |
| 14147 | 'PRG4_No1' | 'GHYFWMLSPFSPPSPAR' | 'Proteoglycan 4' |
| 14148 | 'PRG4_No2' | 'GLPNVVTSAISLPNIR' | 'Proteoglycan 4' |
| 14149 | 'PRG4_No3' | 'DATCNCDYNCQHYMECCPDFKR' | 'Proteoglycan 4' |
| 14150 | 'PRG4_No4' | 'DAGYPKPIFK' | 'Proteoglycan 4' |
| 14151 | 'PRG4_No5' | 'RITEVWGIPSPIDTVFTR' | 'Proteoglycan 4' |
| 14152 | 'PRG4_No6' | 'YKNWPESVYFFK' | 'Proteoglycan 4' |
| 14153 | 'PRG4_No7' | 'GFGGLTGQIVAALSTAK' | 'Proteoglycan 4' |
| 14154 | 'PRG4_No8' | 'DATCNCDYNCQHYMECCPDFKR' | 'Proteoglycan 4' |
| 14155 | 'PRG4_No9' | 'VPNQGIIINPMLSDETNICNGKPVDGLTTLR' | 'Proteoglycan 4' |
| 14156 | 'PRG4_No10' | 'KPDGYDYYAFSK' | 'Proteoglycan 4' |
| 14157 | 'PRG4_No11' | 'PALNYPVYGETTQVR' | 'Proteoglycan 4' |
| 14158 | 'PRG4_No12' | 'RGGSIQQYIYK' | 'Proteoglycan 4' |
| 14159 | 'PRG4_No13' | 'TFFFKDSQYWR' | 'Proteoglycan 4' |
| 14160 | 'PRG4_No14' | 'LAYQDKGVLHNEVK' | 'Proteoglycan 4' |
| 14161 | 'PRG4_No15' | 'DQYYNIDVPSR' | 'Proteoglycan 4' |
| 14162 | 'PRG4_No16' | 'PVDGLTTLR' | 'Proteoglycan 4' |
| 14163 | 'PRG4_No17' | 'AIGPSQTHTIR' | 'Proteoglycan 4' |
| 14164 | 'PRG4_No18' | 'DATCNCDYNCQHYMECCPDFK' | 'Proteoglycan 4' |
| 14165 | 'PRG4_No19' | 'NWPESVYFFK' | 'Proteoglycan 4' |
| 14166 | 'PRG4_No20' | 'FTNDIKDAGYPKPIFK' | 'Proteoglycan 4' |
| 14167 | 'PRG4_No21' | 'VCTAELSCK' | 'Proteoglycan 4' |
| 14168 | 'PRG4_No22' | 'TITTTEIMNKPEETAKPK' | 'Proteoglycan 4' |
| 14169 | 'PRG4_No23' | 'KPDGYDYYAFSKDQYYNIDVPSR' | 'Proteoglycan 4' |

| | A | B | C |
|---|---|---|---|
| 1 | **Peptide Name** | **Peptide Sequence** | **Peptide Description** |
| 14170 | 'PRG4_No24' | 'ETSLTVNKETTVETK' | 'Proteoglycan 4' |
| 14171 | 'PRG4_No25' | 'GGSIQQYIYKQEPVQK' | 'Proteoglycan 4' |
| 14172 | 'PRG4_No26' | 'VPNQGIIINPMLSDETNICNGKPVDGLTTLR' | 'Proteoglycan 4' |
| 14173 | 'PRG4_No27' | 'GHYFWMLSPFSPPSPAR' | 'Proteoglycan 4' |
| 14174 | 'PRG4_No28' | 'DATCNCDYNCQHYMECCPDFK' | 'Proteoglycan 4' |
| 14175 | 'PRG4_No29' | 'ITEVWGIPSPIDTVFTR' | 'Proteoglycan 4' |
| 14176 | 'PRG4_No30' | 'SEDAGGAEGETPHMLLR' | 'Proteoglycan 4' |
| 14177 | 'PRG4_No31' | 'TITTTEIMNKPEETAKPK' | 'Proteoglycan 4' |
| 14178 | 'PRG4_No32' | 'NWPESVYFFKR' | 'Proteoglycan 4' |
| 14179 | 'PRG4_No33' | 'SEDAGGAEGETPHMLLR' | 'Proteoglycan 4' |
| 14180 | 'PRG4_No34' | 'RGGSIQQYIYKQEPVQK' | 'Proteoglycan 4' |
| 14181 | 'PRG4_No35' | 'RPALNYPVYGETTQVR' | 'Proteoglycan 4' |
| 14182 | 'PRG4_No36' | 'FTNDIKDAGYPK' | 'Proteoglycan 4' |
| 14183 | 'PRG4_No37' | 'GVLHNEVK' | 'Proteoglycan 4' |
| 14184 | 'PRG4_No38' | 'GGSIQQYIYK' | 'Proteoglycan 4' |
| 14185 | 'EXT2_No1' | 'FASVFGTMPLK' | 'Exostosin-2' |
| 14186 | 'EXT2_No2' | 'LPADSPIPER' | 'Exostosin-2' |
| 14187 | 'EXT2_No3' | 'ACLFVPSIDVLNQNTLR' | 'Exostosin-2' |
| 14188 | 'EXT2_No4' | 'MSDVYSILQSIPQR' | 'Exostosin-2' |
| 14189 | 'EXT2_No5' | 'YFNYLYTYK' | 'Exostosin-2' |
| 14190 | 'ATP2A3_No1' | 'MEAAHLLPAADVLR' | 'Sarcoplasmic/endoplasmic reticulum calcium ATPase 3' |
| 14191 | 'ATP2A3_No2' | 'IRDWGSGSDTLR' | 'Sarcoplasmic/endoplasmic reticulum calcium ATPase 3' |
| 14192 | 'ATP2A3_No3' | 'NAESAIEALKEYEPEMGK' | 'Sarcoplasmic/endoplasmic reticulum calcium ATPase 3' |
| 14193 | 'ATP2A3_No4' | 'SMSVYCTPTRPHPTGQGSK' | 'Sarcoplasmic/endoplasmic reticulum calcium ATPase 3' |
| 14194 | 'ATP2A3_No5' | 'VDQSILTGESVSVTK' | 'Sarcoplasmic/endoplasmic reticulum calcium ATPase 3' |
| 14195 | 'ATP2A3_No6' | 'MNVFDTDLQALSR' | 'Sarcoplasmic/endoplasmic reticulum calcium ATPase 3' |
| 14196 | 'ATP2A3_No7' | 'AVGVAVATGLHTELGK' | 'Sarcoplasmic/endoplasmic reticulum calcium ATPase 3' |
| 14197 | 'ATP2A3_No8' | 'LGIFGDTEDVAGK' | 'Sarcoplasmic/endoplasmic reticulum calcium ATPase 3' |
| 14198 | 'ATP2A3_No9' | 'SMSVYCTPTRPHPTGQGSK' | 'Sarcoplasmic/endoplasmic reticulum calcium ATPase 3' |
| 14199 | 'ATP2A3_No10' | 'HFSVTAEGGLSPAQVTGAR' | 'Sarcoplasmic/endoplasmic reticulum calcium ATPase 3' |
| 14200 | 'ATP2A3_No11' | 'YGPNELPSEEGK' | 'Sarcoplasmic/endoplasmic reticulum calcium ATPase 3' |
| 14201 | 'ATP2A3_No12' | 'CSEDNPLFAGIDCEVFESR' | 'Sarcoplasmic/endoplasmic reticulum calcium ATPase 3' |
| 14202 | 'ATP2A3_No13' | 'MNVFDTDLQALSR' | 'Sarcoplasmic/endoplasmic reticulum calcium ATPase 3' |
| 14203 | 'ATP2A3_No14' | 'NAESAIEALKEYEPEMGK' | 'Sarcoplasmic/endoplasmic reticulum calcium ATPase 3' |
| 14204 | 'ATP2A3_No15' | 'RLGIFGDTEDVAGK' | 'Sarcoplasmic/endoplasmic reticulum calcium ATPase 3' |
| 14205 | 'SYTL4_No1' | 'TLQFSVWHHGR' | 'Synaptotagmin-like protein 4' |
| 14206 | 'SYTL4_No2' | 'YEQQTQSLVVHVK' | 'Synaptotagmin-like protein 4' |
| 14207 | 'SYTL4_No3' | 'ISAESPTGLPSHK' | 'Synaptotagmin-like protein 4' |
| 14208 | 'SYTL4_No4' | 'SVIDLRPEDVVHESGSLGDR' | 'Synaptotagmin-like protein 4' |
| 14209 | 'KCTD12_No1' | 'CTVVSVPDSLLWR' | 'BTB/POZ domain-containing protein KCTD12' |
| 14210 | 'KCTD12_No2' | 'MFTQQQPQELAR' | 'BTB/POZ domain-containing protein KCTD12' |
| 14211 | 'KCTD12_No3' | 'DLQLVLPDYFPER' | 'BTB/POZ domain-containing protein KCTD12' |
| 14212 | 'KCTD12_No4' | 'EAEYFELPELVR' | 'BTB/POZ domain-containing protein KCTD12' |
| 14213 | 'ZG16B_No1' | 'VSVGLLLVK' | 'Zymogen granule protein 16 homolog B' |

| | A | B | C |
|---|---|---|---|
| 1 | **Peptide Name** | **Peptide Sequence** | **Peptide Description** |
| 14214 | 'ZG16B_No2' | 'LGALGGNTQEVTLQPGEYITK' | 'Zymogen granule protein 16 homolog B' |
| 14215 | 'ZG16B_No3' | 'YFSTTEDYDHEITGLR' | 'Zymogen granule protein 16 homolog B' |
| 14216 | 'CMTM5' | 'DRHPEEGVVAELQGFAVDK' | 'CKLF-like MARVEL transmembrane domain-containing protein 5' |
| 14217 | 'ABHD14B' | 'FSSETWQNLGTLHR' | 'Protein ABHD14B' |
| 14218 | 'CPB2_No1' | 'YSFTIELR' | 'Carboxypeptidase B2' |
| 14219 | 'CPB2_No2' | 'SFYANNHCIGTDLNR' | 'Carboxypeptidase B2' |
| 14220 | 'CPB2_No3' | 'DTGTYGFLLPER' | 'Carboxypeptidase B2' |
| 14221 | 'CPB2_No4' | 'YPLYVLK' | 'Carboxypeptidase B2' |
| 14222 | 'CPB2_No5' | 'NAIWIDCGIHAR' | 'Carboxypeptidase B2' |
| 14223 | 'CPB2_No6' | 'SKDHEELSLVASEAVR' | 'Carboxypeptidase B2' |
| 14224 | 'CPB2_No7' | 'IAWHVIR' | 'Carboxypeptidase B2' |
| 14225 | 'CPB2_No8' | 'AYISMHSYSQHIVFPYSYTR' | 'Carboxypeptidase B2' |
| 14226 | 'CPB2_No9' | 'LVDFYVMPVVNVDGYDYSWKK' | 'Carboxypeptidase B2' |
| 14227 | 'CPB2_No10' | 'YTHGHGSETLYLAPGGGDDWIYDLGIK' | 'Carboxypeptidase B2' |
| 14228 | 'CPB2_No11' | 'AYISMHSYSQHIVFPYSYTR' | 'Carboxypeptidase B2' |
| 14229 | 'CPB2_No12' | 'AVASFLRR' | 'Carboxypeptidase B2' |
| 14230 | 'CPB2_No13' | 'IHIGSSFEK' | 'Carboxypeptidase B2' |
| 14231 | 'CPB2_No14' | 'IHIGSSFEKYPLYVLK' | 'Carboxypeptidase B2' |
| 14232 | 'CPB2_No15' | 'SKSKDHEELSLVASEAVR' | 'Carboxypeptidase B2' |
| 14233 | 'CPB2_No16' | 'YIKPTCR' | 'Carboxypeptidase B2' |
| 14234 | 'CPB2_No17' | 'EAFAAVSK' | 'Carboxypeptidase B2' |
| 14235 | 'CPB2_No18' | 'DHEELSLVASEAVR' | 'Carboxypeptidase B2' |
| 14236 | 'CPB2_No19' | 'LVDFYVMPVVNVDGYDYSWKK' | 'Carboxypeptidase B2' |
| 14237 | 'CPB2_No20' | 'SFYANNHCIGTDLNRNFASK' | 'Carboxypeptidase B2' |
| 14238 | 'CPB2_No21' | 'HWCEEGASSSSCSETYCGLYPESEPEVK' | 'Carboxypeptidase B2' |
| 14239 | 'CPB2_No22' | 'EAFAAVSKIAWHVIRNV' | 'Carboxypeptidase B2' |
| 14240 | 'MEGF10_No1' | 'NDAVCSPVDGSCTCK' | 'Multiple epidermal growth factor-like domains protein 10' |
| 14241 | 'MEGF10_No2' | 'CELPCQDGTYGLNCAER' | 'Multiple epidermal growth factor-like domains protein 10' |
| 14242 | 'MEGF10_No3' | 'AGWHGVDCSIR' | 'Multiple epidermal growth factor-like domains protein 10' |
| 14243 | 'CNDP1_No1' | 'EWVAIESDSVQPVPR' | 'Beta-Ala-His dipeptidase' |
| 14244 | 'CNDP1_No2' | 'AIHLDLEEYR' | 'Beta-Ala-His dipeptidase' |
| 14245 | 'CNDP1_No3' | 'FLFDTKEEILMHLWR' | 'Beta-Ala-His dipeptidase' |
| 14246 | 'CNDP1_No4' | 'VFQYIDLHQDEFVQTLK' | 'Beta-Ala-His dipeptidase' |
| 14247 | 'CNDP1_No5' | 'EEILMHLWR' | 'Beta-Ala-His dipeptidase' |
| 14248 | 'CNDP1_No6' | 'MFQEIVHK' | 'Beta-Ala-His dipeptidase' |
| 14249 | 'CNDP1_No7' | 'MMAVAADTLQR' | 'Beta-Ala-His dipeptidase' |
| 14250 | 'CNDP1_No8' | 'GDGWLTDPYVLTEVDGK' | 'Beta-Ala-His dipeptidase' |
| 14251 | 'CNDP1_No9' | 'SVVLIPLGAVDDGEHSQNEK' | 'Beta-Ala-His dipeptidase' |
| 14252 | 'CNDP1_No10' | 'MMAVAADTLQR' | 'Beta-Ala-His dipeptidase' |
| 14253 | 'CNDP1_No11' | 'FIIEGMEEAGSVALEELVEKEKDR' | 'Beta-Ala-His dipeptidase' |
| 14254 | 'CNDP1_No12' | 'MMAVAADTLQR' | 'Beta-Ala-His dipeptidase' |
| 14255 | 'CNDP1_No13' | 'MMAVAADTLQR' | 'Beta-Ala-His dipeptidase' |
| 14256 | 'CNDP1_No14' | 'HLEDVFSKR' | 'Beta-Ala-His dipeptidase' |
| 14257 | 'CNDP1_No15' | 'ALEQDLPVNIK' | 'Beta-Ala-His dipeptidase' |

| | A | B | C |
|---|---|---|---|
| 1 | **Peptide Name** | **Peptide Sequence** | **Peptide Description** |
| 14258 | 'CNDP1_No16' | 'HLEDVFSK' | 'Beta-Ala-His dipeptidase' |
| 14259 | 'CNDP1_No17' | 'YPSLSIHGIEGAFDEPGTK' | 'Beta-Ala-His dipeptidase' |
| 14260 | 'CNDP1_No18' | 'MVVSMTLGLHPWIANIDDTQYLAAK' | 'Beta-Ala-His dipeptidase' |
| 14261 | 'CNDP1_No19' | 'TVFGTEPDMIR' | 'Beta-Ala-His dipeptidase' |
| 14262 | 'CNDP1_No20' | 'TVFGTEPDMIR' | 'Beta-Ala-His dipeptidase' |
| 14263 | 'CNDP1_No21' | 'TVFGTEPDMIRDGSTIPIAK' | 'Beta-Ala-His dipeptidase' |
| 14264 | 'CNDP1_No22' | 'GATDNKGPVLAWINAVSAFR' | 'Beta-Ala-His dipeptidase' |
| 14265 | 'CNDP1_No23' | 'FLFDTKEEILMHLWR' | 'Beta-Ala-His dipeptidase' |
| 14266 | 'CNDP1_No24' | 'GTVCFYGHLDVQPADR' | 'Beta-Ala-His dipeptidase' |
| 14267 | 'CNDP1_No25' | 'FIIEGMEEAGSVALEELVEKEK' | 'Beta-Ala-His dipeptidase' |
| 14268 | 'CNDP1_No26' | 'EEILMHLWR' | 'Beta-Ala-His dipeptidase' |
| 14269 | 'CNDP1_No27' | 'FIIEGMEEAGSVALEELVEK' | 'Beta-Ala-His dipeptidase' |
| 14270 | 'CNDP1_No28' | 'KPAITYGTR' | 'Beta-Ala-His dipeptidase' |
| 14271 | 'CNDP1_No29' | 'MVVSMTLGLHPWIANIDDTQYLAAK' | 'Beta-Ala-His dipeptidase' |
| 14272 | 'CNDP1_No30' | 'FIIEGMEEAGSVALEELVEK' | 'Beta-Ala-His dipeptidase' |
| 14273 | 'CNDP1_No31' | 'FIIEGMEEAGSVALEELVEKEK' | 'Beta-Ala-His dipeptidase' |
| 14274 | 'CNDP1_No32' | 'FIIEGMEEAGSVALEELVEKEKDR' | 'Beta-Ala-His dipeptidase' |
| 14275 | 'CNDP1_No33' | 'TVFGTEPDMIRDGSTIPIAK' | 'Beta-Ala-His dipeptidase' |
| 14276 | 'CNDP1_No34' | 'INRWNYIEGTK' | 'Beta-Ala-His dipeptidase' |
| 14277 | 'CNDP1_No35' | 'SVVLIPLGAVDDGEHSQNEKINR' | 'Beta-Ala-His dipeptidase' |
| 14278 | 'SERPINB12_No1' | 'LSMFVLLPSHSK' | 'Serpin B12' |
| 14279 | 'SERPINB12_No2' | 'ADLTGISPSPNLYLSK' | 'Serpin B12' |
| 14280 | 'SERPINB12_No3' | 'TFVEVDENGTQAAAATGAVVSER' | 'Serpin B12' |
| 14281 | 'SERPINB12_No4' | 'IGFIEEVK' | 'Serpin B12' |
| 14282 | 'PGLYRP2_No1' | 'PSLSHLLSQYYGAGVAR' | 'N-acetylmuramoyl-L-alanine amidase' |
| 14283 | 'PGLYRP2_No2' | 'AGLLRPDYALLGHR' | 'N-acetylmuramoyl-L-alanine amidase' |
| 14284 | 'PGLYRP2_No3' | 'DTLPSCAVR' | 'N-acetylmuramoyl-L-alanine amidase' |
| 14285 | 'PGLYRP2_No4' | 'GCPDVQASLPDAK' | 'N-acetylmuramoyl-L-alanine amidase' |
| 14286 | 'PGLYRP2_No5' | 'EFTEAFLGCPAIHPR' | 'N-acetylmuramoyl-L-alanine amidase' |
| 14287 | 'PGLYRP2_No6' | 'DGSPDVTTADIGANTPDATK' | 'N-acetylmuramoyl-L-alanine amidase' |
| 14288 | 'PGLYRP2_No7' | 'GSQTQSHPDLGTEGCWDQLSAPR' | 'N-acetylmuramoyl-L-alanine amidase' |
| 14289 | 'PGLYRP2_No8' | 'TWPHFTATVKPR' | 'N-acetylmuramoyl-L-alanine amidase' |
| 14290 | 'PGLYRP2_No9' | 'GWHWVGAHTLGHNSR' | 'N-acetylmuramoyl-L-alanine amidase' |
| 14291 | 'PGLYRP2_No10' | 'QNGAALTSASILAQQVWGTLVLLQR' | 'N-acetylmuramoyl-L-alanine amidase' |
| 14292 | 'PGLYRP2_No11' | 'HTASAWLMSAPNSGPHNR' | 'N-acetylmuramoyl-L-alanine amidase' |
| 14293 | 'PGLYRP2_No12' | 'HTASAWLMSAPNSGPHNR' | 'N-acetylmuramoyl-L-alanine amidase' |
| 14294 | 'PGLYRP2_No13' | 'QNGAALTSASILAQQVWGTLVLLQR' | 'N-acetylmuramoyl-L-alanine amidase' |
| 14295 | 'PGLYRP2_No14' | 'TPEPRPSLSHLLSQYYGAGVAR' | 'N-acetylmuramoyl-L-alanine amidase' |
| 14296 | 'PGLYRP2_No15' | 'WGAAPYR' | 'N-acetylmuramoyl-L-alanine amidase' |
| 14297 | 'PGLYRP2_No16' | 'TVRDTLPSCAVR' | 'N-acetylmuramoyl-L-alanine amidase' |
| 14298 | 'PGLYRP2_No17' | 'TDCPGDALFDLLR' | 'N-acetylmuramoyl-L-alanine amidase' |
| 14299 | 'PGLYRP2_No18' | 'HDVREGKEYGVVLAPDGSTVAVEPLLAGLEAGLQGR' | 'N-acetylmuramoyl-L-alanine amidase' |
| 14300 | 'PGLYRP2_No19' | 'RVINLPLDSMAAPWETGDTFPDVVAIAPDVR' | 'N-acetylmuramoyl-L-alanine amidase' |
| 14301 | 'PGLYRP2_No20' | 'TWPHFTATVK' | 'N-acetylmuramoyl-L-alanine amidase' |

| | A | B | C |
|---|---|---|---|
| 1 | **Peptide Name** | **Peptide Sequence** | **Peptide Description** |
| 14302 | 'PGLYRP2_No21' | 'EYGVVLAPDGSTVAVEPLLAGLEAGLQGR' | 'N-acetylmuramoyl-L-alanine amidase' |
| 14303 | 'PGLYRP2_No22' | 'EGKEYGVVLAPDGSTVAVEPLLAGLEAGLQGR' | 'N-acetylmuramoyl-L-alanine amidase' |
| 14304 | 'PGLYRP2_No23' | 'TRHTASAWLMSAPNSGPHNR' | 'N-acetylmuramoyl-L-alanine amidase' |
| 14305 | 'PGLYRP2_No24' | 'DGSPDVTTADIGANTPDATKGCPDVQASLPDAK' | 'N-acetylmuramoyl-L-alanine amidase' |
| 14306 | 'PGLYRP2_No25' | 'QLVRTDCPGDALFDLLR' | 'N-acetylmuramoyl-L-alanine amidase' |
| 14307 | 'PGLYRP2_No26' | 'YHQDTQGWGDIGYSFVVGSDGYVYEGR' | 'N-acetylmuramoyl-L-alanine amidase' |
| 14308 | 'PGLYRP2_No27' | 'TRHTASAWLMSAPNSGPHNR' | 'N-acetylmuramoyl-L-alanine amidase' |
| 14309 | 'PGLYRP2_No28' | 'VINLPLDSMAAPWETGDTFPDVVAIAPDVR' | 'N-acetylmuramoyl-L-alanine amidase' |
| 14310 | 'PGLYRP2_No29' | 'VINLPLDSMAAPWETGDTFPDVVAIAPDVR' | 'N-acetylmuramoyl-L-alanine amidase' |
| 14311 | 'PEBP4_No1' | 'IQGQELSAYQAPSPPAHSGFHR' | 'Phosphatidylethanolamine-binding protein 4' |
| 14312 | 'PEBP4_No2' | 'HWLVTDIK' | 'Phosphatidylethanolamine-binding protein 4' |
| 14313 | 'PEBP4_No3' | 'ITSWMEPIVK' | 'Phosphatidylethanolamine-binding protein 4' |
| 14314 | 'PEBP4_No4' | 'YQFFVYLQEGK' | 'Phosphatidylethanolamine-binding protein 4' |
| 14315 | 'MYADM_No1' | 'ALTQPLGLLR' | 'Myeloid-associated differentiation marker' |
| 14316 | 'MYADM_No2' | 'TTITTTTTSSSGLGSPMIVGSPR' | 'Myeloid-associated differentiation marker' |
| 14317 | 'MYADM_No3' | 'TTITTTTTSSSGLGSPMIVGSPR' | 'Myeloid-associated differentiation marker' |
| 14318 | 'CNN2_No1' | 'TWIEGLTGLSIGPDFQK' | 'Calponin-2' |
| 14319 | 'CNN2_No2' | 'SMQNWHQLENLSNFIK' | 'Calponin-2' |
| 14320 | 'CNN2_No3' | 'YDPQKEAELR' | 'Calponin-2' |
| 14321 | 'CNN2_No4' | 'SMQNWHQLENLSNFIK' | 'Calponin-2' |
| 14322 | 'CNN2_No5' | 'DGTILCTLMNK' | 'Calponin-2' |
| 14323 | 'CNN2_No6' | 'TKGLQSGVDIGVK' | 'Calponin-2' |
| 14324 | 'PARK7_No1' | 'DVVICPDASLEDAKK' | 'Protein/nucleic acid deglycase DJ-1' |
| 14325 | 'PARK7_No2' | 'VTVAGLAGKDPVQCSR' | 'Protein/nucleic acid deglycase DJ-1' |
| 14326 | 'PARK7_No3' | 'GAEEMETVIPVDVMR' | 'Protein/nucleic acid deglycase DJ-1' |
| 14327 | 'PARK7_No4' | 'EGPYDVVVLPGGNLGAQNLSESAAVK' | 'Protein/nucleic acid deglycase DJ-1' |
| 14328 | 'PARK7_No5' | 'KGLIAAICAGPTALLAHEIGFGSK' | 'Protein/nucleic acid deglycase DJ-1' |
| 14329 | 'PHB2_No1' | 'VLPSIVNEVLK' | 'Prohibitin-2' |
| 14330 | 'PHB2_No2' | 'IVQAEGEAEAAK' | 'Prohibitin-2' |
| 14331 | 'PHB2_No3' | 'IGGVQQDTILAEGLHFR' | 'Prohibitin-2' |
| 14332 | 'IL12RB2' | 'LILYKFDR' | 'Interleukin-12 receptor subunit beta-2' |
| 14333 | 'MGLL' | 'LTVPFLLLQGSADR' | 'Monoglyceride lipase' |
| 14334 | 'OLFM1_No1' | 'CICTVVAPQQTMCSR' | 'Noelin' |
| 14335 | 'OLFM1_No2' | 'FQSHIIIR' | 'Noelin' |
| 14336 | 'OLFM1_No3' | 'FGSWMTDPLAPEGDNR' | 'Noelin' |
| 14337 | 'OLFM1_No4' | 'LTGISDPVTVK' | 'Noelin' |
| 14338 | 'OLFM1_No5' | 'VWYMDGYHNNR' | 'Noelin' |
| 14339 | 'OLFM1_No6' | 'MDELRPLIPVLEEYKADAK' | 'Noelin' |
| 14340 | 'OLFM1_No7' | 'LDPVSLQTLQTWNTSYPK' | 'Noelin' |
| 14341 | 'OLFM1_No8' | 'MDELRPLIPVLEEYKADAK' | 'Noelin' |
| 14342 | 'OLFM1_No9' | 'YSHISMLDYNPK' | 'Noelin' |
| 14343 | 'OLFM1_No10' | 'DLQYVEKMENQMK' | 'Noelin' |
| 14344 | 'OLFM1_No11' | 'CICTVVAPQQTMCSR' | 'Noelin' |
| 14345 | 'RARRES2_No1' | 'LVHCPIETQVLR' | 'Retinoic acid receptor responder protein 2' |

| | A | B | C |
|---|---|---|---|
| 1 | **Peptide Name** | **Peptide Sequence** | **Peptide Description** |
| 14346 | 'RARRES2_No2' | 'AGEDPHSFYFPGQFAFSK' | 'Retinoic acid receptor responder protein 2' |
| 14347 | 'RARRES2_No3' | 'GLQVALEEFHK' | 'Retinoic acid receptor responder protein 2' |
| 14348 | 'RARRES2_No4' | 'RGLQVALEEFHK' | 'Retinoic acid receptor responder protein 2' |
| 14349 | 'MYOC_No1' | 'SGEGDTGCGELVWVGEPLTLR' | 'Myocilin' |
| 14350 | 'MYOC_No2' | 'KLFAWDNLNMVTYDIK' | 'Myocilin' |
| 14351 | 'MYOC_No3' | 'YSSMIDYNPLEK' | 'Myocilin' |
| 14352 | 'MYOC_No4' | 'DPKPTYPYTQETTWR' | 'Myocilin' |
| 14353 | 'MYOC_No5' | 'YSSMIDYNPLEK' | 'Myocilin' |
| 14354 | 'MYOC_No6' | 'LNPENLELEQTWETNIR' | 'Myocilin' |
| 14355 | 'AKAP9' | 'LIFLDSIK' | 'A-kinase anchor protein 9' |
| 14356 | 'TUBA1C' | 'AVCMLSNTTAVAEAWAR' | 'Tubulin alpha-1C chain' |
| 14357 | 'CORO1B' | 'DADPILISLR' | 'Coronin-1B' |
| 14358 | 'ERP44_No1' | 'VANILHDDCAFLSAFGDVSKPER' | 'Endoplasmic reticulum resident protein 44' |
| 14359 | 'ERP44_No2' | 'TPADCPVIAIDSFR' | 'Endoplasmic reticulum resident protein 44' |
| 14360 | 'ERP44_No3' | 'VDCDQHSDIAQR' | 'Endoplasmic reticulum resident protein 44' |
| 14361 | 'FUCA2_No1' | 'FFNANQWADIFQASGAK' | 'Plasma alpha-L-fucosidase' |
| 14362 | 'FUCA2_No2' | 'VNGEAIYETHTWR' | 'Plasma alpha-L-fucosidase' |
| 14363 | 'FUCA2_No3' | 'LVYAIFLK' | 'Plasma alpha-L-fucosidase' |
| 14364 | 'FUCA2_No4' | 'EAGISDYLTIEELVK' | 'Plasma alpha-L-fucosidase' |
| 14365 | 'FUCA2_No5' | 'WPTSGQLFLGHPK' | 'Plasma alpha-L-fucosidase' |
| 14366 | 'MENT_No1' | 'IILEDENDAMADADR' | 'Protein MENT' |
| 14367 | 'MENT_No2' | 'LVLMPWGPWHCHCK' | 'Protein MENT' |
| 14368 | 'MENT_No3' | 'TEHKPCTYQQCPCNR' | 'Protein MENT' |
| 14369 | 'MENT_No4' | 'SSAINEEDGSSEEGVVINAGK' | 'Protein MENT' |
| 14370 | 'MENT_No5' | 'IILEDENDAMADADR' | 'Protein MENT' |
| 14371 | 'MENT_No6' | 'FIANSQEPEIR' | 'Protein MENT' |
| 14372 | 'ACAT2_No1' | 'TIIGSFNGALAAVPVQDLGSTVIK' | 'Acetyl-CoA acetyltransferase, cytosolic' |
| 14373 | 'ACAT2_No2' | 'VNIEGGAIALGHPLGASGCR' | 'Acetyl-CoA acetyltransferase, cytosolic' |
| 14374 | 'COLEC11_No1' | 'AIGEMDNQVSQLTSELK' | 'Collectin-11' |
| 14375 | 'COLEC11_No2' | 'YADAQLSCQGR' | 'Collectin-11' |
| 14376 | 'COLEC11_No3' | 'GGTLSMPKDEAANGLMAAYLAQAGLAR' | 'Collectin-11' |
| 14377 | 'COLEC11_No4' | 'RYADAQLSCQGR' | 'Collectin-11' |
| 14378 | 'COLEC11_No5' | 'VFIGINDLEKEGAFVYSDHSPMR' | 'Collectin-11' |
| 14379 | 'COLEC11_No6' | 'AIGEMDNQVSQLTSELK' | 'Collectin-11' |
| 14380 | 'COLEC11_No7' | 'EGAFVYSDHSPMR' | 'Collectin-11' |
| 14381 | 'COLEC11_No8' | 'IYLLVKEEKR' | 'Collectin-11' |
| 14382 | 'COLEC11_No9' | 'IYLLVKEEK' | 'Collectin-11' |
| 14383 | 'COLEC11_No10' | 'DEAANGLMAAYLAQAGLAR' | 'Collectin-11' |
| 14384 | 'COLEC11_No11' | 'KAIGEMDNQVSQLTSELK' | 'Collectin-11' |
| 14385 | 'COLEC11_No12' | 'KAIGEMDNQVSQLTSELK' | 'Collectin-11' |
| 14386 | 'COLEC11_No13' | 'VFIGINDLEKEGAFVYSDHSPMR' | 'Collectin-11' |
| 14387 | 'COLEC11_No14' | 'GGTLSMPKDEAANGLMAAYLAQAGLAR' | 'Collectin-11' |
| 14388 | 'COLEC11_No15' | 'DEAANGLMAAYLAQAGLAR' | 'Collectin-11' |
| 14389 | 'COLEC11_No16' | 'EGAFVYSDHSPMR' | 'Collectin-11' |

| | A | B | C |
|---|---|---|---|
| 1 | **Peptide Name** | **Peptide Sequence** | **Peptide Description** |
| 14390 | 'COLEC11_No17' | 'AIGEMDNQVSQLTSELKFIK' | 'Collectin-11' |
| 14391 | 'CFHR5_No1' | 'WNPEVDCTEKR' | 'Complement factor H-related protein 5' |
| 14392 | 'CFHR5_No2' | 'CLDPCVVSEENMNK' | 'Complement factor H-related protein 5' |
| 14393 | 'CFHR5_No3' | 'AMISSPPFR' | 'Complement factor H-related protein 5' |
| 14394 | 'CFHR5_No4' | 'WNPEVDCTEK' | 'Complement factor H-related protein 5' |
| 14395 | 'CFHR5_No5' | 'CLDPCVVSEENMNKNNIQLK' | 'Complement factor H-related protein 5' |
| 14396 | 'CFHR5_No6' | 'VGSDSVQCYQFGWSPNFPTCK' | 'Complement factor H-related protein 5' |
| 14397 | 'CFHR5_No7' | 'GWSTPPICSFTK' | 'Complement factor H-related protein 5' |
| 14398 | 'CFHR5_No8' | 'KIQCVDGEWTTLPTCVEQVK' | 'Complement factor H-related protein 5' |
| 14399 | 'CFHR5_No9' | 'IQCVDGEWTTLPTCVEQVK' | 'Complement factor H-related protein 5' |
| 14400 | 'CFHR5_No10' | 'LQGSVTVTCR' | 'Complement factor H-related protein 5' |
| 14401 | 'CFHR5_No11' | 'GECHVPILEANVDAQPK' | 'Complement factor H-related protein 5' |
| 14402 | 'CFHR5_No12' | 'SCGPPPQLSNGEVK' | 'Complement factor H-related protein 5' |
| 14403 | 'CFHR5_No13' | 'AICQEGKFEYPICE' | 'Complement factor H-related protein 5' |
| 14404 | 'CFHR5_No14' | 'VAVLCKENYLLPEAK' | 'Complement factor H-related protein 5' |
| 14405 | 'CFHR5_No15' | 'CLDPCVVSEENMNK' | 'Complement factor H-related protein 5' |
| 14406 | 'CFHR5_No16' | 'TGDAVEFQCKFPHK' | 'Complement factor H-related protein 5' |
| 14407 | 'CFHR5_No17' | 'CLDPCVVSEENMNKNNIQLK' | 'Complement factor H-related protein 5' |
| 14408 | 'CFHR5_No18' | 'ENYLLPEAK' | 'Complement factor H-related protein 5' |
| 14409 | 'CFHR5_No19' | 'MCSFPFVK' | 'Complement factor H-related protein 5' |
| 14410 | 'CFHR5_No20' | 'TCGYIPELEYGYVQPSVPPYQHGVSVEVNCR' | 'Complement factor H-related protein 5' |
| 14411 | 'CFHR5_No21' | 'AMISSPPFR' | 'Complement factor H-related protein 5' |
| 14412 | 'CADM1_No1' | 'YFCQLYTDPPQESYTTITVLVPPR' | 'Cell adhesion molecule 1' |
| 14413 | 'CADM1_No2' | 'EGDALELTCEAIGKPQPVMVTWVR' | 'Cell adhesion molecule 1' |
| 14414 | 'RTN4R_No1' | 'CCQPDAADK' | 'Reticulon-4 receptor' |
| 14415 | 'RTN4R_No2' | 'VTTSCPQQGLQAVPVGIPAASQR' | 'Reticulon-4 receptor' |
| 14416 | 'NIF3L1_No1' | 'ALMQVVDFLSR' | 'NIF3-like protein 1' |
| 14417 | 'NIF3L1_No2' | 'INIILSETDRDPLQVV' | 'NIF3-like protein 1' |
| 14418 | 'NIF3L1_No3' | 'TEILSLEKPLLLHTGMGR' | 'NIF3-like protein 1' |
| 14419 | 'UBA5' | 'LLLFDYDK' | 'Ubiquitin-like modifier-activating enzyme 5' |
| 14420 | 'HDHD2_No1' | 'LLLDGAPLIAIHK' | 'Haloacid dehalogenase-like hydrolase domain-containing protein 2' |
| 14421 | 'HDHD2_No2' | 'TFFLEALR' | 'Haloacid dehalogenase-like hydrolase domain-containing protein 2' |
| 14422 | 'RAB1B_No1' | 'MGPGAASGGERPNLK' | 'Ras-related protein Rab-1B' |
| 14423 | 'RAB1B_No2' | 'MGPGAASGGERPNLK' | 'Ras-related protein Rab-1B' |
| 14424 | 'RAB1B_No3' | 'MNPEYDYLFK' | 'Ras-related protein Rab-1B' |
| 14425 | 'RAB1B_No4' | 'QWLQEIDR' | 'Ras-related protein Rab-1B' |
| 14426 | 'RAB1B_No5' | 'MNPEYDYLFK' | 'Ras-related protein Rab-1B' |
| 14427 | 'SH3BGRL3_No1' | 'IQYQLVDISQDNALRDEMR' | 'SH3 domain-binding glutamic acid-rich-like protein 3' |
| 14428 | 'SH3BGRL3_No2' | 'IQYQLVDISQDNALRDEMR' | 'SH3 domain-binding glutamic acid-rich-like protein 3' |
| 14429 | 'SH3BGRL3_No3' | 'RIQYQLVDISQDNALRDEMR' | 'SH3 domain-binding glutamic acid-rich-like protein 3' |
| 14430 | 'DPEP2_No1' | 'NVPDDILQLLK' | 'Dipeptidase 2' |
| 14431 | 'DPEP2_No2' | 'ALEVSQAPVIFSHSAAR' | 'Dipeptidase 2' |
| 14432 | 'DPEP2_No3' | 'GWSEEELQGVLR' | 'Dipeptidase 2' |
| 14433 | 'DPEP2_No4' | 'TFYMLGVR' | 'Dipeptidase 2' |

| | A | B | C |
|---|---|---|---|
| 1 | **Peptide Name** | **Peptide Sequence** | **Peptide Description** |
| 14434 | 'DPEP2_No5' | 'DFPLVDGHNDLPLVLR' | 'Dipeptidase 2' |
| 14435 | 'DPEP2_No6' | 'LTLEQIDLIR' | 'Dipeptidase 2' |
| 14436 | 'DPEP2_No7' | 'DGLVGAQFWSAYVPCQTQDR' | 'Dipeptidase 2' |
| 14437 | 'DPEP2_No8' | 'LACLIGVEGGHSLDNSLSILR' | 'Dipeptidase 2' |
| 14438 | 'DPEP2_No9' | 'NVPDDILQLLKK' | 'Dipeptidase 2' |
| 14439 | 'DPEP2_No10' | 'YLTLTHTCNTPWAESSAK' | 'Dipeptidase 2' |
| 14440 | 'TUBB1_No1' | 'YLTVACIFR' | 'Tubulin beta-1 chain' |
| 14441 | 'TUBB1_No2' | 'AVLVDLEPGTMDSIR' | 'Tubulin beta-1 chain' |
| 14442 | 'TUBB1_No3' | 'ALSVAELTQQMFDAR' | 'Tubulin beta-1 chain' |
| 14443 | 'TUBB1_No4' | 'VSEHFSAMFK' | 'Tubulin beta-1 chain' |
| 14444 | 'TUBB1_No5' | 'AVLEEDEEVTEEAEMEPEDKGH' | 'Tubulin beta-1 chain' |
| 14445 | 'TUBB1_No6' | 'FWEMIGEEHGIDLAGSDR' | 'Tubulin beta-1 chain' |
| 14446 | 'TUBB1_No7' | 'EIVHIQIGQCGNQIGAK' | 'Tubulin beta-1 chain' |
| 14447 | 'TUBB1_No8' | 'NSSCFVEWIPNNVK' | 'Tubulin beta-1 chain' |
| 14448 | 'TUBB1_No9' | 'LHFFMPGFAPLTAQGSQQYR' | 'Tubulin beta-1 chain' |
| 14449 | 'TUBB1_No10' | 'LGALFQPDSFVHGNSGAGNNWAK' | 'Tubulin beta-1 chain' |
| 14450 | 'TUBB1_No11' | 'LHFFMPGFAPLTAQGSQQYR' | 'Tubulin beta-1 chain' |
| 14451 | 'TUBB1_No12' | 'AVLEEDEEVTEEAEMEPEDKGH' | 'Tubulin beta-1 chain' |
| 14452 | 'TUBB1_No13' | 'LTTPTYGDLNHLVSLTMSGITTSLR' | 'Tubulin beta-1 chain' |
| 14453 | 'TUBB1_No14' | 'GHYTEGAELIENVLEVVR' | 'Tubulin beta-1 chain' |
| 14454 | 'TUBB1_No15' | 'VSEHFSAMFK' | 'Tubulin beta-1 chain' |
| 14455 | 'TUBB1_No16' | 'IMNSFSVMPSPK' | 'Tubulin beta-1 chain' |
| 14456 | 'TUBB1_No17' | 'IMNSFSVMPSPK' | 'Tubulin beta-1 chain' |
| 14457 | 'TUBB1_No18' | 'AVLVDLEPGTMDSIR' | 'Tubulin beta-1 chain' |
| 14458 | 'TUBB1_No19' | 'GLSMAATFIGNNTAIQEIFNR' | 'Tubulin beta-1 chain' |
| 14459 | 'TUBB1_No20' | 'ALSVAELTQQMFDAR' | 'Tubulin beta-1 chain' |
| 14460 | 'TUBB1_No21' | 'ISVYYNEAYGRK' | 'Tubulin beta-1 chain' |
| 14461 | 'TUBB1_No22' | 'MREIVHIQIGQCGNQIGAK' | 'Tubulin beta-1 chain' |
| 14462 | 'TUBB1_No23' | 'EVDQQLLSVQTR' | 'Tubulin beta-1 chain' |
| 14463 | 'TUBB1_No24' | 'VSEHFSAMFKR' | 'Tubulin beta-1 chain' |
| 14464 | 'GLIPR2_No1' | 'NYNFQQPGFTSGTGHFTAMVWK' | 'Golgi-associated plant pathogenesis-related protein 1' |
| 14465 | 'GLIPR2_No2' | 'YFPAGNVVNEGFFEENVLPPKK' | 'Golgi-associated plant pathogenesis-related protein 1' |
| 14466 | 'GLIPR2_No3' | 'GQCGENLAWASYDQTGK' | 'Golgi-associated plant pathogenesis-related protein 1' |
| 14467 | 'GLIPR2_No4' | 'YFPAGNVVNEGFFEENVLPPK' | 'Golgi-associated plant pathogenesis-related protein 1' |
| 14468 | 'EHD1_No1' | 'LLPLEEHYR' | 'EH domain-containing protein 1' |
| 14469 | 'EHD1_No2' | 'LNAFGNAFLNR' | 'EH domain-containing protein 1' |
| 14470 | 'EHD1_No3' | 'VYIGSFWSHPLLIPDNRK' | 'EH domain-containing protein 1' |
| 14471 | 'EHD1_No4' | 'EPELFQTVAEGLR' | 'EH domain-containing protein 1' |
| 14472 | 'EHD1_No5' | 'LLDTVDDMLANDIAR' | 'EH domain-containing protein 1' |
| 14473 | 'EHD1_No6' | 'ELVNNLGEIYQK' | 'EH domain-containing protein 1' |
| 14474 | 'EHD1_No7' | 'LADVDKDGLLDDEEFALANHLIK' | 'EH domain-containing protein 1' |
| 14475 | 'EHD1_No8' | 'VYIGSFWSHPLLIPDNR' | 'EH domain-containing protein 1' |
| 14476 | 'EHD1_No9' | 'LLDTVDDMLANDIAR' | 'EH domain-containing protein 1' |
| 14477 | 'EHD1_No10' | 'LDISDEFSEVIK' | 'EH domain-containing protein 1' |

| | A | B | C |
|---|---|---|---|
| 1 | **Peptide Name** | **Peptide Sequence** | **Peptide Description** |
| 14478 | 'EHD1_No11' | 'ADQIETQQLMR' | 'EH domain-containing protein 1' |
| 14479 | 'EHD1_No12' | 'EHQISPGDFPSLR' | 'EH domain-containing protein 1' |
| 14480 | 'MVB12B' | 'HISLTLPATFR' | 'Multivesicular body subunit 12B' |
| 14481 | 'MMRN2_No1' | 'VWFELTQGSITK' | 'Multimerin-2' |
| 14482 | 'MMRN2_No2' | 'TPVCTTGQGSGSTATVFAMAELQK' | 'Multimerin-2' |
| 14483 | 'MMRN2_No3' | 'SLSGTAFGGFLMFK' | 'Multimerin-2' |
| 14484 | 'MMRN2_No4' | 'SLSGTAFGGFLMFK' | 'Multimerin-2' |
| 14485 | 'MMRN2_No5' | 'VWFELTQGSITKR' | 'Multimerin-2' |
| 14486 | 'MMRN2_No6' | 'TPVCTTGQGSGSTATVFAMAELQK' | 'Multimerin-2' |
| 14487 | 'COG4' | 'DYHQQFR' | 'Conserved oligomeric Golgi complex subunit 4' |
| 14488 | 'CDHR5_No1' | 'VFVSVLDVNDNAPEFPFK' | 'Cadherin-related family member 5' |
| 14489 | 'CDHR5_No2' | 'GINQPIIYSIFR' | 'Cadherin-related family member 5' |
| 14490 | 'CDHR5_No3' | 'IQAQDPEFSDLNSAITYR' | 'Cadherin-related family member 5' |
| 14491 | 'CDHR5_No4' | 'SLLEAQLLCQSGGTLVTQLR' | 'Cadherin-related family member 5' |
| 14492 | 'CDHR5_No5' | 'GAGAGVVVKDAAAPSQPLR' | 'Cadherin-related family member 5' |
| 14493 | 'PARVB_No1' | 'LQTVLEAVHDLLRPR' | 'Beta-parvin' |
| 14494 | 'PARVB_No2' | 'VLYNLFTK' | 'Beta-parvin' |
| 14495 | 'PARVB_No3' | 'DAFDTLFDHAPDKLSVVK' | 'Beta-parvin' |
| 14496 | 'PARVB_No4' | 'QLEEDLYDGQVLQK' | 'Beta-parvin' |
| 14497 | 'PARVB_No5' | 'TMIDPTSKEDPK' | 'Beta-parvin' |
| 14498 | 'PARVB_No6' | 'LNVAEVTQSEIGQK' | 'Beta-parvin' |
| 14499 | 'PARVB_No7' | 'ARPEDVVNLDLK' | 'Beta-parvin' |
| 14500 | 'PARVB_No8' | 'EGLLHSSHISEELTTTTEMMMGR' | 'Beta-parvin' |
| 14501 | 'PARVB_No9' | 'QLEEDLYDGQVLQK' | 'Beta-parvin' |
| 14502 | 'PARVB_No10' | 'DAFDTLFDHAPDKLSVVKK' | 'Beta-parvin' |
| 14503 | 'PARVB_No11' | 'QKLQTVLEAVHDLLRPR' | 'Beta-parvin' |
| 14504 | 'PARVB_No12' | 'WSVDSIHGK' | 'Beta-parvin' |
| 14505 | 'PARVB_No13' | 'DAFDTLFDHAPDK' | 'Beta-parvin' |
| 14506 | 'PARVB_No14' | 'LPEHVTVQVVVVR' | 'Beta-parvin' |
| 14507 | 'PARVB_No15' | 'TMIDPTSKEDPKFK' | 'Beta-parvin' |
| 14508 | 'PARVB_No16' | 'QKLQTVLEAVHDLLRPR' | 'Beta-parvin' |
| 14509 | 'CD248_No1' | 'LLWIGLQR' | 'Endosialin' |
| 14510 | 'CD248_No2' | 'GASLLCVKQPEGGVGWSR' | 'Endosialin' |
| 14511 | 'CD248_No3' | 'AGPLCLGTGCSPDNGGCEHECVEEVDGHVSCR' | 'Endosialin' |
| 14512 | 'CD248_No4' | 'TFLEAWR' | 'Endosialin' |
| 14513 | 'CD248_No5' | 'VDSLVGAGPASR' | 'Endosialin' |
| 14514 | 'APMAP_No1' | 'LFENQLVGPESIAHIGDVMFTGTADGR' | 'Adipocyte plasma membrane-associated protein' |
| 14515 | 'APMAP_No2' | 'LFSQETVMK' | 'Adipocyte plasma membrane-associated protein' |
| 14516 | 'APMAP_No3' | 'VLLDQLR' | 'Adipocyte plasma membrane-associated protein' |
| 14517 | 'APMAP_No4' | 'GLFEVNPWKR' | 'Adipocyte plasma membrane-associated protein' |
| 14518 | 'APMAP_No5' | 'VVKLENGEIETIAR' | 'Adipocyte plasma membrane-associated protein' |
| 14519 | 'APMAP_No6' | 'SLHDPDGLVATYISEVHEHDGHLYLGSFR' | 'Adipocyte plasma membrane-associated protein' |
| 14520 | 'APMAP_No7' | 'TRDDEPVCGRPLGIR' | 'Adipocyte plasma membrane-associated protein' |
| 14521 | 'APMAP_No8' | 'GLFEVNPWK' | 'Adipocyte plasma membrane-associated protein' |

| | A | B | C |
|---|---|---|---|
| 1 | **Peptide Name** | **Peptide Sequence** | **Peptide Description** |
| 14522 | 'APMAP_No9' | 'LLLSSETPIEGK' | 'Adipocyte plasma membrane-associated protein' |
| 14523 | 'APMAP_No10' | 'EPPLLLGVLHPNTK' | 'Adipocyte plasma membrane-associated protein' |
| 14524 | 'APMAP_No11' | 'RDYLLLVMEGTDDGR' | 'Adipocyte plasma membrane-associated protein' |
| 14525 | 'APMAP_No12' | 'LLEYDTVTR' | 'Adipocyte plasma membrane-associated protein' |
| 14526 | 'APMAP_No13' | 'LENGEIETIAR' | 'Adipocyte plasma membrane-associated protein' |
| 14527 | 'APMAP_No14' | 'LFSQETVMK' | 'Adipocyte plasma membrane-associated protein' |
| 14528 | 'APMAP_No15' | 'LFENQLVGPESIAHIGDVMFTGTADGR' | 'Adipocyte plasma membrane-associated protein' |
| 14529 | 'APMAP_No16' | 'VYVSGLMK' | 'Adipocyte plasma membrane-associated protein' |
| 14530 | 'APMAP_No17' | 'GGADLFVENMPGFPDNIR' | 'Adipocyte plasma membrane-associated protein' |
| 14531 | 'APMAP_No18' | 'YSLVLELSDSGAFR' | 'Adipocyte plasma membrane-associated protein' |
| 14532 | 'BPIFA1_No1' | 'LQVNTPLVGASLLR' | 'BPI fold-containing family A member 1' |
| 14533 | 'BPIFA1_No2' | 'VTSVIPGLNNIIDIK' | 'BPI fold-containing family A member 1' |
| 14534 | 'IL1RAP_No1' | 'VAFPLEVVQK' | 'Interleukin-1 receptor accessory protein' |
| 14535 | 'IL1RAP_No2' | 'QDRDLEEPINFR' | 'Interleukin-1 receptor accessory protein' |
| 14536 | 'IL1RAP_No3' | 'NEVWWTIDGK' | 'Interleukin-1 receptor accessory protein' |
| 14537 | 'IL1RAP_No4' | 'ITCPNVDGYFPSSVKPTITWYMGCYK' | 'Interleukin-1 receptor accessory protein' |
| 14538 | 'IL1RAP_No5' | 'QIQVFEDEPAR' | 'Interleukin-1 receptor accessory protein' |
| 14539 | 'IL1RAP_No6' | 'KPDDITIDVTINESISHSR' | 'Interleukin-1 receptor accessory protein' |
| 14540 | 'IL1RAP_No7' | 'QDRDLEEPINFR' | 'Interleukin-1 receptor accessory protein' |
| 14541 | 'IL1RAP_No8' | 'NAVPPVIHSPNDHVVYEK' | 'Interleukin-1 receptor accessory protein' |
| 14542 | 'IL1RAP_No9' | 'DSCFNSPMKLPVHK' | 'Interleukin-1 receptor accessory protein' |
| 14543 | 'IL1RAP_No10' | 'TEDETRTQILSIKK' | 'Interleukin-1 receptor accessory protein' |
| 14544 | 'IL1RAP_No11' | 'QDRDLEEPINFRLPENR' | 'Interleukin-1 receptor accessory protein' |
| 14545 | 'IL1RAP_No12' | 'CPLFEHFLK' | 'Interleukin-1 receptor accessory protein' |
| 14546 | 'IL1RAP_No13' | 'LYIEYGIQR' | 'Interleukin-1 receptor accessory protein' |
| 14547 | 'IL1RAP_No14' | 'QDRDLEEPINFRLPENR' | 'Interleukin-1 receptor accessory protein' |
| 14548 | 'IL1RAP_No15' | 'DSCFNSPMKLPVHK' | 'Interleukin-1 receptor accessory protein' |
| 14549 | 'IL1RAP_No16' | 'CDDWGLDTMR' | 'Interleukin-1 receptor accessory protein' |
| 14550 | 'SEMA4B_No1' | 'ISLPLGSEERPFLR' | 'Semaphorin-4B' |
| 14551 | 'SEMA4B_No2' | 'ETQQWYTVTHPVPTPR' | 'Semaphorin-4B' |
| 14552 | 'SEMA4B_No3' | 'VPGLHHTYDVLFLGTGDGR' | 'Semaphorin-4B' |
| 14553 | 'SEMA4B_No4' | 'GTTEGSAVCVFTMK' | 'Semaphorin-4B' |
| 14554 | 'SEMA4B_No5' | 'HVSLYQPQLATRPWIQDIEGASAK' | 'Semaphorin-4B' |
| 14555 | 'CD93_No1' | 'WSEGPCGSPGSPGSNIEGFVCK' | 'Complement component C1q receptor' |
| 14556 | 'CD93_No2' | 'EKAPDVFDWGSSGPLCVSPK' | 'Complement component C1q receptor' |
| 14557 | 'CD93_No3' | 'FWIGLQR' | 'Complement component C1q receptor' |
| 14558 | 'CD93_No4' | 'LSAAEAQNHCNQNGGNLATVK' | 'Complement component C1q receptor' |
| 14559 | 'CD93_No5' | 'LLDDLVTCASR' | 'Complement component C1q receptor' |
| 14560 | 'CD93_No6' | 'APDVFDWGSSGPLCVSPK' | 'Complement component C1q receptor' |
| 14561 | 'SPINK5_No1' | 'FFQSLDGIMFINK' | 'Serine protease inhibitor Kazal-type 5' |
| 14562 | 'SPINK5_No2' | 'SSNPEQDVCSAFRPFVR' | 'Serine protease inhibitor Kazal-type 5' |
| 14563 | 'SPINK5_No3' | 'AVFLTEALER' | 'Serine protease inhibitor Kazal-type 5' |
| 14564 | 'CASS4_No1' | 'KLSLPEIPSYGFLVPR' | 'Cas scaffolding protein family member 4' |
| 14565 | 'CASS4_No2' | 'GVHGTACNLTDSNLQNR' | 'Cas scaffolding protein family member 4' |

| | A | B | C |
|---|---|---|---|
| 1 | **Peptide Name** | **Peptide Sequence** | **Peptide Description** |
| 14566 | 'CASS4_No3' | 'GTFPLDEDVSYKVPSSFLIPR' | 'Cas scaffolding protein family member 4' |
| 14567 | 'CRTAC1_No1' | 'GNQGFNNNWLR' | 'Cartilage acidic protein 1' |
| 14568 | 'CRTAC1_No2' | 'GVASLFAGR' | 'Cartilage acidic protein 1' |
| 14569 | 'CRTAC1_No3' | 'VDIVYGNWNGPHR' | 'Cartilage acidic protein 1' |
| 14570 | 'CRTAC1_No4' | 'GVSVGPILSSSASDIFCDNENGPNFLFHNR' | 'Cartilage acidic protein 1' |
| 14571 | 'CRTAC1_No5' | 'DGKVDIVYGNWNGPHR' | 'Cartilage acidic protein 1' |
| 14572 | 'CRTAC1_No6' | 'IIDGGSGYLCEMEPVAHFGLGK' | 'Cartilage acidic protein 1' |
| 14573 | 'CRTAC1_No7' | 'FSMPSPVR' | 'Cartilage acidic protein 1' |
| 14574 | 'CRTAC1_No8' | 'REHGDPLIEELNPGDALEPEGR' | 'Cartilage acidic protein 1' |
| 14575 | 'CRTAC1_No9' | 'WEDILSDEVNVAR' | 'Cartilage acidic protein 1' |
| 14576 | 'CRTAC1_No10' | 'EHGDPLIEELNPGDALEPEGR' | 'Cartilage acidic protein 1' |
| 14577 | 'CRTAC1_No11' | 'FSMPSPVR' | 'Cartilage acidic protein 1' |
| 14578 | 'CRTAC1_No12' | 'DKPVCVNTYGSYR' | 'Cartilage acidic protein 1' |
| 14579 | 'CRTAC1_No13' | 'GILALRDVAAEAGVSK' | 'Cartilage acidic protein 1' |
| 14580 | 'CRTAC1_No14' | 'NNRWEDILSDEVNVAR' | 'Cartilage acidic protein 1' |
| 14581 | 'CRTAC1_No15' | 'GDGTFVDAAASAGVDDPHQHGR' | 'Cartilage acidic protein 1' |
| 14582 | 'CRTAC1_No16' | 'RLVNIAVDER' | 'Cartilage acidic protein 1' |
| 14583 | 'CRTAC1_No17' | 'NVASGEMNSVLEILYPR' | 'Cartilage acidic protein 1' |
| 14584 | 'CRTAC1_No18' | 'GVALADFNRDGK' | 'Cartilage acidic protein 1' |
| 14585 | 'CRTAC1_No19' | 'SSPYYALRDR' | 'Cartilage acidic protein 1' |
| 14586 | 'CRTAC1_No20' | 'FRNNRWEDILSDEVNVAR' | 'Cartilage acidic protein 1' |
| 14587 | 'CRTAC1_No21' | 'GILALRDVAAEAGVSKYTGGR' | 'Cartilage acidic protein 1' |
| 14588 | 'CRTAC1_No22' | 'IIDGGSGYLCEMEPVAHFGLGK' | 'Cartilage acidic protein 1' |
| 14589 | 'CRTAC1_No23' | 'DEASSVEVTWPDGK' | 'Cartilage acidic protein 1' |
| 14590 | 'CRTAC1_No24' | 'NVASGEMNSVLEILYPR' | 'Cartilage acidic protein 1' |
| 14591 | 'RTN4_No1' | 'HQAQIDHYLGLANK' | 'Reticulon-4' |
| 14592 | 'RTN4_No2' | 'YSNSALGHVNCTIK' | 'Reticulon-4' |
| 14593 | 'RTN4_No3' | 'GPLPAAPPVAPER' | 'Reticulon-4' |
| 14594 | 'RTN4_No4' | 'AYLESEVAISEELVQK' | 'Reticulon-4' |
| 14595 | 'XPNPEP1_No1' | 'VGVDPLIIPTDYWK' | 'Xaa-Pro aminopeptidase 1' |
| 14596 | 'XPNPEP1_No2' | 'GSLTFEPLTLVPIQTK' | 'Xaa-Pro aminopeptidase 1' |
| 14597 | 'PDLIM7_No1' | 'LQGGKDFNVPLSISR' | 'PDZ and LIM domain protein 7' |
| 14598 | 'PDLIM7_No2' | 'VVLEGPAPWGFR' | 'PDZ and LIM domain protein 7' |
| 14599 | 'PDLIM7_No3' | 'YLVALGHAYHPEEFVCSQCGK' | 'PDZ and LIM domain protein 7' |
| 14600 | 'PDLIM7_No4' | 'YAPDKTSTVLTR' | 'PDZ and LIM domain protein 7' |
| 14601 | 'SAR1A' | 'LVFLGLDNAGK' | 'GTP-binding protein SAR1a' |
| 14602 | 'GKN1_No1' | 'EVMPSIQSLDALVK' | 'Gastrokine-1' |
| 14603 | 'GKN1_No2' | 'GLMYSVNPNKVDDLSK' | 'Gastrokine-1' |
| 14604 | 'GKN1_No3' | 'MNKEVMPSIQSLDALVK' | 'Gastrokine-1' |
| 14605 | 'SACM1L_No1' | 'TNVIQSLLAR' | 'Phosphatidylinositide phosphatase SAC1' |
| 14606 | 'SACM1L_No2' | 'HFDSQVIIYGK' | 'Phosphatidylinositide phosphatase SAC1' |
| 14607 | 'SACM1L_No3' | 'LSNTSPEFQEMSLLER' | 'Phosphatidylinositide phosphatase SAC1' |
| 14608 | 'CBLN4_No1' | 'CLVVCDSNPATDSK' | 'Cerebellin-4' |
| 14609 | 'CBLN4_No2' | 'EAATNGVLLYLDKEDK' | 'Cerebellin-4' |

| | A | B | C |
|---|---|---|---|
| 1 | **Peptide Name** | **Peptide Sequence** | **Peptide Description** |
| 14610 | 'STAB1_No1' | 'ILLGPEGVPLQR' | 'Stabilin-1' |
| 14611 | 'STAB1_No2' | 'LGGQEVATLNPTTR' | 'Stabilin-1' |
| 14612 | 'STAB1_No3' | 'GGCSENAECVPGSLGTHHCTCHK' | 'Stabilin-1' |
| 14613 | 'STAB1_No4' | 'TTFVTHVPCTSCAAIK' | 'Stabilin-1' |
| 14614 | 'STAB1_No5' | 'ANNIAANGVFHVVTGLR' | 'Stabilin-1' |
| 14615 | 'STAB1_No6' | 'GGCHTDALCSYVGPGQSR' | 'Stabilin-1' |
| 14616 | 'STAB1_No7' | 'QHIIAGQHILEDTR' | 'Stabilin-1' |
| 14617 | 'STAB1_No8' | 'LGFAGDGYQCSPIDPCR' | 'Stabilin-1' |
| 14618 | 'STAB1_No9' | 'ACYGHLLHEVQK' | 'Stabilin-1' |
| 14619 | 'STAB1_No10' | 'LIYLFTAGLSK' | 'Stabilin-1' |
| 14620 | 'TUBA8_No1' | 'INDDDSFTTFFSETGNGK' | 'Tubulin alpha-8 chain' |
| 14621 | 'TUBA8_No2' | 'IHFPLVTYAPIISAEK' | 'Tubulin alpha-8 chain' |
| 14622 | 'TUBA8_No3' | 'AVMIDLEPTVVDEVR' | 'Tubulin alpha-8 chain' |
| 14623 | 'TUBA8_No4' | 'ESIDLVLDR' | 'Tubulin alpha-8 chain' |
| 14624 | 'TUBA8_No5' | 'AVMIDLEPTVVDEVR' | 'Tubulin alpha-8 chain' |
| 14625 | 'B3GNT2_No1' | 'KPFLLLAIK' | 'N-acetyllactosaminide beta-1,3-N-acetylglucosaminyltransferase 2' |
| 14626 | 'B3GNT2_No2' | 'KPQEMIDIWSQLQSAHLK' | 'N-acetyllactosaminide beta-1,3-N-acetylglucosaminyltransferase 2' |
| 14627 | 'B3GNT2_No3' | 'LYHITDQVHLYPIDDVYTGMCLQK' | 'N-acetyllactosaminide beta-1,3-N-acetylglucosaminyltransferase 2' |
| 14628 | 'B3GNT2_No4' | 'NNICSYVDLMLVHSR' | 'N-acetyllactosaminide beta-1,3-N-acetylglucosaminyltransferase 2' |
| 14629 | 'B3GNT2_No5' | 'NNICSYVDLMLVHSR' | 'N-acetyllactosaminide beta-1,3-N-acetylglucosaminyltransferase 2' |
| 14630 | 'ERAP1_No1' | 'ASLINNAFQLVSIGK' | 'Endoplasmic reticulum aminopeptidase 1' |
| 14631 | 'ERAP1_No2' | 'NPVGYPLAWQFLR' | 'Endoplasmic reticulum aminopeptidase 1' |
| 14632 | 'ERAP1_No3' | 'TQEFPQILTLIGR' | 'Endoplasmic reticulum aminopeptidase 1' |
| 14633 | 'ERAP1_No4' | 'TDVLILPEEVEWIK' | 'Endoplasmic reticulum aminopeptidase 1' |
| 14634 | 'ERAP1_No5' | 'GSDGAPDTGYLWHVPLTFITSK' | 'Endoplasmic reticulum aminopeptidase 1' |
| 14635 | 'ERAP1_No6' | 'FELGSSSIAHMVMGTTNQFSTR' | 'Endoplasmic reticulum aminopeptidase 1' |
| 14636 | 'ERAP1_No7' | 'GACILNMLR' | 'Endoplasmic reticulum aminopeptidase 1' |
| 14637 | 'ERAP1_No8' | 'SVTVAEGLIEDHFDVTVK' | 'Endoplasmic reticulum aminopeptidase 1' |
| 14638 | 'ERAP1_No9' | 'LSEEPLQVLEHPR' | 'Endoplasmic reticulum aminopeptidase 1' |
| 14639 | 'ADA2_No1' | 'DIPIEVCPISNQVLK' | 'Adenosine deaminase 2' |
| 14640 | 'ADA2_No2' | 'WILLEDYR' | 'Adenosine deaminase 2' |
| 14641 | 'ADA2_No3' | 'TLIFPPSMHFFQAK' | 'Adenosine deaminase 2' |
| 14642 | 'ADA2_No4' | 'LLPVYELSGEHHDEEWSVK' | 'Adenosine deaminase 2' |
| 14643 | 'ADA2_No5' | 'WILLEDYRK' | 'Adenosine deaminase 2' |
| 14644 | 'ADA2_No6' | 'LPYFFHAGETDWQGTSIDR' | 'Adenosine deaminase 2' |
| 14645 | 'ADA2_No7' | 'LVLNTKEELANER' | 'Adenosine deaminase 2' |
| 14646 | 'ADA2_No8' | 'YSTLLESEKNTFMEIWK' | 'Adenosine deaminase 2' |
| 14647 | 'EHD3_No1' | 'MQDQLQAQDFSK' | 'EH domain-containing protein 3' |
| 14648 | 'EHD3_No2' | 'ELVNNLAEIYGR' | 'EH domain-containing protein 3' |
| 14649 | 'EHD3_No3' | 'LFEAEEQDLFR' | 'EH domain-containing protein 3' |
| 14650 | 'EHD3_No4' | 'DKPMYDEIFYTLSPVDGK' | 'EH domain-containing protein 3' |
| 14651 | 'EHD3_No5' | 'LADIDKDGMLDDDEFALANHLIK' | 'EH domain-containing protein 3' |
| 14652 | 'EHD3_No6' | 'DPEVFQTVSEGLK' | 'EH domain-containing protein 3' |
| 14653 | 'EHD3_No7' | 'MFSWLGTDDRR' | 'EH domain-containing protein 3' |

| | A | B | C |
|---|---|---|---|
| 1 | **Peptide Name** | **Peptide Sequence** | **Peptide Description** |
| 14654 | 'EHD3_No8' | 'LADIDKDGMLDDDEFALANHLIK' | 'EH domain-containing protein 3' |
| 14655 | 'EHD3_No9' | 'IVNTPEVIR' | 'EH domain-containing protein 3' |
| 14656 | 'EHD3_No10' | 'KDPEVFQTVSEGLKK' | 'EH domain-containing protein 3' |
| 14657 | 'C1RL_No1' | 'VLSYVDWIK' | 'Complement C1r subcomponent-like protein' |
| 14658 | 'C1RL_No2' | 'GSEAINAPGDNPAK' | 'Complement C1r subcomponent-like protein' |
| 14659 | 'C1RL_No3' | 'VQNHCQEPYYQAAAAGALTCATPGTWK' | 'Complement C1r subcomponent-like protein' |
| 14660 | 'C1RL_No4' | 'LGNFPWQAFTSIHGR' | 'Complement C1r subcomponent-like protein' |
| 14661 | 'C1RL_No5' | 'GQESSTDIKAPEGFAVR' | 'Complement C1r subcomponent-like protein' |
| 14662 | 'C1RL_No6' | 'QRPEVFSDNMFCVGDETQR' | 'Complement C1r subcomponent-like protein' |
| 14663 | 'C1RL_No7' | 'DRQDGEEVLQCMPVCGR' | 'Complement C1r subcomponent-like protein' |
| 14664 | 'C1RL_No8' | 'QRPEVFSDNMFCVGDETQR' | 'Complement C1r subcomponent-like protein' |
| 14665 | 'C1RL_No9' | 'QRPEVFSDNMFCVGDETQR' | 'Complement C1r subcomponent-like protein' |
| 14666 | 'C1RL_No10' | 'VVVHPDYR' | 'Complement C1r subcomponent-like protein' |
| 14667 | 'C1RL_No11' | 'RQRPEVFSDNMFCVGDETQR' | 'Complement C1r subcomponent-like protein' |
| 14668 | 'C1RL_No12' | 'AKLGNFPWQAFTSIHGR' | 'Complement C1r subcomponent-like protein' |
| 14669 | 'C1RL_No13' | 'QRPEVFSDNMFCVGDETQR' | 'Complement C1r subcomponent-like protein' |
| 14670 | 'C1RL_No14' | 'GGGALLGDR' | 'Complement C1r subcomponent-like protein' |
| 14671 | 'C1RL_No15' | 'QDGEEVLQCMPVCGR' | 'Complement C1r subcomponent-like protein' |
| 14672 | 'C1RL_No16' | 'QDGEEVLQCMPVCGR' | 'Complement C1r subcomponent-like protein' |
| 14673 | 'C1RL_No17' | 'VQNHCQEPYYQAAAAGALTCATPGTWKDR' | 'Complement C1r subcomponent-like protein' |
| 14674 | 'C1RL_No18' | 'EACNAWLQKR' | 'Complement C1r subcomponent-like protein' |
| 14675 | 'C1RL_No19' | 'DRQDGEEVLQCMPVCGR' | 'Complement C1r subcomponent-like protein' |
| 14676 | 'C1RL_No20' | 'RQRPEVFSDNMFCVGDETQR' | 'Complement C1r subcomponent-like protein' |
| 14677 | 'CALML5_No1' | 'AGELTPEEEAQYKK' | 'Calmodulin-like protein 5' |
| 14678 | 'CALML5_No2' | 'AFSAVDTDGNGTINAQELGAALK' | 'Calmodulin-like protein 5' |
| 14679 | 'ABRACL_No1' | 'MNVDHEVNLLVEEIHR' | 'Costars family protein ABRACL' |
| 14680 | 'ABRACL_No2' | 'MNVDHEVNLLVEEIHR' | 'Costars family protein ABRACL' |
| 14681 | 'CNTN3_No1' | 'HGLVYSSAELK' | 'Contactin-3' |
| 14682 | 'CNTN3_No2' | 'YVCMVQTGVDSVSSAADLIVR' | 'Contactin-3' |
| 14683 | 'CNTN3_No3' | 'IEVQFPETLPAAK' | 'Contactin-3' |
| 14684 | 'CNTN3_No4' | 'TPFSVGWQTVTTVPEVIDGK' | 'Contactin-3' |
| 14685 | 'CNTN3_No5' | 'TSAELVLPIKEDYIIEVK' | 'Contactin-3' |
| 14686 | 'CNTN3_No6' | 'VLLNWEQVK' | 'Contactin-3' |
| 14687 | 'CNTN3_No7' | 'LQFAYLENFK' | 'Contactin-3' |
| 14688 | 'CNTN3_No8' | 'NWDTGTYQCFATNSLGTIVSR' | 'Contactin-3' |
| 14689 | 'GMPR2_No1' | 'GHIISDGGCSCPGDVAK' | 'GMP reductase 2' |
| 14690 | 'GMPR2_No2' | 'LDFKDVLLRPK' | 'GMP reductase 2' |
| 14691 | 'DKK3_No1' | 'GLLFPVCTPLPVEGELCHDPASR' | 'Dickkopf-related protein 3' |
| 14692 | 'DKK3_No2' | 'SAVEEMEAEEAAAK' | 'Dickkopf-related protein 3' |
| 14693 | 'DKK3_No3' | 'EVEELMEDTQHK' | 'Dickkopf-related protein 3' |
| 14694 | 'DKK3_No4' | 'DSECCGDQLCVWGHCTK' | 'Dickkopf-related protein 3' |
| 14695 | 'EXTL2_No1' | 'YSNIMISQFGFPYANYK' | 'Exostosin-like 2' |
| 14696 | 'EXTL2_No2' | 'APDELWNSLGPHPIPVIFK' | 'Exostosin-like 2' |
| 14697 | 'EXTL2_No3' | 'LVNIYDSMPLR' | 'Exostosin-like 2' |

| | A | B | C |
|---|---|---|---|
| 1 | **Peptide Name** | **Peptide Sequence** | **Peptide Description** |
| 14698 | 'EXTL2_No4' | 'QPAAVHALIDDTQNCDDIAMNFIIAK' | 'Exostosin-like 2' |
| 14699 | 'EXTL2_No5' | 'TSGIFVKPVNMDNLEK' | 'Exostosin-like 2' |
| 14700 | 'EXTL2_No6' | 'VIVVWNNIGEK' | 'Exostosin-like 2' |
| 14701 | 'EXTL2_No7' | 'YLELFQR' | 'Exostosin-like 2' |
| 14702 | 'GRHPR' | 'GVAGAHGLLCLLSDHVDKR' | 'Glyoxylate reductase/hydroxypyruvate reductase' |
| 14703 | 'BIN2_No1' | 'AIVWNNDLLWEDYEEK' | 'Bridging integrator 2' |
| 14704 | 'BIN2_No2' | 'IGCYVTIFQNISNLR' | 'Bridging integrator 2' |
| 14705 | 'BIN2_No3' | 'VSETLQEIYSSEWDGHEELK' | 'Bridging integrator 2' |
| 14706 | 'BIN2_No4' | 'FEQSASNFYQQQAEGHK' | 'Bridging integrator 2' |
| 14707 | 'BIN2_No5' | 'TSLEVSPNPEPPEKPVR' | 'Bridging integrator 2' |
| 14708 | 'BIN2_No6' | 'LNHNLYEVMSK' | 'Bridging integrator 2' |
| 14709 | 'FBLN5_No1' | 'YPGAYYIFQIK' | 'Fibulin-5' |
| 14710 | 'FBLN5_No2' | 'DQPFTILYR' | 'Fibulin-5' |
| 14711 | 'FBLN5_No3' | 'QSGQCLDIDECR' | 'Fibulin-5' |
| 14712 | 'FBLN5_No4' | 'QTGPISATLVMTRPIK' | 'Fibulin-5' |
| 14713 | 'FBLN5_No5' | 'SVPADIFQMQATTR' | 'Fibulin-5' |
| 14714 | 'FBLN5_No6' | 'CIDPIRCEEPYLR' | 'Fibulin-5' |
| 14715 | 'FBLN5_No7' | 'QTGPISATLVMTRPIK' | 'Fibulin-5' |
| 14716 | 'FBLN5_No8' | 'QTGPISATLVMTRPIK' | 'Fibulin-5' |
| 14717 | 'FBLN5_No9' | 'SVPADIFQMQATTR' | 'Fibulin-5' |
| 14718 | 'FBLN5_No10' | 'GPYSNPYSTPYSGPYPAAAPPLSAPNYPTISR' | 'Fibulin-5' |
| 14719 | 'TJP2' | 'SNPSAVAGNETPGASTK' | 'Tight junction protein ZO-2' |
| 14720 | 'VTI1B' | 'YAPLSFRNPMMSK' | 'Vesicle transport through interaction with t-SNAREs homolog 1B' |
| 14721 | 'MARCO_No1' | 'NSWGHHDCSHEEDAGVECSV' | 'Macrophage receptor MARCO' |
| 14722 | 'MARCO_No2' | 'LQVLQAQLTWVR' | 'Macrophage receptor MARCO' |
| 14723 | 'NIPSNAP3A' | 'LVGVFHTEYGALNR' | 'Protein NipSnap homolog 3A' |
| 14724 | 'DMBT1' | 'FGQGSGPIVLDDVR' | 'Deleted in malignant brain tumors 1 protein' |
| 14725 | 'FETUB_No1' | 'LVVLPFPK' | 'Fetuin-B' |
| 14726 | 'FETUB_No2' | 'SQASSCSLQSSDSVPVGLCK' | 'Fetuin-B' |
| 14727 | 'FETUB_No3' | 'GSVQYLPDLDDKNSQEK' | 'Fetuin-B' |
| 14728 | 'FETUB_No4' | 'IYMTCPDCPSSIPTDSSNHQVLEAATESLAK' | 'Fetuin-B' |
| 14729 | 'FETUB_No5' | 'AIFYMNNPSR' | 'Fetuin-B' |
| 14730 | 'FETUB_No6' | 'ASSQWVVGPSYFVEYLIK' | 'Fetuin-B' |
| 14731 | 'FETUB_No7' | 'AIFYMNNPSR' | 'Fetuin-B' |
| 14732 | 'FETUB_No8' | 'IYMTCPDCPSSIPTDSSNHQVLEAATESLAK' | 'Fetuin-B' |
| 14733 | 'FETUB_No9' | 'KIYMTCPDCPSSIPTDSSNHQVLEAATESLAK' | 'Fetuin-B' |
| 14734 | 'FETUB_No10' | 'TAECPGPAQNASPLVLPP' | 'Fetuin-B' |
| 14735 | 'FETUB_No11' | 'KKIYMTCPDCPSSIPTDSSNHQVLEAATESLAK' | 'Fetuin-B' |
| 14736 | 'FETUB_No12' | 'KIYMTCPDCPSSIPTDSSNHQVLEAATESLAK' | 'Fetuin-B' |
| 14737 | 'FETUB_No13' | 'IFFESVYGQCK' | 'Fetuin-B' |
| 14738 | 'FETUB_No14' | 'LNRVNDAQEYR' | 'Fetuin-B' |
| 14739 | 'FETUB_No15' | 'VNDAQEYR' | 'Fetuin-B' |
| 14740 | 'FETUB_No16' | 'YNNENTSKQYSLFK' | 'Fetuin-B' |
| 14741 | 'FETUB_No17' | 'VLYLAAYNCTLRPVSK' | 'Fetuin-B' |

| | A | B | C |
|---|---|---|---|
| 1 | **Peptide Name** | **Peptide Sequence** | **Peptide Description** |
| 14742 | 'PCYOX1_No1' | 'FLNEMIAPVMR' | 'Prenylcysteine oxidase 1' |
| 14743 | 'PCYOX1_No2' | 'MHMWVEDVLDK' | 'Prenylcysteine oxidase 1' |
| 14744 | 'PCYOX1_No3' | 'YGFQSLR' | 'Prenylcysteine oxidase 1' |
| 14745 | 'PCYOX1_No4' | 'YQSHDYAFSSVEK' | 'Prenylcysteine oxidase 1' |
| 14746 | 'PCYOX1_No5' | 'EKEDPEPSTDGTYVWK' | 'Prenylcysteine oxidase 1' |
| 14747 | 'PCYOX1_No6' | 'IFSQETLTK' | 'Prenylcysteine oxidase 1' |
| 14748 | 'PCYOX1_No7' | 'SNLISGSVMYIEEK' | 'Prenylcysteine oxidase 1' |
| 14749 | 'PCYOX1_No8' | 'LVCSGLLQASK' | 'Prenylcysteine oxidase 1' |
| 14750 | 'PCYOX1_No9' | 'WNGHTDMIDQDGLYEK' | 'Prenylcysteine oxidase 1' |
| 14751 | 'PCYOX1_No10' | 'MYEVVYQIGTETR' | 'Prenylcysteine oxidase 1' |
| 14752 | 'PCYOX1_No11' | 'SDFYDIVLVATPLNR' | 'Prenylcysteine oxidase 1' |
| 14753 | 'PCYOX1_No12' | 'SNLISGSVMYIEEK' | 'Prenylcysteine oxidase 1' |
| 14754 | 'PCYOX1_No13' | 'SDFYDIVLVATPLNRK' | 'Prenylcysteine oxidase 1' |
| 14755 | 'PCYOX1_No14' | 'IDLFEREEVGGR' | 'Prenylcysteine oxidase 1' |
| 14756 | 'PCYOX1_No15' | 'MHMWVEDVLDK' | 'Prenylcysteine oxidase 1' |
| 14757 | 'PCYOX1_No16' | 'MHMWVEDVLDK' | 'Prenylcysteine oxidase 1' |
| 14758 | 'PCYOX1_No17' | 'APPDKIAIIGAGIGGTSAAYYLR' | 'Prenylcysteine oxidase 1' |
| 14759 | 'PCYOX1_No18' | 'WNGHTDMIDQDGLYEK' | 'Prenylcysteine oxidase 1' |
| 14760 | 'PCYOX1_No19' | 'MHMWVEDVLDK' | 'Prenylcysteine oxidase 1' |
| 14761 | 'PCYOX1_No20' | 'CPSIILHDR' | 'Prenylcysteine oxidase 1' |
| 14762 | 'PCYOX1_No21' | 'IAIIGAGIGGTSAAYYLR' | 'Prenylcysteine oxidase 1' |
| 14763 | 'PCYOX1_No22' | 'KPWLAYPHYKPPEK' | 'Prenylcysteine oxidase 1' |
| 14764 | 'PCYOX1_No23' | 'LFLSYDYAVK' | 'Prenylcysteine oxidase 1' |
| 14765 | 'PCYOX1_No24' | 'FLNEMIAPVMR' | 'Prenylcysteine oxidase 1' |
| 14766 | 'PCYOX1_No25' | 'FLNEMIAPVMR' | 'Prenylcysteine oxidase 1' |
| 14767 | 'CD84_No1' | 'IHALGPNYNLVISDLR' | 'SLAM family member 5' |
| 14768 | 'CD84_No2' | 'TSVAYVTPGDSETAPVVTVTHR' | 'SLAM family member 5' |
| 14769 | 'SH3BGRL2_No1' | 'ESNTVFSFLGLKPR' | 'SH3 domain-binding glutamic acid-rich-like protein 2' |
| 14770 | 'SH3BGRL2_No2' | 'YCGDYDSFFESK' | 'SH3 domain-binding glutamic acid-rich-like protein 2' |
| 14771 | 'GNPTG_No1' | 'RDPSPVSGPVHLFR' | 'N-acetylglucosamine-1-phosphotransferase subunit gamma' |
| 14772 | 'GNPTG_No2' | 'VVEEPNAFGVNNPFLPQASR' | 'N-acetylglucosamine-1-phosphotransferase subunit gamma' |
| 14773 | 'GNPTG_No3' | 'QWDQVEQDLADELITPQGHEK' | 'N-acetylglucosamine-1-phosphotransferase subunit gamma' |
| 14774 | 'GNPTG_No4' | 'TLFEDAGYLK' | 'N-acetylglucosamine-1-phosphotransferase subunit gamma' |
| 14775 | 'GNPTG_No5' | 'DPSPVSGPVHLFR' | 'N-acetylglucosamine-1-phosphotransferase subunit gamma' |
| 14776 | 'DBNL_No1' | 'NGPALQEAYVR' | 'Drebrin-like protein' |
| 14777 | 'DBNL_No2' | 'AEEDVEPECIMEK' | 'Drebrin-like protein' |
| 14778 | 'DBNL_No3' | 'VAGTGEGGLEEMVEELNSGK' | 'Drebrin-like protein' |
| 14779 | 'DBNL_No4' | 'FVLINWTGEGVNDVRK' | 'Drebrin-like protein' |
| 14780 | 'DBNL_No5' | 'FQDVGPQAPVGSVYQK' | 'Drebrin-like protein' |
| 14781 | 'DBNL_No6' | 'AMSTTSISSPQPGK' | 'Drebrin-like protein' |
| 14782 | 'SERPINA10_No1' | 'GLHLQALKPTKPGLLPSLFK' | 'Protein Z-dependent protease inhibitor' |
| 14783 | 'SERPINA10_No2' | 'ETSNFGFSLLR' | 'Protein Z-dependent protease inhibitor' |
| 14784 | 'SERPINA10_No3' | 'GKWLTPFDPVFTEVDTFHLDKYK' | 'Protein Z-dependent protease inhibitor' |
| 14785 | 'SERPINA10_No4' | 'LILVDYILFK' | 'Protein Z-dependent protease inhibitor' |

| | A | B | C |
|---|---|---|---|
| 1 | **Peptide Name** | **Peptide Sequence** | **Peptide Description** |
| 14786 | 'SERPINA10_No5' | 'IFSPFADLSELSATGR' | 'Protein Z-dependent protease inhibitor' |
| 14787 | 'SERPINA10_No6' | 'LFDEINPETK' | 'Protein Z-dependent protease inhibitor' |
| 14788 | 'SERPINA10_No7' | 'YEMHELLR' | 'Protein Z-dependent protease inhibitor' |
| 14789 | 'SERPINA10_No8' | 'NLELGLTQGSFAFIHK' | 'Protein Z-dependent protease inhibitor' |
| 14790 | 'SERPINA10_No9' | 'FASTFDKNFR' | 'Protein Z-dependent protease inhibitor' |
| 14791 | 'SERPINA10_No10' | 'WLTPFDPVFTEVDTFHLDKYK' | 'Protein Z-dependent protease inhibitor' |
| 14792 | 'SERPINA10_No11' | 'YFDTECVPMNFR' | 'Protein Z-dependent protease inhibitor' |
| 14793 | 'SERPINA10_No12' | 'FKLDQKYEMHELLR' | 'Protein Z-dependent protease inhibitor' |
| 14794 | 'SERPINA10_No13' | 'YFDTECVPMNFR' | 'Protein Z-dependent protease inhibitor' |
| 14795 | 'SERPINA10_No14' | 'NLELGLTQGSFAFIHKDFDVK' | 'Protein Z-dependent protease inhibitor' |
| 14796 | 'SERPINA10_No15' | 'LDQKYEMHELLR' | 'Protein Z-dependent protease inhibitor' |
| 14797 | 'SERPINA10_No16' | 'PGLLPSLFK' | 'Protein Z-dependent protease inhibitor' |
| 14798 | 'SERPINA10_No17' | 'TIKVPMMYGAGK' | 'Protein Z-dependent protease inhibitor' |
| 14799 | 'SERPINA10_No18' | 'NMEVFFPK' | 'Protein Z-dependent protease inhibitor' |
| 14800 | 'SERPINA10_No19' | 'TVIEVDER' | 'Protein Z-dependent protease inhibitor' |
| 14801 | 'SERPINA10_No20' | 'GTEAVAGILSEITAYSMPPVIK' | 'Protein Z-dependent protease inhibitor' |
| 14802 | 'SERPINA10_No21' | 'ETSNFGFSLLRK' | 'Protein Z-dependent protease inhibitor' |
| 14803 | 'SERPINA10_No22' | 'VPMMYGAGK' | 'Protein Z-dependent protease inhibitor' |
| 14804 | 'SERPINA10_No23' | 'YEMHELLR' | 'Protein Z-dependent protease inhibitor' |
| 14805 | 'SERPINA10_No24' | 'FKLDQKYEMHELLR' | 'Protein Z-dependent protease inhibitor' |
| 14806 | 'SERPINA10_No25' | 'LDQKYEMHELLR' | 'Protein Z-dependent protease inhibitor' |
| 14807 | 'SERPINA10_No26' | 'GTEAVAGILSEITAYSMPPVIK' | 'Protein Z-dependent protease inhibitor' |
| 14808 | 'SERPINA10_No27' | 'NMEVFFPK' | 'Protein Z-dependent protease inhibitor' |
| 14809 | 'SERPINA10_No28' | 'RIFSPFADLSELSATGR' | 'Protein Z-dependent protease inhibitor' |
| 14810 | 'TRHDE_No1' | 'LLIDQLIR' | 'Thyrotropin-releasing hormone-degrading ectoenzyme' |
| 14811 | 'TRHDE_No2' | 'GLQDYLTIHK' | 'Thyrotropin-releasing hormone-degrading ectoenzyme' |
| 14812 | 'TRHDE_No3' | 'AGLIDDAFSLAR' | 'Thyrotropin-releasing hormone-degrading ectoenzyme' |
| 14813 | 'CORO1C_No1' | 'NADPILISLK' | 'Coronin-1C' |
| 14814 | 'CORO1C_No2' | 'SIKDTICNQDER' | 'Coronin-1C' |
| 14815 | 'CORO1C_No3' | 'VGIVAWHPTAR' | 'Coronin-1C' |
| 14816 | 'CORO1C_No4' | 'AIFLADGNVFTTGFSR' | 'Coronin-1C' |
| 14817 | 'CORO1C_No5' | 'RGLDVNKCEIAR' | 'Coronin-1C' |
| 14818 | 'CORO1C_No6' | 'YFEITDESPYVHYLNTFSSK' | 'Coronin-1C' |
| 14819 | 'CORO1C_No7' | 'KCEPIIMTVPR' | 'Coronin-1C' |
| 14820 | 'CORO1C_No8' | 'KCEPIIMTVPR' | 'Coronin-1C' |
| 14821 | 'CORO1C_No9' | 'GLDVNKCEIAR' | 'Coronin-1C' |
| 14822 | 'CORO1C_No10' | 'KTTDTASVQNEAKLDEILK' | 'Coronin-1C' |
| 14823 | 'CORO1C_No11' | 'HVFGQAVKNDQCYDDIR' | 'Coronin-1C' |
| 14824 | 'NOTCH3_No1' | 'ELQCPVGVPCQQTPR' | 'Neurogenic locus notch homolog protein 3' |
| 14825 | 'NOTCH3_No2' | 'GVCQSSVVAGTAR' | 'Neurogenic locus notch homolog protein 3' |
| 14826 | 'NOTCH3_No3' | 'CACPPGLSGPSCR' | 'Neurogenic locus notch homolog protein 3' |
| 14827 | 'NOTCH3_No4' | 'NVDDCSPDPCHHGR' | 'Neurogenic locus notch homolog protein 3' |
| 14828 | 'NOTCH3_No5' | 'PAPLAPFFR' | 'Neurogenic locus notch homolog protein 3' |
| 14829 | 'NOTCH3_No6' | 'VASFYCACPMGK' | 'Neurogenic locus notch homolog protein 3' |

| | A | B | C |
|---|---|---|---|
| 1 | **Peptide Name** | **Peptide Sequence** | **Peptide Description** |
| 14830 | 'PROCR_No1' | 'TLAFPLTIR' | 'Endothelial protein C receptor' |
| 14831 | 'PROCR_No2' | 'LHMLQISYFR' | 'Endothelial protein C receptor' |
| 14832 | 'PROCR_No3' | 'EFLEDTCVQYVQK' | 'Endothelial protein C receptor' |
| 14833 | 'PROCR_No4' | 'TQSGLQSYLLQFHGLVR' | 'Endothelial protein C receptor' |
| 14834 | 'MINPP1_No1' | 'LASLFPALFSR' | 'Multiple inositol polyphosphate phosphatase 1' |
| 14835 | 'MINPP1_No2' | 'DILQSCQTSEECELAR' | 'Multiple inositol polyphosphate phosphatase 1' |
| 14836 | 'MINPP1_No3' | 'VLPLAYSQETVSFYEDLK' | 'Multiple inositol polyphosphate phosphatase 1' |
| 14837 | 'MINPP1_No4' | 'SSCTLFQDIFQHLDK' | 'Multiple inositol polyphosphate phosphatase 1' |
| 14838 | 'MINPP1_No5' | 'VLPLAYSQETVSFYEDLKNHYK' | 'Multiple inositol polyphosphate phosphatase 1' |
| 14839 | 'MINPP1_No6' | 'DPVASSLSPYFGTK' | 'Multiple inositol polyphosphate phosphatase 1' |
| 14840 | 'MINPP1_No7' | 'QLHGLLQAR' | 'Multiple inositol polyphosphate phosphatase 1' |
| 14841 | 'MINPP1_No8' | 'DKEPLTAYNYK' | 'Multiple inositol polyphosphate phosphatase 1' |
| 14842 | 'MINPP1_No9' | 'VLEYLNDLK' | 'Multiple inositol polyphosphate phosphatase 1' |
| 14843 | 'MINPP1_No10' | 'SPWCDVFDIDDAK' | 'Multiple inositol polyphosphate phosphatase 1' |
| 14844 | 'MINPP1_No11' | 'TGPEMQNILK' | 'Multiple inositol polyphosphate phosphatase 1' |
| 14845 | 'PA2G4_No1' | 'LVKPGNQNTQVTEAWNK' | 'Proliferation-associated protein 2G4' |
| 14846 | 'PA2G4_No2' | 'HELLQPFNVLYEK' | 'Proliferation-associated protein 2G4' |
| 14847 | 'PA2G4_No3' | 'SGEDEQQEQTIAEDLVVTK' | 'Proliferation-associated protein 2G4' |
| 14848 | 'HPSE_No1' | 'VFQVVESTRPGK' | 'Heparanase' |
| 14849 | 'HPSE_No2' | 'AGGEVIDSVTWHHYYLNGR' | 'Heparanase' |
| 14850 | 'HPSE_No3' | 'YLLRPLGPHGLLSK' | 'Heparanase' |
| 14851 | 'HPSE_No4' | 'LYGPDVGQPR' | 'Heparanase' |
| 14852 | 'VDAC3_No1' | 'VNNASLIGLGYTQTLRPGVK' | 'Voltage-dependent anion-selective channel protein 3' |
| 14853 | 'VDAC3_No2' | 'SCSGVEFSTSGHAYTDTGK' | 'Voltage-dependent anion-selective channel protein 3' |
| 14854 | 'VDAC3_No3' | 'AADFQLHTHVNDGTEFGGSIYQK' | 'Voltage-dependent anion-selective channel protein 3' |
| 14855 | 'VSIG4_No1' | 'GSDPVTIFLR' | 'V-set and immunoglobulin domain-containing protein 4' |
| 14856 | 'VSIG4_No2' | 'GDVNLPCTYDPLQGYTQVLVK' | 'V-set and immunoglobulin domain-containing protein 4' |
| 14857 | 'VSIG4_No3' | 'GSPPISYIWYK' | 'V-set and immunoglobulin domain-containing protein 4' |
| 14858 | 'VSIG4_No4' | 'SHYTCEVTWQTPDGNQVVR' | 'V-set and immunoglobulin domain-containing protein 4' |
| 14859 | 'VSIG4_No5' | 'PAVIADSGSYFCTAK' | 'V-set and immunoglobulin domain-containing protein 4' |
| 14860 | 'TMED7' | 'SVIDYQTHFR' | 'Transmembrane emp24 domain-containing protein 7' |
| 14861 | 'TLN1_No1' | 'MVAAATNNLCEAANAAVQGHASQEK' | 'Talin-1' |
| 14862 | 'TLN1_No2' | 'ILAQATSDLVNAIK' | 'Talin-1' |
| 14863 | 'TLN1_No3' | 'TLAESALQLLYTAK' | 'Talin-1' |
| 14864 | 'TLN1_No4' | 'AGALQCSPSDAYTK' | 'Talin-1' |
| 14865 | 'TLN1_No5' | 'TMLESAGGLIQTAR' | 'Talin-1' |
| 14866 | 'TLN1_No6' | 'VLGEAMTGISQNAK' | 'Talin-1' |
| 14867 | 'TLN1_No7' | 'ASVPTIQDQASAMQLSQCAK' | 'Talin-1' |
| 14868 | 'TLN1_No8' | 'FLPSELRDEH' | 'Talin-1' |
| 14869 | 'TLN1_No9' | 'QVAASTAQLLVACK' | 'Talin-1' |
| 14870 | 'TLN1_No10' | 'TLSHPQQMALLDQTK' | 'Talin-1' |
| 14871 | 'TLN1_No11' | 'AVAEQIPLLVQGVR' | 'Talin-1' |
| 14872 | 'TLN1_No12' | 'LLAALLEDEGGSGRPLLQAAK' | 'Talin-1' |
| 14873 | 'TLN1_No13' | 'VAGSVTELIQAAEAMK' | 'Talin-1' |

| | A | B | C |
|---|---|---|---|
| 1 | **Peptide Name** | **Peptide Sequence** | **Peptide Description** |
| 14874 | 'TLN1_No14' | 'DPPSWSVLAGHSR' | 'Talin-1' |
| 14875 | 'TLN1_No15' | 'IGITNHDEYSLVR' | 'Talin-1' |
| 14876 | 'TLN1_No16' | 'ALDGAFTEENR' | 'Talin-1' |
| 14877 | 'TLN1_No17' | 'GLAGAVSELLR' | 'Talin-1' |
| 14878 | 'TLN1_No18' | 'EQGVEEHETLLLR' | 'Talin-1' |
| 14879 | 'TLN1_No19' | 'VSHVLAALQAGNR' | 'Talin-1' |
| 14880 | 'TLN1_No20' | 'AAAFEEQENETVVVK' | 'Talin-1' |
| 14881 | 'TLN1_No21' | 'ASAGPQPLLVQSCK' | 'Talin-1' |
| 14882 | 'TLN1_No22' | 'PAAVAAENEEIGSHIK' | 'Talin-1' |
| 14883 | 'TLN1_No23' | 'GSQAQPDSPSAQLALIAASQSFLQPGGK' | 'Talin-1' |
| 14884 | 'TLN1_No24' | 'VLGEAMTGISQNAK' | 'Talin-1' |
| 14885 | 'TLN1_No25' | 'VVAPTISSPVCQEQLVEAGR' | 'Talin-1' |
| 14886 | 'TLN1_No26' | 'ASVPTIQDQASAMQLSQCAK' | 'Talin-1' |
| 14887 | 'TLN1_No27' | 'MVGGIAQIIAAQEEMLRK' | 'Talin-1' |
| 14888 | 'TLN1_No28' | 'EVANSTANLVK' | 'Talin-1' |
| 14889 | 'TLN1_No29' | 'IPEAPAGPPSDFGLFLSDDDPKK' | 'Talin-1' |
| 14890 | 'TLN1_No30' | 'VSQMAQYFEPLTLAAVGAASK' | 'Talin-1' |
| 14891 | 'TLN1_No31' | 'EVIQEWNLTNIK' | 'Talin-1' |
| 14892 | 'TLN1_No32' | 'VLVQNAAGSQEK' | 'Talin-1' |
| 14893 | 'TLN1_No33' | 'EADESLNFEEQILEAAK' | 'Talin-1' |
| 14894 | 'TLN1_No34' | 'VMVTNVTSLLK' | 'Talin-1' |
| 14895 | 'TLN1_No35' | 'QEDVIATANLSR' | 'Talin-1' |
| 14896 | 'TLN1_No36' | 'LASEAKPAAVAAENEEIGSHIK' | 'Talin-1' |
| 14897 | 'TLN1_No37' | 'DLDQASLAAVSQQLAPR' | 'Talin-1' |
| 14898 | 'TLN1_No38' | 'GVGAAATAVTQALNELLQHVK' | 'Talin-1' |
| 14899 | 'TLN1_No39' | 'AAMEPIVISAK' | 'Talin-1' |
| 14900 | 'TLN1_No40' | 'DKAPGQLECETAIAALNSCLR' | 'Talin-1' |
| 14901 | 'TLN1_No41' | 'TLREQGVEEHETLLLR' | 'Talin-1' |
| 14902 | 'TLN1_No42' | 'SNTSPEELGPLANQLTSDYGR' | 'Talin-1' |
| 14903 | 'TLN1_No43' | 'LKPLPGETMEK' | 'Talin-1' |
| 14904 | 'TLN1_No44' | 'LNEAAAGLNQAATELVQASR' | 'Talin-1' |
| 14905 | 'TLN1_No45' | 'QAAASATQTIAAAQHAASTPK' | 'Talin-1' |
| 14906 | 'TLN1_No46' | 'VEHGSVALPAIMR' | 'Talin-1' |
| 14907 | 'TLN1_No47' | 'AGALQCSPSDAYTKK' | 'Talin-1' |
| 14908 | 'TLN1_No48' | 'AVEGCVSASQAATEDGQLLR' | 'Talin-1' |
| 14909 | 'TLN1_No49' | 'LGAASLGAEDPETQVVLINAVK' | 'Talin-1' |
| 14910 | 'TLN1_No50' | 'NCGQMSEIEAK' | 'Talin-1' |
| 14911 | 'TLN1_No51' | 'FFYSDQNVDSR' | 'Talin-1' |
| 14912 | 'TLN1_No52' | 'VQELGHGCAALVTK' | 'Talin-1' |
| 14913 | 'TLN1_No53' | 'QVAASTAQLLVACK' | 'Talin-1' |
| 14914 | 'TLN1_No54' | 'FFYSDQNVDSRDPVQLNLLYVQAR' | 'Talin-1' |
| 14915 | 'TLN1_No55' | 'LHTDDELNWLDHGR' | 'Talin-1' |
| 14916 | 'TLN1_No56' | 'AGFLDLKDFLPK' | 'Talin-1' |
| 14917 | 'TLN1_No57' | 'VGAIPANALDDGQWSQGLISAAR' | 'Talin-1' |

| | A | B | C |
|---|---|---|---|
| 1 | **Peptide Name** | **Peptide Sequence** | **Peptide Description** |
| 14918 | 'TLN1_No58' | 'TYGVSFFLVK' | 'Talin-1' |
| 14919 | 'TLN1_No59' | 'EVIQEWNLTNIKR' | 'Talin-1' |
| 14920 | 'TLN1_No60' | 'ALDYYMLR' | 'Talin-1' |
| 14921 | 'TLN1_No61' | 'TMQFEPSTMVYDACR' | 'Talin-1' |
| 14922 | 'TLN1_No62' | 'SKDHFGLEGDEESTMLEDSVSPK' | 'Talin-1' |
| 14923 | 'TLN1_No63' | 'EAAYHPEVAPDVR' | 'Talin-1' |
| 14924 | 'TLN1_No64' | 'VMVTNVTSLLK' | 'Talin-1' |
| 14925 | 'TLN1_No65' | 'DPVQLNLLYVQAR' | 'Talin-1' |
| 14926 | 'TLN1_No66' | 'QKLHTDDELNWLDHGR' | 'Talin-1' |
| 14927 | 'TLN1_No67' | 'TMLESAGGLIQTAR' | 'Talin-1' |
| 14928 | 'TLN1_No68' | 'TKEVIQEWNLTNIKR' | 'Talin-1' |
| 14929 | 'TLN1_No69' | 'SGASGPENFQVGSMPPAQQQITSGQMHR' | 'Talin-1' |
| 14930 | 'TLN1_No70' | 'ALGDLISATK' | 'Talin-1' |
| 14931 | 'TLN1_No71' | 'AVTDSINQLITMCTQQAPGQK' | 'Talin-1' |
| 14932 | 'TLN1_No72' | 'IPEAPAGPPSDFGLFLSDDDPK' | 'Talin-1' |
| 14933 | 'TLN1_No73' | 'AQEACGPLEMDSALSVVQNLEK' | 'Talin-1' |
| 14934 | 'TLN1_No74' | 'VGDDPAVWQLK' | 'Talin-1' |
| 14935 | 'TLN1_No75' | 'ALEATTEHIRQELAVFCSPEPPAK' | 'Talin-1' |
| 14936 | 'TLN1_No76' | 'TLREQGVEEHETLLLRR' | 'Talin-1' |
| 14937 | 'TLN1_No77' | 'ILAQATSDLVNAIKADAEGESDLENSRK' | 'Talin-1' |
| 14938 | 'TLN1_No78' | 'TLSHPQQMALLDQTK' | 'Talin-1' |
| 14939 | 'TLN1_No79' | 'EGTETFADHREGILK' | 'Talin-1' |
| 14940 | 'TLN1_No80' | 'STVLQQQYNR' | 'Talin-1' |
| 14941 | 'TLN1_No81' | 'LLSDSLPPSTGTFQEAQSR' | 'Talin-1' |
| 14942 | 'TLN1_No82' | 'VSQMAQYFEPLTLAAVGAASK' | 'Talin-1' |
| 14943 | 'TLN1_No83' | 'MATNAAAQNAIK' | 'Talin-1' |
| 14944 | 'TLN1_No84' | 'ALDGAFTEENRAQCR' | 'Talin-1' |
| 14945 | 'TLN1_No85' | 'KFFYSDQNVDSRDPVQLNLLYVQAR' | 'Talin-1' |
| 14946 | 'TLN1_No86' | 'SGASGPENFQVGSMPPAQQQITSGQMHR' | 'Talin-1' |
| 14947 | 'TLN1_No87' | 'AIAVTVQEMVTK' | 'Talin-1' |
| 14948 | 'TLN1_No88' | 'QKLHTDDELNWLDHGR' | 'Talin-1' |
| 14949 | 'TLN1_No89' | 'ELMEEKKEEITGTLR' | 'Talin-1' |
| 14950 | 'TLN1_No90' | 'TEDSGLQTQVIAAATQCALSTSQLVACTK' | 'Talin-1' |
| 14951 | 'TLN1_No91' | 'DDILNGSHPVSFDK' | 'Talin-1' |
| 14952 | 'TLN1_No92' | 'HRVQELGHGCAALVTK' | 'Talin-1' |
| 14953 | 'TLN1_No93' | 'TVTDMLMTICAR' | 'Talin-1' |
| 14954 | 'TLN1_No94' | 'ADAEGESDLENSR' | 'Talin-1' |
| 14955 | 'TLN1_No95' | 'SKDHFGLEGDEESTMLEDSVSPKK' | 'Talin-1' |
| 14956 | 'TLN1_No96' | 'LGAASLGAEDPETQVVLINAVKDVAK' | 'Talin-1' |
| 14957 | 'TLN1_No97' | 'VAGSVTELIQAAEAMK' | 'Talin-1' |
| 14958 | 'TLN1_No98' | 'VGKVEHGSVALPAIMR' | 'Talin-1' |
| 14959 | 'TLN1_No99' | 'AVTDSINQLITMCTQQAPGQKECDNALR' | 'Talin-1' |
| 14960 | 'TLN1_No100' | 'QAAASATQTIAAAQHAASTPK' | 'Talin-1' |
| 14961 | 'TLN1_No101' | 'ACKEAAYHPEVAPDVR' | 'Talin-1' |

| | A | B | C |
|---|---|---|---|
| 1 | **Peptide Name** | **Peptide Sequence** | **Peptide Description** |
| 14962 | 'TLN1_No102' | 'ACEFAGFQCQIQFGPHNEQK' | 'Talin-1' |
| 14963 | 'TLN1_No103' | 'MVAAATNNLCEAANAAVQGHASQEK' | 'Talin-1' |
| 14964 | 'TLN1_No104' | 'MVGGIAQIIAAQEEMLRK' | 'Talin-1' |
| 14965 | 'TLN1_No105' | 'TMQFEPSTMVYDACR' | 'Talin-1' |
| 14966 | 'TLN1_No106' | 'MVGGIAQIIAAQEEMLRK' | 'Talin-1' |
| 14967 | 'TLN1_No107' | 'TKEVIQEWNLTNIK' | 'Talin-1' |
| 14968 | 'TLN1_No108' | 'MVGGIAQIIAAQEEMLRK' | 'Talin-1' |
| 14969 | 'TLN1_No109' | 'ERIPEAPAGPPSDFGLFLSDDDPKK' | 'Talin-1' |
| 14970 | 'TLN1_No110' | 'AAAFEEQENETVVVKEK' | 'Talin-1' |
| 14971 | 'TLN1_No111' | 'ANQAIQMACQSLGEPGCTQAQVLSAATIVAK' | 'Talin-1' |
| 14972 | 'TLN1_No112' | 'FGQDFSTFLEAGVEMAGQAPSQEDR' | 'Talin-1' |
| 14973 | 'TLN1_No113' | 'ERIPEAPAGPPSDFGLFLSDDDPK' | 'Talin-1' |
| 14974 | 'TLN1_No114' | 'AATAPLLEAVDNLSAFASNPEFSSIPAQISPEGR' | 'Talin-1' |
| 14975 | 'TLN1_No115' | 'QELAVFCSPEPPAK' | 'Talin-1' |
| 14976 | 'TLN1_No116' | 'ELMEEKKEEITGTLR' | 'Talin-1' |
| 14977 | 'TLN1_No117' | 'MVGGIAQIIAAQEEMLR' | 'Talin-1' |
| 14978 | 'TLN1_No118' | 'ALAVNPRDPPSWSVLAGHSR' | 'Talin-1' |
| 14979 | 'TLN1_No119' | 'QFVQSAKEVANSTANLVK' | 'Talin-1' |
| 14980 | 'TLN1_No120' | 'SVAQRTEDSGLQTQVIAAATQCALSTSQLVACTK' | 'Talin-1' |
| 14981 | 'TLN1_No121' | 'TSTPEDFIR' | 'Talin-1' |
| 14982 | 'TLN1_No122' | 'EQGVEEHETLLLRR' | 'Talin-1' |
| 14983 | 'TLN1_No123' | 'SKDHFGLEGDEESTMLEDSVSPK' | 'Talin-1' |
| 14984 | 'TLN1_No124' | 'LASEAKPAAVAAENEEIGSHIKHR' | 'Talin-1' |
| 14985 | 'TLN1_No125' | 'AIAVTVQEMVTK' | 'Talin-1' |
| 14986 | 'TLN1_No126' | 'MVAAATNNLCEAANAAVQGHASQEKLISSAK' | 'Talin-1' |
| 14987 | 'TLN1_No127' | 'ANQAIQMACQSLGEPGCTQAQVLSAATIVAK' | 'Talin-1' |
| 14988 | 'TLN1_No128' | 'ILAQATSDLVNAIKADAEGESDLENSR' | 'Talin-1' |
| 14989 | 'TLN1_No129' | 'VGKVEHGSVALPAIMR' | 'Talin-1' |
| 14990 | 'TLN1_No130' | 'APGQLECETAIAALNSCLR' | 'Talin-1' |
| 14991 | 'TLN1_No131' | 'RVAGSVTELIQAAEAMK' | 'Talin-1' |
| 14992 | 'TLN1_No132' | 'AKPKEADESLNFEEQILEAAK' | 'Talin-1' |
| 14993 | 'TLN1_No133' | 'QELAVFCSPEPPAK' | 'Talin-1' |
| 14994 | 'TLN1_No134' | 'MVAAATNNLCEAANAAVQGHASQEKLISSAK' | 'Talin-1' |
| 14995 | 'TLN1_No135' | 'VLVEDTKVLVQNAAGSQEKLAQAAQSSVATITR' | 'Talin-1' |
| 14996 | 'TLN1_No136' | 'ALSTDPAAPNLKSQLAAAAR' | 'Talin-1' |
| 14997 | 'TLN1_No137' | 'MVGGIAQIIAAQEEMLR' | 'Talin-1' |
| 14998 | 'TLN1_No138' | 'RVAGSVTELIQAAEAMK' | 'Talin-1' |
| 14999 | 'TLN1_No139' | 'TLAESALQLLYTAKEAGGNPK' | 'Talin-1' |
| 15000 | 'TLN1_No140' | 'NLGTALAELR' | 'Talin-1' |
| 15001 | 'TLN1_No141' | 'QFVQSAKEVANSTANLVK' | 'Talin-1' |
| 15002 | 'TLN1_No142' | 'VEHGSVALPAIMR' | 'Talin-1' |
| 15003 | 'TLN1_No143' | 'MVGGIAQIIAAQEEMLR' | 'Talin-1' |
| 15004 | 'TLN1_No144' | 'AVTDSINQLITMCTQQAPGQK' | 'Talin-1' |
| 15005 | 'TLN1_No145' | 'EGISQEALHTQMLTAVQEISHLIEPLANAAR' | 'Talin-1' |

| | A | B | C |
|---|---|---|---|
| 1 | **Peptide Name** | **Peptide Sequence** | **Peptide Description** |
| 15006 | 'TLN1_No146' | 'VLVEDTKVLVQNAAGSQEK' | 'Talin-1' |
| 15007 | 'TLN1_No147' | 'LAQAAQSSVATITR' | 'Talin-1' |
| 15008 | 'HYOU1_No1' | 'LSAASTWLEDEGVGATTVMLK' | 'Hypoxia up-regulated protein 1' |
| 15009 | 'HYOU1_No2' | 'VLQLINDNTATALSYGVFR' | 'Hypoxia up-regulated protein 1' |
| 15010 | 'HYOU1_No3' | 'EVEEEPGIHSLK' | 'Hypoxia up-regulated protein 1' |
| 15011 | 'HYOU1_No4' | 'LQDLTLRDLEK' | 'Hypoxia up-regulated protein 1' |
| 15012 | 'HYOU1_No5' | 'KLCQGLFFR' | 'Hypoxia up-regulated protein 1' |
| 15013 | 'HYOU1_No6' | 'AANSLEAFIFETQDK' | 'Hypoxia up-regulated protein 1' |
| 15014 | 'HYOU1_No7' | 'LSAASTWLEDEGVGATTVMLK' | 'Hypoxia up-regulated protein 1' |
| 15015 | 'HYOU1_No8' | 'VEFEELCADLFER' | 'Hypoxia up-regulated protein 1' |
| 15016 | 'HYOU1_No9' | 'LYQPEYQEVSTEEQR' | 'Hypoxia up-regulated protein 1' |
| 15017 | 'HYOU1_No10' | 'AEAGPEGVAPAPEGEKK' | 'Hypoxia up-regulated protein 1' |
| 15018 | 'HYOU1_No11' | 'LCQGLFFR' | 'Hypoxia up-regulated protein 1' |
| 15019 | 'HYOU1_No12' | 'MMALDREVQYLLNK' | 'Hypoxia up-regulated protein 1' |
| 15020 | 'ANGPTL3_No1' | 'ILANGLLQLGHGLK' | 'Angiopoietin-related protein 3' |
| 15021 | 'ANGPTL3_No2' | 'HDGIPAECTTIYNR' | 'Angiopoietin-related protein 3' |
| 15022 | 'ANGPTL3_No3' | 'LDGEFWLGLEK' | 'Angiopoietin-related protein 3' |
| 15023 | 'ANGPTL3_No4' | 'LNIFDQSFYDLSLQTSEIKEEEK' | 'Angiopoietin-related protein 3' |
| 15024 | 'ANGPTL3_No5' | 'FAMLDDVK' | 'Angiopoietin-related protein 3' |
| 15025 | 'ANGPTL3_No6' | 'TSIQEPTEISLSSKPR' | 'Angiopoietin-related protein 3' |
| 15026 | 'ANGPTL3_No7' | 'DLVFSTWDHK' | 'Angiopoietin-related protein 3' |
| 15027 | 'ANGPTL3_No8' | 'QDNSIKDLLQTVEDQYK' | 'Angiopoietin-related protein 3' |
| 15028 | 'ANGPTL3_No9' | 'NVKHDGIPAECTTIYNR' | 'Angiopoietin-related protein 3' |
| 15029 | 'ANGPTL3_No10' | 'YLEEQLTNLIQNQPETPEHPEVTSLK' | 'Angiopoietin-related protein 3' |
| 15030 | 'ANGPTL3_No11' | 'IELEDWK' | 'Angiopoietin-related protein 3' |
| 15031 | 'ANGPTL3_No12' | 'DLLQTVEDQYK' | 'Angiopoietin-related protein 3' |
| 15032 | 'ANGPTL3_No13' | 'MLIHPTDSESFE' | 'Angiopoietin-related protein 3' |
| 15033 | 'LYVE1_No1' | 'NGVGVLIWK' | 'Lymphatic vessel endothelial hyaluronic acid receptor 1' |
| 15034 | 'LYVE1_No2' | 'LLGLSLAGKDQVETALK' | 'Lymphatic vessel endothelial hyaluronic acid receptor 1' |
| 15035 | 'LYVE1_No3' | 'LLGLSLAGK' | 'Lymphatic vessel endothelial hyaluronic acid receptor 1' |
| 15036 | 'LYVE1_No4' | 'IMGITLVSK' | 'Lymphatic vessel endothelial hyaluronic acid receptor 1' |
| 15037 | 'LYVE1_No5' | 'IMGITLVSK' | 'Lymphatic vessel endothelial hyaluronic acid receptor 1' |
| 15038 | 'LYVE1_No6' | 'AEELSIQVSCR' | 'Lymphatic vessel endothelial hyaluronic acid receptor 1' |
| 15039 | 'F11R_No1' | 'VTFLPTGITFK' | 'Junctional adhesion molecule A' |
| 15040 | 'F11R_No2' | 'NGYGTPMTSNAVR' | 'Junctional adhesion molecule A' |
| 15041 | 'F11R_No3' | 'AVLTCSEQDGSPPSEYTWFK' | 'Junctional adhesion molecule A' |
| 15042 | 'EMILIN1_No1' | 'LQQEATEHATESEER' | 'EMILIN-1' |
| 15043 | 'EMILIN1_No2' | 'LVGSGLHTVEAAGEAR' | 'EMILIN-1' |
| 15044 | 'EMILIN1_No3' | 'APAPASAPPGPSEELLR' | 'EMILIN-1' |
| 15045 | 'EMILIN1_No4' | 'VQQLEEQVQSLTK' | 'EMILIN-1' |
| 15046 | 'FCGBP_No1' | 'VNGVLTALPVSVADGR' | 'IgGFc-binding protein' |
| 15047 | 'FCGBP_No2' | 'AIGYATAADCGR' | 'IgGFc-binding protein' |
| 15048 | 'FCGBP_No3' | 'APGWDPLCWDECR' | 'IgGFc-binding protein' |
| 15049 | 'FCGBP_No4' | 'LPVVLANGQIR' | 'IgGFc-binding protein' |

| | A | B | C |
|---|---|---|---|
| 1 | **Peptide Name** | **Peptide Sequence** | **Peptide Description** |
| 15050 | 'FCGBP_No5' | 'TVLSPVEPSCEGMQCAAGQR' | 'IgGFc-binding protein' |
| 15051 | 'FCGBP_No6' | 'TPDGSLLVR' | 'IgGFc-binding protein' |
| 15052 | 'FCGBP_No7' | 'SPANCPLSCPANSR' | 'IgGFc-binding protein' |
| 15053 | 'FCGBP_No8' | 'SLAAYTAACQAAGVAVKPWR' | 'IgGFc-binding protein' |
| 15054 | 'FCGBP_No9' | 'GCVLDVCMGGGDRDILCK' | 'IgGFc-binding protein' |
| 15055 | 'FCGBP_No10' | 'LRVPAAYAGSLCGLCGNYNQDPADDLK' | 'IgGFc-binding protein' |
| 15056 | 'FCGBP_No11' | 'AGCVAESTAVCR' | 'IgGFc-binding protein' |
| 15057 | 'FCGBP_No12' | 'YDLAFVVASQATK' | 'IgGFc-binding protein' |
| 15058 | 'FCGBP_No13' | 'VTASSPVAVLSGHSCAQK' | 'IgGFc-binding protein' |
| 15059 | 'FCGBP_No14' | 'DPCHGVTCRPQETCK' | 'IgGFc-binding protein' |
| 15060 | 'FCGBP_No15' | 'TCQGSCAALSGLTGCTTR' | 'IgGFc-binding protein' |
| 15061 | 'FCGBP_No16' | 'GNPAVSYVR' | 'IgGFc-binding protein' |
| 15062 | 'FCGBP_No17' | 'FAVLQENVAWGNGR' | 'IgGFc-binding protein' |
| 15063 | 'FCGBP_No18' | 'HTTCNHVVEQLLPTSAWGTHYVVPTLASQSR' | 'IgGFc-binding protein' |
| 15064 | 'FCGBP_No19' | 'SVPGCEGVALVVAQTK' | 'IgGFc-binding protein' |
| 15065 | 'FCGBP_No20' | 'AISGLTIDGHAVGAK' | 'IgGFc-binding protein' |
| 15066 | 'FCGBP_No21' | 'YQKEEFCGLLSSPTGPLSSCHK' | 'IgGFc-binding protein' |
| 15067 | 'FCGBP_No22' | 'NPQGPFATCQAVLSPSEYFR' | 'IgGFc-binding protein' |
| 15068 | 'FCGBP_No23' | 'VVVCQEHSCKPGQVCQPSGGILSCVTK' | 'IgGFc-binding protein' |
| 15069 | 'FCGBP_No24' | 'VTLQPYNVAQLQSSVDLSGSK' | 'IgGFc-binding protein' |
| 15070 | 'FCGBP_No25' | 'VAYDLVYYVR' | 'IgGFc-binding protein' |
| 15071 | 'FCGBP_No26' | 'LCGACGNFDGDQTNDWHDSQEKPAMEK' | 'IgGFc-binding protein' |
| 15072 | 'FCGBP_No27' | 'NTGREEFLTAFLQNYQLAYSK' | 'IgGFc-binding protein' |
| 15073 | 'FCGBP_No28' | 'ALASYVAACQAAGVVIEDWR' | 'IgGFc-binding protein' |
| 15074 | 'FCGBP_No29' | 'TVLSPVEPSCEGMQCAAGQR' | 'IgGFc-binding protein' |
| 15075 | 'FCGBP_No30' | 'CSVQNGLLGCYPDR' | 'IgGFc-binding protein' |
| 15076 | 'FCGBP_No31' | 'VDVTLPSSYHGAVCGLCGNMDR' | 'IgGFc-binding protein' |
| 15077 | 'FCGBP_No32' | 'CLANGGIHYITLDGR' | 'IgGFc-binding protein' |
| 15078 | 'FCGBP_No33' | 'VDVTLPSSYHGAVCGLCGNMDR' | 'IgGFc-binding protein' |
| 15079 | 'FCGBP_No34' | 'VPSSYAEALCGLCGNFNGDPADDLALR' | 'IgGFc-binding protein' |
| 15080 | 'FCGBP_No35' | 'GEVGFVLVDNQR' | 'IgGFc-binding protein' |
| 15081 | 'FCGBP_No36' | 'VYDLHGSCSYVLAQVCHPKPGDEDFSIVLEK' | 'IgGFc-binding protein' |
| 15082 | 'FCGBP_No37' | 'CPGLQNTIPWYR' | 'IgGFc-binding protein' |
| 15083 | 'FCGBP_No38' | 'TDSFCPLHCPAHSHYSICTR' | 'IgGFc-binding protein' |
| 15084 | 'FCGBP_No39' | 'KYQKEEFCGLLSSPTGPLSSCHK' | 'IgGFc-binding protein' |
| 15085 | 'FCGBP_No40' | 'YYPLGQTFYPGPGCDSLCR' | 'IgGFc-binding protein' |
| 15086 | 'FCGBP_No41' | 'GMVCQEHSCKPGQVCQPSGGILSCVTK' | 'IgGFc-binding protein' |
| 15087 | 'FCGBP_No42' | 'LPVSLSEGR' | 'IgGFc-binding protein' |
| 15088 | 'FCGBP_No43' | 'RCECGPGGHVTCQEGAACGPHEECR' | 'IgGFc-binding protein' |
| 15089 | 'FCGBP_No44' | 'LRVPAAYAASLCGLCGNYNQDPADDLK' | 'IgGFc-binding protein' |
| 15090 | 'FCGBP_No45' | 'FDFMGTCVYVLAQTCGTRPGLHR' | 'IgGFc-binding protein' |
| 15091 | 'FCGBP_No46' | 'YYPLGEVFYPGPECER' | 'IgGFc-binding protein' |
| 15092 | 'FCGBP_No47' | 'LLFDGDAHLLMSIPSPFR' | 'IgGFc-binding protein' |
| 15093 | 'FCGBP_No48' | 'EGGEVSCEPSSCGPHETCR' | 'IgGFc-binding protein' |

| | A | B | C |
|---|---|---|---|
| 1 | **Peptide Name** | **Peptide Sequence** | **Peptide Description** |
| 15094 | 'FCGBP_No49' | 'VVAEVQICHGK' | 'IgGFc-binding protein' |
| 15095 | 'FCGBP_No50' | 'ASQHGSDVVIETDFGLR' | 'IgGFc-binding protein' |
| 15096 | 'FCGBP_No51' | 'VPAAYAGSLCGLCGNYNQDPADDLK' | 'IgGFc-binding protein' |
| 15097 | 'FCGBP_No52' | 'FDFQGTCNYVLATTGCPGVSTQGLTPFTVTTK' | 'IgGFc-binding protein' |
| 15098 | 'FCGBP_No53' | 'LLISSLSESPASVSILSQADNTSKK' | 'IgGFc-binding protein' |
| 15099 | 'FCGBP_No54' | 'CSCSSSSGLTCQAAGCPPGR' | 'IgGFc-binding protein' |
| 15100 | 'FCGBP_No55' | 'GCVLDVCMGGGDRDILCK' | 'IgGFc-binding protein' |
| 15101 | 'FCGBP_No56' | 'AYSHSVSLTR' | 'IgGFc-binding protein' |
| 15102 | 'FCGBP_No57' | 'VVVCQEHSCKPGQVCQPSGGILSCVNK' | 'IgGFc-binding protein' |
| 15103 | 'FCGBP_No58' | 'CECGPGGHVTCQEGAACGPHEECR' | 'IgGFc-binding protein' |
| 15104 | 'FCGBP_No59' | 'GMVCQEHSCKPGQVCQPSGGILSCVTK' | 'IgGFc-binding protein' |
| 15105 | 'FCGBP_No60' | 'LCGACGNFDGDQTNDWHDSQEKPAMEK' | 'IgGFc-binding protein' |
| 15106 | 'FCGBP_No61' | 'PAGWQVGGAQGCGECVSK' | 'IgGFc-binding protein' |
| 15107 | 'FCGBP_No62' | 'SRLPVSLSEGR' | 'IgGFc-binding protein' |
| 15108 | 'FCGBP_No63' | 'VLVENEHRGSQTVSYTR' | 'IgGFc-binding protein' |
| 15109 | 'FCGBP_No64' | 'GGGQAANALAFGNSWQEETRPGCGATEPGDCPK' | 'IgGFc-binding protein' |
| 15110 | 'FCGBP_No65' | 'RFDFMGTCTYLLVGSCGQNAALPAFR' | 'IgGFc-binding protein' |
| 15111 | 'FCGBP_No66' | 'RVSYVGLVTVR' | 'IgGFc-binding protein' |
| 15112 | 'FCGBP_No67' | 'TEAVGQVHIFFQDGMVTLTPNK' | 'IgGFc-binding protein' |
| 15113 | 'FCGBP_No68' | 'AVGGKPAGWQVGGAQGCGECVSK' | 'IgGFc-binding protein' |
| 15114 | 'FCGBP_No69' | 'VSYVGLVTVR' | 'IgGFc-binding protein' |
| 15115 | 'FCGBP_No70' | 'PGDEDFSIVLEK' | 'IgGFc-binding protein' |
| 15116 | 'FCGBP_No71' | 'LEDGVQACHATGCGR' | 'IgGFc-binding protein' |
| 15117 | 'FCGBP_No72' | 'EGCVCDAGFVLSGDTCVPVGQCGCLHDGR' | 'IgGFc-binding protein' |
| 15118 | 'FCGBP_No73' | 'AGVQVWLGANGK' | 'IgGFc-binding protein' |
| 15119 | 'FCGBP_No74' | 'LDDGDYLCEDGCQNNCPACTPGQAQHYEGDRLCGMLTK' | 'IgGFc-binding protein' |
| 15120 | 'FCGBP_No75' | 'LDSLVAQQLQSK' | 'IgGFc-binding protein' |
| 15121 | 'FCGBP_No76' | 'LDGPFAVCHDTLDPR' | 'IgGFc-binding protein' |
| 15122 | 'FCGBP_No77' | 'KFDFQGTCNYVLATTGCPGVSTQGLTPFTVTTK' | 'IgGFc-binding protein' |
| 15123 | 'FCGBP_No78' | 'TEAVGQVHIFFQDGMVTLTPNK' | 'IgGFc-binding protein' |
| 15124 | 'FCGBP_No79' | 'VTVNGVDMKLPVVLANGQIR' | 'IgGFc-binding protein' |
| 15125 | 'COLEC10_No1' | 'DEAANTLIADYVAK' | 'Collectin-10' |
| 15126 | 'COLEC10_No2' | 'FVGQLDISIAR' | 'Collectin-10' |
| 15127 | 'COLEC10_No3' | 'GGMLAMPKDEAANTLIADYVAK' | 'Collectin-10' |
| 15128 | 'COLEC10_No4' | 'NVIAGIRETEEK' | 'Collectin-10' |
| 15129 | 'COLEC10_No5' | 'VFIGVNDLER' | 'Collectin-10' |
| 15130 | 'COLEC10_No6' | 'ETEEKFYYIVQEEK' | 'Collectin-10' |
