## Supplemental Material 4 for "Identification of candidate biomarkers and molecular networks associated with Pulmonary Arterial Hypertension using machine learning and plasma multi-Omics analysis"

# Z10

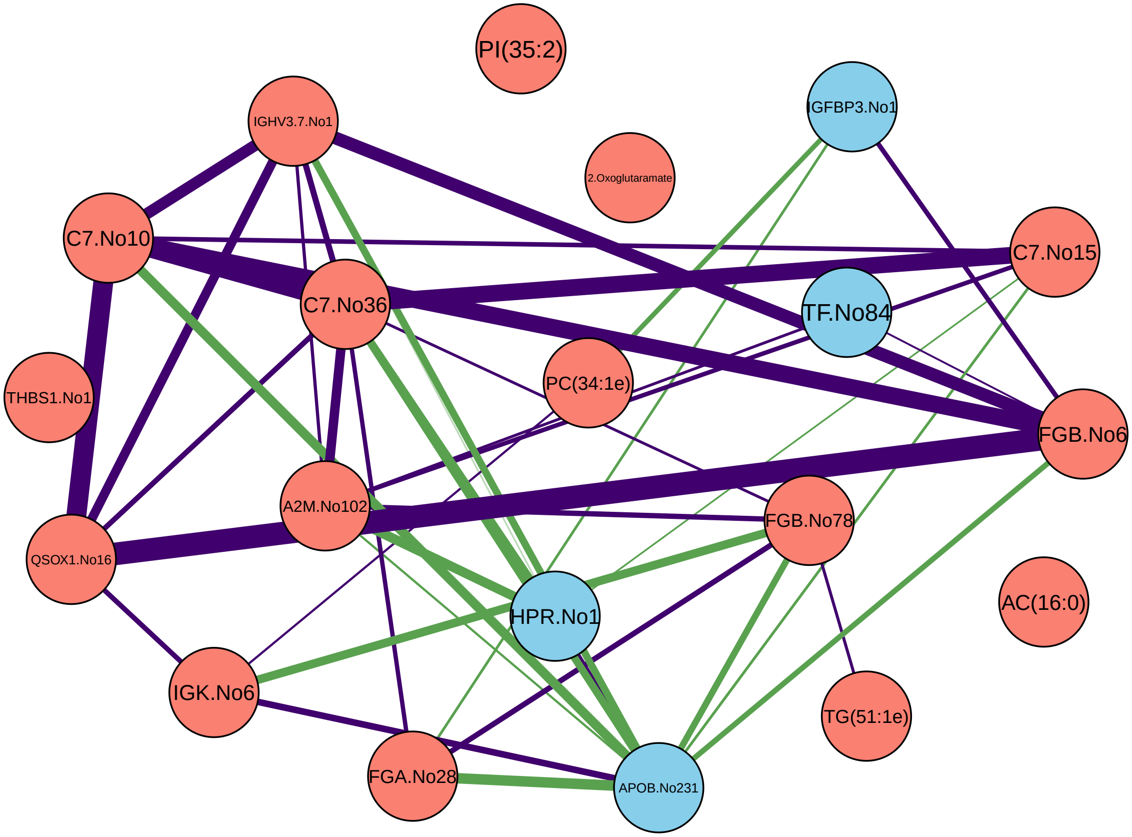
PI(35:2)
IGFBP3.No1
IGHV3.7.No1
2.Oxoglutaramate
C7.No10
C7.No15
C7.No36
TF.No84
PC(34:1e)
THBS1.No1
FGB.No6
A2M.No102
FGB.No78
QSOX1.No16
HPR.No1
AC(16:0)
IGK.No6
TG(51:1e)
FGA.No28
APOB.No231

Z12

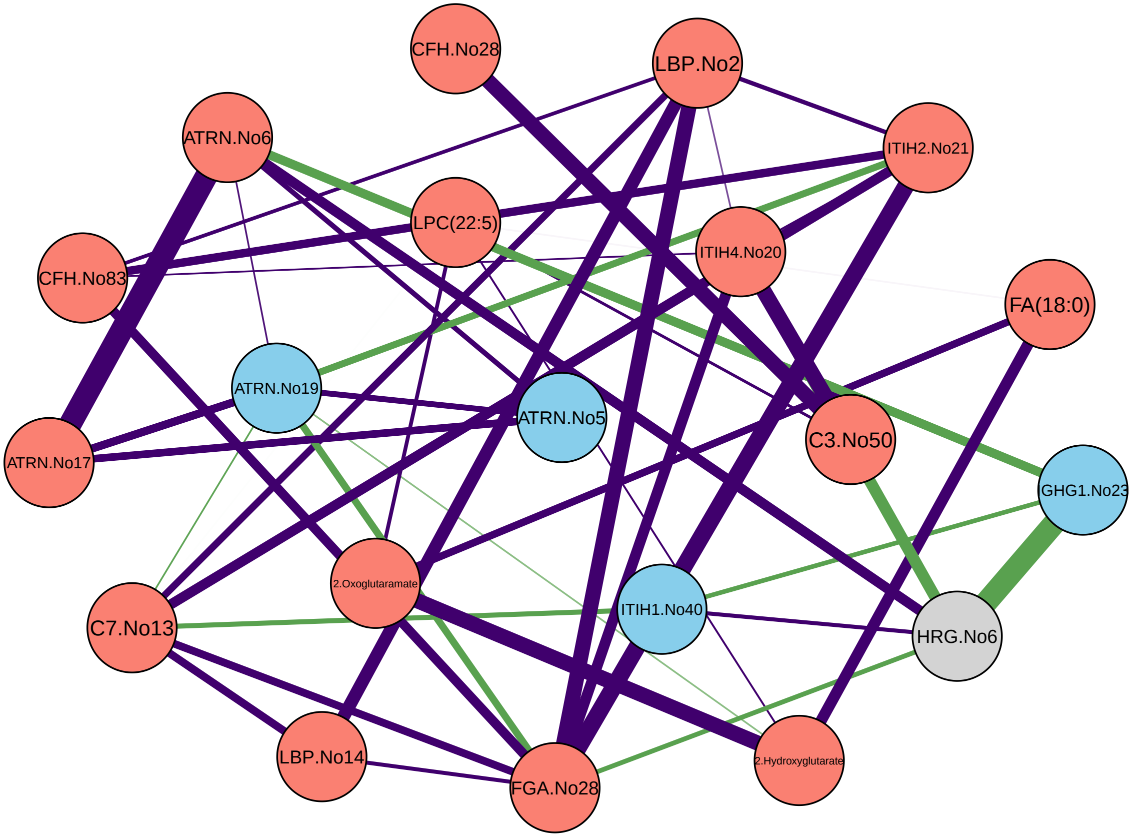

CFH.No28
LBP.No2
ATRN.No6
ITIH2.No21
LPC(22:5)
ITIH4.No20
CFH.No83
FA(18:0)
ATRN.No19
ATRN.No5
C3.No50
ATRN.No17
GHG1.No23
2.Oxoglutaramate
ITIH1.No40
HRG.No6
C7.No13
LBP.No14
FGA.No28
2.Hydroxyglutarate

Z24

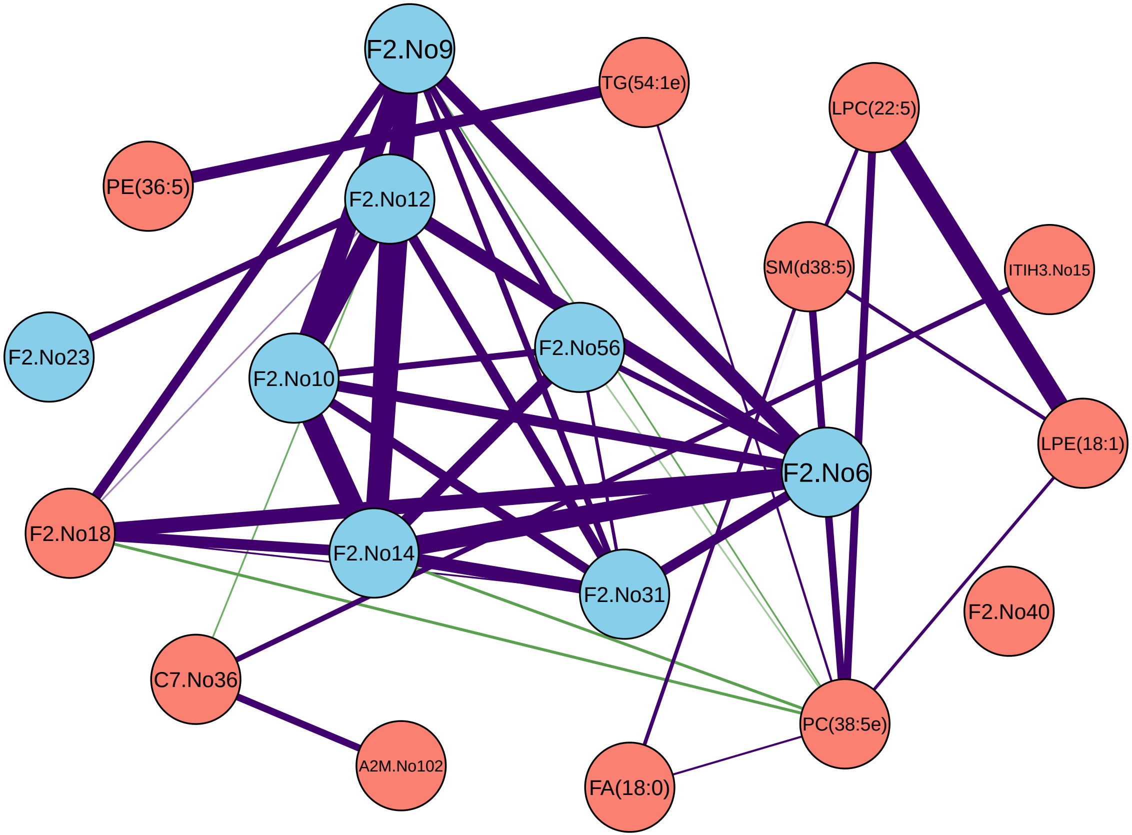

F2.No9
TG(54:1e)
LPC(22:5)
PE(36:5)
F2.No12
SM(d38:5)
ITIH3.No15
F2.No23
F2.No56
F2.No10
LPE(18:1)
F2.No6
F2.No18
F2.No14
F2.No31
F2.No40
C7.No36
PC(38:5e)
A2M.No102
FA(18:0)

# Z58

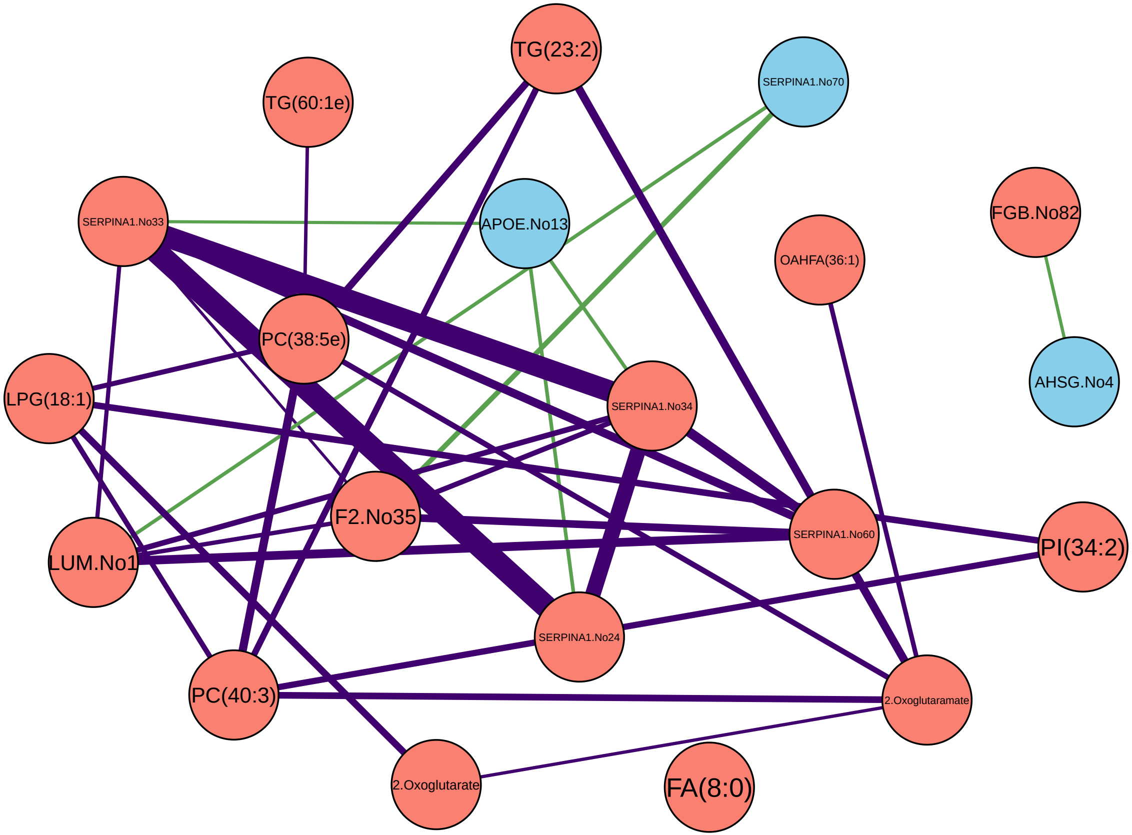
TG(23:2)
SERPINA1.No70
TG(60:1e)
SERPINA1.No33
APOE.No13
FGB.No82
OAHFA(36:1)
PC(38:5e)
AHSG.No4
LPG(18:1)
SERPINA1.No34
F2.No35
SERPINA1.No60
LUM.No1
PI(34:2)
SERPINA1.No24
PC(40:3)
2.Oxoglutaramate
2.Oxoglutarate
FA(8:0)

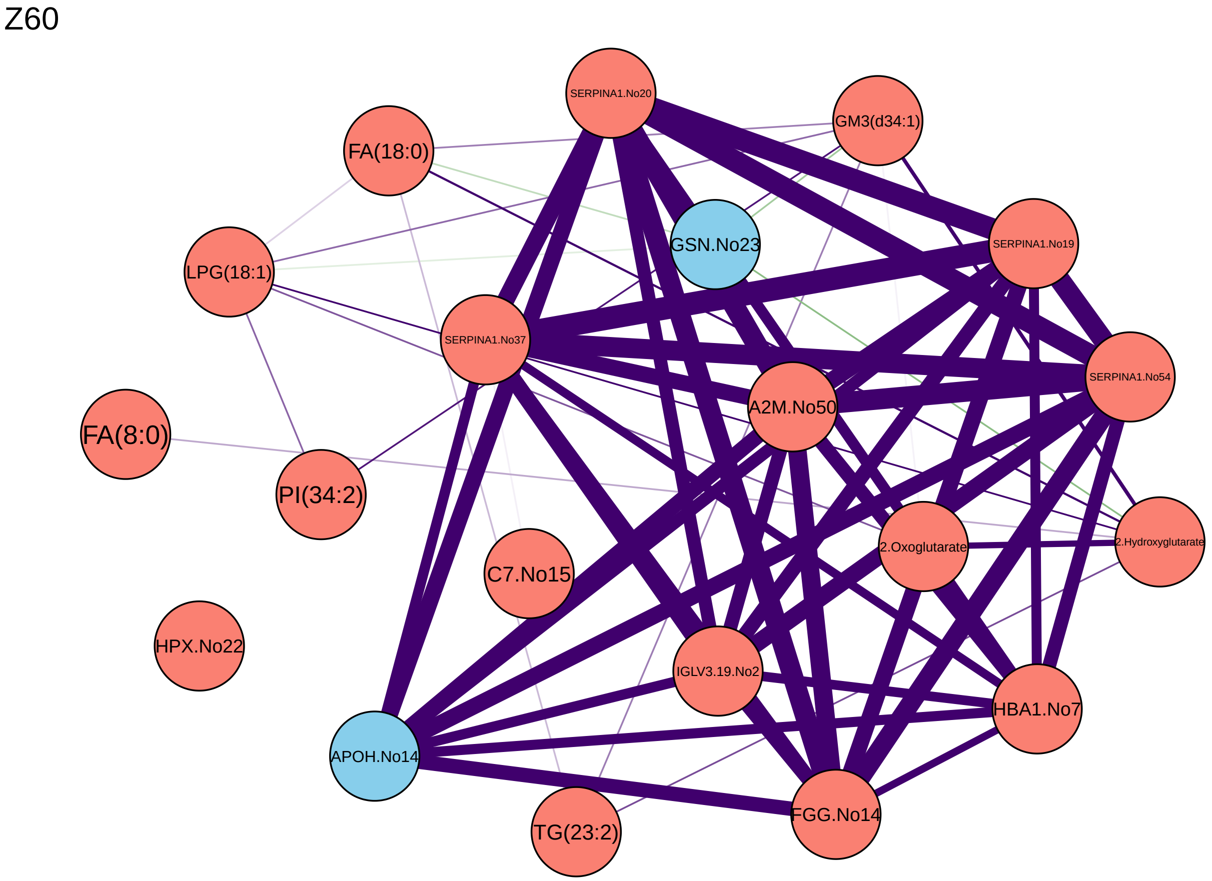

Z60
SERPINA1.No20
GM3(d34:1)
FA(18:0)
GSN.No23
LPG(18:1)
SERPINA1.No19
SERPINA1.No37
SERPINA1.No54
A2M.No50
FA(8:0)
PI(34:2)
2.Hydroxyglutarate
2.Oxoglutarate
C7.No15
HPX.No22
IGLV3.19.No2
HBA1.No7
APOH.No14
FGG.No14
TG(23:2)

Z74

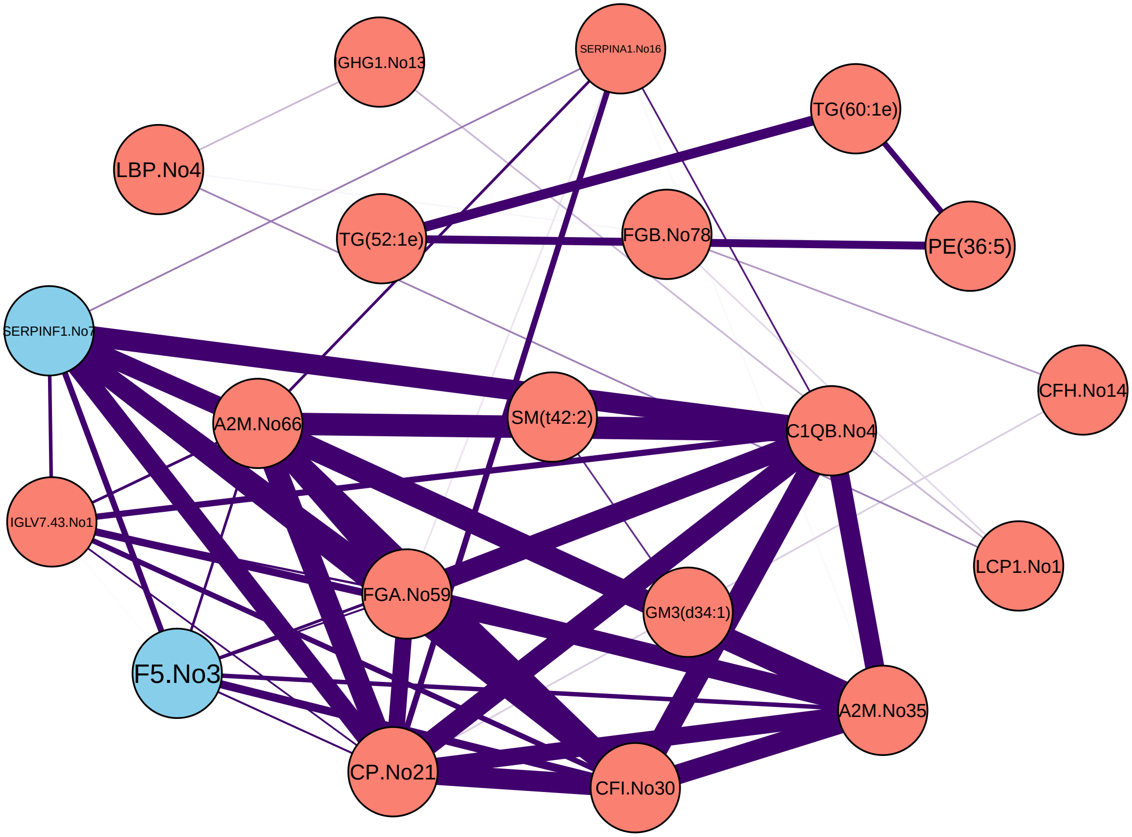

GHG1.No13
SERPINA1.No16
TG(60:1e)
LBP.No4
TG(52:1e)
FGB.No78
PE(36:5)
SERPINF1.No7
CFH.No14
A2M.No66
SM(t42:2)
C1QB.No4
IGLV7.43.No1
LCP1.No1
FGA.No59
GM3(d34:1)
F5.No3
A2M.No35
CP.No21
CFI.No30

Z81

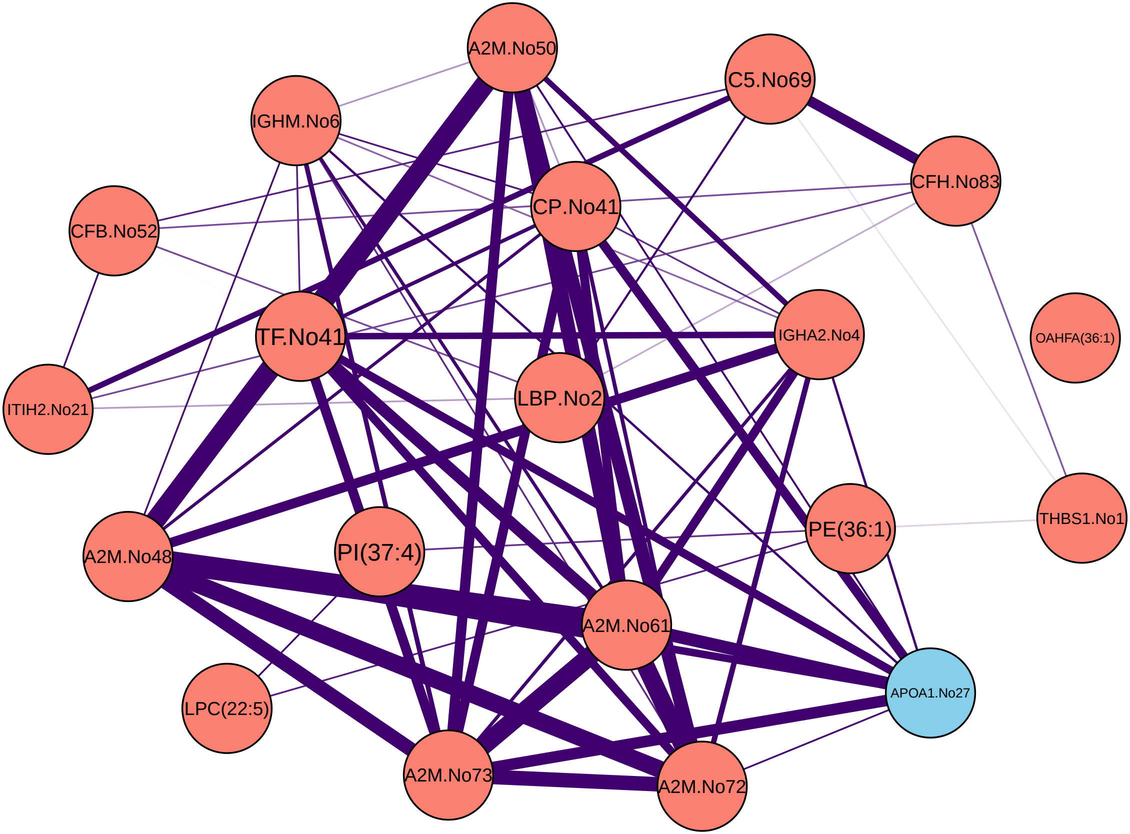

A2M.No50
C5.No69
IGHM.No6
CFH.No83
CP.No41
CFB.No52
IGHA2.No4
OAHFA(36:1)
TF.No41
ITIH2.No21
LBP.No2
THBS1.No1
PE(36:1)
A2M.No48
PI(37:4)
A2M.No61
APOA1.No27
LPC(22:5)
A2M.No73
A2M.No72

Z86

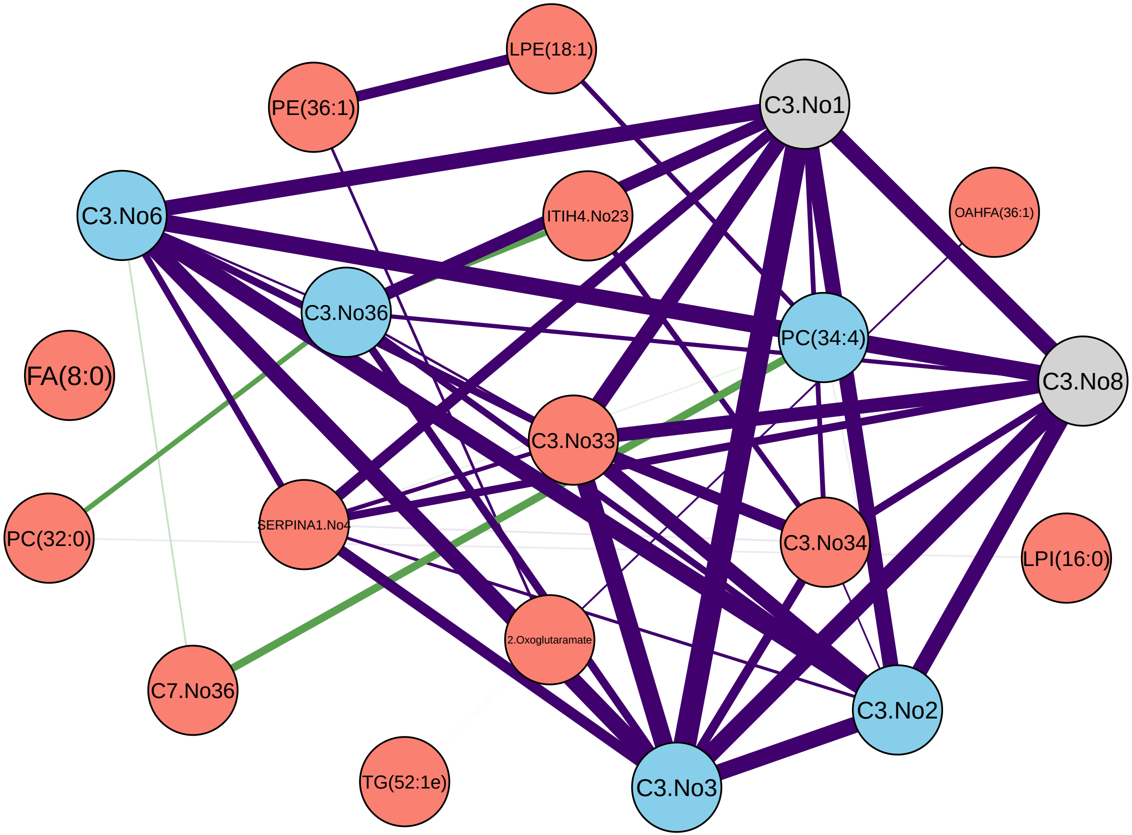

LPE(18:1)
PE(36:1)
C3.No1
C3.No6
ITIH4.No23
OAHFA(36:1)
C3.No36
PC(34:4)
FA(8:0)
C3.No8
C3.No33
PC(32:0)
SERPINA1.No4
C3.No34
LPI(16:0)
2.Oxoglutaramate
C7.No36
C3.No2
TG(52:1e)
C3.No3

Z100

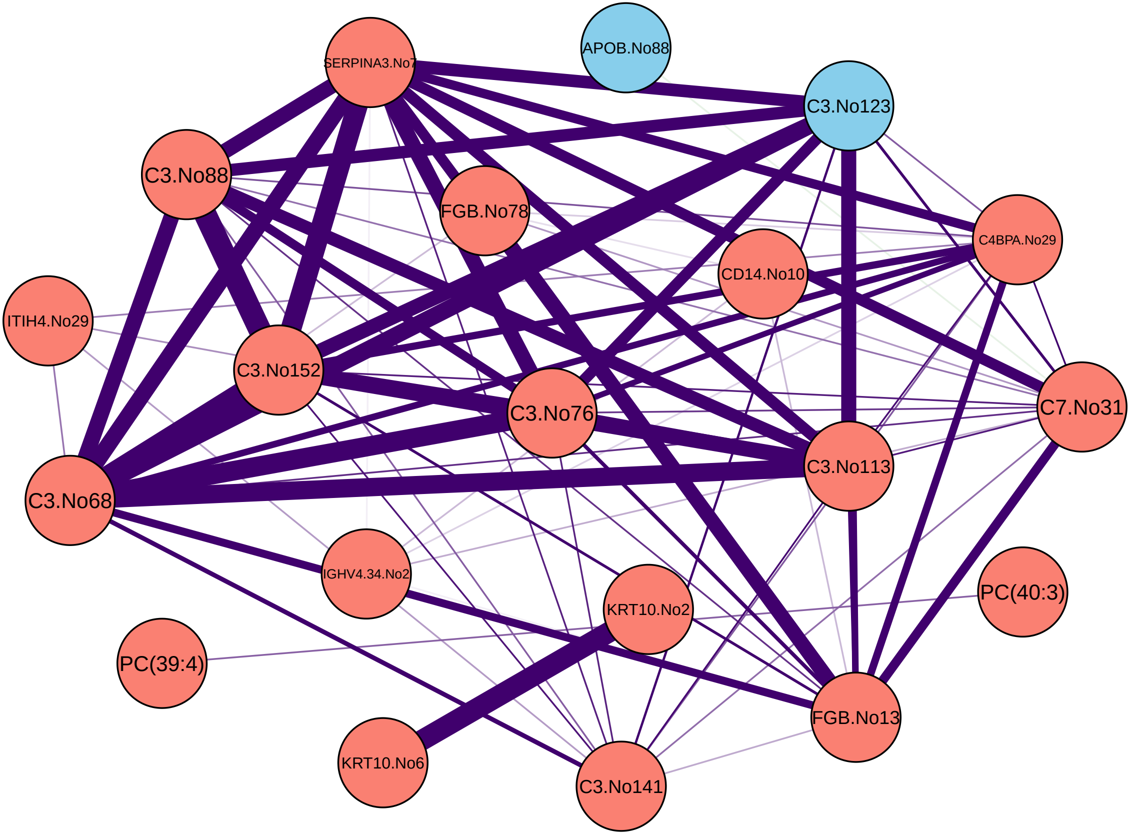
SERPINA3.No7
APOB.No88
C3.No123
C3.No88
FGB.No78
C4BPA.No29
CD14.No10
ITIH4.No29
C3.No152
C3.No76
C7.No31
C3.No68
C3.No113
IGHV4.34.No2
KRT10.No2
PC(40:3)
PC(39:4)
FGB.No13
KRT10.No6
C3.No141

Z133

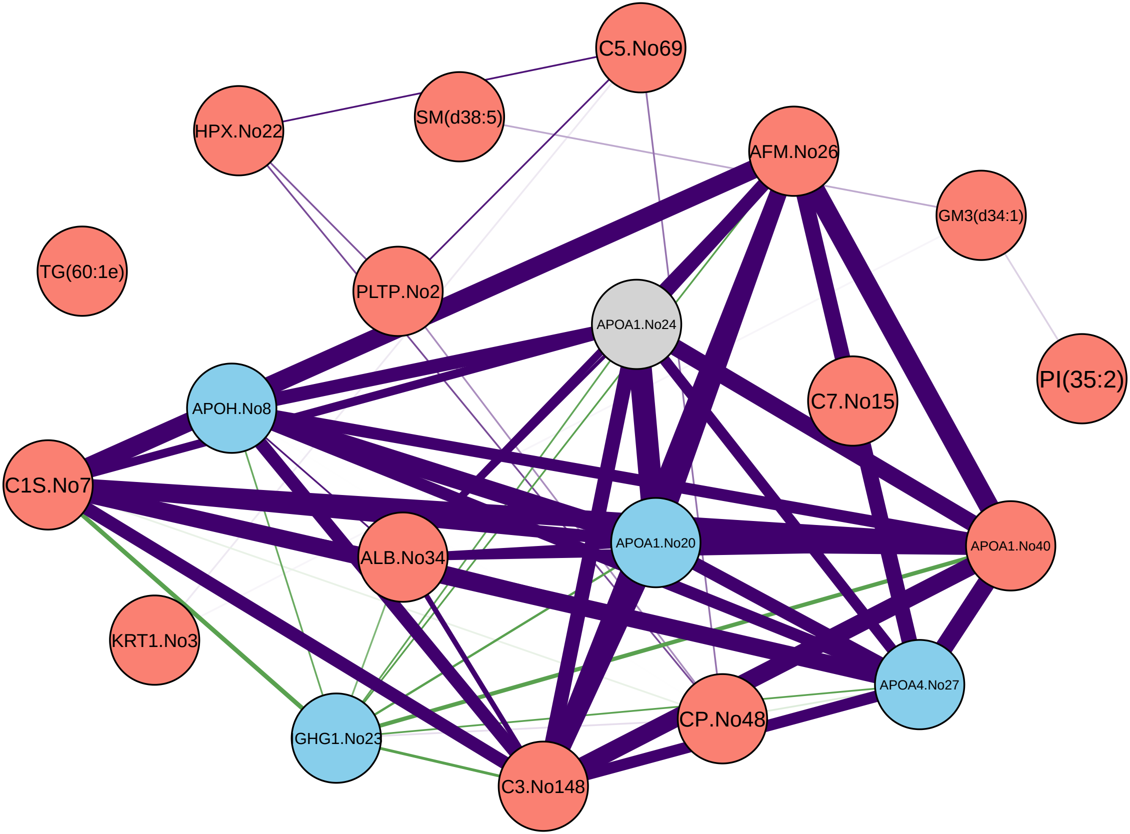
C5.No69
HPX.No22
SM(d38:5)
AFM.No26
GM3(d34:1)
TG(60:1e)
PLTP.No2
APOA1.No24
PI(35:2)
C7.No15
APOH.No8
C1S.No7
APOA1.No20
APOA1.No40
ALB.No34
KRT1.No3
APOA4.No27
CP.No48
GHG1.No23
C3.No148

Z152

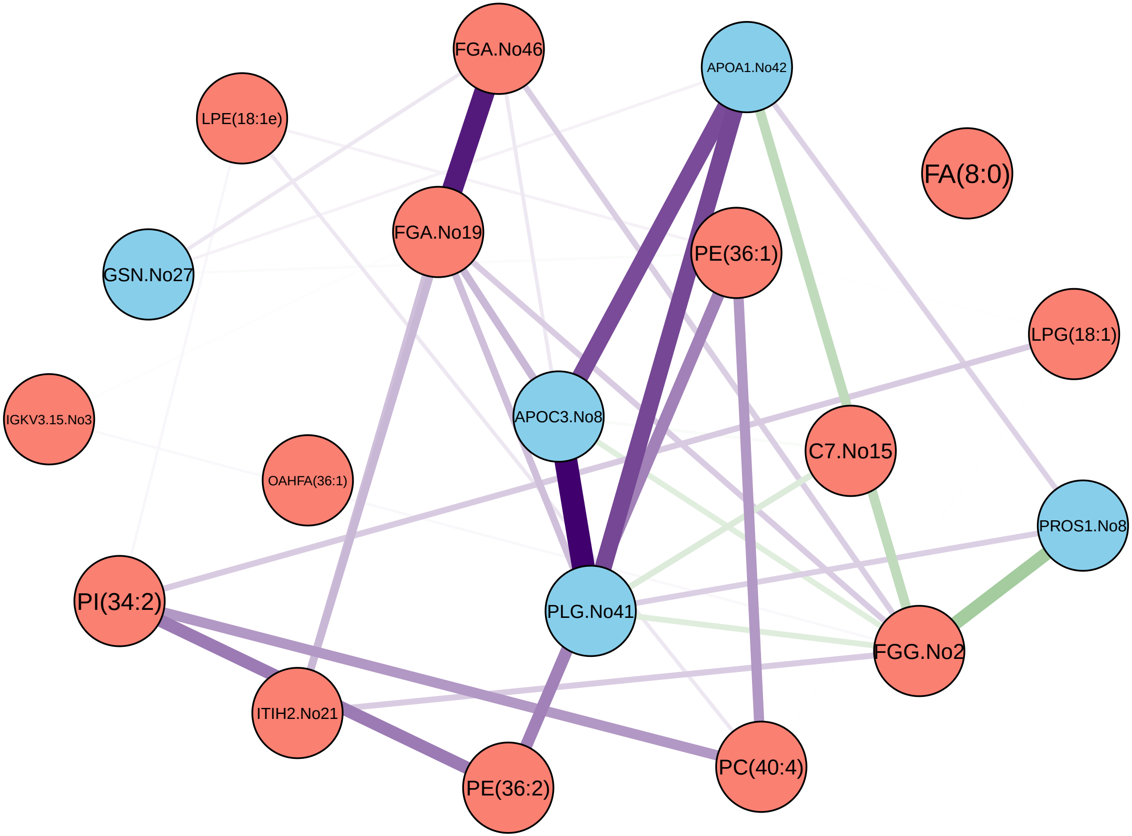

FGA.No46
APOA1.No42
LPE(18:1e)
FA(8:0)
FGA.No19
PE(36:1)
GSN.No27
LPG(18:1)
APOC3.No8
IGKV3.15.No3
C7.No15
OAHFA(36:1)
PROS1.No8
PI(34:2)
PLG.No41
FGG.No2
ITIH2.No21
PC(40:4)
PE(36:2)

Z159

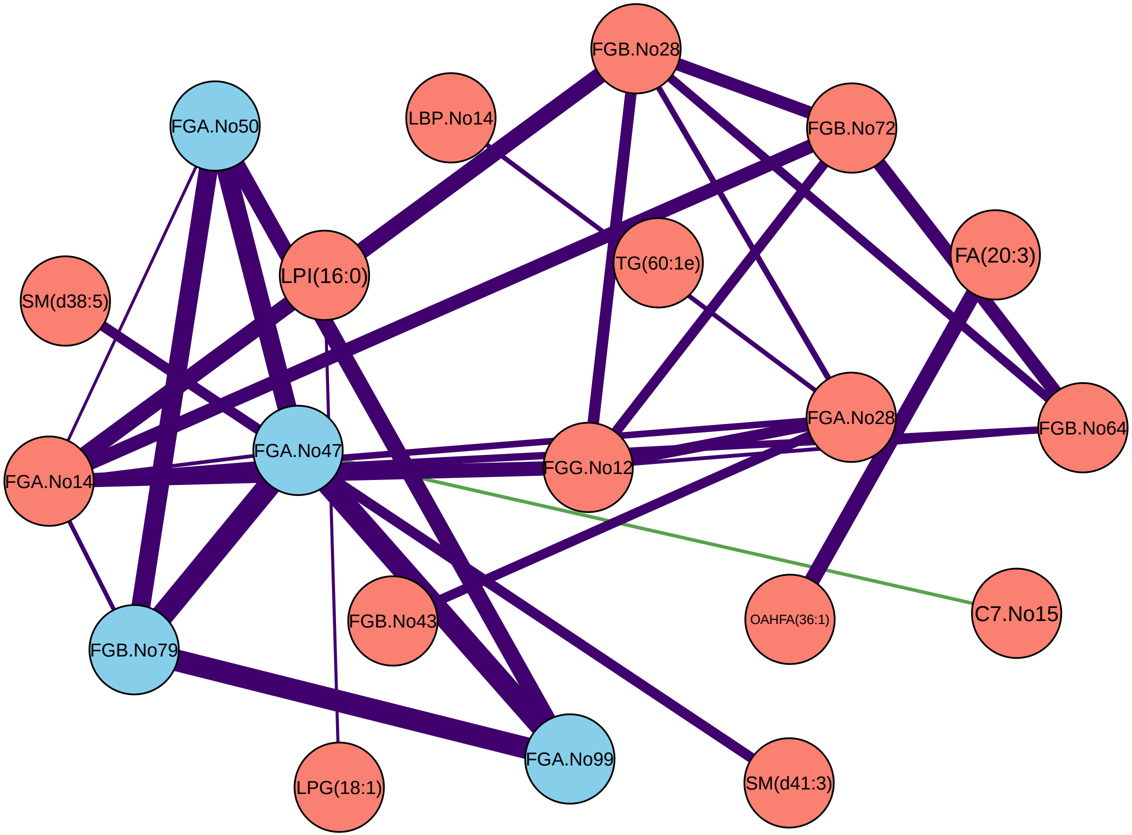

FGB.No28
FGA.No50
LBP.No14
FGB.No72
LPI(16:0)
TG(60:1e)
FA(20:3)
SM(d38:5)
FGA.No28
FGB.No64
FGA.No47
FGA.No14
FGG.No12
FGB.No43
OAHFA(36:1)
C7.No15
FGB.No79
FGA.No99
LPG(18:1)
SM(d41:3)

Z176

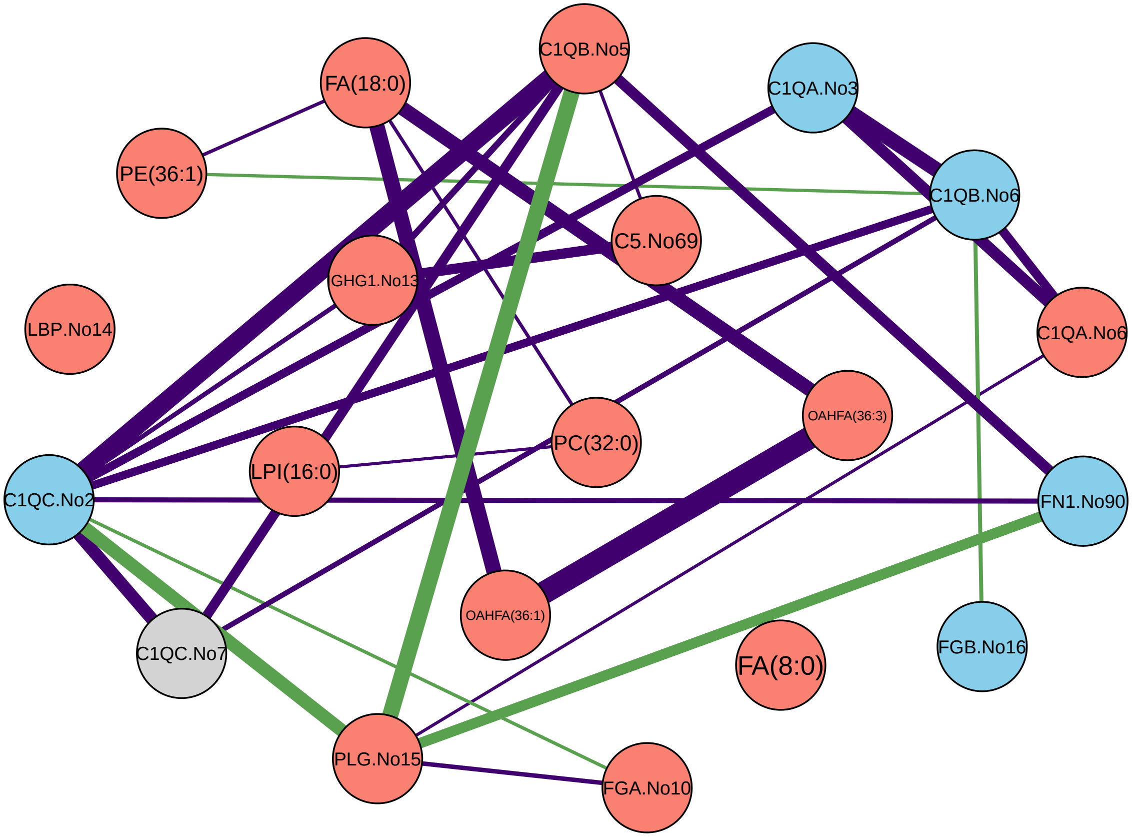

C1QB.No5
FA(18:0)
C1QA.No3
PE(36:1)
C1QB.No6
C5.No69
GHG1.No13
LBP.No14
C1QA.No6
OAHFA(36:3)
PC(32:0)
LPI(16:0)
C1QC.No2
FN1.No90
OAHFA(36:1)
C1QC.No7
FA(8:0)
FGB.No16
PLG.No15
FGA.No10

Z182

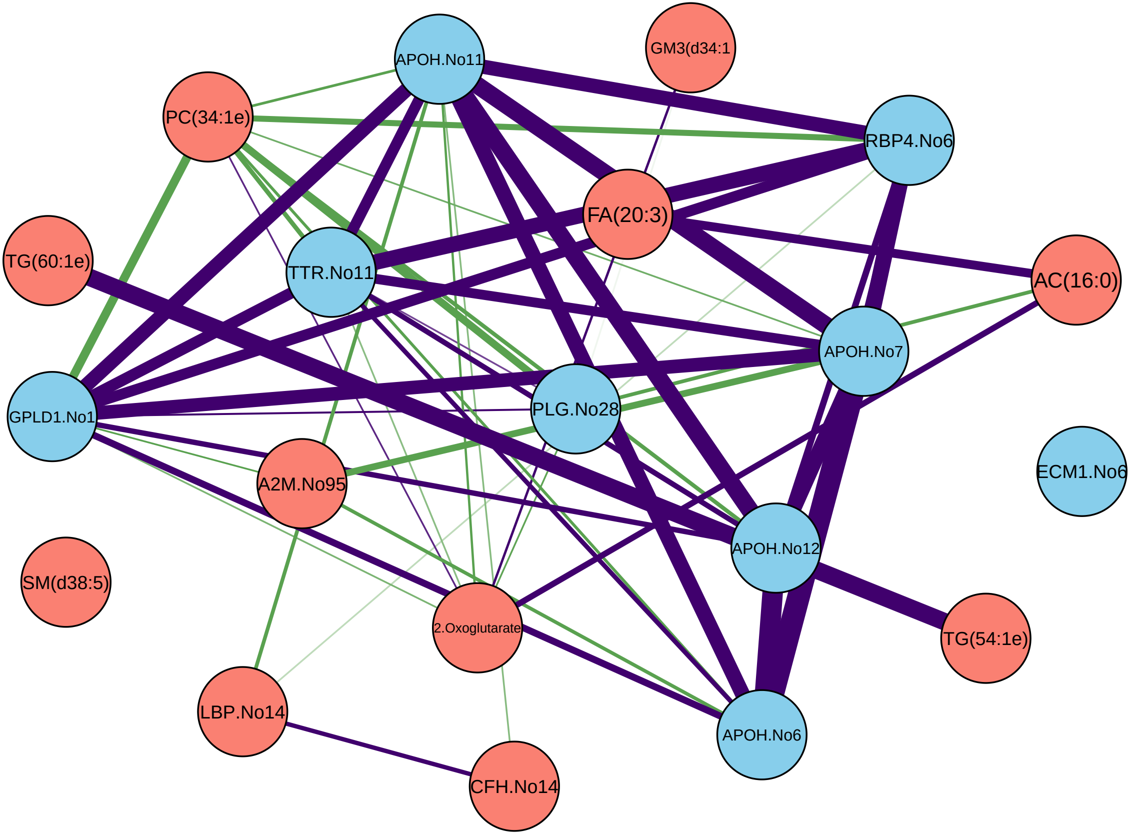

APOH.No11
GM3(d34:1
PC(34:1e)
RBP4.No6
FA(20:3)
TG(60:1e)
TTR.No11
AC(16:0)
APOH.No7
GPLD1.No1
PLG.No28
ECM1.No6
A2M.No95
APOH.No12
SM(d38:5)
2.Oxoglutarate
TG(54:1e)
LBP.No14
APOH.No6
CFH.No14

Z209

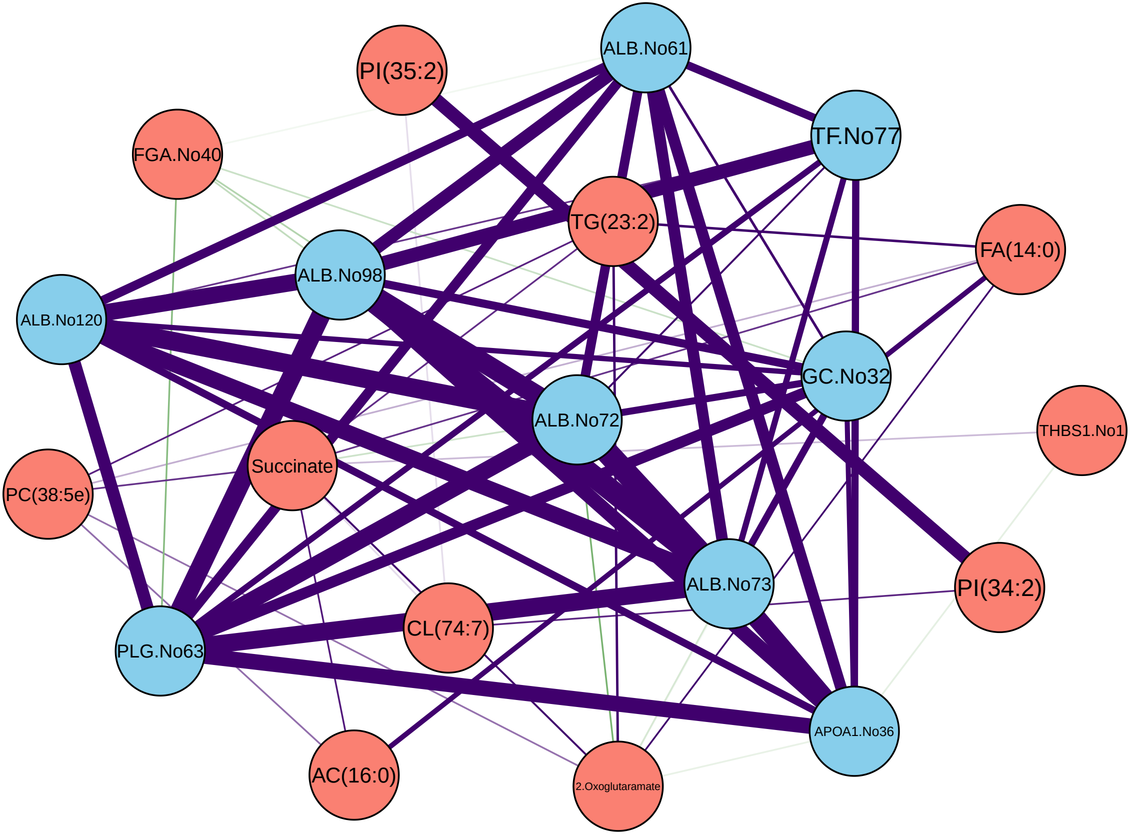
ALB.No61
PI(35:2)
TF.No77
FGA.No40
TG(23:2)
FA(14:0)
ALB.No98
ALB.No120
GC.No32
ALB.No72
THBS1.No1
Succinate
PC(38:5e)
ALB.No73
PI(34:2)
CL(74:7)
PLG.No63
APOA1.No36
AC(16:0)
2.Oxoglutaramate

Z233

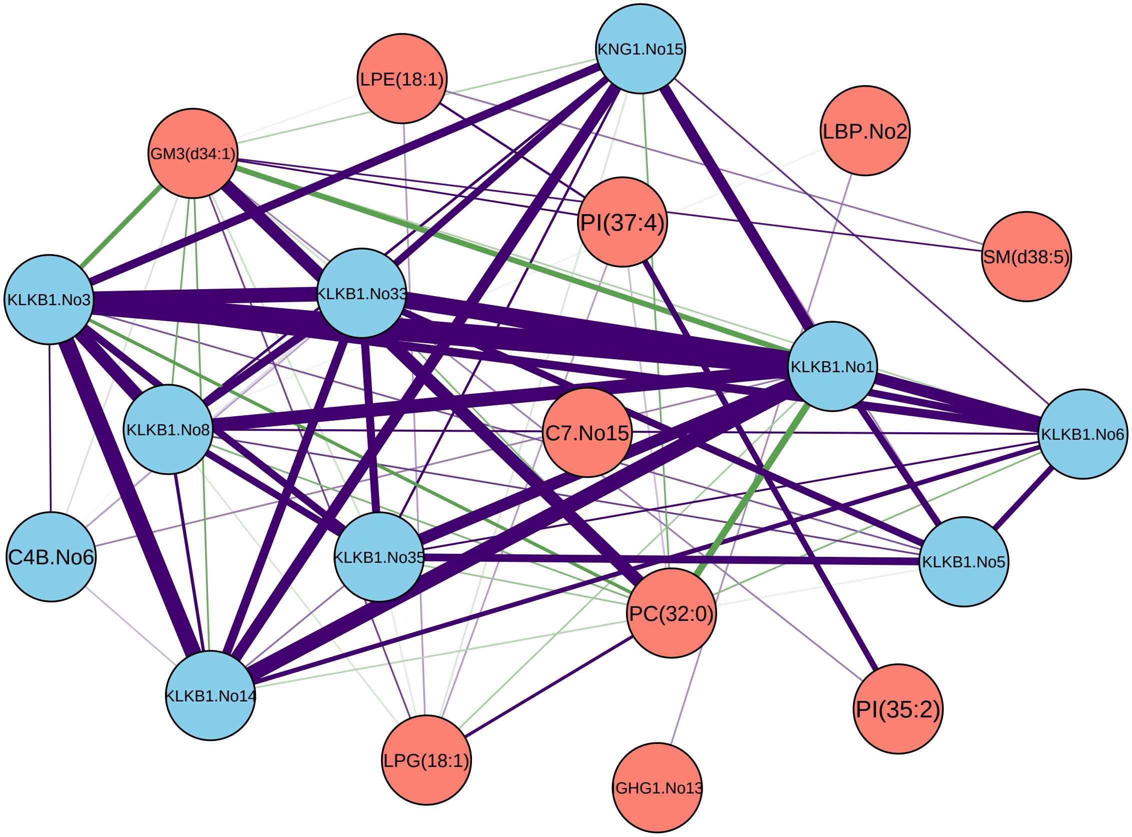
KNG1.No15
LPE(18:1)
LBP.No2
GM3(d34:1)
PI(37:4)
SM(d38:5)
KLKB1.No3
KLKB1.No33
KLKB1.No1
KLKB1.No8
C7.No15
KLKB1.No6
C4B.No6
KLKB1.No35
KLKB1.No5
PC(32:0)
KLKB1.No14
PI(35:2)
LPG(18:1)
GHG1.No13

Z261

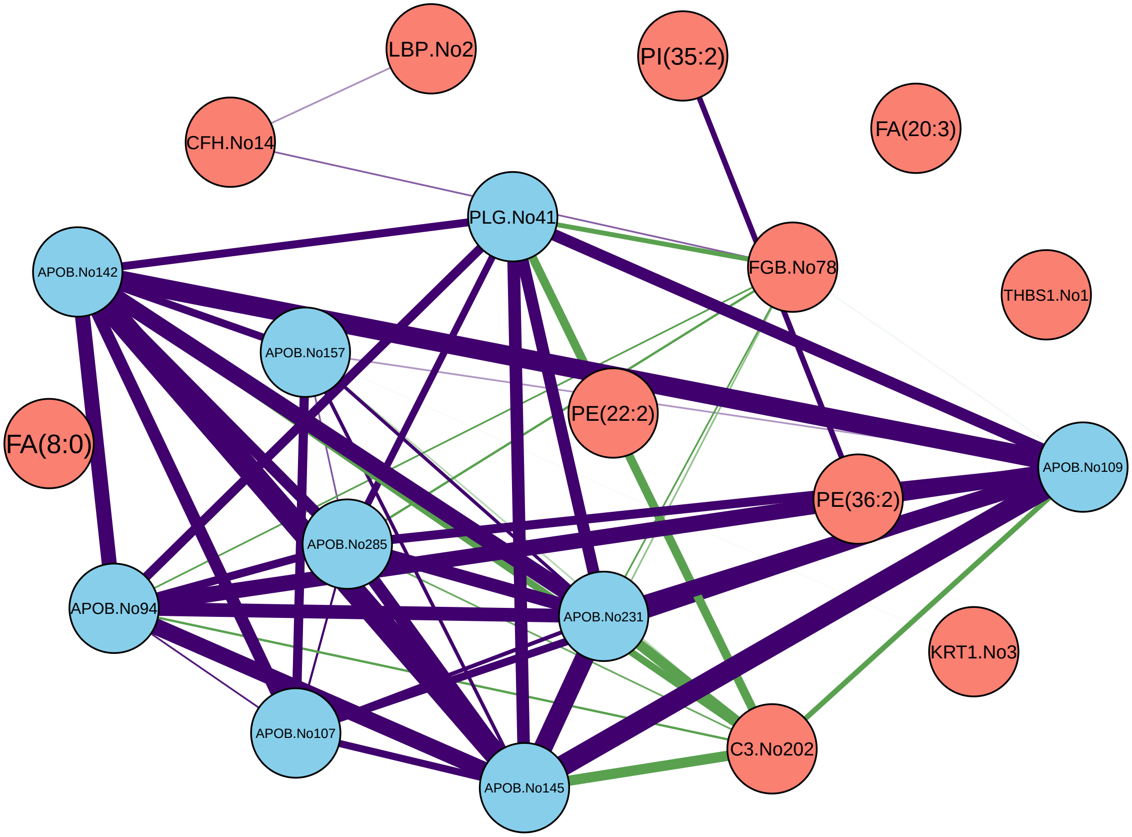
LBP.No2
PI(35:2)
CFH.No14
FA(20:3)
PLG.No41
APOB.No142
FGB.No78
THBS1.No1
APOB.No157
PE(22:2)
FA(8:0)
APOB.No109
PE(36:2)
APOB.No285
APOB.No94
APOB.No231
KRT1.No3
APOB.No107
C3.No202
APOB.No145

Z308

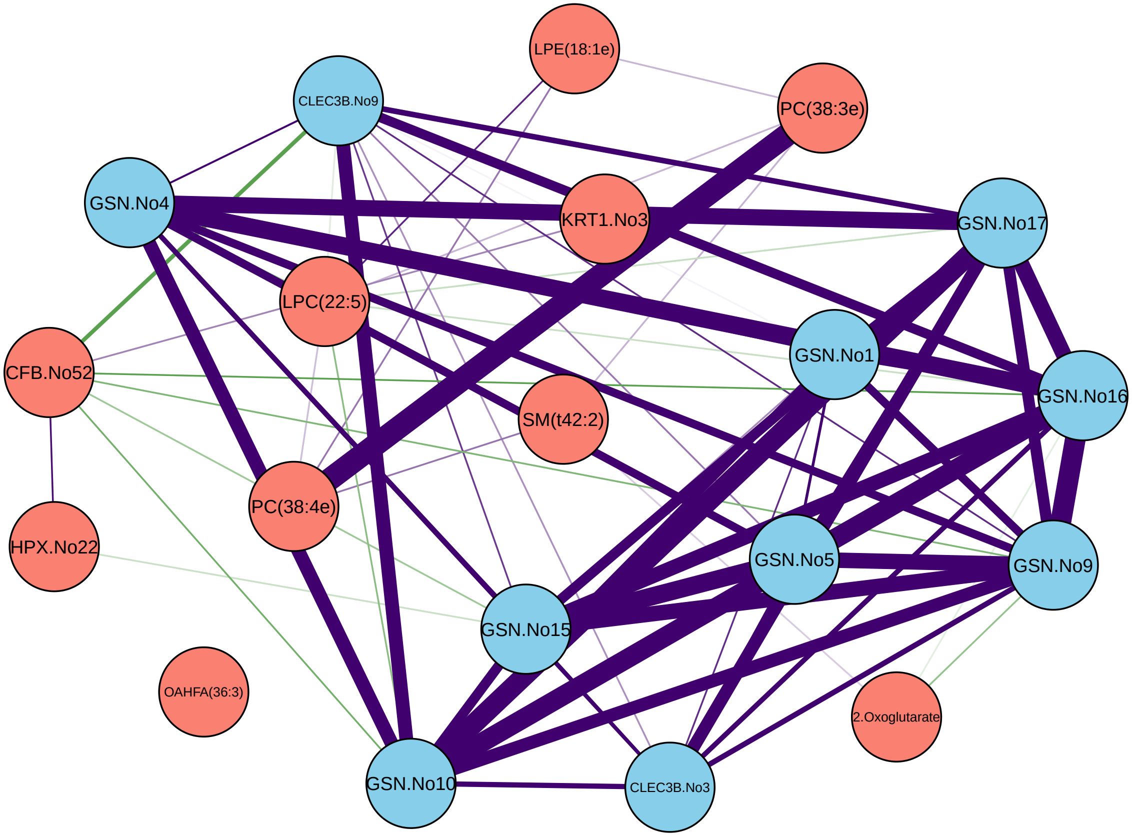
LPE(18:1e)
CLEC3B.No9
PC(38:3e)
GSN.No4
KRT1.No3
GSN.No17
LPC(22:5)
CFB.No52
GSN.No1
GSN.No16
SM(t42:2)
PC(38:4e)
HPX.No22
GSN.No5
GSN.No9
GSN.No15
OAHFA(36:3)
2.Oxoglutarate
GSN.No10
CLEC3B.No3

Z323

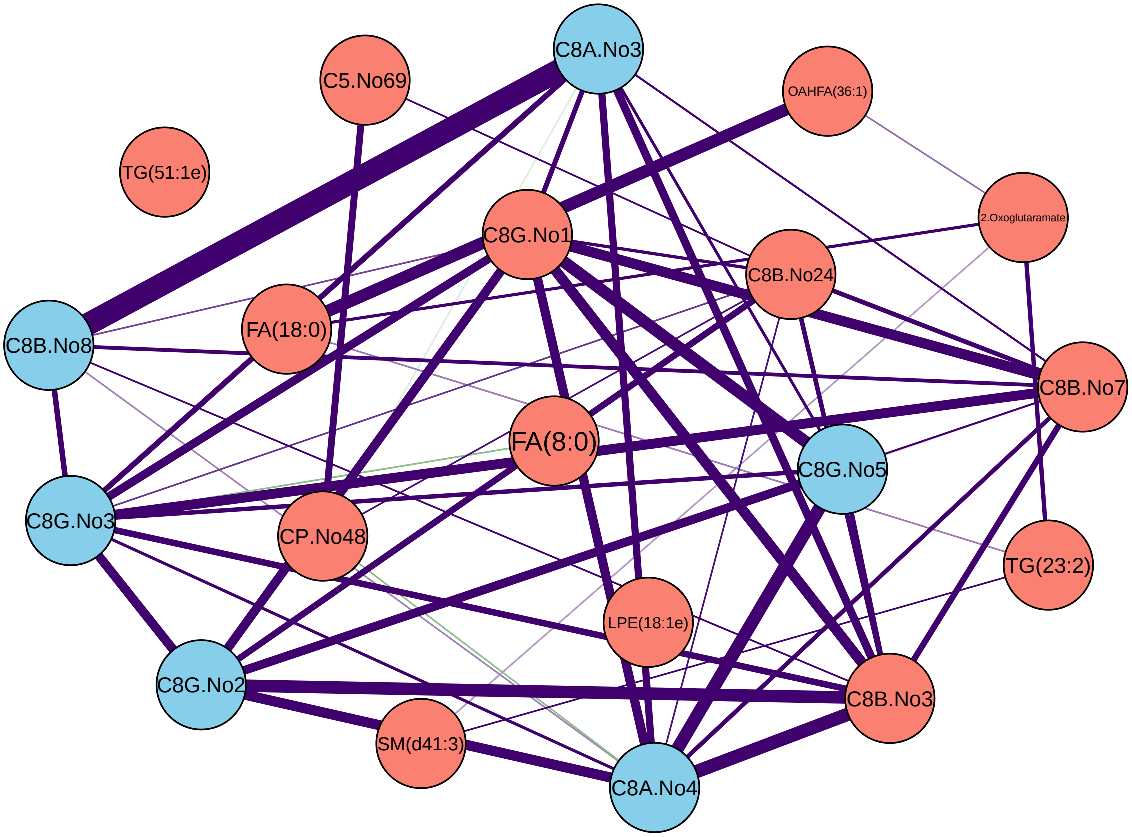

C8A.No3
C5.No69
OAHFA(36:1)
TG(51:1e)
2.Oxoglutaramate
C8G.No1
C8B.No24
C8B.No8
FA(18:0)
C8B.No7
FA(8:0)
C8G.No5
C8G.No3
CP.No48
TG(23:2)
LPE(18:1e)
C8G.No2
C8B.No3
SM(d41:3)
C8A.No4

Z338

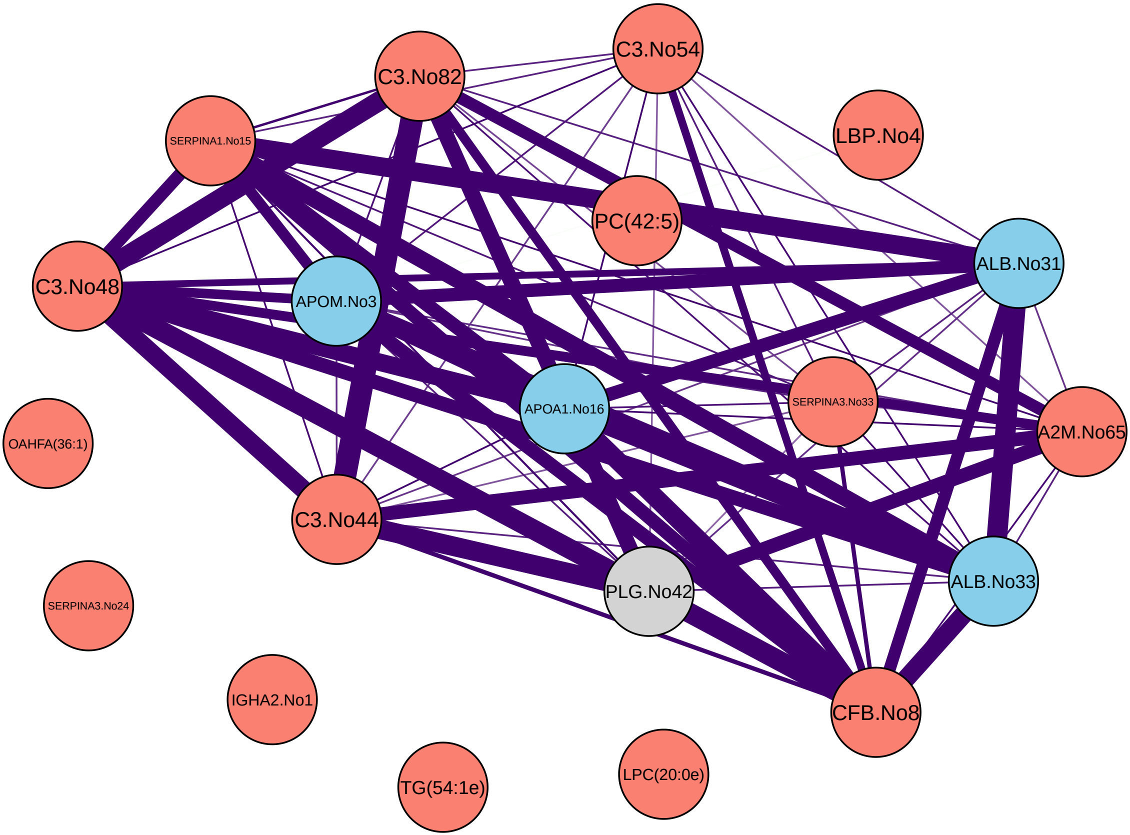

C3.No54
C3.No82
SERPINA1.No15
LBP.No4
PC(42:5)
ALB.No31
C3.No48
APOM.No3
APOA1.No16
SERPINA3.No33
A2M.No65
OAHFA(36:1)
C3.No44
PLG.No42
ALB.No33
SERPINA3.No24
IGHA2.No1
CFB.No8
TG(54:1e)
LPC(20:0e)

Z341

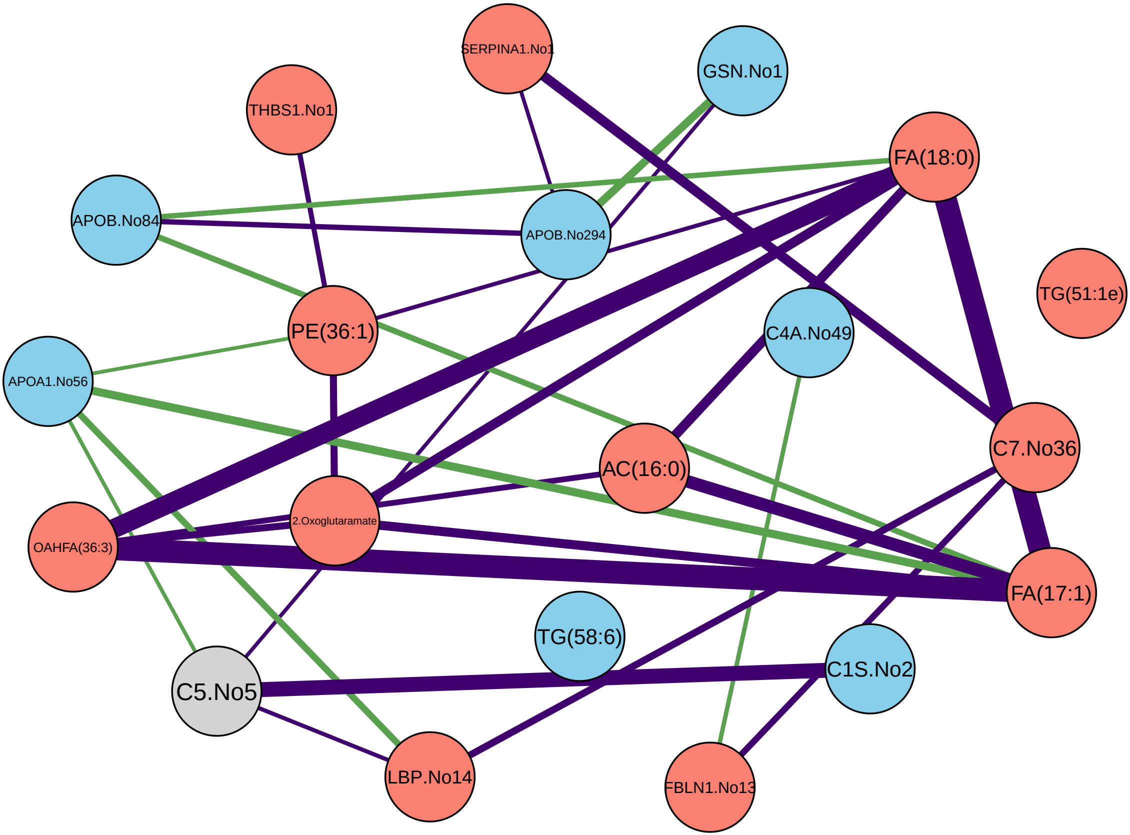
SERPINA1.No1
GSN.No1
THBS1.No1
FA(18:0)
APOB.No84
APOB.No294
TG(51:1e)
C4A.No49
PE(36:1)
APOA1.No56
AC(16:0)
C7.No36
2.Oxoglutaramate
OAHFA(36:3)
FA(17:1)
TG(58:6)
C1S.No2
C5.No5
LBP.No14
FBLN1.No13

Z344

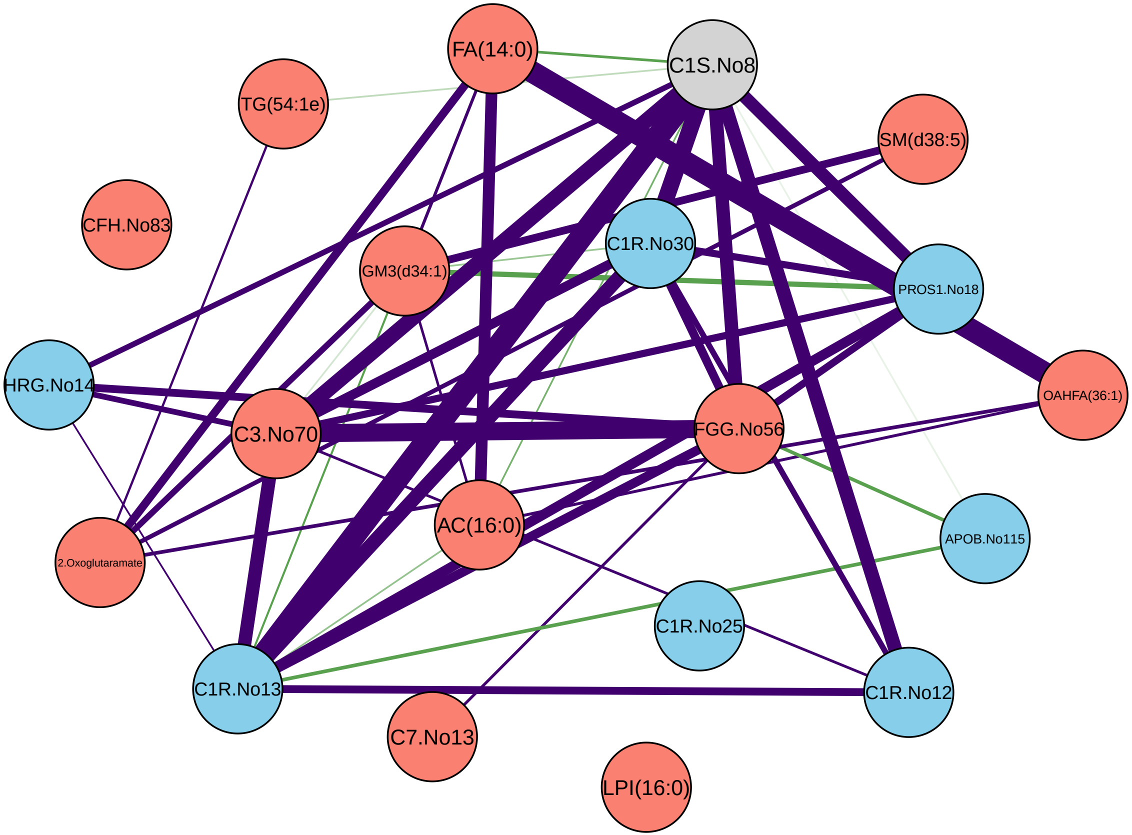
FA(14:0)
C1S.No8
TG(54:1e)
SM(d38:5)
CFH.No83
C1R.No30
GM3(d34:1)
PROS1.No18
HRG.No14
OAHFA(36:1)
C3.No70
FGG.No56
AC(16:0)
APOB.No115
2.Oxoglutaramate
C1R.No25
C1R.No13
C1R.No12
C7.No13
LPI(16:0)

Z380

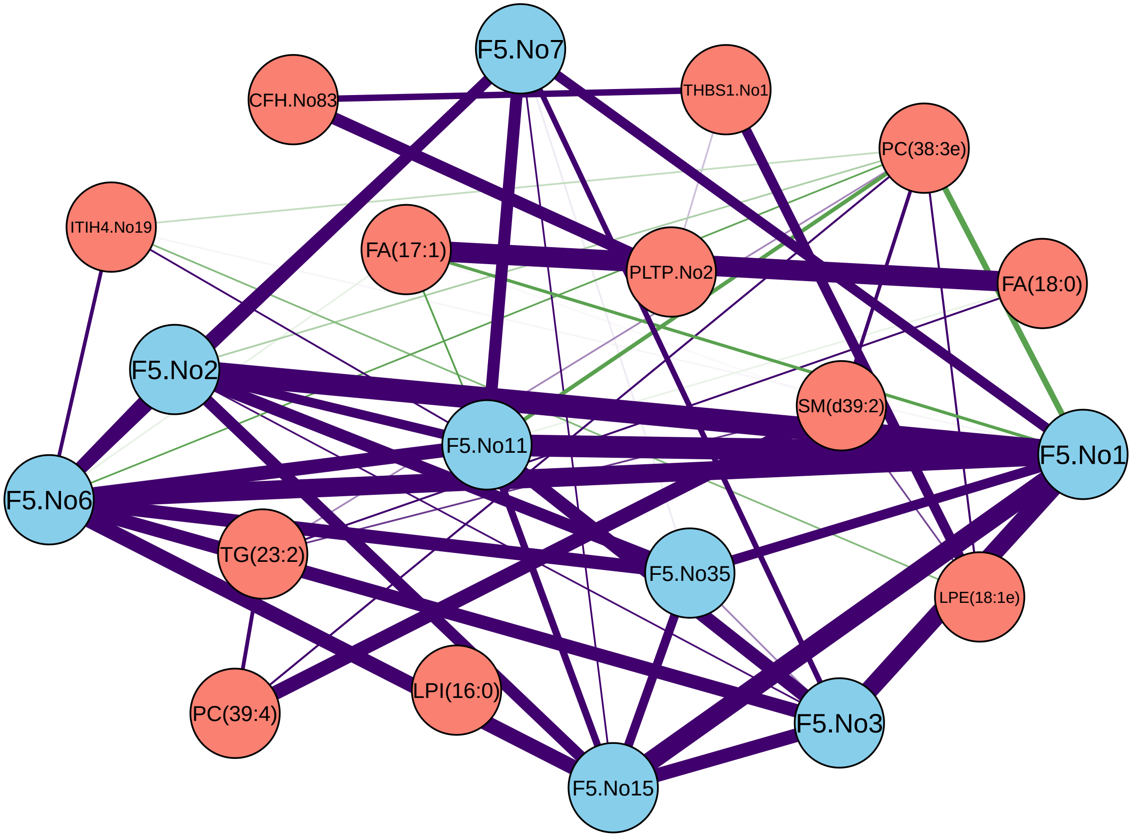

F5.No7
CFH.No83
THBS1.No1
PC(38:3e)
ITIH4.No19
FA(17:1)
PLTP.No2
FA(18:0)
F5.No2
SM(d39:2)
F5.No11
F5.No1
F5.No6
TG(23:2)
F5.No35
LPE(18:1e)
PC(39:4)
LPI(16:0)
F5.No3
F5.No15

Z384

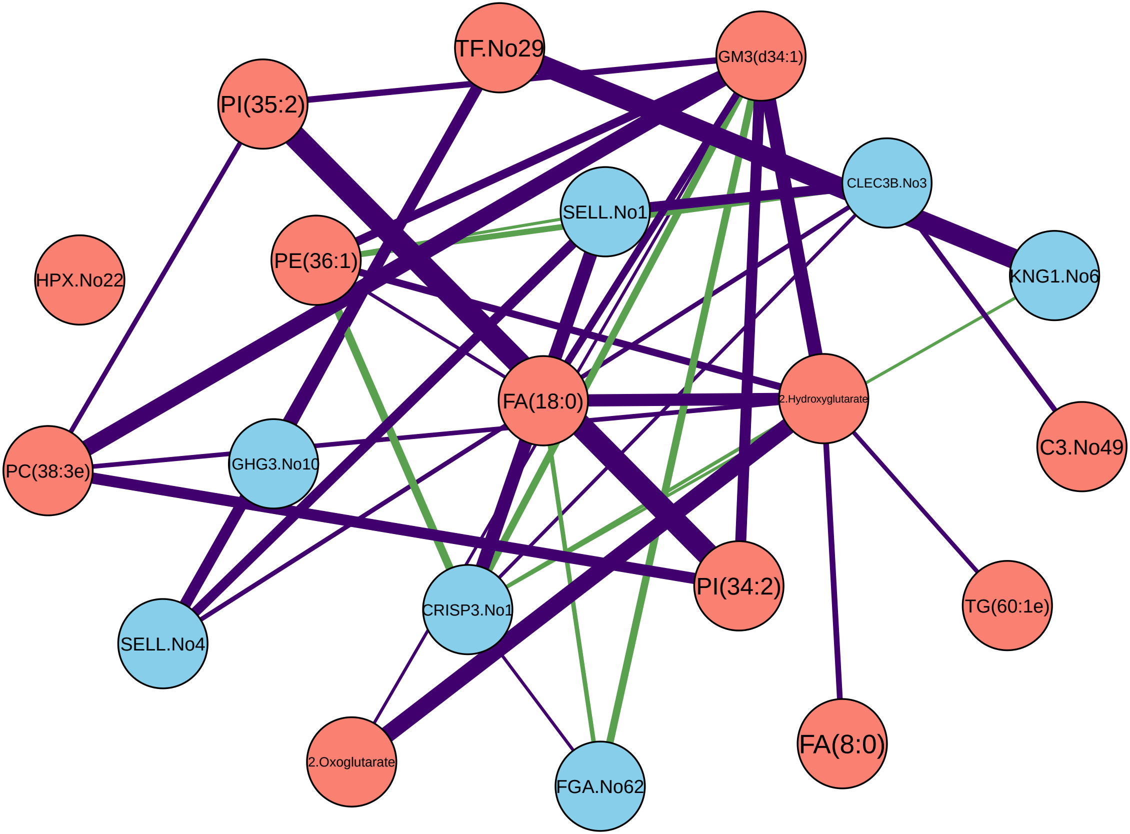
TF.No29
GM3(d34:1)
PI(35:2)
CLEC3B.No3
SELL.No1
HPX.No22
PE(36:1)
KNG1.No6
FA(18:0)
2.Hydroxyglutarate
C3.No49
PC(38:3e)
GHG3.No10
PI(34:2)
TG(60:1e)
SELL.No4
CRISP3.No1
2.Oxoglutarate
FGA.No62
FA(8:0)

Z414

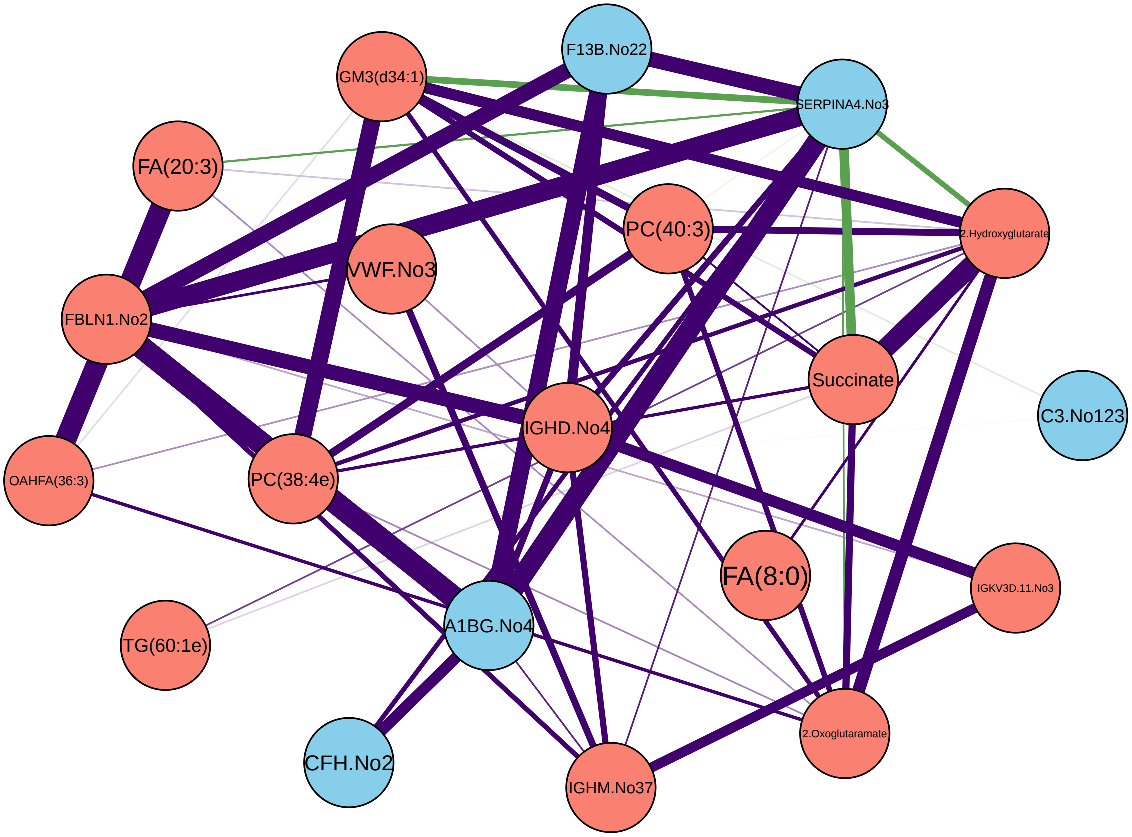
F13B.No22
GM3(d34:1)
SERPINA4.No3
FA(20:3)
PC(40:3)
2.Hydroxyglutarate
VWF.No3
FBLN1.No2
Succinate
C3.No123
IGHD.No4
OAHFA(36:3)
PC(38:4e)
FA(8:0)
IGKV3D.11.No3
TG(60:1e)
A1BG.No4
CFH.No2
2.Oxoglutaramate
IGHM.No37

Z441

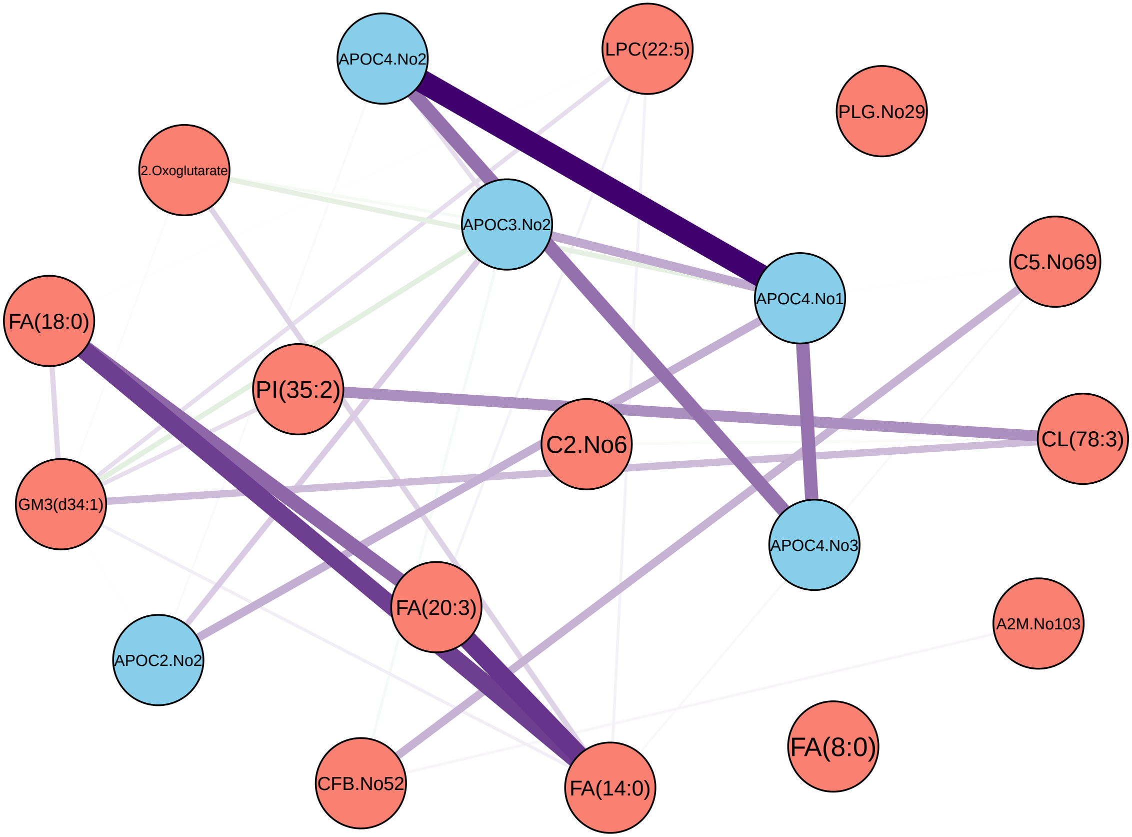
APOC4.No2
LPC(22:5)
PLG.No29
2.Oxoglutarate
APOC3.No2
APOC4.No1
C5.No69
FA(18:0)
PI(35:2)
C2.No6
CL(78:3)
GM3(d34:1)
APOC4.No3
FA(20:3)
A2M.No103
APOC2.No2
FA(8:0)
CFB.No52
FA(14:0)

Z459

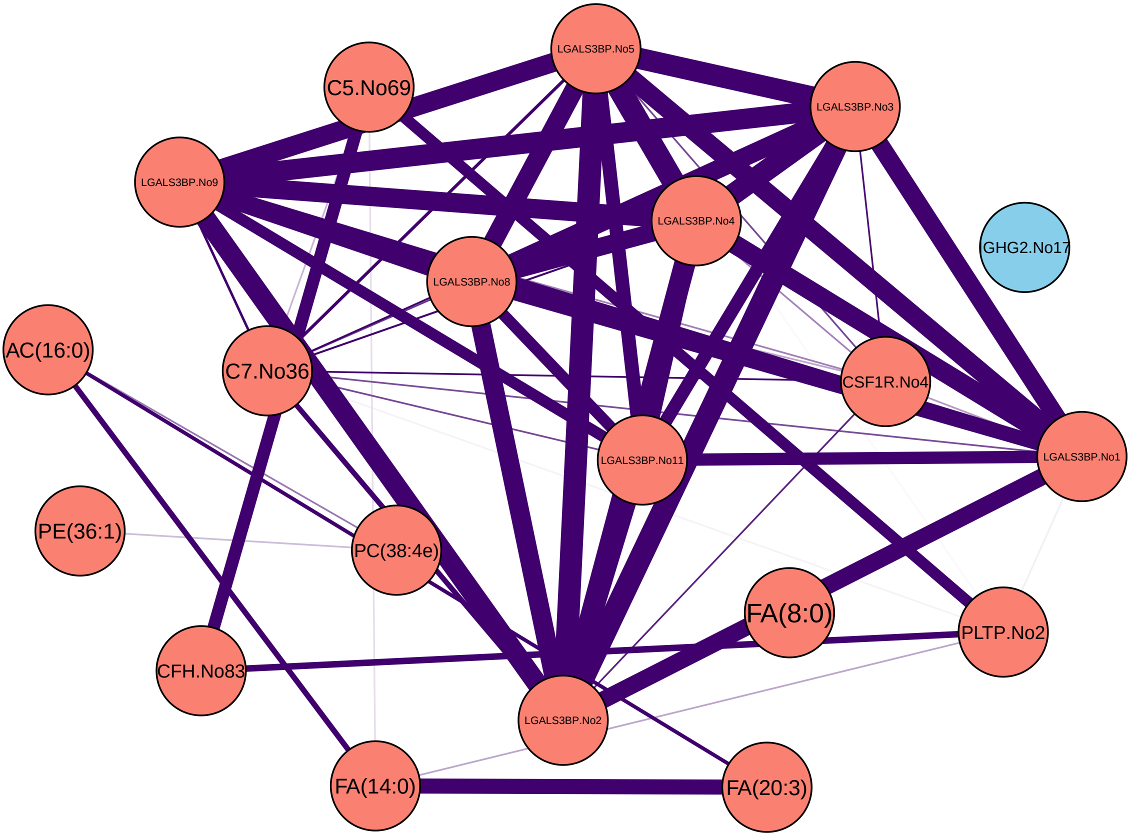
LGALS3BP.No5
C5.No69
LGALS3BP.No3
LGALS3BP.No9
LGALS3BP.No4
GHG2.No17
LGALS3BP.No8
AC(16:0)
C7.No36
CSF1R.No4
LGALS3BP.No11
LGALS3BP.No1
PE(36:1)
PC(38:4e)
FA(8:0)
PLTP.No2
CFH.No83
LGALS3BP.No2
FA(14:0)
FA(20:3)

Z472

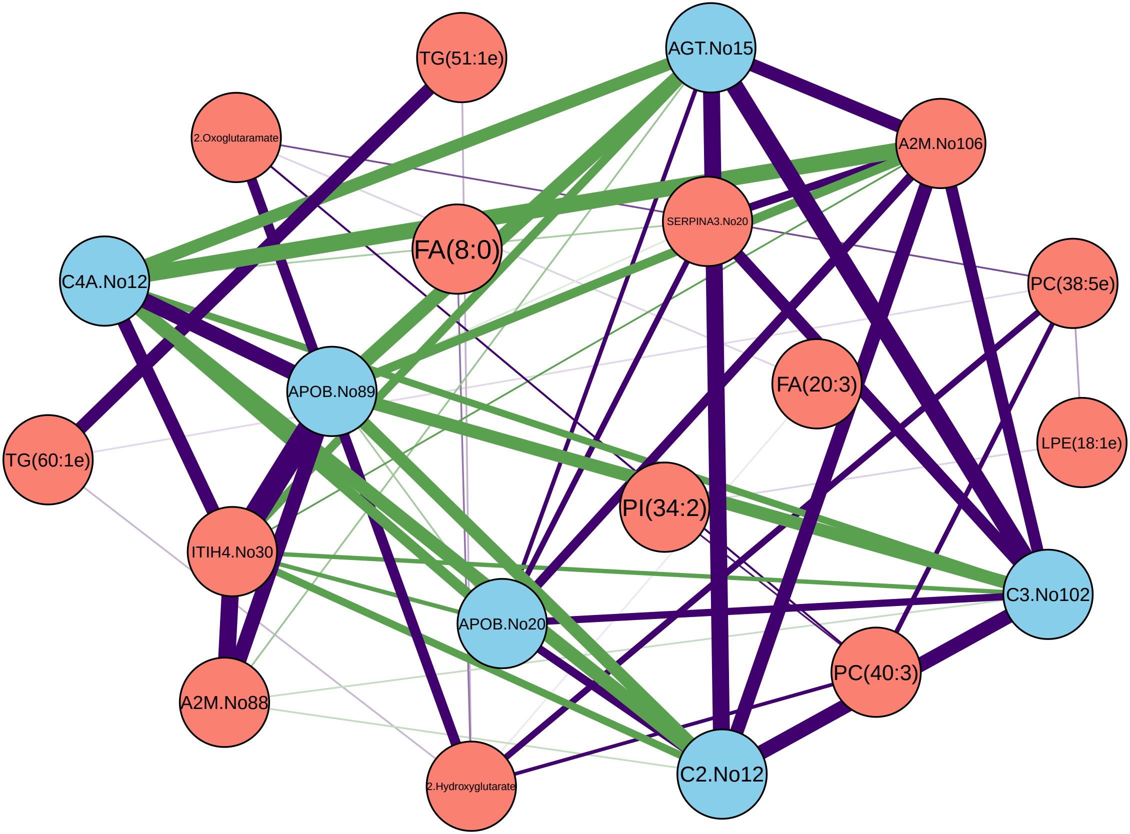
TG(51:1e)
AGT.No15
2.Oxoglutaramate
A2M.No106
SERPINA3.No20
FA(8:0)
C4A.No12
PC(38:5e)
FA(20:3)
APOB.No89
LPE(18:1e)
TG(60:1e)
PI(34:2)
ITIH4.No30
C3.No102
APOB.No20
PC(40:3)
A2M.No88
C2.No12
2.Hydroxyglutarate

Z499

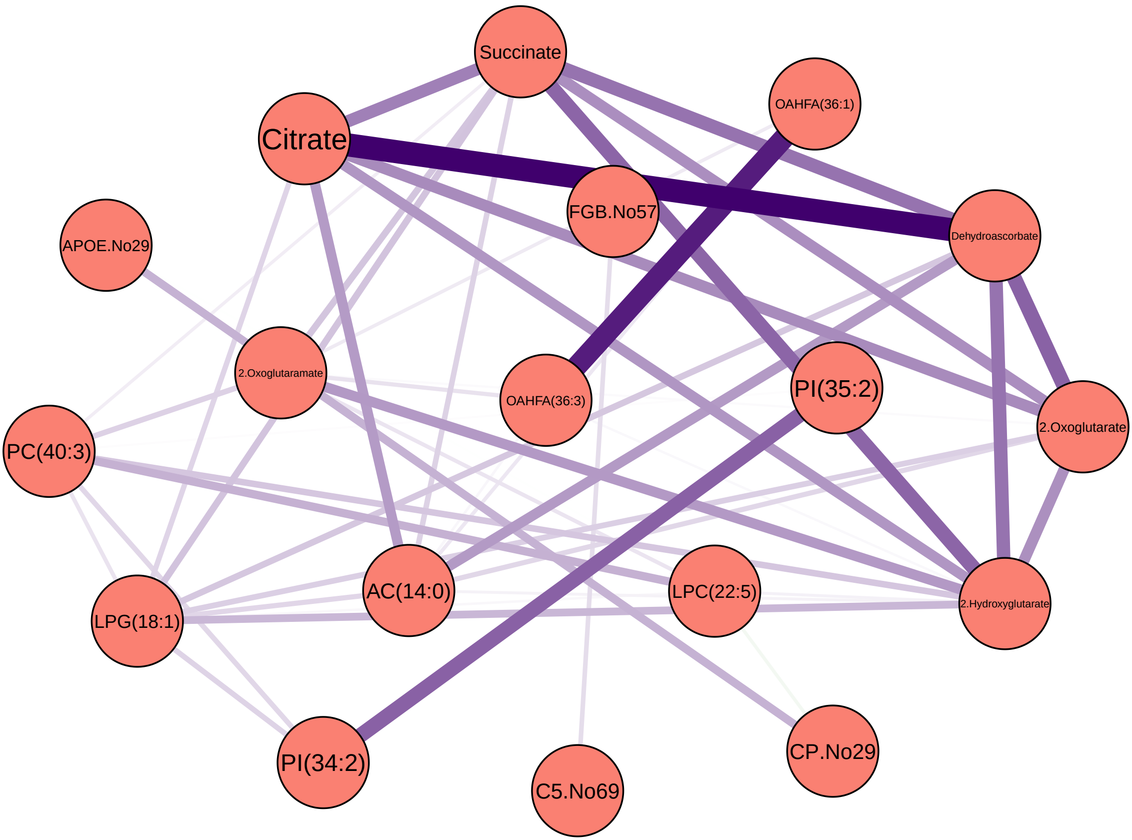

Succinate
OAHFA(36:1)
Citrate
FGB.No57
Dehydroascorbate
APOE.No29
2.Oxoglutaramate
OAHFA(36:3)
PI(35:2)
2.Oxoglutarate
PC(40:3)
AC(14:0)
LPC(22:5)
2.Hydroxyglutarate
LPG(18:1)
PI(34:2)
C5.No69
CP.No29

Z509

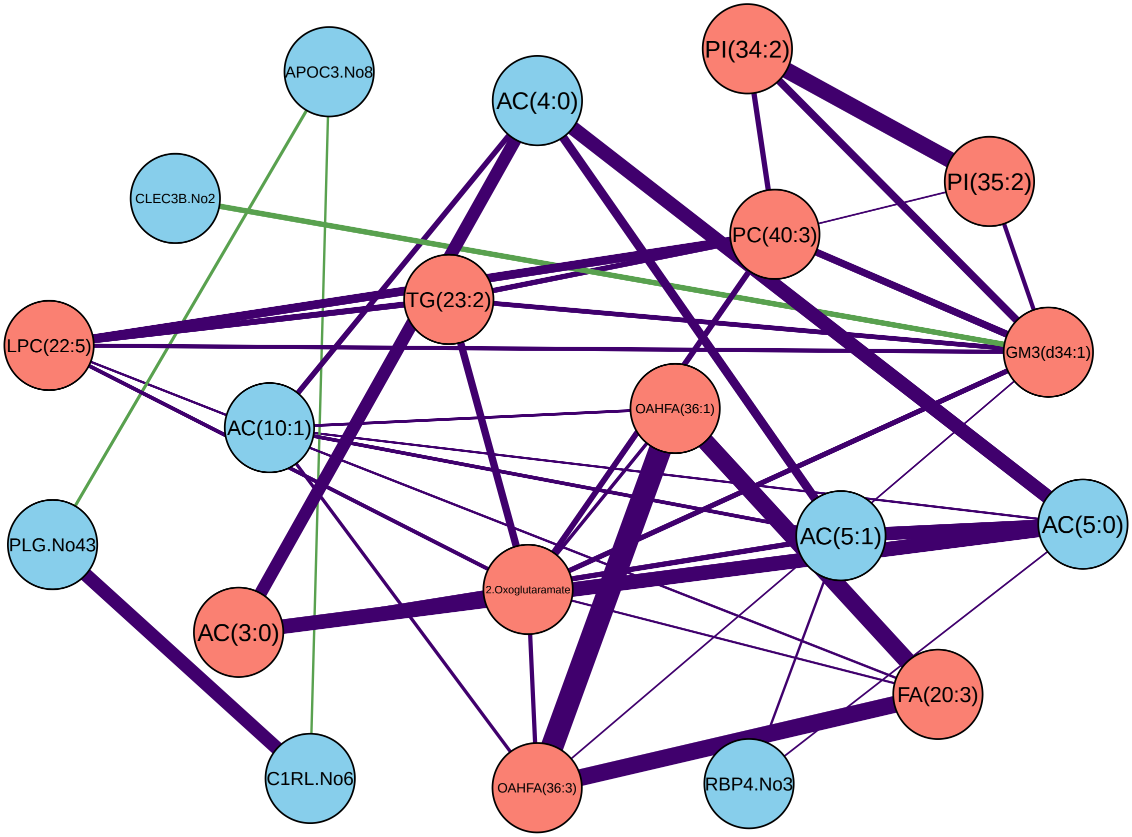

APOC3.No8
AC(4:0)
PI(34:2)
CLEC3B.No2
PI(35:2)
PC(40:3)
TG(23:2)
LPC(22:5)
GM3(d34:1)
OAHFA(36:1)
AC(10:1)
PLG.No43
AC(5:0)
AC(5:1)
2.Oxoglutaramate
AC(3:0)
FA(20:3)
C1RL.No6
OAHFA(36:3)
RBP4.No3

Z510

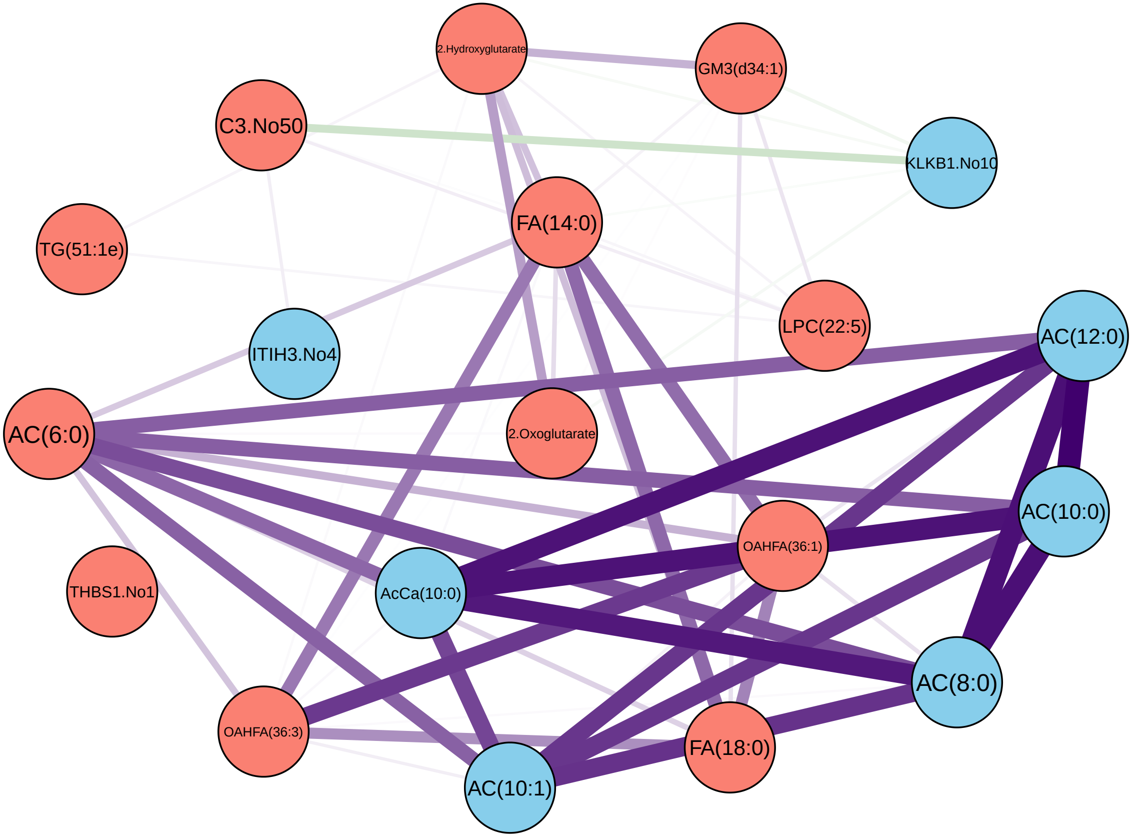

2.Hydroxyglutarate
GM3(d34:1)
C3.No50
KLKB1.No10
TG(51:1e)
FA(14:0)
LPC(22:5)
AC(12:0)
ITIH3.No4
AC(6:0)
2.Oxoglutarate
AC(10:0)
OAHFA(36:1)
THBS1.No1
AcCa(10:0)
AC(8:0)
OAHFA(36:3)
FA(18:0)
AC(10:1)

Z514

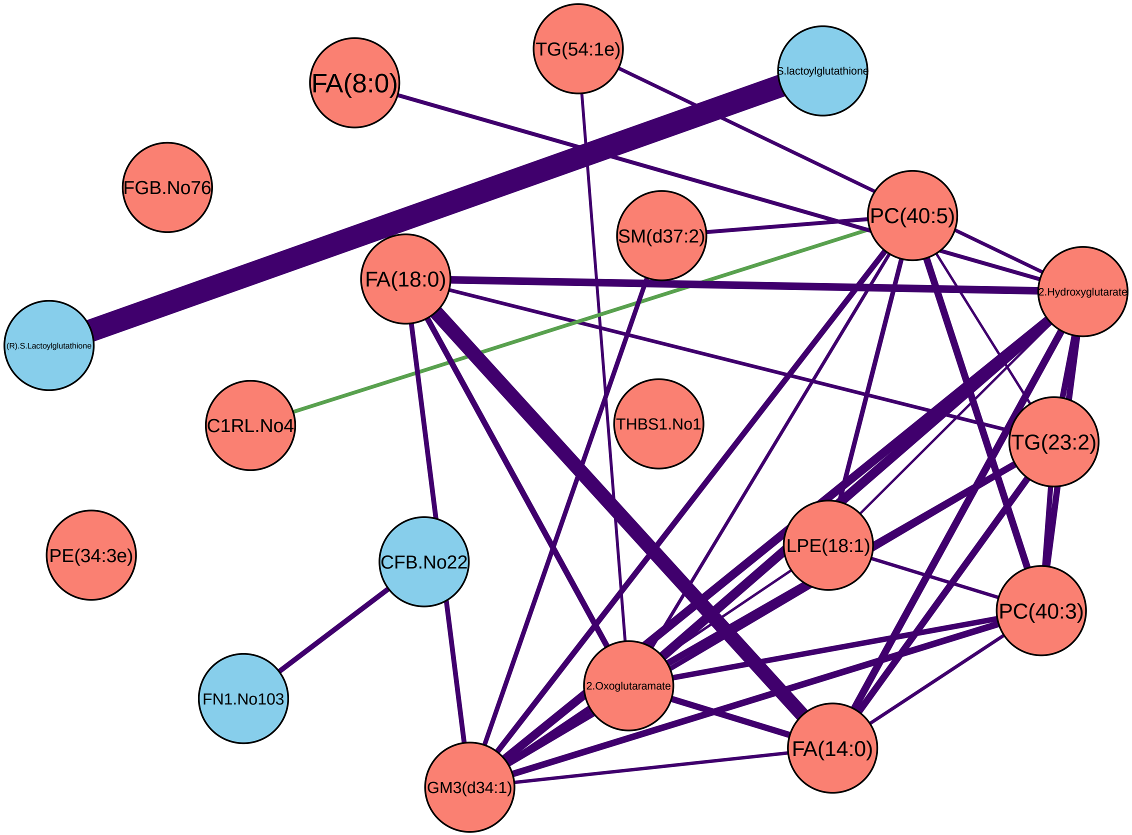

TG(54:1e)
S.lactoylglutathione
FA(8:0)
FGB.No76
SM(d37:2)
PC(40:5)
FA(18:0)
2.Hydroxyglutarate
(R).S.Lactoylglutathione
C1RL.No4
THBS1.No1
TG(23:2)
PE(34:3e)
CFB.No22
LPE(18:1)
PC(40:3)
FN1.No103
2.Oxoglutaramate
FA(14:0)
GM3(d34:1)

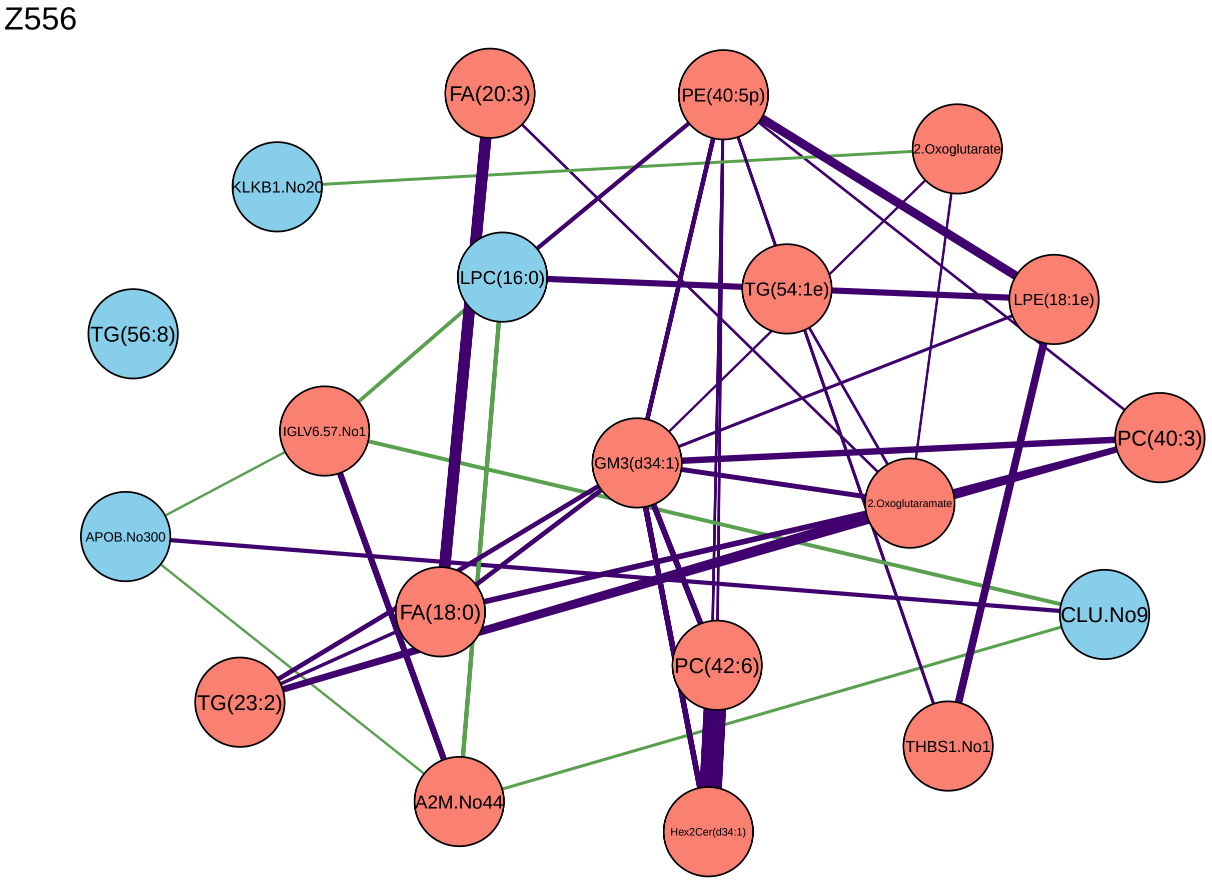
Z556
FA(20:3)
PE(40:5p)
2.Oxoglutarate
KLKB1.No20
LPC(16:0)
TG(54:1e)
LPE(18:1e)
TG(56:8)
IGLV6.57.No1
GM3(d34:1)
PC(40:3)
2.Oxoglutaramate
APOB.No300
FA(18:0)
CLU.No9
PC(42:6)
TG(23:2)
THBS1.No1
A2M.No44
Hex2Cer(d34:1)

Z559

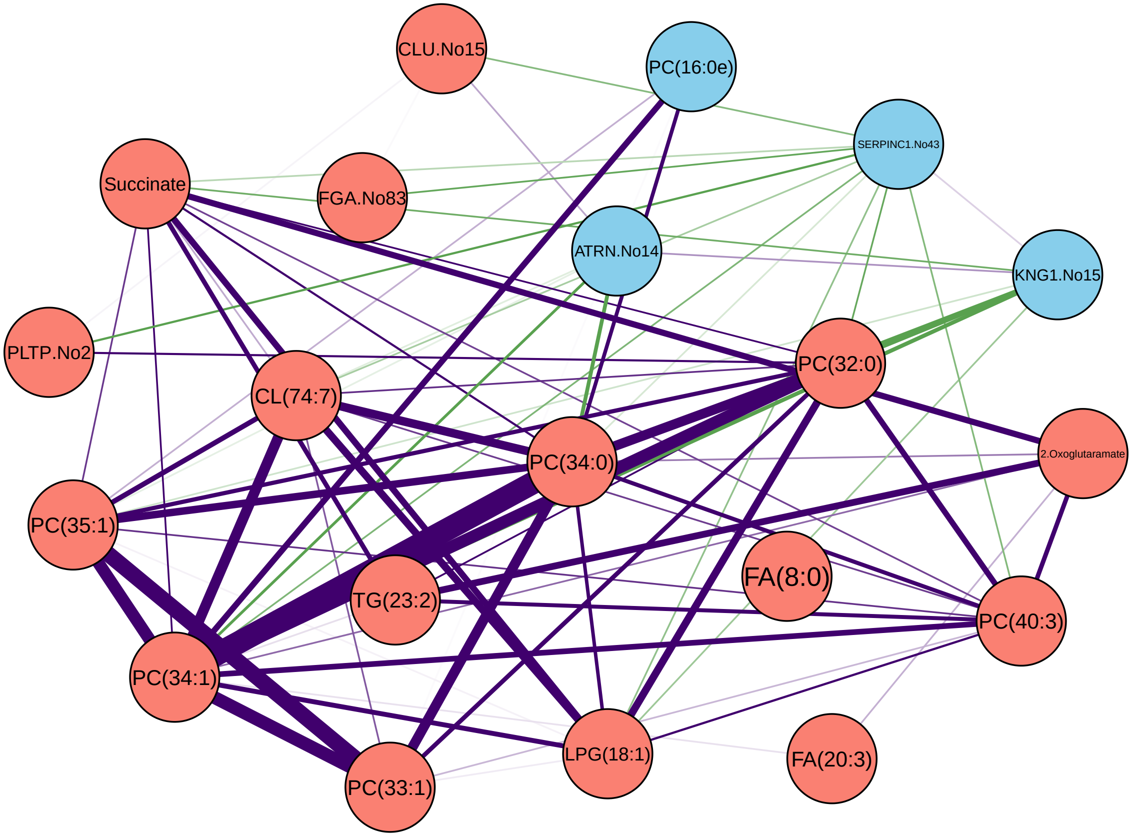

CLU.No15
PC(16:0e)
SERPINC1.No43
Succinate
FGA.No83
ATRN.No14
KNG1.No15
PLTP.No2
PC(32:0)
CL(74:7)
PC(34:0)
2.Oxoglutaramate
PC(35:1)
FA(8:0)
TG(23:2)
PC(40:3)
PC(34:1)
LPG(18:1)
FA(20:3)
PC(33:1)

Z581

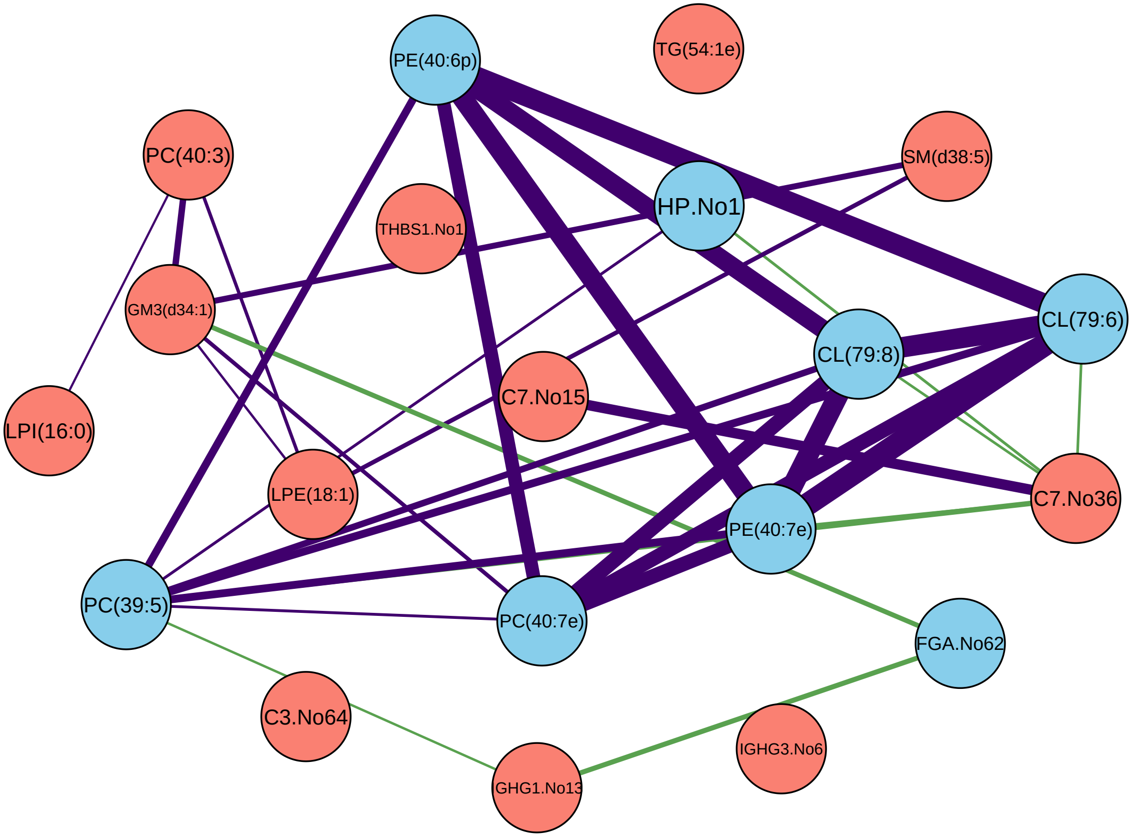
PE(40:6p)
TG(54:1e)
PC(40:3)
SM(d38:5)
THBS1.No1
HP.No1
GM3(d34:1)
CL(79:6)
CL(79:8)
LPI(16:0)
C7.No15
C7.No36
LPE(18:1)
PE(40:7e)
PC(39:5)
PC(40:7e)
FGA.No62
C3.No64
IGHG3.No6
GHG1.No13

Z582

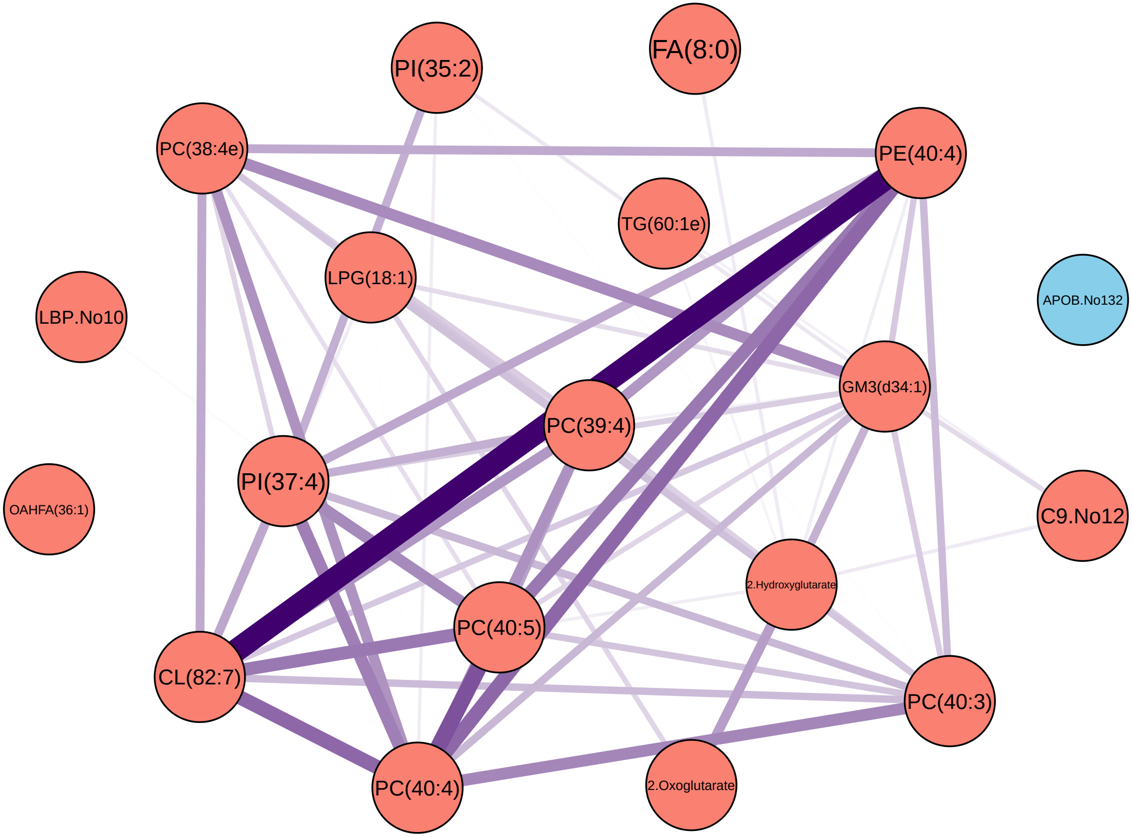
PI(35:2)
FA(8:0)
PC(38:4e)
PE(40:4)
TG(60:1e)
LPG(18:1)
APOB.No132
LBP.No10
GM3(d34:1)
PC(39:4)
PI(37:4)
OAHFA(36:1)
C9.No12
2.Hydroxyglutarate
PC(40:5)
CL(82:7)
PC(40:3)
PC(40:4)
2.Oxoglutarate

Z584

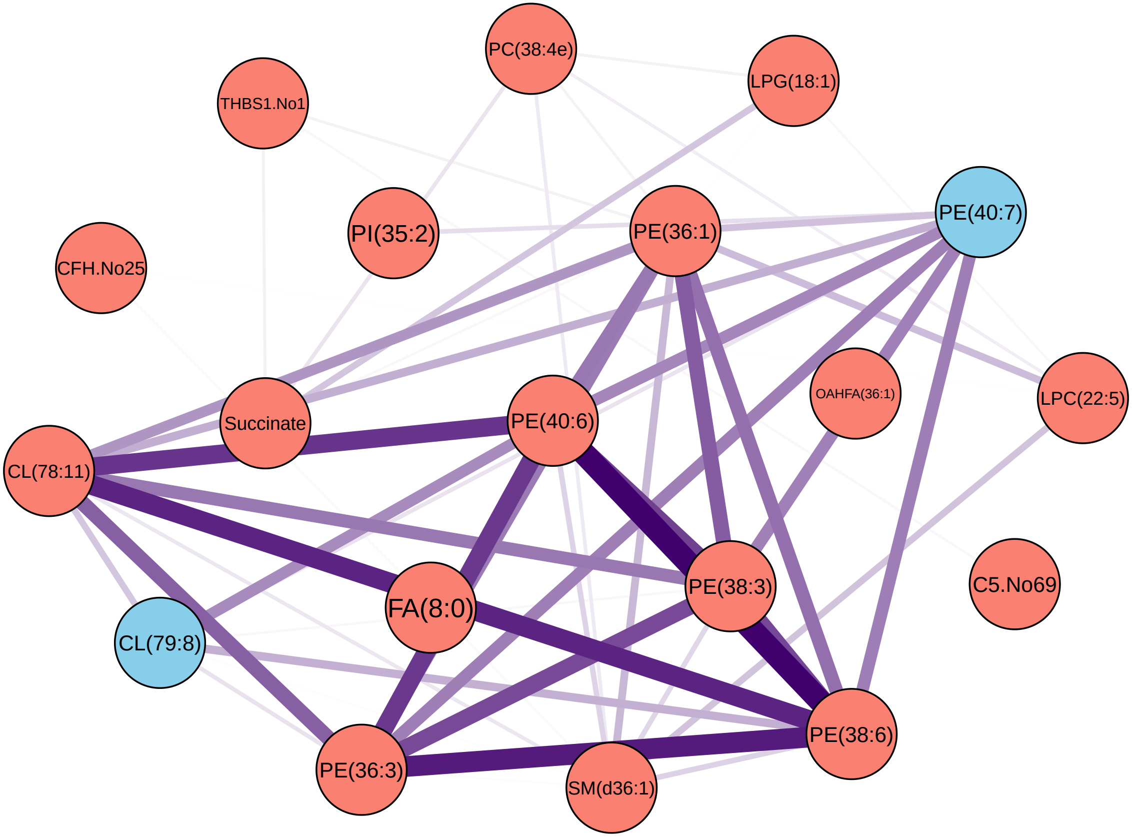
PC(38:4e)
LPG(18:1)
THBS1.No1
PI(35:2)
PE(36:1)
PE(40:7)
CFH.No25
OAHFA(36:1)
LPC(22:5)
Succinate
PE(40:6)
CL(78:11)
PE(38:3)
C5.No69
FA(8:0)
CL(79:8)
PE(38:6)
PE(36:3)
SM(d36:1)

Z586

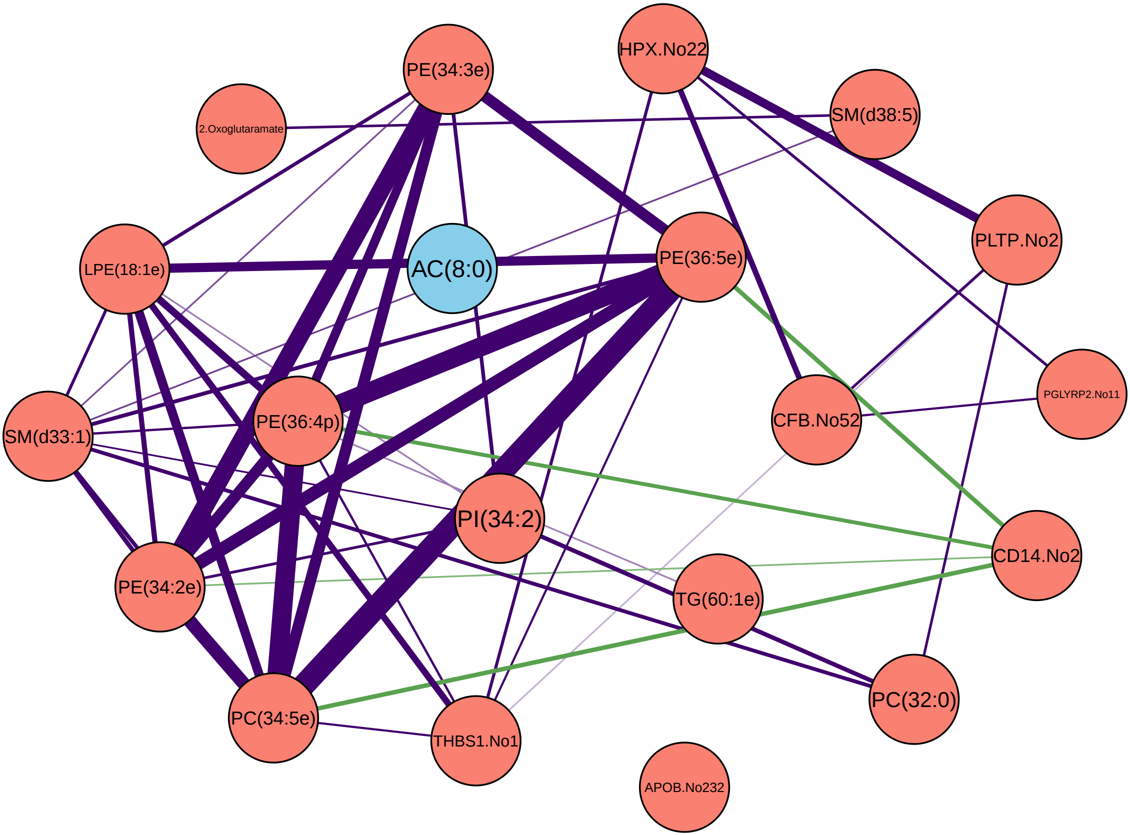

PE(34:3e)
HPX.No22
2.Oxoglutaramate
SM(d38:5)
LPE(18:1e)
AC(8:0)
PE(36:5e)
PLTP.No2
SM(d33:1)
PE(36:4p)
CFB.No52
PGLYRP2.No11
PI(34:2)
CD14.No2
PE(34:2e)
TG(60:1e)
PC(34:5e)
THBS1.No1
PC(32:0)
APOB.No232

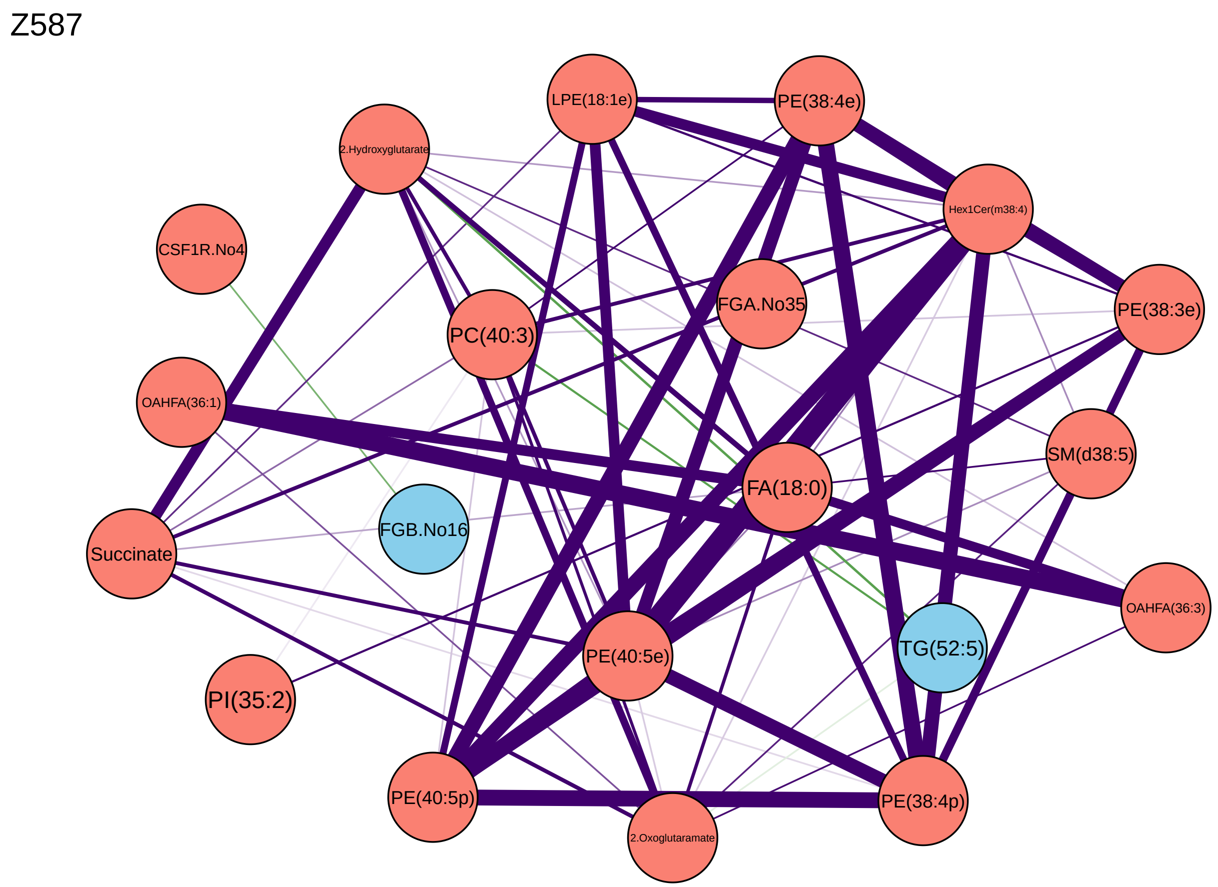
Z587
LPE(18:1e)
PE(38:4e)
2.Hydroxyglutarate
Hex1Cer(m38:4)
CSF1R.No4
FGA.No35
PE(38:3e)
PC(40:3)
OAHFA(36:1)
SM(d38:5)
FA(18:0)
FGB.No16
Succinate
OAHFA(36:3)
TG(52:5)
PE(40:5e)
PI(35:2)
PE(40:5p)
PE(38:4p)
2.Oxoglutaramate

Z589

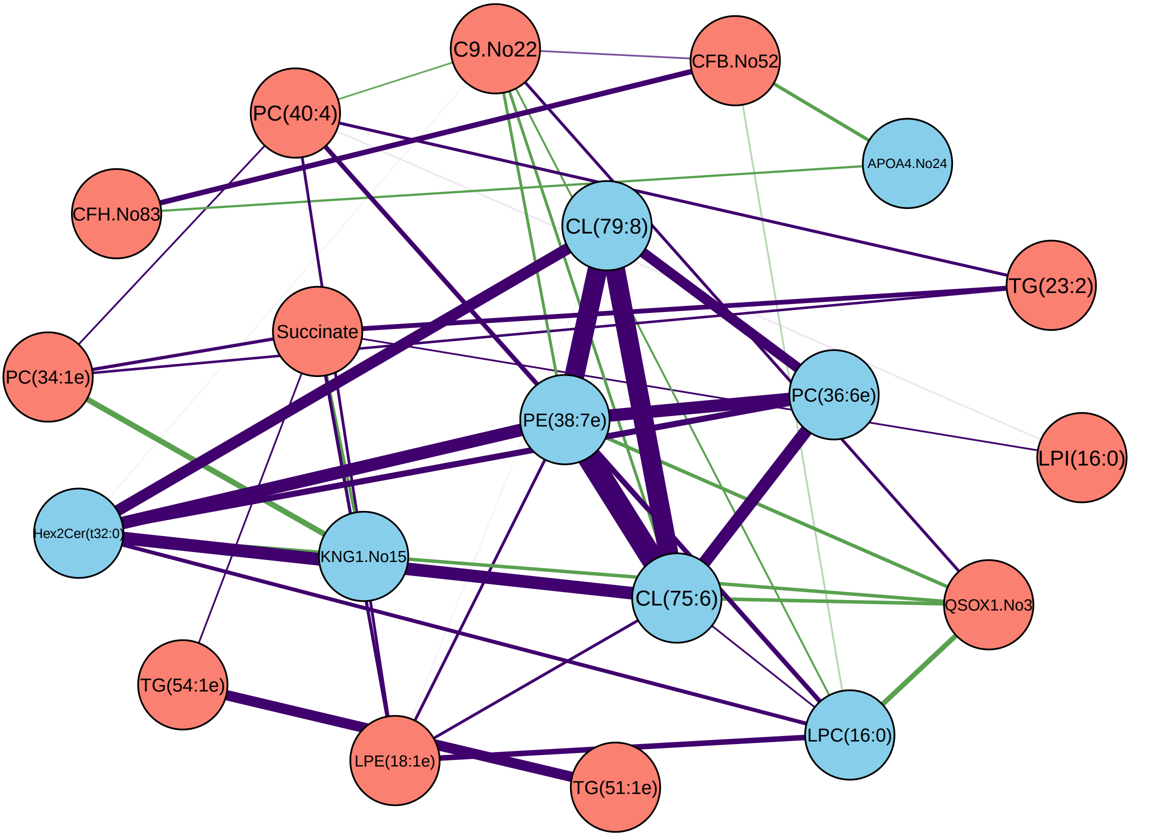
C9.No22
CFB.No52
PC(40:4)
APOA4.No24
CFH.No83
CL(79:8)
TG(23:2)
Succinate
PC(34:1e)
PC(36:6e)
PE(38:7e)
LPI(16:0)
Hex2Cer(t32:0)
KNG1.No15
CL(75:6)
QSOX1.No3
TG(54:1e)
LPC(16:0)
LPE(18:1e)
TG(51:1e)

Z593

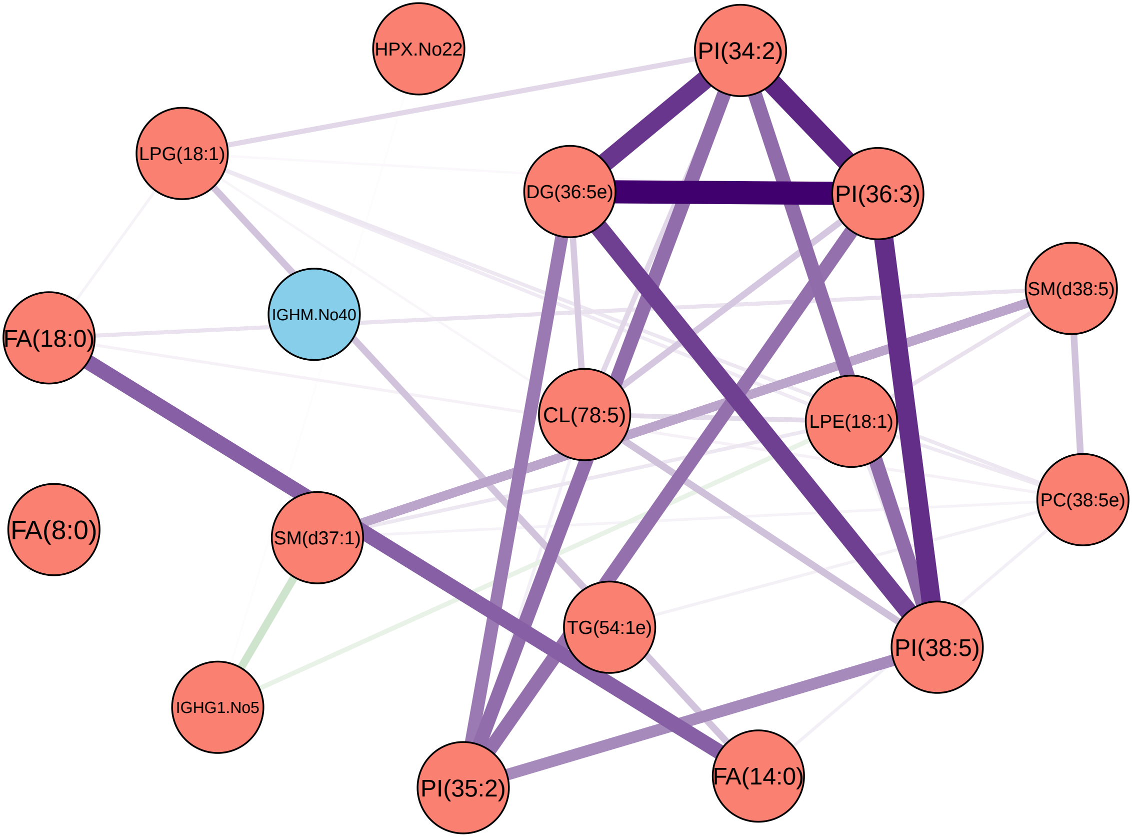
HPX.No22
PI(34:2)
LPG(18:1)
DG(36:5e)
PI(36:3)
SM(d38:5)
FA(18:0)
IGHM.No40
CL(78:5)
LPE(18:1)
PC(38:5e)
FA(8:0)
SM(d37:1)
TG(54:1e)
PI(38:5)
IGHG1.No5
PI(35:2)
FA(14:0)

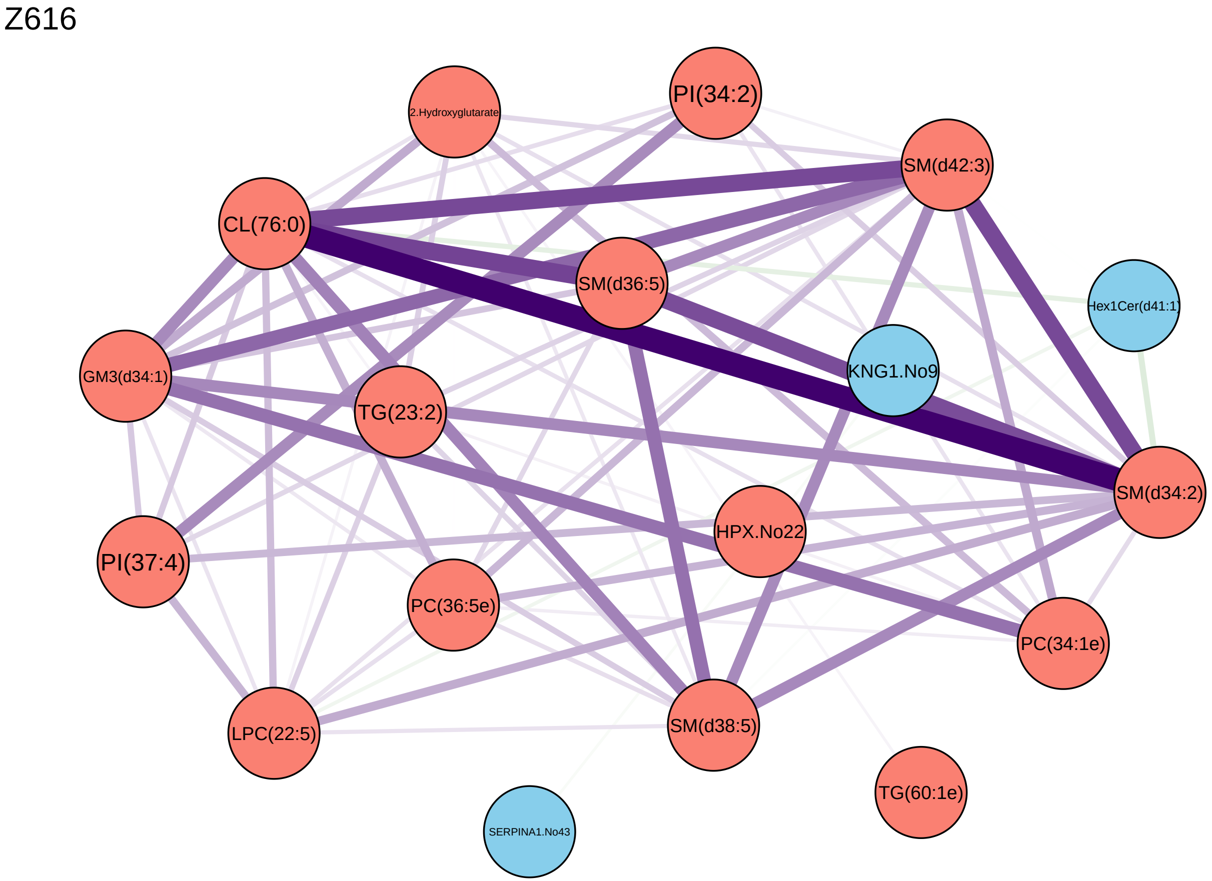
Z616
2.Hydroxyglutarate
PI(34:2)
SM(d42:3)
CL(76:0)
SM(d36:5)
Hex1Cer(d41:1)
GM3(d34:1)
KNG1.No9
TG(23:2)
SM(d34:2)
HPX.No22
PI(37:4)
PC(36:5e)
PC(34:1e)
LPC(22:5)
SM(d38:5)
TG(60:1e)
SERPINA1.No43

Z620

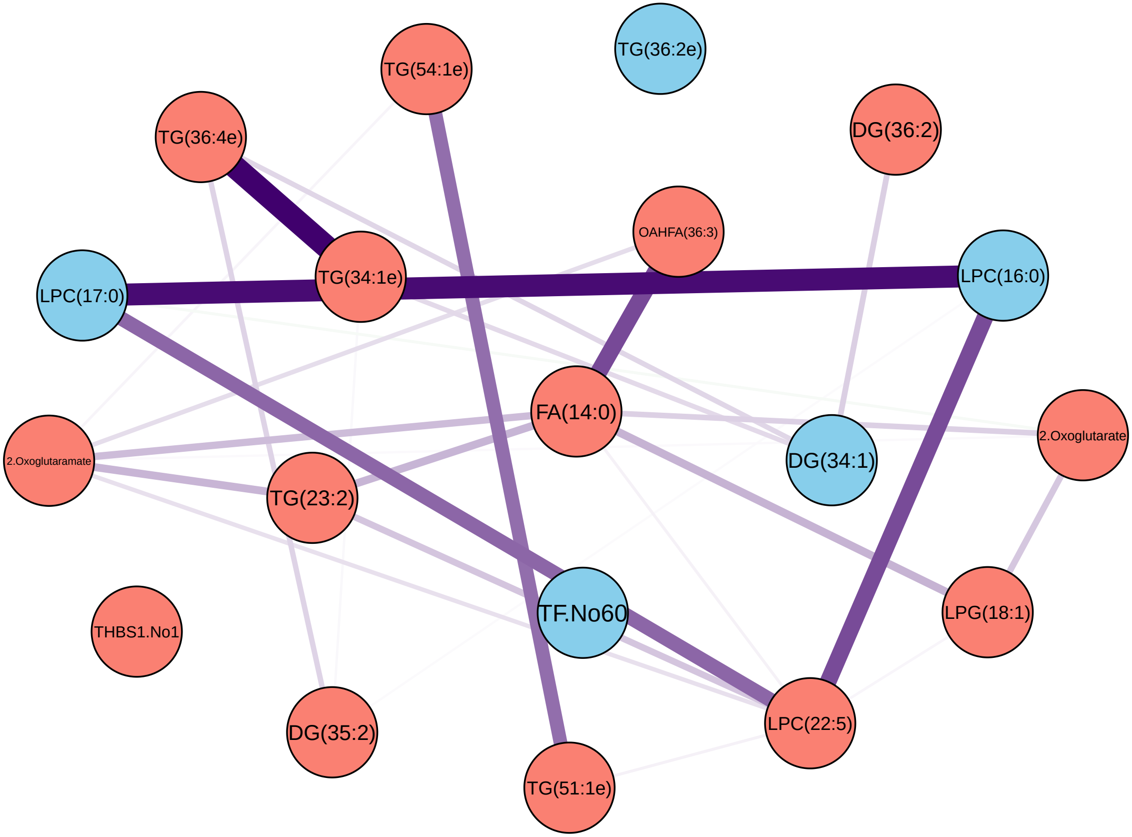
TG(54:1e)
TG(36:2e)
TG(36:4e)
DG(36:2)
OAHFA(36:3)
TG(34:1e)
LPC(17:0)
LPC(16:0)
FA(14:0)
2.Oxoglutaramate
2.Oxoglutarate
DG(34:1)
TG(23:2)
TF.No60
LPG(18:1)
THBS1.No1
DG(35:2)
LPC(22:5)
TG(51:1e)

Z627

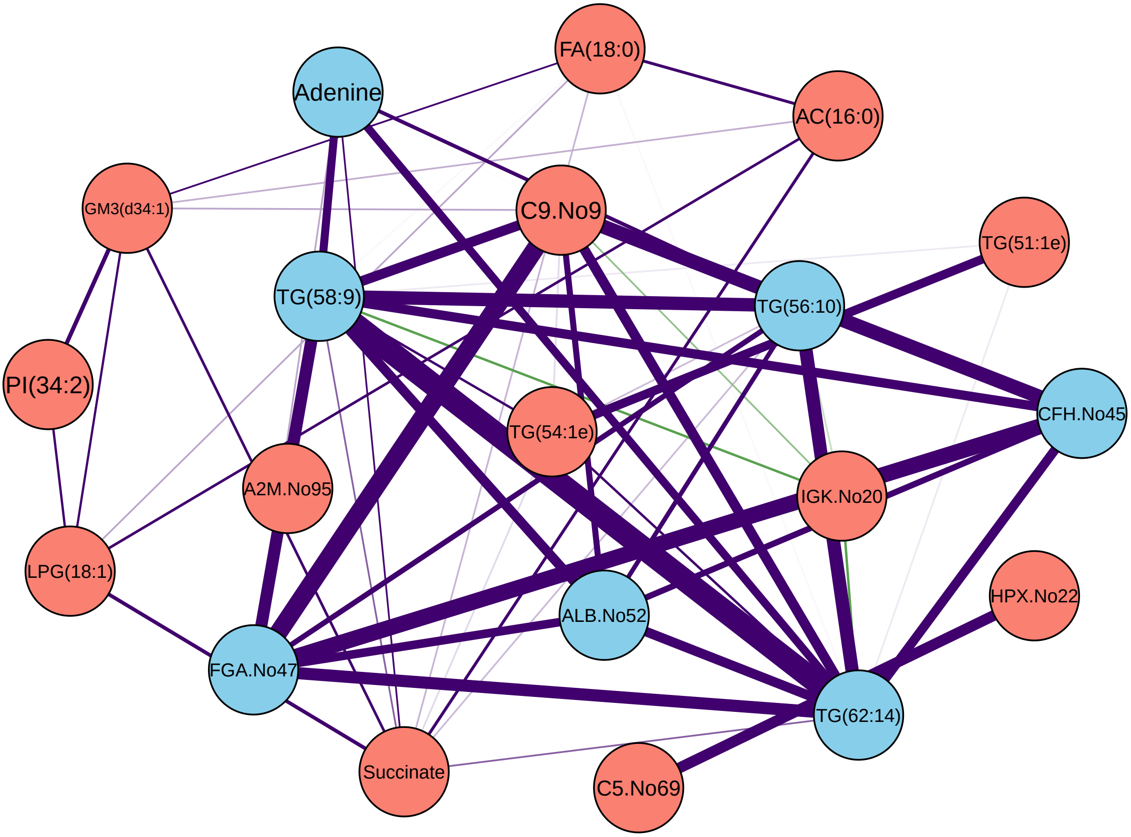
FA(18:0)
Adenine
AC(16:0)
GM3(d34:1)
C9.No9
TG(51:1e)
TG(58:9)
TG(56:10)
PI(34:2)
CFH.No45
TG(54:1e)
A2M.No95
IGK.No20
LPG(18:1)
HPX.No22
ALB.No52
FGA.No47
TG(62:14)
Succinate
C5.No69
